# Environment-wide association study (EWAS) on cardiometabolic traits: A systematic assessment of the association of lifestyle variables on a longitudinal setting

**DOI:** 10.1101/2021.03.22.21254099

**Authors:** Alaitz Poveda, Yan Chen, Hugo Pomares-Millan, Azra Kurbasic, Chirag J Patel, Frida Renström, Göran Hallmans, Ingegerd Johansson, Paul W. Franks

## Abstract

The present study aims to assess the over-time association of ∼300 lifestyle exposures with nine cardiometabolic traits with the ultimate aim of identifying exposures/exposure groups that could inform lifestyle interventions aiming at controlling cardiometabolic diseases. The analyses were undertaken in a longitudinal sample comprising >31000 adults living in northern Sweden. Linear mixed models were used to assess the average associations of lifestyle exposures and linear regression models were used to test association with 10-year change of the cardiometabolic traits. ‘Physical activity’ and ‘General Health’ were the exposure categories containing the highest number of ‘tentative signals’ in analyses assessing the average association of lifestyle variables, while ‘Tobacco use’ was the top-category for the 10-year change association analyses. Thirteen modifiable variables showed a consistent average association among the majority of cardiometabolic traits. These variables belonged to four main groups: i) Smoking, ii) Diet (secoisolariciresinol intake and brewed coffee), iii) Leisure time physical activity and iv) a group of variables more specific to the Swedish lifestyle (snuff status, hunting/fishing during leisure time and boiled coffee). Interestingly, sweet drinks, fish intake and salt content, all lifestyle exposures frequently mentioned in public health recommendations were not broadly associated with the analysed cardiometabolic traits.

---

The majority of non-communicable diseases are caused by the complex interplay of genetic and environmental factors. In the last decades, major progress has been made in discovering genetic loci predisposing for these diseases, facilitated by genome-wide association studies (GWAS). These studies allow high-troughput and systematic screening of millions of variants against quantitative traits or hard disease endpoints. Unlike population genetics, there are no standard environment ‘chips’ that capture multiple environment exposures simultaneously. Therefore, environmental epidemiology typically involves approaches where hypothesized associations between specific environmental exposures and disease traits are tested. These studies are limited by the expectations and knowledge about the hypothesized relationships they seek to test, which may cause bias and inhibit discovery (1).

Environment-wide association studies (EWAS) represent an approach through which multiple environmental factors can be systematically screened for their associations with disease traits in a manner that is to a large degree agnostic to prior knowledge about disease associations; in this sense the EWAS approach is similar to GWAS. EWAS was first described in the published literature in a 2010 paper reporting associations analyses between metabolites and type 2 diabetes (2). Later, EWAS was used to identify nutrients, environmental contaminants, and prescribed drugs (3-9) associated with disease and disease complications. Almost all published EWAS have used cross-sectional epidemiological data to assess exposures at a fixed time point without consideration of the impact of exposures throughout an individual’s lifetime. Longitudinal data analyses may help us understand the associations among exposures and changes in cardiometabolic traits over time.

The present study sought to assess the over-time association of more than 300 lifestyle exposures (e.g. food items, sleep habits, physical activity, psicosocial factors) with nine cardiometabolic traits (i.e. BMI, blood lipids, blood glucose, and blood pressure) and use these results to identify target lifestyle exposures/exposure groups that could inform lifestyle interventions focused on controlling cardiometabolic diseases.

## METHODS

### Participants

The analyses reported here were undertaken using data from the Västerbotten Intervention Programme (VIP) (10). VIP is a prospective, population-based cohort study originally designed as a long-term project intended for health promotion among the general population in Västerbotten county (approx. 254,000 inhabitants), northern Sweden. Since 1985, adults residing in Västerbotten have been invited to undergo a clinical examination and complete lifestyle questionnaires during the years of their 30^th^, 40^th^, 50^th^, and 60^th^ birthdays.

A sub-cohort of VIP (*n*=88,614) was used in the present analyses. Participants with non-Swedish origin (*n*=14,629) were excluded from the analyses as the different cultural and lifestyle habits and disease predisposition of non-Swedish participants may cause confounding by population stratification in EWAS analyses. Participants with diagnosed diabetes and cardiovascular diseases (*n*=3,025) were also excluded to minimize bias attributable to diagnostic labelling and medications. The final dataset comprised 31,362 partipants including 67,738 health examinations performed between 1990 and 2013. Written informed consent was obtained from all living participants as part of the VIP. The study was approved by the Regional Ethical Review Board in Umeå.

### Clinical measurements

Nine cardiometabolic traits were analysed in the study: body mass index (BMI), systolic and diastolic blood pressures (SBP and DBP, respectively), fasting and 2h glucose, total cholesterol, triglycerides, HDL cholesterol and LDL cholesterol. Clinical measures in VIP are described in detail elsewhere (10). In brief, participants’ weight (in kg) and height (in cm) were measured using calibrated scale and stadiometer, with participants wearing light clothing and no shoes. BMI was calculated as body weight in kilograms divided by height in meters squared. SBP and DBP were measured once, after 5-min rest, with the participant in a recumbent position using either manual or automated sphygmomanometers. Capillary blood was drawn after overnight fasting and a second blood sample was drawn two hours after the administration of a 75-gram oral glucose load. Blood glucose, total cholesterol and triacylglycerol levels were then measured using a Reflotron bench-top analyzer (Roche Diagnostics Scandinavia AB). HDL cholesterol was measured in a subgroup of participants and LDL cholesterol was estimated using the Friedewald formula (11). The measurement for lipids and blood pressure changed in September 2009. From this date onwards, blood pressure was measured twice in a sitting position and averaged, and total cholesterol and triglyceride levels were analysed using clinical chemical analysis in the laboratory. Thus, validated conversion equations were used to align the lipid and blood pressure measurements taken before and after September 2009 (12). For participants on lipid and/or blood pressure lowering medications, lipid and/or blood pressure levels were corrected by adding published constants (+0.208□mmol/l for triglycerides, +1.347□mmol/l for total cholesterol, −0.060□mmol/l for HDL cholesterol, +1.290□mmol/l for LDL cholesterol, +15 mmHg for SBP and +10 mmHg for DBP) (13, 14). Values of cardiometabolic traits located outside the normal range suggested by VIP data managers (see Suplementary Material) were considered outliers and excluded.

### Lifestyle assessments

Participants were asked to complete a self-administered questionnaire during each visit that included questions about socio-economic factors, physical/mental health, quality of life, social network and support, working conditions, and alcohol/tobacco consumption. Physical activity was assessed through a modified version of the International Physical Activity Questionnaire (15, 16). A validated semi-quantitative food frequency questionnaire (FFQ) designed to capture habitual diet over the last year was used to capture information on various dietary factors (17). Up to the mid-1990s, the FFQ consisted of 84 different foods items/groups, but it was reduced to 66 items in 1996 by combining similar line items and by removing items that provided minimal unique information. For the current analysis, matching food items from different FFQ versions were combined in new variables and all analyses including dietary variables were adjusted for FFQ version. In the FFQ, participants indicated how often they consumed foods and beverages on a nine-point frequency scale. Information on average portion size of meat and fish, vegetables, potatoes, rice and pasta was also gathered. Nutrient and energy content were calculated based on the Swedish Food Composition Database (18) based on meal frequency and portion size. Food intake level (FIL) was calculated as total energy intake divided by estimated basal metabolic rate. Participants with more than 10% FFQ data missing, one or more portion indication missing, or a seemingly implausible total energy intake (the top 2.5% and bottom 5% of FIL in the original VIP dataset) were excluded from the analyses. Implausible values for other lifestyle variables (see Supplementary Material) were also removed from the analyses. Lifestyle variables were grouped in 10 different categories to facilitate understanding of the results: i) alcohol consumption; ii) non-alcoholic beverage consumption; iii) food; iv) nutrients; v) general health; vi) physical activity & fitness; vii) psychosocial; viii) sleep; ix) social conditions; x) tobacco use.

### Statistical analysis

To internally validate our findings, the final dataset was randomly divided into a training (*n*=15,823) and a testing (*n*=15,539) set. The training set was used to scan for lifestyle variables associated with any of the nine cardiometabolic traits and the testing set to validate any associations discovered in the training set (Figure 1).

**Fig 1.**
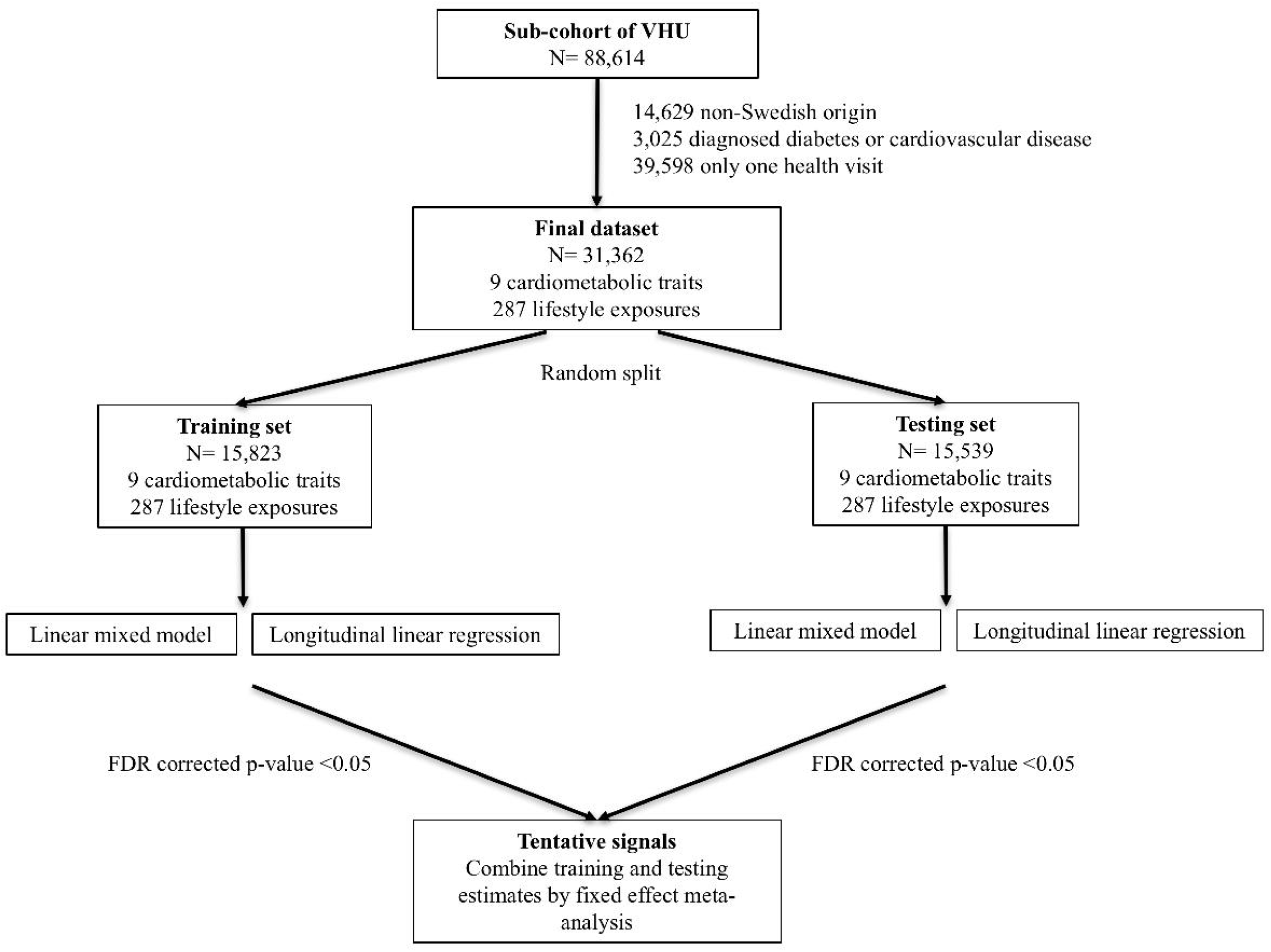
Flow chart of the method followed in the study

Lifestyle variables were treated either as continuous or as categorical variables; thus, ordinal variables were treated as continuous variables. For categorical variables with more than two levels dummy variables were created and dichotomized. All numeric lifestyle variables were z-score standardized in order to compare the association effects of the different lifestyle variables. Similarly, for categorical variables, levels were harmonized from low to high, using the lowest one as reference. Thirty-eight categorical variables that had 90% of the observations belonging to one category were excluded from the analyses. In total, the analyses included 242 numeric and 45 categorical lifestyle variables. Dietary variables were regressed on total energy intake and their residuals along with total energy intake were included in the analyses of these variables to account for potential confounding by total energy intake (19). Models with glycaemic or lipid traits as the dependent variables were additionally adjusted for fasting status. All models (except models having BMI as outcome) were adjusted for BMI.

#### Average lifestyle associations

Linear mixed models were used to estimate an average linear effect of the lifestyle exposures on the cardiometabolic traits. The models were adjusted for age, age^2^, sex, follow-up time, FFQ version (where appropriate), total energy intake (TEI; where appropriate), BMI (where appropriate) and fasting status (where appropriate).

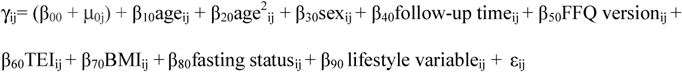

where γ_ij_ represents a cardiometabolic trait value at visit i for participant j, β00 is the fixed intercept, μ_0j_ represents different random intercepts for each participant, the rest of the β estimates are the estimated fixed effect size parameters for each corresponding variable, and ε represents error.

#### Long-term lifestyle associations

Linear regression models were used to test if the lifestyle variables were associated with 10-year changes in the cardiometabolic traits:

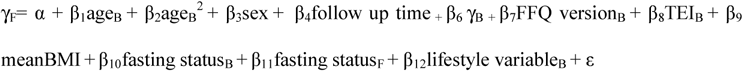

where γ_F_ represents the value of the cardiometabolic trait at follow-up and γ_B_ the value at baseline, α is the intercept, β_i_ represent the estimated effect size parameter for each corresponding variable. Age_B,_ FFQ version_B,_ TEI_B,_ fasting status_B_ and lifestyle variable_B_ are the age, FFQ version, TEI, fasting status and lifestyle variable values at baseline; fasting status_F_ is the fasting status value at follow up; meanBMI is the average BMI of the baseline and followup BMI values, and ε represents error.

#### Tentative signals

The Benjamini and Hochberg (20) False Discovery Rate (FDR) was used to correct for multiple testing. Associations of lifestyle variables were considered “tentative signals” if they achieved significance at *P*_*FDR*_ <0.05 after multiple testing correction in both training and testing sets, and showed the same direction of association (either positive or negative) in both sets. Estimates obtained in the training and testing sets were meta-analyzed using a fixed effect model to obtain overall estimates (i.e. effect size, standard error and *P*). Overall estimates were used in the description of the results.

#### Correlation patterns

Correlations between ‘tentative signals’ on the linear mixed and/or longitudinal linear regression analyses were calculated and visualized using a heatmap. A hierarchical clustering algorithm was used to arrange lifestyle variables, so that the pair of variables with higher correlations appear closer in the heatmap.

*Prioritization of modifiable lifestyle variables*

‘Tentative signals’ for each of the cardiometabolic traits were gathered and prioritized to identify target lifestyle exposures and exposure groups in which lifestyle interventions aiming at controlling cardiometabolic diseases may focus. First, variance explained for each lifestyle variable (and covariates) was estimated and variables were rank-ordered within each lifestyle category for each of the nine outcome traits. In the linear mixed models, marginal (fixed terms) variance explained was used. The top-ranked variables (five per category per trait) were identified and the top-ranked variables represented in the majority of the cardiometabolic traits (at least five traits) were prioritized. Target groups were evaluated using a hierarchical clustering algorithm based on correlations between the prioritized variables and visualized in a heatmap. Non-modifiable variables were excluded from the prioritization and clustering step as these variables could not be affected by a lifestyle intervention.

Statistical analyses were performed using *R* software versions 3.5.2 and 3.6.1 (21) (see Supplementary Material for the specific packages used for analyses).

## RESULTS

Descriptive characteristics of the training and testing sets of the study population are summarized in Table 1 and S1 and S2. Mean age of participants was 47.7 years in both sets with 50.8% of women in the training set and 50.5% in the testing set.

**Table 1.** Summary of participant characteristics in training (*n*_*max*_= 15 823) and testing (*n*_*max*_= 15 539) sets

| Variables | Training set |  | Testing set |  |
| --- | --- | --- | --- | --- |
|  | Number of observations | Mean (SD) | Number of observations | Mean (SD) |
| Age (years) | 34 234 | 47.71±8.91 | 33504 | 47.72±8.92 |
| Height (cm) | 34 103 | 172.1±9.2 | 33373 | 172.0±9.2 |
| Weight (kg) | 34 084 | 76.42±14.59 | 33358 | 76.41±14.46 |
| BMI (kg/m<sup>2</sup>) | 34 068 | 25.72±4.00 | 33345 | 25.74±3.98 |
| Total cholesterol (mmol/l) | 33 968 | 5.51±1.09 | 33213 | 5.51±1.09 |
| Triglycerides (mmol/l) | 29 735 | 1.39±0.75 | 29170 | 1.40±0.81 |
| HDL-C (mmol/l) | 13 977 | 1.40±0.47 | 13826 | 1.39±0.47 |
| LDL-C (mmol/l) | 13 908 | 3.86±1.03 | 13741 | 3.86±1.03 |
| Fasting glucose (mmol/l) | 34 043 | 5.39±0.75 | 33296 | 5.39±0.75 |
| 2-h glucose (mmol/l) | 32 826 | 6.56±1.50 | 32125 | 6.57±1.50 |
| SBP (mmHg) | 33 950 | 126.2±17.4 | 33243 | 126.7±17.6 |
| DBP (mmHg) | 33 929 | 78.77±11.28 | 33231 | 78.89±11.31 |

BMI: Body mass index; HDL-C: High-density lipoprotein cholesterol; LDL-C: Low-density lipoprotein cholesterol; SBP: Systolic blood pressure; DBP: Diastolic blood pressure; SD: Standard deviation.

### Average lifestyle associations

167 out of 287 lifestyle variables were considered tentative signals for BMI (S3), 49 for SBP (S4), 37 for DBP (S5), 87 for total cholesterol (S6), 108 for triglycerides (S7), 69 for HDL cholesterol (S8), 21 for LDL cholesterol (S9), 43 for fasting glucose (S10) and 58 for 2h glucose (S11). ‘Physical activity’ and ‘General health’ were the top-categories for BMI (Figure 2) and ‘General health’ for blood pressure traits (Figure 3). Regarding lipids, ‘Beverage’, ‘Nutrients’ and ‘Physical activity’ were the categories with the highest number of ‘tentative signals’ for total and LDL cholesterol (Figures 4A and 4D), while ‘Physical activity’, ‘Tobacco use’ and ‘General health’ were the top-categories for triglycerides (Figure 4B), and ‘Alcohol’ for HDL cholesterol (Figure 4C). For glucose traits, ‘Physical activity’, ‘General health’ and ‘Tobacco use’ were the top-categories (Figure 5).

**Fig 2.**
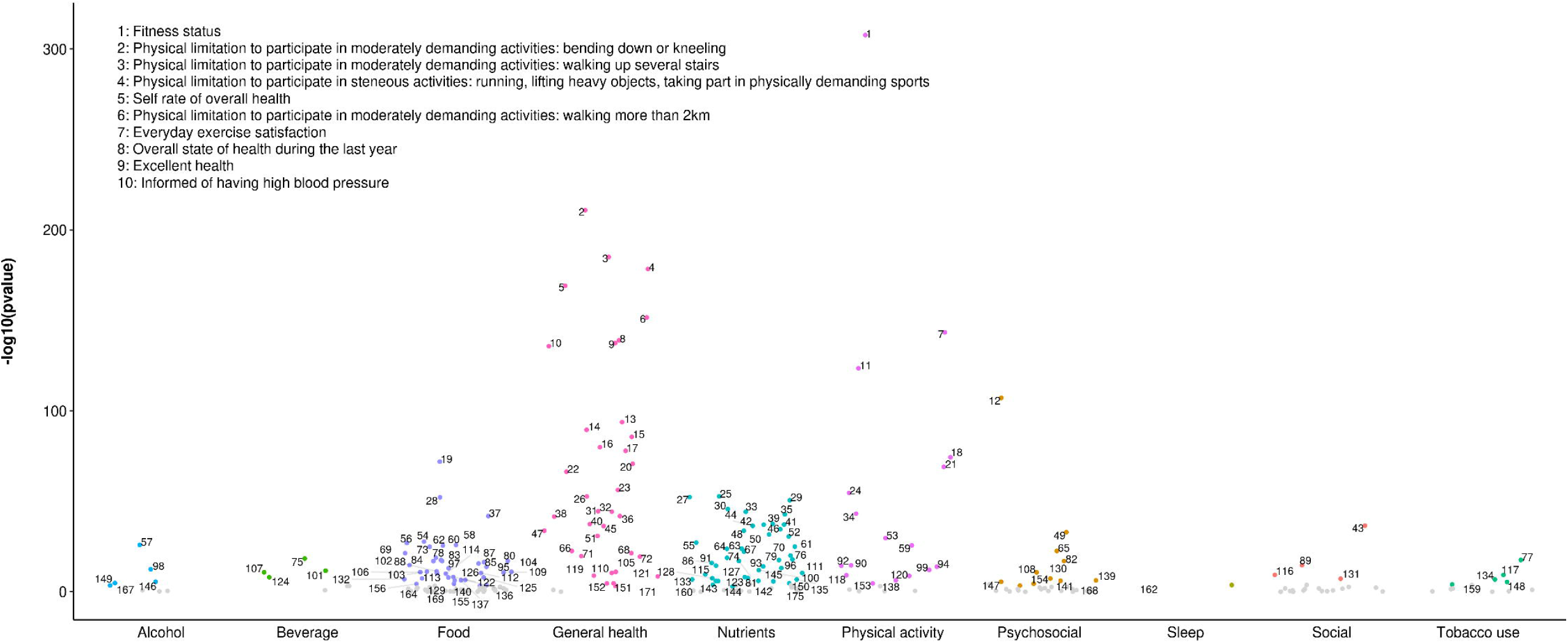
‘Manhattan plot’ representing the distribution of *P* values of the association of lifestyle variables and BMI by lifestyle category ‘Tentative signals’ are coloured and number labeled in the figure and the top 10 variables are spelled out. See S25 for references to the labels

**Fig 3.**
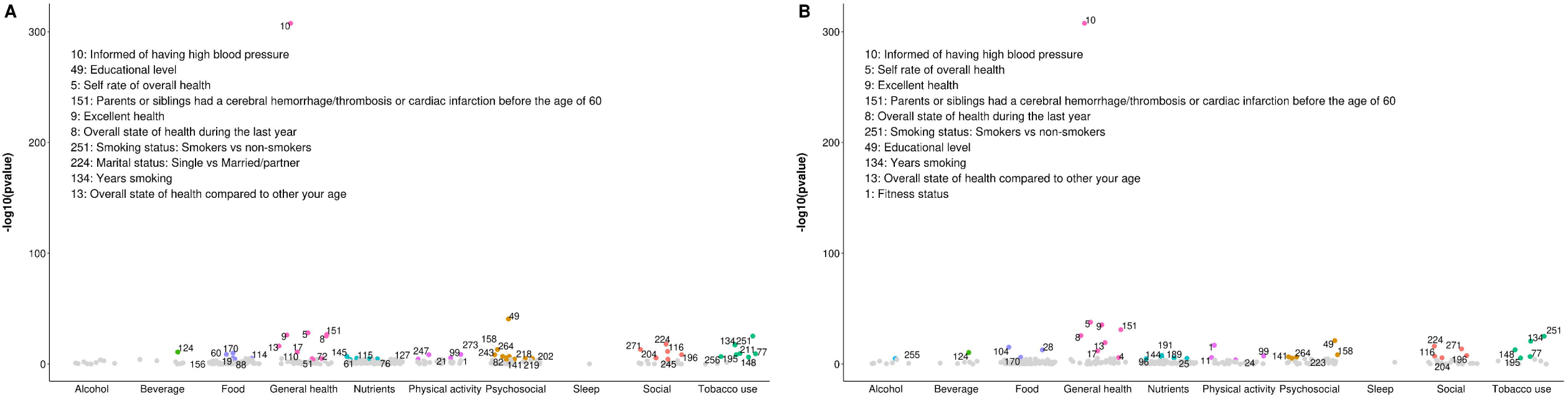
‘Manhattan plot’ representing the distribution of *P* values of the association of lifestyle variables and blood pressure traits by lifestyle category. A) Systolic blood pressure and B) Diastolic blood pressure ‘Tentative signals’ are coloured and number labeled in the figure and the top 10 variables are spelled out. See S25 for references to the labels

**Fig 4.**
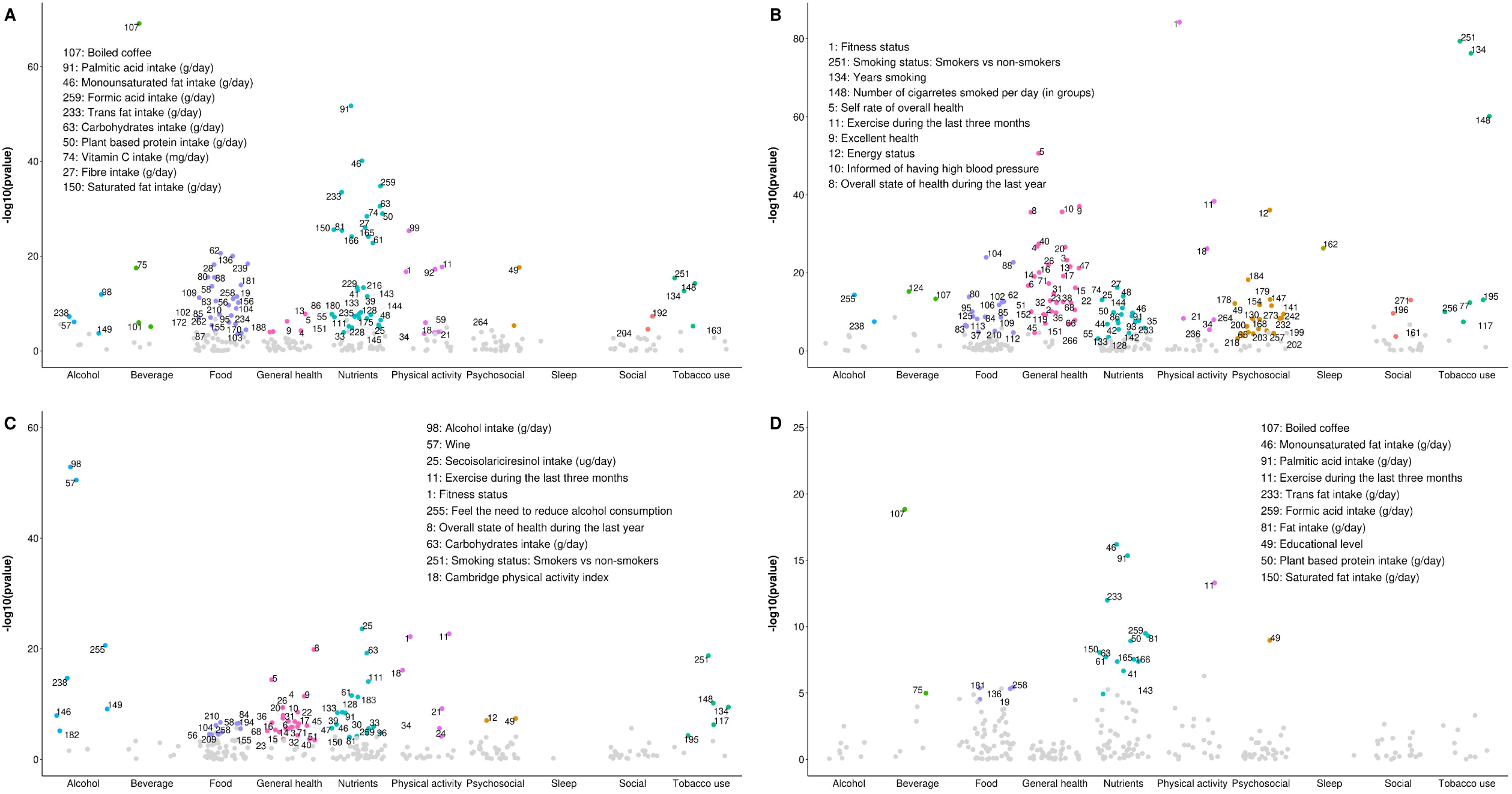
‘Manhattan plot’ representing the distribution of *P* values of the association of lifestyle variables and lipid traits by lifestyle category. A) Total cholesterol, B) Triglycerides, C) HDL cholesterol and D) LDL cholesterol ‘Tentative signals’ are coloured and number labeled in the figure and the top 10 variables are spelled out. See S25 for references to the labels

**Fig 5.**
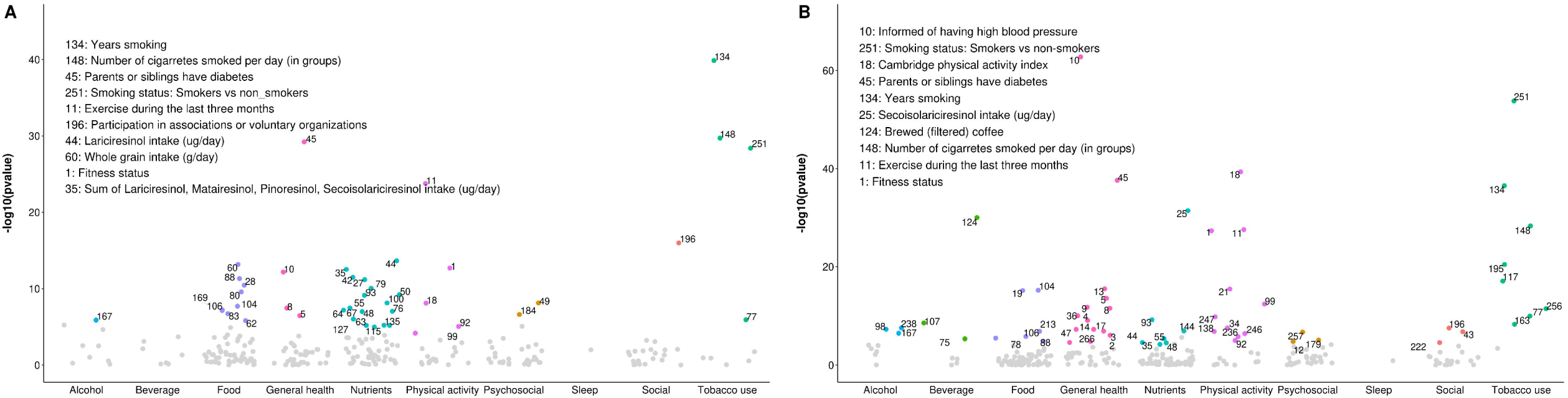
‘Manhattan plot’ representing the distribution of *P* values of the association of lifestyle variables and glucose traits by lifestyle category. A) Fasting glucose and B) 2h glucose ‘Tentative signals’ are coloured and number labeled in the figure and the top 10 variables are spelled out. See S25 for references to the labels

### Long-term lifestyle associations

After multiple testing correction, 35 lifestyle variables showed a tentative association with 10-year change in BMI (S12), 3 with change in SBP and DBP (S13-S14), 15 with change in total cholesterol (S15), 10 in triglycerides (S16), none in HDL and LDL cholesterol (S17-S18), 5 in fasting glucose (S19) and 8 in 2h glucose (S20). The majority of the ‘tentative signals’ were in the ‘Tobacco use’ category for BMI, lipids and fasting glucose, while for blood pressure traits the top-category was ‘General health’ and for 2h glucose, ‘Physical activity’, ‘Food’, and ‘General health’ were the top-categories.

### Correlation patterns

Patterns of correlations were identified among lifestyle variables showing tentative association with any of the cardiometabolic traits based on the correlation heatmap (Figure 6). Variables related to meat and fish consumption, sodium, calcium, vitamin B12, and total and animal based protein intake appeared in close proximity showing correlations around 0.5. Variables describing fat consumption and fatty acid intakes were grouped together showing a high positive correlation. Variables assessing vegetable, fibre and fruit intake, plant lignans, whole grain intake, and carbohydrates intake also appear near each other in the heatmap showing high positive correlations between them and negative correlations with fat related variables. Variables in ‘Psychosocial’ category and ‘General health’ variables were grouped together.

**Fig 6.**
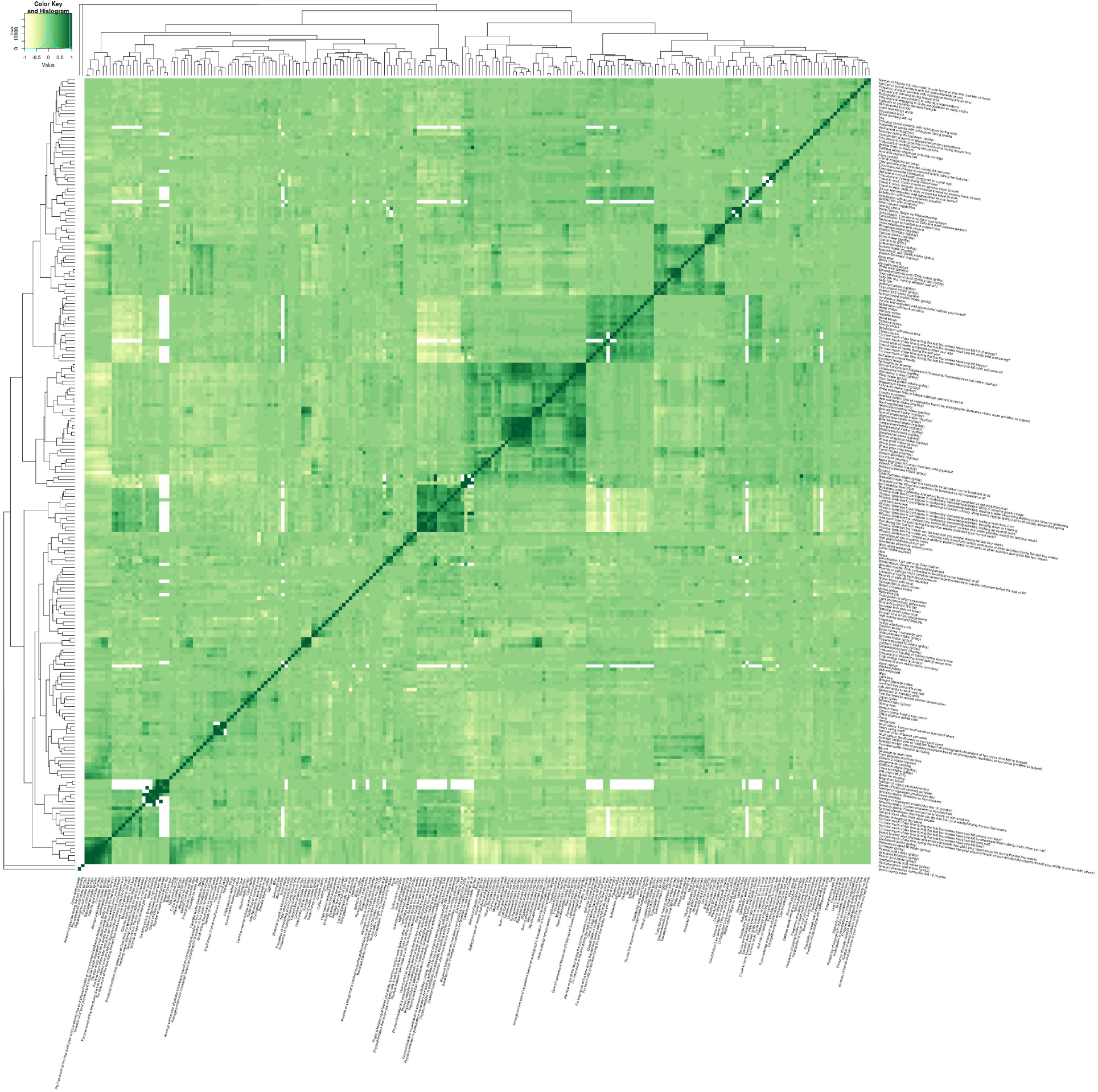
Heat map showing all the correlations for tentative signals Pairs of factors where correlations could not be computed are shown in white

### Prioritization of modifiable lifestyle variables

#### Average lifestyle associations

Sixteen variables were prioritized among all the ‘tentative signals’ as they showed the most consistent associations across all the cardiometabolic traits (top-ranked in at least 5 out of 9 cardiometabolic traits) (S21). Three of these variables (‘Informed of having a high blood pressure’, ‘Overall state of health during the last year’ and ‘Educational level’) were considered non-modifiable and excluded. The rest of the variables were included in a hierarchical clustering algorithm which identified four main target groups suitable for interventions (Figure 7). The first group included smoking related variables and were in general positively associated with BMI, fasting glucose, total cholesterol and triglycerides and negatively with blood pressure traits, HDL cholesterol and 2h glucose (S21).

**Fig 7.**
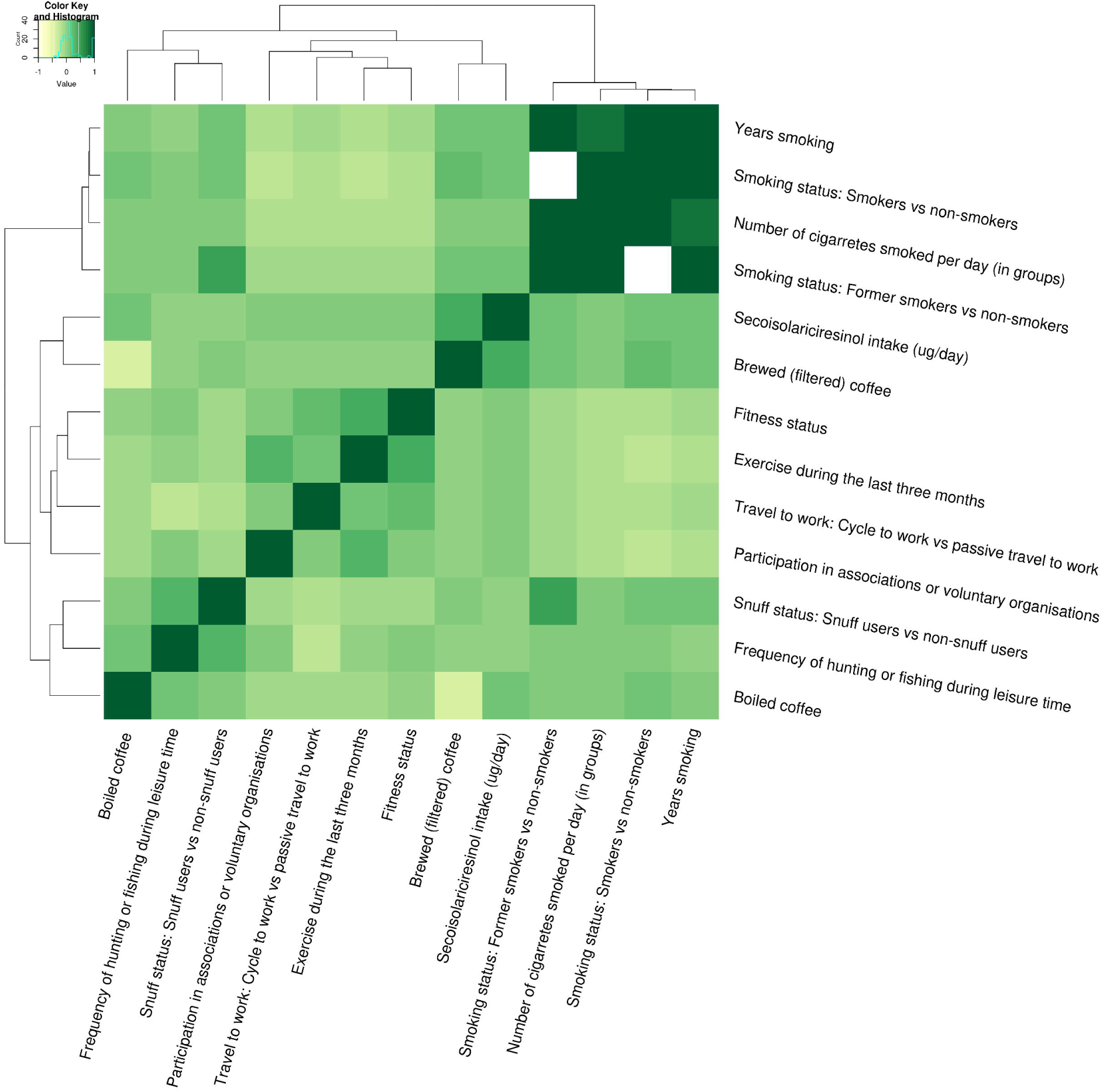
Heat map showing clusters of correlations between top-ranked modifiable lifestyle variables

The second group included two diet variables, the plant lignand ‘Secoisolariciresinol intake (µg/day)’, which was negatively associated with BMI, DBP and triglycerides and positively with HDL cholesterol; and ‘Brewed (filtered) coffee’, which was negatively associated with BMI, blood pressure traits, triglycerides and 2h glucose. The third group included physical activity related variables (e.g. ‘Exercise during the last three months’). These variables were in general negatively associated with all cardiometabolic traits except with HDL-C with which they showed a positive association. Altough this exposure group included variables related to leisure time physical activity, no variables related to the amount of physical activity at work were included in this group. The fourth group was composed of three lifestyle variables which could be linked to the Swedish lifestyle (specially northern Swedish lifestyle), ‘Snuff status: Snuff users vs non-snuff users’, ‘Frequency of hunting or fishing during leisure time’ and ‘Boiled coffee’. These three variables did not show a clear common pattern of associations with cardiometabolic traits.

In general, BMI showed more shared tentative signals with 2h glucose and HDL-cholesterol than with the rest of cardiometabolic traits and triglycerides, BMI and 2h glucose were the cardiometabolic traits sharing the highest number of tentative signals with the rest of cardiometabolic traits (S22).

#### Long-term lifestyle associations

None of the ‘tentative signals’ showed a consistent association with the majority of cardiometabolic traits (5 out 9 traits) (S23). However, four variables in the ‘Tobacco use’ category showed a consistent positive associations with 10-year changes in at least three cardiometabolic traits (BMI, total cholesterol, triglycerides and/or fasting glucose).

Among all the cardiometabolic traits BMI and lipid traits shared the highest number of tentative signals (S24).

## DISCUSSION

Although EWAS analyses have been reported previously, this is the first study to integrate repeated exposures and outcome assessments, which allow us to make inferences about long-term exposure to these risk factors. Here, we systematically and agnostically assessed average (across the study’s follow-up time) and 10-year associations between 287 lifestyle variables and 9 cardiometabolic traits. In analyses assessing average association of lifestyle variables, ‘Physical activity’ and ‘General Health’ were the categories containing the highest number of ‘tentative signals’ and 13 modifiable variables were prioritized for lifestyle interventions focused on controlling cardiometabolic diseases. A cluster analyses grouped these 13 variables into four main target groups: i) Smoking, ii) Diet (specifically secoisolariciresinol intake and brewed coffee), iii) Leisure time physical activity and iv) a group more specific to the lifestyle in Sweden (including snuff, boiled coffee and frequency of hunting and fishing).

For 10-year associations, ‘Tobacco use’ was the category including the highest number of ‘tentative signals’ for the majority of the cardiometabolic traits. No modifiable lifestyle variable was consistently associated with the majority of cardiometabolic traits but four variables in the ‘Tobacco use’ category were consistently associated with at least three of the analysed cardiometabolic traits (BMI, total cholesterol, triglycerides and/or fasting glucose).

Smoking and physical activity correspond to two of the most well-known modifiable risk factors for cardiometabolic diseases. According to a study analysing the burden of disease caused by physical inactivity, worldwide, 6% of the burden of coronary heart disease and 7% of type 2 diabetes was caused by physical inactivity (22). On the other hand, smoking alters lipid metabolism and glucose homeostasis through the increase in lipolysis, insulin resistance and tissue lipotoxicity (23, 24) and smoking cessation restores, at least in part, the mentioned metabolic alterations. However, in our study the association of smoking with cardiometabolic traits was not only restricted to the average effect across the studied period but we also found a remarkable association of variables included in the ‘Tobacco use’ category and cardiometabolic traits in the 10 years of follow-up.

Among the prioritized dietary variables, boiled (unfiltered) coffee but not brewed (filtered) coffee was found positively associated with lipid traits, specifically with total cholesterol, triglycerides, and LDL cholesterol. Previous metanalyses have also identified associations between unfiltered coffee and dose-dependent increase of plasma concentrations of total and LDL cholesterol (25, 26). The effects of coffee in the lipid profile are probably caused by two diterpenes (i.e. kahweol and cafestol), which sometimes get trapped in the filter used to make coffee which can explain the differential effects of filtered and unfiltered coffee (26). On the other hand, brewed (filtered) coffee was found negatively associated with BMI, blood pressure, triglycerides, and 2h glucose in the present study which is in agreement with previous studies showing an inverse association between habitual coffee intake and risk of several cardiometabolic diseases (27, 28).

Plant lignans (biphenolic compounds found in tea, coffee, whole-grain products, berries, vegetables, fruit, nuts and seeds) were among the top-tentative signals for fasting and 2h glucose, showing a negative association with both traits and secoisolariciresinol intake was a prioritized variable. Previous studies have suggested that lignans and their metabolites may protect against cardiovascular disease and metabolic syndrome by reducing lipid concentrations, lowering blood pressure, and decreasing oxidative stress and inflammation (29). A study conducted in Finland found that men with high serum concentrations of enterolactone (a lignan produced by the intestinal microflora) had a lower risk of acute coronary events than men with lower concentrations (30).

An interesting observation emerging from our analysis is that several variables that are featured in public health recommendations were not broadly associated with the cardiometabolic traits studied here. Recommended dietary patterns emphasize the importance of limiting the consumption of sugar-rich products, particularly sweet drinks (31). However, variables related to sweets and sweet drink consumption (e.g. “Sodas, soft drinks, juice” and “Sweets”) were not identified as tentative signals for any of the cardiometabolic traits. Salt content is also usually limited in diets recommended to lower risk of cardiometabolic diseases but “Sodium intake” was not consistently associated with cardiometabolic traits, being identified as a tentative signal only for BMI, total and HDL cholesterol. In the same way, fish and shellfish are frequently recommended in healthy dietary patterns but “Lean fish” and “Shellfish” variables were not tentative signals for any cardiometabolic traits and “Fatty fish” was associated with lipid traits except for LDL cholesterol.

There are also limitations to the present study. The present sample is limited to a Swedish population between 30-70 years and thus caution should be used when extrapolating the findings to other countries and age groups, specially since lifestyle variables affecting cardiometabolic traits in Swedish population might differ from other populations. Dietary variables were characterized using a FFQ, which suffer from systematic and random measurement errors. However, to minimize this source of error the FFQ used in this study was validated against repeated 24 hour recalls (17). VIP cohort is exceptionally well-powered for analyses of the nature performed here and there were, consequently, a large number of associations that passed conventional statistical thresholds. Most of these statistically robust associations emerged due to the complex correlation structure (Figure 6) found within the set of exposure variables. EWAS analyses assess the association of each lifestyle variable individually and do not take into account the complex interrelations that exist between these variables. Although the results were internally validated, ideally these findings would need to be replicated in independent samples. However, it is important to keep in mind the difficulty to standardize lifestyle data, owing to the variety of ways in which these exposures are captured and the manner in which they are arrayed, which complicates the task of finding a replication dataset focusing on the same set of lifestyle variables.

In conclusion, using an EWAS approach in a large prospective Swedish cohort a large number of associations between lifestyle exposures and cardiometabolic traits were identified. Thirteen modifiable exposures were consistently top-ranked among the majority of cardiometabolic traits and were identified as target lifestyle exposures that could inform lifestyle interventions aiming at controlling cardiometabolic diseases. These variables belonged to four target groups: i) Smoking, ii) Diet (specifically plant lignans and brewed (filtered) coffee) and iii) Leisure time physical activity and iv) a group of lifestyle more specific to the Swedish lifestyle. Interestingly, sweet drinks, fish intake and salt content, all lifestyle exposures frequently mentioned in public health recommendations were not broadly associated with the analysed cardiometabolic traits.

## Data Availability

The data generated and analyzed are not publicly available due to Swedish legislation but are available from the Department of Biobank Research, Umea University, on reasonable request and with appropriate approvals

## Ackowledgements

We thank the participants, health professionals and data managers involved in the Västerbotten Intervention Programme.

