## Supplementary material for "Environment-wide association study (EWAS) on cardiometabolic traits: A systematic assessment of the association of lifestyle variables on a longitudinal setting"

**Limit values proposed by VIP for exclusions on cardiometabolic traits**

Height: <130 cm or >210 cm

Weight: <35 kg

BMI: <15 kg/m^2^ or >70 kg/m^2^

Systolic blood pressure: <20 or >300

Diastolic blood pressure: <20 or >250

Total cholesterol: <0.5 mmol/l or >15 mmol/l

Triglycerides: <0.15 mmol/l or >20 mmol/l. Triglycerides values lower than 0.8 mmol/l were additionally excluded due to the sensitivity of the Reflotron benchtop analyzer.

HDL-cholesterol: <0.15 mmol/l or>7 mmol/l

LDL-cholesterol: Not defined. LDL cholesterol values lower than 0.5 mmol/l and higher than 13 mmol/l were excluded.

Fasting glucose: <1 mmol/l or >25 mmol/l. Fasting glucose values lower than 2 mmol/l were additionally excluded as they were considered biologically implausible.

2h glucose: <1 mmol/l or >35 mmol/l. 2h glucose values lower than 2 mmol/l were additionally excluded as they were considered biologically implausible.

**Values considered implausible for certain lifestyle variables**

Distance to work in kilometers (one way): All answers beyond 200 km were excluded.

Grams of tobacco smoked per week: All answers equal or beyond 350 gr/week were excluded.

Arachidonic acid (ARA) intake (g/day): All answers equal or beyond 0.9 gr/day were excluded.

Eicosapentaenoic acid (EPA) intake (g/day): All answers equal or beyond 2 gr/day were excluded.

Sodium intake (mg/day): All answers equal or beyond 10000 mg/day were excluded.

**R packages used in the analyses**

Data manipulation: psych (1), data.table (2), plyr (3), dplyr (4) and tidyverse (5)

EWAS analyses: getopt (6) and nlme (7)

Metanalysis: meta (8)

R^2^ estimation: piecewiseSEM (9)

Correlation estimation: polycor (10)

Data visualization: ggplot2 (11), ggrepel (12), gridExtra (13), RColorBrewer (14) and gplots (15)
