## Supplementary tables for "Environment-wide association study (EWAS) on cardiometabolic traits: A systematic assessment of the association of lifestyle variables on a longitudinal setting"

**Supplementary Table 1**. Descriptive statistics of the numeric lifestyle variables in training and testing sets

| | Training set | | | | | | Testing set | | | | | |
|---|---|---|---|---|---|---|---|---|---|---|---|---|
| **Variables** | **N of observations** | **Mean** | **SD** | **Median** | **Min** | **Max** | **N of observations** | **Mean** | **SD** | **Median** | **Min** | **Max** |
| Distance to work in kilometers (one way) | 29107 | 13.84 | 17.26 | 6.00 | 0.00 | 188.00 | 28369 | 14.14 | 17.71 | 6.00 | 0.00 | 165.00 |
| Number of cigarretes smoked per day | 21861 | 0.31 | 2.15 | 0.00 | 0.00 | 50.00 | 21431 | 0.32 | 2.14 | 0.00 | 0.00 | 40.00 |
| Number of cigars smoked per day | 21309 | 0.01 | 0.28 | 0.00 | 0.00 | 20.00 | 20855 | 0.01 | 0.26 | 0.00 | 0.00 | 25.00 |
| Grams of tobacco smoked per week | 21324 | 0.08 | 2.47 | 0.00 | 0.00 | 175.00 | 20876 | 0.10 | 3.01 | 0.00 | 0.00 | 275.00 |
| Years smoking | 29638 | 9.38 | 13.07 | 0.00 | 0.00 | 51.30 | 28849 | 9.47 | 13.18 | 0.00 | 0.00 | 52.00 |
| Years using snuff | 31829 | 4.47 | 9.37 | 0.00 | 0.00 | 50.00 | 31140 | 4.56 | 9.46 | 0.00 | 0.00 | 51.00 |
| Food intake level | 31996 | 1.08 | 0.39 | 1.03 | 0.09 | 17.97 | 31316 | 1.08 | 0.38 | 1.02 | 0.14 | 9.81 |
| Total energy intake (kcal/day) | 29792 | 1757.84 | 541.91 | 1675.70 | 663.16 | 4363.79 | 29020 | 1751.35 | 538.41 | 1668.31 | 718.98 | 4332.38 |
| Total protein intake (g/day) | 29792 | 63.97 | 20.48 | 60.94 | 11.28 | 197.75 | 29020 | 63.77 | 20.38 | 60.79 | 18.21 | 201.49 |
| Animal based protein intake (g/day) | 29792 | 45.45 | 16.45 | 42.97 | 0.27 | 166.33 | 29020 | 45.35 | 16.46 | 42.81 | 0.13 | 178.47 |
| Plant based protein intake (g/day) | 29792 | 18.52 | 7.00 | 17.45 | 1.35 | 71.98 | 29020 | 18.42 | 6.91 | 17.32 | 0.49 | 72.43 |
| Carbohydrates intake (g/day) | 29792 | 210.02 | 70.61 | 199.72 | 18.93 | 663.42 | 29020 | 208.85 | 69.78 | 198.78 | 17.72 | 636.13 |
| Sucrose intake (g/day) | 29792 | 27.55 | 16.16 | 23.89 | 0.77 | 155.39 | 29020 | 27.30 | 16.17 | 23.53 | 1.33 | 178.93 |
| Disaccharides intake (g/day) | 29792 | 50.79 | 22.81 | 46.72 | 4.05 | 226.17 | 29020 | 50.36 | 22.66 | 46.36 | 6.04 | 242.42 |
| Monosaccharides intake (g/day) | 29792 | 25.17 | 11.84 | 23.21 | 1.87 | 129.95 | 29020 | 24.93 | 11.76 | 22.87 | 2.48 | 135.93 |
| Fibre intake (g/day) | 29792 | 18.87 | 7.11 | 17.90 | 1.31 | 68.86 | 29020 | 18.77 | 7.07 | 17.81 | 2.00 | 78.20 |
| Whole grain intake (g/day) | 29792 | 69.22 | 35.10 | 64.09 | 0.00 | 299.39 | 29020 | 68.77 | 35.03 | 63.26 | 0.00 | 307.12 |
| Alcohol intake (g/day) | 29792 | 4.40 | 4.62 | 3.21 | 0.00 | 73.14 | 29020 | 4.47 | 4.86 | 3.20 | 0.00 | 123.62 |
| Fat intake (g/day) | 29792 | 68.55 | 26.25 | 63.98 | 13.87 | 297.24 | 29020 | 68.42 | 26.56 | 63.72 | 10.52 | 269.06 |
| Saturated fat intake (g/day) | 29792 | 28.66 | 12.17 | 26.42 | 2.97 | 151.84 | 29020 | 28.61 | 12.26 | 26.36 | 3.44 | 133.40 |
| Monounsaturated fat intake (g/day) | 29792 | 23.48 | 9.24 | 21.79 | 4.18 | 101.43 | 29020 | 23.40 | 9.33 | 21.61 | 3.05 | 96.31 |
| Polyunsaturated fat intake (g/day) | 29792 | 10.81 | 5.09 | 9.73 | 1.90 | 74.51 | 29020 | 10.80 | 5.09 | 9.70 | 2.24 | 69.12 |
| Trans fat intake (g/day) | 29792 | 1.76 | 1.25 | 1.42 | 0.11 | 12.08 | 29020 | 1.74 | 1.23 | 1.41 | 0.01 | 15.33 |
| Cholesterol intake (g/day) | 29792 | 0.20 | 0.08 | 0.18 | 0.01 | 0.90 | 29020 | 0.20 | 0.08 | 0.18 | 0.01 | 0.81 |
| Formic acid intaje (g/day) | 29792 | 3.27 | 1.65 | 2.94 | 0.11 | 21.71 | 29020 | 3.26 | 1.66 | 2.93 | 0.29 | 19.28 |
| Palmitic acid intake (g/day) | 29792 | 14.54 | 5.80 | 13.46 | 1.97 | 73.01 | 29020 | 14.51 | 5.85 | 13.43 | 2.05 | 62.10 |
| Linoleic acid intake (g/day) | 29792 | 7.85 | 4.07 | 6.94 | 1.48 | 59.71 | 29020 | 7.82 | 4.05 | 6.91 | 1.47 | 52.66 |
| Linolenic acid intake (g/day) | 29792 | 1.60 | 0.73 | 1.46 | 0.26 | 9.48 | 29020 | 1.60 | 0.73 | 1.46 | 0.30 | 9.27 |
| Arachidonic acid (ARA) intake (g/day) | 29791 | 0.09 | 0.05 | 0.08 | 0.00 | 0.63 | 29019 | 0.09 | 0.05 | 0.08 | 0.00 | 0.68 |
| Eicosapentaenoic acid (EPA) intake (g/day) | 29791 | 0.07 | 0.06 | 0.06 | 0.00 | 1.23 | 29020 | 0.07 | 0.06 | 0.05 | 0.00 | 1.43 |
| Docosahexaenoic acid (DHA) intake (g/day) | 29792 | 0.14 | 0.12 | 0.12 | 0.00 | 4.40 | 29020 | 0.14 | 0.12 | 0.12 | 0.00 | 3.07 |
| Pentadecanoic acid intake (g/day) | 29792 | 0.23 | 0.15 | 0.20 | 0.00 | 2.03 | 29020 | 0.23 | 0.15 | 0.20 | 0.00 | 1.70 |
| Heptadecanoic acid intake (g/day) | 29792 | 0.13 | 0.09 | 0.11 | 0.00 | 1.18 | 29020 | 0.13 | 0.09 | 0.11 | 0.00 | 0.99 |
| Magnesium intake (mg/day) | 29792 | 292.79 | 88.60 | 280.67 | 76.03 | 1154.65 | 29020 | 291.92 | 87.89 | 279.72 | 89.58 | 790.94 |
| Phosphate intake (mg/day) | 29792 | 1180.92 | 394.98 | 1122.19 | 249.07 | 3757.68 | 29020 | 1176.71 | 392.71 | 1121.59 | 320.01 | 3740.93 |
| Vitamin B3 intake (mg/day) | 29792 | 15.12 | 5.07 | 14.23 | 3.08 | 55.12 | 29020 | 15.08 | 5.04 | 14.23 | 4.04 | 61.00 |
| Selenium intake (ug/day) | 29792 | 22.74 | 8.25 | 21.55 | 3.00 | 102.17 | 29020 | 22.64 | 8.22 | 21.33 | 3.92 | 128.78 |
| Zinc intake (mg/day) | 29792 | 8.20 | 2.68 | 7.80 | 1.41 | 28.59 | 29020 | 8.18 | 2.67 | 7.77 | 1.36 | 23.23 |
| Vitamin A intake (mg/day) | 29792 | 0.61 | 0.33 | 0.55 | 0.03 | 4.35 | 29020 | 0.61 | 0.32 | 0.54 | 0.07 | 5.98 |
| Beta-carotene intake (mg/day) | 29792 | 4.28 | 4.59 | 2.75 | 0.08 | 45.85 | 29020 | 4.22 | 4.55 | 2.69 | 0.07 | 44.68 |
| Tiamin intake (mg/day) | 29792 | 1.21 | 0.41 | 1.15 | 0.32 | 4.38 | 29020 | 1.21 | 0.41 | 1.15 | 0.17 | 4.05 |
| Folic acid intake (ug/day) | 29792 | 233.37 | 90.59 | 218.87 | 42.18 | 938.70 | 29020 | 232.77 | 90.80 | 217.73 | 48.22 | 1052.67 |
| Vitamin B2 intake (ug/day) | 29792 | 1.44 | 0.52 | 1.37 | 0.31 | 5.20 | 29020 | 1.43 | 0.52 | 1.36 | 0.26 | 5.01 |
| Vitamin B6 intake (mg/day) | 29792 | 1.92 | 0.65 | 1.82 | 0.48 | 9.29 | 29020 | 1.91 | 0.64 | 1.81 | 0.45 | 6.36 |
| Vitamin B12 intake (ug/day) | 29792 | 4.68 | 2.13 | 4.29 | 0.03 | 23.79 | 29020 | 4.65 | 2.12 | 4.27 | 0.18 | 28.27 |

| | | | | | | | | | | |
|---|---|---|---|---|---|---|---|---|---|---|
| Vitamin C intake (mg/day) | 29792 | 74.21 | 45.69 | 63.51 | 0.71 | 535.69 | 29020 | 73.72 | 45.20 | 62.90 | 3.21 | 464.93 |
| Vitamin D intake (ug/day) | 29792 | 5.41 | 2.17 | 5.06 | 0.63 | 26.17 | 29020 | 5.38 | 2.17 | 5.03 | 0.72 | 33.19 |
| Vitamin E intake (mg/day) | 29792 | 6.38 | 2.31 | 6.00 | 1.61 | 26.07 | 29020 | 6.36 | 2.33 | 5.98 | 1.63 | 31.05 |
| Vitamin K intake (ug/day) | 29792 | 3.93 | 2.38 | 3.34 | 0.10 | 27.77 | 29020 | 3.88 | 2.33 | 3.30 | 0.00 | 29.11 |
| Iron intake (mg/day) | 29792 | 12.26 | 4.34 | 11.54 | 1.88 | 46.73 | 29020 | 12.20 | 4.28 | 11.49 | 2.71 | 44.92 |
| Iodine intake (ug/day) | 29792 | 121.46 | 50.51 | 113.43 | 5.53 | 641.77 | 29020 | 121.15 | 50.64 | 112.77 | 17.02 | 649.79 |
| Calcium intake (mg/day) | 29792 | 786.95 | 342.62 | 734.03 | 92.10 | 3244.76 | 29020 | 782.64 | 339.40 | 730.57 | 100.61 | 3102.59 |
| Potassium intake (mg/day) | 29792 | 3192.42 | 1001.31 | 3059.09 | 685.88 | 14480.73 | 29020 | 3182.10 | 991.63 | 3040.47 | 835.48 | 9714.26 |
| Beta-sitosterol intake (mg/day) | 29792 | 147.43 | 52.14 | 139.68 | 20.40 | 603.68 | 29020 | 146.66 | 51.63 | 139.16 | 8.44 | 504.10 |
| Beta-sitostanol intake (mg/day) | 29792 | 8.39 | 3.64 | 7.77 | 0.38 | 31.64 | 29020 | 8.35 | 3.61 | 7.69 | 0.14 | 34.95 |
| Campesterol intake (mg/day) | 29792 | 56.16 | 22.79 | 52.25 | 6.30 | 240.08 | 29020 | 55.91 | 22.79 | 51.91 | 2.54 | 256.68 |
| Campestanol intake (mg/day) | 29792 | 5.46 | 2.65 | 4.98 | 0.13 | 25.04 | 29020 | 5.43 | 2.63 | 4.94 | 0.05 | 25.75 |
| Stigmasterol intake (mg/day) | 29792 | 11.52 | 4.16 | 10.90 | 1.62 | 41.66 | 29020 | 11.47 | 4.13 | 10.86 | 0.76 | 50.25 |
| Sum of phytosterols intake (mg/day) | 29792 | 228.97 | 81.52 | 216.68 | 31.19 | 923.76 | 29020 | 227.82 | 80.87 | 215.57 | 11.92 | 797.71 |
| Enterodiol intake (ug/day) | 29792 | 0.01 | 0.02 | 0.00 | 0.00 | 0.16 | 29020 | 0.01 | 0.02 | 0.00 | 0.00 | 0.16 |
| Enterolactone intake (ug/day) | 29792 | 5.54 | 5.68 | 4.03 | 0.00 | 62.29 | 29020 | 5.50 | 5.65 | 3.83 | 0.00 | 62.01 |
| Equol intake (ug/day) | 29792 | 0.26 | 0.39 | 0.06 | 0.00 | 3.72 | 29020 | 0.25 | 0.38 | 0.06 | 0.00 | 3.61 |
| Lariciresinol intake (ug/day) | 29792 | 508.86 | 212.57 | 475.76 | 50.79 | 2638.93 | 29020 | 506.95 | 211.29 | 474.72 | 42.82 | 2377.42 |
| Matairesinol intake (ug/day) | 29792 | 40.78 | 19.58 | 37.85 | 1.63 | 163.86 | 29020 | 40.69 | 19.61 | 37.53 | 2.55 | 178.21 |
| Medioresinol intake (ug/day) | 29792 | 509.16 | 288.94 | 454.50 | 0.23 | 2478.11 | 29020 | 506.79 | 289.02 | 450.35 | 0.66 | 2726.08 |
| Pinoresinol intake (ug/day) | 29792 | 394.00 | 190.00 | 363.42 | 11.70 | 2372.68 | 29020 | 392.88 | 189.22 | 363.82 | 12.20 | 1827.69 |
| Secoisolariciresinol intake (ug/day) | 29792 | 140.39 | 48.34 | 134.91 | 16.47 | 725.99 | 29020 | 140.31 | 48.08 | 134.46 | 13.07 | 668.70 |
| Syringaresinol intake (ug/day) | 29792 | 1331.81 | 700.47 | 1205.52 | 0.23 | 6101.68 | 29020 | 1326.26 | 701.07 | 1193.52 | 1.26 | 6603.59 |
| Sum of all lignans intake (ug/day) | 29792 | 2930.82 | 1373.91 | 2706.35 | 208.63 | 12267.07 | 29020 | 2919.65 | 1372.77 | 2686.25 | 174.30 | 12827.25 |
| Sum of Lariciresinol, Matairesinol, Pinoresinol, Secoisolariciresino | 29792 | 1084.03 | 449.69 | 1014.77 | 107.80 | 5749.52 | 29020 | 1080.84 | 447.02 | 1012.01 | 87.72 | 4791.20 |
| Sodium intake (mg/day) | 29791 | 2074.94 | 730.70 | 1955.62 | 446.15 | 8629.67 | 29019 | 2064.96 | 726.19 | 1945.70 | 490.91 | 8372.63 |
| Bregott on bread | 29792 | 9.49 | 10.97 | 5.00 | 0.00 | 60.00 | 29020 | 9.31 | 10.90 | 5.00 | 0.00 | 60.00 |
| Butter on bread | 29792 | 1.14 | 5.94 | 0.00 | 0.00 | 80.00 | 29020 | 1.25 | 6.22 | 0.00 | 0.00 | 80.00 |
| Low fat margarine on bread | 29792 | 6.66 | 9.93 | 0.56 | 0.00 | 60.00 | 29020 | 6.58 | 9.92 | 0.40 | 0.00 | 60.00 |
| Margarine on bread | 29792 | 0.61 | 3.36 | 0.00 | 0.00 | 60.00 | 29020 | 0.59 | 3.34 | 0.00 | 0.00 | 60.00 |
| Butter for cooking | 29792 | 2.65 | 6.07 | 0.06 | 0.00 | 100.00 | 29020 | 2.56 | 5.92 | 0.06 | 0.00 | 100.00 |
| Margarine for cooking | 29792 | 8.15 | 9.57 | 7.00 | 0.00 | 100.00 | 29020 | 8.34 | 9.81 | 7.00 | 0.00 | 100.00 |
| Oil for cooking | 29792 | 4.52 | 6.53 | 2.00 | 0.00 | 80.00 | 29020 | 4.54 | 6.51 | 2.00 | 0.00 | 80.00 |
| Salad dressing with oil | 29792 | 3.14 | 5.30 | 1.82 | 0.00 | 96.00 | 29020 | 3.03 | 5.18 | 1.82 | 0.00 | 96.00 |
| Cream, creme fraiche, sour cream | 29792 | 5.48 | 6.39 | 3.50 | 0.00 | 160.00 | 29020 | 5.54 | 6.62 | 3.50 | 0.00 | 160.00 |
| Whole grain crisp bread | 29792 | 27.94 | 21.46 | 21.00 | 0.00 | 104.00 | 29020 | 27.92 | 21.68 | 18.20 | 0.00 | 104.00 |
| Whole grain soft bread | 29792 | 31.57 | 34.63 | 20.52 | 0.00 | 264.00 | 29020 | 31.30 | 34.70 | 20.52 | 0.00 | 264.00 |
| White (soft) bread, thin crisp bread | 29792 | 22.45 | 27.54 | 12.78 | 0.00 | 298.00 | 29020 | 22.07 | 26.99 | 12.78 | 0.00 | 247.50 |
| Coffee rolls/buns, rusk | 29792 | 12.10 | 15.83 | 6.16 | 0.00 | 176.00 | 29020 | 11.78 | 15.41 | 5.88 | 0.00 | 176.00 |
| Cheese 28% | 29792 | 12.00 | 13.22 | 7.20 | 0.00 | 96.00 | 29020 | 12.04 | 13.22 | 7.20 | 0.00 | 96.00 |
| Cheese 10-17% | 29792 | 5.31 | 9.86 | 1.52 | 0.00 | 84.00 | 29020 | 5.34 | 9.99 | 1.52 | 0.00 | 84.00 |
| Soft cheese | 7163 | 1.00 | 3.30 | 0.05 | 0.00 | 55.00 | 6910 | 0.93 | 2.96 | 0.05 | 0.00 | 72.00 |
| Soft whey cheese | 7163 | 0.79 | 3.07 | 0.00 | 0.00 | 60.00 | 6910 | 0.89 | 3.56 | 0.01 | 0.00 | 60.00 |
| Sausage, liver pate on bread | 29792 | 4.81 | 6.82 | 2.13 | 0.00 | 152.10 | 29020 | 4.78 | 6.82 | 2.10 | 0.00 | 139.22 |
| Meat on bread | 29792 | 4.56 | 6.06 | 2.10 | 0.00 | 112.00 | 29020 | 4.58 | 6.27 | 2.10 | 0.00 | 92.00 |
| Oatflake, whole wheat, rye or barley porridge | 29792 | 35.20 | 54.31 | 16.00 | 0.00 | 800.60 | 29020 | 35.52 | 55.92 | 16.00 | 0.00 | 800.00 |
| Rosehip, sweet syrup soup | 29792 | 9.30 | 20.74 | 0.53 | 0.00 | 500.00 | 29020 | 9.15 | 21.35 | 0.53 | 0.00 | 700.00 |
| Sour milk, yoghurt (3% fat) | 29792 | 94.92 | 106.82 | 39.20 | 0.00 | 1200.00 | 29020 | 94.41 | 107.71 | 39.20 | 0.00 | 1200.00 |
| Sour milk, yoghurt (low fat) | 29792 | 53.96 | 90.60 | 18.00 | 0.00 | 1120.00 | 29020 | 53.76 | 90.83 | 18.00 | 0.00 | 1120.00 |

| | | | | | | | | | | | |
|---|---|---|---|---|---|---|---|---|---|---|---|
| Fiber cereals | 29792 | 10.57 | 13.75 | 3.60 | 0.00 | 180.00 | 29020 | 10.64 | 14.15 | 3.60 | 0.00 | 172.00 |
| Corn flakes | 29792 | 3.51 | 5.82 | 1.12 | 0.00 | 68.00 | 29020 | 3.37 | 5.71 | 1.12 | 0.00 | 88.00 |
| Berries (fresh or frozen) | 29792 | 8.66 | 14.05 | 5.20 | 0.00 | 260.00 | 29020 | 8.74 | 14.04 | 5.20 | 0.00 | 260.00 |
| Apple, pear, peach, orange, mandarin and grapefruit | 29792 | 109.64 | 102.24 | 80.50 | 0.00 | 920.00 | 29020 | 107.94 | 101.19 | 80.50 | 0.00 | 920.00 |
| Banana | 29792 | 42.35 | 45.05 | 37.80 | 0.00 | 420.00 | 29020 | 42.35 | 45.67 | 37.80 | 0.00 | 420.00 |
| Root vegetables, carrot | 29792 | 35.13 | 45.20 | 18.00 | 0.00 | 416.00 | 29020 | 34.53 | 44.61 | 18.00 | 0.00 | 416.00 |
| Tomato, cucumber | 29792 | 36.68 | 41.46 | 23.40 | 0.00 | 380.00 | 29020 | 37.38 | 42.64 | 23.40 | 0.00 | 380.00 |
| White cabbage, lettuce, lettuce cabbage, spinach, borecole | 29792 | 27.26 | 35.48 | 17.16 | 0.00 | 333.32 | 29020 | 27.48 | 36.05 | 17.16 | 0.00 | 483.40 |
| Mixed frozen vegetables | 7163 | 8.04 | 12.75 | 4.80 | 0.00 | 250.00 | 6910 | 7.77 | 12.43 | 4.80 | 0.00 | 250.00 |
| Boiled or baked potato | 29792 | 115.86 | 95.49 | 86.40 | 0.00 | 1280.00 | 29020 | 116.05 | 94.78 | 86.40 | 0.00 | 1280.00 |
| Fried potatoes, pommes frites | 29792 | 13.40 | 15.86 | 10.72 | 0.00 | 487.50 | 29020 | 13.40 | 16.17 | 10.72 | 0.00 | 600.00 |
| Mashed potato | 7163 | 13.67 | 14.28 | 14.00 | 0.00 | 300.00 | 6910 | 13.68 | 14.38 | 14.00 | 0.00 | 210.00 |
| Potato salad | 5720 | 3.71 | 6.39 | 0.44 | 0.00 | 190.00 | 5519 | 3.62 | 5.69 | 0.44 | 0.00 | 68.40 |
| Rice | 29792 | 30.39 | 28.82 | 19.60 | 0.00 | 720.00 | 29020 | 30.10 | 28.67 | 19.60 | 0.00 | 600.00 |
| Pasta | 29792 | 42.80 | 36.28 | 28.00 | 0.00 | 500.00 | 29020 | 42.98 | 36.62 | 28.00 | 0.00 | 750.00 |
| Brown beans, pea soup | 29792 | 10.95 | 19.64 | 0.83 | 0.00 | 687.50 | 29020 | 10.74 | 18.24 | 0.83 | 0.00 | 687.50 |
| Blota (broth + bread) | 7163 | 0.69 | 2.05 | 0.24 | 0.00 | 80.00 | 6910 | 0.69 | 2.30 | 0.24 | 0.00 | 80.00 |
| Pancake, waffle, Swedish dumpling | 29792 | 23.43 | 18.83 | 21.20 | 0.00 | 375.00 | 29020 | 23.35 | 19.35 | 21.20 | 0.00 | 375.00 |
| Pizza | 29792 | 15.69 | 14.81 | 12.00 | 0.00 | 325.00 | 29020 | 15.57 | 14.85 | 12.00 | 0.00 | 375.00 |
| Minced meat dishes | 29792 | 20.94 | 17.42 | 17.50 | 0.00 | 500.00 | 29020 | 20.78 | 17.04 | 17.50 | 0.00 | 312.50 |
| Meat stew | 29792 | 17.45 | 15.45 | 14.00 | 0.00 | 210.00 | 29020 | 17.64 | 16.44 | 14.00 | 0.00 | 700.00 |
| Steak, chop, e.g. | 29792 | 15.28 | 12.67 | 12.00 | 0.00 | 250.00 | 29020 | 15.39 | 12.96 | 12.00 | 0.00 | 200.00 |
| Bacon | 29792 | 3.71 | 4.21 | 2.72 | 0.00 | 150.00 | 29020 | 3.76 | 4.25 | 2.72 | 0.00 | 105.00 |
| Sausage as main dish | 29792 | 11.96 | 10.55 | 10.50 | 0.00 | 200.00 | 29020 | 12.02 | 10.97 | 10.50 | 0.00 | 300.00 |
| Hamburger | 29792 | 7.01 | 6.81 | 8.80 | 0.00 | 300.00 | 29020 | 6.90 | 6.43 | 8.80 | 0.00 | 110.00 |
| White meat (poultry) | 29792 | 15.09 | 13.69 | 12.00 | 0.00 | 300.00 | 29020 | 15.16 | 14.23 | 12.00 | 0.00 | 375.00 |
| Blood based food | 7163 | 7.82 | 8.52 | 0.57 | 0.00 | 132.30 | 6910 | 7.77 | 8.68 | 0.57 | 0.00 | 132.30 |
| Liver, kidney | 7163 | 2.32 | 4.07 | 0.23 | 0.00 | 50.40 | 6910 | 2.20 | 4.01 | 0.23 | 0.00 | 52.50 |
| Lean fish (e.g. perch, bass, cod) | 29792 | 11.46 | 10.34 | 11.20 | 0.00 | 275.00 | 29020 | 11.38 | 10.02 | 11.20 | 0.00 | 275.00 |
| Fatty fish (e.g. herring, whitefish, salmon) | 29792 | 7.48 | 8.82 | 4.80 | 0.00 | 200.00 | 29020 | 7.47 | 8.99 | 4.80 | 0.00 | 300.00 |
| Shellfish (e.g. shrimps, scallops) | 7163 | 2.05 | 2.70 | 0.21 | 0.00 | 24.84 | 6910 | 2.05 | 2.98 | 0.21 | 0.00 | 69.00 |
| Salty fish | 29792 | 1.12 | 2.80 | 0.18 | 0.00 | 250.00 | 29020 | 1.07 | 2.34 | 0.18 | 0.00 | 70.00 |
| Smoked fish/meat | 29792 | 1.05 | 2.42 | 0.08 | 0.00 | 62.50 | 29020 | 1.05 | 2.37 | 0.08 | 0.00 | 62.50 |
| Ice cream | 29792 | 6.61 | 7.85 | 5.20 | 0.00 | 260.00 | 29020 | 6.53 | 7.37 | 5.20 | 0.00 | 162.50 |
| Sweets | 29792 | 5.68 | 7.03 | 3.36 | 0.00 | 160.00 | 29020 | 5.66 | 6.89 | 3.36 | 0.00 | 160.00 |
| Sugar, honey, marmelade, jam | 29792 | 10.92 | 15.05 | 4.76 | 0.00 | 115.00 | 29020 | 10.92 | 15.17 | 4.40 | 0.00 | 115.00 |
| Cookies, pastry | 29792 | 8.82 | 12.04 | 4.48 | 0.00 | 156.00 | 29020 | 8.62 | 11.66 | 4.48 | 0.00 | 184.00 |
| Chips, popcorn, salted nuts | 29792 | 2.70 | 3.32 | 2.16 | 0.00 | 87.50 | 29020 | 2.73 | 3.51 | 2.16 | 0.00 | 87.50 |
| Low fat milk (0,5%) | 29792 | 56.07 | 130.79 | 0.00 | 0.00 | 1040.00 | 29020 | 54.69 | 128.02 | 0.00 | 0.00 | 1040.00 |
| Milk, sour milk (1,5%) | 29792 | 139.22 | 170.83 | 68.40 | 0.00 | 1040.00 | 29020 | 138.15 | 169.66 | 68.40 | 0.00 | 1040.00 |
| Milk, sour milk (3%) | 29792 | 25.46 | 93.22 | 0.00 | 0.00 | 1400.00 | 29020 | 25.81 | 95.19 | 0.00 | 0.00 | 1400.00 |
| Sodas, soft drinks, juice | 29792 | 87.21 | 126.97 | 37.33 | 0.00 | 2108.00 | 29020 | 86.06 | 127.03 | 37.33 | 0.00 | 2022.00 |
| Brewed (filtered) coffee | 29792 | 307.72 | 205.51 | 375.00 | 0.00 | 800.00 | 29020 | 307.90 | 205.49 | 375.00 | 0.00 | 800.00 |
| Boiled coffee | 29792 | 98.47 | 175.20 | 0.45 | 0.00 | 800.00 | 29020 | 98.47 | 174.91 | 0.45 | 0.00 | 800.00 |
| Tea | 29792 | 152.87 | 199.17 | 90.00 | 0.00 | 1000.00 | 29020 | 150.72 | 197.45 | 90.00 | 0.00 | 1000.00 |
| Light beer | 29792 | 26.86 | 64.54 | 0.99 | 0.00 | 1320.00 | 29020 | 26.89 | 63.56 | 0.99 | 0.00 | 825.00 |
| Medium beer | 29792 | 21.63 | 45.22 | 0.99 | 0.00 | 875.00 | 29020 | 21.71 | 46.28 | 0.99 | 0.00 | 1400.00 |
| Strong beer | 29792 | 19.25 | 31.66 | 1.20 | 0.00 | 962.50 | 29020 | 19.39 | 34.04 | 1.20 | 0.00 | 1250.00 |
| Wine | 29792 | 18.72 | 25.96 | 18.00 | 0.00 | 687.50 | 29020 | 19.05 | 26.86 | 18.00 | 0.00 | 562.50 |

| | | | | | | | | | | | |
|---|---|---|---|---|---|---|---|---|---|---|---|
| Liquor, spirits | 29792 | 2.25 | 4.07 | 0.25 | 0.00 | 84.00 | 29020 | 2.33 | 4.83 | 0.25 | 0.00 | 210.00 |

**Supplementary Table 2**. Descriptive statistics of the categorical and ordinal lifestyle variables in training and testing sets

| Variables | Categories | N of observations in training set | N of observations in testing set |
|---|---|---|---|
| Gender | 1= Male | 16860 | 16572 |
| | 2= Female | 17374 | 16932 |
| Fasting status | 1= 8 hours or more | 29777 | 29148 |
| | 2= less than 8 hours | 887 | 831 |
| Educational level | 1= Elementary school + nine-year (compulsor | 5495 | 5376 |
| | 2= Folk high school equivalent to nine-year (compulsory) school + junior secondary school + girls' school + vocational (training) school | 6738 | 6554 |
| | 3= Folk high school equivalent to upper secor | 12346 | 12164 |
| | 4= University education/college | 9397 | 9136 |
| Work shifts/weekends | 1= No | 23892 | 23162 |
| | 2= Yes | 8473 | 8575 |
| Long-term sickness | 1= No | 26154 | 25474 |
| | 2= Yes | 5376 | 5289 |
| Permanent employment | 1= No | 8340 | 8334 |
| | 2= Yes | 21038 | 20283 |
| Self-employed | 1= No | 25988 | 25187 |
| | 2= Yes | 3017 | 3039 |
| Overall state of health compared to other your age | 1= Worse | 1144 | 1120 |
| | 2= About the same | 13724 | 13313 |
| | 3= Better | 1995 | 1982 |
| Overall state of health during the last year | 1= Poor | 402 | 325 |
| | 2= Fairly poor | 1525 | 1550 |
| | 3= Tolerably | 6393 | 6170 |
| | 4= Fairly good | 16015 | 15428 |
| | 5= Very good | 9284 | 9397 |
| Parents or siblings had a cerebral hemorrhage/thrombosis or cardiac infarction before the age of 60 | 1= No | 26493 | 25855 |
| | 2= Yes | 6544 | 6485 |
| Parents or siblings have diabetes | 1= No | 26357 | 25688 |
| | 2= Yes | 6723 | 6666 |
| Informed of having high blood pressure | 1= No | 26368 | 25656 |
| | 2= Yes | 7441 | 7398 |
| Self rate of overall health | 1= Poor | 202 | 223 |
| | 2= Fairly good | 2076 | 2010 |
| | 3= Good | 5170 | 5024 |
| | 4= Very good | 4951 | 4849 |
| | 5= Excellent | 2090 | 2196 |
| Self rate of overall health compared to a year ago | 1= Much worse than a year ago | 162 | 152 |
| | 2= A little worse than a year ago | 1535 | 1457 |
| | 3= About the same | 10463 | 10381 |
| | 4= A little better than a year ago | 1659 | 1615 |
| | 5= Much better than a year ago | 559 | 564 |
| Physical limitation to participate in steneous activities: running, lifting heavy objects, taking part in physically demanding sports | 1= No, not limited at all | 6158 | 6095 |
| | 2= Yes, a little limited | 5818 | 5730 |
| | 3= Yes, very limited | 2482 | 2450 |
| Physical limitation to participate in moderately demanding activities: moving a table, vacuuming, walking in the forest or gardening | 1= No, not limited at all | 11910 | 11755 |
| | 2= Yes, a little limited | 2209 | 2196 |
| | 3= Yes, very limited | 336 | 321 |
| Physical limitation to participate in moderately demanding activities: lifting or carrying grocery bags | 1= No, not limited at all | 11644 | 11548 |

| | | | |
|---|---|---|---|
| | 2= Yes, a little limited | 2323 | 2274 |
| | 3= Yes, very limited | 474 | 441 |
| Physical limitation to participate in moderately demanding activities: walking up several stairs | 1= No, not limited at all | 11946 | 11724 |
| | 2= Yes, a little limited | 2134 | 2138 |
| | 3= Yes, very limited | 377 | 394 |
| Physical limitation to participate in moderately demanding activities: bending down or kneeling | 1= No, not limited at all | 10815 | 10630 |
| | 2= Yes, a little limited | 3056 | 3036 |
| | 3= Yes, very limited | 571 | 577 |
| Physical limitation to participate in moderately demanding activities: walking more than 2 km | 1= No, not limited at all | 12398 | 12240 |
| | 2= Yes, a little limited | 1557 | 1520 |
| | 3= Yes, very limited | 508 | 503 |
| Physical limitation that reduced the normal time spent at work or in other activities during the last four weeks | 1= No | 12896 | 12661 |
| | 2= Yes | 1590 | 1604 |
| Physical limitation that made you do less than you wanted during the last four weeks | 1= No | 11619 | 11454 |
| | 2= Yes | 2850 | 2813 |
| Physical limitation that made you not being able to perform certain work tasks or other activities during the last four weeks | 1= No | 12268 | 12071 |
| | 2= Yes | 2145 | 2140 |
| Physical limitation that limited your ability to perform certain work tasks or other activities during the last four weeks | 1= No | 12455 | 12264 |
| | 2= Yes | 1986 | 1952 |
| Emotional problems that made you do less than you wanted during the last four weeks | 1= No | 12659 | 12491 |
| | 2= Yes | 1826 | 1776 |
| Extent to what your physical and emotional health disrupted your usual social life during the last four weeks | 1= Not al all | 11221 | 11015 |
| | 2= A little | 1846 | 1872 |
| | 3= Moderately | 991 | 979 |
| | 4= Much | 334 | 342 |
| | 5= Very much | 75 | 60 |
| Pain during the last four weeks | 1= None | 4748 | 4790 |
| | 2= Very little | 2902 | 2796 |
| | 3= Little | 2202 | 2167 |
| | 4= Moderate | 3659 | 3509 |
| | 5= Severe | 847 | 893 |
| | 6= Very severe | 100 | 87 |
| How much has the pain during the last four weeks disturbed your normal work? | 1= Not at all | 8018 | 8005 |
| | 2= A little | 3074 | 3035 |
| | 3= Moderately | 2333 | 2186 |
| | 4= Much | 818 | 823 |
| | 5= Very much | 176 | 168 |
| For how much of the time during the last four weeks have you felt really alert and strong? | 1= None of the time | 676 | 629 |
| | 2= A little of the time | 1177 | 1215 |
| | 3= Part of the time | 2126 | 2094 |
| | 4= Much of the time | 2545 | 2430 |
| | 5= Most of the time | 5979 | 5962 |
| | 6= All of the time | 1924 | 1889 |
| For how much of the time during the last four weeks have you felt very nervous? | 1= None of the time | 10756 | 10552 |
| | 2= A little of the time | 2702 | 2702 |
| | 3= Part of the time | 614 | 598 |
| | 4= Much of the time | 274 | 251 |
| | 5= Most of the time | 47 | 68 |
| | 6= All of the time | 54 | 77 |
| For how much of the time during the last four weeks have you felt so depressed that nothing could cheer you up? | 1= None of the time | 12106 | 11941 |
| | 2= A little of the time | 1556 | 1518 |
| | 3= Part of the time | 497 | 452 |
| | 4= Much of the time | 196 | 204 |

| | | | |
|---|---|---|---|
| | 5= Most of the time | 51 | 80 |
| | 6= All of the time | 57 | 66 |
| For how much of the time during the last four weeks have you felt calm and serene? | 1= None of the time | 272 | 295 |
| | 2= A little of the time | 698 | 712 |
| | 3= Part of the time | 1402 | 1379 |
| | 4= Much of the time | 2083 | 2032 |
| | 5= Most of the time | 6666 | 6584 |
| | 6= All of the time | 3325 | 3252 |
| For how much of the time during the last four weeks have you felt full of energy? | 1= None of the time | 737 | 728 |
| | 2= A little of the time | 1441 | 1510 |
| | 3= Part of the time | 2517 | 2404 |
| | 4= Much of the time | 3404 | 3432 |
| | 5= Most of the time | 4963 | 4829 |
| | 6= All of the time | 1367 | 1330 |
| For how much of the time during the last four weeks have you felt gloomy and sad? | 1= None of the time | 8789 | 8707 |
| | 2= A little of the time | 3999 | 3905 |
| | 3= Part of the time | 1035 | 1052 |
| | 4= Much of the time | 410 | 352 |
| | 5= Most of the time | 124 | 136 |
| | 6= All of the time | 76 | 83 |
| For how much of the time during the last four weeks have you felt worn out? | 1= None of the time | 5684 | 5571 |
| | 2= A little of the time | 4712 | 4669 |
| | 3= Part of the time | 2171 | 2110 |
| | 4= Much of the time | 1149 | 1149 |
| | 5= Most of the time | 507 | 479 |
| | 6= All of the time | 201 | 225 |
| For how much of the time during the last four weeks have you felt happy? | 1= None of the time | 244 | 284 |
| | 2= A little of the time | 986 | 948 |
| | 3= Part of the time | 1996 | 1940 |
| | 4= Much of the time | 2695 | 2669 |
| | 5= Most of the time | 6477 | 6377 |
| | 6= All of the time | 2054 | 2022 |
| For how much of the time during the last four weeks have you felt tired? | 1= None of the time | 2019 | 1948 |
| | 2= A little of the time | 5665 | 5598 |
| | 3= Part of the time | 3249 | 3190 |
| | 4= Much of the time | 2205 | 2199 |
| | 5= Most of the time | 906 | 901 |
| | 6= All of the time | 422 | 413 |
| For how much of the time during the last four weeks has your physical health or your emotional problems limited your ability to interact with others? | 1= None of the time | 10720 | 10496 |
| | 2= A little of the time | 2094 | 2150 |
| | 3= Part of the time | 1203 | 1184 |
| | 4= Most of the time | 294 | 272 |
| | 5= All of the time | 69 | 66 |
| Get sick more often than other people | 1= Not at all true | 10763 | 10574 |
| | 2= Not very true | 2380 | 2392 |
| | 3= Unsure | 845 | 844 |
| | 4= Mostly true | 361 | 364 |
| | 5= Altogether true | 87 | 62 |
| As healthy as anyone | 1= Not at all true | 635 | 622 |
| | 2= Not very true | 1059 | 1020 |
| | 3= Unsure | 1430 | 1499 |
| | 4= Mostly true | 4560 | 4585 |
| | 5= Totally true | 6707 | 6471 |

| | | | |
|---|---|---|---|
| Worsen in health in the future | 1= Not at all true | 5124 | 5129 |
| | 2= Not very true | 3265 | 3190 |
| | 3= Unsure | 4481 | 4315 |
| | 4= Mostly true | 1247 | 1299 |
| | 5= Totally true | 268 | 259 |
| Excellent health | 1= Not at all true | 720 | 734 |
| | 2= Not very true | 1577 | 1512 |
| | 3= Unsure | 1659 | 1665 |
| | 4= Mostly true | 7041 | 6666 |
| | 5= Totally true | 3436 | 3665 |
| Satisfaction with home and family situation | 1= Very poor … 7= Excellent | 83 | 94 |
| | | 248 | 269 |
| | | 609 | 615 |
| | | 1292 | 1285 |
| | | 2814 | 2720 |
| | | 6394 | 6224 |
| | | 13205 | 13015 |
| Satisfaction with accomodation | 1= Very poor … 7= Excellent | 40 | 36 |
| | | 110 | 117 |
| | | 353 | 380 |
| | | 987 | 1030 |
| | | 2455 | 2456 |
| | | 6051 | 5921 |
| | | 14672 | 14302 |
| Satisfaction with work situation | 1= Very poor … 7= Excellent | 806 | 857 |
| | | 923 | 910 |
| | | 1618 | 1664 |
| | | 3330 | 3245 |
| | | 5453 | 5295 |
| | | 6401 | 6082 |
| | | 5814 | 5851 |
| Satisfaction with economy | 1= Very poor … 7= Excellent | 321 | 375 |
| | | 633 | 617 |
| | | 1235 | 1257 |
| | | 2860 | 2900 |
| | | 5031 | 4988 |
| | | 7205 | 6809 |
| | | 7362 | 7265 |
| Satisfaction with leisure time | 1= Very poor … 7= Excellent | 193 | 207 |
| | | 530 | 542 |
| | | 1121 | 1125 |
| | | 2407 | 2404 |
| | | 4500 | 4462 |
| | | 6955 | 6847 |
| | | 8906 | 8594 |
| Hearing status | 1= Very poor … 7= Excellent | 162 | 180 |
| | | 630 | 579 |
| | | 1788 | 1895 |
| | | 2779 | 2703 |
| | | 4933 | 4916 |
| | | 6676 | 6559 |
| | | 7699 | 7400 |
| Vision status | 1= Very poor … 7= Excellent | 114 | 133 |

| | | | |
|---|---|---|---|
| | | 541 | 536 |
| | | 2365 | 2492 |
| | | 5221 | 5117 |
| | | 7479 | 7383 |
| | | 5833 | 5454 |
| | | 3090 | 3077 |
| Memory status | 1= Very poor … 7= Excellent | 112 | 111 |
| | | 375 | 351 |
| | | 1190 | 1267 |
| | | 3103 | 3097 |
| | | 6454 | 6225 |
| | | 8609 | 8352 |
| | | 4761 | 4754 |
| Fitness status | 1= Very poor … 7= Excellent | 431 | 449 |
| | | 1138 | 1130 |
| | | 2734 | 2843 |
| | | 5702 | 5682 |
| | | 7426 | 7304 |
| | | 5060 | 4661 |
| | | 2144 | 2128 |
| Appetite status | 1= Very poor … 7= Excellent | 23 | 7 |
| | | 71 | 57 |
| | | 257 | 279 |
| | | 1080 | 1101 |
| | | 2598 | 2578 |
| | | 6349 | 6189 |
| | | 14267 | 14004 |
| Mood status | 1= Very poor … 7= Excellent | 54 | 45 |
| | | 270 | 291 |
| | | 930 | 871 |
| | | 2592 | 2545 |
| | | 5376 | 5329 |
| | | 8611 | 8393 |
| | | 6797 | 6723 |
| Energy status | 1= Very poor … 7= Excellent | 229 | 232 |
| | | 809 | 828 |
| | | 1847 | 1776 |
| | | 3988 | 3916 |
| | | 7052 | 6963 |
| | | 7152 | 6819 |
| | | 3545 | 3637 |
| Patience status | 1= Very poor … 7= Excellent | 106 | 108 |
| | | 439 | 432 |
| | | 1305 | 1292 |
| | | 3073 | 3071 |
| | | 6189 | 6164 |
| | | 8355 | 7897 |
| | | 5163 | 5240 |
| Confidence status | 1= Very poor … 7= Excellent | 122 | 131 |
| | | 408 | 422 |
| | | 1010 | 1000 |
| | | 2721 | 2786 |
| | | 5957 | 5837 |

| | | | |
|---|---|---|---|
| | | 8647 | 8431 |
| | | 5758 | 5574 |
| Sleep status | 1= Very poor … 7= Excellent | 505 | 508 |
| | | 1083 | 1144 |
| | | 2061 | 2038 |
| | | 3009 | 3010 |
| | | 4603 | 4392 |
| | | 5903 | 5583 |
| | | 7501 | 7567 |
| Do you feel important and appreciated outside your home? | 1= Very poor … 7= Excellent | 118 | 128 |
| | | 266 | 255 |
| | | 636 | 636 |
| | | 2324 | 2377 |
| | | 6037 | 5965 |
| | | 9476 | 9243 |
| | | 5799 | 5631 |
| Do you feel important and appreciated in your home? | 1= Very poor … 7= Excellent | 99 | 111 |
| | | 172 | 148 |
| | | 406 | 384 |
| | | 1395 | 1335 |
| | | 3206 | 3087 |
| | | 7831 | 7689 |
| | | 11379 | 11288 |
| Number of social contacts with the same interests as you | 1= No one | 212 | 218 |
| | 2= 1-2 persons | 2038 | 2027 |
| | 3= 3-5 persons | 8069 | 8009 |
| | 4= 6-10 persons | 9771 | 9448 |
| | 5= 11-15 persons | 4039 | 4010 |
| | 6= >15 persons | 9775 | 9443 |
| Number of social interactions during a normal week | 1= No one | 39 | 37 |
| | 2= 1-2 persons | 1209 | 1233 |
| | 3= 3-5 persons | 5525 | 5436 |
| | 4= 6-10 persons | 8094 | 7914 |
| | 5= 11-15 persons | 4963 | 4818 |
| | 6= >15 persons | 13774 | 13403 |
| Would you say that the number of people that you meet in your everyday life is enough or would you like to meet more or fewer people? | 1= Fewer | 453 | 427 |
| | 2= Sufficiently enough | 29080 | 28212 |
| | 3= More | 4424 | 4581 |
| Number of friends that can come to your home at any time and feel at home | 1= No one | 652 | 643 |
| | 2= 1-2 persons | 3711 | 3667 |
| | 3= 3-5 persons | 11193 | 10974 |
| | 4= 6-10 persons | 10400 | 10209 |
| | 5= 11-15 persons | 3440 | 3272 |
| | 6= >15 persons | 4085 | 3982 |
| Number of people with whom you can speak openly | 1= No one | 525 | 509 |
| | 2= 1-2 persons | 5883 | 5881 |
| | 3= 3-5 persons | 13736 | 13541 |
| | 4= 6-10 persons | 8800 | 8617 |
| | 5= 11-15 persons | 2543 | 2340 |
| | 6= >15 persons | 2457 | 2307 |
| Support from others | 1= No | 1180 | 1236 |
| | 2= Yes, but I do not need it | 3908 | 3896 |
| | 3= Yes | 28901 | 28111 |

| | | | |
|---|---|---|---|
| Close relationship with anyone | 1= No | 737 | 752 |
| | 2= Not sure | 3007 | 3039 |
| | 3= Yes | 29824 | 29009 |
| Receive hugs to comfort and support you | 1= No | 5658 | 5449 |
| | 2= Yes | 27701 | 27138 |
| People to ask for help apart from the ones at home | 1= No | 3431 | 3435 |
| | 2= Yes | 30431 | 29680 |
| Participation in associations or voluntary organisations | 1= No | 12427 | 12265 |
| | 2= Yes | 20978 | 20426 |
| Frequency of engaging in clubs, associations or study circles | 1= 1-2 times per year | 3930 | 3887 |
| | 2= 1-2 times per month | 7107 | 6881 |
| | 3= 1-2 times per week | 9905 | 9628 |
| | 4= Every day | 554 | 536 |
| Participation in sports or physical exercise associations | 1= No | 5001 | 4887 |
| | 2= Yes | 5738 | 5497 |
| Participation in study circles | 1= No | 9158 | 8868 |
| | 2= Yes | 1581 | 1516 |
| Participation in other association | 1= No | 5838 | 5533 |
| | 2= Yes | 4901 | 4851 |
| High physical demand from job | 1= No as good as never | 9863 | 9633 |
| | 2= No rarely | 7763 | 7631 |
| | 3= Yes sometimes | 11421 | 11149 |
| | 4= Yes often | 4514 | 4470 |
| Job demands to work very fast | 1= No as good as never | 1838 | 1696 |
| | 2= No rarely | 6369 | 6272 |
| | 3= Yes sometimes | 18968 | 18541 |
| | 4= Yes often | 6305 | 6304 |
| High mental demand from job | 1= No as good as never | 5516 | 5275 |
| | 2= No rarely | 16462 | 16135 |
| | 3= Yes sometimes | 7981 | 7973 |
| | 4= Yes often | 3271 | 3206 |
| Enough time for job assignments | 1= No as good as never | 1165 | 1167 |
| | 2= No rarely | 6125 | 6010 |
| | 3= Yes sometimes | 13156 | 12890 |
| | 4= Yes often | 12886 | 12594 |
| Contradictory demands in job | 1= No as good as never | 4884 | 4736 |
| | 2= No rarely | 12016 | 11767 |
| | 3= Yes sometimes | 13171 | 12974 |
| | 4= Yes often | 3192 | 3070 |
| Learn new things at job | 1= No as good as never | 880 | 845 |
| | 2= No rarely | 3841 | 3823 |
| | 3= Yes sometimes | 18057 | 17712 |
| | 4= Yes often | 10642 | 10355 |
| Skill demand from job | 1= No as good as never | 453 | 445 |
| | 2= No rarely | 2038 | 2003 |
| | 3= Yes sometimes | 14604 | 14121 |
| | 4= Yes often | 16299 | 16115 |
| Ingenuity or creativity demand from job | 1= No as good as never | 545 | 514 |
| | 2= No rarely | 2686 | 2680 |
| | 3= Yes sometimes | 14977 | 14638 |
| | 4= Yes often | 15123 | 14825 |
| Repetitive job | 1= No as good as never | 1067 | 1114 |
| | 2= No rarely | 6628 | 6496 |

| | | | |
|---|---|---|---|
| | 3= Yes sometimes | 12446 | 12142 |
| | 4= Yes often | 13288 | 12993 |
| Control over planning and execution of the workday | 1= No as good as never | 638 | 622 |
| | 2= No rarely | 2648 | 2598 |
| | 3= Yes sometimes | 11250 | 11099 |
| | 4= Yes often | 18966 | 18482 |
| Control over own work assignment | 1= No as good as never | 1819 | 1763 |
| | 2= No rarely | 6444 | 6570 |
| | 3= Yes sometimes | 13167 | 12673 |
| | 4= Yes often | 12039 | 11785 |
| Possibility to speak with colleagues during breaks | 1= No, I do not have breaks with colleagues | 3451 | 3458 |
| | 2= No, I do not have breaks | 762 | 682 |
| | 3= Yes, most of the time | 11698 | 11548 |
| | 4= Yes, always | 16310 | 15842 |
| Possibility to leave your work for a while to speak with a colleague | 1= No, it is totally impossible | 1755 | 1725 |
| | 2= Only for urgent matters | 3127 | 3099 |
| | 3= Yes, sometimes | 9672 | 9428 |
| | 4= Yes, most of the time | 17463 | 17073 |
| Frequent social contacts with colleagues during work | 1= Seldom or never | 991 | 1009 |
| | 2= No, I mostly work alone | 2820 | 2865 |
| | 3= One or a few times per month | 1325 | 1240 |
| | 4= Yes, a lot | 26861 | 26175 |
| Frequncy of social contacts with colleagues during leisure time | 1= Seldom or never | 8243 | 8119 |
| | 2= One or more times per year | 12223 | 11843 |
| | 3= One or more times per month | 7842 | 7762 |
| | 4= One or more times per week | 3146 | 3037 |
| Last time a colleague visited you at home | 1= I have never been visited by a colleague | 3549 | 3557 |
| | 2= More than a year ago | 4985 | 4866 |
| | 3= One to twelve months ago | 10626 | 10333 |
| | 4= One to four weeks ago | 12712 | 12437 |
| Sedentary or standing work | 1= No | 23935 | 23220 |
| | 2= Yes | 8643 | 8636 |
| Light but partly physically active work | 1= No | 26366 | 25977 |
| | 2= Yes | 6212 | 5879 |
| Light and physically active work | 1= No | 24694 | 24223 |
| | 2= Yes | 7884 | 7633 |
| Sometimes physically straining work | 1= No | 23167 | 22626 |
| | 2= Yes | 9411 | 9230 |
| Frequency of walking during leisure time | 1= Never | 1331 | 1355 |
| | 2= 1-2 times a month | 4769 | 4535 |
| | 3= 3-4 times a month | 6254 | 6409 |
| | 4= 2-3 times a week | 12566 | 12187 |
| | 5= Every day | 7448 | 7149 |
| Frequency of cycling during leisure time | 1= Never | 5719 | 5509 |
| | 2= 1-2 times a month | 7124 | 6955 |
| | 3= 3-4 times a month | 4964 | 5017 |
| | 4= 2-3 times a week | 6094 | 5902 |
| | 5= Every day | 3612 | 3353 |
| Frequency of dancing during leisure time | 1= Never | 12173 | 11787 |
| | 2= 1-2 times a month | 5567 | 5498 |
| | 3= 3-4 times a month | 1308 | 1199 |
| | 4= 2-3 times a week | 461 | 400 |
| | 5= Every day | 5 | 3 |

| | | | |
|---|---|---|---|
| Frequency of shoverling snow during leisure time | 1= Never | 2424 | 2381 |
| | 2= 1-2 times a month | 4667 | 4568 |
| | 3= 3-4 times a month | 5829 | 5644 |
| | 4= 2-3 times a week | 6911 | 6801 |
| | 5= Every day | 1230 | 1110 |
| Frequency of gardening during leisure time | 1= Never | 2099 | 2001 |
| | 2= 1-2 times a month | 4054 | 4034 |
| | 3= 3-4 times a month | 5582 | 5399 |
| | 4= 2-3 times a week | 8282 | 7964 |
| | 5= Every day | 990 | 914 |
| Frequency of hunting or fishing during leisure time | 1= Never | 7560 | 7262 |
| | 2= 1-2 times a month | 6158 | 6074 |
| | 3= 3-4 times a month | 4421 | 4357 |
| | 4= 2-3 times a week | 2146 | 2026 |
| | 5= Every day | 130 | 107 |
| Frequency of picking berries or mushrooms during leisure time | 1= Never | 3433 | 3373 |
| | 2= 1-2 times a month | 8563 | 8344 |
| | 3= 3-4 times a month | 5473 | 5336 |
| | 4= 2-3 times a week | 3079 | 2939 |
| | 5= Every day | 294 | 275 |
| Changed everyday exercise during the last year | 1= Decreased a lot | 1474 | 1477 |
| | 2= Decreased somewhat | 3830 | 3871 |
| | 3= As before | 12106 | 11834 |
| | 4= Increased somewhat | 4113 | 3801 |
| | 5= Increased a lot | 666 | 627 |
| Everyday exercise satisfaction | 1= Not at all | 2852 | 2811 |
| | 2= Rather poorly | 4754 | 4702 |
| | 3= Partly | 10664 | 10321 |
| | 4= Completely | 3860 | 3704 |
| Exercise during the last three months | 1= Never | 13129 | 12896 |
| | 2= Every now and then- not regularly | 7747 | 7761 |
| | 3= 1-2 times/week | 5693 | 5308 |
| | 4= 2-3 times/week | 4703 | 4572 |
| | 5= More than 3 times/week | 1963 | 1948 |
| If you exercise, change in exercise habits during the last year | 1= Decreased a lot | 1720 | 1828 |
| | 2= Decreased somewhat | 3453 | 3383 |
| | 3= As before | 10089 | 9673 |
| | 4= Increased somewhat | 3205 | 3125 |
| | 5= Increased a lot | 587 | 553 |
| Amount of exercise during the last 12 months | 1= Sedentary leisure time | 915 | 926 |
| | 2= Moderate exercise in leisure time | 5932 | 5855 |
| | 3= Moderate, regular exercise in leisure time | 3024 | 2941 |
| | 4= Regular exercise | 1570 | 1539 |
| Time spent in a week in moderately steneous activities | 1= No time at all | 271 | 290 |
| | 2= Not more than 1 hour per week | 1247 | 1185 |
| | 3= 1-3 hours per week | 3942 | 3837 |
| | 4= More than 3 hours, but less than 5 hours pe | 3000 | 3016 |
| | 5= 5 hours per week or more | 2635 | 2572 |
| Risk of sleeping while sitting and reading | 1= None | 5287 | 5183 |
| | 2= Little | 3449 | 3465 |
| | 3= Moderate | 1914 | 1893 |
| | 4= Big | 569 | 535 |
| Risk of sleeping while watching TV | 1= None | 1708 | 1763 |

| | | | |
|---|---|---|---|
| | 2= Little | 3588 | 3577 |
| | 3= Moderate | 3910 | 3857 |
| | 4= Big | 2136 | 1997 |
| Risk of sleeping while sitting inactive in a public place | 1= None | 6325 | 6344 |
| | 2= Little | 3537 | 3454 |
| | 3= Moderate | 1172 | 1106 |
| | 4= Big | 230 | 218 |
| Risk of sleeping as a passenger in a car for one hour without break | 1= None | 4694 | 4586 |
| | 2= Little | 3431 | 3466 |
| | 3= Moderate | 2094 | 2024 |
| | 4= Big | 1007 | 1001 |
| Risk of sleeping while lying down resting in the afternoon | 1= None | 1029 | 1097 |
| | 2= Little | 2291 | 2271 |
| | 3= Moderate | 3547 | 3517 |
| | 4= Big | 4427 | 4278 |
| Risk of sleeping while sitting still after having lunch | 1= None | 6938 | 6972 |
| | 2= Little | 2898 | 2759 |
| | 3= Moderate | 1225 | 1194 |
| | 4= Big | 241 | 245 |
| Snore during sleep | 1= No, never | 827 | 837 |
| | 2= No, almost never | 2209 | 2177 |
| | 3= Yes, sometimes | 5413 | 5331 |
| | 4= Yes, almost always | 1475 | 1457 |
| | 5= Yes, always | 644 | 665 |
| Breath-holds during sleep | 1= No, never | 5721 | 5687 |
| | 2= No, almost never | 492 | 473 |
| | 3= Yes, sometimes | 1335 | 1266 |
| | 4= Yes, almost always | 203 | 167 |
| | 5= Yes, always | 187 | 183 |
| Teetotaler | 1= No | 19574 | 19155 |
| | 2= Yes | 2237 | 2058 |
| Feel the need to reduce alcohol consumption | 1= No | 26631 | 25914 |
| | 2= Yes | 3836 | 3919 |
| Feel uneasy or guilty because of your way of drinking | 1= No | 17367 | 16930 |
| | 2= Yes | 2217 | 2222 |
| Frequency of alcohol consumption | 1= Never | 1011 | 1044 |
| | 2= 1 time/month or more seldom | 3149 | 3092 |
| | 3= 2-4 times/month | 5687 | 5557 |
| | 4= 2-3 times/week | 1526 | 1522 |
| | 5= 4 times/week or more | 111 | 123 |
| Amount of alcohol drunk in a day | 1= 0-2 glasses | 5954 | 5777 |
| | 2= 3-4 glasses | 3969 | 3956 |
| | 3= 5-6 glasses | 873 | 894 |
| | 4= 7-9 glasses | 145 | 121 |
| | 5= 10 glasses or more | 26 | 18 |
| Frequency of drinking six or more glasses at the same occasion | 1= Never | 6448 | 6226 |
| | 2= More seldom than once a month | 3662 | 3716 |
| | 3= Every month | 854 | 825 |
| | 4= Every week | 207 | 208 |
| | 5= Daily or almost daily | 2 | 5 |
| Times during last year that you felt guilty because of your drinking | 1= Never | 10102 | 9865 |
| | 2= More seldom than once a month | 966 | 996 |
| | 3= Every month | 64 | 65 |

| | | | |
|---|---|---|---|
| | 4= Every week | 20 | 19 |
| | 5= Daily or almost daily | 8 | 8 |
| Number of cigarretes smoked per day (in groups) | 1=0 | 21290 | 20834 |
| | 2= 1-4 | 1063 | 1126 |
| | 3= 5-14 | 3274 | 3254 |
| | 4= 15-24 | 1500 | 1461 |
| | 5= >25 | 112 | 126 |
| Number of snuff boxes per week | 1= 0 | 27098 | 26407 |
| | 2= Less than 2 | 1854 | 1934 |
| | 3= 2 to 4 | 2331 | 2329 |
| | 4= More than 4 but less than 7 | 1103 | 1066 |
| | 5= 7 or more | 340 | 302 |
| Cambridge physical activity index | 1= Inactive | 4712 | 4864 |
| | 2= Moderatively inactive | 10193 | 9707 |
| | 3= Moderatively active | 9346 | 9163 |
| | 4= Active | 7918 | 7718 |
| FFQ version | AC00= Optically readable; 66 food items | 3434 | 3219 |
| | AC03= Optically readable; 66 food items | 3067 | 3028 |
| | AC05= Optically readable; 66 food items | 8324 | 8304 |
| | AC11= Optically readable; 66 food items | 3198 | 3056 |
| | AC4= Optically readable; 64 food items | 2606 | 2628 |
| | AC5= Optically readable; 65 food items | 1481 | 1451 |
| | AC6= Optically readable; 65 food items | 2710 | 2712 |
| | APRI= Manually readable; 84 food items | 1912 | 1841 |
| | BAS6= Optically readable; 84 food items | 1180 | 1099 |
| | BASG= Optically readable; 84 food items | 2254 | 2111 |
| | BASN= Optically readable; 84 food items | 2890 | 2861 |
| Average portion size of potatoes/rice/pasta based on photographic illustration of four sizes (smallest to largest) | 1= A | 1630 | 1567 |
| | 2= B | 12065 | 11683 |
| | 3= C | 13500 | 13149 |
| | 4= D | 2597 | 2621 |
| Average portion size of meat/fish based on photographic illustration of four sizes (smallest to largest) | 1= A | 1179 | 1114 |
| | 2= B | 13208 | 12836 |
| | 3= C | 13332 | 12975 |
| | 4= D | 2073 | 2095 |
| Average portion size of vegetables based on photographic illustration of four sizes (smallest to largest) | 1= A | 4164 | 4081 |
| | 2= B | 8451 | 8210 |
| | 3= C | 10195 | 9760 |
| | 4= D | 6982 | 6969 |
| Eat breakfast from 2000 | 1= No | 1451 | 1546 |
| | 2= Yes | 15683 | 15183 |
| Marital status | 1= Single | 3175 | 3185 |
| | 2= Married/partner | 28003 | 27300 |
| | 3= Divorced/separated | 2394 | 2398 |
| | 4= Widow/widower | 426 | 373 |
| Cohabitation | 1= Live alone | 3393 | 3429 |
| | 2= Only one adult (spouse, partner) | 11904 | 11713 |
| | 3= Only children | 1559 | 1533 |
| | 4= Adult and children | 16229 | 15706 |
| | 5= Other/others | 464 | 432 |
| Smoking status | 1= Non smokers | 16764 | 16287 |
| | 2= Smokers | 4910 | 5005 |
| | 3= Former smokers | 7477 | 7143 |

| | | | |
|---|---|---|---|
| | 4= Former occasional smokers | 3321 | 3359 |
| Snuff status | 1= Non-snuff users | 23245 | 22703 |
| | 2= Snuff users | 5628 | 5631 |
| | 3= Former snuff users | 3853 | 3704 |
| Breakfast habits | 1= Not breakfast at all | 1008 | 1078 |
| | 2= Only coffee/tea for breakfast | 913 | 925 |
| | 3= Coffee/tea and wheat buns and rusk for bre | 606 | 525 |
| | 4= Porridge w/o sandwich for breakfast | 3245 | 3248 |
| | 5= Gruel w/o sandwich for breakfast | 457 | 448 |
| Travel to work | 1= Passive travel (by car or bus) | 18672 | 18337 |
| | 2= Walk | 2907 | 2927 |
| | 3= Cycle | 6734 | 6381 |
| | 4= Irregular travel mode to work | 3004 | 2956 |

**Supplementary Table 3**. Linear mixed model association results for BMI

| | | Training set | | | | | | Testing set | | | | | | Metanalysis | | | | | R2 | | |
|---|---|---|---|---|---|---|---|---|---|---|---|---|---|---|---|---|---|---|---|---|---|
| **Description** | **Group** | **N** | **Effect estimate** | **S.E.** | **p-value** | **p-value$_{FDR}$** | **p-value rank** | **N** | **Effect estimate** | **S.E.** | **p-value** | **p-value$_{FDR}$** | **p-value rank** | **Effect estimate** | **S.E.** | **p-value** | **$I^2$** | **Q p-value** | **marginal R2** | **conditional R2** | **R2 rank** |
| Fitness status | Physical activity | 24593 | -0.86 | 0.02 | <2.22E-308 | **<2.22E-308** | 1 | 24154 | -0.87 | 0.02 | <2.22E-308 | **<2.22E-308** | 1 | -0.86 | 0.01 | <2.22E-308 | 0.00 | 7.41E-01 | 0.08 | 0.84 | 1 |
| Physical limitation to participate in moderately demanding activities: bending down or kneeling | General health | 14429 | 0.95 | 0.03 | 4.57E-105 | **6.53E-103** | 2 | 14232 | 1.02 | 0.03 | 1.68E-108 | **1.59E-106** | 3 | 0.99 | 0.02 | 1.34E-211 | 0.50 | 1.56E-01 | 0.07 | 0.80 | 4 |
| Physical limitation to participate in moderately demanding activities: walking up several stairs | General health | 14444 | 0.91 | 0.03 | 2.58E-98 | **2.46E-96** | 3 | 14245 | 0.88 | 0.03 | 3.16E-89 | **1.79E-87** | 5 | 0.89 | 0.02 | 9.34E-186 | 0.00 | 5.02E-01 | 0.06 | 0.81 | 13 |
| Physical limitation to participate in steneous activities: running, lifting heavy objects, taking part in physically demanding sports | General health | 14445 | 0.86 | 0.03 | 4.58E-89 | **3.27E-87** | 4 | 14264 | 0.91 | 0.03 | 5.15E-92 | **3.66E-90** | 4 | 0.88 | 0.02 | 3.96E-179 | 0.21 | 2.60E-01 | 0.06 | 0.80 | 22 |
| Self rate of overall health | General health | 14476 | -0.81 | 0.03 | 6.97E-85 | **3.99E-83** | 5 | 14289 | -0.85 | 0.03 | 7.81E-87 | **3.70E-85** | 6 | -0.83 | 0.02 | 8.47E-170 | 0.00 | 3.81E-01 | 0.06 | 0.81 | 39 |
| Everyday exercise satisfaction | Physical activity | 21994 | -0.36 | 0.02 | 7.12E-78 | **3.40E-76** | 6 | 21404 | -0.34 | 0.02 | 3.08E-68 | **7.95E-67** | 11 | -0.35 | 0.01 | 4.48E-144 | 0.00 | 4.50E-01 | 0.06 | 0.83 | 10 |
| Physical limitation to participate in moderately demanding activities: walking more than 2 km | General health | 14450 | 0.74 | 0.03 | 2.61E-75 | **1.07E-73** | 7 | 14252 | 0.79 | 0.03 | 6.17E-79 | **2.50E-77** | 7 | 0.77 | 0.02 | 2.54E-152 | 0.25 | 2.47E-01 | 0.05 | 0.81 | 129 |
| Overall state of health during the last year | General health | 33471 | -0.26 | 0.01 | 3.92E-72 | **1.40E-70** | 8 | 32722 | -0.26 | 0.01 | 1.75E-69 | **5.51E-68** | 9 | -0.26 | 0.01 | 1.08E-139 | 0.00 | 9.58E-01 | 0.06 | 0.83 | 9 |
| Exercise during the last three months | Physical activity | 33086 | -0.26 | 0.01 | 1.16E-69 | **3.70E-68** | 9 | 32342 | -0.24 | 0.02 | 1.17E-56 | **2.55E-55** | 13 | -0.25 | 0.01 | 3.09E-124 | 0.00 | 3.40E-01 | 0.06 | 0.83 | 11 |
| Informed of having high blood pressure | General health | 33649 | 0.67 | 0.04 | 1.74E-69 | **4.99E-68** | 10 | 32899 | 0.67 | 0.04 | 6.69E-69 | **1.90E-67** | 10 | 0.67 | 0.03 | 1.82E-136 | 0.00 | 9.46E-01 | 0.06 | 0.83 | 7 |
| Excellent health | General health | 14421 | -0.69 | 0.03 | 2.48E-68 | **6.46E-67** | 11 | 14231 | -0.74 | 0.03 | 8.30E-72 | **2.95E-70** | 8 | -0.72 | 0.02 | 3.24E-138 | 0.00 | 3.21E-01 | 0.05 | 0.80 | 130 |
| Energy status | Psychosocial | 24582 | -0.32 | 0.02 | 2.63E-57 | **6.27E-56** | 12 | 24128 | -0.31 | 0.02 | 1.74E-52 | **3.29E-51** | 15 | -0.32 | 0.01 | 7.17E-108 | 0.00 | 6.01E-01 | 0.04 | 0.85 | 134 |
| Cambridge physical activity index | Physical activity | 32023 | -0.21 | 0.01 | 8.81E-46 | **1.94E-44** | 13 | 31315 | -0.18 | 0.02 | 1.28E-31 | **1.45E-30** | 25 | -0.19 | 0.01 | 4.65E-75 | 0.62 | 1.03E-01 | 0.06 | 0.83 | 15 |
| Overall state of health compared to other your age | General health | 16780 | -0.38 | 0.03 | 3.21E-45 | **6.56E-44** | 14 | 16341 | -0.40 | 0.03 | 4.23E-51 | **7.51E-50** | 16 | -0.39 | 0.02 | 1.67E-94 | 0.00 | 5.54E-01 | 0.06 | 0.83 | 29 |
| Pain during the last four weeks | General health | 14444 | 0.52 | 0.03 | 1.35E-43 | **2.57E-42** | 15 | 14229 | 0.54 | 0.03 | 1.56E-44 | **2.46E-43** | 18 | 0.53 | 0.02 | 2.46E-86 | 0.00 | 6.72E-01 | 0.03 | 0.80 | 142 |
| How much has the pain during the last four weeks disturbed your normal work? | General health | 14405 | 0.50 | 0.03 | 7.32E-41 | **1.31E-39** | 16 | 14205 | 0.52 | 0.03 | 1.47E-41 | **2.20E-40** | 19 | 0.51 | 0.02 | 1.23E-80 | 0.00 | 6.86E-01 | 0.03 | 0.80 | 144 |
| As healthy as anyone | General health | 14379 | -0.48 | 0.03 | 1.71E-39 | **2.88E-38** | 17 | 14185 | -0.51 | 0.03 | 5.96E-41 | **8.47E-40** | 20 | -0.49 | 0.02 | 1.23E-78 | 0.00 | 5.33E-01 | 0.03 | 0.80 | 145 |
| Physical limitation to participate in moderately demanding activities: moving a table, vacuuming, walking in the forest or gardening | General health | 14442 | 0.48 | 0.03 | 1.12E-38 | **1.78E-37** | 18 | 14261 | 0.61 | 0.03 | 9.85E-54 | **2.00E-52** | 14 | 0.54 | 0.02 | 3.18E-90 | 0.87 | 5.26E-03 | 0.03 | 0.81 | 136 |
| For how much of the time during the last four weeks have you felt really alert and strong? | General health | 14414 | -0.47 | 0.03 | 1.10E-36 | **1.66E-35** | 19 | 14206 | -0.48 | 0.03 | 1.72E-36 | **2.22E-35** | 22 | -0.47 | 0.02 | 1.99E-71 | 0.00 | 8.46E-01 | 0.03 | 0.81 | 146 |
| Travel to work: Cycle to work vs passive travel to work | Physical activity | 25302 | -0.58 | 0.05 | 5.38E-36 | **7.69E-35** | 20 | 24613 | -0.58 | 0.05 | 1.57E-35 | **1.94E-34** | 23 | -0.58 | 0.03 | 9.40E-70 | 0.00 | 9.65E-01 | 0.06 | 0.83 | 6 |
| Physical limitation that made you do less than you wanted during the last four weeks | General health | 14456 | 1.05 | 0.08 | 1.93E-31 | **2.62E-30** | 21 | 14256 | 1.22 | 0.09 | 1.46E-37 | **1.97E-36** | 21 | 1.13 | 0.06 | 4.32E-67 | 0.50 | 1.57E-01 | 0.03 | 0.80 | 147 |
| For how much of the time during the last four weeks have you felt full of energy? | General health | 14416 | -0.42 | 0.03 | 8.67E-31 | **1.13E-29** | 22 | 14220 | -0.37 | 0.03 | 2.46E-24 | **1.79E-23** | 39 | -0.40 | 0.02 | 2.17E-53 | 0.00 | 3.39E-01 | 0.02 | 0.81 | 151 |
| Average portion size of meat/fish based on photographic illustration of four sizes (smallest to largest) | Food | 29676 | 0.19 | 0.02 | 2.34E-30 | **2.79E-29** | 23 | 28910 | 0.24 | 0.02 | 8.09E-45 | **1.35E-43** | 17 | 0.21 | 0.01 | 1.16E-72 | 0.72 | 6.07E-02 | 0.06 | 0.84 | 38 |
| Frequency of walking during leisure time | Physical activity | 32232 | -0.17 | 0.02 | 2.34E-30 | **2.79E-29** | 24 | 31500 | -0.16 | 0.02 | 9.37E-27 | **8.06E-26** | 33 | -0.17 | 0.01 | 2.32E-55 | 0.00 | 7.15E-01 | 0.06 | 0.83 | 19 |
| For how much of the time during the last four weeks have you felt worn out? | General health | 14411 | 0.41 | 0.03 | 5.01E-30 | **5.73E-29** | 25 | 14191 | 0.41 | 0.03 | 1.21E-28 | **1.15E-27** | 30 | 0.41 | 0.02 | 5.66E-57 | 0.00 | 9.95E-01 | 0.03 | 0.81 | 150 |
| Frequency of cycling during leisure time | Physical activity | 27392 | -0.19 | 0.02 | 1.14E-28 | **1.26E-27** | 26 | 26617 | -0.15 | 0.02 | 1.82E-17 | **9.25E-17** | 56 | -0.17 | 0.01 | 7.13E-44 | 0.65 | 9.33E-02 | 0.06 | 0.83 | 17 |
| Average portion size of potatoes/rice/pasta based on photographic illustration of four sizes (smallest to largest) | Food | 29676 | 0.18 | 0.02 | 6.27E-28 | **6.65E-27** | 27 | 28910 | 0.18 | 0.02 | 1.01E-26 | **8.40E-26** | 34 | 0.18 | 0.01 | 6.22E-53 | 0.00 | 8.57E-01 | 0.05 | 0.84 | 41 |
| Fibre intake (g/day) | Nutrients | 29676 | -0.18 | 0.02 | 1.24E-25 | **1.27E-24** | 28 | 28910 | -0.20 | 0.02 | 3.33E-29 | **3.26E-28** | 29 | -0.19 | 0.01 | 4.73E-53 | 0.00 | 5.63E-01 | 0.05 | 0.84 | 46 |
| For how much of the time during the last four weeks have you felt tired? | General health | 14453 | 0.38 | 0.03 | 1.30E-25 | **1.28E-24** | 29 | 14237 | 0.35 | 0.03 | 2.79E-21 | **1.73E-20** | 46 | 0.36 | 0.02 | 3.22E-45 | 0.00 | 5.42E-01 | 0.02 | 0.81 | 152 |
| Secoisolariciresinol intake (ug/day) | Nutrients | 29676 | -0.17 | 0.02 | 2.74E-25 | **2.61E-24** | 30 | 28910 | -0.18 | 0.02 | 5.88E-30 | **6.18E-29** | 27 | -0.18 | 0.01 | 1.82E-53 | 0.00 | 5.71E-01 | 0.05 | 0.84 | 47 |
| Get sick more often than other people | General health | 14424 | 0.35 | 0.03 | 1.16E-23 | **1.07E-22** | 31 | 14224 | 0.34 | 0.03 | 1.60E-20 | **9.30E-20** | 49 | 0.34 | 0.02 | 1.52E-42 | 0.00 | 7.46E-01 | 0.02 | 0.76 | 154 |
| Arachidonic acid (ARA) intake (g/day) | Nutrients | 29675 | 0.14 | 0.02 | 9.68E-21 | **8.42E-20** | 32 | 28909 | 0.17 | 0.02 | 1.20E-27 | **1.07E-26** | 32 | 0.16 | 0.01 | 1.96E-46 | 0.21 | 2.59E-01 | 0.05 | 0.84 | 48 |
| Animal based protein intake (g/day) | Nutrients | 29676 | 0.14 | 0.01 | 9.71E-21 | **8.42E-20** | 33 | 28910 | 0.18 | 0.01 | 6.35E-33 | **7.51E-32** | 24 | 0.16 | 0.01 | 2.73E-51 | 0.69 | 7.41E-02 | 0.05 | 0.84 | 43 |
| Participation in sports or physical exercise associations | Social | 10733 | -0.63 | 0.07 | 2.49E-20 | **2.09E-19** | 34 | 10375 | -0.65 | 0.07 | 1.56E-18 | **8.03E-18** | 55 | -0.64 | 0.05 | 3.13E-37 | 0.00 | 8.63E-01 | 0.03 | 0.86 | 143 |
| Physical limitation that made you not being able to perform certain work tasks or other activities during the last four weeks | General health | 14400 | 0.90 | 0.09 | 3.88E-20 | **3.17E-19** | 35 | 14200 | 1.09 | 0.10 | 1.05E-26 | **8.51E-26** | 35 | 0.99 | 0.07 | 5.86E-45 | 0.53 | 1.45E-01 | 0.02 | 0.81 | 153 |
| Long-term sickness | General health | 31418 | 0.37 | 0.04 | 1.87E-19 | **1.49E-18** | 36 | 30658 | 0.38 | 0.04 | 2.49E-20 | **1.38E-19** | 51 | 0.38 | 0.03 | 3.91E-38 | 0.00 | 8.40E-01 | 0.06 | 0.83 | 33 |
| Vitamin D intake (ug/day) | Nutrients | 29676 | 0.13 | 0.01 | 6.25E-19 | **4.83E-18** | 37 | 28910 | 0.14 | 0.01 | 1.46E-20 | **8.66E-20** | 48 | 0.13 | 0.01 | 7.65E-38 | 0.00 | 8.26E-01 | 0.05 | 0.84 | 54 |
| Sodium intake (mg/day) | Nutrients | 29675 | 0.14 | 0.02 | 6.62E-19 | **4.98E-18** | 38 | 28909 | 0.17 | 0.02 | 3.69E-28 | **3.38E-27** | 31 | 0.15 | 0.01 | 6.34E-45 | 0.55 | 1.37E-01 | 0.05 | 0.84 | 44 |
| Sum of Lariciresinol, Matairesinol, Pinoresinol, Secoisolariciresinol intake (ug/day) | Nutrients | 29676 | -0.15 | 0.02 | 6.91E-19 | **5.07E-18** | 39 | 28910 | -0.18 | 0.02 | 1.21E-26 | **9.55E-26** | 36 | -0.16 | 0.01 | 1.69E-43 | 0.42 | 1.90E-01 | 0.05 | 0.84 | 51 |
| Physical limitation that limited your ability to perform certain work tasks or other activities during the last four weeks | General health | 14428 | 0.87 | 0.09 | 2.53E-18 | **1.81E-17** | 40 | 14205 | 1.11 | 0.10 | 5.96E-26 | **4.58E-25** | 37 | 0.98 | 0.07 | 3.02E-42 | 0.68 | 7.48E-02 | 0.02 | 0.80 | 155 |
| Worsen in health in the future | General health | 14373 | 0.29 | 0.03 | 1.63E-16 | **1.14E-15** | 41 | 14181 | 0.30 | 0.03 | 1.19E-16 | **5.74E-16** | 59 | 0.29 | 0.02 | 1.43E-31 | 0.00 | 8.21E-01 | 0.02 | 0.80 | 158 |
| Pinoresinol intake (ug/day) | Nutrients | 29676 | -0.13 | 0.02 | 1.85E-16 | **1.26E-15** | 42 | 28910 | -0.16 | 0.02 | 2.78E-23 | **1.92E-22** | 41 | -0.15 | 0.01 | 8.99E-38 | 0.36 | 2.11E-01 | 0.05 | 0.84 | 56 |
| Parents or siblings have diabetes | General health | 32933 | 0.37 | 0.04 | 2.30E-16 | **1.53E-15** | 43 | 32207 | 0.43 | 0.04 | 1.62E-22 | **1.10E-21** | 42 | 0.40 | 0.03 | 4.88E-37 | 0.00 | 3.19E-01 | 0.06 | 0.83 | 18 |
| Lariciresinol intake (ug/day) | Nutrients | 29676 | -0.13 | 0.02 | 7.67E-16 | **4.96E-15** | 44 | 28910 | -0.16 | 0.02 | 2.33E-23 | **1.66E-22** | 40 | -0.15 | 0.01 | 3.69E-37 | 0.48 | 1.66E-01 | 0.05 | 0.84 | 58 |
| Physical limitation to participate in moderately demanding activities: lifting or carrying grocery bags | General health | 14428 | 0.29 | 0.03 | 7.80E-16 | **4.96E-15** | 45 | 14252 | 0.35 | 0.04 | 2.01E-20 | **1.14E-19** | 50 | 0.32 | 0.02 | 1.79E-34 | 0.28 | 2.40E-01 | 0.02 | 0.81 | 157 |
| Vitamin B3 intake (mg/day) | Nutrients | 29676 | 0.12 | 0.02 | 1.11E-15 | **6.92E-15** | 46 | 28910 | 0.16 | 0.02 | 9.93E-25 | **7.42E-24** | 38 | 0.14 | 0.01 | 3.23E-38 | 0.62 | 1.06E-01 | 0.05 | 0.84 | 49 |
| Wine | Alcohol | 29676 | -0.14 | 0.02 | 1.36E-15 | **8.28E-15** | 47 | 28910 | -0.12 | 0.02 | 1.10E-12 | **4.17E-12** | 75 | -0.13 | 0.01 | 1.27E-26 | 0.00 | 5.10E-01 | 0.05 | 0.84 | 59 |
| Sausage as main dish | Food | 29676 | 0.12 | 0.02 | 1.45E-15 | **8.65E-15** | 48 | 28910 | 0.17 | 0.01 | 1.28E-29 | **1.29E-28** | 28 | 0.14 | 0.01 | 1.53E-42 | 0.79 | 3.04E-02 | 0.05 | 0.84 | 50 |
| Folic acid intake (ug/day) | Nutrients | 29676 | -0.14 | 0.02 | 2.64E-15 | **1.54E-14** | 49 | 28910 | -0.16 | 0.02 | 4.85E-21 | **2.93E-20** | 47 | -0.15 | 0.01 | 1.85E-34 | 0.21 | 2.62E-01 | 0.05 | 0.84 | 63 |
| Plant based protein intake (g/day) | Nutrients | 29676 | -0.12 | 0.02 | 3.81E-15 | **2.18E-14** | 50 | 28910 | -0.14 | 0.02 | 5.45E-19 | **2.87E-18** | 54 | -0.13 | 0.01 | 2.01E-32 | 0.00 | 4.78E-01 | 0.05 | 0.84 | 60 |
| Monounsaturated fat intake (g/day) | Nutrients | 29676 | 0.12 | 0.02 | 7.06E-15 | **3.95E-14** | 51 | 28910 | 0.15 | 0.02 | 1.78E-22 | **1.18E-21** | 43 | 0.14 | 0.01 | 2.59E-35 | 0.49 | 1.60E-01 | 0.05 | 0.84 | 55 |
| Changed everyday exercise during the last year | Physical activity | 22054 | -0.14 | 0.02 | 7.18E-15 | **3.95E-14** | 52 | 21476 | -0.15 | 0.02 | 3.78E-17 | **1.88E-16** | 57 | -0.14 | 0.01 | 2.16E-30 | 0.00 | 6.71E-01 | 0.06 | 0.83 | 37 |
| Breakfast habits: Porridge w/o sandwich for breakfast vs not breakfast at all | Food | 4240 | -1.21 | 0.16 | 3.52E-14 | **1.90E-13** | 53 | 4318 | -1.09 | 0.15 | 5.11E-13 | **2.01E-12** | 72 | -1.15 | 0.11 | 1.38E-25 | 0.00 | 5.62E-01 | 0.04 | 0.82 | 133 |
| Educational level | Psychosocial | 33817 | -0.18 | 0.02 | 6.51E-14 | **3.45E-13** | 54 | 33074 | -0.23 | 0.02 | 7.15E-22 | **4.62E-21** | 44 | -0.20 | 0.02 | 1.17E-33 | 0.59 | 1.21E-01 | 0.06 | 0.83 | 12 |
| Monosaccharides intake (g/day) | Nutrients | 29676 | -0.12 | 0.02 | 1.19E-13 | **6.18E-13** | 55 | 28910 | -0.14 | 0.02 | 7.32E-16 | **3.20E-15** | 65 | -0.13 | 0.01 | 6.89E-28 | 0.00 | 6.19E-01 | 0.05 | 0.84 | 68 |
| Meat stew | Food | 29676 | 0.11 | 0.01 | 3.46E-13 | **1.77E-12** | 56 | 28910 | 0.12 | 0.01 | 4.01E-16 | **1.81E-15** | 63 | 0.11 | 0.01 | 1.11E-27 | 0.00 | 5.90E-01 | 0.05 | 0.84 | 62 |
| If you exercise, change in exercise habits during the last year | Physical activity | 18940 | -0.14 | 0.02 | 9.30E-13 | **4.67E-12** | 57 | 18460 | -0.15 | 0.02 | 3.10E-15 | **1.32E-14** | 67 | -0.15 | 0.01 | 2.33E-26 | 0.00 | 5.49E-01 | 0.06 | 0.83 | 32 |
| Smoking status: Former smokers vs non-smokers | Tobacco use | 24136 | 0.46 | 0.07 | 3.28E-12 | **1.62E-11** | 58 | 23338 | 0.35 | 0.07 | 7.37E-08 | **2.09E-07** | 100 | 0.41 | 0.05 | 2.61E-18 | 0.25 | 2.48E-01 | 0.06 | 0.84 | 31 |
| Total protein intake (g/day) | Nutrients | 29676 | 0.11 | 0.02 | 3.57E-12 | **1.73E-11** | 59 | 28910 | 0.14 | 0.02 | 2.35E-21 | **1.48E-20** | 45 | 0.13 | 0.01 | 2.93E-31 | 0.69 | 7.41E-02 | 0.05 | 0.84 | 57 |
| Permanent employment | Psychosocial | 29220 | -0.25 | 0.04 | 6.00E-12 | **2.86E-11** | 60 | 28464 | -0.26 | 0.04 | 7.02E-13 | **2.69E-12** | 74 | -0.26 | 0.03 | 2.78E-23 | 0.00 | 8.07E-01 | 0.07 | 0.82 | 5 |
| Matairesinol intake (ug/day) | Nutrients | 29676 | -0.11 | 0.02 | 6.92E-12 | **3.24E-11** | 61 | 28910 | -0.12 | 0.02 | 3.60E-14 | **1.51E-13** | 68 | -0.11 | 0.01 | 1.88E-24 | 0.00 | 5.93E-01 | 0.05 | 0.84 | 67 |
| Whole grain intake (g/day) | Food | 29676 | -0.10 | 0.01 | 1.28E-11 | **5.91E-11** | 62 | 28910 | -0.12 | 0.01 | 1.84E-16 | **8.69E-16** | 60 | -0.11 | 0.01 | 2.90E-26 | 0.18 | 2.70E-01 | 0.05 | 0.84 | 70 |
| Fiber cereals | Food | 29676 | -0.10 | 0.01 | 1.56E-11 | **7.06E-11** | 63 | 28910 | -0.08 | 0.02 | 7.00E-08 | **2.01E-07** | 99 | -0.09 | 0.01 | 9.09E-18 | 0.00 | 3.61E-01 | 0.05 | 0.84 | 75 |
| Steak, chop, e.g. | Food | 29676 | 0.10 | 0.01 | 1.82E-11 | **8.14E-11** | 64 | 28910 | 0.13 | 0.01 | 4.10E-19 | **2.24E-18** | 52 | 0.12 | 0.01 | 1.94E-28 | 0.63 | 1.02E-01 | 0.05 | 0.84 | 61 |
| Eat breakfast from 2000 | Food | 17120 | -0.65 | 0.10 | 2.80E-11 | **1.23E-10** | 65 | 16711 | -0.45 | 0.10 | 4.71E-06 | **1.10E-05** | 122 | -0.55 | 0.07 | 1.90E-15 | 0.54 | 1.40E-01 | 0.02 | 0.85 | 156 |
| Stigmasterol intake (mg/day) | Nutrients | 29676 | -0.11 | 0.02 | 3.36E-11 | **1.46E-10** | 66 | 28910 | -0.12 | 0.02 | 2.57E-13 | **1.03E-12** | 71 | -0.11 | 0.01 | 6.13E-23 | 0.00 | 6.33E-01 | 0.05 | 0.84 | 76 |
| Boiled coffee | Beverage | 29676 | 0.11 | 0.02 | 5.34E-11 | **2.28E-10** | 67 | 28910 | 0.05 | 0.02 | 3.00E-03 | **5.39E-03** | 158 | 0.08 | 0.01 | 1.78E-11 | 0.85 | 9.71E-03 | 0.05 | 0.84 | 82 |
| Carbohydrates intake (g/day) | Nutrients | 29676 | -0.10 | 0.02 | 5.85E-11 | **2.46E-10** | 68 | 28910 | -0.13 | 0.02 | 2.56E-15 | **1.10E-14** | 66 | -0.11 | 0.01 | 1.55E-24 | 0.00 | 3.35E-01 | 0.05 | 0.84 | 69 |
| Tea | Beverage | 29676 | -0.11 | 0.02 | 7.30E-11 | **3.02E-10** | 69 | 28910 | -0.11 | 0.02 | 9.50E-10 | **3.21E-09** | 84 | -0.11 | 0.01 | 4.19E-19 | 0.00 | 7.67E-01 | 0.05 | 0.84 | 66 |
| For how much of the time during the last four weeks has your physical health or your emotional problems limited your ability to interact with others? | General health | 14367 | 0.22 | 0.03 | 8.69E-11 | **3.55E-10** | 70 | 14156 | 0.26 | 0.03 | 6.86E-13 | **2.67E-12** | 73 | 0.24 | 0.02 | 4.28E-22 | 0.00 | 5.09E-01 | 0.02 | 0.80 | 160 |
| Frequency of picking berries or mushrooms during leisure time | Physical activity | 20723 | -0.14 | 0.02 | 9.95E-11 | **4.00E-10** | 71 | 20146 | -0.10 | 0.02 | 3.40E-06 | **8.24E-06** | 117 | -0.12 | 0.02 | 3.88E-15 | 0.40 | 1.95E-01 | 0.05 | 0.83 | 45 |
| Vitamin B12 intake (ug/day) | Nutrients | 29676 | 0.09 | 0.01 | 1.01E-10 | **4.00E-10** | 72 | 28910 | 0.10 | 0.01 | 1.52E-11 | **5.46E-11** | 79 | 0.09 | 0.01 | 9.42E-21 | 0.00 | 8.40E-01 | 0.05 | 0.84 | 72 |
| Breakfast habits: Gruel w/o sandwich for breakfast vs not breakfast at all | Food | 1457 | -1.79 | 0.26 | 1.10E-10 | **4.32E-10** | 73 | 1521 | -1.73 | 0.26 | 2.49E-10 | **8.53E-10** | 83 | -1.76 | 0.18 | 1.54E-19 | 0.00 | 8.73E-01 | 0.08 | 0.88 | 2 |
| Cholesterol intake (g/day) | Nutrients | 29676 | 0.10 | 0.02 | 1.59E-10 | **6.16E-10** | 74 | 28910 | 0.13 | 0.02 | 4.53E-17 | **2.22E-16** | 58 | 0.12 | 0.01 | 1.20E-25 | 0.47 | 1.71E-01 | 0.05 | 0.84 | 64 |
| Emotional problems that made you do less than you wanted during the last four weeks | General health | 14473 | 0.64 | 0.10 | 2.08E-10 | **7.95E-10** | 75 | 14256 | 0.70 | 0.10 | 2.83E-11 | **1.01E-10** | 80 | 0.67 | 0.07 | 3.71E-20 | 0.00 | 6.67E-01 | 0.02 | 0.80 | 162 |
| Oatflake, whole wheat, rye or barley porridge | Food | 29676 | -0.10 | 0.02 | 5.35E-10 | **2.01E-09** | 76 | 28910 | -0.09 | 0.02 | 3.71E-09 | **1.17E-08** | 90 | -0.09 | 0.01 | 1.14E-17 | 0.00 | 8.10E-01 | 0.05 | 0.84 | 79 |
| Satisfaction with leisure time | Psychosocial | 24570 | -0.12 | 0.02 | 8.79E-10 | **3.26E-09** | 77 | 24138 | -0.12 | 0.02 | 2.20E-09 | **7.18E-09** | 87 | -0.12 | 0.01 | 1.08E-17 | 0.00 | 9.31E-01 | 0.03 | 0.84 | 135 |
| Bacon | Food | 29676 | 0.09 | 0.01 | 9.44E-10 | **3.46E-09** | 78 | 28910 | 0.13 | 0.01 | 4.38E-19 | **2.35E-18** | 53 | 0.11 | 0.01 | 1.56E-26 | 0.72 | 5.98E-02 | 0.05 | 0.84 | 65 |
| Frequency of dancing during leisure time | Physical activity | 19406 | -0.13 | 0.02 | 1.90E-09 | **6.89E-09** | 79 | 18777 | -0.12 | 0.02 | 2.18E-07 | **6.00E-07** | 103 | -0.12 | 0.02 | 2.51E-15 | 0.00 | 5.67E-01 | 0.05 | 0.82 | 42 |
| Extent to what your physical and emotional health disrupted your usual social life during the last four weeks | General health | 14454 | 0.20 | 0.03 | 2.57E-09 | **9.19E-09** | 80 | 14255 | 0.29 | 0.03 | 5.40E-16 | **2.40E-15** | 64 | 0.24 | 0.02 | 2.79E-23 | 0.69 | 7.11E-02 | 0.02 | 0.81 | 159 |
| Root vegetables, carrot | Food | 29676 | -0.09 | 0.02 | 5.81E-09 | **2.05E-08** | 81 | 28910 | -0.10 | 0.02 | 1.33E-09 | **4.38E-09** | 86 | -0.10 | 0.01 | 4.25E-17 | 0.00 | 8.90E-01 | 0.05 | 0.84 | 92 |
| Physical limitation that reduced the normal time spent at work or in other activities during the last four weeks | General health | 14473 | 0.62 | 0.10 | 7.44E-09 | **2.60E-08** | 82 | 14253 | 0.81 | 0.11 | 2.40E-13 | **9.74E-13** | 70 | 0.71 | 0.07 | 1.97E-20 | 0.41 | 1.91E-01 | 0.02 | 0.80 | 161 |
| Beta-carotene intake (mg/day) | Nutrients | 29676 | -0.09 | 0.02 | 7.54E-09 | **2.60E-08** | 83 | 28910 | -0.10 | 0.02 | 2.98E-09 | **9.50E-09** | 89 | -0.10 | 0.01 | 1.22E-16 | 0.00 | 9.33E-01 | 0.05 | 0.84 | 95 |
| Salad dressing with oil | Food | 29676 | -0.09 | 0.01 | 8.45E-09 | **2.88E-08** | 84 | 28910 | -0.09 | 0.01 | 5.15E-09 | **1.61E-08** | 91 | -0.09 | 0.01 | 2.33E-16 | 0.00 | 9.57E-01 | 0.05 | 0.84 | 87 |
| Fat intake (g/day) | Nutrients | 29676 | 0.09 | 0.02 | 1.73E-08 | **5.84E-08** | 85 | 28910 | 0.10 | 0.02 | 8.14E-11 | **2.85E-10** | 81 | 0.09 | 0.01 | 9.47E-18 | 0.00 | 5.37E-01 | 0.05 | 0.84 | 80 |
| Sum of all lignans intake (ug/day) | Nutrients | 29676 | -0.09 | 0.02 | 1.95E-08 | **6.50E-08** | 86 | 28910 | -0.11 | 0.02 | 7.74E-12 | **2.86E-11** | 77 | -0.10 | 0.01 | 1.26E-18 | 0.00 | 3.80E-01 | 0.05 | 0.84 | 78 |
| Minced meat dishes | Food | 29676 | 0.08 | 0.01 | 4.02E-08 | **1.31E-07** | 87 | 28910 | 0.12 | 0.01 | 3.22E-16 | **1.47E-15** | 62 | 0.10 | 0.01 | 4.44E-22 | 0.72 | 6.07E-02 | 0.05 | 0.84 | 71 |
| Beta-sitosterol intake (mg/day) | Nutrients | 29676 | -0.09 | 0.02 | 4.02E-08 | **1.31E-07** | 88 | 28910 | -0.11 | 0.02 | 8.84E-12 | **3.22E-11** | 78 | -0.10 | 0.01 | 3.05E-18 | 0.00 | 3.54E-01 | 0.05 | 0.84 | 84 |
| Distance to work in kilometers (one way) | Physical activity | 28982 | 0.09 | 0.02 | 5.80E-08 | **1.86E-07** | 89 | 28250 | 0.09 | 0.02 | 5.20E-08 | **1.51E-07** | 98 | 0.09 | 0.01 | 1.52E-14 | 0.00 | 9.92E-01 | 0.06 | 0.83 | 21 |
| Average portion size of vegetables based on photographic illustration of four sizes (smallest to largest) | Food | 29676 | -0.09 | 0.02 | 1.00E-07 | **3.19E-07** | 90 | 28910 | -0.07 | 0.02 | 1.58E-05 | **3.55E-05** | 126 | -0.08 | 0.01 | 9.09E-12 | 0.00 | 4.80E-01 | 0.05 | 0.84 | 53 |

| | | | | | | | | | | | | | | | | | | | | | |
|---|---|---|---|---|---|---|---|---|---|---|---|---|---|---|---|---|---|---|---|---|---|
| Vitamin C intake (mg/day) | Nutrients | 29676 | -0.08 | 0.02 | 1.04E-07 | **3.28E-07** | 91 | 28910 | -0.11 | 0.02 | 1.03E-13 | **4.24E-13** | 69 | -0.10 | 0.01 | 1.79E-19 | 0.54 | 1.41E-01 | 0.05 | 0.84 | 83 |
| Frequency of hunting or fishing during leisure time | Physical activity | 20298 | 0.13 | 0.03 | 1.36E-07 | **4.24E-07** | 92 | 19707 | 0.12 | 0.03 | 1.25E-06 | **3.17E-06** | 112 | 0.13 | 0.02 | 8.35E-13 | 0.00 | 7.85E-01 | 0.06 | 0.83 | 34 |
| Palmitic acid intake (g/day) | Nutrients | 29676 | 0.08 | 0.02 | 2.42E-07 | **7.45E-07** | 93 | 28910 | 0.09 | 0.02 | 2.66E-09 | **8.60E-09** | 88 | 0.09 | 0.01 | 3.80E-15 | 0.00 | 5.96E-01 | 0.05 | 0.84 | 81 |
| White cabbage, lettuce, lettuce cabbage, spinach, borecole | Food | 29676 | -0.08 | 0.02 | 2.47E-07 | **7.50E-07** | 94 | 28910 | -0.11 | 0.02 | 3.93E-12 | **1.47E-11** | 76 | -0.10 | 0.01 | 1.18E-17 | 0.38 | 2.04E-01 | 0.05 | 0.84 | 89 |
| Alcohol intake (g/day) | Alcohol | 29676 | -0.10 | 0.02 | 3.00E-07 | **9.04E-07** | 95 | 28910 | -0.09 | 0.02 | 2.50E-07 | **6.77E-07** | 105 | -0.09 | 0.01 | 3.62E-13 | 0.00 | 9.35E-01 | 0.05 | 0.84 | 74 |
| Travel to work: Walk to work vs passive travel to work | Physical activity | 21492 | -0.32 | 0.06 | 4.66E-07 | **1.39E-06** | 96 | 21177 | -0.22 | 0.06 | 4.99E-04 | **1.00E-03** | 141 | -0.27 | 0.04 | 1.73E-09 | 0.21 | 2.60E-01 | 0.06 | 0.82 | 23 |
| Tomato, cucumber | Food | 29676 | -0.08 | 0.02 | 5.29E-07 | **1.56E-06** | 97 | 28910 | -0.09 | 0.02 | 5.39E-09 | **1.66E-08** | 92 | -0.09 | 0.01 | 1.69E-14 | 0.00 | 5.58E-01 | 0.05 | 0.84 | 96 |
| Fried potatoes, pommes frites | Food | 29676 | 0.07 | 0.02 | 6.12E-07 | **1.79E-06** | 98 | 28910 | 0.07 | 0.01 | 1.71E-06 | **4.30E-06** | 113 | 0.07 | 0.01 | 5.02E-12 | 0.00 | 7.65E-01 | 0.05 | 0.84 | 90 |
| Selenium intake (ug/day) | Nutrients | 29676 | 0.08 | 0.02 | 8.07E-07 | **2.33E-06** | 99 | 28910 | 0.09 | 0.02 | 1.83E-08 | **5.46E-08** | 95 | 0.08 | 0.01 | 8.14E-14 | 0.00 | 6.22E-01 | 0.05 | 0.84 | 77 |
| Banana | Food | 29676 | -0.07 | 0.02 | 8.85E-07 | **2.53E-06** | 100 | 28910 | -0.04 | 0.02 | 1.99E-02 | **3.15E-02** | 179 | -0.05 | 0.01 | 2.81E-07 | 0.69 | 7.29E-02 | 0.05 | 0.84 | 111 |
| Participation in other association | Social | 10733 | 0.33 | 0.07 | 9.93E-07 | **2.81E-06** | 101 | 10375 | 0.46 | 0.07 | 1.77E-10 | **6.14E-10** | 82 | 0.39 | 0.05 | 2.12E-15 | 0.45 | 1.77E-01 | 0.03 | 0.86 | 148 |
| Low fat milk (0,5%) | Beverage | 29676 | 0.07 | 0.02 | 1.16E-06 | **3.26E-06** | 102 | 28910 | 0.08 | 0.02 | 4.18E-07 | **1.09E-06** | 109 | 0.08 | 0.01 | 2.28E-12 | 0.00 | 8.78E-01 | 0.05 | 0.84 | 86 |
| Pancake, waffle, Swedish dumpling | Food | 29676 | 0.07 | 0.02 | 1.26E-06 | **3.49E-06** | 103 | 28910 | 0.09 | 0.02 | 1.38E-08 | **4.18E-08** | 94 | 0.08 | 0.01 | 1.01E-13 | 0.00 | 5.55E-01 | 0.05 | 0.84 | 85 |
| Magnesium intake (mg/day) | Nutrients | 29676 | -0.08 | 0.02 | 1.29E-06 | **3.54E-06** | 104 | 28910 | -0.10 | 0.02 | 1.03E-09 | **3.45E-09** | 85 | -0.09 | 0.01 | 1.02E-14 | 0.00 | 3.57E-01 | 0.05 | 0.84 | 108 |
| Possibility to speak with colleagues during breaks | Psychosocial | 32095 | -0.07 | 0.01 | 1.33E-06 | **3.62E-06** | 105 | 31415 | -0.07 | 0.01 | 3.35E-06 | **8.19E-06** | 116 | -0.07 | 0.01 | 1.99E-11 | 0.00 | 9.26E-01 | 0.06 | 0.83 | 35 |
| Light and physically active work | Physical activity | 32431 | -0.15 | 0.03 | 1.71E-06 | **4.62E-06** | 106 | 31717 | -0.13 | 0.03 | 9.09E-05 | **1.99E-04** | 130 | -0.14 | 0.02 | 7.37E-10 | 0.00 | 5.88E-01 | 0.06 | 0.83 | 26 |
| Years using snuff | Tobacco use | 31689 | 0.12 | 0.02 | 1.82E-06 | **4.86E-06** | 107 | 31005 | 0.10 | 0.02 | 6.03E-05 | **1.34E-04** | 128 | 0.11 | 0.02 | 5.04E-10 | 0.00 | 6.46E-01 | 0.06 | 0.83 | 25 |
| Sedentary or standing work | Physical activity | 32431 | 0.17 | 0.03 | 1.97E-06 | **5.22E-06** | 108 | 31717 | 0.08 | 0.04 | 1.79E-02 | **2.90E-02** | 175 | 0.12 | 0.02 | 4.58E-07 | 0.64 | 9.39E-02 | 0.06 | 0.83 | 27 |
| Brewed (filtered) coffee | Beverage | 29676 | -0.08 | 0.02 | 2.13E-06 | **5.59E-06** | 109 | 28910 | -0.06 | 0.02 | 7.39E-04 | **1.47E-03** | 143 | -0.07 | 0.01 | 9.49E-09 | 0.00 | 3.37E-01 | 0.05 | 0.84 | 107 |
| For how much of the time during the last four weeks have you felt gloomy and sad? | General health | 14420 | 0.16 | 0.03 | 2.45E-06 | **6.38E-06** | 110 | 14223 | 0.16 | 0.03 | 3.76E-06 | **8.97E-06** | 119 | 0.16 | 0.02 | 4.01E-11 | 0.00 | 9.81E-01 | 0.02 | 0.81 | 163 |
| Boiled or baked potato | Food | 29676 | 0.07 | 0.02 | 2.72E-06 | **7.01E-06** | 111 | 28910 | 0.05 | 0.01 | 7.53E-04 | **1.47E-03** | 145 | 0.06 | 0.01 | 1.24E-08 | 0.00 | 3.33E-01 | 0.05 | 0.84 | 105 |
| For how much of the time during the last four weeks have you felt happy? | General health | 14439 | -0.16 | 0.03 | 3.13E-06 | **7.98E-06** | 112 | 14228 | -0.14 | 0.03 | 9.16E-05 | **1.99E-04** | 131 | -0.15 | 0.02 | 1.25E-09 | 0.00 | 6.47E-01 | 0.02 | 0.81 | 165 |
| Sour milk, yoghurt (3% fat) | Food | 29676 | -0.07 | 0.01 | 4.16E-06 | **1.05E-05** | 113 | 28910 | -0.07 | 0.01 | 2.82E-06 | **6.97E-06** | 115 | -0.07 | 0.01 | 5.15E-11 | 0.00 | 9.46E-01 | 0.05 | 0.84 | 100 |
| Participation in study circles | Social | 10733 | 0.41 | 0.09 | 5.32E-06 | **1.34E-05** | 114 | 10375 | 0.29 | 0.10 | 2.36E-03 | **4.30E-03** | 156 | 0.35 | 0.07 | 6.30E-08 | 0.00 | 3.92E-01 | 0.03 | 0.86 | 149 |
| For how much of the time during the last four weeks have you felt so depressed that nothing could cheer you up? | General health | 14450 | 0.15 | 0.03 | 5.90E-06 | **1.47E-05** | 115 | 14248 | 0.13 | 0.03 | 1.35E-04 | **2.90E-04** | 132 | 0.14 | 0.02 | 3.37E-09 | 0.00 | 6.63E-01 | 0.02 | 0.81 | 166 |
| Years smoking | Tobacco use | 29497 | 0.12 | 0.03 | 6.09E-06 | **1.50E-05** | 116 | 28721 | 0.08 | 0.03 | 3.84E-03 | **6.78E-03** | 161 | 0.10 | 0.02 | 1.55E-07 | 0.23 | 2.55E-01 | 0.06 | 0.83 | 30 |
| Apple, pear, peach, orange, mandarin and grapefruit | Food | 29676 | -0.08 | 0.02 | 8.32E-06 | **2.03E-05** | 117 | 28910 | -0.09 | 0.02 | 3.37E-07 | **9.03E-07** | 106 | -0.08 | 0.01 | 1.39E-11 | 0.00 | 6.48E-01 | 0.05 | 0.84 | 104 |
| Mashed potato | Food | 7086 | 0.26 | 0.04 | 1.07E-05 | **2.60E-05** | 118 | 6838 | 0.20 | 0.04 | 5.03E-03 | **8.71E-03** | 164 | 0.24 | 0.03 | 1.25E-07 | 0.05 | 3.05E-01 | 0.04 | 0.75 | 131 |
| Sum of phytosterols intake (mg/day) | Nutrients | 29676 | -0.07 | 0.02 | 1.15E-05 | **2.76E-05** | 119 | 28910 | -0.09 | 0.02 | 1.38E-08 | **4.18E-08** | 93 | -0.08 | 0.01 | 1.10E-12 | 0.00 | 3.70E-01 | 0.05 | 0.84 | 99 |
| Margarine for cooking | Food | 29676 | 0.06 | 0.01 | 1.18E-05 | **2.81E-05** | 120 | 28910 | 0.07 | 0.01 | 3.82E-07 | **1.01E-06** | 108 | 0.07 | 0.01 | 2.24E-11 | 0.00 | 6.25E-01 | 0.05 | 0.84 | 102 |
| Sucrose intake (g/day) | Nutrients | 29676 | -0.07 | 0.02 | 2.15E-05 | **5.08E-05** | 121 | 28910 | -0.08 | 0.02 | 3.68E-07 | **9.76E-07** | 107 | -0.07 | 0.01 | 4.08E-11 | 0.00 | 5.61E-01 | 0.05 | 0.84 | 88 |
| Self rate of overall health compared to a year ago | General health | 14365 | -0.14 | 0.03 | 3.08E-05 | **7.21E-05** | 122 | 14157 | -0.19 | 0.03 | 4.18E-08 | **1.22E-07** | 97 | -0.16 | 0.02 | 9.69E-12 | 0.19 | 2.66E-01 | 0.02 | 0.81 | 164 |
| Salty fish | Food | 29676 | 0.06 | 0.01 | 3.32E-05 | **7.73E-05** | 123 | 28910 | 0.08 | 0.01 | 2.31E-08 | **6.83E-08** | 96 | 0.07 | 0.01 | 7.42E-12 | 0.35 | 2.16E-01 | 0.05 | 0.84 | 91 |
| Hamburger | Food | 29676 | 0.06 | 0.02 | 3.78E-05 | **8.73E-05** | 124 | 28910 | 0.13 | 0.02 | 2.63E-16 | **1.22E-15** | 61 | 0.10 | 0.01 | 2.99E-18 | 0.88 | 4.15E-03 | 0.05 | 0.84 | 73 |
| Sausage, liver pate on bread | Food | 29676 | 0.06 | 0.02 | 4.09E-05 | **9.37E-05** | 125 | 28910 | 0.08 | 0.02 | 1.73E-07 | **4.80E-07** | 102 | 0.07 | 0.01 | 4.20E-11 | 0.00 | 4.27E-01 | 0.05 | 0.84 | 94 |
| Marital status: Single vs Divorced/separated | Social | 5550 | -0.46 | 0.11 | 4.47E-05 | **1.01E-04** | 126 | 5551 | -0.53 | 0.11 | 2.22E-06 | **5.54E-06** | 114 | -0.49 | 0.08 | 4.54E-10 | 0.00 | 6.55E-01 | 0.06 | 0.80 | 8 |
| Syringaresinol intake (ug/day) | Nutrients | 29676 | -0.06 | 0.02 | 4.51E-05 | **1.01E-04** | 127 | 28910 | -0.07 | 0.02 | 1.04E-06 | **2.66E-06** | 111 | -0.07 | 0.01 | 2.33E-10 | 0.00 | 5.66E-01 | 0.05 | 0.84 | 97 |
| Breakfast habits: Coffee/tea and wheat buns or rusk for breakfast vs not breakfast at all | Food | 1606 | -0.84 | 0.20 | 4.53E-05 | **1.01E-04** | 128 | 1597 | -0.85 | 0.23 | 2.62E-04 | **5.40E-04** | 138 | -0.84 | 0.15 | 4.26E-08 | 0.00 | 9.64E-01 | 0.05 | 0.78 | 128 |
| Number of cigarretes smoked per day (in groups) | Tobacco use | 27140 | 0.10 | 0.02 | 4.73E-05 | **1.05E-04** | 129 | 26696 | 0.06 | 0.02 | 1.43E-02 | **2.35E-02** | 173 | 0.08 | 0.02 | 3.99E-06 | 0.22 | 2.57E-01 | 0.06 | 0.83 | 40 |
| Job demands to work very fast | Psychosocial | 33324 | -0.06 | 0.01 | 5.96E-05 | **1.31E-04** | 130 | 32667 | -0.05 | 0.01 | 2.20E-04 | **4.62E-04** | 135 | -0.06 | 0.01 | 4.97E-08 | 0.00 | 8.37E-01 | 0.06 | 0.83 | 20 |
| Mixed frozen vegetables | Food | 7086 | 0.22 | 0.04 | 6.98E-05 | **1.52E-04** | 131 | 6838 | 0.21 | 0.04 | 4.05E-03 | **7.09E-03** | 162 | 0.21 | 0.03 | 3.88E-07 | 0.00 | 8.60E-01 | 0.04 | 0.73 | 132 |
| Vitamin A intake (mg/day) | Nutrients | 29676 | 0.06 | 0.02 | 9.04E-05 | **1.96E-04** | 132 | 28910 | 0.05 | 0.02 | 2.32E-03 | **4.24E-03** | 155 | 0.05 | 0.01 | 8.74E-07 | 0.00 | 5.16E-01 | 0.05 | 0.84 | 120 |
| Work shifts/weekends | Psychosocial | 32226 | 0.14 | 0.04 | 1.22E-04 | **2.62E-04** | 133 | 31608 | 0.12 | 0.04 | 1.04E-03 | **2.02E-03** | 146 | 0.13 | 0.03 | 4.79E-07 | 0.00 | 6.74E-01 | 0.06 | 0.83 | 28 |
| White meat (poultry) | Food | 29676 | 0.06 | 0.02 | 1.44E-04 | **3.08E-04** | 134 | 28910 | 0.08 | 0.02 | 8.46E-08 | **2.38E-07** | 101 | 0.07 | 0.01 | 9.82E-11 | 0.22 | 2.56E-01 | 0.05 | 0.84 | 98 |
| Snuff status: Former snuff users vs non-snuff users | Tobacco use | 26976 | 0.27 | 0.07 | 2.14E-04 | **4.54E-04** | 135 | 26290 | 0.11 | 0.07 | 1.14E-01 | 1.52E-01 | 213 | 0.19 | 0.05 | 1.86E-04 | 0.55 | 1.34E-01 | - | - | - |
| Repetitive job | Psychosocial | 33276 | 0.06 | 0.02 | 2.60E-04 | **5.46E-04** | 136 | 32600 | 0.03 | 0.02 | 5.07E-02 | 7.42E-02 | 194 | 0.04 | 0.01 | 7.23E-05 | 0.29 | 2.35E-01 | - | - | - |
| Travel to work: Irregular travel mode to work vs passive travel to work | Physical activity | 21589 | -0.21 | 0.06 | 2.97E-04 | **6.20E-04** | 137 | 21208 | -0.13 | 0.06 | 1.80E-02 | **2.91E-02** | 176 | -0.17 | 0.04 | 2.51E-05 | 0.00 | 3.35E-01 | 0.06 | 0.84 | 14 |
| Sugar, honey, marmelade, jam | Food | 29676 | -0.06 | 0.02 | 3.22E-04 | **6.68E-04** | 138 | 28910 | -0.04 | 0.02 | 2.51E-02 | **3.88E-02** | 184 | -0.05 | 0.01 | 3.72E-05 | 0.00 | 3.32E-01 | 0.05 | 0.84 | 117 |
| Berries (fresh or frozen) | Food | 29676 | -0.05 | 0.01 | 3.69E-04 | **7.58E-04** | 139 | 28910 | -0.07 | 0.01 | 3.51E-06 | **8.45E-06** | 118 | -0.06 | 0.01 | 6.79E-09 | 0.00 | 4.35E-01 | 0.05 | 0.84 | 112 |
| Frequency of gardening during leisure time | Physical activity | 20880 | -0.07 | 0.02 | 4.26E-04 | **8.71E-04** | 140 | 20186 | -0.03 | 0.02 | 2.21E-01 | 2.79E-01 | 225 | -0.05 | 0.01 | 8.02E-04 | 0.63 | 1.01E-01 | - | - | - |
| Pizza | Food | 29676 | 0.05 | 0.02 | 5.32E-04 | **1.08E-03** | 141 | 28910 | 0.07 | 0.02 | 3.96E-06 | **9.37E-06** | 120 | 0.06 | 0.01 | 1.12E-08 | 0.00 | 4.15E-01 | 0.05 | 0.84 | 106 |
| Low fat margarine on bread | Food | 29676 | 0.05 | 0.01 | 5.74E-04 | **1.16E-03** | 142 | 28910 | 0.03 | 0.01 | 1.89E-02 | **3.02E-02** | 178 | 0.04 | 0.01 | 4.29E-05 | 0.00 | 4.26E-01 | 0.05 | 0.84 | 121 |
| Receive hugs to comfort and support you | Social | 33213 | -0.14 | 0.04 | 6.47E-04 | **1.29E-03** | 143 | 32440 | -0.08 | 0.04 | 5.87E-02 | 8.50E-02 | 196 | -0.11 | 0.03 | 1.70E-04 | 0.07 | 2.99E-01 | - | - | - |
| Brown beans, pea soup | Food | 29676 | -0.05 | 0.01 | 6.93E-04 | **1.38E-03** | 144 | 28910 | -0.05 | 0.01 | 2.14E-04 | **4.54E-04** | 134 | -0.05 | 0.01 | 5.25E-07 | 0.00 | 8.38E-01 | 0.05 | 0.84 | 116 |
| Satisfaction with economy | Psychosocial | 24605 | -0.07 | 0.02 | 8.54E-04 | **1.68E-03** | 145 | 24168 | -0.07 | 0.02 | 2.81E-04 | **5.74E-04** | 139 | -0.07 | 0.01 | 8.36E-07 | 0.00 | 8.34E-01 | 0.03 | 0.84 | 137 |
| Eicosapentaenoic acid (EPA) intake (g/day) | Nutrients | 29675 | 0.05 | 0.02 | 9.12E-04 | **1.79E-03** | 146 | 28910 | 0.07 | 0.02 | 7.15E-06 | **1.64E-05** | 124 | 0.06 | 0.01 | 3.50E-08 | 0.00 | 3.88E-01 | 0.05 | 0.84 | 101 |
| Do you feel important and appreciated in your home? | Psychosocial | 24447 | -0.07 | 0.02 | 9.59E-04 | **1.87E-03** | 147 | 24000 | 0.01 | 0.02 | 7.48E-01 | 7.97E-01 | 266 | -0.03 | 0.01 | 3.44E-02 | 0.85 | 1.05E-02 | - | - | - |
| Equol intake (ug/day) | Nutrients | 29676 | 0.05 | 0.02 | 9.73E-04 | **1.88E-03** | 148 | 28910 | 0.05 | 0.02 | 3.19E-04 | **6.47E-04** | 140 | 0.05 | 0.01 | 1.07E-06 | 0.00 | 8.38E-01 | 0.05 | 0.84 | 109 |
| Beta-sitostanol intake (mg/day) | Nutrients | 29676 | -0.05 | 0.01 | 9.79E-04 | **1.88E-03** | 149 | 28910 | -0.07 | 0.01 | 4.71E-06 | **1.10E-05** | 121 | -0.06 | 0.01 | 2.58E-08 | 0.00 | 3.64E-01 | 0.05 | 0.84 | 110 |
| Vitamin B6 intake (mg/day) | Nutrients | 29676 | 0.05 | 0.02 | 1.19E-03 | **2.27E-03** | 150 | 28910 | 0.08 | 0.02 | 9.52E-07 | **2.46E-06** | 110 | 0.06 | 0.01 | 8.31E-09 | 0.25 | 2.49E-01 | 0.05 | 0.84 | 93 |
| Participation in associations or voluntary organisations | Social | 33259 | -0.10 | 0.03 | 1.23E-03 | **2.32E-03** | 151 | 32545 | 0.00 | 0.03 | 9.45E-01 | 9.49E-01 | 283 | -0.05 | 0.02 | 1.89E-02 | 0.80 | 2.62E-02 | - | - | - |
| Liquor, spirits | Alcohol | 29676 | 0.06 | 0.02 | 1.41E-03 | **2.66E-03** | 152 | 28910 | 0.05 | 0.02 | 3.19E-03 | **5.69E-03** | 159 | 0.05 | 0.01 | 1.54E-05 | 0.00 | 6.55E-01 | 0.05 | 0.84 | 118 |
| Docosahexaenoic acid (DHA) intake (g/day) | Nutrients | 29676 | 0.05 | 0.02 | 1.93E-03 | **3.60E-03** | 153 | 28910 | 0.07 | 0.02 | 1.48E-05 | **3.37E-05** | 125 | 0.06 | 0.01 | 1.54E-07 | 0.00 | 3.62E-01 | 0.05 | 0.84 | 103 |
| Campestanol intake (mg/day) | Nutrients | 29676 | -0.05 | 0.01 | 1.94E-03 | **3.60E-03** | 154 | 28910 | -0.06 | 0.01 | 1.64E-05 | **3.68E-05** | 127 | -0.05 | 0.01 | 1.62E-07 | 0.00 | 3.93E-01 | 0.05 | 0.84 | 113 |
| Pasta | Food | 29676 | 0.05 | 0.02 | 2.01E-03 | **3.71E-03** | 155 | 28910 | 0.03 | 0.02 | 2.85E-02 | **4.37E-02** | 185 | 0.04 | 0.01 | 1.91E-04 | 0.00 | 5.15E-01 | 0.05 | 0.84 | 124 |
| Sleep status | Sleep | 24623 | -0.06 | 0.02 | 2.75E-03 | **5.04E-03** | 156 | 24199 | -0.05 | 0.02 | 2.18E-02 | **3.43E-02** | 180 | -0.05 | 0.01 | 1.82E-04 | 0.00 | 6.34E-01 | 0.03 | 0.84 | 141 |
| Grams of tobacco smoked per week | Tobacco use | 21247 | -0.07 | 0.02 | 2.92E-03 | **5.31E-03** | 157 | 20790 | 0.01 | 0.03 | 8.00E-01 | 8.38E-01 | 271 | -0.04 | 0.02 | 3.59E-02 | 0.78 | 3.34E-02 | - | - | - |
| Patience status | Psychosocial | 24589 | -0.06 | 0.02 | 3.23E-03 | **5.85E-03** | 158 | 24161 | -0.07 | 0.02 | 2.50E-04 | **5.18E-04** | 137 | -0.07 | 0.01 | 2.94E-06 | 0.00 | 6.32E-01 | 0.03 | 0.85 | 138 |
| Saturated fat intake (g/day) | Nutrients | 29676 | 0.05 | 0.02 | 3.31E-03 | **5.95E-03** | 159 | 28910 | 0.05 | 0.02 | 1.93E-03 | **3.61E-03** | 152 | 0.05 | 0.01 | 1.96E-05 | 0.00 | 9.21E-01 | 0.05 | 0.84 | 119 |
| Parents or siblings had a cerebral hemorrhage/thrombosis or cardiac infarction before the age of 60 | General health | 32888 | 0.13 | 0.05 | 3.60E-03 | **6.43E-03** | 160 | 32193 | 0.14 | 0.05 | 1.93E-03 | **3.61E-03** | 151 | 0.14 | 0.03 | 2.13E-05 | 0.00 | 8.90E-01 | 0.06 | 0.83 | 24 |
| For how much of the time during the last four weeks have you felt calm and serene? | General health | 14433 | -0.10 | 0.03 | 3.65E-03 | **6.49E-03** | 161 | 14242 | -0.11 | 0.03 | 2.19E-03 | **4.04E-03** | 154 | -0.10 | 0.02 | 2.44E-05 | 0.00 | 8.57E-01 | 0.02 | 0.81 | 167 |
| Milk, sour milk (1,5%) | Beverage | 29676 | 0.04 | 0.02 | 3.69E-03 | **6.52E-03** | 162 | 28910 | 0.03 | 0.02 | 6.13E-02 | 8.72E-02 | 199 | 0.04 | 0.01 | 7.23E-04 | 0.00 | 4.77E-01 | - | - | - |
| Medium beer | Alcohol | 29676 | -0.04 | 0.02 | 4.45E-03 | **7.81E-03** | 163 | 28910 | -0.06 | 0.02 | 1.67E-04 | **3.56E-04** | 133 | -0.05 | 0.01 | 2.80E-06 | 0.00 | 5.72E-01 | 0.05 | 0.84 | 114 |
| Enterodiol intake (ug/day) | Nutrients | 29676 | 0.04 | 0.01 | 4.87E-03 | **8.45E-03** | 164 | 28910 | 0.04 | 0.01 | 1.04E-02 | **1.74E-02** | 170 | 0.04 | 0.01 | 1.43E-04 | 0.00 | 8.57E-01 | 0.05 | 0.84 | 122 |
| Medioresinol intake (ug/day) | Nutrients | 29676 | -0.04 | 0.02 | 4.88E-03 | **8.45E-03** | 165 | 28910 | -0.06 | 0.02 | 8.30E-05 | **1.83E-04** | 129 | -0.05 | 0.01 | 1.81E-06 | 0.00 | 4.29E-01 | 0.05 | 0.84 | 115 |
| Sodas, soft drinks, juice | Beverage | 29676 | 0.04 | 0.02 | 5.20E-03 | **8.95E-03** | 166 | 28910 | -0.01 | 0.01 | 4.76E-01 | 5.45E-01 | 248 | 0.02 | 0.01 | 1.41E-01 | 0.84 | 1.31E-02 | - | - | - |
| Snuff status: Snuff users vs non-snuff users | Tobacco use | 28739 | 0.19 | 0.07 | 5.76E-03 | **9.87E-03** | 167 | 28205 | 0.04 | 0.07 | 5.41E-01 | 6.08E-01 | 253 | 0.12 | 0.05 | 1.59E-02 | 0.54 | 1.39E-01 | - | - | - |
| Mood status | Psychosocial | 24589 | -0.05 | 0.02 | 8.24E-03 | **1.40E-02** | 168 | 24154 | -0.05 | 0.02 | 1.66E-02 | **2.71E-02** | 174 | -0.05 | 0.01 | 3.68E-04 | 0.00 | 8.63E-01 | 0.03 | 0.84 | 140 |
| Teetotaler | Alcohol | 21681 | 0.20 | 0.08 | 9.98E-03 | **1.69E-02** | 169 | 21098 | 0.21 | 0.08 | 1.13E-02 | **1.86E-02** | 172 | 0.20 | 0.06 | 3.02E-04 | 0.00 | 9.87E-01 | 0.05 | 0.83 | 52 |
| Number of social interactions during a normal week | Social | 33458 | -0.04 | 0.02 | 1.22E-02 | **2.06E-02** | 170 | 32694 | -0.01 | 0.02 | 3.37E-01 | 4.03E-01 | 238 | -0.03 | 0.01 | 1.39E-02 | 0.12 | 2.86E-01 | - | - | - |
| Number of cigarretes smoked per day | Tobacco use | 21765 | -0.06 | 0.02 | 1.35E-02 | **2.25E-02** | 171 | 21328 | -0.08 | 0.03 | 2.04E-03 | **3.80E-03** | 153 | -0.07 | 0.02 | 8.77E-05 | 0.00 | 6.30E-01 | 0.06 | 0.84 | 16 |
| Frequent social contacts with colleagues during work | Psychosocial | 31875 | -0.04 | 0.01 | 1.39E-02 | **2.31E-02** | 172 | 31180 | -0.05 | 0.01 | 7.43E-04 | **1.47E-03** | 144 | -0.04 | 0.01 | 3.70E-05 | 0.00 | 5.21E-01 | 0.06 | 0.83 | 36 |
| Polyunsaturated fat intake (g/day) | Nutrients | 29676 | 0.04 | 0.01 | 1.52E-02 | **2.51E-02** | 173 | 28910 | 0.03 | 0.01 | 3.17E-02 | **4.79E-02** | 188 | 0.03 | 0.01 | 1.22E-03 | 0.00 | 8.41E-01 | 0.05 | 0.84 | 125 |
| Iodine intake (ug/day) | Nutrients | 29676 | 0.04 | 0.02 | 1.58E-02 | **2.59E-02** | 174 | 28910 | 0.02 | 0.02 | 2.67E-01 | 3.27E-01 | 232 | 0.03 | 0.01 | 1.28E-02 | 0.00 | 3.52E-01 | - | - | - |
| Control over own work assignment | Psychosocial | 33316 | 0.04 | 0.01 | 1.70E-02 | **2.78E-02** | 175 | 32647 | 0.02 | 0.01 | 1.98E-01 | 2.52E-01 | 223 | 0.03 | 0.01 | 9.26E-03 | 0.00 | 4.46E-01 | - | - | - |
| Total energy intake (kcal/day) | Nutrients | 29676 | 0.04 | 0.02 | 2.00E-02 | **3.26E-02** | 176 | 28910 | 0.09 | 0.02 | 5.24E-06 | **1.21E-05** | 123 | 0.07 | 0.01 | 1.12E-06 | 0.59 | 1.17E-01 | 0.05 | 0.84 | 126 |
| Skill demand from job | Psychosocial | 33240 | -0.03 | 0.02 | 2.07E-02 | **3.34E-02** | 177 | 32538 | -0.03 | 0.02 | 4.52E-02 | 6.68E-02 | 192 | -0.03 | 0.01 | 2.26E-03 | 0.00 | 8.46E-01 | - | - | - |
| Cream, creme fraiche, sour cream | Food | 29676 | -0.03 | 0.01 | 2.38E-02 | **3.83E-02** | 178 | 28910 | -0.03 | 0.01 | 5.81E-02 | 8.47E-02 | 195 | -0.03 | 0.01 | 3.35E-03 | 0.00 | 7.67E-01 | - | - | - |
| Snore during sleep | Sleep | 10559 | 1.05 | 0.04 | 2.40E-02 | **3.83E-02** | 179 | 10456 | 1.05 | 0.04 | 1.23E-146 | **1.75E-144** | 2 | 1.05 | 0.03 | - | 0.00 | 9.96E-01 | 0.08 | 0.08 | 3 |
| Hearing status | General health | 24624 | -0.05 | 0.02 | 2.49E-02 | **3.96E-02** | 180 | 24189 | -0.06 | 0.02 | 1.01E-02 | **1.69E-02** | 169 | -0.05 | 0.02 | 6.58E-04 | 0.00 | 8.27E-01 | 0.03 | 0.84 | 139 |
| Rosehip, sweet syrup soup | Food | 29676 | -0.03 | 0.01 | 2.56E-02 | **4.04E-02** | 181 | 28910 | 0.00 | 0.01 | 9.24E-01 | 9.34E-01 | 281 | -0.02 | 0.01 | 1.05E-01 | 0.58 | 1.25E-01 | - | - | - |
| For how much of the time during the last four weeks have you felt very nervous? | General health | 14434 | 0.07 | 0.03 | 2.66E-02 | **4.18E-02** | 182 | 14235 | 0.00 | 0.03 | 9.17E-01 | 9.31E-01 | 279 | 0.04 | 0.02 | 9.47E-02 | 0.54 | 1.41E-01 | - | - | - |
| Frequncy of social contacts with colleagues during leisure time | Psychosocial | 31333 | 0.03 | 0.01 | 2.73E-02 | **4.27E-02** | 183 | 30648 | 0.00 | 0.01 | 7.36E-01 | 7.95E-01 | 263 | 0.01 | 0.01 | 1.84E-01 | 0.69 | 7.28E-02 | - | - | - |
| Coffee rolls/buns, rusk | Food | 29676 | -0.03 | 0.02 | 2.77E-02 | **4.31E-02** | 184 | 28910 | -0.02 | 0.02 | 2.08E-01 | 2.63E-01 | 224 | -0.03 | 0.01 | 1.43E-02 | 0.00 | 5.12E-01 | - | - | - |
| Whole grain soft bread | Food | 29676 | -0.03 | 0.01 | 2.81E-02 | **4.34E-02** | 185 | 28910 | -0.04 | 0.01 | 6.24E-03 | **1.07E-02** | 166 | -0.04 | 0.01 | 4.90E-04 | 0.00 | 6.98E-01 | 0.05 | 0.84 | 123 |

| | | | | | | | | | | | | | | | | | | | | | |
|---|---|---|---|---|---|---|---|---|---|---|---|---|---|---|---|---|---|---|---|---|---|
| Amount of exercise during the last 12 months | Physical activity | 11430 | -0.75 | 0.04 | 3.21E-02 | **4.93E-02** | 186 | 11250 | -0.82 | 0.04 | 3.03E-02 | **4.60E-02** | 187 | -0.78 | 0.03 | - | 0.22 | 2.58E-01 | 0.05 | 0.05 | 127 |
| Milk, sour milk (3%) | Beverage | 29676 | -0.03 | 0.02 | 3.29E-02 | 5.04E-02 | 187 | 28910 | -0.04 | 0.02 | 8.09E-03 | **1.37E-02** | 168 | -0.04 | 0.01 | 7.22E-04 | 0.00 | 7.17E-01 | - | - | - |
| Smoked fish/meat | Food | 29676 | 0.03 | 0.01 | 3.42E-02 | 5.20E-02 | 188 | 28910 | 0.02 | 0.01 | 2.61E-01 | 3.23E-01 | 229 | 0.02 | 0.01 | 2.14E-02 | 0.00 | 4.98E-01 | - | - | - |
| Heptadecanoic acid intake (g/day) | Nutrients | 29676 | -0.03 | 0.02 | 3.82E-02 | 5.72E-02 | 189 | 28910 | -0.05 | 0.02 | 1.78E-03 | **3.38E-03** | 150 | -0.04 | 0.01 | 2.35E-04 | 0.00 | 4.68E-01 | - | - | - |
| Pentadecanoic acid intake (g/day) | Nutrients | 29676 | -0.03 | 0.02 | 3.82E-02 | 5.72E-02 | 190 | 28910 | -0.05 | 0.02 | 1.78E-03 | **3.38E-03** | 149 | -0.04 | 0.01 | 2.35E-04 | 0.00 | 4.68E-01 | - | - | - |
| Soft cheese | Food | 7086 | 0.09 | 0.04 | 3.82E-02 | 5.72E-02 | 191 | 6838 | 0.12 | 0.04 | 3.53E-02 | 5.30E-02 | 189 | 0.10 | 0.03 | 1.67E-03 | 0.00 | 6.59E-01 | - | - | - |
| Time spent in a week in moderately steneous activities | Physical activity | 11084 | -0.58 | 0.04 | 4.21E-02 | 6.28E-02 | 192 | 10891 | -0.59 | 0.04 | 4.27E-02 | 6.38E-02 | 190 | -0.58 | 0.03 | - | 0.00 | 9.17E-01 | - | - | - |
| Vitamin B2 intake (ug/day) | Nutrients | 29676 | 0.03 | 0.02 | 4.50E-02 | 6.67E-02 | 193 | 28910 | 0.04 | 0.02 | 2.20E-02 | **3.44E-02** | 182 | 0.03 | 0.01 | 2.39E-03 | 0.00 | 8.41E-01 | - | - | - |
| Blota (broth + bread) | Food | 7086 | 0.09 | 0.04 | 5.16E-02 | 7.60E-02 | 194 | 6838 | 0.09 | 0.04 | 8.96E-02 | 1.21E-01 | 210 | 0.09 | 0.03 | 7.38E-03 | 0.00 | 9.96E-01 | - | - | - |
| Zinc intake (mg/day) | Nutrients | 29676 | 0.03 | 0.02 | 5.20E-02 | 7.62E-02 | 195 | 28910 | 0.06 | 0.02 | 2.23E-04 | **4.67E-04** | 136 | 0.04 | 0.01 | 6.96E-05 | 0.37 | 2.09E-01 | - | - | - |
| Amount of alcohol drunk in a day | Alcohol | 10956 | 0.49 | 0.04 | 5.33E-02 | 7.77E-02 | 196 | 10757 | 0.49 | 0.04 | 1.88E-30 | **2.05E-29** | 26 | 0.49 | 0.03 | - | 0.00 | 9.05E-01 | - | - | - |
| Do you feel important and appreciated outside your home? | Psychosocial | 24614 | -0.04 | 0.02 | 5.49E-02 | 7.96E-02 | 197 | 24192 | -0.02 | 0.02 | 2.40E-01 | 3.01E-01 | 226 | -0.03 | 0.01 | 2.87E-02 | 0.00 | 5.96E-01 | - | - | - |
| Lean fish (e.g. perch, bass, cod) | Food | 29676 | 0.03 | 0.01 | 5.51E-02 | 7.96E-02 | 198 | 28910 | 0.02 | 0.01 | 1.82E-01 | 2.35E-01 | 220 | 0.02 | 0.01 | 2.15E-02 | 0.00 | 6.75E-01 | - | - | - |
| Frequency of alcohol consumption | Alcohol | 11473 | -0.40 | 0.04 | 6.14E-02 | 8.83E-02 | 199 | 11326 | -0.41 | 0.04 | 6.14E-02 | 8.72E-02 | 200 | -0.40 | 0.03 | - | 0.00 | 8.73E-01 | - | - | - |
| Frequency of drinking six or more glasses at the same occasion | Alcohol | 11162 | 0.42 | 0.04 | 6.35E-02 | 9.09E-02 | 200 | 10971 | 0.37 | 0.04 | 7.43E-02 | 1.04E-01 | 202 | 0.39 | 0.03 | - | 0.00 | 3.93E-01 | - | - | - |
| Marital status: Single vs Married/partner | Social | 31026 | -0.11 | 0.06 | 7.30E-02 | 1.04E-01 | 201 | 30346 | 0.00 | 0.06 | 9.35E-01 | 9.41E-01 | 282 | -0.06 | 0.04 | 1.85E-01 | 0.32 | 2.26E-01 | - | - | - |
| People to ask for help apart from the ones at home | Social | 33705 | -0.08 | 0.05 | 7.59E-02 | 1.07E-01 | 202 | 32961 | 0.03 | 0.05 | 5.50E-01 | 6.13E-01 | 255 | -0.03 | 0.03 | 4.01E-01 | 0.64 | 9.41E-02 | - | - | - |
| Potato salad | Food | 5678 | 0.09 | 0.05 | 8.15E-02 | 1.15E-01 | 203 | 5481 | -0.01 | 0.05 | 8.03E-01 | 8.38E-01 | 272 | 0.04 | 0.03 | 2.63E-01 | 0.55 | 1.35E-01 | - | - | - |
| Phosphate intake (mg/day) | Nutrients | 29676 | 0.03 | 0.01 | 8.22E-02 | 1.15E-01 | 204 | 28910 | 0.03 | 0.01 | 4.44E-02 | 6.61E-02 | 191 | 0.03 | 0.01 | 8.03E-03 | 0.00 | 8.54E-01 | - | - | - |
| Appetite status | Psychosocial | 24605 | 0.03 | 0.02 | 8.37E-02 | 1.17E-01 | 205 | 24172 | 0.04 | 0.02 | 2.19E-02 | **3.43E-02** | 181 | 0.04 | 0.01 | 4.45E-03 | 0.00 | 6.93E-01 | - | - | - |
| Whole grain crisp bread | Food | 29676 | -0.03 | 0.02 | 8.66E-02 | 1.20E-01 | 206 | 28910 | -0.05 | 0.02 | 1.39E-03 | **2.66E-03** | 148 | -0.04 | 0.01 | 5.17E-04 | 0.10 | 2.93E-01 | - | - | - |
| Rice | Food | 29676 | 0.03 | 0.02 | 9.73E-02 | 1.34E-01 | 207 | 28910 | -0.02 | 0.01 | 1.84E-01 | 2.37E-01 | 221 | 0.00 | 0.01 | 8.29E-01 | 0.78 | 3.46E-02 | - | - | - |
| Risk of sleeping while watching TV | Sleep | 11330 | 0.22 | 0.04 | 1.11E-01 | 1.52E-01 | 208 | 11183 | 0.31 | 0.04 | 8.00E-02 | 1.10E-01 | 206 | 0.27 | 0.03 | - | 0.64 | 9.76E-02 | - | - | - |
| Light beer | Alcohol | 29676 | -0.02 | 0.02 | 1.11E-01 | 1.52E-01 | 209 | 28910 | -0.04 | 0.02 | 1.08E-02 | **1.79E-02** | 171 | -0.03 | 0.01 | 3.48E-03 | 0.00 | 4.75E-01 | - | - | - |
| Soft whey cheese | Food | 7086 | -0.07 | 0.04 | 1.13E-01 | 1.54E-01 | 210 | 6838 | -0.11 | 0.04 | 4.64E-02 | 6.82E-02 | 193 | -0.09 | 0.03 | 6.36E-03 | 0.00 | 4.89E-01 | - | - | - |
| Frequency of shoverling snow during leisure time | Physical activity | 20940 | -0.03 | 0.02 | 1.42E-01 | 1.93E-01 | 211 | 20378 | 0.00 | 0.02 | 9.59E-01 | 9.59E-01 | 284 | -0.02 | 0.02 | 3.17E-01 | 0.14 | 2.82E-01 | - | - | - |
| Liver, kidney | Food | 7086 | 0.06 | 0.04 | 1.44E-01 | 1.94E-01 | 212 | 6838 | 0.05 | 0.04 | 2.64E-01 | 3.25E-01 | 231 | 0.06 | 0.03 | 6.15E-02 | 0.00 | 8.51E-01 | - | - | - |
| Risk of sleeping while sitting still after having lunch | Sleep | 11290 | 0.17 | 0.04 | 1.46E-01 | 1.96E-01 | 213 | 11158 | 0.21 | 0.04 | 2.23E-07 | **6.10E-07** | 104 | 0.19 | 0.03 | - | 0.00 | 4.84E-01 | - | - | - |
| Confidence status | Psychosocial | 24582 | -0.03 | 0.02 | 1.54E-01 | 2.06E-01 | 214 | 24138 | -0.03 | 0.02 | 1.31E-01 | 1.72E-01 | 216 | -0.03 | 0.01 | 3.80E-02 | 0.00 | 9.38E-01 | - | - | - |
| Linolenic acid intake (g/day) | Nutrients | 29676 | 0.02 | 0.01 | 1.69E-01 | 2.25E-01 | 215 | 28910 | 0.00 | 0.01 | 7.44E-01 | 7.97E-01 | 264 | 0.01 | 0.01 | 2.29E-01 | 0.00 | 4.57E-01 | - | - | - |
| Cheese 28% | Food | 29676 | -0.02 | 0.01 | 1.77E-01 | 2.34E-01 | 216 | 28910 | -0.02 | 0.01 | 1.51E-01 | 1.96E-01 | 219 | -0.02 | 0.01 | 4.89E-02 | 0.00 | 9.52E-01 | - | - | - |
| Satisfaction with work situation | Psychosocial | 24304 | -0.02 | 0.02 | 1.78E-01 | 2.34E-01 | 217 | 23862 | -0.06 | 0.02 | 2.86E-03 | **5.18E-03** | 157 | -0.04 | 0.01 | 2.19E-03 | 0.25 | 2.48E-01 | - | - | - |
| Self-employed | Psychosocial | 28859 | 0.08 | 0.06 | 1.91E-01 | 2.50E-01 | 218 | 28081 | 0.02 | 0.06 | 7.95E-01 | 8.38E-01 | 269 | 0.05 | 0.04 | 2.67E-01 | 0.00 | 4.59E-01 | - | - | - |
| Number of friends that can come to your home at any time and feel at home | Social | 33337 | 0.02 | 0.02 | 2.11E-01 | 2.76E-01 | 219 | 32601 | 0.04 | 0.02 | 2.47E-02 | **3.83E-02** | 183 | 0.03 | 0.01 | 1.38E-02 | 0.00 | 4.60E-01 | - | - | - |
| Strong beer | Alcohol | 29676 | 0.02 | 0.02 | 2.12E-01 | 2.76E-01 | 220 | 28910 | 0.00 | 0.02 | 9.02E-01 | 9.24E-01 | 277 | 0.01 | 0.01 | 3.38E-01 | 0.00 | 4.18E-01 | - | - | - |
| Risk of sleeping while lying down resting in the afternoon | Sleep | 11282 | 0.10 | 0.04 | 2.35E-01 | 3.04E-01 | 221 | 11152 | 0.11 | 0.04 | 5.37E-03 | **9.24E-03** | 165 | 0.11 | 0.03 | - | 0.00 | 8.59E-01 | - | - | - |
| Risk of sleeping while sitting inactive in a public place | Sleep | 11252 | 0.10 | 0.04 | 2.40E-01 | 3.09E-01 | 222 | 11111 | 0.21 | 0.04 | - | - | 286 | 0.15 | 0.03 | - | 0.76 | 4.24E-02 | - | - | - |
| Support from others | Social | 33829 | -0.02 | 0.01 | 2.51E-01 | 3.22E-01 | 223 | 33088 | -0.04 | 0.01 | 4.92E-03 | **8.58E-03** | 163 | -0.03 | 0.01 | 5.33E-03 | 0.32 | 2.27E-01 | - | - | - |
| Cheese 10-17% | Food | 29676 | -0.02 | 0.01 | 2.53E-01 | 3.24E-01 | 224 | 28910 | 0.00 | 0.01 | 7.93E-01 | 8.38E-01 | 268 | -0.01 | 0.01 | 5.35E-01 | 0.00 | 3.20E-01 | - | - | - |
| Would you say that the number of people that you meet in your everyday life is enough or would you like to meet more or fewer people? | Social | 33799 | -0.02 | 0.01 | 2.55E-01 | 3.24E-01 | 225 | 33065 | -0.04 | 0.01 | 1.23E-03 | **2.38E-03** | 147 | -0.03 | 0.01 | 2.05E-03 | 0.55 | 1.35E-01 | - | - | - |
| Disaccharides intake (g/day) | Nutrients | 29676 | -0.02 | 0.02 | 2.57E-01 | 3.26E-01 | 226 | 28910 | -0.05 | 0.02 | 3.83E-03 | **6.78E-03** | 160 | -0.03 | 0.01 | 4.39E-03 | 0.35 | 2.16E-01 | - | - | - |
| Cohabitation: Live alone vs Adult and children | Social | 19514 | -0.08 | 0.07 | 2.59E-01 | 3.27E-01 | 227 | 19042 | 0.07 | 0.07 | 2.94E-01 | 3.57E-01 | 234 | 0.00 | 0.05 | 9.59E-01 | 0.58 | 1.24E-01 | - | - | - |
| Satisfaction with accomodation | Psychosocial | 24625 | 0.02 | 0.02 | 2.72E-01 | 3.41E-01 | 228 | 24199 | 0.02 | 0.02 | 3.37E-01 | 4.03E-01 | 237 | 0.02 | 0.01 | 1.45E-01 | 0.00 | 9.21E-01 | - | - | - |
| Sweets | Food | 29676 | 0.02 | 0.02 | 3.00E-01 | 3.75E-01 | 229 | 28910 | 0.01 | 0.02 | 3.81E-01 | 4.43E-01 | 244 | 0.01 | 0.01 | 1.76E-01 | 0.00 | 9.04E-01 | - | - | - |
| Cohabitation: Live alone vs Only children | Social | 4936 | -0.12 | 0.12 | 3.04E-01 | 3.77E-01 | 230 | 4933 | -0.21 | 0.11 | 5.95E-02 | 8.53E-02 | 198 | -0.17 | 0.08 | 3.85E-02 | 0.00 | 5.67E-01 | - | - | - |
| Cohabitation: Live alone vs Only one adult (spouse, partner) | Social | 15246 | -0.07 | 0.07 | 3.04E-01 | 3.77E-01 | 231 | 15091 | 0.07 | 0.07 | 3.51E-01 | 4.14E-01 | 241 | 0.00 | 0.05 | 9.45E-01 | 0.48 | 1.66E-01 | - | - | - |
| Ingenuity or creativity demand from job | Psychosocial | 33179 | 0.02 | 0.02 | 3.14E-01 | 3.87E-01 | 232 | 32511 | -0.03 | 0.02 | 8.47E-02 | 1.15E-01 | 209 | -0.01 | 0.01 | 6.24E-01 | 0.73 | 5.30E-02 | - | - | - |
| Marital status: Single vs Widow/widower | Social | 3588 | -0.27 | 0.27 | 3.20E-01 | 3.93E-01 | 233 | 3540 | -0.21 | 0.26 | 4.19E-01 | 4.85E-01 | 245 | -0.24 | 0.19 | 2.03E-01 | 0.00 | 8.90E-01 | - | - | - |
| Feel uneasy or guilty because of your way of drinking | Alcohol | 19472 | -0.07 | 0.07 | 3.39E-01 | 4.15E-01 | 234 | 19045 | -0.07 | 0.07 | 2.96E-01 | 3.58E-01 | 235 | -0.07 | 0.05 | 1.57E-01 | 0.00 | 9.52E-01 | - | - | - |
| Sour milk, yoghurt (low fat) | Food | 29676 | -0.01 | 0.01 | 3.49E-01 | 4.24E-01 | 235 | 28910 | -0.01 | 0.01 | 5.74E-01 | 6.35E-01 | 257 | -0.01 | 0.01 | 2.89E-01 | 0.00 | 7.89E-01 | - | - | - |
| Last time a colleague visited you at home | Psychosocial | 31751 | 0.01 | 0.01 | 3.51E-01 | 4.25E-01 | 236 | 31081 | 0.01 | 0.01 | 3.75E-01 | 4.40E-01 | 242 | 0.01 | 0.01 | 1.98E-01 | 0.00 | 9.82E-01 | - | - | - |
| Corn flakes | Food | 29676 | 0.01 | 0.01 | 3.56E-01 | 4.29E-01 | 237 | 28910 | 0.01 | 0.01 | 4.92E-01 | 5.59E-01 | 250 | 0.01 | 0.01 | 2.54E-01 | 0.00 | 8.71E-01 | - | - | - |
| Smoking status: Former occasional smokers vs non-smokers | Tobacco use | 20016 | -0.06 | 0.07 | 3.61E-01 | 4.33E-01 | 238 | 19568 | -0.10 | 0.07 | 1.38E-01 | 1.81E-01 | 217 | -0.08 | 0.05 | 8.92E-02 | 0.00 | 7.02E-01 | - | - | - |
| Light but partly physically active work | Physical activity | 32431 | 0.03 | 0.03 | 3.71E-01 | 4.45E-01 | 239 | 31717 | 0.06 | 0.03 | 7.90E-02 | 1.09E-01 | 205 | 0.05 | 0.02 | 6.18E-02 | 0.00 | 5.29E-01 | - | - | - |
| Risk of sleeping as a passenger in a car for one hour without break | Sleep | 11214 | 0.06 | 0.04 | 3.75E-01 | 4.47E-01 | 240 | 11066 | 0.09 | 0.04 | - | - | 287 | 0.07 | 0.03 | - | 0.00 | 5.66E-01 | - | - | - |
| Ice cream | Food | 29676 | 0.01 | 0.01 | 3.85E-01 | 4.57E-01 | 241 | 28910 | 0.01 | 0.01 | 3.44E-01 | 4.07E-01 | 240 | 0.01 | 0.01 | 2.00E-01 | 0.00 | 9.45E-01 | - | - | - |
| Contradictory demands in job | Psychosocial | 33110 | -0.01 | 0.01 | 3.94E-01 | 4.66E-01 | 242 | 32405 | 0.01 | 0.01 | 3.79E-01 | 4.43E-01 | 243 | 0.00 | 0.01 | 9.88E-01 | 0.33 | 2.21E-01 | - | - | - |
| Feel the need to reduce alcohol consumption | Alcohol | 30342 | -0.04 | 0.05 | 4.13E-01 | 4.86E-01 | 243 | 29717 | 0.07 | 0.05 | 1.23E-01 | 1.63E-01 | 214 | 0.02 | 0.03 | 6.14E-01 | 0.64 | 9.42E-02 | - | - | - |
| Bregott on bread | Food | 29676 | -0.01 | 0.02 | 4.20E-01 | 4.90E-01 | 244 | 28910 | -0.01 | 0.02 | 6.07E-01 | 6.65E-01 | 259 | -0.01 | 0.01 | 3.50E-01 | 0.00 | 8.37E-01 | - | - | - |
| Cookies, pastry | Food | 29676 | -0.01 | 0.02 | 4.20E-01 | 4.90E-01 | 245 | 28910 | -0.03 | 0.02 | 8.19E-02 | 1.12E-01 | 207 | -0.02 | 0.01 | 7.17E-02 | 0.00 | 5.10E-01 | - | - | - |
| Risk of sleeping while sitting and reading | Sleep | 11207 | 0.05 | 0.04 | 4.26E-01 | 4.95E-01 | 246 | 11065 | 0.11 | 0.04 | 6.36E-03 | **1.08E-02** | 167 | 0.08 | 0.03 | - | 0.10 | 2.91E-01 | - | - | - |
| Butter on bread | Food | 29676 | 0.01 | 0.01 | 4.30E-01 | 4.98E-01 | 247 | 28910 | -0.02 | 0.01 | 1.00E-01 | 1.35E-01 | 211 | -0.01 | 0.01 | 5.35E-01 | 0.66 | 8.61E-02 | - | - | - |
| Times during last year that you felt guilty because of your drinking | Alcohol | 11149 | -0.05 | 0.04 | 4.33E-01 | 4.99E-01 | 248 | 10944 | -0.03 | 0.04 | - | - | 285 | -0.04 | 0.03 | - | 0.00 | 7.81E-01 | - | - | - |
| Control over planning and execution of the workday | Psychosocial | 33348 | 0.01 | 0.01 | 4.63E-01 | 5.30E-01 | 249 | 32656 | 0.02 | 0.01 | 1.43E-01 | 1.86E-01 | 218 | 0.02 | 0.01 | 1.20E-01 | 0.00 | 6.01E-01 | - | - | - |
| High physical demand from job | Physical activity | 33405 | 0.01 | 0.02 | 4.63E-01 | 5.30E-01 | 250 | 32738 | 0.01 | 0.02 | 7.13E-01 | 7.76E-01 | 261 | 0.01 | 0.01 | 4.36E-01 | 0.00 | 7.95E-01 | - | - | - |
| Enough time for job assignments | Psychosocial | 33179 | -0.01 | 0.01 | 4.68E-01 | 5.33E-01 | 251 | 32516 | -0.01 | 0.01 | 5.87E-01 | 6.46E-01 | 258 | -0.01 | 0.01 | 3.69E-01 | 0.00 | 9.03E-01 | - | - | - |
| Meat on bread | Food | 29676 | 0.01 | 0.01 | 4.89E-01 | 5.55E-01 | 252 | 28910 | 0.01 | 0.01 | 5.46E-01 | 6.10E-01 | 254 | 0.01 | 0.01 | 3.60E-01 | 0.00 | 9.45E-01 | - | - | - |
| Fatty fish (e.g. herring, whitefish, salmon) | Food | 29676 | 0.01 | 0.01 | 5.01E-01 | 5.67E-01 | 253 | 28910 | 0.03 | 0.01 | 7.49E-02 | 1.05E-01 | 203 | 0.02 | 0.01 | 8.24E-02 | 0.00 | 4.36E-01 | - | - | - |
| White (soft) bread, thin crisp bread | Food | 29676 | 0.01 | 0.01 | 5.06E-01 | 5.70E-01 | 254 | 28910 | 0.03 | 0.01 | 2.94E-02 | **4.49E-02** | 186 | 0.02 | 0.01 | 4.40E-02 | 0.11 | 2.88E-01 | - | - | - |
| Cohabitation: Live alone vs Other/others | Social | 3844 | -0.12 | 0.19 | 5.19E-01 | 5.82E-01 | 255 | 3844 | 0.30 | 0.18 | 1.04E-01 | 1.39E-01 | 212 | 0.09 | 0.13 | 4.74E-01 | 0.61 | 1.10E-01 | - | - | - |
| Learn new things at job | Psychosocial | 33267 | -0.01 | 0.01 | 5.39E-01 | 6.02E-01 | 256 | 32589 | -0.03 | 0.01 | 1.86E-02 | **2.99E-02** | 177 | -0.02 | 0.01 | 3.64E-02 | 0.35 | 2.15E-01 | - | - | - |
| Calcium intake (mg/day) | Nutrients | 29676 | -0.01 | 0.02 | 5.60E-01 | 6.23E-01 | 257 | 28910 | -0.02 | 0.02 | 1.29E-01 | 1.70E-01 | 215 | -0.02 | 0.01 | 1.37E-01 | 0.00 | 5.07E-01 | - | - | - |
| Blood based food | Food | 7086 | 0.02 | 0.04 | 5.89E-01 | 6.52E-01 | 258 | 6838 | -0.01 | 0.04 | 7.50E-01 | 7.97E-01 | 267 | 0.00 | 0.03 | 8.78E-01 | 0.00 | 5.30E-01 | - | - | - |
| Sometimes physically straining work | Physical activity | 32431 | -0.02 | 0.03 | 5.95E-01 | 6.57E-01 | 259 | 31717 | -0.02 | 0.03 | 5.68E-01 | 6.30E-01 | 256 | -0.02 | 0.02 | 4.36E-01 | 0.00 | 9.76E-01 | - | - | - |
| Close relationship with anyone | Social | 33420 | -0.01 | 0.01 | 6.15E-01 | 6.77E-01 | 260 | 32655 | 0.01 | 0.01 | 5.13E-01 | 5.80E-01 | 251 | 0.00 | 0.01 | 9.13E-01 | 0.00 | 4.13E-01 | - | - | - |
| Breakfast habits: Only coffee/tea for breakfast vs not breakfast at all | Food | 1914 | -0.08 | 0.17 | 6.34E-01 | 6.93E-01 | 261 | 1997 | -0.63 | 0.18 | 7.27E-04 | **1.45E-03** | 142 | -0.34 | 0.13 | 7.57E-03 | 0.78 | 3.12E-02 | - | - | - |
| Oil for cooking | Food | 29676 | -0.01 | 0.01 | 6.35E-01 | 6.93E-01 | 262 | 28910 | -0.03 | 0.01 | 7.76E-02 | 1.08E-01 | 204 | -0.02 | 0.01 | 1.12E-01 | 0.00 | 3.66E-01 | - | - | - |
| Number of people with whom you can speak openly | Social | 33786 | -0.01 | 0.02 | 6.63E-01 | 7.19E-01 | 263 | 33041 | 0.02 | 0.02 | 3.05E-01 | 3.67E-01 | 236 | 0.00 | 0.01 | 6.84E-01 | 0.07 | 2.99E-01 | - | - | - |
| Trans fat intake (g/day) | Nutrients | 29676 | -0.01 | 0.02 | 6.64E-01 | 7.19E-01 | 264 | 28910 | -0.02 | 0.02 | 2.70E-01 | 3.29E-01 | 233 | -0.02 | 0.01 | 2.76E-01 | 0.00 | 6.37E-01 | - | - | - |
| Butter for cooking | Food | 29676 | -0.01 | 0.01 | 6.70E-01 | 7.23E-01 | 265 | 28910 | 0.00 | 0.01 | 7.97E-01 | 8.38E-01 | 270 | 0.00 | 0.01 | 6.30E-01 | 0.00 | 9.01E-01 | - | - | - |
| High mental demand from job | Psychosocial | 33075 | -0.01 | 0.01 | 6.79E-01 | 7.30E-01 | 266 | 32445 | 0.00 | 0.02 | 8.45E-01 | 8.76E-01 | 274 | 0.00 | 0.01 | 8.74E-01 | 0.00 | 6.67E-01 | - | - | - |
| Vision status | General health | 24600 | -0.01 | 0.02 | 6.92E-01 | 7.41E-01 | 267 | 24149 | 0.00 | 0.02 | 8.35E-01 | 8.68E-01 | 273 | 0.00 | 0.01 | 8.95E-01 | 0.00 | 6.69E-01 | - | - | - |
| Linoleic acid intake (g/day) | Nutrients | 29676 | 0.01 | 0.01 | 6.98E-01 | 7.44E-01 | 268 | 28910 | -0.01 | 0.01 | 7.30E-01 | 7.91E-01 | 262 | 0.00 | 0.01 | 9.76E-01 | 0.00 | 6.04E-01 | - | - | - |
| Number of snuff boxes per week | Tobacco use | 32580 | -0.01 | 0.02 | 6.99E-01 | 7.44E-01 | 269 | 31895 | -0.01 | 0.02 | 7.44E-01 | 7.97E-01 | 265 | -0.01 | 0.01 | 6.14E-01 | 0.00 | 9.73E-01 | - | - | - |
| Enterolactone intake (ug/day) | Nutrients | 29676 | -0.01 | 0.01 | 7.29E-01 | 7.73E-01 | 270 | 28910 | -0.01 | 0.01 | 5.29E-01 | 5.96E-01 | 252 | -0.01 | 0.01 | 4.90E-01 | 0.00 | 8.43E-01 | - | - | - |
| Shellfish (e.g. shrimps, scallops) | Food | 7086 | 0.01 | 0.04 | 7.44E-01 | 7.86E-01 | 271 | 6838 | 0.03 | 0.04 | 4.88E-01 | 5.57E-01 | 249 | 0.02 | 0.03 | 4.55E-01 | 0.00 | 7.64E-01 | - | - | - |
| Tiamin intake (mg/day) | Nutrients | 29676 | 0.00 | 0.02 | 7.70E-01 | 8.10E-01 | 272 | 28910 | 0.00 | 0.02 | 9.11E-01 | 9.31E-01 | 278 | 0.00 | 0.01 | 7.75E-01 | 0.00 | 9.00E-01 | - | - | - |
| Campesterol intake (mg/day) | Nutrients | 29676 | 0.00 | 0.01 | 7.75E-01 | 8.12E-01 | 273 | 28910 | -0.02 | 0.01 | 2.47E-01 | 3.08E-01 | 228 | -0.01 | 0.01 | 3.07E-01 | 0.00 | 5.39E-01 | - | - | - |
| Number of cigars smoked per day | Tobacco use | 21233 | -0.01 | 0.02 | 7.85E-01 | 8.19E-01 | 274 | 20767 | 0.00 | 0.02 | 8.90E-01 | 9.19E-01 | 275 | 0.00 | 0.02 | 9.27E-01 | 0.00 | 7.70E-01 | - | - | - |
| Iron intake (mg/day) | Nutrients | 29676 | 0.00 | 0.02 | 8.20E-01 | 8.52E-01 | 275 | 28910 | 0.03 | 0.02 | 6.25E-02 | 8.83E-02 | 201 | 0.01 | 0.01 | 2.47E-01 | 0.54 | 1.40E-01 | - | - | - |
| Formic acid intaje (g/day) | Nutrients | 29676 | 0.00 | 0.02 | 8.40E-01 | 8.68E-01 | 276 | 28910 | -0.01 | 0.02 | 7.02E-01 | 7.67E-01 | 260 | 0.00 | 0.01 | 6.79E-01 | 0.00 | 9.00E-01 | - | - | - |
| Smoking status: Smokers vs non-smokers | Tobacco use | 21577 | -0.02 | 0.08 | 8.41E-01 | 8.68E-01 | 277 | 21200 | -0.06 | 0.08 | 4.61E-01 | 5.30E-01 | 247 | -0.04 | 0.06 | 5.06E-01 | 0.00 | 7.06E-01 | - | - | - |
| Frequency of engaging in clubs, associations or study circles | Social | 21389 | 0.00 | 0.02 | 8.46E-01 | 8.70E-01 | 278 | 20833 | -0.02 | 0.02 | 3.41E-01 | 4.06E-01 | 239 | -0.01 | 0.01 | 4.27E-01 | 0.00 | 5.77E-01 | - | - | - |
| Satisfaction with home and family situation | Psychosocial | 24603 | 0.00 | 0.02 | 8.57E-01 | 8.79E-01 | 279 | 24180 | 0.04 | 0.02 | 5.91E-02 | 8.52E-02 | 197 | 0.02 | 0.01 | 1.45E-01 | 0.32 | 2.25E-01 | - | - | - |
| Number of social contacts with the same interests as you | Social | 33748 | 0.00 | 0.02 | 8.62E-01 | 8.80E-01 | 280 | 33000 | 0.03 | 0.02 | 8.28E-02 | 1.13E-01 | 208 | 0.02 | 0.01 | 1.81E-01 | 0.20 | 2.63E-01 | - | - | - |

| | | | | | | | | | | | | | | | | | | | | | |
|---|---|---|---|---|---|---|---|---|---|---|---|---|---|---|---|---|---|---|---|---|---|
| Vitamin E intake (mg/day) | Nutrients | 29676 | 0.00 | 0.01 | 8.78E-01 | 8.94E-01 | 281 | 28910 | 0.02 | 0.01 | 2.45E-01 | 3.07E-01 | 227 | 0.01 | 0.01 | 3.51E-01 | 0.00 | 4.78E-01 | - | - | - |
| Chips, popcorn, salted nuts | Food | 29676 | 0.00 | 0.02 | 9.03E-01 | 9.16E-01 | 282 | 28910 | 0.00 | 0.02 | 9.01E-01 | 9.24E-01 | 276 | 0.00 | 0.01 | 8.62E-01 | 0.00 | 9.98E-01 | - | - | - |
| Memory status | Psychosocial | 24562 | 0.00 | 0.02 | 9.18E-01 | 9.28E-01 | 283 | 24114 | 0.02 | 0.02 | 2.62E-01 | 3.23E-01 | 230 | 0.01 | 0.01 | 3.86E-01 | 0.00 | 4.72E-01 | - | - | - |
| Possibility to leave your work for a while to speak with a colleague | Psychosocial | 31894 | 0.00 | 0.02 | 9.31E-01 | 9.37E-01 | 284 | 31214 | -0.02 | 0.02 | 1.86E-01 | 2.37E-01 | 222 | -0.01 | 0.01 | 3.22E-01 | 0.00 | 3.77E-01 | - | - | - |
| Potassium intake (mg/day) | Nutrients | 29676 | 0.00 | 0.02 | 9.41E-01 | 9.45E-01 | 285 | 28910 | -0.01 | 0.02 | 4.20E-01 | 4.85E-01 | 246 | -0.01 | 0.01 | 5.34E-01 | 0.00 | 6.05E-01 | - | - | - |
| Margarine on bread | Food | 29676 | 0.00 | 0.01 | 9.64E-01 | 9.64E-01 | 286 | 28910 | 0.00 | 0.01 | 9.18E-01 | 9.31E-01 | 280 | 0.00 | 0.01 | 9.16E-01 | 0.00 | 9.67E-01 | - | - | - |
| Breath-holds during sleep | Sleep | 7930 | 0.70 | 0.05 | - | - | 287 | 7771 | 0.80 | 0.05 | 5.83E-65 | **1.38E-63** | 12 | 0.75 | 0.03 | - | 0.56 | 1.34E-01 | - | - | - |

**Supplementary Table 4**. Linear mixed model association results for SBP

| | | Training set | | | | | | Testing set | | | | | | Metanalysis | | | | | R2 | | |
|---|---|---|---|---|---|---|---|---|---|---|---|---|---|---|---|---|---|---|---|---|---|
| **Description** | **Group** | **N** | **Effect estimate** | **S.E.** | **p-value** | **p-value$_{FDR}$** | **p-value rank** | **N** | **Effect estimate** | **S.E.** | **p-value** | **p-value$_{FDR}$** | **p-value rank** | **Effect estimate** | **S.E.** | **p-value** | **$I^2$** | **Q p-value** | **marginal R2** | **conditional R2** | **R2 rank** |
| Informed of having high blood pressure | General health | 33454 | 15.96 | 0.19 | <2.22E-308 | **<2.22E-308** | 1 | 32729 | 16.07 | 0.20 | <2.22E-308 | **<2.22E-308** | 1 | 16.01 | 0.14 | <2.22E-308 | 0.00 | 6.87E-01 | 0.35 | 0.59 | 1 |
| Educational level | Psychosocial | 33621 | -0.93 | 0.10 | 1.18E-19 | **1.62E-17** | 2 | 32904 | -1.06 | 0.11 | 1.17E-23 | **1.63E-21** | 2 | -0.99 | 0.07 | 1.69E-41 | 0.00 | 3.95E-01 | 0.21 | 0.61 | 4 |
| Self rate of overall health | General health | 14450 | -1.13 | 0.14 | 6.04E-15 | **4.12E-13** | 4 | 14267 | -1.19 | 0.14 | 1.43E-15 | **1.00E-13** | 4 | -1.16 | 0.10 | 6.17E-29 | 0.00 | 7.76E-01 | 0.19 | 0.66 | 44 |
| Parents or siblings had a cerebral hemorrhage/thrombosis or cardiac infarction before the age of 60 | General health | 32696 | 1.57 | 0.23 | 1.11E-11 | **3.80E-10** | 8 | 32026 | 2.02 | 0.23 | 8.14E-18 | **7.57E-16** | 3 | 1.79 | 0.16 | 1.58E-27 | 0.46 | 1.72E-01 | 0.21 | 0.61 | 5 |
| Excellent health | General health | 14395 | -1.10 | 0.14 | 2.62E-14 | **1.43E-12** | 5 | 14209 | -1.12 | 0.14 | 3.30E-14 | **1.84E-12** | 5 | -1.11 | 0.10 | 5.81E-27 | 0.00 | 9.15E-01 | 0.19 | 0.66 | 45 |
| Overall state of health during the last year | General health | 33274 | -0.67 | 0.08 | 2.95E-16 | **2.68E-14** | 3 | 32554 | -0.57 | 0.08 | 1.50E-11 | **4.65E-10** | 9 | -0.62 | 0.06 | 4.36E-26 | 0.00 | 3.78E-01 | 0.21 | 0.61 | 9 |
| Smoking status: Smokers vs non-smokers | Tobacco use | 21442 | -2.24 | 0.30 | 4.41E-14 | **2.01E-12** | 6 | 21080 | -2.20 | 0.30 | 1.82E-13 | **8.49E-12** | 6 | -2.22 | 0.21 | 5.48E-26 | 0.00 | 9.21E-01 | 0.20 | 0.61 | 31 |
| Marital status: Single vs Married/partner | Social | 30840 | -1.77 | 0.31 | 9.58E-09 | **2.91E-07** | 9 | 30191 | -2.10 | 0.31 | 1.29E-11 | **4.50E-10** | 8 | -1.93 | 0.22 | 9.36E-19 | 0.00 | 4.51E-01 | 0.21 | 0.61 | 10 |
| Years smoking | Tobacco use | 29331 | -0.75 | 0.11 | 8.68E-12 | **3.39E-10** | 7 | 28570 | -0.61 | 0.11 | 5.94E-08 | **1.38E-06** | 12 | -0.68 | 0.08 | 4.16E-18 | 0.00 | 3.87E-01 | 0.21 | 0.61 | 11 |
| Overall state of health compared to other your age | General health | 16623 | -0.54 | 0.12 | 2.97E-06 | **5.41E-05** | 15 | 16201 | -0.86 | 0.12 | 6.23E-13 | **2.48E-11** | 7 | -0.70 | 0.08 | 5.35E-17 | 0.73 | 5.49E-02 | 0.19 | 0.55 | 43 |
| Cohabitation: Live alone vs Adult and children | Social | 19387 | -1.77 | 0.31 | 1.24E-08 | **3.39E-07** | 10 | 18932 | -1.56 | 0.32 | 7.29E-07 | **1.45E-05** | 14 | -1.67 | 0.22 | 4.97E-14 | 0.00 | 6.37E-01 | 0.20 | 0.59 | 35 |
| Repetitive job | Psychosocial | 33085 | 0.34 | 0.09 | 8.57E-05 | **8.66E-04** | 27 | 32435 | 0.59 | 0.09 | 2.62E-11 | **7.30E-10** | 10 | 0.46 | 0.06 | 7.47E-14 | 0.75 | 4.67E-02 | 0.20 | 0.61 | 14 |
| Marital status: Single vs Divorced/separated | Social | 5519 | -2.30 | 0.47 | 8.69E-07 | **1.69E-05** | 14 | 5525 | -2.30 | 0.47 | 9.61E-07 | **1.67E-05** | 16 | -2.30 | 0.33 | 3.83E-12 | 0.00 | 9.93E-01 | 0.22 | 0.59 | 3 |
| As healthy as anyone | General health | 14352 | -0.55 | 0.14 | 6.88E-05 | **7.23E-04** | 26 | 14164 | -0.81 | 0.14 | 2.02E-08 | **5.12E-07** | 11 | -0.68 | 0.10 | 1.22E-11 | 0.39 | 2.00E-01 | 0.19 | 0.66 | 46 |
| Brewed (filtered) coffee | Beverage | 29503 | -0.47 | 0.09 | 1.97E-07 | **4.88E-06** | 11 | 28758 | -0.41 | 0.09 | 1.30E-05 | **1.74E-04** | 20 | -0.44 | 0.06 | 1.30E-11 | 0.00 | 6.08E-01 | 0.20 | 0.62 | 21 |
| Whole grain crisp bread | Food | 29503 | 0.37 | 0.09 | 2.76E-05 | **3.64E-04** | 20 | 28758 | 0.45 | 0.09 | 8.60E-07 | **1.60E-05** | 15 | 0.41 | 0.06 | 1.21E-10 | 0.00 | 5.55E-01 | 0.20 | 0.62 | 20 |
| Smoking status: Former occasional smokers vs non-smokers | Tobacco use | 19897 | -1.30 | 0.31 | 3.38E-05 | **4.19E-04** | 22 | 19463 | -1.49 | 0.31 | 2.02E-06 | **3.32E-05** | 17 | -1.40 | 0.22 | 3.13E-10 | 0.00 | 6.72E-01 | 0.20 | 0.62 | 30 |
| Smoking status: Former smokers vs non-smokers | Tobacco use | 23995 | -1.32 | 0.26 | 2.74E-07 | **6.24E-06** | 12 | 23218 | -0.97 | 0.27 | 2.81E-04 | **2.38E-03** | 33 | -1.15 | 0.18 | 4.97E-10 | 0.00 | 3.35E-01 | 0.21 | 0.62 | 7 |
| Whole grain intake (g/day) | Food | 29503 | 0.38 | 0.09 | 8.93E-06 | **1.43E-04** | 17 | 28758 | 0.36 | 0.09 | 4.66E-05 | **5.20E-04** | 25 | 0.37 | 0.06 | 1.71E-09 | 0.00 | 8.61E-01 | 0.20 | 0.62 | 22 |
| Frequency of hunting or fishing during leisure time | Physical activity | 20135 | 0.49 | 0.12 | 2.80E-05 | **3.64E-04** | 21 | 19570 | 0.51 | 0.12 | 1.76E-05 | **2.23E-04** | 22 | 0.50 | 0.08 | 2.01E-09 | 0.00 | 8.89E-01 | 0.20 | 0.60 | 34 |
| Participation in associations or voluntary organisations | Social | 33064 | -0.85 | 0.17 | 4.98E-07 | **1.05E-05** | 13 | 32378 | -0.57 | 0.17 | 8.52E-04 | **5.80E-03** | 41 | -0.71 | 0.12 | 3.16E-09 | 0.24 | 2.51E-01 | 0.20 | 0.61 | 13 |
| High physical demand from job | Physical activity | 33213 | 0.36 | 0.09 | 6.45E-05 | **7.04E-04** | 25 | 32573 | 0.39 | 0.09 | 1.31E-05 | **1.74E-04** | 21 | 0.38 | 0.06 | 3.50E-09 | 0.00 | 7.76E-01 | 0.20 | 0.61 | 12 |
| Snuff status: Snuff users vs non-snuff users | Tobacco use | 28567 | -1.25 | 0.28 | 8.88E-06 | **1.43E-04** | 16 | 28056 | -1.14 | 0.29 | 1.03E-04 | **9.87E-04** | 29 | -1.20 | 0.20 | 3.76E-09 | 0.00 | 7.76E-01 | 0.20 | 0.61 | 17 |
| Memory status | Psychosocial | 24481 | 0.45 | 0.10 | 1.47E-05 | **2.22E-04** | 18 | 24035 | 0.42 | 0.11 | 7.39E-05 | **7.64E-04** | 27 | 0.43 | 0.07 | 4.37E-09 | 0.00 | 8.43E-01 | 0.20 | 0.64 | 39 |
| Contradictory demands in job | Psychosocial | 32920 | -0.28 | 0.08 | 7.88E-04 | **6.00E-03** | 35 | 32241 | -0.35 | 0.08 | 3.29E-05 | **4.00E-04** | 23 | -0.31 | 0.06 | 1.12E-07 | 0.00 | 5.49E-01 | 0.20 | 0.61 | 15 |
| High mental demand from job | Psychosocial | 32886 | 0.32 | 0.08 | 1.22E-04 | **1.11E-03** | 30 | 32282 | 0.31 | 0.09 | 3.02E-04 | **2.48E-03** | 34 | 0.31 | 0.06 | 1.34E-07 | 0.00 | 9.05E-01 | 0.20 | 0.61 | 18 |
| Medioresinol intake (ug/day) | Nutrients | 29503 | 0.31 | 0.09 | 2.57E-04 | **2.20E-03** | 32 | 28758 | 0.33 | 0.09 | 1.42E-04 | **1.28E-03** | 31 | 0.32 | 0.06 | 1.34E-07 | 0.00 | 8.69E-01 | 0.20 | 0.62 | 23 |
| Number of snuff boxes per week | Tobacco use | 32391 | -0.40 | 0.10 | 2.28E-05 | **3.27E-04** | 19 | 31732 | -0.31 | 0.10 | 1.58E-03 | **8.66E-03** | 51 | -0.36 | 0.07 | 1.61E-07 | 0.00 | 4.95E-01 | 0.21 | 0.61 | 6 |
| Number of cigarretes smoked per day (in groups) | Tobacco use | 26972 | -0.42 | 0.11 | 6.03E-05 | **6.86E-04** | 24 | 26557 | -0.33 | 0.11 | 1.88E-03 | **9.69E-03** | 54 | -0.38 | 0.07 | 4.66E-07 | 0.00 | 5.48E-01 | 0.20 | 0.61 | 32 |
| Satisfaction with leisure time | Psychosocial | 24490 | 0.40 | 0.10 | 9.76E-05 | **9.18E-04** | 29 | 24059 | 0.30 | 0.10 | 3.71E-03 | **1.64E-02** | 63 | 0.35 | 0.07 | 1.47E-06 | 0.00 | 5.17E-01 | 0.20 | 0.64 | 36 |
| Cohabitation: Live alone vs Only children | Social | 4915 | -1.79 | 0.53 | 7.15E-04 | **5.74E-03** | 34 | 4907 | -1.71 | 0.53 | 1.24E-03 | **7.35E-03** | 46 | -1.75 | 0.37 | 2.91E-06 | 0.00 | 9.05E-01 | 0.23 | 0.59 | 2 |
| Travel to work: Cycle to work vs passive travel to work | Physical activity | 25152 | -0.62 | 0.23 | 7.17E-03 | **2.92E-02** | 67 | 24490 | -0.93 | 0.24 | 8.31E-05 | **8.28E-04** | 28 | -0.77 | 0.17 | 2.99E-06 | 0.00 | 3.45E-01 | 0.20 | 0.61 | 16 |
| Average portion size of meat/fish based on photographic illustration of four sizes (smallest to largest) | Food | 29503 | -0.28 | 0.09 | 2.66E-03 | **1.54E-02** | 47 | 28758 | -0.34 | 0.10 | 4.69E-04 | **3.45E-03** | 38 | -0.31 | 0.07 | 4.39E-06 | 0.00 | 6.77E-01 | 0.20 | 0.62 | 33 |
| Syringaresinol intake (ug/day) | Nutrients | 29503 | 0.27 | 0.09 | 1.80E-03 | **1.22E-02** | 40 | 28758 | 0.29 | 0.09 | 9.91E-04 | **6.43E-03** | 43 | 0.28 | 0.06 | 5.80E-06 | 0.00 | 8.62E-01 | 0.20 | 0.62 | 24 |
| White meat (poultry) | Food | 29503 | -0.27 | 0.09 | 1.85E-03 | **1.22E-02** | 41 | 28758 | -0.30 | 0.09 | 1.04E-03 | **6.62E-03** | 44 | -0.28 | 0.06 | 6.25E-06 | 0.00 | 8.51E-01 | 0.20 | 0.62 | 27 |
| Satisfaction with home and family situation | Psychosocial | 24521 | 0.31 | 0.10 | 1.79E-03 | **1.22E-02** | 39 | 24100 | 0.33 | 0.10 | 1.37E-03 | **7.99E-03** | 48 | 0.32 | 0.07 | 7.84E-06 | 0.00 | 9.08E-01 | 0.20 | 0.64 | 37 |
| Learn new things at job | Psychosocial | 33076 | -0.24 | 0.08 | 2.90E-03 | **1.64E-02** | 48 | 32427 | -0.28 | 0.08 | 9.62E-04 | **6.39E-03** | 42 | -0.26 | 0.06 | 9.04E-06 | 0.00 | 7.95E-01 | 0.20 | 0.61 | 19 |
| Physical limitation to participate in moderately demanding activities: walking up several stairs | General health | 14417 | 0.30 | 0.14 | 3.29E-02 | 8.89E-02 | 101 | 14223 | 0.60 | 0.15 | 3.97E-05 | **4.62E-04** | 24 | 0.45 | 0.10 | 1.05E-05 | 0.54 | 1.40E-01 | - | - | - |
| Salad dressing with oil | Food | 29503 | -0.15 | 0.09 | 7.23E-02 | 1.65E-01 | 119 | 28758 | -0.39 | 0.09 | 8.48E-06 | **1.32E-04** | 18 | -0.27 | 0.06 | 1.10E-05 | 0.73 | 5.34E-02 | - | - | - |
| Cholesterol intake (g/day) | Nutrients | 29503 | -0.27 | 0.09 | 2.50E-03 | **1.48E-02** | 46 | 28758 | -0.29 | 0.09 | 1.41E-03 | **8.01E-03** | 49 | -0.28 | 0.06 | 1.11E-05 | 0.00 | 8.70E-01 | 0.20 | 0.62 | 25 |
| Emotional problems that made you do less than you wanted during the last four weeks | General health | 14446 | -1.28 | 0.41 | 1.89E-03 | **1.22E-02** | 42 | 14234 | -1.31 | 0.42 | 2.07E-03 | **1.01E-02** | 57 | -1.29 | 0.29 | 1.21E-05 | 0.00 | 9.64E-01 | 0.19 | 0.66 | 47 |
| For how much of the time during the last four weeks have you felt gloomy and sad? | General health | 14393 | -0.42 | 0.14 | 2.41E-03 | **1.48E-02** | 44 | 14201 | -0.43 | 0.14 | 2.28E-03 | **1.09E-02** | 58 | -0.42 | 0.10 | 1.68E-05 | 0.00 | 9.56E-01 | 0.19 | 0.67 | 48 |
| Boiled or baked potato | Food | 29503 | 0.21 | 0.09 | 1.54E-02 | 5.39E-02 | 78 | 28758 | 0.32 | 0.09 | 2.59E-04 | **2.26E-03** | 32 | 0.27 | 0.06 | 1.81E-05 | 0.00 | 3.61E-01 | - | - | - |
| Beta-sitostanol intake (mg/day) | Nutrients | 29503 | 0.25 | 0.09 | 3.39E-03 | **1.68E-02** | 55 | 28758 | 0.27 | 0.09 | 1.83E-03 | **9.62E-03** | 53 | 0.26 | 0.06 | 1.92E-05 | 0.00 | 8.55E-01 | 0.20 | 0.62 | 26 |
| Eat breakfast from 2000 | Food | 17082 | -1.51 | 0.45 | 7.92E-04 | **6.00E-03** | 36 | 16684 | -1.14 | 0.44 | 1.05E-02 | **3.75E-02** | 78 | -1.32 | 0.32 | 2.93E-05 | 0.00 | 5.52E-01 | 0.19 | 0.66 | 42 |
| Vitamin B6 intake (mg/day) | Nutrients | 29503 | 0.17 | 0.09 | 5.72E-02 | 1.41E-01 | 111 | 28758 | 0.37 | 0.09 | 6.29E-05 | **6.75E-04** | 26 | 0.27 | 0.06 | 3.19E-05 | 0.57 | 1.27E-01 | - | - | - |
| Fitness status | Physical activity | 24511 | -0.30 | 0.11 | 4.81E-03 | **2.19E-02** | 60 | 24074 | -0.33 | 0.11 | 2.35E-03 | **1.11E-02** | 59 | -0.32 | 0.08 | 3.44E-05 | 0.00 | 8.35E-01 | 0.20 | 0.64 | 40 |
| Satisfaction with economy | Psychosocial | 24524 | 0.31 | 0.10 | 3.09E-03 | **1.65E-02** | 51 | 24088 | 0.31 | 0.11 | 3.96E-03 | **1.71E-02** | 64 | 0.31 | 0.08 | 3.63E-05 | 0.00 | 9.96E-01 | 0.20 | 0.64 | 38 |
| Hearing status | General health | 24542 | 0.15 | 0.11 | 1.53E-01 | 2.96E-01 | 141 | 24110 | 0.49 | 0.11 | 9.38E-06 | **1.38E-04** | 19 | 0.32 | 0.08 | 3.69E-05 | 0.79 | 3.07E-02 | - | - | - |
| Frequency of engaging in clubs, associations or study circles | Social | 21249 | -0.27 | 0.10 | 6.63E-03 | **2.80E-02** | 64 | 20712 | -0.32 | 0.10 | 1.95E-03 | **9.88E-03** | 55 | -0.29 | 0.07 | 4.00E-05 | 0.00 | 7.57E-01 | 0.21 | 0.61 | 8 |
| Carbohydrates intake (g/day) | Nutrients | 29503 | 0.20 | 0.09 | 3.09E-02 | 8.56E-02 | 98 | 28758 | 0.33 | 0.09 | 3.32E-04 | **2.57E-03** | 36 | 0.26 | 0.06 | 5.09E-05 | 0.11 | 2.88E-01 | - | - | - |
| Animal based protein intake (g/day) | Nutrients | 29503 | -0.29 | 0.09 | 8.44E-04 | **6.23E-03** | 37 | 28758 | -0.21 | 0.09 | 1.85E-02 | 5.79E-02 | 89 | -0.25 | 0.06 | 5.50E-05 | 0.00 | 5.14E-01 | - | - | - |
| Changed everyday exercise during the last year | Physical activity | 21883 | 0.35 | 0.09 | 2.13E-04 | **1.88E-03** | 31 | 21328 | 0.19 | 0.10 | 4.71E-02 | 1.17E-01 | 112 | 0.27 | 0.07 | 5.51E-05 | 0.28 | 2.37E-01 | - | - | - |
| Iodine intake (ug/day) | Nutrients | 29503 | -0.34 | 0.09 | 9.28E-05 | **9.05E-04** | 28 | 28758 | -0.15 | 0.09 | 8.96E-02 | 1.85E-01 | 135 | -0.25 | 0.06 | 6.83E-05 | 0.57 | 1.29E-01 | - | - | - |
| Sum of all lignans intake (ug/day) | Nutrients | 29503 | 0.22 | 0.09 | 1.28E-02 | **4.53E-02** | 77 | 28758 | 0.28 | 0.09 | 1.67E-03 | **8.93E-03** | 52 | 0.25 | 0.06 | 6.93E-05 | 0.00 | 6.11E-01 | 0.20 | 0.62 | 29 |
| Boiled coffee | Beverage | 29503 | 0.30 | 0.09 | 1.35E-03 | **9.69E-03** | 38 | 28758 | 0.23 | 0.09 | 1.58E-02 | 5.07E-02 | 87 | 0.26 | 0.07 | 6.95E-05 | 0.00 | 6.01E-01 | - | - | - |
| Selenium intake (ug/day) | Nutrients | 29503 | -0.31 | 0.09 | 3.84E-04 | **3.18E-03** | 33 | 28758 | -0.18 | 0.09 | 4.17E-02 | 1.08E-01 | 108 | -0.25 | 0.06 | 7.38E-05 | 0.04 | 3.06E-01 | - | - | - |
| Low fat margarine on bread | Food | 29503 | 0.25 | 0.09 | 4.00E-03 | **1.88E-02** | 58 | 28758 | 0.24 | 0.09 | 6.46E-03 | **2.61E-02** | 69 | 0.24 | 0.06 | 7.44E-05 | 0.00 | 9.40E-01 | 0.20 | 0.62 | 28 |
| Worsen in health in the future | General health | 14347 | 0.36 | 0.14 | 9.26E-03 | **3.51E-02** | 72 | 14161 | 0.42 | 0.14 | 2.88E-03 | **1.34E-02** | 60 | 0.39 | 0.10 | 7.96E-05 | 0.00 | 7.51E-01 | 0.19 | 0.66 | 49 |
| Do you feel important and appreciated in your home? | Psychosocial | 24365 | 0.23 | 0.10 | 2.22E-02 | 7.05E-02 | 86 | 23921 | 0.34 | 0.10 | 1.10E-03 | **6.84E-03** | 45 | 0.29 | 0.07 | 9.02E-05 | 0.00 | 4.60E-01 | - | - | - |
| Enough time for job assignments | Psychosocial | 32990 | 0.18 | 0.08 | 2.73E-02 | 8.10E-02 | 92 | 32353 | 0.26 | 0.08 | 1.24E-03 | **7.35E-03** | 47 | 0.22 | 0.06 | 1.24E-04 | 0.00 | 4.51E-01 | - | - | - |
| Parents or siblings have diabetes | General health | 32738 | 0.42 | 0.23 | 6.41E-02 | 1.50E-01 | 116 | 32038 | 0.82 | 0.23 | 4.10E-04 | **3.09E-03** | 37 | 0.62 | 0.16 | 1.44E-04 | 0.32 | 2.25E-01 | - | - | - |
| Average portion size of vegetables based on photographic illustration of four sizes (smallest to largest) | Food | 29503 | -0.27 | 0.09 | 3.49E-03 | **1.70E-02** | 56 | 28758 | -0.23 | 0.10 | 1.53E-02 | 5.01E-02 | 85 | -0.25 | 0.07 | 1.54E-04 | 0.00 | 7.64E-01 | - | - | - |
| Satisfaction with accomodation | Psychosocial | 24544 | 0.26 | 0.10 | 8.16E-03 | **3.23E-02** | 69 | 24120 | 0.27 | 0.10 | 8.11E-03 | **3.14E-02** | 72 | 0.27 | 0.07 | 1.82E-04 | 0.00 | 9.64E-01 | 0.20 | 0.64 | 41 |
| Teetotaler | Alcohol | 21512 | 1.46 | 0.35 | 3.85E-05 | **4.57E-04** | 23 | 20953 | 0.41 | 0.37 | 2.74E-01 | 4.29E-01 | 178 | 0.96 | 0.26 | 1.87E-04 | 0.76 | 4.05E-02 | - | - | - |
| Fat intake (g/day) | Nutrients | 29503 | -0.13 | 0.09 | 1.52E-01 | 2.96E-01 | 140 | 28758 | -0.35 | 0.09 | 1.18E-04 | **1.09E-03** | 30 | -0.24 | 0.06 | 2.03E-04 | 0.68 | 7.88E-02 | - | - | - |
| Matairesinol intake (ug/day) | Nutrients | 29503 | 0.19 | 0.09 | 3.10E-02 | 8.56E-02 | 99 | 28758 | 0.27 | 0.09 | 3.22E-03 | **1.45E-02** | 62 | 0.23 | 0.06 | 3.17E-04 | 0.00 | 5.46E-01 | - | - | - |
| Mood status | Psychosocial | 24507 | 0.27 | 0.10 | 7.29E-03 | **2.93E-02** | 68 | 24074 | 0.25 | 0.10 | 1.65E-02 | 5.22E-02 | 88 | 0.26 | 0.07 | 3.24E-04 | 0.00 | 8.70E-01 | - | - | - |
| Skill demand from job | Psychosocial | 33048 | -0.25 | 0.08 | 2.48E-03 | **1.48E-02** | 45 | 32375 | -0.17 | 0.09 | 4.75E-02 | 1.17E-01 | 113 | -0.21 | 0.06 | 3.90E-04 | 0.00 | 4.79E-01 | - | - | - |
| Frequncy of social contacts with colleagues during leisure time | Psychosocial | 31152 | 0.24 | 0.08 | 3.34E-03 | **1.68E-02** | 54 | 30494 | 0.17 | 0.08 | 3.97E-02 | 1.05E-01 | 106 | 0.21 | 0.06 | 4.10E-04 | 0.00 | 5.52E-01 | - | - | - |
| Fibre intake (g/day) | Nutrients | 29503 | 0.15 | 0.10 | 1.29E-01 | 2.60E-01 | 135 | 28758 | 0.35 | 0.10 | 4.92E-04 | **3.52E-03** | 39 | 0.24 | 0.07 | 4.33E-04 | 0.52 | 1.50E-01 | - | - | - |
| Ice cream | Food | 29503 | -0.25 | 0.08 | 3.15E-03 | **1.65E-02** | 52 | 28758 | -0.17 | 0.09 | 4.70E-02 | 1.17E-01 | 111 | -0.21 | 0.06 | 4.58E-04 | 0.00 | 5.34E-01 | - | - | - |
| Physical limitation to participate in steneous activities: running, lifting heavy objects, taking part in physically demanding sports | General health | 14418 | 0.36 | 0.14 | 1.15E-02 | **4.13E-02** | 76 | 14242 | 0.36 | 0.15 | 1.54E-02 | 5.01E-02 | 86 | 0.36 | 0.10 | 4.63E-04 | 0.00 | 9.80E-01 | - | - | - |
| Saturated fat intake (g/day) | Nutrients | 29503 | -0.16 | 0.09 | 8.07E-02 | 1.76E-01 | 125 | 28758 | -0.29 | 0.09 | 1.49E-03 | **8.29E-03** | 50 | -0.22 | 0.06 | 5.18E-04 | 0.09 | 2.95E-01 | - | - | - |
| Exercise during the last three months | Physical activity | 32891 | -0.23 | 0.08 | 6.97E-03 | **2.88E-02** | 66 | 32175 | -0.18 | 0.09 | 3.18E-02 | 8.88E-02 | 100 | -0.20 | 0.06 | 6.04E-04 | 0.00 | 7.23E-01 | - | - | - |
| Total energy intake (kcal/day) | Nutrients | 29503 | -0.23 | 0.10 | 2.42E-02 | 7.42E-02 | 89 | 28758 | -0.27 | 0.11 | 9.44E-03 | **3.53E-02** | 74 | -0.25 | 0.07 | 6.11E-04 | 0.00 | 7.82E-01 | - | - | - |
| Would you say that the number of people that you meet in your everyday life is enough or would you like to meet more or fewer people? | Social | 33605 | -0.19 | 0.08 | 1.65E-02 | 5.63E-02 | 80 | 32895 | -0.20 | 0.08 | 1.48E-02 | **4.92E-02** | 84 | -0.19 | 0.06 | 6.30E-04 | 0.00 | 9.62E-01 | - | - | - |
| Pasta | Food | 29503 | -0.23 | 0.09 | 9.25E-03 | **3.51E-02** | 71 | 28758 | -0.20 | 0.09 | 2.91E-02 | 8.29E-02 | 98 | -0.22 | 0.06 | 7.08E-04 | 0.00 | 7.93E-01 | - | - | - |
| Everyday exercise satisfaction | Physical activity | 21824 | 0.28 | 0.10 | 5.05E-03 | **2.26E-02** | 61 | 21258 | 0.19 | 0.10 | 5.57E-02 | 1.34E-01 | 116 | 0.24 | 0.07 | 8.28E-04 | 0.00 | 5.55E-01 | - | - | - |
| Total protein intake (g/day) | Nutrients | 29503 | -0.26 | 0.09 | 3.01E-03 | **1.64E-02** | 50 | 28758 | -0.15 | 0.09 | 8.80E-02 | 1.85E-01 | 131 | -0.21 | 0.06 | 9.15E-04 | 0.00 | 3.96E-01 | - | - | - |
| Frequency of dancing during leisure time | Physical activity | 19247 | -0.17 | 0.11 | 1.06E-01 | 2.19E-01 | 132 | 18645 | -0.34 | 0.11 | 2.04E-03 | **1.01E-02** | 56 | -0.25 | 0.08 | 9.35E-04 | 0.15 | 2.79E-01 | - | - | - |
| Fiber cereals | Food | 29503 | -0.22 | 0.09 | 1.11E-02 | **4.04E-02** | 75 | 28758 | -0.19 | 0.09 | 3.29E-02 | 9.09E-02 | 101 | -0.20 | 0.06 | 9.38E-04 | 0.00 | 8.12E-01 | - | - | - |
| Sodas, soft drinks, juice | Beverage | 29503 | 0.24 | 0.09 | 6.05E-03 | **2.66E-02** | 62 | 28758 | 0.17 | 0.09 | 5.87E-02 | 1.39E-01 | 118 | 0.21 | 0.06 | 1.02E-03 | 0.00 | 5.74E-01 | - | - | - |
| Participation in sports or physical exercise associations | Social | 10706 | -0.73 | 0.32 | 2.08E-02 | 6.69E-02 | 85 | 10359 | -0.75 | 0.33 | 2.06E-02 | 6.39E-02 | 90 | -0.74 | 0.23 | 1.07E-03 | 0.00 | 9.61E-01 | - | - | - |
| Sausage, liver pate on bread | Food | 29503 | 0.25 | 0.09 | 3.00E-03 | **1.64E-02** | 49 | 28758 | 0.14 | 0.09 | 1.03E-01 | 2.05E-01 | 139 | 0.20 | 0.06 | 1.09E-03 | 0.00 | 3.68E-01 | - | - | - |
| Coffee rolls/buns, rusk | Food | 29503 | 0.25 | 0.09 | 4.59E-03 | **2.13E-02** | 59 | 28758 | 0.16 | 0.09 | 8.33E-02 | 1.82E-01 | 128 | 0.20 | 0.06 | 1.20E-03 | 0.00 | 4.65E-01 | - | - | - |
| Strong beer | Alcohol | 29503 | 0.25 | 0.09 | 6.28E-03 | **2.72E-02** | 63 | 28758 | 0.17 | 0.09 | 6.52E-02 | 1.48E-01 | 123 | 0.21 | 0.07 | 1.21E-03 | 0.00 | 5.34E-01 | - | - | - |
| Frequency of walking during leisure time | Physical activity | 32041 | 0.16 | 0.09 | 6.30E-02 | 1.50E-01 | 115 | 31334 | 0.24 | 0.09 | 7.02E-03 | **2.76E-02** | 71 | 0.20 | 0.06 | 1.31E-03 | 0.00 | 5.29E-01 | - | - | - |
| Campestanol intake (mg/day) | Nutrients | 29503 | 0.19 | 0.09 | 2.36E-02 | 7.33E-02 | 88 | 28758 | 0.20 | 0.09 | 2.41E-02 | 7.04E-02 | 94 | 0.20 | 0.06 | 1.40E-03 | 0.00 | 9.75E-01 | - | - | - |
| Banana | Food | 29503 | 0.16 | 0.09 | 5.89E-02 | 1.44E-01 | 112 | 28758 | 0.23 | 0.09 | 9.49E-03 | **3.53E-02** | 75 | 0.20 | 0.06 | 1.57E-03 | 0.00 | 5.84E-01 | - | - | - |

| | | | | | | | | | | | | | | | | | | | | | |
|---|---|---|---|---|---|---|---|---|---|---|---|---|---|---|---|---|---|---|---|---|---|
| Heptadecanoic acid intake (g/day) | Nutrients | 29503 | -0.19 | 0.09 | 3.04E-02 | 8.56E-02 | 94 | 28758 | -0.20 | 0.09 | 2.42E-02 | 7.04E-02 | 96 | -0.20 | 0.06 | 1.79E-03 | 0.00 | 9.26E-01 | - | - | - |
| Pentadecanoic acid intake (g/day) | Nutrients | 29503 | -0.19 | 0.09 | 3.04E-02 | 8.56E-02 | 95 | 28758 | -0.20 | 0.09 | 2.42E-02 | 7.04E-02 | 95 | -0.20 | 0.06 | 1.79E-03 | 0.00 | 9.26E-01 | - | - | - |
| Secoisolariciresinol intake (ug/day) | Nutrients | 29503 | -0.27 | 0.09 | 3.54E-03 | **1.70E-02** | 57 | 28758 | -0.13 | 0.09 | 1.62E-01 | 2.89E-01 | 156 | -0.20 | 0.07 | 2.22E-03 | 0.10 | 2.93E-01 | - | - | - |
| Pain during the last four weeks | General health | 14417 | 0.23 | 0.14 | 9.19E-02 | 1.98E-01 | 127 | 14207 | 0.38 | 0.14 | 8.43E-03 | **3.22E-02** | 73 | 0.30 | 0.10 | 2.29E-03 | 0.00 | 4.78E-01 | - | - | - |
| Sour milk, yoghurt (3% fat) | Food | 29503 | -0.20 | 0.08 | 1.72E-02 | 5.81E-02 | 81 | 28758 | -0.16 | 0.09 | 5.81E-02 | 1.39E-01 | 117 | -0.18 | 0.06 | 2.46E-03 | 0.00 | 7.60E-01 | - | - | - |
| Palmitic acid intake (g/day) | Nutrients | 29503 | -0.15 | 0.09 | 8.46E-02 | 1.83E-01 | 126 | 28758 | -0.23 | 0.09 | 1.04E-02 | **3.75E-02** | 77 | -0.19 | 0.06 | 2.50E-03 | 0.00 | 5.32E-01 | - | - | - |
| Rice | Food | 29503 | -0.21 | 0.09 | 1.63E-02 | 5.63E-02 | 79 | 28758 | -0.16 | 0.09 | 6.78E-02 | 1.51E-01 | 125 | -0.18 | 0.06 | 2.76E-03 | 0.00 | 6.99E-01 | - | - | - |
| Get sick more often than other people | General health | 14398 | 0.19 | 0.14 | 1.72E-01 | 3.27E-01 | 144 | 14203 | 0.40 | 0.14 | 4.12E-03 | **1.74E-02** | 66 | 0.29 | 0.10 | 2.84E-03 | 0.18 | 2.68E-01 | - | - | - |
| Cheese 28% | Food | 29503 | -0.16 | 0.08 | 6.15E-02 | 1.47E-01 | 114 | 28758 | -0.19 | 0.09 | 2.54E-02 | 7.30E-02 | 97 | -0.17 | 0.06 | 3.73E-03 | 0.00 | 7.70E-01 | - | - | - |
| How much has the pain during the last four weeks disturbed your normal work? | General health | 14378 | 0.28 | 0.14 | 4.34E-02 | 1.13E-01 | 105 | 14183 | 0.28 | 0.14 | 5.00E-02 | 1.21E-01 | 115 | 0.28 | 0.10 | 4.88E-03 | 0.00 | 9.92E-01 | - | - | - |
| Cream, creme fraiche, sour cream | Food | 29503 | -0.12 | 0.08 | 1.51E-01 | 2.96E-01 | 139 | 28758 | -0.22 | 0.09 | 1.20E-02 | **4.20E-02** | 80 | -0.17 | 0.06 | 5.35E-03 | 0.00 | 4.34E-01 | - | - | - |
| If you exercise, change in exercise habits during the last year | Physical activity | 18788 | 0.28 | 0.10 | 6.68E-03 | **2.80E-02** | 65 | 18335 | 0.13 | 0.10 | 2.27E-01 | 3.64E-01 | 174 | 0.20 | 0.07 | 5.41E-03 | 0.08 | 2.98E-01 | - | - | - |
| Extent to what your physical and emotional health disrupted your usual social life during the last four weeks | General health | 14427 | -0.30 | 0.14 | 2.68E-02 | 8.05E-02 | 91 | 14233 | -0.24 | 0.14 | 8.82E-02 | 1.85E-01 | 132 | -0.27 | 0.10 | 5.50E-03 | 0.00 | 7.41E-01 | - | - | - |
| Monosaccharides intake (g/day) | Nutrients | 29503 | 0.09 | 0.09 | 3.59E-01 | 5.65E-01 | 172 | 28758 | 0.29 | 0.10 | 3.21E-03 | **1.45E-02** | 61 | 0.18 | 0.07 | 6.67E-03 | 0.54 | 1.41E-01 | - | - | - |
| Ingenuity or creativity demand from job | Psychosocial | 32988 | -0.15 | 0.08 | 7.25E-02 | 1.65E-01 | 120 | 32348 | -0.17 | 0.09 | 4.12E-02 | 1.07E-01 | 107 | -0.16 | 0.06 | 6.70E-03 | 0.00 | 8.44E-01 | - | - | - |
| Self rate of overall health compared to a year ago | General health | 14338 | -0.35 | 0.14 | 9.97E-03 | **3.68E-02** | 74 | 14135 | -0.17 | 0.14 | 2.23E-01 | 3.60E-01 | 173 | -0.26 | 0.10 | 7.02E-03 | 0.00 | 3.47E-01 | - | - | - |
| Formic acid intaje (g/day) | Nutrients | 29503 | -0.13 | 0.09 | 1.27E-01 | 2.58E-01 | 134 | 28758 | -0.21 | 0.09 | 2.26E-02 | 6.86E-02 | 92 | -0.17 | 0.06 | 7.23E-03 | 0.00 | 5.75E-01 | - | - | - |
| Calcium intake (mg/day) | Nutrients | 29503 | -0.19 | 0.09 | 3.08E-02 | 8.56E-02 | 97 | 28758 | -0.14 | 0.09 | 1.16E-01 | 2.26E-01 | 143 | -0.16 | 0.06 | 8.16E-03 | 0.00 | 7.06E-01 | - | - | - |
| Number of cigarretes smoked per day | Tobacco use | 21637 | -0.19 | 0.11 | 7.74E-02 | 1.72E-01 | 123 | 21218 | -0.21 | 0.11 | 5.00E-02 | 1.21E-01 | 114 | -0.20 | 0.07 | 8.44E-03 | 0.00 | 8.76E-01 | - | - | - |
| Sometimes physically straining work | Physical activity | 32240 | 0.48 | 0.18 | 8.85E-03 | **3.45E-02** | 70 | 31557 | 0.20 | 0.19 | 2.81E-01 | 4.38E-01 | 179 | 0.34 | 0.13 | 8.72E-03 | 0.12 | 2.87E-01 | - | - | - |
| Plant based protein intake (g/day) | Nutrients | 29503 | 0.15 | 0.09 | 1.01E-01 | 2.13E-01 | 129 | 28758 | 0.19 | 0.09 | 3.91E-02 | 1.04E-01 | 105 | 0.17 | 0.06 | 8.92E-03 | 0.00 | 7.44E-01 | - | - | - |
| Tea | Beverage | 29503 | -0.18 | 0.09 | 4.91E-02 | 1.23E-01 | 109 | 28758 | -0.16 | 0.10 | 8.91E-02 | 1.85E-01 | 134 | -0.17 | 0.07 | 9.45E-03 | 0.00 | 8.75E-01 | - | - | - |
| Light beer | Alcohol | 29503 | -0.10 | 0.09 | 2.57E-01 | 4.47E-01 | 157 | 28758 | -0.23 | 0.09 | 1.18E-02 | **4.16E-02** | 79 | -0.16 | 0.06 | 1.04E-02 | 0.06 | 3.03E-01 | - | - | - |
| Last time a colleague visited you at home | Psychosocial | 31566 | 0.15 | 0.08 | 6.43E-02 | 1.50E-01 | 117 | 30922 | 0.15 | 0.08 | 7.62E-02 | 1.67E-01 | 127 | 0.15 | 0.06 | 1.04E-02 | 0.00 | 9.77E-01 | - | - | - |
| Light but partly physically active work | Physical activity | 32240 | -0.62 | 0.20 | 1.93E-03 | **1.22E-02** | 43 | 31557 | -0.10 | 0.21 | 6.36E-01 | 7.76E-01 | 228 | -0.37 | 0.14 | 1.06E-02 | 0.70 | 6.86E-02 | - | - | - |
| Sedentary or standing work | Physical activity | 32240 | -0.21 | 0.19 | 2.65E-01 | 4.56E-01 | 159 | 31557 | -0.49 | 0.20 | 1.24E-02 | **4.26E-02** | 81 | -0.35 | 0.14 | 1.08E-02 | 0.00 | 3.17E-01 | - | - | - |
| Fatty fish (e.g. herring, whitefish, salmon) | Food | 29503 | -0.14 | 0.09 | 1.15E-01 | 2.37E-01 | 133 | 28758 | -0.18 | 0.09 | 4.36E-02 | 1.12E-01 | 109 | -0.16 | 0.06 | 1.12E-02 | 0.00 | 7.33E-01 | - | - | - |
| Berries (fresh or frozen) | Food | 29503 | -0.18 | 0.08 | 3.05E-02 | 8.56E-02 | 96 | 28758 | -0.11 | 0.09 | 1.97E-01 | 3.29E-01 | 167 | -0.15 | 0.06 | 1.43E-02 | 0.00 | 5.58E-01 | - | - | - |
| Physical limitation to participate in moderately demanding activities: bending down or kneeling | General health | 14402 | 0.01 | 0.14 | 9.42E-01 | 9.60E-01 | 268 | 14210 | 0.50 | 0.15 | 7.75E-04 | **5.41E-03** | 40 | 0.25 | 0.10 | 1.62E-02 | 0.82 | 1.73E-02 | - | - | - |
| Frequency of picking berries or mushrooms during leisure time | Physical activity | 20560 | 0.28 | 0.11 | 9.71E-03 | **3.63E-02** | 73 | 20005 | 0.09 | 0.11 | 4.39E-01 | 5.97E-01 | 205 | 0.18 | 0.08 | 1.67E-02 | 0.36 | 2.11E-01 | - | - | - |
| Sum of phytosterols intake (mg/day) | Nutrients | 29503 | 0.15 | 0.09 | 8.04E-02 | 1.76E-01 | 124 | 28758 | 0.14 | 0.09 | 1.18E-01 | 2.28E-01 | 144 | 0.15 | 0.06 | 1.91E-02 | 0.00 | 9.18E-01 | - | - | - |
| Vitamin B3 intake (mg/day) | Nutrients | 29503 | -0.20 | 0.09 | 1.97E-02 | 6.42E-02 | 84 | 28758 | -0.07 | 0.09 | 4.12E-01 | 5.78E-01 | 198 | -0.14 | 0.06 | 2.49E-02 | 0.07 | 2.99E-01 | - | - | - |
| Years using snuff | Tobacco use | 31504 | -0.16 | 0.11 | 1.41E-01 | 2.80E-01 | 137 | 30850 | -0.18 | 0.11 | 1.10E-01 | 2.16E-01 | 142 | -0.17 | 0.08 | 2.99E-02 | 0.00 | 8.99E-01 | - | - | - |
| Docosahexaenoic acid (DHA) intake (g/day) | Nutrients | 29503 | -0.12 | 0.09 | 1.83E-01 | 3.45E-01 | 145 | 28758 | -0.16 | 0.09 | 8.47E-02 | 1.83E-01 | 129 | -0.13 | 0.06 | 3.12E-02 | 0.00 | 7.53E-01 | - | - | - |
| Corn flakes | Food | 29503 | -0.18 | 0.08 | 3.00E-02 | 8.56E-02 | 93 | 28758 | -0.07 | 0.09 | 3.93E-01 | 5.58E-01 | 195 | -0.13 | 0.06 | 3.15E-02 | 0.00 | 3.67E-01 | - | - | - |
| Monounsaturated fat intake (g/day) | Nutrients | 29503 | -0.09 | 0.09 | 3.34E-01 | 5.43E-01 | 167 | 28758 | -0.19 | 0.09 | 3.89E-02 | 1.04E-01 | 104 | -0.14 | 0.07 | 3.30E-02 | 0.00 | 4.19E-01 | - | - | - |
| For how much of the time during the last four weeks have you felt so depressed that nothing could cheer you up? | General health | 14423 | -0.28 | 0.14 | 4.13E-02 | 1.09E-01 | 103 | 14226 | -0.13 | 0.14 | 3.48E-01 | 5.19E-01 | 187 | -0.21 | 0.10 | 3.42E-02 | 0.00 | 4.48E-01 | - | - | - |
| Vitamin B2 intake (ug/day) | Nutrients | 29503 | -0.18 | 0.09 | 4.18E-02 | 1.10E-01 | 104 | 28758 | -0.08 | 0.09 | 3.45E-01 | 5.19E-01 | 185 | -0.13 | 0.06 | 3.43E-02 | 0.00 | 4.57E-01 | - | - | - |
| Travel to work: Walk to work vs passive travel to work | Physical activity | 21382 | 0.37 | 0.31 | 2.34E-01 | 4.21E-01 | 152 | 21079 | 0.57 | 0.32 | 7.24E-02 | 1.60E-01 | 126 | 0.47 | 0.22 | 3.52E-02 | 0.00 | 6.50E-01 | - | - | - |
| Physical limitation to participate in moderately demanding activities: walking more than 2 km | General health | 14423 | 0.11 | 0.14 | 4.25E-01 | 6.20E-01 | 187 | 14231 | 0.31 | 0.14 | 2.98E-02 | 8.40E-02 | 99 | 0.21 | 0.10 | 3.66E-02 | 0.01 | 3.15E-01 | - | - | - |
| Wine | Alcohol | 29503 | -0.19 | 0.09 | 4.41E-02 | 1.14E-01 | 106 | 28758 | -0.09 | 0.09 | 3.57E-01 | 5.23E-01 | 190 | -0.14 | 0.07 | 3.71E-02 | 0.00 | 4.55E-01 | - | - | - |
| Rosehip, sweet syrup soup | Food | 29503 | 0.05 | 0.08 | 5.12E-01 | 6.79E-01 | 206 | 28758 | 0.19 | 0.08 | 2.19E-02 | 6.71E-02 | 91 | 0.12 | 0.06 | 3.76E-02 | 0.27 | 2.43E-01 | - | - | - |
| Lariciresinol intake (ug/day) | Nutrients | 29503 | 0.04 | 0.09 | 6.36E-01 | 7.73E-01 | 224 | 28758 | 0.23 | 0.10 | 1.48E-02 | **4.92E-02** | 83 | 0.14 | 0.07 | 4.12E-02 | 0.50 | 1.57E-01 | - | - | - |
| Average portion size of potatoes/rice/pasta based on photographic illustration of four sizes (smallest to largest) | Food | 29503 | -0.18 | 0.10 | 6.01E-02 | 1.45E-01 | 113 | 28758 | -0.09 | 0.10 | 3.40E-01 | 5.16E-01 | 184 | -0.14 | 0.07 | 4.43E-02 | 0.00 | 5.28E-01 | - | - | - |
| Self-employed | Psychosocial | 28672 | -0.67 | 0.31 | 3.24E-02 | 8.84E-02 | 100 | 27917 | -0.22 | 0.32 | 4.91E-01 | 6.49E-01 | 211 | -0.45 | 0.22 | 4.49E-02 | 0.03 | 3.10E-01 | - | - | - |
| Meat on bread | Food | 29503 | 0.10 | 0.08 | 2.34E-01 | 4.21E-01 | 151 | 28758 | 0.14 | 0.09 | 1.03E-01 | 2.05E-01 | 140 | 0.12 | 0.06 | 4.63E-02 | 0.00 | 7.42E-01 | - | - | - |
| Campesterol intake (mg/day) | Nutrients | 29503 | 0.20 | 0.08 | 1.90E-02 | 6.33E-02 | 82 | 28758 | 0.04 | 0.09 | 6.56E-01 | 7.92E-01 | 231 | 0.12 | 0.06 | 4.66E-02 | 0.43 | 1.87E-01 | - | - | - |
| Beta-sitosterol intake (mg/day) | Nutrients | 29503 | 0.10 | 0.09 | 2.48E-01 | 4.40E-01 | 154 | 28758 | 0.15 | 0.09 | 1.01E-01 | 2.04E-01 | 138 | 0.13 | 0.06 | 4.85E-02 | 0.00 | 7.14E-01 | - | - | - |
| For how much of the time during the last four weeks has your physical health or your emotional problems limited your ability to interact with others? | General health | 14341 | -0.18 | 0.14 | 1.86E-01 | 3.46E-01 | 147 | 14134 | -0.20 | 0.14 | 1.64E-01 | 2.90E-01 | 158 | -0.19 | 0.10 | 5.51E-02 | 0.00 | 9.45E-01 | - | - | - |
| Linoleic acid intake (g/day) | Nutrients | 29503 | -0.05 | 0.08 | 5.63E-01 | 7.19E-01 | 214 | 28758 | -0.18 | 0.09 | 3.75E-02 | 1.01E-01 | 103 | -0.11 | 0.06 | 6.20E-02 | 0.15 | 2.77E-01 | - | - | - |
| Potassium intake (mg/day) | Nutrients | 29503 | -0.01 | 0.09 | 9.14E-01 | 9.41E-01 | 265 | 28758 | 0.27 | 0.10 | 5.40E-03 | **2.25E-02** | 67 | 0.13 | 0.07 | 6.21E-02 | 0.77 | 3.87E-02 | - | - | - |
| Frequency of cycling during leisure time | Physical activity | 27222 | -0.08 | 0.09 | 3.60E-01 | 5.65E-01 | 173 | 26469 | -0.16 | 0.09 | 8.91E-02 | 1.85E-01 | 133 | -0.12 | 0.07 | 6.55E-02 | 0.00 | 5.63E-01 | - | - | - |
| Stigmasterol intake (mg/day) | Nutrients | 29503 | 0.10 | 0.09 | 2.84E-01 | 4.81E-01 | 161 | 28758 | 0.14 | 0.09 | 1.39E-01 | 2.54E-01 | 153 | 0.12 | 0.07 | 7.19E-02 | 0.00 | 7.56E-01 | - | - | - |
| Oil for cooking | Food | 29503 | -0.09 | 0.09 | 3.15E-01 | 5.19E-01 | 166 | 28758 | -0.14 | 0.09 | 1.23E-01 | 2.33E-01 | 146 | -0.11 | 0.06 | 7.23E-02 | 0.00 | 6.88E-01 | - | - | - |
| For how much of the time during the last four weeks have you felt happy? | General health | 14412 | 0.30 | 0.14 | 2.66E-02 | 8.05E-02 | 90 | 14206 | 0.04 | 0.14 | 7.77E-01 | 8.64E-01 | 251 | 0.17 | 0.10 | 7.44E-02 | 0.46 | 1.75E-01 | - | - | - |
| Zinc intake (mg/day) | Nutrients | 29503 | -0.21 | 0.09 | 1.94E-02 | 6.39E-02 | 83 | 28758 | -0.01 | 0.09 | 8.91E-01 | 9.11E-01 | 273 | -0.11 | 0.06 | 7.61E-02 | 0.57 | 1.26E-01 | - | - | - |
| Margarine for cooking | Food | 29503 | 0.17 | 0.09 | 4.53E-02 | 1.16E-01 | 107 | 28758 | 0.04 | 0.09 | 6.27E-01 | 7.75E-01 | 225 | 0.11 | 0.06 | 7.62E-02 | 0.09 | 2.94E-01 | - | - | - |
| Number of social contacts with the same interests as you | Social | 33555 | -0.10 | 0.09 | 2.48E-01 | 4.40E-01 | 153 | 32831 | -0.12 | 0.09 | 1.78E-01 | 3.07E-01 | 161 | -0.11 | 0.06 | 7.71E-02 | 0.00 | 8.78E-01 | - | - | - |
| Breakfast habits: Gruel w/o sandwich for breakfast vs not breakfast at all | Food | 1448 | 0.95 | 1.04 | 3.60E-01 | 5.65E-01 | 174 | 1508 | 1.46 | 0.94 | 1.23E-01 | 2.33E-01 | 147 | 1.23 | 0.70 | 7.86E-02 | 0.00 | 7.15E-01 | - | - | - |
| Milk, sour milk (1,5%) | Beverage | 29503 | -0.11 | 0.09 | 1.86E-01 | 3.46E-01 | 146 | 28758 | -0.10 | 0.09 | 2.53E-01 | 4.02E-01 | 176 | -0.11 | 0.06 | 8.10E-02 | 0.00 | 9.19E-01 | - | - | - |
| Cheese 10-17% | Food | 29503 | 0.02 | 0.08 | 8.28E-01 | 8.91E-01 | 255 | 28758 | 0.19 | 0.08 | 2.40E-02 | 7.04E-02 | 93 | 0.10 | 0.06 | 8.43E-02 | 0.54 | 1.42E-01 | - | - | - |
| Bacon | Food | 29503 | -0.09 | 0.08 | 2.87E-01 | 4.83E-01 | 162 | 28758 | -0.12 | 0.09 | 1.78E-01 | 3.07E-01 | 162 | -0.10 | 0.06 | 8.88E-02 | 0.00 | 8.31E-01 | - | - | - |
| Eicosapentaenoic acid (EPA) intake (g/day) | Nutrients | 29502 | -0.07 | 0.09 | 3.93E-01 | 6.00E-01 | 179 | 28758 | -0.14 | 0.09 | 1.19E-01 | 2.29E-01 | 145 | -0.11 | 0.06 | 8.94E-02 | 0.00 | 5.99E-01 | - | - | - |
| For how much of the time during the last four weeks have you felt tired? | General health | 14426 | -0.25 | 0.14 | 7.67E-02 | 1.72E-01 | 121 | 14216 | -0.09 | 0.14 | 5.44E-01 | 6.96E-01 | 218 | -0.17 | 0.10 | 9.09E-02 | 0.00 | 4.19E-01 | - | - | - |
| Phosphate intake (mg/day) | Nutrients | 29503 | -0.18 | 0.09 | 3.90E-02 | 1.04E-01 | 102 | 28758 | -0.03 | 0.09 | 7.68E-01 | 8.57E-01 | 249 | -0.10 | 0.06 | 9.24E-02 | 0.34 | 2.18E-01 | - | - | - |
| Receive hugs to comfort and support you | Social | 33017 | -0.40 | 0.22 | 7.69E-02 | 1.72E-01 | 122 | 32274 | -0.13 | 0.23 | 5.81E-01 | 7.23E-01 | 224 | -0.26 | 0.16 | 9.87E-02 | 0.00 | 4.00E-01 | - | - | - |
| Apple, pear, peach, orange, mandarin and grapefruit | Food | 29503 | -0.04 | 0.09 | 6.99E-01 | 8.12E-01 | 233 | 28758 | 0.27 | 0.10 | 6.04E-03 | **2.48E-02** | 68 | 0.11 | 0.07 | 1.01E-01 | 0.80 | 2.54E-02 | - | - | - |
| Pinoresinol intake (ug/day) | Nutrients | 29503 | 0.05 | 0.09 | 5.47E-01 | 7.04E-01 | 212 | 28758 | 0.16 | 0.09 | 8.59E-02 | 1.84E-01 | 130 | 0.11 | 0.07 | 1.03E-01 | 0.00 | 4.18E-01 | - | - | - |
| Control over planning and execution of the workday | Psychosocial | 33157 | -0.13 | 0.08 | 1.04E-01 | 2.17E-01 | 131 | 32491 | -0.06 | 0.08 | 5.01E-01 | 6.56E-01 | 213 | -0.09 | 0.06 | 1.03E-01 | 0.00 | 5.08E-01 | - | - | - |
| Feel uneasy or guilty because of your way of drinking | Alcohol | 19320 | -0.28 | 0.34 | 4.05E-01 | 6.07E-01 | 181 | 18924 | -0.48 | 0.34 | 1.55E-01 | 2.79E-01 | 155 | -0.38 | 0.24 | 1.11E-01 | 0.00 | 6.74E-01 | - | - | - |
| Vitamin C intake (mg/day) | Nutrients | 29503 | -0.05 | 0.09 | 5.46E-01 | 7.04E-01 | 211 | 28758 | 0.25 | 0.09 | 3.99E-03 | **1.71E-02** | 65 | 0.10 | 0.06 | 1.13E-01 | 0.84 | 1.31E-02 | - | - | - |
| Magnesium intake (mg/day) | Nutrients | 29503 | -0.02 | 0.09 | 8.39E-01 | 8.91E-01 | 254 | 28758 | 0.23 | 0.09 | 1.44E-02 | **4.89E-02** | 82 | 0.10 | 0.07 | 1.19E-01 | 0.72 | 5.78E-02 | - | - | - |
| Polyunsaturated fat intake (g/day) | Nutrients | 29503 | -0.03 | 0.09 | 7.27E-01 | 8.22E-01 | 241 | 28758 | -0.16 | 0.09 | 6.65E-02 | 1.50E-01 | 124 | -0.09 | 0.06 | 1.26E-01 | 0.13 | 2.84E-01 | - | - | - |
| Sum of Lariciresinol, Matairesinol, Pinoresinol, Secoisolariciresinol intake (ug/day) | Nutrients | 29503 | 0.02 | 0.09 | 8.01E-01 | 8.72E-01 | 251 | 28758 | 0.18 | 0.09 | 6.16E-02 | 1.41E-01 | 122 | 0.10 | 0.07 | 1.37E-01 | 0.26 | 2.45E-01 | - | - | - |
| Sugar, honey, marmelade, jam | Food | 29503 | -0.10 | 0.09 | 2.55E-01 | 4.47E-01 | 155 | 28758 | -0.08 | 0.09 | 3.53E-01 | 5.21E-01 | 189 | -0.09 | 0.06 | 1.43E-01 | 0.00 | 8.94E-01 | - | - | - |
| Minced meat dishes | Food | 29503 | -0.16 | 0.08 | 5.18E-02 | 1.28E-01 | 110 | 28758 | -0.01 | 0.09 | 9.23E-01 | 9.33E-01 | 276 | -0.09 | 0.06 | 1.44E-01 | 0.40 | 1.97E-01 | - | - | - |
| Frequent social contacts with colleagues during work | Psychosocial | 31690 | -0.06 | 0.08 | 5.00E-01 | 6.72E-01 | 202 | 31022 | -0.11 | 0.08 | 1.67E-01 | 2.91E-01 | 160 | -0.08 | 0.06 | 1.47E-01 | 0.00 | 6.11E-01 | - | - | - |
| Physical limitation to participate in moderately demanding activities: moving a table, vacuuming, walking in the forest or gardening | General health | 14415 | 0.05 | 0.14 | 7.09E-01 | 8.15E-01 | 237 | 14239 | 0.24 | 0.14 | 9.48E-02 | 1.95E-01 | 136 | 0.14 | 0.10 | 1.52E-01 | 0.00 | 3.47E-01 | - | - | - |
| Physical limitation that reduced the normal time spent at work or in other activities during the last four weeks | General health | 14446 | 0.31 | 0.44 | 4.70E-01 | 6.48E-01 | 198 | 14231 | 0.56 | 0.44 | 2.01E-01 | 3.32E-01 | 168 | 0.44 | 0.31 | 1.57E-01 | 0.00 | 6.87E-01 | - | - | - |
| Iron intake (mg/day) | Nutrients | 29503 | 0.04 | 0.09 | 6.49E-01 | 7.80E-01 | 227 | 28758 | 0.14 | 0.09 | 1.31E-01 | 2.45E-01 | 148 | 0.09 | 0.06 | 1.67E-01 | 0.00 | 4.46E-01 | - | - | - |
| Light and physically active work | Physical activity | 32240 | 0.15 | 0.19 | 4.21E-01 | 6.20E-01 | 185 | 31557 | 0.22 | 0.19 | 2.55E-01 | 4.02E-01 | 177 | 0.18 | 0.13 | 1.71E-01 | 0.00 | 7.99E-01 | - | - | - |
| Participation in study circles | Social | 10706 | 0.72 | 0.43 | 9.79E-02 | 2.09E-01 | 128 | 10359 | 0.12 | 0.44 | 7.92E-01 | 8.70E-01 | 254 | 0.42 | 0.31 | 1.72E-01 | 0.00 | 3.31E-01 | - | - | - |
| Sweets | Food | 29503 | -0.07 | 0.09 | 4.45E-01 | 6.30E-01 | 193 | 28758 | -0.10 | 0.09 | 2.50E-01 | 3.99E-01 | 175 | -0.09 | 0.06 | 1.77E-01 | 0.00 | 7.77E-01 | - | - | - |
| For how much of the time during the last four weeks have you felt very nervous? | General health | 14407 | 0.25 | 0.14 | 6.88E-02 | 1.59E-01 | 118 | 14213 | 0.01 | 0.14 | 9.65E-01 | 9.65E-01 | 279 | 0.13 | 0.10 | 1.82E-01 | 0.35 | 2.14E-01 | - | - | - |
| Disaccharides intake (g/day) | Nutrients | 29503 | 0.10 | 0.09 | 2.80E-01 | 4.77E-01 | 160 | 28758 | 0.07 | 0.09 | 4.24E-01 | 5.86E-01 | 201 | 0.09 | 0.07 | 1.83E-01 | 0.00 | 8.54E-01 | - | - | - |
| Energy status | Psychosocial | 24500 | 0.20 | 0.10 | 4.78E-02 | 1.21E-01 | 108 | 24048 | -0.02 | 0.11 | 8.54E-01 | 8.96E-01 | 266 | 0.10 | 0.07 | 1.98E-01 | 0.56 | 1.30E-01 | - | - | - |
| For how much of the time during the last four weeks have you felt really alert and strong? | General health | 14387 | -0.05 | 0.14 | 7.13E-01 | 8.15E-01 | 239 | 14184 | -0.20 | 0.14 | 1.49E-01 | 2.70E-01 | 154 | -0.13 | 0.10 | 2.03E-01 | 0.00 | 4.38E-01 | - | - | - |
| Hamburger | Food | 29503 | -0.09 | 0.09 | 2.93E-01 | 4.88E-01 | 164 | 28758 | -0.07 | 0.09 | 4.58E-01 | 6.12E-01 | 209 | -0.08 | 0.06 | 2.04E-01 | 0.00 | 8.43E-01 | - | - | - |
| Number of social interactions during a normal week | Social | 33263 | 0.04 | 0.08 | 6.27E-01 | 7.68E-01 | 223 | 32526 | 0.11 | 0.09 | 1.88E-01 | 3.17E-01 | 166 | 0.08 | 0.06 | 2.05E-01 | 0.00 | 5.48E-01 | - | - | - |
| Feel the need to reduce alcohol consumption | Alcohol | 30165 | 0.27 | 0.26 | 3.04E-01 | 5.03E-01 | 165 | 29576 | 0.20 | 0.27 | 4.45E-01 | 6.00E-01 | 207 | 0.24 | 0.19 | 2.05E-01 | 0.00 | 8.54E-01 | - | - | - |
| Job demands to work very fast | Psychosocial | 33132 | -0.06 | 0.08 | 4.31E-01 | 6.20E-01 | 188 | 32504 | -0.08 | 0.08 | 3.19E-01 | 4.89E-01 | 182 | -0.07 | 0.06 | 2.07E-01 | 0.00 | 8.77E-01 | - | - | - |
| Sucrose intake (g/day) | Nutrients | 29503 | 0.10 | 0.09 | 2.89E-01 | 4.83E-01 | 163 | 28758 | 0.07 | 0.09 | 4.79E-01 | 6.37E-01 | 210 | 0.08 | 0.06 | 2.10E-01 | 0.00 | 8.14E-01 | - | - | - |
| White (soft) bread, thin crisp bread | Food | 29503 | 0.11 | 0.09 | 1.95E-01 | 3.57E-01 | 149 | 28758 | 0.04 | 0.09 | 6.79E-01 | 8.07E-01 | 234 | 0.08 | 0.06 | 2.24E-01 | 0.00 | 5.41E-01 | - | - | - |
| White cabbage, lettuce, lettuce cabbage, spinach, borecole | Food | 29503 | -0.15 | 0.09 | 1.01E-01 | 2.13E-01 | 130 | 28758 | 0.00 | 0.09 | 9.57E-01 | 9.61E-01 | 278 | -0.08 | 0.06 | 2.25E-01 | 0.18 | 2.70E-01 | - | - | - |
| Potato salad | Food | 5586 | 0.14 | 0.19 | 4.65E-01 | 6.45E-01 | 196 | 5409 | 0.19 | 0.19 | 3.53E-01 | 5.21E-01 | 188 | 0.17 | 0.13 | 2.25E-01 | 0.00 | 8.43E-01 | - | - | - |

| | | | | | | | | | | | | | | | | | | | | | |
|---|---|---|---|---|---|---|---|---|---|---|---|---|---|---|---|---|---|---|---|---|---|
| Possibility to leave your work for a while to speak with a colleague | Psychosocial | 31708 | -0.02 | 0.09 | 8.41E-01 | 8.91E-01 | 256 | 31054 | -0.13 | 0.09 | 1.32E-01 | 2.45E-01 | 150 | -0.07 | 0.06 | 2.31E-01 | 0.00 | 3.49E-01 | - | - | - |
| Frequency of gardening during leisure time | Physical activity | 20714 | 0.15 | 0.10 | 1.57E-01 | 3.00E-01 | 143 | 20046 | 0.03 | 0.10 | 7.88E-01 | 8.69E-01 | 253 | 0.09 | 0.07 | 2.31E-01 | 0.00 | 4.23E-01 | - | - | - |
| Number of people with whom you can speak openly | Social | 33591 | 0.07 | 0.08 | 4.32E-01 | 6.20E-01 | 189 | 32872 | 0.08 | 0.09 | 3.84E-01 | 5.52E-01 | 194 | 0.07 | 0.06 | 2.42E-01 | 0.00 | 9.42E-01 | - | - | - |
| Sodium intake (mg/day) | Nutrients | 29502 | -0.07 | 0.09 | 4.14E-01 | 6.15E-01 | 184 | 28757 | -0.07 | 0.09 | 4.24E-01 | 5.86E-01 | 202 | -0.07 | 0.06 | 2.53E-01 | 0.00 | 9.99E-01 | - | - | - |
| Enterolactone intake (ug/day) | Nutrients | 29503 | -0.06 | 0.08 | 4.79E-01 | 6.57E-01 | 199 | 28758 | -0.08 | 0.09 | 3.79E-01 | 5.50E-01 | 192 | -0.07 | 0.06 | 2.62E-01 | 0.00 | 8.94E-01 | - | - | - |
| Physical limitation to participate in moderately demanding activities: lifting or carrying grocery bags | General health | 14401 | 0.11 | 0.14 | 4.32E-01 | 6.20E-01 | 190 | 14230 | 0.11 | 0.15 | 4.41E-01 | 5.97E-01 | 206 | 0.11 | 0.10 | 2.71E-01 | 0.00 | 9.97E-01 | - | - | - |
| Patience status | Psychosocial | 24507 | 0.03 | 0.10 | 7.78E-01 | 8.49E-01 | 250 | 24081 | 0.13 | 0.10 | 2.06E-01 | 3.38E-01 | 170 | 0.08 | 0.07 | 2.77E-01 | 0.00 | 4.81E-01 | - | - | - |
| Breggott on bread | Food | 29503 | -0.01 | 0.09 | 8.70E-01 | 9.10E-01 | 261 | 28758 | -0.12 | 0.09 | 1.82E-01 | 3.09E-01 | 164 | -0.07 | 0.06 | 2.94E-01 | 0.00 | 4.00E-01 | - | - | - |
| Do you feel important and appreciated outside your home? | Psychosocial | 24532 | -0.04 | 0.10 | 6.90E-01 | 8.12E-01 | 230 | 24113 | 0.20 | 0.10 | 6.01E-02 | 1.41E-01 | 119 | 0.08 | 0.07 | 3.03E-01 | 0.62 | 1.05E-01 | - | - | - |
| Lean fish (e.g. perch, bass, cod) | Food | 29503 | -0.19 | 0.08 | 2.25E-02 | 7.05E-02 | 87 | 28758 | 0.07 | 0.09 | 3.93E-01 | 5.58E-01 | 196 | -0.06 | 0.06 | 3.04E-01 | 0.80 | 2.71E-02 | - | - | - |
| Folic acid intake (ug/day) | Nutrients | 29503 | -0.05 | 0.10 | 5.95E-01 | 7.40E-01 | 219 | 28758 | 0.20 | 0.10 | 4.57E-02 | 1.16E-01 | 110 | 0.07 | 0.07 | 3.13E-01 | 0.69 | 7.11E-02 | - | - | - |
| Tiamin intake (mg/day) | Nutrients | 29503 | -0.02 | 0.09 | 8.06E-01 | 8.73E-01 | 252 | 28758 | 0.15 | 0.09 | 9.73E-02 | 1.98E-01 | 137 | 0.06 | 0.06 | 3.29E-01 | 0.46 | 1.73E-01 | - | - | - |
| Cohabitation: Live alone vs Only one adult (spouse, partner) | Social | 15176 | -0.03 | 0.35 | 9.22E-01 | 9.46E-01 | 266 | 15028 | -0.45 | 0.35 | 2.01E-01 | 3.32E-01 | 169 | -0.24 | 0.25 | 3.29E-01 | 0.00 | 4.06E-01 | - | - | - |
| Number of cigars smoked per day | Tobacco use | 21106 | 0.08 | 0.10 | 4.66E-01 | 6.45E-01 | 197 | 20659 | -0.23 | 0.11 | 3.33E-02 | 9.11E-02 | 102 | -0.07 | 0.07 | 3.32E-01 | 0.76 | 4.23E-02 | - | - | - |
| Vitamin D intake (ug/day) | Nutrients | 29503 | 0.01 | 0.08 | 8.96E-01 | 9.26E-01 | 264 | 28758 | -0.13 | 0.09 | 1.39E-01 | 2.54E-01 | 152 | -0.06 | 0.06 | 3.45E-01 | 0.24 | 2.51E-01 | - | - | - |
| Breakfast habits: Coffee/tea and wheat buns or rusk for breakfast vs not breakfast at all | Food | 1596 | -0.73 | 0.84 | 3.87E-01 | 5.94E-01 | 178 | 1585 | -0.38 | 0.80 | 6.37E-01 | 7.76E-01 | 229 | -0.55 | 0.58 | 3.47E-01 | 0.00 | 7.66E-01 | - | - | - |
| Support from others | Social | 33634 | 0.03 | 0.08 | 7.32E-01 | 8.22E-01 | 243 | 32917 | 0.08 | 0.08 | 3.32E-01 | 5.06E-01 | 183 | 0.05 | 0.06 | 3.55E-01 | 0.00 | 6.51E-01 | - | - | - |
| Beta-carotene intake (mg/day) | Nutrients | 29503 | -0.06 | 0.09 | 5.20E-01 | 6.83E-01 | 207 | 28758 | -0.06 | 0.09 | 5.18E-01 | 6.72E-01 | 215 | -0.06 | 0.07 | 3.62E-01 | 0.00 | 9.89E-01 | - | - | - |
| Equol intake (ug/day) | Nutrients | 29503 | 0.04 | 0.09 | 6.72E-01 | 8.01E-01 | 229 | 28758 | 0.08 | 0.09 | 3.84E-01 | 5.52E-01 | 193 | 0.06 | 0.06 | 3.62E-01 | 0.00 | 7.43E-01 | - | - | - |
| Trans fat intake (g/day) | Nutrients | 29503 | 0.11 | 0.12 | 3.34E-01 | 5.43E-01 | 168 | 28758 | 0.04 | 0.12 | 7.66E-01 | 8.57E-01 | 248 | 0.08 | 0.09 | 3.69E-01 | 0.00 | 6.44E-01 | - | - | - |
| Linolenic acid intake (g/day) | Nutrients | 29503 | 0.03 | 0.08 | 7.12E-01 | 8.15E-01 | 238 | 28758 | -0.14 | 0.09 | 1.10E-01 | 2.16E-01 | 141 | -0.05 | 0.06 | 3.94E-01 | 0.49 | 1.61E-01 | - | - | - |
| Oatflake, whole wheat, rye or barley porridge | Food | 29503 | -0.13 | 0.09 | 1.34E-01 | 2.68E-01 | 136 | 28758 | 0.03 | 0.09 | 7.53E-01 | 8.56E-01 | 245 | -0.05 | 0.06 | 3.94E-01 | 0.38 | 2.02E-01 | - | - | - |
| Possibility to speak with colleagues during breaks | Psychosocial | 31906 | -0.11 | 0.08 | 1.91E-01 | 3.53E-01 | 148 | 31255 | 0.01 | 0.08 | 9.03E-01 | 9.16E-01 | 275 | -0.05 | 0.06 | 3.98E-01 | 0.01 | 3.16E-01 | - | - | - |
| Alcohol intake (g/day) | Alcohol | 29503 | -0.09 | 0.10 | 3.50E-01 | 5.58E-01 | 171 | 28758 | -0.02 | 0.10 | 8.02E-01 | 8.77E-01 | 255 | -0.06 | 0.07 | 4.00E-01 | 0.00 | 6.32E-01 | - | - | - |
| Margarine on bread | Food | 29503 | -0.03 | 0.08 | 7.39E-01 | 8.27E-01 | 244 | 28758 | -0.07 | 0.08 | 3.94E-01 | 5.58E-01 | 197 | -0.05 | 0.06 | 4.04E-01 | 0.00 | 7.08E-01 | - | - | - |
| Root vegetables, carrot | Food | 29503 | -0.04 | 0.09 | 6.37E-01 | 7.73E-01 | 225 | 28758 | -0.06 | 0.09 | 4.95E-01 | 6.51E-01 | 212 | -0.05 | 0.07 | 4.15E-01 | 0.00 | 8.73E-01 | - | - | - |
| Vision status | General health | 24518 | 0.08 | 0.10 | 4.33E-01 | 6.20E-01 | 191 | 24069 | 0.04 | 0.10 | 7.18E-01 | 8.32E-01 | 241 | 0.06 | 0.07 | 4.17E-01 | 0.00 | 7.71E-01 | - | - | - |
| Enterodiol intake (ug/day) | Nutrients | 29503 | 0.07 | 0.09 | 4.05E-01 | 6.07E-01 | 182 | 28758 | 0.03 | 0.09 | 7.68E-01 | 8.57E-01 | 250 | 0.05 | 0.06 | 4.22E-01 | 0.00 | 7.11E-01 | - | - | - |
| Grams of tobacco smoked per week | Tobacco use | 21120 | -0.06 | 0.10 | 5.72E-01 | 7.24E-01 | 215 | 20682 | -0.06 | 0.11 | 5.75E-01 | 7.22E-01 | 222 | -0.06 | 0.07 | 4.26E-01 | 0.00 | 9.92E-01 | - | - | - |
| Low fat milk (0,5%) | Beverage | 29503 | 0.02 | 0.09 | 8.54E-01 | 9.00E-01 | 259 | 28758 | 0.08 | 0.09 | 3.47E-01 | 5.19E-01 | 186 | 0.05 | 0.06 | 4.31E-01 | 0.00 | 5.85E-01 | - | - | - |
| Appetite status | Psychosocial | 24523 | 0.02 | 0.10 | 8.79E-01 | 9.13E-01 | 263 | 24092 | -0.13 | 0.10 | 2.11E-01 | 3.44E-01 | 171 | -0.05 | 0.07 | 4.42E-01 | 0.00 | 3.18E-01 | - | - | - |
| Participation in other association | Social | 10706 | 0.05 | 0.31 | 8.77E-01 | 9.13E-01 | 262 | 10359 | 0.29 | 0.32 | 3.58E-01 | 5.23E-01 | 191 | 0.17 | 0.22 | 4.51E-01 | 0.00 | 5.83E-01 | - | - | - |
| Whole grain soft bread | Food | 29503 | 0.07 | 0.09 | 4.12E-01 | 6.15E-01 | 183 | 28758 | 0.02 | 0.09 | 8.38E-01 | 8.86E-01 | 264 | 0.05 | 0.06 | 4.66E-01 | 0.00 | 6.70E-01 | - | - | - |
| Arachidonic acid (ARA) intake (g/day) | Nutrients | 29502 | -0.07 | 0.09 | 4.22E-01 | 6.20E-01 | 186 | 28757 | -0.02 | 0.09 | 8.28E-01 | 8.85E-01 | 261 | -0.05 | 0.06 | 4.67E-01 | 0.00 | 6.86E-01 | - | - | - |
| Permanent employment | Psychosocial | 29033 | 0.07 | 0.20 | 7.30E-01 | 8.22E-01 | 242 | 28299 | -0.28 | 0.20 | 1.65E-01 | 2.90E-01 | 159 | -0.10 | 0.14 | 4.68E-01 | 0.34 | 2.18E-01 | - | - | - |
| Breakfast habits: Porridge w/o sandwich for breakfast vs not breakfast at all | Food | 4220 | -0.31 | 0.67 | 6.40E-01 | 7.73E-01 | 226 | 4296 | -0.35 | 0.63 | 5.80E-01 | 7.23E-01 | 223 | -0.33 | 0.46 | 4.69E-01 | 0.00 | 9.69E-01 | - | - | - |
| Butter for cooking | Food | 29503 | -0.12 | 0.09 | 1.56E-01 | 3.00E-01 | 142 | 28758 | 0.04 | 0.09 | 6.83E-01 | 8.07E-01 | 236 | -0.04 | 0.06 | 4.70E-01 | 0.40 | 1.98E-01 | - | - | - |
| For how much of the time during the last four weeks have you felt worn out? | General health | 14384 | 0.16 | 0.14 | 2.56E-01 | 4.47E-01 | 156 | 14169 | -0.02 | 0.14 | 8.98E-01 | 9.14E-01 | 274 | 0.07 | 0.10 | 4.70E-01 | 0.00 | 3.74E-01 | - | - | - |
| Chips, popcorn, salted nuts | Food | 29503 | -0.05 | 0.09 | 5.56E-01 | 7.12E-01 | 213 | 28758 | -0.04 | 0.09 | 6.72E-01 | 8.07E-01 | 232 | -0.04 | 0.06 | 4.73E-01 | 0.00 | 9.14E-01 | - | - | - |
| Confidence status | Psychosocial | 24500 | -0.04 | 0.10 | 7.01E-01 | 8.12E-01 | 235 | 24058 | -0.07 | 0.11 | 5.39E-01 | 6.92E-01 | 217 | -0.05 | 0.07 | 4.81E-01 | 0.00 | 8.63E-01 | - | - | - |
| Long-term sickness | General health | 31236 | 0.09 | 0.23 | 6.97E-01 | 8.12E-01 | 231 | 30499 | -0.33 | 0.23 | 1.63E-01 | 2.90E-01 | 157 | -0.11 | 0.16 | 4.84E-01 | 0.38 | 2.05E-01 | - | - | - |
| Pizza | Food | 29503 | -0.05 | 0.09 | 5.79E-01 | 7.27E-01 | 217 | 28758 | -0.04 | 0.09 | 6.93E-01 | 8.12E-01 | 238 | -0.04 | 0.06 | 5.01E-01 | 0.00 | 9.16E-01 | - | - | - |
| Distance to work in kilometers (one way) | Physical activity | 28813 | -0.01 | 0.09 | 9.30E-01 | 9.51E-01 | 267 | 28111 | 0.09 | 0.09 | 3.08E-01 | 4.75E-01 | 181 | 0.04 | 0.06 | 5.13E-01 | 0.00 | 4.32E-01 | - | - | - |
| Medium beer | Alcohol | 29503 | -0.07 | 0.09 | 4.39E-01 | 6.24E-01 | 192 | 28758 | -0.01 | 0.09 | 8.91E-01 | 9.11E-01 | 272 | -0.04 | 0.06 | 5.17E-01 | 0.00 | 6.55E-01 | - | - | - |
| Vitamin B12 intake (ug/day) | Nutrients | 29503 | -0.06 | 0.08 | 4.90E-01 | 6.69E-01 | 200 | 28758 | -0.01 | 0.09 | 8.73E-01 | 9.01E-01 | 270 | -0.04 | 0.06 | 5.45E-01 | 0.00 | 7.13E-01 | - | - | - |
| People to ask for help apart from the ones at home | Social | 33508 | 0.16 | 0.27 | 5.43E-01 | 7.04E-01 | 210 | 32792 | 0.06 | 0.27 | 8.25E-01 | 8.85E-01 | 259 | 0.11 | 0.19 | 5.57E-01 | 0.00 | 7.87E-01 | - | - | - |
| Brown beans, pea soup | Food | 29503 | 0.00 | 0.08 | 9.94E-01 | 9.98E-01 | 271 | 28758 | -0.07 | 0.09 | 4.13E-01 | 5.78E-01 | 199 | -0.03 | 0.06 | 5.63E-01 | 0.00 | 5.61E-01 | - | - | - |
| Salty fish | Food | 29503 | 0.08 | 0.08 | 3.49E-01 | 5.58E-01 | 170 | 28758 | -0.02 | 0.09 | 8.59E-01 | 8.97E-01 | 267 | 0.03 | 0.06 | 5.76E-01 | 0.00 | 4.39E-01 | - | - | - |
| Cookies, pastry | Food | 29503 | 0.00 | 0.09 | 9.98E-01 | 9.98E-01 | 272 | 28758 | -0.07 | 0.09 | 4.36E-01 | 5.97E-01 | 204 | -0.03 | 0.06 | 5.85E-01 | 0.00 | 5.79E-01 | - | - | - |
| Steak, chop, e.g. | Food | 29503 | -0.02 | 0.08 | 7.76E-01 | 8.49E-01 | 249 | 28758 | -0.04 | 0.09 | 6.31E-01 | 7.76E-01 | 227 | -0.03 | 0.06 | 5.90E-01 | 0.00 | 8.84E-01 | - | - | - |
| Sour milk, yoghurt (low fat) | Food | 29503 | 0.04 | 0.08 | 6.21E-01 | 7.67E-01 | 221 | 28758 | 0.02 | 0.09 | 8.11E-01 | 8.77E-01 | 258 | 0.03 | 0.06 | 6.02E-01 | 0.00 | 8.61E-01 | - | - | - |
| Satisfaction with work situation | Psychosocial | 24226 | -0.05 | 0.10 | 6.27E-01 | 7.68E-01 | 222 | 23786 | -0.02 | 0.10 | 8.09E-01 | 8.77E-01 | 257 | -0.04 | 0.07 | 6.05E-01 | 0.00 | 8.67E-01 | - | - | - |
| Sleep status | Sleep | 24541 | 0.08 | 0.10 | 4.51E-01 | 6.32E-01 | 194 | 24119 | -0.16 | 0.11 | 1.33E-01 | 2.45E-01 | 151 | -0.04 | 0.07 | 6.10E-01 | 0.61 | 1.09E-01 | - | - | - |
| Pancake, waffle, Swedish dumpling | Food | 29503 | 0.03 | 0.09 | 7.01E-01 | 8.12E-01 | 234 | 28758 | 0.02 | 0.09 | 8.37E-01 | 8.86E-01 | 262 | 0.03 | 0.06 | 6.75E-01 | 0.00 | 9.03E-01 | - | - | - |
| Marital status: Single vs Widow/widower | Social | 3565 | 0.36 | 0.98 | 7.18E-01 | 8.16E-01 | 240 | 3525 | 0.22 | 0.99 | 8.27E-01 | 8.85E-01 | 260 | 0.29 | 0.70 | 6.81E-01 | 0.00 | 9.21E-01 | - | - | - |
| Butter on bread | Food | 29503 | 0.02 | 0.08 | 8.42E-01 | 8.91E-01 | 258 | 28758 | 0.03 | 0.08 | 7.17E-01 | 8.32E-01 | 240 | 0.02 | 0.06 | 6.92E-01 | 0.00 | 9.06E-01 | - | - | - |
| Breakfast habits: Only coffee/tea for breakfast vs not breakfast at all | Food | 1910 | 0.74 | 0.84 | 3.80E-01 | 5.86E-01 | 177 | 1988 | -0.25 | 0.82 | 7.58E-01 | 8.56E-01 | 247 | 0.23 | 0.59 | 6.94E-01 | 0.00 | 3.98E-01 | - | - | - |
| Close relationship with anyone | Social | 33225 | 0.07 | 0.08 | 3.74E-01 | 5.80E-01 | 176 | 32486 | -0.03 | 0.08 | 7.24E-01 | 8.34E-01 | 242 | 0.02 | 0.06 | 7.03E-01 | 0.00 | 3.80E-01 | - | - | - |
| Frequency of shoverling snow during leisure time | Physical activity | 20775 | -0.08 | 0.11 | 4.52E-01 | 6.32E-01 | 195 | 20235 | 0.03 | 0.11 | 7.81E-01 | 8.65E-01 | 252 | -0.03 | 0.08 | 7.31E-01 | 0.00 | 4.69E-01 | - | - | - |
| Work shifts/weekends | Psychosocial | 32038 | -0.29 | 0.20 | 1.51E-01 | 2.96E-01 | 138 | 31449 | 0.38 | 0.20 | 6.05E-02 | 1.41E-01 | 120 | 0.04 | 0.14 | 7.62E-01 | 0.82 | 1.91E-02 | - | - | - |
| Cohabitation: Live alone vs Other/others | Social | 3825 | -0.51 | 0.83 | 5.42E-01 | 7.04E-01 | 209 | 3823 | 0.17 | 0.85 | 8.38E-01 | 8.86E-01 | 263 | -0.17 | 0.60 | 7.70E-01 | 0.00 | 5.66E-01 | - | - | - |
| Cambridge physical activity index | Physical activity | 31834 | 0.00 | 0.08 | 9.64E-01 | 9.75E-01 | 270 | 31155 | 0.04 | 0.09 | 6.47E-01 | 7.85E-01 | 230 | 0.02 | 0.06 | 7.73E-01 | 0.00 | 7.20E-01 | - | - | - |
| Travel to work: Irregular travel mode to work vs passive travel to work | Physical activity | 21470 | -0.20 | 0.30 | 4.93E-01 | 6.70E-01 | 201 | 21105 | 0.09 | 0.30 | 7.54E-01 | 8.56E-01 | 246 | -0.06 | 0.21 | 7.89E-01 | 0.00 | 4.81E-01 | - | - | - |
| For how much of the time during the last four weeks have you felt calm and serene? | General health | 14406 | 0.02 | 0.14 | 8.67E-01 | 9.10E-01 | 260 | 14220 | 0.03 | 0.14 | 8.46E-01 | 8.90E-01 | 265 | 0.03 | 0.10 | 7.98E-01 | 0.00 | 9.82E-01 | - | - | - |
| Physical limitation that made you do less than you wanted during the last four weeks | General health | 14429 | -0.13 | 0.35 | 6.98E-01 | 8.12E-01 | 232 | 14234 | 0.27 | 0.35 | 4.50E-01 | 6.04E-01 | 208 | 0.06 | 0.25 | 8.03E-01 | 0.00 | 4.17E-01 | - | - | - |
| Milk, sour milk (3%) | Beverage | 29503 | 0.10 | 0.09 | 2.64E-01 | 4.56E-01 | 158 | 28758 | -0.07 | 0.09 | 4.26E-01 | 5.86E-01 | 203 | 0.01 | 0.06 | 8.08E-01 | 0.45 | 1.77E-01 | - | - | - |
| Number of friends that can come to your home at any time and feel at home | Social | 33143 | 0.06 | 0.09 | 5.21E-01 | 6.83E-01 | 208 | 32434 | -0.04 | 0.09 | 6.82E-01 | 8.07E-01 | 235 | 0.01 | 0.06 | 8.62E-01 | 0.00 | 4.58E-01 | - | - | - |
| Smoked fish/meat | Food | 29503 | 0.05 | 0.08 | 5.73E-01 | 7.24E-01 | 216 | 28758 | -0.03 | 0.08 | 7.12E-01 | 8.31E-01 | 239 | 0.01 | 0.06 | 8.82E-01 | 0.00 | 5.11E-01 | - | - | - |
| Sausage as main dish | Food | 29503 | 0.03 | 0.09 | 7.02E-01 | 8.12E-01 | 236 | 28758 | -0.05 | 0.09 | 5.58E-01 | 7.11E-01 | 219 | -0.01 | 0.06 | 8.90E-01 | 0.00 | 4.93E-01 | - | - | - |
| Liquor, spirits | Alcohol | 29503 | -0.05 | 0.10 | 5.81E-01 | 7.27E-01 | 218 | 28758 | 0.06 | 0.09 | 5.16E-01 | 6.72E-01 | 214 | 0.01 | 0.07 | 9.30E-01 | 0.00 | 3.96E-01 | - | - | - |
| Snuff status: Former snuff users vs non-snuff users | Tobacco use | 26807 | 0.21 | 0.32 | 5.03E-01 | 6.72E-01 | 204 | 26154 | -0.26 | 0.33 | 4.19E-01 | 5.84E-01 | 200 | -0.02 | 0.23 | 9.33E-01 | 0.09 | 2.95E-01 | - | - | - |
| Tomato, cucumber | Food | 29503 | -0.06 | 0.09 | 5.03E-01 | 6.72E-01 | 203 | 28758 | 0.05 | 0.09 | 5.69E-01 | 7.18E-01 | 221 | -0.01 | 0.06 | 9.34E-01 | 0.00 | 3.81E-01 | - | - | - |
| Vitamin E intake (mg/day) | Nutrients | 29503 | 0.03 | 0.08 | 7.50E-01 | 8.32E-01 | 246 | 28758 | -0.04 | 0.09 | 6.76E-01 | 8.07E-01 | 233 | 0.00 | 0.06 | 9.49E-01 | 0.00 | 6.02E-01 | - | - | - |
| Control over own work assignment | Psychosocial | 33124 | 0.01 | 0.08 | 9.46E-01 | 9.60E-01 | 269 | 32482 | -0.01 | 0.08 | 8.73E-01 | 9.01E-01 | 269 | 0.00 | 0.06 | 9.49E-01 | 0.00 | 8.72E-01 | - | - | - |
| Physical limitation that limited your ability to perform certain work tasks or other activities during the last four weeks | General health | 14401 | 0.00 | 0.40 | 9.98E-01 | 9.98E-01 | 273 | 14183 | 0.03 | 0.41 | 9.44E-01 | 9.51E-01 | 277 | 0.01 | 0.28 | 9.60E-01 | 0.00 | 9.61E-01 | - | - | - |
| For how much of the time during the last four weeks have you felt full of energy? | General health | 14389 | 0.07 | 0.14 | 5.96E-01 | 7.40E-01 | 220 | 14198 | -0.07 | 0.14 | 6.28E-01 | 7.75E-01 | 226 | 0.00 | 0.10 | 9.70E-01 | 0.00 | 4.73E-01 | - | - | - |
| Meat stew | Food | 29503 | -0.06 | 0.08 | 5.05E-01 | 6.72E-01 | 205 | 28758 | 0.05 | 0.09 | 5.29E-01 | 6.83E-01 | 216 | 0.00 | 0.06 | 9.72E-01 | 0.00 | 3.59E-01 | - | - | - |
| Fried potatoes, pommes frites | Food | 29503 | -0.02 | 0.09 | 8.42E-01 | 8.91E-01 | 257 | 28758 | 0.02 | 0.09 | 8.06E-01 | 8.77E-01 | 256 | 0.00 | 0.06 | 9.74E-01 | 0.00 | 7.53E-01 | - | - | - |
| Physical limitation that made you not being able to perform certain work tasks or other activities during the last four weeks | General health | 14373 | -0.11 | 0.39 | 7.75E-01 | 8.49E-01 | 248 | 14178 | 0.12 | 0.39 | 7.51E-01 | 8.56E-01 | 244 | 0.00 | 0.28 | 9.86E-01 | 0.00 | 6.69E-01 | - | - | - |
| Vitamin A intake (mg/day) | Nutrients | 29503 | 0.03 | 0.09 | 7.69E-01 | 8.49E-01 | 247 | 28758 | -0.03 | 0.10 | 7.51E-01 | 8.56E-01 | 243 | 0.00 | 0.07 | 9.89E-01 | 0.00 | 6.65E-01 | - | - | - |
| Amount of alcohol drunk in a day | Alcohol | 10932 | 0.54 | 0.17 | - | - | 277 | 10737 | 0.88 | 0.17 | 3.20E-07 | **6.87E-06** | 13 | 0.71 | 0.12 | - | 0.49 | 1.62E-01 | - | - | - |
| Snore during sleep | Sleep | 10534 | 0.63 | 0.17 | - | - | 286 | 10436 | 0.62 | 0.17 | 3.23E-04 | **2.57E-03** | 35 | 0.62 | 0.12 | - | 0.00 | 9.83E-01 | - | - | - |
| Risk of sleeping while sitting and reading | Sleep | 11181 | -0.54 | 0.16 | - | - | 280 | 11044 | -0.43 | 0.16 | 6.64E-03 | **2.65E-02** | 70 | -0.49 | 0.11 | - | 0.00 | 6.39E-01 | - | - | - |
| Breath-holds during sleep | Sleep | 7912 | 0.24 | 0.20 | - | - | 287 | 7758 | 0.52 | 0.20 | 9.85E-03 | **3.62E-02** | 76 | 0.37 | 0.14 | - | 0.00 | 3.19E-01 | - | - | - |
| Risk of sleeping while sitting still after having lunch | Sleep | 11263 | -0.47 | 0.16 | - | - | 285 | 11137 | -0.30 | 0.16 | 6.10E-02 | 1.41E-01 | 121 | -0.39 | 0.11 | - | 0.00 | 4.40E-01 | - | - | - |
| Shellfish (e.g. shrimps, scallops) | Food | 6988 | -0.16 | 0.17 | 3.47E-01 | 5.58E-01 | 169 | 6762 | -0.32 | 0.17 | 1.31E-01 | 2.45E-01 | 149 | -0.24 | 0.12 | - | 0.00 | 5.04E-01 | - | - | - |
| Risk of sleeping while lying down resting in the afternoon | Sleep | 11255 | -0.58 | 0.16 | - | - | 284 | 11131 | -0.21 | 0.16 | 1.80E-01 | 3.09E-01 | 163 | -0.40 | 0.11 | - | 0.62 | 1.05E-01 | - | - | - |
| Mixed frozen vegetables | Food | 6988 | 0.07 | 0.17 | 6.67E-01 | 7.99E-01 | 228 | 6762 | 0.27 | 0.17 | 1.83E-01 | 3.10E-01 | 165 | 0.17 | 0.12 | - | 0.00 | 4.03E-01 | - | - | - |
| Soft whey cheese | Food | 6988 | 0.14 | 0.16 | 4.00E-01 | 6.07E-01 | 180 | 6762 | -0.24 | 0.17 | 2.22E-01 | 3.60E-01 | 172 | -0.04 | 0.12 | - | 0.63 | 1.01E-01 | - | - | - |
| Blood based food | Food | 6988 | -0.05 | 0.16 | 7.47E-01 | 8.32E-01 | 245 | 6762 | 0.20 | 0.17 | 3.05E-01 | 4.73E-01 | 180 | 0.07 | 0.12 | - | 0.12 | 2.86E-01 | - | - | - |
| Soft cheese | Food | 6988 | 0.21 | 0.16 | 2.07E-01 | 3.76E-01 | 150 | 6762 | -0.10 | 0.16 | 5.65E-01 | 7.17E-01 | 220 | 0.06 | 0.12 | - | 0.47 | 1.69E-01 | - | - | - |
| Blota (broth + bread) | Food | 6988 | 0.58 | 0.16 | 3.20E-03 | **1.65E-02** | 53 | 6762 | 0.07 | 0.16 | 6.92E-01 | 8.12E-01 | 237 | 0.33 | 0.12 | - | 0.79 | 2.78E-02 | - | - | - |
| Liver, kidney | Food | 6988 | 0.15 | 0.16 | 3.69E-01 | 5.76E-01 | 175 | 6762 | 0.03 | 0.17 | 8.64E-01 | 9.00E-01 | 268 | 0.09 | 0.12 | - | 0.00 | 6.00E-01 | - | - | - |
| Mashed potato | Food | 6988 | -0.04 | 0.17 | 8.25E-01 | 8.90E-01 | 253 | 6762 | -0.03 | 0.17 | 8.75E-01 | 9.01E-01 | 271 | -0.03 | 0.12 | - | 0.00 | 9.71E-01 | - | - | - |
| Time spent in a week in moderately steneous activities | Physical activity | 11059 | -0.23 | 0.16 | - | - | 274 | 10871 | -0.20 | 0.16 | - | - | 280 | -0.22 | 0.11 | - | 0.00 | 9.04E-01 | - | - | - |

| | | | | | | | | | | | | | | | | | | | | | |
|---|---|---|---|---|---|---|---|---|---|---|---|---|---|---|---|---|---|---|---|---|---|
| Amount of exercise during the last 12 months | Physical activity | 11403 | -0.41 | 0.16 | - | - | 275 | 11231 | -0.33 | 0.16 | - | - | 281 | -0.37 | 0.11 | - | 0.00 | 7.23E-01 | - | - | - |
| Frequency of alcohol consumption | Alcohol | 11446 | -0.28 | 0.16 | - | - | 276 | 11305 | 0.00 | 0.16 | - | - | 282 | -0.14 | 0.11 | - | 0.37 | 2.09E-01 | - | - | - |
| Frequency of drinking six or more glasses at the same occasion | Alcohol | 11137 | 0.59 | 0.17 | - | - | 278 | 10952 | 0.92 | 0.17 | - | - | 283 | 0.75 | 0.12 | - | 0.48 | 1.64E-01 | - | - | - |
| Times during last year that you felt guilty because of your drinking | Alcohol | 11124 | 0.09 | 0.16 | - | - | 279 | 10924 | 0.23 | 0.16 | - | - | 284 | 0.16 | 0.11 | - | 0.00 | 5.35E-01 | - | - | - |
| Risk of sleeping while watching TV | Sleep | 11303 | 0.19 | 0.16 | - | - | 281 | 11162 | 0.16 | 0.16 | - | - | 285 | 0.17 | 0.11 | - | 0.00 | 9.06E-01 | - | - | - |
| Risk of sleeping while sitting inactive in a public place | Sleep | 11227 | -0.58 | 0.16 | - | - | 282 | 11090 | -0.42 | 0.16 | - | - | 286 | -0.50 | 0.11 | - | 0.00 | 4.69E-01 | - | - | - |
| Risk of sleeping as a passenger in a car for one hour without break | Sleep | 11188 | -0.97 | 0.16 | - | - | 283 | 11045 | -0.64 | 0.16 | - | - | 287 | -0.81 | 0.11 | - | 0.52 | 1.49E-01 | - | - | - |

**Supplementary Table 5**. Linear mixed model association results for DBP

| | | Training set | | | | | | Testing set | | | | | | Metanalysis | | | | | R2 | | |
|---|---|---|---|---|---|---|---|---|---|---|---|---|---|---|---|---|---|---|---|---|---|
| **Description** | **Group** | **N** | **Effect estimate** | **S.E.** | **p-value** | **p-value$_{FDR}$** | **p-value rank** | **N** | **Effect estimate** | **S.E.** | **p-value** | **p-value$_{FDR}$** | **p-value rank** | **Effect estimate** | **S.E.** | **p-value** | **$I^2$** | **Q p-value** | **marginal R2** | **conditional R2** | **R2 rank** |
| Informed of having high blood pressure | General health | 33435 | 10.90 | 0.13 | <2.22E-308 | **<2.22E-308** | 1 | 32717 | 10.76 | 0.13 | <2.22E-308 | **<2.22E-308** | 1 | 10.83 | 0.09 | <2.22E-308 | 0.00 | 4.31E-01 | 0.34 | 0.55 | 1 |
| Self rate of overall health | General health | 14438 | -0.82 | 0.09 | 2.59E-19 | **3.53E-17** | 2 | 14266 | -0.87 | 0.09 | 7.83E-21 | **7.28E-19** | 3 | -0.85 | 0.06 | 1.67E-38 | 0.00 | 6.91E-01 | 0.18 | 0.59 | 26 |
| Excellent health | General health | 14384 | -0.73 | 0.09 | 8.27E-16 | **5.65E-14** | 4 | 14208 | -0.90 | 0.09 | 3.38E-22 | **4.71E-20** | 2 | -0.82 | 0.06 | 3.71E-36 | 0.48 | 1.66E-01 | 0.18 | 0.59 | 28 |
| Parents or siblings had a cerebral hemorrhage/thrombosis or cardiac infarction before the age of 60 | General health | 32678 | 1.27 | 0.15 | 6.35E-17 | **5.78E-15** | 3 | 32014 | 1.27 | 0.15 | 1.70E-16 | **1.19E-14** | 4 | 1.27 | 0.11 | 8.09E-32 | 0.00 | 9.94E-01 | 0.18 | 0.57 | 11 |
| Overall state of health during the last year | General health | 33255 | -0.41 | 0.05 | 4.85E-14 | **2.29E-12** | 5 | 32542 | -0.42 | 0.06 | 6.24E-14 | **2.90E-12** | 6 | -0.41 | 0.04 | 2.08E-26 | 0.00 | 9.02E-01 | 0.18 | 0.57 | 15 |
| Smoking status: Smokers vs non-smokers | Tobacco use | 21432 | -1.45 | 0.19 | 7.22E-14 | **2.82E-12** | 7 | 21071 | -1.43 | 0.19 | 1.42E-13 | **5.68E-12** | 7 | -1.44 | 0.14 | 6.96E-26 | 0.00 | 9.57E-01 | 0.18 | 0.57 | 27 |
| Educational level | Psychosocial | 33602 | -0.43 | 0.07 | 9.85E-11 | **3.36E-09** | 8 | 32893 | -0.49 | 0.07 | 9.35E-13 | **2.90E-11** | 9 | -0.46 | 0.05 | 6.68E-22 | 0.00 | 5.74E-01 | 0.18 | 0.57 | 12 |
| Years smoking | Tobacco use | 29313 | -0.54 | 0.07 | 5.04E-14 | **2.29E-12** | 6 | 28558 | -0.43 | 0.07 | 4.06E-09 | **1.03E-07** | 11 | -0.49 | 0.05 | 2.19E-21 | 0.13 | 2.83E-01 | 0.18 | 0.57 | 22 |
| Overall state of health compared to other your age | General health | 16616 | -0.42 | 0.08 | 1.16E-07 | **2.27E-06** | 14 | 16190 | -0.62 | 0.08 | 1.49E-14 | **8.34E-13** | 5 | -0.52 | 0.06 | 4.51E-20 | 0.70 | 6.88E-02 | 0.16 | 0.54 | 37 |
| Fitness status | Physical activity | 24494 | -0.44 | 0.07 | 1.66E-10 | **5.04E-09** | 9 | 24064 | -0.41 | 0.07 | 1.04E-08 | **2.24E-07** | 13 | -0.43 | 0.05 | 1.03E-17 | 0.00 | 7.04E-01 | 0.18 | 0.60 | 25 |
| Marital status: Single vs Married/partner | Social | 30820 | -0.94 | 0.20 | 3.54E-06 | **6.17E-05** | 15 | 30180 | -1.47 | 0.20 | 6.87E-13 | **2.39E-11** | 8 | -1.20 | 0.14 | 6.70E-17 | 0.70 | 6.73E-02 | 0.18 | 0.57 | 18 |
| Average portion size of vegetables based on photographic illustration of four sizes (smallest to largest) | Food | 29486 | -0.35 | 0.06 | 7.57E-09 | **1.88E-07** | 11 | 28746 | -0.36 | 0.06 | 1.43E-08 | **2.85E-07** | 14 | -0.36 | 0.04 | 5.72E-16 | 0.00 | 9.72E-01 | 0.18 | 0.58 | 32 |
| Cohabitation: Live alone vs Adult and children | Social | 19380 | -1.13 | 0.21 | 1.07E-07 | **2.26E-06** | 13 | 18924 | -1.18 | 0.21 | 3.58E-08 | **6.24E-07** | 16 | -1.15 | 0.15 | 1.96E-14 | 0.00 | 8.65E-01 | 0.18 | 0.56 | 16 |
| Number of cigarretes smoked per day (in groups) | Tobacco use | 26959 | -0.38 | 0.07 | 2.73E-08 | **6.22E-07** | 12 | 26547 | -0.34 | 0.07 | 8.85E-07 | **1.37E-05** | 18 | -0.36 | 0.05 | 1.27E-13 | 0.00 | 6.97E-01 | 0.18 | 0.57 | 30 |
| Average portion size of potatoes/rice/pasta based on photographic illustration of four sizes (smallest to largest) | Food | 29486 | -0.38 | 0.06 | 1.56E-09 | **4.27E-08** | 10 | 28746 | -0.29 | 0.07 | 1.16E-05 | **1.47E-04** | 22 | -0.34 | 0.05 | 1.53E-13 | 0.14 | 2.82E-01 | 0.18 | 0.58 | 33 |
| As healthy as anyone | General health | 14341 | -0.30 | 0.09 | 5.74E-04 | **4.90E-03** | 32 | 14163 | -0.59 | 0.09 | 5.35E-11 | **1.49E-09** | 10 | -0.44 | 0.06 | 1.30E-12 | 0.82 | 1.92E-02 | 0.18 | 0.59 | 34 |
| Brewed (filtered) coffee | Beverage | 29486 | -0.24 | 0.06 | 8.14E-05 | **1.11E-03** | 20 | 28746 | -0.33 | 0.06 | 5.17E-08 | **8.49E-07** | 17 | -0.28 | 0.04 | 3.52E-11 | 0.24 | 2.51E-01 | 0.18 | 0.59 | 4 |
| Repetitive job | Psychosocial | 33066 | 0.16 | 0.06 | 6.02E-03 | **2.93E-02** | 56 | 32423 | 0.32 | 0.06 | 2.24E-08 | **4.17E-07** | 15 | 0.24 | 0.04 | 4.06E-09 | 0.76 | 3.95E-02 | 0.18 | 0.57 | 19 |
| Iodine intake (ug/day) | Nutrients | 29486 | -0.26 | 0.06 | 3.62E-06 | **6.17E-05** | 16 | 28746 | -0.20 | 0.06 | 7.87E-04 | **5.32E-03** | 41 | -0.23 | 0.04 | 1.51E-08 | 0.00 | 4.07E-01 | 0.18 | 0.59 | 6 |
| Participation in associations or voluntary organisations | Social | 33044 | -0.43 | 0.11 | 1.27E-04 | **1.51E-03** | 23 | 32366 | -0.47 | 0.11 | 4.68E-05 | **4.67E-04** | 28 | -0.45 | 0.08 | 2.32E-08 | 0.00 | 8.23E-01 | 0.18 | 0.57 | 20 |
| Frequency of hunting or fishing during leisure time | Physical activity | 20124 | 0.30 | 0.08 | 1.79E-04 | **2.03E-03** | 24 | 19561 | 0.32 | 0.08 | 8.06E-05 | **7.76E-04** | 29 | 0.31 | 0.06 | 5.45E-08 | 0.00 | 8.49E-01 | 0.16 | 0.57 | 36 |
| Marital status: Single vs Divorced/separated | Social | 5517 | -0.86 | 0.31 | 4.94E-03 | **2.59E-02** | 52 | 5522 | -1.47 | 0.31 | 1.88E-06 | **2.76E-05** | 19 | -1.16 | 0.22 | 8.57E-08 | 0.50 | 1.58E-01 | 0.20 | 0.55 | 3 |
| Smoking status: Former smokers vs non-smokers | Tobacco use | 23981 | -0.77 | 0.17 | 4.11E-06 | **6.59E-05** | 17 | 23210 | -0.48 | 0.17 | 5.37E-03 | **2.42E-02** | 62 | -0.63 | 0.12 | 1.59E-07 | 0.33 | 2.21E-01 | 0.18 | 0.58 | 17 |
| Satisfaction with economy | Psychosocial | 24507 | 0.30 | 0.07 | 1.09E-05 | **1.66E-04** | 18 | 24078 | 0.20 | 0.07 | 3.77E-03 | **1.84E-02** | 57 | 0.25 | 0.05 | 2.35E-07 | 0.02 | 3.12E-01 | 0.18 | 0.60 | 29 |
| High mental demand from job | Psychosocial | 32867 | 0.18 | 0.06 | 9.95E-04 | **7.76E-03** | 35 | 32270 | 0.22 | 0.06 | 1.36E-04 | **1.18E-03** | 32 | 0.20 | 0.04 | 5.12E-07 | 0.00 | 6.72E-01 | 0.18 | 0.57 | 21 |
| Whole grain crisp bread | Food | 29486 | 0.24 | 0.06 | 6.11E-05 | **8.78E-04** | 19 | 28746 | 0.18 | 0.06 | 2.99E-03 | **1.50E-02** | 55 | 0.21 | 0.04 | 7.71E-07 | 0.00 | 5.01E-01 | 0.18 | 0.59 | 5 |
| Exercise during the last three months | Physical activity | 32872 | -0.20 | 0.06 | 3.94E-04 | **3.84E-03** | 28 | 32163 | -0.19 | 0.06 | 8.01E-04 | **5.32E-03** | 42 | -0.19 | 0.04 | 1.07E-06 | 0.00 | 9.37E-01 | 0.18 | 0.57 | 24 |
| Total energy intake (kcal/day) | Nutrients | 29486 | -0.21 | 0.07 | 2.15E-03 | **1.43E-02** | 41 | 28746 | -0.26 | 0.07 | 1.51E-04 | **1.28E-03** | 33 | -0.24 | 0.05 | 1.27E-06 | 0.00 | 5.74E-01 | 0.18 | 0.59 | 10 |
| Physical limitation to participate in steneous activities: running, lifting heavy objects, taking part in physically demanding sports | General health | 14406 | 0.25 | 0.09 | 6.14E-03 | **2.93E-02** | 57 | 14241 | 0.38 | 0.09 | 4.45E-05 | **4.67E-04** | 25 | 0.31 | 0.06 | 1.42E-06 | 0.01 | 3.14E-01 | 0.18 | 0.59 | 35 |
| Cohabitation: Live alone vs Only children | Social | 4913 | -1.15 | 0.35 | 9.07E-04 | **7.28E-03** | 34 | 4904 | -1.16 | 0.34 | 7.16E-04 | **5.19E-03** | 38 | -1.16 | 0.24 | 2.15E-06 | 0.00 | 9.79E-01 | 0.20 | 0.56 | 2 |
| Snuff status: Snuff users vs non-snuff users | Tobacco use | 28550 | -0.72 | 0.18 | 8.89E-05 | **1.16E-03** | 21 | 28045 | -0.50 | 0.19 | 8.10E-03 | **3.18E-02** | 71 | -0.62 | 0.13 | 3.23E-06 | 0.00 | 4.04E-01 | 0.18 | 0.57 | 31 |
| Phosphate intake (mg/day) | Nutrients | 29486 | -0.16 | 0.06 | 3.68E-03 | **2.09E-02** | 48 | 28746 | -0.21 | 0.06 | 2.39E-04 | **1.96E-03** | 34 | -0.19 | 0.04 | 3.41E-06 | 0.00 | 5.46E-01 | 0.18 | 0.59 | 9 |
| Physical limitation to participate in moderately demanding activities: walking up several stairs | General health | 14405 | 0.21 | 0.09 | 1.72E-02 | 6.54E-02 | 72 | 14222 | 0.37 | 0.09 | 4.56E-05 | **4.67E-04** | 26 | 0.29 | 0.06 | 4.96E-06 | 0.36 | 2.11E-01 | - | - | - |
| Frequncy of social contacts with colleagues during leisure time | Psychosocial | 31132 | 0.20 | 0.06 | 2.43E-04 | **2.55E-03** | 26 | 30483 | 0.15 | 0.06 | 6.41E-03 | **2.75E-02** | 65 | 0.18 | 0.04 | 5.95E-06 | 0.00 | 5.32E-01 | 0.18 | 0.57 | 14 |
| Selenium intake (ug/day) | Nutrients | 29486 | -0.19 | 0.06 | 8.24E-04 | **6.81E-03** | 33 | 28746 | -0.18 | 0.06 | 2.70E-03 | **1.40E-02** | 54 | -0.19 | 0.04 | 7.15E-06 | 0.00 | 8.54E-01 | 0.18 | 0.59 | 8 |
| Feel the need to reduce alcohol consumption | Alcohol | 30145 | 0.52 | 0.17 | 2.92E-03 | **1.81E-02** | 44 | 29566 | 0.56 | 0.18 | 1.36E-03 | **8.27E-03** | 46 | 0.54 | 0.12 | 1.25E-05 | 0.00 | 8.62E-01 | 0.18 | 0.58 | 13 |
| How much has the pain during the last four weeks disturbed your normal work? | General health | 14367 | 0.15 | 0.09 | 8.12E-02 | 2.02E-01 | 108 | 14182 | 0.39 | 0.09 | 1.04E-05 | **1.38E-04** | 21 | 0.27 | 0.06 | 1.32E-05 | 0.73 | 5.26E-02 | - | - | - |
| Secoisolariciresinol intake (ug/day) | Nutrients | 29486 | -0.18 | 0.06 | 3.06E-03 | **1.86E-02** | 45 | 28746 | -0.19 | 0.06 | 1.57E-03 | **9.15E-03** | 48 | -0.19 | 0.04 | 1.51E-05 | 0.00 | 8.61E-01 | 0.18 | 0.59 | 7 |
| For how much of the time during the last four weeks have you felt really alert and strong? | General health | 14376 | -0.16 | 0.09 | 5.93E-02 | 1.65E-01 | 98 | 14183 | -0.37 | 0.09 | 2.39E-05 | **2.90E-04** | 23 | -0.27 | 0.06 | 1.51E-05 | 0.65 | 8.98E-02 | - | - | - |
| Enough time for job assignments | Psychosocial | 32971 | 0.21 | 0.05 | 9.86E-05 | **1.22E-03** | 22 | 32341 | 0.12 | 0.05 | 2.81E-02 | 8.96E-02 | 87 | 0.16 | 0.04 | 1.57E-05 | 0.25 | 2.48E-01 | - | - | - |
| Satisfaction with accomodation | Psychosocial | 24527 | 0.24 | 0.06 | 2.08E-04 | **2.27E-03** | 25 | 24110 | 0.15 | 0.07 | 2.22E-02 | 7.93E-02 | 78 | 0.20 | 0.05 | 2.13E-05 | 0.00 | 3.36E-01 | - | - | - |
| Boiled or baked potato | Food | 29486 | 0.12 | 0.06 | 3.20E-02 | 1.08E-01 | 81 | 28746 | 0.22 | 0.06 | 1.30E-04 | **1.17E-03** | 31 | 0.17 | 0.04 | 2.58E-05 | 0.35 | 2.15E-01 | - | - | - |
| Contradictory demands in job | Psychosocial | 32901 | -0.11 | 0.05 | 3.93E-02 | 1.23E-01 | 87 | 32229 | -0.22 | 0.06 | 1.03E-04 | **9.60E-04** | 30 | -0.16 | 0.04 | 2.77E-05 | 0.43 | 1.85E-01 | - | - | - |
| Average portion size of meat/fish based on photographic illustration of four sizes (smallest to largest) | Food | 29486 | -0.15 | 0.06 | 1.35E-02 | 5.30E-02 | 69 | 28746 | -0.22 | 0.06 | 7.66E-04 | **5.32E-03** | 40 | -0.19 | 0.04 | 3.85E-05 | 0.00 | 4.86E-01 | - | - | - |
| Memory status | Psychosocial | 24464 | 0.24 | 0.07 | 4.09E-04 | **3.85E-03** | 29 | 24025 | 0.15 | 0.07 | 2.30E-02 | 8.07E-02 | 79 | 0.20 | 0.05 | 3.88E-05 | 0.00 | 3.92E-01 | - | - | - |
| Pain during the last four weeks | General health | 14406 | 0.11 | 0.09 | 2.17E-01 | 3.98E-01 | 149 | 14206 | 0.40 | 0.09 | 6.06E-06 | **8.45E-05** | 20 | 0.25 | 0.06 | 4.43E-05 | 0.82 | 1.74E-02 | - | - | - |
| Get sick more often than other people | General health | 14387 | 0.14 | 0.09 | 1.09E-01 | 2.47E-01 | 120 | 14202 | 0.36 | 0.09 | 4.43E-05 | **4.67E-04** | 24 | 0.25 | 0.06 | 5.69E-05 | 0.69 | 7.18E-02 | - | - | - |
| Eat breakfast from 2000 | Food | 17069 | -1.00 | 0.28 | 4.30E-04 | **3.91E-03** | 30 | 16679 | -0.60 | 0.28 | 3.17E-02 | 9.67E-02 | 91 | -0.80 | 0.20 | 6.26E-05 | 0.00 | 3.17E-01 | - | - | - |
| Number of snuff boxes per week | Tobacco use | 32372 | -0.22 | 0.06 | 5.13E-04 | **4.52E-03** | 31 | 31721 | -0.14 | 0.06 | 3.16E-02 | 9.67E-02 | 90 | -0.18 | 0.05 | 6.65E-05 | 0.00 | 3.77E-01 | - | - | - |
| Strong beer | Alcohol | 29486 | 0.13 | 0.06 | 3.06E-02 | 1.06E-01 | 79 | 28746 | 0.21 | 0.06 | 6.40E-04 | **4.83E-03** | 37 | 0.17 | 0.04 | 8.14E-05 | 0.00 | 3.69E-01 | - | - | - |
| Worsen in health in the future | General health | 14336 | 0.28 | 0.09 | 1.40E-03 | **1.03E-02** | 37 | 14160 | 0.19 | 0.09 | 2.66E-02 | 8.87E-02 | 83 | 0.24 | 0.06 | 1.27E-04 | 0.00 | 4.96E-01 | - | - | - |
| Fiber cereals | Food | 29486 | -0.12 | 0.06 | 3.81E-02 | 1.23E-01 | 85 | 28746 | -0.20 | 0.06 | 8.26E-04 | **5.36E-03** | 43 | -0.16 | 0.04 | 1.36E-04 | 0.00 | 3.37E-01 | - | - | - |
| Frequency of walking during leisure time | Physical activity | 32022 | 0.15 | 0.06 | 7.51E-03 | **3.26E-02** | 63 | 31322 | 0.16 | 0.06 | 7.46E-03 | **3.06E-02** | 67 | 0.15 | 0.04 | 1.56E-04 | 0.00 | 9.59E-01 | 0.18 | 0.57 | 23 |
| Participation in sports or physical exercise associations | Social | 10696 | -0.60 | 0.20 | 2.74E-03 | **1.74E-02** | 43 | 10356 | -0.48 | 0.21 | 2.20E-02 | 7.93E-02 | 76 | -0.54 | 0.14 | 1.79E-04 | 0.00 | 6.67E-01 | - | - | - |
| Cambridge physical activity index | Physical activity | 31817 | -0.09 | 0.06 | 9.39E-02 | 2.21E-01 | 116 | 31143 | -0.21 | 0.06 | 2.80E-04 | **2.20E-03** | 35 | -0.15 | 0.04 | 1.85E-04 | 0.51 | 1.54E-01 | - | - | - |
| White cabbage, lettuce, lettuce cabbage, spinach, borecole | Food | 29486 | -0.17 | 0.06 | 3.61E-03 | **2.09E-02** | 47 | 28746 | -0.14 | 0.06 | 1.87E-02 | 7.15E-02 | 73 | -0.16 | 0.04 | 1.94E-04 | 0.00 | 7.39E-01 | - | - | - |
| Salad dressing with oil | Food | 29486 | -0.14 | 0.06 | 1.37E-02 | 5.30E-02 | 70 | 28746 | -0.16 | 0.06 | 5.89E-03 | **2.57E-02** | 64 | -0.15 | 0.04 | 2.27E-04 | 0.00 | 7.96E-01 | - | - | - |
| Would you say that the number of people that you meet in your everyday life is enough or would you like to meet more or fewer people? | Social | 33585 | -0.15 | 0.05 | 3.42E-03 | **2.03E-02** | 46 | 32883 | -0.12 | 0.05 | 2.37E-02 | 8.17E-02 | 81 | -0.14 | 0.04 | 2.40E-04 | 0.00 | 6.58E-01 | - | - | - |
| White meat (poultry) | Food | 29486 | -0.16 | 0.06 | 5.41E-03 | **2.74E-02** | 54 | 28746 | -0.14 | 0.06 | 2.22E-02 | 7.93E-02 | 77 | -0.15 | 0.04 | 3.31E-04 | 0.00 | 7.73E-01 | - | - | - |
| Magnesium intake (mg/day) | Nutrients | 29486 | -0.14 | 0.06 | 1.99E-02 | 7.35E-02 | 74 | 28746 | -0.16 | 0.06 | 7.74E-03 | **3.13E-02** | 69 | -0.15 | 0.04 | 4.23E-04 | 0.00 | 7.72E-01 | - | - | - |
| Skill demand from job | Psychosocial | 33029 | -0.17 | 0.06 | 1.78E-03 | **1.21E-02** | 40 | 32363 | -0.10 | 0.06 | 6.51E-02 | 1.61E-01 | 113 | -0.14 | 0.04 | 4.27E-04 | 0.00 | 3.84E-01 | - | - | - |
| Frequency of picking berries or mushrooms during leisure time | Physical activity | 20550 | 0.22 | 0.07 | 2.36E-03 | **1.53E-02** | 42 | 19994 | 0.14 | 0.08 | 6.43E-02 | 1.60E-01 | 112 | 0.18 | 0.05 | 5.22E-04 | 0.00 | 4.23E-01 | - | - | - |
| Vitamin B2 intake (ug/day) | Nutrients | 29486 | -0.09 | 0.06 | 1.20E-01 | 2.59E-01 | 125 | 28746 | -0.19 | 0.06 | 1.03E-03 | **6.50E-03** | 44 | -0.14 | 0.04 | 6.63E-04 | 0.38 | 2.04E-01 | - | - | - |
| Travel to work: Cycle to work vs passive travel to work | Physical activity | 25135 | -0.30 | 0.15 | 4.85E-02 | 1.44E-01 | 92 | 24482 | -0.44 | 0.16 | 4.81E-03 | **2.24E-02** | 60 | -0.37 | 0.11 | 7.21E-04 | 0.00 | 5.23E-01 | - | - | - |
| Parents or siblings have diabetes | General health | 32719 | 0.29 | 0.15 | 5.37E-02 | 1.56E-01 | 94 | 32026 | 0.43 | 0.15 | 4.42E-03 | **2.09E-02** | 59 | 0.36 | 0.11 | 7.44E-04 | 0.00 | 5.03E-01 | - | - | - |
| Frequency of engaging in clubs, associations or study circles | Social | 21236 | -0.17 | 0.07 | 1.06E-02 | **4.38E-02** | 66 | 20707 | -0.15 | 0.07 | 3.28E-02 | 9.83E-02 | 93 | -0.16 | 0.05 | 8.97E-04 | 0.00 | 8.04E-01 | - | - | - |
| Pasta | Food | 29486 | -0.16 | 0.06 | 5.27E-03 | **2.71E-02** | 53 | 28746 | -0.11 | 0.06 | 6.72E-02 | 1.64E-01 | 114 | -0.14 | 0.04 | 1.06E-03 | 0.00 | 5.23E-01 | - | - | - |
| Do you feel important and appreciated in your home? | Psychosocial | 24348 | 0.12 | 0.07 | 7.49E-02 | 1.93E-01 | 106 | 23912 | 0.19 | 0.07 | 4.96E-03 | **2.27E-02** | 61 | 0.15 | 0.05 | 1.20E-03 | 0.00 | 4.45E-01 | - | - | - |
| Smoking status: Former occasional smokers vs non-smokers | Tobacco use | 19890 | -0.52 | 0.20 | 1.14E-02 | **4.59E-02** | 68 | 19456 | -0.40 | 0.21 | 5.18E-02 | 1.33E-01 | 109 | -0.46 | 0.15 | 1.55E-03 | 0.00 | 6.92E-01 | - | - | - |
| Satisfaction with leisure time | Psychosocial | 24473 | 0.19 | 0.07 | 4.93E-03 | **2.59E-02** | 51 | 24049 | 0.11 | 0.07 | 1.01E-01 | 2.19E-01 | 128 | 0.15 | 0.05 | 1.59E-03 | 0.00 | 4.26E-01 | - | - | - |
| For how much of the time during the last four weeks have you felt gloomy and sad? | General health | 14382 | -0.23 | 0.09 | 7.21E-03 | **3.23E-02** | 61 | 14200 | -0.15 | 0.09 | 8.54E-02 | 1.95E-01 | 122 | -0.19 | 0.06 | 1.80E-03 | 0.00 | 4.99E-01 | - | - | - |
| Number of cigarretes smoked per day | Tobacco use | 21630 | -0.17 | 0.07 | 1.38E-02 | 5.30E-02 | 71 | 21211 | -0.14 | 0.07 | 5.29E-02 | 1.34E-01 | 110 | -0.15 | 0.05 | 1.84E-03 | 0.00 | 7.38E-01 | - | - | - |
| Vitamin B3 intake (mg/day) | Nutrients | 29486 | -0.16 | 0.06 | 5.66E-03 | **2.81E-02** | 55 | 28746 | -0.10 | 0.06 | 1.07E-01 | 2.29E-01 | 130 | -0.13 | 0.04 | 1.88E-03 | 0.00 | 4.41E-01 | - | - | - |
| Sodium intake (mg/day) | Nutrients | 29485 | -0.18 | 0.06 | 1.73E-03 | **1.21E-02** | 39 | 28745 | -0.07 | 0.06 | 2.35E-01 | 3.90E-01 | 168 | -0.13 | 0.04 | 2.12E-03 | 0.44 | 1.81E-01 | - | - | - |
| Oatflake, whole wheat, rye or barley porridge | Food | 29486 | -0.19 | 0.06 | 1.02E-03 | **7.76E-03** | 36 | 28746 | -0.06 | 0.06 | 3.15E-01 | 4.75E-01 | 185 | -0.13 | 0.04 | 2.27E-03 | 0.60 | 1.15E-01 | - | - | - |
| Learn new things at job | Psychosocial | 33057 | -0.06 | 0.05 | 2.75E-01 | 4.70E-01 | 160 | 32415 | -0.18 | 0.06 | 1.20E-03 | **7.46E-03** | 45 | -0.12 | 0.04 | 2.30E-03 | 0.58 | 1.22E-01 | - | - | - |
| Sodas, soft drinks, juice | Beverage | 29486 | 0.12 | 0.06 | 3.88E-02 | 1.23E-01 | 86 | 28746 | 0.13 | 0.06 | 2.67E-02 | 8.87E-02 | 84 | 0.13 | 0.04 | 2.48E-03 | 0.00 | 8.81E-01 | - | - | - |
| Teetotaler | Alcohol | 21501 | 0.65 | 0.24 | 6.94E-03 | **3.16E-02** | 60 | 20942 | 0.39 | 0.25 | 1.23E-01 | 2.48E-01 | 138 | 0.53 | 0.17 | 2.53E-03 | 0.00 | 4.56E-01 | - | - | - |
| Rice | Food | 29486 | -0.21 | 0.06 | 3.27E-04 | **3.31E-03** | 27 | 28746 | -0.04 | 0.06 | 5.17E-01 | 6.68E-01 | 216 | -0.12 | 0.04 | 2.56E-03 | 0.76 | 3.96E-02 | - | - | - |
| Satisfaction with home and family situation | Psychosocial | 24504 | 0.14 | 0.06 | 3.09E-02 | 1.06E-01 | 80 | 24090 | 0.14 | 0.07 | 3.93E-02 | 1.12E-01 | 98 | 0.14 | 0.05 | 2.85E-03 | 0.00 | 9.74E-01 | - | - | - |
| Root vegetables, carrot | Food | 29486 | -0.07 | 0.06 | 2.78E-01 | 4.72E-01 | 161 | 28746 | -0.20 | 0.06 | 1.69E-03 | **9.63E-03** | 49 | -0.13 | 0.04 | 3.01E-03 | 0.55 | 1.35E-01 | - | - | - |
| Beta-carotene intake (mg/day) | Nutrients | 29486 | -0.07 | 0.06 | 2.47E-01 | 4.36E-01 | 153 | 28746 | -0.19 | 0.06 | 2.25E-03 | **1.26E-02** | 50 | -0.13 | 0.04 | 3.08E-03 | 0.48 | 1.67E-01 | - | - | - |
| Cream, creme fraiche, sour cream | Food | 29486 | -0.08 | 0.06 | 1.30E-01 | 2.72E-01 | 131 | 28746 | -0.15 | 0.06 | 7.46E-03 | **3.06E-02** | 68 | -0.12 | 0.04 | 3.14E-03 | 0.00 | 3.94E-01 | - | - | - |
| Self rate of overall health compared to a year ago | General health | 14326 | -0.23 | 0.09 | 7.45E-03 | **3.26E-02** | 62 | 14134 | -0.13 | 0.09 | 1.36E-01 | 2.65E-01 | 143 | -0.18 | 0.06 | 3.15E-03 | 0.00 | 4.05E-01 | - | - | - |
| Folic acid intake (ug/day) | Nutrients | 29486 | -0.13 | 0.06 | 3.74E-02 | 1.23E-01 | 83 | 28746 | -0.14 | 0.07 | 3.65E-02 | 1.07E-01 | 95 | -0.13 | 0.05 | 3.18E-03 | 0.00 | 9.55E-01 | - | - | - |
| Medioresinol intake (ug/day) | Nutrients | 29486 | 0.16 | 0.06 | 4.49E-03 | **2.45E-02** | 50 | 28746 | 0.08 | 0.06 | 1.93E-01 | 3.41E-01 | 158 | 0.12 | 0.04 | 3.25E-03 | 0.10 | 2.93E-01 | - | - | - |
| Margarine for cooking | Food | 29486 | 0.12 | 0.06 | 3.79E-02 | 1.23E-01 | 84 | 28746 | 0.12 | 0.06 | 3.70E-02 | 1.08E-01 | 96 | 0.12 | 0.04 | 3.26E-03 | 0.00 | 9.60E-01 | - | - | - |
| Sausage, liver pate on bread | Food | 29486 | 0.15 | 0.06 | 6.23E-03 | **2.93E-02** | 58 | 28746 | 0.08 | 0.06 | 1.71E-01 | 3.17E-01 | 151 | 0.12 | 0.04 | 3.56E-03 | 0.00 | 3.55E-01 | - | - | - |
| Last time a colleague visited you at home | Psychosocial | 31546 | 0.14 | 0.05 | 9.79E-03 | **4.11E-02** | 65 | 30910 | 0.08 | 0.06 | 1.46E-01 | 2.83E-01 | 144 | 0.11 | 0.04 | 4.20E-03 | 0.00 | 4.43E-01 | - | - | - |
| Breakfast habits: Porridge w/o sandwich for breakfast vs not breakfast at all | Food | 4217 | -1.10 | 0.43 | 1.12E-02 | **4.57E-02** | 67 | 4294 | -0.62 | 0.40 | 1.26E-01 | 2.50E-01 | 140 | -0.84 | 0.30 | 4.32E-03 | 0.00 | 4.21E-01 | - | - | - |

| | | | | | | | | | | | | | | | | | | | | | |
|---|---|---|---|---|---|---|---|---|---|---|---|---|---|---|---|---|---|---|---|---|---|
| Ingenuity or creativity demand from job | Psychosocial | 32969 | -0.09 | 0.06 | 1.09E-01 | 2.47E-01 | 119 | 32336 | -0.13 | 0.06 | 1.90E-02 | 7.15E-02 | 74 | -0.11 | 0.04 | 5.29E-03 | 0.00 | 5.81E-01 | - | - | - |
| For how much of the time during the last four weeks have you felt full of energy? | General health | 14378 | -0.08 | 0.09 | 3.57E-01 | 5.46E-01 | 178 | 14197 | -0.26 | 0.09 | 2.69E-03 | **1.40E-02** | 53 | -0.17 | 0.06 | 5.50E-03 | 0.55 | 1.37E-01 | - | - | - |
| Ice cream | Food | 29486 | -0.16 | 0.06 | 3.95E-03 | **2.20E-02** | 49 | 28746 | -0.06 | 0.06 | 3.37E-01 | 5.01E-01 | 187 | -0.11 | 0.04 | 6.11E-03 | 0.42 | 1.91E-01 | - | - | - |
| Monounsaturated fat intake (g/day) | Nutrients | 29486 | -0.11 | 0.06 | 7.29E-02 | 1.90E-01 | 105 | 28746 | -0.13 | 0.06 | 4.27E-02 | 1.16E-01 | 103 | -0.12 | 0.04 | 6.97E-03 | 0.00 | 8.36E-01 | - | - | - |
| Total protein intake (g/day) | Nutrients | 29486 | -0.10 | 0.06 | 8.13E-02 | 2.02E-01 | 110 | 28746 | -0.12 | 0.06 | 4.02E-02 | 1.12E-01 | 100 | -0.11 | 0.04 | 7.36E-03 | 0.00 | 7.98E-01 | - | - | - |
| Physical limitation to participate in moderately demanding activities: bending down or kneeling | General health | 14390 | 0.03 | 0.09 | 7.06E-01 | 7.94E-01 | 242 | 14209 | 0.31 | 0.09 | 7.25E-04 | **5.19E-03** | 39 | 0.17 | 0.06 | 8.08E-03 | 0.79 | 3.05E-02 | - | - | - |
| Physical limitation that reduced the normal time spent at work or in other activities during the last four weeks | General health | 14434 | 0.18 | 0.27 | 5.08E-01 | 6.70E-01 | 207 | 14230 | 0.84 | 0.27 | 2.41E-03 | **1.32E-02** | 51 | 0.51 | 0.19 | 8.76E-03 | 0.65 | 9.10E-02 | - | - | - |
| Frequency of dancing during leisure time | Physical activity | 19236 | -0.14 | 0.07 | 6.15E-02 | 1.66E-01 | 101 | 18634 | -0.14 | 0.08 | 7.06E-02 | 1.70E-01 | 116 | -0.14 | 0.05 | 9.29E-03 | 0.00 | 9.94E-01 | - | - | - |
| Everyday exercise satisfaction | Physical activity | 21813 | 0.21 | 0.07 | 1.51E-03 | **1.08E-02** | 38 | 21247 | 0.03 | 0.07 | 6.69E-01 | 7.91E-01 | 236 | 0.12 | 0.05 | 1.01E-02 | 0.73 | 5.62E-02 | - | - | - |
| Number of people with whom you can speak openly | Social | 33571 | 0.12 | 0.06 | 3.50E-02 | 1.17E-01 | 82 | 32860 | 0.09 | 0.06 | 1.29E-01 | 2.54E-01 | 141 | 0.10 | 0.04 | 1.02E-02 | 0.00 | 6.97E-01 | - | - | - |
| Physical limitation to participate in moderately demanding activities: moving a table, vacuuming, walking in the forest or gardening | General health | 14403 | 0.04 | 0.09 | 6.60E-01 | 7.70E-01 | 234 | 14238 | 0.29 | 0.09 | 1.40E-03 | **8.33E-03** | 47 | 0.16 | 0.06 | 1.04E-02 | 0.75 | 4.73E-02 | - | - | - |
| Linoleic acid intake (g/day) | Nutrients | 29486 | -0.11 | 0.06 | 5.61E-02 | 1.59E-01 | 96 | 28746 | -0.10 | 0.06 | 9.59E-02 | 2.12E-01 | 126 | -0.10 | 0.04 | 1.14E-02 | 0.00 | 8.90E-01 | - | - | - |
| Cholesterol intake (g/day) | Nutrients | 29486 | -0.09 | 0.06 | 1.21E-01 | 2.59E-01 | 127 | 28746 | -0.12 | 0.06 | 4.39E-02 | 1.17E-01 | 105 | -0.11 | 0.04 | 1.18E-02 | 0.00 | 7.21E-01 | - | - | - |
| Cheese 28% | Food | 29486 | -0.10 | 0.06 | 6.82E-02 | 1.83E-01 | 102 | 28746 | -0.10 | 0.06 | 8.51E-02 | 1.95E-01 | 121 | -0.10 | 0.04 | 1.22E-02 | 0.00 | 9.69E-01 | - | - | - |
| Mood status | Psychosocial | 24490 | 0.18 | 0.07 | 7.92E-03 | **3.38E-02** | 64 | 24064 | 0.06 | 0.07 | 3.89E-01 | 5.48E-01 | 197 | 0.12 | 0.05 | 1.25E-02 | 0.36 | 2.13E-01 | - | - | - |
| Animal based protein intake (g/day) | Nutrients | 29486 | -0.09 | 0.06 | 1.02E-01 | 2.38E-01 | 117 | 28746 | -0.11 | 0.06 | 5.76E-02 | 1.45E-01 | 111 | -0.10 | 0.04 | 1.25E-02 | 0.00 | 8.29E-01 | - | - | - |
| Energy status | Psychosocial | 24483 | 0.01 | 0.07 | 9.38E-01 | 9.56E-01 | 268 | 24038 | -0.25 | 0.07 | 2.84E-04 | **2.20E-03** | 36 | -0.12 | 0.05 | 1.29E-02 | 0.86 | 8.09E-03 | - | - | - |
| Sugar, honey, marmelade, jam | Food | 29486 | -0.11 | 0.06 | 5.69E-02 | 1.60E-01 | 97 | 28746 | -0.10 | 0.06 | 1.11E-01 | 2.35E-01 | 132 | -0.10 | 0.04 | 1.33E-02 | 0.00 | 8.50E-01 | - | - | - |
| Sour milk, yoghurt (3% fat) | Food | 29486 | -0.08 | 0.06 | 1.49E-01 | 3.00E-01 | 134 | 28746 | -0.12 | 0.06 | 3.96E-02 | 1.12E-01 | 99 | -0.10 | 0.04 | 1.35E-02 | 0.00 | 6.39E-01 | - | - | - |
| Tiamin intake (mg/day) | Nutrients | 29486 | -0.10 | 0.06 | 9.00E-02 | 2.16E-01 | 114 | 28746 | -0.11 | 0.06 | 7.30E-02 | 1.74E-01 | 117 | -0.10 | 0.04 | 1.37E-02 | 0.00 | 9.12E-01 | - | - | - |
| Low fat margarine on bread | Food | 29486 | 0.08 | 0.06 | 1.51E-01 | 3.00E-01 | 137 | 28746 | 0.12 | 0.06 | 4.38E-02 | 1.17E-01 | 104 | 0.10 | 0.04 | 1.48E-02 | 0.00 | 6.64E-01 | - | - | - |
| Boiled coffee | Beverage | 29486 | 0.11 | 0.06 | 7.15E-02 | 1.88E-01 | 104 | 28746 | 0.10 | 0.06 | 1.02E-01 | 2.20E-01 | 129 | 0.11 | 0.04 | 1.50E-02 | 0.00 | 9.27E-01 | - | - | - |
| Rosehip, sweet syrup soup | Food | 29486 | 0.03 | 0.06 | 6.27E-01 | 7.58E-01 | 226 | 28746 | 0.17 | 0.06 | 3.01E-03 | **1.50E-02** | 56 | 0.10 | 0.04 | 1.51E-02 | 0.68 | 7.65E-02 | - | - | - |
| Physical limitation that made you do less than you wanted during the last four weeks | General health | 14417 | 0.15 | 0.22 | 4.98E-01 | 6.67E-01 | 204 | 14233 | 0.61 | 0.22 | 5.71E-03 | **2.53E-02** | 63 | 0.38 | 0.15 | 1.52E-02 | 0.56 | 1.33E-01 | - | - | - |
| Oil for cooking | Food | 29486 | -0.10 | 0.06 | 7.03E-02 | 1.86E-01 | 103 | 28746 | -0.09 | 0.06 | 1.08E-01 | 2.31E-01 | 131 | -0.10 | 0.04 | 1.57E-02 | 0.00 | 9.09E-01 | - | - | - |
| Heptadecanoic acid intake (g/day) | Nutrients | 29486 | -0.08 | 0.06 | 1.80E-01 | 3.43E-01 | 142 | 28746 | -0.12 | 0.06 | 4.15E-02 | 1.14E-01 | 101 | -0.10 | 0.04 | 1.71E-02 | 0.00 | 6.04E-01 | - | - | - |
| Pentadecanoic acid intake (g/day) | Nutrients | 29486 | -0.08 | 0.06 | 1.80E-01 | 3.43E-01 | 143 | 28746 | -0.12 | 0.06 | 4.15E-02 | 1.14E-01 | 102 | -0.10 | 0.04 | 1.71E-02 | 0.00 | 6.04E-01 | - | - | - |
| Receive hugs to comfort and support you | Social | 32997 | -0.30 | 0.15 | 4.50E-02 | 1.36E-01 | 90 | 32262 | -0.20 | 0.15 | 1.85E-01 | 3.33E-01 | 155 | -0.25 | 0.11 | 1.82E-02 | 0.00 | 6.54E-01 | - | - | - |
| Changed everyday exercise during the last year | Physical activity | 21872 | 0.14 | 0.06 | 2.44E-02 | 8.88E-02 | 75 | 21317 | 0.07 | 0.07 | 2.83E-01 | 4.41E-01 | 178 | 0.11 | 0.05 | 1.83E-02 | 0.00 | 4.19E-01 | - | - | - |
| Milk, sour milk (1,5%) | Beverage | 29486 | -0.04 | 0.06 | 4.81E-01 | 6.56E-01 | 200 | 28746 | -0.16 | 0.06 | 7.90E-03 | **3.15E-02** | 70 | -0.10 | 0.04 | 1.85E-02 | 0.50 | 1.56E-01 | - | - | - |
| Physical limitation to participate in moderately demanding activities: walking more than 2 km | General health | 14411 | 0.10 | 0.09 | 2.49E-01 | 4.36E-01 | 156 | 14230 | 0.19 | 0.09 | 3.04E-02 | 9.52E-02 | 89 | 0.15 | 0.06 | 1.91E-02 | 0.00 | 4.61E-01 | - | - | - |
| Number of social interactions during a normal week | Social | 33243 | 0.10 | 0.06 | 6.04E-02 | 1.65E-01 | 100 | 32514 | 0.08 | 0.06 | 1.57E-01 | 2.97E-01 | 148 | 0.09 | 0.04 | 1.97E-02 | 0.00 | 7.63E-01 | - | - | - |
| Calcium intake (mg/day) | Nutrients | 29486 | -0.06 | 0.06 | 3.27E-01 | 5.10E-01 | 175 | 28746 | -0.14 | 0.06 | 2.04E-02 | 7.60E-02 | 75 | -0.10 | 0.04 | 2.05E-02 | 0.00 | 3.26E-01 | - | - | - |
| Meat on bread | Food | 29486 | 0.06 | 0.06 | 2.83E-01 | 4.72E-01 | 164 | 28746 | 0.13 | 0.06 | 2.75E-02 | 8.96E-02 | 85 | 0.09 | 0.04 | 2.09E-02 | 0.00 | 4.11E-01 | - | - | - |
| Sleep status | Sleep | 24524 | -0.03 | 0.07 | 6.74E-01 | 7.76E-01 | 237 | 24109 | -0.20 | 0.07 | 4.09E-03 | **1.97E-02** | 58 | -0.11 | 0.05 | 2.09E-02 | 0.68 | 7.86E-02 | - | - | - |
| Whole grain intake (g/day) | Food | 29486 | 0.13 | 0.06 | 1.82E-02 | 6.81E-02 | 73 | 28746 | 0.05 | 0.06 | 3.89E-01 | 5.48E-01 | 198 | 0.09 | 0.04 | 2.17E-02 | 0.05 | 3.05E-01 | - | - | - |
| Vision status | General health | 24501 | -0.03 | 0.07 | 6.13E-01 | 7.50E-01 | 223 | 24059 | -0.18 | 0.07 | 6.57E-03 | **2.78E-02** | 66 | -0.11 | 0.05 | 2.34E-02 | 0.60 | 1.13E-01 | - | - | - |
| High physical demand from job | Physical activity | 33194 | 0.09 | 0.06 | 1.16E-01 | 2.57E-01 | 123 | 32561 | 0.09 | 0.06 | 1.12E-01 | 2.35E-01 | 133 | 0.09 | 0.04 | 2.55E-02 | 0.00 | 9.82E-01 | - | - | - |
| Fatty fish (e.g. herring, whitefish, salmon) | Food | 29486 | -0.10 | 0.06 | 9.34E-02 | 2.21E-01 | 115 | 28746 | -0.08 | 0.06 | 1.49E-01 | 2.86E-01 | 145 | -0.09 | 0.04 | 2.72E-02 | 0.00 | 8.91E-01 | - | - | - |
| Docosahexaenoic acid (DHA) intake (g/day) | Nutrients | 29486 | -0.12 | 0.06 | 4.02E-02 | 1.25E-01 | 88 | 28746 | -0.06 | 0.06 | 3.01E-01 | 4.61E-01 | 182 | -0.09 | 0.04 | 2.81E-02 | 0.00 | 4.96E-01 | - | - | - |
| Physical limitation that made you not being able to perform certain work tasks or other activities during the last four weeks | General health | 14361 | 0.20 | 0.24 | 4.10E-01 | 5.99E-01 | 186 | 14177 | 0.56 | 0.24 | 2.31E-02 | 8.07E-02 | 80 | 0.38 | 0.17 | 2.87E-02 | 0.07 | 3.00E-01 | - | - | - |
| Coffee rolls/buns, rusk | Food | 29486 | 0.09 | 0.06 | 1.27E-01 | 2.71E-01 | 128 | 28746 | 0.09 | 0.06 | 1.20E-01 | 2.44E-01 | 137 | 0.09 | 0.04 | 2.93E-02 | 0.00 | 9.55E-01 | - | - | - |
| Tomato, cucumber | Food | 29486 | -0.08 | 0.06 | 1.70E-01 | 3.32E-01 | 140 | 28746 | -0.11 | 0.06 | 8.62E-02 | 1.96E-01 | 123 | -0.09 | 0.04 | 2.94E-02 | 0.00 | 7.81E-01 | - | - | - |
| Emotional problems that made you do less than you wanted during the last four weeks | General health | 14434 | -0.70 | 0.26 | 6.54E-03 | **3.03E-02** | 59 | 14233 | -0.08 | 0.26 | 7.58E-01 | 8.57E-01 | 245 | -0.40 | 0.18 | 3.06E-02 | 0.65 | 9.04E-02 | - | - | - |
| Zinc intake (mg/day) | Nutrients | 29486 | -0.06 | 0.06 | 3.26E-01 | 5.10E-01 | 174 | 28746 | -0.13 | 0.06 | 3.82E-02 | 1.10E-01 | 97 | -0.09 | 0.04 | 3.18E-02 | 0.00 | 4.19E-01 | - | - | - |
| Palmitic acid intake (g/day) | Nutrients | 29486 | -0.07 | 0.06 | 2.10E-01 | 3.92E-01 | 146 | 28746 | -0.11 | 0.06 | 7.56E-02 | 1.77E-01 | 119 | -0.09 | 0.04 | 3.24E-02 | 0.00 | 6.92E-01 | - | - | - |
| Corn flakes | Food | 29486 | -0.07 | 0.06 | 2.37E-01 | 4.28E-01 | 151 | 28746 | -0.10 | 0.06 | 8.77E-02 | 1.97E-01 | 124 | -0.08 | 0.04 | 4.15E-02 | 0.00 | 6.89E-01 | - | - | - |
| Physical limitation to participate in moderately demanding activities: lifting or carrying grocery bags | General health | 14389 | 0.09 | 0.09 | 3.01E-01 | 4.90E-01 | 167 | 14229 | 0.16 | 0.09 | 7.00E-02 | 1.70E-01 | 115 | 0.13 | 0.06 | 4.43E-02 | 0.00 | 5.74E-01 | - | - | - |
| Syringaresinol intake (ug/day) | Nutrients | 29486 | 0.13 | 0.06 | 2.68E-02 | 9.63E-02 | 76 | 28746 | 0.04 | 0.06 | 5.49E-01 | 6.96E-01 | 220 | 0.08 | 0.04 | 4.50E-02 | 0.20 | 2.64E-01 | - | - | - |
| Polyunsaturated fat intake (g/day) | Nutrients | 29486 | -0.09 | 0.06 | 1.30E-01 | 2.72E-01 | 130 | 28746 | -0.07 | 0.06 | 2.03E-01 | 3.55E-01 | 159 | -0.08 | 0.04 | 4.83E-02 | 0.00 | 8.88E-01 | - | - | - |
| Berries (fresh or frozen) | Food | 29486 | -0.08 | 0.06 | 1.74E-01 | 3.36E-01 | 141 | 28746 | -0.08 | 0.06 | 1.59E-01 | 2.97E-01 | 149 | -0.08 | 0.04 | 5.03E-02 | 0.00 | 9.50E-01 | - | - | - |
| Travel to work: Walk to work vs passive travel to work | Physical activity | 21368 | 0.11 | 0.21 | 5.95E-01 | 7.39E-01 | 219 | 21069 | 0.46 | 0.21 | 2.79E-02 | 8.96E-02 | 86 | 0.28 | 0.15 | 5.51E-02 | 0.31 | 2.30E-01 | - | - | - |
| Cheese 10-17% | Food | 29486 | 0.01 | 0.05 | 8.15E-01 | 8.86E-01 | 251 | 28746 | 0.14 | 0.06 | 1.34E-02 | 5.21E-02 | 72 | 0.07 | 0.04 | 5.79E-02 | 0.61 | 1.09E-01 | - | - | - |
| Pizza | Food | 29486 | 0.07 | 0.06 | 2.64E-01 | 4.53E-01 | 159 | 28746 | 0.10 | 0.06 | 1.16E-01 | 2.38E-01 | 136 | 0.08 | 0.04 | 5.80E-02 | 0.00 | 7.29E-01 | - | - | - |
| Physical limitation that limited your ability to perform certain work tasks or other activities during the last four weeks | General health | 14389 | 0.23 | 0.25 | 3.67E-01 | 5.50E-01 | 182 | 14182 | 0.45 | 0.25 | 7.85E-02 | 1.82E-01 | 120 | 0.33 | 0.18 | 6.01E-02 | 0.00 | 5.35E-01 | - | - | - |
| For how much of the time during the last four weeks have you felt tired? | General health | 14415 | 0.03 | 0.09 | 6.97E-01 | 7.90E-01 | 241 | 14215 | 0.19 | 0.09 | 2.82E-02 | 8.96E-02 | 88 | 0.11 | 0.06 | 6.80E-02 | 0.40 | 1.98E-01 | - | - | - |
| Eicosapentaenoic acid (EPA) intake (g/day) | Nutrients | 29485 | -0.10 | 0.06 | 8.13E-02 | 2.02E-01 | 109 | 28746 | -0.05 | 0.06 | 4.20E-01 | 5.74E-01 | 204 | -0.08 | 0.04 | 6.98E-02 | 0.00 | 5.27E-01 | - | - | - |
| Butter on bread | Food | 29486 | 0.09 | 0.05 | 8.34E-02 | 2.05E-01 | 111 | 28746 | 0.05 | 0.06 | 4.17E-01 | 5.74E-01 | 202 | 0.07 | 0.04 | 7.12E-02 | 0.00 | 5.26E-01 | - | - | - |
| Do you feel important and appreciated outside your home? | Psychosocial | 24515 | 0.05 | 0.07 | 4.66E-01 | 6.39E-01 | 199 | 24103 | 0.12 | 0.07 | 7.51E-02 | 1.77E-01 | 118 | 0.08 | 0.05 | 7.72E-02 | 0.00 | 4.48E-01 | - | - | - |
| Hearing status | General health | 24525 | 0.10 | 0.07 | 1.61E-01 | 3.19E-01 | 138 | 24100 | 0.08 | 0.07 | 2.79E-01 | 4.41E-01 | 175 | 0.09 | 0.05 | 7.88E-02 | 0.00 | 8.31E-01 | - | - | - |
| Liquor, spirits | Alcohol | 29486 | 0.02 | 0.06 | 7.74E-01 | 8.52E-01 | 248 | 28746 | 0.13 | 0.06 | 3.19E-02 | 9.67E-02 | 92 | 0.08 | 0.04 | 8.08E-02 | 0.39 | 2.01E-01 | - | - | - |
| For how much of the time during the last four weeks have you felt calm and serene? | General health | 14395 | -0.09 | 0.09 | 3.05E-01 | 4.90E-01 | 170 | 14219 | -0.13 | 0.09 | 1.50E-01 | 2.86E-01 | 146 | -0.11 | 0.06 | 8.13E-02 | 0.00 | 7.63E-01 | - | - | - |
| Pancake, waffle, Swedish dumpling | Food | 29486 | -0.10 | 0.06 | 8.94E-02 | 2.16E-01 | 113 | 28746 | -0.04 | 0.06 | 4.64E-01 | 6.16E-01 | 210 | -0.07 | 0.04 | 8.40E-02 | 0.00 | 5.09E-01 | - | - | - |
| For how much of the time during the last four weeks have you felt worn out? | General health | 14373 | 0.10 | 0.09 | 2.63E-01 | 4.53E-01 | 158 | 14168 | 0.11 | 0.09 | 1.92E-01 | 3.41E-01 | 157 | 0.11 | 0.06 | 8.65E-02 | 0.00 | 8.91E-01 | - | - | - |
| Bacon | Food | 29486 | -0.05 | 0.06 | 3.77E-01 | 5.60E-01 | 184 | 28746 | -0.09 | 0.06 | 1.24E-01 | 2.48E-01 | 139 | -0.07 | 0.04 | 8.76E-02 | 0.00 | 6.29E-01 | - | - | - |
| For how much of the time during the last four weeks have you felt so depressed that nothing could cheer you up? | General health | 14412 | -0.16 | 0.09 | 5.45E-02 | 1.57E-01 | 95 | 14225 | -0.04 | 0.09 | 6.66E-01 | 7.91E-01 | 235 | -0.10 | 0.06 | 9.45E-02 | 0.09 | 2.94E-01 | - | - | - |
| Beta-sitosterol intake (mg/day) | Nutrients | 29486 | -0.07 | 0.06 | 2.16E-01 | 3.98E-01 | 148 | 28746 | -0.07 | 0.06 | 2.81E-01 | 4.41E-01 | 177 | -0.07 | 0.04 | 1.01E-01 | 0.00 | 9.27E-01 | - | - | - |
| Carbohydrates intake (g/day) | Nutrients | 29486 | 0.07 | 0.06 | 2.28E-01 | 4.15E-01 | 150 | 28746 | 0.07 | 0.06 | 2.83E-01 | 4.41E-01 | 179 | 0.07 | 0.04 | 1.07E-01 | 0.00 | 9.43E-01 | - | - | - |
| Grams of tobacco smoked per week | Tobacco use | 21113 | -0.11 | 0.07 | 1.11E-01 | 2.47E-01 | 122 | 20675 | -0.04 | 0.07 | 5.56E-01 | 6.99E-01 | 222 | -0.08 | 0.05 | 1.19E-01 | 0.00 | 4.95E-01 | - | - | - |
| Tea | Beverage | 29486 | -0.12 | 0.06 | 4.60E-02 | 1.38E-01 | 91 | 28746 | -0.01 | 0.06 | 8.65E-01 | 9.08E-01 | 265 | -0.07 | 0.04 | 1.22E-01 | 0.38 | 2.04E-01 | - | - | - |
| Alcohol intake (g/day) | Alcohol | 29486 | 0.01 | 0.06 | 8.30E-01 | 8.96E-01 | 253 | 28746 | 0.13 | 0.06 | 4.83E-02 | 1.26E-01 | 106 | 0.07 | 0.05 | 1.24E-01 | 0.36 | 2.10E-01 | - | - | - |
| Disaccharides intake (g/day) | Nutrients | 29486 | -0.01 | 0.06 | 9.22E-01 | 9.43E-01 | 267 | 28746 | -0.13 | 0.06 | 3.56E-02 | 1.06E-01 | 94 | -0.07 | 0.04 | 1.24E-01 | 0.51 | 1.51E-01 | - | - | - |
| Control over planning and execution of the workday | Psychosocial | 33138 | -0.08 | 0.05 | 1.32E-01 | 2.74E-01 | 132 | 32479 | -0.03 | 0.05 | 5.52E-01 | 6.97E-01 | 221 | -0.06 | 0.04 | 1.36E-01 | 0.00 | 5.28E-01 | - | - | - |
| If you exercise, change in exercise habits during the last year | Physical activity | 18780 | 0.14 | 0.07 | 4.45E-02 | 1.36E-01 | 89 | 18326 | 0.00 | 0.07 | 9.86E-01 | 9.93E-01 | 276 | 0.07 | 0.05 | 1.47E-01 | 0.48 | 1.64E-01 | - | - | - |
| Smoked fish/meat | Food | 29486 | -0.09 | 0.05 | 8.91E-02 | 2.16E-01 | 112 | 28746 | -0.02 | 0.06 | 7.64E-01 | 8.57E-01 | 248 | -0.06 | 0.04 | 1.52E-01 | 0.00 | 3.35E-01 | - | - | - |
| Linolenic acid intake (g/day) | Nutrients | 29486 | -0.06 | 0.06 | 2.94E-01 | 4.86E-01 | 165 | 28746 | -0.06 | 0.06 | 3.31E-01 | 4.97E-01 | 186 | -0.06 | 0.04 | 1.53E-01 | 0.00 | 9.72E-01 | - | - | - |
| Frequency of cycling during leisure time | Physical activity | 27210 | -0.05 | 0.06 | 4.31E-01 | 6.13E-01 | 192 | 26459 | -0.08 | 0.06 | 2.28E-01 | 3.81E-01 | 167 | -0.06 | 0.04 | 1.60E-01 | 0.00 | 7.53E-01 | - | - | - |
| Support from others | Social | 33614 | 0.10 | 0.05 | 4.97E-02 | 1.46E-01 | 93 | 32905 | 0.00 | 0.05 | 9.99E-01 | 9.99E-01 | 279 | 0.05 | 0.04 | 1.61E-01 | 0.47 | 1.70E-01 | - | - | - |
| Sometimes physically straining work | Physical activity | 32223 | 0.14 | 0.12 | 2.56E-01 | 4.46E-01 | 157 | 31545 | 0.11 | 0.12 | 3.98E-01 | 5.55E-01 | 200 | 0.12 | 0.09 | 1.61E-01 | 0.00 | 8.50E-01 | - | - | - |
| Sum of Lariciresinol, Matairesinol, Pinoresinol, Secoisolariciresinol intake (ug/day) | Nutrients | 29486 | -0.04 | 0.06 | 4.60E-01 | 6.37E-01 | 197 | 28746 | -0.08 | 0.06 | 2.17E-01 | 3.65E-01 | 166 | -0.06 | 0.04 | 1.65E-01 | 0.00 | 7.13E-01 | - | - | - |
| Apple, pear, peach, orange, mandarin and grapefruit | Food | 29486 | -0.10 | 0.06 | 1.10E-01 | 2.47E-01 | 121 | 28746 | -0.02 | 0.06 | 7.29E-01 | 8.43E-01 | 241 | -0.06 | 0.04 | 1.65E-01 | 0.00 | 3.86E-01 | - | - | - |
| Control over own work assignment | Psychosocial | 33105 | -0.05 | 0.05 | 3.20E-01 | 5.05E-01 | 173 | 32470 | -0.05 | 0.06 | 3.38E-01 | 5.01E-01 | 188 | -0.05 | 0.04 | 1.67E-01 | 0.00 | 9.89E-01 | - | - | - |
| Formic acid intaje (g/day) | Nutrients | 29486 | -0.04 | 0.06 | 4.85E-01 | 6.58E-01 | 201 | 28746 | -0.07 | 0.06 | 2.12E-01 | 3.60E-01 | 164 | -0.06 | 0.04 | 1.70E-01 | 0.00 | 6.85E-01 | - | - | - |
| Fat intake (g/day) | Nutrients | 29486 | -0.04 | 0.06 | 4.89E-01 | 6.58E-01 | 203 | 28746 | -0.08 | 0.06 | 2.14E-01 | 3.62E-01 | 165 | -0.06 | 0.04 | 1.73E-01 | 0.00 | 6.83E-01 | - | - | - |
| Potato salad | Food | 5584 | 0.09 | 0.13 | 5.03E-01 | 6.69E-01 | 205 | 5407 | 0.18 | 0.13 | 2.44E-01 | 4.02E-01 | 169 | 0.13 | 0.09 | 1.73E-01 | 0.00 | 6.40E-01 | - | - | - |
| Beta-sitostanol intake (mg/day) | Nutrients | 29486 | 0.09 | 0.06 | 1.19E-01 | 2.59E-01 | 124 | 28746 | 0.02 | 0.06 | 7.43E-01 | 8.53E-01 | 243 | 0.05 | 0.04 | 1.78E-01 | 0.00 | 3.94E-01 | - | - | - |
| Vitamin B6 intake (mg/day) | Nutrients | 29486 | 0.03 | 0.06 | 6.57E-01 | 7.70E-01 | 230 | 28746 | 0.09 | 0.06 | 1.55E-01 | 2.94E-01 | 147 | 0.06 | 0.04 | 1.90E-01 | 0.00 | 4.80E-01 | - | - | - |
| Vitamin E intake (mg/day) | Nutrients | 29486 | -0.06 | 0.06 | 3.05E-01 | 4.90E-01 | 169 | 28746 | -0.05 | 0.06 | 4.11E-01 | 5.71E-01 | 201 | -0.05 | 0.04 | 1.91E-01 | 0.00 | 8.98E-01 | - | - | - |
| Arachidonic acid (ARA) intake (g/day) | Nutrients | 29485 | -0.09 | 0.06 | 1.20E-01 | 2.59E-01 | 126 | 28745 | -0.02 | 0.06 | 7.83E-01 | 8.64E-01 | 253 | -0.05 | 0.04 | 1.91E-01 | 0.00 | 3.76E-01 | - | - | - |
| Feel uneasy or guilty because of your way of drinking | Alcohol | 19309 | 0.24 | 0.23 | 3.04E-01 | 4.90E-01 | 168 | 18914 | 0.19 | 0.23 | 4.22E-01 | 5.74E-01 | 205 | 0.21 | 0.16 | 1.95E-01 | 0.00 | 8.75E-01 | - | - | - |
| Years using snuff | Tobacco use | 31487 | 0.07 | 0.07 | 3.39E-01 | 5.25E-01 | 176 | 30839 | 0.06 | 0.07 | 3.81E-01 | 5.42E-01 | 196 | 0.07 | 0.05 | 1.95E-01 | 0.00 | 9.69E-01 | - | - | - |
| Lariciresinol intake (ug/day) | Nutrients | 29486 | -0.05 | 0.06 | 4.47E-01 | 6.25E-01 | 195 | 28746 | -0.07 | 0.06 | 2.93E-01 | 4.51E-01 | 181 | -0.06 | 0.04 | 2.01E-01 | 0.00 | 8.24E-01 | - | - | - |
| Cohabitation: Live alone vs Only one adult (spouse, partner) | Social | 15166 | -0.11 | 0.22 | 6.00E-01 | 7.41E-01 | 221 | 15022 | -0.28 | 0.22 | 2.06E-01 | 3.59E-01 | 160 | -0.20 | 0.15 | 2.06E-01 | 0.00 | 6.01E-01 | - | - | - |
| Self-employed | Psychosocial | 28658 | -0.36 | 0.21 | 8.02E-02 | 2.02E-01 | 107 | 27905 | 0.00 | 0.21 | 9.98E-01 | 9.99E-01 | 278 | -0.18 | 0.15 | 2.14E-01 | 0.34 | 2.18E-01 | - | - | - |
| Lean fish (e.g. perch, bass, cod) | Food | 29486 | -0.12 | 0.06 | 2.84E-02 | 1.01E-01 | 77 | 28746 | 0.03 | 0.06 | 6.31E-01 | 7.62E-01 | 231 | -0.05 | 0.04 | 2.19E-01 | 0.72 | 6.04E-02 | - | - | - |

| | | | | | | | | | | | | | | | | | | | | | |
|---|---|---|---|---|---|---|---|---|---|---|---|---|---|---|---|---|---|---|---|---|---|
| Stigmasterol intake (mg/day) | Nutrients | 29486 | -0.01 | 0.06 | 8.15E-01 | 8.86E-01 | 250 | 28746 | -0.09 | 0.06 | 1.30E-01 | 2.54E-01 | 142 | -0.05 | 0.04 | 2.21E-01 | 0.00 | 3.56E-01 | - | - | - |
| Sum of all lignans intake (ug/day) | Nutrients | 29486 | 0.09 | 0.06 | 1.29E-01 | 2.72E-01 | 129 | 28746 | 0.01 | 0.06 | 8.48E-01 | 9.00E-01 | 263 | 0.05 | 0.04 | 2.22E-01 | 0.00 | 3.56E-01 | - | - | - |
| Brown beans, pea soup | Food | 29486 | -0.02 | 0.06 | 6.52E-01 | 7.70E-01 | 228 | 28746 | -0.07 | 0.06 | 2.09E-01 | 3.60E-01 | 161 | -0.05 | 0.04 | 2.30E-01 | 0.00 | 5.59E-01 | - | - | - |
| Cookies, pastry | Food | 29486 | -0.02 | 0.06 | 7.53E-01 | 8.32E-01 | 247 | 28746 | -0.08 | 0.06 | 1.65E-01 | 3.06E-01 | 150 | -0.05 | 0.04 | 2.32E-01 | 0.00 | 4.38E-01 | - | - | - |
| Saturated fat intake (g/day) | Nutrients | 29486 | -0.02 | 0.06 | 7.06E-01 | 7.94E-01 | 243 | 28746 | -0.08 | 0.06 | 1.88E-01 | 3.36E-01 | 156 | -0.05 | 0.04 | 2.34E-01 | 0.00 | 4.97E-01 | - | - | - |
| Vitamin D intake (ug/day) | Nutrients | 29486 | -0.04 | 0.06 | 5.27E-01 | 6.89E-01 | 209 | 28746 | -0.06 | 0.06 | 3.05E-01 | 4.63E-01 | 184 | -0.05 | 0.04 | 2.42E-01 | 0.00 | 7.71E-01 | - | - | - |
| Matairesinol intake (ug/day) | Nutrients | 29486 | 0.08 | 0.06 | 1.50E-01 | 3.00E-01 | 136 | 28746 | 0.01 | 0.06 | 8.55E-01 | 9.03E-01 | 264 | 0.05 | 0.04 | 2.47E-01 | 0.00 | 3.83E-01 | - | - | - |
| Potassium intake (mg/day) | Nutrients | 29486 | -0.05 | 0.06 | 4.13E-01 | 5.99E-01 | 187 | 28746 | -0.05 | 0.06 | 4.20E-01 | 5.74E-01 | 203 | -0.05 | 0.04 | 2.51E-01 | 0.00 | 9.94E-01 | - | - | - |
| Confidence status | Psychosocial | 24483 | 0.00 | 0.07 | 9.90E-01 | 9.93E-01 | 271 | 24048 | -0.11 | 0.07 | 1.15E-01 | 2.38E-01 | 135 | -0.05 | 0.05 | 2.67E-01 | 0.20 | 2.64E-01 | - | - | - |
| Light beer | Alcohol | 29486 | 0.02 | 0.06 | 7.36E-01 | 8.24E-01 | 244 | 28746 | -0.12 | 0.06 | 5.11E-02 | 1.32E-01 | 108 | -0.05 | 0.04 | 2.68E-01 | 0.63 | 1.01E-01 | - | - | - |
| Breakfast habits: Only coffee/tea for breakfast vs not breakfast at all | Food | 1909 | -0.11 | 0.55 | 8.46E-01 | 8.99E-01 | 257 | 1986 | -0.75 | 0.55 | 1.74E-01 | 3.19E-01 | 152 | -0.43 | 0.39 | 2.70E-01 | 0.00 | 4.12E-01 | - | - | - |
| Banana | Food | 29486 | 0.06 | 0.06 | 3.17E-01 | 5.03E-01 | 172 | 28746 | 0.03 | 0.06 | 5.83E-01 | 7.16E-01 | 227 | 0.05 | 0.04 | 2.71E-01 | 0.00 | 7.63E-01 | - | - | - |
| Number of friends that can come to your home at any time and feel at home | Social | 33124 | 0.08 | 0.06 | 1.42E-01 | 2.91E-01 | 133 | 32422 | 0.00 | 0.06 | 9.62E-01 | 9.76E-01 | 275 | 0.04 | 0.04 | 2.78E-01 | 0.00 | 3.22E-01 | - | - | - |
| Job demands to work very fast | Psychosocial | 33113 | -0.05 | 0.05 | 3.79E-01 | 5.60E-01 | 185 | 32492 | -0.03 | 0.06 | 5.66E-01 | 7.01E-01 | 225 | -0.04 | 0.04 | 3.03E-01 | 0.00 | 8.35E-01 | - | - | - |
| For how much of the time during the last four weeks have you felt very nervous? | General health | 14396 | 0.11 | 0.09 | 1.84E-01 | 3.49E-01 | 144 | 14212 | 0.01 | 0.09 | 9.25E-01 | 9.53E-01 | 271 | 0.06 | 0.06 | 3.12E-01 | 0.00 | 3.84E-01 | - | - | - |
| Butter for cooking | Food | 29486 | -0.11 | 0.06 | 6.03E-02 | 1.65E-01 | 99 | 28746 | 0.03 | 0.06 | 6.13E-01 | 7.47E-01 | 229 | -0.04 | 0.04 | 3.25E-01 | 0.64 | 9.34E-02 | - | - | - |
| Vitamin A intake (mg/day) | Nutrients | 29486 | 0.05 | 0.06 | 4.36E-01 | 6.17E-01 | 193 | 28746 | 0.04 | 0.06 | 5.60E-01 | 7.01E-01 | 223 | 0.04 | 0.04 | 3.35E-01 | 0.00 | 8.96E-01 | - | - | - |
| Breakfast habits: Gruel w/o sandwich for breakfast vs not breakfast at all | Food | 1445 | 0.30 | 0.68 | 6.59E-01 | 7.70E-01 | 231 | 1505 | 0.59 | 0.65 | 3.64E-01 | 5.29E-01 | 192 | 0.45 | 0.47 | 3.35E-01 | 0.00 | 7.58E-01 | - | - | - |
| Sum of phytosterols intake (mg/day) | Nutrients | 29486 | -0.03 | 0.06 | 5.73E-01 | 7.29E-01 | 214 | 28746 | -0.05 | 0.06 | 4.40E-01 | 5.96E-01 | 206 | -0.04 | 0.04 | 3.46E-01 | 0.00 | 8.73E-01 | - | - | - |
| Sausage as main dish | Food | 29486 | -0.08 | 0.06 | 1.69E-01 | 3.32E-01 | 139 | 28746 | 0.00 | 0.06 | 9.57E-01 | 9.75E-01 | 273 | -0.04 | 0.04 | 3.46E-01 | 0.01 | 3.15E-01 | - | - | - |
| Low fat milk (0,5%) | Beverage | 29486 | 0.01 | 0.06 | 8.60E-01 | 9.01E-01 | 259 | 28746 | 0.07 | 0.06 | 2.45E-01 | 4.02E-01 | 170 | 0.04 | 0.04 | 3.49E-01 | 0.00 | 4.77E-01 | - | - | - |
| Equol intake (ug/day) | Nutrients | 29486 | 0.03 | 0.06 | 5.66E-01 | 7.29E-01 | 211 | 28746 | 0.04 | 0.06 | 4.57E-01 | 6.12E-01 | 208 | 0.04 | 0.04 | 3.53E-01 | 0.00 | 8.94E-01 | - | - | - |
| Light and physically active work | Physical activity | 32223 | 0.11 | 0.12 | 3.63E-01 | 5.48E-01 | 181 | 31545 | 0.05 | 0.13 | 7.03E-01 | 8.17E-01 | 240 | 0.08 | 0.09 | 3.58E-01 | 0.00 | 7.20E-01 | - | - | - |
| Medium beer | Alcohol | 29486 | -0.06 | 0.06 | 3.40E-01 | 5.25E-01 | 177 | 28746 | -0.02 | 0.06 | 7.72E-01 | 8.57E-01 | 250 | -0.04 | 0.04 | 3.77E-01 | 0.00 | 6.43E-01 | - | - | - |
| Vitamin B12 intake (ug/day) | Nutrients | 29486 | -0.04 | 0.06 | 4.57E-01 | 6.36E-01 | 196 | 28746 | -0.03 | 0.06 | 6.26E-01 | 7.60E-01 | 230 | -0.03 | 0.04 | 3.83E-01 | 0.00 | 8.65E-01 | - | - | - |
| Participation in study circles | Social | 10696 | 0.12 | 0.28 | 6.64E-01 | 7.72E-01 | 235 | 10356 | -0.48 | 0.28 | 9.01E-02 | 2.01E-01 | 125 | -0.17 | 0.20 | 3.85E-01 | 0.57 | 1.28E-01 | - | - | - |
| Iron intake (mg/day) | Nutrients | 29486 | 0.01 | 0.06 | 8.90E-01 | 9.14E-01 | 266 | 28746 | 0.07 | 0.06 | 2.81E-01 | 4.41E-01 | 176 | 0.04 | 0.04 | 3.94E-01 | 0.00 | 4.99E-01 | - | - | - |
| Vitamin C intake (mg/day) | Nutrients | 29486 | -0.02 | 0.06 | 6.94E-01 | 7.89E-01 | 240 | 28746 | 0.09 | 0.06 | 1.13E-01 | 2.35E-01 | 134 | 0.03 | 0.04 | 4.09E-01 | 0.50 | 1.59E-01 | - | - | - |
| Frequent social contacts with colleagues during work | Psychosocial | 31671 | 0.05 | 0.05 | 3.58E-01 | 5.46E-01 | 179 | 31010 | 0.01 | 0.06 | 8.18E-01 | 8.78E-01 | 260 | 0.03 | 0.04 | 4.14E-01 | 0.00 | 6.31E-01 | - | - | - |
| Minced meat dishes | Food | 29486 | 0.00 | 0.06 | 9.93E-01 | 9.93E-01 | 273 | 28746 | -0.07 | 0.06 | 2.48E-01 | 4.04E-01 | 171 | -0.03 | 0.04 | 4.17E-01 | 0.00 | 4.11E-01 | - | - | - |
| Fried potatoes, pommes frites | Food | 29486 | 0.01 | 0.06 | 8.46E-01 | 8.99E-01 | 256 | 28746 | 0.05 | 0.06 | 3.45E-01 | 5.06E-01 | 190 | 0.03 | 0.04 | 4.21E-01 | 0.00 | 5.95E-01 | - | - | - |
| Whole grain soft bread | Food | 29486 | 0.01 | 0.06 | 8.51E-01 | 9.00E-01 | 258 | 28746 | -0.08 | 0.06 | 1.79E-01 | 3.24E-01 | 154 | -0.03 | 0.04 | 4.23E-01 | 0.17 | 2.73E-01 | - | - | - |
| Enterolactone intake (ug/day) | Nutrients | 29486 | -0.02 | 0.06 | 6.59E-01 | 7.70E-01 | 232 | 28746 | -0.04 | 0.06 | 4.87E-01 | 6.38E-01 | 213 | -0.03 | 0.04 | 4.23E-01 | 0.00 | 8.50E-01 | - | - | - |
| Breakfast habits: Coffee/tea and wheat buns or rusk for breakfast vs not breakfast at all | Food | 1595 | -0.32 | 0.56 | 5.71E-01 | 7.29E-01 | 213 | 1584 | -0.31 | 0.56 | 5.80E-01 | 7.16E-01 | 226 | -0.31 | 0.40 | 4.28E-01 | 0.00 | 9.89E-01 | - | - | - |
| Enterodiol intake (ug/day) | Nutrients | 29486 | 0.03 | 0.06 | 6.18E-01 | 7.52E-01 | 224 | 28746 | 0.04 | 0.06 | 5.36E-01 | 6.86E-01 | 218 | 0.03 | 0.04 | 4.30E-01 | 0.00 | 9.24E-01 | - | - | - |
| Pinoresinol intake (ug/day) | Nutrients | 29486 | -0.01 | 0.06 | 8.07E-01 | 8.85E-01 | 249 | 28746 | -0.05 | 0.06 | 3.81E-01 | 5.42E-01 | 195 | -0.03 | 0.04 | 4.32E-01 | 0.00 | 6.47E-01 | - | - | - |
| Monosaccharides intake (g/day) | Nutrients | 29486 | -0.05 | 0.06 | 4.18E-01 | 5.99E-01 | 190 | 28746 | -0.01 | 0.06 | 8.36E-01 | 8.91E-01 | 262 | -0.03 | 0.04 | 4.69E-01 | 0.00 | 6.77E-01 | - | - | - |
| Distance to work in kilometers (one way) | Physical activity | 28796 | 0.03 | 0.06 | 5.92E-01 | 7.39E-01 | 218 | 28101 | 0.03 | 0.06 | 6.49E-01 | 7.77E-01 | 233 | 0.03 | 0.04 | 4.83E-01 | 0.00 | 9.60E-01 | - | - | - |
| Wine | Alcohol | 29486 | -0.08 | 0.06 | 1.93E-01 | 3.63E-01 | 145 | 28746 | 0.02 | 0.06 | 7.34E-01 | 8.47E-01 | 242 | -0.03 | 0.04 | 4.87E-01 | 0.25 | 2.49E-01 | - | - | - |
| Light but partly physically active work | Physical activity | 32223 | -0.15 | 0.13 | 2.46E-01 | 4.36E-01 | 152 | 31545 | 0.03 | 0.14 | 8.29E-01 | 8.86E-01 | 261 | -0.07 | 0.10 | 4.94E-01 | 0.00 | 3.37E-01 | - | - | - |
| Permanent employment | Psychosocial | 29019 | 0.21 | 0.13 | 1.08E-01 | 2.47E-01 | 118 | 28287 | -0.09 | 0.13 | 5.04E-01 | 6.58E-01 | 214 | 0.06 | 0.09 | 4.97E-01 | 0.61 | 1.09E-01 | - | - | - |
| People to ask for help apart from the ones at home | Social | 33488 | 0.11 | 0.18 | 5.18E-01 | 6.80E-01 | 208 | 32780 | 0.05 | 0.18 | 7.67E-01 | 8.57E-01 | 249 | 0.08 | 0.13 | 5.04E-01 | 0.00 | 8.08E-01 | - | - | - |
| Sucrose intake (g/day) | Nutrients | 29486 | 0.05 | 0.06 | 4.19E-01 | 5.99E-01 | 191 | 28746 | 0.01 | 0.06 | 9.02E-01 | 9.42E-01 | 267 | 0.03 | 0.04 | 5.06E-01 | 0.00 | 6.35E-01 | - | - | - |
| Bregott on bread | Food | 29486 | 0.01 | 0.06 | 8.89E-01 | 9.14E-01 | 265 | 28746 | -0.06 | 0.06 | 2.90E-01 | 4.49E-01 | 180 | -0.03 | 0.04 | 5.23E-01 | 0.00 | 3.92E-01 | - | - | - |
| Campestanol intake (mg/day) | Nutrients | 29486 | 0.05 | 0.06 | 3.70E-01 | 5.52E-01 | 183 | 28746 | 0.00 | 0.06 | 9.86E-01 | 9.93E-01 | 277 | 0.03 | 0.04 | 5.29E-01 | 0.00 | 5.23E-01 | - | - | - |
| Cohabitation: Live alone vs Other/others | Social | 3824 | -0.44 | 0.54 | 4.15E-01 | 5.99E-01 | 189 | 3819 | -0.04 | 0.55 | 9.46E-01 | 9.70E-01 | 272 | -0.24 | 0.38 | 5.29E-01 | 0.00 | 6.02E-01 | - | - | - |
| Salty fish | Food | 29486 | 0.00 | 0.05 | 9.87E-01 | 9.93E-01 | 270 | 28746 | 0.05 | 0.06 | 3.80E-01 | 5.42E-01 | 194 | 0.02 | 0.04 | 5.39E-01 | 0.00 | 5.31E-01 | - | - | - |
| Steak, chop, e.g. | Food | 29486 | 0.06 | 0.06 | 2.81E-01 | 4.72E-01 | 162 | 28746 | -0.02 | 0.06 | 7.90E-01 | 8.64E-01 | 255 | 0.02 | 0.04 | 5.55E-01 | 0.00 | 3.47E-01 | - | - | - |
| Participation in other association | Social | 10696 | -0.06 | 0.20 | 7.50E-01 | 8.32E-01 | 246 | 10356 | 0.23 | 0.21 | 2.57E-01 | 4.14E-01 | 173 | 0.08 | 0.14 | 5.75E-01 | 0.07 | 3.00E-01 | - | - | - |
| Trans fat intake (g/day) | Nutrients | 29486 | -0.01 | 0.08 | 8.84E-01 | 9.14E-01 | 263 | 28746 | -0.05 | 0.08 | 5.15E-01 | 6.68E-01 | 215 | -0.03 | 0.06 | 5.76E-01 | 0.00 | 7.15E-01 | - | - | - |
| Sedentary or standing work | Physical activity | 32223 | -0.06 | 0.13 | 6.44E-01 | 7.70E-01 | 227 | 31545 | -0.04 | 0.13 | 7.60E-01 | 8.57E-01 | 246 | -0.05 | 0.09 | 5.87E-01 | 0.00 | 9.15E-01 | - | - | - |
| Appetite status | Psychosocial | 24506 | 0.01 | 0.06 | 8.40E-01 | 8.99E-01 | 255 | 24082 | -0.06 | 0.07 | 3.73E-01 | 5.40E-01 | 193 | -0.02 | 0.05 | 6.31E-01 | 0.00 | 4.38E-01 | - | - | - |
| Meat stew | Food | 29486 | -0.06 | 0.06 | 2.83E-01 | 4.72E-01 | 163 | 28746 | 0.02 | 0.06 | 6.74E-01 | 7.93E-01 | 237 | -0.02 | 0.04 | 6.37E-01 | 0.10 | 2.93E-01 | - | - | - |
| Chips, popcorn, salted nuts | Food | 29486 | -0.02 | 0.06 | 6.72E-01 | 7.76E-01 | 236 | 28746 | -0.01 | 0.06 | 8.09E-01 | 8.72E-01 | 258 | -0.02 | 0.04 | 6.37E-01 | 0.00 | 9.03E-01 | - | - | - |
| Number of cigars smoked per day | Tobacco use | 21099 | -0.03 | 0.07 | 6.84E-01 | 7.84E-01 | 238 | 20652 | -0.02 | 0.07 | 7.99E-01 | 8.68E-01 | 257 | -0.02 | 0.05 | 6.39E-01 | 0.00 | 9.20E-01 | - | - | - |
| For how much of the time during the last four weeks has your physical health or your emotional problems limited your ability to interact with others? | General health | 14330 | -0.07 | 0.09 | 4.14E-01 | 5.99E-01 | 188 | 14133 | 0.01 | 0.09 | 8.66E-01 | 9.08E-01 | 266 | -0.03 | 0.06 | 6.44E-01 | 0.00 | 4.87E-01 | - | - | - |
| Sweets | Food | 29486 | 0.07 | 0.06 | 2.48E-01 | 4.36E-01 | 154 | 28746 | -0.03 | 0.06 | 5.97E-01 | 7.31E-01 | 228 | 0.02 | 0.04 | 6.49E-01 | 0.29 | 2.35E-01 | - | - | - |
| Hamburger | Food | 29486 | 0.00 | 0.06 | 9.91E-01 | 9.93E-01 | 272 | 28746 | 0.04 | 0.06 | 5.38E-01 | 6.86E-01 | 219 | 0.02 | 0.04 | 6.63E-01 | 0.00 | 6.63E-01 | - | - | - |
| Campesterol intake (mg/day) | Nutrients | 29486 | 0.03 | 0.06 | 5.89E-01 | 7.39E-01 | 217 | 28746 | 0.00 | 0.06 | 9.58E-01 | 9.75E-01 | 274 | 0.02 | 0.04 | 6.71E-01 | 0.00 | 7.35E-01 | - | - | - |
| Close relationship with anyone | Social | 33205 | 0.02 | 0.05 | 6.60E-01 | 7.70E-01 | 233 | 32474 | -0.06 | 0.05 | 3.05E-01 | 4.63E-01 | 183 | -0.02 | 0.04 | 6.84E-01 | 0.07 | 2.99E-01 | - | - | - |
| White (soft) bread, thin crisp bread | Food | 29486 | -0.03 | 0.06 | 5.75E-01 | 7.29E-01 | 215 | 28746 | 0.07 | 0.06 | 2.53E-01 | 4.11E-01 | 172 | 0.02 | 0.04 | 6.92E-01 | 0.32 | 2.27E-01 | - | - | - |
| Fibre intake (g/day) | Nutrients | 29486 | 0.04 | 0.06 | 5.30E-01 | 6.89E-01 | 210 | 28746 | -0.01 | 0.07 | 9.22E-01 | 9.53E-01 | 270 | 0.02 | 0.05 | 7.02E-01 | 0.00 | 6.11E-01 | - | - | - |
| Number of social contacts with the same interests as you | Social | 33535 | 0.01 | 0.06 | 8.38E-01 | 8.99E-01 | 254 | 32819 | -0.04 | 0.06 | 4.59E-01 | 6.12E-01 | 209 | -0.02 | 0.04 | 7.10E-01 | 0.00 | 5.01E-01 | - | - | - |
| For how much of the time during the last four weeks have you felt happy? | General health | 14401 | 0.12 | 0.09 | 1.49E-01 | 3.00E-01 | 135 | 14205 | -0.08 | 0.09 | 3.47E-01 | 5.06E-01 | 191 | 0.02 | 0.06 | 7.18E-01 | 0.65 | 9.17E-02 | - | - | - |
| Sour milk, yoghurt (low fat) | Food | 29486 | 0.04 | 0.06 | 4.39E-01 | 6.18E-01 | 194 | 28746 | -0.02 | 0.06 | 7.73E-01 | 8.57E-01 | 251 | 0.01 | 0.04 | 7.24E-01 | 0.00 | 4.56E-01 | - | - | - |
| Travel to work: Irregular travel mode to work vs passive travel to work | Physical activity | 21454 | -0.18 | 0.20 | 3.61E-01 | 5.48E-01 | 180 | 21096 | 0.09 | 0.20 | 6.58E-01 | 7.85E-01 | 234 | -0.05 | 0.14 | 7.33E-01 | 0.00 | 3.39E-01 | - | - | - |
| Satisfaction with work situation | Psychosocial | 24209 | 0.04 | 0.06 | 4.89E-01 | 6.58E-01 | 202 | 23776 | -0.07 | 0.07 | 2.59E-01 | 4.15E-01 | 174 | -0.01 | 0.05 | 7.66E-01 | 0.40 | 1.97E-01 | - | - | - |
| Margarine on bread | Food | 29486 | 0.00 | 0.05 | 9.48E-01 | 9.62E-01 | 269 | 28746 | 0.02 | 0.05 | 7.74E-01 | 8.57E-01 | 252 | 0.01 | 0.04 | 8.04E-01 | 0.00 | 8.73E-01 | - | - | - |
| Patience status | Psychosocial | 24490 | 0.03 | 0.07 | 6.87E-01 | 7.85E-01 | 239 | 24071 | -0.05 | 0.07 | 4.49E-01 | 6.05E-01 | 207 | -0.01 | 0.05 | 8.07E-01 | 0.00 | 4.11E-01 | - | - | - |
| Possibility to speak with colleagues during breaks | Psychosocial | 31887 | -0.03 | 0.06 | 6.13E-01 | 7.50E-01 | 222 | 31243 | 0.01 | 0.06 | 7.96E-01 | 8.67E-01 | 256 | -0.01 | 0.04 | 8.56E-01 | 0.00 | 5.90E-01 | - | - | - |
| Plant based protein intake (g/day) | Nutrients | 29486 | 0.01 | 0.06 | 8.87E-01 | 9.14E-01 | 264 | 28746 | 0.01 | 0.06 | 9.11E-01 | 9.45E-01 | 269 | 0.01 | 0.04 | 8.58E-01 | 0.00 | 9.85E-01 | - | - | - |
| Frequency of shoverling snow during leisure time | Physical activity | 20765 | 0.03 | 0.07 | 6.57E-01 | 7.70E-01 | 229 | 20224 | -0.05 | 0.07 | 4.84E-01 | 6.38E-01 | 211 | -0.01 | 0.05 | 8.64E-01 | 0.00 | 4.17E-01 | - | - | - |
| Possibility to leave your work for a while to speak with a colleague | Psychosocial | 31688 | 0.06 | 0.06 | 3.12E-01 | 4.97E-01 | 171 | 31042 | -0.07 | 0.06 | 2.11E-01 | 3.60E-01 | 163 | -0.01 | 0.04 | 8.81E-01 | 0.61 | 1.09E-01 | - | - | - |
| Long-term sickness | General health | 31217 | 0.07 | 0.15 | 6.20E-01 | 7.52E-01 | 225 | 30486 | -0.05 | 0.15 | 7.63E-01 | 8.57E-01 | 247 | 0.02 | 0.11 | 8.86E-01 | 0.00 | 5.74E-01 | - | - | - |
| Snuff status: Former snuff users vs non-snuff users | Tobacco use | 26796 | 0.11 | 0.21 | 5.95E-01 | 7.39E-01 | 220 | 26144 | -0.09 | 0.21 | 6.84E-01 | 7.98E-01 | 239 | 0.01 | 0.15 | 9.24E-01 | 0.00 | 5.08E-01 | - | - | - |
| Frequency of gardening during leisure time | Physical activity | 20704 | 0.05 | 0.07 | 4.62E-01 | 6.37E-01 | 198 | 20035 | -0.06 | 0.07 | 3.91E-01 | 5.49E-01 | 199 | 0.00 | 0.05 | 9.40E-01 | 0.21 | 2.60E-01 | - | - | - |
| Extent to what your physical and emotional health disrupted your usual social life during the last four weeks | General health | 14416 | -0.11 | 0.09 | 2.13E-01 | 3.96E-01 | 147 | 14232 | 0.12 | 0.09 | 1.75E-01 | 3.19E-01 | 153 | 0.00 | 0.06 | 9.44E-01 | 0.71 | 6.54E-02 | - | - | - |
| Work shifts/weekends | Psychosocial | 32019 | -0.04 | 0.13 | 7.41E-01 | 8.26E-01 | 245 | 31436 | 0.04 | 0.13 | 7.54E-01 | 8.57E-01 | 244 | 0.00 | 0.09 | 9.89E-01 | 0.00 | 6.49E-01 | - | - | - |
| Milk, sour milk (3%) | Beverage | 29486 | 0.03 | 0.06 | 5.67E-01 | 7.29E-01 | 212 | 28746 | -0.03 | 0.06 | 5.64E-01 | 7.01E-01 | 224 | 0.00 | 0.04 | 9.93E-01 | 0.00 | 4.16E-01 | - | - | - |
| Marital status: Single vs Widow/widower | Social | 3563 | 0.42 | 0.62 | 5.05E-01 | 6.69E-01 | 206 | 3523 | -0.46 | 0.65 | 4.85E-01 | 6.38E-01 | 212 | 0.00 | 0.45 | 9.99E-01 | 0.00 | 3.34E-01 | - | - | - |
| Amount of alcohol drunk in a day | Alcohol | 10924 | 0.42 | 0.10 | - | - | 277 | 10736 | 0.61 | 0.11 | 6.84E-09 | **1.59E-07** | 12 | 0.51 | 0.07 | - | 0.43 | 1.85E-01 | - | - | - |
| Snore during sleep | Sleep | 10526 | 0.53 | 0.10 | - | - | 286 | 10435 | 0.43 | 0.11 | 4.62E-05 | **4.67E-04** | 27 | 0.48 | 0.07 | - | 0.00 | 5.11E-01 | - | - | - |
| Breath-holds during sleep | Sleep | 7905 | 0.47 | 0.12 | - | - | 287 | 7757 | 0.37 | 0.12 | 2.51E-03 | **1.34E-02** | 52 | 0.42 | 0.09 | - | 0.00 | 5.69E-01 | - | - | - |
| Risk of sleeping while sitting still after having lunch | Sleep | 11254 | -0.25 | 0.10 | - | - | 285 | 11136 | -0.22 | 0.10 | 2.61E-02 | 8.87E-02 | 82 | -0.24 | 0.07 | - | 0.00 | 7.93E-01 | - | - | - |
| Risk of sleeping while sitting and reading | Sleep | 11172 | -0.15 | 0.10 | - | - | 280 | 11043 | -0.19 | 0.10 | 4.83E-02 | 1.26E-01 | 107 | -0.17 | 0.07 | - | 0.00 | 7.31E-01 | - | - | - |
| Soft whey cheese | Food | 6985 | 0.12 | 0.11 | 2.95E-01 | 4.86E-01 | 166 | 6760 | -0.25 | 0.12 | 9.66E-02 | 2.12E-01 | 127 | -0.06 | 0.08 | - | 0.81 | 2.07E-02 | - | - | - |
| Mixed frozen vegetables | Food | 6985 | 0.02 | 0.12 | 8.61E-01 | 9.01E-01 | 260 | 6760 | 0.18 | 0.12 | 2.09E-01 | 3.60E-01 | 162 | 0.10 | 0.08 | - | 0.00 | 3.43E-01 | - | - | - |
| Shellfish (e.g. shrimps, scallops) | Food | 6985 | -0.02 | 0.12 | 8.65E-01 | 9.01E-01 | 262 | 6760 | -0.13 | 0.12 | 3.43E-01 | 5.06E-01 | 189 | -0.07 | 0.08 | - | 0.00 | 5.15E-01 | - | - | - |
| Blota (broth + bread) | Food | 6985 | 0.28 | 0.11 | 2.98E-02 | 1.04E-01 | 78 | 6760 | 0.08 | 0.12 | 5.19E-01 | 6.68E-01 | 217 | 0.18 | 0.08 | - | 0.29 | 2.35E-01 | - | - | - |
| Mashed potato | Food | 6985 | 0.02 | 0.12 | 8.61E-01 | 9.01E-01 | 261 | 6760 | 0.06 | 0.12 | 6.39E-01 | 7.69E-01 | 232 | 0.04 | 0.08 | - | 0.00 | 8.07E-01 | - | - | - |
| Soft cheese | Food | 6985 | 0.14 | 0.11 | 2.49E-01 | 4.36E-01 | 155 | 6760 | -0.05 | 0.12 | 6.80E-01 | 7.97E-01 | 238 | 0.04 | 0.08 | - | 0.25 | 2.48E-01 | - | - | - |
| Liver, kidney | Food | 6985 | 0.03 | 0.11 | 8.24E-01 | 8.93E-01 | 252 | 6760 | -0.03 | 0.12 | 7.87E-01 | 8.64E-01 | 254 | 0.00 | 0.08 | - | 0.00 | 7.16E-01 | - | - | - |
| Risk of sleeping while lying down resting in the afternoon | Sleep | 11246 | -0.17 | 0.10 | - | - | 284 | 11130 | 0.02 | 0.10 | 8.10E-01 | 8.72E-01 | 259 | -0.08 | 0.07 | - | 0.50 | 1.58E-01 | - | - | - |
| Blood based food | Food | 6985 | -0.07 | 0.11 | 5.77E-01 | 7.29E-01 | 216 | 6760 | -0.01 | 0.12 | 9.10E-01 | 9.45E-01 | 268 | -0.04 | 0.08 | - | 0.00 | 7.54E-01 | - | - | - |
| Time spent in a week in moderately steneous activities | Physical activity | 11050 | -0.35 | 0.10 | - | - | 274 | 10870 | -0.24 | 0.10 | - | - | 280 | -0.30 | 0.07 | - | 0.00 | 4.21E-01 | - | - | - |

| | | | | | | | | | | | | | | | | | | | | | |
|---|---|---|---|---|---|---|---|---|---|---|---|---|---|---|---|---|---|---|---|---|---|
| Amount of exercise during the last 12 months | Physical activity | 11394 | -0.46 | 0.10 | - | - | 275 | 11230 | -0.42 | 0.10 | - | - | 281 | -0.44 | 0.07 | - | 0.00 | 7.64E-01 | - | - | - |
| Frequency of alcohol consumption | Alcohol | 11437 | -0.02 | 0.10 | - | - | 276 | 11304 | 0.19 | 0.10 | - | - | 282 | 0.08 | 0.07 | - | 0.56 | 1.32E-01 | - | - | - |
| Frequency of drinking six or more glasses at the same occasion | Alcohol | 11128 | 0.42 | 0.10 | - | - | 278 | 10952 | 0.67 | 0.11 | - | - | 283 | 0.54 | 0.07 | - | 0.64 | 9.42E-02 | - | - | - |
| Times during last year that you felt guilty because of your drinking | Alcohol | 11115 | 0.05 | 0.10 | - | - | 279 | 10924 | 0.27 | 0.10 | - | - | 284 | 0.16 | 0.07 | - | 0.60 | 1.16E-01 | - | - | - |
| Risk of sleeping while watching TV | Sleep | 11294 | 0.07 | 0.10 | - | - | 281 | 11161 | 0.16 | 0.10 | - | - | 285 | 0.11 | 0.07 | - | 0.00 | 4.87E-01 | - | - | - |
| Risk of sleeping while sitting inactive in a public place | Sleep | 11218 | -0.27 | 0.10 | - | - | 282 | 11089 | -0.19 | 0.10 | - | - | 286 | -0.23 | 0.07 | - | 0.00 | 5.83E-01 | - | - | - |
| Risk of sleeping as a passenger in a car for one hour without break | Sleep | 11179 | -0.41 | 0.10 | - | - | 283 | 11044 | -0.39 | 0.10 | - | - | 287 | -0.40 | 0.07 | - | 0.00 | 9.25E-01 | - | - | - |

**Supplementary Table 6**. Linear mixed model association results for total cholesterol

| | | Training set | | | | | | Testing set | | | | | | Metanalysis | | | | | R2 | | |
|---|---|---|---|---|---|---|---|---|---|---|---|---|---|---|---|---|---|---|---|---|---|
| **Description** | **Group** | **N** | **Effect estimate** | **S.E.** | **p-value** | **p-value$_{FDR}$** | **p-value rank** | **N** | **Effect estimate** | **S.E.** | **p-value** | **p-value$_{FDR}$** | **p-value rank** | **Effect estimate** | **S.E.** | **p-value** | **$I^2$** | **Q p-value** | **marginal R2** | **conditional R2** | **R2 rank** |
| Boiled coffee | Beverage | 27757 | 0.08 | 0.01 | 9.41E-36 | **2.54E-33** | 1 | 27040 | 0.08 | 0.01 | 7.33E-36 | **2.03E-33** | 1 | 0.08 | 0.00 | 7.71E-70 | 0.00 | 9.36E-01 | 0.10 | 0.59 | 1 |
| Palmitic acid intake (g/day) | Nutrients | 27757 | 0.07 | 0.01 | 4.39E-28 | **5.92E-26** | 2 | 27040 | 0.06 | 0.01 | 4.45E-26 | **6.17E-24** | 2 | 0.07 | 0.00 | 1.95E-52 | 0.00 | 7.96E-01 | 0.10 | 0.59 | 2 |
| Monounsaturated fat intake (g/day) | Nutrients | 27757 | 0.06 | 0.01 | 7.61E-22 | **6.85E-20** | 3 | 27040 | 0.06 | 0.01 | 1.19E-20 | **1.10E-18** | 3 | 0.06 | 0.00 | 7.75E-41 | 0.00 | 9.31E-01 | 0.10 | 0.59 | 3 |
| Formic acid intake (g/day) | Nutrients | 27757 | 0.06 | 0.01 | 3.18E-20 | **2.14E-18** | 4 | 27040 | 0.05 | 0.01 | 5.44E-17 | **2.15E-15** | 7 | 0.05 | 0.00 | 1.59E-35 | 0.00 | 5.77E-01 | 0.10 | 0.59 | 5 |
| Trans fat intake (g/day) | Nutrients | 27757 | 0.08 | 0.01 | 1.59E-19 | **8.56E-18** | 5 | 27040 | 0.07 | 0.01 | 2.27E-16 | **6.99E-15** | 9 | 0.07 | 0.01 | 3.20E-34 | 0.00 | 5.99E-01 | 0.10 | 0.59 | 4 |
| Carbohydrates intake (g/day) | Nutrients | 27757 | -0.05 | 0.01 | 9.55E-15 | **2.87E-13** | 9 | 27040 | -0.05 | 0.01 | 3.05E-18 | **2.11E-16** | 4 | -0.05 | 0.00 | 2.87E-31 | 0.00 | 4.47E-01 | 0.10 | 0.59 | 7 |
| Plant based protein intake (g/day) | Nutrients | 27757 | -0.05 | 0.01 | 7.30E-15 | **2.46E-13** | 8 | 27040 | -0.05 | 0.01 | 1.90E-16 | **6.57E-15** | 8 | -0.05 | 0.00 | 1.09E-29 | 0.00 | 6.87E-01 | 0.10 | 0.59 | 6 |
| Vitamin C intake (mg/day) | Nutrients | 27757 | -0.04 | 0.01 | 5.06E-13 | **1.07E-11** | 12 | 27040 | -0.05 | 0.01 | 6.19E-18 | **3.43E-16** | 5 | -0.05 | 0.00 | 3.89E-29 | 0.11 | 2.89E-01 | 0.10 | 0.59 | 8 |
| Fibre intake (g/day) | Nutrients | 27757 | -0.05 | 0.01 | 5.16E-13 | **1.07E-11** | 13 | 27040 | -0.05 | 0.01 | 2.23E-15 | **6.18E-14** | 10 | -0.05 | 0.00 | 9.37E-27 | 0.00 | 5.56E-01 | 0.10 | 0.59 | 9 |
| Saturated fat intake (g/day) | Nutrients | 27757 | 0.05 | 0.01 | 8.46E-14 | **2.28E-12** | 10 | 27040 | 0.05 | 0.01 | 4.39E-14 | **1.01E-12** | 12 | 0.05 | 0.00 | 2.54E-26 | 0.00 | 9.24E-01 | 0.10 | 0.59 | 12 |
| Fat intake (g/day) | Nutrients | 27757 | 0.04 | 0.01 | 2.27E-13 | **5.58E-12** | 11 | 27040 | 0.05 | 0.01 | 2.43E-14 | **6.12E-13** | 11 | 0.05 | 0.00 | 3.88E-26 | 0.00 | 7.87E-01 | 0.10 | 0.59 | 11 |
| Frequency of hunting or fishing during leisure time | Physical activity | 17739 | 0.06 | 0.01 | 1.72E-10 | **2.73E-09** | 17 | 17210 | 0.08 | 0.01 | 1.35E-17 | **6.22E-16** | 6 | 0.07 | 0.01 | 4.39E-26 | 0.55 | 1.35E-01 | 0.09 | 0.57 | 62 |
| Pentadecanoic acid intake (g/day) | Nutrients | 27757 | 0.05 | 0.01 | 3.95E-15 | **1.53E-13** | 6 | 27040 | 0.04 | 0.01 | 2.13E-11 | **3.68E-10** | 16 | 0.04 | 0.00 | 7.56E-25 | 0.00 | 4.27E-01 | 0.10 | 0.59 | 14 |
| Heptadecanoic acid intake (g/day) | Nutrients | 27757 | 0.05 | 0.01 | 3.95E-15 | **1.53E-13** | 7 | 27040 | 0.04 | 0.01 | 2.13E-11 | **3.68E-10** | 15 | 0.04 | 0.00 | 7.56E-25 | 0.00 | 4.27E-01 | 0.10 | 0.59 | 15 |
| Cholesterol intake (g/day) | Nutrients | 27757 | 0.04 | 0.01 | 2.77E-11 | **4.67E-10** | 16 | 27040 | 0.05 | 0.01 | 7.66E-14 | **1.63E-12** | 13 | 0.04 | 0.00 | 1.64E-23 | 0.00 | 5.40E-01 | 0.10 | 0.59 | 16 |
| Breakfast habits: Porridge w/o sandwich for breakfast vs not breakfast at all | Food | 4159 | -0.31 | 0.04 | 2.57E-12 | **4.95E-11** | 14 | 4229 | -0.27 | 0.04 | 1.33E-10 | **2.04E-09** | 18 | -0.29 | 0.03 | 2.49E-21 | 0.00 | 5.12E-01 | 0.08 | 0.60 | 66 |
| Banana | Food | 27757 | -0.04 | 0.01 | 8.02E-10 | **9.85E-09** | 22 | 27040 | -0.04 | 0.01 | 1.48E-12 | **2.93E-11** | 14 | -0.04 | 0.00 | 9.69E-21 | 0.00 | 4.35E-01 | 0.10 | 0.59 | 13 |
| Bregott on bread | Food | 27757 | 0.04 | 0.01 | 5.21E-12 | **9.38E-11** | 15 | 27040 | 0.03 | 0.01 | 1.03E-08 | **8.92E-08** | 32 | 0.04 | 0.00 | 4.16E-19 | 0.00 | 4.37E-01 | 0.10 | 0.59 | 18 |
| Average portion size of potatoes/rice/pasta based on photographic illustration of four sizes (smallest to largest) | Food | 27757 | -0.04 | 0.01 | 3.23E-10 | **4.85E-09** | 18 | 27040 | -0.04 | 0.01 | 3.31E-10 | **4.58E-09** | 20 | -0.04 | 0.00 | 6.18E-19 | 0.00 | 9.65E-01 | 0.07 | 0.55 | 78 |
| Exercise during the last three months | Physical activity | 30003 | -0.04 | 0.01 | 3.47E-10 | **4.93E-09** | 19 | 29318 | -0.04 | 0.01 | 9.67E-10 | **1.07E-08** | 25 | -0.04 | 0.00 | 1.92E-18 | 0.00 | 9.29E-01 | 0.07 | 0.55 | 71 |
| Educational level | Psychosocial | 30365 | -0.04 | 0.01 | 6.64E-10 | **8.54E-09** | 21 | 29654 | -0.04 | 0.01 | 6.15E-10 | **7.69E-09** | 22 | -0.04 | 0.01 | 2.32E-18 | 0.00 | 9.93E-01 | 0.07 | 0.54 | 74 |
| Tea | Beverage | 27757 | -0.04 | 0.01 | 5.15E-10 | **6.95E-09** | 20 | 27040 | -0.04 | 0.01 | 1.10E-09 | **1.17E-08** | 26 | -0.04 | 0.00 | 3.20E-18 | 0.00 | 9.68E-01 | 0.10 | 0.59 | 10 |
| Frequency of picking berries or mushrooms during leisure time | Physical activity | 18004 | 0.05 | 0.01 | 3.04E-09 | **3.16E-08** | 26 | 17506 | 0.05 | 0.01 | 3.20E-10 | **4.58E-09** | 19 | 0.05 | 0.01 | 5.58E-18 | 0.00 | 8.21E-01 | 0.09 | 0.57 | 65 |
| Fitness status | Physical activity | 24265 | -0.04 | 0.01 | 2.42E-09 | **2.72E-08** | 24 | 23834 | -0.04 | 0.01 | 1.41E-09 | **1.40E-08** | 28 | -0.04 | 0.00 | 1.89E-17 | 0.00 | 9.47E-01 | 0.07 | 0.57 | 75 |
| Fiber cereals | Food | 27757 | -0.03 | 0.01 | 7.44E-08 | **7.44E-07** | 27 | 27040 | -0.04 | 0.01 | 5.55E-10 | **7.33E-09** | 21 | -0.03 | 0.00 | 2.75E-16 | 0.00 | 5.03E-01 | 0.10 | 0.59 | 17 |
| Eat breakfast from 2000 | Food | 16919 | -0.14 | 0.03 | 5.37E-07 | **5.00E-06** | 29 | 16508 | -0.18 | 0.03 | 6.99E-11 | **1.14E-09** | 17 | -0.16 | 0.02 | 3.00E-16 | 0.00 | 3.43E-01 | 0.10 | 0.56 | 61 |
| Smoking status: Smokers vs non-smokers | Tobacco use | 19296 | 0.11 | 0.02 | 1.07E-07 | **1.04E-06** | 28 | 18958 | 0.13 | 0.02 | 6.39E-10 | **7.69E-09** | 23 | 0.12 | 0.01 | 4.24E-16 | 0.00 | 5.70E-01 | 0.07 | 0.54 | 67 |
| Number of cigarretes smoked per day (in groups) | Tobacco use | 24912 | 0.04 | 0.01 | 9.72E-07 | **8.20E-06** | 32 | 24558 | 0.04 | 0.01 | 8.38E-10 | **9.67E-09** | 24 | 0.04 | 0.01 | 5.91E-15 | 0.00 | 3.93E-01 | 0.07 | 0.55 | 69 |
| Cream, creme fraiche, sour cream | Food | 27757 | 0.04 | 0.01 | 9.60E-10 | **1.13E-08** | 23 | 27040 | 0.03 | 0.01 | 1.63E-06 | **1.01E-05** | 45 | 0.03 | 0.00 | 1.23E-14 | 0.00 | 3.39E-01 | 0.10 | 0.59 | 28 |
| Bacon | Food | 27757 | 0.03 | 0.01 | 6.41E-07 | **5.77E-06** | 30 | 27040 | 0.03 | 0.01 | 5.99E-09 | **5.35E-08** | 31 | 0.03 | 0.00 | 2.30E-14 | 0.00 | 5.42E-01 | 0.10 | 0.59 | 22 |
| Iron intake (mg/day) | Nutrients | 27757 | -0.04 | 0.01 | 2.81E-09 | **3.03E-08** | 25 | 27040 | -0.03 | 0.01 | 2.19E-06 | **1.32E-05** | 46 | -0.03 | 0.00 | 4.14E-14 | 0.00 | 4.25E-01 | 0.10 | 0.59 | 24 |
| Calcium intake (mg/day) | Nutrients | 27757 | -0.03 | 0.01 | 3.81E-06 | **2.94E-05** | 35 | 27040 | -0.04 | 0.01 | 1.32E-09 | **1.35E-08** | 27 | -0.03 | 0.00 | 4.41E-14 | 0.15 | 2.78E-01 | 0.10 | 0.59 | 20 |
| Vitamin D intake (ug/day) | Nutrients | 27757 | 0.03 | 0.01 | 1.50E-06 | **1.19E-05** | 34 | 27040 | 0.03 | 0.01 | 1.62E-08 | **1.32E-07** | 34 | 0.03 | 0.00 | 1.44E-13 | 0.00 | 5.22E-01 | 0.10 | 0.59 | 21 |
| Years smoking | Tobacco use | 26509 | 0.03 | 0.01 | 8.23E-06 | **5.85E-05** | 38 | 25785 | 0.04 | 0.01 | 3.29E-09 | **3.04E-08** | 30 | 0.04 | 0.01 | 2.18E-13 | 0.06 | 3.03E-01 | 0.07 | 0.54 | 77 |
| Alcohol intake (g/day) | Alcohol | 27757 | 0.03 | 0.01 | 1.26E-05 | **8.54E-05** | 40 | 27040 | 0.04 | 0.01 | 1.26E-08 | **1.06E-07** | 33 | 0.03 | 0.00 | 1.15E-12 | 0.00 | 3.45E-01 | 0.10 | 0.59 | 19 |
| Equol intake (ug/day) | Nutrients | 27757 | -0.03 | 0.01 | 8.68E-07 | **7.56E-06** | 31 | 27040 | -0.03 | 0.01 | 7.93E-07 | **5.36E-06** | 41 | -0.03 | 0.00 | 3.18E-12 | 0.00 | 9.69E-01 | 0.10 | 0.59 | 27 |
| Average portion size of meat/fish based on photographic illustration of four sizes (smallest to largest) | Food | 27757 | 0.03 | 0.01 | 5.23E-06 | **3.93E-05** | 36 | 27040 | 0.04 | 0.01 | 1.17E-07 | **8.98E-07** | 36 | 0.03 | 0.00 | 3.33E-12 | 0.00 | 5.58E-01 | 0.07 | 0.55 | 81 |
| Margarine for cooking | Food | 27757 | 0.02 | 0.01 | 2.18E-04 | **1.18E-03** | 50 | 27040 | 0.04 | 0.01 | 1.47E-09 | **1.40E-08** | 29 | 0.03 | 0.00 | 5.73E-12 | 0.65 | 9.17E-02 | 0.10 | 0.59 | 23 |
| Butter for cooking | Food | 27757 | 0.03 | 0.01 | 9.08E-06 | **6.29E-05** | 39 | 27040 | 0.03 | 0.01 | 2.23E-07 | **1.63E-06** | 38 | 0.03 | 0.00 | 1.03E-11 | 0.00 | 6.16E-01 | 0.10 | 0.59 | 29 |
| Oatflake, whole wheat, rye or barley porridge | Food | 27757 | -0.03 | 0.01 | 5.99E-06 | **4.37E-05** | 37 | 27040 | -0.03 | 0.01 | 1.05E-06 | **6.63E-06** | 44 | -0.03 | 0.00 | 2.90E-11 | 0.00 | 7.79E-01 | 0.10 | 0.59 | 25 |
| Low fat margarine on bread | Food | 27757 | -0.03 | 0.01 | 1.39E-06 | **1.13E-05** | 33 | 27040 | -0.03 | 0.01 | 1.00E-05 | **5.34E-05** | 51 | -0.03 | 0.00 | 6.25E-11 | 0.00 | 7.95E-01 | 0.10 | 0.59 | 39 |
| Meat stew | Food | 27757 | 0.02 | 0.01 | 4.92E-05 | **3.24E-04** | 41 | 27040 | 0.03 | 0.01 | 8.80E-07 | **5.80E-06** | 42 | 0.03 | 0.00 | 2.24E-10 | 0.00 | 5.19E-01 | 0.10 | 0.59 | 26 |
| Average portion size of vegetables based on photographic illustration of four sizes (smallest to largest) | Food | 27757 | -0.02 | 0.01 | 3.60E-04 | **1.72E-03** | 56 | 27040 | -0.03 | 0.01 | 4.07E-07 | **2.89E-06** | 39 | -0.03 | 0.00 | 1.06E-09 | 0.15 | 2.78E-01 | 0.07 | 0.55 | 82 |
| Vitamin B3 intake (mg/day) | Nutrients | 27757 | 0.02 | 0.01 | 1.58E-04 | **8.91E-04** | 48 | 27040 | 0.03 | 0.01 | 1.02E-05 | **5.34E-05** | 53 | 0.02 | 0.00 | 7.09E-09 | 0.00 | 6.29E-01 | 0.10 | 0.59 | 30 |
| Docosahexaenoic acid (DHA) intake (g/day) | Nutrients | 27757 | 0.02 | 0.01 | 5.29E-04 | **2.30E-03** | 62 | 27040 | 0.03 | 0.01 | 4.10E-06 | **2.36E-05** | 48 | 0.02 | 0.00 | 1.22E-08 | 0.00 | 3.77E-01 | 0.10 | 0.59 | 34 |
| Overall state of health compared to other your age | General health | 16031 | -0.03 | 0.01 | 7.28E-05 | **4.37E-04** | 45 | 15555 | -0.03 | 0.01 | 5.75E-05 | **2.61E-04** | 61 | -0.03 | 0.01 | 1.60E-08 | 0.00 | 9.60E-01 | 0.09 | 0.52 | 64 |
| Beta-carotene intake (mg/day) | Nutrients | 27757 | -0.02 | 0.01 | 1.81E-04 | **9.98E-04** | 49 | 27040 | -0.03 | 0.01 | 2.34E-05 | **1.16E-04** | 56 | -0.02 | 0.00 | 1.74E-08 | 0.00 | 7.07E-01 | 0.10 | 0.59 | 38 |
| Fatty fish (e.g. herring, whitefish, salmon) | Food | 27757 | 0.02 | 0.01 | 6.55E-04 | **2.76E-03** | 64 | 27040 | 0.03 | 0.01 | 5.74E-06 | **3.18E-05** | 50 | 0.02 | 0.00 | 1.99E-08 | 0.00 | 4.09E-01 | 0.10 | 0.59 | 36 |
| Total energy intake (kcal/day) | Nutrients | 27757 | -0.02 | 0.01 | 8.02E-04 | **3.23E-03** | 67 | 27040 | -0.03 | 0.01 | 5.41E-06 | **3.06E-05** | 49 | -0.03 | 0.00 | 2.34E-08 | 0.00 | 3.91E-01 | 0.10 | 0.59 | 60 |
| Sour milk, yoghurt (low fat) | Food | 27757 | -0.02 | 0.01 | 7.22E-05 | **4.37E-04** | 44 | 27040 | -0.02 | 0.01 | 1.25E-04 | **5.33E-04** | 64 | -0.02 | 0.00 | 3.39E-08 | 0.00 | 9.44E-01 | 0.10 | 0.59 | 42 |
| Vitamin B2 intake (ug/day) | Nutrients | 27757 | -0.02 | 0.01 | 3.27E-04 | **1.60E-03** | 55 | 27040 | -0.03 | 0.01 | 3.00E-05 | **1.43E-04** | 58 | -0.02 | 0.00 | 4.05E-08 | 0.00 | 6.53E-01 | 0.10 | 0.59 | 35 |
| Number of friends that can come to your home at any time and feel at home | Social | 30199 | 0.02 | 0.01 | 9.98E-04 | **3.90E-03** | 69 | 29532 | 0.03 | 0.01 | 1.02E-05 | **5.34E-05** | 52 | 0.02 | 0.00 | 5.16E-08 | 0.00 | 4.17E-01 | 0.07 | 0.55 | 80 |
| Campesterol intake (mg/day) | Nutrients | 27757 | 0.02 | 0.01 | 8.11E-05 | **4.76E-04** | 46 | 27040 | 0.02 | 0.01 | 1.85E-04 | **7.65E-04** | 67 | 0.02 | 0.00 | 5.60E-08 | 0.00 | 9.10E-01 | 0.10 | 0.59 | 32 |
| Strong beer | Alcohol | 27757 | 0.02 | 0.01 | 2.85E-04 | **1.42E-03** | 54 | 27040 | 0.03 | 0.01 | 5.25E-05 | **2.42E-04** | 60 | 0.02 | 0.00 | 5.84E-08 | 0.00 | 7.45E-01 | 0.10 | 0.59 | 31 |
| Monosaccharides intake (g/day) | Nutrients | 27757 | -0.02 | 0.01 | 6.87E-03 | **2.11E-02** | 88 | 27040 | -0.03 | 0.01 | 7.68E-07 | **5.32E-06** | 40 | -0.02 | 0.00 | 6.98E-08 | 0.63 | 1.01E-01 | 0.10 | 0.59 | 46 |
| Fried potatoes, pommes frites | Food | 27757 | 0.02 | 0.01 | 5.44E-05 | **3.50E-04** | 42 | 27040 | 0.02 | 0.01 | 4.21E-04 | **1.60E-03** | 73 | 0.02 | 0.00 | 9.10E-08 | 0.00 | 6.80E-01 | 0.10 | 0.59 | 33 |
| Root vegetables, carrot | Food | 27757 | -0.02 | 0.01 | 3.64E-04 | **1.72E-03** | 57 | 27040 | -0.02 | 0.01 | 6.55E-05 | **2.93E-04** | 62 | -0.02 | 0.00 | 9.19E-08 | 0.00 | 7.41E-01 | 0.10 | 0.59 | 43 |
| Eicosapentaenoic acid (EPA) intake (g/day) | Nutrients | 27756 | 0.02 | 0.01 | 1.45E-03 | **5.50E-03** | 71 | 27040 | 0.03 | 0.01 | 1.38E-05 | **6.93E-05** | 54 | 0.02 | 0.00 | 1.06E-07 | 0.00 | 3.76E-01 | 0.10 | 0.59 | 40 |
| Folic acid intake (ug/day) | Nutrients | 27757 | -0.02 | 0.01 | 2.74E-04 | **1.42E-03** | 51 | 27040 | -0.02 | 0.01 | 1.38E-04 | **5.79E-04** | 66 | -0.02 | 0.00 | 1.38E-07 | 0.00 | 8.69E-01 | 0.10 | 0.59 | 47 |
| Steak, chop, e.g. | Food | 27757 | 0.01 | 0.01 | 3.94E-02 | 9.24E-02 | 115 | 27040 | 0.03 | 0.01 | 1.28E-07 | **9.59E-07** | 37 | 0.02 | 0.00 | 2.24E-07 | 0.81 | 2.08E-02 | - | - | - |
| Polyunsaturated fat intake (g/day) | Nutrients | 27757 | 0.02 | 0.01 | 2.84E-04 | **1.42E-03** | 53 | 27040 | 0.02 | 0.01 | 2.99E-04 | **1.20E-03** | 69 | 0.02 | 0.00 | 3.00E-07 | 0.00 | 9.76E-01 | 0.10 | 0.59 | 44 |
| Arachidonic acid (ARA) intake (g/day) | Nutrients | 27756 | 0.01 | 0.01 | 1.96E-02 | 5.14E-02 | 103 | 27039 | 0.03 | 0.01 | 1.02E-06 | **6.56E-06** | 43 | 0.02 | 0.00 | 3.50E-07 | 0.71 | 6.51E-02 | - | - | - |
| Self rate of overall health | General health | 14290 | -0.03 | 0.01 | 2.67E-03 | **9.24E-03** | 78 | 14107 | -0.03 | 0.01 | 4.72E-05 | **2.22E-04** | 59 | -0.03 | 0.01 | 5.64E-07 | 0.00 | 4.33E-01 | 0.07 | 0.58 | 85 |
| Wine | Alcohol | 27757 | 0.02 | 0.01 | 1.57E-03 | **5.79E-03** | 73 | 27040 | 0.02 | 0.01 | 1.25E-04 | **5.33E-04** | 65 | 0.02 | 0.00 | 7.56E-07 | 0.00 | 6.16E-01 | 0.10 | 0.59 | 48 |
| Tomato, cucumber | Food | 27757 | -0.02 | 0.01 | 2.77E-04 | **1.42E-03** | 52 | 27040 | -0.02 | 0.01 | 9.86E-04 | **3.41E-03** | 80 | -0.02 | 0.00 | 9.46E-07 | 0.00 | 8.41E-01 | 0.10 | 0.59 | 49 |
| Low fat milk (0,5%) | Beverage | 27757 | -0.02 | 0.01 | 7.01E-04 | **2.91E-03** | 65 | 27040 | -0.02 | 0.01 | 4.39E-04 | **1.62E-03** | 75 | -0.02 | 0.00 | 1.05E-06 | 0.00 | 9.06E-01 | 0.10 | 0.59 | 53 |
| If you exercise, change in exercise habits during the last year | Physical activity | 16768 | -0.03 | 0.01 | 9.82E-04 | **3.90E-03** | 68 | 16339 | -0.03 | 0.01 | 3.19E-04 | **1.25E-03** | 71 | -0.03 | 0.01 | 1.08E-06 | 0.00 | 8.51E-01 | 0.09 | 0.57 | 63 |
| Rice | Food | 27757 | -0.02 | 0.01 | 4.21E-04 | **1.91E-03** | 59 | 27040 | -0.02 | 0.01 | 1.57E-03 | **5.23E-03** | 83 | -0.02 | 0.00 | 2.24E-06 | 0.00 | 8.02E-01 | 0.10 | 0.59 | 50 |
| Medium beer | Alcohol | 27757 | 0.01 | 0.01 | 2.01E-02 | 5.18E-02 | 104 | 27040 | 0.03 | 0.01 | 1.38E-05 | **6.93E-05** | 55 | 0.02 | 0.00 | 2.46E-06 | 0.53 | 1.46E-01 | - | - | - |
| Secoisolariciresinol intake (ug/day) | Nutrients | 27757 | 0.02 | 0.01 | 4.51E-04 | **2.00E-03** | 61 | 27040 | 0.02 | 0.01 | 2.10E-03 | **6.70E-03** | 87 | 0.02 | 0.00 | 3.23E-06 | 0.00 | 7.61E-01 | 0.10 | 0.59 | 41 |
| Whole grain crisp bread | Food | 27757 | 0.02 | 0.01 | 3.58E-03 | **1.18E-02** | 82 | 27040 | 0.02 | 0.01 | 3.10E-04 | **1.23E-03** | 70 | 0.02 | 0.00 | 4.09E-06 | 0.00 | 6.02E-01 | 0.10 | 0.59 | 37 |
| High mental demand from job | Psychosocial | 29607 | 0.02 | 0.01 | 4.13E-04 | **1.91E-03** | 58 | 29003 | 0.02 | 0.01 | 3.21E-03 | **9.78E-03** | 91 | 0.02 | 0.00 | 4.61E-06 | 0.00 | 6.89E-01 | 0.07 | 0.55 | 76 |
| Snuff status: Former snuff users vs non-snuff users | Tobacco use | 24195 | 0.08 | 0.02 | 4.24E-04 | **1.91E-03** | 60 | 23607 | 0.06 | 0.02 | 3.86E-03 | **1.16E-02** | 92 | 0.07 | 0.02 | 5.80E-06 | 0.00 | 6.39E-01 | 0.07 | 0.55 | 72 |
| Sucrose intake (g/day) | Nutrients | 27757 | -0.02 | 0.01 | 4.10E-03 | **1.32E-02** | 84 | 27040 | -0.02 | 0.01 | 4.27E-04 | **1.60E-03** | 74 | -0.02 | 0.00 | 6.29E-06 | 0.00 | 6.17E-01 | 0.10 | 0.59 | 45 |
| Milk, sour milk (3%) | Beverage | 27757 | 0.02 | 0.01 | 9.09E-03 | **2.61E-02** | 94 | 27040 | 0.02 | 0.01 | 1.89E-04 | **7.72E-04** | 68 | 0.02 | 0.00 | 7.68E-06 | 0.00 | 3.95E-01 | 0.10 | 0.59 | 54 |
| Linolenic acid intake (g/day) | Nutrients | 27757 | 0.02 | 0.01 | 1.52E-03 | **5.72E-03** | 72 | 27040 | 0.02 | 0.01 | 2.20E-03 | **6.85E-03** | 89 | 0.02 | 0.00 | 1.05E-05 | 0.00 | 9.62E-01 | 0.10 | 0.59 | 51 |
| High physical demand from job | Physical activity | 29929 | 0.01 | 0.01 | 1.18E-01 | 2.01E-01 | 158 | 29288 | 0.03 | 0.01 | 3.70E-06 | **2.18E-05** | 47 | 0.02 | 0.00 | 1.15E-05 | 0.78 | 3.13E-02 | - | - | - |
| Sodium intake (mg/day) | Nutrients | 27756 | 0.02 | 0.01 | 2.58E-03 | **9.04E-03** | 77 | 27039 | 0.02 | 0.01 | 1.56E-03 | **5.23E-03** | 82 | 0.02 | 0.00 | 1.25E-05 | 0.00 | 8.94E-01 | 0.10 | 0.59 | 52 |
| Cohabitation: Live alone vs Only children | Social | 4519 | -0.11 | 0.04 | 2.00E-03 | **7.22E-03** | 74 | 4516 | -0.11 | 0.04 | 4.23E-03 | **1.23E-02** | 95 | -0.11 | 0.03 | 2.58E-05 | 0.00 | 8.60E-01 | 0.07 | 0.52 | 86 |
| Breakfast habits: Only coffee/tea for breakfast vs not breakfast at all | Food | 1888 | -0.14 | 0.06 | 1.60E-02 | **4.31E-02** | 100 | 1959 | -0.20 | 0.06 | 5.96E-04 | **2.12E-03** | 78 | -0.17 | 0.04 | 3.57E-05 | 0.00 | 4.58E-01 | 0.05 | 0.48 | 87 |
| Last time a colleague visited you at home | Psychosocial | 29429 | 0.01 | 0.01 | 1.20E-01 | 2.03E-01 | 159 | 28815 | 0.02 | 0.01 | 2.54E-05 | **1.24E-04** | 57 | 0.02 | 0.00 | 4.69E-05 | 0.72 | 5.79E-02 | - | - | - |
| Parents or siblings had a cerebral hemorrhage/thrombosis or cardiac infarction before the age of 60 | General health | 29740 | 0.05 | 0.02 | 2.33E-03 | **8.29E-03** | 76 | 29118 | 0.04 | 0.02 | 7.73E-03 | **2.10E-02** | 102 | 0.05 | 0.01 | 5.44E-05 | 0.00 | 7.82E-01 | 0.07 | 0.54 | 79 |
| Sugar, honey, marmelade, jam | Food | 27757 | 0.02 | 0.01 | 1.10E-02 | **3.13E-02** | 95 | 27040 | 0.02 | 0.01 | 1.59E-03 | **5.24E-03** | 84 | 0.02 | 0.00 | 5.61E-05 | 0.00 | 6.51E-01 | 0.10 | 0.59 | 57 |
| Total protein intake (g/day) | Nutrients | 27757 | -0.02 | 0.01 | 6.22E-04 | **2.67E-03** | 63 | 27040 | -0.01 | 0.01 | 2.95E-02 | 6.39E-02 | 128 | -0.02 | 0.00 | 7.39E-05 | 0.00 | 3.90E-01 | - | - | - |
| Frequency of gardening during leisure time | Physical activity | 18121 | 0.02 | 0.01 | 3.59E-02 | 8.59E-02 | 113 | 17550 | 0.03 | 0.01 | 4.97E-04 | **1.81E-03** | 76 | 0.02 | 0.01 | 7.85E-05 | 0.00 | 3.32E-01 | - | - | - |
| Physical limitation to participate in steneous activities: running, lifting heavy objects, taking part in physically demanding sports | General health | 14260 | 0.03 | 0.01 | 2.00E-03 | **7.22E-03** | 75 | 14082 | 0.02 | 0.01 | 1.31E-02 | **3.28E-02** | 111 | 0.02 | 0.01 | 8.09E-05 | 0.00 | 6.73E-01 | 0.07 | 0.59 | 84 |
| Travel to work: Cycle to work vs passive travel to work | Physical activity | 22876 | -0.05 | 0.02 | 4.45E-03 | **1.41E-02** | 85 | 22241 | -0.04 | 0.02 | 7.72E-03 | **2.10E-02** | 101 | -0.05 | 0.01 | 9.83E-05 | 0.00 | 8.97E-01 | 0.07 | 0.55 | 70 |
| Smoking status: Former smokers vs non-smokers | Tobacco use | 21829 | 0.04 | 0.02 | 3.18E-02 | 7.87E-02 | 109 | 21070 | 0.06 | 0.02 | 8.17E-04 | **2.86E-03** | 79 | 0.05 | 0.01 | 1.02E-04 | 0.00 | 3.96E-01 | - | - | - |
| Satisfaction with home and family situation | Psychosocial | 24276 | 0.02 | 0.01 | 7.44E-04 | **3.05E-03** | 66 | 23859 | 0.01 | 0.01 | 3.48E-02 | 7.15E-02 | 135 | 0.02 | 0.00 | 1.05E-04 | 0.00 | 3.77E-01 | - | - | - |
| Excellent health | General health | 14238 | -0.02 | 0.01 | 5.21E-03 | **1.62E-02** | 87 | 14050 | -0.02 | 0.01 | 7.22E-03 | **2.00E-02** | 100 | -0.02 | 0.01 | 1.06E-04 | 0.00 | 9.44E-01 | 0.07 | 0.58 | 83 |

| | | | | | | | | | | | | | | | | | | | | | |
|---|---|---|---|---|---|---|---|---|---|---|---|---|---|---|---|---|---|---|---|---|---|
| Medioresinol intake (ug/day) | Nutrients | 27757 | 0.02 | 0.01 | 5.02E-03 | **1.58E-02** | 86 | 27040 | 0.02 | 0.01 | 8.52E-03 | **2.27E-02** | 104 | 0.02 | 0.00 | 1.21E-04 | 0.00 | 9.27E-01 | 0.10 | 0.59 | 55 |
| Boiled or baked potato | Food | 27757 | 0.01 | 0.01 | 1.90E-02 | 5.03E-02 | 102 | 27040 | 0.02 | 0.01 | 2.13E-03 | **6.70E-03** | 88 | 0.02 | 0.00 | 1.29E-04 | 0.00 | 5.93E-01 | - | - | - |
| Cambridge physical activity index | Physical activity | 29044 | -0.02 | 0.01 | 7.53E-03 | **2.19E-02** | 93 | 28387 | -0.02 | 0.01 | 6.83E-03 | **1.93E-02** | 98 | -0.02 | 0.00 | 1.43E-04 | 0.00 | 9.80E-01 | 0.07 | 0.55 | 73 |
| Salad dressing with oil | Food | 27757 | -0.01 | 0.01 | 1.15E-02 | **3.23E-02** | 96 | 27040 | -0.02 | 0.01 | 5.81E-03 | **1.67E-02** | 96 | -0.02 | 0.00 | 1.86E-04 | 0.00 | 8.65E-01 | 0.10 | 0.59 | 59 |
| Liquor, spirits | Alcohol | 27757 | 0.02 | 0.01 | 7.40E-03 | **2.17E-02** | 92 | 27040 | 0.02 | 0.01 | 9.07E-03 | **2.39E-02** | 105 | 0.02 | 0.00 | 1.87E-04 | 0.00 | 8.73E-01 | 0.10 | 0.59 | 56 |
| Beta-sitostanol intake (mg/day) | Nutrients | 27757 | 0.02 | 0.01 | 2.93E-03 | **1.00E-02** | 79 | 27040 | 0.01 | 0.01 | 2.30E-02 | 5.33E-02 | 119 | 0.02 | 0.00 | 2.03E-04 | 0.00 | 6.44E-01 | - | - | - |
| Frequency of cycling during leisure time | Physical activity | 24855 | -0.02 | 0.01 | 4.09E-03 | **1.32E-02** | 83 | 24149 | -0.02 | 0.01 | 1.77E-02 | **4.27E-02** | 115 | -0.02 | 0.00 | 2.08E-04 | 0.00 | 7.40E-01 | 0.07 | 0.55 | 68 |
| Rosehip, sweet syrup soup | Food | 27757 | -0.02 | 0.01 | 1.09E-04 | **6.24E-04** | 47 | 27040 | -0.01 | 0.01 | 1.76E-01 | 2.68E-01 | 182 | -0.01 | 0.00 | 2.22E-04 | 0.69 | 7.45E-02 | - | - | - |
| Salty fish | Food | 27757 | 0.01 | 0.01 | 7.39E-03 | **2.17E-02** | 91 | 27040 | 0.01 | 0.01 | 1.24E-02 | **3.13E-02** | 109 | 0.01 | 0.00 | 2.48E-04 | 0.00 | 9.94E-01 | 0.10 | 0.59 | 58 |
| Feel the need to reduce alcohol consumption | Alcohol | 27882 | 0.02 | 0.02 | 2.03E-01 | 3.07E-01 | 179 | 27306 | 0.07 | 0.02 | 1.10E-04 | **4.85E-04** | 63 | 0.05 | 0.01 | 2.67E-04 | 0.70 | 6.98E-02 | - | - | - |
| Sometimes physically straining work | Physical activity | 29412 | 0.02 | 0.01 | 1.16E-01 | 2.01E-01 | 155 | 28750 | 0.05 | 0.01 | 3.50E-04 | **1.35E-03** | 72 | 0.03 | 0.01 | 2.74E-04 | 0.50 | 1.56E-01 | - | - | - |
| Number of social contacts with the same interests as you | Social | 30264 | 0.02 | 0.01 | 7.00E-03 | **2.12E-02** | 89 | 29576 | 0.01 | 0.01 | 2.20E-02 | 5.21E-02 | 117 | 0.02 | 0.00 | 4.20E-04 | 0.00 | 7.81E-01 | - | - | - |
| Apple, pear, peach, orange, mandarin and grapefruit | Food | 27757 | -0.01 | 0.01 | 7.17E-02 | 1.42E-01 | 136 | 27040 | -0.02 | 0.01 | 1.49E-03 | **5.11E-03** | 81 | -0.02 | 0.00 | 4.46E-04 | 0.00 | 3.16E-01 | - | - | - |
| Stigmasterol intake (mg/day) | Nutrients | 27757 | -0.01 | 0.01 | 4.38E-02 | 9.69E-02 | 122 | 27040 | -0.02 | 0.01 | 5.86E-03 | **1.67E-02** | 97 | -0.01 | 0.00 | 7.49E-04 | 0.00 | 5.86E-01 | - | - | - |
| Enough time for job assignments | Psychosocial | 29719 | 0.02 | 0.01 | 7.13E-03 | **2.14E-02** | 90 | 29083 | 0.01 | 0.01 | 3.96E-02 | 8.01E-02 | 137 | 0.01 | 0.00 | 7.83E-04 | 0.00 | 6.61E-01 | - | - | - |
| White (soft) bread, thin crisp bread | Food | 27757 | 0.02 | 0.01 | 1.29E-02 | **3.60E-02** | 97 | 27040 | 0.01 | 0.01 | 2.69E-02 | 5.91E-02 | 126 | 0.01 | 0.00 | 8.90E-04 | 0.00 | 8.58E-01 | - | - | - |
| Changed everyday exercise during the last year | Physical activity | 19041 | -0.02 | 0.01 | 3.03E-03 | **1.02E-02** | 80 | 18522 | -0.01 | 0.01 | 8.30E-02 | 1.39E-01 | 165 | -0.02 | 0.01 | 9.01E-04 | 0.00 | 3.79E-01 | - | - | - |
| Frequncy of social contacts with colleagues during leisure time | Psychosocial | 29002 | 0.01 | 0.01 | 3.99E-02 | 9.28E-02 | 116 | 28377 | 0.02 | 0.01 | 9.22E-03 | **2.41E-02** | 106 | 0.01 | 0.00 | 9.88E-04 | 0.00 | 6.94E-01 | - | - | - |
| Minced meat dishes | Food | 27757 | 0.01 | 0.01 | 3.84E-02 | 9.10E-02 | 114 | 27040 | 0.01 | 0.01 | 1.42E-02 | **3.51E-02** | 112 | 0.01 | 0.00 | 1.38E-03 | 0.00 | 7.87E-01 | - | - | - |
| Cohabitation: Live alone vs Other/others | Social | 3466 | 0.11 | 0.06 | 6.44E-02 | 1.30E-01 | 133 | 3449 | 0.17 | 0.06 | 7.85E-03 | **2.11E-02** | 103 | 0.14 | 0.04 | 1.47E-03 | 0.00 | 5.35E-01 | - | - | - |
| Would you say that the number of people that you meet in your everyday life is enough or would you like to meet more or fewer people? | Social | 30300 | -0.02 | 0.01 | 6.79E-05 | **4.27E-04** | 43 | 29608 | 0.00 | 0.01 | 6.18E-01 | 6.96E-01 | 246 | -0.01 | 0.00 | 1.51E-03 | 0.84 | 1.38E-02 | - | - | - |
| Years using snuff | Tobacco use | 28552 | 0.01 | 0.01 | 1.79E-01 | 2.77E-01 | 175 | 27936 | 0.02 | 0.01 | 1.97E-03 | **6.34E-03** | 86 | 0.02 | 0.01 | 1.70E-03 | 0.35 | 2.16E-01 | - | - | - |
| Pasta | Food | 27757 | -0.01 | 0.01 | 2.02E-02 | 5.18E-02 | 105 | 27040 | -0.01 | 0.01 | 4.02E-02 | 8.08E-02 | 138 | -0.01 | 0.00 | 1.97E-03 | 0.00 | 8.58E-01 | - | - | - |
| Physical limitation to participate in moderately demanding activities: lifting or carrying grocery bags | General health | 14244 | 0.02 | 0.01 | 2.70E-02 | 6.88E-02 | 106 | 14069 | 0.02 | 0.01 | 3.24E-02 | 6.90E-02 | 130 | 0.02 | 0.01 | 2.09E-03 | 0.00 | 9.64E-01 | - | - | - |
| Marital status: Single vs Divorced/separated | Social | 5016 | -0.04 | 0.03 | 2.32E-01 | 3.42E-01 | 183 | 4980 | -0.11 | 0.03 | 1.62E-03 | **5.28E-03** | 85 | -0.07 | 0.02 | 2.09E-03 | 0.48 | 1.66E-01 | - | - | - |
| Breakfast habits: Gruel w/o sandwich for breakfast vs not breakfast at all | Food | 1429 | -0.16 | 0.07 | 3.45E-02 | 8.33E-02 | 112 | 1488 | -0.15 | 0.07 | 2.71E-02 | 5.91E-02 | 127 | -0.16 | 0.05 | 2.21E-03 | 0.00 | 9.84E-01 | - | - | - |
| Number of people with whom you can speak openly | Social | 30289 | 0.01 | 0.01 | 1.49E-02 | **4.07E-02** | 99 | 29595 | 0.01 | 0.01 | 5.87E-02 | 1.08E-01 | 150 | 0.01 | 0.00 | 2.22E-03 | 0.00 | 7.07E-01 | - | - | - |
| Permanent employment | Psychosocial | 25825 | 0.03 | 0.01 | 1.44E-02 | **3.97E-02** | 98 | 25108 | 0.03 | 0.01 | 6.56E-02 | 1.17E-01 | 155 | 0.03 | 0.01 | 2.43E-03 | 0.00 | 6.68E-01 | - | - | - |
| Sausage as main dish | Food | 27757 | 0.02 | 0.01 | 3.41E-03 | **1.14E-02** | 81 | 27040 | 0.01 | 0.01 | 2.04E-01 | 3.01E-01 | 188 | 0.01 | 0.00 | 3.03E-03 | 0.28 | 2.37E-01 | - | - | - |
| Participation in associations or voluntary organisations | Social | 30118 | -0.02 | 0.01 | 9.66E-02 | 1.76E-01 | 148 | 29472 | -0.03 | 0.01 | 1.16E-02 | **2.97E-02** | 108 | -0.03 | 0.01 | 3.08E-03 | 0.00 | 5.38E-01 | - | - | - |
| Satisfaction with work situation | Psychosocial | 23979 | -0.01 | 0.01 | 2.96E-02 | 7.47E-02 | 107 | 23548 | -0.01 | 0.01 | 4.48E-02 | 8.69E-02 | 143 | -0.01 | 0.00 | 3.11E-03 | 0.00 | 9.01E-01 | - | - | - |
| Distance to work in kilometers (one way) | Physical activity | 26228 | 0.02 | 0.01 | 1.33E-03 | **5.15E-03** | 70 | 25523 | 0.01 | 0.01 | 3.36E-01 | 4.42E-01 | 209 | 0.01 | 0.00 | 3.19E-03 | 0.61 | 1.12E-01 | - | - | - |
| For how much of the time during the last four weeks have you felt worn out? | General health | 14226 | 0.02 | 0.01 | 5.68E-02 | 1.19E-01 | 129 | 14009 | 0.02 | 0.01 | 2.51E-02 | 5.66E-02 | 123 | 0.02 | 0.01 | 3.37E-03 | 0.00 | 8.09E-01 | - | - | - |
| Physical limitation to participate in moderately demanding activities: moving a table, vacuuming, walking in the forest or gardening | General health | 14257 | 0.02 | 0.01 | 5.89E-02 | 1.22E-01 | 130 | 14079 | 0.02 | 0.01 | 2.62E-02 | 5.80E-02 | 125 | 0.02 | 0.01 | 3.64E-03 | 0.00 | 8.03E-01 | - | - | - |
| Frequency of engaging in clubs, associations or study circles | Social | 19002 | -0.01 | 0.01 | 8.01E-02 | 1.53E-01 | 141 | 18524 | -0.02 | 0.01 | 2.32E-02 | 5.33E-02 | 120 | -0.01 | 0.01 | 4.50E-03 | 0.00 | 7.05E-01 | - | - | - |
| Sweets | Food | 27757 | 0.01 | 0.01 | 4.79E-02 | 1.04E-01 | 124 | 27040 | 0.01 | 0.01 | 4.42E-02 | 8.69E-02 | 140 | 0.01 | 0.00 | 4.78E-03 | 0.00 | 9.84E-01 | - | - | - |
| Frequency of shoverling snow during leisure time | Physical activity | 18196 | 0.01 | 0.01 | 1.08E-01 | 1.90E-01 | 153 | 17692 | 0.02 | 0.01 | 2.40E-02 | 5.45E-02 | 122 | 0.02 | 0.01 | 6.26E-03 | 0.00 | 6.47E-01 | - | - | - |
| Enterolactone intake (ug/day) | Nutrients | 27757 | -0.01 | 0.01 | 8.53E-02 | 1.60E-01 | 144 | 27040 | -0.01 | 0.01 | 3.43E-02 | 7.13E-02 | 132 | -0.01 | 0.00 | 6.71E-03 | 0.00 | 7.64E-01 | - | - | - |
| Cheese 10-17% | Food | 27757 | -0.01 | 0.01 | 4.32E-02 | 9.64E-02 | 121 | 27040 | -0.01 | 0.01 | 7.96E-02 | 1.36E-01 | 162 | -0.01 | 0.00 | 7.57E-03 | 0.00 | 8.65E-01 | - | - | - |
| Energy status | Psychosocial | 24253 | -0.01 | 0.01 | 5.59E-02 | 1.18E-01 | 128 | 23808 | -0.01 | 0.01 | 6.26E-02 | 1.13E-01 | 154 | -0.01 | 0.00 | 7.62E-03 | 0.00 | 9.70E-01 | - | - | - |
| Informed of having high blood pressure | General health | 30326 | 0.03 | 0.01 | 6.89E-02 | 1.38E-01 | 135 | 29642 | 0.03 | 0.01 | 5.43E-02 | 1.03E-01 | 146 | 0.03 | 0.01 | 8.12E-03 | 0.00 | 9.43E-01 | - | - | - |
| Sleep status | Sleep | 24295 | -0.01 | 0.01 | 2.98E-01 | 4.20E-01 | 192 | 23879 | -0.02 | 0.01 | 7.04E-03 | **1.97E-02** | 99 | -0.01 | 0.00 | 8.25E-03 | 0.27 | 2.42E-01 | - | - | - |
| Physical limitation to participate in moderately demanding activities: walking up several stairs | General health | 14260 | 0.02 | 0.01 | 4.20E-02 | 9.64E-02 | 117 | 14063 | 0.01 | 0.01 | 9.33E-02 | 1.54E-01 | 168 | 0.02 | 0.01 | 8.63E-03 | 0.00 | 8.09E-01 | - | - | - |
| Frequency of dancing during leisure time | Physical activity | 17010 | -0.01 | 0.01 | 2.53E-01 | 3.66E-01 | 187 | 16479 | -0.02 | 0.01 | 1.06E-02 | **2.74E-02** | 107 | -0.02 | 0.01 | 8.82E-03 | 0.00 | 3.21E-01 | - | - | - |
| Snuff status: Snuff users vs non-snuff users | Tobacco use | 25844 | 0.01 | 0.02 | 5.35E-01 | 6.54E-01 | 221 | 25360 | 0.06 | 0.02 | 2.45E-03 | **7.54E-03** | 90 | 0.04 | 0.01 | 9.90E-03 | 0.66 | 8.80E-02 | - | - | - |
| Self-employed | Psychosocial | 25825 | -0.04 | 0.02 | 8.07E-02 | 1.53E-01 | 142 | 25108 | -0.04 | 0.02 | 5.76E-02 | 1.07E-01 | 149 | -0.04 | 0.02 | 9.93E-03 | 0.00 | 9.28E-01 | - | - | - |
| Overall state of health during the last year | General health | 30317 | -0.01 | 0.01 | 4.26E-02 | 9.64E-02 | 119 | 29635 | -0.01 | 0.01 | 1.09E-01 | 1.79E-01 | 169 | -0.01 | 0.00 | 1.03E-02 | 0.00 | 7.71E-01 | - | - | - |
| Butter on bread | Food | 27757 | 0.01 | 0.01 | 9.84E-02 | 1.77E-01 | 149 | 27040 | 0.01 | 0.01 | 6.06E-02 | 1.10E-01 | 152 | 0.01 | 0.00 | 1.26E-02 | 0.00 | 8.68E-01 | - | - | - |
| Margarine on bread | Food | 27757 | -0.01 | 0.01 | 1.00E-01 | 1.79E-01 | 151 | 27040 | -0.01 | 0.01 | 6.08E-02 | 1.10E-01 | 153 | -0.01 | 0.00 | 1.29E-02 | 0.00 | 8.64E-01 | - | - | - |
| Skill demand from job | Psychosocial | 29782 | 0.00 | 0.01 | 5.43E-01 | 6.61E-01 | 222 | 29113 | -0.02 | 0.01 | 4.11E-03 | **1.22E-02** | 93 | -0.01 | 0.00 | 1.41E-02 | 0.61 | 1.08E-01 | - | - | - |
| Sour milk, yoghurt (3% fat) | Food | 27757 | -0.01 | 0.01 | 3.29E-01 | 4.56E-01 | 194 | 27040 | -0.01 | 0.01 | 1.24E-02 | **3.13E-02** | 110 | -0.01 | 0.00 | 1.42E-02 | 0.17 | 2.74E-01 | - | - | - |
| Chips, popcorn, salted nuts | Food | 27757 | 0.01 | 0.01 | 4.26E-02 | 9.64E-02 | 118 | 27040 | 0.01 | 0.01 | 1.53E-01 | 2.40E-01 | 177 | 0.01 | 0.00 | 1.45E-02 | 0.00 | 6.79E-01 | - | - | - |
| Self rate of overall health compared to a year ago | General health | 14183 | -0.02 | 0.01 | 1.71E-02 | **4.57E-02** | 101 | 13976 | -0.01 | 0.01 | 2.92E-01 | 4.08E-01 | 198 | -0.01 | 0.01 | 1.49E-02 | 0.00 | 3.45E-01 | - | - | - |
| Corn flakes | Food | 27757 | -0.01 | 0.01 | 7.92E-02 | 1.53E-01 | 140 | 27040 | -0.01 | 0.01 | 9.23E-02 | 1.53E-01 | 167 | -0.01 | 0.00 | 1.50E-02 | 0.00 | 9.74E-01 | - | - | - |
| Pain during the last four weeks | General health | 14260 | 0.01 | 0.01 | 2.39E-01 | 3.49E-01 | 185 | 14048 | 0.02 | 0.01 | 2.57E-02 | 5.74E-02 | 124 | 0.01 | 0.01 | 1.59E-02 | 0.00 | 4.53E-01 | - | - | - |
| For how much of the time during the last four weeks have you felt full of energy? | General health | 14231 | -0.01 | 0.01 | 1.68E-01 | 2.66E-01 | 171 | 14038 | -0.02 | 0.01 | 4.46E-02 | 8.69E-02 | 142 | -0.01 | 0.01 | 1.66E-02 | 0.00 | 6.56E-01 | - | - | - |
| Brewed (filtered) coffee | Beverage | 27757 | -0.01 | 0.01 | 2.06E-01 | 3.07E-01 | 181 | 27040 | -0.01 | 0.01 | 3.44E-02 | 7.13E-02 | 133 | -0.01 | 0.00 | 1.70E-02 | 0.00 | 5.37E-01 | - | - | - |
| Pancake, waffle, Swedish dumpling | Food | 27757 | 0.01 | 0.01 | 1.73E-01 | 2.72E-01 | 172 | 27040 | 0.01 | 0.01 | 4.72E-02 | 9.07E-02 | 144 | 0.01 | 0.00 | 1.81E-02 | 0.00 | 6.50E-01 | - | - | - |
| How much has the pain during the last four weeks disturbed your normal work? | General health | 14221 | 0.01 | 0.01 | 2.91E-01 | 4.14E-01 | 190 | 14024 | 0.02 | 0.01 | 2.33E-02 | 5.33E-02 | 121 | 0.01 | 0.01 | 1.87E-02 | 0.00 | 3.87E-01 | - | - | - |
| Sedentary or standing work | Physical activity | 29412 | -0.02 | 0.01 | 1.10E-01 | 1.93E-01 | 154 | 28750 | -0.02 | 0.01 | 8.69E-02 | 1.45E-01 | 166 | -0.02 | 0.01 | 1.92E-02 | 0.00 | 9.41E-01 | - | - | - |
| Physical limitation that reduced the normal time spent at work or in other activities during the last four weeks | General health | 14287 | 0.02 | 0.03 | 4.25E-01 | 5.52E-01 | 208 | 14073 | 0.06 | 0.03 | 1.61E-02 | **3.91E-02** | 114 | 0.04 | 0.02 | 2.30E-02 | 0.22 | 2.57E-01 | - | - | - |
| Vitamin E intake (mg/day) | Nutrients | 27757 | 0.01 | 0.01 | 3.10E-01 | 4.34E-01 | 193 | 27040 | 0.01 | 0.01 | 3.45E-02 | 7.13E-02 | 134 | 0.01 | 0.00 | 2.71E-02 | 0.00 | 4.32E-01 | - | - | - |
| Contradictory demands in job | Psychosocial | 29676 | -0.01 | 0.01 | 3.12E-02 | 7.80E-02 | 108 | 29002 | -0.01 | 0.01 | 3.91E-01 | 4.84E-01 | 224 | -0.01 | 0.00 | 3.30E-02 | 0.00 | 3.62E-01 | - | - | - |
| Milk, sour milk (1,5%) | Beverage | 27757 | 0.01 | 0.01 | 4.61E-02 | 1.01E-01 | 123 | 27040 | 0.01 | 0.01 | 3.22E-01 | 4.39E-01 | 203 | 0.01 | 0.00 | 3.41E-02 | 0.00 | 4.92E-01 | - | - | - |
| For how much of the time during the last four weeks have you felt really alert and strong? | General health | 14229 | -0.02 | 0.01 | 5.36E-02 | 1.14E-01 | 127 | 14026 | -0.01 | 0.01 | 2.87E-01 | 4.04E-01 | 197 | -0.01 | 0.01 | 3.41E-02 | 0.00 | 5.39E-01 | - | - | - |
| Syringaresinol intake (ug/day) | Nutrients | 27757 | 0.01 | 0.01 | 8.64E-02 | 1.60E-01 | 146 | 27040 | 0.01 | 0.01 | 2.04E-01 | 3.01E-01 | 187 | 0.01 | 0.00 | 3.46E-02 | 0.00 | 7.68E-01 | - | - | - |
| Pinoresinol intake (ug/day) | Nutrients | 27757 | -0.01 | 0.01 | 1.49E-01 | 2.41E-01 | 167 | 27040 | -0.01 | 0.01 | 1.33E-01 | 2.12E-01 | 174 | -0.01 | 0.00 | 3.75E-02 | 0.00 | 9.54E-01 | - | - | - |
| Berries (fresh or frozen) | Food | 27757 | -0.01 | 0.01 | 1.76E-01 | 2.73E-01 | 174 | 27040 | -0.01 | 0.01 | 1.17E-01 | 1.90E-01 | 170 | -0.01 | 0.00 | 3.89E-02 | 0.00 | 8.74E-01 | - | - | - |
| Cohabitation: Live alone vs Only one adult (spouse, partner) | Social | 14031 | 0.05 | 0.02 | 4.29E-02 | 9.64E-02 | 120 | 13857 | 0.02 | 0.02 | 3.72E-01 | 4.66E-01 | 221 | 0.03 | 0.02 | 3.96E-02 | 0.00 | 4.15E-01 | - | - | - |
| As healthy as anyone | General health | 14197 | -0.01 | 0.01 | 4.83E-01 | 6.13E-01 | 213 | 14003 | -0.02 | 0.01 | 3.02E-02 | 6.49E-02 | 129 | -0.01 | 0.01 | 4.25E-02 | 0.09 | 2.96E-01 | - | - | - |
| Memory status | Psychosocial | 24234 | 0.01 | 0.01 | 1.30E-01 | 2.18E-01 | 161 | 23795 | 0.01 | 0.01 | 1.82E-01 | 2.76E-01 | 183 | 0.01 | 0.00 | 4.42E-02 | 0.00 | 8.94E-01 | - | - | - |
| Frequent social contacts with colleagues during work | Psychosocial | 29536 | -0.01 | 0.01 | 3.38E-02 | 8.25E-02 | 110 | 28893 | 0.00 | 0.01 | 4.86E-01 | 5.68E-01 | 237 | -0.01 | 0.00 | 4.63E-02 | 0.02 | 3.12E-01 | - | - | - |
| Light beer | Alcohol | 27757 | 0.01 | 0.01 | 8.49E-02 | 1.60E-01 | 143 | 27040 | 0.01 | 0.01 | 2.86E-01 | 4.04E-01 | 196 | 0.01 | 0.00 | 4.79E-02 | 0.00 | 6.61E-01 | - | - | - |
| White cabbage, lettuce, lettuce cabbage, spinach, borecole | Food | 27757 | -0.01 | 0.01 | 6.47E-02 | 1.30E-01 | 134 | 27040 | -0.01 | 0.01 | 3.47E-01 | 4.45E-01 | 216 | -0.01 | 0.00 | 4.80E-02 | 0.00 | 5.32E-01 | - | - | - |
| Cohabitation: Live alone vs Adult and children | Social | 17503 | -0.04 | 0.02 | 7.57E-02 | 1.48E-01 | 138 | 17095 | -0.02 | 0.02 | 3.35E-01 | 4.42E-01 | 208 | -0.03 | 0.02 | 5.31E-02 | 0.00 | 5.57E-01 | - | - | - |
| Teetotaler | Alcohol | 19025 | -0.03 | 0.03 | 3.54E-01 | 4.82E-01 | 198 | 18510 | -0.05 | 0.03 | 7.40E-02 | 1.29E-01 | 159 | -0.04 | 0.02 | 5.58E-02 | 0.00 | 5.30E-01 | - | - | - |
| Vitamin B6 intake (mg/day) | Nutrients | 27757 | -0.01 | 0.01 | 1.39E-01 | 2.32E-01 | 162 | 27040 | -0.01 | 0.01 | 2.34E-01 | 3.38E-01 | 192 | -0.01 | 0.00 | 5.90E-02 | 0.00 | 8.47E-01 | - | - | - |
| Zinc intake (mg/day) | Nutrients | 27757 | -0.01 | 0.01 | 2.19E-01 | 3.25E-01 | 182 | 27040 | -0.01 | 0.01 | 1.67E-01 | 2.57E-01 | 180 | -0.01 | 0.00 | 6.52E-02 | 0.00 | 9.00E-01 | - | - | - |
| Appetite status | Psychosocial | 24277 | 0.00 | 0.01 | 7.83E-01 | 8.53E-01 | 247 | 23852 | 0.01 | 0.01 | 2.03E-02 | **4.86E-02** | 116 | 0.01 | 0.00 | 6.60E-02 | 0.52 | 1.49E-01 | - | - | - |
| Marital status: Single vs Married/partner | Social | 27706 | -0.02 | 0.02 | 4.47E-01 | 5.69E-01 | 211 | 27035 | -0.04 | 0.02 | 7.74E-02 | 1.34E-01 | 160 | -0.03 | 0.02 | 7.34E-02 | 0.00 | 4.82E-01 | - | - | - |
| Do you feel important and appreciated in your home? | Psychosocial | 24120 | 0.01 | 0.01 | 1.74E-01 | 2.72E-01 | 173 | 23684 | 0.01 | 0.01 | 2.62E-01 | 3.72E-01 | 195 | 0.01 | 0.00 | 7.95E-02 | 0.00 | 8.69E-01 | - | - | - |
| Physical limitation that made you do less than you wanted during the last four weeks | General health | 14270 | 0.03 | 0.02 | 1.40E-01 | 2.32E-01 | 163 | 14075 | 0.02 | 0.02 | 3.41E-01 | 4.42E-01 | 214 | 0.03 | 0.01 | 8.58E-02 | 0.00 | 7.13E-01 | - | - | - |
| For how much of the time during the last four weeks have you felt calm and serene? | General health | 14249 | -0.01 | 0.01 | 4.02E-01 | 5.32E-01 | 204 | 14060 | -0.01 | 0.01 | 1.23E-01 | 1.99E-01 | 171 | -0.01 | 0.00 | 9.20E-02 | 0.00 | 6.17E-01 | - | - | - |
| Sum of all lignans intake (ug/day) | Nutrients | 27757 | 0.01 | 0.01 | 1.68E-01 | 2.66E-01 | 170 | 27040 | 0.01 | 0.01 | 3.27E-01 | 4.42E-01 | 204 | 0.01 | 0.00 | 9.48E-02 | 0.00 | 7.89E-01 | - | - | - |
| Breakfast habits: Coffee/tea and wheat buns or rusk for breakfast vs not breakfast at all | Food | 1575 | -0.10 | 0.06 | 8.91E-02 | 1.64E-01 | 147 | 1555 | -0.04 | 0.06 | 5.17E-01 | 5.97E-01 | 240 | -0.07 | 0.04 | 9.61E-02 | 0.00 | 4.53E-01 | - | - | - |
| Campestanol intake (mg/day) | Nutrients | 27757 | 0.01 | 0.01 | 1.46E-01 | 2.38E-01 | 166 | 27040 | 0.01 | 0.01 | 3.72E-01 | 4.66E-01 | 220 | 0.01 | 0.00 | 9.64E-02 | 0.00 | 7.03E-01 | - | - | - |
| Possibility to speak with colleagues during breaks | Psychosocial | 29731 | -0.01 | 0.01 | 3.39E-02 | 8.25E-02 | 111 | 29109 | 0.00 | 0.01 | 8.45E-01 | 8.83E-01 | 265 | -0.01 | 0.00 | 1.01E-01 | 0.46 | 1.75E-01 | - | - | - |
| Linoleic acid intake (g/day) | Nutrients | 27757 | 0.01 | 0.01 | 1.43E-01 | 2.35E-01 | 165 | 27040 | 0.00 | 0.01 | 4.03E-01 | 4.92E-01 | 227 | 0.01 | 0.00 | 1.03E-01 | 0.00 | 6.65E-01 | - | - | - |
| Possibility to leave your work for a while to speak with a colleague | Psychosocial | 29548 | 0.01 | 0.01 | 2.06E-01 | 3.07E-01 | 180 | 28927 | 0.01 | 0.01 | 3.45E-01 | 4.45E-01 | 215 | 0.01 | 0.00 | 1.18E-01 | 0.00 | 8.26E-01 | - | - | - |
| Pizza | Food | 27757 | 0.00 | 0.01 | 8.61E-01 | 9.22E-01 | 252 | 27040 | -0.01 | 0.01 | 4.46E-02 | 8.69E-02 | 141 | -0.01 | 0.00 | 1.24E-01 | 0.41 | 1.92E-01 | - | - | - |
| For how much of the time during the last four weeks have you felt so depressed that nothing could cheer you up? | General health | 14265 | 0.01 | 0.01 | 1.60E-01 | 2.55E-01 | 169 | 14067 | 0.01 | 0.01 | 4.56E-01 | 5.37E-01 | 235 | 0.01 | 0.00 | 1.28E-01 | 0.00 | 6.43E-01 | - | - | - |
| Lean fish (e.g. perch, bass, cod) | Food | 27757 | 0.00 | 0.01 | 7.87E-01 | 8.53E-01 | 249 | 27040 | 0.01 | 0.01 | 1.43E-02 | **3.51E-02** | 113 | 0.01 | 0.00 | 1.31E-01 | 0.74 | 5.15E-02 | - | - | - |
| Light but partly physically active work | Physical activity | 29412 | -0.02 | 0.01 | 1.97E-01 | 2.99E-01 | 178 | 28750 | -0.01 | 0.01 | 4.01E-01 | 4.91E-01 | 226 | -0.02 | 0.01 | 1.32E-01 | 0.00 | 7.57E-01 | - | - | - |
| Worsen in health in the future | General health | 14191 | 0.01 | 0.01 | 4.20E-01 | 5.48E-01 | 207 | 14000 | 0.01 | 0.01 | 1.92E-01 | 2.89E-01 | 184 | 0.01 | 0.00 | 1.35E-01 | 0.00 | 7.21E-01 | - | - | - |
| For how much of the time during the last four weeks have you felt happy? | General health | 14254 | -0.01 | 0.01 | 2.74E-01 | 3.93E-01 | 188 | 14046 | -0.01 | 0.01 | 3.15E-01 | 4.32E-01 | 202 | -0.01 | 0.01 | 1.38E-01 | 0.00 | 9.50E-01 | - | - | - |

| | | | | | | | | | | | | | | | | | | | | | |
|---|---|---|---|---|---|---|---|---|---|---|---|---|---|---|---|---|---|---|---|---|---|
| Brown beans, pea soup | Food | 27757 | -0.01 | 0.01 | 1.24E-01 | 2.10E-01 | 160 | 27040 | 0.00 | 0.01 | 6.15E-01 | 6.95E-01 | 245 | -0.01 | 0.00 | 1.47E-01 | 0.00 | 4.75E-01 | - | - | - |
| Vitamin B12 intake (ug/day) | Nutrients | 27757 | 0.00 | 0.01 | 9.49E-01 | 9.82E-01 | 261 | 27040 | 0.01 | 0.01 | 3.34E-02 | 7.07E-02 | 131 | 0.01 | 0.00 | 1.48E-01 | 0.59 | 1.19E-01 | - | - | - |
| Magnesium intake (mg/day) | Nutrients | 27757 | 0.00 | 0.01 | 5.34E-01 | 6.54E-01 | 220 | 27040 | -0.01 | 0.01 | 1.53E-01 | 2.40E-01 | 176 | -0.01 | 0.00 | 1.48E-01 | 0.00 | 5.59E-01 | - | - | - |
| Travel to work: Walk to work vs passive travel to work | Physical activity | 19510 | 0.01 | 0.02 | 5.10E-01 | 6.35E-01 | 217 | 19248 | 0.03 | 0.02 | 1.75E-01 | 2.68E-01 | 181 | 0.02 | 0.02 | 1.53E-01 | 0.00 | 6.28E-01 | - | - | - |
| Enterodiol intake (ug/day) | Nutrients | 27757 | -0.01 | 0.01 | 1.86E-01 | 2.85E-01 | 176 | 27040 | 0.00 | 0.01 | 4.90E-01 | 5.68E-01 | 239 | -0.01 | 0.00 | 1.54E-01 | 0.00 | 6.63E-01 | - | - | - |
| Mood status | Psychosocial | 24261 | 0.01 | 0.01 | 1.18E-01 | 2.01E-01 | 157 | 23834 | 0.00 | 0.01 | 6.56E-01 | 7.32E-01 | 248 | 0.01 | 0.00 | 1.55E-01 | 0.00 | 4.28E-01 | - | - | - |
| Vitamin A intake (mg/day) | Nutrients | 27757 | 0.00 | 0.01 | 6.32E-01 | 7.35E-01 | 232 | 27040 | 0.01 | 0.01 | 1.34E-01 | 2.12E-01 | 175 | 0.01 | 0.00 | 1.63E-01 | 0.00 | 4.67E-01 | - | - | - |
| Whole grain intake (g/day) | Food | 27757 | 0.00 | 0.01 | 9.92E-01 | 9.96E-01 | 268 | 27040 | -0.01 | 0.01 | 5.27E-02 | 1.01E-01 | 145 | -0.01 | 0.00 | 1.72E-01 | 0.47 | 1.69E-01 | - | - | - |
| Satisfaction with accomodation | Psychosocial | 24297 | 0.00 | 0.01 | 8.98E-01 | 9.48E-01 | 256 | 23880 | 0.01 | 0.01 | 7.30E-02 | 1.28E-01 | 158 | 0.01 | 0.00 | 1.74E-01 | 0.28 | 2.39E-01 | - | - | - |
| Sodas, soft drinks, juice | Beverage | 27757 | 0.01 | 0.01 | 7.47E-02 | 1.47E-01 | 137 | 27040 | 0.00 | 0.01 | 9.13E-01 | 9.40E-01 | 269 | 0.01 | 0.00 | 1.76E-01 | 0.27 | 2.43E-01 | - | - | - |
| Physical limitation that limited your ability to perform certain work tasks or other activities during the last four weeks | General health | 14243 | 0.03 | 0.02 | 2.98E-01 | 4.20E-01 | 191 | 14027 | 0.02 | 0.02 | 4.09E-01 | 4.97E-01 | 228 | 0.02 | 0.02 | 1.87E-01 | 0.00 | 8.81E-01 | - | - | - |
| Physical limitation to participate in moderately demanding activities: walking more than 2 km | General health | 14265 | 0.00 | 0.01 | 6.64E-01 | 7.54E-01 | 238 | 14069 | 0.01 | 0.01 | 1.57E-01 | 2.44E-01 | 178 | 0.01 | 0.01 | 1.91E-01 | 0.00 | 4.84E-01 | - | - | - |
| Iodine intake (ug/day) | Nutrients | 27757 | 0.00 | 0.01 | 5.88E-01 | 7.02E-01 | 226 | 27040 | 0.01 | 0.01 | 2.39E-01 | 3.43E-01 | 193 | 0.01 | 0.00 | 2.25E-01 | 0.00 | 6.47E-01 | - | - | - |
| Vision status | General health | 24272 | -0.01 | 0.01 | 4.11E-01 | 5.41E-01 | 205 | 23828 | -0.01 | 0.01 | 3.94E-01 | 4.85E-01 | 225 | -0.01 | 0.00 | 2.36E-01 | 0.00 | 9.83E-01 | - | - | - |
| For how much of the time during the last four weeks has your physical health or your emotional problems limited your ability to interact with others? | General health | 14184 | 0.01 | 0.01 | 4.97E-01 | 6.23E-01 | 215 | 13978 | 0.01 | 0.01 | 3.40E-01 | 4.42E-01 | 211 | 0.01 | 0.01 | 2.48E-01 | 0.00 | 8.45E-01 | - | - | - |
| Sum of phytosterols intake (mg/day) | Nutrients | 27757 | 0.01 | 0.01 | 2.33E-01 | 3.42E-01 | 184 | 27040 | 0.00 | 0.01 | 6.64E-01 | 7.33E-01 | 251 | 0.00 | 0.00 | 2.49E-01 | 0.00 | 5.97E-01 | - | - | - |
| For how much of the time during the last four weeks have you felt very nervous? | General health | 14250 | 0.01 | 0.01 | 1.43E-01 | 2.35E-01 | 164 | 14053 | 0.00 | 0.01 | 8.80E-01 | 9.13E-01 | 267 | 0.01 | 0.01 | 2.51E-01 | 0.00 | 3.55E-01 | - | - | - |
| Selenium intake (ug/day) | Nutrients | 27757 | 0.00 | 0.01 | 8.98E-01 | 9.48E-01 | 255 | 27040 | 0.01 | 0.01 | 7.79E-02 | 1.34E-01 | 161 | 0.00 | 0.00 | 2.53E-01 | 0.45 | 1.77E-01 | - | - | - |
| Physical limitation that made you not being able to perform certain work tasks or other activities during the last four weeks | General health | 14214 | -0.01 | 0.02 | 7.43E-01 | 8.23E-01 | 244 | 14019 | 0.04 | 0.02 | 5.45E-02 | 1.03E-01 | 147 | 0.02 | 0.02 | 2.57E-01 | 0.61 | 1.11E-01 | - | - | - |
| Participation in other association | Social | 10603 | 0.01 | 0.02 | 6.20E-01 | 7.24E-01 | 231 | 10245 | -0.04 | 0.02 | 3.60E-02 | 7.33E-02 | 136 | -0.02 | 0.01 | 2.62E-01 | 0.71 | 6.54E-02 | - | - | - |
| Parents or siblings have diabetes | General health | 29781 | 0.02 | 0.02 | 1.96E-01 | 2.98E-01 | 177 | 29132 | -0.05 | 0.02 | 4.18E-03 | **1.23E-02** | 94 | -0.01 | 0.01 | 2.62E-01 | 0.88 | 3.32E-03 | - | - | - |
| People to ask for help apart from the ones at home | Social | 30220 | 0.02 | 0.02 | 3.50E-01 | 4.80E-01 | 197 | 29534 | 0.01 | 0.02 | 5.59E-01 | 6.40E-01 | 242 | 0.01 | 0.01 | 2.84E-01 | 0.00 | 8.01E-01 | - | - | - |
| For how much of the time during the last four weeks have you felt gloomy and sad? | General health | 14235 | 0.00 | 0.01 | 6.60E-01 | 7.52E-01 | 237 | 14041 | 0.02 | 0.01 | 5.55E-02 | 1.04E-01 | 148 | 0.01 | 0.01 | 2.97E-01 | 0.64 | 9.48E-02 | - | - | - |
| Frequency of walking during leisure time | Physical activity | 29233 | 0.00 | 0.01 | 7.78E-01 | 8.53E-01 | 246 | 28569 | 0.01 | 0.01 | 8.23E-02 | 1.39E-01 | 163 | 0.00 | 0.00 | 3.06E-01 | 0.51 | 1.52E-01 | - | - | - |
| Potassium intake (mg/day) | Nutrients | 27757 | 0.00 | 0.01 | 6.43E-01 | 7.36E-01 | 236 | 27040 | -0.01 | 0.01 | 3.30E-01 | 4.42E-01 | 205 | 0.00 | 0.00 | 3.11E-01 | 0.00 | 7.13E-01 | - | - | - |
| Learn new things at job | Psychosocial | 29793 | 0.00 | 0.01 | 7.84E-01 | 8.53E-01 | 248 | 29144 | 0.01 | 0.01 | 2.58E-01 | 3.68E-01 | 194 | 0.00 | 0.00 | 3.20E-01 | 0.00 | 5.44E-01 | - | - | - |
| Support from others | Social | 30320 | 0.00 | 0.01 | 6.42E-01 | 7.36E-01 | 234 | 29635 | 0.01 | 0.01 | 7.09E-02 | 1.25E-01 | 157 | 0.00 | 0.00 | 3.43E-01 | 0.61 | 1.08E-01 | - | - | - |
| Animal based protein intake (g/day) | Nutrients | 27757 | 0.00 | 0.01 | 9.88E-01 | 9.96E-01 | 266 | 27040 | 0.01 | 0.01 | 1.96E-01 | 2.93E-01 | 185 | 0.00 | 0.00 | 3.57E-01 | 0.00 | 3.64E-01 | - | - | - |
| Lariciresinol intake (ug/day) | Nutrients | 27757 | 0.00 | 0.01 | 7.89E-01 | 8.53E-01 | 250 | 27040 | -0.01 | 0.01 | 3.40E-01 | 4.42E-01 | 212 | 0.00 | 0.00 | 3.89E-01 | 0.00 | 6.23E-01 | - | - | - |
| Close relationship with anyone | Social | 30277 | 0.01 | 0.01 | 1.17E-01 | 2.01E-01 | 156 | 29586 | 0.00 | 0.01 | 7.31E-01 | 7.85E-01 | 258 | 0.00 | 0.00 | 3.92E-01 | 0.46 | 1.76E-01 | - | - | - |
| Control over planning and execution of the workday | Psychosocial | 29878 | 0.01 | 0.01 | 1.02E-01 | 1.81E-01 | 152 | 29216 | 0.00 | 0.01 | 6.55E-01 | 7.32E-01 | 247 | 0.00 | 0.00 | 4.01E-01 | 0.54 | 1.40E-01 | - | - | - |
| For how much of the time during the last four weeks have you felt tired? | General health | 14269 | 0.00 | 0.01 | 8.57E-01 | 9.22E-01 | 251 | 14055 | 0.01 | 0.01 | 3.57E-01 | 4.53E-01 | 218 | 0.00 | 0.01 | 4.36E-01 | 0.00 | 5.99E-01 | - | - | - |
| Sum of Lariciresinol, Matairesinol, Pinoresinol, Secoisolariciresinol intake (ug/day) | Nutrients | 27757 | 0.00 | 0.01 | 7.31E-01 | 8.12E-01 | 243 | 27040 | 0.00 | 0.01 | 4.54E-01 | 5.37E-01 | 234 | 0.00 | 0.00 | 4.41E-01 | 0.00 | 7.71E-01 | - | - | - |
| Number of snuff boxes per week | Tobacco use | 29321 | -0.01 | 0.01 | 6.40E-02 | 1.30E-01 | 132 | 28690 | 0.01 | 0.01 | 4.35E-01 | 5.20E-01 | 231 | 0.00 | 0.00 | 4.47E-01 | 0.71 | 6.28E-02 | - | - | - |
| Control over own work assignment | Psychosocial | 29856 | 0.00 | 0.01 | 7.52E-01 | 8.29E-01 | 245 | 29214 | 0.00 | 0.01 | 4.61E-01 | 5.41E-01 | 236 | 0.00 | 0.00 | 4.56E-01 | 0.00 | 7.66E-01 | - | - | - |
| Repetitive job | Psychosocial | 29805 | 0.00 | 0.01 | 4.92E-01 | 6.20E-01 | 214 | 29164 | 0.01 | 0.01 | 8.27E-02 | 1.39E-01 | 164 | 0.00 | 0.00 | 4.59E-01 | 0.66 | 8.67E-02 | - | - | - |
| Participation in study circles | Social | 10603 | -0.01 | 0.03 | 7.26E-01 | 8.10E-01 | 242 | 10245 | -0.02 | 0.03 | 5.19E-01 | 5.97E-01 | 241 | -0.01 | 0.02 | 4.82E-01 | 0.00 | 8.32E-01 | - | - | - |
| Beta-sitosterol intake (mg/day) | Nutrients | 27757 | 0.00 | 0.01 | 9.66E-01 | 9.92E-01 | 263 | 27040 | -0.01 | 0.01 | 3.40E-01 | 4.42E-01 | 213 | 0.00 | 0.00 | 4.84E-01 | 0.00 | 5.17E-01 | - | - | - |
| Satisfaction with leisure time | Psychosocial | 24243 | 0.00 | 0.01 | 5.67E-01 | 6.83E-01 | 224 | 23819 | 0.00 | 0.01 | 6.93E-01 | 7.56E-01 | 254 | 0.00 | 0.00 | 4.94E-01 | 0.00 | 9.00E-01 | - | - | - |
| Feel uneasy or guilty because of your way of drinking | Alcohol | 17149 | -0.05 | 0.03 | 7.90E-02 | 1.53E-01 | 139 | 16775 | 0.02 | 0.03 | 4.33E-01 | 5.20E-01 | 229 | -0.01 | 0.02 | 5.02E-01 | 0.69 | 7.13E-02 | - | - | - |
| Everyday exercise satisfaction | Physical activity | 18991 | 0.00 | 0.01 | 5.93E-01 | 7.06E-01 | 227 | 18458 | 0.00 | 0.01 | 6.88E-01 | 7.54E-01 | 253 | 0.00 | 0.01 | 5.08E-01 | 0.00 | 9.24E-01 | - | - | - |
| Extent to what your physical and emotional health disrupted your usual social life during the last four weeks | General health | 14270 | 0.00 | 0.01 | 9.11E-01 | 9.57E-01 | 257 | 14073 | 0.01 | 0.01 | 2.99E-01 | 4.14E-01 | 200 | 0.00 | 0.01 | 5.13E-01 | 0.00 | 4.15E-01 | - | - | - |
| Sausage, liver pate on bread | Food | 27757 | 0.00 | 0.01 | 4.15E-01 | 5.44E-01 | 206 | 27040 | 0.00 | 0.01 | 9.44E-01 | 9.55E-01 | 274 | 0.00 | 0.00 | 5.29E-01 | 0.00 | 6.01E-01 | - | - | - |
| Ingenuity or creativity demand from job | Psychosocial | 29713 | 0.00 | 0.01 | 5.32E-01 | 6.54E-01 | 219 | 29074 | -0.01 | 0.01 | 1.31E-01 | 2.09E-01 | 173 | 0.00 | 0.00 | 5.34E-01 | 0.56 | 1.30E-01 | - | - | - |
| Tiamin intake (mg/day) | Nutrients | 27757 | 0.00 | 0.01 | 6.04E-01 | 7.15E-01 | 228 | 27040 | 0.00 | 0.01 | 7.27E-01 | 7.85E-01 | 256 | 0.00 | 0.00 | 5.39E-01 | 0.00 | 9.09E-01 | - | - | - |
| Cookies, pastry | Food | 27757 | 0.01 | 0.01 | 3.71E-01 | 4.94E-01 | 203 | 27040 | 0.00 | 0.01 | 9.52E-01 | 9.59E-01 | 275 | 0.00 | 0.00 | 5.54E-01 | 0.00 | 5.01E-01 | - | - | - |
| Satisfaction with economy | Psychosocial | 24277 | 0.00 | 0.01 | 9.90E-01 | 9.96E-01 | 267 | 23847 | -0.01 | 0.01 | 4.35E-01 | 5.20E-01 | 232 | 0.00 | 0.00 | 5.75E-01 | 0.00 | 5.87E-01 | - | - | - |
| Get sick more often than other people | General health | 14241 | 0.00 | 0.01 | 9.97E-01 | 9.97E-01 | 270 | 14042 | -0.01 | 0.01 | 4.34E-01 | 5.20E-01 | 230 | 0.00 | 0.01 | 5.83E-01 | 0.00 | 5.76E-01 | - | - | - |
| White meat (poultry) | Food | 27757 | 0.00 | 0.01 | 4.31E-01 | 5.54E-01 | 210 | 27040 | 0.00 | 0.01 | 9.87E-01 | 9.87E-01 | 277 | 0.00 | 0.00 | 5.83E-01 | 0.00 | 5.72E-01 | - | - | - |
| Hearing status | General health | 24296 | 0.00 | 0.01 | 6.43E-01 | 7.36E-01 | 235 | 23868 | 0.00 | 0.01 | 7.69E-01 | 8.16E-01 | 261 | 0.00 | 0.00 | 5.92E-01 | 0.00 | 9.02E-01 | - | - | - |
| Long-term sickness | General health | 29453 | -0.02 | 0.02 | 2.53E-01 | 3.66E-01 | 186 | 28749 | 0.01 | 0.02 | 6.61E-01 | 7.32E-01 | 250 | -0.01 | 0.01 | 6.17E-01 | 0.20 | 2.64E-01 | - | - | - |
| Work shifts/weekends | Psychosocial | 29454 | -0.01 | 0.01 | 3.69E-01 | 4.93E-01 | 202 | 28866 | 0.00 | 0.01 | 8.11E-01 | 8.54E-01 | 263 | 0.00 | 0.01 | 6.44E-01 | 0.00 | 4.20E-01 | - | - | - |
| Receive hugs to comfort and support you | Social | 30081 | 0.01 | 0.02 | 4.99E-01 | 6.23E-01 | 216 | 29388 | 0.00 | 0.02 | 9.68E-01 | 9.71E-01 | 276 | 0.01 | 0.01 | 6.52E-01 | 0.00 | 6.13E-01 | - | - | - |
| Ice cream | Food | 27757 | 0.00 | 0.01 | 4.28E-01 | 5.53E-01 | 209 | 27040 | -0.01 | 0.01 | 1.62E-01 | 2.51E-01 | 179 | 0.00 | 0.00 | 6.72E-01 | 0.58 | 1.21E-01 | - | - | - |
| Meat on bread | Food | 27757 | -0.01 | 0.01 | 3.29E-01 | 4.56E-01 | 195 | 27040 | 0.00 | 0.01 | 6.82E-01 | 7.50E-01 | 252 | 0.00 | 0.00 | 6.84E-01 | 0.00 | 3.29E-01 | - | - | - |
| Disaccharides intake (g/day) | Nutrients | 27757 | 0.00 | 0.01 | 6.34E-01 | 7.35E-01 | 233 | 27040 | -0.01 | 0.01 | 3.02E-01 | 4.17E-01 | 201 | 0.00 | 0.00 | 7.01E-01 | 0.12 | 2.85E-01 | - | - | - |
| Cheese 28% | Food | 27757 | 0.00 | 0.01 | 6.78E-01 | 7.63E-01 | 240 | 27040 | 0.00 | 0.01 | 9.28E-01 | 9.43E-01 | 272 | 0.00 | 0.00 | 7.19E-01 | 0.00 | 8.21E-01 | - | - | - |
| Participation in sports or physical exercise associations | Social | 10603 | 0.00 | 0.02 | 8.87E-01 | 9.47E-01 | 253 | 10245 | 0.01 | 0.02 | 7.29E-01 | 7.85E-01 | 257 | 0.00 | 0.01 | 7.31E-01 | 0.00 | 8.83E-01 | - | - | - |
| Emotional problems that made you do less than you wanted during the last four weeks | General health | 14287 | 0.04 | 0.02 | 1.56E-01 | 2.51E-01 | 168 | 14074 | -0.02 | 0.03 | 3.38E-01 | 4.42E-01 | 210 | 0.01 | 0.02 | 7.40E-01 | 0.65 | 9.29E-02 | - | - | - |
| Matairesinol intake (ug/day) | Nutrients | 27757 | 0.00 | 0.01 | 7.24E-01 | 8.10E-01 | 241 | 27040 | 0.00 | 0.01 | 9.30E-01 | 9.43E-01 | 273 | 0.00 | 0.00 | 7.54E-01 | 0.00 | 8.54E-01 | - | - | - |
| Phosphate intake (mg/day) | Nutrients | 27757 | 0.00 | 0.01 | 8.93E-01 | 9.48E-01 | 254 | 27040 | 0.00 | 0.01 | 5.72E-01 | 6.52E-01 | 243 | 0.00 | 0.00 | 7.63E-01 | 0.00 | 6.20E-01 | - | - | - |
| Oil for cooking | Food | 27757 | 0.00 | 0.01 | 9.24E-01 | 9.63E-01 | 258 | 27040 | 0.00 | 0.01 | 6.07E-01 | 6.89E-01 | 244 | 0.00 | 0.00 | 7.68E-01 | 0.00 | 6.65E-01 | - | - | - |
| Smoking status: Former occasional smokers vs non-smokers | Tobacco use | 18070 | 0.00 | 0.02 | 9.27E-01 | 9.63E-01 | 260 | 17715 | 0.01 | 0.02 | 7.93E-01 | 8.38E-01 | 262 | 0.00 | 0.02 | 8.01E-01 | 0.00 | 9.06E-01 | - | - | - |
| Smoked fish/meat | Food | 27757 | 0.00 | 0.01 | 9.85E-01 | 9.96E-01 | 265 | 27040 | 0.00 | 0.01 | 7.37E-01 | 7.88E-01 | 259 | 0.00 | 0.00 | 8.25E-01 | 0.00 | 8.00E-01 | - | - | - |
| Patience status | Psychosocial | 24260 | 0.00 | 0.01 | 6.12E-01 | 7.19E-01 | 230 | 23841 | 0.00 | 0.01 | 4.54E-01 | 5.37E-01 | 233 | 0.00 | 0.00 | 8.63E-01 | 0.00 | 3.75E-01 | - | - | - |
| Marital status: Single vs Widow/widower | Social | 3194 | 0.00 | 0.07 | 9.62E-01 | 9.92E-01 | 262 | 3096 | 0.01 | 0.07 | 8.44E-01 | 8.83E-01 | 264 | 0.01 | 0.05 | 8.65E-01 | 0.00 | 9.14E-01 | - | - | - |
| Travel to work: Irregular travel mode to work vs passive travel to work | Physical activity | 19540 | -0.02 | 0.02 | 3.58E-01 | 4.84E-01 | 200 | 19202 | 0.01 | 0.02 | 4.89E-01 | 5.68E-01 | 238 | 0.00 | 0.02 | 8.79E-01 | 0.23 | 2.54E-01 | - | - | - |
| Number of social interactions during a normal week | Social | 30316 | 0.00 | 0.01 | 6.10E-01 | 7.19E-01 | 229 | 29621 | 0.00 | 0.01 | 7.64E-01 | 8.14E-01 | 260 | 0.00 | 0.00 | 8.80E-01 | 0.00 | 5.67E-01 | - | - | - |
| Confidence status | Psychosocial | 24253 | -0.01 | 0.01 | 2.87E-01 | 4.11E-01 | 189 | 23818 | 0.01 | 0.01 | 3.88E-01 | 4.82E-01 | 223 | 0.00 | 0.00 | 8.88E-01 | 0.46 | 1.73E-01 | - | - | - |
| Light and physically active work | Physical activity | 29412 | 0.01 | 0.01 | 4.47E-01 | 5.69E-01 | 212 | 28750 | -0.01 | 0.01 | 3.33E-01 | 4.42E-01 | 207 | 0.00 | 0.01 | 8.88E-01 | 0.33 | 2.21E-01 | - | - | - |
| Whole grain soft bread | Food | 27757 | 0.00 | 0.01 | 5.87E-01 | 7.02E-01 | 225 | 27040 | 0.00 | 0.01 | 7.17E-01 | 7.79E-01 | 255 | 0.00 | 0.00 | 8.93E-01 | 0.00 | 5.23E-01 | - | - | - |
| Physical limitation to participate in moderately demanding activities: bending down or kneeling | General health | 14245 | 0.00 | 0.01 | 9.78E-01 | 9.96E-01 | 264 | 14049 | 0.00 | 0.01 | 9.24E-01 | 9.43E-01 | 271 | 0.00 | 0.01 | 9.31E-01 | 0.00 | 9.61E-01 | - | - | - |
| Job demands to work very fast | Psychosocial | 29850 | -0.01 | 0.01 | 3.46E-01 | 4.76E-01 | 196 | 29223 | 0.01 | 0.01 | 2.97E-01 | 4.14E-01 | 199 | 0.00 | 0.00 | 9.43E-01 | 0.49 | 1.60E-01 | - | - | - |
| Hamburger | Food | 27757 | 0.00 | 0.01 | 9.92E-01 | 9.96E-01 | 269 | 27040 | 0.00 | 0.01 | 9.17E-01 | 9.41E-01 | 270 | 0.00 | 0.00 | 9.47E-01 | 0.00 | 9.35E-01 | - | - | - |
| Do you feel important and appreciated outside your home? | Psychosocial | 24287 | 0.00 | 0.01 | 9.24E-01 | 9.63E-01 | 259 | 23872 | 0.00 | 0.01 | 9.09E-01 | 9.40E-01 | 268 | 0.00 | 0.00 | 9.89E-01 | 0.00 | 8.83E-01 | - | - | - |
| Coffee rolls/buns, rusk | Food | 27757 | 0.01 | 0.01 | 3.56E-01 | 4.84E-01 | 199 | 27040 | -0.01 | 0.01 | 3.55E-01 | 4.53E-01 | 217 | 0.00 | 0.00 | 9.95E-01 | 0.41 | 1.91E-01 | - | - | - |
| Amount of alcohol drunk in a day | Alcohol | 10825 | 0.04 | 0.01 | - | - | 274 | 10621 | 0.05 | 0.01 | 1.11E-07 | **8.76E-07** | 35 | 0.05 | 0.01 | - | 0.03 | 3.09E-01 | - | - | - |
| Snore during sleep | Sleep | 10436 | 0.03 | 0.01 | - | - | 283 | 10318 | 0.04 | 0.01 | 5.27E-04 | **1.89E-03** | 77 | 0.04 | 0.01 | - | 0.00 | 9.49E-01 | - | - | - |
| Soft cheese | Food | 5467 | 0.03 | 0.01 | 9.86E-02 | 1.77E-01 | 150 | 5262 | -0.06 | 0.02 | 2.26E-02 | 5.31E-02 | 118 | -0.01 | 0.01 | - | 0.93 | 1.03E-04 | - | - | - |
| Times during last year that you felt guilty because of your drinking | Alcohol | 11016 | 0.01 | 0.01 | - | - | 276 | 10805 | 0.02 | 0.01 | 4.20E-02 | 8.36E-02 | 139 | 0.01 | 0.01 | - | 0.00 | 4.51E-01 | - | - | - |
| Blota (broth + bread) | Food | 5467 | 0.03 | 0.01 | 8.60E-02 | 1.60E-01 | 145 | 5262 | 0.04 | 0.02 | 6.05E-02 | 1.10E-01 | 151 | 0.03 | 0.01 | - | 0.00 | 4.95E-01 | - | - | - |
| Breath-holds during sleep | Sleep | 7835 | 0.02 | 0.01 | - | - | 284 | 7671 | 0.02 | 0.01 | 6.89E-02 | 1.22E-01 | 156 | 0.02 | 0.01 | - | 0.00 | 8.50E-01 | - | - | - |
| Risk of sleeping while sitting still after having lunch | Sleep | 11155 | -0.01 | 0.01 | - | - | 282 | 11016 | -0.01 | 0.01 | 1.24E-01 | 2.00E-01 | 172 | -0.01 | 0.01 | - | 0.00 | 6.34E-01 | - | - | - |
| Risk of sleeping while lying down resting in the afternoon | Sleep | 11147 | -0.01 | 0.01 | - | - | 281 | 11013 | -0.01 | 0.01 | 1.99E-01 | 2.97E-01 | 186 | -0.01 | 0.01 | - | 0.00 | 6.43E-01 | - | - | - |
| Mixed frozen vegetables | Food | 5467 | 0.01 | 0.01 | 5.27E-01 | 6.53E-01 | 218 | 5262 | -0.02 | 0.01 | 2.16E-01 | 3.16E-01 | 189 | -0.01 | 0.01 | - | 0.56 | 1.33E-01 | - | - | - |
| Soft whey cheese | Food | 5467 | -0.03 | 0.01 | 4.96E-02 | 1.07E-01 | 125 | 5262 | -0.02 | 0.02 | 2.18E-01 | 3.17E-01 | 190 | -0.03 | 0.01 | - | 0.00 | 6.35E-01 | - | - | - |
| Risk of sleeping while sitting and reading | Sleep | 11070 | -0.02 | 0.01 | - | - | 277 | 10926 | -0.01 | 0.01 | 2.23E-01 | 3.23E-01 | 191 | -0.02 | 0.01 | - | 0.00 | 4.63E-01 | - | - | - |
| Mashed potato | Food | 5467 | -0.01 | 0.01 | 5.52E-01 | 6.69E-01 | 223 | 5262 | 0.02 | 0.02 | 3.30E-01 | 4.42E-01 | 206 | 0.00 | 0.01 | - | 0.34 | 2.19E-01 | - | - | - |
| Potato salad | Food | 5467 | 0.01 | 0.01 | 3.64E-01 | 4.89E-01 | 201 | 5262 | 0.02 | 0.02 | 3.69E-01 | 4.66E-01 | 219 | 0.01 | 0.01 | - | 0.00 | 9.47E-01 | - | - | - |
| Shellfish (e.g. shrimps, scallops) | Food | 5467 | -0.01 | 0.01 | 6.78E-01 | 7.63E-01 | 239 | 5262 | -0.01 | 0.01 | 3.78E-01 | 4.72E-01 | 222 | -0.01 | 0.01 | - | 0.00 | 6.92E-01 | - | - | - |
| Blood based food | Food | 5467 | -0.03 | 0.02 | 6.08E-02 | 1.25E-01 | 131 | 5262 | -0.01 | 0.02 | 6.60E-01 | 7.32E-01 | 249 | -0.02 | 0.01 | - | 0.22 | 2.58E-01 | - | - | - |
| Liver, kidney | Food | 5467 | -0.03 | 0.02 | 5.19E-02 | 1.11E-01 | 126 | 5262 | 0.00 | 0.02 | 8.54E-01 | 8.89E-01 | 266 | -0.02 | 0.01 | - | 0.64 | 9.72E-02 | - | - | - |
| Time spent in a week in moderately steneous activities | Physical activity | 10952 | -0.02 | 0.01 | - | - | 271 | 10752 | -0.01 | 0.01 | - | - | 278 | -0.02 | 0.01 | - | 0.00 | 6.59E-01 | - | - | - |
| Amount of exercise during the last 12 months | Physical activity | 11290 | -0.03 | 0.01 | - | - | 272 | 11106 | -0.02 | 0.01 | - | - | 279 | -0.03 | 0.01 | - | 0.00 | 5.53E-01 | - | - | - |
| Frequency of alcohol consumption | Alcohol | 11334 | 0.03 | 0.01 | - | - | 273 | 11181 | 0.06 | 0.01 | - | - | 280 | 0.04 | 0.01 | - | 0.65 | 8.97E-02 | - | - | - |

| | | | | | | | | | | | | | | | | | | | | | |
|---|---|---|---|---|---|---|---|---|---|---|---|---|---|---|---|---|---|---|---|---|---|
| Frequency of drinking six or more glasses at the same occasion | Alcohol | 11029 | 0.06 | 0.01 | - | - | 275 | 10832 | 0.07 | 0.01 | - | - | 281 | 0.06 | 0.01 | - | 0.00 | 4.18E-01 | - | - | - |
| Risk of sleeping while watching TV | Sleep | 11192 | 0.01 | 0.01 | - | - | 278 | 11040 | 0.00 | 0.01 | - | - | 282 | 0.01 | 0.01 | - | 0.00 | 4.34E-01 | - | - | - |
| Risk of sleeping while sitting inactive in a public place | Sleep | 11118 | -0.04 | 0.01 | - | - | 279 | 10970 | -0.02 | 0.01 | - | - | 283 | -0.03 | 0.01 | - | 0.00 | 3.60E-01 | - | - | - |
| Risk of sleeping as a passenger in a car for one hour without break | Sleep | 11078 | -0.02 | 0.01 | - | - | 280 | 10924 | -0.02 | 0.01 | - | - | 284 | -0.02 | 0.01 | - | 0.00 | 9.24E-01 | - | - | - |

**Supplementary Table 7**. Linear mixed model association results for triglycerides

| Description | Group | Training set | | | | | | Testing set | | | | | | Metanalysis | | | | | R2 | | |
|---|---|---|---|---|---|---|---|---|---|---|---|---|---|---|---|---|---|---|---|---|---|
| | | N | Effect estimate | S.E. | p-value | p-value$_{FDR}$ | p-value rank | N | Effect estimate | S.E. | p-value | p-value$_{FDR}$ | p-value rank | Effect estimate | S.E. | p-value | $I^2$ | Q p-value | marginal R2 | conditional R2 | R2 rank |
| Fitness status | Physical activity | 22637 | -0.08 | 0.00 | 5.16E-54 | **1.39E-51** | 1 | 22252 | -0.06 | 0.01 | 1.39E-33 | **9.30E-32** | 4 | -0.07 | 0.00 | 5.45E-85 | 0.75 | 4.35E-02 | 0.12 | 0.51 | 2 |
| Smoking status: Smokers vs non-smokers | Tobacco use | 17378 | 0.20 | 0.01 | 2.44E-43 | **3.29E-41** | 2 | 17125 | 0.20 | 0.02 | 1.46E-38 | **1.95E-36** | 2 | 0.20 | 0.01 | 4.28E-80 | 0.00 | 8.46E-01 | 0.12 | 0.50 | 37 |
| Years smoking | Tobacco use | 23963 | 0.07 | 0.01 | 2.82E-40 | **2.53E-38** | 3 | 23359 | 0.07 | 0.01 | 1.40E-38 | **1.95E-36** | 1 | 0.07 | 0.00 | 5.33E-77 | 0.00 | 5.84E-01 | 0.12 | 0.50 | 43 |
| Number of cigarretes smoked per day (in groups) | Tobacco use | 22457 | 0.06 | 0.01 | 1.16E-28 | **6.28E-27** | 5 | 22197 | 0.07 | 0.01 | 2.93E-34 | **2.62E-32** | 3 | 0.06 | 0.00 | 7.53E-61 | 0.36 | 2.13E-01 | 0.12 | 0.49 | 39 |
| Self rate of overall health | General health | 13401 | -0.08 | 0.01 | 6.47E-32 | **4.37E-30** | 4 | 13248 | -0.07 | 0.01 | 2.91E-21 | **1.30E-19** | 6 | -0.08 | 0.00 | 2.24E-51 | 0.61 | 1.11E-01 | 0.12 | 0.55 | 65 |
| Exercise during the last three months | Physical activity | 27108 | -0.05 | 0.00 | 6.40E-28 | **2.88E-26** | 6 | 26546 | -0.03 | 0.00 | 1.35E-13 | **2.78E-12** | 13 | -0.04 | 0.00 | 4.22E-39 | 0.73 | 5.25E-02 | 0.12 | 0.49 | 64 |
| Excellent health | General health | 13347 | -0.07 | 0.01 | 1.79E-22 | **6.90E-21** | 7 | 13187 | -0.06 | 0.01 | 6.30E-17 | **1.95E-15** | 8 | -0.06 | 0.00 | 9.26E-38 | 0.00 | 4.37E-01 | 0.11 | 0.56 | 82 |
| Energy status | Psychosocial | 22625 | -0.04 | 0.00 | 1.02E-19 | **2.74E-18** | 10 | 22235 | -0.04 | 0.01 | 9.66E-19 | **3.70E-17** | 7 | -0.04 | 0.00 | 7.93E-37 | 0.00 | 9.40E-01 | 0.12 | 0.50 | 38 |
| Informed of having high blood pressure | General health | 27421 | 0.08 | 0.01 | 1.58E-15 | **3.05E-14** | 14 | 26845 | 0.11 | 0.01 | 2.93E-23 | **1.57E-21** | 5 | 0.10 | 0.01 | 2.29E-36 | 0.73 | 5.47E-02 | 0.12 | 0.49 | 59 |
| Overall state of health during the last year | General health | 27414 | -0.04 | 0.00 | 9.00E-22 | **2.70E-20** | 9 | 26840 | -0.04 | 0.00 | 3.52E-16 | **8.57E-15** | 11 | -0.04 | 0.00 | 2.76E-36 | 0.00 | 6.49E-01 | 0.12 | 0.49 | 61 |
| Long-term sickness | General health | 26643 | 0.10 | 0.01 | 1.63E-17 | **3.66E-16** | 12 | 26037 | 0.09 | 0.01 | 2.67E-12 | **4.78E-11** | 15 | 0.09 | 0.01 | 3.59E-28 | 0.00 | 5.49E-01 | 0.12 | 0.50 | 60 |
| Physical limitation to participate in steneous activities: running, lifting heavy objects, taking part in physically demanding sports | General health | 13370 | 0.06 | 0.01 | 7.69E-19 | **1.89E-17** | 11 | 13224 | 0.05 | 0.01 | 1.43E-10 | **1.60E-09** | 24 | 0.06 | 0.00 | 1.62E-27 | 0.65 | 8.95E-02 | 0.11 | 0.55 | 84 |
| For how much of the time during the last four weeks have you felt really alert and strong? | General health | 13340 | -0.06 | 0.01 | 2.85E-17 | **5.93E-16** | 13 | 13169 | -0.05 | 0.01 | 9.76E-12 | **1.45E-10** | 18 | -0.05 | 0.00 | 2.55E-27 | 0.15 | 2.77E-01 | 0.11 | 0.56 | 83 |
| Sleep status | Sleep | 22667 | -0.03 | 0.00 | 5.20E-12 | **6.69E-11** | 21 | 22296 | -0.04 | 0.01 | 6.56E-17 | **1.95E-15** | 9 | -0.04 | 0.00 | 4.95E-27 | 0.35 | 2.17E-01 | 0.12 | 0.51 | 41 |
| Cambridge physical activity index | Physical activity | 26213 | -0.04 | 0.00 | 5.30E-22 | **1.79E-20** | 8 | 25679 | -0.03 | 0.00 | 7.22E-08 | **4.50E-07** | 43 | -0.03 | 0.00 | 6.43E-27 | 0.85 | 9.60E-03 | 0.12 | 0.49 | 63 |
| Average portion size of vegetables based on photographic illustration of four sizes (smallest to largest) | Food | 25076 | -0.03 | 0.00 | 2.50E-10 | **2.32E-09** | 29 | 24452 | -0.04 | 0.01 | 1.39E-16 | **3.73E-15** | 10 | -0.04 | 0.00 | 1.07E-24 | 0.68 | 7.69E-02 | 0.12 | 0.50 | 55 |
| Physical limitation to participate in moderately demanding activities: walking up several stairs | General health | 13372 | 0.05 | 0.01 | 2.39E-14 | **3.80E-13** | 17 | 13203 | 0.05 | 0.01 | 3.21E-11 | **4.53E-10** | 19 | 0.05 | 0.00 | 5.11E-24 | 0.00 | 6.14E-01 | 0.11 | 0.55 | 86 |
| Eat breakfast from 2000 | Food | 15989 | -0.16 | 0.02 | 3.85E-15 | **6.93E-14** | 15 | 15634 | -0.13 | 0.02 | 5.01E-10 | **4.97E-09** | 27 | -0.15 | 0.01 | 1.93E-23 | 0.00 | 3.17E-01 | 0.11 | 0.46 | 81 |
| For how much of the time during the last four weeks have you felt full of energy? | General health | 13344 | -0.05 | 0.01 | 6.26E-15 | **1.06E-13** | 16 | 13178 | -0.04 | 0.01 | 1.28E-09 | **1.23E-08** | 28 | -0.05 | 0.00 | 7.51E-23 | 0.28 | 2.39E-01 | 0.11 | 0.55 | 85 |
| Overall state of health compared to other your age | General health | 14005 | -0.04 | 0.01 | 5.76E-12 | **7.07E-11** | 22 | 13611 | -0.05 | 0.01 | 5.57E-12 | **9.33E-11** | 16 | -0.04 | 0.00 | 2.47E-22 | 0.00 | 5.01E-01 | 0.11 | 0.49 | 87 |
| Physical limitation to participate in moderately demanding activities: lifting or carrying grocery bags | General health | 13360 | 0.04 | 0.01 | 3.20E-10 | **2.88E-09** | 30 | 13213 | 0.05 | 0.01 | 2.20E-13 | **4.22E-12** | 14 | 0.05 | 0.00 | 6.21E-22 | 0.06 | 3.02E-01 | 0.11 | 0.56 | 88 |
| How much has the pain during the last four weeks disturbed your normal work? | General health | 13333 | 0.05 | 0.01 | 3.34E-13 | **4.74E-12** | 19 | 13165 | 0.04 | 0.01 | 3.33E-09 | **2.79E-08** | 32 | 0.05 | 0.00 | 8.11E-21 | 0.00 | 3.92E-01 | 0.11 | 0.55 | 95 |
| As healthy as anyone | General health | 13306 | -0.04 | 0.01 | 9.15E-11 | **9.15E-10** | 27 | 13148 | -0.04 | 0.01 | 1.22E-10 | **1.42E-09** | 23 | -0.04 | 0.00 | 6.51E-20 | 0.00 | 8.86E-01 | 0.11 | 0.55 | 93 |
| Physical limitation to participate in moderately demanding activities: moving a table, vacuuming, walking in the forest or gardening | General health | 13366 | 0.04 | 0.01 | 3.52E-10 | **3.06E-09** | 31 | 13218 | 0.05 | 0.01 | 3.60E-11 | **4.80E-10** | 20 | 0.04 | 0.00 | 8.04E-20 | 0.00 | 6.20E-01 | 0.11 | 0.55 | 89 |
| Appetite status | Psychosocial | 22651 | -0.04 | 0.00 | 5.83E-14 | **8.74E-13** | 18 | 22268 | -0.02 | 0.00 | 5.27E-07 | **2.57E-06** | 55 | -0.03 | 0.00 | 5.81E-19 | 0.60 | 1.13E-01 | 0.12 | 0.50 | 42 |
| Physical limitation that reduced the normal time spent at work or in other activities during the last four weeks | General health | 13397 | 0.14 | 0.02 | 3.21E-11 | **3.77E-10** | 23 | 13212 | 0.12 | 0.02 | 2.52E-08 | **1.83E-07** | 37 | 0.13 | 0.01 | 5.31E-18 | 0.00 | 4.78E-01 | 0.11 | 0.55 | 92 |
| Physical limitation to participate in moderately demanding activities: walking more than 2 km | General health | 13379 | 0.04 | 0.01 | 6.80E-11 | **7.06E-10** | 26 | 13207 | 0.04 | 0.01 | 4.47E-08 | **2.92E-07** | 41 | 0.04 | 0.00 | 1.83E-17 | 0.00 | 5.57E-01 | 0.11 | 0.55 | 99 |
| Fibre intake (g/day) | Nutrients | 25076 | -0.02 | 0.00 | 5.25E-06 | **2.12E-05** | 67 | 24452 | -0.04 | 0.01 | 1.30E-13 | **2.78E-12** | 12 | -0.03 | 0.00 | 5.61E-17 | 0.82 | 1.85E-02 | 0.12 | 0.51 | 7 |
| Pain during the last four weeks | General health | 13371 | 0.05 | 0.01 | 2.45E-12 | **3.31E-11** | 20 | 13188 | 0.03 | 0.01 | 2.34E-06 | **1.01E-05** | 61 | 0.04 | 0.00 | 6.69E-17 | 0.56 | 1.30E-01 | 0.11 | 0.55 | 96 |
| Brewed (filtered) coffee | Beverage | 25076 | -0.03 | 0.00 | 8.56E-09 | **6.60E-08** | 35 | 24452 | -0.03 | 0.00 | 1.22E-08 | **9.37E-08** | 35 | -0.03 | 0.00 | 5.73E-16 | 0.00 | 7.96E-01 | 0.12 | 0.51 | 5 |
| For how much of the time during the last four weeks have you felt tired? | General health | 13381 | 0.04 | 0.01 | 6.04E-11 | **6.52E-10** | 25 | 13195 | 0.03 | 0.01 | 3.52E-06 | **1.45E-05** | 65 | 0.04 | 0.00 | 1.96E-15 | 0.39 | 1.99E-01 | 0.11 | 0.55 | 91 |
| Feel the need to reduce alcohol consumption | Alcohol | 25184 | 0.07 | 0.01 | 9.63E-08 | **6.34E-07** | 41 | 24724 | 0.08 | 0.01 | 8.21E-09 | **6.47E-08** | 34 | 0.08 | 0.01 | 4.73E-15 | 0.00 | 5.82E-01 | 0.12 | 0.49 | 67 |
| Folic acid intake (ug/day) | Nutrients | 25076 | -0.02 | 0.00 | 2.22E-06 | **1.05E-05** | 57 | 24452 | -0.03 | 0.01 | 3.12E-10 | **3.22E-09** | 26 | -0.03 | 0.00 | 9.64E-15 | 0.51 | 1.53E-01 | 0.12 | 0.51 | 10 |
| Fiber cereals | Food | 25076 | -0.02 | 0.00 | 4.44E-06 | **1.89E-05** | 63 | 24452 | -0.03 | 0.00 | 1.79E-10 | **1.91E-09** | 25 | -0.02 | 0.00 | 1.39E-14 | 0.61 | 1.12E-01 | 0.12 | 0.51 | 3 |
| Boiled coffee | Beverage | 25076 | 0.03 | 0.00 | 9.00E-08 | **6.07E-07** | 40 | 24452 | 0.03 | 0.01 | 9.82E-08 | **5.72E-07** | 46 | 0.03 | 0.00 | 4.55E-14 | 0.00 | 7.87E-01 | 0.12 | 0.51 | 4 |
| Skill demand from job | Psychosocial | 26904 | -0.02 | 0.00 | 2.39E-07 | **1.34E-06** | 48 | 26361 | -0.03 | 0.00 | 4.85E-08 | **3.10E-07** | 42 | -0.02 | 0.00 | 6.69E-14 | 0.00 | 5.79E-01 | 0.11 | 0.51 | 68 |
| Snuff status: Snuff users vs non-snuff users | Tobacco use | 23376 | 0.07 | 0.01 | 3.41E-07 | **1.84E-06** | 50 | 22940 | 0.08 | 0.01 | 3.91E-08 | **2.62E-07** | 40 | 0.07 | 0.01 | 8.15E-14 | 0.00 | 5.10E-01 | 0.12 | 0.51 | 56 |
| Vitamin C intake (mg/day) | Nutrients | 25076 | -0.02 | 0.00 | 1.76E-07 | **1.08E-06** | 44 | 24452 | -0.03 | 0.00 | 9.01E-08 | **5.36E-07** | 45 | -0.02 | 0.00 | 8.42E-14 | 0.00 | 6.91E-01 | 0.12 | 0.51 | 8 |
| Cohabitation: Live alone vs Adult and children | Social | 15575 | -0.08 | 0.02 | 3.82E-06 | **1.76E-05** | 58 | 15262 | -0.11 | 0.02 | 2.22E-09 | **1.98E-08** | 30 | -0.09 | 0.01 | 9.24E-14 | 0.39 | 2.02E-01 | 0.13 | 0.50 | 1 |
| Physical limitation that made you not being able to perform certain work tasks or other activities during the last four weeks | General health | 13328 | 0.11 | 0.02 | 1.00E-08 | **7.53E-08** | 36 | 13157 | 0.09 | 0.02 | 2.34E-06 | **1.01E-05** | 62 | 0.10 | 0.01 | 1.29E-13 | 0.00 | 5.37E-01 | 0.11 | 0.55 | 101 |
| Breakfast habits: Porridge w/o sandwich for breakfast vs not breakfast at all | Food | 3706 | -0.15 | 0.03 | 8.26E-06 | **3.18E-05** | 70 | 3780 | -0.21 | 0.04 | 2.66E-09 | **2.30E-08** | 31 | -0.18 | 0.02 | 2.29E-13 | 0.42 | 1.88E-01 | 0.11 | 0.47 | 108 |
| Physical limitation that limited your ability to perform certain work tasks or other activities during the last four weeks | General health | 13353 | 0.11 | 0.02 | 2.70E-08 | **1.87E-07** | 39 | 13166 | 0.09 | 0.02 | 1.98E-06 | **8.86E-06** | 60 | 0.10 | 0.01 | 2.71E-13 | 0.00 | 6.57E-01 | 0.11 | 0.55 | 104 |
| Fried potatoes, pommes frites | Food | 25076 | 0.03 | 0.00 | 7.61E-09 | **6.04E-08** | 34 | 24452 | 0.02 | 0.00 | 7.42E-06 | **2.80E-05** | 71 | 0.02 | 0.00 | 3.49E-13 | 0.00 | 4.58E-01 | 0.12 | 0.51 | 6 |
| Smoking status: Former smokers vs non-smokers | Tobacco use | 19677 | 0.06 | 0.01 | 1.68E-06 | **8.10E-06** | 56 | 19034 | 0.07 | 0.01 | 3.55E-08 | **2.44E-07** | 39 | 0.06 | 0.01 | 4.14E-13 | 0.00 | 3.88E-01 | 0.12 | 0.49 | 49 |
| For how much of the time during the last four weeks have you felt worn out? | General health | 13337 | 0.04 | 0.01 | 4.69E-11 | **5.27E-10** | 24 | 13150 | 0.02 | 0.01 | 3.66E-04 | **1.02E-03** | 96 | 0.03 | 0.00 | 4.33E-13 | 0.77 | 3.51E-02 | 0.11 | 0.56 | 90 |
| Physical limitation that made you do less than you wanted during the last four weeks | General health | 13380 | 0.10 | 0.02 | 1.34E-09 | **1.13E-08** | 32 | 13211 | 0.07 | 0.02 | 3.82E-05 | **1.35E-04** | 76 | 0.09 | 0.01 | 4.42E-13 | 0.39 | 2.00E-01 | 0.11 | 0.55 | 97 |
| Satisfaction with work situation | Psychosocial | 22366 | -0.03 | 0.00 | 1.25E-08 | **9.10E-08** | 37 | 21975 | -0.02 | 0.00 | 8.74E-06 | **3.25E-05** | 72 | -0.02 | 0.00 | 6.59E-13 | 0.00 | 4.68E-01 | 0.12 | 0.51 | 40 |
| Frequent social contacts with colleagues during work | Psychosocial | 26668 | -0.02 | 0.00 | 3.64E-09 | **2.98E-08** | 33 | 26147 | -0.02 | 0.00 | 5.96E-05 | **2.05E-04** | 78 | -0.02 | 0.00 | 1.58E-12 | 0.00 | 3.20E-01 | 0.11 | 0.50 | 74 |
| Apple, pear, peach, orange, mandarin and grapefruit | Food | 25076 | -0.02 | 0.00 | 1.44E-06 | **7.19E-06** | 54 | 24452 | -0.03 | 0.01 | 2.02E-07 | **1.09E-06** | 50 | -0.02 | 0.00 | 1.59E-12 | 0.00 | 5.82E-01 | 0.12 | 0.51 | 12 |
| Patience status | Psychosocial | 22634 | -0.02 | 0.00 | 5.56E-06 | **2.21E-05** | 68 | 22265 | -0.03 | 0.00 | 7.58E-08 | **4.62E-07** | 44 | -0.02 | 0.00 | 2.53E-12 | 0.00 | 4.67E-01 | 0.12 | 0.50 | 46 |
| Secoisolariciresinol intake (ug/day) | Nutrients | 25076 | -0.02 | 0.00 | 3.85E-06 | **1.76E-05** | 59 | 24452 | -0.02 | 0.00 | 4.96E-07 | **2.46E-06** | 54 | -0.02 | 0.00 | 9.95E-12 | 0.00 | 5.95E-01 | 0.12 | 0.51 | 11 |
| For how much of the time during the last four weeks has your physical health or your emotional problems limited your ability to interact with others? | General health | 13299 | 0.04 | 0.01 | 2.09E-10 | **2.02E-09** | 28 | 13121 | 0.02 | 0.01 | 3.58E-03 | **8.07E-03** | 119 | 0.03 | 0.00 | 3.15E-11 | 0.83 | 1.61E-02 | 0.11 | 0.55 | 94 |
| Physical limitation to participate in moderately demanding activities: bending down or kneeling | General health | 13355 | 0.03 | 0.01 | 1.62E-05 | **6.07E-05** | 72 | 13189 | 0.03 | 0.01 | 9.56E-07 | **4.42E-06** | 58 | 0.03 | 0.00 | 7.75E-11 | 0.00 | 5.48E-01 | 0.11 | 0.55 | 105 |
| Tomato, cucumber | Food | 25076 | -0.02 | 0.00 | 4.28E-06 | **1.87E-05** | 62 | 24452 | -0.02 | 0.00 | 4.44E-06 | **1.77E-05** | 67 | -0.02 | 0.00 | 8.56E-11 | 0.00 | 7.82E-01 | 0.12 | 0.51 | 19 |
| Worsen in health in the future | General health | 13305 | 0.02 | 0.01 | 2.81E-04 | **8.52E-04** | 89 | 13142 | 0.04 | 0.01 | 2.77E-08 | **1.96E-07** | 38 | 0.03 | 0.00 | 9.16E-11 | 0.59 | 1.18E-01 | 0.11 | 0.56 | 102 |
| Number of snuff boxes per week | Tobacco use | 26510 | 0.02 | 0.00 | 9.69E-05 | **3.31E-04** | 79 | 25976 | 0.03 | 0.01 | 1.10E-07 | **6.29E-07** | 47 | 0.02 | 0.00 | 1.03E-10 | 0.39 | 2.02E-01 | 0.12 | 0.50 | 62 |
| Plant based protein intake (g/day) | Nutrients | 25076 | -0.01 | 0.00 | 5.36E-03 | **1.26E-02** | 115 | 24452 | -0.03 | 0.00 | 1.19E-10 | **1.42E-09** | 22 | -0.02 | 0.00 | 1.33E-10 | 0.88 | 4.63E-03 | 0.12 | 0.51 | 17 |
| Get sick more often than other people | General health | 13351 | 0.03 | 0.01 | 8.80E-07 | **4.57E-06** | 52 | 13182 | 0.03 | 0.01 | 4.06E-05 | **1.41E-04** | 77 | 0.03 | 0.00 | 1.61E-10 | 0.00 | 6.61E-01 | 0.11 | 0.56 | 100 |
| Monounsaturated fat intake (g/day) | Nutrients | 25076 | 0.02 | 0.00 | 1.20E-03 | **3.15E-03** | 103 | 24452 | 0.03 | 0.01 | 3.66E-09 | **2.97E-08** | 33 | 0.02 | 0.00 | 1.77E-10 | 0.78 | 3.19E-02 | 0.12 | 0.51 | 9 |
| Participation in associations or voluntary organisations | Social | 27219 | -0.04 | 0.01 | 4.32E-05 | **1.55E-04** | 75 | 26685 | -0.05 | 0.01 | 8.85E-07 | **4.16E-06** | 57 | -0.04 | 0.01 | 2.40E-10 | 0.00 | 3.77E-01 | 0.11 | 0.49 | 72 |
| Satisfaction with economy | Psychosocial | 22648 | -0.03 | 0.00 | 2.34E-07 | **1.34E-06** | 47 | 22264 | -0.02 | 0.01 | 2.46E-04 | **7.24E-04** | 91 | -0.02 | 0.00 | 3.61E-10 | 0.00 | 3.52E-01 | 0.12 | 0.51 | 44 |
| For how much of the time during the last four weeks have you felt calm and serene? | General health | 13361 | -0.04 | 0.01 | 1.71E-08 | **1.21E-07** | 38 | 13201 | -0.02 | 0.01 | 1.85E-03 | **4.56E-03** | 109 | -0.03 | 0.00 | 4.29E-10 | 0.67 | 8.29E-02 | 0.11 | 0.56 | 106 |
| Beta-carotene intake (mg/day) | Nutrients | 25076 | -0.02 | 0.00 | 3.11E-04 | **9.34E-04** | 90 | 24452 | -0.03 | 0.00 | 1.30E-07 | **7.29E-07** | 48 | -0.02 | 0.00 | 4.60E-10 | 0.51 | 1.54E-01 | 0.12 | 0.51 | 20 |
| For how much of the time during the last four weeks have you felt happy? | General health | 13367 | -0.03 | 0.01 | 1.42E-06 | **7.19E-06** | 53 | 13187 | -0.03 | 0.01 | 9.43E-05 | **3.01E-04** | 83 | -0.03 | 0.00 | 6.14E-10 | 0.00 | 5.81E-01 | 0.11 | 0.58 | 103 |
| Total energy intake (kcal/day) | Nutrients | 25076 | -0.02 | 0.01 | 4.83E-06 | **1.98E-05** | 66 | 24452 | -0.02 | 0.01 | 6.18E-05 | **2.10E-04** | 79 | -0.02 | 0.00 | 1.24E-09 | 0.00 | 8.37E-01 | 0.12 | 0.51 | 36 |
| Pizza | Food | 25076 | 0.02 | 0.00 | 5.66E-06 | **2.21E-05** | 69 | 24452 | 0.02 | 0.01 | 6.61E-05 | **2.21E-04** | 80 | 0.02 | 0.00 | 1.52E-09 | 0.00 | 9.14E-01 | 0.12 | 0.51 | 15 |
| Root vegetables, carrot | Food | 25076 | -0.02 | 0.00 | 6.35E-04 | **1.78E-03** | 96 | 24452 | -0.03 | 0.00 | 2.45E-07 | **1.26E-06** | 52 | -0.02 | 0.00 | 1.81E-09 | 0.54 | 1.42E-01 | 0.12 | 0.51 | 21 |
| Margarine for cooking | Food | 25076 | 0.02 | 0.00 | 1.58E-06 | **7.76E-06** | 55 | 24452 | 0.02 | 0.00 | 3.11E-04 | **8.85E-04** | 94 | 0.02 | 0.00 | 2.33E-09 | 0.00 | 5.37E-01 | 0.12 | 0.51 | 13 |
| Sodas, soft drinks, juice | Beverage | 25076 | 0.01 | 0.00 | 7.29E-02 | 1.27E-01 | 155 | 24452 | 0.03 | 0.01 | 7.66E-12 | **1.21E-10** | 17 | 0.02 | 0.00 | 2.48E-09 | 0.93 | 1.33E-04 | - | - | - |
| Educational level | Psychosocial | 27448 | -0.02 | 0.00 | 4.48E-06 | **1.89E-05** | 64 | 26855 | -0.02 | 0.01 | 2.09E-04 | **6.29E-04** | 89 | -0.02 | 0.00 | 3.91E-09 | 0.00 | 7.18E-01 | 0.11 | 0.49 | 78 |
| Frequency of cycling during leisure time | Physical activity | 22336 | -0.03 | 0.00 | 1.17E-07 | **7.53E-07** | 42 | 21741 | -0.01 | 0.01 | 4.00E-03 | **8.79E-03** | 122 | -0.02 | 0.00 | 4.74E-09 | 0.52 | 1.50E-01 | 0.12 | 0.49 | 66 |
| Job demands to work very fast | Psychosocial | 26964 | -0.02 | 0.00 | 4.57E-06 | **1.90E-05** | 65 | 26462 | -0.02 | 0.00 | 2.55E-04 | **7.42E-04** | 92 | -0.02 | 0.00 | 4.89E-09 | 0.00 | 6.95E-01 | 0.11 | 0.50 | 75 |
| Enough time for job assignments | Psychosocial | 26844 | 0.02 | 0.00 | 1.84E-04 | **5.77E-04** | 86 | 26333 | 0.02 | 0.00 | 5.68E-06 | **2.21E-05** | 69 | 0.02 | 0.00 | 5.80E-09 | 0.00 | 4.07E-01 | 0.11 | 0.51 | 69 |
| White cabbage, lettuce, lettuce cabbage, spinach, borecole | Food | 25076 | -0.02 | 0.00 | 1.95E-04 | **6.05E-04** | 87 | 24452 | -0.02 | 0.00 | 6.43E-06 | **2.46E-05** | 70 | -0.02 | 0.00 | 6.77E-09 | 0.00 | 4.25E-01 | 0.12 | 0.51 | 24 |
| Mood status | Psychosocial | 22635 | -0.02 | 0.00 | 8.14E-05 | **2.82E-04** | 78 | 22253 | -0.02 | 0.00 | 2.49E-05 | **9.01E-05** | 74 | -0.02 | 0.00 | 8.33E-09 | 0.00 | 7.46E-01 | 0.12 | 0.50 | 45 |
| Travel to work: Cycle to work vs passive travel to work | Physical activity | 20568 | -0.06 | 0.01 | 1.47E-07 | **9.22E-07** | 43 | 20079 | -0.04 | 0.01 | 6.32E-03 | **1.27E-02** | 133 | -0.05 | 0.01 | 9.84E-09 | 0.55 | 1.35E-01 | 0.12 | 0.51 | 57 |
| Extent to what your physical and emotional health disrupted your usual social life during the last four weeks | General health | 13381 | 0.03 | 0.01 | 2.11E-07 | **1.27E-06** | 45 | 13214 | 0.02 | 0.01 | 5.97E-03 | **1.22E-02** | 131 | 0.03 | 0.00 | 1.46E-08 | 0.64 | 9.69E-02 | 0.11 | 0.56 | 107 |
| Sum of Lariciresinol, Matairesinol, Pinoresinol, Secoisolariciresinol intake (ug/day) | Nutrients | 25076 | -0.01 | 0.00 | 3.56E-03 | **8.66E-03** | 111 | 24452 | -0.03 | 0.01 | 2.30E-07 | **1.21E-06** | 51 | -0.02 | 0.00 | 1.59E-08 | 0.70 | 6.78E-02 | 0.12 | 0.51 | 22 |
| Lariciresinol intake (ug/day) | Nutrients | 25076 | -0.01 | 0.00 | 3.66E-03 | **8.83E-03** | 112 | 24452 | -0.03 | 0.01 | 2.67E-07 | **1.35E-06** | 53 | -0.02 | 0.00 | 1.85E-08 | 0.70 | 6.98E-02 | 0.12 | 0.51 | 23 |
| Strong beer | Alcohol | 25076 | 0.01 | 0.00 | 8.18E-03 | **1.84E-02** | 120 | 24452 | 0.03 | 0.01 | 1.37E-07 | **7.49E-07** | 49 | 0.02 | 0.00 | 3.18E-08 | 0.76 | 4.08E-02 | 0.12 | 0.51 | 14 |
| Years using snuff | Tobacco use | 25813 | 0.02 | 0.01 | 1.18E-04 | **3.87E-04** | 82 | 25288 | 0.02 | 0.01 | 7.80E-05 | **2.58E-04** | 81 | 0.02 | 0.00 | 3.61E-08 | 0.00 | 7.75E-01 | 0.12 | 0.51 | 58 |
| Magnesium intake (mg/day) | Nutrients | 25076 | -0.01 | 0.00 | 2.45E-02 | **4.94E-02** | 134 | 24452 | -0.03 | 0.01 | 1.46E-08 | **1.09E-07** | 36 | -0.02 | 0.00 | 3.80E-08 | 0.86 | 8.41E-03 | 0.12 | 0.51 | 33 |
| Palmitic acid intake (g/day) | Nutrients | 25076 | 0.01 | 0.00 | 5.34E-03 | **1.26E-02** | 114 | 24452 | 0.02 | 0.00 | 8.50E-07 | **4.07E-06** | 56 | 0.02 | 0.00 | 6.66E-08 | 0.65 | 9.11E-02 | 0.12 | 0.51 | 28 |
| Parents or siblings had a cerebral hemorrhage/thrombosis or cardiac infarction before the age of 60 | General health | 26881 | 0.06 | 0.01 | 2.21E-07 | **1.30E-06** | 46 | 26343 | 0.03 | 0.01 | 2.31E-02 | **4.19E-02** | 148 | 0.05 | 0.01 | 8.80E-08 | 0.70 | 6.56E-02 | 0.11 | 0.49 | 73 |
| Parents or siblings have diabetes | General health | 26915 | 0.05 | 0.01 | 4.03E-06 | **1.81E-05** | 60 | 26354 | 0.03 | 0.01 | 4.92E-03 | **1.06E-02** | 125 | 0.04 | 0.01 | 1.21E-07 | 0.14 | 2.81E-01 | 0.11 | 0.49 | 80 |
| Trans fat intake (g/day) | Nutrients | 25076 | 0.03 | 0.01 | 4.17E-06 | **1.84E-05** | 61 | 24452 | 0.02 | 0.01 | 5.09E-03 | **1.07E-02** | 128 | 0.03 | 0.01 | 1.37E-07 | 0.21 | 2.60E-01 | 0.12 | 0.51 | 18 |
| Marital status: Single vs Married/partner | Social | 25030 | -0.02 | 0.02 | 1.24E-01 | 1.89E-01 | 177 | 24474 | -0.10 | 0.02 | 1.80E-09 | **1.67E-08** | 29 | -0.06 | 0.01 | 1.76E-07 | 0.91 | 7.71E-04 | - | - | - |
| Sour milk, yoghurt (3% fat) | Food | 25076 | -0.01 | 0.00 | 6.96E-04 | **1.92E-03** | 98 | 24452 | -0.02 | 0.00 | 8.24E-05 | **2.69E-04** | 82 | -0.02 | 0.00 | 2.43E-07 | 0.00 | 5.55E-01 | 0.12 | 0.51 | 16 |
| Self-employed | Psychosocial | 23118 | -0.06 | 0.02 | 4.50E-04 | **1.33E-03** | 91 | 22500 | -0.06 | 0.02 | 2.90E-04 | **8.37E-04** | 93 | -0.06 | 0.01 | 4.72E-07 | 0.00 | 7.90E-01 | 0.12 | 0.50 | 53 |
| Oatflake, whole wheat, rye or barley porridge | Food | 25076 | -0.01 | 0.00 | 6.48E-04 | **1.81E-03** | 97 | 24452 | -0.02 | 0.00 | 1.98E-04 | **6.03E-04** | 88 | -0.02 | 0.00 | 4.89E-07 | 0.00 | 6.76E-01 | 0.12 | 0.51 | 25 |

| | | | | | | | | | | | | | | | | | | | | | |
|---|---|---|---|---|---|---|---|---|---|---|---|---|---|---|---|---|---|---|---|---|---|
| High mental demand from job | Psychosocial | 26747 | 0.02 | 0.00 | 1.97E-05 | **7.19E-05** | 74 | 26245 | 0.01 | 0.00 | 6.15E-03 | **1.25E-02** | 132 | 0.02 | 0.00 | 5.68E-07 | 0.00 | 4.00E-01 | 0.11 | 0.50 | 77 |
| Number of social interactions during a normal week | Social | 27407 | -0.02 | 0.00 | 3.65E-07 | **1.93E-06** | 51 | 26822 | -0.01 | 0.00 | 7.64E-02 | 1.15E-01 | 178 | -0.02 | 0.00 | 7.26E-07 | 0.78 | 3.43E-02 | - | - | - |
| Sodium intake (mg/day) | Nutrients | 25075 | 0.02 | 0.00 | 2.87E-07 | **1.58E-06** | 49 | 24451 | 0.01 | 0.00 | 1.12E-01 | 1.60E-01 | 188 | 0.02 | 0.00 | 1.29E-06 | 0.82 | 1.97E-02 | - | - | - |
| Pinoresinol intake (ug/day) | Nutrients | 25076 | -0.01 | 0.00 | 2.44E-02 | **4.94E-02** | 132 | 24452 | -0.02 | 0.00 | 3.04E-06 | **1.30E-05** | 63 | -0.02 | 0.00 | 1.44E-06 | 0.73 | 5.64E-02 | 0.12 | 0.51 | 27 |
| Hamburger | Food | 25076 | 0.01 | 0.00 | 2.54E-02 | 5.03E-02 | 136 | 24452 | 0.02 | 0.00 | 3.25E-06 | **1.36E-05** | 64 | 0.02 | 0.00 | 1.58E-06 | 0.72 | 5.70E-02 | - | - | - |
| Fat intake (g/day) | Nutrients | 25076 | 0.01 | 0.00 | 2.51E-02 | 5.02E-02 | 135 | 24452 | 0.02 | 0.00 | 5.19E-06 | **2.04E-05** | 68 | 0.02 | 0.00 | 2.11E-06 | 0.70 | 6.93E-02 | - | - | - |
| Do you feel important and appreciated in your home? | Psychosocial | 22507 | -0.02 | 0.00 | 8.91E-04 | **2.41E-03** | 100 | 22113 | -0.02 | 0.00 | 7.69E-04 | **1.96E-03** | 105 | -0.02 | 0.00 | 2.28E-06 | 0.00 | 8.98E-01 | 0.12 | 0.50 | 47 |
| Monosaccharides intake (g/day) | Nutrients | 25076 | -0.02 | 0.00 | 1.34E-04 | **4.30E-04** | 84 | 24452 | -0.01 | 0.01 | 5.95E-03 | **1.22E-02** | 130 | -0.02 | 0.00 | 2.96E-06 | 0.00 | 5.77E-01 | 0.12 | 0.51 | 32 |
| Frequency of shoverling snow during leisure time | Physical activity | 16198 | -0.02 | 0.01 | 5.74E-04 | **1.63E-03** | 95 | 15762 | -0.02 | 0.01 | 2.13E-03 | **5.14E-03** | 111 | -0.02 | 0.00 | 3.93E-06 | 0.00 | 9.89E-01 | 0.11 | 0.46 | 98 |
| Do you feel important and appreciated outside your home? | Psychosocial | 22658 | -0.02 | 0.00 | 1.14E-04 | **3.79E-04** | 81 | 22288 | -0.01 | 0.00 | 1.26E-02 | **2.36E-02** | 142 | -0.02 | 0.00 | 6.39E-06 | 0.00 | 3.83E-01 | 0.12 | 0.50 | 52 |
| Fatty fish (e.g. herring, whitefish, salmon) | Food | 25076 | -0.02 | 0.00 | 4.64E-04 | **1.35E-03** | 92 | 24452 | -0.01 | 0.00 | 4.96E-03 | **1.06E-02** | 126 | -0.01 | 0.00 | 7.59E-06 | 0.00 | 7.41E-01 | 0.12 | 0.51 | 31 |
| Sausage, liver pate on bread | Food | 25076 | 0.01 | 0.00 | 8.88E-04 | **2.41E-03** | 99 | 24452 | 0.01 | 0.00 | 5.03E-03 | **1.06E-02** | 127 | 0.01 | 0.00 | 1.40E-05 | 0.00 | 8.25E-01 | 0.12 | 0.51 | 29 |
| Confidence status | Psychosocial | 22631 | -0.02 | 0.00 | 1.57E-03 | **4.00E-03** | 106 | 22238 | -0.01 | 0.01 | 3.51E-03 | **7.97E-03** | 118 | -0.02 | 0.00 | 1.69E-05 | 0.00 | 9.31E-01 | 0.12 | 0.51 | 51 |
| Sausage as main dish | Food | 25076 | 0.01 | 0.00 | 9.04E-03 | **2.00E-02** | 122 | 24452 | 0.02 | 0.00 | 5.72E-04 | **1.53E-03** | 100 | 0.01 | 0.00 | 1.99E-05 | 0.00 | 4.85E-01 | 0.12 | 0.51 | 30 |
| Possibility to speak with colleagues during breaks | Psychosocial | 26856 | -0.02 | 0.00 | 1.58E-05 | **6.01E-05** | 71 | 26350 | -0.01 | 0.00 | 1.21E-01 | 1.71E-01 | 189 | -0.01 | 0.00 | 2.21E-05 | 0.67 | 8.05E-02 | - | - | - |
| Emotional problems that made you do less than you wanted during the last four weeks | General health | 13396 | 0.08 | 0.02 | 4.36E-05 | **1.55E-04** | 76 | 13212 | 0.04 | 0.02 | 6.65E-02 | 1.03E-01 | 173 | 0.06 | 0.01 | 2.30E-05 | 0.57 | 1.27E-01 | - | - | - |
| Vision status | General health | 22647 | -0.02 | 0.00 | 1.30E-03 | **3.36E-03** | 104 | 22253 | -0.01 | 0.01 | 5.77E-03 | **1.20E-02** | 129 | -0.01 | 0.00 | 2.31E-05 | 0.00 | 8.12E-01 | 0.12 | 0.50 | 50 |
| Everyday exercise satisfaction | Physical activity | 16920 | -0.02 | 0.01 | 9.80E-05 | **3.31E-04** | 80 | 16461 | -0.01 | 0.01 | 5.05E-02 | 8.20E-02 | 165 | -0.02 | 0.00 | 2.53E-05 | 0.22 | 2.59E-01 | - | - | - |
| Potassium intake (mg/day) | Nutrients | 25076 | -0.01 | 0.00 | 8.03E-02 | 1.37E-01 | 158 | 24452 | -0.02 | 0.01 | 1.80E-05 | **6.60E-05** | 73 | -0.01 | 0.00 | 2.65E-05 | 0.74 | 5.09E-02 | - | - | - |
| Vitamin A intake (mg/day) | Nutrients | 25076 | 0.02 | 0.01 | 2.33E-03 | **5.88E-03** | 107 | 24452 | 0.02 | 0.01 | 3.80E-03 | **8.43E-03** | 120 | 0.02 | 0.00 | 2.66E-05 | 0.00 | 9.88E-01 | 0.12 | 0.51 | 26 |
| Satisfaction with leisure time | Psychosocial | 22616 | -0.02 | 0.00 | 1.47E-04 | **4.67E-04** | 85 | 22241 | -0.01 | 0.00 | 3.58E-02 | 6.04E-02 | 159 | -0.01 | 0.00 | 2.77E-05 | 0.20 | 2.63E-01 | - | - | - |
| Ingenuity or creativity demand from job | Psychosocial | 26836 | -0.01 | 0.00 | 1.06E-03 | **2.82E-03** | 102 | 26319 | -0.01 | 0.00 | 8.90E-03 | **1.70E-02** | 140 | -0.01 | 0.00 | 2.91E-05 | 0.00 | 7.79E-01 | 0.11 | 0.50 | 79 |
| Contradictory demands in job | Psychosocial | 26809 | -0.01 | 0.00 | 4.66E-04 | **1.35E-03** | 93 | 26254 | -0.01 | 0.00 | 2.29E-02 | **4.17E-02** | 147 | -0.01 | 0.00 | 3.82E-05 | 0.00 | 4.92E-01 | 0.11 | 0.50 | 70 |
| Tea | Beverage | 25076 | -0.02 | 0.00 | 1.90E-05 | **7.04E-05** | 73 | 24452 | -0.01 | 0.01 | 1.70E-01 | 2.31E-01 | 198 | -0.01 | 0.00 | 4.55E-05 | 0.72 | 5.97E-02 | - | - | - |
| Permanent employment | Psychosocial | 23118 | -0.03 | 0.01 | 6.16E-03 | **1.43E-02** | 116 | 22500 | -0.03 | 0.01 | 2.59E-03 | **6.03E-03** | 115 | -0.03 | 0.01 | 5.02E-05 | 0.00 | 7.08E-01 | 0.12 | 0.50 | 54 |
| Self rate of overall health compared to a year ago | General health | 13303 | -0.03 | 0.01 | 1.19E-04 | **3.87E-04** | 83 | 13125 | -0.01 | 0.01 | 6.51E-02 | 1.01E-01 | 172 | -0.02 | 0.00 | 5.10E-05 | 0.48 | 1.64E-01 | - | - | - |
| Selenium intake (ug/day) | Nutrients | 25076 | -0.01 | 0.00 | 2.40E-01 | 3.26E-01 | 199 | 24452 | -0.02 | 0.00 | 3.80E-06 | **1.54E-05** | 66 | -0.01 | 0.00 | 6.36E-05 | 0.85 | 9.29E-03 | - | - | - |
| Whole grain intake (g/day) | Food | 25076 | -0.01 | 0.00 | 4.93E-02 | 8.99E-02 | 148 | 24452 | -0.02 | 0.00 | 1.92E-04 | **5.92E-04** | 87 | -0.01 | 0.00 | 6.96E-05 | 0.49 | 1.62E-01 | - | - | - |
| Milk, sour milk (1,5%) | Beverage | 25076 | 0.02 | 0.00 | 7.57E-05 | **2.65E-04** | 77 | 24452 | 0.01 | 0.00 | 1.39E-01 | 1.93E-01 | 193 | 0.01 | 0.00 | 8.95E-05 | 0.60 | 1.13E-01 | - | - | - |
| Ice cream | Food | 25076 | -0.01 | 0.00 | 9.32E-02 | 1.57E-01 | 160 | 24452 | -0.02 | 0.00 | 9.44E-05 | **3.01E-04** | 84 | -0.01 | 0.00 | 9.60E-05 | 0.65 | 9.11E-02 | - | - | - |
| Cohabitation: Live alone vs Only one adult (spouse, partner) | Social | 12904 | -0.03 | 0.02 | 6.33E-02 | 1.12E-01 | 153 | 12759 | -0.07 | 0.02 | 1.75E-04 | **5.53E-04** | 85 | -0.05 | 0.01 | 9.61E-05 | 0.57 | 1.27E-01 | - | - | - |
| Liquor, spirits | Alcohol | 25076 | 0.01 | 0.00 | 6.85E-02 | 1.20E-01 | 154 | 24452 | 0.02 | 0.00 | 2.46E-04 | **7.24E-04** | 90 | 0.01 | 0.00 | 1.03E-04 | 0.41 | 1.94E-01 | - | - | - |
| Satisfaction with accomodation | Psychosocial | 22669 | -0.01 | 0.00 | 3.96E-02 | 7.32E-02 | 146 | 22295 | -0.02 | 0.00 | 5.77E-04 | **1.53E-03** | 101 | -0.01 | 0.00 | 1.09E-04 | 0.10 | 2.92E-01 | - | - | - |
| Learn new things at job | Psychosocial | 26915 | -0.01 | 0.00 | 2.00E-02 | **4.16E-02** | 130 | 26393 | -0.01 | 0.00 | 1.60E-03 | **4.01E-03** | 107 | -0.01 | 0.00 | 1.20E-04 | 0.00 | 4.47E-01 | 0.11 | 0.50 | 76 |
| Number of social contacts with the same interests as you | Social | 27361 | -0.01 | 0.00 | 1.46E-03 | **3.77E-03** | 105 | 26781 | -0.01 | 0.00 | 2.91E-02 | 5.09E-02 | 153 | -0.01 | 0.00 | 1.33E-04 | 0.00 | 5.90E-01 | - | - | - |
| Receive hugs to comfort and support you | Social | 27190 | -0.03 | 0.01 | 8.14E-03 | **1.84E-02** | 119 | 26607 | -0.03 | 0.01 | 8.56E-03 | **1.66E-02** | 138 | -0.03 | 0.01 | 1.93E-04 | 0.00 | 8.80E-01 | 0.11 | 0.49 | 71 |
| Light beer | Alcohol | 25076 | -0.02 | 0.00 | 5.30E-04 | **1.52E-03** | 94 | 24452 | -0.01 | 0.01 | 1.08E-01 | 1.55E-01 | 186 | -0.01 | 0.00 | 2.75E-04 | 0.27 | 2.43E-01 | - | - | - |
| Carbohydrates intake (g/day) | Nutrients | 25076 | -0.01 | 0.00 | 9.24E-02 | 1.57E-01 | 159 | 24452 | -0.02 | 0.00 | 4.89E-04 | **1.32E-03** | 99 | -0.01 | 0.00 | 3.12E-04 | 0.50 | 1.58E-01 | - | - | - |
| Docosahexaenoic acid (DHA) intake (g/day) | Nutrients | 25076 | -0.01 | 0.00 | 1.37E-02 | **2.92E-02** | 127 | 24452 | -0.01 | 0.00 | 8.31E-03 | **1.64E-02** | 136 | -0.01 | 0.00 | 3.18E-04 | 0.00 | 7.84E-01 | 0.12 | 0.51 | 34 |
| Campesterol intake (mg/day) | Nutrients | 25076 | 0.01 | 0.00 | 5.07E-02 | 9.12E-02 | 150 | 24452 | 0.01 | 0.00 | 1.84E-03 | **4.56E-03** | 108 | 0.01 | 0.00 | 3.83E-04 | 0.00 | 3.41E-01 | - | - | - |
| Cookies, pastry | Food | 25076 | -0.02 | 0.00 | 2.67E-04 | **8.19E-04** | 88 | 24452 | -0.01 | 0.00 | 2.15E-01 | 2.84E-01 | 203 | -0.01 | 0.00 | 4.30E-04 | 0.59 | 1.19E-01 | - | - | - |
| Satisfaction with home and family situation | Psychosocial | 22646 | -0.01 | 0.00 | 9.11E-03 | **2.00E-02** | 123 | 22277 | -0.01 | 0.00 | 2.53E-02 | **4.52E-02** | 150 | -0.01 | 0.00 | 6.01E-04 | 0.00 | 8.59E-01 | 0.12 | 0.50 | 48 |
| Eicosapentaenoic acid (EPA) intake (g/day) | Nutrients | 25075 | -0.01 | 0.00 | 1.42E-02 | **2.99E-02** | 128 | 24452 | -0.01 | 0.00 | 1.97E-02 | **3.62E-02** | 146 | -0.01 | 0.00 | 7.14E-04 | 0.00 | 9.67E-01 | 0.12 | 0.51 | 35 |
| Saturated fat intake (g/day) | Nutrients | 25076 | 0.01 | 0.00 | 1.33E-01 | 2.01E-01 | 179 | 24452 | 0.02 | 0.00 | 8.35E-04 | **2.11E-03** | 106 | 0.01 | 0.00 | 7.24E-04 | 0.50 | 1.58E-01 | - | - | - |
| Tiamin intake (mg/day) | Nutrients | 25076 | 0.00 | 0.00 | 4.92E-01 | 5.85E-01 | 227 | 24452 | -0.02 | 0.00 | 2.61E-05 | **9.33E-05** | 75 | -0.01 | 0.00 | 7.97E-04 | 0.86 | 8.53E-03 | - | - | - |
| Frequency of walking during leisure time | Physical activity | 26424 | -0.01 | 0.00 | 3.53E-03 | **8.66E-03** | 110 | 25889 | -0.01 | 0.00 | 9.13E-02 | 1.34E-01 | 182 | -0.01 | 0.00 | 1.00E-03 | 0.00 | 4.62E-01 | - | - | - |
| Sugar, honey, marmelade, jam | Food | 25076 | 0.01 | 0.00 | 8.02E-02 | 1.37E-01 | 157 | 24452 | 0.01 | 0.00 | 3.81E-03 | **8.43E-03** | 121 | 0.01 | 0.00 | 1.15E-03 | 0.00 | 3.52E-01 | - | - | - |
| Linolenic acid intake (g/day) | Nutrients | 25076 | 0.01 | 0.00 | 2.05E-01 | 2.90E-01 | 191 | 24452 | 0.02 | 0.00 | 7.61E-04 | **1.96E-03** | 104 | 0.01 | 0.00 | 1.30E-03 | 0.62 | 1.07E-01 | - | - | - |
| Vitamin D intake (ug/day) | Nutrients | 25076 | 0.01 | 0.00 | 5.10E-02 | 9.12E-02 | 151 | 24452 | 0.01 | 0.00 | 9.12E-03 | **1.73E-02** | 141 | 0.01 | 0.00 | 1.36E-03 | 0.00 | 5.57E-01 | - | - | - |
| Stigmasterol intake (mg/day) | Nutrients | 25076 | -0.01 | 0.00 | 1.16E-01 | 1.82E-01 | 171 | 24452 | -0.01 | 0.00 | 3.17E-03 | **7.27E-03** | 117 | -0.01 | 0.00 | 1.58E-03 | 0.16 | 2.74E-01 | - | - | - |
| Banana | Food | 25076 | -0.01 | 0.00 | 2.61E-02 | 5.12E-02 | 138 | 24452 | -0.01 | 0.00 | 2.93E-02 | 5.10E-02 | 154 | -0.01 | 0.00 | 1.86E-03 | 0.00 | 9.00E-01 | - | - | - |
| If you exercise, change in exercise habits during the last year | Physical activity | 14937 | -0.01 | 0.01 | 8.84E-03 | **1.97E-02** | 121 | 14574 | -0.01 | 0.01 | 8.95E-02 | 1.33E-01 | 180 | -0.01 | 0.00 | 2.01E-03 | 0.00 | 6.56E-01 | - | - | - |
| Cohabitation: Live alone vs Other/others | Social | 3117 | -0.12 | 0.05 | 7.81E-03 | **1.79E-02** | 118 | 3110 | -0.10 | 0.06 | 1.11E-01 | 1.59E-01 | 187 | -0.11 | 0.04 | 2.04E-03 | 0.00 | 7.36E-01 | - | - | - |
| Formic acid intaje (g/day) | Nutrients | 25076 | 0.01 | 0.00 | 1.84E-01 | 2.65E-01 | 187 | 24452 | 0.01 | 0.00 | 2.01E-03 | **4.89E-03** | 110 | 0.01 | 0.00 | 2.07E-03 | 0.45 | 1.78E-01 | - | - | - |
| Frequency of hunting or fishing during leisure time | Physical activity | 15770 | 0.01 | 0.01 | 2.69E-02 | 5.19E-02 | 139 | 15341 | 0.01 | 0.01 | 3.21E-02 | 5.48E-02 | 157 | 0.01 | 0.00 | 2.08E-03 | 0.00 | 9.00E-01 | - | - | - |
| For how much of the time during the last four weeks have you felt very nervous? | General health | 13360 | 0.01 | 0.01 | 3.17E-02 | 5.97E-02 | 142 | 13192 | 0.01 | 0.01 | 3.06E-02 | 5.28E-02 | 155 | 0.01 | 0.00 | 2.30E-03 | 0.00 | 9.41E-01 | - | - | - |
| Snuff status: Former snuff users vs non-snuff users | Tobacco use | 21911 | 0.05 | 0.02 | 9.03E-04 | **2.41E-03** | 101 | 21411 | 0.01 | 0.02 | 3.55E-01 | 4.39E-01 | 217 | 0.03 | 0.01 | 2.35E-03 | 0.62 | 1.05E-01 | - | - | - |
| Polyunsaturated fat intake (g/day) | Nutrients | 25076 | 0.01 | 0.00 | 9.92E-02 | 1.65E-01 | 162 | 24452 | 0.01 | 0.00 | 7.86E-03 | **1.56E-02** | 135 | 0.01 | 0.00 | 2.58E-03 | 0.00 | 4.03E-01 | - | - | - |
| Cohabitation: Live alone vs Only children | Social | 4068 | -0.06 | 0.03 | 3.18E-02 | 5.97E-02 | 144 | 4047 | -0.07 | 0.04 | 3.86E-02 | 6.43E-02 | 161 | -0.07 | 0.02 | 3.03E-03 | 0.00 | 7.52E-01 | - | - | - |
| Memory status | Psychosocial | 22614 | -0.01 | 0.00 | 2.49E-01 | 3.37E-01 | 200 | 22217 | -0.02 | 0.01 | 2.52E-03 | **5.92E-03** | 114 | -0.01 | 0.00 | 3.42E-03 | 0.47 | 1.69E-01 | - | - | - |
| Whole grain crisp bread | Food | 25076 | 0.01 | 0.00 | 2.90E-02 | 5.55E-02 | 141 | 24452 | 0.01 | 0.00 | 6.40E-02 | 1.00E-01 | 171 | 0.01 | 0.00 | 4.23E-03 | 0.00 | 9.04E-01 | - | - | - |
| Sour milk, yoghurt (low fat) | Food | 25076 | 0.00 | 0.00 | 3.14E-01 | 4.04E-01 | 210 | 24452 | -0.01 | 0.00 | 2.27E-03 | **5.44E-03** | 112 | -0.01 | 0.00 | 4.97E-03 | 0.59 | 1.18E-01 | - | - | - |
| For how much of the time during the last four weeks have you felt so depressed that nothing could cheer you up? | General health | 13374 | 0.02 | 0.01 | 1.05E-02 | **2.26E-02** | 125 | 13205 | 0.01 | 0.01 | 1.95E-01 | 2.59E-01 | 202 | 0.01 | 0.00 | 6.05E-03 | 0.00 | 3.94E-01 | - | - | - |
| White (soft) bread, thin crisp bread | Food | 25076 | 0.01 | 0.00 | 1.08E-01 | 1.74E-01 | 168 | 24452 | 0.01 | 0.01 | 2.78E-02 | **4.90E-02** | 152 | 0.01 | 0.00 | 7.49E-03 | 0.00 | 6.05E-01 | - | - | - |
| Berries (fresh or frozen) | Food | 25076 | -0.01 | 0.00 | 1.57E-01 | 2.33E-01 | 182 | 24452 | -0.01 | 0.00 | 1.65E-02 | **3.05E-02** | 145 | -0.01 | 0.00 | 7.77E-03 | 0.00 | 4.15E-01 | - | - | - |
| Calcium intake (mg/day) | Nutrients | 25076 | 0.00 | 0.00 | 6.48E-01 | 7.42E-01 | 236 | 24452 | -0.02 | 0.00 | 7.17E-04 | **1.86E-03** | 103 | -0.01 | 0.00 | 8.63E-03 | 0.79 | 2.91E-02 | - | - | - |
| Bacon | Food | 25076 | 0.01 | 0.00 | 1.63E-02 | **3.40E-02** | 129 | 24452 | 0.01 | 0.00 | 2.48E-01 | 3.20E-01 | 208 | 0.01 | 0.00 | 1.09E-02 | 0.00 | 4.29E-01 | - | - | - |
| Wine | Alcohol | 25076 | -0.01 | 0.00 | 1.02E-02 | **2.22E-02** | 124 | 24452 | 0.00 | 0.00 | 3.43E-01 | 4.25E-01 | 216 | -0.01 | 0.00 | 1.13E-02 | 0.08 | 2.97E-01 | - | - | - |
| Number of people with whom you can speak openly | Social | 27381 | -0.01 | 0.00 | 1.13E-01 | 1.80E-01 | 169 | 26798 | -0.01 | 0.00 | 4.33E-02 | 7.17E-02 | 162 | -0.01 | 0.00 | 1.13E-02 | 0.00 | 6.68E-01 | - | - | - |
| Salad dressing with oil | Food | 25076 | -0.01 | 0.00 | 1.02E-01 | 1.67E-01 | 165 | 24452 | -0.01 | 0.00 | 5.52E-02 | 8.79E-02 | 168 | -0.01 | 0.00 | 1.23E-02 | 0.00 | 7.69E-01 | - | - | - |
| Matairesinol intake (ug/day) | Nutrients | 25076 | 0.00 | 0.00 | 4.56E-01 | 5.48E-01 | 225 | 24452 | -0.01 | 0.00 | 4.25E-03 | **9.25E-03** | 123 | -0.01 | 0.00 | 1.29E-02 | 0.61 | 1.10E-01 | - | - | - |
| Participation in sports or physical exercise associations | Social | 9986 | -0.04 | 0.01 | 7.31E-03 | **1.69E-02** | 117 | 9682 | -0.01 | 0.02 | 5.15E-01 | 6.01E-01 | 230 | -0.03 | 0.01 | 1.53E-02 | 0.43 | 1.85E-01 | - | - | - |
| For how much of the time during the last four weeks have you felt gloomy and sad? | General health | 13344 | 0.01 | 0.01 | 2.17E-01 | 3.04E-01 | 193 | 13186 | 0.01 | 0.01 | 3.27E-02 | 5.55E-02 | 158 | 0.01 | 0.00 | 1.76E-02 | 0.00 | 4.96E-01 | - | - | - |
| Pasta | Food | 25076 | 0.01 | 0.00 | 2.89E-03 | **7.15E-03** | 109 | 24452 | 0.00 | 0.00 | 8.05E-01 | 8.53E-01 | 253 | 0.01 | 0.00 | 1.92E-02 | 0.71 | 6.28E-02 | - | - | - |
| Total protein intake (g/day) | Nutrients | 25076 | 0.00 | 0.00 | 9.40E-01 | 9.52E-01 | 266 | 24452 | -0.02 | 0.00 | 4.38E-04 | **1.20E-03** | 98 | -0.01 | 0.00 | 1.95E-02 | 0.86 | 8.55E-03 | - | - | - |
| Butter on bread | Food | 25076 | 0.01 | 0.00 | 2.69E-02 | 5.19E-02 | 140 | 24452 | 0.00 | 0.00 | 3.10E-01 | 3.93E-01 | 212 | 0.01 | 0.00 | 2.08E-02 | 0.00 | 4.46E-01 | - | - | - |
| Disaccharides intake (g/day) | Nutrients | 25076 | 0.00 | 0.00 | 4.05E-01 | 4.99E-01 | 219 | 24452 | 0.01 | 0.01 | 1.37E-02 | **2.56E-02** | 144 | 0.01 | 0.00 | 2.22E-02 | 0.35 | 2.15E-01 | - | - | - |
| Support from others | Social | 27408 | 0.00 | 0.00 | 3.12E-01 | 4.04E-01 | 208 | 26834 | -0.01 | 0.00 | 2.51E-02 | **4.51E-02** | 149 | -0.01 | 0.00 | 2.39E-02 | 0.00 | 3.32E-01 | - | - | - |
| Pancake, waffle, Swedish dumpling | Food | 25076 | 0.01 | 0.00 | 2.60E-02 | 5.12E-02 | 137 | 24452 | 0.00 | 0.01 | 3.75E-01 | 4.59E-01 | 219 | 0.01 | 0.00 | 2.55E-02 | 0.00 | 3.83E-01 | - | - | - |
| Sweets | Food | 25076 | 0.01 | 0.00 | 1.37E-01 | 2.06E-01 | 180 | 24452 | 0.01 | 0.00 | 9.78E-02 | 1.43E-01 | 183 | 0.01 | 0.00 | 2.66E-02 | 0.00 | 8.51E-01 | - | - | - |
| Repetitive job | Psychosocial | 26934 | 0.00 | 0.00 | 5.64E-01 | 6.57E-01 | 232 | 26405 | 0.01 | 0.00 | 8.49E-03 | **1.66E-02** | 137 | 0.01 | 0.00 | 2.78E-02 | 0.59 | 1.20E-01 | - | - | - |
| Chips, popcorn, salted nuts | Food | 25076 | 0.01 | 0.00 | 5.72E-02 | 1.02E-01 | 152 | 24452 | 0.01 | 0.00 | 2.74E-01 | 3.51E-01 | 209 | 0.01 | 0.00 | 3.28E-02 | 0.00 | 6.08E-01 | - | - | - |
| Changed everyday exercise during the last year | Physical activity | 16970 | -0.01 | 0.00 | 9.37E-02 | 1.57E-01 | 161 | 16520 | -0.01 | 0.01 | 1.92E-01 | 2.57E-01 | 200 | -0.01 | 0.00 | 3.40E-02 | 0.00 | 8.90E-01 | - | - | - |
| Beta-sitosterol intake (mg/day) | Nutrients | 25076 | -0.01 | 0.00 | 2.35E-01 | 3.22E-01 | 197 | 24452 | -0.01 | 0.00 | 6.85E-02 | 1.05E-01 | 175 | -0.01 | 0.00 | 3.49E-02 | 0.00 | 5.98E-01 | - | - | - |
| Meat stew | Food | 25076 | 0.00 | 0.00 | 2.79E-01 | 3.71E-01 | 203 | 24452 | 0.01 | 0.00 | 5.25E-02 | 8.42E-02 | 167 | 0.01 | 0.00 | 3.49E-02 | 0.00 | 4.87E-01 | - | - | - |
| Iron intake (mg/day) | Nutrients | 25076 | 0.00 | 0.00 | 6.82E-01 | 7.74E-01 | 238 | 24452 | -0.01 | 0.01 | 8.86E-03 | **1.70E-02** | 139 | -0.01 | 0.00 | 3.77E-02 | 0.63 | 1.00E-01 | - | - | - |
| Frequency of gardening during leisure time | Physical activity | 16128 | -0.01 | 0.01 | 2.28E-01 | 3.16E-01 | 195 | 15655 | -0.01 | 0.01 | 8.31E-02 | 1.24E-01 | 179 | -0.01 | 0.00 | 4.03E-02 | 0.00 | 6.16E-01 | - | - | - |
| Brown beans, pea soup | Food | 25076 | -0.01 | 0.00 | 1.14E-01 | 1.81E-01 | 170 | 24452 | -0.01 | 0.00 | 1.94E-01 | 2.58E-01 | 201 | -0.01 | 0.00 | 4.09E-02 | 0.00 | 9.15E-01 | - | - | - |
| Distance to work in kilometers (one way) | Physical activity | 23589 | 0.01 | 0.00 | 1.16E-01 | 1.82E-01 | 172 | 23009 | 0.01 | 0.00 | 2.40E-01 | 3.11E-01 | 207 | 0.01 | 0.00 | 5.10E-02 | 0.00 | 8.45E-01 | - | - | - |
| Low fat milk (0,5%) | Beverage | 25076 | 0.01 | 0.00 | 1.19E-01 | 1.85E-01 | 174 | 24452 | 0.01 | 0.00 | 2.38E-01 | 3.09E-01 | 206 | 0.01 | 0.00 | 5.19E-02 | 0.00 | 8.42E-01 | - | - | - |
| Work shifts/weekends | Psychosocial | 26612 | 0.02 | 0.01 | 3.80E-02 | 7.07E-02 | 145 | 26136 | 0.01 | 0.01 | 5.52E-01 | 6.27E-01 | 236 | 0.01 | 0.01 | 5.39E-02 | 0.00 | 3.31E-01 | - | - | - |
| Control over own work assignment | Psychosocial | 26971 | 0.00 | 0.00 | 9.21E-01 | 9.45E-01 | 263 | 26456 | 0.01 | 0.00 | 3.11E-03 | **7.19E-03** | 116 | 0.01 | 0.00 | 5.51E-02 | 0.80 | 2.43E-02 | - | - | - |
| Lean fish (e.g. perch, bass, cod) | Food | 25076 | 0.00 | 0.00 | 9.07E-01 | 9.45E-01 | 257 | 24452 | -0.01 | 0.00 | 6.59E-03 | **1.32E-02** | 134 | -0.01 | 0.00 | 5.54E-02 | 0.73 | 5.35E-02 | - | - | - |
| Linoleic acid intake (g/day) | Nutrients | 25076 | 0.00 | 0.00 | 4.24E-01 | 5.18E-01 | 221 | 24452 | 0.01 | 0.00 | 5.20E-02 | 8.40E-02 | 166 | 0.01 | 0.00 | 5.67E-02 | 0.00 | 3.76E-01 | - | - | - |
| Would you say that the number of people that you meet in your everyday life is enough or would you like to meet more or fewer people? | Social | 27397 | 0.01 | 0.00 | 5.01E-02 | 9.09E-02 | 149 | 26810 | 0.00 | 0.00 | 4.99E-01 | 5.87E-01 | 228 | 0.01 | 0.00 | 5.73E-02 | 0.00 | 4.10E-01 | - | - | - |

| | | | | | | | | | | | | | | | | | | | | | |
|---|---|---|---|---|---|---|---|---|---|---|---|---|---|---|---|---|---|---|---|---|---|
| Frequency of picking berries or mushrooms during leisure time | Physical activity | 16019 | -0.01 | 0.01 | 1.04E-01 | 1.69E-01 | 166 | 15601 | -0.01 | 0.01 | 3.17E-01 | 3.99E-01 | 213 | -0.01 | 0.00 | 6.01E-02 | 0.00 | 7.44E-01 | - | - | - |
| Feel uneasy or guilty because of your way of drinking | Alcohol | 15256 | 0.02 | 0.02 | 3.89E-01 | 4.84E-01 | 217 | 14943 | 0.03 | 0.02 | 7.01E-02 | 1.07E-01 | 176 | 0.02 | 0.01 | 6.25E-02 | 0.00 | 4.56E-01 | - | - | - |
| White meat (poultry) | Food | 25076 | 0.00 | 0.00 | 4.49E-01 | 5.42E-01 | 224 | 24452 | -0.01 | 0.00 | 5.54E-02 | 8.79E-02 | 169 | -0.01 | 0.00 | 6.31E-02 | 0.00 | 3.75E-01 | - | - | - |
| Medioresinol intake (ug/day) | Nutrients | 25076 | 0.01 | 0.00 | 2.44E-02 | **4.94E-02** | 133 | 24452 | 0.00 | 0.00 | 8.40E-01 | 8.79E-01 | 256 | 0.01 | 0.00 | 7.34E-02 | 0.47 | 1.68E-01 | - | - | - |
| Bregott on bread | Food | 25076 | 0.00 | 0.00 | 3.68E-01 | 4.62E-01 | 215 | 24452 | 0.02 | 0.00 | 4.29E-04 | **1.18E-03** | 97 | 0.01 | 0.00 | 8.28E-02 | 0.90 | 1.39E-03 | - | - | - |
| Enterodiol intake (ug/day) | Nutrients | 25076 | 0.01 | 0.00 | 2.22E-01 | 3.10E-01 | 194 | 24452 | 0.01 | 0.00 | 2.26E-01 | 2.96E-01 | 204 | 0.01 | 0.00 | 8.57E-02 | 0.00 | 9.51E-01 | - | - | - |
| Hearing status | General health | 22666 | 0.00 | 0.01 | 6.00E-01 | 6.95E-01 | 233 | 22284 | -0.01 | 0.01 | 5.87E-02 | 9.25E-02 | 170 | -0.01 | 0.00 | 9.07E-02 | 0.00 | 3.20E-01 | - | - | - |
| Boiled or baked potato | Food | 25076 | 0.01 | 0.00 | 1.97E-01 | 2.81E-01 | 189 | 24452 | 0.01 | 0.00 | 2.79E-01 | 3.56E-01 | 210 | 0.01 | 0.00 | 9.25E-02 | 0.00 | 9.29E-01 | - | - | - |
| Close relationship with anyone | Social | 27375 | 0.00 | 0.00 | 7.44E-01 | 8.30E-01 | 242 | 26789 | -0.01 | 0.00 | 4.61E-03 | **9.96E-03** | 124 | -0.01 | 0.00 | 9.28E-02 | 0.81 | 2.12E-02 | - | - | - |
| Enterolactone intake (ug/day) | Nutrients | 25076 | 0.00 | 0.00 | 3.12E-01 | 4.04E-01 | 207 | 24452 | -0.01 | 0.00 | 1.70E-01 | 2.31E-01 | 197 | -0.01 | 0.00 | 9.42E-02 | 0.00 | 7.49E-01 | - | - | - |
| Phosphate intake (mg/day) | Nutrients | 25076 | 0.00 | 0.00 | 2.90E-01 | 3.81E-01 | 205 | 24452 | -0.02 | 0.00 | 3.49E-04 | **9.84E-04** | 95 | -0.01 | 0.00 | 9.82E-02 | 0.91 | 8.26E-04 | - | - | - |
| Vitamin B2 intake (ug/day) | Nutrients | 25076 | 0.00 | 0.00 | 3.37E-01 | 4.29E-01 | 212 | 24452 | -0.02 | 0.00 | 7.06E-04 | **1.86E-03** | 102 | -0.01 | 0.00 | 1.11E-01 | 0.90 | 1.69E-03 | - | - | - |
| Smoking status: Former occasional smokers vs non-smokers | Tobacco use | 16209 | 0.03 | 0.02 | 4.87E-02 | 8.95E-02 | 147 | 15924 | 0.00 | 0.02 | 7.98E-01 | 8.51E-01 | 251 | 0.02 | 0.01 | 1.13E-01 | 0.30 | 2.31E-01 | - | - | - |
| Coffee rolls/buns, rusk | Food | 25076 | -0.01 | 0.00 | 1.19E-01 | 1.85E-01 | 173 | 24452 | 0.00 | 0.00 | 5.41E-01 | 6.19E-01 | 234 | -0.01 | 0.00 | 1.19E-01 | 0.00 | 5.39E-01 | - | - | - |
| Teetotaler | Alcohol | 16952 | -0.03 | 0.02 | 1.08E-01 | 1.74E-01 | 167 | 16506 | -0.01 | 0.02 | 6.69E-01 | 7.41E-01 | 242 | -0.02 | 0.01 | 1.36E-01 | 0.00 | 4.60E-01 | - | - | - |
| Beta-sitostanol intake (mg/day) | Nutrients | 25076 | 0.01 | 0.00 | 2.27E-02 | **4.69E-02** | 131 | 24452 | 0.00 | 0.00 | 7.85E-01 | 8.42E-01 | 250 | 0.00 | 0.00 | 1.36E-01 | 0.67 | 8.10E-02 | - | - | - |
| Frequncy of social contacts with colleagues during leisure time | Psychosocial | 26185 | 0.00 | 0.00 | 4.40E-01 | 5.33E-01 | 223 | 25680 | -0.01 | 0.00 | 1.91E-01 | 2.57E-01 | 199 | 0.00 | 0.00 | 1.47E-01 | 0.00 | 6.50E-01 | - | - | - |
| Zinc intake (mg/day) | Nutrients | 25076 | 0.00 | 0.00 | 3.63E-01 | 4.58E-01 | 214 | 24452 | -0.02 | 0.00 | 2.40E-03 | **5.70E-03** | 113 | 0.00 | 0.00 | 1.64E-01 | 0.88 | 4.41E-03 | - | - | - |
| Campestanol intake (mg/day) | Nutrients | 25076 | 0.01 | 0.00 | 1.35E-02 | **2.89E-02** | 126 | 24452 | 0.00 | 0.00 | 5.23E-01 | 6.07E-01 | 231 | 0.00 | 0.00 | 1.67E-01 | 0.78 | 3.19E-02 | - | - | - |
| Rice | Food | 25076 | 0.01 | 0.00 | 2.73E-03 | **6.83E-03** | 108 | 24452 | -0.01 | 0.00 | 2.35E-01 | 3.07E-01 | 205 | 0.00 | 0.00 | 1.71E-01 | 0.88 | 3.52E-03 | - | - | - |
| Low fat margarine on bread | Food | 25076 | 0.01 | 0.00 | 1.99E-01 | 2.83E-01 | 190 | 24452 | 0.00 | 0.00 | 5.42E-01 | 6.19E-01 | 235 | 0.00 | 0.00 | 1.76E-01 | 0.00 | 6.68E-01 | - | - | - |
| Corn flakes | Food | 25076 | 0.00 | 0.00 | 9.27E-01 | 9.49E-01 | 264 | 24452 | -0.01 | 0.00 | 3.78E-02 | 6.32E-02 | 160 | 0.00 | 0.00 | 1.77E-01 | 0.60 | 1.14E-01 | - | - | - |
| Sum of all lignans intake (ug/day) | Nutrients | 25076 | 0.00 | 0.00 | 8.00E-01 | 8.74E-01 | 247 | 24452 | -0.01 | 0.00 | 2.72E-02 | **4.83E-02** | 151 | 0.00 | 0.00 | 1.89E-01 | 0.69 | 7.27E-02 | - | - | - |
| Cheese 10-17% | Food | 25076 | -0.01 | 0.00 | 1.26E-01 | 1.92E-01 | 178 | 24452 | 0.00 | 0.00 | 8.00E-01 | 8.51E-01 | 252 | 0.00 | 0.00 | 1.95E-01 | 0.00 | 3.96E-01 | - | - | - |
| Frequency of engaging in clubs, associations or study circles | Social | 17085 | -0.01 | 0.01 | 1.84E-01 | 2.65E-01 | 186 | 16622 | 0.00 | 0.01 | 7.38E-01 | 8.05E-01 | 245 | 0.00 | 0.00 | 2.24E-01 | 0.00 | 5.28E-01 | - | - | - |
| Rosehip, sweet syrup soup | Food | 25076 | 0.01 | 0.00 | 1.24E-01 | 1.89E-01 | 176 | 24452 | 0.00 | 0.00 | 9.28E-01 | 9.42E-01 | 264 | 0.00 | 0.00 | 2.36E-01 | 0.00 | 3.25E-01 | - | - | - |
| Arachidonic acid (ARA) intake (g/day) | Nutrients | 25075 | 0.00 | 0.00 | 4.35E-01 | 5.29E-01 | 222 | 24451 | 0.00 | 0.00 | 4.01E-01 | 4.84E-01 | 221 | 0.00 | 0.00 | 2.53E-01 | 0.00 | 9.32E-01 | - | - | - |
| Steak, chop, e.g. | Food | 25076 | 0.00 | 0.00 | 7.57E-01 | 8.38E-01 | 244 | 24452 | 0.01 | 0.00 | 5.03E-02 | 8.20E-02 | 164 | 0.00 | 0.00 | 2.69E-01 | 0.63 | 9.98E-02 | - | - | - |
| Control over planning and execution of the workday | Psychosocial | 26995 | -0.01 | 0.00 | 7.81E-02 | 1.35E-01 | 156 | 26448 | 0.00 | 0.00 | 7.56E-01 | 8.14E-01 | 249 | 0.00 | 0.00 | 2.74E-01 | 0.50 | 1.57E-01 | - | - | - |
| Vitamin B6 intake (mg/day) | Nutrients | 25076 | 0.00 | 0.00 | 8.38E-01 | 9.12E-01 | 248 | 24452 | -0.01 | 0.00 | 6.77E-02 | 1.04E-01 | 174 | 0.00 | 0.00 | 2.76E-01 | 0.54 | 1.39E-01 | - | - | - |
| Iodine intake (ug/day) | Nutrients | 25076 | 0.01 | 0.00 | 3.94E-03 | **9.41E-03** | 113 | 24452 | -0.01 | 0.00 | 1.25E-01 | 1.75E-01 | 191 | 0.00 | 0.00 | 2.82E-01 | 0.89 | 2.04E-03 | - | - | - |
| Average portion size of potatoes/rice/pasta based on photographic illustration of four sizes (smallest to largest) | Food | 25076 | 0.00 | 0.00 | 9.16E-01 | 9.45E-01 | 260 | 24452 | -0.01 | 0.01 | 9.04E-02 | 1.34E-01 | 181 | 0.00 | 0.00 | 2.84E-01 | 0.42 | 1.88E-01 | - | - | - |
| Cholesterol intake (g/day) | Nutrients | 25076 | 0.00 | 0.00 | 8.94E-01 | 9.45E-01 | 255 | 24452 | 0.01 | 0.00 | 1.64E-01 | 2.24E-01 | 196 | 0.00 | 0.00 | 2.94E-01 | 0.00 | 3.54E-01 | - | - | - |
| Heptadecanoic acid intake (g/day) | Nutrients | 25076 | 0.00 | 0.00 | 8.92E-01 | 9.45E-01 | 254 | 24452 | 0.01 | 0.00 | 1.06E-01 | 1.54E-01 | 185 | 0.00 | 0.00 | 3.14E-01 | 0.38 | 2.04E-01 | - | - | - |
| Pentadecanoic acid intake (g/day) | Nutrients | 25076 | 0.00 | 0.00 | 8.92E-01 | 9.45E-01 | 253 | 24452 | 0.01 | 0.00 | 1.06E-01 | 1.54E-01 | 184 | 0.00 | 0.00 | 3.14E-01 | 0.38 | 2.04E-01 | - | - | - |
| Sedentary or standing work | Physical activity | 26559 | 0.02 | 0.01 | 1.24E-01 | 1.89E-01 | 175 | 26014 | 0.00 | 0.01 | 8.15E-01 | 8.60E-01 | 254 | 0.01 | 0.01 | 3.33E-01 | 0.33 | 2.22E-01 | - | - | - |
| Frequency of dancing during leisure time | Physical activity | 15120 | 0.00 | 0.01 | 4.79E-01 | 5.73E-01 | 226 | 14694 | 0.00 | 0.01 | 5.32E-01 | 6.15E-01 | 232 | 0.00 | 0.00 | 3.45E-01 | 0.00 | 9.99E-01 | - | - | - |
| Minced meat dishes | Food | 25076 | 0.00 | 0.00 | 4.04E-01 | 4.99E-01 | 218 | 24452 | 0.00 | 0.00 | 6.96E-01 | 7.68E-01 | 243 | 0.00 | 0.00 | 3.81E-01 | 0.00 | 7.73E-01 | - | - | - |
| Margarine on bread | Food | 25076 | -0.01 | 0.00 | 2.08E-01 | 2.93E-01 | 192 | 24452 | 0.00 | 0.00 | 9.17E-01 | 9.38E-01 | 262 | 0.00 | 0.00 | 3.93E-01 | 0.00 | 3.52E-01 | - | - | - |
| Butter for cooking | Food | 25076 | 0.00 | 0.00 | 7.70E-01 | 8.48E-01 | 245 | 24452 | 0.01 | 0.00 | 1.44E-01 | 1.98E-01 | 194 | 0.00 | 0.00 | 4.26E-01 | 0.37 | 2.08E-01 | - | - | - |
| Whole grain soft bread | Food | 25076 | 0.00 | 0.00 | 5.62E-01 | 6.57E-01 | 231 | 24452 | -0.01 | 0.00 | 7.42E-02 | 1.12E-01 | 177 | 0.00 | 0.00 | 4.33E-01 | 0.66 | 8.81E-02 | - | - | - |
| Salty fish | Food | 25076 | 0.00 | 0.00 | 6.57E-01 | 7.48E-01 | 237 | 24452 | 0.00 | 0.00 | 5.04E-01 | 5.90E-01 | 229 | 0.00 | 0.00 | 4.38E-01 | 0.00 | 8.34E-01 | - | - | - |
| Sometimes physically straining work | Physical activity | 26559 | -0.01 | 0.01 | 3.86E-01 | 4.82E-01 | 216 | 26014 | 0.00 | 0.01 | 8.63E-01 | 8.89E-01 | 259 | -0.01 | 0.01 | 4.50E-01 | 0.00 | 6.45E-01 | - | - | - |
| Cream, creme fraiche, sour cream | Food | 25076 | 0.00 | 0.00 | 8.64E-01 | 9.33E-01 | 250 | 24452 | 0.00 | 0.00 | 3.64E-01 | 4.48E-01 | 218 | 0.00 | 0.00 | 4.55E-01 | 0.00 | 5.87E-01 | - | - | - |
| Participation in study circles | Social | 9986 | -0.01 | 0.02 | 6.03E-01 | 6.96E-01 | 234 | 9682 | -0.01 | 0.02 | 6.01E-01 | 6.73E-01 | 239 | -0.01 | 0.01 | 4.61E-01 | 0.00 | 9.69E-01 | - | - | - |
| Number of friends that can come to your home at any time and feel at home | Social | 27298 | 0.00 | 0.00 | 3.32E-01 | 4.25E-01 | 211 | 26741 | 0.00 | 0.00 | 9.77E-01 | 9.81E-01 | 267 | 0.00 | 0.00 | 4.62E-01 | 0.00 | 5.28E-01 | - | - | - |
| Travel to work: Walk to work vs passive travel to work | Physical activity | 17695 | -0.02 | 0.02 | 1.83E-01 | 2.65E-01 | 185 | 17519 | 0.01 | 0.02 | 7.27E-01 | 7.99E-01 | 244 | -0.01 | 0.01 | 4.62E-01 | 0.26 | 2.44E-01 | - | - | - |
| Meat on bread | Food | 25076 | 0.00 | 0.00 | 2.73E-01 | 3.65E-01 | 202 | 24452 | 0.00 | 0.00 | 9.25E-01 | 9.42E-01 | 263 | 0.00 | 0.00 | 4.64E-01 | 0.00 | 4.12E-01 | - | - | - |
| Light but partly physically active work | Physical activity | 26559 | 0.01 | 0.01 | 3.61E-01 | 4.58E-01 | 213 | 26014 | 0.00 | 0.01 | 9.92E-01 | 9.92E-01 | 268 | 0.01 | 0.01 | 5.05E-01 | 0.00 | 5.33E-01 | - | - | - |
| High physical demand from job | Physical activity | 27039 | 0.00 | 0.00 | 2.91E-01 | 3.81E-01 | 206 | 26519 | 0.00 | 0.00 | 8.62E-01 | 8.89E-01 | 258 | 0.00 | 0.00 | 5.08E-01 | 0.00 | 4.00E-01 | - | - | - |
| Vitamin B12 intake (ug/day) | Nutrients | 25076 | 0.00 | 0.00 | 2.82E-01 | 3.74E-01 | 204 | 24452 | 0.00 | 0.00 | 8.46E-01 | 8.82E-01 | 257 | 0.00 | 0.00 | 5.13E-01 | 0.00 | 3.81E-01 | - | - | - |
| Breakfast habits: Gruel w/o sandwich for breakfast vs not breakfast at all | Food | 1239 | -0.01 | 0.06 | 8.82E-01 | 9.45E-01 | 251 | 1306 | -0.06 | 0.07 | 4.12E-01 | 4.93E-01 | 224 | -0.03 | 0.05 | 5.16E-01 | 0.00 | 5.99E-01 | - | - | - |
| Participation in other association | Social | 9986 | 0.01 | 0.01 | 6.38E-01 | 7.33E-01 | 235 | 9682 | 0.01 | 0.02 | 6.69E-01 | 7.41E-01 | 241 | 0.01 | 0.01 | 5.25E-01 | 0.00 | 9.99E-01 | - | - | - |
| Medium beer | Alcohol | 25076 | 0.00 | 0.00 | 5.26E-01 | 6.20E-01 | 229 | 24452 | -0.01 | 0.00 | 1.44E-01 | 1.98E-01 | 195 | 0.00 | 0.00 | 5.87E-01 | 0.55 | 1.35E-01 | - | - | - |
| Vitamin B3 intake (mg/day) | Nutrients | 25076 | 0.00 | 0.00 | 2.65E-01 | 3.56E-01 | 201 | 24452 | -0.01 | 0.00 | 4.86E-02 | 8.00E-02 | 163 | 0.00 | 0.00 | 6.00E-01 | 0.79 | 2.75E-02 | - | - | - |
| Smoked fish/meat | Food | 25076 | 0.00 | 0.00 | 9.69E-01 | 9.73E-01 | 269 | 24452 | 0.00 | 0.00 | 4.66E-01 | 5.53E-01 | 226 | 0.00 | 0.00 | 6.06E-01 | 0.00 | 6.06E-01 | - | - | - |
| Average portion size of meat/fish based on photographic illustration of four sizes (smallest to largest) | Food | 25076 | 0.00 | 0.00 | 7.19E-01 | 8.12E-01 | 239 | 24452 | 0.00 | 0.01 | 7.47E-01 | 8.08E-01 | 248 | 0.00 | 0.00 | 6.29E-01 | 0.00 | 9.96E-01 | - | - | - |
| People to ask for help apart from the ones at home | Social | 27322 | 0.00 | 0.01 | 9.32E-01 | 9.49E-01 | 265 | 26739 | -0.01 | 0.02 | 5.54E-01 | 6.27E-01 | 237 | 0.00 | 0.01 | 6.43E-01 | 0.00 | 7.06E-01 | - | - | - |
| Sum of phytosterols intake (mg/day) | Nutrients | 25076 | 0.00 | 0.00 | 9.06E-01 | 9.45E-01 | 256 | 24452 | 0.00 | 0.00 | 5.86E-01 | 6.60E-01 | 238 | 0.00 | 0.00 | 6.48E-01 | 0.00 | 7.50E-01 | - | - | - |
| Marital status: Single vs Divorced/separated | Social | 4510 | 0.03 | 0.02 | 1.62E-01 | 2.39E-01 | 183 | 4471 | -0.08 | 0.03 | 1.26E-02 | **2.36E-02** | 143 | -0.01 | 0.02 | 6.49E-01 | 0.87 | 4.69E-03 | - | - | - |
| Animal based protein intake (g/day) | Nutrients | 25076 | 0.01 | 0.00 | 2.36E-01 | 3.22E-01 | 198 | 24452 | 0.00 | 0.00 | 5.36E-01 | 6.17E-01 | 233 | 0.00 | 0.00 | 6.56E-01 | 0.37 | 2.08E-01 | - | - | - |
| Breakfast habits: Only coffee/tea for breakfast vs not breakfast at all | Food | 1717 | 0.03 | 0.05 | 5.00E-01 | 5.92E-01 | 228 | 1786 | -0.01 | 0.07 | 8.63E-01 | 8.89E-01 | 260 | 0.02 | 0.04 | 6.64E-01 | 0.00 | 5.86E-01 | - | - | - |
| Possibility to leave your work for a while to speak with a colleague | Psychosocial | 26685 | 0.00 | 0.00 | 9.41E-01 | 9.52E-01 | 267 | 26178 | 0.00 | 0.00 | 4.72E-01 | 5.58E-01 | 227 | 0.00 | 0.00 | 6.70E-01 | 0.00 | 5.60E-01 | - | - | - |
| Last time a colleague visited you at home | Psychosocial | 26570 | 0.00 | 0.00 | 9.20E-01 | 9.45E-01 | 262 | 26076 | 0.00 | 0.00 | 6.16E-01 | 6.88E-01 | 240 | 0.00 | 0.00 | 6.82E-01 | 0.00 | 7.60E-01 | - | - | - |
| Cheese 28% | Food | 25076 | 0.00 | 0.00 | 7.28E-01 | 8.16E-01 | 241 | 24452 | 0.00 | 0.00 | 8.66E-01 | 8.89E-01 | 261 | 0.00 | 0.00 | 7.12E-01 | 0.00 | 9.10E-01 | - | - | - |
| Marital status: Single vs Widow/widower | Social | 2848 | -0.01 | 0.05 | 9.15E-01 | 9.45E-01 | 259 | 2776 | 0.06 | 0.07 | 4.39E-01 | 5.23E-01 | 225 | 0.01 | 0.04 | 7.30E-01 | 0.00 | 4.83E-01 | - | - | - |
| Breakfast habits: Coffee/tea and wheat buns or rusk for breakfast vs not breakfast at all | Food | 1375 | 0.01 | 0.05 | 7.97E-01 | 8.74E-01 | 246 | 1367 | -0.06 | 0.07 | 4.01E-01 | 4.84E-01 | 222 | -0.01 | 0.04 | 7.38E-01 | 0.00 | 4.15E-01 | - | - | - |
| Syringaresinol intake (ug/day) | Nutrients | 25076 | 0.01 | 0.00 | 1.54E-01 | 2.30E-01 | 181 | 24452 | -0.01 | 0.00 | 2.91E-01 | 3.70E-01 | 211 | 0.00 | 0.00 | 7.41E-01 | 0.67 | 8.14E-02 | - | - | - |
| Oil for cooking | Food | 25076 | 0.00 | 0.00 | 7.22E-01 | 8.13E-01 | 240 | 24452 | 0.00 | 0.00 | 4.04E-01 | 4.86E-01 | 223 | 0.00 | 0.00 | 7.59E-01 | 0.00 | 3.94E-01 | - | - | - |
| Alcohol intake (g/day) | Alcohol | 25076 | -0.01 | 0.00 | 2.35E-01 | 3.22E-01 | 196 | 24452 | 0.01 | 0.01 | 1.32E-01 | 1.84E-01 | 192 | 0.00 | 0.00 | 8.71E-01 | 0.73 | 5.59E-02 | - | - | - |
| Equol intake (ug/day) | Nutrients | 25076 | 0.00 | 0.00 | 5.28E-01 | 6.20E-01 | 230 | 24452 | 0.00 | 0.00 | 3.95E-01 | 4.81E-01 | 220 | 0.00 | 0.00 | 9.06E-01 | 0.10 | 2.92E-01 | - | - | - |
| Sucrose intake (g/day) | Nutrients | 25076 | -0.01 | 0.00 | 3.17E-02 | 5.97E-02 | 143 | 24452 | 0.01 | 0.00 | 3.08E-02 | 5.29E-02 | 156 | 0.00 | 0.00 | 9.11E-01 | 0.89 | 2.33E-03 | - | - | - |
| Milk, sour milk (3%) | Beverage | 25076 | 0.00 | 0.00 | 8.48E-01 | 9.20E-01 | 249 | 24452 | 0.00 | 0.00 | 7.40E-01 | 8.05E-01 | 246 | 0.00 | 0.00 | 9.35E-01 | 0.00 | 7.08E-01 | - | - | - |
| Vitamin E intake (mg/day) | Nutrients | 25076 | 0.00 | 0.00 | 9.49E-01 | 9.56E-01 | 268 | 24452 | 0.00 | 0.00 | 9.68E-01 | 9.75E-01 | 266 | 0.00 | 0.00 | 9.41E-01 | 0.00 | 9.89E-01 | - | - | - |
| Travel to work: Irregular travel mode to work vs passive travel to work | Physical activity | 17666 | 0.00 | 0.02 | 9.78E-01 | 9.78E-01 | 270 | 17448 | 0.00 | 0.02 | 9.42E-01 | 9.52E-01 | 265 | 0.00 | 0.01 | 9.44E-01 | 0.00 | 9.73E-01 | - | - | - |
| Light and physically active work | Physical activity | 26559 | 0.00 | 0.01 | 9.12E-01 | 9.45E-01 | 258 | 26014 | 0.00 | 0.01 | 8.27E-01 | 8.69E-01 | 255 | 0.00 | 0.01 | 9.46E-01 | 0.00 | 8.14E-01 | - | - | - |
| Amount of alcohol drunk in a day | Alcohol | 10015 | 0.04 | 0.01 | - | - | 274 | 9869 | 0.06 | 0.01 | 3.76E-11 | **4.80E-10** | 21 | 0.05 | 0.01 | - | 0.00 | 3.19E-01 | - | - | - |
| Snore during sleep | Sleep | 9666 | 0.03 | 0.01 | - | - | 283 | 9587 | 0.04 | 0.01 | 1.34E-06 | **6.07E-06** | 59 | 0.03 | 0.01 | - | 0.17 | 2.72E-01 | - | - | - |
| Breath-holds during sleep | Sleep | 7214 | 0.02 | 0.01 | - | - | 284 | 7101 | 0.04 | 0.01 | 1.91E-04 | **5.92E-04** | 86 | 0.03 | 0.01 | - | 0.39 | 2.02E-01 | - | - | - |
| Times during last year that you felt guilty because of your drinking | Alcohol | 10193 | 0.01 | 0.01 | - | - | 276 | 10039 | 0.01 | 0.01 | 1.21E-01 | 1.71E-01 | 190 | 0.01 | 0.01 | - | 0.00 | 5.98E-01 | - | - | - |
| Risk of sleeping while lying down resting in the afternoon | Sleep | 10314 | 0.01 | 0.01 | - | - | 281 | 10219 | 0.01 | 0.01 | 3.37E-01 | 4.22E-01 | 214 | 0.01 | 0.01 | - | 0.00 | 8.45E-01 | - | - | - |
| Risk of sleeping while sitting and reading | Sleep | 10244 | -0.02 | 0.01 | - | - | 277 | 10131 | -0.01 | 0.01 | 3.39E-01 | 4.23E-01 | 215 | -0.01 | 0.01 | - | 0.00 | 3.61E-01 | - | - | - |
| Risk of sleeping while sitting still after having lunch | Sleep | 10323 | 0.00 | 0.01 | - | - | 282 | 10221 | 0.00 | 0.01 | 7.42E-01 | 8.05E-01 | 247 | 0.00 | 0.01 | - | 0.00 | 6.77E-01 | - | - | - |
| Mashed potato | Food | 4303 | 0.03 | 0.01 | 1.01E-01 | 1.67E-01 | 163 | 4154 | 0.00 | 0.01 | - | - | 274 | 0.02 | 0.01 | - | 0.70 | 7.01E-02 | - | - | - |
| Liver, kidney | Food | 4303 | -0.03 | 0.01 | 1.01E-01 | 1.67E-01 | 164 | 4154 | 0.01 | 0.01 | - | - | 278 | -0.01 | 0.01 | - | 0.84 | 1.22E-02 | - | - | - |
| Potato salad | Food | 4303 | 0.02 | 0.01 | 1.67E-01 | 2.45E-01 | 184 | 4154 | 0.00 | 0.01 | - | - | 275 | 0.01 | 0.01 | - | 0.41 | 1.94E-01 | - | - | - |
| Blota (broth + bread) | Food | 4303 | 0.02 | 0.01 | 1.84E-01 | 2.65E-01 | 188 | 4154 | 0.02 | 0.02 | - | - | 276 | 0.02 | 0.01 | - | 0.00 | 9.61E-01 | - | - | - |
| Blood based food | Food | 4303 | -0.01 | 0.01 | 3.13E-01 | 4.04E-01 | 209 | 4154 | 0.00 | 0.01 | - | - | 277 | -0.01 | 0.01 | - | 0.09 | 2.94E-01 | - | - | - |
| Soft whey cheese | Food | 4303 | -0.01 | 0.01 | 4.08E-01 | 5.01E-01 | 220 | 4154 | -0.01 | 0.01 | - | - | 272 | -0.01 | 0.01 | - | 0.00 | 8.11E-01 | - | - | - |
| Soft cheese | Food | 4303 | 0.00 | 0.01 | 7.56E-01 | 8.38E-01 | 243 | 4154 | -0.03 | 0.01 | - | - | 271 | -0.01 | 0.01 | - | 0.68 | 7.73E-02 | - | - | - |
| Mixed frozen vegetables | Food | 4303 | 0.00 | 0.01 | 8.91E-01 | 9.45E-01 | 252 | 4154 | -0.01 | 0.01 | - | - | 273 | 0.00 | 0.01 | - | 0.00 | 7.98E-01 | - | - | - |
| Shellfish (e.g. shrimps, scallops) | Food | 4303 | 0.00 | 0.01 | 9.20E-01 | 9.45E-01 | 261 | 4154 | -0.02 | 0.01 | - | - | 279 | -0.01 | 0.01 | - | 0.58 | 1.22E-01 | - | - | - |
| Time spent in a week in moderately steneous activities | Physical activity | 10121 | -0.05 | 0.01 | - | - | 271 | 9970 | -0.03 | 0.01 | - | - | 269 | -0.04 | 0.01 | - | 0.60 | 1.12E-01 | - | - | - |
| Amount of exercise during the last 12 months | Physical activity | 10452 | -0.06 | 0.01 | - | - | 272 | 10309 | -0.06 | 0.01 | - | - | 270 | -0.06 | 0.01 | - | 0.00 | 9.99E-01 | - | - | - |
| Frequency of alcohol consumption | Alcohol | 10492 | 0.00 | 0.01 | - | - | 273 | 10377 | 0.01 | 0.01 | - | - | 280 | 0.00 | 0.01 | - | 0.34 | 2.19E-01 | - | - | - |

| | | | | | | | | | | | | | | | | | | | | | |
|---|---|---|---|---|---|---|---|---|---|---|---|---|---|---|---|---|---|---|---|---|---|
| Frequency of drinking six or more glasses at the same occasion | Alcohol | 10206 | 0.06 | 0.01 | - | - | 275 | 10062 | 0.05 | 0.01 | - | - | 281 | 0.06 | 0.01 | - | 0.00 | 6.55E-01 | - | - | - |
| Risk of sleeping while watching TV | Sleep | 10358 | -0.02 | 0.01 | - | - | 278 | 10244 | 0.00 | 0.01 | - | - | 282 | -0.01 | 0.01 | - | 0.59 | 1.18E-01 | - | - | - |
| Risk of sleeping while sitting inactive in a public place | Sleep | 10287 | -0.01 | 0.01 | - | - | 279 | 10177 | -0.01 | 0.01 | - | - | 283 | -0.01 | 0.01 | - | 0.00 | 4.35E-01 | - | - | - |
| Risk of sleeping as a passenger in a car for one hour without break | Sleep | 10253 | -0.01 | 0.01 | - | - | 280 | 10134 | 0.00 | 0.01 | - | - | 284 | -0.01 | 0.01 | - | 0.00 | 4.93E-01 | - | - | - |

**Supplementary Table 8**. Linear mixed model association results for HDL cholesterol

| Description | Group | Training set | | | | | | Testing set | | | | | | Metanalysis | | | | | R2 | | |
|---|---|---|---|---|---|---|---|---|---|---|---|---|---|---|---|---|---|---|---|---|---|
| | | N | Effect estimate | S.E. | p-value | p-value$_{FDR}$ | p-value rank | N | Effect estimate | S.E. | p-value | p-value$_{FDR}$ | p-value rank | Effect estimate | S.E. | p-value | $I^2$ | Q p-value | marginal R2 | conditional R2 | R2 rank |
| Alcohol intake (g/day) | Alcohol | 11888 | 0.05 | 0.00 | 3.95E-28 | **1.04E-25** | 1 | 11685 | 0.04 | 0.00 | 3.43E-27 | **4.75E-25** | 2 | 0.04 | 0.00 | 1.33E-53 | 0.00 | 8.48E-01 | 0.17 | 0.36 | 4 |
| Wine | Alcohol | 11888 | 0.04 | 0.00 | 3.35E-25 | **4.40E-23** | 2 | 11685 | 0.04 | 0.00 | 8.76E-28 | **2.43E-25** | 1 | 0.04 | 0.00 | 3.00E-51 | 0.00 | 6.68E-01 | 0.17 | 0.36 | 5 |
| Secoisolariciresinol intake (ug/day) | Nutrients | 11888 | 0.03 | 0.00 | 3.02E-11 | **1.13E-09** | 7 | 11685 | 0.03 | 0.00 | 8.55E-15 | **5.92E-13** | 4 | 0.03 | 0.00 | 2.49E-24 | 0.00 | 3.59E-01 | 0.17 | 0.35 | 10 |
| Exercise during the last three months | Physical activity | 12872 | 0.03 | 0.00 | 3.77E-13 | **2.48E-11** | 4 | 12726 | 0.03 | 0.00 | 7.84E-12 | **3.10E-10** | 7 | 0.03 | 0.00 | 1.92E-23 | 0.00 | 8.44E-01 | 0.16 | 0.36 | 42 |
| Fitness status | Physical activity | 11710 | 0.03 | 0.00 | 1.17E-14 | **1.03E-12** | 3 | 11473 | 0.03 | 0.00 | 6.06E-10 | **1.68E-08** | 10 | 0.03 | 0.00 | 6.29E-23 | 0.00 | 3.68E-01 | 0.18 | 0.34 | 2 |
| Feel the need to reduce alcohol consumption | Alcohol | 12150 | 0.08 | 0.01 | 2.52E-11 | **1.10E-09** | 6 | 11933 | 0.08 | 0.01 | 1.59E-11 | **5.52E-10** | 8 | 0.08 | 0.01 | 2.45E-21 | 0.00 | 9.99E-01 | 0.16 | 0.35 | 37 |
| Overall state of health during the last year | General health | 13139 | 0.03 | 0.00 | 1.70E-12 | **8.92E-11** | 5 | 12978 | 0.02 | 0.00 | 1.06E-09 | **2.67E-08** | 11 | 0.03 | 0.00 | 1.33E-20 | 0.00 | 5.01E-01 | 0.16 | 0.36 | 38 |
| Carbohydrates intake (g/day) | Nutrients | 11888 | -0.02 | 0.00 | 3.04E-09 | **1.00E-07** | 8 | 11685 | -0.03 | 0.00 | 2.28E-12 | **1.05E-10** | 6 | -0.03 | 0.00 | 6.17E-20 | 0.00 | 3.52E-01 | 0.17 | 0.36 | 8 |
| Smoking status: Smokers vs non-smokers | Tobacco use | 8222 | -0.06 | 0.01 | 8.85E-07 | **1.45E-05** | 16 | 8146 | -0.10 | 0.01 | 4.24E-15 | **3.91E-13** | 3 | -0.08 | 0.01 | 1.75E-19 | 0.78 | 3.10E-02 | 0.15 | 0.34 | 49 |
| Cambridge physical activity index | Physical activity | 12359 | 0.02 | 0.00 | 9.61E-08 | **2.11E-06** | 12 | 12212 | 0.03 | 0.00 | 9.39E-11 | **2.89E-09** | 9 | 0.02 | 0.00 | 7.19E-17 | 0.00 | 3.67E-01 | 0.16 | 0.36 | 43 |
| Strong beer | Alcohol | 11888 | 0.02 | 0.00 | 3.78E-08 | **9.04E-07** | 11 | 11685 | 0.02 | 0.00 | 1.02E-08 | **2.02E-07** | 14 | 0.02 | 0.00 | 2.08E-15 | 0.00 | 7.80E-01 | 0.17 | 0.36 | 7 |
| Self rate of overall health | General health | 9521 | 0.02 | 0.00 | 2.67E-08 | **7.03E-07** | 10 | 9215 | 0.02 | 0.00 | 2.99E-08 | **5.52E-07** | 15 | 0.02 | 0.00 | 3.81E-15 | 0.00 | 9.65E-01 | - | - | - |
| Sucrose intake (g/day) | Nutrients | 11888 | -0.02 | 0.00 | 3.67E-07 | **6.90E-06** | 14 | 11685 | -0.03 | 0.00 | 3.81E-09 | **8.79E-08** | 12 | -0.02 | 0.00 | 8.63E-15 | 0.00 | 5.19E-01 | 0.17 | 0.36 | 9 |
| Cholesterol intake (g/day) | Nutrients | 11888 | 0.02 | 0.00 | 5.47E-07 | **9.59E-06** | 15 | 11685 | 0.02 | 0.00 | 1.14E-06 | **1.75E-05** | 18 | 0.02 | 0.00 | 2.87E-12 | 0.00 | 9.88E-01 | 0.17 | 0.36 | 11 |
| Excellent health | General health | 9497 | 0.02 | 0.00 | 3.27E-07 | **6.62E-06** | 13 | 9187 | 0.02 | 0.00 | 2.57E-06 | **3.75E-05** | 19 | 0.02 | 0.00 | 3.81E-12 | 0.00 | 6.98E-01 | - | - | - |
| Disaccharides intake (g/day) | Nutrients | 11888 | -0.02 | 0.00 | 1.86E-05 | **1.96E-04** | 25 | 11685 | -0.03 | 0.00 | 3.89E-08 | **6.74E-07** | 16 | -0.02 | 0.00 | 5.18E-12 | 0.00 | 3.32E-01 | 0.17 | 0.36 | 6 |
| Number of cigarretes smoked per day (in groups) | Tobacco use | 10646 | -0.01 | 0.00 | 8.13E-04 | **3.62E-03** | 59 | 10566 | -0.03 | 0.00 | 4.52E-09 | **9.63E-08** | 13 | -0.02 | 0.00 | 6.77E-11 | 0.68 | 7.68E-02 | 0.16 | 0.35 | 46 |
| Years smoking | Tobacco use | 11339 | -0.02 | 0.00 | 6.96E-05 | **5.77E-04** | 31 | 11163 | -0.02 | 0.00 | 9.28E-07 | **1.51E-05** | 17 | -0.02 | 0.00 | 3.61E-10 | 0.00 | 4.40E-01 | 0.15 | 0.36 | 50 |
| Physical limitation to participate in steneous activities: running, lifting heavy objects, taking part in physically demanding sports | General health | 9500 | -0.02 | 0.00 | 2.92E-06 | **3.65E-05** | 21 | 9193 | -0.02 | 0.00 | 3.50E-05 | **3.87E-04** | 25 | -0.02 | 0.00 | 4.24E-10 | 0.00 | 6.83E-01 | - | - | - |
| Travel to work: Cycle to work vs passive travel to work | Physical activity | 9689 | 0.05 | 0.01 | 1.67E-06 | **2.32E-05** | 19 | 9485 | 0.04 | 0.01 | 7.97E-05 | **7.61E-04** | 29 | 0.05 | 0.01 | 6.76E-10 | 0.00 | 5.07E-01 | 0.16 | 0.34 | 36 |
| Liquor, spirits | Alcohol | 11888 | 0.02 | 0.00 | 1.98E-08 | **5.78E-07** | 9 | 11685 | 0.01 | 0.00 | 1.60E-03 | **7.40E-03** | 60 | 0.02 | 0.00 | 7.88E-10 | 0.74 | 5.18E-02 | 0.17 | 0.36 | 12 |
| Eicosapentaenoic acid (EPA) intake (g/day) | Nutrients | 11887 | 0.02 | 0.00 | 1.28E-06 | **1.88E-05** | 18 | 11685 | 0.01 | 0.00 | 4.33E-04 | **2.88E-03** | 41 | 0.02 | 0.00 | 2.95E-09 | 0.00 | 4.23E-01 | 0.17 | 0.36 | 23 |
| Informed of having high blood pressure | General health | 13134 | -0.04 | 0.01 | 2.31E-05 | **2.34E-04** | 26 | 12974 | -0.04 | 0.01 | 3.22E-05 | **3.72E-04** | 24 | -0.04 | 0.01 | 2.98E-09 | 0.00 | 9.93E-01 | 0.16 | 0.36 | 44 |
| Docosahexaenoic acid (DHA) intake (g/day) | Nutrients | 11888 | 0.02 | 0.00 | 1.22E-06 | **1.88E-05** | 17 | 11685 | 0.01 | 0.00 | 5.61E-04 | **3.38E-03** | 46 | 0.02 | 0.00 | 3.60E-09 | 0.00 | 4.23E-01 | 0.17 | 0.36 | 25 |
| Palmitic acid intake (g/day) | Nutrients | 11888 | 0.02 | 0.00 | 3.11E-05 | **3.03E-04** | 27 | 11685 | 0.02 | 0.00 | 3.18E-05 | **3.72E-04** | 23 | 0.02 | 0.00 | 3.93E-09 | 0.00 | 9.63E-01 | 0.17 | 0.36 | 18 |
| For how much of the time during the last four weeks have you felt full of energy? | General health | 9498 | 0.02 | 0.00 | 2.48E-04 | **1.48E-03** | 44 | 9179 | 0.02 | 0.00 | 1.08E-05 | **1.49E-04** | 20 | 0.02 | 0.00 | 1.08E-08 | 0.00 | 5.61E-01 | - | - | - |
| Medium beer | Alcohol | 11888 | 0.02 | 0.00 | 1.40E-04 | **9.92E-04** | 37 | 11685 | 0.02 | 0.00 | 2.02E-05 | **2.67E-04** | 21 | 0.02 | 0.00 | 1.17E-08 | 0.00 | 7.01E-01 | 0.17 | 0.36 | 20 |
| For how much of the time during the last four weeks have you felt really alert and strong? | General health | 9496 | 0.02 | 0.00 | 2.41E-04 | **1.47E-03** | 43 | 9166 | 0.02 | 0.00 | 4.09E-05 | **4.35E-04** | 26 | 0.02 | 0.00 | 3.63E-08 | 0.00 | 7.50E-01 | - | - | - |
| Educational level | Psychosocial | 13161 | 0.01 | 0.00 | 3.18E-04 | **1.74E-03** | 48 | 12982 | 0.02 | 0.00 | 3.00E-05 | **3.72E-04** | 22 | 0.02 | 0.00 | 3.91E-08 | 0.00 | 6.60E-01 | 0.16 | 0.36 | 39 |
| Energy status | Psychosocial | 11708 | 0.01 | 0.00 | 2.12E-04 | **1.41E-03** | 39 | 11463 | 0.02 | 0.00 | 1.21E-04 | **1.08E-03** | 31 | 0.01 | 0.00 | 9.50E-08 | 0.00 | 8.60E-01 | 0.18 | 0.34 | 3 |
| How much has the pain during the last four weeks disturbed your normal work? | General health | 9495 | -0.02 | 0.00 | 2.24E-04 | **1.41E-03** | 41 | 9167 | -0.02 | 0.00 | 1.57E-04 | **1.21E-03** | 36 | -0.02 | 0.00 | 1.23E-07 | 0.00 | 9.87E-01 | - | - | - |
| For how much of the time during the last four weeks have you felt tired? | General health | 9522 | -0.02 | 0.00 | 2.19E-04 | **1.41E-03** | 40 | 9190 | -0.02 | 0.00 | 1.84E-04 | **1.34E-03** | 38 | -0.02 | 0.00 | 1.40E-07 | 0.00 | 9.66E-01 | 0.22 | 0.22 | 1 |
| Fatty fish (e.g. herring, whitefish, salmon) | Food | 11888 | 0.02 | 0.00 | 2.78E-06 | **3.65E-05** | 20 | 11685 | 0.01 | 0.00 | 8.86E-03 | **2.82E-02** | 87 | 0.01 | 0.00 | 2.06E-07 | 0.47 | 1.70E-01 | 0.17 | 0.36 | 27 |
| Get sick more often than other people | General health | 9503 | -0.02 | 0.00 | 8.40E-05 | **6.50E-04** | 34 | 9182 | -0.01 | 0.00 | 7.29E-04 | **4.21E-03** | 48 | -0.01 | 0.00 | 2.24E-07 | 0.00 | 6.63E-01 | - | - | - |
| White cabbage, lettuce, lettuce cabbage, spinach, borecole | Food | 11888 | 0.02 | 0.00 | 5.39E-05 | **4.89E-04** | 29 | 11685 | 0.01 | 0.00 | 1.36E-03 | **6.61E-03** | 56 | 0.01 | 0.00 | 2.80E-07 | 0.00 | 6.61E-01 | 0.17 | 0.36 | 28 |
| As healthy as anyone | General health | 9477 | 0.01 | 0.00 | 2.07E-03 | **7.89E-03** | 69 | 9156 | 0.02 | 0.00 | 4.25E-05 | **4.36E-04** | 27 | 0.01 | 0.00 | 3.54E-07 | 0.00 | 4.35E-01 | - | - | - |
| Coffee rolls/buns, rusk | Food | 11888 | -0.02 | 0.00 | 9.82E-05 | **7.17E-04** | 36 | 11685 | -0.02 | 0.00 | 9.72E-04 | **5.23E-03** | 50 | -0.02 | 0.00 | 3.58E-07 | 0.00 | 7.01E-01 | 0.17 | 0.36 | 13 |
| Vitamin B3 intake (mg/day) | Nutrients | 11888 | 0.01 | 0.00 | 7.51E-04 | **3.40E-03** | 58 | 11685 | 0.01 | 0.00 | 1.71E-04 | **1.28E-03** | 37 | 0.01 | 0.00 | 4.72E-07 | 0.00 | 7.29E-01 | 0.17 | 0.36 | 31 |
| Years using snuff | Tobacco use | 12455 | 0.02 | 0.00 | 1.53E-05 | **1.75E-04** | 23 | 12299 | 0.01 | 0.00 | 6.25E-03 | **2.25E-02** | 77 | 0.01 | 0.00 | 5.42E-07 | 0.09 | 2.95E-01 | 0.16 | 0.39 | 35 |
| Average portion size of vegetables based on photographic illustration of four sizes (smallest to largest) | Food | 11888 | 0.01 | 0.00 | 4.74E-04 | **2.50E-03** | 49 | 11685 | 0.02 | 0.00 | 4.36E-04 | **2.88E-03** | 42 | 0.01 | 0.00 | 7.12E-07 | 0.00 | 9.27E-01 | 0.16 | 0.35 | 41 |
| Physical limitation to participate in moderately demanding activities: lifting or carrying grocery bags | General health | 9481 | -0.02 | 0.00 | 2.25E-04 | **1.41E-03** | 42 | 9182 | -0.01 | 0.00 | 1.00E-03 | **5.23E-03** | 53 | -0.02 | 0.00 | 7.85E-07 | 0.00 | 7.17E-01 | - | - | - |
| Parents or siblings have diabetes | General health | 12837 | -0.03 | 0.01 | 5.76E-04 | **2.91E-03** | 52 | 12703 | -0.03 | 0.01 | 6.28E-04 | **3.70E-03** | 47 | -0.03 | 0.01 | 1.22E-06 | 0.00 | 9.86E-01 | 0.16 | 0.36 | 45 |
| Sodium intake (mg/day) | Nutrients | 11887 | 0.01 | 0.00 | 1.02E-03 | **4.48E-03** | 60 | 11684 | 0.01 | 0.00 | 3.49E-04 | **2.42E-03** | 40 | 0.01 | 0.00 | 1.25E-06 | 0.00 | 7.66E-01 | 0.17 | 0.36 | 29 |
| Physical limitation to participate in moderately demanding activities: walking up several stairs | General health | 9494 | -0.01 | 0.00 | 3.01E-04 | **1.68E-03** | 47 | 9177 | -0.01 | 0.00 | 1.46E-03 | **6.99E-03** | 58 | -0.01 | 0.00 | 1.47E-06 | 0.00 | 7.61E-01 | - | - | - |
| Physical limitation that made you do less than you wanted during the last four weeks | General health | 9511 | -0.04 | 0.01 | 9.23E-05 | **6.93E-04** | 35 | 9191 | -0.03 | 0.01 | 4.20E-03 | **1.64E-02** | 71 | -0.04 | 0.01 | 1.59E-06 | 0.00 | 4.05E-01 | - | - | - |
| Cream, creme fraiche, sour cream | Food | 11888 | 0.02 | 0.00 | 8.36E-06 | **9.99E-05** | 22 | 11685 | 0.01 | 0.00 | 2.73E-02 | 7.42E-02 | 102 | 0.01 | 0.00 | 2.19E-06 | 0.57 | 1.25E-01 | - | - | - |
| Selenium intake (ug/day) | Nutrients | 11888 | 0.01 | 0.00 | 2.81E-04 | **1.61E-03** | 46 | 11685 | 0.01 | 0.00 | 2.40E-03 | **1.02E-02** | 65 | 0.01 | 0.00 | 2.33E-06 | 0.00 | 7.35E-01 | 0.17 | 0.36 | 30 |
| Participation in sports or physical exercise associations | Social | 5971 | 0.04 | 0.01 | 1.76E-05 | **1.93E-04** | 24 | 5640 | 0.02 | 0.01 | 1.98E-02 | 5.48E-02 | 100 | 0.03 | 0.01 | 2.38E-06 | 0.48 | 1.64E-01 | - | - | - |
| Frequency of cycling during leisure time | Physical activity | 10272 | 0.02 | 0.00 | 7.90E-05 | **6.29E-04** | 33 | 10000 | 0.01 | 0.00 | 7.00E-03 | **2.40E-02** | 81 | 0.01 | 0.00 | 2.46E-06 | 0.00 | 4.11E-01 | 0.15 | 0.38 | 48 |
| Arachidonic acid (ARA) intake (g/day) | Nutrients | 11887 | 0.01 | 0.00 | 4.24E-03 | **1.47E-02** | 76 | 11685 | 0.02 | 0.00 | 1.44E-04 | **1.17E-03** | 34 | 0.01 | 0.00 | 2.65E-06 | 0.00 | 4.46E-01 | 0.17 | 0.36 | 26 |
| Physical limitation that reduced the normal time spent at work or in other activities during the last four weeks | General health | 9520 | -0.05 | 0.01 | 6.42E-04 | **3.19E-03** | 53 | 9189 | -0.04 | 0.01 | 1.36E-03 | **6.61E-03** | 57 | -0.04 | 0.01 | 2.85E-06 | 0.00 | 8.16E-01 | - | - | - |
| Bacon | Food | 11888 | 0.01 | 0.00 | 7.02E-05 | **5.77E-04** | 32 | 11685 | 0.01 | 0.00 | 8.77E-03 | **2.82E-02** | 86 | 0.01 | 0.00 | 2.86E-06 | 0.00 | 3.77E-01 | 0.17 | 0.36 | 19 |
| Physical limitation to participate in moderately demanding activities: walking more than 2 km | General health | 9498 | -0.02 | 0.00 | 1.66E-04 | **1.15E-03** | 38 | 9186 | -0.01 | 0.00 | 4.59E-03 | **1.77E-02** | 72 | -0.01 | 0.00 | 2.86E-06 | 0.00 | 4.81E-01 | - | - | - |
| Worsen in health in the future | General health | 9467 | -0.01 | 0.00 | 2.93E-03 | **1.08E-02** | 71 | 9162 | -0.01 | 0.00 | 4.91E-04 | **3.03E-03** | 44 | -0.01 | 0.00 | 4.74E-06 | 0.00 | 6.84E-01 | - | - | - |
| Light beer | Alcohol | 11888 | 0.02 | 0.01 | 1.15E-03 | **4.86E-03** | 62 | 11685 | 0.02 | 0.01 | 1.94E-03 | **8.48E-03** | 63 | 0.02 | 0.00 | 7.04E-06 | 0.00 | 9.80E-01 | 0.16 | 0.36 | 32 |
| Physical limitation to participate in moderately demanding activities: moving a table, vacuuming, walking in the forest or gardening | General health | 9493 | -0.02 | 0.00 | 2.68E-04 | **1.57E-03** | 45 | 9187 | -0.01 | 0.00 | 7.36E-03 | **2.44E-02** | 83 | -0.01 | 0.00 | 7.41E-06 | 0.00 | 4.50E-01 | - | - | - |
| For how much of the time during the last four weeks has your physical health or your emotional problems limited your ability to interact with others? | General health | 9455 | -0.01 | 0.00 | 6.84E-04 | **3.33E-03** | 54 | 9144 | -0.01 | 0.00 | 5.00E-03 | **1.90E-02** | 73 | -0.01 | 0.00 | 1.15E-05 | 0.00 | 6.09E-01 | - | - | - |
| Meat stew | Food | 11888 | 0.01 | 0.00 | 3.62E-03 | **1.31E-02** | 72 | 11685 | 0.01 | 0.00 | 1.05E-03 | **5.36E-03** | 54 | 0.01 | 0.00 | 1.19E-05 | 0.00 | 8.42E-01 | 0.17 | 0.36 | 24 |
| Monounsaturated fat intake (g/day) | Nutrients | 11888 | 0.01 | 0.00 | 1.80E-02 | **4.88E-02** | 97 | 11685 | 0.02 | 0.00 | 1.50E-04 | **1.18E-03** | 35 | 0.01 | 0.00 | 1.46E-05 | 0.17 | 2.72E-01 | 0.16 | 0.36 | 33 |
| Rosehip, sweet syrup soup | Food | 11888 | -0.02 | 0.00 | 4.44E-05 | **4.17E-04** | 28 | 11685 | -0.01 | 0.00 | 3.89E-02 | 9.26E-02 | 116 | -0.01 | 0.00 | 1.48E-05 | 0.55 | 1.38E-01 | - | - | - |
| Formic acid intaje (g/day) | Nutrients | 11888 | 0.01 | 0.00 | 7.01E-04 | **3.35E-03** | 55 | 11685 | 0.01 | 0.00 | 6.68E-03 | **2.32E-02** | 79 | 0.01 | 0.00 | 1.57E-05 | 0.00 | 6.57E-01 | 0.17 | 0.36 | 22 |
| Butter for cooking | Food | 11888 | 0.01 | 0.00 | 1.41E-03 | **5.52E-03** | 67 | 11685 | 0.01 | 0.00 | 6.70E-03 | **2.32E-02** | 80 | 0.01 | 0.00 | 3.00E-05 | 0.00 | 7.10E-01 | 0.17 | 0.36 | 21 |
| Cookies, pastry | Food | 11888 | -0.01 | 0.00 | 1.58E-02 | **4.39E-02** | 95 | 11685 | -0.02 | 0.00 | 4.92E-04 | **3.03E-03** | 45 | -0.01 | 0.00 | 3.01E-05 | 0.00 | 4.54E-01 | 0.17 | 0.36 | 15 |
| Sugar, honey, marmelade, jam | Food | 11888 | -0.01 | 0.00 | 3.64E-03 | **1.31E-02** | 73 | 11685 | -0.01 | 0.00 | 3.00E-03 | **1.20E-02** | 69 | -0.01 | 0.00 | 3.26E-05 | 0.00 | 9.45E-01 | 0.17 | 0.36 | 14 |
| Pain during the last four weeks | General health | 9517 | -0.01 | 0.00 | 4.32E-03 | **1.48E-02** | 77 | 9181 | -0.01 | 0.00 | 3.44E-03 | **1.36E-02** | 70 | -0.01 | 0.00 | 4.29E-05 | 0.00 | 9.58E-01 | - | - | - |
| Snuff status: Snuff users vs non-snuff users | Tobacco use | 11098 | 0.04 | 0.01 | 1.31E-03 | **5.30E-03** | 65 | 11031 | 0.03 | 0.01 | 1.20E-02 | **3.70E-02** | 90 | 0.03 | 0.01 | 5.20E-05 | 0.00 | 6.03E-01 | 0.16 | 0.39 | 34 |
| Fat intake (g/day) | Nutrients | 11888 | 0.01 | 0.00 | 1.10E-02 | **3.14E-02** | 92 | 11685 | 0.01 | 0.00 | 1.84E-03 | **8.35E-03** | 61 | 0.01 | 0.00 | 6.51E-05 | 0.00 | 6.35E-01 | 0.17 | 0.36 | 16 |
| Frequency of walking during leisure time | Physical activity | 12497 | 0.01 | 0.00 | 1.17E-03 | **4.87E-03** | 63 | 12358 | 0.01 | 0.00 | 1.74E-02 | **4.98E-02** | 96 | 0.01 | 0.00 | 6.87E-05 | 0.00 | 5.49E-01 | 0.15 | 0.35 | 47 |
| Folic acid intake (ug/day) | Nutrients | 11888 | 0.01 | 0.00 | 4.75E-04 | **2.50E-03** | 50 | 11685 | 0.01 | 0.00 | 4.03E-02 | 9.39E-02 | 118 | 0.01 | 0.00 | 8.02E-05 | 0.00 | 3.49E-01 | - | - | - |
| Skill demand from job | Psychosocial | 12897 | 0.01 | 0.00 | 5.55E-04 | **2.86E-03** | 51 | 12729 | 0.01 | 0.00 | 3.45E-02 | 8.53E-02 | 112 | 0.01 | 0.00 | 8.18E-05 | 0.00 | 3.47E-01 | - | - | - |
| Heptadecanoic acid intake (g/day) | Nutrients | 11888 | 0.01 | 0.00 | 7.35E-04 | **3.39E-03** | 56 | 11685 | 0.01 | 0.00 | 3.19E-02 | 8.11E-02 | 108 | 0.01 | 0.00 | 9.10E-05 | 0.00 | 4.04E-01 | - | - | - |
| Pentadecanoic acid intake (g/day) | Nutrients | 11888 | 0.01 | 0.00 | 7.35E-04 | **3.39E-03** | 57 | 11685 | 0.01 | 0.00 | 3.19E-02 | 8.11E-02 | 109 | 0.01 | 0.00 | 9.10E-05 | 0.00 | 4.04E-01 | - | - | - |
| Saturated fat intake (g/day) | Nutrients | 11888 | 0.01 | 0.00 | 4.67E-03 | **1.56E-02** | 78 | 11685 | 0.01 | 0.00 | 7.12E-03 | **2.41E-02** | 82 | 0.01 | 0.00 | 9.45E-05 | 0.00 | 9.54E-01 | 0.17 | 0.36 | 17 |
| Appetite status | Psychosocial | 11718 | 0.01 | 0.00 | 4.54E-02 | 1.04E-01 | 115 | 11491 | 0.01 | 0.00 | 4.60E-04 | **2.96E-03** | 43 | 0.01 | 0.00 | 1.06E-04 | 0.22 | 2.59E-01 | - | - | - |
| Animal based protein intake (g/day) | Nutrients | 11888 | 0.01 | 0.00 | 2.30E-02 | 6.05E-02 | 100 | 11685 | 0.01 | 0.00 | 1.96E-03 | **8.48E-03** | 64 | 0.01 | 0.00 | 1.53E-04 | 0.00 | 5.12E-01 | - | - | - |
| Overall state of health compared to other your age | General health | 3622 | 0.03 | 0.01 | 1.21E-03 | **4.99E-03** | 64 | 3775 | 0.02 | 0.01 | 4.43E-02 | 1.02E-01 | 120 | 0.02 | 0.01 | 1.90E-04 | 0.00 | 3.84E-01 | - | - | - |
| Physical limitation that made you not being able to perform certain work tasks or other activities during the last four weeks | General health | 9470 | -0.03 | 0.01 | 4.67E-03 | **1.56E-02** | 79 | 9158 | -0.03 | 0.01 | 1.56E-02 | **4.55E-02** | 95 | -0.03 | 0.01 | 2.05E-04 | 0.00 | 7.37E-01 | - | - | - |
| Long-term sickness | General health | 12769 | -0.03 | 0.01 | 6.94E-03 | **2.15E-02** | 85 | 12639 | -0.03 | 0.01 | 1.09E-02 | **3.38E-02** | 89 | -0.03 | 0.01 | 2.07E-04 | 0.00 | 9.05E-01 | 0.16 | 0.36 | 40 |
| Average portion size of meat/fish based on photographic illustration of four sizes (smallest to largest) | Food | 11888 | 0.01 | 0.00 | 1.97E-01 | 3.18E-01 | 163 | 11685 | 0.02 | 0.00 | 1.06E-04 | **9.83E-04** | 30 | 0.01 | 0.00 | 2.95E-04 | 0.72 | 5.72E-02 | - | - | - |
| For how much of the time during the last four weeks have you felt worn out? | General health | 9498 | -0.01 | 0.00 | 8.41E-03 | **2.49E-02** | 89 | 9157 | -0.01 | 0.00 | 1.51E-02 | **4.55E-02** | 91 | -0.01 | 0.00 | 3.37E-04 | 0.00 | 8.73E-01 | - | - | - |
| White meat (poultry) | Food | 11888 | 0.01 | 0.00 | 1.58E-01 | 2.69E-01 | 154 | 11685 | 0.01 | 0.00 | 2.40E-04 | **1.70E-03** | 39 | 0.01 | 0.00 | 3.54E-04 | 0.64 | 9.72E-02 | - | - | - |
| Cheese 28% | Food | 11888 | 0.01 | 0.00 | 5.24E-02 | 1.17E-01 | 118 | 11685 | 0.01 | 0.00 | 1.91E-03 | **8.48E-03** | 62 | 0.01 | 0.00 | 3.75E-04 | 0.00 | 3.86E-01 | - | - | - |
| Extent to what your physical and emotional health disrupted your usual social life during the last four weeks | General health | 9517 | -0.01 | 0.00 | 2.39E-02 | 6.24E-02 | 101 | 9195 | -0.01 | 0.00 | 6.47E-03 | **2.30E-02** | 78 | -0.01 | 0.00 | 4.12E-04 | 0.00 | 7.65E-01 | - | - | - |
| Steak, chop, e.g. | Food | 11888 | 0.01 | 0.00 | 9.80E-03 | **2.85E-02** | 90 | 11685 | 0.01 | 0.00 | 1.79E-02 | 5.05E-02 | 98 | 0.01 | 0.00 | 4.59E-04 | 0.00 | 9.48E-01 | - | - | - |
| Smoked fish/meat | Food | 11888 | 0.00 | 0.00 | 2.02E-01 | 3.21E-01 | 166 | 11685 | 0.01 | 0.00 | 1.42E-04 | **1.17E-03** | 33 | 0.01 | 0.00 | 4.71E-04 | 0.74 | 4.82E-02 | - | - | - |
| Brewed (filtered) coffee | Beverage | 11888 | 0.01 | 0.00 | 8.79E-02 | 1.76E-01 | 131 | 11685 | 0.01 | 0.00 | 1.16E-03 | **5.82E-03** | 55 | 0.01 | 0.00 | 4.84E-04 | 0.24 | 2.53E-01 | - | - | - |
| Number of snuff boxes per week | Tobacco use | 12805 | 0.01 | 0.00 | 1.12E-03 | **4.83E-03** | 61 | 12664 | 0.01 | 0.00 | 9.63E-02 | 1.90E-01 | 140 | 0.01 | 0.00 | 4.93E-04 | 0.20 | 2.63E-01 | - | - | - |
| Tomato, cucumber | Food | 11888 | 0.01 | 0.00 | 7.45E-03 | **2.25E-02** | 87 | 11685 | 0.01 | 0.00 | 3.08E-02 | 8.08E-02 | 105 | 0.01 | 0.00 | 6.19E-04 | 0.00 | 7.38E-01 | - | - | - |
| Physical limitation that limited your ability to perform certain work tasks or other activities during the last four weeks | General health | 9487 | -0.03 | 0.01 | 3.99E-03 | **1.42E-02** | 74 | 9163 | -0.02 | 0.01 | 5.15E-02 | 1.18E-01 | 121 | -0.03 | 0.01 | 6.30E-04 | 0.00 | 4.77E-01 | - | - | - |
| Trans fat intake (g/day) | Nutrients | 11888 | 0.02 | 0.01 | 6.45E-03 | **2.04E-02** | 83 | 11685 | 0.02 | 0.01 | 3.73E-02 | 8.99E-02 | 115 | 0.02 | 0.01 | 6.73E-04 | 0.00 | 6.58E-01 | - | - | - |

| | | | | | | | | | | | | | | | | | | | | | |
|---|---|---|---|---|---|---|---|---|---|---|---|---|---|---|---|---|---|---|---|---|---|
| Do you feel important and appreciated outside your home? | Psychosocial | 11721 | 0.01 | 0.00 | 4.15E-02 | 9.75E-02 | 112 | 11486 | 0.01 | 0.00 | 6.13E-03 | **2.23E-02** | 76 | 0.01 | 0.00 | 7.37E-04 | 0.00 | 5.98E-01 | - | - | - |
| Learn new things at job | Psychosocial | 12915 | 0.01 | 0.00 | 2.24E-03 | **8.42E-03** | 70 | 12722 | 0.01 | 0.00 | 9.50E-02 | 1.89E-01 | 139 | 0.01 | 0.00 | 8.27E-04 | 0.00 | 3.28E-01 | - | - | - |
| Sodas, soft drinks, juice | Beverage | 11888 | -0.01 | 0.00 | 4.61E-02 | 1.04E-01 | 116 | 11685 | -0.01 | 0.00 | 7.40E-03 | **2.44E-02** | 84 | -0.01 | 0.00 | 9.74E-04 | 0.00 | 5.99E-01 | - | - | - |
| Salty fish | Food | 11888 | 0.01 | 0.00 | 1.21E-02 | **3.44E-02** | 93 | 11685 | 0.01 | 0.00 | 3.24E-02 | 8.17E-02 | 110 | 0.01 | 0.00 | 9.80E-04 | 0.00 | 9.65E-01 | - | - | - |
| Frequent social contacts with colleagues during work | Psychosocial | 12812 | 0.01 | 0.00 | 1.40E-03 | **5.52E-03** | 66 | 12623 | 0.01 | 0.00 | 1.49E-01 | 2.62E-01 | 158 | 0.01 | 0.00 | 1.02E-03 | 0.33 | 2.21E-01 | - | - | - |
| Participation in other association | Social | 5971 | -0.01 | 0.01 | 1.78E-01 | 2.95E-01 | 159 | 5640 | -0.03 | 0.01 | 9.75E-04 | **5.23E-03** | 51 | -0.02 | 0.01 | 1.03E-03 | 0.52 | 1.48E-01 | - | - | - |
| Low fat margarine on bread | Food | 11888 | -0.01 | 0.00 | 1.98E-03 | **7.64E-03** | 68 | 11685 | -0.01 | 0.00 | 1.36E-01 | 2.44E-01 | 154 | -0.01 | 0.00 | 1.09E-03 | 0.13 | 2.84E-01 | - | - | - |
| Corn flakes | Food | 11888 | -0.01 | 0.00 | 1.58E-02 | **4.39E-02** | 94 | 11685 | -0.01 | 0.00 | 2.96E-02 | 7.90E-02 | 104 | -0.01 | 0.00 | 1.17E-03 | 0.00 | 9.04E-01 | - | - | - |
| Do you feel important and appreciated in your home? | Psychosocial | 11641 | 0.01 | 0.00 | 1.88E-01 | 3.08E-01 | 161 | 11399 | 0.01 | 0.00 | 1.49E-03 | **6.99E-03** | 59 | 0.01 | 0.00 | 1.65E-03 | 0.48 | 1.64E-01 | - | - | - |
| Total protein intake (g/day) | Nutrients | 11888 | 0.01 | 0.00 | 9.70E-02 | 1.90E-01 | 134 | 11685 | 0.01 | 0.00 | 6.04E-03 | **2.23E-02** | 75 | 0.01 | 0.00 | 1.93E-03 | 0.00 | 4.09E-01 | - | - | - |
| Sum of Lariciresinol, Matairesinol, Pinoresinol, Secoisolariciresinol intake (ug/day) | Nutrients | 11888 | 0.01 | 0.00 | 6.22E-03 | **1.99E-02** | 82 | 11685 | 0.01 | 0.00 | 1.12E-01 | 2.13E-01 | 146 | 0.01 | 0.00 | 2.12E-03 | 0.00 | 4.48E-01 | - | - | - |
| Breakfast habits: Only coffee/tea for breakfast vs not breakfast at all | Food | 871 | 0.05 | 0.04 | 1.49E-01 | 2.61E-01 | 150 | 928 | 0.09 | 0.03 | 5.39E-03 | **2.02E-02** | 74 | 0.07 | 0.02 | 2.42E-03 | 0.00 | 3.75E-01 | - | - | - |
| Average portion size of potatoes/rice/pasta based on photographic illustration of four sizes (smallest to largest) | Food | 11888 | -0.01 | 0.00 | 9.86E-03 | **2.85E-02** | 91 | 11685 | -0.01 | 0.00 | 1.14E-01 | 2.13E-01 | 148 | -0.01 | 0.00 | 3.13E-03 | 0.00 | 5.11E-01 | - | - | - |
| Vitamin B12 intake (ug/day) | Nutrients | 11888 | 0.01 | 0.00 | 4.21E-02 | 9.81E-02 | 113 | 11685 | 0.01 | 0.00 | 3.29E-02 | 8.22E-02 | 111 | 0.01 | 0.00 | 3.24E-03 | 0.00 | 9.07E-01 | - | - | - |
| Monosaccharides intake (g/day) | Nutrients | 11888 | -0.01 | 0.00 | 2.55E-01 | 3.81E-01 | 176 | 11685 | -0.01 | 0.00 | 2.56E-03 | **1.06E-02** | 67 | -0.01 | 0.00 | 3.49E-03 | 0.47 | 1.70E-01 | - | - | - |
| Satisfaction with economy | Psychosocial | 11719 | 0.01 | 0.00 | 2.52E-02 | 6.49E-02 | 102 | 11477 | 0.01 | 0.00 | 6.13E-02 | 1.33E-01 | 128 | 0.01 | 0.00 | 3.62E-03 | 0.00 | 8.23E-01 | - | - | - |
| Breakfast habits: Porridge w/o sandwich for breakfast vs not breakfast at all | Food | 1854 | 0.04 | 0.03 | 1.12E-01 | 2.06E-01 | 143 | 1823 | 0.08 | 0.03 | 1.03E-02 | **3.24E-02** | 88 | 0.06 | 0.02 | 3.69E-03 | 0.00 | 3.81E-01 | - | - | - |
| Magnesium intake (mg/day) | Nutrients | 11888 | 0.01 | 0.00 | 4.86E-03 | **1.58E-02** | 81 | 11685 | 0.01 | 0.00 | 2.20E-01 | 3.57E-01 | 170 | 0.01 | 0.00 | 3.97E-03 | 0.13 | 2.84E-01 | - | - | - |
| Parents or siblings had a cerebral hemorrhage/thrombosis or cardiac infarction before the age of 60 | General health | 12842 | -0.01 | 0.01 | 1.52E-01 | 2.64E-01 | 152 | 12699 | -0.03 | 0.01 | 7.92E-03 | **2.58E-02** | 85 | -0.02 | 0.01 | 3.99E-03 | 0.00 | 3.66E-01 | - | - | - |
| Minced meat dishes | Food | 11888 | 0.00 | 0.00 | 4.66E-01 | 5.92E-01 | 207 | 11685 | 0.01 | 0.00 | 9.84E-04 | **5.23E-03** | 52 | 0.01 | 0.00 | 4.33E-03 | 0.69 | 7.05E-02 | - | - | - |
| Milk, sour milk (1,5%) | Beverage | 11888 | -0.01 | 0.00 | 6.35E-02 | 1.37E-01 | 122 | 11685 | -0.01 | 0.00 | 2.93E-02 | 7.87E-02 | 103 | -0.01 | 0.00 | 4.38E-03 | 0.00 | 7.83E-01 | - | - | - |
| Job demands to work very fast | Psychosocial | 12940 | 0.01 | 0.00 | 3.00E-02 | 7.52E-02 | 105 | 12774 | 0.01 | 0.00 | 6.50E-02 | 1.38E-01 | 131 | 0.01 | 0.00 | 4.51E-03 | 0.00 | 8.32E-01 | - | - | - |
| Equol intake (ug/day) | Nutrients | 11888 | -0.01 | 0.00 | 4.22E-03 | **1.47E-02** | 75 | 11685 | 0.00 | 0.00 | 2.73E-01 | 4.17E-01 | 181 | -0.01 | 0.00 | 4.59E-03 | 0.27 | 2.42E-01 | - | - | - |
| Matairesinol intake (ug/day) | Nutrients | 11888 | 0.01 | 0.00 | 6.54E-03 | **2.05E-02** | 84 | 11685 | 0.00 | 0.00 | 2.47E-01 | 3.85E-01 | 177 | 0.01 | 0.00 | 5.86E-03 | 0.13 | 2.84E-01 | - | - | - |
| Travel to work: Walk to work vs passive travel to work | Physical activity | 8657 | 0.02 | 0.01 | 8.44E-02 | 1.73E-01 | 128 | 8640 | 0.03 | 0.01 | 3.09E-02 | 8.08E-02 | 106 | 0.03 | 0.01 | 5.91E-03 | 0.00 | 8.05E-01 | - | - | - |
| Snuff status: Former snuff users vs non-snuff users | Tobacco use | 10540 | 0.02 | 0.01 | 1.27E-01 | 2.26E-01 | 147 | 10284 | 0.03 | 0.01 | 1.74E-02 | **4.98E-02** | 97 | 0.02 | 0.01 | 6.07E-03 | 0.00 | 4.99E-01 | - | - | - |
| Ingenuity or creativity demand from job | Psychosocial | 12871 | 0.02 | 0.00 | 5.91E-05 | **5.18E-04** | 30 | 12695 | 0.00 | 0.00 | 8.65E-01 | 9.01E-01 | 266 | 0.01 | 0.00 | 6.10E-03 | 0.88 | 3.19E-03 | - | - | - |
| Number of social contacts with the same interests as you | Social | 13133 | 0.00 | 0.00 | 3.69E-01 | 5.01E-01 | 194 | 12960 | 0.01 | 0.00 | 2.96E-03 | **1.20E-02** | 68 | 0.01 | 0.00 | 6.43E-03 | 0.55 | 1.36E-01 | - | - | - |
| Total energy intake (kcal/day) | Nutrients | 11888 | -0.01 | 0.00 | 7.80E-03 | **2.33E-02** | 88 | 11685 | -0.01 | 0.00 | 2.46E-01 | 3.85E-01 | 176 | -0.01 | 0.00 | 6.74E-03 | 0.08 | 2.96E-01 | - | - | - |
| Hearing status | General health | 11723 | 0.01 | 0.00 | 3.70E-02 | 8.86E-02 | 110 | 11495 | 0.01 | 0.00 | 8.48E-02 | 1.74E-01 | 135 | 0.01 | 0.00 | 6.99E-03 | 0.00 | 8.32E-01 | - | - | - |
| Brown beans, pea soup | Food | 11888 | 0.00 | 0.00 | 1.63E-01 | 2.77E-01 | 155 | 11685 | 0.01 | 0.00 | 1.56E-02 | **4.55E-02** | 93 | 0.01 | 0.00 | 7.75E-03 | 0.00 | 4.00E-01 | - | - | - |
| Memory status | Psychosocial | 11694 | 0.01 | 0.00 | 3.21E-02 | 7.88E-02 | 107 | 11449 | 0.01 | 0.00 | 1.09E-01 | 2.07E-01 | 145 | 0.01 | 0.00 | 7.89E-03 | 0.00 | 7.36E-01 | - | - | - |
| Salad dressing with oil | Food | 11888 | 0.01 | 0.00 | 1.06E-01 | 2.01E-01 | 137 | 11685 | 0.01 | 0.00 | 3.15E-02 | 8.11E-02 | 107 | 0.01 | 0.00 | 7.91E-03 | 0.00 | 6.59E-01 | - | - | - |
| Physical limitation to participate in moderately demanding activities: bending down or kneeling | General health | 9486 | -0.01 | 0.00 | 1.08E-01 | 2.03E-01 | 140 | 9167 | -0.01 | 0.00 | 4.03E-02 | 9.39E-02 | 119 | -0.01 | 0.00 | 9.62E-03 | 0.00 | 7.36E-01 | - | - | - |
| Boiled or baked potato | Food | 11888 | -0.01 | 0.00 | 8.40E-02 | 1.73E-01 | 127 | 11685 | -0.01 | 0.00 | 5.36E-02 | 1.20E-01 | 124 | -0.01 | 0.00 | 9.75E-03 | 0.00 | 8.58E-01 | - | - | - |
| Plant based protein intake (g/day) | Nutrients | 11888 | -0.01 | 0.00 | 4.31E-02 | 9.95E-02 | 114 | 11685 | -0.01 | 0.00 | 1.07E-01 | 2.06E-01 | 144 | -0.01 | 0.00 | 1.00E-02 | 0.00 | 8.16E-01 | - | - | - |
| Everyday exercise satisfaction | Physical activity | 4455 | 0.01 | 0.01 | 7.03E-02 | 1.49E-01 | 124 | 4584 | 0.01 | 0.01 | 8.91E-02 | 1.81E-01 | 136 | 0.01 | 0.01 | 1.31E-02 | 0.00 | 9.12E-01 | - | - | - |
| Satisfaction with leisure time | Psychosocial | 11700 | 0.01 | 0.00 | 6.44E-02 | 1.38E-01 | 123 | 11465 | 0.01 | 0.00 | 9.76E-02 | 1.90E-01 | 142 | 0.01 | 0.00 | 1.31E-02 | 0.00 | 9.30E-01 | - | - | - |
| Permanent employment | Psychosocial | 11542 | 0.03 | 0.01 | 4.80E-03 | **1.58E-02** | 80 | 11421 | 0.01 | 0.01 | 5.27E-01 | 6.54E-01 | 223 | 0.02 | 0.01 | 1.40E-02 | 0.57 | 1.27E-01 | - | - | - |
| Pinoresinol intake (ug/day) | Nutrients | 11888 | 0.01 | 0.00 | 1.70E-02 | **4.65E-02** | 96 | 11685 | 0.00 | 0.00 | 2.92E-01 | 4.30E-01 | 188 | 0.01 | 0.00 | 1.43E-02 | 0.00 | 3.69E-01 | - | - | - |
| Teetotaler | Alcohol | 4461 | -0.05 | 0.03 | 7.34E-02 | 1.54E-01 | 125 | 4598 | -0.04 | 0.03 | 9.66E-02 | 1.90E-01 | 141 | -0.04 | 0.02 | 1.46E-02 | 0.00 | 9.40E-01 | - | - | - |
| Low fat milk (0,5%) | Beverage | 11888 | -0.01 | 0.00 | 4.10E-02 | 9.73E-02 | 111 | 11685 | -0.01 | 0.00 | 1.66E-01 | 2.90E-01 | 159 | -0.01 | 0.00 | 1.49E-02 | 0.00 | 6.85E-01 | - | - | - |
| Calcium intake (mg/day) | Nutrients | 11888 | -0.01 | 0.00 | 1.06E-01 | 2.01E-01 | 138 | 11685 | -0.01 | 0.00 | 6.70E-02 | 1.41E-01 | 132 | -0.01 | 0.00 | 1.50E-02 | 0.00 | 8.31E-01 | - | - | - |
| Phosphate intake (mg/day) | Nutrients | 11888 | 0.01 | 0.00 | 3.07E-02 | 7.62E-02 | 106 | 11685 | 0.01 | 0.00 | 2.13E-01 | 3.53E-01 | 167 | 0.01 | 0.00 | 1.54E-02 | 0.00 | 5.54E-01 | - | - | - |
| Vitamin C intake (mg/day) | Nutrients | 11888 | -0.01 | 0.00 | 1.50E-01 | 2.61E-01 | 151 | 11685 | -0.01 | 0.00 | 5.23E-02 | 1.18E-01 | 122 | -0.01 | 0.00 | 1.72E-02 | 0.00 | 6.86E-01 | - | - | - |
| For how much of the time during the last four weeks have you felt happy? | General health | 9510 | 0.00 | 0.00 | 9.75E-01 | 9.83E-01 | 261 | 9184 | 0.01 | 0.00 | 9.46E-04 | **5.23E-03** | 49 | 0.01 | 0.00 | 1.79E-02 | 0.83 | 1.50E-02 | - | - | - |
| Patience status | Psychosocial | 11710 | 0.00 | 0.00 | 3.54E-01 | 4.82E-01 | 193 | 11472 | 0.01 | 0.00 | 1.56E-02 | **4.55E-02** | 94 | 0.01 | 0.00 | 1.91E-02 | 0.18 | 2.70E-01 | - | - | - |
| Frequncy of social contacts with colleagues during leisure time | Psychosocial | 12557 | 0.01 | 0.00 | 1.67E-01 | 2.81E-01 | 156 | 12375 | 0.01 | 0.00 | 6.20E-02 | 1.33E-01 | 129 | 0.01 | 0.00 | 2.16E-02 | 0.00 | 7.28E-01 | - | - | - |
| Frequency of shoverling snow during leisure time | Physical activity | 4229 | 0.01 | 0.01 | 4.29E-01 | 5.58E-01 | 202 | 4354 | 0.02 | 0.01 | 1.82E-02 | 5.08E-02 | 99 | 0.01 | 0.01 | 2.49E-02 | 0.17 | 2.73E-01 | - | - | - |
| Beta-carotene intake (mg/day) | Nutrients | 11888 | 0.01 | 0.00 | 5.11E-02 | 1.15E-01 | 117 | 11685 | 0.01 | 0.00 | 2.41E-01 | 3.81E-01 | 175 | 0.01 | 0.00 | 2.63E-02 | 0.00 | 6.19E-01 | - | - | - |
| Cohabitation: Live alone vs Only one adult (spouse, partner) | Social | 7413 | -0.03 | 0.01 | 3.42E-02 | 8.33E-02 | 108 | 7269 | -0.01 | 0.01 | 3.11E-01 | 4.48E-01 | 192 | -0.02 | 0.01 | 2.74E-02 | 0.00 | 4.20E-01 | - | - | - |
| Sweets | Food | 11888 | -0.01 | 0.00 | 1.03E-01 | 2.00E-01 | 135 | 11685 | -0.01 | 0.00 | 1.40E-01 | 2.51E-01 | 155 | -0.01 | 0.00 | 2.81E-02 | 0.00 | 9.22E-01 | - | - | - |
| Feel uneasy or guilty because of your way of drinking | Alcohol | 3979 | 0.05 | 0.03 | 5.95E-02 | 1.29E-01 | 121 | 4103 | 0.03 | 0.03 | 2.18E-01 | 3.57E-01 | 168 | 0.04 | 0.02 | 2.84E-02 | 0.00 | 6.00E-01 | - | - | - |
| Sour milk, yoghurt (3% fat) | Food | 11888 | 0.01 | 0.00 | 7.32E-03 | **2.24E-02** | 86 | 11685 | 0.00 | 0.00 | 7.13E-01 | 8.02E-01 | 246 | 0.01 | 0.00 | 2.85E-02 | 0.61 | 1.11E-01 | - | - | - |
| Number of social interactions during a normal week | Social | 13146 | 0.00 | 0.00 | 2.49E-01 | 3.75E-01 | 175 | 12975 | 0.01 | 0.00 | 5.25E-02 | 1.18E-01 | 123 | 0.01 | 0.00 | 2.91E-02 | 0.00 | 5.68E-01 | - | - | - |
| Number of friends that can come to your home at any time and feel at home | Social | 13085 | 0.00 | 0.00 | 2.15E-01 | 3.38E-01 | 167 | 12938 | 0.01 | 0.00 | 7.69E-02 | 1.60E-01 | 133 | 0.01 | 0.00 | 3.37E-02 | 0.00 | 6.92E-01 | - | - | - |
| Participation in associations or voluntary organisations | Social | 13009 | 0.01 | 0.01 | 2.78E-01 | 4.09E-01 | 178 | 12868 | 0.01 | 0.01 | 6.05E-02 | 1.32E-01 | 127 | 0.01 | 0.01 | 3.67E-02 | 0.00 | 5.60E-01 | - | - | - |
| Campesterol intake (mg/day) | Nutrients | 11888 | -0.01 | 0.00 | 1.98E-01 | 3.18E-01 | 164 | 11685 | -0.01 | 0.00 | 1.13E-01 | 2.13E-01 | 147 | -0.01 | 0.00 | 4.28E-02 | 0.00 | 8.01E-01 | - | - | - |
| Work shifts/weekends | Psychosocial | 12800 | -0.01 | 0.01 | 1.06E-01 | 2.01E-01 | 139 | 12633 | -0.01 | 0.01 | 2.22E-01 | 3.58E-01 | 172 | -0.01 | 0.01 | 4.47E-02 | 0.00 | 7.92E-01 | - | - | - |
| Iodine intake (ug/day) | Nutrients | 11888 | 0.01 | 0.00 | 1.90E-01 | 3.08E-01 | 162 | 11685 | 0.01 | 0.00 | 1.28E-01 | 2.33E-01 | 153 | 0.01 | 0.00 | 4.56E-02 | 0.00 | 8.50E-01 | - | - | - |
| Margarine for cooking | Food | 11888 | -0.01 | 0.00 | 1.90E-02 | 5.09E-02 | 98 | 11685 | 0.00 | 0.00 | 6.71E-01 | 7.83E-01 | 237 | -0.01 | 0.00 | 4.71E-02 | 0.43 | 1.86E-01 | - | - | - |
| Banana | Food | 11888 | 0.00 | 0.00 | 3.00E-01 | 4.27E-01 | 185 | 11685 | -0.01 | 0.00 | 9.40E-02 | 1.89E-01 | 138 | -0.01 | 0.00 | 5.57E-02 | 0.00 | 6.40E-01 | - | - | - |
| Lean fish (e.g. perch, bass, cod) | Food | 11888 | 0.01 | 0.00 | 1.15E-01 | 2.10E-01 | 144 | 11685 | 0.00 | 0.00 | 2.87E-01 | 4.26E-01 | 186 | 0.01 | 0.00 | 6.09E-02 | 0.00 | 7.39E-01 | - | - | - |
| Root vegetables, carrot | Food | 11888 | 0.01 | 0.00 | 8.68E-02 | 1.76E-01 | 130 | 11685 | 0.00 | 0.00 | 3.67E-01 | 5.14E-01 | 198 | 0.01 | 0.00 | 6.24E-02 | 0.00 | 5.99E-01 | - | - | - |
| Enterodiol intake (ug/day) | Nutrients | 11888 | -0.01 | 0.00 | 1.16E-01 | 2.11E-01 | 145 | 11685 | 0.00 | 0.00 | 3.01E-01 | 4.41E-01 | 189 | -0.01 | 0.00 | 6.41E-02 | 0.00 | 7.40E-01 | - | - | - |
| Smoking status: Former occasional smokers vs non-smokers | Tobacco use | 7883 | 0.01 | 0.01 | 2.78E-01 | 4.09E-01 | 179 | 7749 | 0.02 | 0.01 | 1.28E-01 | 2.33E-01 | 152 | 0.02 | 0.01 | 6.62E-02 | 0.00 | 7.37E-01 | - | - | - |
| Mood status | Psychosocial | 11706 | 0.00 | 0.00 | 9.17E-01 | 9.57E-01 | 252 | 11472 | 0.01 | 0.00 | 1.53E-02 | **4.55E-02** | 92 | 0.00 | 0.00 | 7.95E-02 | 0.65 | 9.24E-02 | - | - | - |
| Repetitive job | Psychosocial | 12917 | -0.01 | 0.00 | 1.28E-01 | 2.27E-01 | 149 | 12743 | 0.00 | 0.00 | 3.47E-01 | 4.89E-01 | 197 | 0.00 | 0.00 | 8.15E-02 | 0.00 | 6.89E-01 | - | - | - |
| Satisfaction with work situation | Psychosocial | 11586 | 0.01 | 0.00 | 8.58E-02 | 1.75E-01 | 129 | 11329 | 0.00 | 0.00 | 4.78E-01 | 6.10E-01 | 217 | 0.00 | 0.00 | 8.37E-02 | 0.00 | 4.94E-01 | - | - | - |
| Cohabitation: Live alone vs Adult and children | Social | 6470 | -0.03 | 0.01 | 2.20E-02 | 5.85E-02 | 99 | 6517 | 0.00 | 0.02 | 9.39E-01 | 9.50E-01 | 273 | -0.02 | 0.01 | 9.22E-02 | 0.59 | 1.19E-01 | - | - | - |
| White (soft) bread, thin crisp bread | Food | 11888 | -0.01 | 0.00 | 2.01E-01 | 3.20E-01 | 165 | 11685 | -0.01 | 0.01 | 2.75E-01 | 4.19E-01 | 182 | -0.01 | 0.00 | 9.33E-02 | 0.00 | 9.08E-01 | - | - | - |
| Sour milk, yoghurt (low fat) | Food | 11888 | -0.01 | 0.00 | 2.76E-02 | 7.05E-02 | 103 | 11685 | 0.00 | 0.00 | 9.38E-01 | 9.50E-01 | 272 | 0.00 | 0.00 | 9.67E-02 | 0.53 | 1.47E-01 | - | - | - |
| Vitamin A intake (mg/day) | Nutrients | 11888 | -0.01 | 0.01 | 3.02E-01 | 4.28E-01 | 186 | 11685 | -0.01 | 0.01 | 2.02E-01 | 3.39E-01 | 165 | -0.01 | 0.00 | 1.02E-01 | 0.00 | 8.90E-01 | - | - | - |
| Vision status | General health | 11715 | 0.00 | 0.00 | 2.41E-01 | 3.66E-01 | 173 | 11475 | 0.00 | 0.00 | 2.72E-01 | 4.17E-01 | 180 | 0.00 | 0.00 | 1.08E-01 | 0.00 | 9.86E-01 | - | - | - |
| Cheese 10-17% | Food | 11888 | 0.00 | 0.00 | 8.02E-01 | 8.64E-01 | 244 | 11685 | -0.01 | 0.00 | 3.91E-02 | 9.26E-02 | 117 | -0.01 | 0.00 | 1.09E-01 | 0.43 | 1.84E-01 | - | - | - |
| Polyunsaturated fat intake (g/day) | Nutrients | 11888 | 0.00 | 0.00 | 2.43E-01 | 3.67E-01 | 174 | 11685 | 0.00 | 0.00 | 2.78E-01 | 4.20E-01 | 183 | 0.00 | 0.00 | 1.11E-01 | 0.00 | 9.86E-01 | - | - | - |
| Sedentary or standing work | Physical activity | 12648 | -0.01 | 0.01 | 2.16E-01 | 3.38E-01 | 168 | 12476 | -0.01 | 0.01 | 3.12E-01 | 4.48E-01 | 193 | -0.01 | 0.01 | 1.12E-01 | 0.00 | 8.82E-01 | - | - | - |
| Meat on bread | Food | 11888 | -0.01 | 0.00 | 1.27E-01 | 2.26E-01 | 148 | 11685 | 0.00 | 0.00 | 4.88E-01 | 6.17E-01 | 219 | 0.00 | 0.00 | 1.14E-01 | 0.00 | 5.78E-01 | - | - | - |
| People to ask for help apart from the ones at home | Social | 13092 | 0.00 | 0.01 | 9.28E-01 | 9.65E-01 | 253 | 12928 | 0.03 | 0.01 | 2.23E-02 | 6.11E-02 | 101 | 0.01 | 0.01 | 1.21E-01 | 0.65 | 9.20E-02 | - | - | - |
| Last time a colleague visited you at home | Psychosocial | 12774 | 0.00 | 0.00 | 3.81E-01 | 5.14E-01 | 195 | 12607 | 0.01 | 0.00 | 1.93E-01 | 3.29E-01 | 163 | 0.00 | 0.00 | 1.24E-01 | 0.00 | 7.54E-01 | - | - | - |
| Possibility to speak with colleagues during breaks | Psychosocial | 12892 | 0.01 | 0.00 | 5.89E-02 | 1.29E-01 | 120 | 12728 | 0.00 | 0.00 | 7.84E-01 | 8.46E-01 | 256 | 0.00 | 0.00 | 1.25E-01 | 0.23 | 2.56E-01 | - | - | - |
| Control over own work assignment | Psychosocial | 12934 | -0.01 | 0.00 | 1.09E-01 | 2.03E-01 | 141 | 12771 | 0.00 | 0.00 | 6.03E-01 | 7.17E-01 | 233 | 0.00 | 0.00 | 1.32E-01 | 0.00 | 4.49E-01 | - | - | - |
| Tiamin intake (mg/day) | Nutrients | 11888 | 0.01 | 0.00 | 1.25E-01 | 2.25E-01 | 146 | 11685 | 0.00 | 0.00 | 5.69E-01 | 6.98E-01 | 226 | 0.00 | 0.00 | 1.35E-01 | 0.00 | 5.05E-01 | - | - | - |
| Fiber cereals | Food | 11888 | 0.01 | 0.00 | 5.50E-02 | 1.22E-01 | 119 | 11685 | 0.00 | 0.00 | 8.75E-01 | 9.01E-01 | 269 | 0.00 | 0.00 | 1.37E-01 | 0.33 | 2.21E-01 | - | - | - |
| Butter on bread | Food | 11888 | 0.00 | 0.00 | 9.65E-01 | 9.81E-01 | 258 | 11685 | 0.01 | 0.00 | 3.61E-02 | 8.85E-02 | 113 | 0.00 | 0.00 | 1.51E-01 | 0.57 | 1.27E-01 | - | - | - |
| Boiled coffee | Beverage | 11888 | 0.00 | 0.00 | 2.90E-01 | 4.19E-01 | 182 | 11685 | 0.00 | 0.00 | 3.44E-01 | 4.86E-01 | 196 | 0.00 | 0.00 | 1.56E-01 | 0.00 | 9.47E-01 | - | - | - |
| Lariciresinol intake (ug/day) | Nutrients | 11888 | 0.01 | 0.00 | 9.30E-02 | 1.84E-01 | 133 | 11685 | 0.00 | 0.00 | 7.69E-01 | 8.41E-01 | 253 | 0.00 | 0.00 | 1.57E-01 | 0.00 | 3.40E-01 | - | - | - |
| Pancake, waffle, Swedish dumpling | Food | 11888 | 0.00 | 0.00 | 7.20E-01 | 8.10E-01 | 234 | 11685 | -0.01 | 0.00 | 1.02E-01 | 1.98E-01 | 143 | 0.00 | 0.00 | 1.65E-01 | 0.00 | 3.50E-01 | - | - | - |
| Support from others | Social | 13148 | 0.00 | 0.00 | 5.08E-01 | 6.31E-01 | 212 | 12981 | 0.00 | 0.00 | 1.95E-01 | 3.30E-01 | 164 | 0.00 | 0.00 | 1.67E-01 | 0.00 | 6.54E-01 | - | - | - |
| Whole grain intake (g/day) | Food | 11888 | 0.00 | 0.00 | 9.08E-01 | 9.51E-01 | 251 | 11685 | -0.01 | 0.00 | 6.26E-02 | 1.33E-01 | 130 | 0.00 | 0.00 | 1.68E-01 | 0.37 | 2.09E-01 | - | - | - |
| Receive hugs to comfort and support you | Social | 13007 | -0.02 | 0.01 | 1.03E-01 | 2.00E-01 | 136 | 12848 | 0.00 | 0.01 | 7.74E-01 | 8.41E-01 | 255 | -0.01 | 0.01 | 1.72E-01 | 0.00 | 3.51E-01 | - | - | - |
| Marital status: Single vs Married/partner | Social | 11909 | -0.02 | 0.01 | 2.32E-01 | 3.55E-01 | 172 | 11750 | -0.01 | 0.01 | 4.63E-01 | 6.03E-01 | 210 | -0.01 | 0.01 | 1.72E-01 | 0.00 | 7.52E-01 | - | - | - |
| Light and physically active work | Physical activity | 12648 | 0.00 | 0.01 | 9.67E-01 | 9.81E-01 | 259 | 12476 | -0.02 | 0.01 | 5.60E-02 | 1.24E-01 | 125 | -0.01 | 0.01 | 1.72E-01 | 0.44 | 1.80E-01 | - | - | - |
| Vitamin B2 intake (ug/day) | Nutrients | 11888 | 0.00 | 0.00 | 4.15E-01 | 5.43E-01 | 201 | 11685 | 0.00 | 0.00 | 2.81E-01 | 4.23E-01 | 184 | 0.00 | 0.00 | 1.82E-01 | 0.00 | 8.32E-01 | - | - | - |
| Close relationship with anyone | Social | 13104 | 0.00 | 0.00 | 3.93E-01 | 5.21E-01 | 198 | 12945 | 0.00 | 0.00 | 3.10E-01 | 4.48E-01 | 191 | 0.00 | 0.00 | 1.86E-01 | 0.00 | 9.15E-01 | - | - | - |

| | | | | | | | | | | | | | | | | | | | | | |
|---|---|---|---|---|---|---|---|---|---|---|---|---|---|---|---|---|---|---|---|---|---|
| Frequency of picking berries or mushrooms during leisure time | Physical activity | 4195 | 0.01 | 0.01 | 3.31E-01 | 4.58E-01 | 190 | 4291 | 0.01 | 0.01 | 3.71E-01 | 5.16E-01 | 199 | 0.01 | 0.01 | 1.87E-01 | 0.00 | 9.25E-01 | - | - | - |
| For how much of the time during the last four weeks have you felt calm and serene? | General health | 9508 | 0.00 | 0.00 | 8.88E-01 | 9.38E-01 | 249 | 9194 | 0.01 | 0.00 | 9.09E-02 | 1.84E-01 | 137 | 0.00 | 0.00 | 1.92E-01 | 0.18 | 2.70E-01 | - | - | - |
| Linolenic acid intake (g/day) | Nutrients | 11888 | 0.00 | 0.00 | 3.14E-01 | 4.39E-01 | 188 | 11685 | 0.00 | 0.00 | 4.65E-01 | 6.03E-01 | 213 | 0.00 | 0.00 | 2.18E-01 | 0.00 | 8.67E-01 | - | - | - |
| Marital status: Single vs Widow/widower | Social | 1469 | 0.08 | 0.04 | 3.46E-02 | 8.34E-02 | 109 | 1497 | -0.01 | 0.04 | 7.72E-01 | 8.41E-01 | 254 | 0.03 | 0.03 | 2.22E-01 | 0.67 | 7.98E-02 | - | - | - |
| Stigmasterol intake (mg/day) | Nutrients | 11888 | 0.01 | 0.00 | 1.54E-01 | 2.65E-01 | 153 | 11685 | 0.00 | 0.00 | 7.84E-01 | 8.46E-01 | 257 | 0.00 | 0.00 | 2.25E-01 | 0.00 | 4.27E-01 | - | - | - |
| Berries (fresh or frozen) | Food | 11888 | 0.00 | 0.00 | 5.93E-01 | 6.96E-01 | 224 | 11685 | 0.00 | 0.00 | 2.31E-01 | 3.70E-01 | 173 | 0.00 | 0.00 | 2.25E-01 | 0.00 | 6.19E-01 | - | - | - |
| Light but partly physically active work | Physical activity | 12648 | 0.01 | 0.01 | 3.83E-01 | 5.14E-01 | 196 | 12476 | 0.01 | 0.01 | 4.05E-01 | 5.53E-01 | 203 | 0.01 | 0.01 | 2.28E-01 | 0.00 | 9.83E-01 | - | - | - |
| Travel to work: Irregular travel mode to work vs passive travel to work | Physical activity | 8583 | 0.00 | 0.01 | 8.68E-01 | 9.20E-01 | 248 | 8416 | 0.02 | 0.01 | 1.26E-01 | 2.30E-01 | 151 | 0.01 | 0.01 | 2.29E-01 | 0.00 | 3.35E-01 | - | - | - |
| Smoking status: Former smokers vs non-smokers | Tobacco use | 9596 | 0.01 | 0.01 | 2.25E-01 | 3.47E-01 | 170 | 9306 | 0.00 | 0.01 | 6.47E-01 | 7.60E-01 | 235 | 0.01 | 0.01 | 2.32E-01 | 0.00 | 6.11E-01 | - | - | - |
| Pasta | Food | 11888 | -0.01 | 0.00 | 7.47E-02 | 1.56E-01 | 126 | 11685 | 0.00 | 0.00 | 8.68E-01 | 9.01E-01 | 267 | 0.00 | 0.00 | 2.42E-01 | 0.46 | 1.75E-01 | - | - | - |
| Rice | Food | 11888 | 0.00 | 0.00 | 8.49E-01 | 9.04E-01 | 247 | 11685 | -0.01 | 0.00 | 1.41E-01 | 2.51E-01 | 156 | 0.00 | 0.00 | 2.49E-01 | 0.00 | 3.51E-01 | - | - | - |
| Would you say that the number of people that you meet in your everyday life is enough or would you like to meet more or fewer people? | Social | 13139 | 0.00 | 0.00 | 3.88E-01 | 5.18E-01 | 197 | 12971 | 0.00 | 0.00 | 4.64E-01 | 6.03E-01 | 211 | 0.00 | 0.00 | 2.59E-01 | 0.00 | 9.36E-01 | - | - | - |
| Tea | Beverage | 11888 | 0.00 | 0.00 | 6.29E-01 | 7.22E-01 | 229 | 11685 | 0.00 | 0.00 | 2.67E-01 | 4.14E-01 | 179 | 0.00 | 0.00 | 2.63E-01 | 0.00 | 6.47E-01 | - | - | - |
| Linoleic acid intake (g/day) | Nutrients | 11888 | 0.00 | 0.00 | 2.97E-01 | 4.27E-01 | 183 | 11685 | 0.00 | 0.00 | 6.18E-01 | 7.32E-01 | 234 | 0.00 | 0.00 | 2.72E-01 | 0.00 | 7.19E-01 | - | - | - |
| Participation in study circles | Social | 5971 | 0.01 | 0.01 | 4.03E-01 | 5.33E-01 | 199 | 5640 | 0.01 | 0.01 | 4.74E-01 | 6.09E-01 | 215 | 0.01 | 0.01 | 2.72E-01 | 0.00 | 9.30E-01 | - | - | - |
| Margarine on bread | Food | 11888 | 0.00 | 0.00 | 5.39E-01 | 6.54E-01 | 217 | 11685 | 0.01 | 0.00 | 3.69E-02 | 8.97E-02 | 114 | 0.00 | 0.00 | 2.80E-01 | 0.72 | 5.88E-02 | - | - | - |
| Sausage as main dish | Food | 11888 | 0.00 | 0.00 | 3.49E-01 | 4.78E-01 | 192 | 11685 | 0.00 | 0.00 | 5.87E-01 | 7.10E-01 | 229 | 0.00 | 0.00 | 2.94E-01 | 0.00 | 7.93E-01 | - | - | - |
| Vitamin D intake (ug/day) | Nutrients | 11888 | 0.00 | 0.00 | 5.05E-01 | 6.30E-01 | 211 | 11685 | 0.00 | 0.00 | 4.24E-01 | 5.73E-01 | 205 | 0.00 | 0.00 | 3.01E-01 | 0.00 | 9.07E-01 | - | - | - |
| Oil for cooking | Food | 11888 | 0.00 | 0.00 | 2.17E-01 | 3.38E-01 | 169 | 11685 | 0.00 | 0.00 | 8.62E-01 | 9.01E-01 | 263 | 0.00 | 0.00 | 3.11E-01 | 0.00 | 4.68E-01 | - | - | - |
| Zinc intake (mg/day) | Nutrients | 11888 | 0.00 | 0.00 | 4.42E-01 | 5.70E-01 | 204 | 11685 | 0.00 | 0.00 | 5.14E-01 | 6.42E-01 | 222 | 0.00 | 0.00 | 3.15E-01 | 0.00 | 9.44E-01 | - | - | - |
| Distance to work in kilometers (one way) | Physical activity | 11132 | -0.01 | 0.00 | 1.72E-01 | 2.87E-01 | 158 | 10920 | 0.00 | 0.00 | 9.63E-01 | 9.64E-01 | 276 | 0.00 | 0.00 | 3.15E-01 | 0.00 | 3.54E-01 | - | - | - |
| If you exercise, change in exercise habits during the last year | Physical activity | 3920 | 0.01 | 0.01 | 1.10E-01 | 2.04E-01 | 142 | 4067 | 0.00 | 0.01 | 8.79E-01 | 9.02E-01 | 270 | 0.01 | 0.01 | 3.20E-01 | 0.37 | 2.06E-01 | - | - | - |
| For how much of the time during the last four weeks have you felt gloomy and sad? | General health | 9505 | 0.00 | 0.00 | 4.10E-01 | 5.39E-01 | 200 | 9177 | 0.00 | 0.00 | 5.65E-01 | 6.95E-01 | 225 | 0.00 | 0.00 | 3.22E-01 | 0.00 | 8.54E-01 | - | - | - |
| Ice cream | Food | 11888 | 0.00 | 0.00 | 7.58E-01 | 8.38E-01 | 238 | 11685 | -0.01 | 0.00 | 8.16E-02 | 1.69E-01 | 134 | 0.00 | 0.00 | 3.32E-01 | 0.54 | 1.39E-01 | - | - | - |
| Changed everyday exercise during the last year | Physical activity | 4471 | 0.01 | 0.01 | 2.84E-01 | 4.15E-01 | 180 | 4604 | 0.00 | 0.01 | 7.64E-01 | 8.40E-01 | 252 | 0.01 | 0.01 | 3.38E-01 | 0.00 | 5.70E-01 | - | - | - |
| Sum of all lignans intake (ug/day) | Nutrients | 11888 | 0.01 | 0.00 | 1.72E-01 | 2.87E-01 | 157 | 11685 | 0.00 | 0.00 | 9.49E-01 | 9.55E-01 | 275 | 0.00 | 0.00 | 3.49E-01 | 0.00 | 3.18E-01 | - | - | - |
| Cohabitation: Live alone vs Only children | Social | 2072 | -0.03 | 0.02 | 2.30E-01 | 3.54E-01 | 171 | 2136 | 0.00 | 0.02 | 8.62E-01 | 9.01E-01 | 264 | -0.01 | 0.02 | 3.59E-01 | 0.00 | 4.26E-01 | - | - | - |
| Sum of phytosterols intake (mg/day) | Nutrients | 11888 | 0.00 | 0.00 | 9.70E-01 | 9.81E-01 | 260 | 11685 | -0.01 | 0.00 | 2.12E-01 | 3.53E-01 | 166 | 0.00 | 0.00 | 3.72E-01 | 0.00 | 3.83E-01 | - | - | - |
| Breakfast habits: Coffee/tea and wheat buns or rusk for breakfast vs not breakfast at all | Food | 606 | 0.00 | 0.03 | 9.87E-01 | 9.91E-01 | 262 | 609 | 0.05 | 0.04 | 1.92E-01 | 3.28E-01 | 162 | 0.02 | 0.03 | 3.85E-01 | 0.02 | 3.11E-01 | - | - | - |
| Potassium intake (mg/day) | Nutrients | 11888 | 0.00 | 0.00 | 2.72E-01 | 4.04E-01 | 177 | 11685 | 0.00 | 0.00 | 9.40E-01 | 9.50E-01 | 274 | 0.00 | 0.00 | 3.98E-01 | 0.00 | 4.81E-01 | - | - | - |
| Sausage, liver pate on bread | Food | 11888 | 0.00 | 0.00 | 4.98E-01 | 6.24E-01 | 210 | 11685 | 0.01 | 0.00 | 5.81E-02 | 1.28E-01 | 126 | 0.00 | 0.00 | 4.10E-01 | 0.70 | 6.62E-02 | - | - | - |
| Self-employed | Psychosocial | 11542 | 0.01 | 0.01 | 4.54E-01 | 5.83E-01 | 205 | 11421 | 0.01 | 0.01 | 6.86E-01 | 7.89E-01 | 241 | 0.01 | 0.01 | 4.14E-01 | 0.00 | 8.10E-01 | - | - | - |
| Frequency of dancing during leisure time | Physical activity | 3884 | 0.00 | 0.01 | 6.82E-01 | 7.76E-01 | 231 | 3948 | -0.01 | 0.01 | 5.05E-01 | 6.36E-01 | 220 | 0.00 | 0.01 | 4.45E-01 | 0.00 | 8.66E-01 | - | - | - |
| Whole grain crisp bread | Food | 11888 | 0.00 | 0.00 | 4.78E-01 | 6.04E-01 | 208 | 11685 | 0.00 | 0.00 | 7.21E-01 | 8.02E-01 | 249 | 0.00 | 0.00 | 4.48E-01 | 0.00 | 8.15E-01 | - | - | - |
| Chips, popcorn, salted nuts | Food | 11888 | 0.00 | 0.00 | 5.91E-01 | 6.96E-01 | 222 | 11685 | 0.01 | 0.00 | 1.23E-01 | 2.28E-01 | 150 | 0.00 | 0.00 | 4.61E-01 | 0.53 | 1.45E-01 | - | - | - |
| Frequency of engaging in clubs, associations or study circles | Social | 7894 | 0.00 | 0.00 | 7.83E-01 | 8.58E-01 | 240 | 7757 | 0.00 | 0.01 | 4.61E-01 | 6.03E-01 | 209 | 0.00 | 0.00 | 4.78E-01 | 0.00 | 7.34E-01 | - | - | - |
| Emotional problems that made you do less than you wanted during the last four weeks | General health | 9524 | 0.00 | 0.01 | 8.30E-01 | 8.87E-01 | 246 | 9196 | -0.01 | 0.01 | 4.44E-01 | 5.97E-01 | 206 | -0.01 | 0.01 | 4.88E-01 | 0.00 | 6.95E-01 | - | - | - |
| Frequency of hunting or fishing during leisure time | Physical activity | 4154 | 0.00 | 0.01 | 9.62E-01 | 9.81E-01 | 256 | 4228 | 0.01 | 0.01 | 3.72E-01 | 5.16E-01 | 200 | 0.00 | 0.01 | 5.00E-01 | 0.00 | 5.58E-01 | - | - | - |
| Vitamin E intake (mg/day) | Nutrients | 11888 | 0.00 | 0.00 | 9.34E-01 | 9.67E-01 | 254 | 11685 | 0.00 | 0.00 | 3.78E-01 | 5.20E-01 | 201 | 0.00 | 0.00 | 5.00E-01 | 0.00 | 5.65E-01 | - | - | - |
| Oatflake, whole wheat, rye or barley porridge | Food | 11888 | 0.00 | 0.00 | 5.67E-01 | 6.78E-01 | 220 | 11685 | 0.00 | 0.00 | 7.17E-01 | 8.02E-01 | 247 | 0.00 | 0.00 | 5.06E-01 | 0.00 | 8.97E-01 | - | - | - |
| Fibre intake (g/day) | Nutrients | 11888 | 0.00 | 0.00 | 9.64E-01 | 9.81E-01 | 257 | 11685 | 0.00 | 0.00 | 3.17E-01 | 4.53E-01 | 194 | 0.00 | 0.00 | 5.07E-01 | 0.00 | 4.53E-01 | - | - | - |
| Pizza | Food | 11888 | 0.01 | 0.00 | 1.85E-01 | 3.03E-01 | 160 | 11685 | 0.00 | 0.00 | 6.76E-01 | 7.83E-01 | 238 | 0.00 | 0.00 | 5.08E-01 | 0.33 | 2.21E-01 | - | - | - |
| Campestanol intake (mg/day) | Nutrients | 11888 | 0.00 | 0.00 | 8.10E-01 | 8.70E-01 | 245 | 11685 | -0.01 | 0.00 | 2.35E-01 | 3.74E-01 | 174 | 0.00 | 0.00 | 5.13E-01 | 0.04 | 3.07E-01 | - | - | - |
| Fried potatoes, pommes frites | Food | 11888 | 0.00 | 0.00 | 5.92E-01 | 6.96E-01 | 223 | 11685 | 0.00 | 0.00 | 7.19E-01 | 8.02E-01 | 248 | 0.00 | 0.00 | 5.27E-01 | 0.00 | 9.00E-01 | - | - | - |
| Satisfaction with accomodation | Psychosocial | 11730 | -0.01 | 0.00 | 2.95E-02 | 7.46E-02 | 104 | 11494 | 0.01 | 0.00 | 1.74E-01 | 3.01E-01 | 160 | 0.00 | 0.00 | 5.28E-01 | 0.84 | 1.28E-02 | - | - | - |
| Possibility to leave your work for a while to speak with a colleague | Psychosocial | 12819 | 0.00 | 0.00 | 3.15E-01 | 4.39E-01 | 189 | 12633 | 0.00 | 0.00 | 8.61E-01 | 9.01E-01 | 262 | 0.00 | 0.00 | 5.53E-01 | 0.00 | 4.07E-01 | - | - | - |
| Vitamin B6 intake (mg/day) | Nutrients | 11888 | 0.00 | 0.00 | 5.11E-01 | 6.31E-01 | 213 | 11685 | 0.00 | 0.00 | 8.65E-01 | 9.01E-01 | 265 | 0.00 | 0.00 | 5.55E-01 | 0.00 | 7.37E-01 | - | - | - |
| High mental demand from job | Psychosocial | 12821 | 0.00 | 0.00 | 6.12E-01 | 7.09E-01 | 227 | 12670 | 0.00 | 0.00 | 7.45E-01 | 8.26E-01 | 250 | 0.00 | 0.00 | 5.55E-01 | 0.00 | 9.01E-01 | - | - | - |
| Beta-sitostanol intake (mg/day) | Nutrients | 11888 | 0.00 | 0.00 | 7.52E-01 | 8.35E-01 | 237 | 11685 | 0.00 | 0.00 | 2.47E-01 | 3.85E-01 | 178 | 0.00 | 0.00 | 5.63E-01 | 0.09 | 2.93E-01 | - | - | - |
| Marital status: Single vs Divorced/separated | Social | 2288 | 0.01 | 0.02 | 4.92E-01 | 6.19E-01 | 209 | 2332 | 0.00 | 0.02 | 9.28E-01 | 9.49E-01 | 271 | 0.01 | 0.01 | 6.00E-01 | 0.00 | 6.50E-01 | - | - | - |
| Satisfaction with home and family situation | Psychosocial | 11718 | 0.00 | 0.00 | 8.00E-01 | 8.64E-01 | 243 | 11485 | 0.00 | 0.00 | 3.09E-01 | 4.48E-01 | 190 | 0.00 | 0.00 | 6.04E-01 | 0.00 | 3.63E-01 | - | - | - |
| Beta-sitosterol intake (mg/day) | Nutrients | 11888 | 0.00 | 0.00 | 7.50E-01 | 8.35E-01 | 236 | 11685 | 0.00 | 0.00 | 2.86E-01 | 4.26E-01 | 185 | 0.00 | 0.00 | 6.10E-01 | 0.00 | 3.23E-01 | - | - | - |
| For how much of the time during the last four weeks have you felt so depressed that nothing could cheer you up? | General health | 9517 | 0.00 | 0.00 | 3.07E-01 | 4.32E-01 | 187 | 9193 | 0.00 | 0.00 | 7.61E-01 | 8.40E-01 | 251 | 0.00 | 0.00 | 6.15E-01 | 0.00 | 3.44E-01 | - | - | - |
| High physical demand from job | Physical activity | 12978 | 0.00 | 0.00 | 7.62E-01 | 8.39E-01 | 239 | 12805 | 0.00 | 0.00 | 6.96E-01 | 7.96E-01 | 242 | 0.00 | 0.00 | 6.24E-01 | 0.00 | 9.45E-01 | - | - | - |
| Frequency of gardening during leisure time | Physical activity | 4184 | 0.00 | 0.01 | 5.55E-01 | 6.67E-01 | 218 | 4275 | 0.01 | 0.01 | 2.19E-01 | 3.57E-01 | 169 | 0.00 | 0.01 | 6.36E-01 | 0.39 | 2.01E-01 | - | - | - |
| Sleep status | Sleep | 11718 | 0.00 | 0.00 | 5.30E-01 | 6.48E-01 | 215 | 11494 | 0.01 | 0.00 | 1.89E-01 | 3.26E-01 | 161 | 0.00 | 0.00 | 6.44E-01 | 0.48 | 1.67E-01 | - | - | - |
| Control over planning and execution of the workday | Psychosocial | 12951 | 0.00 | 0.00 | 5.55E-01 | 6.67E-01 | 219 | 12766 | 0.00 | 0.00 | 9.64E-01 | 9.64E-01 | 277 | 0.00 | 0.00 | 6.52E-01 | 0.00 | 7.02E-01 | - | - | - |
| Contradictory demands in job | Psychosocial | 12877 | 0.00 | 0.00 | 2.86E-01 | 4.15E-01 | 181 | 12678 | 0.00 | 0.00 | 6.47E-01 | 7.60E-01 | 236 | 0.00 | 0.00 | 6.61E-01 | 0.14 | 2.82E-01 | - | - | - |
| Cohabitation: Live alone vs Other/others | Social | 1673 | -0.04 | 0.04 | 3.38E-01 | 4.66E-01 | 191 | 1729 | 0.01 | 0.04 | 7.13E-01 | 8.02E-01 | 245 | -0.01 | 0.03 | 6.93E-01 | 0.00 | 3.42E-01 | - | - | - |
| Number of people with whom you can speak openly | Social | 13134 | 0.00 | 0.00 | 6.07E-01 | 7.06E-01 | 226 | 12964 | 0.00 | 0.00 | 2.87E-01 | 4.26E-01 | 187 | 0.00 | 0.00 | 7.06E-01 | 0.20 | 2.63E-01 | - | - | - |
| Milk, sour milk (3%) | Beverage | 11888 | 0.00 | 0.00 | 7.87E-01 | 8.59E-01 | 241 | 11685 | 0.00 | 0.00 | 8.36E-01 | 8.88E-01 | 261 | 0.00 | 0.00 | 7.35E-01 | 0.00 | 9.72E-01 | - | - | - |
| Enough time for job assignments | Psychosocial | 12874 | 0.00 | 0.00 | 7.96E-01 | 8.64E-01 | 242 | 12712 | 0.00 | 0.00 | 8.29E-01 | 8.87E-01 | 259 | 0.00 | 0.00 | 7.37E-01 | 0.00 | 9.77E-01 | - | - | - |
| Sometimes physically straining work | Physical activity | 12648 | 0.00 | 0.01 | 7.41E-01 | 8.30E-01 | 235 | 12476 | 0.01 | 0.01 | 4.22E-01 | 5.73E-01 | 204 | 0.00 | 0.01 | 7.43E-01 | 0.00 | 4.22E-01 | - | - | - |
| Bregott on bread | Food | 11888 | 0.00 | 0.00 | 4.58E-01 | 5.84E-01 | 206 | 11685 | -0.01 | 0.00 | 2.21E-01 | 3.57E-01 | 171 | 0.00 | 0.00 | 7.48E-01 | 0.49 | 1.63E-01 | - | - | - |
| Whole grain soft bread | Food | 11888 | 0.00 | 0.00 | 5.95E-01 | 6.96E-01 | 225 | 11685 | 0.00 | 0.00 | 3.26E-01 | 4.64E-01 | 195 | 0.00 | 0.00 | 7.65E-01 | 0.14 | 2.82E-01 | - | - | - |
| Breakfast habits: Gruel w/o sandwich for breakfast vs not breakfast at all | Food | 526 | -0.01 | 0.04 | 9.05E-01 | 9.51E-01 | 250 | 533 | 0.02 | 0.05 | 6.00E-01 | 7.17E-01 | 231 | 0.01 | 0.03 | 7.87E-01 | 0.00 | 6.36E-01 | - | - | - |
| Iron intake (mg/day) | Nutrients | 11888 | 0.00 | 0.00 | 9.57E-01 | 9.81E-01 | 255 | 11685 | 0.00 | 0.00 | 6.79E-01 | 7.83E-01 | 240 | 0.00 | 0.00 | 8.05E-01 | 0.00 | 7.36E-01 | - | - | - |
| Self rate of overall health compared to a year ago | General health | 9434 | 0.00 | 0.00 | 6.36E-01 | 7.27E-01 | 230 | 9121 | 0.00 | 0.00 | 4.66E-01 | 6.03E-01 | 214 | 0.00 | 0.00 | 8.54E-01 | 0.00 | 3.94E-01 | - | - | - |
| Confidence status | Psychosocial | 11701 | 0.00 | 0.00 | 9.94E-01 | 9.94E-01 | 263 | 11452 | 0.00 | 0.00 | 8.09E-01 | 8.69E-01 | 258 | 0.00 | 0.00 | 8.63E-01 | 0.00 | 8.66E-01 | - | - | - |
| Medioresinol intake (ug/day) | Nutrients | 11888 | 0.00 | 0.00 | 6.26E-01 | 7.22E-01 | 228 | 11685 | 0.00 | 0.00 | 4.64E-01 | 6.03E-01 | 212 | 0.00 | 0.00 | 8.74E-01 | 0.00 | 3.87E-01 | - | - | - |
| Enterolactone intake (ug/day) | Nutrients | 11888 | 0.00 | 0.00 | 7.16E-01 | 8.09E-01 | 233 | 11685 | 0.00 | 0.00 | 5.46E-01 | 6.75E-01 | 224 | 0.00 | 0.00 | 8.75E-01 | 0.00 | 4.92E-01 | - | - | - |
| Syringaresinol intake (ug/day) | Nutrients | 11888 | 0.00 | 0.00 | 5.35E-01 | 6.51E-01 | 216 | 11685 | 0.00 | 0.00 | 3.97E-01 | 5.44E-01 | 202 | 0.00 | 0.00 | 8.85E-01 | 0.08 | 2.98E-01 | - | - | - |
| Eat breakfast from 2000 | Food | 9738 | 0.01 | 0.01 | 5.70E-01 | 6.78E-01 | 221 | 9439 | -0.01 | 0.01 | 4.86E-01 | 6.17E-01 | 218 | 0.00 | 0.01 | 9.19E-01 | 0.00 | 3.71E-01 | - | - | - |
| Hamburger | Food | 11888 | 0.00 | 0.00 | 6.97E-01 | 7.90E-01 | 232 | 11685 | 0.00 | 0.00 | 6.00E-01 | 7.17E-01 | 230 | 0.00 | 0.00 | 9.23E-01 | 0.00 | 5.18E-01 | - | - | - |
| For how much of the time during the last four weeks have you felt very nervous? | General health | 9505 | 0.01 | 0.00 | 9.05E-02 | 1.80E-01 | 132 | 9186 | -0.01 | 0.00 | 1.15E-01 | 2.13E-01 | 149 | 0.00 | 0.00 | 9.32E-01 | 0.82 | 1.96E-02 | - | - | - |
| Apple, pear, peach, orange, mandarin and grapefruit | Food | 11888 | 0.00 | 0.00 | 4.32E-01 | 5.59E-01 | 203 | 11685 | 0.00 | 0.00 | 4.46E-01 | 5.97E-01 | 207 | 0.00 | 0.00 | 9.64E-01 | 0.17 | 2.74E-01 | - | - | - |
| Times during last year that you felt guilty because of your drinking | Alcohol | 8472 | 0.02 | 0.00 | - | - | 278 | 8117 | 0.03 | 0.00 | 5.73E-13 | **3.18E-11** | 5 | 0.03 | 0.00 | - | 0.18 | 2.69E-01 | - | - | - |
| Snore during sleep | Sleep | 8010 | -0.01 | 0.00 | - | - | 283 | 7753 | -0.02 | 0.00 | 5.10E-05 | **5.04E-04** | 28 | -0.01 | 0.00 | - | 0.72 | 5.72E-02 | - | - | - |
| Amount of alcohol drunk in a day | Alcohol | 8330 | 0.02 | 0.00 | - | - | 276 | 7989 | 0.02 | 0.00 | 1.40E-04 | **1.17E-03** | 32 | 0.02 | 0.00 | - | 0.00 | 7.81E-01 | - | - | - |
| Risk of sleeping while lying down resting in the afternoon | Sleep | 8575 | 0.00 | 0.00 | - | - | 281 | 8307 | -0.01 | 0.00 | 2.52E-03 | **1.06E-02** | 66 | -0.01 | 0.00 | - | 0.51 | 1.54E-01 | - | - | - |
| Breath-holds during sleep | Sleep | 6011 | 0.00 | 0.01 | - | - | 284 | 5752 | -0.01 | 0.01 | 1.44E-01 | 2.54E-01 | 157 | -0.01 | 0.00 | - | 0.00 | 5.95E-01 | - | - | - |
| Potato salad | Food | 1269 | -0.02 | 0.02 | - | - | 270 | 1309 | 0.01 | 0.01 | 4.55E-01 | 6.03E-01 | 208 | 0.00 | 0.01 | - | 0.49 | 1.63E-01 | - | - | - |
| Liver, kidney | Food | 1269 | 0.02 | 0.02 | - | - | 273 | 1309 | -0.01 | 0.01 | 4.75E-01 | 6.09E-01 | 216 | 0.00 | 0.01 | - | 0.39 | 2.02E-01 | - | - | - |
| Mixed frozen vegetables | Food | 1269 | -0.01 | 0.01 | - | - | 268 | 1309 | 0.01 | 0.01 | 5.14E-01 | 6.42E-01 | 221 | 0.00 | 0.01 | - | 0.00 | 4.06E-01 | - | - | - |
| Soft cheese | Food | 1269 | 0.01 | 0.01 | - | - | 266 | 1309 | -0.01 | 0.01 | 5.74E-01 | 7.01E-01 | 227 | 0.00 | 0.01 | - | 0.00 | 3.62E-01 | - | - | - |
| Shellfish (e.g. shrimps, scallops) | Food | 1269 | 0.02 | 0.02 | - | - | 274 | 1309 | -0.01 | 0.02 | 5.79E-01 | 7.04E-01 | 228 | 0.00 | 0.01 | - | 0.20 | 2.62E-01 | - | - | - |
| Risk of sleeping while sitting still after having lunch | Sleep | 8581 | 0.00 | 0.00 | - | - | 282 | 8304 | 0.00 | 0.00 | 6.01E-01 | 7.17E-01 | 232 | 0.00 | 0.00 | - | 0.00 | 6.93E-01 | - | - | - |
| Risk of sleeping while sitting and reading | Sleep | 8518 | 0.00 | 0.00 | - | - | 279 | 8237 | 0.00 | 0.00 | 6.77E-01 | 7.83E-01 | 239 | 0.00 | 0.00 | - | 0.00 | 8.50E-01 | - | - | - |
| Mashed potato | Food | 1269 | 0.01 | 0.01 | - | - | 269 | 1309 | 0.01 | 0.02 | 7.01E-01 | 7.99E-01 | 243 | 0.01 | 0.01 | - | 0.00 | 7.35E-01 | - | - | - |
| Blood based food | Food | 1269 | 0.01 | 0.02 | - | - | 272 | 1309 | 0.01 | 0.01 | 7.07E-01 | 8.02E-01 | 244 | 0.01 | 0.01 | - | 0.00 | 8.88E-01 | - | - | - |
| Blota (broth + bread) | Food | 1269 | 0.01 | 0.02 | - | - | 271 | 1309 | 0.00 | 0.02 | 8.35E-01 | 8.88E-01 | 260 | 0.00 | 0.01 | - | 0.00 | 5.29E-01 | - | - | - |
| Soft whey cheese | Food | 1269 | -0.01 | 0.02 | - | - | 267 | 1309 | 0.00 | 0.01 | 8.75E-01 | 9.01E-01 | 268 | 0.00 | 0.01 | - | 0.00 | 7.47E-01 | - | - | - |
| Risk of sleeping while watching TV | Sleep | 8612 | 0.00 | 0.00 | 2.99E-01 | 4.27E-01 | 184 | 8326 | 0.00 | 0.00 | - | - | 282 | 0.00 | 0.00 | - | 0.00 | 9.15E-01 | - | - | - |
| Risk of sleeping as a passenger in a car for one hour without break | Sleep | 8518 | 0.00 | 0.00 | 5.28E-01 | 6.48E-01 | 214 | 8240 | 0.00 | 0.00 | - | - | 284 | 0.00 | 0.00 | - | 0.00 | 3.58E-01 | - | - | - |
| Time spent in a week in moderately steneous activities | Physical activity | 8400 | 0.03 | 0.00 | - | - | 264 | 8083 | 0.02 | 0.00 | - | - | 278 | 0.02 | 0.00 | - | 0.00 | 3.80E-01 | - | - | - |

| | | | | | | | | | | | | | | | | | | | | | |
|---|---|---|---|---|---|---|---|---|---|---|---|---|---|---|---|---|---|---|---|---|---|
| Amount of exercise during the last 12 months | Physical activity | 8665 | 0.02 | 0.00 | - | - | 265 | 8359 | 0.03 | 0.00 | - | - | 279 | 0.03 | 0.00 | - | 0.00 | 3.64E-01 | - | - | - |
| Frequency of alcohol consumption | Alcohol | 8706 | 0.05 | 0.00 | - | - | 275 | 8418 | 0.06 | 0.00 | - | - | 280 | 0.06 | 0.00 | - | 0.72 | 5.82E-02 | - | - | - |
| Frequency of drinking six or more glasses at the same occasion | Alcohol | 8481 | 0.03 | 0.00 | - | - | 277 | 8135 | 0.04 | 0.00 | - | - | 281 | 0.03 | 0.00 | - | 0.57 | 1.26E-01 | - | - | - |
| Risk of sleeping while sitting inactive in a public place | Sleep | 8556 | 0.00 | 0.00 | - | - | 280 | 8279 | 0.00 | 0.00 | - | - | 283 | 0.00 | 0.00 | - | 0.00 | 7.34E-01 | - | - | - |

**Supplementary Table 9**. Linear mixed model association results for LDL cholesterol

| | | Training set | | | | | | Testing set | | | | | | Metanalysis | | | | | R2 | | |
|---|---|---|---|---|---|---|---|---|---|---|---|---|---|---|---|---|---|---|---|---|---|
| **Description** | **Group** | **N** | **Effect estimate** | **S.E.** | **p-value** | **p-value$_{FDR}$** | **p-value rank** | **N** | **Effect estimate** | **S.E.** | **p-value** | **p-value$_{FDR}$** | **p-value rank** | **Effect estimate** | **S.E.** | **p-value** | **$I^2$** | **Q p-value** | **marginal R2** | **conditional R2** | **R2 rank** |
| Boiled coffee | Beverage | 11841 | 0.07 | 0.01 | 8.01E-12 | **1.05E-09** | 2 | 11617 | 0.06 | 0.01 | 2.55E-09 | **7.06E-07** | 1 | 0.06 | 0.01 | 1.41E-19 | 0.00 | 5.42E-01 | 0.08 | 0.55 | 1 |
| Monounsaturated fat intake (g/day) | Nutrients | 11841 | 0.07 | 0.01 | 1.46E-12 | **3.83E-10** | 1 | 11617 | 0.05 | 0.01 | 2.59E-06 | **2.39E-04** | 3 | 0.06 | 0.01 | 6.31E-17 | 0.60 | 1.15E-01 | 0.08 | 0.55 | 2 |
| Palmitic acid intake (g/day) | Nutrients | 11841 | 0.06 | 0.01 | 4.98E-11 | **4.36E-09** | 3 | 11617 | 0.05 | 0.01 | 9.10E-07 | **1.26E-04** | 2 | 0.05 | 0.01 | 4.44E-16 | 0.26 | 2.46E-01 | 0.08 | 0.55 | 3 |
| Exercise during the last three months | Physical activity | 12815 | -0.05 | 0.01 | 3.16E-10 | **1.66E-08** | 5 | 12653 | -0.04 | 0.01 | 1.41E-05 | **9.79E-04** | 4 | -0.05 | 0.01 | 4.98E-14 | 0.43 | 1.84E-01 | 0.04 | 0.51 | 19 |
| Trans fat intake (g/day) | Nutrients | 11841 | 0.10 | 0.02 | 1.32E-10 | **8.65E-09** | 4 | 11617 | 0.06 | 0.02 | 2.70E-04 | **6.80E-03** | 11 | 0.08 | 0.01 | 1.02E-12 | 0.74 | 4.81E-02 | 0.08 | 0.55 | 4 |
| Formic acid intaje (g/day) | Nutrients | 11841 | 0.05 | 0.01 | 3.54E-07 | **1.16E-05** | 8 | 11617 | 0.04 | 0.01 | 1.51E-04 | **5.67E-03** | 7 | 0.04 | 0.01 | 3.27E-10 | 0.00 | 3.66E-01 | 0.08 | 0.55 | 5 |
| Fat intake (g/day) | Nutrients | 11841 | 0.05 | 0.01 | 5.32E-07 | **1.55E-05** | 9 | 11617 | 0.04 | 0.01 | 1.64E-04 | **5.67E-03** | 8 | 0.04 | 0.01 | 4.86E-10 | 0.00 | 4.22E-01 | 0.08 | 0.55 | 7 |
| Educational level | Psychosocial | 13104 | -0.05 | 0.01 | 1.48E-06 | **3.88E-05** | 10 | 12906 | -0.04 | 0.01 | 1.42E-04 | **5.67E-03** | 6 | -0.04 | 0.01 | 1.07E-09 | 0.00 | 4.92E-01 | 0.04 | 0.51 | 20 |
| Plant based protein intake (g/day) | Nutrients | 11841 | -0.05 | 0.01 | 4.69E-08 | **2.05E-06** | 6 | 11617 | -0.03 | 0.01 | 1.98E-03 | **2.19E-02** | 25 | -0.04 | 0.01 | 1.19E-09 | 0.60 | 1.14E-01 | 0.08 | 0.55 | 6 |
| Saturated fat intake (g/day) | Nutrients | 11841 | 0.04 | 0.01 | 7.41E-06 | **1.30E-04** | 15 | 11617 | 0.03 | 0.01 | 2.63E-04 | **6.80E-03** | 10 | 0.04 | 0.01 | 8.74E-09 | 0.00 | 5.78E-01 | 0.08 | 0.55 | 11 |
| Iron intake (mg/day) | Nutrients | 11841 | -0.05 | 0.01 | 5.66E-08 | **2.13E-06** | 7 | 11617 | -0.03 | 0.01 | 8.31E-03 | 5.90E-02 | 39 | -0.04 | 0.01 | 9.07E-09 | 0.72 | 6.10E-02 | - | - | - |
| Carbohydrates intake (g/day) | Nutrients | 11841 | -0.04 | 0.01 | 1.35E-05 | **2.21E-04** | 16 | 11617 | -0.03 | 0.01 | 3.36E-04 | **6.87E-03** | 13 | -0.04 | 0.01 | 1.91E-08 | 0.00 | 6.31E-01 | 0.08 | 0.55 | 12 |
| Cholesterol intake (g/day) | Nutrients | 11841 | 0.04 | 0.01 | 7.06E-05 | **9.77E-04** | 19 | 11617 | 0.04 | 0.01 | 1.07E-04 | **5.67E-03** | 5 | 0.04 | 0.01 | 2.85E-08 | 0.00 | 9.60E-01 | 0.08 | 0.55 | 18 |
| Heptadecanoic acid intake (g/day) | Nutrients | 11841 | 0.04 | 0.01 | 7.26E-06 | **1.30E-04** | 13 | 11617 | 0.03 | 0.01 | 1.12E-03 | **1.35E-02** | 23 | 0.04 | 0.01 | 4.24E-08 | 0.00 | 3.97E-01 | 0.08 | 0.55 | 10 |
| Pentadecanoic acid intake (g/day) | Nutrients | 11841 | 0.04 | 0.01 | 7.26E-06 | **1.30E-04** | 14 | 11617 | 0.03 | 0.01 | 1.12E-03 | **1.35E-02** | 22 | 0.04 | 0.01 | 4.24E-08 | 0.00 | 3.97E-01 | 0.08 | 0.55 | 9 |
| Vitamin D intake (ug/day) | Nutrients | 11841 | 0.03 | 0.01 | 5.59E-05 | **8.16E-04** | 18 | 11617 | 0.03 | 0.01 | 1.00E-03 | **1.32E-02** | 21 | 0.03 | 0.01 | 2.17E-07 | 0.00 | 6.59E-01 | 0.08 | 0.55 | 8 |
| Fitness status | Physical activity | 11676 | -0.05 | 0.01 | 2.20E-06 | **5.26E-05** | 11 | 11410 | -0.02 | 0.01 | 1.94E-02 | 9.60E-02 | 56 | -0.03 | 0.01 | 5.20E-07 | 0.64 | 9.65E-02 | - | - | - |
| Cream, creme fraiche, sour cream | Food | 11841 | 0.03 | 0.01 | 1.54E-03 | **1.39E-02** | 29 | 11617 | 0.03 | 0.01 | 6.96E-04 | **1.12E-02** | 17 | 0.03 | 0.01 | 3.54E-06 | 0.00 | 8.40E-01 | 0.08 | 0.55 | 13 |
| Banana | Food | 11841 | -0.03 | 0.01 | 1.37E-03 | **1.33E-02** | 27 | 11617 | -0.03 | 0.01 | 9.60E-04 | **1.32E-02** | 20 | -0.03 | 0.01 | 4.26E-06 | 0.00 | 9.27E-01 | 0.08 | 0.55 | 16 |
| Bregott on bread | Food | 11841 | 0.04 | 0.01 | 1.43E-05 | **2.22E-04** | 17 | 11617 | 0.02 | 0.01 | 3.42E-02 | 1.50E-01 | 63 | 0.03 | 0.01 | 4.59E-06 | 0.58 | 1.25E-01 | - | - | - |
| Butter for cooking | Food | 11841 | 0.03 | 0.01 | 2.11E-04 | **2.64E-03** | 21 | 11617 | 0.02 | 0.01 | 5.42E-03 | **4.69E-02** | 32 | 0.03 | 0.01 | 4.69E-06 | 0.00 | 4.71E-01 | 0.08 | 0.55 | 17 |
| Fibre intake (g/day) | Nutrients | 11841 | -0.04 | 0.01 | 1.27E-04 | **1.67E-03** | 20 | 11617 | -0.03 | 0.01 | 9.84E-03 | 6.49E-02 | 42 | -0.03 | 0.01 | 5.44E-06 | 0.00 | 4.05E-01 | - | - | - |
| Tea | Beverage | 11841 | -0.03 | 0.01 | 4.22E-03 | **2.92E-02** | 38 | 11617 | -0.03 | 0.01 | 7.26E-04 | **1.12E-02** | 18 | -0.03 | 0.01 | 1.03E-05 | 0.00 | 6.86E-01 | 0.08 | 0.55 | 15 |
| Equol intake (ug/day) | Nutrients | 11841 | -0.03 | 0.01 | 9.37E-04 | **9.86E-03** | 25 | 11617 | -0.03 | 0.01 | 4.03E-03 | **3.72E-02** | 30 | -0.03 | 0.01 | 1.19E-05 | 0.00 | 8.39E-01 | 0.08 | 0.55 | 14 |
| Margarine for cooking | Food | 11841 | 0.02 | 0.01 | 9.45E-03 | 5.65E-02 | 44 | 11617 | 0.03 | 0.01 | 3.47E-04 | **6.87E-03** | 14 | 0.03 | 0.01 | 1.32E-05 | 0.00 | 4.54E-01 | - | - | - |
| Vitamin C intake (mg/day) | Nutrients | 11841 | -0.03 | 0.01 | 2.69E-04 | **3.22E-03** | 22 | 11617 | -0.02 | 0.01 | 1.37E-02 | 7.80E-02 | 48 | -0.03 | 0.01 | 1.47E-05 | 0.00 | 4.40E-01 | - | - | - |
| Eat breakfast from 2000 | Food | 9720 | -0.12 | 0.04 | 1.25E-03 | **1.26E-02** | 26 | 9408 | -0.09 | 0.03 | 6.72E-03 | 5.17E-02 | 36 | -0.10 | 0.02 | 2.77E-05 | 0.00 | 6.40E-01 | - | - | - |
| Average portion size of meat/fish based on photographic illustration of four sizes (smallest to largest) | Food | 11841 | 0.03 | 0.01 | 3.42E-03 | **2.43E-02** | 37 | 11617 | 0.03 | 0.01 | 2.91E-03 | **2.99E-02** | 26 | 0.03 | 0.01 | 2.99E-05 | 0.00 | 9.26E-01 | 0.04 | 0.51 | 21 |
| Parents or siblings had a cerebral hemorrhage/thrombosis or cardiac infarction before the age of 60 | General health | 12787 | 0.07 | 0.02 | 8.91E-04 | **9.77E-03** | 24 | 12623 | 0.06 | 0.02 | 1.13E-02 | 6.93E-02 | 45 | 0.07 | 0.02 | 3.38E-05 | 0.00 | 5.96E-01 | - | - | - |
| Fatty fish (e.g. herring, whitefish, salmon) | Food | 11841 | 0.02 | 0.01 | 3.53E-02 | 1.39E-01 | 67 | 11617 | 0.03 | 0.01 | 2.41E-04 | **6.80E-03** | 9 | 0.02 | 0.01 | 4.73E-05 | 0.27 | 2.40E-01 | - | - | - |
| Calcium intake (mg/day) | Nutrients | 11841 | -0.03 | 0.01 | 2.30E-03 | **1.89E-02** | 32 | 11617 | -0.03 | 0.01 | 7.64E-03 | 5.72E-02 | 37 | -0.03 | 0.01 | 5.18E-05 | 0.00 | 8.55E-01 | - | - | - |
| Docosahexaenoic acid (DHA) intake (g/day) | Nutrients | 11841 | 0.02 | 0.01 | 3.45E-02 | 1.38E-01 | 65 | 11617 | 0.03 | 0.01 | 2.95E-04 | **6.80E-03** | 12 | 0.02 | 0.01 | 5.71E-05 | 0.28 | 2.37E-01 | - | - | - |
| Low fat milk (0,5%) | Beverage | 11841 | -0.03 | 0.01 | 5.84E-03 | **3.84E-02** | 40 | 11617 | -0.03 | 0.01 | 6.57E-03 | 5.17E-02 | 35 | -0.03 | 0.01 | 1.08E-04 | 0.00 | 9.55E-01 | - | - | - |
| If you exercise, change in exercise habits during the last year | Physical activity | 3872 | -0.05 | 0.02 | 3.40E-03 | **2.43E-02** | 36 | 4000 | -0.04 | 0.02 | 1.38E-02 | 7.80E-02 | 49 | -0.05 | 0.01 | 1.38E-04 | 0.00 | 7.09E-01 | - | - | - |
| Bacon | Food | 11841 | 0.02 | 0.01 | 1.79E-02 | 8.28E-02 | 57 | 11617 | 0.02 | 0.01 | 2.91E-03 | **2.99E-02** | 27 | 0.02 | 0.01 | 1.60E-04 | 0.00 | 6.45E-01 | - | - | - |
| Eicosapentaenoic acid (EPA) intake (g/day) | Nutrients | 11840 | 0.02 | 0.01 | 5.85E-02 | 1.87E-01 | 82 | 11617 | 0.03 | 0.01 | 5.91E-04 | **1.02E-02** | 16 | 0.02 | 0.01 | 1.80E-04 | 0.27 | 2.42E-01 | - | - | - |
| Average portion size of vegetables based on photographic illustration of four sizes (smallest to largest) | Food | 11841 | -0.03 | 0.01 | 6.88E-04 | **7.87E-03** | 23 | 11617 | -0.02 | 0.01 | 5.94E-02 | 2.11E-01 | 78 | -0.03 | 0.01 | 1.80E-04 | 0.06 | 3.03E-01 | - | - | - |
| Total energy intake (kcal/day) | Nutrients | 11841 | -0.03 | 0.01 | 1.74E-02 | 8.28E-02 | 54 | 11617 | -0.03 | 0.01 | 3.72E-03 | **3.55E-02** | 29 | -0.03 | 0.01 | 1.89E-04 | 0.00 | 7.07E-01 | - | - | - |
| Vitamin B3 intake (mg/day) | Nutrients | 11841 | 0.01 | 0.01 | 9.05E-02 | 2.48E-01 | 96 | 11617 | 0.03 | 0.01 | 4.23E-04 | **7.81E-03** | 15 | 0.02 | 0.01 | 2.33E-04 | 0.43 | 1.84E-01 | - | - | - |
| Brown beans, pea soup | Food | 11841 | -0.02 | 0.01 | 3.08E-03 | **2.31E-02** | 35 | 11617 | -0.02 | 0.01 | 3.03E-02 | 1.35E-01 | 62 | -0.02 | 0.01 | 2.70E-04 | 0.00 | 6.65E-01 | - | - | - |
| Breakfast habits: Porridge w/o sandwich for breakfast vs not breakfast at all | Food | 1845 | -0.18 | 0.07 | 7.01E-03 | **4.50E-02** | 41 | 1806 | -0.15 | 0.07 | 2.06E-02 | 1.00E-01 | 57 | -0.17 | 0.05 | 3.90E-04 | 0.00 | 7.56E-01 | - | - | - |
| Smoking status: Smokers vs non-smokers | Tobacco use | 8184 | 0.07 | 0.03 | 1.74E-02 | 8.28E-02 | 55 | 8090 | 0.07 | 0.03 | 1.07E-02 | 6.73E-02 | 44 | 0.07 | 0.02 | 4.88E-04 | 0.00 | 9.15E-01 | - | - | - |
| Pizza | Food | 11841 | -0.02 | 0.01 | 5.99E-02 | 1.87E-01 | 84 | 11617 | -0.03 | 0.01 | 3.61E-03 | **3.55E-02** | 28 | -0.02 | 0.01 | 7.27E-04 | 0.00 | 4.40E-01 | - | - | - |
| Campesterol intake (mg/day) | Nutrients | 11841 | 0.03 | 0.01 | 4.93E-03 | **3.32E-02** | 39 | 11617 | 0.02 | 0.01 | 5.22E-02 | 1.96E-01 | 73 | 0.02 | 0.01 | 7.50E-04 | 0.00 | 5.73E-01 | - | - | - |
| Cambridge physical activity index | Physical activity | 12306 | -0.02 | 0.01 | 3.47E-02 | 1.38E-01 | 66 | 12142 | -0.02 | 0.01 | 9.64E-03 | 6.49E-02 | 41 | -0.02 | 0.01 | 8.99E-04 | 0.00 | 7.10E-01 | - | - | - |
| Changed everyday exercise during the last year | Physical activity | 4416 | -0.05 | 0.02 | 2.77E-03 | **2.16E-02** | 33 | 4532 | -0.03 | 0.02 | 1.02E-01 | 3.06E-01 | 92 | -0.04 | 0.01 | 1.02E-03 | 0.00 | 3.46E-01 | - | - | - |
| Arachidonic acid (ARA) intake (g/day) | Nutrients | 11840 | 0.02 | 0.01 | 6.07E-02 | 1.87E-01 | 85 | 11617 | 0.02 | 0.01 | 5.86E-03 | **4.78E-02** | 34 | 0.02 | 0.01 | 1.09E-03 | 0.00 | 5.00E-01 | - | - | - |
| Steak, chop, e.g. | Food | 11841 | 0.02 | 0.01 | 2.76E-02 | 1.17E-01 | 62 | 11617 | 0.02 | 0.01 | 1.70E-02 | 8.55E-02 | 55 | 0.02 | 0.01 | 1.18E-03 | 0.00 | 8.51E-01 | - | - | - |
| Cheese 10-17% | Food | 11841 | -0.03 | 0.01 | 1.93E-03 | **1.64E-02** | 30 | 11617 | -0.01 | 0.01 | 1.57E-01 | 4.18E-01 | 103 | -0.02 | 0.01 | 1.28E-03 | 0.21 | 2.62E-01 | - | - | - |
| Polyunsaturated fat intake (g/day) | Nutrients | 11841 | 0.02 | 0.01 | 3.79E-02 | 1.46E-01 | 68 | 11617 | 0.02 | 0.01 | 1.30E-02 | 7.71E-02 | 46 | 0.02 | 0.01 | 1.30E-03 | 0.00 | 7.13E-01 | - | - | - |
| Milk, sour milk (3%) | Beverage | 11841 | 0.01 | 0.01 | 1.70E-01 | 3.73E-01 | 120 | 11617 | 0.03 | 0.01 | 1.34E-03 | **1.54E-02** | 24 | 0.02 | 0.01 | 1.35E-03 | 0.48 | 1.66E-01 | - | - | - |
| Rice | Food | 11841 | -0.04 | 0.01 | 6.33E-06 | **1.30E-04** | 12 | 11617 | 0.00 | 0.01 | 8.69E-01 | 9.44E-01 | 255 | -0.02 | 0.01 | 1.66E-03 | 0.91 | 1.12E-03 | - | - | - |
| Low fat margarine on bread | Food | 11841 | -0.02 | 0.01 | 8.54E-02 | 2.39E-01 | 94 | 11617 | -0.03 | 0.01 | 8.00E-03 | 5.83E-02 | 38 | -0.02 | 0.01 | 2.07E-03 | 0.00 | 4.76E-01 | - | - | - |
| Would you say that the number of people that you meet in your everyday life is enough or would you like to meet more or fewer people? | Social | 13083 | -0.03 | 0.01 | 1.94E-03 | **1.64E-02** | 31 | 12894 | -0.01 | 0.01 | 2.27E-01 | 5.15E-01 | 122 | -0.02 | 0.01 | 2.23E-03 | 0.42 | 1.87E-01 | - | - | - |
| Number of snuff boxes per week | Tobacco use | 12753 | -0.02 | 0.01 | 1.36E-02 | 7.00E-02 | 51 | 12593 | -0.02 | 0.01 | 7.84E-02 | 2.56E-01 | 84 | -0.02 | 0.01 | 2.81E-03 | 0.00 | 6.06E-01 | - | - | - |
| Vitamin E intake (mg/day) | Nutrients | 11841 | 0.02 | 0.01 | 4.92E-02 | 1.74E-01 | 73 | 11617 | 0.02 | 0.01 | 2.44E-02 | 1.14E-01 | 59 | 0.02 | 0.01 | 2.87E-03 | 0.00 | 8.23E-01 | - | - | - |
| Frequency of cycling during leisure time | Physical activity | 10222 | -0.02 | 0.01 | 7.25E-02 | 2.13E-01 | 89 | 9948 | -0.02 | 0.01 | 1.60E-02 | 8.38E-02 | 53 | -0.02 | 0.01 | 2.98E-03 | 0.00 | 6.48E-01 | - | - | - |
| High physical demand from job | Physical activity | 12925 | 0.01 | 0.01 | 1.14E-01 | 2.85E-01 | 105 | 12728 | 0.02 | 0.01 | 8.97E-03 | 6.21E-02 | 40 | 0.02 | 0.01 | 3.10E-03 | 0.00 | 4.44E-01 | - | - | - |
| Meat stew | Food | 11841 | 0.02 | 0.01 | 4.89E-02 | 1.74E-01 | 72 | 11617 | 0.02 | 0.01 | 2.73E-02 | 1.24E-01 | 61 | 0.02 | 0.01 | 3.11E-03 | 0.00 | 9.35E-01 | - | - | - |
| Light beer | Alcohol | 11841 | 0.04 | 0.01 | 1.47E-03 | **1.38E-02** | 28 | 11617 | 0.01 | 0.01 | 3.20E-01 | 6.39E-01 | 138 | 0.03 | 0.01 | 3.15E-03 | 0.58 | 1.21E-01 | - | - | - |
| Satisfaction with home and family situation | Psychosocial | 11685 | 0.02 | 0.01 | 1.15E-02 | 6.42E-02 | 47 | 11422 | 0.01 | 0.01 | 1.34E-01 | 3.67E-01 | 101 | 0.02 | 0.01 | 4.32E-03 | 0.00 | 4.80E-01 | - | - | - |
| Salad dressing with oil | Food | 11841 | -0.01 | 0.01 | 1.10E-01 | 2.80E-01 | 102 | 11617 | -0.02 | 0.01 | 1.57E-02 | 8.37E-02 | 52 | -0.02 | 0.01 | 4.72E-03 | 0.00 | 5.23E-01 | - | - | - |
| Fiber cereals | Food | 11841 | -0.02 | 0.01 | 1.95E-02 | 8.56E-02 | 60 | 11617 | -0.01 | 0.01 | 1.21E-01 | 3.39E-01 | 99 | -0.02 | 0.01 | 5.92E-03 | 0.00 | 5.96E-01 | - | - | - |
| Number of cigarretes smoked per day (in groups) | Tobacco use | 10600 | 0.01 | 0.01 | 1.46E-01 | 3.39E-01 | 113 | 10496 | 0.02 | 0.01 | 1.49E-02 | 8.25E-02 | 50 | 0.02 | 0.01 | 5.97E-03 | 0.00 | 4.84E-01 | - | - | - |
| Whole grain crisp bread | Food | 11841 | 0.02 | 0.01 | 1.28E-02 | 6.77E-02 | 49 | 11617 | 0.01 | 0.01 | 1.86E-01 | 4.47E-01 | 115 | 0.02 | 0.01 | 6.75E-03 | 0.00 | 4.34E-01 | - | - | - |
| Folic acid intake (ug/day) | Nutrients | 11841 | -0.02 | 0.01 | 1.11E-02 | 6.32E-02 | 46 | 11617 | -0.01 | 0.01 | 2.11E-01 | 4.88E-01 | 120 | -0.02 | 0.01 | 7.01E-03 | 0.00 | 3.85E-01 | - | - | - |
| Last time a colleague visited you at home | Psychosocial | 12721 | 0.01 | 0.01 | 1.22E-01 | 3.01E-01 | 107 | 12534 | 0.02 | 0.01 | 2.41E-02 | 1.14E-01 | 58 | 0.02 | 0.01 | 7.33E-03 | 0.00 | 5.92E-01 | - | - | - |
| Number of friends that can come to your home at any time and feel at home | Social | 13029 | 0.01 | 0.01 | 2.26E-01 | 4.28E-01 | 139 | 12861 | 0.02 | 0.01 | 1.31E-02 | 7.71E-02 | 47 | 0.02 | 0.01 | 9.25E-03 | 0.00 | 3.56E-01 | - | - | - |
| Beta-carotene intake (mg/day) | Nutrients | 11841 | -0.02 | 0.01 | 2.26E-02 | 9.73E-02 | 61 | 11617 | -0.01 | 0.01 | 1.72E-01 | 4.35E-01 | 109 | -0.02 | 0.01 | 9.63E-03 | 0.00 | 5.46E-01 | - | - | - |
| Travel to work: Cycle to work vs passive travel to work | Physical activity | 9645 | -0.04 | 0.02 | 9.63E-02 | 2.58E-01 | 98 | 9441 | -0.05 | 0.02 | 4.72E-02 | 1.90E-01 | 69 | -0.05 | 0.02 | 9.92E-03 | 0.00 | 8.14E-01 | - | - | - |
| Tomato, cucumber | Food | 11841 | -0.02 | 0.01 | 1.09E-02 | 6.32E-02 | 45 | 11617 | -0.01 | 0.01 | 2.88E-01 | 6.06E-01 | 131 | -0.02 | 0.01 | 1.06E-02 | 0.08 | 2.97E-01 | - | - | - |
| Minced meat dishes | Food | 11841 | 0.01 | 0.01 | 8.82E-02 | 2.44E-01 | 95 | 11617 | 0.02 | 0.01 | 6.07E-02 | 2.13E-01 | 79 | 0.02 | 0.01 | 1.13E-02 | 0.00 | 9.32E-01 | - | - | - |
| Vitamin B2 intake (ug/day) | Nutrients | 11841 | -0.02 | 0.01 | 1.93E-02 | 8.56E-02 | 59 | 11617 | -0.01 | 0.01 | 2.36E-01 | 5.28E-01 | 123 | -0.02 | 0.01 | 1.22E-02 | 0.00 | 4.36E-01 | - | - | - |
| Berries (fresh or frozen) | Food | 11841 | -0.02 | 0.01 | 1.62E-02 | 8.21E-02 | 52 | 11617 | -0.01 | 0.01 | 2.64E-01 | 5.76E-01 | 127 | -0.01 | 0.01 | 1.23E-02 | 0.00 | 3.81E-01 | - | - | - |
| Root vegetables, carrot | Food | 11841 | -0.02 | 0.01 | 3.08E-02 | 1.27E-01 | 64 | 11617 | -0.01 | 0.01 | 1.77E-01 | 4.36E-01 | 111 | -0.02 | 0.01 | 1.26E-02 | 0.00 | 6.00E-01 | - | - | - |
| Chips, popcorn, salted nuts | Food | 11841 | 0.01 | 0.01 | 1.16E-01 | 2.87E-01 | 106 | 11617 | 0.02 | 0.01 | 5.92E-02 | 2.11E-01 | 77 | 0.01 | 0.01 | 1.43E-02 | 0.00 | 8.60E-01 | - | - | - |
| Appetite status | Psychosocial | 11684 | 0.01 | 0.01 | 9.98E-02 | 2.60E-01 | 101 | 11428 | 0.02 | 0.01 | 7.45E-02 | 2.55E-01 | 81 | 0.02 | 0.01 | 1.53E-02 | 0.00 | 9.15E-01 | - | - | - |
| Sucrose intake (g/day) | Nutrients | 11841 | -0.01 | 0.01 | 5.56E-01 | 7.39E-01 | 198 | 11617 | -0.03 | 0.01 | 4.60E-03 | **4.11E-02** | 31 | -0.02 | 0.01 | 1.57E-02 | 0.61 | 1.10E-01 | - | - | - |
| Sour milk, yoghurt (low fat) | Food | 11841 | -0.02 | 0.01 | 4.19E-02 | 1.55E-01 | 71 | 11617 | -0.01 | 0.01 | 1.87E-01 | 4.47E-01 | 116 | -0.01 | 0.01 | 1.71E-02 | 0.00 | 6.58E-01 | - | - | - |
| Linolenic acid intake (g/day) | Nutrients | 11841 | 0.02 | 0.01 | 5.18E-02 | 1.79E-01 | 76 | 11617 | 0.01 | 0.01 | 1.71E-01 | 4.35E-01 | 108 | 0.01 | 0.01 | 1.87E-02 | 0.00 | 7.19E-01 | - | - | - |
| Boiled or baked potato | Food | 11841 | 0.01 | 0.01 | 2.75E-01 | 4.78E-01 | 149 | 11617 | 0.02 | 0.01 | 2.72E-02 | 1.24E-01 | 60 | 0.02 | 0.01 | 2.00E-02 | 0.00 | 4.15E-01 | - | - | - |
| Number of people with whom you can speak openly | Social | 13077 | 0.01 | 0.01 | 2.14E-01 | 4.28E-01 | 131 | 12887 | 0.02 | 0.01 | 5.16E-02 | 1.96E-01 | 71 | 0.01 | 0.01 | 2.44E-02 | 0.00 | 6.06E-01 | - | - | - |
| Cohabitation: Live alone vs Only children | Social | 2062 | -0.09 | 0.05 | 9.83E-02 | 2.58E-01 | 100 | 2125 | -0.08 | 0.05 | 1.33E-01 | 3.67E-01 | 100 | -0.08 | 0.04 | 2.57E-02 | 0.00 | 8.88E-01 | - | - | - |
| Possibility to leave your work for a while to speak with a colleague | Psychosocial | 12765 | 0.01 | 0.01 | 1.81E-01 | 3.83E-01 | 124 | 12558 | 0.02 | 0.01 | 7.78E-02 | 2.56E-01 | 82 | 0.01 | 0.01 | 2.85E-02 | 0.00 | 7.42E-01 | - | - | - |
| Vitamin A intake (mg/day) | Nutrients | 11841 | 0.02 | 0.01 | 5.74E-02 | 1.87E-01 | 80 | 11617 | 0.01 | 0.01 | 2.36E-01 | 5.28E-01 | 124 | 0.02 | 0.01 | 3.03E-02 | 0.00 | 5.70E-01 | - | - | - |
| Number of social contacts with the same interests as you | Social | 13076 | 0.01 | 0.01 | 5.16E-01 | 7.04E-01 | 192 | 12884 | 0.02 | 0.01 | 1.64E-02 | 8.41E-02 | 54 | 0.01 | 0.01 | 3.17E-02 | 0.37 | 2.09E-01 | - | - | - |
| Sedentary or standing work | Physical activity | 12596 | -0.03 | 0.02 | 1.66E-01 | 3.73E-01 | 117 | 12402 | -0.03 | 0.02 | 1.12E-01 | 3.28E-01 | 95 | -0.03 | 0.01 | 3.57E-02 | 0.00 | 8.72E-01 | - | - | - |
| Frequency of engaging in clubs, associations or study circles | Social | 7851 | -0.01 | 0.01 | 3.23E-01 | 5.32E-01 | 160 | 7701 | -0.02 | 0.01 | 5.47E-02 | 1.99E-01 | 76 | -0.02 | 0.01 | 4.01E-02 | 0.00 | 4.98E-01 | - | - | - |
| Medioresinol intake (ug/day) | Nutrients | 11841 | 0.02 | 0.01 | 7.85E-02 | 2.23E-01 | 91 | 11617 | 0.01 | 0.01 | 2.74E-01 | 5.88E-01 | 129 | 0.01 | 0.01 | 4.30E-02 | 0.00 | 6.54E-01 | - | - | - |
| Fried potatoes, pommes frites | Food | 11841 | 0.02 | 0.01 | 5.79E-02 | 1.87E-01 | 81 | 11617 | 0.01 | 0.01 | 3.52E-01 | 6.72E-01 | 145 | 0.01 | 0.01 | 4.69E-02 | 0.00 | 4.74E-01 | - | - | - |

| | | | | | | | | | | | | | | | | | | | | | |
|---|---|---|---|---|---|---|---|---|---|---|---|---|---|---|---|---|---|---|---|---|---|
| Physical limitation to participate in moderately demanding activities: lifting or carrying grocery bags | General health | 9476 | 0.03 | 0.01 | 1.71E-02 | 8.28E-02 | 53 | 9170 | 0.00 | 0.01 | 6.87E-01 | 9.19E-01 | 207 | 0.01 | 0.01 | 4.86E-02 | 0.52 | 1.47E-01 | - | - | - |
| Salty fish | Food | 11841 | 0.01 | 0.01 | 2.17E-01 | 4.28E-01 | 132 | 11617 | 0.01 | 0.01 | 1.14E-01 | 3.29E-01 | 96 | 0.01 | 0.01 | 4.97E-02 | 0.00 | 6.80E-01 | - | - | - |
| Self rate of overall health | General health | 9516 | -0.02 | 0.01 | 1.40E-01 | 3.34E-01 | 110 | 9203 | -0.01 | 0.01 | 2.02E-01 | 4.74E-01 | 117 | -0.01 | 0.01 | 5.12E-02 | 0.00 | 8.96E-01 | - | - | - |
| Sausage, liver pate on bread | Food | 11841 | -0.01 | 0.01 | 4.86E-01 | 6.93E-01 | 183 | 11617 | -0.02 | 0.01 | 3.85E-02 | 1.67E-01 | 64 | -0.01 | 0.01 | 5.22E-02 | 0.00 | 3.17E-01 | - | - | - |
| Feel the need to reduce alcohol consumption | Alcohol | 12094 | -0.06 | 0.03 | 1.76E-02 | 8.28E-02 | 56 | 11864 | -0.01 | 0.03 | 7.13E-01 | 9.20E-01 | 212 | -0.04 | 0.02 | 5.30E-02 | 0.51 | 1.54E-01 | - | - | - |
| Ice cream | Food | 11841 | 0.03 | 0.01 | 2.79E-03 | **2.16E-02** | 34 | 11617 | 0.00 | 0.01 | 7.25E-01 | 9.20E-01 | 218 | 0.01 | 0.01 | 5.43E-02 | 0.81 | 2.03E-02 | - | - | - |
| Possibility to speak with colleagues during breaks | Psychosocial | 12839 | -0.02 | 0.01 | 4.93E-02 | 1.74E-01 | 74 | 12652 | -0.01 | 0.01 | 4.55E-01 | 7.55E-01 | 167 | -0.01 | 0.01 | 5.45E-02 | 0.00 | 3.94E-01 | - | - | - |
| Control over planning and execution of the workday | Psychosocial | 12897 | 0.01 | 0.01 | 9.78E-02 | 2.58E-01 | 99 | 12688 | 0.01 | 0.01 | 3.00E-01 | 6.15E-01 | 135 | 0.01 | 0.01 | 5.66E-02 | 0.00 | 6.68E-01 | - | - | - |
| Frequency of picking berries or mushrooms during leisure time | Physical activity | 4143 | 0.03 | 0.02 | 7.87E-02 | 2.23E-01 | 93 | 4227 | 0.02 | 0.02 | 3.52E-01 | 6.72E-01 | 144 | 0.02 | 0.01 | 5.71E-02 | 0.00 | 5.57E-01 | - | - | - |
| Enterolactone intake (ug/day) | Nutrients | 11841 | -0.02 | 0.01 | 1.19E-02 | 6.51E-02 | 48 | 11617 | 0.00 | 0.01 | 8.98E-01 | 9.56E-01 | 260 | -0.01 | 0.01 | 5.82E-02 | 0.64 | 9.64E-02 | - | - | - |
| High mental demand from job | Psychosocial | 12767 | 0.01 | 0.01 | 1.44E-01 | 3.39E-01 | 111 | 12593 | 0.01 | 0.01 | 2.23E-01 | 5.12E-01 | 121 | 0.01 | 0.01 | 5.82E-02 | 0.00 | 8.73E-01 | - | - | - |
| Medium beer | Alcohol | 11841 | 0.01 | 0.01 | 2.10E-01 | 4.28E-01 | 128 | 11617 | 0.01 | 0.01 | 1.57E-01 | 4.18E-01 | 104 | 0.01 | 0.01 | 5.93E-02 | 0.00 | 9.07E-01 | - | - | - |
| Snuff status: Former snuff users vs non-snuff users | Tobacco use | 10503 | 0.03 | 0.03 | 2.76E-01 | 4.78E-01 | 152 | 10228 | 0.05 | 0.03 | 1.16E-01 | 3.32E-01 | 97 | 0.04 | 0.02 | 6.06E-02 | 0.00 | 7.17E-01 | - | - | - |
| Rosehip, sweet syrup soup | Food | 11841 | -0.02 | 0.01 | 7.87E-02 | 2.23E-01 | 92 | 11617 | -0.01 | 0.01 | 3.65E-01 | 6.78E-01 | 149 | -0.01 | 0.01 | 6.17E-02 | 0.00 | 5.14E-01 | - | - | - |
| Overall state of health compared to other your age | General health | 3570 | -0.03 | 0.02 | 1.69E-01 | 3.73E-01 | 119 | 3709 | -0.02 | 0.02 | 2.11E-01 | 4.88E-01 | 119 | -0.02 | 0.01 | 6.32E-02 | 0.00 | 9.24E-01 | - | - | - |
| Monosaccharides intake (g/day) | Nutrients | 11841 | -0.01 | 0.01 | 3.63E-01 | 5.77E-01 | 165 | 11617 | -0.02 | 0.01 | 8.88E-02 | 2.76E-01 | 89 | -0.01 | 0.01 | 6.55E-02 | 0.00 | 5.64E-01 | - | - | - |
| Cohabitation: Live alone vs Other/others | Social | 1668 | 0.11 | 0.08 | 2.13E-01 | 4.28E-01 | 129 | 1722 | 0.12 | 0.09 | 1.78E-01 | 4.36E-01 | 113 | 0.11 | 0.06 | 6.66E-02 | 0.00 | 9.29E-01 | - | - | - |
| Overall state of health during the last year | General health | 13083 | -0.02 | 0.01 | 3.84E-02 | 1.46E-01 | 69 | 12901 | 0.00 | 0.01 | 6.15E-01 | 8.94E-01 | 190 | -0.01 | 0.01 | 6.80E-02 | 0.18 | 2.71E-01 | - | - | - |
| Margarine on bread | Food | 11841 | 0.00 | 0.01 | 9.33E-01 | 9.70E-01 | 253 | 11617 | -0.02 | 0.01 | 1.52E-02 | 8.25E-02 | 51 | -0.01 | 0.01 | 6.80E-02 | 0.61 | 1.08E-01 | - | - | - |
| Beta-sitostanol intake (mg/day) | Nutrients | 11841 | 0.01 | 0.01 | 2.31E-01 | 4.29E-01 | 142 | 11617 | 0.01 | 0.01 | 1.68E-01 | 4.35E-01 | 106 | 0.01 | 0.01 | 6.87E-02 | 0.00 | 8.79E-01 | - | - | - |
| Cookies, pastry | Food | 11841 | 0.02 | 0.01 | 4.95E-02 | 1.74E-01 | 75 | 11617 | 0.01 | 0.01 | 5.49E-01 | 8.26E-01 | 183 | 0.01 | 0.01 | 7.09E-02 | 0.00 | 3.28E-01 | - | - | - |
| Total protein intake (g/day) | Nutrients | 11841 | -0.02 | 0.01 | 2.80E-02 | 1.17E-01 | 63 | 11617 | 0.00 | 0.01 | 7.86E-01 | 9.28E-01 | 234 | -0.01 | 0.01 | 7.79E-02 | 0.44 | 1.80E-01 | - | - | - |
| Everyday exercise satisfaction | Physical activity | 4401 | -0.01 | 0.02 | 6.10E-01 | 7.87E-01 | 204 | 4512 | -0.03 | 0.02 | 5.24E-02 | 1.96E-01 | 74 | -0.02 | 0.01 | 8.40E-02 | 0.05 | 3.05E-01 | - | - | - |
| Frequency of dancing during leisure time | Physical activity | 3838 | -0.01 | 0.02 | 6.68E-01 | 8.32E-01 | 210 | 3892 | -0.04 | 0.02 | 4.37E-02 | 1.83E-01 | 66 | -0.02 | 0.01 | 8.53E-02 | 0.24 | 2.52E-01 | - | - | - |
| Selenium intake (ug/day) | Nutrients | 11841 | 0.00 | 0.01 | 9.19E-01 | 9.70E-01 | 247 | 11617 | 0.02 | 0.01 | 1.05E-02 | 6.73E-02 | 43 | 0.01 | 0.01 | 8.62E-02 | 0.72 | 5.70E-02 | - | - | - |
| Average portion size of potatoes/rice/pasta based on photographic illustration of four sizes (smallest to largest) | Food | 11841 | -0.01 | 0.01 | 1.83E-01 | 3.84E-01 | 125 | 11617 | -0.01 | 0.01 | 2.82E-01 | 6.01E-01 | 130 | -0.01 | 0.01 | 8.82E-02 | 0.00 | 8.69E-01 | - | - | - |
| Participation in associations or voluntary organisations | Social | 12953 | -0.04 | 0.02 | 1.29E-02 | 6.77E-02 | 50 | 12791 | 0.00 | 0.02 | 9.12E-01 | 9.56E-01 | 263 | -0.02 | 0.01 | 8.95E-02 | 0.70 | 6.83E-02 | - | - | - |
| Physical limitation to participate in moderately demanding activities: bending down or kneeling | General health | 9482 | 0.00 | 0.01 | 6.97E-01 | 8.37E-01 | 219 | 9155 | -0.02 | 0.01 | 4.67E-02 | 1.90E-01 | 68 | -0.01 | 0.01 | 9.18E-02 | 0.25 | 2.47E-01 | - | - | - |
| Oatflake, whole wheat, rye or barley porridge | Food | 11841 | -0.01 | 0.01 | 5.17E-01 | 7.04E-01 | 193 | 11617 | -0.02 | 0.01 | 7.96E-02 | 2.56E-01 | 86 | -0.01 | 0.01 | 9.29E-02 | 0.00 | 4.12E-01 | - | - | - |
| Sausage as main dish | Food | 11841 | 0.02 | 0.01 | 9.25E-03 | 5.65E-02 | 43 | 11617 | 0.00 | 0.01 | 7.81E-01 | 9.28E-01 | 232 | 0.01 | 0.01 | 9.86E-02 | 0.76 | 4.20E-02 | - | - | - |
| Frequency of gardening during leisure time | Physical activity | 4132 | 0.02 | 0.02 | 2.46E-01 | 4.47E-01 | 145 | 4212 | 0.02 | 0.02 | 2.64E-01 | 5.76E-01 | 126 | 0.02 | 0.01 | 1.07E-01 | 0.00 | 9.87E-01 | - | - | - |
| Stigmasterol intake (mg/day) | Nutrients | 11841 | -0.01 | 0.01 | 1.51E-01 | 3.44E-01 | 115 | 11617 | -0.01 | 0.01 | 4.18E-01 | 7.20E-01 | 160 | -0.01 | 0.01 | 1.11E-01 | 0.00 | 6.71E-01 | - | - | - |
| Pasta | Food | 11841 | -0.03 | 0.01 | 8.18E-03 | 5.12E-02 | 42 | 11617 | 0.00 | 0.01 | 6.48E-01 | 9.06E-01 | 198 | -0.01 | 0.01 | 1.14E-01 | 0.79 | 2.98E-02 | - | - | - |
| Sugar, honey, marmelade, jam | Food | 11841 | 0.02 | 0.01 | 5.97E-02 | 1.87E-01 | 83 | 11617 | 0.00 | 0.01 | 7.36E-01 | 9.22E-01 | 221 | 0.01 | 0.01 | 1.16E-01 | 0.16 | 2.75E-01 | - | - | - |
| Informed of having high blood pressure | General health | 13078 | -0.01 | 0.02 | 5.61E-01 | 7.42E-01 | 199 | 12896 | -0.03 | 0.02 | 1.09E-01 | 3.25E-01 | 93 | -0.02 | 0.01 | 1.24E-01 | 0.00 | 4.65E-01 | - | - | - |
| Strong beer | Alcohol | 11841 | 0.02 | 0.01 | 5.53E-02 | 1.84E-01 | 79 | 11617 | 0.00 | 0.01 | 7.92E-01 | 9.28E-01 | 235 | 0.01 | 0.01 | 1.24E-01 | 0.28 | 2.40E-01 | - | - | - |
| Sometimes physically straining work | Physical activity | 12596 | 0.02 | 0.02 | 2.40E-01 | 4.39E-01 | 144 | 12402 | 0.02 | 0.02 | 3.30E-01 | 6.52E-01 | 140 | 0.02 | 0.01 | 1.28E-01 | 0.00 | 9.00E-01 | - | - | - |
| Vitamin B12 intake (ug/day) | Nutrients | 11841 | 0.00 | 0.01 | 6.18E-01 | 7.89E-01 | 206 | 11617 | 0.01 | 0.01 | 1.00E-01 | 3.06E-01 | 91 | 0.01 | 0.01 | 1.32E-01 | 0.00 | 4.11E-01 | - | - | - |
| Iodine intake (ug/day) | Nutrients | 11841 | 0.00 | 0.01 | 6.96E-01 | 8.37E-01 | 218 | 11617 | 0.02 | 0.01 | 8.39E-02 | 2.67E-01 | 87 | 0.01 | 0.01 | 1.38E-01 | 0.00 | 3.32E-01 | - | - | - |
| Milk, sour milk (1,5%) | Beverage | 11841 | 0.02 | 0.01 | 1.92E-02 | 8.56E-02 | 58 | 11617 | 0.00 | 0.01 | 7.79E-01 | 9.28E-01 | 231 | 0.01 | 0.01 | 1.38E-01 | 0.70 | 6.64E-02 | - | - | - |
| Frequent social contacts with colleagues during work | Psychosocial | 12760 | -0.01 | 0.01 | 1.38E-01 | 3.33E-01 | 108 | 12548 | 0.00 | 0.01 | 5.69E-01 | 8.38E-01 | 188 | -0.01 | 0.01 | 1.45E-01 | 0.00 | 5.23E-01 | - | - | - |
| For how much of the time during the last four weeks have you felt tired? | General health | 9519 | 0.00 | 0.01 | 7.36E-01 | 8.62E-01 | 224 | 9178 | -0.02 | 0.01 | 8.81E-02 | 2.76E-01 | 88 | -0.01 | 0.01 | 1.47E-01 | 0.00 | 3.26E-01 | - | - | - |
| Sodium intake (mg/day) | Nutrients | 11840 | 0.01 | 0.01 | 5.03E-01 | 7.00E-01 | 189 | 11616 | 0.01 | 0.01 | 1.73E-01 | 4.35E-01 | 110 | 0.01 | 0.01 | 1.53E-01 | 0.00 | 6.09E-01 | - | - | - |
| For how much of the time during the last four weeks have you felt worn out? | General health | 9495 | 0.01 | 0.01 | 2.64E-01 | 4.68E-01 | 148 | 9145 | 0.01 | 0.01 | 3.73E-01 | 6.81E-01 | 150 | 0.01 | 0.01 | 1.55E-01 | 0.00 | 8.67E-01 | - | - | - |
| Breakfast habits: Gruel w/o sandwich for breakfast vs not breakfast at all | Food | 525 | -0.13 | 0.11 | 2.59E-01 | 4.63E-01 | 147 | 528 | -0.10 | 0.11 | 3.85E-01 | 6.93E-01 | 154 | -0.11 | 0.08 | 1.55E-01 | 0.00 | 8.60E-01 | - | - | - |
| Syringaresinol intake (ug/day) | Nutrients | 11841 | 0.01 | 0.01 | 2.35E-01 | 4.33E-01 | 143 | 11617 | 0.01 | 0.01 | 4.31E-01 | 7.27E-01 | 164 | 0.01 | 0.01 | 1.62E-01 | 0.00 | 7.89E-01 | - | - | - |
| Breakfast habits: Only coffee/tea for breakfast vs not breakfast at all | Food | 869 | 0.01 | 0.08 | 8.88E-01 | 9.65E-01 | 242 | 921 | -0.15 | 0.08 | 4.46E-02 | 1.84E-01 | 67 | -0.08 | 0.06 | 1.62E-01 | 0.55 | 1.38E-01 | - | - | - |
| Satisfaction with accomodation | Psychosocial | 11697 | 0.01 | 0.01 | 2.31E-01 | 4.29E-01 | 141 | 11431 | 0.01 | 0.01 | 4.59E-01 | 7.56E-01 | 168 | 0.01 | 0.01 | 1.70E-01 | 0.00 | 7.54E-01 | - | - | - |
| Do you feel important and appreciated outside your home? | Psychosocial | 11686 | -0.01 | 0.01 | 1.92E-01 | 4.01E-01 | 126 | 11423 | -0.01 | 0.01 | 5.35E-01 | 8.18E-01 | 181 | -0.01 | 0.01 | 1.74E-01 | 0.00 | 6.23E-01 | - | - | - |
| Sour milk, yoghurt (3% fat) | Food | 11841 | -0.02 | 0.01 | 4.18E-02 | 1.55E-01 | 70 | 11617 | 0.00 | 0.01 | 8.82E-01 | 9.47E-01 | 258 | -0.01 | 0.01 | 1.74E-01 | 0.57 | 1.27E-01 | - | - | - |
| Pinoresinol intake (ug/day) | Nutrients | 11841 | -0.01 | 0.01 | 2.25E-01 | 4.28E-01 | 136 | 11617 | -0.01 | 0.01 | 5.01E-01 | 7.89E-01 | 176 | -0.01 | 0.01 | 1.81E-01 | 0.00 | 7.14E-01 | - | - | - |
| Skill demand from job | Psychosocial | 12843 | -0.01 | 0.01 | 3.71E-01 | 5.81E-01 | 168 | 12652 | -0.01 | 0.01 | 3.21E-01 | 6.39E-01 | 139 | -0.01 | 0.01 | 1.82E-01 | 0.00 | 9.38E-01 | - | - | - |
| Extent to what your physical and emotional health disrupted your usual social life during the last four weeks | General health | 9514 | -0.01 | 0.01 | 2.25E-01 | 4.28E-01 | 137 | 9183 | -0.01 | 0.01 | 5.00E-01 | 7.89E-01 | 175 | -0.01 | 0.01 | 1.83E-01 | 0.00 | 6.90E-01 | - | - | - |
| Sweets | Food | 11841 | 0.01 | 0.01 | 2.98E-01 | 5.03E-01 | 156 | 11617 | 0.01 | 0.01 | 4.00E-01 | 7.06E-01 | 157 | 0.01 | 0.01 | 1.84E-01 | 0.00 | 8.85E-01 | - | - | - |
| Do you feel important and appreciated in your home? | Psychosocial | 11608 | 0.02 | 0.01 | 9.58E-02 | 2.58E-01 | 97 | 11336 | 0.00 | 0.01 | 8.68E-01 | 9.44E-01 | 253 | 0.01 | 0.01 | 1.92E-01 | 0.10 | 2.93E-01 | - | - | - |
| Get sick more often than other people | General health | 9499 | 0.00 | 0.01 | 7.83E-01 | 8.84E-01 | 233 | 9170 | -0.02 | 0.01 | 1.21E-01 | 3.39E-01 | 98 | -0.01 | 0.01 | 1.95E-01 | 0.00 | 3.62E-01 | - | - | - |
| Physical limitation to participate in moderately demanding activities: walking more than 2 km | General health | 9494 | 0.00 | 0.01 | 8.30E-01 | 9.29E-01 | 235 | 9174 | -0.02 | 0.01 | 1.10E-01 | 3.25E-01 | 94 | -0.01 | 0.01 | 1.99E-01 | 0.00 | 3.23E-01 | - | - | - |
| Cohabitation: Live alone vs Adult and children | Social | 6431 | -0.04 | 0.03 | 2.22E-01 | 4.28E-01 | 134 | 6472 | -0.02 | 0.03 | 5.54E-01 | 8.26E-01 | 186 | -0.03 | 0.02 | 1.99E-01 | 0.00 | 6.59E-01 | - | - | - |
| Enterodiol intake (ug/day) | Nutrients | 11841 | -0.01 | 0.01 | 1.38E-01 | 3.33E-01 | 109 | 11617 | 0.00 | 0.01 | 8.03E-01 | 9.28E-01 | 239 | -0.01 | 0.01 | 2.13E-01 | 0.00 | 3.99E-01 | - | - | - |
| Years smoking | Tobacco use | 11291 | 0.00 | 0.01 | 6.89E-01 | 8.35E-01 | 217 | 11093 | 0.01 | 0.01 | 1.79E-01 | 4.36E-01 | 114 | 0.01 | 0.01 | 2.19E-01 | 0.00 | 5.01E-01 | - | - | - |
| Emotional problems that made you do less than you wanted during the last four weeks | General health | 9520 | 0.03 | 0.03 | 2.75E-01 | 4.78E-01 | 150 | 9184 | -0.09 | 0.03 | 5.63E-03 | **4.73E-02** | 33 | -0.03 | 0.02 | 2.21E-01 | 0.87 | 5.49E-03 | - | - | - |
| Sum of phytosterols intake (mg/day) | Nutrients | 11841 | 0.01 | 0.01 | 4.05E-01 | 6.12E-01 | 173 | 11617 | 0.01 | 0.01 | 3.74E-01 | 6.81E-01 | 152 | 0.01 | 0.01 | 2.23E-01 | 0.00 | 9.55E-01 | - | - | - |
| Wine | Alcohol | 11841 | -0.01 | 0.01 | 3.47E-01 | 5.64E-01 | 162 | 11617 | -0.01 | 0.01 | 4.35E-01 | 7.28E-01 | 165 | -0.01 | 0.01 | 2.24E-01 | 0.00 | 9.05E-01 | - | - | - |
| Excellent health | General health | 9494 | -0.01 | 0.01 | 1.48E-01 | 3.42E-01 | 114 | 9175 | 0.00 | 0.01 | 8.22E-01 | 9.33E-01 | 244 | -0.01 | 0.01 | 2.37E-01 | 0.00 | 3.81E-01 | - | - | - |
| Physical limitation to participate in steneous activities: running, lifting heavy objects, taking part in physically demanding sports | General health | 9496 | 0.01 | 0.01 | 2.08E-01 | 4.28E-01 | 127 | 9181 | 0.00 | 0.01 | 7.12E-01 | 9.20E-01 | 211 | 0.01 | 0.01 | 2.47E-01 | 0.00 | 5.31E-01 | - | - | - |
| Brewed (filtered) coffee | Beverage | 11841 | 0.01 | 0.01 | 1.10E-01 | 2.80E-01 | 103 | 11617 | 0.00 | 0.01 | 9.81E-01 | 9.84E-01 | 276 | 0.01 | 0.01 | 2.48E-01 | 0.18 | 2.71E-01 | - | - | - |
| Lean fish (e.g. perch, bass, cod) | Food | 11841 | 0.00 | 0.01 | 7.38E-01 | 8.62E-01 | 225 | 11617 | 0.02 | 0.01 | 4.80E-02 | 1.90E-01 | 70 | 0.01 | 0.01 | 2.48E-01 | 0.63 | 1.01E-01 | - | - | - |
| Travel to work: Walk to work vs passive travel to work | Physical activity | 8626 | 0.04 | 0.03 | 2.26E-01 | 4.28E-01 | 138 | 8587 | 0.01 | 0.03 | 7.99E-01 | 9.28E-01 | 238 | 0.02 | 0.02 | 3.02E-01 | 0.00 | 4.94E-01 | - | - | - |
| For how much of the time during the last four weeks have you felt calm and serene? | General health | 9505 | 0.01 | 0.01 | 2.48E-01 | 4.47E-01 | 146 | 9182 | 0.00 | 0.01 | 7.74E-01 | 9.28E-01 | 229 | 0.01 | 0.01 | 3.07E-01 | 0.00 | 5.35E-01 | - | - | - |
| Satisfaction with work situation | Psychosocial | 11555 | 0.00 | 0.01 | 6.33E-01 | 8.04E-01 | 207 | 11265 | -0.01 | 0.01 | 3.39E-01 | 6.61E-01 | 142 | -0.01 | 0.01 | 3.11E-01 | 0.00 | 7.34E-01 | - | - | - |
| Snuff status: Snuff users vs non-snuff users | Tobacco use | 11048 | -0.03 | 0.03 | 2.81E-01 | 4.83E-01 | 153 | 10961 | -0.01 | 0.03 | 7.24E-01 | 9.20E-01 | 217 | -0.02 | 0.02 | 3.11E-01 | 0.00 | 6.08E-01 | - | - | - |
| Self rate of overall health compared to a year ago | General health | 9429 | -0.02 | 0.01 | 6.12E-02 | 1.87E-01 | 86 | 9109 | 0.00 | 0.01 | 6.44E-01 | 9.06E-01 | 197 | -0.01 | 0.01 | 3.14E-01 | 0.64 | 9.55E-02 | - | - | - |
| Cohabitation: Live alone vs Only one adult (spouse, partner) | Social | 7391 | 0.04 | 0.03 | 1.72E-01 | 3.73E-01 | 121 | 7238 | 0.00 | 0.03 | 9.52E-01 | 9.66E-01 | 273 | 0.02 | 0.02 | 3.18E-01 | 0.00 | 3.49E-01 | - | - | - |
| Confidence status | Psychosocial | 11668 | 0.00 | 0.01 | 7.17E-01 | 8.52E-01 | 221 | 11389 | 0.02 | 0.01 | 7.44E-02 | 2.55E-01 | 80 | 0.01 | 0.01 | 3.18E-01 | 0.57 | 1.28E-01 | - | - | - |
| Mood status | Psychosocial | 11674 | 0.01 | 0.01 | 1.68E-01 | 3.73E-01 | 118 | 11409 | 0.00 | 0.01 | 9.91E-01 | 9.91E-01 | 277 | 0.01 | 0.01 | 3.22E-01 | 0.00 | 3.37E-01 | - | - | - |
| Corn flakes | Food | 11841 | -0.02 | 0.01 | 6.48E-02 | 1.94E-01 | 88 | 11617 | 0.00 | 0.01 | 6.30E-01 | 8.97E-01 | 194 | -0.01 | 0.01 | 3.23E-01 | 0.63 | 1.02E-01 | - | - | - |
| Sum of all lignans intake (ug/day) | Nutrients | 11841 | 0.01 | 0.01 | 4.36E-01 | 6.48E-01 | 177 | 11617 | 0.01 | 0.01 | 5.37E-01 | 8.18E-01 | 182 | 0.01 | 0.01 | 3.23E-01 | 0.00 | 9.16E-01 | - | - | - |
| Secoisolariciresinol intake (ug/day) | Nutrients | 11841 | 0.01 | 0.01 | 4.39E-01 | 6.48E-01 | 178 | 11617 | 0.01 | 0.01 | 5.34E-01 | 8.18E-01 | 180 | 0.01 | 0.01 | 3.23E-01 | 0.00 | 9.17E-01 | - | - | - |
| Frequency of hunting or fishing during leisure time | Physical activity | 4104 | -0.01 | 0.02 | 7.19E-01 | 8.52E-01 | 222 | 4165 | 0.03 | 0.02 | 7.92E-02 | 2.56E-01 | 85 | 0.01 | 0.01 | 3.26E-01 | 0.56 | 1.33E-01 | - | - | - |
| Feel uneasy or guilty because of your way of drinking | Alcohol | 3925 | -0.05 | 0.05 | 3.30E-01 | 5.39E-01 | 161 | 4038 | -0.02 | 0.05 | 7.18E-01 | 9.20E-01 | 214 | -0.04 | 0.04 | 3.45E-01 | 0.00 | 6.65E-01 | - | - | - |
| Control over own work assignment | Psychosocial | 12880 | 0.00 | 0.01 | 9.70E-01 | 9.75E-01 | 261 | 12693 | 0.01 | 0.01 | 1.78E-01 | 4.36E-01 | 112 | 0.01 | 0.01 | 3.57E-01 | 0.00 | 3.25E-01 | - | - | - |
| Linoleic acid intake (g/day) | Nutrients | 11841 | 0.01 | 0.01 | 5.12E-01 | 7.04E-01 | 191 | 11617 | 0.01 | 0.01 | 5.22E-01 | 8.09E-01 | 179 | 0.01 | 0.01 | 3.60E-01 | 0.00 | 9.93E-01 | - | - | - |
| Breakfast habits: Coffee/tea and wheat buns or rusk for breakfast vs not breakfast at all | Food | 603 | -0.03 | 0.09 | 7.22E-01 | 8.52E-01 | 223 | 602 | -0.08 | 0.09 | 3.61E-01 | 6.76E-01 | 148 | -0.06 | 0.06 | 3.62E-01 | 0.00 | 7.02E-01 | - | - | - |
| Parents or siblings have diabetes | General health | 12782 | 0.00 | 0.02 | 9.20E-01 | 9.70E-01 | 248 | 12625 | -0.03 | 0.02 | 1.69E-01 | 4.35E-01 | 107 | -0.01 | 0.02 | 3.70E-01 | 0.09 | 2.94E-01 | - | - | - |
| Energy status | Psychosocial | 11674 | -0.01 | 0.01 | 4.87E-01 | 6.93E-01 | 185 | 11400 | 0.00 | 0.01 | 6.28E-01 | 8.97E-01 | 193 | -0.01 | 0.01 | 4.04E-01 | 0.00 | 8.83E-01 | - | - | - |
| For how much of the time during the last four weeks have you felt very nervous? | General health | 9502 | -0.01 | 0.01 | 3.90E-01 | 6.03E-01 | 170 | 9174 | 0.00 | 0.01 | 7.53E-01 | 9.27E-01 | 225 | -0.01 | 0.01 | 4.05E-01 | 0.00 | 6.99E-01 | - | - | - |
| Potassium intake (mg/day) | Nutrients | 11841 | 0.00 | 0.01 | 9.06E-01 | 9.70E-01 | 244 | 11617 | 0.01 | 0.01 | 2.89E-01 | 6.06E-01 | 132 | 0.01 | 0.01 | 4.11E-01 | 0.00 | 4.96E-01 | - | - | - |
| Marital status: Single vs Widow/widower | Social | 1464 | -0.10 | 0.09 | 2.76E-01 | 4.78E-01 | 151 | 1490 | -0.01 | 0.09 | 9.52E-01 | 9.66E-01 | 272 | -0.05 | 0.06 | 4.26E-01 | 0.00 | 4.56E-01 | - | - | - |
| For how much of the time during the last four weeks has your physical health or your emotional problems limited your ability to interact with others? | General health | 9450 | -0.01 | 0.01 | 2.14E-01 | 4.28E-01 | 130 | 9132 | 0.00 | 0.01 | 9.12E-01 | 9.56E-01 | 264 | -0.01 | 0.01 | 4.29E-01 | 0.00 | 3.29E-01 | - | - | - |
| Marital status: Single vs Married/partner | Social | 11859 | 0.03 | 0.03 | 4.05E-01 | 6.12E-01 | 174 | 11680 | 0.01 | 0.03 | 7.73E-01 | 9.28E-01 | 228 | 0.02 | 0.02 | 4.30E-01 | 0.00 | 6.95E-01 | - | - | - |
| Learn new things at job | Psychosocial | 12862 | -0.01 | 0.01 | 3.64E-01 | 5.77E-01 | 166 | 12644 | 0.02 | 0.01 | 4.35E-02 | 1.83E-01 | 65 | 0.00 | 0.01 | 4.32E-01 | 0.77 | 3.85E-02 | - | - | - |
| Distance to work in kilometers (one way) | Physical activity | 11087 | 0.01 | 0.01 | 2.24E-01 | 4.28E-01 | 135 | 10852 | 0.00 | 0.01 | 9.16E-01 | 9.56E-01 | 265 | 0.01 | 0.01 | 4.35E-01 | 0.00 | 3.49E-01 | - | - | - |
| Participation in sports or physical exercise associations | Social | 5960 | -0.04 | 0.03 | 1.45E-01 | 3.39E-01 | 112 | 5622 | 0.01 | 0.03 | 6.75E-01 | 9.18E-01 | 203 | -0.01 | 0.02 | 4.45E-01 | 0.42 | 1.88E-01 | - | - | - |
| Animal based protein intake (g/day) | Nutrients | 11841 | 0.00 | 0.01 | 9.08E-01 | 9.70E-01 | 245 | 11617 | 0.01 | 0.01 | 3.54E-01 | 6.72E-01 | 146 | 0.00 | 0.01 | 4.66E-01 | 0.00 | 5.61E-01 | - | - | - |
| White meat (poultry) | Food | 11841 | 0.00 | 0.01 | 7.70E-01 | 8.77E-01 | 231 | 11617 | 0.01 | 0.01 | 4.65E-01 | 7.61E-01 | 169 | 0.00 | 0.01 | 4.71E-01 | 0.00 | 7.50E-01 | - | - | - |

| | | | | | | | | | | | | | | | | | | | | | |
|---|---|---|---|---|---|---|---|---|---|---|---|---|---|---|---|---|---|---|---|---|---|
| Satisfaction with economy | Psychosocial | 11686 | 0.00 | 0.01 | 9.75E-01 | 9.75E-01 | 263 | 11415 | -0.01 | 0.01 | 3.33E-01 | 6.55E-01 | 141 | 0.00 | 0.01 | 4.79E-01 | 0.00 | 5.08E-01 | - | - | - |
| Campestanol intake (mg/day) | Nutrients | 11841 | 0.00 | 0.01 | 8.44E-01 | 9.41E-01 | 236 | 11617 | 0.01 | 0.01 | 4.36E-01 | 7.28E-01 | 166 | 0.00 | 0.01 | 4.93E-01 | 0.00 | 6.75E-01 | - | - | - |
| Frequncy of social contacts with colleagues during leisure time | Psychosocial | 12507 | 0.01 | 0.01 | 2.29E-01 | 4.29E-01 | 140 | 12301 | 0.00 | 0.01 | 8.06E-01 | 9.28E-01 | 240 | 0.00 | 0.01 | 4.94E-01 | 0.04 | 3.07E-01 | - | - | - |
| Sleep status | Sleep | 11684 | 0.00 | 0.01 | 9.66E-01 | 9.75E-01 | 260 | 11431 | 0.01 | 0.01 | 3.60E-01 | 6.76E-01 | 147 | 0.00 | 0.01 | 4.98E-01 | 0.00 | 5.36E-01 | - | - | - |
| Matairesinol intake (ug/day) | Nutrients | 11841 | 0.01 | 0.01 | 4.72E-01 | 6.85E-01 | 181 | 11617 | 0.00 | 0.01 | 8.38E-01 | 9.44E-01 | 246 | 0.00 | 0.01 | 5.12E-01 | 0.00 | 7.18E-01 | - | - | - |
| Close relationship with anyone | Social | 13048 | -0.01 | 0.01 | 4.44E-01 | 6.52E-01 | 179 | 12867 | 0.00 | 0.01 | 8.75E-01 | 9.45E-01 | 256 | 0.00 | 0.01 | 5.17E-01 | 0.00 | 6.62E-01 | - | - | - |
| Physical limitation that limited your ability to perform certain work tasks or other activities during the last four weeks | General health | 9484 | 0.03 | 0.03 | 2.90E-01 | 4.92E-01 | 155 | 9151 | -0.06 | 0.03 | 5.21E-02 | 1.96E-01 | 72 | -0.01 | 0.02 | 5.19E-01 | 0.78 | 3.24E-02 | - | - | - |
| Number of social interactions during a normal week | Social | 13089 | 0.00 | 0.01 | 5.85E-01 | 7.58E-01 | 203 | 12898 | 0.00 | 0.01 | 7.18E-01 | 9.20E-01 | 213 | 0.00 | 0.01 | 5.20E-01 | 0.00 | 8.99E-01 | - | - | - |
| Physical limitation to participate in moderately demanding activities: moving a table, vacuuming, walking in the forest or gardening | General health | 9489 | 0.02 | 0.01 | 1.11E-01 | 2.80E-01 | 104 | 9175 | -0.01 | 0.01 | 4.81E-01 | 7.71E-01 | 172 | 0.00 | 0.01 | 5.31E-01 | 0.63 | 1.01E-01 | - | - | - |
| Support from others | Social | 13091 | -0.02 | 0.01 | 5.50E-02 | 1.84E-01 | 78 | 12904 | 0.01 | 0.01 | 2.97E-01 | 6.14E-01 | 134 | 0.00 | 0.01 | 5.38E-01 | 0.77 | 3.60E-02 | - | - | - |
| Tiamin intake (mg/day) | Nutrients | 11841 | 0.00 | 0.01 | 6.87E-01 | 8.35E-01 | 216 | 11617 | 0.01 | 0.01 | 2.02E-01 | 4.74E-01 | 118 | 0.00 | 0.01 | 5.42E-01 | 0.30 | 2.34E-01 | - | - | - |
| Physical limitation that made you not being able to perform certain work tasks or other activities during the last four weeks | General health | 9467 | 0.00 | 0.03 | 8.72E-01 | 9.52E-01 | 241 | 9146 | -0.02 | 0.03 | 4.92E-01 | 7.83E-01 | 174 | -0.01 | 0.02 | 5.47E-01 | 0.00 | 7.11E-01 | - | - | - |
| Frequency of walking during leisure time | Physical activity | 12444 | -0.01 | 0.01 | 1.52E-01 | 3.44E-01 | 116 | 12284 | 0.01 | 0.01 | 5.53E-01 | 8.26E-01 | 185 | 0.00 | 0.01 | 5.48E-01 | 0.51 | 1.53E-01 | - | - | - |
| Physical limitation that made you do less than you wanted during the last four weeks | General health | 9507 | 0.05 | 0.03 | 5.42E-02 | 1.84E-01 | 77 | 9179 | -0.03 | 0.03 | 2.73E-01 | 5.88E-01 | 128 | 0.01 | 0.02 | 5.55E-01 | 0.79 | 3.09E-02 | - | - | - |
| White cabbage, lettuce, lettuce cabbage, spinach, borecole | Food | 11841 | -0.01 | 0.01 | 4.98E-01 | 6.97E-01 | 188 | 11617 | 0.00 | 0.01 | 9.03E-01 | 9.56E-01 | 261 | 0.00 | 0.01 | 5.65E-01 | 0.00 | 7.06E-01 | - | - | - |
| Pain during the last four weeks | General health | 9514 | 0.00 | 0.01 | 9.24E-01 | 9.70E-01 | 250 | 9169 | 0.01 | 0.01 | 4.82E-01 | 7.71E-01 | 173 | 0.00 | 0.01 | 5.72E-01 | 0.00 | 6.65E-01 | - | - | - |
| Work shifts/weekends | Psychosocial | 12749 | -0.02 | 0.02 | 2.83E-01 | 4.83E-01 | 154 | 12557 | 0.01 | 0.02 | 7.73E-01 | 9.28E-01 | 227 | -0.01 | 0.01 | 5.77E-01 | 0.00 | 3.36E-01 | - | - | - |
| Patience status | Psychosocial | 11676 | 0.01 | 0.01 | 3.20E-01 | 5.29E-01 | 159 | 11409 | 0.00 | 0.01 | 8.08E-01 | 9.28E-01 | 241 | 0.00 | 0.01 | 5.91E-01 | 0.00 | 3.83E-01 | - | - | - |
| Sum of Lariciresinol, Matairesinol, Pinoresinol, Secoisolariciresinol intake (ug/day) | Nutrients | 11841 | -0.01 | 0.01 | 5.42E-01 | 7.23E-01 | 197 | 11617 | 0.00 | 0.01 | 8.94E-01 | 9.56E-01 | 259 | 0.00 | 0.01 | 5.97E-01 | 0.00 | 7.40E-01 | - | - | - |
| Job demands to work very fast | Psychosocial | 12887 | 0.00 | 0.01 | 5.77E-01 | 7.51E-01 | 202 | 12696 | 0.00 | 0.01 | 8.53E-01 | 9.44E-01 | 250 | 0.00 | 0.01 | 5.97E-01 | 0.00 | 7.95E-01 | - | - | - |
| Participation in other association | Social | 5960 | 0.02 | 0.03 | 3.77E-01 | 5.87E-01 | 169 | 5622 | -0.04 | 0.03 | 9.68E-02 | 2.98E-01 | 90 | -0.01 | 0.02 | 6.04E-01 | 0.70 | 6.94E-02 | - | - | - |
| Smoking status: Former occasional smokers vs non-smokers | Tobacco use | 7849 | -0.04 | 0.03 | 2.21E-01 | 4.28E-01 | 133 | 7702 | 0.01 | 0.03 | 6.31E-01 | 8.97E-01 | 195 | -0.01 | 0.02 | 6.04E-01 | 0.32 | 2.27E-01 | - | - | - |
| Disaccharides intake (g/day) | Nutrients | 11841 | 0.01 | 0.01 | 4.87E-01 | 6.93E-01 | 184 | 11617 | -0.01 | 0.01 | 1.48E-01 | 4.03E-01 | 102 | 0.00 | 0.01 | 6.06E-01 | 0.57 | 1.29E-01 | - | - | - |
| Travel to work: Irregular travel mode to work vs passive travel to work | Physical activity | 8551 | -0.03 | 0.03 | 3.03E-01 | 5.08E-01 | 157 | 8365 | 0.06 | 0.03 | 7.79E-02 | 2.56E-01 | 83 | 0.01 | 0.02 | 6.22E-01 | 0.75 | 4.74E-02 | - | - | - |
| Alcohol intake (g/day) | Alcohol | 11841 | 0.01 | 0.01 | 5.24E-01 | 7.10E-01 | 194 | 11617 | 0.00 | 0.01 | 9.51E-01 | 9.66E-01 | 270 | 0.00 | 0.01 | 6.24E-01 | 0.00 | 6.80E-01 | - | - | - |
| Light and physically active work | Physical activity | 12596 | -0.02 | 0.02 | 3.94E-01 | 6.06E-01 | 171 | 12402 | 0.00 | 0.02 | 8.61E-01 | 9.44E-01 | 252 | -0.01 | 0.01 | 6.25E-01 | 0.00 | 4.72E-01 | - | - | - |
| Sodas, soft drinks, juice | Beverage | 11841 | 0.02 | 0.01 | 7.27E-02 | 2.13E-01 | 90 | 11617 | -0.01 | 0.01 | 2.58E-01 | 5.71E-01 | 125 | 0.00 | 0.01 | 6.28E-01 | 0.77 | 3.88E-02 | - | - | - |
| Magnesium intake (mg/day) | Nutrients | 11841 | 0.00 | 0.01 | 7.41E-01 | 8.62E-01 | 226 | 11617 | 0.00 | 0.01 | 7.41E-01 | 9.23E-01 | 222 | 0.00 | 0.01 | 6.40E-01 | 0.00 | 9.95E-01 | - | - | - |
| Zinc intake (mg/day) | Nutrients | 11841 | -0.01 | 0.01 | 3.12E-01 | 5.19E-01 | 158 | 11617 | 0.00 | 0.01 | 7.07E-01 | 9.20E-01 | 210 | 0.00 | 0.01 | 6.47E-01 | 0.00 | 3.28E-01 | - | - | - |
| Marital status: Single vs Divorced/separated | Social | 2276 | 0.01 | 0.05 | 7.63E-01 | 8.74E-01 | 229 | 2318 | -0.04 | 0.04 | 3.48E-01 | 6.72E-01 | 143 | -0.01 | 0.03 | 6.48E-01 | 0.00 | 3.81E-01 | - | - | - |
| Contradictory demands in job | Psychosocial | 12823 | -0.01 | 0.01 | 3.63E-01 | 5.77E-01 | 164 | 12603 | 0.00 | 0.01 | 7.77E-01 | 9.28E-01 | 230 | 0.00 | 0.01 | 6.51E-01 | 0.00 | 4.02E-01 | - | - | - |
| Years using snuff | Tobacco use | 12408 | -0.01 | 0.01 | 4.11E-01 | 6.14E-01 | 176 | 12225 | 0.00 | 0.01 | 8.22E-01 | 9.33E-01 | 243 | 0.00 | 0.01 | 6.68E-01 | 0.00 | 4.61E-01 | - | - | - |
| Satisfaction with leisure time | Psychosocial | 11667 | 0.00 | 0.01 | 7.89E-01 | 8.87E-01 | 234 | 11403 | 0.00 | 0.01 | 7.46E-01 | 9.23E-01 | 223 | 0.00 | 0.01 | 6.76E-01 | 0.00 | 9.66E-01 | - | - | - |
| Memory status | Psychosocial | 11659 | 0.00 | 0.01 | 9.58E-01 | 9.75E-01 | 257 | 11386 | 0.01 | 0.01 | 5.21E-01 | 8.09E-01 | 178 | 0.00 | 0.01 | 6.78E-01 | 0.00 | 6.22E-01 | - | - | - |
| Repetitive job | Psychosocial | 12861 | 0.01 | 0.01 | 1.79E-01 | 3.82E-01 | 123 | 12666 | -0.01 | 0.01 | 4.26E-01 | 7.24E-01 | 163 | 0.00 | 0.01 | 6.89E-01 | 0.56 | 1.31E-01 | - | - | - |
| For how much of the time during the last four weeks have you felt full of energy? | General health | 9495 | 0.00 | 0.01 | 9.28E-01 | 9.70E-01 | 251 | 9167 | 0.00 | 0.01 | 6.38E-01 | 9.02E-01 | 196 | 0.00 | 0.01 | 6.91E-01 | 0.00 | 7.88E-01 | - | - | - |
| Beta-sitosterol intake (mg/day) | Nutrients | 11841 | 0.00 | 0.01 | 9.31E-01 | 9.70E-01 | 252 | 11617 | 0.00 | 0.01 | 6.68E-01 | 9.18E-01 | 201 | 0.00 | 0.01 | 7.18E-01 | 0.00 | 8.06E-01 | - | - | - |
| Hamburger | Food | 11841 | 0.01 | 0.01 | 1.73E-01 | 3.74E-01 | 122 | 11617 | -0.01 | 0.01 | 3.74E-01 | 6.81E-01 | 151 | 0.00 | 0.01 | 7.42E-01 | 0.61 | 1.11E-01 | - | - | - |
| Worsen in health in the future | General health | 9463 | 0.00 | 0.01 | 9.63E-01 | 9.75E-01 | 259 | 9150 | -0.01 | 0.01 | 6.10E-01 | 8.94E-01 | 189 | 0.00 | 0.01 | 7.44E-01 | 0.00 | 6.92E-01 | - | - | - |
| For how much of the time during the last four weeks have you felt happy? | General health | 9507 | 0.01 | 0.01 | 5.27E-01 | 7.11E-01 | 195 | 9172 | 0.00 | 0.01 | 8.58E-01 | 9.44E-01 | 251 | 0.00 | 0.01 | 7.48E-01 | 0.00 | 5.65E-01 | - | - | - |
| Vitamin B6 intake (mg/day) | Nutrients | 11841 | -0.01 | 0.01 | 4.07E-01 | 6.12E-01 | 175 | 11617 | 0.00 | 0.01 | 6.90E-01 | 9.19E-01 | 208 | 0.00 | 0.01 | 7.54E-01 | 0.00 | 3.87E-01 | - | - | - |
| Long-term sickness | General health | 12715 | -0.01 | 0.02 | 7.64E-01 | 8.74E-01 | 230 | 12564 | 0.00 | 0.02 | 9.18E-01 | 9.56E-01 | 266 | 0.00 | 0.02 | 7.76E-01 | 0.00 | 8.89E-01 | - | - | - |
| For how much of the time during the last four weeks have you felt so depressed that nothing could cheer you up? | General health | 9514 | 0.00 | 0.01 | 9.13E-01 | 9.70E-01 | 246 | 9181 | 0.00 | 0.01 | 6.16E-01 | 8.94E-01 | 191 | 0.00 | 0.01 | 7.79E-01 | 0.00 | 6.66E-01 | - | - | - |
| People to ask for help apart from the ones at home | Social | 13035 | -0.01 | 0.03 | 7.59E-01 | 8.74E-01 | 228 | 12851 | 0.00 | 0.03 | 9.36E-01 | 9.66E-01 | 268 | -0.01 | 0.02 | 7.84E-01 | 0.00 | 8.74E-01 | - | - | - |
| Light but partly physically active work | Physical activity | 12596 | 0.00 | 0.02 | 9.75E-01 | 9.75E-01 | 262 | 12402 | 0.01 | 0.02 | 6.76E-01 | 9.18E-01 | 204 | 0.00 | 0.02 | 7.86E-01 | 0.00 | 7.50E-01 | - | - | - |
| Whole grain intake (g/day) | Food | 11841 | 0.00 | 0.01 | 9.60E-01 | 9.75E-01 | 258 | 11617 | 0.00 | 0.01 | 6.70E-01 | 9.18E-01 | 202 | 0.00 | 0.01 | 7.93E-01 | 0.00 | 7.34E-01 | - | - | - |
| Receive hugs to comfort and support you | Social | 12951 | 0.02 | 0.02 | 4.82E-01 | 6.93E-01 | 182 | 12769 | -0.01 | 0.02 | 7.21E-01 | 9.20E-01 | 215 | 0.00 | 0.02 | 7.98E-01 | 0.00 | 4.56E-01 | - | - | - |
| Teetotaler | Alcohol | 4406 | 0.00 | 0.05 | 9.51E-01 | 9.75E-01 | 254 | 4527 | 0.02 | 0.05 | 7.81E-01 | 9.28E-01 | 233 | 0.01 | 0.04 | 8.13E-01 | 0.00 | 8.75E-01 | - | - | - |
| Coffee rolls/buns, rusk | Food | 11841 | 0.00 | 0.01 | 9.56E-01 | 9.75E-01 | 255 | 11617 | 0.00 | 0.01 | 7.94E-01 | 9.28E-01 | 236 | 0.00 | 0.01 | 8.23E-01 | 0.00 | 8.84E-01 | - | - | - |
| For how much of the time during the last four weeks have you felt gloomy and sad? | General health | 9502 | -0.01 | 0.01 | 6.15E-01 | 7.89E-01 | 205 | 9165 | 0.00 | 0.01 | 8.50E-01 | 9.44E-01 | 248 | 0.00 | 0.01 | 8.27E-01 | 0.00 | 6.22E-01 | - | - | - |
| How much has the pain during the last four weeks disturbed your normal work? | General health | 9493 | 0.00 | 0.01 | 8.60E-01 | 9.50E-01 | 238 | 9155 | 0.00 | 0.01 | 9.05E-01 | 9.56E-01 | 262 | 0.00 | 0.01 | 8.34E-01 | 0.00 | 9.66E-01 | - | - | - |
| Whole grain soft bread | Food | 11841 | 0.00 | 0.01 | 9.58E-01 | 9.75E-01 | 256 | 11617 | 0.00 | 0.01 | 7.31E-01 | 9.21E-01 | 220 | 0.00 | 0.01 | 8.40E-01 | 0.00 | 7.77E-01 | - | - | - |
| Meat on bread | Food | 11841 | -0.01 | 0.01 | 5.06E-01 | 7.01E-01 | 190 | 11617 | 0.00 | 0.01 | 6.94E-01 | 9.19E-01 | 209 | 0.00 | 0.01 | 8.40E-01 | 0.00 | 4.56E-01 | - | - | - |
| Lariciresinol intake (ug/day) | Nutrients | 11841 | 0.00 | 0.01 | 6.65E-01 | 8.32E-01 | 209 | 11617 | 0.00 | 0.01 | 8.76E-01 | 9.45E-01 | 257 | 0.00 | 0.01 | 8.40E-01 | 0.00 | 6.79E-01 | - | - | - |
| Physical limitation to participate in moderately demanding activities: walking up several stairs | General health | 9490 | 0.00 | 0.01 | 6.69E-01 | 8.32E-01 | 211 | 9165 | -0.01 | 0.01 | 4.78E-01 | 7.71E-01 | 171 | 0.00 | 0.01 | 8.46E-01 | 0.00 | 4.19E-01 | - | - | - |
| Smoked fish/meat | Food | 11841 | 0.01 | 0.01 | 4.95E-01 | 6.97E-01 | 186 | 11617 | -0.01 | 0.01 | 3.03E-01 | 6.17E-01 | 136 | 0.00 | 0.01 | 8.51E-01 | 0.33 | 2.22E-01 | - | - | - |
| Permanent employment | Psychosocial | 11487 | 0.01 | 0.02 | 7.56E-01 | 8.74E-01 | 227 | 11349 | 0.00 | 0.02 | 9.44E-01 | 9.66E-01 | 269 | 0.00 | 0.01 | 8.64E-01 | 0.00 | 7.88E-01 | - | - | - |
| Cheese 28% | Food | 11841 | 0.01 | 0.01 | 5.69E-01 | 7.47E-01 | 200 | 11617 | 0.00 | 0.01 | 7.23E-01 | 9.20E-01 | 216 | 0.00 | 0.01 | 8.76E-01 | 0.00 | 5.14E-01 | - | - | - |
| Liquor, spirits | Alcohol | 11841 | 0.00 | 0.01 | 8.71E-01 | 9.52E-01 | 240 | 11617 | 0.00 | 0.01 | 9.56E-01 | 9.66E-01 | 274 | 0.00 | 0.01 | 8.81E-01 | 0.00 | 9.34E-01 | - | - | - |
| Pancake, waffle, Swedish dumpling | Food | 11841 | -0.01 | 0.01 | 5.71E-01 | 7.47E-01 | 201 | 11617 | 0.01 | 0.01 | 4.25E-01 | 7.24E-01 | 162 | 0.00 | 0.01 | 8.84E-01 | 0.00 | 3.33E-01 | - | - | - |
| Ingenuity or creativity demand from job | Psychosocial | 12817 | 0.00 | 0.01 | 6.74E-01 | 8.32E-01 | 213 | 12617 | 0.00 | 0.01 | 8.21E-01 | 9.33E-01 | 242 | 0.00 | 0.01 | 8.88E-01 | 0.00 | 6.48E-01 | - | - | - |
| White (soft) bread, thin crisp bread | Food | 11841 | 0.01 | 0.01 | 3.98E-01 | 6.08E-01 | 172 | 11617 | -0.01 | 0.01 | 5.04E-01 | 7.89E-01 | 177 | 0.00 | 0.01 | 8.96E-01 | 0.13 | 2.85E-01 | - | - | - |
| Phosphate intake (mg/day) | Nutrients | 11841 | 0.00 | 0.01 | 6.40E-01 | 8.09E-01 | 208 | 11617 | 0.00 | 0.01 | 7.47E-01 | 9.23E-01 | 224 | 0.00 | 0.01 | 9.10E-01 | 0.00 | 5.78E-01 | - | - | - |
| Hearing status | General health | 11689 | 0.00 | 0.01 | 8.51E-01 | 9.44E-01 | 237 | 11432 | 0.00 | 0.01 | 9.51E-01 | 9.66E-01 | 271 | 0.00 | 0.01 | 9.28E-01 | 0.00 | 8.60E-01 | - | - | - |
| Apple, pear, peach, orange, mandarin and grapefruit | Food | 11841 | 0.00 | 0.01 | 6.81E-01 | 8.33E-01 | 214 | 11617 | 0.00 | 0.01 | 7.63E-01 | 9.28E-01 | 226 | 0.00 | 0.01 | 9.30E-01 | 0.00 | 6.15E-01 | - | - | - |
| Enough time for job assignments | Psychosocial | 12819 | 0.01 | 0.01 | 3.59E-01 | 5.77E-01 | 163 | 12637 | -0.01 | 0.01 | 2.92E-01 | 6.08E-01 | 133 | 0.00 | 0.01 | 9.31E-01 | 0.49 | 1.63E-01 | - | - | - |
| Vision status | General health | 11681 | -0.01 | 0.01 | 4.97E-01 | 6.97E-01 | 187 | 11412 | 0.01 | 0.01 | 5.50E-01 | 8.26E-01 | 184 | 0.00 | 0.01 | 9.49E-01 | 0.00 | 3.67E-01 | - | - | - |
| Self-employed | Psychosocial | 11487 | 0.01 | 0.03 | 8.65E-01 | 9.52E-01 | 239 | 11349 | 0.00 | 0.03 | 9.32E-01 | 9.66E-01 | 267 | 0.00 | 0.02 | 9.52E-01 | 0.00 | 8.57E-01 | - | - | - |
| For how much of the time during the last four weeks have you felt really alert and strong? | General health | 9492 | 0.00 | 0.01 | 6.81E-01 | 8.33E-01 | 215 | 9154 | 0.00 | 0.01 | 6.25E-01 | 8.97E-01 | 192 | 0.00 | 0.01 | 9.54E-01 | 0.00 | 5.24E-01 | - | - | - |
| Smoking status: Former smokers vs non-smokers | Tobacco use | 9561 | -0.02 | 0.02 | 3.68E-01 | 5.80E-01 | 167 | 9253 | 0.02 | 0.02 | 4.06E-01 | 7.07E-01 | 159 | 0.00 | 0.02 | 9.55E-01 | 0.33 | 2.21E-01 | - | - | - |
| Physical limitation that reduced the normal time spent at work or in other activities during the last four weeks | General health | 9516 | -0.01 | 0.03 | 7.79E-01 | 8.83E-01 | 232 | 9177 | 0.01 | 0.03 | 7.28E-01 | 9.20E-01 | 219 | 0.00 | 0.02 | 9.57E-01 | 0.00 | 6.56E-01 | - | - | - |
| Frequency of shoverling snow during leisure time | Physical activity | 4174 | -0.01 | 0.02 | 5.30E-01 | 7.11E-01 | 196 | 4287 | 0.01 | 0.02 | 5.65E-01 | 8.37E-01 | 187 | 0.00 | 0.01 | 9.62E-01 | 0.00 | 3.94E-01 | - | - | - |
| As healthy as anyone | General health | 9474 | 0.00 | 0.01 | 9.23E-01 | 9.70E-01 | 249 | 9144 | 0.00 | 0.01 | 9.73E-01 | 9.80E-01 | 275 | 0.00 | 0.01 | 9.64E-01 | 0.00 | 9.27E-01 | - | - | - |
| Participation in study circles | Social | 5960 | 0.01 | 0.04 | 7.04E-01 | 8.41E-01 | 220 | 5622 | -0.02 | 0.04 | 6.79E-01 | 9.18E-01 | 205 | 0.00 | 0.03 | 9.81E-01 | 0.00 | 5.74E-01 | - | - | - |
| Butter on bread | Food | 11841 | 0.00 | 0.01 | 8.95E-01 | 9.68E-01 | 243 | 11617 | 0.00 | 0.01 | 8.68E-01 | 9.44E-01 | 254 | 0.00 | 0.01 | 9.83E-01 | 0.00 | 8.33E-01 | - | - | - |
| Oil for cooking | Food | 11841 | 0.00 | 0.01 | 6.74E-01 | 8.32E-01 | 212 | 11617 | 0.00 | 0.01 | 6.83E-01 | 9.18E-01 | 206 | 0.00 | 0.01 | 9.84E-01 | 0.00 | 5.58E-01 | - | - | - |
| Snore during sleep | Sleep | 8008 | 0.02 | 0.01 | - | - | 283 | 7749 | 0.04 | 0.01 | 8.93E-04 | **1.30E-02** | 19 | 0.03 | 0.01 | - | 0.00 | 3.91E-01 | - | - | - |
| Soft cheese | Food | 1252 | 0.04 | 0.03 | - | - | 266 | 1298 | -0.05 | 0.03 | 5.45E-02 | 1.99E-01 | 75 | -0.01 | 0.02 | - | 0.83 | 1.49E-02 | - | - | - |
| Soft whey cheese | Food | 1252 | -0.05 | 0.03 | - | - | 267 | 1298 | -0.04 | 0.03 | 1.63E-01 | 4.29E-01 | 105 | -0.04 | 0.02 | - | 0.00 | 8.46E-01 | - | - | - |
| Breath-holds during sleep | Sleep | 6008 | -0.01 | 0.01 | - | - | 284 | 5748 | 0.01 | 0.01 | 3.16E-01 | 6.39E-01 | 137 | 0.00 | 0.01 | - | 0.13 | 2.84E-01 | - | - | - |
| Risk of sleeping while sitting still after having lunch | Sleep | 8579 | 0.00 | 0.01 | - | - | 282 | 8299 | -0.01 | 0.01 | 3.77E-01 | 6.82E-01 | 153 | -0.01 | 0.01 | - | 0.00 | 6.45E-01 | - | - | - |
| Potato salad | Food | 1252 | 0.03 | 0.03 | - | - | 270 | 1298 | 0.03 | 0.03 | 3.96E-01 | 7.03E-01 | 156 | 0.03 | 0.02 | - | 0.00 | 9.60E-01 | - | - | - |
| Blood based food | Food | 1252 | -0.11 | 0.03 | - | - | 272 | 1298 | -0.03 | 0.03 | 3.94E-01 | 7.03E-01 | 155 | -0.07 | 0.02 | - | 0.66 | 8.47E-02 | - | - | - |
| Mixed frozen vegetables | Food | 1252 | 0.03 | 0.03 | - | - | 268 | 1298 | -0.03 | 0.03 | 4.06E-01 | 7.07E-01 | 158 | 0.01 | 0.02 | - | 0.47 | 1.71E-01 | - | - | - |
| Times during last year that you felt guilty because of your drinking | Alcohol | 8470 | -0.01 | 0.01 | - | - | 278 | 8112 | -0.01 | 0.01 | 4.19E-01 | 7.20E-01 | 161 | -0.01 | 0.01 | - | 0.00 | 7.75E-01 | - | - | - |
| Risk of sleeping while sitting and reading | Sleep | 8516 | -0.02 | 0.01 | - | - | 279 | 8231 | -0.01 | 0.01 | 4.69E-01 | 7.64E-01 | 170 | -0.01 | 0.01 | - | 0.00 | 3.54E-01 | - | - | - |
| Mashed potato | Food | 1252 | -0.05 | 0.03 | - | - | 269 | 1298 | -0.02 | 0.04 | 6.67E-01 | 9.18E-01 | 199 | -0.03 | 0.02 | - | 0.00 | 4.94E-01 | - | - | - |
| Liver, kidney | Food | 1252 | -0.01 | 0.03 | - | - | 273 | 1298 | 0.01 | 0.03 | 6.67E-01 | 9.18E-01 | 200 | 0.00 | 0.02 | - | 0.00 | 5.37E-01 | - | - | - |
| Blota (broth + bread) | Food | 1252 | 0.05 | 0.04 | - | - | 271 | 1298 | 0.01 | 0.04 | 7.97E-01 | 9.28E-01 | 237 | 0.03 | 0.03 | - | 0.00 | 4.24E-01 | - | - | - |
| Shellfish (e.g. shrimps, scallops) | Food | 1252 | 0.01 | 0.03 | - | - | 274 | 1298 | 0.01 | 0.03 | 8.26E-01 | 9.34E-01 | 245 | 0.01 | 0.02 | - | 0.00 | 9.91E-01 | - | - | - |
| Amount of alcohol drunk in a day | Alcohol | 8328 | 0.01 | 0.01 | - | - | 276 | 7984 | 0.00 | 0.01 | 8.53E-01 | 9.44E-01 | 249 | 0.00 | 0.01 | - | 0.00 | 7.91E-01 | - | - | - |
| Risk of sleeping while lying down resting in the afternoon | Sleep | 8573 | 0.00 | 0.01 | - | - | 281 | 8302 | 0.00 | 0.01 | 8.43E-01 | 9.44E-01 | 247 | 0.00 | 0.01 | - | 0.00 | 8.56E-01 | - | - | - |
| Risk of sleeping as a passenger in a car for one hour without break | Sleep | 8516 | -0.02 | 0.01 | 6.18E-02 | 1.87E-01 | 87 | 8235 | -0.03 | 0.01 | - | - | 284 | -0.02 | 0.01 | - | 0.00 | 4.83E-01 | - | - | - |
| Risk of sleeping while watching TV | Sleep | 8610 | 0.01 | 0.01 | 4.60E-01 | 6.73E-01 | 180 | 8321 | -0.01 | 0.01 | - | - | 282 | 0.00 | 0.01 | - | 0.00 | 3.59E-01 | - | - | - |
| Time spent in a week in moderately steneous activities | Physical activity | 8399 | -0.02 | 0.01 | - | - | 264 | 8078 | -0.01 | 0.01 | - | - | 278 | -0.01 | 0.01 | - | 0.00 | 7.36E-01 | - | - | - |

| | | | | | | | | | | | | | | | | | | | | | |
|---|---|---|---|---|---|---|---|---|---|---|---|---|---|---|---|---|---|---|---|---|---|
| Amount of exercise during the last 12 months | Physical activity | 8662 | -0.03 | 0.01 | - | - | 265 | 8354 | -0.01 | 0.01 | - | - | 279 | -0.02 | 0.01 | - | 0.24 | 2.51E-01 | - | - | - |
| Frequency of alcohol consumption | Alcohol | 8704 | -0.02 | 0.01 | - | - | 275 | 8413 | -0.01 | 0.01 | - | - | 280 | -0.02 | 0.01 | - | 0.00 | 3.78E-01 | - | - | - |
| Frequency of drinking six or more glasses at the same occasion | Alcohol | 8479 | 0.00 | 0.01 | - | - | 277 | 8130 | -0.01 | 0.01 | - | - | 281 | 0.00 | 0.01 | - | 0.00 | 5.26E-01 | - | - | - |
| Risk of sleeping while sitting inactive in a public place | Sleep | 8554 | -0.03 | 0.01 | - | - | 280 | 8274 | -0.02 | 0.01 | - | - | 283 | -0.03 | 0.01 | - | 0.00 | 7.42E-01 | - | - | - |

**Supplementary Table 10**. Linear mixed model association results for fasting glucose

| Description | Group | Training set | | | | | | Testing set | | | | | | Metanalysis | | | | | R2 | | |
|---|---|---|---|---|---|---|---|---|---|---|---|---|---|---|---|---|---|---|---|---|---|
| | | N | Effect estimate | S.E. | p-value | p-value$_{FDR}$ | p-value rank | N | Effect estimate | S.E. | p-value | p-value$_{FDR}$ | p-value rank | Effect estimate | S.E. | p-value | $I^2$ | Q p-value | marginal R2 | conditional R2 | R2 rank |
| Years smoking | Tobacco use | 26512 | 0.05 | 0.00 | 1.28E-26 | **3.46E-24** | 1 | 25794 | 0.04 | 0.00 | 3.38E-16 | **4.68E-14** | 2 | 0.05 | 0.00 | 1.34E-40 | 0.63 | 1.01E-01 | 0.07 | 0.25 | 27 |
| Number of cigarretes smoked per day (in groups) | Tobacco use | 24922 | 0.04 | 0.00 | 2.18E-18 | **2.93E-16** | 2 | 24552 | 0.04 | 0.00 | 8.33E-14 | **7.69E-12** | 3 | 0.04 | 0.00 | 1.93E-30 | 0.00 | 3.83E-01 | 0.06 | 0.25 | 28 |
| Parents or siblings have diabetes | General health | 29796 | 0.08 | 0.01 | 9.72E-13 | **6.53E-11** | 4 | 29133 | 0.10 | 0.01 | 2.83E-19 | **7.85E-17** | 1 | 0.09 | 0.01 | 5.99E-30 | 0.55 | 1.35E-01 | 0.06 | 0.25 | 31 |
| Smoking status: Smokers vs non-smokers | Tobacco use | 19306 | 0.12 | 0.01 | 9.01E-18 | **8.08E-16** | 3 | 18963 | 0.10 | 0.01 | 3.82E-13 | **2.12E-11** | 5 | 0.11 | 0.01 | 4.07E-29 | 0.03 | 3.11E-01 | 0.06 | 0.26 | 30 |
| Exercise during the last three months | Physical activity | 30012 | -0.03 | 0.00 | 1.50E-12 | **8.09E-11** | 5 | 29322 | -0.03 | 0.00 | 1.64E-13 | **1.13E-11** | 4 | -0.03 | 0.00 | 1.73E-24 | 0.00 | 7.17E-01 | 0.06 | 0.25 | 33 |
| Participation in associations or voluntary organisations | Social | 30131 | -0.05 | 0.01 | 3.13E-09 | **1.40E-07** | 6 | 29475 | -0.05 | 0.01 | 6.07E-09 | **2.40E-07** | 7 | -0.05 | 0.01 | 1.03E-16 | 0.00 | 9.35E-01 | 0.06 | 0.25 | 32 |
| Lariciresinol intake (ug/day) | Nutrients | 27762 | -0.03 | 0.00 | 3.29E-08 | **1.11E-06** | 8 | 27043 | -0.03 | 0.00 | 1.37E-07 | **3.79E-06** | 10 | -0.03 | 0.00 | 2.26E-14 | 0.00 | 9.78E-01 | 0.08 | 0.29 | 3 |
| Whole grain intake (g/day) | Food | 27762 | -0.02 | 0.00 | 9.85E-07 | **1.89E-05** | 14 | 27043 | -0.03 | 0.00 | 1.05E-08 | **3.65E-07** | 8 | -0.02 | 0.00 | 6.70E-14 | 0.00 | 4.60E-01 | 0.08 | 0.29 | 4 |
| Fitness status | Physical activity | 24274 | -0.03 | 0.01 | 2.61E-07 | **5.41E-06** | 13 | 23843 | -0.03 | 0.01 | 1.57E-07 | **3.96E-06** | 11 | -0.03 | 0.00 | 2.01E-13 | 0.00 | 8.94E-01 | 0.05 | 0.26 | 41 |
| Sum of Lariciresinol, Matairesinol, Pinoresinol, Secoisolariciresinol intake (ug/day) | Nutrients | 27762 | -0.02 | 0.00 | 1.33E-07 | **3.59E-06** | 10 | 27043 | -0.02 | 0.00 | 4.70E-07 | **1.08E-05** | 12 | -0.02 | 0.00 | 3.01E-13 | 0.00 | 9.83E-01 | 0.08 | 0.29 | 5 |
| Informed of having high blood pressure | General health | 30337 | 0.05 | 0.01 | 1.94E-05 | **2.48E-04** | 21 | 29645 | 0.06 | 0.01 | 3.27E-09 | **1.51E-07** | 6 | 0.05 | 0.01 | 6.53E-13 | 0.37 | 2.07E-01 | 0.06 | 0.25 | 34 |
| Pinoresinol intake (ug/day) | Nutrients | 27762 | -0.02 | 0.00 | 1.84E-07 | **4.44E-06** | 11 | 27043 | -0.02 | 0.00 | 3.75E-06 | **6.49E-05** | 16 | -0.02 | 0.00 | 3.29E-12 | 0.00 | 7.84E-01 | 0.08 | 0.29 | 7 |
| Eat breakfast from 2000 | Food | 16929 | -0.12 | 0.02 | 1.98E-07 | **4.44E-06** | 12 | 16524 | -0.10 | 0.02 | 4.55E-06 | **7.25E-05** | 17 | -0.11 | 0.02 | 4.76E-12 | 0.00 | 5.87E-01 | 0.06 | 0.32 | 38 |
| Fibre intake (g/day) | Nutrients | 27762 | -0.02 | 0.00 | 1.20E-05 | **1.70E-04** | 19 | 27043 | -0.03 | 0.00 | 8.45E-08 | **2.60E-06** | 9 | -0.02 | 0.00 | 6.56E-12 | 0.00 | 4.01E-01 | 0.08 | 0.29 | 8 |
| Average portion size of potatoes/rice/pasta based on photographic illustration of four sizes (smallest to largest) | Food | 27762 | 0.02 | 0.00 | 3.56E-06 | **5.98E-05** | 16 | 27043 | 0.02 | 0.01 | 2.20E-06 | **4.06E-05** | 15 | 0.02 | 0.00 | 3.52E-11 | 0.00 | 8.37E-01 | 0.06 | 0.25 | 39 |
| Beta-sitosterol intake (mg/day) | Nutrients | 27762 | -0.03 | 0.00 | 9.56E-09 | **3.67E-07** | 7 | 27043 | -0.02 | 0.00 | 7.15E-04 | **5.66E-03** | 35 | -0.02 | 0.00 | 9.00E-11 | 0.58 | 1.24E-01 | 0.08 | 0.29 | 10 |
| Fiber cereals | Food | 27762 | -0.02 | 0.00 | 6.27E-06 | **9.93E-05** | 17 | 27043 | -0.02 | 0.00 | 9.90E-06 | **1.37E-04** | 20 | -0.02 | 0.00 | 2.63E-10 | 0.00 | 9.43E-01 | 0.08 | 0.29 | 9 |
| Plant based protein intake (g/day) | Nutrients | 27762 | -0.02 | 0.00 | 2.85E-05 | **3.25E-04** | 23 | 27043 | -0.02 | 0.00 | 4.71E-06 | **7.25E-05** | 18 | -0.02 | 0.00 | 6.06E-10 | 0.00 | 6.80E-01 | 0.08 | 0.29 | 11 |
| Magnesium intake (mg/day) | Nutrients | 27762 | -0.02 | 0.00 | 7.06E-05 | **6.33E-04** | 30 | 27043 | -0.02 | 0.00 | 2.05E-06 | **4.06E-05** | 14 | -0.02 | 0.00 | 7.50E-10 | 0.00 | 5.00E-01 | 0.08 | 0.29 | 12 |
| Sum of phytosterols intake (mg/day) | Nutrients | 27762 | -0.02 | 0.00 | 7.83E-08 | **2.34E-06** | 9 | 27043 | -0.01 | 0.00 | 5.89E-03 | **3.02E-02** | 54 | -0.02 | 0.00 | 7.39E-09 | 0.67 | 8.24E-02 | 0.08 | 0.29 | 13 |
| Educational level | Psychosocial | 30377 | -0.02 | 0.00 | 4.71E-05 | **4.69E-04** | 27 | 29655 | -0.02 | 0.00 | 4.00E-05 | **5.27E-04** | 21 | -0.02 | 0.00 | 7.40E-09 | 0.00 | 9.20E-01 | 0.06 | 0.25 | 36 |
| Cambridge physical activity index | Physical activity | 29051 | -0.02 | 0.00 | 2.66E-05 | **3.25E-04** | 22 | 28391 | -0.02 | 0.00 | 7.57E-05 | **9.11E-04** | 23 | -0.02 | 0.00 | 7.88E-09 | 0.00 | 9.36E-01 | 0.06 | 0.25 | 37 |
| Average portion size of vegetables based on photographic illustration of four sizes (smallest to largest) | Food | 27762 | -0.01 | 0.00 | 1.45E-03 | **7.37E-03** | 53 | 27043 | -0.02 | 0.00 | 1.71E-06 | **3.63E-05** | 13 | -0.02 | 0.00 | 1.99E-08 | 0.35 | 2.14E-01 | 0.06 | 0.25 | 40 |
| Monosaccharides intake (g/day) | Nutrients | 27762 | -0.02 | 0.00 | 1.29E-05 | **1.73E-04** | 20 | 27043 | -0.02 | 0.00 | 6.17E-04 | **5.34E-03** | 32 | -0.02 | 0.00 | 3.40E-08 | 0.00 | 5.87E-01 | 0.08 | 0.29 | 14 |
| Overall state of health during the last year | General health | 30328 | -0.02 | 0.00 | 3.00E-06 | **5.38E-05** | 15 | 29638 | -0.01 | 0.00 | 1.92E-03 | **1.33E-02** | 40 | -0.02 | 0.00 | 3.48E-08 | 0.03 | 3.11E-01 | 0.06 | 0.25 | 35 |
| Whole grain soft bread | Food | 27762 | -0.02 | 0.00 | 2.90E-05 | **3.25E-04** | 24 | 27043 | -0.02 | 0.00 | 5.31E-04 | **4.74E-03** | 31 | -0.02 | 0.00 | 6.07E-08 | 0.00 | 6.99E-01 | 0.08 | 0.29 | 16 |
| Matairesinol intake (ug/day) | Nutrients | 27762 | -0.02 | 0.00 | 7.85E-05 | **6.60E-04** | 32 | 27043 | -0.02 | 0.00 | 2.25E-04 | **2.40E-03** | 26 | -0.02 | 0.00 | 6.54E-08 | 0.00 | 9.47E-01 | 0.08 | 0.29 | 15 |
| Apple, pear, peach, orange, mandarin and grapefruit | Food | 27762 | -0.02 | 0.00 | 6.00E-05 | **5.71E-04** | 28 | 27043 | -0.02 | 0.00 | 3.21E-04 | **3.15E-03** | 28 | -0.02 | 0.00 | 7.18E-08 | 0.00 | 8.58E-01 | 0.08 | 0.29 | 18 |
| Sum of all lignans intake (ug/day) | Nutrients | 27762 | -0.02 | 0.00 | 7.50E-05 | **6.51E-04** | 31 | 27043 | -0.02 | 0.00 | 3.30E-04 | **3.15E-03** | 29 | -0.02 | 0.00 | 9.13E-08 | 0.00 | 8.83E-01 | 0.08 | 0.29 | 19 |
| Folic acid intake (ug/day) | Nutrients | 27762 | -0.02 | 0.00 | 3.62E-05 | **3.74E-04** | 26 | 27043 | -0.02 | 0.00 | 7.01E-04 | **5.66E-03** | 34 | -0.02 | 0.00 | 9.95E-08 | 0.00 | 6.83E-01 | 0.08 | 0.29 | 20 |
| Oatflake, whole wheat, rye or barley porridge | Food | 27762 | -0.01 | 0.00 | 3.38E-03 | **1.49E-02** | 61 | 27043 | -0.02 | 0.00 | 7.89E-06 | **1.15E-04** | 19 | -0.02 | 0.00 | 1.89E-07 | 0.29 | 2.34E-01 | 0.08 | 0.29 | 17 |
| Appetite status | Psychosocial | 24286 | -0.02 | 0.00 | 1.65E-04 | **1.23E-03** | 36 | 23861 | -0.02 | 0.00 | 4.09E-04 | **3.77E-03** | 30 | -0.02 | 0.00 | 2.40E-07 | 0.00 | 9.27E-01 | 0.05 | 0.27 | 42 |
| Self rate of overall health | General health | 14299 | -0.03 | 0.01 | 1.49E-04 | **1.14E-03** | 35 | 14122 | -0.02 | 0.01 | 6.43E-04 | **5.40E-03** | 33 | -0.03 | 0.00 | 3.40E-07 | 0.00 | 8.27E-01 | - | - | - |
| Stigmasterol intake (mg/day) | Nutrients | 27762 | -0.02 | 0.00 | 1.20E-04 | **9.75E-04** | 33 | 27043 | -0.01 | 0.00 | 2.16E-03 | **1.46E-02** | 41 | -0.02 | 0.00 | 9.64E-07 | 0.00 | 6.51E-01 | 0.08 | 0.29 | 21 |
| Smoking status: Former smokers vs non-smokers | Tobacco use | 21827 | 0.04 | 0.01 | 3.52E-04 | **2.22E-03** | 42 | 21080 | 0.04 | 0.01 | 9.84E-04 | **7.36E-03** | 37 | 0.04 | 0.01 | 1.18E-06 | 0.00 | 8.87E-01 | 0.06 | 0.26 | 29 |
| Teetotaler | Alcohol | 19025 | -0.05 | 0.02 | 1.23E-03 | **6.50E-03** | 50 | 18496 | -0.06 | 0.02 | 2.92E-04 | **2.99E-03** | 27 | -0.06 | 0.01 | 1.30E-06 | 0.00 | 7.03E-01 | 0.08 | 0.25 | 1 |
| Breakfast habits: Porridge w/o sandwich for breakfast vs not breakfast at all | Food | 4167 | -0.09 | 0.03 | 2.88E-03 | **1.38E-02** | 56 | 4232 | -0.11 | 0.03 | 1.42E-04 | **1.57E-03** | 25 | -0.10 | 0.02 | 1.54E-06 | 0.00 | 6.00E-01 | 0.07 | 0.31 | 26 |
| Liquor, spirits | Alcohol | 27762 | 0.02 | 0.00 | 3.13E-05 | **3.36E-04** | 25 | 27043 | 0.01 | 0.00 | 2.42E-02 | 8.44E-02 | 79 | 0.01 | 0.00 | 5.66E-06 | 0.45 | 1.77E-01 | - | - | - |
| Beta-sitostanol intake (mg/day) | Nutrients | 27762 | -0.02 | 0.00 | 1.35E-04 | **1.07E-03** | 34 | 27043 | -0.01 | 0.00 | 1.08E-02 | **4.98E-02** | 60 | -0.01 | 0.00 | 6.18E-06 | 0.00 | 4.26E-01 | 0.08 | 0.29 | 24 |
| Campestanol intake (mg/day) | Nutrients | 27762 | -0.01 | 0.00 | 5.28E-04 | **3.16E-03** | 45 | 27043 | -0.01 | 0.00 | 3.67E-03 | **2.21E-02** | 46 | -0.01 | 0.00 | 6.39E-06 | 0.00 | 7.68E-01 | 0.08 | 0.29 | 23 |
| Carbohydrates intake (g/day) | Nutrients | 27762 | -0.02 | 0.00 | 3.18E-04 | **2.16E-03** | 39 | 27043 | -0.01 | 0.00 | 5.78E-03 | **3.02E-02** | 53 | -0.01 | 0.00 | 6.52E-06 | 0.00 | 6.16E-01 | 0.08 | 0.29 | 22 |
| Permanent employment | Psychosocial | 25839 | -0.04 | 0.01 | 8.22E-06 | **1.23E-04** | 18 | 25115 | -0.02 | 0.01 | 7.06E-02 | 1.76E-01 | 111 | -0.03 | 0.01 | 8.51E-06 | 0.70 | 6.73E-02 | - | - | - |
| Frequency of picking berries or mushrooms during leisure time | Physical activity | 18001 | -0.02 | 0.01 | 3.54E-04 | **2.22E-03** | 43 | 17492 | -0.01 | 0.01 | 6.88E-03 | **3.40E-02** | 56 | -0.02 | 0.00 | 8.89E-06 | 0.00 | 5.64E-01 | 0.08 | 0.25 | 2 |
| Syringaresinol intake (ug/day) | Nutrients | 27762 | -0.01 | 0.00 | 9.64E-04 | **5.41E-03** | 48 | 27043 | -0.01 | 0.00 | 3.49E-03 | **2.19E-02** | 44 | -0.01 | 0.00 | 1.06E-05 | 0.00 | 8.63E-01 | 0.08 | 0.29 | 25 |
| White cabbage, lettuce, lettuce cabbage, spinach, borecole | Food | 27762 | -0.02 | 0.00 | 1.78E-04 | **1.29E-03** | 37 | 27043 | -0.01 | 0.00 | 1.63E-02 | 6.47E-02 | 70 | -0.01 | 0.00 | 1.25E-05 | 0.00 | 3.93E-01 | - | - | - |
| Alcohol intake (g/day) | Alcohol | 27762 | 0.02 | 0.00 | 3.33E-04 | **2.19E-03** | 41 | 27043 | 0.01 | 0.00 | 1.72E-02 | 6.71E-02 | 71 | 0.01 | 0.00 | 2.27E-05 | 0.00 | 4.37E-01 | - | - | - |
| Frequency of hunting or fishing during leisure time | Physical activity | 17736 | -0.02 | 0.01 | 2.84E-03 | **1.38E-02** | 55 | 17201 | -0.02 | 0.01 | 8.05E-03 | **3.84E-02** | 58 | -0.02 | 0.00 | 6.70E-05 | 0.00 | 8.43E-01 | 0.08 | 0.24 | 6 |
| Root vegetables, carrot | Food | 27762 | -0.01 | 0.00 | 3.22E-03 | **1.47E-02** | 59 | 27043 | -0.01 | 0.00 | 1.20E-02 | 5.10E-02 | 65 | -0.01 | 0.00 | 1.11E-04 | 0.00 | 8.19E-01 | - | - | - |
| Beta-carotene intake (mg/day) | Nutrients | 27762 | -0.01 | 0.00 | 4.06E-03 | **1.73E-02** | 63 | 27043 | -0.01 | 0.00 | 1.10E-02 | 5.01E-02 | 61 | -0.01 | 0.00 | 1.26E-04 | 0.00 | 8.76E-01 | - | - | - |
| Breakfast habits: Gruel w/o sandwich for breakfast vs not breakfast at all | Food | 1434 | -0.11 | 0.05 | 1.91E-02 | 5.89E-02 | 87 | 1485 | -0.14 | 0.05 | 2.69E-03 | **1.73E-02** | 43 | -0.12 | 0.03 | 1.58E-04 | 0.00 | 6.01E-01 | - | - | - |
| Excellent health | General health | 14247 | -0.02 | 0.01 | 3.12E-03 | **1.45E-02** | 58 | 14065 | -0.02 | 0.01 | 1.79E-02 | 6.79E-02 | 73 | -0.02 | 0.00 | 1.62E-04 | 0.00 | 7.20E-01 | - | - | - |
| Tea | Beverage | 27762 | -0.02 | 0.00 | 2.56E-04 | **1.81E-03** | 38 | 27043 | -0.01 | 0.00 | 1.17E-01 | 2.58E-01 | 126 | -0.01 | 0.00 | 1.99E-04 | 0.49 | 1.59E-01 | - | - | - |
| Breakfast habits: Coffee/tea and wheat buns or rusk for breakfast vs not breakfast at all | Food | 1580 | -0.11 | 0.04 | 7.25E-03 | **2.83E-02** | 69 | 1554 | -0.10 | 0.04 | 1.27E-02 | 5.33E-02 | 66 | -0.11 | 0.03 | 2.46E-04 | 0.00 | 9.35E-01 | - | - | - |
| Coffee rolls/buns, rusk | Food | 27762 | -0.01 | 0.00 | 1.58E-02 | 5.20E-02 | 81 | 27043 | -0.01 | 0.00 | 5.48E-03 | **2.94E-02** | 50 | -0.01 | 0.00 | 2.47E-04 | 0.00 | 7.44E-01 | - | - | - |
| Fat intake (g/day) | Nutrients | 27762 | 0.01 | 0.00 | 7.20E-03 | **2.83E-02** | 68 | 27043 | 0.01 | 0.00 | 1.31E-02 | 5.43E-02 | 67 | 0.01 | 0.00 | 2.56E-04 | 0.00 | 9.42E-01 | - | - | - |
| Secoisolariciresinol intake (ug/day) | Nutrients | 27762 | -0.01 | 0.00 | 5.31E-02 | 1.25E-01 | 114 | 27043 | -0.02 | 0.00 | 1.17E-03 | **8.55E-03** | 38 | -0.01 | 0.00 | 2.66E-04 | 0.00 | 3.22E-01 | - | - | - |
| Frequncy of social contacts with colleagues during leisure time | Psychosocial | 29009 | -0.01 | 0.00 | 1.84E-02 | 5.82E-02 | 85 | 28382 | -0.01 | 0.00 | 6.19E-03 | **3.12E-02** | 55 | -0.01 | 0.00 | 3.23E-04 | 0.00 | 7.27E-01 | - | - | - |
| Cholesterol intake (g/day) | Nutrients | 27762 | 0.02 | 0.00 | 8.99E-04 | **5.14E-03** | 47 | 27043 | 0.01 | 0.00 | 8.28E-02 | 2.03E-01 | 113 | 0.01 | 0.00 | 3.25E-04 | 0.10 | 2.91E-01 | - | - | - |
| Hamburger | Food | 27762 | 0.01 | 0.00 | 1.54E-03 | **7.68E-03** | 54 | 27043 | 0.01 | 0.00 | 6.30E-02 | 1.63E-01 | 107 | 0.01 | 0.00 | 3.55E-04 | 0.00 | 3.93E-01 | - | - | - |
| Zinc intake (mg/day) | Nutrients | 27762 | -0.01 | 0.00 | 8.79E-03 | **3.28E-02** | 72 | 27043 | -0.01 | 0.00 | 1.76E-02 | 6.76E-02 | 72 | -0.01 | 0.00 | 4.08E-04 | 0.00 | 9.20E-01 | - | - | - |
| Fried potatoes, pommes frites | Food | 27762 | 0.01 | 0.00 | 4.77E-03 | **1.98E-02** | 64 | 27043 | 0.01 | 0.00 | 3.24E-02 | 1.03E-01 | 86 | 0.01 | 0.00 | 4.41E-04 | 0.00 | 6.60E-01 | - | - | - |
| Saturated fat intake (g/day) | Nutrients | 27762 | 0.01 | 0.00 | 8.78E-03 | **3.28E-02** | 71 | 27043 | 0.01 | 0.00 | 2.20E-02 | 7.97E-02 | 76 | 0.01 | 0.00 | 5.08E-04 | 0.00 | 8.67E-01 | - | - | - |
| High physical demand from job | Physical activity | 29938 | 0.01 | 0.00 | 1.06E-03 | **5.84E-03** | 49 | 29291 | 0.01 | 0.00 | 1.08E-01 | 2.43E-01 | 123 | 0.01 | 0.00 | 5.38E-04 | 0.24 | 2.51E-01 | - | - | - |
| Number of social contacts with the same interests as you | Social | 30278 | -0.01 | 0.00 | 1.16E-02 | **4.11E-02** | 76 | 29582 | -0.01 | 0.00 | 1.96E-02 | 7.23E-02 | 75 | -0.01 | 0.00 | 5.88E-04 | 0.00 | 9.34E-01 | - | - | - |
| Rosehip, sweet syrup soup | Food | 27762 | 0.01 | 0.00 | 3.35E-03 | **1.49E-02** | 60 | 27043 | 0.01 | 0.00 | 6.75E-02 | 1.71E-01 | 109 | 0.01 | 0.00 | 7.18E-04 | 0.00 | 4.76E-01 | - | - | - |
| Sodas, soft drinks, juice | Beverage | 27762 | 0.01 | 0.00 | 8.46E-02 | 1.75E-01 | 129 | 27043 | 0.01 | 0.00 | 2.30E-03 | **1.52E-02** | 42 | 0.01 | 0.00 | 8.04E-04 | 0.04 | 3.09E-01 | - | - | - |
| Cohabitation: Live alone vs Adult and children | Social | 17509 | -0.05 | 0.02 | 1.23E-03 | **6.50E-03** | 51 | 17093 | -0.02 | 0.02 | 1.52E-01 | 2.98E-01 | 141 | -0.04 | 0.01 | 8.46E-04 | 0.27 | 2.43E-01 | - | - | - |
| Enterodiol intake (ug/day) | Nutrients | 27762 | 0.01 | 0.00 | 5.25E-02 | 1.25E-01 | 113 | 27043 | 0.01 | 0.00 | 5.27E-03 | **2.94E-02** | 49 | 0.01 | 0.00 | 8.66E-04 | 0.00 | 5.02E-01 | - | - | - |
| Frequent social contacts with colleagues during work | Psychosocial | 29543 | -0.01 | 0.00 | 1.26E-02 | **4.34E-02** | 78 | 28896 | -0.01 | 0.00 | 2.72E-02 | 9.19E-02 | 82 | -0.01 | 0.00 | 8.71E-04 | 0.00 | 8.88E-01 | - | - | - |
| Breakfast habits: Only coffee/tea for breakfast vs not breakfast at all | Food | 1892 | 0.10 | 0.04 | 1.43E-02 | **4.85E-02** | 79 | 1960 | 0.10 | 0.04 | 3.25E-02 | 1.03E-01 | 87 | 0.10 | 0.03 | 1.14E-03 | 0.00 | 9.29E-01 | - | - | - |
| Vitamin A intake (mg/day) | Nutrients | 27762 | 0.00 | 0.01 | 4.26E-01 | 5.65E-01 | 203 | 27043 | 0.02 | 0.01 | 1.14E-04 | **1.32E-03** | 24 | 0.01 | 0.00 | 1.16E-03 | 0.80 | 2.58E-02 | - | - | - |
| Light but partly physically active work | Physical activity | 29420 | -0.02 | 0.01 | 8.48E-02 | 1.75E-01 | 130 | 28753 | -0.03 | 0.01 | 3.91E-03 | **2.31E-02** | 47 | -0.03 | 0.01 | 1.19E-03 | 0.00 | 3.76E-01 | - | - | - |
| For how much of the time during the last four weeks have you felt very nervous? | General health | 14259 | 0.02 | 0.01 | 3.22E-04 | **2.16E-03** | 40 | 14068 | 0.01 | 0.01 | 3.74E-01 | 5.51E-01 | 188 | 0.02 | 0.00 | 1.30E-03 | 0.72 | 6.02E-02 | - | - | - |
| Tiamin intake (mg/day) | Nutrients | 27762 | -0.01 | 0.00 | 2.07E-01 | 3.35E-01 | 166 | 27043 | -0.02 | 0.00 | 9.67E-04 | **7.36E-03** | 36 | -0.01 | 0.00 | 1.41E-03 | 0.56 | 1.30E-01 | - | - | - |
| White (soft) bread, thin crisp bread | Food | 27762 | 0.01 | 0.00 | 5.34E-02 | 1.25E-01 | 115 | 27043 | 0.01 | 0.00 | 1.16E-02 | 5.01E-02 | 64 | 0.01 | 0.00 | 1.68E-03 | 0.00 | 6.29E-01 | - | - | - |
| Last time a colleague visited you at home | Psychosocial | 29436 | -0.01 | 0.00 | 1.41E-01 | 2.52E-01 | 150 | 28818 | -0.01 | 0.00 | 3.56E-03 | **2.19E-02** | 45 | -0.01 | 0.00 | 2.02E-03 | 0.12 | 2.85E-01 | - | - | - |
| Medioresinol intake (ug/day) | Nutrients | 27762 | -0.01 | 0.00 | 1.88E-02 | 5.87E-02 | 86 | 27043 | -0.01 | 0.00 | 4.94E-02 | 1.38E-01 | 99 | -0.01 | 0.00 | 2.24E-03 | 0.00 | 8.37E-01 | - | - | - |
| For how much of the time during the last four weeks have you felt worn out? | General health | 14235 | 0.02 | 0.01 | 2.57E-02 | 7.32E-02 | 94 | 14024 | 0.01 | 0.01 | 4.10E-02 | 1.18E-01 | 96 | 0.01 | 0.00 | 2.49E-03 | 0.00 | 9.32E-01 | - | - | - |
| Cheese 10-17% | Food | 27762 | -0.02 | 0.00 | 6.15E-05 | **5.71E-04** | 29 | 27043 | 0.00 | 0.00 | 8.97E-01 | 9.51E-01 | 259 | -0.01 | 0.00 | 2.82E-03 | 0.86 | 7.46E-03 | - | - | - |
| Palmitic acid intake (g/day) | Nutrients | 27762 | 0.01 | 0.00 | 3.23E-02 | 8.66E-02 | 100 | 27043 | 0.01 | 0.00 | 3.80E-02 | 1.15E-01 | 92 | 0.01 | 0.00 | 2.87E-03 | 0.00 | 9.92E-01 | - | - | - |
| Feel uneasy or guilty because of your way of drinking | Alcohol | 17158 | -0.03 | 0.02 | 3.55E-02 | 9.33E-02 | 102 | 16761 | -0.03 | 0.02 | 3.47E-02 | 1.09E-01 | 88 | -0.03 | 0.01 | 2.89E-03 | 0.00 | 9.90E-01 | - | - | - |
| Wine | Alcohol | 27762 | 0.01 | 0.00 | 2.28E-02 | 6.67E-02 | 92 | 27043 | 0.01 | 0.00 | 5.79E-02 | 1.57E-01 | 102 | 0.01 | 0.00 | 3.12E-03 | 0.00 | 8.37E-01 | - | - | - |
| Physical limitation to participate in moderately demanding activities: walking up several stairs | General health | 14270 | 0.01 | 0.01 | 1.01E-01 | 1.96E-01 | 138 | 14077 | 0.02 | 0.01 | 1.15E-02 | 5.01E-02 | 63 | 0.01 | 0.01 | 3.24E-03 | 0.00 | 5.10E-01 | - | - | - |
| Banana | Food | 27762 | 0.00 | 0.00 | 9.25E-01 | 9.39E-01 | 265 | 27043 | -0.02 | 0.00 | 4.21E-05 | **5.30E-04** | 22 | -0.01 | 0.00 | 3.78E-03 | 0.88 | 3.74E-03 | - | - | - |
| Energy status | Psychosocial | 24262 | -0.01 | 0.00 | 3.25E-02 | 8.66E-02 | 101 | 23817 | -0.01 | 0.01 | 5.36E-02 | 1.47E-01 | 101 | -0.01 | 0.00 | 4.00E-03 | 0.00 | 9.07E-01 | - | - | - |
| Animal based protein intake (g/day) | Nutrients | 27762 | 0.01 | 0.00 | 1.19E-02 | **4.14E-02** | 77 | 27043 | 0.01 | 0.00 | 1.40E-01 | 2.87E-01 | 133 | 0.01 | 0.00 | 4.55E-03 | 0.00 | 4.97E-01 | - | - | - |
| If you exercise, change in exercise habits during the last year | Physical activity | 16772 | -0.01 | 0.01 | 3.74E-02 | 9.51E-02 | 105 | 16333 | -0.01 | 0.01 | 5.92E-02 | 1.58E-01 | 104 | -0.01 | 0.00 | 5.02E-03 | 0.00 | 8.91E-01 | - | - | - |
| Patience status | Psychosocial | 24269 | -0.01 | 0.00 | 5.54E-03 | **2.22E-02** | 67 | 23850 | -0.01 | 0.01 | 2.42E-01 | 4.06E-01 | 165 | -0.01 | 0.00 | 5.09E-03 | 0.18 | 2.69E-01 | - | - | - |
| Worsen in health in the future | General health | 14200 | 0.01 | 0.01 | 2.06E-01 | 3.35E-01 | 165 | 14015 | 0.02 | 0.01 | 8.57E-03 | **4.02E-02** | 59 | 0.01 | 0.00 | 6.13E-03 | 0.03 | 3.10E-01 | - | - | - |
| Distance to work in kilometers (one way) | Physical activity | 26229 | -0.01 | 0.00 | 5.17E-03 | **2.11E-02** | 66 | 25530 | 0.00 | 0.00 | 3.22E-01 | 4.90E-01 | 182 | -0.01 | 0.00 | 7.10E-03 | 0.36 | 2.13E-01 | - | - | - |

| | | | | | | | | | | | | | | | | | | | | | |
|---|---|---|---|---|---|---|---|---|---|---|---|---|---|---|---|---|---|---|---|---|---|
| Milk, sour milk (3%) | Beverage | 27762 | 0.01 | 0.00 | 9.62E-02 | 1.90E-01 | 136 | 27043 | 0.01 | 0.00 | 3.17E-02 | 1.03E-01 | 85 | 0.01 | 0.00 | 7.25E-03 | 0.00 | 6.81E-01 | - | - | - |
| Vitamin D intake (ug/day) | Nutrients | 27762 | 0.01 | 0.00 | 5.24E-02 | 1.25E-01 | 112 | 27043 | 0.01 | 0.00 | 6.49E-02 | 1.66E-01 | 108 | 0.01 | 0.00 | 7.41E-03 | 0.00 | 9.94E-01 | - | - | - |
| Learn new things at job | Psychosocial | 29801 | -0.01 | 0.00 | 3.08E-03 | **1.45E-02** | 57 | 29148 | 0.00 | 0.00 | 4.70E-01 | 6.55E-01 | 199 | -0.01 | 0.00 | 8.63E-03 | 0.58 | 1.23E-01 | - | - | - |
| Participation in other association | Social | 10604 | 0.03 | 0.02 | 2.50E-02 | 7.22E-02 | 93 | 10260 | 0.02 | 0.02 | 1.42E-01 | 2.87E-01 | 136 | 0.03 | 0.01 | 8.72E-03 | 0.00 | 5.78E-01 | - | - | - |
| Potassium intake (mg/day) | Nutrients | 27762 | -0.01 | 0.00 | 1.00E-01 | 1.96E-01 | 137 | 27043 | -0.01 | 0.00 | 3.87E-02 | 1.15E-01 | 93 | -0.01 | 0.00 | 8.82E-03 | 0.00 | 7.25E-01 | - | - | - |
| For how much of the time during the last four weeks have you felt tired? | General health | 14278 | 0.01 | 0.01 | 8.59E-02 | 1.75E-01 | 132 | 14070 | 0.01 | 0.01 | 4.73E-02 | 1.34E-01 | 98 | 0.01 | 0.00 | 8.95E-03 | 0.00 | 8.15E-01 | - | - | - |
| Campesterol intake (mg/day) | Nutrients | 27762 | -0.01 | 0.00 | 6.33E-04 | **3.70E-03** | 46 | 27043 | 0.00 | 0.00 | 8.38E-01 | 9.08E-01 | 255 | -0.01 | 0.00 | 9.15E-03 | 0.80 | 2.64E-02 | - | - | - |
| Receive hugs to comfort and support you | Social | 30091 | -0.02 | 0.01 | 6.10E-02 | 1.40E-01 | 117 | 29391 | -0.02 | 0.01 | 7.13E-02 | 1.76E-01 | 112 | -0.02 | 0.01 | 9.31E-03 | 0.00 | 9.94E-01 | - | - | - |
| For how much of the time during the last four weeks has your physical health or your emotional problems limited your ability to interact with others? | General health | 14193 | 0.02 | 0.01 | 8.93E-03 | **3.29E-02** | 73 | 13993 | 0.01 | 0.01 | 3.00E-01 | 4.70E-01 | 176 | 0.01 | 0.00 | 9.32E-03 | 0.16 | 2.76E-01 | - | - | - |
| Sleep status | Sleep | 24303 | -0.01 | 0.00 | 1.64E-01 | 2.84E-01 | 155 | 23888 | -0.01 | 0.01 | 2.44E-02 | 8.44E-02 | 80 | -0.01 | 0.00 | 1.02E-02 | 0.00 | 5.25E-01 | - | - | - |
| Frequency of engaging in clubs, associations or study circles | Social | 19008 | 0.00 | 0.01 | 4.08E-01 | 5.45E-01 | 201 | 18522 | -0.01 | 0.01 | 5.49E-03 | **2.94E-02** | 51 | -0.01 | 0.00 | 1.05E-02 | 0.46 | 1.73E-01 | - | - | - |
| Steak, chop, e.g. | Food | 27762 | 0.01 | 0.00 | 1.10E-02 | **3.94E-02** | 75 | 27043 | 0.00 | 0.00 | 3.40E-01 | 5.11E-01 | 184 | 0.01 | 0.00 | 1.25E-02 | 0.13 | 2.85E-01 | - | - | - |
| Cohabitation: Live alone vs Only one adult (spouse, partner) | Social | 14042 | -0.04 | 0.02 | 2.05E-02 | 6.19E-02 | 89 | 13853 | -0.02 | 0.02 | 2.40E-01 | 4.05E-01 | 164 | -0.03 | 0.01 | 1.38E-02 | 0.00 | 4.03E-01 | - | - | - |
| Repetitive job | Psychosocial | 29814 | 0.01 | 0.00 | 6.21E-02 | 1.41E-01 | 118 | 29170 | 0.01 | 0.00 | 1.07E-01 | 2.43E-01 | 122 | 0.01 | 0.00 | 1.39E-02 | 0.00 | 8.78E-01 | - | - | - |
| Formic acid intaje (g/day) | Nutrients | 27762 | 0.01 | 0.00 | 1.45E-01 | 2.56E-01 | 152 | 27043 | 0.01 | 0.00 | 4.26E-02 | 1.22E-01 | 97 | 0.01 | 0.00 | 1.40E-02 | 0.00 | 6.53E-01 | - | - | - |
| Physical limitation to participate in steneous activities: running, lifting heavy objects, taking part in physically demanding sports | General health | 14269 | 0.01 | 0.01 | 3.75E-02 | 9.51E-02 | 106 | 14096 | 0.01 | 0.01 | 1.67E-01 | 3.14E-01 | 147 | 0.01 | 0.01 | 1.41E-02 | 0.00 | 6.38E-01 | - | - | - |
| Would you say that the number of people that you meet in your everyday life is enough or would you like to meet more or fewer people? | Social | 30313 | 0.00 | 0.00 | 3.10E-01 | 4.41E-01 | 189 | 29611 | -0.01 | 0.00 | 1.49E-02 | 6.05E-02 | 68 | -0.01 | 0.00 | 1.53E-02 | 0.07 | 2.98E-01 | - | - | - |
| Vitamin B12 intake (ug/day) | Nutrients | 27762 | 0.01 | 0.00 | 3.62E-02 | 9.36E-02 | 104 | 27043 | 0.01 | 0.00 | 1.88E-01 | 3.45E-01 | 151 | 0.01 | 0.00 | 1.54E-02 | 0.00 | 6.16E-01 | - | - | - |
| Cookies, pastry | Food | 27762 | -0.01 | 0.00 | 8.40E-02 | 1.75E-01 | 128 | 27043 | -0.01 | 0.00 | 9.75E-02 | 2.25E-01 | 120 | -0.01 | 0.00 | 1.67E-02 | 0.00 | 9.94E-01 | - | - | - |
| For how much of the time during the last four weeks have you felt so depressed that nothing could cheer you up? | General health | 14274 | 0.01 | 0.01 | 3.17E-02 | 8.62E-02 | 99 | 14081 | 0.01 | 0.01 | 2.27E-01 | 3.94E-01 | 159 | 0.01 | 0.00 | 1.71E-02 | 0.00 | 5.30E-01 | - | - | - |
| Years using snuff | Tobacco use | 28561 | -0.02 | 0.00 | 5.04E-04 | **3.08E-03** | 44 | 27938 | 0.00 | 0.01 | 8.38E-01 | 9.08E-01 | 254 | -0.01 | 0.00 | 1.81E-02 | 0.85 | 1.04E-02 | - | - | - |
| Frequency of walking during leisure time | Physical activity | 29243 | -0.01 | 0.00 | 1.07E-01 | 2.03E-01 | 141 | 28570 | -0.01 | 0.00 | 8.51E-02 | 2.05E-01 | 114 | -0.01 | 0.00 | 1.84E-02 | 0.00 | 9.16E-01 | - | - | - |
| Chips, popcorn, salted nuts | Food | 27762 | 0.01 | 0.00 | 1.66E-02 | 5.31E-02 | 84 | 27043 | 0.00 | 0.00 | 3.71E-01 | 5.51E-01 | 185 | 0.01 | 0.00 | 1.90E-02 | 0.04 | 3.08E-01 | - | - | - |
| Physical limitation to participate in moderately demanding activities: walking more than 2 km | General health | 14276 | 0.01 | 0.01 | 8.11E-02 | 1.73E-01 | 126 | 14084 | 0.01 | 0.01 | 1.17E-01 | 2.58E-01 | 125 | 0.01 | 0.00 | 1.92E-02 | 0.00 | 9.13E-01 | - | - | - |
| Margarine on bread | Food | 27762 | 0.01 | 0.00 | 1.74E-01 | 2.97E-01 | 157 | 27043 | 0.01 | 0.00 | 5.08E-02 | 1.41E-01 | 100 | 0.01 | 0.00 | 1.95E-02 | 0.00 | 6.49E-01 | - | - | - |
| Support from others | Social | 30333 | -0.01 | 0.00 | 2.18E-01 | 3.43E-01 | 171 | 29639 | -0.01 | 0.00 | 3.95E-02 | 1.17E-01 | 94 | -0.01 | 0.00 | 2.04E-02 | 0.00 | 5.38E-01 | - | - | - |
| As healthy as anyone | General health | 14205 | -0.01 | 0.01 | 6.22E-02 | 1.41E-01 | 119 | 14018 | -0.01 | 0.01 | 1.64E-01 | 3.12E-01 | 146 | -0.01 | 0.00 | 2.11E-02 | 0.00 | 7.59E-01 | - | - | - |
| Monounsaturated fat intake (g/day) | Nutrients | 27762 | 0.01 | 0.00 | 8.35E-02 | 1.75E-01 | 127 | 27043 | 0.01 | 0.00 | 1.31E-01 | 2.80E-01 | 130 | 0.01 | 0.00 | 2.18E-02 | 0.00 | 9.18E-01 | - | - | - |
| Sausage, liver pate on bread | Food | 27762 | 0.01 | 0.00 | 7.31E-02 | 1.60E-01 | 123 | 27043 | 0.01 | 0.00 | 1.50E-01 | 2.98E-01 | 139 | 0.01 | 0.00 | 2.21E-02 | 0.00 | 8.37E-01 | - | - | - |
| Long-term sickness | General health | 29463 | 0.03 | 0.01 | 3.82E-03 | **1.66E-02** | 62 | 28755 | 0.00 | 0.01 | 7.69E-01 | 8.73E-01 | 244 | 0.02 | 0.01 | 2.23E-02 | 0.69 | 7.21E-02 | - | - | - |
| Physical limitation to participate in moderately demanding activities: bending down or kneeling | General health | 14254 | 0.01 | 0.01 | 4.77E-02 | 1.16E-01 | 110 | 14064 | 0.01 | 0.01 | 2.18E-01 | 3.89E-01 | 155 | 0.01 | 0.01 | 2.26E-02 | 0.00 | 6.16E-01 | - | - | - |
| Phosphate intake (mg/day) | Nutrients | 27762 | -0.01 | 0.00 | 7.95E-02 | 1.71E-01 | 125 | 27043 | -0.01 | 0.00 | 1.58E-01 | 3.02E-01 | 145 | -0.01 | 0.00 | 2.49E-02 | 0.00 | 8.47E-01 | - | - | - |
| Close relationship with anyone | Social | 30290 | -0.01 | 0.00 | 9.40E-02 | 1.87E-01 | 135 | 29589 | -0.01 | 0.00 | 1.53E-01 | 2.98E-01 | 142 | -0.01 | 0.00 | 2.80E-02 | 0.00 | 8.87E-01 | - | - | - |
| Mood status | Psychosocial | 24269 | -0.01 | 0.00 | 1.59E-02 | 5.20E-02 | 82 | 23844 | 0.00 | 0.01 | 5.01E-01 | 6.81E-01 | 204 | -0.01 | 0.00 | 2.81E-02 | 0.31 | 2.28E-01 | - | - | - |
| Ice cream | Food | 27762 | 0.00 | 0.00 | 5.78E-01 | 6.94E-01 | 224 | 27043 | -0.01 | 0.00 | 1.13E-02 | 5.01E-02 | 62 | -0.01 | 0.00 | 3.06E-02 | 0.51 | 1.52E-01 | - | - | - |
| Tomato, cucumber | Food | 27762 | -0.01 | 0.00 | 1.38E-01 | 2.51E-01 | 148 | 27043 | -0.01 | 0.00 | 1.54E-01 | 2.99E-01 | 143 | -0.01 | 0.00 | 3.97E-02 | 0.00 | 9.96E-01 | - | - | - |
| For how much of the time during the last four weeks have you felt calm and serene? | General health | 14258 | -0.01 | 0.01 | 3.09E-02 | 8.47E-02 | 98 | 14075 | 0.00 | 0.01 | 4.94E-01 | 6.74E-01 | 203 | -0.01 | 0.00 | 4.27E-02 | 0.04 | 3.07E-01 | - | - | - |
| Minced meat dishes | Food | 27762 | 0.01 | 0.00 | 6.96E-02 | 1.53E-01 | 122 | 27043 | 0.00 | 0.00 | 3.01E-01 | 4.70E-01 | 177 | 0.01 | 0.00 | 4.30E-02 | 0.00 | 6.02E-01 | - | - | - |
| Berries (fresh or frozen) | Food | 27762 | 0.00 | 0.00 | 4.04E-01 | 5.43E-01 | 200 | 27043 | -0.01 | 0.00 | 4.06E-02 | 1.18E-01 | 95 | -0.01 | 0.00 | 4.32E-02 | 0.00 | 3.70E-01 | - | - | - |
| Sour milk, yoghurt (3% fat) | Food | 27762 | -0.01 | 0.00 | 9.13E-02 | 1.83E-01 | 134 | 27043 | -0.01 | 0.00 | 2.55E-01 | 4.24E-01 | 166 | -0.01 | 0.00 | 4.49E-02 | 0.00 | 7.27E-01 | - | - | - |
| For how much of the time during the last four weeks have you felt really alert and strong? | General health | 14238 | -0.01 | 0.01 | 1.40E-01 | 2.52E-01 | 149 | 14041 | -0.01 | 0.01 | 1.76E-01 | 3.29E-01 | 148 | -0.01 | 0.00 | 4.52E-02 | 0.00 | 9.52E-01 | - | - | - |
| Salad dressing with oil | Food | 27762 | 0.00 | 0.00 | 3.31E-01 | 4.66E-01 | 191 | 27043 | -0.01 | 0.00 | 6.30E-02 | 1.63E-01 | 106 | -0.01 | 0.00 | 4.63E-02 | 0.00 | 5.10E-01 | - | - | - |
| Participation in sports or physical exercise associations | Social | 10604 | -0.02 | 0.02 | 1.33E-01 | 2.43E-01 | 147 | 10260 | -0.02 | 0.02 | 1.92E-01 | 3.49E-01 | 152 | -0.02 | 0.01 | 4.70E-02 | 0.00 | 8.84E-01 | - | - | - |
| People to ask for help apart from the ones at home | Social | 30231 | -0.01 | 0.01 | 6.41E-01 | 7.47E-01 | 231 | 29538 | -0.03 | 0.01 | 1.89E-02 | 7.08E-02 | 74 | -0.02 | 0.01 | 4.82E-02 | 0.45 | 1.77E-01 | - | - | - |
| Low fat milk (0,5%) | Beverage | 27762 | 0.00 | 0.00 | 2.78E-01 | 4.15E-01 | 180 | 27043 | 0.01 | 0.00 | 8.60E-02 | 2.05E-01 | 116 | 0.01 | 0.00 | 4.85E-02 | 0.00 | 6.28E-01 | - | - | - |
| For how much of the time during the last four weeks have you felt full of energy? | General health | 14240 | -0.02 | 0.01 | 2.59E-02 | 7.32E-02 | 95 | 14053 | 0.00 | 0.01 | 6.03E-01 | 7.49E-01 | 223 | -0.01 | 0.00 | 4.97E-02 | 0.29 | 2.35E-01 | - | - | - |
| Arachidonic acid (ARA) intake (g/day) | Nutrients | 27761 | 0.01 | 0.00 | 1.63E-02 | 5.28E-02 | 83 | 27042 | 0.00 | 0.00 | 7.51E-01 | 8.59E-01 | 242 | 0.01 | 0.00 | 5.07E-02 | 0.51 | 1.52E-01 | - | - | - |
| Bregott on bread | Food | 27762 | 0.00 | 0.00 | 4.86E-01 | 6.14E-01 | 213 | 27043 | 0.01 | 0.00 | 3.72E-02 | 1.14E-01 | 90 | 0.01 | 0.00 | 5.15E-02 | 0.04 | 3.08E-01 | - | - | - |
| Get sick more often than other people | General health | 14250 | 0.00 | 0.01 | 4.94E-01 | 6.18E-01 | 215 | 14057 | 0.01 | 0.01 | 3.70E-02 | 1.14E-01 | 89 | 0.01 | 0.00 | 5.19E-02 | 0.05 | 3.04E-01 | - | - | - |
| Vitamin C intake (mg/day) | Nutrients | 27762 | 0.00 | 0.00 | 2.60E-01 | 3.95E-01 | 177 | 27043 | -0.01 | 0.00 | 1.23E-01 | 2.68E-01 | 127 | -0.01 | 0.00 | 5.99E-02 | 0.00 | 7.39E-01 | - | - | - |
| Frequency of shoverling snow during leisure time | Physical activity | 18197 | -0.01 | 0.01 | 2.15E-01 | 3.42E-01 | 169 | 17676 | -0.01 | 0.01 | 1.58E-01 | 3.02E-01 | 144 | -0.01 | 0.00 | 6.08E-02 | 0.00 | 8.90E-01 | - | - | - |
| Butter on bread | Food | 27762 | 0.01 | 0.00 | 1.00E-02 | **3.65E-02** | 74 | 27043 | 0.00 | 0.00 | 9.56E-01 | 9.70E-01 | 273 | 0.01 | 0.00 | 6.90E-02 | 0.70 | 6.82E-02 | - | - | - |
| Travel to work: Walk to work vs passive travel to work | Physical activity | 19526 | 0.00 | 0.02 | 7.84E-01 | 8.47E-01 | 249 | 19250 | -0.04 | 0.02 | 2.42E-02 | 8.44E-02 | 78 | -0.02 | 0.01 | 7.43E-02 | 0.49 | 1.60E-01 | - | - | - |
| Iron intake (mg/day) | Nutrients | 27762 | 0.00 | 0.00 | 8.81E-01 | 9.15E-01 | 258 | 27043 | -0.01 | 0.00 | 1.57E-02 | 6.32E-02 | 69 | -0.01 | 0.00 | 7.52E-02 | 0.63 | 1.01E-01 | - | - | - |
| Sour milk, yoghurt (low fat) | Food | 27762 | -0.01 | 0.00 | 2.03E-01 | 3.34E-01 | 163 | 27043 | -0.01 | 0.00 | 2.15E-01 | 3.87E-01 | 154 | -0.01 | 0.00 | 7.56E-02 | 0.00 | 9.90E-01 | - | - | - |
| Vitamin B2 intake (ug/day) | Nutrients | 27762 | -0.01 | 0.00 | 1.65E-01 | 2.84E-01 | 156 | 27043 | 0.00 | 0.00 | 2.78E-01 | 4.42E-01 | 174 | -0.01 | 0.00 | 7.94E-02 | 0.00 | 8.61E-01 | - | - | - |
| Possibility to speak with colleagues during breaks | Psychosocial | 29740 | -0.01 | 0.00 | 2.12E-02 | 6.32E-02 | 90 | 29114 | 0.00 | 0.00 | 9.21E-01 | 9.56E-01 | 267 | -0.01 | 0.00 | 8.49E-02 | 0.58 | 1.25E-01 | - | - | - |
| Vitamin E intake (mg/day) | Nutrients | 27762 | -0.01 | 0.00 | 1.01E-01 | 1.96E-01 | 139 | 27043 | 0.00 | 0.00 | 4.37E-01 | 6.21E-01 | 195 | -0.01 | 0.00 | 8.56E-02 | 0.00 | 5.63E-01 | - | - | - |
| Corn flakes | Food | 27762 | 0.00 | 0.00 | 5.78E-01 | 6.94E-01 | 223 | 27043 | -0.01 | 0.00 | 6.22E-02 | 1.63E-01 | 105 | -0.01 | 0.00 | 9.05E-02 | 0.00 | 3.37E-01 | - | - | - |
| Job demands to work very fast | Psychosocial | 29859 | 0.00 | 0.00 | 8.72E-01 | 9.12E-01 | 257 | 29226 | 0.01 | 0.00 | 2.50E-02 | 8.56E-02 | 81 | 0.01 | 0.00 | 9.16E-02 | 0.55 | 1.38E-01 | - | - | - |
| Linoleic acid intake (g/day) | Nutrients | 27762 | -0.01 | 0.00 | 3.89E-02 | 9.77E-02 | 107 | 27043 | 0.00 | 0.00 | 7.86E-01 | 8.85E-01 | 246 | -0.01 | 0.00 | 9.31E-02 | 0.34 | 2.18E-01 | - | - | - |
| Strong beer | Alcohol | 27762 | 0.01 | 0.00 | 1.03E-01 | 1.99E-01 | 140 | 27043 | 0.00 | 0.00 | 4.77E-01 | 6.60E-01 | 200 | 0.01 | 0.00 | 9.53E-02 | 0.00 | 5.40E-01 | - | - | - |
| Sugar, honey, marmelade, jam | Food | 27762 | 0.00 | 0.00 | 6.66E-01 | 7.61E-01 | 235 | 27043 | 0.01 | 0.00 | 5.52E-03 | **2.94E-02** | 52 | 0.01 | 0.00 | 1.07E-01 | 0.81 | 2.15E-02 | - | - | - |
| Sucrose intake (g/day) | Nutrients | 27762 | 0.00 | 0.00 | 4.49E-01 | 5.82E-01 | 206 | 27043 | 0.01 | 0.00 | 1.28E-01 | 2.75E-01 | 129 | 0.01 | 0.00 | 1.10E-01 | 0.00 | 5.64E-01 | - | - | - |
| Enterolactone intake (ug/day) | Nutrients | 27762 | 0.00 | 0.00 | 5.42E-01 | 6.66E-01 | 218 | 27043 | 0.01 | 0.00 | 9.48E-02 | 2.22E-01 | 118 | 0.00 | 0.00 | 1.10E-01 | 0.00 | 4.33E-01 | - | - | - |
| Heptadecanoic acid intake (g/day) | Nutrients | 27762 | 0.00 | 0.00 | 2.85E-01 | 4.15E-01 | 184 | 27043 | 0.01 | 0.00 | 2.39E-01 | 4.05E-01 | 161 | 0.01 | 0.00 | 1.13E-01 | 0.00 | 9.15E-01 | - | - | - |
| Pentadecanoic acid intake (g/day) | Nutrients | 27762 | 0.00 | 0.00 | 2.85E-01 | 4.15E-01 | 185 | 27043 | 0.01 | 0.00 | 2.39E-01 | 4.05E-01 | 162 | 0.01 | 0.00 | 1.13E-01 | 0.00 | 9.15E-01 | - | - | - |
| Snuff status: Former snuff users vs non-snuff users | Tobacco use | 24204 | -0.03 | 0.01 | 3.57E-02 | 9.33E-02 | 103 | 23610 | 0.00 | 0.01 | 8.98E-01 | 9.51E-01 | 261 | -0.02 | 0.01 | 1.13E-01 | 0.48 | 1.66E-01 | - | - | - |
| Boiled or baked potato | Food | 27762 | 0.00 | 0.00 | 6.72E-01 | 7.61E-01 | 237 | 27043 | 0.01 | 0.00 | 6.79E-02 | 1.71E-01 | 110 | 0.01 | 0.00 | 1.16E-01 | 0.05 | 3.06E-01 | - | - | - |
| Number of snuff boxes per week | Tobacco use | 29333 | 0.00 | 0.00 | 6.79E-01 | 7.64E-01 | 239 | 28689 | 0.01 | 0.00 | 7.81E-03 | **3.79E-02** | 57 | 0.01 | 0.00 | 1.23E-01 | 0.79 | 2.73E-02 | - | - | - |
| Bacon | Food | 27762 | 0.01 | 0.00 | 1.38E-03 | **7.14E-03** | 52 | 27043 | 0.00 | 0.00 | 2.61E-01 | 4.24E-01 | 169 | 0.00 | 0.00 | 1.26E-01 | 0.89 | 2.48E-03 | - | - | - |
| Pizza | Food | 27762 | 0.00 | 0.00 | 8.50E-01 | 8.94E-01 | 256 | 27043 | 0.01 | 0.00 | 2.21E-02 | 7.97E-02 | 77 | 0.00 | 0.00 | 1.46E-01 | 0.68 | 7.53E-02 | - | - | - |
| Marital status: Single vs Married/partner | Social | 27721 | -0.03 | 0.02 | 3.08E-02 | 8.47E-02 | 97 | 27040 | 0.00 | 0.02 | 9.07E-01 | 9.51E-01 | 264 | -0.02 | 0.01 | 1.47E-01 | 0.61 | 1.09E-01 | - | - | - |
| Oil for cooking | Food | 27762 | -0.01 | 0.00 | 5.91E-02 | 1.37E-01 | 116 | 27043 | 0.00 | 0.00 | 9.04E-01 | 9.51E-01 | 262 | 0.00 | 0.00 | 1.49E-01 | 0.33 | 2.22E-01 | - | - | - |
| Sausage as main dish | Food | 27762 | 0.01 | 0.00 | 1.93E-02 | 5.89E-02 | 88 | 27043 | 0.00 | 0.00 | 7.06E-01 | 8.31E-01 | 234 | 0.00 | 0.00 | 1.56E-01 | 0.72 | 5.76E-02 | - | - | - |
| Skill demand from job | Psychosocial | 29791 | -0.01 | 0.00 | 4.79E-02 | 1.16E-01 | 111 | 29116 | 0.00 | 0.00 | 9.79E-01 | 9.85E-01 | 275 | 0.00 | 0.00 | 1.61E-01 | 0.49 | 1.63E-01 | - | - | - |
| Do you feel important and appreciated in your home? | Psychosocial | 24127 | 0.00 | 0.00 | 5.92E-01 | 6.99E-01 | 228 | 23692 | -0.01 | 0.01 | 1.47E-01 | 2.96E-01 | 138 | 0.00 | 0.00 | 1.63E-01 | 0.00 | 5.08E-01 | - | - | - |
| Travel to work: Cycle to work vs passive travel to work | Physical activity | 22879 | -0.01 | 0.01 | 5.80E-01 | 6.94E-01 | 225 | 22233 | -0.02 | 0.01 | 1.51E-01 | 2.98E-01 | 140 | -0.01 | 0.01 | 1.64E-01 | 0.00 | 5.11E-01 | - | - | - |
| Salty fish | Food | 27762 | 0.01 | 0.00 | 1.53E-02 | 5.16E-02 | 80 | 27043 | 0.00 | 0.00 | 5.60E-01 | 7.19E-01 | 216 | 0.00 | 0.00 | 1.67E-01 | 0.77 | 3.80E-02 | - | - | - |
| Self rate of overall health compared to a year ago | General health | 14192 | -0.01 | 0.01 | 3.13E-01 | 4.43E-01 | 190 | 13991 | -0.01 | 0.01 | 3.74E-01 | 5.51E-01 | 187 | -0.01 | 0.00 | 1.79E-01 | 0.00 | 9.41E-01 | - | - | - |
| Trans fat intake (g/day) | Nutrients | 27762 | 0.00 | 0.01 | 8.05E-01 | 8.66E-01 | 250 | 27043 | 0.01 | 0.01 | 9.35E-02 | 2.21E-01 | 117 | 0.01 | 0.00 | 1.79E-01 | 0.07 | 3.01E-01 | - | - | - |
| Docosahexaenoic acid (DHA) intake (g/day) | Nutrients | 27762 | 0.01 | 0.00 | 4.79E-03 | **1.98E-02** | 65 | 27043 | 0.00 | 0.00 | 3.05E-01 | 4.74E-01 | 178 | 0.00 | 0.00 | 1.82E-01 | 0.86 | 7.17E-03 | - | - | - |
| Enough time for job assignments | Psychosocial | 29729 | -0.01 | 0.00 | 2.22E-01 | 3.47E-01 | 172 | 29090 | 0.00 | 0.00 | 5.12E-01 | 6.91E-01 | 205 | 0.00 | 0.00 | 1.82E-01 | 0.00 | 7.03E-01 | - | - | - |
| Hearing status | General health | 24304 | 0.00 | 0.01 | 8.20E-01 | 8.72E-01 | 253 | 23877 | -0.01 | 0.01 | 1.01E-01 | 2.32E-01 | 121 | 0.00 | 0.00 | 1.90E-01 | 0.02 | 3.13E-01 | - | - | - |
| Changed everyday exercise during the last year | Physical activity | 19041 | -0.01 | 0.00 | 1.94E-01 | 3.25E-01 | 161 | 18508 | 0.00 | 0.00 | 5.82E-01 | 7.32E-01 | 220 | 0.00 | 0.00 | 1.90E-01 | 0.00 | 6.02E-01 | - | - | - |
| Control over own work assignment | Psychosocial | 29866 | 0.00 | 0.00 | 8.95E-01 | 9.20E-01 | 261 | 29219 | 0.01 | 0.00 | 8.54E-02 | 2.05E-01 | 115 | 0.00 | 0.00 | 1.95E-01 | 0.23 | 2.55E-01 | - | - | - |
| Sweets | Food | 27762 | 0.00 | 0.00 | 5.69E-01 | 6.94E-01 | 220 | 27043 | 0.01 | 0.00 | 2.14E-01 | 3.87E-01 | 153 | 0.00 | 0.00 | 2.03E-01 | 0.00 | 6.19E-01 | - | - | - |
| Do you feel important and appreciated outside your home? | Psychosocial | 24295 | -0.01 | 0.00 | 3.02E-01 | 4.34E-01 | 187 | 23881 | 0.00 | 0.01 | 4.83E-01 | 6.66E-01 | 201 | 0.00 | 0.00 | 2.19E-01 | 0.00 | 8.25E-01 | - | - | - |
| Cohabitation: Live alone vs Other/others | Social | 3468 | -0.05 | 0.04 | 2.04E-01 | 3.34E-01 | 164 | 3446 | -0.02 | 0.05 | 7.08E-01 | 8.31E-01 | 236 | -0.04 | 0.03 | 2.22E-01 | 0.00 | 6.04E-01 | - | - | - |
| Vitamin B6 intake (mg/day) | Nutrients | 27762 | 0.00 | 0.00 | 6.74E-01 | 7.61E-01 | 238 | 27043 | -0.01 | 0.00 | 3.77E-02 | 1.15E-01 | 91 | 0.00 | 0.00 | 2.57E-01 | 0.69 | 7.32E-02 | - | - | - |
| Frequency of cycling during leisure time | Physical activity | 24860 | 0.00 | 0.00 | 3.96E-01 | 5.41E-01 | 197 | 24146 | 0.00 | 0.00 | 4.69E-01 | 6.55E-01 | 198 | 0.00 | 0.00 | 2.66E-01 | 0.00 | 9.43E-01 | - | - | - |
| Calcium intake (mg/day) | Nutrients | 27762 | -0.01 | 0.00 | 1.22E-01 | 2.28E-01 | 144 | 27043 | 0.00 | 0.00 | 9.61E-01 | 9.71E-01 | 274 | 0.00 | 0.00 | 2.78E-01 | 0.18 | 2.70E-01 | - | - | - |
| Light beer | Alcohol | 27762 | 0.00 | 0.00 | 4.71E-01 | 6.01E-01 | 211 | 27043 | 0.00 | 0.00 | 4.29E-01 | 6.13E-01 | 194 | 0.00 | 0.00 | 2.86E-01 | 0.00 | 9.40E-01 | - | - | - |
| Lean fish (e.g. perch, bass, cod) | Food | 27762 | 0.00 | 0.00 | 6.84E-01 | 7.66E-01 | 240 | 27043 | 0.01 | 0.00 | 2.62E-01 | 4.24E-01 | 171 | 0.00 | 0.00 | 2.87E-01 | 0.00 | 5.91E-01 | - | - | - |
| Smoking status: Former occasional smokers vs non-smokers | Tobacco use | 18070 | 0.01 | 0.01 | 6.32E-01 | 7.39E-01 | 230 | 17710 | 0.02 | 0.01 | 3.07E-01 | 4.74E-01 | 179 | 0.01 | 0.01 | 2.88E-01 | 0.00 | 7.04E-01 | - | - | - |

| | | | | | | | | | | | | | | | | | | | | | |
|---|---|---|---|---|---|---|---|---|---|---|---|---|---|---|---|---|---|---|---|---|---|
| Number of social interactions during a normal week | Social | 30329 | -0.01 | 0.00 | 8.52E-02 | 1.75E-01 | 131 | 29625 | 0.00 | 0.00 | 8.02E-01 | 8.96E-01 | 247 | 0.00 | 0.00 | 2.89E-01 | 0.47 | 1.68E-01 | - | - | - |
| Contradictory demands in job | Psychosocial | 29686 | 0.00 | 0.00 | 4.02E-01 | 5.43E-01 | 199 | 29006 | 0.00 | 0.00 | 5.30E-01 | 6.99E-01 | 209 | 0.00 | 0.00 | 2.99E-01 | 0.00 | 8.93E-01 | - | - | - |
| Polyunsaturated fat intake (g/day) | Nutrients | 27762 | -0.01 | 0.00 | 1.21E-01 | 2.28E-01 | 142 | 27043 | 0.00 | 0.00 | 8.98E-01 | 9.51E-01 | 260 | 0.00 | 0.00 | 3.02E-01 | 0.27 | 2.43E-01 | - | - | - |
| Meat on bread | Food | 27762 | -0.01 | 0.00 | 2.92E-02 | 8.18E-02 | 96 | 27043 | 0.00 | 0.00 | 4.14E-01 | 6.00E-01 | 191 | 0.00 | 0.00 | 3.11E-01 | 0.77 | 3.60E-02 | - | - | - |
| Eicosapentaenoic acid (EPA) intake (g/day) | Nutrients | 27761 | 0.01 | 0.00 | 7.41E-03 | **2.85E-02** | 70 | 27043 | -0.01 | 0.00 | 1.80E-01 | 3.34E-01 | 149 | 0.00 | 0.00 | 3.13E-01 | 0.87 | 4.79E-03 | - | - | - |
| Pasta | Food | 27762 | -0.01 | 0.00 | 2.18E-01 | 3.43E-01 | 170 | 27043 | 0.00 | 0.00 | 8.69E-01 | 9.33E-01 | 258 | 0.00 | 0.00 | 3.16E-01 | 0.00 | 4.63E-01 | - | - | - |
| Frequency of dancing during leisure time | Physical activity | 17005 | 0.00 | 0.01 | 3.56E-01 | 4.98E-01 | 192 | 16467 | 0.00 | 0.01 | 6.24E-01 | 7.64E-01 | 226 | 0.00 | 0.00 | 3.16E-01 | 0.00 | 7.66E-01 | - | - | - |
| Marital status: Single vs Divorced/separated | Social | 5018 | -0.03 | 0.02 | 1.56E-01 | 2.72E-01 | 154 | 4982 | 0.00 | 0.03 | 9.17E-01 | 9.55E-01 | 266 | -0.02 | 0.02 | 3.19E-01 | 0.03 | 3.10E-01 | - | - | - |
| Whole grain crisp bread | Food | 27762 | 0.00 | 0.00 | 4.19E-01 | 5.58E-01 | 202 | 27043 | 0.00 | 0.00 | 5.58E-01 | 7.19E-01 | 215 | 0.00 | 0.00 | 3.23E-01 | 0.00 | 8.91E-01 | - | - | - |
| Physical limitation that limited your ability to perform certain work tasks or other activities during the last four weeks | General health | 14252 | 0.02 | 0.02 | 3.99E-01 | 5.41E-01 | 198 | 14042 | 0.01 | 0.02 | 5.99E-01 | 7.47E-01 | 222 | 0.01 | 0.01 | 3.32E-01 | 0.00 | 8.27E-01 | - | - | - |
| Fatty fish (e.g. herring, whitefish, salmon) | Food | 27762 | 0.01 | 0.00 | 2.17E-02 | 6.42E-02 | 91 | 27043 | 0.00 | 0.00 | 3.26E-01 | 4.93E-01 | 183 | 0.00 | 0.00 | 3.32E-01 | 0.81 | 2.14E-02 | - | - | - |
| Brown beans, pea soup | Food | 27762 | 0.00 | 0.00 | 8.88E-01 | 9.19E-01 | 260 | 27043 | 0.01 | 0.00 | 2.21E-01 | 3.90E-01 | 157 | 0.00 | 0.00 | 3.44E-01 | 0.00 | 4.30E-01 | - | - | - |
| Cream, creme fraiche, sour cream | Food | 27762 | 0.00 | 0.00 | 2.80E-01 | 4.15E-01 | 181 | 27043 | 0.00 | 0.00 | 8.16E-01 | 9.04E-01 | 250 | 0.00 | 0.00 | 3.48E-01 | 0.00 | 5.60E-01 | - | - | - |
| Control over planning and execution of the workday | Psychosocial | 29888 | -0.01 | 0.00 | 6.94E-02 | 1.53E-01 | 121 | 29220 | 0.00 | 0.00 | 5.97E-01 | 7.47E-01 | 221 | 0.00 | 0.00 | 3.53E-01 | 0.63 | 9.94E-02 | - | - | - |
| High mental demand from job | Psychosocial | 29618 | 0.00 | 0.00 | 3.88E-01 | 5.34E-01 | 195 | 29008 | -0.01 | 0.00 | 3.04E-02 | 1.00E-01 | 84 | 0.00 | 0.00 | 3.68E-01 | 0.78 | 3.15E-02 | - | - | - |
| Frequency of gardening during leisure time | Physical activity | 18123 | 0.00 | 0.00 | 5.05E-01 | 6.29E-01 | 216 | 17532 | 0.00 | 0.01 | 5.55E-01 | 7.18E-01 | 214 | 0.00 | 0.00 | 3.74E-01 | 0.00 | 9.63E-01 | - | - | - |
| Butter for cooking | Food | 27762 | 0.00 | 0.00 | 4.49E-01 | 5.82E-01 | 205 | 27043 | 0.00 | 0.00 | 6.20E-01 | 7.64E-01 | 224 | 0.00 | 0.00 | 3.74E-01 | 0.00 | 8.64E-01 | - | - | - |
| Average portion size of meat/fish based on photographic illustration of four sizes (smallest to largest) | Food | 27762 | 0.01 | 0.00 | 2.81E-01 | 4.15E-01 | 182 | 27043 | 0.00 | 0.00 | 9.39E-01 | 9.60E-01 | 271 | 0.00 | 0.00 | 4.05E-01 | 0.00 | 4.91E-01 | - | - | - |
| White meat (poultry) | Food | 27762 | 0.01 | 0.00 | 2.27E-01 | 3.53E-01 | 173 | 27043 | 0.00 | 0.00 | 9.45E-01 | 9.63E-01 | 272 | 0.00 | 0.00 | 4.11E-01 | 0.00 | 3.74E-01 | - | - | - |
| Margarine for cooking | Food | 27762 | 0.00 | 0.00 | 9.59E-01 | 9.63E-01 | 268 | 27043 | 0.01 | 0.00 | 2.64E-01 | 4.26E-01 | 172 | 0.00 | 0.00 | 4.18E-01 | 0.00 | 4.42E-01 | - | - | - |
| Physical limitation to participate in moderately demanding activities: lifting or carrying grocery bags | General health | 14254 | 0.00 | 0.01 | 6.49E-01 | 7.51E-01 | 232 | 14084 | 0.00 | 0.01 | 5.18E-01 | 6.93E-01 | 207 | 0.00 | 0.01 | 4.36E-01 | 0.00 | 8.88E-01 | - | - | - |
| For how much of the time during the last four weeks have you felt gloomy and sad? | General health | 14244 | 0.01 | 0.01 | 2.73E-01 | 4.11E-01 | 178 | 14056 | 0.00 | 0.01 | 9.81E-01 | 9.85E-01 | 276 | 0.00 | 0.00 | 4.39E-01 | 0.00 | 4.37E-01 | - | - | - |
| Light and physically active work | Physical activity | 29420 | 0.00 | 0.01 | 9.04E-01 | 9.22E-01 | 263 | 28753 | 0.01 | 0.01 | 2.19E-01 | 3.89E-01 | 156 | 0.01 | 0.01 | 4.43E-01 | 0.00 | 3.33E-01 | - | - | - |
| Satisfaction with economy | Psychosocial | 24286 | 0.00 | 0.01 | 8.12E-01 | 8.70E-01 | 251 | 23856 | 0.00 | 0.01 | 4.02E-01 | 5.86E-01 | 189 | 0.00 | 0.00 | 4.49E-01 | 0.00 | 6.65E-01 | - | - | - |
| Self-employed | Psychosocial | 25839 | 0.01 | 0.02 | 4.66E-01 | 5.97E-01 | 210 | 25115 | 0.01 | 0.02 | 7.46E-01 | 8.57E-01 | 241 | 0.01 | 0.01 | 4.56E-01 | 0.00 | 7.77E-01 | - | - | - |
| Satisfaction with home and family situation | Psychosocial | 24285 | 0.00 | 0.00 | 7.73E-01 | 8.38E-01 | 248 | 23868 | 0.00 | 0.01 | 4.52E-01 | 6.38E-01 | 196 | 0.00 | 0.00 | 4.64E-01 | 0.00 | 7.35E-01 | - | - | - |
| Work shifts/weekends | Psychosocial | 29464 | 0.01 | 0.01 | 5.88E-01 | 6.97E-01 | 227 | 28871 | -0.02 | 0.01 | 1.11E-01 | 2.48E-01 | 124 | -0.01 | 0.01 | 4.66E-01 | 0.57 | 1.29E-01 | - | - | - |
| Possibility to leave your work for a while to speak with a colleague | Psychosocial | 29555 | -0.01 | 0.00 | 1.25E-01 | 2.32E-01 | 145 | 28932 | 0.00 | 0.00 | 5.78E-01 | 7.32E-01 | 218 | 0.00 | 0.00 | 4.73E-01 | 0.53 | 1.43E-01 | - | - | - |
| Ingenuity or creativity demand from job | Psychosocial | 29719 | -0.01 | 0.00 | 2.01E-01 | 3.34E-01 | 162 | 29077 | 0.00 | 0.00 | 7.78E-01 | 8.80E-01 | 245 | 0.00 | 0.00 | 4.74E-01 | 0.17 | 2.74E-01 | - | - | - |
| Smoked fish/meat | Food | 27762 | 0.00 | 0.00 | 7.48E-01 | 8.22E-01 | 245 | 27043 | 0.00 | 0.00 | 5.32E-01 | 6.99E-01 | 210 | 0.00 | 0.00 | 5.07E-01 | 0.00 | 8.19E-01 | - | - | - |
| Confidence status | Psychosocial | 24262 | 0.00 | 0.00 | 5.70E-01 | 6.94E-01 | 221 | 23827 | 0.00 | 0.01 | 7.18E-01 | 8.35E-01 | 238 | 0.00 | 0.00 | 5.10E-01 | 0.00 | 8.89E-01 | - | - | - |
| Boiled coffee | Beverage | 27762 | 0.00 | 0.00 | 9.57E-01 | 9.63E-01 | 267 | 27043 | 0.00 | 0.00 | 3.72E-01 | 5.51E-01 | 186 | 0.00 | 0.00 | 5.11E-01 | 0.00 | 5.44E-01 | - | - | - |
| Pain during the last four weeks | General health | 14269 | 0.01 | 0.01 | 2.33E-01 | 3.60E-01 | 174 | 14063 | 0.00 | 0.01 | 7.64E-01 | 8.71E-01 | 243 | 0.00 | 0.00 | 5.16E-01 | 0.09 | 2.96E-01 | - | - | - |
| Travel to work: Irregular travel mode to work vs passive travel to work | Physical activity | 19549 | 0.02 | 0.02 | 2.73E-01 | 4.11E-01 | 179 | 19201 | 0.00 | 0.02 | 8.27E-01 | 9.05E-01 | 252 | 0.01 | 0.01 | 5.31E-01 | 0.00 | 3.55E-01 | - | - | - |
| Disaccharides intake (g/day) | Nutrients | 27762 | 0.00 | 0.00 | 8.34E-01 | 8.80E-01 | 255 | 27043 | 0.01 | 0.00 | 2.61E-01 | 4.24E-01 | 168 | 0.00 | 0.00 | 5.32E-01 | 0.00 | 3.39E-01 | - | - | - |
| Sedentary or standing work | Physical activity | 29420 | 0.01 | 0.01 | 3.89E-01 | 5.34E-01 | 196 | 28753 | 0.00 | 0.01 | 9.91E-01 | 9.91E-01 | 277 | 0.00 | 0.01 | 5.44E-01 | 0.00 | 5.41E-01 | - | - | - |
| Iodine intake (ug/day) | Nutrients | 27762 | 0.00 | 0.00 | 9.72E-01 | 9.72E-01 | 269 | 27043 | 0.00 | 0.00 | 4.02E-01 | 5.86E-01 | 190 | 0.00 | 0.00 | 5.46E-01 | 0.00 | 5.60E-01 | - | - | - |
| Rice | Food | 27762 | 0.00 | 0.00 | 7.30E-01 | 8.08E-01 | 243 | 27043 | -0.01 | 0.00 | 2.27E-01 | 3.94E-01 | 160 | 0.00 | 0.00 | 5.55E-01 | 0.19 | 2.68E-01 | - | - | - |
| Physical limitation to participate in moderately demanding activities: moving a table, vacuuming, walking in the forest or gardening | General health | 14267 | 0.00 | 0.01 | 4.81E-01 | 6.11E-01 | 212 | 14094 | 0.00 | 0.01 | 9.05E-01 | 9.51E-01 | 263 | 0.00 | 0.00 | 5.57E-01 | 0.00 | 6.84E-01 | - | - | - |
| Selenium intake (ug/day) | Nutrients | 27762 | 0.01 | 0.00 | 1.32E-01 | 2.42E-01 | 146 | 27043 | 0.00 | 0.00 | 4.67E-01 | 6.55E-01 | 197 | 0.00 | 0.00 | 5.57E-01 | 0.59 | 1.17E-01 | - | - | - |
| Meat stew | Food | 27762 | 0.00 | 0.00 | 3.73E-01 | 5.18E-01 | 194 | 27043 | 0.00 | 0.00 | 9.30E-01 | 9.57E-01 | 269 | 0.00 | 0.00 | 5.61E-01 | 0.00 | 4.97E-01 | - | - | - |
| Feel the need to reduce alcohol consumption | Alcohol | 27900 | 0.01 | 0.01 | 3.07E-01 | 4.39E-01 | 188 | 27310 | 0.00 | 0.01 | 8.27E-01 | 9.05E-01 | 253 | 0.01 | 0.01 | 5.65E-01 | 0.00 | 3.83E-01 | - | - | - |
| For how much of the time during the last four weeks have you felt happy? | General health | 14263 | 0.00 | 0.01 | 5.73E-01 | 6.94E-01 | 222 | 14061 | 0.00 | 0.01 | 8.22E-01 | 9.05E-01 | 251 | 0.00 | 0.00 | 5.74E-01 | 0.00 | 8.19E-01 | - | - | - |
| Satisfaction with work situation | Psychosocial | 23991 | -0.01 | 0.00 | 2.09E-01 | 3.35E-01 | 168 | 23556 | 0.00 | 0.01 | 6.26E-01 | 7.64E-01 | 227 | 0.00 | 0.00 | 5.75E-01 | 0.33 | 2.21E-01 | - | - | - |
| How much has the pain during the last four weeks disturbed your normal work? | General health | 14229 | 0.01 | 0.01 | 2.50E-01 | 3.82E-01 | 176 | 14039 | 0.00 | 0.01 | 6.73E-01 | 8.11E-01 | 230 | 0.00 | 0.00 | 5.92E-01 | 0.18 | 2.70E-01 | - | - | - |
| Equol intake (ug/day) | Nutrients | 27762 | 0.00 | 0.00 | 8.96E-01 | 9.20E-01 | 262 | 27043 | 0.00 | 0.00 | 5.33E-01 | 6.99E-01 | 211 | 0.00 | 0.00 | 5.98E-01 | 0.00 | 7.20E-01 | - | - | - |
| Linolenic acid intake (g/day) | Nutrients | 27762 | -0.01 | 0.00 | 7.86E-02 | 1.70E-01 | 124 | 27043 | 0.01 | 0.00 | 2.56E-01 | 4.24E-01 | 167 | 0.00 | 0.00 | 6.28E-01 | 0.76 | 4.16E-02 | - | - | - |
| Extent to what your physical and emotional health disrupted your usual social life during the last four weeks | General health | 14278 | 0.01 | 0.01 | 2.44E-01 | 3.74E-01 | 175 | 14088 | 0.00 | 0.01 | 5.51E-01 | 7.17E-01 | 213 | 0.00 | 0.00 | 6.68E-01 | 0.35 | 2.15E-01 | - | - | - |
| Physical limitation that reduced the normal time spent at work or in other activities during the last four weeks | General health | 14297 | 0.01 | 0.02 | 6.61E-01 | 7.60E-01 | 234 | 14087 | -0.02 | 0.02 | 3.08E-01 | 4.74E-01 | 180 | -0.01 | 0.02 | 6.84E-01 | 0.06 | 3.02E-01 | - | - | - |
| Total protein intake (g/day) | Nutrients | 27762 | 0.00 | 0.00 | 3.62E-01 | 5.04E-01 | 193 | 27043 | 0.00 | 0.00 | 6.94E-01 | 8.31E-01 | 231 | 0.00 | 0.00 | 7.00E-01 | 0.00 | 3.60E-01 | - | - | - |
| Brewed (filtered) coffee | Beverage | 27762 | 0.00 | 0.00 | 6.16E-01 | 7.24E-01 | 229 | 27043 | 0.00 | 0.00 | 2.90E-01 | 4.59E-01 | 175 | 0.00 | 0.00 | 7.11E-01 | 0.19 | 2.67E-01 | - | - | - |
| Satisfaction with accomodation | Psychosocial | 24307 | 0.00 | 0.00 | 6.70E-01 | 7.61E-01 | 236 | 23888 | 0.00 | 0.01 | 9.34E-01 | 9.58E-01 | 270 | 0.00 | 0.00 | 7.17E-01 | 0.00 | 8.11E-01 | - | - | - |
| Milk, sour milk (1,5%) | Beverage | 27762 | 0.00 | 0.00 | 8.15E-01 | 8.70E-01 | 252 | 27043 | 0.00 | 0.00 | 8.05E-01 | 8.96E-01 | 249 | 0.00 | 0.00 | 7.34E-01 | 0.00 | 9.86E-01 | - | - | - |
| Emotional problems that made you do less than you wanted during the last four weeks | General health | 14297 | 0.02 | 0.02 | 2.96E-01 | 4.28E-01 | 186 | 14089 | -0.03 | 0.02 | 1.25E-01 | 2.71E-01 | 128 | 0.00 | 0.01 | 7.41E-01 | 0.70 | 6.74E-02 | - | - | - |
| Total energy intake (kcal/day) | Nutrients | 27762 | 0.00 | 0.01 | 5.43E-01 | 6.66E-01 | 219 | 27043 | 0.00 | 0.01 | 8.39E-01 | 9.08E-01 | 256 | 0.00 | 0.00 | 7.66E-01 | 0.00 | 5.69E-01 | - | - | - |
| Marital status: Single vs Widow/widower | Social | 3199 | 0.03 | 0.05 | 4.90E-01 | 6.16E-01 | 214 | 3098 | -0.02 | 0.05 | 7.15E-01 | 8.35E-01 | 237 | 0.01 | 0.03 | 7.71E-01 | 0.00 | 4.68E-01 | - | - | - |
| Vitamin B3 intake (mg/day) | Nutrients | 27762 | 0.01 | 0.00 | 6.93E-02 | 1.53E-01 | 120 | 27043 | -0.01 | 0.00 | 1.40E-01 | 2.87E-01 | 132 | 0.00 | 0.00 | 7.75E-01 | 0.81 | 2.01E-02 | - | - | - |
| Medium beer | Alcohol | 27762 | 0.00 | 0.00 | 5.83E-01 | 6.94E-01 | 226 | 27043 | 0.00 | 0.00 | 8.68E-01 | 9.33E-01 | 257 | 0.00 | 0.00 | 7.78E-01 | 0.00 | 6.18E-01 | - | - | - |
| Sometimes physically straining work | Physical activity | 29420 | 0.01 | 0.01 | 4.49E-01 | 5.82E-01 | 207 | 28753 | 0.00 | 0.01 | 7.03E-01 | 8.31E-01 | 233 | 0.00 | 0.01 | 7.82E-01 | 0.00 | 4.23E-01 | - | - | - |
| Memory status | Psychosocial | 24243 | 0.00 | 0.00 | 7.66E-01 | 8.35E-01 | 247 | 23805 | 0.00 | 0.01 | 9.27E-01 | 9.57E-01 | 268 | 0.00 | 0.00 | 7.82E-01 | 0.00 | 8.87E-01 | - | - | - |
| Number of friends that can come to your home at any time and feel at home | Social | 30212 | 0.00 | 0.00 | 4.50E-01 | 5.82E-01 | 208 | 29536 | 0.00 | 0.00 | 6.96E-01 | 8.31E-01 | 232 | 0.00 | 0.00 | 7.87E-01 | 0.00 | 4.20E-01 | - | - | - |
| Physical limitation that made you not being able to perform certain work tasks or other activities during the last four weeks | General health | 14224 | 0.00 | 0.02 | 9.45E-01 | 9.56E-01 | 266 | 14034 | -0.01 | 0.02 | 7.06E-01 | 8.31E-01 | 235 | 0.00 | 0.01 | 8.29E-01 | 0.00 | 7.52E-01 | - | - | - |
| Cohabitation: Live alone vs Only children | Social | 4521 | 0.00 | 0.03 | 9.05E-01 | 9.22E-01 | 264 | 4514 | -0.01 | 0.03 | 6.50E-01 | 7.87E-01 | 229 | 0.00 | 0.02 | 8.33E-01 | 0.00 | 6.75E-01 | - | - | - |
| Sodium intake (mg/day) | Nutrients | 27761 | 0.01 | 0.00 | 8.99E-02 | 1.82E-01 | 133 | 27042 | -0.01 | 0.00 | 1.40E-01 | 2.87E-01 | 134 | 0.00 | 0.00 | 8.41E-01 | 0.80 | 2.52E-02 | - | - | - |
| Cheese 28% | Food | 27762 | 0.00 | 0.00 | 8.33E-01 | 8.80E-01 | 254 | 27043 | 0.00 | 0.00 | 6.35E-01 | 7.71E-01 | 228 | 0.00 | 0.00 | 8.60E-01 | 0.00 | 6.25E-01 | - | - | - |
| Satisfaction with leisure time | Psychosocial | 24252 | 0.00 | 0.00 | 7.33E-01 | 8.08E-01 | 244 | 23827 | 0.00 | 0.01 | 9.13E-01 | 9.54E-01 | 265 | 0.00 | 0.00 | 8.66E-01 | 0.00 | 7.52E-01 | - | - | - |
| Low fat margarine on bread | Food | 27762 | 0.00 | 0.00 | 4.41E-01 | 5.81E-01 | 204 | 27043 | 0.00 | 0.00 | 5.50E-01 | 7.17E-01 | 212 | 0.00 | 0.00 | 8.88E-01 | 0.00 | 3.34E-01 | - | - | - |
| Physical limitation that made you do less than you wanted during the last four weeks | General health | 14280 | 0.01 | 0.02 | 4.64E-01 | 5.97E-01 | 209 | 14090 | -0.01 | 0.02 | 5.68E-01 | 7.25E-01 | 217 | 0.00 | 0.01 | 9.04E-01 | 0.00 | 3.57E-01 | - | - | - |
| Everyday exercise satisfaction | Physical activity | 18991 | 0.01 | 0.00 | 1.83E-01 | 3.07E-01 | 160 | 18444 | -0.01 | 0.00 | 2.39E-01 | 4.05E-01 | 163 | 0.00 | 0.00 | 9.04E-01 | 0.68 | 7.60E-02 | - | - | - |
| Snuff status: Snuff users vs non-snuff users | Tobacco use | 25853 | -0.02 | 0.01 | 2.08E-01 | 3.35E-01 | 167 | 25358 | 0.02 | 0.01 | 1.41E-01 | 2.87E-01 | 135 | 0.00 | 0.01 | 9.06E-01 | 0.73 | 5.31E-02 | - | - | - |
| Number of people with whom you can speak openly | Social | 30301 | 0.00 | 0.00 | 6.86E-01 | 7.66E-01 | 241 | 29599 | 0.00 | 0.00 | 5.80E-01 | 7.32E-01 | 219 | 0.00 | 0.00 | 9.25E-01 | 0.00 | 4.97E-01 | - | - | - |
| Parents or siblings had a cerebral hemorrhage/thrombosis or cardiac infarction before the age of 60 | General health | 29750 | 0.00 | 0.01 | 8.81E-01 | 9.15E-01 | 259 | 29121 | 0.00 | 0.01 | 8.02E-01 | 8.96E-01 | 248 | 0.00 | 0.01 | 9.47E-01 | 0.00 | 7.76E-01 | - | - | - |
| Pancake, waffle, Swedish dumpling | Food | 27762 | 0.01 | 0.00 | 4.54E-02 | 1.12E-01 | 109 | 27043 | -0.01 | 0.00 | 2.99E-02 | 9.98E-02 | 83 | 0.00 | 0.00 | 9.49E-01 | 0.89 | 3.15E-03 | - | - | - |
| Vision status | General health | 24281 | 0.00 | 0.01 | 7.53E-01 | 8.23E-01 | 246 | 23837 | 0.00 | 0.01 | 7.21E-01 | 8.36E-01 | 239 | 0.00 | 0.00 | 9.81E-01 | 0.00 | 6.34E-01 | - | - | - |
| Participation in study circles | Social | 10604 | 0.01 | 0.02 | 5.25E-01 | 6.51E-01 | 217 | 10260 | -0.01 | 0.02 | 5.14E-01 | 6.91E-01 | 206 | 0.00 | 0.02 | 9.90E-01 | 0.00 | 3.62E-01 | - | - | - |
| Amount of alcohol drunk in a day | Alcohol | 10836 | 0.04 | 0.01 | - | - | 273 | 10637 | 0.03 | 0.01 | 1.48E-03 | **1.05E-02** | 39 | 0.03 | 0.01 | - | 0.00 | 4.47E-01 | - | - | - |
| Breath-holds during sleep | Sleep | 7839 | 0.02 | 0.01 | - | - | 283 | 7680 | 0.03 | 0.01 | 4.21E-03 | **2.43E-02** | 48 | 0.03 | 0.01 | - | 0.00 | 6.55E-01 | - | - | - |
| Snore during sleep | Sleep | 10447 | 0.02 | 0.01 | - | - | 282 | 10334 | 0.02 | 0.01 | 5.83E-02 | 1.57E-01 | 103 | 0.02 | 0.01 | - | 0.00 | 9.57E-01 | - | - | - |
| Risk of sleeping while sitting still after having lunch | Sleep | 11164 | 0.00 | 0.01 | - | - | 281 | 11034 | -0.01 | 0.01 | 9.54E-02 | 2.22E-01 | 119 | 0.00 | 0.01 | - | 0.61 | 1.08E-01 | - | - | - |
| Overall state of health compared to other your age | General health | - | - | - | - | - | - | 15543 | -0.01 | 0.01 | 1.32E-01 | 2.80E-01 | 131 | - | - | - | - | - | - | - | - |
| Potato salad | Food | 5468 | 0.01 | 0.01 | 1.76E-01 | 2.98E-01 | 159 | 5254 | 0.02 | 0.01 | 1.42E-01 | 2.87E-01 | 137 | 0.01 | 0.01 | - | 0.00 | 7.64E-01 | - | - | - |
| Soft cheese | Food | 5468 | -0.01 | 0.01 | 2.82E-01 | 4.15E-01 | 183 | 5254 | 0.01 | 0.01 | 1.87E-01 | 3.45E-01 | 150 | 0.00 | 0.01 | - | 0.73 | 5.21E-02 | - | - | - |
| Mashed potato | Food | 5468 | 0.01 | 0.01 | 1.53E-01 | 2.69E-01 | 153 | 5254 | 0.01 | 0.01 | 2.26E-01 | 3.94E-01 | 158 | 0.01 | 0.01 | - | 0.00 | 9.99E-01 | - | - | - |
| Liver, kidney | Food | 5468 | 0.01 | 0.01 | 1.74E-01 | 2.97E-01 | 158 | 5254 | 0.01 | 0.01 | 2.62E-01 | 4.24E-01 | 170 | 0.01 | 0.01 | - | 0.00 | 9.39E-01 | - | - | - |
| Mixed frozen vegetables | Food | 5468 | 0.00 | 0.01 | 6.89E-01 | 7.66E-01 | 242 | 5254 | -0.01 | 0.01 | 2.77E-01 | 4.42E-01 | 173 | 0.00 | 0.01 | - | 0.29 | 2.37E-01 | - | - | - |
| Risk of sleeping while sitting and reading | Sleep | 11080 | 0.00 | 0.01 | - | - | 276 | 10943 | -0.01 | 0.01 | 3.20E-01 | 4.90E-01 | 181 | -0.01 | 0.01 | - | 0.00 | 7.66E-01 | - | - | - |
| Times during last year that you felt guilty because of your drinking | Alcohol | 11026 | 0.01 | 0.01 | - | - | 275 | 10821 | 0.01 | 0.01 | 4.22E-01 | 6.09E-01 | 192 | 0.01 | 0.01 | - | 0.00 | 6.70E-01 | - | - | - |
| Blood based food | Food | 5468 | 0.02 | 0.01 | 4.23E-02 | 1.05E-01 | 108 | 5254 | -0.01 | 0.01 | 4.26E-01 | 6.11E-01 | 193 | 0.01 | 0.01 | - | 0.80 | 2.37E-02 | - | - | - |
| Risk of sleeping while lying down resting in the afternoon | Sleep | 11156 | 0.01 | 0.01 | - | - | 280 | 11029 | -0.01 | 0.01 | 4.89E-01 | 6.71E-01 | 202 | 0.00 | 0.01 | - | 0.07 | 3.01E-01 | - | - | - |
| Soft whey cheese | Food | 5468 | -0.01 | 0.01 | 1.22E-01 | 2.28E-01 | 143 | 5254 | 0.01 | 0.01 | 5.22E-01 | 6.95E-01 | 208 | 0.00 | 0.01 | - | 0.65 | 9.31E-02 | - | - | - |
| Shellfish (e.g. shrimps, scallops) | Food | 5468 | 0.00 | 0.01 | 6.50E-01 | 7.51E-01 | 233 | 5254 | 0.00 | 0.01 | 6.23E-01 | 7.64E-01 | 225 | 0.00 | 0.01 | - | 0.00 | 4.80E-01 | - | - | - |
| Blota (broth + bread) | Food | 5468 | 0.01 | 0.01 | 1.43E-01 | 2.54E-01 | 151 | 5254 | 0.00 | 0.01 | 7.41E-01 | 8.55E-01 | 240 | 0.01 | 0.01 | - | 0.43 | 1.86E-01 | - | - | - |
| Time spent in a week in moderately steneous activities | Physical activity | 10961 | -0.02 | 0.01 | - | - | 270 | 10769 | -0.02 | 0.01 | - | - | 278 | -0.02 | 0.01 | - | 0.00 | 9.84E-01 | - | - | - |
| Amount of exercise during the last 12 months | Physical activity | 11301 | -0.03 | 0.01 | - | - | 271 | 11123 | -0.03 | 0.01 | - | - | 279 | -0.03 | 0.01 | - | 0.00 | 5.21E-01 | - | - | - |
| Frequency of alcohol consumption | Alcohol | 11344 | 0.02 | 0.01 | - | - | 272 | 11199 | 0.02 | 0.01 | - | - | 280 | 0.02 | 0.01 | - | 0.00 | 9.29E-01 | - | - | - |

| | | | | | | | | | | | | | | | | | | | | | |
|---|---|---|---|---|---|---|---|---|---|---|---|---|---|---|---|---|---|---|---|---|---|
| Frequency of drinking six or more glasses at the same occasion | Alcohol | 11039 | 0.03 | 0.01 | - | - | 274 | 10848 | 0.03 | 0.01 | - | - | 281 | 0.03 | 0.01 | - | 0.00 | 8.20E-01 | - | - | - |
| Risk of sleeping while watching TV | Sleep | 11202 | 0.00 | 0.01 | - | - | 277 | 11058 | 0.00 | 0.01 | - | - | 282 | 0.00 | 0.01 | - | 0.00 | 6.98E-01 | - | - | - |
| Risk of sleeping while sitting inactive in a public place | Sleep | 11127 | 0.00 | 0.01 | - | - | 278 | 10988 | 0.00 | 0.01 | - | - | 283 | 0.00 | 0.01 | - | 0.00 | 9.49E-01 | - | - | - |
| Risk of sleeping as a passenger in a car for one hour without break | Sleep | 11087 | 0.01 | 0.01 | - | - | 279 | 10942 | -0.01 | 0.01 | - | - | 284 | 0.00 | 0.01 | - | 0.45 | 1.77E-01 | - | - | - |

**Supplementary Table 11**. Linear mixed model association results for 2h glucose

| | | Training set | | | | | | Testing set | | | | | | Metanalysis | | | | | R2 | | |
|---|---|---|---|---|---|---|---|---|---|---|---|---|---|---|---|---|---|---|---|---|---|
| **Description** | **Group** | **N** | **Effect estimate** | **S.E.** | **p-value** | **p-value$_{FDR}$** | **p-value rank** | **N** | **Effect estimate** | **S.E.** | **p-value** | **p-value$_{FDR}$** | **p-value rank** | **Effect estimate** | **S.E.** | **p-value** | **$I^2$** | **Q p-value** | **marginal R2** | **conditional R2** | **R2 rank** |
| Informed of having high blood pressure | General health | 29289 | 0.25 | 0.02 | 2.47E-31 | **6.67E-29** | 1 | 28637 | 0.26 | 0.02 | 6.16E-34 | **1.70E-31** | 1 | 0.26 | 0.02 | 1.75E-63 | 0.00 | 6.83E-01 | 0.09 | 0.40 | 23 |
| Smoking status: Smokers vs non-smokers | Tobacco use | 18623 | -0.31 | 0.03 | 4.49E-27 | **6.06E-25** | 2 | 18325 | -0.32 | 0.03 | 3.74E-29 | **5.18E-27** | 2 | -0.31 | 0.02 | 1.68E-54 | 0.00 | 8.23E-01 | 0.09 | 0.41 | 21 |
| Cambridge physical activity index | Physical activity | 28071 | -0.08 | 0.01 | 9.15E-22 | **6.18E-20** | 4 | 27436 | -0.08 | 0.01 | 5.68E-20 | **3.93E-18** | 4 | -0.08 | 0.01 | 4.48E-40 | 0.00 | 8.26E-01 | 0.08 | 0.40 | 28 |
| Parents or siblings have diabetes | General health | 28782 | 0.22 | 0.02 | 8.65E-22 | **6.18E-20** | 3 | 28158 | 0.20 | 0.02 | 2.83E-18 | **1.57E-16** | 5 | 0.21 | 0.02 | 2.38E-38 | 0.00 | 5.59E-01 | 0.08 | 0.40 | 26 |
| Years smoking | Tobacco use | 25591 | -0.09 | 0.01 | 7.24E-17 | **2.79E-15** | 7 | 24922 | -0.10 | 0.01 | 3.30E-22 | **3.05E-20** | 3 | -0.09 | 0.01 | 3.08E-37 | 0.00 | 3.28E-01 | 0.09 | 0.40 | 24 |
| Secoisolariciresinol intake (ug/day) | Nutrients | 26853 | -0.08 | 0.01 | 3.06E-18 | **1.65E-16** | 5 | 26173 | -0.07 | 0.01 | 1.38E-15 | **4.79E-14** | 8 | -0.08 | 0.01 | 3.66E-32 | 0.00 | 5.97E-01 | 0.10 | 0.41 | 6 |
| Brewed (filtered) coffee | Beverage | 26853 | -0.07 | 0.01 | 7.33E-15 | **2.20E-13** | 9 | 26173 | -0.08 | 0.01 | 1.48E-17 | **6.82E-16** | 6 | -0.07 | 0.01 | 9.43E-31 | 0.00 | 5.58E-01 | 0.10 | 0.41 | 7 |
| Number of cigarretes smoked per day (in groups) | Tobacco use | 24067 | -0.07 | 0.01 | 1.33E-13 | **3.58E-12** | 10 | 23715 | -0.08 | 0.01 | 3.75E-17 | **1.48E-15** | 7 | -0.08 | 0.01 | 4.66E-29 | 0.00 | 4.56E-01 | 0.09 | 0.40 | 22 |
| Exercise during the last three months | Physical activity | 28980 | -0.07 | 0.01 | 1.72E-15 | **5.81E-14** | 8 | 28336 | -0.07 | 0.01 | 2.15E-14 | **6.63E-13** | 9 | -0.07 | 0.01 | 2.66E-28 | 0.00 | 8.66E-01 | 0.08 | 0.40 | 29 |
| Fitness status | Physical activity | 23543 | -0.09 | 0.01 | 3.73E-17 | **1.68E-15** | 6 | 23107 | -0.07 | 0.01 | 1.20E-12 | **3.31E-11** | 10 | -0.08 | 0.01 | 4.59E-28 | 0.00 | 3.74E-01 | 0.07 | 0.41 | 45 |
| Snuff status: Snuff users vs non-snuff users | Tobacco use | 24955 | -0.19 | 0.03 | 8.45E-13 | **2.07E-11** | 11 | 24459 | -0.17 | 0.03 | 5.30E-10 | **1.13E-08** | 13 | -0.18 | 0.02 | 3.28E-21 | 0.00 | 5.38E-01 | 0.08 | 0.40 | 27 |
| Years using snuff | Tobacco use | 27589 | -0.07 | 0.01 | 8.89E-11 | **1.85E-09** | 13 | 26969 | -0.06 | 0.01 | 1.39E-08 | **2.41E-07** | 16 | -0.06 | 0.01 | 7.97E-18 | 0.00 | 5.91E-01 | 0.08 | 0.40 | 30 |
| Overall state of health compared to other your age | General health | 15456 | -0.07 | 0.01 | 2.91E-11 | **6.55E-10** | 12 | 14990 | -0.05 | 0.01 | 9.88E-07 | **1.19E-05** | 23 | -0.06 | 0.01 | 3.00E-16 | 0.33 | 2.21E-01 | 0.11 | 0.43 | 1 |
| Travel to work: Cycle to work vs passive travel to work | Physical activity | 22133 | -0.12 | 0.02 | 2.43E-07 | **3.27E-06** | 20 | 21508 | -0.15 | 0.02 | 1.66E-10 | **4.18E-09** | 11 | -0.14 | 0.02 | 3.32E-16 | 0.00 | 3.44E-01 | 0.08 | 0.40 | 44 |
| Average portion size of vegetables based on photographic illustration of four sizes (smallest to largest) | Food | 26853 | -0.05 | 0.01 | 9.55E-09 | **1.72E-07** | 15 | 26173 | -0.05 | 0.01 | 1.15E-08 | **2.11E-07** | 15 | -0.05 | 0.01 | 5.78E-16 | 0.00 | 9.90E-01 | 0.08 | 0.40 | 36 |
| Average portion size of meat/fish based on photographic illustration of four sizes (smallest to largest) | Food | 26853 | -0.05 | 0.01 | 2.18E-07 | **3.10E-06** | 19 | 26173 | -0.06 | 0.01 | 4.28E-10 | **9.87E-09** | 12 | -0.06 | 0.01 | 6.78E-16 | 0.00 | 4.18E-01 | 0.08 | 0.40 | 35 |
| Self rate of overall health | General health | 13833 | -0.07 | 0.01 | 9.53E-08 | **1.51E-06** | 17 | 13669 | -0.08 | 0.01 | 5.42E-08 | **8.83E-07** | 17 | -0.08 | 0.01 | 2.55E-14 | 0.00 | 9.04E-01 | - | - | - |
| Frequency of hunting or fishing during leisure time | Physical activity | 17139 | -0.05 | 0.01 | 1.72E-05 | **1.79E-04** | 26 | 16623 | -0.07 | 0.01 | 2.19E-09 | **4.34E-08** | 14 | -0.06 | 0.01 | 3.75E-13 | 0.35 | 2.14E-01 | 0.10 | 0.40 | 3 |
| Excellent health | General health | 13784 | -0.07 | 0.01 | 1.30E-07 | **1.96E-06** | 18 | 13618 | -0.07 | 0.01 | 2.60E-06 | **2.88E-05** | 25 | -0.07 | 0.01 | 1.66E-12 | 0.00 | 6.93E-01 | - | - | - |
| Overall state of health during the last year | General health | 29280 | -0.05 | 0.01 | 7.32E-08 | **1.23E-06** | 16 | 28633 | -0.04 | 0.01 | 7.00E-06 | **7.45E-05** | 26 | -0.04 | 0.01 | 2.78E-12 | 0.00 | 5.61E-01 | 0.08 | 0.40 | 32 |
| Number of snuff boxes per week | Tobacco use | 28325 | -0.05 | 0.01 | 2.73E-07 | **3.51E-06** | 21 | 27700 | -0.05 | 0.01 | 2.57E-06 | **2.88E-05** | 24 | -0.05 | 0.01 | 3.32E-12 | 0.00 | 8.12E-01 | 0.08 | 0.40 | 31 |
| Get sick more often than other people | General health | 13788 | 0.07 | 0.01 | 1.69E-06 | **2.08E-05** | 22 | 13610 | 0.06 | 0.01 | 1.25E-05 | **1.15E-04** | 30 | 0.06 | 0.01 | 9.27E-11 | 0.00 | 7.91E-01 | - | - | - |
| Smoking status: Former smokers vs non-smokers | Tobacco use | 21103 | -0.10 | 0.02 | 5.85E-05 | **5.64E-04** | 28 | 20390 | -0.12 | 0.02 | 3.14E-07 | **4.35E-06** | 20 | -0.11 | 0.02 | 1.07E-10 | 0.00 | 4.27E-01 | 0.09 | 0.41 | 25 |
| High physical demand from job | Physical activity | 28915 | -0.05 | 0.01 | 7.75E-09 | **1.49E-07** | 14 | 28301 | -0.03 | 0.01 | 1.07E-03 | **5.60E-03** | 53 | -0.04 | 0.01 | 1.53E-10 | 0.67 | 8.00E-02 | 0.08 | 0.40 | 38 |
| Magnesium intake (mg/day) | Nutrients | 26853 | -0.03 | 0.01 | 3.56E-04 | **2.70E-03** | 35 | 26173 | -0.05 | 0.01 | 2.21E-07 | **3.33E-06** | 18 | -0.04 | 0.01 | 6.21E-10 | 0.25 | 2.47E-01 | 0.09 | 0.42 | 10 |
| Physical limitation to participate in steneous activities: running, lifting heavy objects, taking part in physically demanding sports | General health | 13800 | 0.07 | 0.01 | 2.77E-06 | **3.26E-05** | 23 | 13646 | 0.06 | 0.01 | 6.45E-05 | **4.58E-04** | 39 | 0.06 | 0.01 | 7.96E-10 | 0.00 | 6.44E-01 | - | - | - |
| Boiled coffee | Beverage | 26853 | -0.04 | 0.01 | 2.29E-05 | **2.29E-04** | 27 | 26173 | -0.04 | 0.01 | 2.91E-05 | **2.44E-04** | 33 | -0.04 | 0.01 | 2.67E-09 | 0.00 | 9.89E-01 | 0.09 | 0.41 | 8 |
| Snuff status: Former snuff users vs non-snuff users | Tobacco use | 23367 | -0.10 | 0.03 | 9.65E-04 | **5.79E-03** | 45 | 22806 | -0.15 | 0.03 | 6.39E-07 | **8.05E-06** | 22 | -0.13 | 0.02 | 4.90E-09 | 0.32 | 2.26E-01 | 0.08 | 0.41 | 43 |
| Strong beer | Alcohol | 26853 | -0.04 | 0.01 | 1.15E-04 | **1.00E-03** | 31 | 26173 | -0.04 | 0.01 | 6.06E-05 | **4.42E-04** | 38 | -0.04 | 0.01 | 2.67E-08 | 0.00 | 8.96E-01 | 0.09 | 0.41 | 9 |
| Frequency of cycling during leisure time | Physical activity | 24025 | -0.04 | 0.01 | 6.51E-05 | **6.06E-04** | 29 | 23332 | -0.04 | 0.01 | 1.14E-04 | **7.70E-04** | 41 | -0.04 | 0.01 | 2.81E-08 | 0.00 | 9.62E-01 | 0.08 | 0.40 | 42 |
| Participation in associations or voluntary organisations | Social | 29095 | -0.05 | 0.02 | 4.94E-03 | **2.30E-02** | 58 | 28483 | -0.09 | 0.02 | 4.50E-07 | **5.94E-06** | 21 | -0.07 | 0.01 | 2.90E-08 | 0.62 | 1.07E-01 | 0.08 | 0.40 | 33 |
| Alcohol intake (g/day) | Alcohol | 26853 | -0.04 | 0.01 | 5.31E-06 | **5.97E-05** | 24 | 26173 | -0.03 | 0.01 | 1.62E-03 | **7.74E-03** | 58 | -0.04 | 0.01 | 5.13E-08 | 0.00 | 3.19E-01 | 0.09 | 0.42 | 11 |
| Physical limitation to participate in moderately demanding activities: moving a table, vacuuming, walking in the forest or gardening | General health | 13798 | 0.03 | 0.01 | 1.23E-02 | **4.76E-02** | 70 | 13643 | 0.07 | 0.01 | 2.28E-07 | **3.33E-06** | 19 | 0.05 | 0.01 | 5.34E-08 | 0.74 | 4.84E-02 | - | - | - |
| Physical limitation to participate in moderately demanding activities: lifting or carrying grocery bags | General health | 13785 | 0.06 | 0.01 | 6.76E-05 | **6.08E-04** | 30 | 13635 | 0.05 | 0.01 | 2.15E-04 | **1.35E-03** | 44 | 0.05 | 0.01 | 5.43E-08 | 0.00 | 8.43E-01 | - | - | - |
| Total energy intake (kcal/day) | Nutrients | 26853 | -0.03 | 0.01 | 1.76E-03 | **9.49E-03** | 50 | 26173 | -0.05 | 0.01 | 1.21E-05 | **1.15E-04** | 29 | -0.04 | 0.01 | 1.11E-07 | 0.00 | 3.81E-01 | 0.09 | 0.42 | 20 |
| As healthy as anyone | General health | 13746 | -0.05 | 0.01 | 6.79E-04 | **4.51E-03** | 40 | 13572 | -0.06 | 0.01 | 4.52E-05 | **3.68E-04** | 34 | -0.05 | 0.01 | 1.24E-07 | 0.00 | 5.98E-01 | - | - | - |
| Rosehip, sweet syrup soup | Food | 26853 | 0.03 | 0.01 | 3.74E-04 | **2.73E-03** | 37 | 26173 | 0.03 | 0.01 | 9.86E-05 | **6.83E-04** | 40 | 0.03 | 0.01 | 1.37E-07 | 0.00 | 8.19E-01 | 0.09 | 0.42 | 14 |
| Sedentary or standing work | Physical activity | 28425 | 0.06 | 0.02 | 1.87E-03 | **9.72E-03** | 52 | 27780 | 0.09 | 0.02 | 1.46E-05 | **1.30E-04** | 31 | 0.07 | 0.01 | 1.42E-07 | 0.00 | 3.78E-01 | 0.08 | 0.40 | 40 |
| Participation in sports or physical exercise associations | Social | 10280 | -0.14 | 0.03 | 7.67E-06 | **8.28E-05** | 25 | 9964 | -0.09 | 0.03 | 3.51E-03 | **1.50E-02** | 64 | -0.11 | 0.02 | 1.64E-07 | 0.15 | 2.79E-01 | 0.07 | 0.47 | 48 |
| Ingenuity or creativity demand from job | Psychosocial | 28706 | -0.03 | 0.01 | 9.58E-04 | **5.79E-03** | 44 | 28098 | -0.04 | 0.01 | 4.91E-05 | **3.78E-04** | 36 | -0.03 | 0.01 | 1.95E-07 | 0.00 | 5.79E-01 | 0.08 | 0.40 | 37 |
| Teetotaler | Alcohol | 18376 | 0.12 | 0.03 | 3.60E-04 | **2.70E-03** | 36 | 17868 | 0.13 | 0.04 | 2.57E-04 | **1.55E-03** | 46 | 0.12 | 0.02 | 3.30E-07 | 0.00 | 8.65E-01 | 0.10 | 0.40 | 2 |
| Sometimes physically straining work | Physical activity | 28425 | -0.06 | 0.02 | 3.84E-03 | **1.85E-02** | 56 | 27780 | -0.08 | 0.02 | 1.74E-05 | **1.51E-04** | 32 | -0.07 | 0.01 | 3.87E-07 | 0.06 | 3.03E-01 | 0.08 | 0.40 | 41 |
| Physical limitation to participate in moderately demanding activities: walking up several stairs | General health | 13801 | 0.04 | 0.01 | 1.31E-02 | **4.92E-02** | 72 | 13628 | 0.06 | 0.01 | 7.86E-06 | **8.06E-05** | 27 | 0.05 | 0.01 | 8.72E-07 | 0.53 | 1.43E-01 | - | - | - |
| Apple, pear, peach, orange, mandarin and grapefruit | Food | 26853 | -0.03 | 0.01 | 9.86E-04 | **5.79E-03** | 46 | 26173 | -0.03 | 0.01 | 4.56E-04 | **2.58E-03** | 49 | -0.03 | 0.01 | 1.53E-06 | 0.00 | 8.62E-01 | 0.09 | 0.42 | 15 |
| Frequency of shoverling snow during leisure time | Physical activity | 17583 | -0.04 | 0.01 | 2.42E-04 | **1.92E-03** | 34 | 17086 | -0.03 | 0.01 | 1.89E-03 | **8.73E-03** | 60 | -0.04 | 0.01 | 1.62E-06 | 0.00 | 7.22E-01 | 0.10 | 0.39 | 4 |
| Potassium intake (mg/day) | Nutrients | 26853 | -0.02 | 0.01 | 2.36E-02 | 7.77E-02 | 82 | 26173 | -0.04 | 0.01 | 1.02E-05 | **1.01E-04** | 28 | -0.03 | 0.01 | 2.38E-06 | 0.57 | 1.26E-01 | - | - | - |
| Eat breakfast from 2000 | Food | 16387 | 0.13 | 0.04 | 4.00E-03 | **1.90E-02** | 57 | 16006 | 0.16 | 0.04 | 2.26E-04 | **1.39E-03** | 45 | 0.14 | 0.03 | 3.27E-06 | 0.00 | 6.23E-01 | 0.06 | 0.48 | 49 |
| Lariciresinol intake (ug/day) | Nutrients | 26853 | -0.03 | 0.01 | 7.30E-04 | **4.58E-03** | 43 | 26173 | -0.03 | 0.01 | 1.72E-03 | **8.09E-03** | 59 | -0.03 | 0.01 | 4.11E-06 | 0.00 | 8.72E-01 | 0.09 | 0.42 | 16 |
| Tea | Beverage | 26853 | 0.03 | 0.01 | 1.80E-03 | **9.53E-03** | 51 | 26173 | 0.03 | 0.01 | 7.83E-04 | **4.25E-03** | 51 | 0.03 | 0.01 | 4.62E-06 | 0.00 | 8.54E-01 | 0.09 | 0.41 | 12 |
| Skill demand from job | Psychosocial | 28777 | -0.02 | 0.01 | 9.82E-03 | **4.02E-02** | 66 | 28136 | -0.03 | 0.01 | 1.93E-04 | **1.25E-03** | 43 | -0.03 | 0.01 | 8.39E-06 | 0.00 | 3.98E-01 | 0.08 | 0.40 | 39 |
| Frequency of picking berries or mushrooms during leisure time | Physical activity | 17397 | -0.04 | 0.01 | 5.67E-04 | **4.03E-03** | 38 | 16906 | -0.03 | 0.01 | 5.27E-03 | **2.15E-02** | 68 | -0.03 | 0.01 | 1.01E-05 | 0.00 | 6.75E-01 | 0.10 | 0.40 | 5 |
| Hamburger | Food | 26853 | 0.03 | 0.01 | 2.31E-04 | **1.89E-03** | 33 | 26173 | 0.02 | 0.01 | 1.46E-02 | **4.92E-02** | 82 | 0.03 | 0.01 | 1.44E-05 | 0.00 | 3.99E-01 | 0.09 | 0.41 | 13 |
| Physical limitation to participate in moderately demanding activities: bending down or kneeling | General health | 13788 | 0.04 | 0.01 | 1.14E-02 | **4.51E-02** | 68 | 13614 | 0.05 | 0.01 | 3.58E-04 | **2.06E-03** | 48 | 0.04 | 0.01 | 1.61E-05 | 0.00 | 4.36E-01 | - | - | - |
| Energy status | Psychosocial | 23530 | -0.03 | 0.01 | 2.11E-03 | **1.06E-02** | 54 | 23079 | -0.03 | 0.01 | 2.51E-03 | **1.10E-02** | 63 | -0.03 | 0.01 | 1.62E-05 | 0.00 | 9.80E-01 | 0.07 | 0.42 | 47 |
| Vision status | General health | 23549 | 0.03 | 0.01 | 5.90E-04 | **4.08E-03** | 39 | 23101 | 0.03 | 0.01 | 1.19E-02 | **4.28E-02** | 77 | 0.03 | 0.01 | 2.53E-05 | 0.00 | 5.31E-01 | 0.07 | 0.42 | 46 |
| Sum of Lariciresinol, Matairesinol, Pinoresinol, Secoisolariciresinol intake (ug/day) | Nutrients | 26853 | -0.03 | 0.01 | 1.46E-03 | **8.19E-03** | 48 | 26173 | -0.03 | 0.01 | 5.84E-03 | **2.34E-02** | 69 | -0.03 | 0.01 | 2.66E-05 | 0.00 | 7.68E-01 | 0.09 | 0.42 | 17 |
| Number of social contacts with the same interests as you | Social | 29241 | -0.02 | 0.01 | 1.27E-02 | **4.81E-02** | 71 | 28579 | -0.03 | 0.01 | 5.56E-04 | **3.08E-03** | 50 | -0.03 | 0.01 | 2.67E-05 | 0.00 | 4.81E-01 | 0.08 | 0.41 | 34 |
| Monosaccharides intake (g/day) | Nutrients | 26853 | -0.03 | 0.01 | 1.32E-03 | **7.59E-03** | 47 | 26173 | -0.03 | 0.01 | 7.27E-03 | **2.70E-02** | 74 | -0.03 | 0.01 | 3.04E-05 | 0.00 | 7.24E-01 | 0.09 | 0.42 | 18 |
| Iron intake (mg/day) | Nutrients | 26853 | 0.03 | 0.01 | 6.87E-04 | **4.51E-03** | 41 | 26173 | 0.02 | 0.01 | 2.02E-02 | 6.23E-02 | 90 | 0.03 | 0.01 | 5.23E-05 | 0.00 | 4.57E-01 | - | - | - |
| Folic acid intake (ug/day) | Nutrients | 26853 | -0.02 | 0.01 | 9.77E-03 | **4.02E-02** | 65 | 26173 | -0.03 | 0.01 | 2.03E-03 | **9.21E-03** | 61 | -0.03 | 0.01 | 6.12E-05 | 0.00 | 7.13E-01 | 0.09 | 0.42 | 19 |
| Satisfaction with economy | Psychosocial | 23556 | 0.02 | 0.01 | 6.32E-02 | 1.60E-01 | 107 | 23123 | 0.04 | 0.01 | 1.55E-04 | **1.02E-03** | 42 | 0.03 | 0.01 | 6.70E-05 | 0.47 | 1.71E-01 | - | - | - |
| Feel the need to reduce alcohol consumption | Alcohol | 26940 | -0.06 | 0.03 | 2.39E-02 | 7.78E-02 | 83 | 26406 | -0.09 | 0.03 | 1.00E-03 | **5.35E-03** | 52 | -0.07 | 0.02 | 8.74E-05 | 0.00 | 4.66E-01 | - | - | - |
| Liquor, spirits | Alcohol | 26853 | -0.02 | 0.01 | 7.36E-02 | 1.81E-01 | 110 | 26173 | -0.03 | 0.01 | 2.74E-04 | **1.61E-03** | 47 | -0.03 | 0.01 | 1.07E-04 | 0.30 | 2.32E-01 | - | - | - |
| Bacon | Food | 26853 | -0.02 | 0.01 | 2.17E-02 | 7.62E-02 | 77 | 26173 | -0.03 | 0.01 | 1.51E-03 | **7.47E-03** | 56 | -0.02 | 0.01 | 1.12E-04 | 0.00 | 5.21E-01 | - | - | - |
| Number of social interactions during a normal week | Social | 29286 | -0.03 | 0.01 | 1.60E-03 | **8.82E-03** | 49 | 28620 | -0.02 | 0.01 | 2.59E-02 | 7.73E-02 | 93 | -0.02 | 0.01 | 1.39E-04 | 0.00 | 5.25E-01 | - | - | - |
| Learn new things at job | Psychosocial | 28790 | -0.01 | 0.01 | 2.09E-01 | 3.67E-01 | 154 | 28169 | -0.04 | 0.01 | 5.42E-05 | **4.06E-04** | 37 | -0.02 | 0.01 | 1.91E-04 | 0.75 | 4.67E-02 | - | - | - |
| For how much of the time during the last four weeks have you felt really alert and strong? | General health | 13778 | -0.03 | 0.01 | 3.95E-02 | 1.12E-01 | 95 | 13595 | -0.04 | 0.01 | 1.62E-03 | **7.74E-03** | 57 | -0.04 | 0.01 | 2.28E-04 | 0.00 | 4.26E-01 | - | - | - |
| Work shifts/weekends | Psychosocial | 28447 | -0.05 | 0.02 | 2.58E-02 | 8.28E-02 | 84 | 27898 | -0.06 | 0.02 | 3.53E-03 | **1.50E-02** | 65 | -0.05 | 0.01 | 2.73E-04 | 0.00 | 6.28E-01 | - | - | - |
| Smoking status: Former occasional smokers vs non-smokers | Tobacco use | 17493 | -0.03 | 0.03 | 3.59E-01 | 5.48E-01 | 176 | 17155 | -0.12 | 0.03 | 4.65E-05 | **3.68E-04** | 35 | -0.08 | 0.02 | 4.00E-04 | 0.80 | 2.67E-02 | - | - | - |
| White cabbage, lettuce, lettuce cabbage, spinach, borecole | Food | 26853 | -0.02 | 0.01 | 1.06E-02 | **4.28E-02** | 67 | 26173 | -0.02 | 0.01 | 1.65E-02 | 5.50E-02 | 83 | -0.02 | 0.01 | 4.60E-04 | 0.00 | 9.24E-01 | - | - | - |
| Medium beer | Alcohol | 26853 | -0.03 | 0.01 | 7.02E-04 | **4.51E-03** | 42 | 26173 | -0.01 | 0.01 | 1.22E-01 | 2.58E-01 | 131 | -0.02 | 0.01 | 4.74E-04 | 0.40 | 1.98E-01 | - | - | - |
| Physical limitation to participate in moderately demanding activities: walking more than 2 km | General health | 13809 | 0.04 | 0.01 | 1.22E-02 | **4.76E-02** | 69 | 13638 | 0.03 | 0.01 | 1.72E-02 | 5.62E-02 | 84 | 0.03 | 0.01 | 5.43E-04 | 0.00 | 9.47E-01 | - | - | - |
| Phosphate intake (mg/day) | Nutrients | 26853 | -0.02 | 0.01 | 8.80E-02 | 2.07E-01 | 115 | 26173 | -0.03 | 0.01 | 1.50E-03 | **7.47E-03** | 55 | -0.02 | 0.01 | 5.66E-04 | 0.10 | 2.92E-01 | - | - | - |
| High mental demand from job | Psychosocial | 28608 | 0.02 | 0.01 | 2.30E-02 | 7.66E-02 | 81 | 28033 | 0.02 | 0.01 | 1.11E-02 | **4.04E-02** | 76 | 0.02 | 0.01 | 6.67E-04 | 0.00 | 8.36E-01 | - | - | - |
| Job demands to work very fast | Psychosocial | 28837 | -0.02 | 0.01 | 4.11E-02 | 1.15E-01 | 96 | 28244 | -0.02 | 0.01 | 5.95E-03 | **2.35E-02** | 70 | -0.02 | 0.01 | 7.02E-04 | 0.00 | 6.12E-01 | - | - | - |
| Beta-carotene intake (mg/day) | Nutrients | 26853 | -0.03 | 0.01 | 2.59E-03 | **1.27E-02** | 55 | 26173 | -0.02 | 0.01 | 1.02E-01 | 2.30E-01 | 123 | -0.02 | 0.01 | 1.01E-03 | 0.00 | 3.31E-01 | - | - | - |
| Root vegetables, carrot | Food | 26853 | -0.03 | 0.01 | 2.10E-03 | **1.06E-02** | 53 | 26173 | -0.01 | 0.01 | 1.26E-01 | 2.63E-01 | 133 | -0.02 | 0.01 | 1.13E-03 | 0.16 | 2.75E-01 | - | - | - |
| Permanent employment | Psychosocial | 24920 | -0.05 | 0.02 | 2.24E-02 | 7.62E-02 | 79 | 24230 | -0.05 | 0.02 | 2.89E-02 | 8.35E-02 | 96 | -0.05 | 0.01 | 1.58E-03 | 0.00 | 9.46E-01 | - | - | - |
| Meat on bread | Food | 26853 | 0.01 | 0.01 | 1.09E-01 | 2.36E-01 | 122 | 26173 | 0.02 | 0.01 | 4.40E-03 | **1.85E-02** | 66 | 0.02 | 0.01 | 1.64E-03 | 0.00 | 3.80E-01 | - | - | - |
| Tiamin intake (mg/day) | Nutrients | 26853 | -0.02 | 0.01 | 7.94E-02 | 1.91E-01 | 112 | 26173 | -0.02 | 0.01 | 7.32E-03 | **2.70E-02** | 75 | -0.02 | 0.01 | 1.73E-03 | 0.00 | 5.03E-01 | - | - | - |
| Wine | Alcohol | 26853 | -0.03 | 0.01 | 5.39E-03 | **2.47E-02** | 59 | 26173 | -0.01 | 0.01 | 1.10E-01 | 2.41E-01 | 126 | -0.02 | 0.01 | 1.94E-03 | 0.00 | 4.06E-01 | - | - | - |
| Participation in study circles | Social | 10280 | 0.08 | 0.04 | 5.34E-02 | 1.41E-01 | 102 | 9964 | 0.10 | 0.04 | 1.72E-02 | 5.62E-02 | 85 | 0.09 | 0.03 | 2.29E-03 | 0.00 | 7.42E-01 | - | - | - |
| Pizza | Food | 26853 | 0.02 | 0.01 | 3.84E-02 | 1.12E-01 | 93 | 26173 | 0.02 | 0.01 | 2.50E-02 | 7.52E-02 | 92 | 0.02 | 0.01 | 2.30E-03 | 0.00 | 9.00E-01 | - | - | - |
| Marital status: Single vs Divorced/separated | Social | 4826 | -0.11 | 0.05 | 2.26E-02 | 7.62E-02 | 80 | 4779 | -0.10 | 0.05 | 4.48E-02 | 1.19E-01 | 104 | -0.10 | 0.03 | 2.41E-03 | 0.00 | 8.79E-01 | - | - | - |
| Everyday exercise satisfaction | Physical activity | 18342 | -0.01 | 0.01 | 2.35E-01 | 4.03E-01 | 157 | 17821 | -0.03 | 0.01 | 2.29E-03 | **1.02E-02** | 62 | -0.02 | 0.01 | 2.83E-03 | 0.45 | 1.79E-01 | - | - | - |
| Physical limitation that limited your ability to perform certain work tasks or other activities during the last four weeks | General health | 13786 | 0.06 | 0.04 | 1.31E-01 | 2.67E-01 | 132 | 13594 | 0.11 | 0.04 | 6.91E-03 | **2.62E-02** | 73 | 0.08 | 0.03 | 2.91E-03 | 0.00 | 3.86E-01 | - | - | - |
| For how much of the time during the last four weeks have you felt full of energy? | General health | 13778 | -0.03 | 0.01 | 6.74E-02 | 1.69E-01 | 108 | 13605 | -0.03 | 0.01 | 1.83E-02 | 5.82E-02 | 87 | -0.03 | 0.01 | 3.05E-03 | 0.00 | 7.00E-01 | - | - | - |
| Self rate of overall health compared to a year ago | General health | 13729 | -0.02 | 0.01 | 9.89E-02 | 2.26E-01 | 118 | 13545 | -0.03 | 0.01 | 1.34E-02 | **4.59E-02** | 80 | -0.03 | 0.01 | 3.56E-03 | 0.00 | 5.46E-01 | - | - | - |
| Satisfaction with accomodation | Psychosocial | 23574 | 0.03 | 0.01 | 7.18E-03 | **3.18E-02** | 61 | 23150 | 0.01 | 0.01 | 1.61E-01 | 3.20E-01 | 138 | 0.02 | 0.01 | 3.79E-03 | 0.00 | 3.68E-01 | - | - | - |
| Parents or siblings had a cerebral hemorrhage/thrombosis or cardiac infarction before the age of 60 | General health | 28732 | 0.05 | 0.02 | 2.23E-02 | 7.62E-02 | 78 | 28136 | 0.04 | 0.02 | 7.35E-02 | 1.77E-01 | 115 | 0.05 | 0.02 | 3.94E-03 | 0.00 | 7.32E-01 | - | - | - |

| | | | | | | | | | | | | | | | | | | | | | |
|---|---|---|---|---|---|---|---|---|---|---|---|---|---|---|---|---|---|---|---|---|---|
| Worsen in health in the future | General health | 13738 | 0.03 | 0.01 | 2.81E-02 | 8.88E-02 | 85 | 13570 | 0.03 | 0.01 | 6.16E-02 | 1.54E-01 | 110 | 0.03 | 0.01 | 4.02E-03 | 0.00 | 8.30E-01 | - | - | - |
| Long-term sickness | General health | 28459 | 0.06 | 0.02 | 7.47E-03 | **3.25E-02** | 62 | 27785 | 0.03 | 0.02 | 1.79E-01 | 3.39E-01 | 145 | 0.05 | 0.02 | 4.43E-03 | 0.00 | 3.53E-01 | - | - | - |
| Physical limitation that made you not being able to perform certain work tasks or other activities during the last four weeks | General health | 13759 | 0.07 | 0.04 | 6.11E-02 | 1.56E-01 | 106 | 13586 | 0.08 | 0.04 | 3.36E-02 | 9.39E-02 | 99 | 0.08 | 0.03 | 4.69E-03 | 0.00 | 8.53E-01 | - | - | - |
| Frequency of dancing during leisure time | Physical activity | 16432 | -0.01 | 0.01 | 4.39E-01 | 6.31E-01 | 188 | 15912 | -0.03 | 0.01 | 1.29E-03 | **6.60E-03** | 54 | -0.02 | 0.01 | 4.83E-03 | 0.67 | 8.16E-02 | - | - | - |
| White (soft) bread, thin crisp bread | Food | 26853 | 0.01 | 0.01 | 1.42E-01 | 2.81E-01 | 136 | 26173 | 0.02 | 0.01 | 1.22E-02 | **4.35E-02** | 78 | 0.02 | 0.01 | 4.97E-03 | 0.00 | 4.60E-01 | - | - | - |
| Margarine for cooking | Food | 26853 | 0.01 | 0.01 | 2.51E-01 | 4.15E-01 | 163 | 26173 | 0.02 | 0.01 | 6.04E-03 | **2.36E-02** | 71 | 0.02 | 0.01 | 5.94E-03 | 0.23 | 2.56E-01 | - | - | - |
| Stigmasterol intake (mg/day) | Nutrients | 26853 | -0.02 | 0.01 | 4.81E-02 | 1.30E-01 | 100 | 26173 | -0.02 | 0.01 | 5.83E-02 | 1.50E-01 | 107 | -0.02 | 0.01 | 6.21E-03 | 0.00 | 9.58E-01 | - | - | - |
| Feel uneasy or guilty because of your way of drinking | Alcohol | 16580 | -0.05 | 0.03 | 9.35E-02 | 2.16E-01 | 117 | 16213 | -0.07 | 0.03 | 3.28E-02 | 9.27E-02 | 98 | -0.06 | 0.02 | 7.07E-03 | 0.00 | 7.36E-01 | - | - | - |
| Satisfaction with home and family situation | Psychosocial | 23556 | 0.01 | 0.01 | 1.28E-01 | 2.64E-01 | 130 | 23131 | 0.02 | 0.01 | 2.29E-02 | 6.98E-02 | 91 | 0.02 | 0.01 | 7.32E-03 | 0.00 | 5.82E-01 | - | - | - |
| Cheese 28% | Food | 26853 | -0.02 | 0.01 | 3.84E-02 | 1.12E-01 | 92 | 26173 | -0.02 | 0.01 | 8.68E-02 | 1.99E-01 | 121 | -0.02 | 0.01 | 7.45E-03 | 0.00 | 8.09E-01 | - | - | - |
| Breakfast habits: Porridge w/o sandwich for breakfast vs not breakfast at all | Food | 4009 | 0.24 | 0.06 | 1.83E-04 | **1.55E-03** | 32 | 4071 | 0.01 | 0.06 | 9.12E-01 | 9.49E-01 | 266 | 0.11 | 0.04 | 7.88E-03 | 0.86 | 7.80E-03 | - | - | - |
| Pentadecanoic acid intake (g/day) | Nutrients | 26853 | -0.02 | 0.01 | 3.04E-02 | 9.33E-02 | 88 | 26173 | -0.01 | 0.01 | 1.15E-01 | 2.45E-01 | 128 | -0.02 | 0.01 | 8.16E-03 | 0.00 | 6.76E-01 | - | - | - |
| Heptadecanoic acid intake (g/day) | Nutrients | 26853 | -0.02 | 0.01 | 3.04E-02 | 9.33E-02 | 87 | 26173 | -0.01 | 0.01 | 1.15E-01 | 2.45E-01 | 129 | -0.02 | 0.01 | 8.16E-03 | 0.00 | 6.76E-01 | - | - | - |
| Berries (fresh or frozen) | Food | 26853 | -0.02 | 0.01 | 5.49E-02 | 1.44E-01 | 103 | 26173 | -0.01 | 0.01 | 1.08E-01 | 2.41E-01 | 124 | -0.02 | 0.01 | 1.26E-02 | 0.00 | 8.23E-01 | - | - | - |
| Possibility to leave your work for a while to speak with a colleague | Psychosocial | 28553 | 0.02 | 0.01 | 5.67E-03 | **2.55E-02** | 60 | 27968 | 0.01 | 0.01 | 5.16E-01 | 6.50E-01 | 220 | 0.02 | 0.01 | 1.51E-02 | 0.54 | 1.40E-01 | - | - | - |
| Chips, popcorn, salted nuts | Food | 26853 | -0.01 | 0.01 | 4.70E-01 | 6.57E-01 | 193 | 26173 | -0.02 | 0.01 | 6.71E-03 | **2.58E-02** | 72 | -0.02 | 0.01 | 1.53E-02 | 0.50 | 1.58E-01 | - | - | - |
| Participation in other association | Social | 10280 | 0.08 | 0.03 | 9.14E-03 | **3.86E-02** | 64 | 9964 | 0.02 | 0.03 | 4.38E-01 | 5.97E-01 | 203 | 0.05 | 0.02 | 1.65E-02 | 0.40 | 1.97E-01 | - | - | - |
| Contradictory demands in job | Psychosocial | 28680 | -0.01 | 0.01 | 2.04E-01 | 3.60E-01 | 153 | 28037 | -0.02 | 0.01 | 3.51E-02 | 9.64E-02 | 101 | -0.01 | 0.01 | 1.71E-02 | 0.00 | 5.44E-01 | - | - | - |
| Meat stew | Food | 26853 | 0.00 | 0.01 | 5.80E-01 | 7.31E-01 | 214 | 26173 | -0.02 | 0.01 | 4.70E-03 | **1.94E-02** | 67 | -0.01 | 0.01 | 1.73E-02 | 0.62 | 1.05E-01 | - | - | - |
| Repetitive job | Psychosocial | 28799 | 0.02 | 0.01 | 7.80E-03 | **3.34E-02** | 63 | 28189 | 0.01 | 0.01 | 4.98E-01 | 6.41E-01 | 213 | 0.01 | 0.01 | 1.79E-02 | 0.48 | 1.64E-01 | - | - | - |
| Disaccharides intake (g/day) | Nutrients | 26853 | -0.02 | 0.01 | 1.09E-01 | 2.36E-01 | 123 | 26173 | -0.02 | 0.01 | 8.45E-02 | 1.97E-01 | 119 | -0.02 | 0.01 | 1.87E-02 | 0.00 | 9.19E-01 | - | - | - |
| Average portion size of potatoes/rice/pasta based on photographic illustration of four sizes (smallest to largest) | Food | 26853 | -0.01 | 0.01 | 2.22E-01 | 3.84E-01 | 156 | 26173 | -0.02 | 0.01 | 3.68E-02 | 9.99E-02 | 102 | -0.02 | 0.01 | 1.94E-02 | 0.00 | 5.32E-01 | - | - | - |
| Boiled or baked potato | Food | 26853 | 0.02 | 0.01 | 5.78E-02 | 1.49E-01 | 104 | 26173 | 0.01 | 0.01 | 1.65E-01 | 3.20E-01 | 142 | 0.01 | 0.01 | 2.01E-02 | 0.00 | 7.23E-01 | - | - | - |
| How much has the pain during the last four weeks disturbed your normal work? | General health | 13767 | 0.02 | 0.01 | 7.89E-02 | 1.91E-01 | 111 | 13591 | 0.02 | 0.01 | 1.30E-01 | 2.68E-01 | 134 | 0.02 | 0.01 | 2.06E-02 | 0.00 | 8.71E-01 | - | - | - |
| Frequency of engaging in clubs, associations or study circles | Social | 18362 | -0.01 | 0.01 | 3.59E-01 | 5.48E-01 | 177 | 17941 | -0.02 | 0.01 | 1.92E-02 | 6.06E-02 | 88 | -0.02 | 0.01 | 2.14E-02 | 0.02 | 3.12E-01 | - | - | - |
| Pain during the last four weeks | General health | 13805 | 0.03 | 0.01 | 3.43E-02 | 1.03E-01 | 90 | 13614 | 0.02 | 0.01 | 2.68E-01 | 4.39E-01 | 169 | 0.02 | 0.01 | 2.23E-02 | 0.00 | 4.81E-01 | - | - | - |
| Appetite status | Psychosocial | 23555 | -0.01 | 0.01 | 1.64E-01 | 3.05E-01 | 145 | 23124 | -0.02 | 0.01 | 7.09E-02 | 1.74E-01 | 113 | -0.02 | 0.01 | 2.38E-02 | 0.00 | 7.62E-01 | - | - | - |
| Hearing status | General health | 23574 | 0.02 | 0.01 | 4.13E-02 | 1.15E-01 | 97 | 23138 | 0.01 | 0.01 | 2.50E-01 | 4.17E-01 | 166 | 0.02 | 0.01 | 2.40E-02 | 0.00 | 5.31E-01 | - | - | - |
| Salad dressing with oil | Food | 26853 | -0.01 | 0.01 | 1.14E-01 | 2.45E-01 | 126 | 26173 | -0.01 | 0.01 | 1.09E-01 | 2.41E-01 | 125 | -0.01 | 0.01 | 2.45E-02 | 0.00 | 9.94E-01 | - | - | - |
| Breakfast habits: Gruel w/o sandwich for breakfast vs not breakfast at all | Food | 1360 | 0.19 | 0.10 | 5.31E-02 | 1.41E-01 | 101 | 1434 | 0.12 | 0.10 | 2.30E-01 | 3.88E-01 | 163 | 0.15 | 0.07 | 2.60E-02 | 0.00 | 6.19E-01 | - | - | - |
| Marital status: Single vs Widow/widower | Social | 3050 | -0.15 | 0.10 | 1.28E-01 | 2.64E-01 | 131 | 2954 | -0.16 | 0.10 | 1.10E-01 | 2.41E-01 | 127 | -0.16 | 0.07 | 2.75E-02 | 0.00 | 9.21E-01 | - | - | - |
| Coffee rolls/buns, rusk | Food | 26853 | 0.02 | 0.01 | 8.34E-02 | 1.98E-01 | 114 | 26173 | 0.01 | 0.01 | 1.81E-01 | 3.39E-01 | 146 | 0.01 | 0.01 | 2.99E-02 | 0.00 | 7.86E-01 | - | - | - |
| Do you feel important and appreciated in your home? | Psychosocial | 23407 | 0.02 | 0.01 | 1.04E-01 | 2.36E-01 | 119 | 22960 | 0.01 | 0.01 | 1.64E-01 | 3.20E-01 | 141 | 0.01 | 0.01 | 3.29E-02 | 0.00 | 8.78E-01 | - | - | - |
| Satisfaction with work situation | Psychosocial | 23275 | 0.02 | 0.01 | 1.16E-01 | 2.46E-01 | 127 | 22836 | 0.01 | 0.01 | 1.62E-01 | 3.20E-01 | 139 | 0.01 | 0.01 | 3.57E-02 | 0.00 | 9.12E-01 | - | - | - |
| Physical limitation that reduced the normal time spent at work or in other activities during the last four weeks | General health | 13828 | 0.06 | 0.04 | 1.44E-01 | 2.85E-01 | 137 | 13638 | 0.06 | 0.04 | 1.35E-01 | 2.77E-01 | 135 | 0.06 | 0.03 | 3.67E-02 | 0.00 | 9.82E-01 | - | - | - |
| Vitamin B3 intake (mg/day) | Nutrients | 26853 | -0.01 | 0.01 | 2.36E-01 | 4.03E-01 | 158 | 26173 | -0.02 | 0.01 | 7.69E-02 | 1.84E-01 | 116 | -0.01 | 0.01 | 3.68E-02 | 0.00 | 6.75E-01 | - | - | - |
| Milk, sour milk (3%) | Beverage | 26853 | -0.01 | 0.01 | 1.09E-01 | 2.36E-01 | 124 | 26173 | -0.01 | 0.01 | 1.83E-01 | 3.39E-01 | 150 | -0.01 | 0.01 | 3.79E-02 | 0.00 | 8.68E-01 | - | - | - |
| Travel to work: Irregular travel mode to work vs passive travel to work | Physical activity | 18883 | -0.02 | 0.03 | 4.63E-01 | 6.51E-01 | 192 | 18581 | -0.07 | 0.03 | 3.21E-02 | 9.15E-02 | 97 | -0.05 | 0.02 | 4.09E-02 | 0.00 | 3.28E-01 | - | - | - |
| Plant based protein intake (g/day) | Nutrients | 26853 | 0.02 | 0.01 | 6.93E-02 | 1.72E-01 | 109 | 26173 | 0.01 | 0.01 | 2.85E-01 | 4.61E-01 | 171 | 0.01 | 0.01 | 4.11E-02 | 0.00 | 6.02E-01 | - | - | - |
| Beta-sitosterol intake (mg/day) | Nutrients | 26853 | -0.02 | 0.01 | 4.25E-02 | 1.17E-01 | 98 | 26173 | -0.01 | 0.01 | 4.02E-01 | 5.82E-01 | 190 | -0.01 | 0.01 | 4.25E-02 | 0.00 | 4.02E-01 | - | - | - |
| Memory status | Psychosocial | 23515 | 0.02 | 0.01 | 1.69E-02 | 6.08E-02 | 75 | 23067 | 0.00 | 0.01 | 6.52E-01 | 7.55E-01 | 239 | 0.01 | 0.01 | 4.44E-02 | 0.47 | 1.72E-01 | - | - | - |
| Cream, creme fraiche, sour cream | Food | 26853 | -0.01 | 0.01 | 1.58E-01 | 2.99E-01 | 142 | 26173 | -0.01 | 0.01 | 1.63E-01 | 3.20E-01 | 140 | -0.01 | 0.01 | 4.72E-02 | 0.00 | 9.74E-01 | - | - | - |
| Physical limitation that made you do less than you wanted during the last four weeks | General health | 13812 | 0.04 | 0.03 | 2.61E-01 | 4.28E-01 | 165 | 13639 | 0.06 | 0.03 | 9.48E-02 | 2.15E-01 | 122 | 0.05 | 0.02 | 4.84E-02 | 0.00 | 6.88E-01 | - | - | - |
| Lean fish (e.g. perch, bass, cod) | Food | 26853 | 0.02 | 0.01 | 1.62E-02 | 5.93E-02 | 74 | 26173 | 0.00 | 0.01 | 7.23E-01 | 8.14E-01 | 246 | 0.01 | 0.01 | 4.85E-02 | 0.50 | 1.56E-01 | - | - | - |
| Palmitic acid intake (g/day) | Nutrients | 26853 | -0.01 | 0.01 | 1.38E-01 | 2.79E-01 | 134 | 26173 | -0.01 | 0.01 | 2.02E-01 | 3.52E-01 | 159 | -0.01 | 0.01 | 5.13E-02 | 0.00 | 8.80E-01 | - | - | - |
| Low fat milk (0,5%) | Beverage | 26853 | 0.02 | 0.01 | 5.79E-02 | 1.49E-01 | 105 | 26173 | 0.01 | 0.01 | 4.24E-01 | 5.97E-01 | 194 | 0.01 | 0.01 | 5.66E-02 | 0.00 | 4.39E-01 | - | - | - |
| Ice cream | Food | 26853 | -0.01 | 0.01 | 1.55E-01 | 2.97E-01 | 141 | 26173 | -0.01 | 0.01 | 2.86E-01 | 4.61E-01 | 172 | -0.01 | 0.01 | 7.85E-02 | 0.00 | 7.99E-01 | - | - | - |
| Vitamin B12 intake (ug/day) | Nutrients | 26853 | 0.02 | 0.01 | 1.39E-02 | 5.16E-02 | 73 | 26173 | 0.00 | 0.01 | 9.88E-01 | 9.92E-01 | 275 | 0.01 | 0.01 | 8.34E-02 | 0.67 | 8.08E-02 | - | - | - |
| Vitamin A intake (mg/day) | Nutrients | 26853 | 0.01 | 0.01 | 5.18E-01 | 6.97E-01 | 200 | 26173 | 0.02 | 0.01 | 7.26E-02 | 1.76E-01 | 114 | 0.01 | 0.01 | 8.43E-02 | 0.00 | 4.16E-01 | - | - | - |
| Tomato, cucumber | Food | 26853 | -0.01 | 0.01 | 2.76E-01 | 4.47E-01 | 167 | 26173 | -0.01 | 0.01 | 1.82E-01 | 3.39E-01 | 148 | -0.01 | 0.01 | 8.68E-02 | 0.00 | 8.55E-01 | - | - | - |
| For how much of the time during the last four weeks has your physical health or your emotional problems limited your ability to interact with others? | General health | 13736 | 0.01 | 0.01 | 6.10E-01 | 7.59E-01 | 217 | 13549 | 0.03 | 0.01 | 5.86E-02 | 1.50E-01 | 108 | 0.02 | 0.01 | 8.96E-02 | 0.00 | 3.24E-01 | - | - | - |
| Educational level | Psychosocial | 29334 | -0.01 | 0.01 | 1.41E-01 | 2.81E-01 | 135 | 28653 | -0.01 | 0.01 | 3.76E-01 | 5.64E-01 | 184 | -0.01 | 0.01 | 9.53E-02 | 0.00 | 6.83E-01 | - | - | - |
| Formic acid intaje (g/day) | Nutrients | 26853 | -0.01 | 0.01 | 1.37E-01 | 2.79E-01 | 133 | 26173 | -0.01 | 0.01 | 4.27E-01 | 5.97E-01 | 197 | -0.01 | 0.01 | 1.07E-01 | 0.00 | 6.24E-01 | - | - | - |
| Cohabitation: Live alone vs Adult and children | Social | 16938 | -0.05 | 0.03 | 8.29E-02 | 1.98E-01 | 113 | 16529 | -0.02 | 0.03 | 6.03E-01 | 7.19E-01 | 232 | -0.04 | 0.02 | 1.10E-01 | 0.00 | 3.95E-01 | - | - | - |
| Enough time for job assignments | Psychosocial | 28722 | 0.00 | 0.01 | 8.90E-01 | 9.50E-01 | 253 | 28105 | 0.02 | 0.01 | 3.39E-02 | 9.39E-02 | 100 | 0.01 | 0.01 | 1.12E-01 | 0.50 | 1.58E-01 | - | - | - |
| Marital status: Single vs Married/partner | Social | 26750 | -0.05 | 0.03 | 9.14E-02 | 2.13E-01 | 116 | 26125 | -0.02 | 0.03 | 5.87E-01 | 7.07E-01 | 230 | -0.04 | 0.02 | 1.14E-01 | 0.00 | 4.19E-01 | - | - | - |
| Corn flakes | Food | 26853 | 0.01 | 0.01 | 5.19E-01 | 6.97E-01 | 201 | 26173 | 0.01 | 0.01 | 1.15E-01 | 2.45E-01 | 130 | 0.01 | 0.01 | 1.17E-01 | 0.00 | 5.04E-01 | - | - | - |
| Sleep status | Sleep | 23572 | -0.01 | 0.01 | 4.14E-01 | 6.17E-01 | 181 | 23149 | -0.01 | 0.01 | 1.87E-01 | 3.42E-01 | 152 | -0.01 | 0.01 | 1.31E-01 | 0.00 | 7.20E-01 | - | - | - |
| Pinoresinol intake (ug/day) | Nutrients | 26853 | -0.01 | 0.01 | 1.85E-01 | 3.34E-01 | 149 | 26173 | -0.01 | 0.01 | 4.25E-01 | 5.97E-01 | 195 | -0.01 | 0.01 | 1.33E-01 | 0.00 | 7.10E-01 | - | - | - |
| People to ask for help apart from the ones at home | Social | 29192 | -0.04 | 0.03 | 1.87E-01 | 3.36E-01 | 150 | 28541 | -0.02 | 0.03 | 4.26E-01 | 5.97E-01 | 196 | -0.03 | 0.02 | 1.34E-01 | 0.00 | 7.10E-01 | - | - | - |
| Breakfast habits: Coffee/tea and wheat buns or rusk for breakfast vs not breakfast at all | Food | 1514 | 0.11 | 0.08 | 2.02E-01 | 3.60E-01 | 151 | 1501 | 0.07 | 0.09 | 4.29E-01 | 5.97E-01 | 199 | 0.09 | 0.06 | 1.41E-01 | 0.00 | 7.55E-01 | - | - | - |
| Fatty fish (e.g. herring, whitefish, salmon) | Food | 26853 | 0.00 | 0.01 | 6.84E-01 | 8.21E-01 | 225 | 26173 | -0.02 | 0.01 | 1.31E-02 | **4.59E-02** | 79 | -0.01 | 0.01 | 1.42E-01 | 0.76 | 4.12E-02 | - | - | - |
| Fiber cereals | Food | 26853 | -0.01 | 0.01 | 4.34E-01 | 6.31E-01 | 185 | 26173 | -0.01 | 0.01 | 1.97E-01 | 3.49E-01 | 156 | -0.01 | 0.01 | 1.43E-01 | 0.00 | 7.13E-01 | - | - | - |
| Pancake, waffle, Swedish dumpling | Food | 26853 | -0.01 | 0.01 | 2.41E-01 | 4.10E-01 | 159 | 26173 | -0.01 | 0.01 | 3.71E-01 | 5.62E-01 | 183 | -0.01 | 0.01 | 1.44E-01 | 0.00 | 8.42E-01 | - | - | - |
| Brown beans, pea soup | Food | 26853 | 0.00 | 0.01 | 9.73E-01 | 9.80E-01 | 268 | 26173 | -0.02 | 0.01 | 4.37E-02 | 1.18E-01 | 103 | -0.01 | 0.01 | 1.49E-01 | 0.50 | 1.59E-01 | - | - | - |
| Sugar, honey, marmelade, jam | Food | 26853 | -0.02 | 0.01 | 3.92E-02 | 1.12E-01 | 94 | 26173 | 0.00 | 0.01 | 9.62E-01 | 9.76E-01 | 273 | -0.01 | 0.01 | 1.53E-01 | 0.55 | 1.37E-01 | - | - | - |
| Would you say that the number of people that you meet in your everyday life is enough or would you like to meet more or fewer people? | Social | 29274 | -0.02 | 0.01 | 1.75E-02 | 6.23E-02 | 76 | 28604 | 0.00 | 0.01 | 7.10E-01 | 8.02E-01 | 245 | -0.01 | 0.01 | 1.54E-01 | 0.73 | 5.27E-02 | - | - | - |
| Rice | Food | 26853 | 0.01 | 0.01 | 4.22E-01 | 6.23E-01 | 183 | 26173 | 0.01 | 0.01 | 2.30E-01 | 3.88E-01 | 164 | 0.01 | 0.01 | 1.57E-01 | 0.00 | 7.76E-01 | - | - | - |
| Travel to work: Walk to work vs passive travel to work | Physical activity | 18878 | -0.04 | 0.03 | 1.84E-01 | 3.34E-01 | 148 | 18619 | -0.02 | 0.03 | 5.01E-01 | 6.41E-01 | 216 | -0.03 | 0.02 | 1.58E-01 | 0.00 | 6.39E-01 | - | - | - |
| Bregott on bread | Food | 26853 | -0.01 | 0.01 | 1.64E-01 | 3.05E-01 | 144 | 26173 | 0.00 | 0.01 | 5.80E-01 | 7.02E-01 | 229 | -0.01 | 0.01 | 1.68E-01 | 0.00 | 5.55E-01 | - | - | - |
| Margarine on bread | Food | 26853 | 0.01 | 0.01 | 5.11E-01 | 6.97E-01 | 197 | 26173 | 0.01 | 0.01 | 1.99E-01 | 3.49E-01 | 157 | 0.01 | 0.01 | 1.70E-01 | 0.00 | 6.59E-01 | - | - | - |
| Pasta | Food | 26853 | -0.01 | 0.01 | 4.33E-01 | 6.31E-01 | 184 | 26173 | -0.01 | 0.01 | 2.50E-01 | 4.17E-01 | 165 | -0.01 | 0.01 | 1.71E-01 | 0.00 | 7.92E-01 | - | - | - |
| For how much of the time during the last four weeks have you felt tired? | General health | 13816 | 0.01 | 0.01 | 5.85E-01 | 7.31E-01 | 216 | 13619 | 0.02 | 0.01 | 1.65E-01 | 3.20E-01 | 143 | 0.01 | 0.01 | 1.72E-01 | 0.00 | 5.48E-01 | - | - | - |
| Breakfast habits: Only coffee/tea for breakfast vs not breakfast at all | Food | 1817 | 0.00 | 0.09 | 9.58E-01 | 9.73E-01 | 264 | 1871 | -0.17 | 0.09 | 6.06E-02 | 1.54E-01 | 109 | -0.08 | 0.06 | 1.81E-01 | 0.43 | 1.86E-01 | - | - | - |
| Light but partly physically active work | Physical activity | 28425 | 0.02 | 0.02 | 2.53E-01 | 4.16E-01 | 164 | 27780 | 0.01 | 0.02 | 4.99E-01 | 6.41E-01 | 215 | 0.02 | 0.02 | 1.97E-01 | 0.00 | 7.51E-01 | - | - | - |
| Sour milk, yoghurt (3% fat) | Food | 26853 | -0.01 | 0.01 | 3.32E-01 | 5.24E-01 | 171 | 26173 | -0.01 | 0.01 | 3.94E-01 | 5.80E-01 | 188 | -0.01 | 0.01 | 1.97E-01 | 0.00 | 9.39E-01 | - | - | - |
| Trans fat intake (g/day) | Nutrients | 26853 | 0.00 | 0.01 | 9.32E-01 | 9.72E-01 | 259 | 26173 | -0.02 | 0.01 | 8.52E-02 | 1.97E-01 | 120 | -0.01 | 0.01 | 2.00E-01 | 0.25 | 2.48E-01 | - | - | - |
| Equol intake (ug/day) | Nutrients | 26853 | 0.01 | 0.01 | 1.09E-01 | 2.36E-01 | 125 | 26173 | 0.00 | 0.01 | 8.46E-01 | 8.91E-01 | 263 | 0.01 | 0.01 | 2.04E-01 | 0.00 | 3.20E-01 | - | - | - |
| Iodine intake (ug/day) | Nutrients | 26853 | 0.00 | 0.01 | 8.36E-01 | 9.35E-01 | 241 | 26173 | -0.02 | 0.01 | 5.62E-02 | 1.47E-01 | 106 | -0.01 | 0.01 | 2.31E-01 | 0.56 | 1.33E-01 | - | - | - |
| Last time a colleague visited you at home | Psychosocial | 28435 | 0.00 | 0.01 | 8.73E-01 | 9.43E-01 | 246 | 27859 | -0.01 | 0.01 | 1.23E-01 | 2.58E-01 | 132 | -0.01 | 0.01 | 2.32E-01 | 0.00 | 3.24E-01 | - | - | - |
| Number of friends that can come to your home at any time and feel at home | Social | 29173 | 0.01 | 0.01 | 5.31E-01 | 7.00E-01 | 205 | 28532 | -0.02 | 0.01 | 1.97E-02 | 6.14E-02 | 89 | -0.01 | 0.01 | 2.33E-01 | 0.77 | 3.58E-02 | - | - | - |
| Frequency of walking during leisure time | Physical activity | 28242 | 0.01 | 0.01 | 4.38E-01 | 6.31E-01 | 186 | 27612 | -0.02 | 0.01 | 1.34E-02 | **4.59E-02** | 81 | -0.01 | 0.01 | 2.36E-01 | 0.81 | 2.12E-02 | - | - | - |
| Sum of phytosterols intake (mg/day) | Nutrients | 26853 | -0.01 | 0.01 | 1.18E-01 | 2.48E-01 | 128 | 26173 | 0.00 | 0.01 | 9.32E-01 | 9.56E-01 | 270 | -0.01 | 0.01 | 2.42E-01 | 0.08 | 2.97E-01 | - | - | - |
| Monounsaturated fat intake (g/day) | Nutrients | 26853 | -0.01 | 0.01 | 5.09E-01 | 6.97E-01 | 196 | 26173 | -0.01 | 0.01 | 3.23E-01 | 5.00E-01 | 179 | -0.01 | 0.01 | 2.44E-01 | 0.00 | 8.14E-01 | - | - | - |
| Vitamin D intake (ug/day) | Nutrients | 26853 | 0.01 | 0.01 | 1.71E-01 | 3.15E-01 | 147 | 26173 | 0.00 | 0.01 | 7.93E-01 | 8.54E-01 | 257 | 0.01 | 0.01 | 2.48E-01 | 0.00 | 4.37E-01 | - | - | - |
| Carbohydrates intake (g/day) | Nutrients | 26853 | 0.01 | 0.01 | 4.95E-01 | 6.85E-01 | 195 | 26173 | 0.01 | 0.01 | 3.45E-01 | 5.28E-01 | 181 | 0.01 | 0.01 | 2.50E-01 | 0.00 | 8.52E-01 | - | - | - |
| Confidence status | Psychosocial | 23531 | 0.00 | 0.01 | 6.80E-01 | 8.20E-01 | 224 | 23094 | -0.01 | 0.01 | 2.29E-01 | 3.88E-01 | 162 | -0.01 | 0.01 | 2.54E-01 | 0.00 | 5.75E-01 | - | - | - |
| Control over own work assignment | Psychosocial | 28850 | 0.00 | 0.01 | 8.14E-01 | 9.23E-01 | 238 | 28237 | 0.01 | 0.01 | 1.74E-01 | 3.35E-01 | 144 | 0.01 | 0.01 | 2.61E-01 | 0.00 | 4.24E-01 | - | - | - |
| Calcium intake (mg/day) | Nutrients | 26853 | 0.00 | 0.01 | 7.88E-01 | 9.14E-01 | 233 | 26173 | -0.01 | 0.01 | 2.09E-01 | 3.62E-01 | 160 | -0.01 | 0.01 | 2.83E-01 | 0.00 | 4.81E-01 | - | - | - |
| Campesterol intake (mg/day) | Nutrients | 26853 | 0.00 | 0.01 | 6.94E-01 | 8.28E-01 | 226 | 26173 | 0.02 | 0.01 | 6.38E-02 | 1.58E-01 | 112 | 0.01 | 0.01 | 3.03E-01 | 0.60 | 1.12E-01 | - | - | - |
| For how much of the time during the last four weeks have you felt calm and serene? | General health | 13794 | -0.01 | 0.01 | 4.88E-01 | 6.79E-01 | 194 | 13627 | -0.01 | 0.01 | 4.46E-01 | 5.99E-01 | 206 | -0.01 | 0.01 | 3.03E-01 | 0.00 | 9.59E-01 | - | - | - |
| Frequncy of social contacts with colleagues during leisure time | Psychosocial | 28026 | 0.00 | 0.01 | 8.79E-01 | 9.43E-01 | 251 | 27437 | -0.01 | 0.01 | 1.92E-01 | 3.47E-01 | 153 | -0.01 | 0.01 | 3.06E-01 | 0.00 | 4.12E-01 | - | - | - |
| Sweets | Food | 26853 | -0.01 | 0.01 | 5.17E-01 | 6.97E-01 | 199 | 26173 | -0.01 | 0.01 | 4.27E-01 | 5.97E-01 | 198 | -0.01 | 0.01 | 3.08E-01 | 0.00 | 9.20E-01 | - | - | - |
| Self-employed | Psychosocial | 24920 | -0.07 | 0.03 | 3.37E-02 | 1.02E-01 | 89 | 24230 | 0.02 | 0.03 | 4.88E-01 | 6.41E-01 | 210 | -0.02 | 0.02 | 3.17E-01 | 0.75 | 4.57E-02 | - | - | - |
| Cholesterol intake (g/day) | Nutrients | 26853 | 0.00 | 0.01 | 9.55E-01 | 9.73E-01 | 263 | 26173 | -0.01 | 0.01 | 1.83E-01 | 3.39E-01 | 149 | -0.01 | 0.01 | 3.26E-01 | 0.00 | 3.68E-01 | - | - | - |

| | | | | | | | | | | | | | | | | | | | | | |
|---|---|---|---|---|---|---|---|---|---|---|---|---|---|---|---|---|---|---|---|---|---|
| Control over planning and execution of the workday | Psychosocial | 28870 | 0.00 | 0.01 | 9.29E-01 | 9.72E-01 | 258 | 28237 | 0.01 | 0.01 | 1.99E-01 | 3.49E-01 | 158 | 0.01 | 0.01 | 3.33E-01 | 0.00 | 3.96E-01 | - | - | - |
| Low fat margarine on bread | Food | 26853 | 0.01 | 0.01 | 5.54E-01 | 7.10E-01 | 210 | 26173 | 0.01 | 0.01 | 4.38E-01 | 5.97E-01 | 202 | 0.01 | 0.01 | 3.34E-01 | 0.00 | 8.95E-01 | - | - | - |
| Extent to what your physical and emotional health disrupted your usual social life during the last four weeks | General health | 13816 | 0.00 | 0.01 | 9.59E-01 | 9.73E-01 | 265 | 13637 | 0.02 | 0.01 | 1.94E-01 | 3.47E-01 | 155 | 0.01 | 0.01 | 3.41E-01 | 0.00 | 3.75E-01 | - | - | - |
| Sum of all lignans intake (ug/day) | Nutrients | 26853 | -0.01 | 0.01 | 3.69E-01 | 5.53E-01 | 179 | 26173 | 0.00 | 0.01 | 6.57E-01 | 7.55E-01 | 241 | -0.01 | 0.01 | 3.42E-01 | 0.00 | 7.50E-01 | - | - | - |
| Arachidonic acid (ARA) intake (g/day) | Nutrients | 26852 | 0.01 | 0.01 | 1.48E-01 | 2.89E-01 | 138 | 26172 | 0.00 | 0.01 | 8.93E-01 | 9.33E-01 | 265 | 0.01 | 0.01 | 3.52E-01 | 0.20 | 2.64E-01 | - | - | - |
| Selenium intake (ug/day) | Nutrients | 26853 | 0.01 | 0.01 | 5.31E-01 | 7.00E-01 | 204 | 26173 | -0.02 | 0.01 | 5.33E-02 | 1.41E-01 | 105 | -0.01 | 0.01 | 3.62E-01 | 0.70 | 6.94E-02 | - | - | - |
| Salty fish | Food | 26853 | 0.02 | 0.01 | 4.66E-02 | 1.27E-01 | 99 | 26173 | -0.01 | 0.01 | 4.35E-01 | 5.97E-01 | 200 | 0.01 | 0.01 | 3.64E-01 | 0.73 | 5.29E-02 | - | - | - |
| Saturated fat intake (g/day) | Nutrients | 26853 | -0.01 | 0.01 | 4.38E-01 | 6.31E-01 | 187 | 26173 | 0.00 | 0.01 | 6.16E-01 | 7.30E-01 | 234 | -0.01 | 0.01 | 3.67E-01 | 0.00 | 8.46E-01 | - | - | - |
| Satisfaction with leisure time | Psychosocial | 23525 | 0.00 | 0.01 | 6.64E-01 | 8.09E-01 | 221 | 23092 | 0.01 | 0.01 | 4.02E-01 | 5.82E-01 | 191 | 0.01 | 0.01 | 3.69E-01 | 0.00 | 7.72E-01 | - | - | - |
| Do you feel important and appreciated outside your home? | Psychosocial | 23570 | 0.00 | 0.01 | 7.69E-01 | 8.95E-01 | 232 | 23142 | 0.01 | 0.01 | 3.55E-01 | 5.41E-01 | 182 | 0.01 | 0.01 | 3.90E-01 | 0.00 | 6.54E-01 | - | - | - |
| Sodas, soft drinks, juice | Beverage | 26853 | 0.00 | 0.01 | 8.75E-01 | 9.43E-01 | 248 | 26173 | 0.01 | 0.01 | 2.97E-01 | 4.72E-01 | 174 | 0.01 | 0.01 | 4.00E-01 | 0.00 | 5.24E-01 | - | - | - |
| Campestanol intake (mg/day) | Nutrients | 26853 | 0.00 | 0.01 | 6.96E-01 | 8.28E-01 | 227 | 26173 | 0.01 | 0.01 | 4.36E-01 | 5.97E-01 | 201 | 0.01 | 0.01 | 4.09E-01 | 0.00 | 7.80E-01 | - | - | - |
| Emotional problems that made you do less than you wanted during the last four weeks | General health | 13830 | -0.09 | 0.04 | 2.83E-02 | 8.88E-02 | 86 | 13638 | 0.04 | 0.04 | 2.88E-01 | 4.61E-01 | 173 | -0.02 | 0.03 | 4.17E-01 | 0.81 | 2.11E-02 | - | - | - |
| Cohabitation: Live alone vs Other/others | Social | 3320 | 0.12 | 0.09 | 1.71E-01 | 3.15E-01 | 146 | 3287 | -0.03 | 0.10 | 7.60E-01 | 8.42E-01 | 249 | 0.05 | 0.07 | 4.21E-01 | 0.24 | 2.50E-01 | - | - | - |
| Polyunsaturated fat intake (g/day) | Nutrients | 26853 | 0.00 | 0.01 | 8.09E-01 | 9.23E-01 | 236 | 26173 | 0.01 | 0.01 | 3.76E-01 | 5.64E-01 | 185 | 0.00 | 0.01 | 4.27E-01 | 0.00 | 6.47E-01 | - | - | - |
| For how much of the time during the last four weeks have you felt gloomy and sad? | General health | 13782 | -0.01 | 0.01 | 5.38E-01 | 7.02E-01 | 206 | 13607 | -0.01 | 0.01 | 6.30E-01 | 7.36E-01 | 237 | -0.01 | 0.01 | 4.37E-01 | 0.00 | 9.28E-01 | - | - | - |
| Medioresinol intake (ug/day) | Nutrients | 26853 | 0.00 | 0.01 | 8.02E-01 | 9.21E-01 | 235 | 26173 | 0.01 | 0.01 | 4.39E-01 | 5.97E-01 | 204 | 0.00 | 0.01 | 4.70E-01 | 0.00 | 7.10E-01 | - | - | - |
| White meat (poultry) | Food | 26853 | -0.01 | 0.01 | 1.08E-01 | 2.36E-01 | 121 | 26173 | 0.01 | 0.01 | 5.28E-01 | 6.59E-01 | 222 | 0.00 | 0.01 | 4.85E-01 | 0.60 | 1.14E-01 | - | - | - |
| Fibre intake (g/day) | Nutrients | 26853 | 0.00 | 0.01 | 7.23E-01 | 8.45E-01 | 231 | 26173 | -0.01 | 0.01 | 5.47E-01 | 6.70E-01 | 226 | 0.00 | 0.01 | 4.99E-01 | 0.00 | 8.58E-01 | - | - | - |
| Eicosapentaenoic acid (EPA) intake (g/day) | Nutrients | 26852 | 0.01 | 0.01 | 1.49E-01 | 2.89E-01 | 139 | 26173 | -0.02 | 0.01 | 1.81E-02 | 5.82E-02 | 86 | 0.00 | 0.01 | 5.23E-01 | 0.86 | 7.03E-03 | - | - | - |
| Close relationship with anyone | Social | 29249 | 0.01 | 0.01 | 1.26E-01 | 2.64E-01 | 129 | 28594 | -0.01 | 0.01 | 5.26E-01 | 6.59E-01 | 221 | 0.00 | 0.01 | 5.24E-01 | 0.57 | 1.26E-01 | - | - | - |
| Mood status | Psychosocial | 23538 | 0.00 | 0.01 | 7.94E-01 | 9.16E-01 | 234 | 23105 | -0.01 | 0.01 | 2.52E-01 | 4.17E-01 | 167 | 0.00 | 0.01 | 5.33E-01 | 0.00 | 3.19E-01 | - | - | - |
| Cookies, pastry | Food | 26853 | -0.01 | 0.01 | 3.47E-01 | 5.41E-01 | 173 | 26173 | 0.00 | 0.01 | 9.29E-01 | 9.56E-01 | 269 | 0.00 | 0.01 | 5.47E-01 | 0.00 | 4.67E-01 | - | - | - |
| Possibility to speak with colleagues during breaks | Psychosocial | 28729 | 0.01 | 0.01 | 2.03E-01 | 3.60E-01 | 152 | 28137 | 0.00 | 0.01 | 6.63E-01 | 7.56E-01 | 243 | 0.00 | 0.01 | 5.49E-01 | 0.31 | 2.29E-01 | - | - | - |
| Vitamin C intake (mg/day) | Nutrients | 26853 | 0.00 | 0.01 | 8.76E-01 | 9.43E-01 | 250 | 26173 | 0.01 | 0.01 | 3.22E-01 | 5.00E-01 | 178 | 0.00 | 0.01 | 5.56E-01 | 0.00 | 4.17E-01 | - | - | - |
| Banana | Food | 26853 | 0.01 | 0.01 | 3.57E-01 | 5.48E-01 | 175 | 26173 | -0.02 | 0.01 | 7.85E-02 | 1.86E-01 | 117 | 0.00 | 0.01 | 5.60E-01 | 0.72 | 5.76E-02 | - | - | - |
| Docosahexaenoic acid (DHA) intake (g/day) | Nutrients | 26853 | 0.01 | 0.01 | 1.59E-01 | 2.99E-01 | 143 | 26173 | -0.02 | 0.01 | 2.67E-02 | 7.87E-02 | 94 | 0.00 | 0.01 | 5.81E-01 | 0.85 | 1.02E-02 | - | - | - |
| Animal based protein intake (g/day) | Nutrients | 26853 | 0.00 | 0.01 | 5.73E-01 | 7.26E-01 | 213 | 26173 | -0.01 | 0.01 | 1.81E-01 | 3.39E-01 | 147 | 0.00 | 0.01 | 5.87E-01 | 0.45 | 1.78E-01 | - | - | - |
| Syringaresinol intake (ug/day) | Nutrients | 26853 | 0.00 | 0.01 | 8.49E-01 | 9.39E-01 | 243 | 26173 | 0.01 | 0.01 | 5.66E-01 | 6.90E-01 | 227 | 0.00 | 0.01 | 5.90E-01 | 0.00 | 7.84E-01 | - | - | - |
| Whole grain intake (g/day) | Food | 26853 | 0.00 | 0.01 | 9.54E-01 | 9.73E-01 | 262 | 26173 | -0.01 | 0.01 | 4.92E-01 | 6.41E-01 | 211 | 0.00 | 0.01 | 6.00E-01 | 0.00 | 6.55E-01 | - | - | - |
| Minced meat dishes | Food | 26853 | 0.00 | 0.01 | 9.06E-01 | 9.59E-01 | 255 | 26173 | -0.01 | 0.01 | 4.06E-01 | 5.82E-01 | 193 | 0.00 | 0.01 | 6.13E-01 | 0.00 | 5.03E-01 | - | - | - |
| Enterolactone intake (ug/day) | Nutrients | 26853 | 0.00 | 0.01 | 9.60E-01 | 9.73E-01 | 266 | 26173 | -0.01 | 0.01 | 5.14E-01 | 6.50E-01 | 219 | 0.00 | 0.01 | 6.21E-01 | 0.00 | 6.68E-01 | - | - | - |
| Sodium intake (mg/day) | Nutrients | 26852 | 0.01 | 0.01 | 2.46E-01 | 4.12E-01 | 161 | 26172 | -0.02 | 0.01 | 6.17E-02 | 1.54E-01 | 111 | 0.00 | 0.01 | 6.21E-01 | 0.78 | 3.20E-02 | - | - | - |
| Fried potatoes, pommes frites | Food | 26853 | 0.01 | 0.01 | 3.56E-01 | 5.48E-01 | 174 | 26173 | 0.00 | 0.01 | 8.26E-01 | 8.76E-01 | 261 | 0.00 | 0.01 | 6.27E-01 | 0.00 | 4.16E-01 | - | - | - |
| Vitamin B2 intake (ug/day) | Nutrients | 26853 | 0.00 | 0.01 | 6.66E-01 | 8.09E-01 | 222 | 26173 | -0.01 | 0.01 | 2.67E-01 | 4.39E-01 | 168 | 0.00 | 0.01 | 6.36E-01 | 0.16 | 2.74E-01 | - | - | - |
| Steak, chop, e.g. | Food | 26853 | 0.00 | 0.01 | 8.51E-01 | 9.39E-01 | 244 | 26173 | 0.00 | 0.01 | 6.28E-01 | 7.36E-01 | 236 | 0.00 | 0.01 | 6.36E-01 | 0.00 | 8.31E-01 | - | - | - |
| Light and physically active work | Physical activity | 28425 | 0.00 | 0.02 | 8.76E-01 | 9.43E-01 | 249 | 27780 | -0.02 | 0.02 | 4.04E-01 | 5.82E-01 | 192 | -0.01 | 0.01 | 6.36E-01 | 0.00 | 4.80E-01 | - | - | - |
| If you exercise, change in exercise habits during the last year | Physical activity | 16204 | -0.01 | 0.01 | 5.22E-01 | 6.98E-01 | 202 | 15786 | 0.00 | 0.01 | 9.92E-01 | 9.92E-01 | 277 | 0.00 | 0.01 | 6.43E-01 | 0.00 | 6.59E-01 | - | - | - |
| Cohabitation: Live alone vs Only one adult (spouse, partner) | Social | 13494 | 0.00 | 0.04 | 9.00E-01 | 9.57E-01 | 254 | 13325 | 0.02 | 0.04 | 6.05E-01 | 7.19E-01 | 233 | 0.01 | 0.02 | 6.49E-01 | 0.00 | 7.82E-01 | - | - | - |
| Matairesinol intake (ug/day) | Nutrients | 26853 | 0.00 | 0.01 | 7.09E-01 | 8.36E-01 | 229 | 26173 | 0.00 | 0.01 | 8.12E-01 | 8.68E-01 | 259 | 0.00 | 0.01 | 6.65E-01 | 0.00 | 9.26E-01 | - | - | - |
| Beta-sitostanol intake (mg/day) | Nutrients | 26853 | 0.00 | 0.01 | 9.45E-01 | 9.73E-01 | 260 | 26173 | 0.00 | 0.01 | 5.97E-01 | 7.15E-01 | 231 | 0.00 | 0.01 | 6.73E-01 | 0.00 | 7.43E-01 | - | - | - |
| Butter for cooking | Food | 26853 | 0.01 | 0.01 | 5.63E-01 | 7.17E-01 | 212 | 26173 | 0.00 | 0.01 | 9.86E-01 | 9.92E-01 | 274 | 0.00 | 0.01 | 6.93E-01 | 0.00 | 6.72E-01 | - | - | - |
| For how much of the time during the last four weeks have you felt worn out? | General health | 13775 | 0.00 | 0.01 | 9.53E-01 | 9.73E-01 | 261 | 13576 | 0.01 | 0.01 | 5.45E-01 | 6.70E-01 | 224 | 0.00 | 0.01 | 7.01E-01 | 0.00 | 6.36E-01 | - | - | - |
| Sour milk, yoghurt (low fat) | Food | 26853 | 0.00 | 0.01 | 8.38E-01 | 9.35E-01 | 242 | 26173 | -0.01 | 0.01 | 4.62E-01 | 6.12E-01 | 209 | 0.00 | 0.01 | 7.08E-01 | 0.00 | 5.06E-01 | - | - | - |
| Enterodiol intake (ug/day) | Nutrients | 26853 | 0.01 | 0.01 | 5.14E-01 | 6.97E-01 | 198 | 26173 | 0.00 | 0.01 | 8.92E-01 | 9.33E-01 | 264 | 0.00 | 0.01 | 7.13E-01 | 0.00 | 5.79E-01 | - | - | - |
| Linolenic acid intake (g/day) | Nutrients | 26853 | 0.00 | 0.01 | 7.02E-01 | 8.31E-01 | 228 | 26173 | 0.01 | 0.01 | 3.91E-01 | 5.79E-01 | 187 | 0.00 | 0.01 | 7.39E-01 | 0.00 | 3.80E-01 | - | - | - |
| Distance to work in kilometers (one way) | Physical activity | 25360 | 0.00 | 0.01 | 8.28E-01 | 9.33E-01 | 239 | 24717 | 0.00 | 0.01 | 8.30E-01 | 8.77E-01 | 262 | 0.00 | 0.01 | 7.60E-01 | 0.00 | 1.00E+00 | - | - | - |
| Total protein intake (g/day) | Nutrients | 26853 | 0.01 | 0.01 | 1.54E-01 | 2.96E-01 | 140 | 26173 | -0.01 | 0.01 | 3.15E-01 | 4.93E-01 | 177 | 0.00 | 0.01 | 7.61E-01 | 0.66 | 8.58E-02 | - | - | - |
| Zinc intake (mg/day) | Nutrients | 26853 | 0.01 | 0.01 | 3.69E-01 | 5.53E-01 | 180 | 26173 | -0.01 | 0.01 | 1.87E-01 | 3.42E-01 | 151 | 0.00 | 0.01 | 7.73E-01 | 0.59 | 1.17E-01 | - | - | - |
| Fat intake (g/day) | Nutrients | 26853 | 0.00 | 0.01 | 9.62E-01 | 9.73E-01 | 267 | 26173 | 0.00 | 0.01 | 7.32E-01 | 8.20E-01 | 247 | 0.00 | 0.01 | 7.82E-01 | 0.00 | 8.35E-01 | - | - | - |
| Vitamin B6 intake (mg/day) | Nutrients | 26853 | 0.01 | 0.01 | 2.74E-01 | 4.45E-01 | 166 | 26173 | -0.01 | 0.01 | 1.42E-01 | 2.89E-01 | 136 | 0.00 | 0.01 | 7.94E-01 | 0.70 | 6.97E-02 | - | - | - |
| Whole grain crisp bread | Food | 26853 | 0.00 | 0.01 | 6.51E-01 | 7.99E-01 | 220 | 26173 | 0.01 | 0.01 | 4.51E-01 | 6.01E-01 | 208 | 0.00 | 0.01 | 8.34E-01 | 0.00 | 3.93E-01 | - | - | - |
| Linoleic acid intake (g/day) | Nutrients | 26853 | 0.00 | 0.01 | 8.75E-01 | 9.43E-01 | 247 | 26173 | 0.00 | 0.01 | 6.54E-01 | 7.55E-01 | 240 | 0.00 | 0.01 | 8.38E-01 | 0.00 | 6.68E-01 | - | - | - |
| For how much of the time during the last four weeks have you felt so depressed that nothing could cheer you up? | General health | 13812 | -0.01 | 0.01 | 5.47E-01 | 7.07E-01 | 209 | 13631 | 0.00 | 0.01 | 7.48E-01 | 8.35E-01 | 248 | 0.00 | 0.01 | 8.40E-01 | 0.00 | 5.14E-01 | - | - | - |
| Support from others | Social | 29289 | 0.00 | 0.01 | 9.82E-01 | 9.82E-01 | 270 | 28632 | 0.00 | 0.01 | 7.63E-01 | 8.42E-01 | 250 | 0.00 | 0.01 | 8.44E-01 | 0.00 | 8.18E-01 | - | - | - |
| Smoked fish/meat | Food | 26853 | 0.00 | 0.01 | 9.19E-01 | 9.65E-01 | 257 | 26173 | 0.00 | 0.01 | 7.69E-01 | 8.42E-01 | 252 | 0.00 | 0.01 | 8.95E-01 | 0.00 | 7.78E-01 | - | - | - |
| Sucrose intake (g/day) | Nutrients | 26853 | 0.00 | 0.01 | 6.49E-01 | 7.99E-01 | 219 | 26173 | 0.00 | 0.01 | 7.66E-01 | 8.42E-01 | 251 | 0.00 | 0.01 | 9.09E-01 | 0.00 | 5.95E-01 | - | - | - |
| Cohabitation: Live alone vs Only children | Social | 4352 | 0.03 | 0.05 | 5.55E-01 | 7.10E-01 | 211 | 4338 | -0.04 | 0.05 | 4.51E-01 | 6.01E-01 | 207 | 0.00 | 0.04 | 9.16E-01 | 0.00 | 3.41E-01 | - | - | - |
| Patience status | Psychosocial | 23539 | -0.01 | 0.01 | 5.28E-01 | 7.00E-01 | 203 | 23113 | 0.00 | 0.01 | 6.26E-01 | 7.36E-01 | 235 | 0.00 | 0.01 | 9.18E-01 | 0.00 | 4.29E-01 | - | - | - |
| Receive hugs to comfort and support you | Social | 29060 | -0.02 | 0.02 | 4.59E-01 | 6.49E-01 | 191 | 28407 | 0.01 | 0.02 | 5.45E-01 | 6.70E-01 | 225 | 0.00 | 0.02 | 9.19E-01 | 0.00 | 3.42E-01 | - | - | - |
| Frequent social contacts with colleagues during work | Psychosocial | 28543 | 0.01 | 0.01 | 3.31E-01 | 5.24E-01 | 170 | 27927 | -0.01 | 0.01 | 3.96E-01 | 5.80E-01 | 189 | 0.00 | 0.01 | 9.26E-01 | 0.40 | 1.98E-01 | - | - | - |
| Light beer | Alcohol | 26853 | -0.02 | 0.01 | 3.73E-02 | 1.11E-01 | 91 | 26173 | 0.02 | 0.01 | 2.75E-02 | 8.01E-02 | 95 | 0.00 | 0.01 | 9.36E-01 | 0.89 | 2.43E-03 | - | - | - |
| Oatflake, whole wheat, rye or barley porridge | Food | 26853 | 0.01 | 0.01 | 2.88E-01 | 4.60E-01 | 169 | 26173 | -0.01 | 0.01 | 3.36E-01 | 5.17E-01 | 180 | 0.00 | 0.01 | 9.43E-01 | 0.51 | 1.52E-01 | - | - | - |
| Sausage as main dish | Food | 26853 | 0.01 | 0.01 | 2.49E-01 | 4.14E-01 | 162 | 26173 | -0.01 | 0.01 | 2.16E-01 | 3.71E-01 | 161 | 0.00 | 0.01 | 9.48E-01 | 0.65 | 9.07E-02 | - | - | - |
| Vitamin E intake (mg/day) | Nutrients | 26853 | 0.00 | 0.01 | 9.19E-01 | 9.65E-01 | 256 | 26173 | 0.00 | 0.01 | 9.91E-01 | 9.92E-01 | 276 | 0.00 | 0.01 | 9.49E-01 | 0.00 | 9.36E-01 | - | - | - |
| Cheese 10-17% | Food | 26853 | 0.00 | 0.01 | 9.76E-01 | 9.80E-01 | 269 | 26173 | 0.00 | 0.01 | 9.51E-01 | 9.71E-01 | 271 | 0.00 | 0.01 | 9.49E-01 | 0.00 | 9.82E-01 | - | - | - |
| For how much of the time during the last four weeks have you felt happy? | General health | 13801 | 0.01 | 0.01 | 4.55E-01 | 6.47E-01 | 190 | 13613 | -0.01 | 0.01 | 5.04E-01 | 6.41E-01 | 218 | 0.00 | 0.01 | 9.52E-01 | 0.00 | 3.17E-01 | - | - | - |
| Changed everyday exercise during the last year | Physical activity | 18389 | 0.00 | 0.01 | 8.52E-01 | 9.39E-01 | 245 | 17883 | 0.00 | 0.01 | 9.18E-01 | 9.49E-01 | 268 | 0.00 | 0.01 | 9.52E-01 | 0.00 | 8.38E-01 | - | - | - |
| Sausage, liver pate on bread | Food | 26853 | 0.01 | 0.01 | 4.42E-01 | 6.32E-01 | 189 | 26173 | -0.01 | 0.01 | 4.95E-01 | 6.41E-01 | 212 | 0.00 | 0.01 | 9.52E-01 | 0.05 | 3.05E-01 | - | - | - |
| Whole grain soft bread | Food | 26853 | 0.00 | 0.01 | 8.29E-01 | 9.33E-01 | 240 | 26173 | 0.00 | 0.01 | 7.69E-01 | 8.42E-01 | 253 | 0.00 | 0.01 | 9.57E-01 | 0.00 | 7.19E-01 | - | - | - |
| Frequency of gardening during leisure time | Physical activity | 17504 | 0.01 | 0.01 | 5.41E-01 | 7.02E-01 | 207 | 16943 | -0.01 | 0.01 | 4.99E-01 | 6.41E-01 | 214 | 0.00 | 0.01 | 9.68E-01 | 0.00 | 3.62E-01 | - | - | - |
| Number of people with whom you can speak openly | Social | 29258 | 0.01 | 0.01 | 3.38E-01 | 5.31E-01 | 172 | 28593 | -0.01 | 0.01 | 3.05E-01 | 4.83E-01 | 175 | 0.00 | 0.01 | 9.69E-01 | 0.49 | 1.61E-01 | - | - | - |
| Milk, sour milk (1,5%) | Beverage | 26853 | 0.00 | 0.01 | 8.80E-01 | 9.43E-01 | 252 | 26173 | 0.00 | 0.01 | 9.15E-01 | 9.49E-01 | 267 | 0.00 | 0.01 | 9.74E-01 | 0.00 | 8.55E-01 | - | - | - |
| Butter on bread | Food | 26853 | 0.00 | 0.01 | 7.21E-01 | 8.45E-01 | 230 | 26173 | 0.00 | 0.01 | 6.90E-01 | 7.84E-01 | 244 | 0.00 | 0.01 | 9.76E-01 | 0.00 | 5.93E-01 | - | - | - |
| Oil for cooking | Food | 26853 | 0.00 | 0.01 | 8.10E-01 | 9.23E-01 | 237 | 26173 | 0.00 | 0.01 | 7.85E-01 | 8.49E-01 | 256 | 0.00 | 0.01 | 9.82E-01 | 0.00 | 7.16E-01 | - | - | - |
| For how much of the time during the last four weeks have you felt very nervous? | General health | 13798 | -0.01 | 0.01 | 5.41E-01 | 7.02E-01 | 208 | 13620 | 0.01 | 0.01 | 5.38E-01 | 6.68E-01 | 223 | 0.00 | 0.01 | 9.99E-01 | 0.00 | 3.85E-01 | - | - | - |
| Soft whey cheese | Food | 5185 | -0.01 | 0.02 | 6.34E-01 | 7.85E-01 | 218 | 5014 | -0.04 | 0.02 | 8.43E-02 | 1.97E-01 | 118 | -0.02 | 0.01 | - | 0.41 | 1.93E-01 | - | - | - |
| Amount of alcohol drunk in a day | Alcohol | 10469 | -0.02 | 0.02 | - | - | 274 | 10282 | -0.02 | 0.02 | 1.56E-01 | 3.16E-01 | 137 | -0.02 | 0.01 | - | 0.00 | 8.18E-01 | - | - | - |
| Shellfish (e.g. shrimps, scallops) | Food | 5185 | -0.01 | 0.02 | 6.73E-01 | 8.15E-01 | 223 | 5014 | -0.03 | 0.02 | 1.94E-01 | 3.47E-01 | 154 | -0.02 | 0.01 | - | 0.00 | 4.35E-01 | - | - | - |
| Potato salad | Food | 5185 | 0.03 | 0.02 | 1.05E-01 | 2.36E-01 | 120 | 5014 | 0.02 | 0.02 | 2.76E-01 | 4.50E-01 | 170 | 0.03 | 0.01 | - | 0.00 | 7.12E-01 | - | - | - |
| Blota (broth + bread) | Food | 5185 | -0.01 | 0.02 | 4.18E-01 | 6.20E-01 | 182 | 5014 | 0.02 | 0.02 | 3.12E-01 | 4.91E-01 | 176 | 0.00 | 0.01 | - | 0.51 | 1.53E-01 | - | - | - |
| Breath-holds during sleep | Sleep | 7573 | 0.03 | 0.02 | - | - | 284 | 7428 | 0.02 | 0.02 | 3.91E-01 | 5.79E-01 | 186 | 0.02 | 0.01 | - | 0.00 | 6.07E-01 | - | - | - |
| Liver, kidney | Food | 5185 | 0.02 | 0.02 | 2.43E-01 | 4.10E-01 | 160 | 5014 | 0.02 | 0.02 | 4.42E-01 | 5.97E-01 | 205 | 0.02 | 0.01 | - | 0.00 | 7.85E-01 | - | - | - |
| Times during last year that you felt guilty because of your drinking | Alcohol | 10642 | -0.03 | 0.02 | - | - | 276 | 10460 | -0.01 | 0.02 | 5.04E-01 | 6.41E-01 | 217 | -0.02 | 0.01 | - | 0.00 | 5.15E-01 | - | - | - |
| Mixed frozen vegetables | Food | 5185 | 0.02 | 0.02 | 3.67E-01 | 5.53E-01 | 178 | 5014 | -0.01 | 0.02 | 5.71E-01 | 6.93E-01 | 228 | 0.00 | 0.01 | - | 0.19 | 2.65E-01 | - | - | - |
| Risk of sleeping while sitting and reading | Sleep | 10701 | -0.03 | 0.02 | - | - | 277 | 10571 | -0.01 | 0.02 | 6.47E-01 | 7.53E-01 | 238 | -0.02 | 0.01 | - | 0.21 | 2.60E-01 | - | - | - |
| Soft cheese | Food | 5185 | -0.01 | 0.02 | 5.82E-01 | 7.31E-01 | 215 | 5014 | 0.01 | 0.02 | 6.63E-01 | 7.56E-01 | 242 | 0.00 | 0.01 | - | 0.00 | 4.63E-01 | - | - | - |
| Risk of sleeping while lying down resting in the afternoon | Sleep | 10778 | 0.01 | 0.02 | - | - | 281 | 10656 | 0.00 | 0.02 | 7.84E-01 | 8.49E-01 | 255 | 0.01 | 0.01 | - | 0.00 | 7.12E-01 | - | - | - |
| Snore during sleep | Sleep | 10086 | 0.05 | 0.02 | - | - | 283 | 9991 | 0.00 | 0.02 | 7.80E-01 | 8.49E-01 | 254 | 0.03 | 0.01 | - | 0.74 | 5.12E-02 | - | - | - |
| Mashed potato | Food | 5185 | 0.02 | 0.02 | 2.14E-01 | 3.73E-01 | 155 | 5014 | 0.00 | 0.02 | 8.12E-01 | 8.68E-01 | 258 | 0.01 | 0.01 | - | 0.19 | 2.68E-01 | - | - | - |
| Risk of sleeping while sitting still after having lunch | Sleep | 10785 | 0.00 | 0.02 | - | - | 282 | 10661 | 0.00 | 0.02 | 8.20E-01 | 8.74E-01 | 260 | 0.00 | 0.01 | - | 0.00 | 9.08E-01 | - | - | - |
| Blood based food | Food | 5185 | 0.02 | 0.02 | 2.85E-01 | 4.58E-01 | 168 | 5014 | 0.00 | 0.02 | 9.53E-01 | 9.71E-01 | 272 | 0.01 | 0.01 | - | 0.00 | 3.92E-01 | - | - | - |
| Time spent in a week in moderately steneous activities | Physical activity | 10582 | -0.08 | 0.02 | - | - | 271 | 10408 | -0.09 | 0.02 | - | - | 278 | -0.09 | 0.01 | - | 0.00 | 6.48E-01 | - | - | - |
| Amount of exercise during the last 12 months | Physical activity | 10910 | -0.10 | 0.02 | - | - | 272 | 10747 | -0.11 | 0.02 | - | - | 279 | -0.11 | 0.01 | - | 0.00 | 5.63E-01 | - | - | - |
| Frequency of alcohol consumption | Alcohol | 10948 | -0.07 | 0.02 | - | - | 273 | 10818 | -0.04 | 0.02 | - | - | 280 | -0.06 | 0.01 | - | 0.41 | 1.91E-01 | - | - | - |

| | | | | | | | | | | | | | | | | | | | | | |
|---|---|---|---|---|---|---|---|---|---|---|---|---|---|---|---|---|---|---|---|---|---|
| Frequency of drinking six or more glasses at the same occasion | Alcohol | 10653 | -0.03 | 0.02 | - | - | 275 | 10484 | 0.00 | 0.02 | - | - | 281 | -0.02 | 0.01 | - | 0.49 | 1.61E-01 | - | - | - |
| Risk of sleeping while watching TV | Sleep | 10815 | -0.01 | 0.02 | - | - | 278 | 10685 | -0.02 | 0.02 | - | - | 282 | -0.02 | 0.01 | - | 0.00 | 7.58E-01 | - | - | - |
| Risk of sleeping while sitting inactive in a public place | Sleep | 10748 | -0.03 | 0.02 | - | - | 279 | 10615 | -0.02 | 0.02 | - | - | 283 | -0.02 | 0.01 | - | 0.00 | 3.94E-01 | - | - | - |
| Risk of sleeping as a passenger in a car for one hour without break | Sleep | 10710 | -0.02 | 0.02 | - | - | 280 | 10571 | -0.02 | 0.02 | - | - | 284 | -0.02 | 0.01 | - | 0.00 | 6.69E-01 | - | - | - |

**Supplementary Table 12**. Longitudinal association results for BMI

| | | Training set | | | | | | Testing set | | | | | | Metanalysis | | | | | R2 | |
|---|---|---|---|---|---|---|---|---|---|---|---|---|---|---|---|---|---|---|---|---|
| **Description** | **Group** | **N** | **Effect estimate** | **S.E.** | **p-value** | **p-value$_{FDR}$** | **p-value rank** | **N** | **Effect estimate** | **S.E.** | **p-value** | **p-value$_{FDR}$** | **p-value rank** | **Effect estimate** | **S.E.** | **p-value** | **$I^2$** | **Q p-value** | **adjusted R2** | **R2 rank** |
| Smoking status: Smokers vs non-smokers | Tobacco use | 9766 | 0.41 | 0.05 | 4.94E-16 | **1.35E-13** | 1 | 9517 | 0.33 | 0.05 | 9.47E-10 | **2.14E-07** | 1 | 0.38 | 0.04 | 4.53E-24 | 0.13 | 2.84E-01 | 0.71 | 5 |
| Educational level | Psychosocial | 14723 | -0.11 | 0.02 | 7.48E-09 | **3.41E-07** | 6 | 14505 | -0.12 | 0.02 | 2.36E-09 | **2.14E-07** | 3 | -0.11 | 0.01 | 9.58E-17 | 0.00 | 7.65E-01 | 0.70 | 25 |
| Cohabitation: Live alone vs Adult and children | Social | 10276 | -0.40 | 0.07 | 2.16E-09 | **1.18E-07** | 5 | 9986 | -0.39 | 0.07 | 4.08E-08 | **2.79E-06** | 4 | -0.39 | 0.05 | 4.45E-16 | 0.00 | 8.99E-01 | 0.70 | 21 |
| Number of snuff boxes per week | Tobacco use | 13957 | 0.11 | 0.02 | 6.15E-08 | **1.86E-06** | 9 | 13756 | 0.12 | 0.02 | 1.84E-09 | **2.14E-07** | 2 | 0.11 | 0.01 | 6.70E-16 | 0.00 | 5.79E-01 | 0.70 | 9 |
| Years smoking | Tobacco use | 12996 | 0.13 | 0.02 | 4.15E-10 | **2.83E-08** | 4 | 12786 | 0.10 | 0.02 | 8.38E-07 | **3.81E-05** | 6 | 0.12 | 0.01 | 2.22E-15 | 0.00 | 4.64E-01 | 0.71 | 6 |
| Overall state of health during the last year | General health | 14646 | -0.11 | 0.02 | 1.16E-08 | **4.53E-07** | 7 | 14389 | -0.10 | 0.02 | 2.84E-07 | **1.55E-05** | 5 | -0.10 | 0.01 | 1.65E-14 | 0.00 | 8.08E-01 | 0.70 | 33 |
| Number of cigarretes smoked per day | Tobacco use | 9491 | 0.14 | 0.02 | 1.41E-10 | **1.93E-08** | 2 | 9247 | 0.09 | 0.02 | 1.88E-04 | **3.43E-03** | 15 | 0.12 | 0.02 | 4.14E-13 | 0.62 | 1.06E-01 | 0.71 | 1 |
| Snuff status: Snuff users vs non-snuff users | Tobacco use | 12437 | 0.28 | 0.05 | 1.03E-07 | **2.56E-06** | 11 | 12280 | 0.27 | 0.06 | 1.34E-06 | **5.23E-05** | 7 | 0.28 | 0.04 | 6.45E-13 | 0.00 | 8.66E-01 | 0.71 | 2 |
| Number of cigarretes smoked per day (in groups) | Tobacco use | 11923 | 0.13 | 0.02 | 3.51E-10 | **2.83E-08** | 3 | 11685 | 0.07 | 0.02 | 2.04E-03 | **1.49E-02** | 37 | 0.10 | 0.01 | 2.41E-11 | 0.77 | 3.71E-02 | 0.71 | 3 |
| Fiber cereals | Food | 13019 | -0.09 | 0.02 | 4.24E-06 | **8.90E-05** | 13 | 12782 | -0.09 | 0.02 | 2.87E-06 | **9.78E-05** | 8 | -0.09 | 0.01 | 5.30E-11 | 0.00 | 8.57E-01 | 0.70 | 7 |
| Sour milk, yoghurt (3% fat) | Food | 13019 | -0.11 | 0.02 | 4.37E-08 | **1.49E-06** | 8 | 12782 | -0.07 | 0.02 | 2.42E-04 | **3.89E-03** | 17 | -0.09 | 0.01 | 8.38E-11 | 0.25 | 2.49E-01 | 0.70 | 8 |
| Cohabitation: Live alone vs Only one adult (spouse, partner) | Social | 4587 | -0.41 | 0.08 | 9.85E-08 | **2.56E-06** | 10 | 4690 | -0.29 | 0.08 | 4.37E-04 | **6.63E-03** | 18 | -0.36 | 0.06 | 2.88E-10 | 0.11 | 2.89E-01 | 0.70 | 34 |
| Satisfaction with economy | Psychosocial | 6917 | -0.12 | 0.03 | 8.89E-06 | **1.62E-04** | 15 | 6787 | -0.10 | 0.03 | 7.55E-04 | **9.18E-03** | 22 | -0.11 | 0.02 | 3.02E-08 | 0.00 | 5.13E-01 | 0.70 | 20 |
| Repetitive job | Psychosocial | 14602 | 0.09 | 0.02 | 1.90E-06 | **4.32E-05** | 12 | 14339 | 0.06 | 0.02 | 2.44E-03 | **1.54E-02** | 42 | 0.07 | 0.01 | 3.04E-08 | 0.18 | 2.70E-01 | 0.70 | 22 |
| Changed everyday exercise during the last year | Physical activity | 14664 | 0.07 | 0.02 | 1.17E-04 | **1.74E-03** | 18 | 14409 | 0.08 | 0.02 | 6.85E-05 | **1.87E-03** | 10 | 0.07 | 0.01 | 3.07E-08 | 0.00 | 8.33E-01 | 0.70 | 28 |
| If you exercise, change in exercise habits during the last year | Physical activity | 12488 | 0.07 | 0.02 | 1.78E-04 | **2.43E-03** | 20 | 12286 | 0.08 | 0.02 | 2.17E-04 | **3.71E-03** | 16 | 0.08 | 0.01 | 1.39E-07 | 0.00 | 9.37E-01 | 0.70 | 35 |
| Fitness status | Physical activity | 6909 | -0.11 | 0.03 | 5.06E-05 | **8.12E-04** | 17 | 6787 | -0.09 | 0.03 | 2.58E-03 | **1.56E-02** | 45 | -0.10 | 0.02 | 5.35E-07 | 0.00 | 5.24E-01 | 0.70 | 29 |
| Cheese 28% | Food | 13019 | -0.09 | 0.02 | 4.88E-06 | **9.52E-05** | 14 | 12782 | -0.05 | 0.02 | 1.48E-02 | 5.19E-02 | 78 | -0.07 | 0.01 | 6.16E-07 | 0.49 | 1.60E-01 | - | - |
| Equol intake (ug/day) | Nutrients | 13019 | 0.07 | 0.02 | 2.01E-04 | **2.50E-03** | 22 | 12782 | 0.06 | 0.02 | 1.42E-03 | **1.30E-02** | 28 | 0.07 | 0.01 | 9.95E-07 | 0.00 | 7.88E-01 | 0.70 | 10 |
| Enterodiol intake (ug/day) | Nutrients | 13019 | 0.07 | 0.02 | 2.85E-04 | **3.24E-03** | 24 | 12782 | 0.07 | 0.02 | 1.05E-03 | **1.19E-02** | 24 | 0.07 | 0.01 | 1.02E-06 | 0.00 | 8.82E-01 | 0.70 | 11 |
| Matairesinol intake (ug/day) | Nutrients | 13019 | -0.07 | 0.02 | 3.36E-04 | **3.67E-03** | 25 | 12782 | -0.07 | 0.02 | 1.29E-03 | **1.30E-02** | 25 | -0.07 | 0.01 | 1.46E-06 | 0.00 | 8.71E-01 | 0.70 | 12 |
| Exercise during the last three months | Physical activity | 14607 | -0.08 | 0.02 | 1.41E-05 | **2.40E-04** | 16 | 14361 | -0.05 | 0.02 | 1.76E-02 | 5.80E-02 | 83 | -0.06 | 0.01 | 1.75E-06 | 0.40 | 1.98E-01 | - | - |
| Appetite status | Psychosocial | 6912 | -0.10 | 0.03 | 2.29E-04 | **2.72E-03** | 23 | 6791 | -0.09 | 0.03 | 2.48E-03 | **1.54E-02** | 44 | -0.09 | 0.02 | 1.98E-06 | 0.00 | 7.09E-01 | 0.70 | 26 |
| Tiamin intake (mg/day) | Nutrients | 13019 | 0.06 | 0.02 | 2.41E-03 | **1.69E-02** | 39 | 12782 | 0.07 | 0.02 | 5.56E-04 | **7.23E-03** | 21 | 0.07 | 0.01 | 4.61E-06 | 0.00 | 7.09E-01 | 0.70 | 13 |
| Informed of having high blood pressure | General health | 14712 | 0.17 | 0.05 | 1.07E-03 | **8.60E-03** | 34 | 14455 | 0.17 | 0.05 | 1.38E-03 | **1.30E-02** | 27 | 0.17 | 0.04 | 4.73E-06 | 0.00 | 9.77E-01 | 0.70 | 30 |
| Sodas, soft drinks, juice | Beverage | 13019 | 0.04 | 0.02 | 3.62E-02 | 1.06E-01 | 93 | 12782 | 0.09 | 0.02 | 1.10E-05 | **3.35E-04** | 9 | 0.07 | 0.01 | 5.26E-06 | 0.67 | 8.39E-02 | - | - |
| Tomato, cucumber | Food | 13019 | 0.08 | 0.02 | 1.91E-04 | **2.49E-03** | 21 | 12782 | 0.06 | 0.02 | 7.43E-03 | **3.27E-02** | 62 | 0.07 | 0.01 | 5.46E-06 | 0.00 | 5.16E-01 | 0.70 | 14 |
| Close relationship with anyone | Social | 14628 | -0.06 | 0.02 | 1.32E-03 | **9.72E-03** | 37 | 14374 | -0.06 | 0.02 | 1.34E-03 | **1.30E-02** | 26 | -0.06 | 0.01 | 5.64E-06 | 0.00 | 9.30E-01 | 0.70 | 32 |
| Work shifts/weekends | Psychosocial | 13831 | 0.15 | 0.04 | 6.58E-04 | **6.42E-03** | 28 | 13576 | 0.13 | 0.04 | 2.74E-03 | **1.62E-02** | 46 | 0.14 | 0.03 | 5.86E-06 | 0.00 | 8.15E-01 | 0.70 | 19 |
| High physical demand from job | Physical activity | 14643 | 0.07 | 0.02 | 1.21E-04 | **1.74E-03** | 19 | 14380 | 0.05 | 0.02 | 1.30E-02 | **4.80E-02** | 74 | 0.06 | 0.01 | 6.98E-06 | 0.00 | 3.86E-01 | 0.70 | 31 |
| Sodium intake (mg/day) | Nutrients | 13019 | 0.05 | 0.02 | 1.69E-02 | 6.78E-02 | 68 | 12782 | 0.08 | 0.02 | 9.51E-05 | **2.16E-03** | 12 | 0.06 | 0.01 | 9.54E-06 | 0.26 | 2.45E-01 | - | - |
| Marital status: Single vs Married/partner | Social | 13757 | -0.17 | 0.06 | 5.18E-03 | **3.07E-02** | 46 | 13512 | -0.22 | 0.06 | 5.07E-04 | **7.23E-03** | 19 | -0.20 | 0.04 | 9.57E-06 | 0.00 | 5.73E-01 | 0.70 | 17 |
| Fried potatoes, pommes frites | Food | 13019 | 0.06 | 0.02 | 1.30E-03 | **9.72E-03** | 36 | 12782 | 0.06 | 0.02 | 2.92E-03 | **1.66E-02** | 48 | 0.06 | 0.01 | 1.17E-05 | 0.00 | 9.28E-01 | 0.70 | 15 |
| Breakfast habits: Porridge w/o sandwich for breakfast vs not breakfast at all | Food | 1747 | -0.27 | 0.12 | 2.50E-02 | 8.30E-02 | 82 | 1697 | -0.49 | 0.13 | 1.48E-04 | **3.11E-03** | 13 | -0.38 | 0.09 | 2.15E-05 | 0.31 | 2.30E-01 | - | - |
| Satisfaction with accomodation | Psychosocial | 6922 | -0.10 | 0.03 | 4.78E-04 | **4.84E-03** | 27 | 6799 | -0.07 | 0.03 | 1.32E-02 | **4.80E-02** | 75 | -0.08 | 0.02 | 2.26E-05 | 0.00 | 5.26E-01 | 0.70 | 24 |
| Arachidonic acid (ARA) intake (g/day) | Nutrients | 13019 | 0.06 | 0.02 | 3.41E-03 | **2.22E-02** | 42 | 12782 | 0.06 | 0.02 | 2.29E-03 | **1.54E-02** | 40 | 0.06 | 0.01 | 2.39E-05 | 0.00 | 8.56E-01 | 0.70 | 16 |
| Satisfaction with home and family situation | Psychosocial | 6918 | -0.08 | 0.03 | 4.66E-03 | **2.83E-02** | 45 | 6795 | -0.09 | 0.03 | 1.74E-03 | **1.44E-02** | 33 | -0.08 | 0.02 | 2.54E-05 | 0.00 | 7.67E-01 | 0.70 | 23 |
| Years using snuff | Tobacco use | 13587 | 0.06 | 0.02 | 1.60E-03 | **1.15E-02** | 38 | 13355 | 0.06 | 0.02 | 5.38E-03 | **2.71E-02** | 54 | 0.06 | 0.01 | 2.61E-05 | 0.00 | 8.64E-01 | 0.71 | 4 |
| Stigmasterol intake (mg/day) | Nutrients | 13019 | 0.07 | 0.02 | 3.82E-04 | **4.02E-03** | 26 | 12782 | 0.05 | 0.02 | 1.90E-02 | 6.11E-02 | 85 | 0.06 | 0.01 | 2.84E-05 | 0.00 | 4.38E-01 | - | - |
| Sleep status | Sleep | 6925 | -0.08 | 0.03 | 3.33E-03 | **2.22E-02** | 41 | 6803 | -0.08 | 0.03 | 2.91E-03 | **1.66E-02** | 47 | -0.08 | 0.02 | 2.90E-05 | 0.00 | 9.07E-01 | 0.70 | 27 |
| Travel to work: Cycle to work vs passive travel to work | Physical activity | 11353 | -0.12 | 0.05 | 8.50E-03 | **4.46E-02** | 52 | 11213 | -0.16 | 0.05 | 1.47E-03 | **1.30E-02** | 30 | -0.14 | 0.03 | 4.17E-05 | 0.00 | 6.18E-01 | 0.70 | 18 |
| Bregott on bread | Food | 13019 | -0.04 | 0.02 | 2.30E-02 | 8.01E-02 | 78 | 12782 | -0.07 | 0.02 | 5.37E-04 | **7.23E-03** | 20 | -0.06 | 0.01 | 5.36E-05 | 0.00 | 3.59E-01 | - | - |
| Beta-carotene intake (mg/day) | Nutrients | 13019 | 0.07 | 0.02 | 1.28E-03 | **9.72E-03** | 35 | 12782 | 0.05 | 0.02 | 1.52E-02 | 5.27E-02 | 79 | 0.06 | 0.01 | 6.24E-05 | 0.00 | 6.23E-01 | - | - |
| Chips, popcorn, salted nuts | Food | 13019 | 0.07 | 0.02 | 9.85E-04 | **8.37E-03** | 31 | 12782 | 0.05 | 0.02 | 2.82E-02 | 8.12E-02 | 95 | 0.06 | 0.01 | 9.73E-05 | 0.00 | 4.86E-01 | - | - |
| Receive hugs to comfort and support you | Social | 14560 | -0.07 | 0.05 | 1.40E-01 | 2.79E-01 | 137 | 14300 | -0.21 | 0.05 | 7.69E-05 | **1.91E-03** | 11 | -0.14 | 0.04 | 1.43E-04 | 0.70 | 6.70E-02 | - | - |
| Wine | Alcohol | 13019 | -0.04 | 0.02 | 2.49E-02 | 8.30E-02 | 81 | 12782 | -0.06 | 0.02 | 2.13E-03 | **1.49E-02** | 39 | -0.05 | 0.01 | 1.79E-04 | 0.00 | 5.09E-01 | - | - |
| Low fat milk (0,5%) | Beverage | 13019 | 0.05 | 0.02 | 1.36E-02 | 6.10E-02 | 61 | 12782 | 0.06 | 0.02 | 5.29E-03 | **2.71E-02** | 53 | 0.05 | 0.01 | 2.06E-04 | 0.00 | 7.61E-01 | - | - |
| Support from others | Social | 14753 | -0.05 | 0.02 | 1.41E-02 | 6.15E-02 | 62 | 14518 | -0.05 | 0.02 | 5.14E-03 | **2.70E-02** | 52 | -0.05 | 0.01 | 2.09E-04 | 0.00 | 7.36E-01 | - | - |
| Root vegetables, carrot | Food | 13019 | 0.06 | 0.02 | 2.82E-03 | **1.92E-02** | 40 | 12782 | 0.05 | 0.02 | 2.50E-02 | 7.49E-02 | 91 | 0.05 | 0.01 | 2.10E-04 | 0.00 | 6.45E-01 | - | - |
| Sour milk, yoghurt (low fat) | Food | 13019 | 0.07 | 0.02 | 7.62E-04 | **7.17E-03** | 29 | 12782 | 0.04 | 0.02 | 7.09E-02 | 1.61E-01 | 120 | 0.05 | 0.01 | 2.35E-04 | 0.07 | 3.00E-01 | - | - |
| Whole grain intake (g/day) | Food | 13019 | -0.03 | 0.02 | 1.45E-01 | 2.84E-01 | 139 | 12782 | -0.08 | 0.02 | 1.71E-04 | **3.34E-03** | 14 | -0.05 | 0.01 | 2.54E-04 | 0.65 | 9.00E-02 | - | - |
| Light beer | Alcohol | 13019 | -0.05 | 0.02 | 1.54E-02 | 6.46E-02 | 65 | 12782 | -0.06 | 0.02 | 6.66E-03 | **3.03E-02** | 60 | -0.05 | 0.01 | 2.85E-04 | 0.00 | 7.83E-01 | - | - |
| Cookies, pastry | Food | 13019 | -0.05 | 0.02 | 1.73E-02 | 6.86E-02 | 69 | 12782 | -0.06 | 0.02 | 5.91E-03 | **2.78E-02** | 58 | -0.05 | 0.01 | 2.90E-04 | 0.00 | 7.37E-01 | - | - |
| Syringaresinol intake (ug/day) | Nutrients | 13019 | -0.05 | 0.02 | 1.10E-02 | 5.46E-02 | 55 | 12782 | -0.05 | 0.02 | 9.85E-03 | **3.96E-02** | 68 | -0.05 | 0.01 | 2.92E-04 | 0.00 | 9.21E-01 | - | - |
| Vitamin B3 intake (mg/day) | Nutrients | 13019 | 0.04 | 0.02 | 4.40E-02 | 1.19E-01 | 100 | 12782 | 0.06 | 0.02 | 1.89E-03 | **1.49E-02** | 34 | 0.05 | 0.01 | 3.14E-04 | 0.00 | 3.96E-01 | - | - |
| Mashed potato | Food | 6555 | 0.06 | 0.03 | 3.14E-02 | 9.73E-02 | 88 | 6423 | 0.08 | 0.03 | 4.15E-03 | **2.22E-02** | 51 | 0.07 | 0.02 | 3.87E-04 | 0.00 | 6.06E-01 | - | - |
| Lariciresinol intake (ug/day) | Nutrients | 13019 | -0.04 | 0.02 | 2.83E-02 | 8.88E-02 | 87 | 12782 | -0.06 | 0.02 | 5.47E-03 | **2.71E-02** | 55 | -0.05 | 0.01 | 4.54E-04 | 0.00 | 6.28E-01 | - | - |
| Vitamin B12 intake (ug/day) | Nutrients | 13019 | 0.05 | 0.02 | 1.67E-02 | 6.78E-02 | 67 | 12782 | 0.05 | 0.02 | 1.12E-02 | **4.35E-02** | 70 | 0.05 | 0.01 | 4.95E-04 | 0.00 | 8.61E-01 | - | - |
| Sum of all lignans intake (ug/day) | Nutrients | 13019 | -0.05 | 0.02 | 1.81E-02 | 7.07E-02 | 70 | 12782 | -0.05 | 0.02 | 1.03E-02 | **4.06E-02** | 69 | -0.05 | 0.01 | 4.96E-04 | 0.00 | 8.31E-01 | - | - |
| Vitamin C intake (mg/day) | Nutrients | 13019 | 0.04 | 0.02 | 7.87E-02 | 1.82E-01 | 118 | 12782 | 0.07 | 0.02 | 1.55E-03 | **1.32E-02** | 32 | 0.05 | 0.01 | 5.36E-04 | 0.11 | 2.88E-01 | - | - |
| Sum of Lariciresinol, Matairesinol, Pinoresinol, Secoisolariciresinol intake (ug/day) | Nutrients | 13019 | -0.04 | 0.02 | 3.24E-02 | 9.82E-02 | 90 | 12782 | -0.06 | 0.02 | 5.85E-03 | **2.78E-02** | 57 | -0.05 | 0.01 | 5.54E-04 | 0.00 | 6.14E-01 | - | - |
| Number of people with whom you can speak openly | Social | 14743 | -0.03 | 0.02 | 9.47E-02 | 2.12E-01 | 122 | 14499 | -0.06 | 0.02 | 1.48E-03 | **1.30E-02** | 31 | -0.05 | 0.01 | 6.56E-04 | 0.23 | 2.55E-01 | - | - |
| Breakfast habits: Gruel w/o sandwich for breakfast vs not breakfast at all | Food | 961 | -0.45 | 0.20 | 2.42E-02 | 8.25E-02 | 80 | 913 | -0.50 | 0.20 | 1.15E-02 | **4.35E-02** | 72 | -0.48 | 0.14 | 7.03E-04 | 0.00 | 8.53E-01 | - | - |
| Whole grain soft bread | Food | 13019 | -0.04 | 0.02 | 2.82E-02 | 8.88E-02 | 86 | 12782 | -0.05 | 0.02 | 9.37E-03 | **3.83E-02** | 66 | -0.05 | 0.01 | 7.19E-04 | 0.00 | 7.22E-01 | - | - |
| Satisfaction with work situation | Psychosocial | 6853 | -0.09 | 0.03 | 1.01E-03 | **8.37E-03** | 33 | 6726 | -0.04 | 0.03 | 1.47E-01 | 2.63E-01 | 153 | -0.07 | 0.02 | 7.31E-04 | 0.34 | 2.20E-01 | - | - |
| Selenium intake (ug/day) | Nutrients | 13019 | 0.05 | 0.02 | 9.90E-03 | 5.01E-02 | 54 | 12782 | 0.04 | 0.02 | 2.88E-02 | 8.18E-02 | 96 | 0.05 | 0.01 | 7.36E-04 | 0.00 | 8.40E-01 | - | - |
| Parents or siblings had a cerebral hemorrhage/thrombosis or cardiac infarction before the age of 60 | General health | 14522 | 0.16 | 0.05 | 9.13E-04 | **8.31E-03** | 30 | 14292 | 0.07 | 0.05 | 1.57E-01 | 2.76E-01 | 155 | 0.12 | 0.03 | 7.55E-04 | 0.40 | 1.98E-01 | - | - |
| Bacon | Food | 13019 | 0.04 | 0.02 | 2.32E-02 | 8.01E-02 | 79 | 12782 | 0.05 | 0.02 | 1.70E-02 | 5.74E-02 | 81 | 0.05 | 0.01 | 9.99E-04 | 0.00 | 8.82E-01 | - | - |
| Total energy intake (kcal/day) | Nutrients | 13019 | -0.03 | 0.02 | 1.22E-01 | 2.54E-01 | 130 | 12782 | -0.07 | 0.02 | 2.00E-03 | **1.49E-02** | 36 | -0.05 | 0.02 | 1.14E-03 | 0.27 | 2.43E-01 | - | - |
| Pentadecanoic acid intake (g/day) | Nutrients | 13019 | -0.05 | 0.02 | 2.05E-02 | 7.28E-02 | 76 | 12782 | -0.05 | 0.02 | 2.46E-02 | 7.47E-02 | 90 | -0.05 | 0.01 | 1.25E-03 | 0.00 | 9.88E-01 | - | - |
| Heptadecanoic acid intake (g/day) | Nutrients | 13019 | -0.05 | 0.02 | 2.05E-02 | 7.28E-02 | 77 | 12782 | -0.05 | 0.02 | 2.46E-02 | 7.47E-02 | 89 | -0.05 | 0.01 | 1.25E-03 | 0.00 | 9.88E-01 | - | - |
| Hamburger | Food | 13019 | 0.06 | 0.02 | 4.09E-03 | **2.54E-02** | 44 | 12782 | 0.03 | 0.02 | 9.73E-02 | 2.04E-01 | 130 | 0.05 | 0.01 | 1.29E-03 | 0.00 | 4.26E-01 | - | - |
| Animal based protein intake (g/day) | Nutrients | 13019 | 0.04 | 0.02 | 4.41E-02 | 1.19E-01 | 101 | 12782 | 0.05 | 0.02 | 1.15E-02 | **4.35E-02** | 71 | 0.04 | 0.01 | 1.36E-03 | 0.00 | 6.67E-01 | - | - |
| Do you feel important and appreciated in your home? | Psychosocial | 6872 | -0.07 | 0.03 | 1.14E-02 | 5.54E-02 | 56 | 6741 | -0.06 | 0.03 | 4.80E-02 | 1.21E-01 | 108 | -0.06 | 0.02 | 1.39E-03 | 0.00 | 7.44E-01 | - | - |
| Pinoresinol intake (ug/day) | Nutrients | 13019 | -0.04 | 0.02 | 4.66E-02 | 1.24E-01 | 103 | 12782 | -0.05 | 0.02 | 1.20E-02 | **4.47E-02** | 73 | -0.05 | 0.01 | 1.49E-03 | 0.00 | 6.67E-01 | - | - |
| Long-term sickness | General health | 13389 | 0.15 | 0.06 | 9.08E-03 | **4.68E-02** | 53 | 13092 | 0.11 | 0.06 | 6.44E-02 | 1.50E-01 | 117 | 0.13 | 0.04 | 1.54E-03 | 0.00 | 6.55E-01 | - | - |
| People to ask for help apart from the ones at home | Social | 14714 | -0.08 | 0.06 | 1.97E-01 | 3.61E-01 | 149 | 14473 | -0.20 | 0.06 | 1.45E-03 | **1.30E-02** | 29 | -0.14 | 0.04 | 1.62E-03 | 0.47 | 1.71E-01 | - | - |
| Excellent health | General health | 191 | -0.25 | 0.24 | 2.95E-01 | 4.74E-01 | 169 | 160 | -0.54 | 0.18 | 2.36E-03 | **1.54E-02** | 41 | -0.44 | 0.14 | 1.82E-03 | 0.00 | 3.31E-01 | - | - |
| Iodine intake (ug/day) | Nutrients | 13019 | 0.03 | 0.02 | 7.86E-02 | 1.82E-01 | 117 | 12782 | 0.05 | 0.02 | 8.00E-03 | **3.41E-02** | 64 | 0.04 | 0.01 | 1.90E-03 | 0.00 | 4.84E-01 | - | - |
| Formic acid intaje (g/day) | Nutrients | 13019 | -0.05 | 0.02 | 1.96E-02 | 7.28E-02 | 73 | 12782 | -0.04 | 0.02 | 4.40E-02 | 1.17E-01 | 103 | -0.04 | 0.01 | 2.08E-03 | 0.00 | 8.67E-01 | - | - |
| Frequency of cycling during leisure time | Physical activity | 12458 | -0.05 | 0.02 | 1.43E-02 | 6.15E-02 | 63 | 12261 | -0.04 | 0.02 | 6.28E-02 | 1.49E-01 | 115 | -0.04 | 0.01 | 2.24E-03 | 0.00 | 7.28E-01 | - | - |
| Minced meat dishes | Food | 13019 | 0.03 | 0.02 | 1.26E-01 | 2.59E-01 | 133 | 12782 | 0.06 | 0.02 | 5.74E-03 | **2.78E-02** | 56 | 0.04 | 0.01 | 2.57E-03 | 0.00 | 3.47E-01 | - | - |
| Tea | Beverage | 13019 | -0.04 | 0.02 | 3.32E-02 | 9.95E-02 | 91 | 12782 | -0.04 | 0.02 | 3.57E-02 | 1.00E-01 | 97 | -0.04 | 0.01 | 2.77E-03 | 0.00 | 9.71E-01 | - | - |
| For how much of the time during the last four weeks have you felt really alert and strong? | General health | 192 | -0.22 | 0.24 | 3.56E-01 | 5.30E-01 | 183 | 162 | -0.52 | 0.17 | 3.12E-03 | **1.74E-02** | 49 | -0.42 | 0.14 | 2.85E-03 | 0.00 | 3.24E-01 | - | - |
| Feel uneasy or guilty because of your way of drinking | Alcohol | 13039 | 0.13 | 0.06 | 3.22E-02 | 9.82E-02 | 89 | 12865 | 0.13 | 0.06 | 4.04E-02 | 1.09E-01 | 101 | 0.13 | 0.04 | 3.03E-03 | 0.00 | 9.87E-01 | - | - |

| | | | | | | | | | | | | | | | | | | | | |
|---|---|---|---|---|---|---|---|---|---|---|---|---|---|---|---|---|---|---|---|---|
| Feel the need to reduce alcohol consumption | Alcohol | 13040 | 0.10 | 0.06 | 1.14E-01 | 2.46E-01 | 127 | 12884 | 0.17 | 0.06 | 9.22E-03 | **3.83E-02** | 65 | 0.13 | 0.05 | 3.20E-03 | 0.00 | 4.44E-01 | - | - |
| Ingenuity or creativity demand from job | Psychosocial | 14554 | -0.04 | 0.02 | 4.08E-02 | 1.16E-01 | 96 | 14307 | -0.04 | 0.02 | 3.63E-02 | 1.01E-01 | 98 | -0.04 | 0.01 | 3.44E-03 | 0.00 | 9.04E-01 | - | - |
| Average portion size of meat/fish based on photographic illustration of four sizes (smallest to largest) | Food | 13019 | 0.04 | 0.02 | 6.18E-02 | 1.55E-01 | 109 | 12782 | 0.05 | 0.02 | 2.39E-02 | 7.41E-02 | 88 | 0.05 | 0.02 | 3.58E-03 | 0.00 | 7.40E-01 | - | - |
| Control over planning and execution of the workday | Psychosocial | 14624 | -0.05 | 0.02 | 1.23E-02 | 5.60E-02 | 60 | 14353 | -0.03 | 0.02 | 1.32E-01 | 2.43E-01 | 148 | -0.04 | 0.01 | 4.34E-03 | 0.00 | 5.26E-01 | - | - |
| Parents or siblings have diabetes | General health | 14548 | 0.09 | 0.05 | 7.27E-02 | 1.74E-01 | 114 | 14317 | 0.11 | 0.05 | 2.60E-02 | 7.70E-02 | 92 | 0.10 | 0.04 | 4.51E-03 | 0.00 | 7.32E-01 | - | - |
| Sausage as main dish | Food | 13019 | 0.04 | 0.02 | 4.35E-02 | 1.19E-01 | 99 | 12782 | 0.04 | 0.02 | 4.64E-02 | 1.20E-01 | 106 | 0.04 | 0.01 | 4.57E-03 | 0.00 | 9.67E-01 | - | - |
| Patience status | Psychosocial | 6913 | -0.06 | 0.03 | 2.52E-02 | 8.30E-02 | 83 | 6792 | -0.05 | 0.03 | 9.26E-02 | 2.01E-01 | 125 | -0.05 | 0.02 | 5.49E-03 | 0.00 | 7.18E-01 | - | - |
| Potassium intake (mg/day) | Nutrients | 13019 | 0.03 | 0.02 | 1.20E-01 | 2.54E-01 | 128 | 12782 | 0.05 | 0.02 | 1.75E-02 | 5.80E-02 | 82 | 0.04 | 0.02 | 5.62E-03 | 0.00 | 5.27E-01 | - | - |
| Shellfish (e.g. shrimps, scallops) | Food | 6555 | 0.09 | 0.03 | 1.01E-03 | **8.37E-03** | 32 | 6423 | 0.02 | 0.03 | 5.75E-01 | 6.85E-01 | 229 | 0.05 | 0.02 | 5.79E-03 | 0.72 | 6.04E-02 | - | - |
| Energy status | Psychosocial | 6910 | -0.05 | 0.03 | 6.60E-02 | 1.61E-01 | 112 | 6780 | -0.06 | 0.03 | 3.88E-02 | 1.07E-01 | 99 | -0.05 | 0.02 | 5.79E-03 | 0.00 | 8.47E-01 | - | - |
| Hearing status | General health | 6917 | -0.04 | 0.03 | 1.42E-01 | 2.81E-01 | 138 | 6801 | -0.07 | 0.03 | 1.46E-02 | 5.16E-02 | 77 | -0.05 | 0.02 | 5.94E-03 | 0.00 | 4.55E-01 | - | - |
| Milk, sour milk (3%) | Beverage | 13019 | 0.05 | 0.02 | 1.65E-02 | 6.78E-02 | 66 | 12782 | 0.03 | 0.02 | 1.45E-01 | 2.61E-01 | 152 | 0.04 | 0.01 | 6.15E-03 | 0.00 | 5.44E-01 | - | - |
| Self rate of overall health | General health | 191 | -0.07 | 0.23 | 7.58E-01 | 8.46E-01 | 243 | 161 | -0.55 | 0.18 | 1.91E-03 | **1.49E-02** | 35 | -0.38 | 0.14 | 6.71E-03 | 0.63 | 9.89E-02 | - | - |
| Potato salad | Food | 5354 | 0.07 | 0.03 | 2.65E-02 | 8.62E-02 | 84 | 5236 | 0.05 | 0.03 | 1.08E-01 | 2.18E-01 | 135 | 0.06 | 0.02 | 6.86E-03 | 0.00 | 6.57E-01 | - | - |
| Butter for cooking | Food | 13019 | 0.01 | 0.02 | 6.12E-01 | 7.49E-01 | 223 | 12782 | 0.07 | 0.02 | 7.74E-04 | **9.18E-03** | 23 | 0.04 | 0.01 | 6.97E-03 | 0.77 | 3.85E-02 | - | - |
| Smoked fish/meat | Food | 13019 | 0.04 | 0.02 | 2.69E-02 | 8.64E-02 | 85 | 12782 | 0.03 | 0.02 | 1.13E-01 | 2.21E-01 | 139 | 0.04 | 0.01 | 7.06E-03 | 0.00 | 6.93E-01 | - | - |
| For how much of the time during the last four weeks have you felt happy? | General health | 192 | -0.32 | 0.23 | 1.72E-01 | 3.23E-01 | 145 | 162 | -0.41 | 0.18 | 2.15E-02 | 6.82E-02 | 86 | -0.38 | 0.14 | 7.36E-03 | 0.00 | 7.58E-01 | - | - |
| Total protein intake (g/day) | Nutrients | 13019 | 0.03 | 0.02 | 1.12E-01 | 2.43E-01 | 126 | 12782 | 0.04 | 0.02 | 2.82E-02 | 8.12E-02 | 94 | 0.04 | 0.01 | 7.70E-03 | 0.00 | 6.26E-01 | - | - |
| Worsen in health in the future | General health | 188 | 0.43 | 0.23 | 6.99E-02 | 1.69E-01 | 113 | 160 | 0.33 | 0.18 | 6.47E-02 | 1.50E-01 | 118 | 0.37 | 0.14 | 9.72E-03 | 0.00 | 7.49E-01 | - | - |
| Low fat margarine on bread | Food | 13019 | 0.02 | 0.02 | 2.39E-01 | 4.11E-01 | 158 | 12782 | 0.05 | 0.02 | 1.38E-02 | **4.94E-02** | 76 | 0.04 | 0.01 | 1.05E-02 | 0.00 | 3.37E-01 | - | - |
| Marital status: Single vs Divorced/separated | Social | 2364 | 0.28 | 0.11 | 1.20E-02 | 5.58E-02 | 58 | 2316 | 0.12 | 0.12 | 3.06E-01 | 4.53E-01 | 184 | 0.21 | 0.08 | 1.09E-02 | 0.00 | 3.46E-01 | - | - |
| Mixed frozen vegetables | Food | 6555 | 0.06 | 0.03 | 3.54E-02 | 1.05E-01 | 92 | 6423 | 0.04 | 0.03 | 1.38E-01 | 2.53E-01 | 149 | 0.05 | 0.02 | 1.10E-02 | 0.00 | 6.94E-01 | - | - |
| Cream, creme fraiche, sour cream | Food | 13019 | -0.02 | 0.02 | 3.04E-01 | 4.80E-01 | 173 | 12782 | -0.05 | 0.02 | 9.39E-03 | **3.83E-02** | 67 | -0.04 | 0.01 | 1.11E-02 | 0.26 | 2.44E-01 | - | - |
| White cabbage, lettuce, lettuce cabbage, spinach, borecole | Food | 13019 | 0.04 | 0.02 | 4.15E-02 | 1.17E-01 | 97 | 12782 | 0.03 | 0.02 | 1.24E-01 | 2.37E-01 | 143 | 0.04 | 0.01 | 1.12E-02 | 0.00 | 7.63E-01 | - | - |
| Everyday exercise satisfaction | Physical activity | 14622 | 0.05 | 0.02 | 8.46E-03 | **4.46E-02** | 51 | 14366 | 0.02 | 0.02 | 3.67E-01 | 5.27E-01 | 190 | 0.03 | 0.01 | 1.13E-02 | 0.25 | 2.47E-01 | - | - |
| Number of cigars smoked per day | Tobacco use | 9118 | 0.03 | 0.02 | 1.80E-01 | 3.34E-01 | 147 | 8905 | 0.05 | 0.02 | 2.74E-02 | 8.03E-02 | 93 | 0.04 | 0.02 | 1.26E-02 | 0.00 | 5.06E-01 | - | - |
| Mood status | Psychosocial | 6912 | -0.05 | 0.03 | 5.23E-02 | 1.36E-01 | 105 | 6790 | -0.04 | 0.03 | 1.18E-01 | 2.28E-01 | 141 | -0.05 | 0.02 | 1.30E-02 | 0.00 | 8.31E-01 | - | - |
| Extent to what your physical and emotional health disrupted your usual social life during the last four weeks | General health | 192 | 0.00 | 0.23 | 9.89E-01 | 9.92E-01 | 272 | 162 | 0.53 | 0.17 | 2.46E-03 | **1.54E-02** | 43 | 0.34 | 0.14 | 1.32E-02 | 0.70 | 6.77E-02 | - | - |
| Participation in study circles | Social | 777 | 0.08 | 0.27 | 7.61E-01 | 8.46E-01 | 244 | 690 | 0.63 | 0.22 | 3.57E-03 | **1.95E-02** | 50 | 0.41 | 0.17 | 1.38E-02 | 0.61 | 1.09E-01 | - | - |
| Eat breakfast from 2000 | Food | 1208 | -0.41 | 0.23 | 7.77E-02 | 1.82E-01 | 116 | 1089 | -0.34 | 0.20 | 9.34E-02 | 2.01E-01 | 126 | -0.37 | 0.15 | 1.52E-02 | 0.00 | 8.38E-01 | - | - |
| Medioresinol intake (ug/day) | Nutrients | 13019 | -0.03 | 0.02 | 7.94E-02 | 1.82E-01 | 119 | 12782 | -0.03 | 0.02 | 9.39E-02 | 2.01E-01 | 127 | -0.03 | 0.01 | 1.53E-02 | 0.00 | 9.94E-01 | - | - |
| Cohabitation: Live alone vs Other/others | Social | 1497 | -0.44 | 0.19 | 1.91E-02 | 7.23E-02 | 72 | 1489 | -0.21 | 0.20 | 2.98E-01 | 4.47E-01 | 182 | -0.33 | 0.14 | 1.54E-02 | 0.00 | 3.98E-01 | - | - |
| Pain during the last four weeks | General health | 191 | 0.08 | 0.24 | 7.39E-01 | 8.45E-01 | 239 | 162 | 0.47 | 0.17 | 6.84E-03 | **3.06E-02** | 61 | 0.33 | 0.14 | 1.60E-02 | 0.45 | 1.77E-01 | - | - |
| Vitamin B6 intake (mg/day) | Nutrients | 13019 | 0.03 | 0.02 | 1.36E-01 | 2.73E-01 | 136 | 12782 | 0.04 | 0.02 | 5.46E-02 | 1.33E-01 | 112 | 0.04 | 0.01 | 1.61E-02 | 0.00 | 7.23E-01 | - | - |
| Control over own work assignment | Psychosocial | 14612 | -0.03 | 0.02 | 6.33E-02 | 1.56E-01 | 111 | 14337 | -0.03 | 0.02 | 1.29E-01 | 2.40E-01 | 146 | -0.03 | 0.01 | 1.67E-02 | 0.00 | 8.56E-01 | - | - |
| Monounsaturated fat intake (g/day) | Nutrients | 13019 | 0.04 | 0.02 | 6.11E-02 | 1.55E-01 | 107 | 12782 | 0.03 | 0.02 | 1.45E-01 | 2.61E-01 | 151 | 0.04 | 0.02 | 1.83E-02 | 0.00 | 8.07E-01 | - | - |
| Learn new things at job | Psychosocial | 14589 | -0.04 | 0.02 | 1.44E-02 | 6.15E-02 | 64 | 14333 | -0.02 | 0.02 | 4.04E-01 | 5.57E-01 | 198 | -0.03 | 0.01 | 1.89E-02 | 0.15 | 2.78E-01 | - | - |
| Cambridge physical activity index | Physical activity | 14345 | -0.04 | 0.02 | 2.00E-02 | 7.28E-02 | 74 | 14054 | -0.02 | 0.02 | 3.35E-01 | 4.92E-01 | 186 | -0.03 | 0.01 | 1.90E-02 | 0.00 | 3.60E-01 | - | - |
| Folic acid intake (ug/day) | Nutrients | 13019 | 0.03 | 0.02 | 9.47E-02 | 2.12E-01 | 121 | 12782 | 0.04 | 0.02 | 1.03E-01 | 2.12E-01 | 132 | 0.03 | 0.01 | 1.95E-02 | 0.00 | 9.87E-01 | - | - |
| Vitamin D intake (ug/day) | Nutrients | 13019 | 0.04 | 0.02 | 5.99E-02 | 1.54E-01 | 106 | 12782 | 0.03 | 0.02 | 1.62E-01 | 2.83E-01 | 156 | 0.03 | 0.01 | 2.00E-02 | 0.00 | 7.72E-01 | - | - |
| For how much of the time during the last four weeks have you felt full of energy? | General health | 191 | -0.16 | 0.24 | 5.03E-01 | 6.57E-01 | 209 | 161 | -0.42 | 0.18 | 1.82E-02 | 5.92E-02 | 84 | -0.33 | 0.14 | 2.07E-02 | 0.00 | 3.71E-01 | - | - |
| As healthy as anyone | General health | 191 | 0.10 | 0.24 | 6.72E-01 | 7.91E-01 | 232 | 161 | -0.54 | 0.17 | 2.07E-03 | **1.49E-02** | 38 | -0.32 | 0.14 | 2.23E-02 | 0.79 | 2.89E-02 | - | - |
| Oil for cooking | Food | 13019 | 0.05 | 0.02 | 8.21E-03 | **4.46E-02** | 49 | 12782 | 0.01 | 0.02 | 5.89E-01 | 6.96E-01 | 231 | 0.03 | 0.01 | 2.24E-02 | 0.52 | 1.50E-01 | - | - |
| Number of friends that can come to your home at any time and feel at home | Social | 14592 | -0.03 | 0.02 | 1.07E-01 | 2.35E-01 | 123 | 14345 | -0.03 | 0.02 | 1.12E-01 | 2.21E-01 | 138 | -0.03 | 0.01 | 2.35E-02 | 0.00 | 9.68E-01 | - | - |
| How much has the pain during the last four weeks disturbed your normal work? | General health | 190 | 0.03 | 0.24 | 9.08E-01 | 9.46E-01 | 261 | 162 | 0.46 | 0.17 | 7.86E-03 | **3.41E-02** | 63 | 0.31 | 0.14 | 2.47E-02 | 0.55 | 1.36E-01 | - | - |
| Average portion size of vegetables based on photographic illustration of four sizes (smallest to largest) | Food | 13019 | -0.02 | 0.02 | 2.61E-01 | 4.36E-01 | 163 | 12782 | -0.04 | 0.02 | 3.93E-02 | 1.07E-01 | 100 | -0.03 | 0.01 | 2.52E-02 | 0.00 | 4.80E-01 | - | - |
| Overall state of health compared to other your age | General health | 12734 | -0.05 | 0.02 | 6.08E-03 | **3.46E-02** | 48 | 12542 | -0.01 | 0.02 | 6.99E-01 | 7.79E-01 | 245 | -0.03 | 0.01 | 2.54E-02 | 0.63 | 1.01E-01 | - | - |
| Polyunsaturated fat intake (g/day) | Nutrients | 13019 | 0.05 | 0.02 | 1.18E-02 | 5.58E-02 | 57 | 12782 | 0.01 | 0.02 | 5.68E-01 | 6.84E-01 | 227 | 0.03 | 0.01 | 2.68E-02 | 0.43 | 1.84E-01 | - | - |
| Liver, kidney | Food | 6555 | 0.06 | 0.03 | 1.87E-02 | 7.17E-02 | 71 | 6423 | 0.02 | 0.03 | 4.63E-01 | 5.99E-01 | 210 | 0.04 | 0.02 | 2.77E-02 | 0.18 | 2.68E-01 | - | - |
| Frequency of shoverling snow during leisure time | Physical activity | 13900 | -0.02 | 0.02 | 2.39E-01 | 4.11E-01 | 159 | 13692 | -0.04 | 0.02 | 5.44E-02 | 1.33E-01 | 111 | -0.03 | 0.01 | 2.92E-02 | 0.00 | 5.65E-01 | - | - |
| Campestanol intake (mg/day) | Nutrients | 13019 | -0.03 | 0.02 | 1.53E-01 | 2.98E-01 | 140 | 12782 | -0.03 | 0.02 | 9.73E-02 | 2.04E-01 | 129 | -0.03 | 0.01 | 2.93E-02 | 0.00 | 8.38E-01 | - | - |
| For how much of the time during the last four weeks have you felt calm and serene? | General health | 191 | -0.21 | 0.23 | 3.67E-01 | 5.36E-01 | 187 | 162 | -0.35 | 0.17 | 4.53E-02 | 1.18E-01 | 105 | -0.30 | 0.14 | 3.08E-02 | 0.00 | 6.33E-01 | - | - |
| Disaccharides intake (g/day) | Nutrients | 13019 | 0.03 | 0.02 | 8.01E-02 | 1.82E-01 | 120 | 12782 | 0.03 | 0.02 | 2.05E-01 | 3.40E-01 | 165 | 0.03 | 0.01 | 3.24E-02 | 0.00 | 7.65E-01 | - | - |
| Liquor, spirits | Alcohol | 13019 | 0.02 | 0.02 | 3.04E-01 | 4.80E-01 | 172 | 12782 | 0.04 | 0.02 | 5.62E-02 | 1.36E-01 | 113 | 0.03 | 0.01 | 3.88E-02 | 0.00 | 5.09E-01 | - | - |
| Linoleic acid intake (g/day) | Nutrients | 13019 | 0.05 | 0.02 | 1.21E-02 | 5.58E-02 | 59 | 12782 | 0.01 | 0.02 | 7.22E-01 | 8.01E-01 | 246 | 0.03 | 0.01 | 3.94E-02 | 0.54 | 1.39E-01 | - | - |
| Monosaccharides intake (g/day) | Nutrients | 13019 | 0.03 | 0.02 | 2.06E-01 | 3.75E-01 | 150 | 12782 | 0.03 | 0.02 | 1.05E-01 | 2.14E-01 | 134 | 0.03 | 0.01 | 4.18E-02 | 0.00 | 7.69E-01 | - | - |
| Steak, chop, e.g. | Food | 13019 | 0.03 | 0.02 | 1.95E-01 | 3.59E-01 | 148 | 12782 | 0.03 | 0.02 | 1.28E-01 | 2.40E-01 | 145 | 0.03 | 0.01 | 4.68E-02 | 0.00 | 8.33E-01 | - | - |
| Physical limitation to participate in steneous activities: running, lifting heavy objects, taking part in physically demanding sports | General health | 190 | 0.14 | 0.24 | 5.77E-01 | 7.19E-01 | 219 | 162 | 0.38 | 0.18 | 4.23E-02 | 1.13E-01 | 102 | 0.29 | 0.15 | 4.86E-02 | 0.00 | 4.33E-01 | - | - |
| White meat (poultry) | Food | 13019 | 0.02 | 0.02 | 3.63E-01 | 5.34E-01 | 185 | 12782 | 0.04 | 0.02 | 5.84E-02 | 1.40E-01 | 114 | 0.03 | 0.01 | 4.91E-02 | 0.00 | 4.64E-01 | - | - |
| Memory status | Psychosocial | 6897 | -0.05 | 0.03 | 6.13E-02 | 1.55E-01 | 108 | 6784 | -0.02 | 0.03 | 3.85E-01 | 5.42E-01 | 194 | -0.04 | 0.02 | 5.08E-02 | 0.00 | 5.06E-01 | - | - |
| Blood based food | Food | 6555 | 0.08 | 0.03 | 3.59E-03 | **2.28E-02** | 43 | 6423 | -0.01 | 0.03 | 8.15E-01 | 8.76E-01 | 254 | 0.04 | 0.02 | 5.23E-02 | 0.79 | 2.89E-02 | - | - |
| Emotional problems that made you do less than you wanted during the last four weeks | General health | 190 | 0.99 | 0.72 | 1.71E-01 | 3.23E-01 | 144 | 161 | 0.68 | 0.50 | 1.75E-01 | 3.02E-01 | 158 | 0.78 | 0.41 | 5.72E-02 | 0.00 | 7.18E-01 | - | - |
| Soft cheese | Food | 6555 | 0.04 | 0.03 | 1.67E-01 | 3.18E-01 | 143 | 6423 | 0.04 | 0.03 | 1.96E-01 | 3.27E-01 | 164 | 0.04 | 0.02 | 5.84E-02 | 0.00 | 9.74E-01 | - | - |
| Cohabitation: Live alone vs Only children | Social | 1952 | -0.14 | 0.12 | 2.53E-01 | 4.27E-01 | 162 | 1922 | -0.22 | 0.14 | 1.18E-01 | 2.28E-01 | 140 | -0.18 | 0.09 | 5.87E-02 | 0.00 | 6.69E-01 | - | - |
| Beta-sitostanol intake (mg/day) | Nutrients | 13019 | -0.02 | 0.02 | 2.95E-01 | 4.74E-01 | 170 | 12782 | -0.03 | 0.02 | 1.03E-01 | 2.12E-01 | 133 | -0.03 | 0.01 | 5.95E-02 | 0.00 | 6.54E-01 | - | - |
| Vitamin E intake (mg/day) | Nutrients | 13019 | 0.02 | 0.02 | 3.22E-01 | 4.93E-01 | 178 | 12782 | 0.03 | 0.02 | 9.41E-02 | 2.01E-01 | 128 | 0.03 | 0.01 | 6.05E-02 | 0.00 | 6.08E-01 | - | - |
| Frequency of walking during leisure time | Physical activity | 14205 | -0.02 | 0.02 | 3.96E-01 | 5.63E-01 | 192 | 13995 | -0.04 | 0.02 | 7.31E-02 | 1.65E-01 | 121 | -0.03 | 0.01 | 6.34E-02 | 0.00 | 4.86E-01 | - | - |
| Cheese 10-17% | Food | 13019 | 0.04 | 0.02 | 4.21E-02 | 1.17E-01 | 98 | 12782 | 0.01 | 0.02 | 5.73E-01 | 6.85E-01 | 228 | 0.03 | 0.01 | 6.36E-02 | 0.01 | 3.15E-01 | - | - |
| Milk, sour milk (1,5%) | Beverage | 13019 | -0.03 | 0.02 | 1.20E-01 | 2.54E-01 | 129 | 12782 | -0.02 | 0.02 | 2.93E-01 | 4.44E-01 | 180 | -0.03 | 0.01 | 6.45E-02 | 0.00 | 7.51E-01 | - | - |
| Frequent social contacts with colleagues during work | Psychosocial | 14058 | -0.01 | 0.02 | 5.32E-01 | 6.78E-01 | 214 | 13814 | -0.04 | 0.02 | 4.46E-02 | 1.17E-01 | 104 | -0.02 | 0.01 | 6.47E-02 | 0.01 | 3.14E-01 | - | - |
| Secoisolariciresinol intake (ug/day) | Nutrients | 13019 | -0.01 | 0.02 | 4.94E-01 | 6.57E-01 | 204 | 12782 | -0.04 | 0.02 | 5.10E-02 | 1.26E-01 | 110 | -0.03 | 0.01 | 6.49E-02 | 0.00 | 3.51E-01 | - | - |
| Participation in sports or physical exercise associations | Social | 777 | -0.42 | 0.18 | 2.04E-02 | 7.28E-02 | 75 | 690 | -0.07 | 0.16 | 6.70E-01 | 7.65E-01 | 239 | -0.22 | 0.12 | 6.54E-02 | 0.54 | 1.39E-01 | - | - |
| Would you say that the number of people that you meet in your everyday life is enough or would you like to meet more or fewer people? | Social | 14748 | 0.01 | 0.02 | 5.21E-01 | 6.68E-01 | 213 | 14495 | 0.04 | 0.02 | 4.74E-02 | 1.21E-01 | 107 | 0.02 | 0.01 | 6.63E-02 | 0.00 | 3.25E-01 | - | - |
| Plant based protein intake (g/day) | Nutrients | 13019 | -0.03 | 0.02 | 1.75E-01 | 3.28E-01 | 146 | 12782 | -0.03 | 0.02 | 2.18E-01 | 3.54E-01 | 168 | -0.03 | 0.01 | 6.72E-02 | 0.00 | 9.59E-01 | - | - |
| Sweets | Food | 13019 | 0.03 | 0.02 | 1.07E-01 | 2.35E-01 | 124 | 12782 | 0.02 | 0.02 | 3.42E-01 | 4.97E-01 | 188 | 0.03 | 0.01 | 6.88E-02 | 0.00 | 6.63E-01 | - | - |
| For how much of the time during the last four weeks have you felt tired? | General health | 191 | 0.18 | 0.24 | 4.53E-01 | 6.21E-01 | 199 | 161 | 0.29 | 0.18 | 9.95E-02 | 2.07E-01 | 131 | 0.25 | 0.14 | 7.48E-02 | 0.00 | 7.10E-01 | - | - |
| Sucrose intake (g/day) | Nutrients | 13019 | 0.03 | 0.02 | 1.10E-01 | 2.41E-01 | 125 | 12782 | 0.02 | 0.02 | 3.69E-01 | 5.28E-01 | 191 | 0.03 | 0.01 | 7.60E-02 | 0.00 | 6.48E-01 | - | - |
| Do you feel important and appreciated outside your home? | Psychosocial | 6919 | -0.03 | 0.03 | 2.27E-01 | 3.98E-01 | 156 | 6799 | -0.04 | 0.03 | 1.94E-01 | 3.25E-01 | 163 | -0.03 | 0.02 | 7.66E-02 | 0.00 | 9.20E-01 | - | - |
| Alcohol intake (g/day) | Alcohol | 13019 | -0.03 | 0.02 | 1.23E-01 | 2.55E-01 | 132 | 12782 | -0.02 | 0.02 | 3.46E-01 | 4.99E-01 | 189 | -0.03 | 0.01 | 7.78E-02 | 0.00 | 6.94E-01 | - | - |
| For how much of the time during the last four weeks have you felt very nervous? | General health | 192 | 0.20 | 0.23 | 3.91E-01 | 5.62E-01 | 190 | 162 | 0.27 | 0.18 | 1.27E-01 | 2.40E-01 | 144 | 0.25 | 0.14 | 8.13E-02 | 0.00 | 8.15E-01 | - | - |
| For how much of the time during the last four weeks have you felt so depressed that nothing could cheer you up? | General health | 192 | 0.48 | 0.23 | 4.04E-02 | 1.16E-01 | 95 | 161 | 0.11 | 0.18 | 5.49E-01 | 6.72E-01 | 223 | 0.24 | 0.14 | 8.25E-02 | 0.38 | 2.05E-01 | - | - |
| Frequency of gardening during leisure time | Physical activity | 13811 | -0.01 | 0.02 | 5.21E-01 | 6.68E-01 | 212 | 13569 | -0.04 | 0.02 | 6.59E-02 | 1.51E-01 | 119 | -0.02 | 0.01 | 8.25E-02 | 0.00 | 3.77E-01 | - | - |
| Brown beans, pea soup | Food | 13019 | 0.02 | 0.02 | 2.72E-01 | 4.43E-01 | 167 | 12782 | 0.03 | 0.02 | 1.86E-01 | 3.14E-01 | 162 | 0.02 | 0.01 | 8.77E-02 | 0.00 | 8.48E-01 | - | - |
| Physical limitation that limited your ability to perform certain work tasks or other activities during the last four weeks | General health | 189 | -0.20 | 0.65 | 7.62E-01 | 8.46E-01 | 245 | 160 | 1.27 | 0.52 | 1.66E-02 | 5.65E-02 | 80 | 0.69 | 0.41 | 8.98E-02 | 0.68 | 7.90E-02 | - | - |
| Grams of tobacco smoked per week | Tobacco use | 9143 | 0.04 | 0.02 | 6.32E-02 | 1.56E-01 | 110 | 8921 | 0.01 | 0.02 | 6.80E-01 | 7.70E-01 | 241 | 0.03 | 0.02 | 1.00E-01 | 0.00 | 3.37E-01 | - | - |
| Butter on bread | Food | 13019 | 0.04 | 0.02 | 4.94E-02 | 1.30E-01 | 104 | 12782 | 0.01 | 0.02 | 7.73E-01 | 8.51E-01 | 248 | 0.02 | 0.01 | 1.06E-01 | 0.25 | 2.49E-01 | - | - |
| Meat stew | Food | 13019 | 0.02 | 0.02 | 2.53E-01 | 4.27E-01 | 161 | 12782 | 0.02 | 0.02 | 2.54E-01 | 3.99E-01 | 174 | 0.02 | 0.01 | 1.06E-01 | 0.00 | 9.72E-01 | - | - |
| Linolenic acid intake (g/day) | Nutrients | 13019 | 0.03 | 0.02 | 7.70E-02 | 1.82E-01 | 115 | 12782 | 0.01 | 0.02 | 6.74E-01 | 7.67E-01 | 240 | 0.02 | 0.01 | 1.17E-01 | 0.00 | 3.58E-01 | - | - |
| Soft whey cheese | Food | 6555 | -0.04 | 0.03 | 1.54E-01 | 2.98E-01 | 141 | 6423 | -0.02 | 0.03 | 4.60E-01 | 5.99E-01 | 209 | -0.03 | 0.02 | 1.24E-01 | 0.00 | 6.46E-01 | - | - |

| | | | | | | | | | | | | | | | | | | | | |
|---|---|---|---|---|---|---|---|---|---|---|---|---|---|---|---|---|---|---|---|---|
| Berries (fresh or frozen) | Food | 13019 | -0.01 | 0.02 | 5.57E-01 | 7.00E-01 | 217 | 12782 | -0.03 | 0.02 | 1.12E-01 | 2.21E-01 | 137 | -0.02 | 0.01 | 1.27E-01 | 0.00 | 4.61E-01 | - | - |
| For how much of the time during the last four weeks has your physical health or your emotional problems limited your ability to interact with others? | General health | 189 | 0.14 | 0.24 | 5.44E-01 | 6.91E-01 | 215 | 161 | 0.26 | 0.18 | 1.49E-01 | 2.65E-01 | 154 | 0.22 | 0.14 | 1.28E-01 | 0.00 | 7.00E-01 | - | - |
| Frequency of dancing during leisure time | Physical activity | 12804 | 0.05 | 0.02 | 8.33E-03 | **4.46E-02** | 50 | 12563 | -0.01 | 0.02 | 5.64E-01 | 6.81E-01 | 226 | 0.02 | 0.01 | 1.31E-01 | 0.80 | 2.50E-02 | - | - |
| Physical limitation that made you do less than you wanted during the last four weeks | General health | 189 | 0.40 | 0.56 | 4.76E-01 | 6.47E-01 | 201 | 161 | 0.60 | 0.45 | 1.81E-01 | 3.11E-01 | 159 | 0.53 | 0.35 | 1.35E-01 | 0.00 | 7.81E-01 | - | - |
| Meat on bread | Food | 13019 | 0.02 | 0.02 | 4.30E-01 | 5.95E-01 | 197 | 12782 | 0.03 | 0.02 | 1.85E-01 | 3.14E-01 | 161 | 0.02 | 0.01 | 1.37E-01 | 0.00 | 6.83E-01 | - | - |
| Docosahexaenoic acid (DHA) intake (g/day) | Nutrients | 13019 | 0.02 | 0.02 | 2.98E-01 | 4.75E-01 | 171 | 12782 | 0.02 | 0.02 | 2.90E-01 | 4.42E-01 | 179 | 0.02 | 0.01 | 1.38E-01 | 0.00 | 9.65E-01 | - | - |
| Light and physically active work | Physical activity | 14390 | -0.04 | 0.04 | 3.32E-01 | 5.06E-01 | 179 | 14098 | -0.05 | 0.04 | 2.66E-01 | 4.13E-01 | 176 | -0.05 | 0.03 | 1.41E-01 | 0.00 | 9.02E-01 | - | - |
| Snuff status: Former snuff users vs non-snuff users | Tobacco use | 11475 | -0.08 | 0.06 | 2.18E-01 | 3.91E-01 | 152 | 11344 | -0.05 | 0.07 | 4.08E-01 | 5.60E-01 | 199 | -0.07 | 0.05 | 1.44E-01 | 0.00 | 7.86E-01 | - | - |
| Smoking status: Former occasional smokers vs non-smokers | Tobacco use | 8461 | 0.05 | 0.06 | 3.95E-01 | 5.63E-01 | 191 | 8290 | 0.08 | 0.07 | 2.23E-01 | 3.60E-01 | 169 | 0.07 | 0.05 | 1.45E-01 | 0.00 | 7.70E-01 | - | - |
| Lean fish (e.g. perch, bass, cod) | Food | 13019 | 0.03 | 0.02 | 1.34E-01 | 2.70E-01 | 135 | 12782 | 0.01 | 0.02 | 5.95E-01 | 6.98E-01 | 233 | 0.02 | 0.01 | 1.47E-01 | 0.00 | 5.13E-01 | - | - |
| Saturated fat intake (g/day) | Nutrients | 13019 | -0.02 | 0.02 | 3.46E-01 | 5.20E-01 | 181 | 12782 | -0.02 | 0.02 | 2.71E-01 | 4.19E-01 | 177 | -0.02 | 0.01 | 1.50E-01 | 0.00 | 8.88E-01 | - | - |
| Rice | Food | 13019 | 0.01 | 0.02 | 6.99E-01 | 8.19E-01 | 233 | 12782 | 0.03 | 0.02 | 9.23E-02 | 2.01E-01 | 123 | 0.02 | 0.01 | 1.50E-01 | 0.00 | 3.41E-01 | - | - |
| Confidence status | Psychosocial | 6908 | -0.03 | 0.03 | 2.25E-01 | 3.97E-01 | 155 | 6791 | -0.02 | 0.03 | 4.17E-01 | 5.67E-01 | 201 | -0.03 | 0.02 | 1.51E-01 | 0.00 | 8.00E-01 | - | - |
| Breakfast habits: Coffee/tea and wheat buns or rusk for breakfast vs not breakfast at all | Food | 1006 | -0.08 | 0.18 | 6.57E-01 | 7.87E-01 | 228 | 981 | -0.25 | 0.17 | 1.30E-01 | 2.41E-01 | 147 | -0.18 | 0.12 | 1.56E-01 | 0.00 | 4.89E-01 | - | - |
| Frequency of hunting or fishing during leisure time | Physical activity | 13433 | 0.01 | 0.02 | 5.93E-01 | 7.31E-01 | 221 | 13231 | 0.03 | 0.02 | 1.41E-01 | 2.56E-01 | 150 | 0.02 | 0.01 | 1.59E-01 | 0.00 | 4.92E-01 | - | - |
| Eicosapentaenoic acid (EPA) intake (g/day) | Nutrients | 13019 | 0.02 | 0.02 | 3.57E-01 | 5.30E-01 | 184 | 12782 | 0.02 | 0.02 | 3.04E-01 | 4.53E-01 | 183 | 0.02 | 0.01 | 1.69E-01 | 0.00 | 9.17E-01 | - | - |
| Corn flakes | Food | 13019 | -0.04 | 0.02 | 3.94E-02 | 1.14E-01 | 94 | 12782 | 0.00 | 0.02 | 8.57E-01 | 8.96E-01 | 261 | -0.02 | 0.01 | 1.74E-01 | 0.59 | 1.19E-01 | - | - |
| Boiled coffee | Beverage | 13019 | 0.02 | 0.02 | 2.20E-01 | 3.92E-01 | 153 | 12782 | 0.01 | 0.02 | 5.08E-01 | 6.36E-01 | 218 | 0.02 | 0.01 | 1.79E-01 | 0.00 | 7.10E-01 | - | - |
| Marital status: Single vs Widow/widower | Social | 1549 | 0.29 | 0.31 | 3.46E-01 | 5.20E-01 | 182 | 1525 | 0.27 | 0.29 | 3.42E-01 | 4.97E-01 | 187 | 0.28 | 0.21 | 1.81E-01 | 0.00 | 9.74E-01 | - | - |
| Possibility to speak with colleagues during breaks | Psychosocial | 14159 | -0.01 | 0.02 | 5.17E-01 | 6.68E-01 | 210 | 13904 | -0.02 | 0.02 | 2.14E-01 | 3.50E-01 | 167 | -0.02 | 0.01 | 1.84E-01 | 0.00 | 6.55E-01 | - | - |
| Skill demand from job | Psychosocial | 14563 | -0.01 | 0.02 | 4.84E-01 | 6.51E-01 | 203 | 14321 | -0.02 | 0.02 | 2.48E-01 | 3.94E-01 | 171 | -0.02 | 0.01 | 1.92E-01 | 0.00 | 7.24E-01 | - | - |
| Blota (broth + bread) | Food | 6555 | 0.05 | 0.03 | 4.53E-02 | 1.21E-01 | 102 | 6423 | -0.01 | 0.03 | 8.38E-01 | 8.87E-01 | 257 | 0.02 | 0.02 | 1.95E-01 | 0.58 | 1.24E-01 | - | - |
| Pasta | Food | 13019 | 0.01 | 0.02 | 7.62E-01 | 8.46E-01 | 246 | 12782 | 0.03 | 0.02 | 1.22E-01 | 2.34E-01 | 142 | 0.02 | 0.01 | 1.96E-01 | 0.00 | 3.66E-01 | - | - |
| Physical limitation that made you not being able to perform certain work tasks or other activities during the last four weeks | General health | 188 | -0.61 | 0.61 | 3.22E-01 | 4.93E-01 | 177 | 160 | 1.01 | 0.44 | 2.38E-02 | 7.41E-02 | 87 | 0.45 | 0.36 | 2.05E-01 | 0.78 | 3.22E-02 | - | - |
| Strong beer | Alcohol | 13019 | 0.01 | 0.02 | 6.35E-01 | 7.67E-01 | 226 | 12782 | 0.03 | 0.02 | 1.84E-01 | 3.14E-01 | 160 | 0.02 | 0.01 | 2.06E-01 | 0.00 | 5.35E-01 | - | - |
| Travel to work: Irregular travel mode to work vs passive travel to work | Physical activity | 9626 | -0.10 | 0.07 | 1.30E-01 | 2.64E-01 | 134 | 9477 | -0.02 | 0.07 | 8.20E-01 | 8.78E-01 | 255 | -0.06 | 0.05 | 2.08E-01 | 0.00 | 3.82E-01 | - | - |
| Participation in other association | Social | 777 | 0.51 | 0.18 | 5.64E-03 | **3.27E-02** | 47 | 690 | -0.11 | 0.16 | 4.65E-01 | 5.99E-01 | 212 | 0.15 | 0.12 | 2.09E-01 | 0.85 | 9.84E-03 | - | - |
| Job demands to work very fast | Psychosocial | 14607 | 0.02 | 0.02 | 2.62E-01 | 4.36E-01 | 164 | 14349 | 0.01 | 0.02 | 5.48E-01 | 6.72E-01 | 222 | 0.02 | 0.01 | 2.21E-01 | 0.00 | 7.31E-01 | - | - |
| Physical limitation to participate in moderately demanding activities: bending down or kneeling | General health | 189 | -0.08 | 0.24 | 7.26E-01 | 8.36E-01 | 237 | 162 | 0.31 | 0.18 | 7.79E-02 | 1.74E-01 | 122 | 0.17 | 0.14 | 2.26E-01 | 0.45 | 1.79E-01 | - | - |
| Sometimes physically straining work | Physical activity | 14390 | 0.04 | 0.04 | 3.09E-01 | 4.80E-01 | 175 | 14098 | 0.02 | 0.04 | 5.78E-01 | 6.86E-01 | 230 | 0.03 | 0.03 | 2.63E-01 | 0.00 | 7.63E-01 | - | - |
| Trans fat intake (g/day) | Nutrients | 13019 | 0.02 | 0.02 | 5.00E-01 | 6.57E-01 | 207 | 12782 | 0.02 | 0.03 | 3.76E-01 | 5.34E-01 | 192 | 0.02 | 0.02 | 2.71E-01 | 0.00 | 8.70E-01 | - | - |
| Salty fish | Food | 13019 | 0.01 | 0.02 | 6.64E-01 | 7.91E-01 | 229 | 12782 | 0.02 | 0.02 | 2.57E-01 | 4.00E-01 | 175 | 0.02 | 0.01 | 2.72E-01 | 0.00 | 6.04E-01 | - | - |
| Sedentary or standing work | Physical activity | 14390 | 0.04 | 0.04 | 3.64E-01 | 5.34E-01 | 186 | 14098 | 0.03 | 0.04 | 5.61E-01 | 6.80E-01 | 225 | 0.03 | 0.03 | 2.90E-01 | 0.00 | 8.33E-01 | - | - |
| Rosehip, sweet syrup soup | Food | 13019 | 0.02 | 0.02 | 4.12E-01 | 5.74E-01 | 196 | 12782 | 0.01 | 0.02 | 5.02E-01 | 6.35E-01 | 214 | 0.01 | 0.01 | 2.91E-01 | 0.00 | 9.34E-01 | - | - |
| Coffee rolls/buns, rusk | Food | 13019 | 0.02 | 0.02 | 2.42E-01 | 4.12E-01 | 160 | 12782 | -0.06 | 0.02 | 6.45E-03 | **2.98E-02** | 59 | -0.01 | 0.01 | 2.93E-01 | 0.87 | 5.56E-03 | - | - |
| Carbohydrates intake (g/day) | Nutrients | 13019 | -0.02 | 0.02 | 4.02E-01 | 5.66E-01 | 194 | 12782 | -0.01 | 0.02 | 5.53E-01 | 6.75E-01 | 224 | -0.01 | 0.01 | 3.10E-01 | 0.00 | 8.79E-01 | - | - |
| Beta-sitosterol intake (mg/day) | Nutrients | 13019 | 0.02 | 0.02 | 3.10E-01 | 4.80E-01 | 176 | 12782 | 0.01 | 0.02 | 6.91E-01 | 7.79E-01 | 242 | 0.01 | 0.01 | 3.14E-01 | 0.00 | 6.75E-01 | - | - |
| Brewed (filtered) coffee | Beverage | 13019 | 0.01 | 0.02 | 4.97E-01 | 6.57E-01 | 206 | 12782 | 0.01 | 0.02 | 4.59E-01 | 5.99E-01 | 208 | 0.01 | 0.01 | 3.16E-01 | 0.00 | 9.49E-01 | - | - |
| Fibre intake (g/day) | Nutrients | 13019 | -0.02 | 0.02 | 4.58E-01 | 6.25E-01 | 200 | 12782 | -0.01 | 0.02 | 5.04E-01 | 6.35E-01 | 216 | -0.01 | 0.01 | 3.18E-01 | 0.00 | 9.75E-01 | - | - |
| Frequncy of social contacts with colleagues during leisure time | Psychosocial | 13865 | 0.00 | 0.02 | 7.91E-01 | 8.68E-01 | 249 | 13610 | 0.02 | 0.02 | 2.48E-01 | 3.94E-01 | 172 | 0.01 | 0.01 | 3.21E-01 | 0.00 | 5.18E-01 | - | - |
| Physical limitation to participate in moderately demanding activities: moving a table, vacuuming, walking in the forest or gardening | General health | 190 | -0.15 | 0.24 | 5.20E-01 | 6.68E-01 | 211 | 162 | 0.30 | 0.18 | 9.24E-02 | 2.01E-01 | 124 | 0.14 | 0.14 | 3.25E-01 | 0.57 | 1.28E-01 | - | - |
| Physical limitation that reduced the normal time spent at work or in other activities during the last four weeks | General health | 190 | -0.83 | 0.75 | 2.67E-01 | 4.40E-01 | 166 | 161 | 1.02 | 0.52 | 5.04E-02 | 1.26E-01 | 109 | 0.42 | 0.43 | 3.26E-01 | 0.76 | 4.13E-02 | - | - |
| Average portion size of potatoes/rice/pasta based on photographic illustration of four sizes (smallest to largest) | Food | 13019 | -0.03 | 0.02 | 1.65E-01 | 3.17E-01 | 142 | 12782 | 0.00 | 0.02 | 9.58E-01 | 9.73E-01 | 269 | -0.01 | 0.02 | 3.36E-01 | 0.00 | 3.16E-01 | - | - |
| Boiled or baked potato | Food | 13019 | -0.02 | 0.02 | 3.40E-01 | 5.16E-01 | 180 | 12782 | -0.01 | 0.02 | 6.99E-01 | 7.79E-01 | 244 | -0.01 | 0.01 | 3.39E-01 | 0.00 | 7.02E-01 | - | - |
| Satisfaction with leisure time | Psychosocial | 6914 | -0.04 | 0.03 | 1.22E-01 | 2.54E-01 | 131 | 6778 | 0.01 | 0.03 | 8.03E-01 | 8.70E-01 | 252 | -0.02 | 0.02 | 3.44E-01 | 0.36 | 2.12E-01 | - | - |
| Permanent employment | Psychosocial | 14487 | 0.02 | 0.04 | 5.94E-01 | 7.31E-01 | 222 | 14266 | -0.08 | 0.04 | 6.42E-02 | 1.50E-01 | 116 | -0.03 | 0.03 | 3.75E-01 | 0.66 | 8.74E-02 | - | - |
| Sugar, honey, marmelade, jam | Food | 13019 | 0.00 | 0.02 | 9.84E-01 | 9.92E-01 | 271 | 12782 | -0.03 | 0.02 | 2.07E-01 | 3.41E-01 | 166 | -0.01 | 0.01 | 3.75E-01 | 0.00 | 3.70E-01 | - | - |
| Self rate of overall health compared to a year ago | General health | 191 | -0.07 | 0.24 | 7.70E-01 | 8.51E-01 | 247 | 160 | -0.15 | 0.17 | 4.01E-01 | 5.55E-01 | 197 | -0.12 | 0.14 | 3.95E-01 | 0.00 | 7.90E-01 | - | - |
| Contradictory demands in job | Psychosocial | 14498 | -0.01 | 0.02 | 5.02E-01 | 6.57E-01 | 208 | 14238 | -0.01 | 0.02 | 6.16E-01 | 7.16E-01 | 235 | -0.01 | 0.01 | 4.05E-01 | 0.00 | 9.19E-01 | - | - |
| Get sick more often than other people | General health | 191 | 0.03 | 0.23 | 9.08E-01 | 9.46E-01 | 262 | 160 | 0.16 | 0.18 | 3.85E-01 | 5.42E-01 | 193 | 0.11 | 0.14 | 4.46E-01 | 0.00 | 6.62E-01 | - | - |
| Teetotaler | Alcohol | 14450 | 0.08 | 0.06 | 2.21E-01 | 3.92E-01 | 154 | 14227 | -0.01 | 0.07 | 8.38E-01 | 8.87E-01 | 256 | 0.03 | 0.04 | 4.50E-01 | 0.00 | 3.25E-01 | - | - |
| Participation in associations or voluntary organisations | Social | 14610 | -0.02 | 0.04 | 5.80E-01 | 7.20E-01 | 220 | 14348 | -0.02 | 0.04 | 6.09E-01 | 7.10E-01 | 234 | -0.02 | 0.03 | 4.51E-01 | 0.00 | 9.92E-01 | - | - |
| Travel to work: Walk to work vs passive travel to work | Physical activity | 9409 | 0.02 | 0.07 | 7.36E-01 | 8.44E-01 | 238 | 9231 | 0.05 | 0.08 | 4.77E-01 | 6.12E-01 | 213 | 0.04 | 0.05 | 4.62E-01 | 0.00 | 7.80E-01 | - | - |
| Light but partly physically active work | Physical activity | 14390 | -0.03 | 0.05 | 4.97E-01 | 6.57E-01 | 205 | 14098 | -0.01 | 0.05 | 7.67E-01 | 8.48E-01 | 247 | -0.02 | 0.03 | 4.87E-01 | 0.00 | 7.97E-01 | - | - |
| Physical limitation to participate in moderately demanding activities: walking more than 2 km | General health | 190 | -0.09 | 0.24 | 7.10E-01 | 8.25E-01 | 235 | 161 | 0.21 | 0.18 | 2.53E-01 | 3.99E-01 | 173 | 0.10 | 0.15 | 4.94E-01 | 0.00 | 3.21E-01 | - | - |
| Enterolactone intake (ug/day) | Nutrients | 13019 | 0.00 | 0.02 | 8.62E-01 | 9.19E-01 | 256 | 12782 | 0.02 | 0.02 | 4.48E-01 | 5.97E-01 | 204 | 0.01 | 0.01 | 5.15E-01 | 0.00 | 6.70E-01 | - | - |
| Phosphate intake (mg/day) | Nutrients | 13019 | 0.00 | 0.02 | 8.77E-01 | 9.28E-01 | 258 | 12782 | 0.02 | 0.02 | 4.48E-01 | 5.97E-01 | 205 | 0.01 | 0.01 | 5.25E-01 | 0.00 | 6.60E-01 | - | - |
| Vitamin B2 intake (ug/day) | Nutrients | 13019 | 0.00 | 0.02 | 9.30E-01 | 9.53E-01 | 265 | 12782 | 0.02 | 0.02 | 4.23E-01 | 5.71E-01 | 202 | 0.01 | 0.01 | 5.36E-01 | 0.00 | 6.04E-01 | - | - |
| Smoking status: Former smokers vs non-smokers | Tobacco use | 10069 | 0.02 | 0.05 | 7.06E-01 | 8.23E-01 | 234 | 9970 | 0.02 | 0.05 | 6.23E-01 | 7.21E-01 | 236 | 0.02 | 0.03 | 5.39E-01 | 0.00 | 9.33E-01 | - | - |
| Enough time for job assignments | Psychosocial | 14555 | 0.01 | 0.02 | 4.51E-01 | 6.21E-01 | 198 | 14282 | 0.00 | 0.02 | 9.24E-01 | 9.48E-01 | 266 | 0.01 | 0.01 | 5.41E-01 | 0.00 | 6.52E-01 | - | - |
| Oatflake, whole wheat, rye or barley porridge | Food | 13019 | -0.01 | 0.02 | 5.74E-01 | 7.18E-01 | 218 | 12782 | -0.01 | 0.02 | 7.84E-01 | 8.56E-01 | 250 | -0.01 | 0.01 | 5.52E-01 | 0.00 | 8.47E-01 | - | - |
| Sum of phytosterols intake (mg/day) | Nutrients | 13019 | 0.02 | 0.02 | 4.12E-01 | 5.74E-01 | 195 | 12782 | 0.00 | 0.02 | 9.89E-01 | 9.90E-01 | 272 | 0.01 | 0.01 | 5.61E-01 | 0.00 | 5.62E-01 | - | - |
| For how much of the time during the last four weeks have you felt worn out? | General health | 191 | -0.22 | 0.24 | 3.76E-01 | 5.43E-01 | 189 | 159 | 0.25 | 0.18 | 1.73E-01 | 3.02E-01 | 157 | 0.08 | 0.14 | 5.68E-01 | 0.57 | 1.27E-01 | - | - |
| Salad dressing with oil | Food | 13019 | 0.00 | 0.02 | 8.43E-01 | 9.06E-01 | 254 | 12782 | -0.01 | 0.02 | 5.44E-01 | 6.72E-01 | 221 | -0.01 | 0.01 | 5.73E-01 | 0.00 | 7.64E-01 | - | - |
| Breakfast habits: Only coffee/tea for breakfast vs not breakfast at all | Food | 809 | 0.00 | 0.31 | 9.96E-01 | 9.96E-01 | 273 | 781 | -0.20 | 0.27 | 4.53E-01 | 5.99E-01 | 206 | -0.11 | 0.20 | 5.76E-01 | 0.00 | 6.17E-01 | - | - |
| Number of social contacts with the same interests as you | Social | 14710 | 0.00 | 0.02 | 8.96E-01 | 9.41E-01 | 260 | 14460 | 0.01 | 0.02 | 5.05E-01 | 6.35E-01 | 217 | 0.01 | 0.01 | 5.79E-01 | 0.00 | 6.95E-01 | - | - |
| Banana | Food | 13019 | 0.00 | 0.02 | 9.68E-01 | 9.79E-01 | 270 | 12782 | -0.02 | 0.02 | 4.64E-01 | 5.99E-01 | 211 | -0.01 | 0.01 | 5.92E-01 | 0.00 | 6.17E-01 | - | - |
| Fatty fish (e.g. herring, whitefish, salmon) | Food | 13019 | 0.01 | 0.02 | 6.15E-01 | 7.49E-01 | 224 | 12782 | 0.00 | 0.02 | 8.13E-01 | 8.76E-01 | 253 | 0.01 | 0.01 | 5.98E-01 | 0.00 | 8.59E-01 | - | - |
| Distance to work in kilometers (one way) | Physical activity | 12915 | -0.01 | 0.02 | 7.20E-01 | 8.33E-01 | 236 | 12750 | 0.02 | 0.02 | 2.84E-01 | 4.36E-01 | 178 | 0.01 | 0.01 | 6.22E-01 | 0.03 | 3.09E-01 | - | - |
| Iron intake (mg/day) | Nutrients | 13019 | 0.02 | 0.02 | 2.72E-01 | 4.43E-01 | 168 | 12782 | -0.01 | 0.02 | 6.56E-01 | 7.53E-01 | 238 | 0.01 | 0.02 | 6.27E-01 | 0.14 | 2.80E-01 | - | - |
| Ice cream | Food | 13019 | 0.02 | 0.02 | 2.08E-01 | 3.77E-01 | 151 | 12782 | -0.01 | 0.02 | 5.35E-01 | 6.64E-01 | 220 | 0.01 | 0.01 | 6.35E-01 | 0.43 | 1.87E-01 | - | - |
| Vision status | General health | 6911 | 0.01 | 0.03 | 7.47E-01 | 8.46E-01 | 240 | 6793 | -0.03 | 0.03 | 3.07E-01 | 4.53E-01 | 185 | -0.01 | 0.02 | 6.38E-01 | 0.00 | 3.36E-01 | - | - |
| Campesterol intake (mg/day) | Nutrients | 13019 | 0.00 | 0.02 | 8.31E-01 | 9.01E-01 | 251 | 12782 | -0.02 | 0.02 | 3.87E-01 | 5.42E-01 | 195 | -0.01 | 0.01 | 6.56E-01 | 0.00 | 4.40E-01 | - | - |
| Apple, pear, peach, orange, mandarin and grapefruit | Food | 13019 | 0.00 | 0.02 | 8.85E-01 | 9.33E-01 | 259 | 12782 | 0.02 | 0.02 | 4.45E-01 | 5.97E-01 | 203 | 0.01 | 0.02 | 6.70E-01 | 0.00 | 5.16E-01 | - | - |
| Number of social interactions during a normal week | Social | 14614 | 0.00 | 0.02 | 9.32E-01 | 9.53E-01 | 267 | 14380 | -0.01 | 0.02 | 5.19E-01 | 6.46E-01 | 219 | -0.01 | 0.01 | 7.00E-01 | 0.00 | 6.00E-01 | - | - |
| Pizza | Food | 13019 | 0.00 | 0.02 | 9.55E-01 | 9.73E-01 | 268 | 12782 | 0.01 | 0.02 | 6.42E-01 | 7.39E-01 | 237 | 0.01 | 0.01 | 7.18E-01 | 0.00 | 7.65E-01 | - | - |
| Possibility to leave your work for a while to speak with a colleague | Psychosocial | 14088 | 0.00 | 0.02 | 8.11E-01 | 8.85E-01 | 250 | 13813 | -0.01 | 0.02 | 4.57E-01 | 5.99E-01 | 207 | 0.00 | 0.01 | 7.30E-01 | 0.00 | 4.84E-01 | - | - |
| Physical limitation to participate in moderately demanding activities: walking up several stairs | General health | 190 | -0.25 | 0.24 | 3.07E-01 | 4.80E-01 | 174 | 160 | 0.22 | 0.19 | 2.31E-01 | 3.71E-01 | 170 | 0.05 | 0.15 | 7.48E-01 | 0.58 | 1.22E-01 | - | - |
| Frequency of engaging in clubs, associations or study circles | Social | 9823 | -0.02 | 0.02 | 2.66E-01 | 4.40E-01 | 165 | 9726 | 0.04 | 0.02 | 1.09E-01 | 2.18E-01 | 136 | 0.00 | 0.02 | 7.62E-01 | 0.73 | 5.39E-02 | - | - |
| Medium beer | Alcohol | 13019 | -0.01 | 0.02 | 6.32E-01 | 7.66E-01 | 225 | 12782 | 0.00 | 0.02 | 9.15E-01 | 9.43E-01 | 265 | 0.00 | 0.01 | 7.86E-01 | 0.00 | 6.82E-01 | - | - |
| Sausage, liver pate on bread | Food | 13019 | 0.01 | 0.02 | 6.67E-01 | 7.91E-01 | 230 | 12782 | 0.00 | 0.02 | 9.51E-01 | 9.69E-01 | 268 | 0.00 | 0.01 | 7.89E-01 | 0.00 | 7.31E-01 | - | - |
| Calcium intake (mg/day) | Nutrients | 13019 | -0.01 | 0.02 | 7.56E-01 | 8.46E-01 | 242 | 12782 | 0.00 | 0.02 | 9.62E-01 | 9.73E-01 | 270 | 0.00 | 0.01 | 7.97E-01 | 0.00 | 8.56E-01 | - | - |
| Margarine for cooking | Food | 13019 | 0.00 | 0.02 | 8.72E-01 | 9.27E-01 | 257 | 12782 | 0.01 | 0.02 | 5.92E-01 | 6.96E-01 | 232 | 0.00 | 0.01 | 7.97E-01 | 0.00 | 6.19E-01 | - | - |
| Whole grain crisp bread | Food | 13019 | -0.01 | 0.02 | 5.49E-01 | 6.94E-01 | 216 | 12782 | 0.01 | 0.02 | 7.94E-01 | 8.64E-01 | 251 | 0.00 | 0.01 | 8.01E-01 | 0.00 | 5.47E-01 | - | - |
| Fat intake (g/day) | Nutrients | 13019 | 0.01 | 0.02 | 6.71E-01 | 7.91E-01 | 231 | 12782 | 0.00 | 0.02 | 9.06E-01 | 9.37E-01 | 264 | 0.00 | 0.01 | 8.22E-01 | 0.00 | 7.04E-01 | - | - |
| Magnesium intake (mg/day) | Nutrients | 13019 | 0.00 | 0.02 | 9.25E-01 | 9.53E-01 | 263 | 12782 | 0.01 | 0.02 | 6.96E-01 | 7.79E-01 | 243 | 0.00 | 0.01 | 8.39E-01 | 0.00 | 7.28E-01 | - | - |
| Zinc intake (mg/day) | Nutrients | 13019 | -0.01 | 0.02 | 6.40E-01 | 7.70E-01 | 227 | 12782 | 0.00 | 0.02 | 8.39E-01 | 8.87E-01 | 258 | 0.00 | 0.01 | 8.43E-01 | 0.00 | 6.38E-01 | - | - |
| For how much of the time during the last four weeks have you felt gloomy and sad? | General health | 190 | -0.30 | 0.25 | 2.29E-01 | 3.98E-01 | 157 | 161 | 0.19 | 0.18 | 2.98E-01 | 4.47E-01 | 181 | 0.02 | 0.14 | 8.85E-01 | 0.60 | 1.12E-01 | - | - |
| Last time a colleague visited you at home | Psychosocial | 14005 | 0.01 | 0.02 | 7.52E-01 | 8.46E-01 | 241 | 13739 | 0.00 | 0.02 | 8.96E-01 | 9.30E-01 | 263 | 0.00 | 0.01 | 8.92E-01 | 0.00 | 7.54E-01 | - | - |

| | | | | | | | | | | | | | | | | | | | | |
|---|---|---|---|---|---|---|---|---|---|---|---|---|---|---|---|---|---|---|---|---|
| Vitamin A intake (mg/day) | Nutrients | 13019 | 0.00 | 0.02 | 8.58E-01 | 9.19E-01 | 255 | 12782 | 0.00 | 0.02 | 9.40E-01 | 9.61E-01 | 267 | 0.00 | 0.02 | 9.40E-01 | 0.00 | 8.59E-01 | - | - |
| Frequency of picking berries or mushrooms during leisure time | Physical activity | 13734 | -0.01 | 0.02 | 4.84E-01 | 6.51E-01 | 202 | 13506 | 0.02 | 0.02 | 4.01E-01 | 5.55E-01 | 196 | 0.00 | 0.01 | 9.43E-01 | 0.16 | 2.75E-01 | - | - |
| Palmitic acid intake (g/day) | Nutrients | 13019 | 0.00 | 0.02 | 9.65E-01 | 9.79E-01 | 269 | 12782 | 0.00 | 0.02 | 8.82E-01 | 9.19E-01 | 262 | 0.00 | 0.01 | 9.44E-01 | 0.00 | 8.91E-01 | - | - |
| White (soft) bread, thin crisp bread | Food | 13019 | 0.00 | 0.02 | 9.32E-01 | 9.53E-01 | 266 | 12782 | 0.00 | 0.02 | 9.90E-01 | 9.90E-01 | 273 | 0.00 | 0.01 | 9.44E-01 | 0.00 | 9.60E-01 | - | - |
| Margarine on bread | Food | 13019 | -0.02 | 0.02 | 3.74E-01 | 5.43E-01 | 188 | 12782 | 0.02 | 0.02 | 4.12E-01 | 5.63E-01 | 200 | 0.00 | 0.01 | 9.46E-01 | 0.31 | 2.28E-01 | - | - |
| Pancake, waffle, Swedish dumpling | Food | 13019 | -0.01 | 0.02 | 7.81E-01 | 8.59E-01 | 248 | 12782 | 0.00 | 0.02 | 8.47E-01 | 8.93E-01 | 259 | 0.00 | 0.01 | 9.47E-01 | 0.00 | 7.40E-01 | - | - |
| High mental demand from job | Psychosocial | 14522 | 0.00 | 0.02 | 9.29E-01 | 9.53E-01 | 264 | 14258 | 0.00 | 0.02 | 9.69E-01 | 9.76E-01 | 271 | 0.00 | 0.01 | 9.70E-01 | 0.00 | 9.28E-01 | - | - |
| Self-employed | Psychosocial | 14364 | -0.01 | 0.06 | 8.36E-01 | 9.02E-01 | 253 | 14131 | 0.02 | 0.07 | 7.83E-01 | 8.56E-01 | 249 | 0.00 | 0.05 | 9.71E-01 | 0.00 | 7.32E-01 | - | - |
| Physical limitation to participate in moderately demanding activities: lifting or carrying grocery bags | General health | 189 | -0.20 | 0.24 | 3.98E-01 | 5.63E-01 | 193 | 162 | 0.12 | 0.18 | 5.04E-01 | 6.35E-01 | 215 | 0.00 | 0.14 | 9.77E-01 | 0.14 | 2.80E-01 | - | - |
| Cholesterol intake (g/day) | Nutrients | 13019 | 0.00 | 0.02 | 8.32E-01 | 9.01E-01 | 252 | 12782 | 0.00 | 0.02 | 8.51E-01 | 8.94E-01 | 260 | 0.00 | 0.01 | 9.81E-01 | 0.00 | 7.78E-01 | - | - |

**Supplementary Table 13**. Longitudinal association results for SBP

| | | Training set | | | | | | Testing set | | | | | | Metanalysis | | | | | R2 | |
|---|---|---|---|---|---|---|---|---|---|---|---|---|---|---|---|---|---|---|---|---|
| Description | Group | N | Effect estimate | S.E. | p-value | p-value$_{FDR}$ | p-value rank | N | Effect estimate | S.E. | p-value | p-value$_{FDR}$ | p-value rank | Effect estimate | S.E. | p-value | $I^2$ | Q p-value | adjusted R2 | R2 rank |
| Informed of having high blood pressure | General health | 14544 | 4.66 | 0.37 | 1.48E-35 | **4.04E-33** | 1 | 14291 | 5.01 | 0.39 | 2.83E-38 | **7.72E-36** | 1 | 4.83 | 0.27 | 2.34E-72 | 0.00 | 5.20E-01 | 0.42 | 1 |
| Educational level | Psychosocial | 14554 | -0.80 | 0.12 | 1.00E-10 | **1.37E-08** | 2 | 14341 | -0.71 | 0.13 | 2.55E-08 | **3.48E-06** | 2 | -0.75 | 0.09 | 1.53E-17 | 0.00 | 6.10E-01 | 0.41 | 2 |
| Parents or siblings had a cerebral hemorrhage/thrombosis or cardiac infarction before the age of 60 | General health | 14356 | 0.97 | 0.31 | 1.55E-03 | **4.24E-02** | 10 | 14131 | 1.35 | 0.32 | 1.86E-05 | **1.69E-03** | 3 | 1.16 | 0.22 | 1.48E-07 | 0.00 | 3.88E-01 | 0.41 | 3 |
| Average portion size of potatoes/rice/pasta based on photographic illustration of four sizes (smallest to largest) | Food | 12871 | -0.57 | 0.14 | 3.29E-05 | **3.00E-03** | 3 | 12635 | -0.39 | 0.14 | 5.17E-03 | 9.40E-02 | 15 | -0.48 | 0.10 | 8.42E-07 | 0.00 | 3.66E-01 | - | - |
| Breakfast habits: Porridge w/o sandwich for breakfast vs not breakfast at all | Food | 1731 | -1.97 | 0.72 | 6.23E-03 | 8.51E-02 | 20 | 1681 | -3.37 | 0.81 | 3.54E-05 | **2.42E-03** | 4 | -2.59 | 0.54 | 1.59E-06 | 0.40 | 1.98E-01 | - | - |
| Total energy intake (kcal/day) | Nutrients | 12871 | -0.51 | 0.14 | 3.52E-04 | **1.60E-02** | 6 | 12635 | -0.43 | 0.15 | 3.96E-03 | 8.73E-02 | 12 | -0.47 | 0.10 | 4.78E-06 | 0.00 | 6.89E-01 | - | - |
| Repetitive job | Psychosocial | 14434 | 0.47 | 0.12 | 1.02E-04 | **6.95E-03** | 4 | 14176 | 0.31 | 0.12 | 1.30E-02 | 1.25E-01 | 26 | 0.39 | 0.09 | 6.15E-06 | 0.00 | 3.58E-01 | - | - |
| Overall state of health compared to other your age | General health | 12575 | -0.46 | 0.13 | 3.25E-04 | **1.60E-02** | 5 | 12392 | -0.32 | 0.13 | 1.43E-02 | 1.30E-01 | 30 | -0.39 | 0.09 | 1.79E-05 | 0.00 | 4.64E-01 | - | - |
| Average portion size of vegetables based on photographic illustration of four sizes (smallest to largest) | Food | 12871 | -0.39 | 0.13 | 2.37E-03 | 5.89E-02 | 11 | 12635 | -0.38 | 0.13 | 4.91E-03 | 9.40E-02 | 14 | -0.38 | 0.09 | 3.47E-05 | 0.00 | 9.22E-01 | - | - |
| Memory status | Psychosocial | 6831 | 0.56 | 0.17 | 9.73E-04 | **2.95E-02** | 9 | 6713 | 0.44 | 0.18 | 1.25E-02 | 1.25E-01 | 22 | 0.50 | 0.12 | 3.96E-05 | 0.00 | 6.27E-01 | - | - |
| Would you say that the number of people that you meet in your everyday life is enough or would you like to meet more or fewer people? | Social | 14579 | -0.33 | 0.12 | 5.63E-03 | 8.51E-02 | 17 | 14332 | -0.37 | 0.12 | 2.53E-03 | 6.91E-02 | 10 | -0.35 | 0.08 | 4.31E-05 | 0.00 | 8.11E-01 | - | - |
| Contradictory demands in job | Psychosocial | 14332 | -0.36 | 0.12 | 2.69E-03 | 6.12E-02 | 12 | 14077 | -0.33 | 0.12 | 7.91E-03 | 1.23E-01 | 17 | -0.34 | 0.09 | 6.22E-05 | 0.00 | 8.61E-01 | - | - |
| Everyday exercise satisfaction | Physical activity | 14456 | 0.42 | 0.12 | 4.81E-04 | **1.88E-02** | 7 | 14203 | 0.25 | 0.12 | 4.69E-02 | 2.81E-01 | 45 | 0.34 | 0.09 | 9.84E-05 | 0.00 | 3.23E-01 | - | - |
| Number of cigarretes smoked per day | Tobacco use | 9385 | 0.42 | 0.14 | 4.02E-03 | 7.84E-02 | 14 | 9140 | 0.39 | 0.15 | 9.92E-03 | 1.25E-01 | 20 | 0.40 | 0.10 | 1.12E-04 | 0.00 | 9.17E-01 | - | - |
| Changed everyday exercise during the last year | Physical activity | 14496 | 0.32 | 0.12 | 6.03E-03 | 8.51E-02 | 19 | 14246 | 0.30 | 0.12 | 1.30E-02 | 1.25E-01 | 27 | 0.31 | 0.08 | 2.15E-04 | 0.00 | 9.03E-01 | - | - |
| Oatflake, whole wheat, rye or barley porridge | Food | 12871 | -0.24 | 0.13 | 5.18E-02 | 2.94E-01 | 48 | 12635 | -0.41 | 0.13 | 1.51E-03 | 5.05E-02 | 7 | -0.32 | 0.09 | 3.10E-04 | 0.00 | 3.58E-01 | - | - |
| If you exercise, change in exercise habits during the last year | Physical activity | 12340 | 0.31 | 0.13 | 1.48E-02 | 1.41E-01 | 28 | 12150 | 0.35 | 0.13 | 8.12E-03 | 1.23E-01 | 18 | 0.33 | 0.09 | 3.30E-04 | 0.00 | 8.15E-01 | - | - |
| Number of snuff boxes per week | Tobacco use | 13796 | 0.24 | 0.13 | 6.03E-02 | 3.29E-01 | 50 | 13597 | 0.42 | 0.13 | 1.40E-03 | 5.05E-02 | 6 | 0.33 | 0.09 | 3.59E-04 | 0.00 | 3.17E-01 | - | - |
| Frequency of gardening during leisure time | Physical activity | 13647 | 0.42 | 0.12 | 6.92E-04 | **2.36E-02** | 8 | 13416 | 0.18 | 0.13 | 1.57E-01 | 5.42E-01 | 79 | 0.30 | 0.09 | 6.20E-04 | 0.45 | 1.79E-01 | - | - |
| Frequency of engaging in clubs, associations or study circles | Social | 9702 | -0.33 | 0.14 | 2.36E-02 | 1.95E-01 | 33 | 9613 | -0.37 | 0.15 | 1.33E-02 | 1.25E-01 | 29 | -0.35 | 0.10 | 8.16E-04 | 0.00 | 8.26E-01 | - | - |
| High physical demand from job | Physical activity | 14475 | 0.33 | 0.12 | 5.52E-03 | 8.51E-02 | 16 | 14217 | 0.23 | 0.12 | 6.52E-02 | 3.56E-01 | 50 | 0.28 | 0.09 | 1.05E-03 | 0.00 | 5.47E-01 | - | - |
| Ice cream | Food | 12871 | -0.19 | 0.13 | 1.34E-01 | 5.32E-01 | 67 | 12635 | -0.41 | 0.13 | 1.62E-03 | 5.05E-02 | 8 | -0.30 | 0.09 | 1.05E-03 | 0.31 | 2.28E-01 | - | - |
| Participation in associations or voluntary organisations | Social | 14442 | -0.74 | 0.25 | 3.30E-03 | 6.93E-02 | 13 | 14186 | -0.41 | 0.26 | 1.14E-01 | 4.63E-01 | 66 | -0.58 | 0.18 | 1.30E-03 | 0.00 | 3.73E-01 | - | - |
| Energy status | Psychosocial | 6844 | 0.38 | 0.17 | 2.58E-02 | 1.99E-01 | 35 | 6709 | 0.40 | 0.18 | 2.35E-02 | 1.89E-01 | 34 | 0.39 | 0.12 | 1.49E-03 | 0.00 | 9.31E-01 | - | - |
| Job demands to work very fast | Psychosocial | 14439 | 0.22 | 0.12 | 6.21E-02 | 3.32E-01 | 51 | 14187 | 0.31 | 0.12 | 1.27E-02 | 1.25E-01 | 24 | 0.26 | 0.09 | 2.10E-03 | 0.00 | 6.24E-01 | - | - |
| Ingenuity or creativity demand from job | Psychosocial | 14388 | -0.25 | 0.12 | 3.54E-02 | 2.30E-01 | 42 | 14144 | -0.27 | 0.12 | 2.95E-02 | 2.24E-01 | 36 | -0.26 | 0.09 | 2.48E-03 | 0.00 | 9.16E-01 | - | - |
| High mental demand from job | Psychosocial | 14356 | 0.32 | 0.12 | 7.30E-03 | 9.06E-02 | 22 | 14096 | 0.18 | 0.12 | 1.41E-01 | 5.14E-01 | 75 | 0.25 | 0.09 | 3.15E-03 | 0.00 | 4.21E-01 | - | - |
| Exercise during the last three months | Physical activity | 14439 | -0.32 | 0.12 | 6.89E-03 | 8.96E-02 | 21 | 14198 | -0.18 | 0.12 | 1.47E-01 | 5.16E-01 | 78 | -0.25 | 0.09 | 3.17E-03 | 0.00 | 4.05E-01 | - | - |
| Cookies, pastry | Food | 12871 | -0.19 | 0.13 | 1.22E-01 | 5.18E-01 | 64 | 12635 | -0.32 | 0.13 | 1.31E-02 | 1.25E-01 | 28 | -0.25 | 0.09 | 4.54E-03 | 0.00 | 4.83E-01 | - | - |
| Meat on bread | Food | 12871 | 0.32 | 0.13 | 1.15E-02 | 1.31E-01 | 24 | 12635 | 0.19 | 0.13 | 1.45E-01 | 5.14E-01 | 77 | 0.25 | 0.09 | 4.66E-03 | 0.00 | 4.77E-01 | - | - |
| Liquor, spirits | Alcohol | 12871 | 0.20 | 0.13 | 1.34E-01 | 5.32E-01 | 69 | 12635 | 0.34 | 0.14 | 1.19E-02 | 1.25E-01 | 21 | 0.27 | 0.09 | 4.66E-03 | 0.00 | 4.50E-01 | - | - |
| Travel to work: Cycle to work vs passive travel to work | Physical activity | 11227 | -0.68 | 0.31 | 2.81E-02 | 1.99E-01 | 37 | 11077 | -0.56 | 0.31 | 7.39E-02 | 3.81E-01 | 53 | -0.62 | 0.22 | 4.81E-03 | 0.00 | 7.93E-01 | - | - |
| Phosphate intake (mg/day) | Nutrients | 12871 | -0.28 | 0.13 | 3.16E-02 | 2.10E-01 | 41 | 12635 | -0.25 | 0.13 | 6.81E-02 | 3.65E-01 | 51 | -0.26 | 0.09 | 4.89E-03 | 0.00 | 8.56E-01 | - | - |
| Learn new things at job | Psychosocial | 14420 | -0.29 | 0.12 | 1.40E-02 | 1.41E-01 | 27 | 14172 | -0.18 | 0.12 | 1.37E-01 | 5.13E-01 | 72 | -0.24 | 0.09 | 5.08E-03 | 0.00 | 5.22E-01 | - | - |
| Snuff status: Snuff users vs non-snuff users | Tobacco use | 12290 | 0.50 | 0.35 | 1.48E-01 | 5.42E-01 | 72 | 12130 | 0.90 | 0.36 | 1.26E-02 | 1.25E-01 | 23 | 0.69 | 0.25 | 5.56E-03 | 0.00 | 4.26E-01 | - | - |
| Pasta | Food | 12871 | -0.29 | 0.13 | 2.84E-02 | 1.99E-01 | 39 | 12635 | -0.22 | 0.14 | 1.07E-01 | 4.58E-01 | 63 | -0.26 | 0.09 | 7.06E-03 | 0.00 | 7.07E-01 | - | - |
| Rice | Food | 12871 | -0.17 | 0.13 | 1.94E-01 | 6.25E-01 | 85 | 12635 | -0.33 | 0.13 | 1.28E-02 | 1.25E-01 | 25 | -0.25 | 0.09 | 7.81E-03 | 0.00 | 3.71E-01 | - | - |
| Satisfaction with home and family situation | Psychosocial | 6853 | 0.38 | 0.17 | 2.54E-02 | 1.99E-01 | 34 | 6724 | 0.26 | 0.18 | 1.43E-01 | 5.14E-01 | 76 | 0.32 | 0.12 | 8.64E-03 | 0.00 | 6.21E-01 | - | - |
| Medium beer | Alcohol | 12871 | 0.11 | 0.13 | 4.02E-01 | 8.44E-01 | 129 | 12635 | 0.38 | 0.13 | 4.16E-03 | 8.73E-02 | 13 | 0.24 | 0.09 | 9.38E-03 | 0.54 | 1.41E-01 | - | - |
| Cheese 28% | Food | 12871 | -0.31 | 0.13 | 1.37E-02 | 1.41E-01 | 26 | 12635 | -0.14 | 0.13 | 2.85E-01 | 6.77E-01 | 115 | -0.23 | 0.09 | 1.20E-02 | 0.00 | 3.42E-01 | - | - |
| Alcohol intake (g/day) | Alcohol | 12871 | 0.17 | 0.13 | 1.92E-01 | 6.23E-01 | 84 | 12635 | 0.30 | 0.13 | 2.43E-02 | 1.89E-01 | 35 | 0.24 | 0.09 | 1.21E-02 | 0.00 | 4.86E-01 | - | - |
| Tea | Beverage | 12871 | -0.35 | 0.13 | 5.76E-03 | 8.51E-02 | 18 | 12635 | -0.10 | 0.13 | 4.50E-01 | 8.28E-01 | 146 | -0.23 | 0.09 | 1.22E-02 | 0.48 | 1.67E-01 | - | - |
| Magnesium intake (mg/day) | Nutrients | 12871 | -0.19 | 0.13 | 1.62E-01 | 5.74E-01 | 77 | 12635 | -0.30 | 0.14 | 3.15E-02 | 2.32E-01 | 37 | -0.24 | 0.10 | 1.23E-02 | 0.00 | 5.69E-01 | - | - |
| Satisfaction with leisure time | Psychosocial | 6849 | 0.33 | 0.17 | 5.13E-02 | 2.94E-01 | 47 | 6708 | 0.28 | 0.18 | 1.14E-01 | 4.63E-01 | 67 | 0.31 | 0.12 | 1.24E-02 | 0.00 | 8.37E-01 | - | - |
| Hamburger | Food | 12871 | 0.17 | 0.13 | 1.85E-01 | 6.17E-01 | 82 | 12635 | 0.28 | 0.13 | 3.31E-02 | 2.32E-01 | 39 | 0.23 | 0.09 | 1.49E-02 | 0.00 | 5.46E-01 | - | - |
| Iodine intake (ug/day) | Nutrients | 12871 | -0.02 | 0.13 | 9.05E-01 | 9.88E-01 | 245 | 12635 | -0.44 | 0.13 | 8.29E-04 | **4.53E-02** | 5 | -0.22 | 0.09 | 1.61E-02 | 0.81 | 2.01E-02 | - | - |
| Fried potatoes, pommes frites | Food | 12871 | 0.25 | 0.13 | 4.72E-02 | 2.80E-01 | 46 | 12635 | 0.17 | 0.13 | 1.81E-01 | 5.73E-01 | 86 | 0.21 | 0.09 | 1.85E-02 | 0.00 | 6.69E-01 | - | - |
| Smoking status: Smokers vs non-smokers | Tobacco use | 9643 | 0.93 | 0.33 | 5.11E-03 | 8.51E-02 | 15 | 9407 | 0.16 | 0.35 | 6.38E-01 | 9.12E-01 | 189 | 0.56 | 0.24 | 1.90E-02 | 0.61 | 1.10E-01 | - | - |
| Participation in other association | Social | 772 | -0.41 | 0.93 | 6.59E-01 | 9.18E-01 | 196 | 689 | -3.18 | 1.06 | 2.82E-03 | 7.01E-02 | 11 | -1.62 | 0.70 | 2.08E-02 | 0.74 | 5.03E-02 | - | - |
| Whole grain intake (g/day) | Food | 12871 | -0.12 | 0.13 | 3.33E-01 | 7.85E-01 | 115 | 12635 | -0.30 | 0.13 | 2.07E-02 | 1.82E-01 | 31 | -0.21 | 0.09 | 2.10E-02 | 0.00 | 3.27E-01 | - | - |
| Eat breakfast from 2000 | Food | 1203 | -1.96 | 1.25 | 1.16E-01 | 5.18E-01 | 61 | 1084 | -2.23 | 1.39 | 1.07E-01 | 4.58E-01 | 64 | -2.08 | 0.93 | 2.47E-02 | 0.00 | 8.84E-01 | - | - |
| Breakfast habits: Coffee/tea and wheat buns or rusk for breakfast vs not breakfast at all | Food | 999 | -0.11 | 1.03 | 9.15E-01 | 9.88E-01 | 252 | 965 | -3.44 | 1.09 | 1.66E-03 | 5.05E-02 | 9 | -1.68 | 0.75 | 2.49E-02 | 0.80 | 2.65E-02 | - | - |
| Support from others | Social | 14584 | -0.28 | 0.12 | 2.07E-02 | 1.76E-01 | 32 | 14354 | -0.10 | 0.12 | 4.21E-01 | 8.20E-01 | 140 | -0.19 | 0.09 | 2.62E-02 | 0.06 | 3.03E-01 | - | - |
| Light but partly physically active work | Physical activity | 14224 | -0.75 | 0.31 | 1.49E-02 | 1.41E-01 | 29 | 13937 | -0.22 | 0.32 | 4.99E-01 | 8.52E-01 | 160 | -0.49 | 0.22 | 2.63E-02 | 0.31 | 2.29E-01 | - | - |
| Physical limitation that made you not being able to perform certain work tasks or other activities during the last four weeks | General health | 188 | 3.64 | 2.89 | 2.09E-01 | 6.34E-01 | 88 | 160 | 4.39 | 2.46 | 7.59E-02 | 3.81E-01 | 54 | 4.08 | 1.87 | 2.94E-02 | 0.00 | 8.43E-01 | - | - |
| Receive hugs to comfort and support you | Social | 14394 | -0.48 | 0.33 | 1.36E-01 | 5.32E-01 | 70 | 14138 | -0.53 | 0.34 | 1.16E-01 | 4.67E-01 | 68 | -0.51 | 0.23 | 3.06E-02 | 0.00 | 9.25E-01 | - | - |
| Frequency of hunting or fishing during leisure time | Physical activity | 13276 | 0.11 | 0.13 | 4.21E-01 | 8.44E-01 | 135 | 13081 | 0.31 | 0.14 | 2.32E-02 | 1.89E-01 | 33 | 0.21 | 0.10 | 3.11E-02 | 0.13 | 2.82E-01 | - | - |
| Brown beans, pea soup | Food | 12871 | 0.21 | 0.13 | 8.72E-02 | 4.07E-01 | 58 | 12635 | 0.17 | 0.13 | 1.86E-01 | 5.73E-01 | 87 | 0.19 | 0.09 | 3.16E-02 | 0.00 | 8.10E-01 | - | - |
| Confidence status | Psychosocial | 6842 | 0.20 | 0.17 | 2.47E-01 | 6.95E-01 | 97 | 6720 | 0.33 | 0.18 | 6.36E-02 | 3.54E-01 | 49 | 0.26 | 0.12 | 3.39E-02 | 0.00 | 5.93E-01 | - | - |
| Chips, popcorn, salted nuts | Food | 12871 | 0.34 | 0.13 | 9.02E-03 | 1.07E-01 | 23 | 12635 | 0.03 | 0.13 | 8.33E-01 | 9.71E-01 | 233 | 0.19 | 0.09 | 4.29E-02 | 0.64 | 9.62E-02 | - | - |
| Breakfast habits: Gruel w/o sandwich for breakfast vs not breakfast at all | Food | 952 | -1.58 | 1.15 | 1.68E-01 | 5.75E-01 | 79 | 901 | -1.97 | 1.33 | 1.39E-01 | 5.13E-01 | 74 | -1.75 | 0.87 | 4.44E-02 | 0.00 | 8.22E-01 | - | - |
| Cohabitation: Live alone vs Only one adult (spouse, partner) | Social | 4529 | 0.89 | 0.52 | 8.62E-02 | 4.07E-01 | 57 | 4629 | 0.57 | 0.53 | 2.83E-01 | 6.77E-01 | 114 | 0.73 | 0.37 | 4.79E-02 | 0.00 | 6.69E-01 | - | - |
| Skill demand from job | Psychosocial | 14395 | -0.11 | 0.12 | 3.57E-01 | 8.24E-01 | 117 | 14158 | -0.23 | 0.12 | 6.15E-02 | 3.50E-01 | 48 | -0.17 | 0.09 | 4.99E-02 | 0.00 | 4.80E-01 | - | - |
| Control over planning and execution of the workday | Psychosocial | 14456 | -0.18 | 0.12 | 1.32E-01 | 5.32E-01 | 66 | 14190 | -0.15 | 0.12 | 2.14E-01 | 6.08E-01 | 96 | -0.17 | 0.09 | 5.15E-02 | 0.00 | 8.79E-01 | - | - |
| Frequency of walking during leisure time | Physical activity | 14040 | 0.01 | 0.13 | 9.24E-01 | 9.88E-01 | 255 | 13839 | 0.35 | 0.13 | 7.09E-03 | 1.21E-01 | 16 | 0.17 | 0.09 | 5.16E-02 | 0.71 | 6.24E-02 | - | - |
| Patience status | Psychosocial | 6847 | 0.26 | 0.17 | 1.21E-01 | 5.18E-01 | 63 | 6722 | 0.21 | 0.18 | 2.40E-01 | 6.44E-01 | 99 | 0.24 | 0.12 | 5.34E-02 | 0.00 | 8.21E-01 | - | - |
| Number of social contacts with the same interests as you | Social | 14542 | -0.24 | 0.12 | 3.93E-02 | 2.50E-01 | 43 | 14297 | -0.07 | 0.12 | 5.44E-01 | 8.59E-01 | 172 | -0.16 | 0.09 | 5.71E-02 | 0.00 | 3.19E-01 | - | - |
| Frequency of picking berries or mushrooms during leisure time | Physical activity | 13571 | 0.07 | 0.13 | 5.98E-01 | 8.97E-01 | 180 | 13356 | 0.28 | 0.13 | 3.30E-02 | 2.32E-01 | 38 | 0.17 | 0.09 | 6.26E-02 | 0.26 | 2.44E-01 | - | - |
| Enough time for job assignments | Psychosocial | 14386 | 0.07 | 0.12 | 5.58E-01 | 8.81E-01 | 169 | 14123 | 0.25 | 0.12 | 4.07E-02 | 2.65E-01 | 42 | 0.16 | 0.09 | 6.51E-02 | 0.12 | 2.88E-01 | - | - |
| Beta-sitosterol intake (mg/day) | Nutrients | 12871 | -0.11 | 0.13 | 3.76E-01 | 8.42E-01 | 122 | 12635 | -0.22 | 0.13 | 9.85E-02 | 4.42E-01 | 59 | -0.16 | 0.09 | 7.40E-02 | 0.00 | 5.71E-01 | - | - |
| Physical limitation that reduced the normal time spent at work or in other activities during the last four weeks | General health | 190 | 2.53 | 3.42 | 4.61E-01 | 8.73E-01 | 144 | 161 | 4.76 | 2.89 | 1.01E-01 | 4.46E-01 | 62 | 3.83 | 2.21 | 8.25E-02 | 0.00 | 6.18E-01 | - | - |
| Physical limitation to participate in moderately demanding activities: lifting or carrying grocery bags | General health | 189 | 1.64 | 1.13 | 1.49E-01 | 5.42E-01 | 75 | 162 | 1.02 | 0.99 | 3.02E-01 | 6.88E-01 | 120 | 1.29 | 0.74 | 8.32E-02 | 0.00 | 6.79E-01 | - | - |
| Physical limitation that made you do less than you wanted during the last four weeks | General health | 189 | 2.92 | 2.59 | 2.62E-01 | 7.11E-01 | 100 | 161 | 3.29 | 2.49 | 1.89E-01 | 5.73E-01 | 89 | 3.11 | 1.80 | 8.34E-02 | 0.00 | 9.18E-01 | - | - |
| Butter on bread | Food | 12871 | 0.10 | 0.12 | 4.24E-01 | 8.44E-01 | 137 | 12635 | 0.21 | 0.13 | 9.87E-02 | 4.42E-01 | 61 | 0.15 | 0.09 | 8.48E-02 | 0.00 | 5.28E-01 | - | - |
| Years smoking | Tobacco use | 12846 | 0.32 | 0.13 | 1.55E-02 | 1.41E-01 | 30 | 12646 | -0.01 | 0.14 | 9.51E-01 | 9.94E-01 | 258 | 0.16 | 0.09 | 8.76E-02 | 0.66 | 8.59E-02 | - | - |
| Blota (broth + bread) | Food | 6466 | 0.31 | 0.18 | 7.84E-02 | 3.82E-01 | 56 | 6344 | 0.11 | 0.18 | 5.30E-01 | 8.59E-01 | 166 | 0.22 | 0.13 | 8.99E-02 | 0.00 | 4.32E-01 | - | - |
| Mood status | Psychosocial | 6846 | 0.17 | 0.17 | 3.21E-01 | 7.75E-01 | 112 | 6719 | 0.25 | 0.18 | 1.64E-01 | 5.56E-01 | 80 | 0.21 | 0.12 | 9.28E-02 | 0.00 | 7.52E-01 | - | - |
| Years using snuff | Tobacco use | 13433 | 0.06 | 0.13 | 6.72E-01 | 9.21E-01 | 199 | 13198 | 0.27 | 0.14 | 4.95E-02 | 2.87E-01 | 47 | 0.16 | 0.10 | 9.49E-02 | 0.20 | 2.63E-01 | - | - |
| Physical limitation that limited your ability to perform certain work tasks or other activities during the last four weeks | General health | 189 | 2.24 | 2.99 | 4.56E-01 | 8.73E-01 | 141 | 160 | 4.69 | 2.93 | 1.12E-01 | 4.63E-01 | 65 | 3.49 | 2.10 | 9.62E-02 | 0.00 | 5.59E-01 | - | - |
| Sum of phytosterols intake (mg/day) | Nutrients | 12871 | -0.08 | 0.13 | 5.18E-01 | 8.81E-01 | 157 | 12635 | -0.22 | 0.13 | 8.67E-02 | 4.15E-01 | 57 | -0.15 | 0.09 | 9.72E-02 | 0.00 | 4.37E-01 | - | - |
| Campesterol intake (mg/day) | Nutrients | 12871 | -0.06 | 0.13 | 6.05E-01 | 8.97E-01 | 183 | 12635 | -0.23 | 0.13 | 7.28E-02 | 3.81E-01 | 52 | -0.15 | 0.09 | 1.05E-01 | 0.00 | 3.53E-01 | - | - |
| Light beer | Alcohol | 12871 | -0.10 | 0.13 | 4.59E-01 | 8.73E-01 | 143 | 12635 | -0.20 | 0.13 | 1.24E-01 | 4.90E-01 | 69 | -0.15 | 0.09 | 1.09E-01 | 0.00 | 5.56E-01 | - | - |
| Vitamin D intake (ug/day) | Nutrients | 12871 | -0.04 | 0.13 | 7.57E-01 | 9.65E-01 | 212 | 12635 | -0.26 | 0.13 | 4.73E-02 | 2.81E-01 | 46 | -0.15 | 0.09 | 1.09E-01 | 0.32 | 2.26E-01 | - | - |
| Permanent employment | Psychosocial | 14319 | 0.69 | 0.28 | 1.20E-02 | 1.31E-01 | 25 | 14107 | -0.09 | 0.29 | 7.47E-01 | 9.47E-01 | 213 | 0.31 | 0.20 | 1.13E-01 | 0.74 | 4.81E-02 | - | - |
| Frequency of shoverling snow during leisure time | Physical activity | 13739 | 0.14 | 0.13 | 2.67E-01 | 7.15E-01 | 102 | 13539 | 0.15 | 0.13 | 2.62E-01 | 6.51E-01 | 110 | 0.14 | 0.09 | 1.15E-01 | 0.00 | 9.75E-01 | - | - |

| | | | | | | | | | | | | | | | | | | | | |
|---|---|---|---|---|---|---|---|---|---|---|---|---|---|---|---|---|---|---|---|---|
| Travel to work: Walk to work vs passive travel to work | Physical activity | 9311 | -0.65 | 0.46 | 1.57E-01 | 5.65E-01 | 76 | 9131 | -0.38 | 0.47 | 4.28E-01 | 8.28E-01 | 141 | -0.52 | 0.33 | 1.17E-01 | 0.00 | 6.76E-01 | - | - |
| Trans fat intake (g/day) | Nutrients | 12871 | -0.22 | 0.16 | 1.67E-01 | 5.75E-01 | 78 | 12635 | -0.13 | 0.16 | 4.32E-01 | 8.28E-01 | 142 | -0.17 | 0.11 | 1.24E-01 | 0.00 | 6.80E-01 | - | - |
| Do you feel important and appreciated in your home? | Psychosocial | 6806 | 0.25 | 0.17 | 1.34E-01 | 5.32E-01 | 68 | 6670 | 0.10 | 0.18 | 5.54E-01 | 8.65E-01 | 175 | 0.18 | 0.12 | 1.36E-01 | 0.00 | 5.39E-01 | - | - |
| Parents or siblings have diabetes | General health | 14380 | 0.23 | 0.32 | 4.78E-01 | 8.81E-01 | 146 | 14154 | 0.45 | 0.33 | 1.70E-01 | 5.59E-01 | 83 | 0.34 | 0.23 | 1.42E-01 | 0.00 | 6.30E-01 | - | - |
| Sometimes physically straining work | Physical activity | 14224 | 0.38 | 0.26 | 1.48E-01 | 5.42E-01 | 73 | 13937 | 0.16 | 0.27 | 5.44E-01 | 8.59E-01 | 173 | 0.28 | 0.19 | 1.43E-01 | 0.00 | 5.71E-01 | - | - |
| Sodium intake (mg/day) | Nutrients | 12871 | 0.01 | 0.13 | 9.68E-01 | 9.94E-01 | 265 | 12635 | -0.28 | 0.13 | 3.93E-02 | 2.62E-01 | 41 | -0.13 | 0.09 | 1.60E-01 | 0.56 | 1.31E-01 | - | - |
| Polyunsaturated fat intake (g/day) | Nutrients | 12871 | -0.09 | 0.13 | 4.96E-01 | 8.81E-01 | 151 | 12635 | -0.17 | 0.13 | 1.90E-01 | 5.73E-01 | 90 | -0.13 | 0.09 | 1.62E-01 | 0.00 | 6.36E-01 | - | - |
| Pain during the last four weeks | General health | 191 | 0.89 | 1.08 | 4.11E-01 | 8.44E-01 | 131 | 162 | 1.09 | 0.97 | 2.62E-01 | 6.51E-01 | 108 | 1.00 | 0.72 | 1.65E-01 | 0.00 | 8.87E-01 | - | - |
| Close relationship with anyone | Social | 14460 | -0.21 | 0.12 | 7.18E-02 | 3.70E-01 | 53 | 14212 | -0.02 | 0.12 | 8.97E-01 | 9.87E-01 | 246 | -0.12 | 0.09 | 1.67E-01 | 0.26 | 2.46E-01 | - | - |
| Linolenic acid intake (g/day) | Nutrients | 12871 | -0.07 | 0.13 | 5.60E-01 | 8.81E-01 | 170 | 12635 | -0.18 | 0.13 | 1.67E-01 | 5.57E-01 | 82 | -0.13 | 0.09 | 1.68E-01 | 0.00 | 5.55E-01 | - | - |
| Snuff status: Former snuff users vs non-snuff users | Tobacco use | 11334 | -0.59 | 0.41 | 1.49E-01 | 5.42E-01 | 74 | 11206 | -0.20 | 0.43 | 6.42E-01 | 9.12E-01 | 190 | -0.40 | 0.30 | 1.73E-01 | 0.00 | 5.04E-01 | - | - |
| Fiber cereals | Food | 12871 | -0.21 | 0.13 | 9.08E-02 | 4.13E-01 | 60 | 12635 | -0.03 | 0.13 | 8.43E-01 | 9.71E-01 | 237 | -0.12 | 0.09 | 1.77E-01 | 0.07 | 3.00E-01 | - | - |
| Disaccharides intake (g/day) | Nutrients | 12871 | 0.01 | 0.13 | 9.33E-01 | 9.88E-01 | 257 | 12635 | -0.27 | 0.13 | 4.35E-02 | 2.76E-01 | 43 | -0.12 | 0.09 | 1.79E-01 | 0.56 | 1.32E-01 | - | - |
| Overall state of health during the last year | General health | 14479 | 0.13 | 0.12 | 2.90E-01 | 7.53E-01 | 105 | 14227 | 0.10 | 0.12 | 4.01E-01 | 8.03E-01 | 136 | 0.12 | 0.09 | 1.79E-01 | 0.00 | 8.93E-01 | - | - |
| Frequncy of social contacts with colleagues during leisure time | Psychosocial | 13704 | -0.03 | 0.12 | 8.37E-01 | 9.80E-01 | 233 | 13459 | -0.21 | 0.13 | 9.14E-02 | 4.30E-01 | 58 | -0.12 | 0.09 | 1.85E-01 | 0.12 | 2.86E-01 | - | - |
| Number of cigarretes smoked per day (in groups) | Tobacco use | 11785 | 0.31 | 0.13 | 2.00E-02 | 1.76E-01 | 31 | 11545 | -0.07 | 0.14 | 6.17E-01 | 9.09E-01 | 183 | 0.13 | 0.09 | 1.86E-01 | 0.74 | 4.79E-02 | - | - |
| Lean fish (e.g. perch, bass, cod) | Food | 12871 | -0.01 | 0.13 | 9.07E-01 | 9.88E-01 | 248 | 12635 | 0.26 | 0.13 | 4.52E-02 | 2.80E-01 | 44 | 0.12 | 0.09 | 1.93E-01 | 0.57 | 1.27E-01 | - | - |
| Whole grain crisp bread | Food | 12871 | 0.24 | 0.13 | 6.40E-02 | 3.36E-01 | 52 | 12635 | -0.01 | 0.13 | 9.65E-01 | 9.94E-01 | 263 | 0.12 | 0.09 | 1.94E-01 | 0.43 | 1.86E-01 | - | - |
| Number of people with whom you can speak openly | Social | 14574 | -0.13 | 0.12 | 2.63E-01 | 7.11E-01 | 101 | 14336 | -0.08 | 0.12 | 4.97E-01 | 8.52E-01 | 159 | -0.11 | 0.08 | 2.02E-01 | 0.00 | 7.71E-01 | - | - |
| Linoleic acid intake (g/day) | Nutrients | 12871 | -0.06 | 0.13 | 6.09E-01 | 8.97E-01 | 184 | 12635 | -0.17 | 0.13 | 1.91E-01 | 5.73E-01 | 91 | -0.12 | 0.09 | 2.02E-01 | 0.00 | 5.56E-01 | - | - |
| White (soft) bread, thin crisp bread | Food | 12871 | 0.28 | 0.13 | 2.65E-02 | 1.99E-01 | 36 | 12635 | -0.06 | 0.13 | 6.32E-01 | 9.12E-01 | 188 | 0.12 | 0.09 | 2.06E-01 | 0.72 | 5.93E-02 | - | - |
| Breakfast habits: Only coffee/tea for breakfast vs not breakfast at all | Food | 804 | -1.05 | 1.71 | 5.39E-01 | 8.81E-01 | 165 | 773 | -2.00 | 1.75 | 2.53E-01 | 6.44E-01 | 107 | -1.51 | 1.22 | 2.15E-01 | 0.00 | 6.97E-01 | - | - |
| Work shifts/weekends | Psychosocial | 13669 | 0.13 | 0.28 | 6.48E-01 | 9.10E-01 | 194 | 13422 | 0.37 | 0.28 | 1.95E-01 | 5.77E-01 | 92 | 0.24 | 0.20 | 2.17E-01 | 0.00 | 5.45E-01 | - | - |
| Cambridge physical activity index | Physical activity | 14179 | -0.01 | 0.12 | 9.06E-01 | 9.88E-01 | 246 | 13893 | -0.21 | 0.12 | 9.86E-02 | 4.42E-01 | 60 | -0.11 | 0.09 | 2.18E-01 | 0.18 | 2.68E-01 | - | - |
| Calcium intake (mg/day) | Nutrients | 12871 | -0.22 | 0.13 | 7.81E-02 | 3.82E-01 | 55 | 12635 | 0.01 | 0.13 | 9.59E-01 | 9.94E-01 | 260 | -0.11 | 0.09 | 2.19E-01 | 0.37 | 2.06E-01 | - | - |
| Soft whey cheese | Food | 6466 | -0.09 | 0.18 | 6.34E-01 | 8.97E-01 | 193 | 6344 | -0.23 | 0.18 | 2.09E-01 | 6.08E-01 | 94 | -0.16 | 0.13 | 2.22E-01 | 0.00 | 5.75E-01 | - | - |
| Feel the need to reduce alcohol consumption | Alcohol | 12897 | 0.34 | 0.41 | 4.00E-01 | 8.44E-01 | 128 | 12744 | 0.36 | 0.41 | 3.88E-01 | 7.91E-01 | 134 | 0.35 | 0.29 | 2.28E-01 | 0.00 | 9.81E-01 | - | - |
| Vitamin C intake (mg/day) | Nutrients | 12871 | -0.01 | 0.13 | 9.64E-01 | 9.94E-01 | 263 | 12635 | 0.24 | 0.13 | 7.67E-02 | 3.81E-01 | 55 | 0.11 | 0.09 | 2.30E-01 | 0.41 | 1.93E-01 | - | - |
| Apple, pear, peach, orange, mandarin and grapefruit | Food | 12871 | -0.16 | 0.14 | 2.39E-01 | 6.87E-01 | 95 | 12635 | -0.07 | 0.14 | 6.18E-01 | 9.09E-01 | 185 | -0.12 | 0.10 | 2.33E-01 | 0.00 | 6.44E-01 | - | - |
| Vision status | General health | 6845 | -0.05 | 0.18 | 7.77E-01 | 9.67E-01 | 219 | 6722 | -0.26 | 0.19 | 1.65E-01 | 5.56E-01 | 81 | -0.15 | 0.13 | 2.45E-01 | 0.00 | 4.18E-01 | - | - |
| Eicosapentaenoic acid (EPA) intake (g/day) | Nutrients | 12871 | -0.11 | 0.13 | 3.69E-01 | 8.32E-01 | 121 | 12635 | -0.10 | 0.13 | 4.63E-01 | 8.28E-01 | 151 | -0.10 | 0.09 | 2.48E-01 | 0.00 | 9.22E-01 | - | - |
| Bacon | Food | 12871 | 0.18 | 0.13 | 1.42E-01 | 5.42E-01 | 71 | 12635 | 0.02 | 0.13 | 8.85E-01 | 9.87E-01 | 241 | 0.10 | 0.09 | 2.48E-01 | 0.00 | 3.59E-01 | - | - |
| Vitamin B2 intake (ug/day) | Nutrients | 12871 | -0.14 | 0.13 | 2.78E-01 | 7.37E-01 | 103 | 12635 | -0.07 | 0.13 | 6.05E-01 | 9.07E-01 | 182 | -0.10 | 0.09 | 2.54E-01 | 0.00 | 7.03E-01 | - | - |
| Satisfaction with work situation | Psychosocial | 6788 | 0.15 | 0.17 | 3.87E-01 | 8.44E-01 | 123 | 6656 | 0.13 | 0.18 | 4.57E-01 | 8.28E-01 | 148 | 0.14 | 0.12 | 2.55E-01 | 0.00 | 9.50E-01 | - | - |
| Steak, chop, e.g. | Food | 12871 | 0.13 | 0.13 | 2.97E-01 | 7.59E-01 | 107 | 12635 | 0.07 | 0.13 | 5.88E-01 | 9.00E-01 | 178 | 0.10 | 0.09 | 2.60E-01 | 0.00 | 7.39E-01 | - | - |
| Wine | Alcohol | 12871 | 0.12 | 0.13 | 3.34E-01 | 7.85E-01 | 116 | 12635 | 0.08 | 0.13 | 5.35E-01 | 8.59E-01 | 167 | 0.10 | 0.09 | 2.60E-01 | 0.00 | 8.17E-01 | - | - |
| Possibility to speak with colleagues during breaks | Psychosocial | 13993 | 0.06 | 0.12 | 6.31E-01 | 8.97E-01 | 191 | 13746 | 0.14 | 0.12 | 2.66E-01 | 6.53E-01 | 111 | 0.10 | 0.09 | 2.63E-01 | 0.00 | 6.41E-01 | - | - |
| Margarine on bread | Food | 12871 | 0.10 | 0.13 | 4.07E-01 | 8.44E-01 | 130 | 12635 | 0.09 | 0.13 | 4.72E-01 | 8.30E-01 | 155 | 0.10 | 0.09 | 2.73E-01 | 0.00 | 9.47E-01 | - | - |
| Cheese 10-17% | Food | 12871 | -0.13 | 0.13 | 3.16E-01 | 7.75E-01 | 111 | 12635 | 0.33 | 0.13 | 9.85E-03 | 1.25E-01 | 19 | 0.10 | 0.09 | 2.79E-01 | 0.85 | 1.08E-02 | - | - |
| Smoked fish/meat | Food | 12871 | 0.23 | 0.12 | 6.00E-02 | 3.29E-01 | 49 | 12635 | -0.05 | 0.13 | 6.75E-01 | 9.28E-01 | 197 | 0.10 | 0.09 | 2.88E-01 | 0.61 | 1.08E-01 | - | - |
| How much has the pain during the last four weeks disturbed your normal work? | General health | 190 | 0.79 | 1.08 | 4.66E-01 | 8.78E-01 | 145 | 162 | 0.71 | 0.97 | 4.64E-01 | 8.28E-01 | 153 | 0.75 | 0.72 | 3.01E-01 | 0.00 | 9.58E-01 | - | - |
| Butter for cooking | Food | 12871 | 0.07 | 0.13 | 5.76E-01 | 8.86E-01 | 177 | 12635 | 0.11 | 0.13 | 3.80E-01 | 7.88E-01 | 129 | 0.09 | 0.09 | 3.11E-01 | 0.00 | 8.11E-01 | - | - |
| Satisfaction with accomodation | Psychosocial | 6857 | 0.08 | 0.17 | 6.50E-01 | 9.10E-01 | 195 | 6728 | 0.17 | 0.18 | 3.36E-01 | 7.34E-01 | 125 | 0.12 | 0.12 | 3.20E-01 | 0.00 | 7.05E-01 | - | - |
| For how much of the time during the last four weeks have you felt gloomy and sad? | General health | 190 | -0.72 | 1.14 | 5.28E-01 | 8.81E-01 | 160 | 161 | -0.74 | 0.98 | 4.51E-01 | 8.28E-01 | 147 | -0.73 | 0.74 | 3.24E-01 | 0.00 | 9.87E-01 | - | - |
| Average portion size of meat/fish based on photographic illustration of four sizes (smallest to largest) | Food | 12871 | -0.10 | 0.14 | 4.90E-01 | 8.81E-01 | 150 | 12635 | -0.10 | 0.14 | 4.96E-01 | 8.52E-01 | 158 | -0.10 | 0.10 | 3.33E-01 | 0.00 | 9.94E-01 | - | - |
| Worsen in health in the future | General health | 188 | 0.63 | 1.08 | 5.62E-01 | 8.81E-01 | 174 | 160 | 0.74 | 1.00 | 4.59E-01 | 8.28E-01 | 149 | 0.69 | 0.73 | 3.47E-01 | 0.00 | 9.40E-01 | - | - |
| Physical limitation to participate in steneous activities: running, lifting heavy objects, taking part in physically demanding sports | General health | 190 | 0.08 | 1.12 | 9.46E-01 | 9.90E-01 | 261 | 162 | 1.25 | 1.03 | 2.28E-01 | 6.34E-01 | 98 | 0.71 | 0.76 | 3.48E-01 | 0.00 | 4.42E-01 | - | - |
| White meat (poultry) | Food | 12871 | 0.14 | 0.14 | 3.03E-01 | 7.65E-01 | 108 | 12635 | -0.30 | 0.13 | 2.15E-02 | 1.83E-01 | 32 | -0.09 | 0.09 | 3.52E-01 | 0.82 | 1.92E-02 | - | - |
| Meat stew | Food | 12871 | 0.00 | 0.12 | 1.00E+00 | 1.00E+00 | 273 | 12635 | -0.17 | 0.13 | 1.79E-01 | 5.73E-01 | 85 | -0.08 | 0.09 | 3.52E-01 | 0.00 | 3.33E-01 | - | - |
| Strong beer | Alcohol | 12871 | 0.06 | 0.13 | 6.71E-01 | 9.21E-01 | 198 | 12635 | 0.12 | 0.14 | 3.74E-01 | 7.88E-01 | 127 | 0.09 | 0.10 | 3.54E-01 | 0.00 | 7.37E-01 | - | - |
| Marital status: Single vs Divorced/separated | Social | 2336 | -0.14 | 0.66 | 8.34E-01 | 9.80E-01 | 232 | 2294 | -0.78 | 0.70 | 2.62E-01 | 6.51E-01 | 109 | -0.44 | 0.48 | 3.56E-01 | 0.00 | 5.02E-01 | - | - |
| Last time a colleague visited you at home | Psychosocial | 13840 | 0.13 | 0.12 | 2.93E-01 | 7.55E-01 | 106 | 13583 | 0.03 | 0.12 | 8.20E-01 | 9.71E-01 | 230 | 0.08 | 0.09 | 3.62E-01 | 0.00 | 5.70E-01 | - | - |
| Lariciresinol intake (ug/day) | Nutrients | 12871 | 0.01 | 0.13 | 9.69E-01 | 9.94E-01 | 266 | 12635 | -0.18 | 0.14 | 1.88E-01 | 5.73E-01 | 88 | -0.08 | 0.09 | 3.74E-01 | 0.00 | 3.32E-01 | - | - |
| Mixed frozen vegetables | Food | 6466 | 0.25 | 0.18 | 1.71E-01 | 5.75E-01 | 81 | 6344 | -0.02 | 0.19 | 9.00E-01 | 9.87E-01 | 249 | 0.12 | 0.13 | 3.74E-01 | 0.09 | 2.93E-01 | - | - |
| Medioresinol intake (ug/day) | Nutrients | 12871 | 0.16 | 0.13 | 2.14E-01 | 6.36E-01 | 92 | 12635 | 0.00 | 0.13 | 9.79E-01 | 9.94E-01 | 268 | 0.08 | 0.09 | 3.83E-01 | 0.00 | 3.76E-01 | - | - |
| Sedentary or standing work | Physical activity | 14224 | 0.02 | 0.27 | 9.37E-01 | 9.88E-01 | 259 | 13937 | 0.33 | 0.28 | 2.46E-01 | 6.44E-01 | 101 | 0.17 | 0.19 | 3.88E-01 | 0.00 | 4.36E-01 | - | - |
| Feel uneasy or guilty because of your way of drinking | Alcohol | 12896 | -0.84 | 0.39 | 3.07E-02 | 2.09E-01 | 40 | 12725 | 0.39 | 0.40 | 3.26E-01 | 7.17E-01 | 124 | -0.24 | 0.28 | 3.90E-01 | 0.80 | 2.69E-02 | - | - |
| Do you feel important and appreciated outside your home? | Psychosocial | 6853 | 0.04 | 0.17 | 8.15E-01 | 9.67E-01 | 229 | 6728 | 0.17 | 0.18 | 3.22E-01 | 7.14E-01 | 123 | 0.10 | 0.12 | 3.92E-01 | 0.00 | 5.82E-01 | - | - |
| Monosaccharides intake (g/day) | Nutrients | 12871 | -0.17 | 0.13 | 2.09E-01 | 6.34E-01 | 89 | 12635 | 0.01 | 0.14 | 9.44E-01 | 9.94E-01 | 256 | -0.08 | 0.10 | 3.94E-01 | 0.00 | 3.55E-01 | - | - |
| Participation in study circles | Social | 772 | -0.13 | 1.35 | 9.21E-01 | 9.88E-01 | 253 | 689 | -1.71 | 1.48 | 2.49E-01 | 6.44E-01 | 105 | -0.85 | 1.00 | 3.94E-01 | 0.00 | 4.32E-01 | - | - |
| Sleep status | Sleep | 6859 | 0.05 | 0.17 | 7.66E-01 | 9.65E-01 | 216 | 6732 | 0.15 | 0.18 | 3.84E-01 | 7.88E-01 | 133 | 0.10 | 0.12 | 4.14E-01 | 0.00 | 6.72E-01 | - | - |
| Sour milk, yoghurt (3% fat) | Food | 12871 | -0.27 | 0.13 | 2.83E-02 | 1.99E-01 | 38 | 12635 | 0.15 | 0.13 | 2.41E-01 | 6.44E-01 | 100 | -0.07 | 0.09 | 4.50E-01 | 0.82 | 1.78E-02 | - | - |
| Oil for cooking | Food | 12871 | -0.03 | 0.13 | 7.96E-01 | 9.67E-01 | 221 | 12635 | -0.11 | 0.13 | 4.13E-01 | 8.11E-01 | 139 | -0.07 | 0.09 | 4.51E-01 | 0.00 | 6.80E-01 | - | - |
| Sodas, soft drinks, juice | Beverage | 12871 | 0.08 | 0.13 | 5.15E-01 | 8.81E-01 | 156 | 12635 | 0.05 | 0.13 | 6.94E-01 | 9.31E-01 | 202 | 0.07 | 0.09 | 4.59E-01 | 0.00 | 8.64E-01 | - | - |
| Frequency of cycling during leisure time | Physical activity | 12306 | 0.04 | 0.13 | 7.64E-01 | 9.65E-01 | 215 | 12120 | 0.10 | 0.13 | 4.64E-01 | 8.28E-01 | 152 | 0.07 | 0.09 | 4.69E-01 | 0.00 | 7.49E-01 | - | - |
| Soft cheese | Food | 6466 | 0.03 | 0.18 | 8.53E-01 | 9.88E-01 | 235 | 6344 | 0.15 | 0.18 | 3.99E-01 | 8.03E-01 | 135 | 0.09 | 0.13 | 4.70E-01 | 0.00 | 6.38E-01 | - | - |
| Sour milk, yoghurt (low fat) | Food | 12871 | -0.25 | 0.13 | 4.51E-02 | 2.80E-01 | 44 | 12635 | 0.13 | 0.13 | 3.00E-01 | 6.88E-01 | 118 | -0.06 | 0.09 | 4.75E-01 | 0.78 | 3.23E-02 | - | - |
| Arachidonic acid (ARA) intake (g/day) | Nutrients | 12871 | 0.16 | 0.13 | 2.08E-01 | 6.34E-01 | 87 | 12635 | -0.04 | 0.13 | 7.83E-01 | 9.47E-01 | 225 | 0.07 | 0.09 | 4.76E-01 | 0.13 | 2.84E-01 | - | - |
| White cabbage, lettuce, lettuce cabbage, spinach, borecole | Food | 12871 | -0.07 | 0.13 | 5.97E-01 | 8.97E-01 | 179 | 12635 | -0.07 | 0.14 | 6.32E-01 | 9.12E-01 | 187 | -0.07 | 0.10 | 4.76E-01 | 0.00 | 9.80E-01 | - | - |
| Self rate of overall health compared to a year ago | General health | 191 | 0.70 | 1.09 | 5.23E-01 | 8.81E-01 | 159 | 160 | -1.44 | 0.95 | 1.33E-01 | 5.13E-01 | 70 | -0.51 | 0.72 | 4.76E-01 | 0.54 | 1.39E-01 | - | - |
| Equol intake (ug/day) | Nutrients | 12871 | -0.16 | 0.13 | 2.11E-01 | 6.34E-01 | 91 | 12635 | 0.04 | 0.13 | 7.84E-01 | 9.47E-01 | 226 | -0.06 | 0.09 | 4.79E-01 | 0.12 | 2.87E-01 | - | - |
| Number of cigars smoked per day | Tobacco use | 9016 | 0.02 | 0.15 | 9.14E-01 | 9.88E-01 | 251 | 8798 | 0.14 | 0.15 | 3.75E-01 | 7.88E-01 | 128 | 0.07 | 0.11 | 4.89E-01 | 0.00 | 5.72E-01 | - | - |
| Pinoresinol intake (ug/day) | Nutrients | 12871 | -0.01 | 0.13 | 9.13E-01 | 9.88E-01 | 250 | 12635 | -0.12 | 0.13 | 3.83E-01 | 7.88E-01 | 131 | -0.06 | 0.09 | 4.92E-01 | 0.00 | 5.83E-01 | - | - |
| Sum of Lariciresinol, Matairesinol, Pinoresinol, Secoisolariciresinol intake (ug/day) | Nutrients | 12871 | 0.02 | 0.13 | 8.74E-01 | 9.88E-01 | 237 | 12635 | -0.15 | 0.13 | 2.51E-01 | 6.44E-01 | 106 | -0.06 | 0.09 | 4.93E-01 | 0.00 | 3.51E-01 | - | - |
| Get sick more often than other people | General health | 191 | -0.84 | 1.07 | 4.36E-01 | 8.50E-01 | 140 | 160 | -0.21 | 0.98 | 8.32E-01 | 9.71E-01 | 232 | -0.50 | 0.72 | 4.93E-01 | 0.00 | 6.67E-01 | - | - |
| Pizza | Food | 12871 | 0.00 | 0.13 | 9.72E-01 | 9.94E-01 | 267 | 12635 | -0.14 | 0.14 | 3.09E-01 | 6.94E-01 | 121 | -0.07 | 0.10 | 4.96E-01 | 0.00 | 4.49E-01 | - | - |
| Docosahexaenoic acid (DHA) intake (g/day) | Nutrients | 12871 | -0.07 | 0.13 | 5.68E-01 | 8.81E-01 | 176 | 12635 | -0.05 | 0.13 | 7.00E-01 | 9.31E-01 | 204 | -0.06 | 0.09 | 4.98E-01 | 0.00 | 9.04E-01 | - | - |
| Margarine for cooking | Food | 12871 | -0.15 | 0.13 | 2.23E-01 | 6.48E-01 | 94 | 12635 | 0.04 | 0.13 | 7.79E-01 | 9.47E-01 | 224 | -0.06 | 0.09 | 4.98E-01 | 0.10 | 2.93E-01 | - | - |
| Control over own work assignment | Psychosocial | 14443 | 0.01 | 0.12 | 9.11E-01 | 9.88E-01 | 249 | 14174 | -0.13 | 0.12 | 2.76E-01 | 6.73E-01 | 112 | -0.06 | 0.09 | 4.99E-01 | 0.00 | 3.89E-01 | - | - |
| For how much of the time during the last four weeks have you felt tired? | General health | 191 | -0.14 | 1.12 | 9.03E-01 | 9.88E-01 | 243 | 161 | -0.75 | 0.99 | 4.49E-01 | 8.28E-01 | 145 | -0.48 | 0.74 | 5.16E-01 | 0.00 | 6.80E-01 | - | - |
| For how much of the time during the last four weeks has your physical health or your emotional problems limited your ability to interact with others? | General health | 189 | -0.41 | 1.08 | 7.03E-01 | 9.31E-01 | 206 | 161 | -0.51 | 0.98 | 6.03E-01 | 9.07E-01 | 181 | -0.47 | 0.73 | 5.21E-01 | 0.00 | 9.46E-01 | - | - |
| Long-term sickness | General health | 13230 | -0.01 | 0.37 | 9.84E-01 | 1.00E+00 | 268 | 12944 | 0.37 | 0.40 | 3.45E-01 | 7.48E-01 | 126 | 0.17 | 0.27 | 5.26E-01 | 0.00 | 4.84E-01 | - | - |
| Vitamin B3 intake (mg/day) | Nutrients | 12871 | 0.26 | 0.13 | 4.62E-02 | 2.80E-01 | 45 | 12635 | -0.15 | 0.13 | 2.47E-01 | 6.44E-01 | 102 | 0.06 | 0.09 | 5.35E-01 | 0.80 | 2.65E-02 | - | - |
| Enterolactone intake (ug/day) | Nutrients | 12871 | -0.15 | 0.12 | 2.17E-01 | 6.37E-01 | 93 | 12635 | 0.05 | 0.13 | 7.01E-01 | 9.31E-01 | 205 | -0.06 | 0.09 | 5.35E-01 | 0.22 | 2.57E-01 | - | - |
| Travel to work: Irregular travel mode to work vs passive travel to work | Physical activity | 9522 | -0.55 | 0.43 | 2.04E-01 | 6.34E-01 | 86 | 9376 | 0.18 | 0.44 | 6.80E-01 | 9.28E-01 | 200 | -0.19 | 0.31 | 5.36E-01 | 0.28 | 2.37E-01 | - | - |
| Bregott on bread | Food | 12871 | 0.02 | 0.13 | 8.84E-01 | 9.88E-01 | 240 | 12635 | -0.13 | 0.13 | 3.02E-01 | 6.88E-01 | 119 | -0.06 | 0.09 | 5.39E-01 | 0.00 | 4.00E-01 | - | - |
| For how much of the time during the last four weeks have you felt full of energy? | General health | 191 | -0.89 | 1.08 | 4.13E-01 | 8.44E-01 | 132 | 161 | -0.08 | 1.00 | 9.39E-01 | 9.94E-01 | 254 | -0.45 | 0.74 | 5.40E-01 | 0.00 | 5.82E-01 | - | - |
| Salty fish | Food | 12871 | -0.11 | 0.13 | 3.95E-01 | 8.44E-01 | 127 | 12635 | 0.00 | 0.13 | 9.86E-01 | 9.94E-01 | 270 | -0.05 | 0.09 | 5.48E-01 | 0.00 | 5.46E-01 | - | - |
| Fibre intake (g/day) | Nutrients | 12871 | 0.09 | 0.13 | 4.97E-01 | 8.81E-01 | 152 | 12635 | 0.02 | 0.14 | 8.91E-01 | 9.87E-01 | 244 | 0.06 | 0.10 | 5.60E-01 | 0.00 | 7.08E-01 | - | - |

| | | | | | | | | | | | | | | | | | | | | |
|---|---|---|---|---|---|---|---|---|---|---|---|---|---|---|---|---|---|---|---|---|
| Hearing status | General health | 6851 | 0.31 | 0.17 | 7.33E-02 | 3.71E-01 | 54 | 6730 | -0.19 | 0.18 | 2.98E-01 | 6.88E-01 | 117 | 0.07 | 0.12 | 5.67E-01 | 0.75 | 4.65E-02 | - | - |
| Physical limitation to participate in moderately demanding activities: walking more than 2 km | General health | 190 | 0.38 | 1.10 | 7.32E-01 | 9.51E-01 | 210 | 161 | 0.47 | 1.04 | 6.49E-01 | 9.12E-01 | 192 | 0.43 | 0.76 | 5.71E-01 | 0.00 | 9.51E-01 | - | - |
| For how much of the time during the last four weeks have you felt very nervous? | General health | 192 | 1.06 | 1.08 | 3.24E-01 | 7.75E-01 | 114 | 162 | -0.14 | 0.98 | 8.90E-01 | 9.87E-01 | 242 | 0.41 | 0.72 | 5.74E-01 | 0.00 | 4.09E-01 | - | - |
| Sugar, honey, marmelade, jam | Food | 12871 | 0.03 | 0.13 | 7.97E-01 | 9.67E-01 | 223 | 12635 | -0.14 | 0.13 | 2.91E-01 | 6.84E-01 | 116 | -0.05 | 0.09 | 5.82E-01 | 0.00 | 3.48E-01 | - | - |
| Boiled or baked potato | Food | 12871 | 0.01 | 0.13 | 9.37E-01 | 9.88E-01 | 258 | 12635 | 0.09 | 0.13 | 4.84E-01 | 8.48E-01 | 156 | 0.05 | 0.09 | 5.85E-01 | 0.00 | 6.56E-01 | - | - |
| Syringaresinol intake (ug/day) | Nutrients | 12871 | 0.12 | 0.13 | 3.61E-01 | 8.24E-01 | 119 | 12635 | -0.02 | 0.13 | 8.72E-01 | 9.87E-01 | 239 | 0.05 | 0.09 | 5.87E-01 | 0.00 | 4.51E-01 | - | - |
| Coffee rolls/buns, rusk | Food | 12871 | 0.02 | 0.13 | 8.89E-01 | 9.88E-01 | 242 | 12635 | 0.08 | 0.13 | 5.27E-01 | 8.59E-01 | 165 | 0.05 | 0.09 | 5.89E-01 | 0.00 | 7.22E-01 | - | - |
| For how much of the time during the last four weeks have you felt calm and serene? | General health | 191 | -0.80 | 1.07 | 4.58E-01 | 8.73E-01 | 142 | 162 | -0.05 | 0.98 | 9.60E-01 | 9.94E-01 | 261 | -0.39 | 0.72 | 5.90E-01 | 0.00 | 6.06E-01 | - | - |
| Corn flakes | Food | 12871 | -0.13 | 0.12 | 3.08E-01 | 7.71E-01 | 109 | 12635 | 0.04 | 0.13 | 7.76E-01 | 9.47E-01 | 222 | -0.05 | 0.09 | 5.94E-01 | 0.00 | 3.60E-01 | - | - |
| Rosehip, sweet syrup soup | Food | 12871 | 0.01 | 0.12 | 9.43E-01 | 9.90E-01 | 260 | 12635 | 0.09 | 0.13 | 4.92E-01 | 8.52E-01 | 157 | 0.05 | 0.09 | 5.97E-01 | 0.00 | 6.57E-01 | - | - |
| Fatty fish (e.g. herring, whitefish, salmon) | Food | 12871 | -0.05 | 0.13 | 7.12E-01 | 9.35E-01 | 208 | 12635 | -0.04 | 0.13 | 7.29E-01 | 9.43E-01 | 211 | -0.05 | 0.09 | 6.13E-01 | 0.00 | 9.93E-01 | - | - |
| Vitamin B6 intake (mg/day) | Nutrients | 12871 | 0.08 | 0.13 | 5.28E-01 | 8.81E-01 | 161 | 12635 | 0.01 | 0.14 | 9.50E-01 | 9.94E-01 | 257 | 0.05 | 0.09 | 6.20E-01 | 0.00 | 6.93E-01 | - | - |
| Frequency of dancing during leisure time | Physical activity | 12649 | 0.05 | 0.13 | 6.68E-01 | 9.21E-01 | 197 | 12424 | -0.15 | 0.13 | 2.48E-01 | 6.44E-01 | 103 | -0.04 | 0.09 | 6.20E-01 | 0.22 | 2.59E-01 | - | - |
| For how much of the time during the last four weeks have you felt so depressed that nothing could cheer you up? | General health | 192 | 0.65 | 1.08 | 5.45E-01 | 8.81E-01 | 166 | 161 | -1.20 | 0.98 | 2.24E-01 | 6.29E-01 | 97 | -0.36 | 0.72 | 6.20E-01 | 0.38 | 2.04E-01 | - | - |
| Physical limitation to participate in moderately demanding activities: moving a table, vacuuming, walking in the forest or gardening | General health | 190 | 0.05 | 1.10 | 9.65E-01 | 9.94E-01 | 264 | 162 | 0.59 | 0.98 | 5.49E-01 | 8.62E-01 | 174 | 0.35 | 0.73 | 6.33E-01 | 0.00 | 7.15E-01 | - | - |
| Iron intake (mg/day) | Nutrients | 12871 | 0.15 | 0.14 | 2.56E-01 | 7.06E-01 | 99 | 12635 | -0.07 | 0.14 | 6.18E-01 | 9.09E-01 | 184 | 0.05 | 0.10 | 6.39E-01 | 0.24 | 2.51E-01 | - | - |
| Fitness status | Physical activity | 6843 | -0.06 | 0.18 | 7.36E-01 | 9.52E-01 | 211 | 6716 | 0.18 | 0.18 | 3.10E-01 | 6.94E-01 | 122 | 0.06 | 0.13 | 6.43E-01 | 0.00 | 3.35E-01 | - | - |
| For how much of the time during the last four weeks have you felt happy? | General health | 192 | -0.98 | 1.07 | 3.62E-01 | 8.24E-01 | 120 | 162 | 0.22 | 0.99 | 8.27E-01 | 9.71E-01 | 231 | -0.33 | 0.73 | 6.46E-01 | 0.00 | 4.13E-01 | - | - |
| Berries (fresh or frozen) | Food | 12871 | 0.05 | 0.13 | 6.91E-01 | 9.29E-01 | 202 | 12635 | 0.03 | 0.13 | 8.16E-01 | 9.71E-01 | 229 | 0.04 | 0.09 | 6.54E-01 | 0.00 | 9.11E-01 | - | - |
| Liver, kidney | Food | 6466 | 0.04 | 0.18 | 8.12E-01 | 9.67E-01 | 227 | 6344 | -0.16 | 0.18 | 3.81E-01 | 7.88E-01 | 130 | -0.06 | 0.13 | 6.58E-01 | 0.00 | 4.28E-01 | - | - |
| Sausage as main dish | Food | 12871 | -0.06 | 0.13 | 6.33E-01 | 8.97E-01 | 192 | 12635 | -0.02 | 0.13 | 8.92E-01 | 9.87E-01 | 245 | -0.04 | 0.09 | 6.61E-01 | 0.00 | 8.15E-01 | - | - |
| Plant based protein intake (g/day) | Nutrients | 12871 | 0.08 | 0.13 | 5.37E-01 | 8.81E-01 | 164 | 12635 | 0.00 | 0.13 | 9.94E-01 | 9.97E-01 | 272 | 0.04 | 0.09 | 6.62E-01 | 0.00 | 6.63E-01 | - | - |
| Low fat milk (0,5%) | Beverage | 12871 | -0.04 | 0.13 | 7.63E-01 | 9.65E-01 | 214 | 12635 | -0.04 | 0.13 | 7.62E-01 | 9.47E-01 | 216 | -0.04 | 0.09 | 6.69E-01 | 0.00 | 9.93E-01 | - | - |
| Potato salad | Food | 5274 | -0.03 | 0.20 | 8.64E-01 | 9.88E-01 | 236 | 5165 | 0.16 | 0.20 | 4.40E-01 | 8.28E-01 | 144 | 0.06 | 0.14 | 6.78E-01 | 0.00 | 5.01E-01 | - | - |
| Zinc intake (mg/day) | Nutrients | 12871 | 0.03 | 0.13 | 7.97E-01 | 9.67E-01 | 222 | 12635 | 0.04 | 0.14 | 7.50E-01 | 9.47E-01 | 214 | 0.04 | 0.10 | 6.84E-01 | 0.00 | 9.59E-01 | - | - |
| Excellent health | General health | 191 | 1.53 | 1.11 | 1.70E-01 | 5.75E-01 | 80 | 160 | -1.76 | 1.00 | 7.85E-02 | 3.83E-01 | 56 | -0.29 | 0.74 | 6.91E-01 | 0.80 | 2.72E-02 | - | - |
| Tomato, cucumber | Food | 12871 | -0.07 | 0.13 | 6.05E-01 | 8.97E-01 | 182 | 12635 | 0.00 | 0.14 | 9.86E-01 | 9.94E-01 | 271 | -0.04 | 0.09 | 7.01E-01 | 0.00 | 7.28E-01 | - | - |
| Fat intake (g/day) | Nutrients | 12871 | -0.07 | 0.13 | 6.01E-01 | 8.97E-01 | 181 | 12635 | 0.00 | 0.13 | 9.97E-01 | 9.97E-01 | 273 | -0.04 | 0.09 | 7.05E-01 | 0.00 | 7.18E-01 | - | - |
| Vitamin A intake (mg/day) | Nutrients | 12871 | 0.12 | 0.14 | 4.14E-01 | 8.44E-01 | 133 | 12635 | -0.04 | 0.15 | 7.71E-01 | 9.47E-01 | 221 | 0.04 | 0.10 | 7.06E-01 | 0.00 | 4.35E-01 | - | - |
| Potassium intake (mg/day) | Nutrients | 12871 | -0.03 | 0.14 | 8.15E-01 | 9.67E-01 | 230 | 12635 | -0.04 | 0.14 | 7.69E-01 | 9.47E-01 | 220 | -0.04 | 0.10 | 7.10E-01 | 0.00 | 9.63E-01 | - | - |
| Formic acid intaje (g/day) | Nutrients | 12871 | -0.15 | 0.13 | 2.55E-01 | 7.06E-01 | 98 | 12635 | 0.09 | 0.13 | 5.24E-01 | 8.59E-01 | 164 | -0.03 | 0.09 | 7.10E-01 | 0.36 | 2.11E-01 | - | - |
| Brewed (filtered) coffee | Beverage | 12871 | 0.19 | 0.13 | 1.23E-01 | 5.18E-01 | 65 | 12635 | -0.28 | 0.13 | 3.39E-02 | 2.32E-01 | 40 | -0.03 | 0.09 | 7.11E-01 | 0.85 | 9.45E-03 | - | - |
| Number of friends that can come to your home at any time and feel at home | Social | 14425 | -0.06 | 0.12 | 6.29E-01 | 8.97E-01 | 190 | 14184 | 0.00 | 0.12 | 9.79E-01 | 9.94E-01 | 267 | -0.03 | 0.09 | 7.14E-01 | 0.00 | 7.51E-01 | - | - |
| Pentadecanoic acid intake (g/day) | Nutrients | 12871 | -0.11 | 0.13 | 3.93E-01 | 8.44E-01 | 125 | 12635 | 0.05 | 0.13 | 7.14E-01 | 9.37E-01 | 207 | -0.03 | 0.09 | 7.21E-01 | 0.00 | 3.91E-01 | - | - |
| Heptadecanoic acid intake (g/day) | Nutrients | 12871 | -0.11 | 0.13 | 3.93E-01 | 8.44E-01 | 126 | 12635 | 0.05 | 0.13 | 7.14E-01 | 9.37E-01 | 208 | -0.03 | 0.09 | 7.21E-01 | 0.00 | 3.91E-01 | - | - |
| Marital status: Single vs Widow/widower | Social | 1530 | 1.60 | 1.74 | 3.58E-01 | 8.24E-01 | 118 | 1514 | -0.64 | 1.66 | 7.02E-01 | 9.31E-01 | 206 | 0.43 | 1.20 | 7.21E-01 | 0.00 | 3.53E-01 | - | - |
| Secoisolariciresinol intake (ug/day) | Nutrients | 12871 | 0.22 | 0.13 | 8.80E-02 | 4.07E-01 | 59 | 12635 | -0.17 | 0.13 | 2.11E-01 | 6.08E-01 | 95 | 0.03 | 0.09 | 7.25E-01 | 0.77 | 3.70E-02 | - | - |
| Folic acid intake (ug/day) | Nutrients | 12871 | -0.08 | 0.13 | 5.77E-01 | 8.86E-01 | 178 | 12635 | 0.01 | 0.14 | 9.43E-01 | 9.94E-01 | 255 | -0.03 | 0.10 | 7.27E-01 | 0.00 | 6.60E-01 | - | - |
| Low fat margarine on bread | Food | 12871 | 0.03 | 0.13 | 8.05E-01 | 9.67E-01 | 225 | 12635 | -0.10 | 0.13 | 4.60E-01 | 8.28E-01 | 150 | -0.03 | 0.09 | 7.36E-01 | 0.00 | 4.83E-01 | - | - |
| Sucrose intake (g/day) | Nutrients | 12871 | 0.16 | 0.13 | 2.10E-01 | 6.34E-01 | 90 | 12635 | -0.11 | 0.13 | 4.03E-01 | 8.03E-01 | 137 | 0.03 | 0.09 | 7.51E-01 | 0.54 | 1.41E-01 | - | - |
| Participation in sports or physical exercise associations | Social | 772 | -0.73 | 0.94 | 4.34E-01 | 8.50E-01 | 139 | 689 | 1.46 | 1.08 | 1.76E-01 | 5.72E-01 | 84 | 0.21 | 0.71 | 7.66E-01 | 0.58 | 1.25E-01 | - | - |
| Marital status: Single vs Married/partner | Social | 13599 | 0.15 | 0.40 | 7.09E-01 | 9.35E-01 | 207 | 13363 | 0.01 | 0.41 | 9.72E-01 | 9.94E-01 | 265 | 0.08 | 0.29 | 7.70E-01 | 0.00 | 8.14E-01 | - | - |
| Number of social interactions during a normal week | Social | 14447 | -0.04 | 0.12 | 7.59E-01 | 9.65E-01 | 213 | 14218 | -0.01 | 0.12 | 9.21E-01 | 9.94E-01 | 251 | -0.02 | 0.09 | 7.72E-01 | 0.00 | 8.87E-01 | - | - |
| Palmitic acid intake (g/day) | Nutrients | 12871 | -0.06 | 0.13 | 6.21E-01 | 8.97E-01 | 187 | 12635 | 0.02 | 0.13 | 9.10E-01 | 9.94E-01 | 250 | -0.03 | 0.09 | 7.82E-01 | 0.00 | 6.71E-01 | - | - |
| Self rate of overall health | General health | 191 | 0.25 | 1.07 | 8.13E-01 | 9.67E-01 | 228 | 161 | 0.16 | 1.02 | 8.76E-01 | 9.87E-01 | 240 | 0.20 | 0.74 | 7.82E-01 | 0.00 | 9.49E-01 | - | - |
| Sweets | Food | 12871 | 0.08 | 0.13 | 5.62E-01 | 8.81E-01 | 173 | 12635 | -0.03 | 0.13 | 8.36E-01 | 9.71E-01 | 235 | 0.03 | 0.09 | 7.86E-01 | 0.00 | 5.80E-01 | - | - |
| Milk, sour milk (1,5%) | Beverage | 12871 | 0.13 | 0.13 | 2.86E-01 | 7.51E-01 | 104 | 12635 | -0.19 | 0.13 | 1.39E-01 | 5.13E-01 | 73 | -0.02 | 0.09 | 7.91E-01 | 0.69 | 7.11E-02 | - | - |
| Satisfaction with economy | Psychosocial | 6851 | -0.07 | 0.17 | 7.02E-01 | 9.31E-01 | 205 | 6716 | 0.14 | 0.18 | 4.37E-01 | 8.28E-01 | 143 | 0.03 | 0.12 | 7.92E-01 | 0.00 | 4.09E-01 | - | - |
| Self-employed | Psychosocial | 14198 | -0.09 | 0.41 | 8.23E-01 | 9.72E-01 | 231 | 13973 | 0.27 | 0.43 | 5.37E-01 | 8.59E-01 | 169 | 0.08 | 0.30 | 7.92E-01 | 0.00 | 5.47E-01 | - | - |
| Grams of tobacco smoked per week | Tobacco use | 9041 | 0.17 | 0.15 | 2.46E-01 | 6.95E-01 | 96 | 8814 | -0.14 | 0.16 | 3.83E-01 | 7.88E-01 | 132 | 0.03 | 0.11 | 7.98E-01 | 0.51 | 1.53E-01 | - | - |
| Extent to what your physical and emotional health disrupted your usual social life during the last four weeks | General health | 192 | -0.76 | 1.08 | 4.80E-01 | 8.81E-01 | 147 | 162 | 0.30 | 0.98 | 7.61E-01 | 9.47E-01 | 215 | -0.18 | 0.72 | 8.02E-01 | 0.00 | 4.66E-01 | - | - |
| Sum of all lignans intake (ug/day) | Nutrients | 12871 | 0.10 | 0.13 | 4.32E-01 | 8.50E-01 | 138 | 12635 | -0.06 | 0.13 | 6.50E-01 | 9.12E-01 | 193 | 0.02 | 0.09 | 8.05E-01 | 0.00 | 3.83E-01 | - | - |
| Smoking status: Former smokers vs non-smokers | Tobacco use | 9952 | 0.20 | 0.31 | 5.34E-01 | 8.81E-01 | 163 | 9860 | -0.10 | 0.33 | 7.66E-01 | 9.47E-01 | 218 | 0.05 | 0.23 | 8.09E-01 | 0.00 | 5.18E-01 | - | - |
| Carbohydrates intake (g/day) | Nutrients | 12871 | 0.03 | 0.13 | 8.40E-01 | 9.80E-01 | 234 | 12635 | -0.07 | 0.13 | 5.82E-01 | 8.98E-01 | 177 | -0.02 | 0.09 | 8.11E-01 | 0.00 | 5.92E-01 | - | - |
| Matairesinol intake (ug/day) | Nutrients | 12871 | 0.02 | 0.13 | 9.06E-01 | 9.88E-01 | 247 | 12635 | -0.06 | 0.13 | 6.49E-01 | 9.12E-01 | 191 | -0.02 | 0.09 | 8.16E-01 | 0.00 | 6.83E-01 | - | - |
| Enterodiol intake (ug/day) | Nutrients | 12871 | 0.00 | 0.12 | 9.89E-01 | 1.00E+00 | 269 | 12635 | -0.04 | 0.13 | 7.38E-01 | 9.47E-01 | 212 | -0.02 | 0.09 | 8.24E-01 | 0.00 | 8.02E-01 | - | - |
| For how much of the time during the last four weeks have you felt really alert and strong? | General health | 192 | -0.32 | 1.11 | 7.71E-01 | 9.65E-01 | 218 | 162 | -0.02 | 0.98 | 9.85E-01 | 9.94E-01 | 269 | -0.15 | 0.74 | 8.36E-01 | 0.00 | 8.36E-01 | - | - |
| People to ask for help apart from the ones at home | Social | 14544 | -0.27 | 0.39 | 4.85E-01 | 8.81E-01 | 149 | 14309 | 0.16 | 0.39 | 6.78E-01 | 9.28E-01 | 199 | -0.06 | 0.28 | 8.38E-01 | 0.00 | 4.32E-01 | - | - |
| Pancake, waffle, Swedish dumpling | Food | 12871 | 0.01 | 0.13 | 9.52E-01 | 9.92E-01 | 262 | 12635 | -0.05 | 0.13 | 7.27E-01 | 9.43E-01 | 209 | -0.02 | 0.09 | 8.42E-01 | 0.00 | 7.70E-01 | - | - |
| Milk, sour milk (3%) | Beverage | 12871 | 0.05 | 0.12 | 6.91E-01 | 9.29E-01 | 203 | 12635 | -0.02 | 0.13 | 9.00E-01 | 9.87E-01 | 248 | 0.02 | 0.09 | 8.43E-01 | 0.00 | 7.14E-01 | - | - |
| Distance to work in kilometers (one way) | Physical activity | 12768 | 0.08 | 0.13 | 5.19E-01 | 8.81E-01 | 158 | 12605 | -0.05 | 0.13 | 6.99E-01 | 9.31E-01 | 203 | 0.02 | 0.09 | 8.43E-01 | 0.00 | 4.68E-01 | - | - |
| Cohabitation: Live alone vs Only children | Social | 1928 | 0.61 | 0.76 | 4.23E-01 | 8.44E-01 | 136 | 1904 | -0.47 | 0.82 | 5.64E-01 | 8.75E-01 | 176 | 0.11 | 0.56 | 8.46E-01 | 0.00 | 3.33E-01 | - | - |
| Vitamin B12 intake (ug/day) | Nutrients | 12871 | 0.07 | 0.13 | 5.50E-01 | 8.81E-01 | 167 | 12635 | -0.04 | 0.13 | 7.29E-01 | 9.43E-01 | 210 | 0.02 | 0.09 | 8.51E-01 | 0.00 | 5.06E-01 | - | - |
| Frequent social contacts with colleagues during work | Psychosocial | 13892 | 0.02 | 0.12 | 8.77E-01 | 9.88E-01 | 238 | 13658 | -0.05 | 0.12 | 6.75E-01 | 9.28E-01 | 198 | -0.02 | 0.09 | 8.57E-01 | 0.00 | 6.82E-01 | - | - |
| Whole grain soft bread | Food | 12871 | -0.08 | 0.13 | 5.05E-01 | 8.81E-01 | 155 | 12635 | 0.06 | 0.13 | 6.62E-01 | 9.22E-01 | 196 | -0.02 | 0.09 | 8.62E-01 | 0.00 | 4.36E-01 | - | - |
| Beta-sitostanol intake (mg/day) | Nutrients | 12871 | 0.11 | 0.13 | 3.91E-01 | 8.44E-01 | 124 | 12635 | -0.08 | 0.13 | 5.19E-01 | 8.59E-01 | 163 | 0.01 | 0.09 | 8.69E-01 | 0.11 | 2.89E-01 | - | - |
| Salad dressing with oil | Food | 12871 | -0.05 | 0.13 | 6.79E-01 | 9.22E-01 | 201 | 12635 | 0.03 | 0.13 | 8.41E-01 | 9.71E-01 | 236 | -0.01 | 0.09 | 8.74E-01 | 0.00 | 6.66E-01 | - | - |
| Mashed potato | Food | 6466 | 0.09 | 0.19 | 6.23E-01 | 8.97E-01 | 188 | 6344 | -0.05 | 0.18 | 7.92E-01 | 9.52E-01 | 227 | 0.02 | 0.13 | 8.74E-01 | 0.00 | 5.93E-01 | - | - |
| Emotional problems that made you do less than you wanted during the last four weeks | General health | 190 | 4.37 | 3.33 | 1.90E-01 | 6.23E-01 | 83 | 161 | -3.57 | 2.76 | 1.98E-01 | 5.82E-01 | 93 | -0.33 | 2.12 | 8.77E-01 | 0.70 | 6.63E-02 | - | - |
| Cohabitation: Live alone vs Other/others | Social | 1478 | 0.09 | 1.07 | 9.31E-01 | 9.88E-01 | 256 | 1475 | -0.32 | 1.08 | 7.64E-01 | 9.47E-01 | 217 | -0.11 | 0.76 | 8.81E-01 | 0.00 | 7.83E-01 | - | - |
| Beta-carotene intake (mg/day) | Nutrients | 12871 | 0.04 | 0.13 | 7.69E-01 | 9.65E-01 | 217 | 12635 | -0.01 | 0.14 | 9.31E-01 | 9.94E-01 | 253 | 0.01 | 0.10 | 8.81E-01 | 0.00 | 7.89E-01 | - | - |
| Physical limitation to participate in moderately demanding activities: walking up several stairs | General health | 190 | -0.55 | 1.10 | 6.16E-01 | 8.97E-01 | 186 | 160 | 0.30 | 1.06 | 7.78E-01 | 9.47E-01 | 223 | -0.11 | 0.76 | 8.86E-01 | 0.00 | 5.77E-01 | - | - |
| Vitamin E intake (mg/day) | Nutrients | 12871 | -0.03 | 0.13 | 8.09E-01 | 9.67E-01 | 226 | 12635 | 0.06 | 0.13 | 6.52E-01 | 9.12E-01 | 195 | 0.01 | 0.09 | 8.86E-01 | 0.00 | 6.23E-01 | - | - |
| Monounsaturated fat intake (g/day) | Nutrients | 12871 | 0.08 | 0.14 | 5.61E-01 | 8.81E-01 | 171 | 12635 | -0.06 | 0.14 | 6.90E-01 | 9.31E-01 | 201 | 0.01 | 0.10 | 8.88E-01 | 0.00 | 4.90E-01 | - | - |
| Selenium intake (ug/day) | Nutrients | 12871 | 0.01 | 0.13 | 9.23E-01 | 9.88E-01 | 254 | 12635 | -0.04 | 0.13 | 7.69E-01 | 9.47E-01 | 219 | -0.01 | 0.09 | 8.92E-01 | 0.00 | 7.81E-01 | - | - |
| Cohabitation: Live alone vs Adult and children | Social | 10162 | 0.35 | 0.43 | 4.14E-01 | 8.44E-01 | 134 | 9884 | -0.28 | 0.44 | 5.16E-01 | 8.59E-01 | 162 | 0.04 | 0.31 | 8.95E-01 | 0.07 | 3.01E-01 | - | - |
| Boiled coffee | Beverage | 12871 | 0.05 | 0.13 | 6.99E-01 | 9.31E-01 | 204 | 12635 | -0.03 | 0.13 | 8.33E-01 | 9.71E-01 | 234 | 0.01 | 0.09 | 8.96E-01 | 0.00 | 6.74E-01 | - | - |
| For how much of the time during the last four weeks have you felt worn out? | General health | 191 | 0.56 | 1.11 | 6.15E-01 | 8.97E-01 | 185 | 159 | -0.61 | 1.00 | 5.40E-01 | 8.59E-01 | 170 | -0.09 | 0.74 | 9.04E-01 | 0.00 | 4.32E-01 | - | - |
| Smoking status: Former occasional smokers vs non-smokers | Tobacco use | 8361 | 0.00 | 0.42 | 9.94E-01 | 1.00E+00 | 271 | 8192 | -0.08 | 0.44 | 8.59E-01 | 9.86E-01 | 238 | -0.04 | 0.31 | 9.07E-01 | 0.00 | 8.94E-01 | - | - |
| Saturated fat intake (g/day) | Nutrients | 12871 | -0.09 | 0.13 | 5.00E-01 | 8.81E-01 | 153 | 12635 | 0.07 | 0.13 | 5.98E-01 | 9.07E-01 | 180 | -0.01 | 0.09 | 9.07E-01 | 0.00 | 3.96E-01 | - | - |
| Root vegetables, carrot | Food | 12871 | 0.03 | 0.13 | 8.01E-01 | 9.67E-01 | 224 | 12635 | -0.01 | 0.14 | 9.21E-01 | 9.94E-01 | 252 | 0.01 | 0.10 | 9.11E-01 | 0.00 | 8.05E-01 | - | - |
| Animal based protein intake (g/day) | Nutrients | 12871 | -0.02 | 0.13 | 9.05E-01 | 9.88E-01 | 244 | 12635 | 0.00 | 0.13 | 9.72E-01 | 9.94E-01 | 266 | -0.01 | 0.09 | 9.12E-01 | 0.00 | 9.53E-01 | - | - |
| Teetotaler | Alcohol | 14283 | -0.10 | 0.40 | 7.93E-01 | 9.67E-01 | 220 | 14064 | 0.06 | 0.42 | 8.90E-01 | 9.87E-01 | 243 | -0.03 | 0.29 | 9.23E-01 | 0.00 | 7.79E-01 | - | - |
| Appetite status | Psychosocial | 6847 | -0.06 | 0.17 | 7.29E-01 | 9.51E-01 | 209 | 6720 | 0.04 | 0.18 | 8.10E-01 | 9.70E-01 | 228 | -0.01 | 0.12 | 9.34E-01 | 0.00 | 6.79E-01 | - | - |
| Sausage, liver pate on bread | Food | 12871 | 0.00 | 0.13 | 9.90E-01 | 1.00E+00 | 270 | 12635 | 0.02 | 0.13 | 9.00E-01 | 9.87E-01 | 247 | 0.01 | 0.09 | 9.37E-01 | 0.00 | 9.21E-01 | - | - |
| Cream, creme fraiche, sour cream | Food | 12871 | -0.13 | 0.13 | 3.14E-01 | 7.75E-01 | 110 | 12635 | 0.15 | 0.13 | 2.48E-01 | 6.44E-01 | 104 | 0.01 | 0.09 | 9.37E-01 | 0.57 | 1.26E-01 | - | - |
| Campestanol intake (mg/day) | Nutrients | 12871 | 0.06 | 0.13 | 6.26E-01 | 8.97E-01 | 189 | 12635 | -0.08 | 0.13 | 5.40E-01 | 8.59E-01 | 171 | -0.01 | 0.09 | 9.38E-01 | 0.00 | 4.36E-01 | - | - |
| Total protein intake (g/day) | Nutrients | 12871 | 0.02 | 0.13 | 8.85E-01 | 9.88E-01 | 241 | 12635 | -0.01 | 0.13 | 9.67E-01 | 9.94E-01 | 264 | 0.01 | 0.09 | 9.40E-01 | 0.00 | 8.96E-01 | - | - |
| Possibility to leave your work for a while to speak with a colleague | Psychosocial | 13922 | -0.09 | 0.12 | 4.82E-01 | 8.81E-01 | 148 | 13654 | 0.11 | 0.13 | 4.07E-01 | 8.05E-01 | 138 | 0.01 | 0.09 | 9.44E-01 | 0.15 | 2.78E-01 | - | - |

| | | | | | | | | | | | | | | | | | | | | |
|---|---|---|---|---|---|---|---|---|---|---|---|---|---|---|---|---|---|---|---|---|
| Shellfish (e.g. shrimps, scallops) | Food | 6466 | 0.11 | 0.18 | 5.61E-01 | 8.81E-01 | 172 | 6344 | -0.09 | 0.19 | 6.19E-01 | 9.09E-01 | 186 | 0.01 | 0.13 | 9.46E-01 | 0.00 | 4.46E-01 | - | - |
| Cholesterol intake (g/day) | Nutrients | 12871 | 0.02 | 0.14 | 8.84E-01 | 9.88E-01 | 239 | 12635 | -0.01 | 0.14 | 9.54E-01 | 9.94E-01 | 259 | 0.01 | 0.10 | 9.48E-01 | 0.00 | 8.86E-01 | - | - |
| As healthy as anyone | General health | 191 | 1.70 | 1.08 | 1.18E-01 | 5.18E-01 | 62 | 161 | -1.46 | 0.97 | 1.37E-01 | 5.13E-01 | 71 | -0.04 | 0.72 | 9.51E-01 | 0.79 | 3.02E-02 | - | - |
| Banana | Food | 12871 | -0.07 | 0.13 | 5.65E-01 | 8.81E-01 | 175 | 12635 | 0.09 | 0.13 | 5.10E-01 | 8.59E-01 | 161 | 0.00 | 0.09 | 9.65E-01 | 0.00 | 3.82E-01 | - | - |
| Minced meat dishes | Food | 12871 | 0.12 | 0.13 | 3.23E-01 | 7.75E-01 | 113 | 12635 | -0.14 | 0.13 | 2.80E-01 | 6.76E-01 | 113 | 0.00 | 0.09 | 9.69E-01 | 0.53 | 1.43E-01 | - | - |
| Blood based food | Food | 6466 | 0.00 | 0.18 | 9.97E-01 | 1.00E+00 | 272 | 6344 | -0.01 | 0.18 | 9.64E-01 | 9.94E-01 | 262 | 0.00 | 0.13 | 9.73E-01 | 0.00 | 9.76E-01 | - | - |
| Tiamin intake (mg/day) | Nutrients | 12871 | 0.09 | 0.13 | 5.04E-01 | 8.81E-01 | 154 | 12635 | -0.10 | 0.14 | 4.69E-01 | 8.30E-01 | 154 | 0.00 | 0.09 | 9.78E-01 | 0.00 | 3.25E-01 | - | - |
| Light and physically active work | Physical activity | 14224 | 0.18 | 0.28 | 5.29E-01 | 8.81E-01 | 162 | 13937 | -0.18 | 0.29 | 5.36E-01 | 8.59E-01 | 168 | 0.00 | 0.20 | 9.84E-01 | 0.00 | 3.77E-01 | - | - |
| Stigmasterol intake (mg/day) | Nutrients | 12871 | 0.05 | 0.13 | 6.78E-01 | 9.22E-01 | 200 | 12635 | -0.06 | 0.13 | 6.51E-01 | 9.12E-01 | 194 | 0.00 | 0.09 | 9.87E-01 | 0.00 | 5.39E-01 | - | - |
| Physical limitation to participate in moderately demanding activities: bending down or kneeling | General health | 189 | -0.66 | 1.12 | 5.55E-01 | 8.81E-01 | 168 | 162 | 0.53 | 0.99 | 5.90E-01 | 9.00E-01 | 179 | 0.01 | 0.74 | 9.89E-01 | 0.00 | 4.24E-01 | - | - |

**Supplementary Table 14**. Longitudinal association results for DBP

| | | Training set | | | | | | Testing set | | | | | | Metanalysis | | | | | R2 | |
|---|---|---|---|---|---|---|---|---|---|---|---|---|---|---|---|---|---|---|---|---|
| **Description** | **Group** | **N** | **Effect estimate** | **S.E.** | **p-value** | **p-value$_{FDR}$** | **p-value rank** | **N** | **Effect estimate** | **S.E.** | **p-value** | **p-value$_{FDR}$** | **p-value rank** | **Effect estimate** | **S.E.** | **p-value** | **$I^2$** | **Q p-value** | **adjusted R2** | **R2 rank** |
| Informed of having high blood pressure | General health | 14534 | 3.88 | 0.24 | 2.72E-58 | **7.41E-56** | 1 | 14269 | 4.32 | 0.24 | 3.46E-69 | **9.45E-67** | 1 | 4.10 | 0.17 | 1.86E-126 | 0.41 | 1.95E-01 | 0.38 | 1 |
| Parents or siblings had a cerebral hemorrhage/thrombosis or cardiac infarction before the age of 60 | General health | 14346 | 0.78 | 0.20 | 8.24E-05 | **5.62E-03** | 4 | 14109 | 0.82 | 0.20 | 4.60E-05 | **4.19E-03** | 3 | 0.80 | 0.14 | 1.46E-08 | 0.00 | 8.86E-01 | 0.37 | 3 |
| Oatflake, whole wheat, rye or barley porridge | Food | 12863 | -0.26 | 0.08 | 1.07E-03 | **4.25E-02** | 6 | 12614 | -0.36 | 0.08 | 1.24E-05 | **1.69E-03** | 2 | -0.31 | 0.06 | 6.79E-08 | 0.00 | 4.02E-01 | 0.37 | 2 |
| Total energy intake (kcal/day) | Nutrients | 12863 | -0.25 | 0.09 | 6.16E-03 | 1.05E-01 | 16 | 12614 | -0.37 | 0.10 | 1.06E-04 | **5.80E-03** | 5 | -0.31 | 0.07 | 3.08E-06 | 0.00 | 3.79E-01 | - | - |
| Frequency of picking berries or mushrooms during leisure time | Physical activity | 13563 | -0.36 | 0.08 | 1.28E-05 | **1.74E-03** | 2 | 13335 | -0.17 | 0.08 | 4.10E-02 | 2.66E-01 | 42 | -0.26 | 0.06 | 5.36E-06 | 0.61 | 1.11E-01 | - | - |
| Overall state of health compared to other your age | General health | 12565 | -0.34 | 0.08 | 3.84E-05 | **3.49E-03** | 3 | 12373 | -0.19 | 0.08 | 2.45E-02 | 2.04E-01 | 32 | -0.27 | 0.06 | 6.20E-06 | 0.37 | 2.07E-01 | - | - |
| Breakfast habits: Porridge w/o sandwich for breakfast vs not breakfast at all | Food | 1732 | -1.49 | 0.47 | 1.66E-03 | 5.03E-02 | 9 | 1677 | -1.57 | 0.52 | 2.40E-03 | **4.67E-02** | 14 | -1.53 | 0.35 | 1.20E-05 | 0.00 | 9.03E-01 | - | - |
| Ice cream | Food | 12863 | -0.22 | 0.08 | 7.23E-03 | 1.16E-01 | 17 | 12614 | -0.26 | 0.08 | 1.50E-03 | **3.42E-02** | 12 | -0.24 | 0.06 | 3.45E-05 | 0.00 | 7.06E-01 | - | - |
| Average portion size of potatoes/rice/pasta based on photographic illustration of four sizes (smallest to largest) | Food | 12863 | -0.28 | 0.09 | 1.99E-03 | 5.10E-02 | 10 | 12614 | -0.25 | 0.09 | 5.84E-03 | 8.39E-02 | 19 | -0.26 | 0.06 | 3.51E-05 | 0.00 | 8.39E-01 | - | - |
| Everyday exercise satisfaction | Physical activity | 14446 | 0.17 | 0.08 | 2.47E-02 | 2.93E-01 | 23 | 14181 | 0.29 | 0.08 | 3.19E-04 | **1.25E-02** | 7 | 0.23 | 0.06 | 3.77E-05 | 0.02 | 3.11E-01 | - | - |
| Meat on bread | Food | 12863 | 0.26 | 0.08 | 1.09E-03 | **4.25E-02** | 7 | 12614 | 0.20 | 0.08 | 1.56E-02 | 1.63E-01 | 26 | 0.23 | 0.06 | 5.62E-05 | 0.00 | 5.83E-01 | - | - |
| Control over planning and execution of the workday | Psychosocial | 14446 | -0.24 | 0.08 | 2.06E-03 | 5.10E-02 | 11 | 14169 | -0.20 | 0.08 | 9.82E-03 | 1.17E-01 | 23 | -0.22 | 0.06 | 6.08E-05 | 0.00 | 7.60E-01 | - | - |
| Number of snuff boxes per week | Tobacco use | 13786 | 0.17 | 0.08 | 3.35E-02 | 3.39E-01 | 27 | 13577 | 0.28 | 0.08 | 9.28E-04 | **2.59E-02** | 9 | 0.23 | 0.06 | 1.27E-04 | 0.00 | 3.71E-01 | - | - |
| Educational level | Psychosocial | 14544 | -0.25 | 0.08 | 1.57E-03 | 5.03E-02 | 8 | 14320 | -0.17 | 0.08 | 3.60E-02 | 2.59E-01 | 38 | -0.21 | 0.06 | 1.95E-04 | 0.00 | 4.71E-01 | - | - |
| Exercise during the last three months | Physical activity | 14429 | -0.22 | 0.08 | 4.72E-03 | 9.90E-02 | 13 | 14176 | -0.17 | 0.08 | 2.98E-02 | 2.31E-01 | 35 | -0.19 | 0.06 | 3.99E-04 | 0.00 | 6.76E-01 | - | - |
| Travel to work: Cycle to work vs passive travel to work | Physical activity | 11221 | -0.47 | 0.20 | 1.71E-02 | 2.33E-01 | 20 | 11060 | -0.53 | 0.20 | 8.77E-03 | 1.12E-01 | 21 | -0.50 | 0.14 | 4.02E-04 | 0.00 | 8.42E-01 | - | - |
| White (soft) bread, thin crisp bread | Food | 12863 | 0.31 | 0.08 | 1.34E-04 | **7.33E-03** | 5 | 12614 | 0.09 | 0.08 | 2.67E-01 | 6.73E-01 | 107 | 0.21 | 0.06 | 4.43E-04 | 0.71 | 6.20E-02 | - | - |
| Fiber cereals | Food | 12863 | -0.24 | 0.08 | 2.85E-03 | 6.48E-02 | 12 | 12614 | -0.14 | 0.08 | 9.67E-02 | 4.30E-01 | 61 | -0.19 | 0.06 | 9.87E-04 | 0.00 | 3.68E-01 | - | - |
| Changed everyday exercise during the last year | Physical activity | 14486 | 0.07 | 0.08 | 3.31E-01 | 7.46E-01 | 121 | 14224 | 0.29 | 0.08 | 2.20E-04 | **1.00E-02** | 6 | 0.18 | 0.05 | 1.04E-03 | 0.74 | 4.95E-02 | - | - |
| Whole grain intake (g/day) | Food | 12863 | -0.19 | 0.08 | 1.87E-02 | 2.43E-01 | 21 | 12614 | -0.19 | 0.08 | 2.27E-02 | 2.04E-01 | 30 | -0.19 | 0.06 | 1.06E-03 | 0.00 | 9.84E-01 | - | - |
| Eat breakfast from 2000 | Food | 1204 | -1.23 | 0.81 | 1.28E-01 | 5.41E-01 | 64 | 1082 | -2.70 | 0.86 | 1.67E-03 | **3.51E-02** | 13 | -1.92 | 0.59 | 1.07E-03 | 0.36 | 2.12E-01 | - | - |
| Frequency of walking during leisure time | Physical activity | 14032 | 0.13 | 0.08 | 1.07E-01 | 4.99E-01 | 58 | 13818 | 0.24 | 0.08 | 3.03E-03 | 5.17E-02 | 16 | 0.19 | 0.06 | 1.26E-03 | 0.00 | 3.23E-01 | - | - |
| Iodine intake (ug/day) | Nutrients | 12863 | -0.10 | 0.08 | 2.36E-01 | 6.87E-01 | 92 | 12614 | -0.28 | 0.08 | 8.90E-04 | **2.59E-02** | 8 | -0.19 | 0.06 | 1.54E-03 | 0.59 | 1.19E-01 | - | - |
| Control over own work assignment | Psychosocial | 14433 | -0.09 | 0.08 | 2.51E-01 | 6.89E-01 | 99 | 14153 | -0.26 | 0.08 | 9.48E-04 | **2.59E-02** | 10 | -0.17 | 0.06 | 1.74E-03 | 0.59 | 1.18E-01 | - | - |
| Light but partly physically active work | Physical activity | 14214 | -0.27 | 0.20 | 1.68E-01 | 6.10E-01 | 75 | 13916 | -0.59 | 0.20 | 3.91E-03 | 5.94E-02 | 18 | -0.43 | 0.14 | 2.68E-03 | 0.18 | 2.70E-01 | - | - |
| If you exercise, change in exercise habits during the last year | Physical activity | 12333 | 0.10 | 0.08 | 2.10E-01 | 6.87E-01 | 83 | 12132 | 0.25 | 0.08 | 3.00E-03 | 5.17E-02 | 15 | 0.18 | 0.06 | 3.01E-03 | 0.37 | 2.09E-01 | - | - |
| High physical demand from job | Physical activity | 14465 | 0.15 | 0.08 | 4.56E-02 | 3.89E-01 | 32 | 14196 | 0.17 | 0.08 | 3.46E-02 | 2.55E-01 | 37 | 0.16 | 0.06 | 3.65E-03 | 0.00 | 9.07E-01 | - | - |
| Magnesium intake (mg/day) | Nutrients | 12863 | -0.15 | 0.09 | 7.64E-02 | 4.53E-01 | 46 | 12614 | -0.20 | 0.09 | 2.26E-02 | 2.04E-01 | 29 | -0.18 | 0.06 | 4.22E-03 | 0.00 | 6.95E-01 | - | - |
| Rice | Food | 12863 | -0.16 | 0.08 | 5.25E-02 | 4.13E-01 | 34 | 12614 | -0.17 | 0.09 | 4.08E-02 | 2.66E-01 | 40 | -0.17 | 0.06 | 4.86E-03 | 0.00 | 9.05E-01 | - | - |
| Job demands to work very fast | Psychosocial | 14430 | 0.13 | 0.08 | 8.02E-02 | 4.61E-01 | 47 | 14166 | 0.18 | 0.08 | 2.55E-02 | 2.05E-01 | 34 | 0.15 | 0.05 | 4.90E-03 | 0.00 | 7.11E-01 | - | - |
| Physical limitation that made you not being able to perform certain work tasks or other activities during the last four weeks | General health | 188 | 2.81 | 1.69 | 9.80E-02 | 4.99E-01 | 52 | 160 | 3.78 | 1.66 | 2.41E-02 | 2.04E-01 | 31 | 3.30 | 1.18 | 5.25E-03 | 0.00 | 6.82E-01 | - | - |
| Satisfaction with home and family situation | Psychosocial | 6846 | 0.13 | 0.11 | 2.45E-01 | 6.89E-01 | 97 | 6714 | 0.30 | 0.11 | 6.26E-03 | 8.54E-02 | 20 | 0.21 | 0.08 | 5.94E-03 | 0.21 | 2.61E-01 | - | - |
| Pasta | Food | 12863 | -0.22 | 0.09 | 8.82E-03 | 1.34E-01 | 18 | 12614 | -0.11 | 0.09 | 2.09E-01 | 6.24E-01 | 90 | -0.17 | 0.06 | 5.96E-03 | 0.00 | 3.48E-01 | - | - |
| Snuff status: Snuff users vs non-snuff users | Tobacco use | 12281 | 0.40 | 0.22 | 7.23E-02 | 4.53E-01 | 42 | 12112 | 0.47 | 0.23 | 3.97E-02 | 2.66E-01 | 39 | 0.44 | 0.16 | 6.48E-03 | 0.00 | 8.24E-01 | - | - |
| Teetotaler | Alcohol | 14273 | -0.58 | 0.26 | 2.30E-02 | 2.85E-01 | 22 | 14042 | -0.41 | 0.27 | 1.31E-01 | 5.04E-01 | 70 | -0.50 | 0.19 | 7.21E-03 | 0.00 | 6.33E-01 | - | - |
| Liquor, spirits | Alcohol | 12863 | 0.10 | 0.09 | 2.40E-01 | 6.87E-01 | 94 | 12614 | 0.23 | 0.09 | 9.06E-03 | 1.12E-01 | 22 | 0.16 | 0.06 | 7.62E-03 | 0.07 | 3.00E-01 | - | - |
| Whole grain soft bread | Food | 12863 | -0.23 | 0.08 | 5.20E-03 | 1.01E-01 | 14 | 12614 | -0.08 | 0.08 | 3.42E-01 | 7.13E-01 | 131 | -0.16 | 0.06 | 7.76E-03 | 0.38 | 2.02E-01 | - | - |
| Would you say that the number of people that you meet in your everyday life is enough or would you like to meet more or fewer people? | Social | 14569 | -0.15 | 0.08 | 4.81E-02 | 3.98E-01 | 33 | 14310 | -0.14 | 0.08 | 8.15E-02 | 3.92E-01 | 55 | -0.14 | 0.05 | 8.51E-03 | 0.00 | 8.88E-01 | - | - |
| Repetitive job | Psychosocial | 14424 | 0.19 | 0.08 | 1.61E-02 | 2.31E-01 | 19 | 14155 | 0.10 | 0.08 | 2.10E-01 | 6.24E-01 | 92 | 0.14 | 0.06 | 9.43E-03 | 0.00 | 4.29E-01 | - | - |
| Memory status | Psychosocial | 6825 | 0.23 | 0.11 | 3.33E-02 | 3.39E-01 | 26 | 6703 | 0.16 | 0.11 | 1.36E-01 | 5.04E-01 | 72 | 0.20 | 0.08 | 1.04E-02 | 0.00 | 6.61E-01 | - | - |
| Alcohol intake (g/day) | Alcohol | 12863 | 0.06 | 0.09 | 5.08E-01 | 8.48E-01 | 161 | 12614 | 0.25 | 0.09 | 3.61E-03 | 5.80E-02 | 17 | 0.15 | 0.06 | 1.19E-02 | 0.61 | 1.08E-01 | - | - |
| Whole grain crisp bread | Food | 12863 | 0.13 | 0.08 | 1.31E-01 | 5.41E-01 | 66 | 12614 | 0.17 | 0.08 | 4.09E-02 | 2.66E-01 | 41 | 0.15 | 0.06 | 1.21E-02 | 0.00 | 6.84E-01 | - | - |
| Learn new things at job | Psychosocial | 14410 | -0.13 | 0.08 | 1.01E-01 | 4.99E-01 | 55 | 14151 | -0.15 | 0.08 | 5.76E-02 | 3.42E-01 | 46 | -0.14 | 0.05 | 1.25E-02 | 0.00 | 8.34E-01 | - | - |
| Blota (broth + bread) | Food | 6463 | 0.24 | 0.12 | 4.16E-02 | 3.84E-01 | 29 | 6332 | 0.18 | 0.12 | 1.38E-01 | 5.04E-01 | 74 | 0.21 | 0.08 | 1.26E-02 | 0.00 | 7.23E-01 | - | - |
| Permanent employment | Psychosocial | 14309 | 0.32 | 0.18 | 7.26E-02 | 4.53E-01 | 43 | 14086 | 0.30 | 0.18 | 1.01E-01 | 4.30E-01 | 64 | 0.31 | 0.13 | 1.51E-02 | 0.00 | 9.34E-01 | - | - |
| Years using snuff | Tobacco use | 13423 | 0.09 | 0.09 | 2.88E-01 | 7.04E-01 | 111 | 13180 | 0.20 | 0.09 | 2.15E-02 | 2.04E-01 | 28 | 0.15 | 0.06 | 1.80E-02 | 0.00 | 3.65E-01 | - | - |
| Phosphate intake (mg/day) | Nutrients | 12863 | -0.18 | 0.08 | 3.06E-02 | 3.34E-01 | 25 | 12614 | -0.10 | 0.09 | 2.70E-01 | 6.73E-01 | 109 | -0.14 | 0.06 | 2.04E-02 | 0.00 | 4.73E-01 | - | - |
| Number of social interactions during a normal week | Social | 14437 | -0.14 | 0.08 | 7.58E-02 | 4.53E-01 | 45 | 14196 | -0.12 | 0.08 | 1.39E-01 | 5.04E-01 | 75 | -0.13 | 0.05 | 2.12E-02 | 0.00 | 8.55E-01 | - | - |
| Hamburger | Food | 12863 | 0.12 | 0.08 | 1.36E-01 | 5.56E-01 | 67 | 12614 | 0.15 | 0.09 | 8.29E-02 | 3.92E-01 | 56 | 0.14 | 0.06 | 2.27E-02 | 0.00 | 8.46E-01 | - | - |
| Participation in associations or voluntary organisations | Social | 14432 | -0.36 | 0.16 | 2.68E-02 | 3.05E-01 | 24 | 14164 | -0.16 | 0.17 | 3.27E-01 | 7.13E-01 | 124 | -0.26 | 0.12 | 2.30E-02 | 0.00 | 4.03E-01 | - | - |
| Soft whey cheese | Food | 6463 | -0.24 | 0.12 | 4.21E-02 | 3.84E-01 | 30 | 6332 | -0.13 | 0.12 | 2.69E-01 | 6.73E-01 | 108 | -0.19 | 0.08 | 2.59E-02 | 0.00 | 5.33E-01 | - | - |
| Frequncy of social contacts with colleagues during leisure time | Psychosocial | 13695 | -0.06 | 0.08 | 4.62E-01 | 8.32E-01 | 150 | 13439 | -0.19 | 0.08 | 1.61E-02 | 1.63E-01 | 27 | -0.12 | 0.06 | 2.71E-02 | 0.31 | 2.29E-01 | - | - |
| Medium beer | Alcohol | 12863 | 0.00 | 0.08 | 9.55E-01 | 9.93E-01 | 262 | 12614 | 0.27 | 0.08 | 1.32E-03 | **3.26E-02** | 11 | 0.13 | 0.06 | 2.74E-02 | 0.82 | 1.94E-02 | - | - |
| Breakfast habits: Only coffee/tea for breakfast vs not breakfast at all | Food | 804 | 1.60 | 1.13 | 1.56E-01 | 5.92E-01 | 72 | 772 | 1.91 | 1.14 | 9.38E-02 | 4.27E-01 | 60 | 1.75 | 0.80 | 2.86E-02 | 0.00 | 8.47E-01 | - | - |
| Chips, popcorn, salted nuts | Food | 12863 | 0.16 | 0.08 | 6.15E-02 | 4.53E-01 | 37 | 12614 | 0.10 | 0.09 | 2.26E-01 | 6.33E-01 | 97 | 0.13 | 0.06 | 2.89E-02 | 0.00 | 6.62E-01 | - | - |
| Cheese 28% | Food | 12863 | -0.15 | 0.08 | 6.38E-02 | 4.53E-01 | 38 | 12614 | -0.10 | 0.08 | 2.44E-01 | 6.54E-01 | 102 | -0.12 | 0.06 | 3.24E-02 | 0.00 | 6.43E-01 | - | - |
| Cookies, pastry | Food | 12863 | -0.10 | 0.08 | 2.14E-01 | 6.87E-01 | 85 | 12614 | -0.15 | 0.08 | 7.51E-02 | 3.92E-01 | 52 | -0.12 | 0.06 | 3.30E-02 | 0.00 | 6.87E-01 | - | - |
| Satisfaction with leisure time | Psychosocial | 6842 | 0.11 | 0.11 | 3.06E-01 | 7.07E-01 | 118 | 6698 | 0.21 | 0.11 | 5.07E-02 | 3.15E-01 | 44 | 0.16 | 0.08 | 3.56E-02 | 0.00 | 5.02E-01 | - | - |
| Physical limitation that reduced the normal time spent at work or in other activities during the last four weeks | General health | 190 | 1.84 | 2.02 | 3.63E-01 | 7.70E-01 | 127 | 161 | 3.90 | 1.96 | 4.85E-02 | 3.08E-01 | 43 | 2.90 | 1.41 | 3.93E-02 | 0.00 | 4.64E-01 | - | - |
| Number of social contacts with the same interests as you | Social | 14532 | -0.05 | 0.08 | 5.56E-01 | 8.48E-01 | 175 | 14275 | -0.18 | 0.08 | 2.46E-02 | 2.04E-01 | 33 | -0.11 | 0.05 | 4.63E-02 | 0.30 | 2.33E-01 | - | - |
| Possibility to speak with colleagues during breaks | Psychosocial | 13983 | 0.13 | 0.08 | 1.07E-01 | 4.99E-01 | 57 | 13725 | 0.09 | 0.08 | 2.34E-01 | 6.39E-01 | 100 | 0.11 | 0.06 | 4.70E-02 | 0.00 | 7.82E-01 | - | - |
| Smoked fish/meat | Food | 12863 | 0.16 | 0.08 | 4.55E-02 | 3.89E-01 | 31 | 12614 | 0.06 | 0.08 | 4.45E-01 | 7.50E-01 | 161 | 0.11 | 0.06 | 4.92E-02 | 0.00 | 3.98E-01 | - | - |
| Physical limitation that limited your ability to perform certain work tasks or other activities during the last four weeks | General health | 189 | 1.17 | 1.76 | 5.09E-01 | 8.48E-01 | 162 | 160 | 4.32 | 1.98 | 3.04E-02 | 2.31E-01 | 36 | 2.56 | 1.32 | 5.15E-02 | 0.30 | 2.34E-01 | - | - |
| Cohabitation: Live alone vs Other/others | Social | 1477 | -0.77 | 0.70 | 2.73E-01 | 6.96E-01 | 107 | 1470 | -1.12 | 0.68 | 1.01E-01 | 4.30E-01 | 63 | -0.95 | 0.49 | 5.20E-02 | 0.00 | 7.20E-01 | - | - |
| Support from others | Social | 14574 | -0.13 | 0.08 | 1.03E-01 | 4.99E-01 | 56 | 14332 | -0.09 | 0.08 | 2.76E-01 | 6.79E-01 | 111 | -0.11 | 0.06 | 5.41E-02 | 0.00 | 7.17E-01 | - | - |
| Frequency of engaging in clubs, associations or study circles | Social | 9697 | -0.11 | 0.09 | 2.41E-01 | 6.87E-01 | 95 | 9604 | -0.15 | 0.10 | 1.20E-01 | 4.90E-01 | 67 | -0.13 | 0.07 | 5.44E-02 | 0.00 | 7.70E-01 | - | - |
| Fried potatoes, pommes frites | Food | 12863 | 0.12 | 0.08 | 1.59E-01 | 5.94E-01 | 73 | 12614 | 0.11 | 0.08 | 2.00E-01 | 6.17E-01 | 88 | 0.11 | 0.06 | 5.70E-02 | 0.00 | 9.42E-01 | - | - |
| Physical limitation that made you do less than you wanted during the last four weeks | General health | 189 | 1.49 | 1.53 | 3.30E-01 | 7.46E-01 | 120 | 161 | 2.94 | 1.69 | 8.31E-02 | 3.92E-01 | 57 | 2.15 | 1.13 | 5.81E-02 | 0.00 | 5.25E-01 | - | - |
| Overall state of health during the last year | General health | 14469 | -0.09 | 0.08 | 2.55E-01 | 6.89E-01 | 100 | 14205 | -0.12 | 0.08 | 1.34E-01 | 5.04E-01 | 71 | -0.10 | 0.06 | 6.27E-02 | 0.00 | 7.84E-01 | - | - |
| Cheese 10-17% | Food | 12863 | -0.10 | 0.08 | 2.27E-01 | 6.87E-01 | 88 | 12614 | 0.32 | 0.08 | 1.05E-04 | **5.80E-03** | 4 | 0.11 | 0.06 | 6.35E-02 | 0.92 | 2.99E-04 | - | - |
| Light beer | Alcohol | 12863 | -0.06 | 0.08 | 4.41E-01 | 8.27E-01 | 145 | 12614 | -0.16 | 0.09 | 6.34E-02 | 3.46E-01 | 50 | -0.11 | 0.06 | 6.45E-02 | 0.00 | 4.30E-01 | - | - |
| Energy status | Psychosocial | 6837 | 0.12 | 0.11 | 2.60E-01 | 6.89E-01 | 102 | 6699 | 0.16 | 0.11 | 1.45E-01 | 5.13E-01 | 77 | 0.14 | 0.08 | 6.76E-02 | 0.00 | 8.07E-01 | - | - |
| Average portion size of vegetables based on photographic illustration of four sizes (smallest to largest) | Food | 12863 | -0.07 | 0.08 | 4.14E-01 | 7.98E-01 | 141 | 12614 | -0.15 | 0.09 | 7.64E-02 | 3.92E-01 | 53 | -0.11 | 0.06 | 6.83E-02 | 0.00 | 4.86E-01 | - | - |
| Pain during the last four weeks | General health | 191 | 0.47 | 0.64 | 4.57E-01 | 8.32E-01 | 148 | 162 | 1.16 | 0.66 | 7.90E-02 | 3.92E-01 | 54 | 0.81 | 0.46 | 7.76E-02 | 0.00 | 4.51E-01 | - | - |
| Self rate of overall health compared to a year ago | General health | 191 | 0.09 | 0.64 | 8.91E-01 | 9.84E-01 | 247 | 160 | -1.62 | 0.65 | 1.33E-02 | 1.51E-01 | 24 | -0.76 | 0.46 | 9.55E-02 | 0.72 | 6.07E-02 | - | - |
| Zinc intake (mg/day) | Nutrients | 12863 | -0.13 | 0.09 | 1.25E-01 | 5.41E-01 | 63 | 12614 | -0.07 | 0.09 | 4.54E-01 | 7.50E-01 | 164 | -0.10 | 0.06 | 1.05E-01 | 0.00 | 5.94E-01 | - | - |
| Contradictory demands in job | Psychosocial | 14323 | -0.13 | 0.08 | 1.01E-01 | 4.99E-01 | 54 | 14057 | -0.05 | 0.08 | 5.23E-01 | 7.87E-01 | 179 | -0.09 | 0.06 | 1.06E-01 | 0.00 | 4.91E-01 | - | - |
| For how much of the time during the last four weeks have you felt gloomy and sad? | General health | 190 | -1.22 | 0.67 | 6.87E-02 | 4.53E-01 | 40 | 161 | -0.30 | 0.67 | 6.55E-01 | 8.63E-01 | 206 | -0.76 | 0.47 | 1.07E-01 | 0.00 | 3.30E-01 | - | - |
| Work shifts/weekends | Psychosocial | 13658 | 0.10 | 0.18 | 5.83E-01 | 8.56E-01 | 186 | 13402 | 0.31 | 0.18 | 8.33E-02 | 3.92E-01 | 58 | 0.20 | 0.13 | 1.08E-01 | 0.00 | 3.98E-01 | - | - |
| Smoking status: Former occasional smokers vs non-smokers | Tobacco use | 8358 | -0.29 | 0.27 | 2.92E-01 | 7.04E-01 | 112 | 8181 | -0.35 | 0.28 | 2.24E-01 | 6.33E-01 | 95 | -0.32 | 0.20 | 1.09E-01 | 0.00 | 8.87E-01 | - | - |
| Pinoresinol intake (ug/day) | Nutrients | 12863 | -0.14 | 0.08 | 9.48E-02 | 4.99E-01 | 51 | 12614 | -0.05 | 0.09 | 5.59E-01 | 8.20E-01 | 186 | -0.10 | 0.06 | 1.09E-01 | 0.00 | 4.52E-01 | - | - |
| Physical limitation to participate in moderately demanding activities: lifting or carrying grocery bags | General health | 189 | 0.99 | 0.66 | 1.38E-01 | 5.56E-01 | 68 | 162 | 0.50 | 0.68 | 4.56E-01 | 7.50E-01 | 165 | 0.75 | 0.47 | 1.13E-01 | 0.00 | 6.10E-01 | - | - |
| Average portion size of meat/fish based on photographic illustration of four sizes (smallest to largest) | Food | 12863 | -0.10 | 0.09 | 2.72E-01 | 6.96E-01 | 106 | 12614 | -0.10 | 0.09 | 2.59E-01 | 6.73E-01 | 104 | -0.10 | 0.06 | 1.15E-01 | 0.00 | 9.70E-01 | - | - |
| Patience status | Psychosocial | 6840 | 0.17 | 0.11 | 1.23E-01 | 5.41E-01 | 61 | 6712 | 0.07 | 0.11 | 5.00E-01 | 7.67E-01 | 178 | 0.12 | 0.08 | 1.16E-01 | 0.00 | 5.46E-01 | - | - |
| Lariciresinol intake (ug/day) | Nutrients | 12863 | -0.11 | 0.09 | 1.77E-01 | 6.25E-01 | 77 | 12614 | -0.07 | 0.09 | 3.90E-01 | 7.30E-01 | 143 | -0.10 | 0.06 | 1.17E-01 | 0.00 | 7.42E-01 | - | - |

| | | | | | | | | | | | | | | | | | | | | |
|---|---|---|---|---|---|---|---|---|---|---|---|---|---|---|---|---|---|---|---|---|
| Breakfast habits: Coffee/tea and wheat buns or rusk for breakfast vs not breakfast at all | Food | 999 | -0.82 | 0.69 | 2.34E-01 | 6.87E-01 | 91 | 964 | -0.70 | 0.72 | 3.27E-01 | 7.13E-01 | 125 | -0.76 | 0.50 | 1.24E-01 | 0.00 | 9.02E-01 | - | - |
| Oil for cooking | Food | 12863 | -0.08 | 0.08 | 3.55E-01 | 7.70E-01 | 124 | 12614 | -0.11 | 0.08 | 2.10E-01 | 6.24E-01 | 91 | -0.09 | 0.06 | 1.24E-01 | 0.00 | 7.97E-01 | - | - |
| Receive hugs to comfort and support you | Social | 14384 | -0.10 | 0.21 | 6.47E-01 | 8.74E-01 | 202 | 14116 | -0.36 | 0.22 | 9.08E-02 | 4.20E-01 | 59 | -0.23 | 0.15 | 1.31E-01 | 0.00 | 3.74E-01 | - | - |
| Strong beer | Alcohol | 12863 | 0.02 | 0.09 | 8.44E-01 | 9.61E-01 | 239 | 12614 | 0.17 | 0.09 | 5.51E-02 | 3.34E-01 | 45 | 0.09 | 0.06 | 1.36E-01 | 0.33 | 2.21E-01 | - | - |
| Sum of Lariciresinol, Matairesinol, Pinoresinol, Secoisolariciresinol intake (ug/day) | Nutrients | 12863 | -0.11 | 0.08 | 1.75E-01 | 6.25E-01 | 76 | 12614 | -0.06 | 0.09 | 4.59E-01 | 7.50E-01 | 167 | -0.09 | 0.06 | 1.37E-01 | 0.00 | 6.75E-01 | - | - |
| Linoleic acid intake (g/day) | Nutrients | 12863 | -0.06 | 0.08 | 4.29E-01 | 8.13E-01 | 144 | 12614 | -0.11 | 0.08 | 2.01E-01 | 6.17E-01 | 89 | -0.09 | 0.06 | 1.45E-01 | 0.00 | 7.13E-01 | - | - |
| Travel to work: Walk to work vs passive travel to work | Physical activity | 9304 | -0.39 | 0.29 | 1.83E-01 | 6.30E-01 | 79 | 9116 | -0.21 | 0.31 | 4.92E-01 | 7.63E-01 | 175 | -0.30 | 0.21 | 1.51E-01 | 0.00 | 6.67E-01 | - | - |
| Eicosapentaenoic acid (EPA) intake (g/day) | Nutrients | 12863 | -0.10 | 0.08 | 2.38E-01 | 6.87E-01 | 93 | 12614 | -0.07 | 0.08 | 3.99E-01 | 7.30E-01 | 149 | -0.08 | 0.06 | 1.52E-01 | 0.00 | 8.27E-01 | - | - |
| Sodas, soft drinks, juice | Beverage | 12863 | 0.06 | 0.08 | 4.79E-01 | 8.32E-01 | 156 | 12614 | 0.11 | 0.09 | 1.91E-01 | 6.03E-01 | 86 | 0.08 | 0.06 | 1.55E-01 | 0.00 | 6.60E-01 | - | - |
| Matairesinol intake (ug/day) | Nutrients | 12863 | -0.15 | 0.08 | 6.98E-02 | 4.53E-01 | 41 | 12614 | -0.01 | 0.09 | 8.65E-01 | 9.67E-01 | 243 | -0.08 | 0.06 | 1.57E-01 | 0.24 | 2.52E-01 | - | - |
| Number of cigars smoked per day | Tobacco use | 9013 | -0.14 | 0.10 | 1.55E-01 | 5.92E-01 | 71 | 8787 | -0.05 | 0.10 | 5.83E-01 | 8.34E-01 | 190 | -0.10 | 0.07 | 1.61E-01 | 0.00 | 5.51E-01 | - | - |
| Enterolactone intake (ug/day) | Nutrients | 12863 | -0.13 | 0.08 | 1.14E-01 | 5.19E-01 | 60 | 12614 | -0.03 | 0.08 | 7.30E-01 | 9.14E-01 | 217 | -0.08 | 0.06 | 1.70E-01 | 0.00 | 3.92E-01 | - | - |
| Sour milk, yoghurt (low fat) | Food | 12863 | -0.14 | 0.08 | 8.10E-02 | 4.61E-01 | 48 | 12614 | -0.01 | 0.08 | 8.73E-01 | 9.67E-01 | 245 | -0.08 | 0.06 | 1.74E-01 | 0.18 | 2.69E-01 | - | - |
| Monosaccharides intake (g/day) | Nutrients | 12863 | -0.09 | 0.09 | 3.01E-01 | 7.04E-01 | 115 | 12614 | -0.08 | 0.09 | 3.80E-01 | 7.30E-01 | 138 | -0.08 | 0.06 | 1.76E-01 | 0.00 | 9.24E-01 | - | - |
| High mental demand from job | Psychosocial | 14346 | 0.16 | 0.08 | 3.78E-02 | 3.68E-01 | 28 | 14075 | -0.02 | 0.08 | 8.43E-01 | 9.67E-01 | 238 | 0.07 | 0.06 | 1.77E-01 | 0.61 | 1.11E-01 | - | - |
| Polyunsaturated fat intake (g/day) | Nutrients | 12863 | -0.06 | 0.08 | 4.89E-01 | 8.39E-01 | 159 | 12614 | -0.10 | 0.08 | 2.29E-01 | 6.33E-01 | 98 | -0.08 | 0.06 | 1.82E-01 | 0.00 | 7.02E-01 | - | - |
| Wine | Alcohol | 12863 | 0.05 | 0.08 | 5.59E-01 | 8.48E-01 | 177 | 12614 | 0.11 | 0.08 | 1.92E-01 | 6.03E-01 | 87 | 0.08 | 0.06 | 1.83E-01 | 0.00 | 6.04E-01 | - | - |
| Steak, chop, e.g. | Food | 12863 | 0.05 | 0.08 | 5.61E-01 | 8.48E-01 | 178 | 12614 | 0.11 | 0.08 | 2.14E-01 | 6.27E-01 | 93 | 0.08 | 0.06 | 1.99E-01 | 0.00 | 6.27E-01 | - | - |
| Frequency of shoverling snow during leisure time | Physical activity | 13730 | -0.03 | 0.08 | 7.52E-01 | 9.48E-01 | 215 | 13517 | -0.13 | 0.08 | 1.30E-01 | 5.04E-01 | 69 | -0.08 | 0.06 | 1.99E-01 | 0.00 | 3.88E-01 | - | - |
| Participation in other association | Social | 773 | 0.00 | 0.60 | 9.97E-01 | 9.98E-01 | 272 | 687 | -1.24 | 0.66 | 5.99E-02 | 3.46E-01 | 47 | -0.57 | 0.45 | 2.03E-01 | 0.48 | 1.64E-01 | - | - |
| Ingenuity or creativity demand from job | Psychosocial | 14378 | -0.06 | 0.08 | 4.63E-01 | 8.32E-01 | 151 | 14123 | -0.08 | 0.08 | 2.89E-01 | 6.85E-01 | 115 | -0.07 | 0.05 | 2.06E-01 | 0.00 | 8.05E-01 | - | - |
| Carbohydrates intake (g/day) | Nutrients | 12863 | -0.03 | 0.08 | 7.43E-01 | 9.48E-01 | 213 | 12614 | -0.13 | 0.09 | 1.41E-01 | 5.06E-01 | 76 | -0.08 | 0.06 | 2.06E-01 | 0.00 | 4.10E-01 | - | - |
| Low fat margarine on bread | Food | 12863 | 0.06 | 0.08 | 4.78E-01 | 8.32E-01 | 155 | 12614 | 0.09 | 0.08 | 2.79E-01 | 6.79E-01 | 112 | 0.07 | 0.06 | 2.06E-01 | 0.00 | 7.82E-01 | - | - |
| Sodium intake (mg/day) | Nutrients | 12863 | -0.02 | 0.08 | 8.50E-01 | 9.61E-01 | 241 | 12614 | -0.14 | 0.09 | 1.12E-01 | 4.62E-01 | 66 | -0.07 | 0.06 | 2.13E-01 | 0.01 | 3.14E-01 | - | - |
| Vitamin B2 intake (ug/day) | Nutrients | 12863 | -0.06 | 0.08 | 4.27E-01 | 8.13E-01 | 143 | 12614 | -0.08 | 0.08 | 3.39E-01 | 7.13E-01 | 129 | -0.07 | 0.06 | 2.16E-01 | 0.00 | 8.97E-01 | - | - |
| For how much of the time during the last four weeks have you felt worn out? | General health | 191 | 1.08 | 0.65 | 9.80E-02 | 4.99E-01 | 53 | 159 | 0.02 | 0.70 | 9.81E-01 | 9.92E-01 | 268 | 0.59 | 0.48 | 2.17E-01 | 0.19 | 2.65E-01 | - | - |
| Potato salad | Food | 5271 | 0.07 | 0.13 | 5.82E-01 | 8.56E-01 | 185 | 5156 | 0.16 | 0.14 | 2.29E-01 | 6.33E-01 | 99 | 0.11 | 0.09 | 2.20E-01 | 0.00 | 6.23E-01 | - | - |
| Skill demand from job | Psychosocial | 14385 | -0.03 | 0.08 | 6.82E-01 | 8.99E-01 | 207 | 14137 | -0.11 | 0.08 | 1.83E-01 | 6.03E-01 | 83 | -0.07 | 0.06 | 2.21E-01 | 0.00 | 5.06E-01 | - | - |
| Vitamin A intake (mg/day) | Nutrients | 12863 | 0.11 | 0.09 | 2.48E-01 | 6.89E-01 | 98 | 12614 | 0.05 | 0.09 | 6.06E-01 | 8.49E-01 | 194 | 0.08 | 0.07 | 2.37E-01 | 0.00 | 6.54E-01 | - | - |
| Meat stew | Food | 12863 | -0.05 | 0.08 | 5.71E-01 | 8.48E-01 | 184 | 12614 | -0.09 | 0.08 | 2.65E-01 | 6.73E-01 | 105 | -0.07 | 0.06 | 2.37E-01 | 0.00 | 6.84E-01 | - | - |
| Milk, sour milk (1,5%) | Beverage | 12863 | 0.16 | 0.08 | 5.29E-02 | 4.13E-01 | 35 | 12614 | -0.03 | 0.08 | 7.59E-01 | 9.23E-01 | 224 | 0.07 | 0.06 | 2.42E-01 | 0.60 | 1.16E-01 | - | - |
| Plant based protein intake (g/day) | Nutrients | 12863 | -0.04 | 0.08 | 6.43E-01 | 8.74E-01 | 199 | 12614 | -0.10 | 0.09 | 2.48E-01 | 6.57E-01 | 103 | -0.07 | 0.06 | 2.54E-01 | 0.00 | 6.16E-01 | - | - |
| Beta-sitosterol intake (mg/day) | Nutrients | 12863 | -0.02 | 0.08 | 7.64E-01 | 9.48E-01 | 219 | 12614 | -0.11 | 0.08 | 1.89E-01 | 6.03E-01 | 85 | -0.07 | 0.06 | 2.57E-01 | 0.00 | 4.66E-01 | - | - |
| Snuff status: Former snuff users vs non-snuff users | Tobacco use | 11326 | -0.15 | 0.26 | 5.68E-01 | 8.48E-01 | 182 | 11190 | -0.28 | 0.27 | 2.99E-01 | 6.95E-01 | 117 | -0.22 | 0.19 | 2.57E-01 | 0.00 | 7.27E-01 | - | - |
| Parents or siblings have diabetes | General health | 14370 | -0.06 | 0.21 | 7.87E-01 | 9.55E-01 | 224 | 14132 | 0.39 | 0.21 | 6.19E-02 | 3.46E-01 | 48 | 0.17 | 0.15 | 2.61E-01 | 0.56 | 1.30E-01 | - | - |
| Margarine on bread | Food | 12863 | 0.07 | 0.08 | 3.65E-01 | 7.70E-01 | 128 | 12614 | 0.06 | 0.08 | 4.95E-01 | 7.63E-01 | 177 | 0.06 | 0.06 | 2.61E-01 | 0.00 | 8.79E-01 | - | - |
| Marital status: Single vs Married/partner | Social | 13588 | 0.29 | 0.26 | 2.71E-01 | 6.96E-01 | 104 | 13343 | 0.13 | 0.27 | 6.31E-01 | 8.52E-01 | 201 | 0.21 | 0.19 | 2.61E-01 | 0.00 | 6.69E-01 | - | - |
| Milk, sour milk (3%) | Beverage | 12863 | -0.08 | 0.08 | 3.02E-01 | 7.04E-01 | 117 | 12614 | -0.04 | 0.08 | 6.13E-01 | 8.49E-01 | 197 | -0.06 | 0.06 | 2.74E-01 | 0.00 | 7.22E-01 | - | - |
| Cohabitation: Live alone vs Only children | Social | 1928 | 0.83 | 0.49 | 9.34E-02 | 4.99E-01 | 50 | 1899 | -0.09 | 0.51 | 8.63E-01 | 9.67E-01 | 242 | 0.39 | 0.36 | 2.75E-01 | 0.40 | 1.98E-01 | - | - |
| Confidence status | Psychosocial | 6835 | 0.05 | 0.11 | 6.39E-01 | 8.74E-01 | 198 | 6710 | 0.12 | 0.11 | 2.81E-01 | 6.79E-01 | 113 | 0.08 | 0.08 | 2.75E-01 | 0.00 | 6.63E-01 | - | - |
| Smoking status: Former smokers vs non-smokers | Tobacco use | 9944 | -0.15 | 0.20 | 4.71E-01 | 8.32E-01 | 152 | 9848 | -0.17 | 0.21 | 4.18E-01 | 7.36E-01 | 154 | -0.16 | 0.14 | 2.79E-01 | 0.00 | 9.42E-01 | - | - |
| Tea | Beverage | 12863 | -0.14 | 0.08 | 8.41E-02 | 4.69E-01 | 49 | 12614 | 0.02 | 0.08 | 8.20E-01 | 9.59E-01 | 232 | -0.06 | 0.06 | 2.83E-01 | 0.47 | 1.70E-01 | - | - |
| Apple, pear, peach, orange, mandarin and grapefruit | Food | 12863 | -0.06 | 0.09 | 4.75E-01 | 8.32E-01 | 154 | 12614 | -0.07 | 0.09 | 4.21E-01 | 7.36E-01 | 156 | -0.07 | 0.06 | 2.83E-01 | 0.00 | 9.40E-01 | - | - |
| Root vegetables, carrot | Food | 12863 | -0.06 | 0.08 | 5.16E-01 | 8.48E-01 | 165 | 12614 | -0.08 | 0.09 | 3.84E-01 | 7.30E-01 | 140 | -0.07 | 0.06 | 2.83E-01 | 0.00 | 8.68E-01 | - | - |
| Beta-carotene intake (mg/day) | Nutrients | 12863 | -0.04 | 0.09 | 6.04E-01 | 8.66E-01 | 190 | 12614 | -0.09 | 0.09 | 3.20E-01 | 7.11E-01 | 123 | -0.07 | 0.06 | 2.86E-01 | 0.00 | 7.30E-01 | - | - |
| Close relationship with anyone | Social | 14450 | -0.09 | 0.08 | 2.71E-01 | 6.96E-01 | 105 | 14190 | -0.03 | 0.08 | 6.91E-01 | 8.90E-01 | 212 | -0.06 | 0.06 | 2.87E-01 | 0.00 | 6.26E-01 | - | - |
| For how much of the time during the last four weeks have you felt full of energy? | General health | 191 | -0.40 | 0.64 | 5.39E-01 | 8.48E-01 | 171 | 161 | -0.58 | 0.68 | 3.97E-01 | 7.30E-01 | 147 | -0.48 | 0.47 | 3.03E-01 | 0.00 | 8.46E-01 | - | - |
| Sometimes physically straining work | Physical activity | 14214 | 0.17 | 0.17 | 3.25E-01 | 7.46E-01 | 119 | 13916 | 0.08 | 0.17 | 6.44E-01 | 8.62E-01 | 204 | 0.12 | 0.12 | 3.04E-01 | 0.00 | 7.23E-01 | - | - |
| Worsen in health in the future | General health | 188 | 0.04 | 0.64 | 9.49E-01 | 9.93E-01 | 258 | 160 | 0.96 | 0.68 | 1.57E-01 | 5.46E-01 | 78 | 0.47 | 0.46 | 3.07E-01 | 0.00 | 3.21E-01 | - | - |
| White cabbage, lettuce, lettuce cabbage, spinach, borecole | Food | 12863 | -0.01 | 0.09 | 9.02E-01 | 9.87E-01 | 249 | 12614 | -0.12 | 0.09 | 1.87E-01 | 6.03E-01 | 84 | -0.06 | 0.06 | 3.12E-01 | 0.00 | 3.91E-01 | - | - |
| Sour milk, yoghurt (3% fat) | Food | 12863 | -0.12 | 0.08 | 1.52E-01 | 5.92E-01 | 70 | 12614 | 0.00 | 0.08 | 9.84E-01 | 9.92E-01 | 270 | -0.06 | 0.06 | 3.13E-01 | 0.03 | 3.09E-01 | - | - |
| Margarine for cooking | Food | 12863 | 0.04 | 0.08 | 5.87E-01 | 8.57E-01 | 187 | 12614 | 0.07 | 0.08 | 3.78E-01 | 7.30E-01 | 137 | 0.06 | 0.06 | 3.15E-01 | 0.00 | 8.03E-01 | - | - |
| Fibre intake (g/day) | Nutrients | 12863 | -0.05 | 0.09 | 5.57E-01 | 8.48E-01 | 176 | 12614 | -0.07 | 0.09 | 4.05E-01 | 7.32E-01 | 150 | -0.06 | 0.06 | 3.17E-01 | 0.00 | 8.53E-01 | - | - |
| Folic acid intake (ug/day) | Nutrients | 12863 | -0.03 | 0.09 | 7.47E-01 | 9.48E-01 | 214 | 12614 | -0.10 | 0.09 | 2.71E-01 | 6.73E-01 | 110 | -0.06 | 0.06 | 3.17E-01 | 0.00 | 5.76E-01 | - | - |
| Cambridge physical activity index | Physical activity | 14169 | -0.04 | 0.08 | 6.22E-01 | 8.72E-01 | 194 | 13872 | -0.07 | 0.08 | 3.57E-01 | 7.30E-01 | 133 | -0.06 | 0.06 | 3.20E-01 | 0.00 | 7.52E-01 | - | - |
| Docosahexaenoic acid (DHA) intake (g/day) | Nutrients | 12863 | -0.07 | 0.08 | 3.71E-01 | 7.70E-01 | 130 | 12614 | -0.04 | 0.08 | 6.12E-01 | 8.49E-01 | 196 | -0.06 | 0.06 | 3.20E-01 | 0.00 | 7.93E-01 | - | - |
| White meat (poultry) | Food | 12863 | 0.04 | 0.09 | 6.64E-01 | 8.84E-01 | 205 | 12614 | -0.15 | 0.08 | 7.48E-02 | 3.92E-01 | 51 | -0.06 | 0.06 | 3.27E-01 | 0.58 | 1.21E-01 | - | - |
| Vitamin E intake (mg/day) | Nutrients | 12863 | 0.07 | 0.08 | 4.15E-01 | 7.98E-01 | 142 | 12614 | 0.05 | 0.08 | 5.69E-01 | 8.26E-01 | 188 | 0.06 | 0.06 | 3.27E-01 | 0.00 | 8.68E-01 | - | - |
| Frequent social contacts with colleagues during work | Psychosocial | 13882 | 0.07 | 0.08 | 3.59E-01 | 7.70E-01 | 126 | 13637 | 0.03 | 0.08 | 6.85E-01 | 8.86E-01 | 211 | 0.05 | 0.06 | 3.48E-01 | 0.00 | 7.26E-01 | - | - |
| Campestanol intake (mg/day) | Nutrients | 12863 | -0.10 | 0.08 | 2.16E-01 | 6.87E-01 | 86 | 12614 | -0.01 | 0.08 | 9.38E-01 | 9.85E-01 | 260 | -0.05 | 0.06 | 3.49E-01 | 0.00 | 4.18E-01 | - | - |
| Disaccharides intake (g/day) | Nutrients | 12863 | 0.00 | 0.08 | 9.95E-01 | 9.98E-01 | 271 | 12614 | -0.11 | 0.08 | 1.78E-01 | 5.93E-01 | 82 | -0.06 | 0.06 | 3.50E-01 | 0.00 | 3.33E-01 | - | - |
| Potassium intake (mg/day) | Nutrients | 12863 | 0.01 | 0.09 | 9.44E-01 | 9.93E-01 | 256 | 12614 | -0.13 | 0.09 | 1.60E-01 | 5.47E-01 | 80 | -0.06 | 0.06 | 3.51E-01 | 0.10 | 2.93E-01 | - | - |
| Number of friends that can come to your home at any time and feel at home | Social | 14416 | -0.04 | 0.08 | 5.63E-01 | 8.48E-01 | 180 | 14162 | -0.06 | 0.08 | 4.63E-01 | 7.53E-01 | 168 | -0.05 | 0.05 | 3.54E-01 | 0.00 | 9.05E-01 | - | - |
| Enterodiol intake (ug/day) | Nutrients | 12863 | -0.07 | 0.08 | 3.96E-01 | 7.78E-01 | 139 | 12614 | -0.04 | 0.08 | 6.64E-01 | 8.63E-01 | 210 | -0.05 | 0.06 | 3.62E-01 | 0.00 | 7.79E-01 | - | - |
| Blood based food | Food | 6463 | -0.13 | 0.12 | 2.58E-01 | 6.89E-01 | 101 | 6332 | -0.02 | 0.12 | 8.95E-01 | 9.67E-01 | 252 | -0.08 | 0.08 | 3.64E-01 | 0.00 | 4.90E-01 | - | - |
| Tomato, cucumber | Food | 12863 | -0.04 | 0.09 | 6.50E-01 | 8.74E-01 | 203 | 12614 | -0.07 | 0.09 | 4.05E-01 | 7.32E-01 | 151 | -0.06 | 0.06 | 3.65E-01 | 0.00 | 7.80E-01 | - | - |
| Frequency of hunting or fishing during leisure time | Physical activity | 13267 | 0.02 | 0.09 | 8.25E-01 | 9.61E-01 | 232 | 13062 | 0.09 | 0.09 | 2.86E-01 | 6.85E-01 | 114 | 0.06 | 0.06 | 3.67E-01 | 0.00 | 5.41E-01 | - | - |
| Feel the need to reduce alcohol consumption | Alcohol | 12887 | 0.17 | 0.26 | 5.28E-01 | 8.48E-01 | 168 | 12725 | 0.17 | 0.27 | 5.23E-01 | 7.87E-01 | 180 | 0.17 | 0.19 | 3.69E-01 | 0.00 | 9.92E-01 | - | - |
| Vitamin C intake (mg/day) | Nutrients | 12863 | 0.08 | 0.08 | 3.70E-01 | 7.70E-01 | 129 | 12614 | 0.03 | 0.09 | 7.24E-01 | 9.14E-01 | 214 | 0.05 | 0.06 | 3.74E-01 | 0.00 | 7.08E-01 | - | - |
| Equol intake (ug/day) | Nutrients | 12863 | -0.08 | 0.08 | 3.01E-01 | 7.04E-01 | 116 | 12614 | -0.02 | 0.08 | 8.52E-01 | 9.67E-01 | 239 | -0.05 | 0.06 | 3.83E-01 | 0.00 | 5.56E-01 | - | - |
| Do you feel important and appreciated outside your home? | Psychosocial | 6846 | 0.05 | 0.11 | 6.61E-01 | 8.84E-01 | 204 | 6718 | 0.09 | 0.11 | 4.31E-01 | 7.40E-01 | 158 | 0.07 | 0.08 | 3.86E-01 | 0.00 | 8.03E-01 | - | - |
| Mood status | Psychosocial | 6839 | 0.06 | 0.11 | 5.66E-01 | 8.48E-01 | 181 | 6709 | 0.07 | 0.11 | 5.26E-01 | 7.87E-01 | 182 | 0.07 | 0.08 | 3.93E-01 | 0.00 | 9.61E-01 | - | - |
| People to ask for help apart from the ones at home | Social | 14534 | 0.39 | 0.25 | 1.24E-01 | 5.41E-01 | 62 | 14287 | -0.09 | 0.25 | 7.34E-01 | 9.14E-01 | 219 | 0.15 | 0.18 | 3.98E-01 | 0.43 | 1.84E-01 | - | - |
| Lean fish (e.g. perch, bass, cod) | Food | 12863 | -0.07 | 0.08 | 3.84E-01 | 7.71E-01 | 136 | 12614 | -0.02 | 0.08 | 7.70E-01 | 9.23E-01 | 227 | -0.05 | 0.06 | 4.07E-01 | 0.00 | 6.94E-01 | - | - |
| Selenium intake (ug/day) | Nutrients | 12863 | -0.02 | 0.08 | 7.66E-01 | 9.48E-01 | 220 | 12614 | -0.07 | 0.08 | 3.81E-01 | 7.30E-01 | 139 | -0.05 | 0.06 | 4.09E-01 | 0.00 | 6.75E-01 | - | - |
| Butter for cooking | Food | 12863 | 0.04 | 0.08 | 5.94E-01 | 8.63E-01 | 188 | 12614 | 0.05 | 0.08 | 5.31E-01 | 7.87E-01 | 184 | 0.05 | 0.06 | 4.13E-01 | 0.00 | 9.39E-01 | - | - |
| Years smoking | Tobacco use | 12837 | 0.00 | 0.08 | 9.58E-01 | 9.93E-01 | 263 | 12626 | -0.11 | 0.09 | 2.21E-01 | 6.33E-01 | 94 | -0.05 | 0.06 | 4.14E-01 | 0.00 | 3.62E-01 | - | - |
| Calcium intake (mg/day) | Nutrients | 12863 | -0.11 | 0.08 | 1.79E-01 | 6.25E-01 | 78 | 12614 | 0.02 | 0.08 | 8.30E-01 | 9.59E-01 | 235 | -0.05 | 0.06 | 4.17E-01 | 0.17 | 2.74E-01 | - | - |
| How much has the pain during the last four weeks disturbed your normal work? | General health | 190 | 0.23 | 0.64 | 7.20E-01 | 9.31E-01 | 211 | 162 | 0.52 | 0.66 | 4.31E-01 | 7.40E-01 | 159 | 0.37 | 0.46 | 4.20E-01 | 0.00 | 7.50E-01 | - | - |
| Self-employed | Psychosocial | 14188 | -0.33 | 0.27 | 2.14E-01 | 6.87E-01 | 84 | 13952 | 0.04 | 0.28 | 8.94E-01 | 9.67E-01 | 251 | -0.15 | 0.19 | 4.22E-01 | 0.00 | 3.38E-01 | - | - |
| Physical limitation to participate in moderately demanding activities: moving a table, vacuuming, walking in the forest or gardening | General health | 190 | -0.18 | 0.64 | 7.84E-01 | 9.55E-01 | 223 | 162 | 0.94 | 0.66 | 1.58E-01 | 5.46E-01 | 79 | 0.37 | 0.46 | 4.27E-01 | 0.31 | 2.27E-01 | - | - |
| Sum of phytosterols intake (mg/day) | Nutrients | 12863 | -0.01 | 0.08 | 9.40E-01 | 9.93E-01 | 255 | 12614 | -0.09 | 0.08 | 2.91E-01 | 6.85E-01 | 116 | -0.05 | 0.06 | 4.28E-01 | 0.00 | 4.83E-01 | - | - |
| Shellfish (e.g. shrimps, scallops) | Food | 6463 | 0.05 | 0.12 | 6.81E-01 | 8.99E-01 | 206 | 6332 | 0.09 | 0.12 | 4.76E-01 | 7.53E-01 | 172 | 0.07 | 0.09 | 4.29E-01 | 0.00 | 8.21E-01 | - | - |
| Vitamin D intake (ug/day) | Nutrients | 12863 | 0.10 | 0.08 | 2.28E-01 | 6.87E-01 | 90 | 12614 | -0.01 | 0.08 | 9.16E-01 | 9.70E-01 | 258 | 0.05 | 0.06 | 4.30E-01 | 0.00 | 3.60E-01 | - | - |
| Soft cheese | Food | 6463 | -0.01 | 0.12 | 9.52E-01 | 9.93E-01 | 260 | 6332 | 0.14 | 0.12 | 2.37E-01 | 6.40E-01 | 101 | 0.06 | 0.08 | 4.35E-01 | 0.00 | 3.73E-01 | - | - |
| Satisfaction with economy | Psychosocial | 6845 | -0.13 | 0.11 | 2.22E-01 | 6.87E-01 | 87 | 6706 | 0.01 | 0.11 | 8.94E-01 | 9.67E-01 | 249 | -0.06 | 0.08 | 4.40E-01 | 0.00 | 3.40E-01 | - | - |
| For how much of the time during the last four weeks has your physical health or your emotional problems limited your ability to interact with others? | General health | 189 | -0.23 | 0.63 | 7.17E-01 | 9.31E-01 | 210 | 161 | -0.49 | 0.67 | 4.71E-01 | 7.53E-01 | 170 | -0.35 | 0.46 | 4.47E-01 | 0.00 | 7.81E-01 | - | - |
| Marital status: Single vs Widow/widower | Social | 1528 | 0.99 | 1.17 | 3.95E-01 | 7.78E-01 | 138 | 1511 | 0.23 | 1.05 | 8.25E-01 | 9.59E-01 | 233 | 0.57 | 0.78 | 4.63E-01 | 0.00 | 6.29E-01 | - | - |
| Physical limitation to participate in steneous activities: running, lifting heavy objects, taking part in physically demanding sports | General health | 190 | 0.08 | 0.66 | 9.07E-01 | 9.87E-01 | 251 | 162 | 0.64 | 0.70 | 3.61E-01 | 7.30E-01 | 135 | 0.34 | 0.48 | 4.76E-01 | 0.00 | 5.58E-01 | - | - |

| | | | | | | | | | | | | | | | | | | | | |
|---|---|---|---|---|---|---|---|---|---|---|---|---|---|---|---|---|---|---|---|---|
| Appetite status | Psychosocial | 6840 | -0.05 | 0.11 | 6.25E-01 | 8.72E-01 | 195 | 6710 | -0.05 | 0.11 | 6.33E-01 | 8.52E-01 | 202 | -0.05 | 0.08 | 4.94E-01 | 0.00 | 9.95E-01 | - | - |
| Satisfaction with accomodation | Psychosocial | 6850 | 0.00 | 0.11 | 9.98E-01 | 9.98E-01 | 273 | 6718 | 0.11 | 0.11 | 3.39E-01 | 7.13E-01 | 128 | 0.05 | 0.08 | 4.99E-01 | 0.00 | 4.99E-01 | - | - |
| Distance to work in kilometers (one way) | Physical activity | 12759 | 0.05 | 0.08 | 5.39E-01 | 8.48E-01 | 170 | 12587 | -0.14 | 0.08 | 1.08E-01 | 4.54E-01 | 65 | -0.04 | 0.06 | 5.00E-01 | 0.60 | 1.14E-01 | - | - |
| Marital status: Single vs Divorced/separated | Social | 2335 | 0.80 | 0.43 | 6.58E-02 | 4.53E-01 | 39 | 2289 | -0.42 | 0.44 | 3.45E-01 | 7.14E-01 | 132 | 0.20 | 0.31 | 5.09E-01 | 0.74 | 5.00E-02 | - | - |
| Fat intake (g/day) | Nutrients | 12863 | 0.02 | 0.08 | 8.30E-01 | 9.61E-01 | 234 | 12614 | 0.06 | 0.09 | 4.81E-01 | 7.54E-01 | 174 | 0.04 | 0.06 | 5.18E-01 | 0.00 | 7.23E-01 | - | - |
| Travel to work: Irregular travel mode to work vs passive travel to work | Physical activity | 9513 | -0.36 | 0.27 | 1.85E-01 | 6.30E-01 | 80 | 9359 | 0.13 | 0.28 | 6.53E-01 | 8.63E-01 | 205 | -0.13 | 0.20 | 5.23E-01 | 0.36 | 2.12E-01 | - | - |
| For how much of the time during the last four weeks have you felt really alert and strong? | General health | 192 | -0.08 | 0.66 | 9.07E-01 | 9.87E-01 | 250 | 162 | -0.52 | 0.67 | 4.36E-01 | 7.44E-01 | 160 | -0.29 | 0.47 | 5.28E-01 | 0.00 | 6.36E-01 | - | - |
| Beta-sitostanol intake (mg/day) | Nutrients | 12863 | -0.05 | 0.08 | 5.03E-01 | 8.48E-01 | 160 | 12614 | -0.02 | 0.08 | 8.33E-01 | 9.59E-01 | 237 | -0.04 | 0.06 | 5.31E-01 | 0.00 | 7.51E-01 | - | - |
| Stigmasterol intake (mg/day) | Nutrients | 12863 | 0.03 | 0.08 | 7.56E-01 | 9.48E-01 | 217 | 12614 | -0.10 | 0.09 | 2.26E-01 | 6.33E-01 | 96 | -0.04 | 0.06 | 5.32E-01 | 0.15 | 2.79E-01 | - | - |
| Number of people with whom you can speak openly | Social | 14564 | 0.01 | 0.08 | 9.32E-01 | 9.93E-01 | 254 | 14314 | -0.07 | 0.08 | 3.39E-01 | 7.13E-01 | 127 | -0.03 | 0.05 | 5.42E-01 | 0.00 | 4.58E-01 | - | - |
| Trans fat intake (g/day) | Nutrients | 12863 | -0.11 | 0.10 | 3.00E-01 | 7.04E-01 | 114 | 12614 | 0.02 | 0.10 | 8.53E-01 | 9.67E-01 | 240 | -0.04 | 0.07 | 5.46E-01 | 0.00 | 3.89E-01 | - | - |
| Sleep status | Sleep | 6852 | 0.01 | 0.11 | 9.23E-01 | 9.93E-01 | 253 | 6722 | -0.10 | 0.11 | 3.42E-01 | 7.13E-01 | 130 | -0.05 | 0.08 | 5.49E-01 | 0.00 | 4.56E-01 | - | - |
| Possibility to leave your work for a while to speak with a colleague | Psychosocial | 13912 | -0.06 | 0.08 | 4.81E-01 | 8.32E-01 | 158 | 13633 | -0.01 | 0.08 | 8.94E-01 | 9.67E-01 | 250 | -0.03 | 0.06 | 5.51E-01 | 0.00 | 6.91E-01 | - | - |
| Do you feel important and appreciated in your home? | Psychosocial | 6799 | 0.08 | 0.11 | 4.72E-01 | 8.32E-01 | 153 | 6660 | 0.01 | 0.11 | 9.10E-01 | 9.70E-01 | 256 | 0.05 | 0.08 | 5.55E-01 | 0.00 | 6.70E-01 | - | - |
| Physical limitation to participate in moderately demanding activities: walking more than 2 km | General health | 190 | 0.43 | 0.65 | 5.10E-01 | 8.48E-01 | 163 | 161 | 0.10 | 0.70 | 8.86E-01 | 9.67E-01 | 248 | 0.28 | 0.48 | 5.60E-01 | 0.00 | 7.33E-01 | - | - |
| Feel uneasy or guilty because of your way of drinking | Alcohol | 12886 | -0.45 | 0.25 | 7.53E-02 | 4.53E-01 | 44 | 12706 | 0.25 | 0.26 | 3.19E-01 | 7.11E-01 | 122 | -0.10 | 0.18 | 5.69E-01 | 0.74 | 5.02E-02 | - | - |
| Rosehip, sweet syrup soup | Food | 12863 | 0.01 | 0.08 | 8.57E-01 | 9.63E-01 | 243 | 12614 | 0.05 | 0.08 | 5.29E-01 | 7.87E-01 | 183 | 0.03 | 0.06 | 5.70E-01 | 0.00 | 7.44E-01 | - | - |
| Berries (fresh or frozen) | Food | 12863 | -0.06 | 0.08 | 4.58E-01 | 8.32E-01 | 149 | 12614 | 0.00 | 0.08 | 9.60E-01 | 9.92E-01 | 264 | -0.03 | 0.06 | 5.72E-01 | 0.00 | 6.28E-01 | - | - |
| Fatty fish (e.g. herring, whitefish, salmon) | Food | 12863 | -0.08 | 0.08 | 3.48E-01 | 7.70E-01 | 122 | 12614 | 0.01 | 0.08 | 8.76E-01 | 9.67E-01 | 246 | -0.03 | 0.06 | 5.75E-01 | 0.00 | 4.43E-01 | - | - |
| Get sick more often than other people | General health | 191 | -0.76 | 0.63 | 2.27E-01 | 6.87E-01 | 89 | 160 | 0.32 | 0.67 | 6.33E-01 | 8.52E-01 | 203 | -0.26 | 0.46 | 5.75E-01 | 0.28 | 2.39E-01 | - | - |
| Liver, kidney | Food | 6463 | 0.00 | 0.12 | 9.92E-01 | 9.98E-01 | 270 | 6332 | -0.10 | 0.12 | 4.16E-01 | 7.36E-01 | 153 | -0.05 | 0.08 | 5.77E-01 | 0.00 | 5.53E-01 | - | - |
| Physical limitation to participate in moderately demanding activities: walking up several stairs | General health | 190 | -0.31 | 0.65 | 6.29E-01 | 8.72E-01 | 197 | 160 | -0.21 | 0.71 | 7.67E-01 | 9.23E-01 | 226 | -0.27 | 0.48 | 5.77E-01 | 0.00 | 9.15E-01 | - | - |
| Sausage as main dish | Food | 12863 | -0.04 | 0.08 | 6.43E-01 | 8.74E-01 | 200 | 12614 | -0.03 | 0.08 | 7.57E-01 | 9.23E-01 | 223 | -0.03 | 0.06 | 5.83E-01 | 0.00 | 9.19E-01 | - | - |
| Heptadecanoic acid intake (g/day) | Nutrients | 12863 | -0.07 | 0.08 | 3.80E-01 | 7.70E-01 | 133 | 12614 | 0.01 | 0.08 | 9.03E-01 | 9.67E-01 | 255 | -0.03 | 0.06 | 5.88E-01 | 0.00 | 4.83E-01 | - | - |
| Pentadecanoic acid intake (g/day) | Nutrients | 12863 | -0.07 | 0.08 | 3.80E-01 | 7.70E-01 | 134 | 12614 | 0.01 | 0.08 | 9.03E-01 | 9.67E-01 | 254 | -0.03 | 0.06 | 5.88E-01 | 0.00 | 4.83E-01 | - | - |
| Saturated fat intake (g/day) | Nutrients | 12863 | -0.01 | 0.08 | 9.20E-01 | 9.93E-01 | 252 | 12614 | 0.07 | 0.09 | 3.87E-01 | 7.30E-01 | 141 | 0.03 | 0.06 | 5.94E-01 | 0.00 | 4.91E-01 | - | - |
| Animal based protein intake (g/day) | Nutrients | 12863 | -0.01 | 0.08 | 9.45E-01 | 9.93E-01 | 257 | 12614 | 0.07 | 0.08 | 4.21E-01 | 7.36E-01 | 155 | 0.03 | 0.06 | 6.07E-01 | 0.00 | 5.33E-01 | - | - |
| Corn flakes | Food | 12863 | 0.00 | 0.08 | 9.87E-01 | 9.98E-01 | 269 | 12614 | -0.06 | 0.08 | 4.52E-01 | 7.50E-01 | 163 | -0.03 | 0.06 | 6.07E-01 | 0.00 | 5.83E-01 | - | - |
| Satisfaction with work situation | Psychosocial | 6781 | -0.05 | 0.11 | 6.45E-01 | 8.74E-01 | 201 | 6646 | -0.03 | 0.11 | 8.08E-01 | 9.55E-01 | 231 | -0.04 | 0.08 | 6.18E-01 | 0.00 | 8.80E-01 | - | - |
| Vitamin B12 intake (ug/day) | Nutrients | 12863 | 0.03 | 0.08 | 7.35E-01 | 9.46E-01 | 212 | 12614 | 0.03 | 0.08 | 7.23E-01 | 9.14E-01 | 213 | 0.03 | 0.06 | 6.24E-01 | 0.00 | 9.86E-01 | - | - |
| Medioresinol intake (ug/day) | Nutrients | 12863 | -0.02 | 0.08 | 7.90E-01 | 9.55E-01 | 225 | 12614 | 0.08 | 0.08 | 3.37E-01 | 7.13E-01 | 126 | 0.03 | 0.06 | 6.30E-01 | 0.00 | 3.83E-01 | - | - |
| Cohabitation: Live alone vs Only one adult (spouse, partner) | Social | 4528 | 0.19 | 0.32 | 5.48E-01 | 8.48E-01 | 172 | 4616 | 0.02 | 0.33 | 9.44E-01 | 9.88E-01 | 261 | 0.11 | 0.23 | 6.33E-01 | 0.00 | 7.12E-01 | - | - |
| Participation in study circles | Social | 773 | 0.15 | 0.87 | 8.63E-01 | 9.66E-01 | 244 | 687 | -0.79 | 0.92 | 3.90E-01 | 7.30E-01 | 145 | -0.30 | 0.63 | 6.40E-01 | 0.00 | 4.59E-01 | - | - |
| Salad dressing with oil | Food | 12863 | -0.05 | 0.08 | 5.10E-01 | 8.48E-01 | 164 | 12614 | 0.00 | 0.08 | 9.95E-01 | 9.96E-01 | 272 | -0.03 | 0.06 | 6.40E-01 | 0.00 | 6.43E-01 | - | - |
| Arachidonic acid (ARA) intake (g/day) | Nutrients | 12863 | 0.04 | 0.08 | 6.18E-01 | 8.72E-01 | 193 | 12614 | 0.01 | 0.08 | 8.77E-01 | 9.67E-01 | 247 | 0.03 | 0.06 | 6.42E-01 | 0.00 | 8.13E-01 | - | - |
| Salty fish | Food | 12863 | -0.02 | 0.08 | 8.29E-01 | 9.61E-01 | 233 | 12614 | -0.04 | 0.08 | 6.59E-01 | 8.63E-01 | 208 | -0.03 | 0.06 | 6.44E-01 | 0.00 | 8.68E-01 | - | - |
| Boiled coffee | Beverage | 12863 | 0.05 | 0.08 | 5.53E-01 | 8.48E-01 | 173 | 12614 | 0.00 | 0.08 | 9.57E-01 | 9.92E-01 | 263 | 0.03 | 0.06 | 6.44E-01 | 0.00 | 7.07E-01 | - | - |
| Butter on bread | Food | 12863 | -0.02 | 0.08 | 8.03E-01 | 9.55E-01 | 229 | 12614 | 0.08 | 0.08 | 3.61E-01 | 7.30E-01 | 134 | 0.03 | 0.06 | 6.47E-01 | 0.00 | 4.07E-01 | - | - |
| For how much of the time during the last four weeks have you felt tired? | General health | 191 | -0.14 | 0.66 | 8.36E-01 | 9.61E-01 | 236 | 161 | -0.30 | 0.67 | 6.59E-01 | 8.63E-01 | 207 | -0.22 | 0.47 | 6.47E-01 | 0.00 | 8.65E-01 | - | - |
| Frequency of cycling during leisure time | Physical activity | 12301 | 0.03 | 0.08 | 7.61E-01 | 9.48E-01 | 218 | 12099 | 0.03 | 0.09 | 7.32E-01 | 9.14E-01 | 218 | 0.03 | 0.06 | 6.47E-01 | 0.00 | 9.73E-01 | - | - |
| Sum of all lignans intake (ug/day) | Nutrients | 12863 | -0.07 | 0.08 | 3.74E-01 | 7.70E-01 | 132 | 12614 | 0.02 | 0.08 | 7.97E-01 | 9.50E-01 | 229 | -0.03 | 0.06 | 6.49E-01 | 0.00 | 4.21E-01 | - | - |
| Bacon | Food | 12863 | 0.05 | 0.08 | 5.70E-01 | 8.48E-01 | 183 | 12614 | 0.01 | 0.08 | 9.52E-01 | 9.92E-01 | 262 | 0.03 | 0.06 | 6.54E-01 | 0.00 | 7.24E-01 | - | - |
| Hearing status | General health | 6845 | 0.10 | 0.11 | 3.55E-01 | 7.70E-01 | 125 | 6720 | -0.03 | 0.11 | 7.64E-01 | 9.23E-01 | 225 | 0.04 | 0.08 | 6.55E-01 | 0.00 | 3.88E-01 | - | - |
| Self rate of overall health | General health | 191 | 0.47 | 0.63 | 4.54E-01 | 8.32E-01 | 147 | 161 | -0.12 | 0.70 | 8.62E-01 | 9.67E-01 | 241 | 0.21 | 0.47 | 6.59E-01 | 0.00 | 5.28E-01 | - | - |
| Cohabitation: Live alone vs Adult and children | Social | 10154 | 0.25 | 0.28 | 3.81E-01 | 7.70E-01 | 135 | 9876 | -0.08 | 0.28 | 7.71E-01 | 9.23E-01 | 228 | 0.08 | 0.20 | 6.74E-01 | 0.00 | 4.11E-01 | - | - |
| Last time a colleague visited you at home | Psychosocial | 13830 | 0.11 | 0.08 | 1.64E-01 | 6.05E-01 | 74 | 13562 | -0.07 | 0.08 | 4.08E-01 | 7.32E-01 | 152 | 0.02 | 0.06 | 6.78E-01 | 0.59 | 1.18E-01 | - | - |
| For how much of the time during the last four weeks have you felt so depressed that nothing could cheer you up? | General health | 192 | -0.16 | 0.64 | 7.99E-01 | 9.55E-01 | 228 | 161 | -0.22 | 0.67 | 7.43E-01 | 9.18E-01 | 221 | -0.19 | 0.46 | 6.81E-01 | 0.00 | 9.50E-01 | - | - |
| Fitness status | Physical activity | 6836 | -0.18 | 0.11 | 1.08E-01 | 4.99E-01 | 59 | 6706 | 0.12 | 0.11 | 3.00E-01 | 6.95E-01 | 118 | -0.03 | 0.08 | 6.81E-01 | 0.71 | 6.17E-02 | - | - |
| Sweets | Food | 12863 | 0.02 | 0.08 | 8.30E-01 | 9.61E-01 | 235 | 12614 | -0.07 | 0.09 | 4.26E-01 | 7.40E-01 | 157 | -0.02 | 0.06 | 6.86E-01 | 0.00 | 4.73E-01 | - | - |
| Physical limitation to participate in moderately demanding activities: bending down or kneeling | General health | 189 | -0.09 | 0.65 | 8.94E-01 | 9.84E-01 | 248 | 162 | 0.48 | 0.67 | 4.75E-01 | 7.53E-01 | 171 | 0.19 | 0.47 | 6.87E-01 | 0.00 | 5.44E-01 | - | - |
| For how much of the time during the last four weeks have you felt very nervous? | General health | 192 | 0.37 | 0.63 | 5.61E-01 | 8.48E-01 | 179 | 162 | -0.03 | 0.67 | 9.64E-01 | 9.92E-01 | 265 | 0.18 | 0.46 | 6.96E-01 | 0.00 | 6.64E-01 | - | - |
| Light and physically active work | Physical activity | 14214 | 0.01 | 0.18 | 9.53E-01 | 9.93E-01 | 261 | 13916 | 0.09 | 0.18 | 6.21E-01 | 8.52E-01 | 199 | 0.05 | 0.13 | 6.98E-01 | 0.00 | 7.55E-01 | - | - |
| Number of cigarretes smoked per day (in groups) | Tobacco use | 11778 | 0.06 | 0.09 | 4.81E-01 | 8.32E-01 | 157 | 11530 | -0.01 | 0.09 | 8.66E-01 | 9.67E-01 | 244 | 0.02 | 0.06 | 7.00E-01 | 0.00 | 5.39E-01 | - | - |
| Sucrose intake (g/day) | Nutrients | 12863 | 0.09 | 0.08 | 2.87E-01 | 7.04E-01 | 110 | 12614 | -0.05 | 0.08 | 5.84E-01 | 8.34E-01 | 191 | 0.02 | 0.06 | 7.05E-01 | 0.22 | 2.56E-01 | - | - |
| Formic acid intaje (g/day) | Nutrients | 12863 | -0.03 | 0.08 | 7.53E-01 | 9.48E-01 | 216 | 12614 | 0.07 | 0.09 | 3.90E-01 | 7.30E-01 | 144 | 0.02 | 0.06 | 7.07E-01 | 0.00 | 4.04E-01 | - | - |
| Boiled or baked potato | Food | 12863 | -0.01 | 0.08 | 8.66E-01 | 9.66E-01 | 245 | 12614 | -0.03 | 0.08 | 7.30E-01 | 9.14E-01 | 216 | -0.02 | 0.06 | 7.18E-01 | 0.00 | 8.98E-01 | - | - |
| Palmitic acid intake (g/day) | Nutrients | 12863 | 0.00 | 0.08 | 9.86E-01 | 9.98E-01 | 268 | 12614 | 0.04 | 0.09 | 6.09E-01 | 8.49E-01 | 195 | 0.02 | 0.06 | 7.31E-01 | 0.00 | 7.05E-01 | - | - |
| Frequency of dancing during leisure time | Physical activity | 12641 | 0.04 | 0.08 | 6.08E-01 | 8.66E-01 | 191 | 12402 | -0.09 | 0.08 | 3.10E-01 | 7.02E-01 | 120 | -0.02 | 0.06 | 7.35E-01 | 0.15 | 2.78E-01 | - | - |
| Cream, creme fraiche, sour cream | Food | 12863 | -0.02 | 0.08 | 7.91E-01 | 9.55E-01 | 226 | 12614 | 0.06 | 0.08 | 4.51E-01 | 7.50E-01 | 162 | 0.02 | 0.06 | 7.37E-01 | 0.00 | 4.68E-01 | - | - |
| Brown beans, pea soup | Food | 12863 | 0.09 | 0.08 | 2.60E-01 | 6.89E-01 | 103 | 12614 | -0.06 | 0.08 | 4.95E-01 | 7.63E-01 | 176 | 0.02 | 0.06 | 7.43E-01 | 0.39 | 2.02E-01 | - | - |
| Excellent health | General health | 191 | 1.78 | 0.64 | 6.09E-03 | 1.05E-01 | 15 | 160 | -1.66 | 0.67 | 1.50E-02 | 1.63E-01 | 25 | 0.15 | 0.46 | 7.54E-01 | 0.93 | 2.20E-04 | - | - |
| Minced meat dishes | Food | 12863 | 0.10 | 0.08 | 2.08E-01 | 6.87E-01 | 82 | 12614 | -0.07 | 0.08 | 3.89E-01 | 7.30E-01 | 142 | 0.02 | 0.06 | 7.61E-01 | 0.55 | 1.35E-01 | - | - |
| Vitamin B6 intake (mg/day) | Nutrients | 12863 | 0.08 | 0.09 | 3.50E-01 | 7.70E-01 | 123 | 12614 | -0.05 | 0.09 | 5.96E-01 | 8.43E-01 | 193 | 0.02 | 0.06 | 7.66E-01 | 0.06 | 3.01E-01 | - | - |
| Smoking status: Smokers vs non-smokers | Tobacco use | 9638 | 0.10 | 0.22 | 6.27E-01 | 8.72E-01 | 196 | 9391 | -0.20 | 0.22 | 3.70E-01 | 7.30E-01 | 136 | -0.04 | 0.15 | 7.79E-01 | 0.00 | 3.27E-01 | - | - |
| Coffee rolls/buns, rusk | Food | 12863 | 0.03 | 0.08 | 6.85E-01 | 8.99E-01 | 208 | 12614 | 0.00 | 0.08 | 9.84E-01 | 9.92E-01 | 271 | 0.02 | 0.06 | 7.83E-01 | 0.00 | 7.66E-01 | - | - |
| Extent to what your physical and emotional health disrupted your usual social life during the last four weeks | General health | 192 | -0.67 | 0.63 | 2.92E-01 | 7.04E-01 | 113 | 162 | 0.47 | 0.67 | 4.77E-01 | 7.53E-01 | 173 | -0.13 | 0.46 | 7.85E-01 | 0.35 | 2.13E-01 | - | - |
| Mixed frozen vegetables | Food | 6463 | 0.13 | 0.12 | 2.80E-01 | 7.01E-01 | 109 | 6332 | -0.18 | 0.12 | 1.36E-01 | 5.04E-01 | 73 | -0.02 | 0.09 | 7.91E-01 | 0.70 | 6.83E-02 | - | - |
| Linolenic acid intake (g/day) | Nutrients | 12863 | 0.02 | 0.08 | 8.49E-01 | 9.61E-01 | 240 | 12614 | -0.05 | 0.08 | 5.66E-01 | 8.26E-01 | 187 | -0.02 | 0.06 | 7.93E-01 | 0.00 | 5.86E-01 | - | - |
| Vision status | General health | 6839 | 0.07 | 0.11 | 5.29E-01 | 8.48E-01 | 169 | 6712 | -0.12 | 0.12 | 3.11E-01 | 7.02E-01 | 121 | -0.02 | 0.08 | 7.94E-01 | 0.26 | 2.45E-01 | - | - |
| As healthy as anyone | General health | 191 | 0.92 | 0.64 | 1.51E-01 | 5.92E-01 | 69 | 161 | -1.25 | 0.66 | 6.22E-02 | 3.46E-01 | 49 | -0.12 | 0.46 | 7.96E-01 | 0.82 | 1.86E-02 | - | - |
| Pancake, waffle, Swedish dumpling | Food | 12863 | 0.07 | 0.08 | 4.13E-01 | 7.98E-01 | 140 | 12614 | -0.04 | 0.09 | 6.28E-01 | 8.52E-01 | 200 | 0.01 | 0.06 | 8.06E-01 | 0.00 | 3.58E-01 | - | - |
| Iron intake (mg/day) | Nutrients | 12863 | 0.01 | 0.09 | 8.75E-01 | 9.71E-01 | 246 | 12614 | -0.04 | 0.09 | 6.18E-01 | 8.52E-01 | 198 | -0.01 | 0.06 | 8.14E-01 | 0.00 | 6.41E-01 | - | - |
| Sedentary or standing work | Physical activity | 14214 | -0.05 | 0.18 | 7.67E-01 | 9.48E-01 | 221 | 13916 | 0.11 | 0.18 | 5.24E-01 | 7.87E-01 | 181 | 0.03 | 0.13 | 8.16E-01 | 0.00 | 5.08E-01 | - | - |
| Low fat milk (0,5%) | Beverage | 12863 | -0.02 | 0.08 | 8.43E-01 | 9.61E-01 | 238 | 12614 | -0.01 | 0.08 | 8.97E-01 | 9.67E-01 | 253 | -0.01 | 0.06 | 8.17E-01 | 0.00 | 9.64E-01 | - | - |
| Frequency of gardening during leisure time | Physical activity | 13639 | 0.10 | 0.08 | 1.88E-01 | 6.35E-01 | 81 | 13393 | -0.08 | 0.08 | 3.07E-01 | 7.02E-01 | 119 | 0.01 | 0.06 | 8.19E-01 | 0.63 | 9.90E-02 | - | - |
| Campesterol intake (mg/day) | Nutrients | 12863 | 0.05 | 0.08 | 5.18E-01 | 8.48E-01 | 166 | 12614 | -0.03 | 0.08 | 7.36E-01 | 9.14E-01 | 220 | 0.01 | 0.06 | 8.20E-01 | 0.00 | 4.88E-01 | - | - |
| Sugar, honey, marmelade, jam | Food | 12863 | 0.04 | 0.08 | 6.01E-01 | 8.66E-01 | 189 | 12614 | -0.07 | 0.08 | 3.97E-01 | 7.30E-01 | 146 | -0.01 | 0.06 | 8.28E-01 | 0.00 | 3.32E-01 | - | - |
| Brewed (filtered) coffee | Beverage | 12863 | 0.16 | 0.08 | 5.54E-02 | 4.20E-01 | 36 | 12614 | -0.14 | 0.08 | 9.81E-02 | 4.30E-01 | 62 | 0.01 | 0.06 | 8.30E-01 | 0.84 | 1.17E-02 | - | - |
| Number of cigarretes smoked per day | Tobacco use | 9382 | 0.10 | 0.09 | 2.75E-01 | 6.96E-01 | 108 | 9127 | -0.08 | 0.10 | 3.98E-01 | 7.30E-01 | 148 | 0.01 | 0.07 | 8.37E-01 | 0.46 | 1.72E-01 | - | - |
| For how much of the time during the last four weeks have you felt calm and serene? | General health | 191 | -0.03 | 0.63 | 9.60E-01 | 9.93E-01 | 264 | 162 | -0.14 | 0.66 | 8.28E-01 | 9.59E-01 | 234 | -0.08 | 0.46 | 8.52E-01 | 0.00 | 9.01E-01 | - | - |
| Monounsaturated fat intake (g/day) | Nutrients | 12863 | 0.02 | 0.09 | 7.95E-01 | 9.55E-01 | 227 | 12614 | 0.00 | 0.09 | 9.80E-01 | 9.92E-01 | 267 | 0.01 | 0.06 | 8.66E-01 | 0.00 | 8.42E-01 | - | - |
| Syringaresinol intake (ug/day) | Nutrients | 12863 | -0.06 | 0.08 | 4.42E-01 | 8.27E-01 | 146 | 12614 | 0.05 | 0.08 | 5.78E-01 | 8.34E-01 | 189 | -0.01 | 0.06 | 8.74E-01 | 0.00 | 3.50E-01 | - | - |
| Enough time for job assignments | Psychosocial | 14376 | -0.02 | 0.08 | 8.07E-01 | 9.55E-01 | 230 | 14102 | 0.00 | 0.08 | 9.75E-01 | 9.92E-01 | 266 | -0.01 | 0.06 | 8.79E-01 | 0.00 | 8.47E-01 | - | - |
| Secoisolariciresinol intake (ug/day) | Nutrients | 12863 | 0.08 | 0.08 | 3.71E-01 | 7.70E-01 | 131 | 12614 | -0.06 | 0.09 | 4.70E-01 | 7.53E-01 | 169 | 0.01 | 0.06 | 8.94E-01 | 0.23 | 2.53E-01 | - | - |
| Pizza | Food | 12863 | 0.02 | 0.09 | 8.08E-01 | 9.55E-01 | 231 | 12614 | -0.04 | 0.09 | 6.62E-01 | 8.63E-01 | 209 | -0.01 | 0.06 | 8.97E-01 | 0.00 | 6.29E-01 | - | - |
| Long-term sickness | General health | 13220 | -0.01 | 0.24 | 9.68E-01 | 9.94E-01 | 266 | 12926 | 0.05 | 0.25 | 8.32E-01 | 9.59E-01 | 236 | 0.02 | 0.18 | 9.06E-01 | 0.00 | 8.57E-01 | - | - |
| Tiamin intake (mg/day) | Nutrients | 12863 | 0.10 | 0.09 | 2.42E-01 | 6.87E-01 | 96 | 12614 | -0.12 | 0.09 | 1.75E-01 | 5.89E-01 | 81 | -0.01 | 0.06 | 9.08E-01 | 0.69 | 7.36E-02 | - | - |
| Participation in sports or physical exercise associations | Social | 773 | -0.52 | 0.61 | 3.87E-01 | 7.71E-01 | 137 | 687 | 0.74 | 0.67 | 2.65E-01 | 6.73E-01 | 106 | 0.05 | 0.45 | 9.11E-01 | 0.50 | 1.59E-01 | - | - |
| Banana | Food | 12863 | 0.03 | 0.08 | 6.90E-01 | 9.02E-01 | 209 | 12614 | -0.02 | 0.09 | 8.02E-01 | 9.52E-01 | 230 | 0.01 | 0.06 | 9.12E-01 | 0.00 | 6.47E-01 | - | - |

| | | | | | | | | | | | | | | | | | | | | |
|---|---|---|---|---|---|---|---|---|---|---|---|---|---|---|---|---|---|---|---|---|
| Mashed potato | Food | 6463 | 0.02 | 0.12 | 8.52E-01 | 9.61E-01 | 242 | 6332 | -0.04 | 0.12 | 7.50E-01 | 9.22E-01 | 222 | -0.01 | 0.09 | 9.25E-01 | 0.00 | 7.21E-01 | - | - |
| Breakfast habits: Gruel w/o sandwich for breakfast vs not breakfast at all | Food | 947 | -0.16 | 0.78 | 8.40E-01 | 9.61E-01 | 237 | 900 | 0.08 | 0.85 | 9.23E-01 | 9.73E-01 | 259 | -0.05 | 0.58 | 9.33E-01 | 0.00 | 8.36E-01 | - | - |
| Grams of tobacco smoked per week | Tobacco use | 9038 | 0.06 | 0.09 | 5.55E-01 | 8.48E-01 | 174 | 8803 | -0.07 | 0.10 | 4.58E-01 | 7.50E-01 | 166 | -0.01 | 0.07 | 9.39E-01 | 0.00 | 3.45E-01 | - | - |
| For how much of the time during the last four weeks have you felt happy? | General health | 192 | -0.04 | 0.63 | 9.50E-01 | 9.93E-01 | 259 | 162 | -0.01 | 0.68 | 9.82E-01 | 9.92E-01 | 269 | -0.03 | 0.46 | 9.51E-01 | 0.00 | 9.79E-01 | - | - |
| Total protein intake (g/day) | Nutrients | 12863 | -0.02 | 0.08 | 7.78E-01 | 9.55E-01 | 222 | 12614 | 0.03 | 0.08 | 7.24E-01 | 9.14E-01 | 215 | 0.00 | 0.06 | 9.64E-01 | 0.00 | 6.53E-01 | - | - |
| Sausage, liver pate on bread | Food | 12863 | 0.00 | 0.08 | 9.64E-01 | 9.93E-01 | 265 | 12614 | -0.01 | 0.08 | 9.16E-01 | 9.70E-01 | 257 | 0.00 | 0.06 | 9.66E-01 | 0.00 | 9.15E-01 | - | - |
| Emotional problems that made you do less than you wanted during the last four weeks | General health | 190 | 1.00 | 1.96 | 6.09E-01 | 8.66E-01 | 192 | 161 | -1.02 | 1.88 | 5.90E-01 | 8.39E-01 | 192 | -0.05 | 1.36 | 9.72E-01 | 0.00 | 4.57E-01 | - | - |
| Bregott on bread | Food | 12863 | 0.05 | 0.08 | 5.21E-01 | 8.48E-01 | 167 | 12614 | -0.05 | 0.08 | 5.44E-01 | 8.03E-01 | 185 | 0.00 | 0.06 | 9.73E-01 | 0.00 | 3.78E-01 | - | - |
| Vitamin B3 intake (mg/day) | Nutrients | 12863 | 0.13 | 0.08 | 1.29E-01 | 5.41E-01 | 65 | 12614 | -0.13 | 0.09 | 1.28E-01 | 5.04E-01 | 68 | 0.00 | 0.06 | 9.88E-01 | 0.78 | 3.16E-02 | - | - |
| Cholesterol intake (g/day) | Nutrients | 12863 | 0.00 | 0.09 | 9.83E-01 | 9.98E-01 | 267 | 12614 | 0.00 | 0.09 | 9.96E-01 | 9.96E-01 | 273 | 0.00 | 0.06 | 9.91E-01 | 0.00 | 9.86E-01 | - | - |

**Supplementary Table 15**. Longitudinal association results for total cholesterol

| | | Training set | | | | | | Testing set | | | | | | Metanalysis | | | | | R2 | |
|---|---|---|---|---|---|---|---|---|---|---|---|---|---|---|---|---|---|---|---|---|
| **Description** | **Group** | **N** | **Effect estimate** | **S.E.** | **p-value** | **p-value$_{FDR}$** | **p-value rank** | **N** | **Effect estimate** | **S.E.** | **p-value** | **p-value$_{FDR}$** | **p-value rank** | **Effect estimate** | **S.E.** | **p-value** | **$I^2$** | **Q p-value** | **adjusted R2** | **R2 rank** |
| Grams of tobacco smoked per week | Tobacco use | 7642 | 0.11 | 0.02 | 2.33E-11 | **9.44E-10** | 4 | 7392 | 0.11 | 0.02 | 2.51E-11 | **2.03E-09** | 1 | 0.11 | 0.01 | 3.11E-21 | 0.00 | 9.85E-01 | 0.33 | 8 |
| Number of cigarretes smoked per day | Tobacco use | 7642 | 0.11 | 0.02 | 2.33E-11 | **9.44E-10** | 5 | 7392 | 0.11 | 0.02 | 2.51E-11 | **2.03E-09** | 2 | 0.11 | 0.01 | 3.11E-21 | 0.00 | 9.85E-01 | 0.33 | 6 |
| Number of cigars smoked per day | Tobacco use | 7642 | 0.11 | 0.02 | 2.33E-11 | **9.44E-10** | 6 | 7392 | 0.11 | 0.02 | 2.51E-11 | **2.03E-09** | 3 | 0.11 | 0.01 | 3.11E-21 | 0.00 | 9.85E-01 | 0.33 | 7 |
| Number of snuff boxes per week | Tobacco use | 11611 | 0.06 | 0.01 | 4.82E-15 | **1.15E-12** | 1 | 11436 | 0.04 | 0.01 | 5.15E-08 | **2.08E-06** | 6 | 0.05 | 0.01 | 4.83E-21 | 0.61 | 1.07E-01 | 0.32 | 12 |
| Snuff status: Snuff users vs non-snuff users | Tobacco use | 10354 | 0.17 | 0.02 | 9.46E-15 | **1.15E-12** | 2 | 10200 | 0.11 | 0.02 | 1.56E-06 | **4.73E-05** | 8 | 0.14 | 0.02 | 5.84E-19 | 0.76 | 4.13E-02 | 0.32 | 14 |
| Frequency of picking berries or mushrooms during leisure time | Physical activity | 11583 | -0.05 | 0.01 | 1.54E-08 | **5.33E-07** | 7 | 11406 | -0.05 | 0.01 | 7.46E-11 | **4.53E-09** | 4 | -0.05 | 0.01 | 7.21E-18 | 0.00 | 5.42E-01 | 0.33 | 9 |
| Years using snuff | Tobacco use | 11312 | 0.06 | 0.01 | 1.43E-13 | **1.16E-11** | 3 | 11099 | 0.03 | 0.01 | 6.35E-05 | **1.54E-03** | 10 | 0.05 | 0.01 | 6.70E-16 | 0.82 | 1.81E-02 | 0.32 | 13 |
| Teetotaler | Alcohol | 12255 | -0.11 | 0.03 | 1.28E-05 | **3.46E-04** | 9 | 12064 | -0.15 | 0.03 | 1.38E-08 | **6.69E-07** | 5 | -0.13 | 0.02 | 1.37E-12 | 0.09 | 2.95E-01 | 0.33 | 10 |
| Strong beer | Alcohol | 11340 | 0.03 | 0.01 | 1.62E-04 | **2.96E-03** | 12 | 11142 | 0.05 | 0.01 | 1.05E-07 | **3.64E-06** | 7 | 0.04 | 0.01 | 1.43E-10 | 0.30 | 2.31E-01 | 0.35 | 1 |
| Breakfast habits: Only coffee/tea for breakfast vs not breakfast at all | Food | 784 | 0.60 | 0.12 | 2.74E-07 | **8.32E-06** | 8 | 749 | 0.35 | 0.11 | 1.52E-03 | **1.95E-02** | 19 | 0.47 | 0.08 | 3.88E-09 | 0.57 | 1.26E-01 | 0.30 | 15 |
| Monounsaturated fat intake (g/day) | Nutrients | 11340 | 0.04 | 0.01 | 1.92E-05 | **4.66E-04** | 10 | 11142 | 0.03 | 0.01 | 1.28E-04 | **2.40E-03** | 13 | 0.04 | 0.01 | 9.90E-09 | 0.00 | 7.56E-01 | 0.35 | 2 |
| Chips, popcorn, salted nuts | Food | 11340 | 0.03 | 0.01 | 1.49E-04 | **2.96E-03** | 11 | 11142 | 0.03 | 0.01 | 1.64E-04 | **2.85E-03** | 14 | 0.03 | 0.01 | 8.84E-08 | 0.00 | 9.89E-01 | 0.35 | 3 |
| Alcohol intake (g/day) | Alcohol | 11340 | 0.02 | 0.01 | 5.75E-03 | **4.82E-02** | 29 | 11142 | 0.03 | 0.01 | 8.97E-05 | **1.98E-03** | 11 | 0.03 | 0.01 | 2.34E-06 | 0.00 | 4.07E-01 | 0.35 | 4 |
| Permanent employment | Psychosocial | 12032 | 0.05 | 0.02 | 4.96E-03 | **4.76E-02** | 25 | 11835 | 0.06 | 0.02 | 5.30E-04 | **8.06E-03** | 16 | 0.06 | 0.01 | 9.14E-06 | 0.00 | 6.36E-01 | 0.33 | 11 |
| Brewed (filtered) coffee | Beverage | 11340 | 0.02 | 0.01 | 1.34E-03 | **1.71E-02** | 19 | 11142 | 0.02 | 0.01 | 2.67E-03 | **2.83E-02** | 23 | 0.02 | 0.01 | 1.12E-05 | 0.00 | 8.70E-01 | 0.35 | 5 |
| Educational level | Psychosocial | 12266 | 0.02 | 0.01 | 3.82E-02 | 1.55E-01 | 59 | 12069 | 0.03 | 0.01 | 1.25E-04 | **2.40E-03** | 12 | 0.02 | 0.01 | 2.99E-05 | 0.37 | 2.06E-01 | - | - |
| Frequency of shoverling snow during leisure time | Physical activity | 11733 | -0.02 | 0.01 | 6.48E-03 | 5.25E-02 | 30 | 11572 | -0.03 | 0.01 | 1.49E-03 | **1.95E-02** | 18 | -0.02 | 0.01 | 3.01E-05 | 0.00 | 7.50E-01 | - | - |
| Sodium intake (mg/day) | Nutrients | 11340 | 0.01 | 0.01 | 9.22E-02 | 2.60E-01 | 86 | 11142 | 0.03 | 0.01 | 2.73E-05 | **7.37E-04** | 9 | 0.02 | 0.01 | 3.17E-05 | 0.68 | 7.67E-02 | - | - |
| Ice cream | Food | 11340 | -0.02 | 0.01 | 5.12E-03 | **4.76E-02** | 26 | 11142 | -0.02 | 0.01 | 5.32E-03 | 5.17E-02 | 25 | -0.02 | 0.01 | 7.85E-05 | 0.00 | 9.06E-01 | - | - |
| Feel uneasy or guilty because of your way of drinking | Alcohol | 11098 | 0.05 | 0.02 | 5.36E-02 | 1.86E-01 | 70 | 10975 | 0.09 | 0.03 | 3.73E-04 | **6.04E-03** | 15 | 0.07 | 0.02 | 1.05E-04 | 0.26 | 2.46E-01 | - | - |
| Feel the need to reduce alcohol consumption | Alcohol | 11101 | 0.05 | 0.03 | 3.83E-02 | 1.55E-01 | 60 | 10983 | 0.09 | 0.03 | 8.94E-04 | **1.28E-02** | 17 | 0.07 | 0.02 | 1.34E-04 | 0.00 | 3.88E-01 | - | - |
| Carbohydrates intake (g/day) | Nutrients | 11340 | -0.03 | 0.01 | 1.71E-04 | **2.96E-03** | 14 | 11142 | -0.01 | 0.01 | 1.13E-01 | 3.10E-01 | 89 | -0.02 | 0.01 | 1.59E-04 | 0.58 | 1.23E-01 | - | - |
| Arachidonic acid (ARA) intake (g/day) | Nutrients | 11340 | 0.03 | 0.01 | 1.32E-03 | **1.71E-02** | 18 | 11142 | 0.02 | 0.01 | 5.70E-02 | 2.04E-01 | 68 | 0.02 | 0.01 | 3.01E-04 | 0.00 | 3.50E-01 | - | - |
| Vitamin B3 intake (mg/day) | Nutrients | 11340 | 0.02 | 0.01 | 2.01E-02 | 1.12E-01 | 43 | 11142 | 0.02 | 0.01 | 6.41E-03 | 5.64E-02 | 27 | 0.02 | 0.01 | 3.56E-04 | 0.00 | 7.71E-01 | - | - |
| Coffee rolls/buns, rusk | Food | 11340 | -0.02 | 0.01 | 6.29E-02 | 2.12E-01 | 72 | 11142 | -0.02 | 0.01 | 2.27E-03 | **2.75E-02** | 20 | -0.02 | 0.01 | 5.01E-04 | 0.00 | 4.15E-01 | - | - |
| Secoisolariciresinol intake (ug/day) | Nutrients | 11340 | 0.02 | 0.01 | 1.00E-02 | 6.76E-02 | 36 | 11142 | 0.02 | 0.01 | 1.92E-02 | 1.17E-01 | 40 | 0.02 | 0.01 | 5.08E-04 | 0.00 | 8.61E-01 | - | - |
| Frequency of gardening during leisure time | Physical activity | 11641 | -0.02 | 0.01 | 4.15E-02 | 1.64E-01 | 61 | 11466 | -0.02 | 0.01 | 4.00E-03 | **4.05E-02** | 24 | -0.02 | 0.01 | 5.09E-04 | 0.00 | 5.46E-01 | - | - |
| Shellfish (e.g. shrimps, scallops) | Food | 5085 | 0.03 | 0.01 | 5.29E-03 | **4.76E-02** | 27 | 4989 | 0.02 | 0.01 | 4.02E-02 | 1.73E-01 | 55 | 0.03 | 0.01 | 6.03E-04 | 0.00 | 6.32E-01 | - | - |
| Cambridge physical activity index | Physical activity | 12073 | 0.02 | 0.01 | 3.78E-02 | 1.55E-01 | 58 | 11847 | 0.02 | 0.01 | 6.63E-03 | 5.64E-02 | 28 | 0.02 | 0.01 | 7.06E-04 | 0.00 | 6.44E-01 | - | - |
| Liquor, spirits | Alcohol | 11340 | 0.01 | 0.01 | 8.62E-02 | 2.51E-01 | 83 | 11142 | 0.02 | 0.01 | 2.49E-03 | **2.75E-02** | 22 | 0.02 | 0.01 | 7.89E-04 | 0.00 | 3.63E-01 | - | - |
| Overall state of health during the last year | General health | 12267 | -0.02 | 0.01 | 2.25E-03 | **2.60E-02** | 21 | 12063 | -0.01 | 0.01 | 1.19E-01 | 3.23E-01 | 90 | -0.02 | 0.01 | 1.10E-03 | 0.11 | 2.90E-01 | - | - |
| Iodine intake (ug/day) | Nutrients | 11340 | 0.02 | 0.01 | 2.33E-02 | 1.18E-01 | 48 | 11142 | 0.02 | 0.01 | 2.00E-02 | 1.19E-01 | 41 | 0.02 | 0.01 | 1.16E-03 | 0.00 | 9.65E-01 | - | - |
| Pasta | Food | 11340 | 0.03 | 0.01 | 1.70E-04 | **2.96E-03** | 13 | 11142 | 0.01 | 0.01 | 3.87E-01 | 5.87E-01 | 160 | 0.02 | 0.01 | 1.17E-03 | 0.77 | 3.70E-02 | - | - |
| Eat breakfast from 2000 | Food | 1192 | -0.22 | 0.08 | 5.67E-03 | **4.82E-02** | 28 | 1065 | -0.15 | 0.08 | 7.74E-02 | 2.51E-01 | 75 | -0.19 | 0.06 | 1.26E-03 | 0.00 | 5.27E-01 | - | - |
| Medium beer | Alcohol | 11340 | 0.02 | 0.01 | 2.12E-02 | 1.12E-01 | 46 | 11142 | 0.02 | 0.01 | 3.80E-02 | 1.73E-01 | 52 | 0.02 | 0.01 | 1.94E-03 | 0.00 | 9.05E-01 | - | - |
| Appetite status | Psychosocial | 6683 | -0.03 | 0.01 | 8.85E-03 | 6.27E-02 | 34 | 6575 | -0.02 | 0.01 | 1.01E-01 | 2.92E-01 | 84 | -0.02 | 0.01 | 2.62E-03 | 0.00 | 4.82E-01 | - | - |
| Exercise during the last three months | Physical activity | 12270 | 0.02 | 0.01 | 1.25E-02 | 7.67E-02 | 39 | 12074 | 0.01 | 0.01 | 8.17E-02 | 2.58E-01 | 77 | 0.02 | 0.01 | 2.70E-03 | 0.00 | 6.05E-01 | - | - |
| Vitamin C intake (mg/day) | Nutrients | 11340 | -0.02 | 0.01 | 2.72E-02 | 1.28E-01 | 51 | 11142 | -0.02 | 0.01 | 4.35E-02 | 1.76E-01 | 60 | -0.02 | 0.01 | 2.80E-03 | 0.00 | 8.97E-01 | - | - |
| Satisfaction with home and family situation | Psychosocial | 6690 | -0.02 | 0.01 | 5.22E-02 | 1.86E-01 | 68 | 6579 | -0.02 | 0.01 | 2.49E-02 | 1.32E-01 | 45 | -0.02 | 0.01 | 3.07E-03 | 0.00 | 8.37E-01 | - | - |
| Vitamin B12 intake (ug/day) | Nutrients | 11340 | 0.02 | 0.01 | 2.20E-02 | 1.14E-01 | 47 | 11142 | 0.02 | 0.01 | 6.06E-02 | 2.13E-01 | 69 | 0.02 | 0.01 | 3.20E-03 | 0.00 | 7.77E-01 | - | - |
| Cohabitation: Live alone vs Other/others | Social | 1260 | -0.12 | 0.07 | 1.11E-01 | 2.98E-01 | 91 | 1225 | -0.19 | 0.08 | 1.20E-02 | 8.75E-02 | 32 | -0.15 | 0.05 | 3.88E-03 | 0.00 | 4.66E-01 | - | - |
| Sucrose intake (g/day) | Nutrients | 11340 | -0.01 | 0.01 | 9.18E-02 | 2.60E-01 | 85 | 11142 | -0.02 | 0.01 | 1.77E-02 | 1.11E-01 | 39 | -0.02 | 0.01 | 4.13E-03 | 0.00 | 6.24E-01 | - | - |
| Tea | Beverage | 11340 | -0.03 | 0.01 | 4.25E-04 | **6.07E-03** | 17 | 11142 | 0.00 | 0.01 | 6.03E-01 | 7.68E-01 | 191 | -0.02 | 0.01 | 4.25E-03 | 0.78 | 3.34E-02 | - | - |
| Soft whey cheese | Food | 5085 | -0.04 | 0.01 | 2.12E-03 | **2.57E-02** | 20 | 4989 | -0.01 | 0.01 | 3.19E-01 | 5.42E-01 | 141 | -0.02 | 0.01 | 4.61E-03 | 0.59 | 1.20E-01 | - | - |
| Animal based protein intake (g/day) | Nutrients | 11340 | 0.02 | 0.01 | 2.50E-03 | **2.77E-02** | 22 | 11142 | 0.01 | 0.01 | 3.28E-01 | 5.42E-01 | 145 | 0.02 | 0.01 | 4.63E-03 | 0.52 | 1.49E-01 | - | - |
| Control over planning and execution of the workday | Psychosocial | 12143 | -0.02 | 0.01 | 4.31E-02 | 1.64E-01 | 64 | 11921 | -0.02 | 0.01 | 4.77E-02 | 1.87E-01 | 62 | -0.02 | 0.01 | 4.64E-03 | 0.00 | 9.82E-01 | - | - |
| Margarine for cooking | Food | 11340 | -0.02 | 0.01 | 4.64E-02 | 1.71E-01 | 65 | 11142 | -0.02 | 0.01 | 5.40E-02 | 2.01E-01 | 65 | -0.02 | 0.01 | 5.59E-03 | 0.00 | 9.58E-01 | - | - |
| Distance to work in kilometers (one way) | Physical activity | 10802 | -0.03 | 0.01 | 2.02E-04 | **3.28E-03** | 15 | 10692 | 0.00 | 0.01 | 9.03E-01 | 9.50E-01 | 231 | -0.02 | 0.01 | 6.51E-03 | 0.84 | 1.12E-02 | - | - |
| Boiled or baked potato | Food | 11340 | -0.02 | 0.01 | 2.79E-02 | 1.28E-01 | 53 | 11142 | -0.01 | 0.01 | 9.97E-02 | 2.92E-01 | 83 | -0.02 | 0.01 | 6.52E-03 | 0.00 | 7.01E-01 | - | - |
| White meat (poultry) | Food | 11340 | 0.01 | 0.01 | 7.88E-02 | 2.36E-01 | 81 | 11142 | 0.02 | 0.01 | 4.08E-02 | 1.73E-01 | 57 | 0.02 | 0.01 | 7.18E-03 | 0.00 | 8.24E-01 | - | - |
| Phosphate intake (mg/day) | Nutrients | 11340 | 0.01 | 0.01 | 1.33E-01 | 3.25E-01 | 99 | 11142 | 0.02 | 0.01 | 2.51E-02 | 1.32E-01 | 46 | 0.02 | 0.01 | 8.10E-03 | 0.00 | 6.09E-01 | - | - |
| Wine | Alcohol | 11340 | 0.01 | 0.01 | 2.09E-01 | 4.27E-01 | 119 | 11142 | 0.02 | 0.01 | 1.40E-02 | 8.95E-02 | 38 | 0.01 | 0.01 | 8.64E-03 | 0.00 | 3.95E-01 | - | - |
| Fried potatoes, pommes frites | Food | 11340 | 0.01 | 0.01 | 2.43E-01 | 4.68E-01 | 126 | 11142 | 0.02 | 0.01 | 1.23E-02 | 8.75E-02 | 34 | 0.01 | 0.01 | 9.25E-03 | 0.00 | 3.52E-01 | - | - |
| Parents or siblings had a cerebral hemorrhage/thrombosis or cardiac infarction before the age of 60 | General health | 12146 | 0.03 | 0.02 | 1.54E-01 | 3.47E-01 | 108 | 11964 | 0.04 | 0.02 | 2.49E-02 | 1.32E-01 | 44 | 0.04 | 0.01 | 9.49E-03 | 0.00 | 5.63E-01 | - | - |
| Parents or siblings have diabetes | General health | 12168 | -0.03 | 0.02 | 1.45E-01 | 3.36E-01 | 105 | 11984 | -0.04 | 0.02 | 4.02E-02 | 1.73E-01 | 56 | -0.04 | 0.01 | 1.30E-02 | 0.00 | 6.84E-01 | - | - |
| Magnesium intake (mg/day) | Nutrients | 11340 | 0.01 | 0.01 | 4.68E-01 | 6.50E-01 | 175 | 11142 | 0.02 | 0.01 | 5.95E-03 | 5.56E-02 | 26 | 0.01 | 0.01 | 1.37E-02 | 0.50 | 1.56E-01 | - | - |
| Hamburger | Food | 11340 | 0.01 | 0.01 | 9.68E-02 | 2.70E-01 | 87 | 11142 | 0.01 | 0.01 | 8.43E-02 | 2.63E-01 | 78 | 0.01 | 0.01 | 1.66E-02 | 0.00 | 9.58E-01 | - | - |
| Meat on bread | Food | 11340 | 0.01 | 0.01 | 8.67E-02 | 2.51E-01 | 84 | 11142 | 0.01 | 0.01 | 9.50E-02 | 2.89E-01 | 80 | 0.01 | 0.01 | 1.68E-02 | 0.00 | 9.81E-01 | - | - |
| Total energy intake (kcal/day) | Nutrients | 11340 | -0.01 | 0.01 | 2.45E-01 | 4.69E-01 | 127 | 11142 | -0.02 | 0.01 | 2.64E-02 | 1.36E-01 | 47 | -0.02 | 0.01 | 1.68E-02 | 0.00 | 4.53E-01 | - | - |
| Vitamin D intake (ug/day) | Nutrients | 11340 | 0.02 | 0.01 | 2.61E-02 | 1.27E-01 | 50 | 11142 | 0.01 | 0.01 | 2.60E-01 | 5.01E-01 | 126 | 0.01 | 0.01 | 1.79E-02 | 0.00 | 4.33E-01 | - | - |
| Sleep status | Sleep | 6697 | -0.01 | 0.01 | 1.54E-01 | 3.47E-01 | 107 | 6587 | -0.02 | 0.01 | 5.58E-02 | 2.02E-01 | 67 | -0.02 | 0.01 | 1.82E-02 | 0.00 | 7.32E-01 | - | - |
| Frequency of dancing during leisure time | Physical activity | 10918 | 0.02 | 0.01 | 2.59E-02 | 1.27E-01 | 49 | 10722 | 0.01 | 0.01 | 3.19E-01 | 5.42E-01 | 140 | 0.01 | 0.01 | 2.25E-02 | 0.00 | 3.85E-01 | - | - |
| Low fat margarine on bread | Food | 11340 | 0.00 | 0.01 | 5.90E-01 | 7.43E-01 | 193 | 11142 | 0.02 | 0.01 | 7.68E-03 | 6.22E-02 | 30 | 0.01 | 0.01 | 2.32E-02 | 0.56 | 1.34E-01 | - | - |
| Cohabitation: Live alone vs Only one adult (spouse, partner) | Social | 3666 | -0.02 | 0.03 | 4.66E-01 | 6.50E-01 | 173 | 3745 | -0.09 | 0.03 | 1.21E-02 | 8.75E-02 | 33 | -0.05 | 0.02 | 2.39E-02 | 0.42 | 1.89E-01 | - | - |
| Plant based protein intake (g/day) | Nutrients | 11340 | -0.02 | 0.01 | 8.07E-03 | 6.13E-02 | 32 | 11142 | 0.00 | 0.01 | 5.80E-01 | 7.64E-01 | 184 | -0.01 | 0.01 | 2.39E-02 | 0.55 | 1.36E-01 | - | - |
| Sodas, soft drinks, juice | Beverage | 11340 | -0.02 | 0.01 | 9.03E-03 | 6.27E-02 | 35 | 11142 | 0.00 | 0.01 | 5.87E-01 | 7.64E-01 | 186 | -0.01 | 0.01 | 2.46E-02 | 0.52 | 1.51E-01 | - | - |
| Tiamin intake (mg/day) | Nutrients | 11340 | 0.01 | 0.01 | 1.69E-01 | 3.62E-01 | 113 | 11142 | 0.01 | 0.01 | 7.35E-02 | 2.41E-01 | 74 | 0.01 | 0.01 | 2.50E-02 | 0.00 | 7.82E-01 | - | - |
| Butter for cooking | Food | 11340 | 0.02 | 0.01 | 8.51E-03 | 6.27E-02 | 33 | 11142 | 0.00 | 0.01 | 6.02E-01 | 7.68E-01 | 190 | 0.01 | 0.01 | 2.64E-02 | 0.56 | 1.32E-01 | - | - |
| Bacon | Food | 11340 | 0.01 | 0.01 | 6.89E-02 | 2.20E-01 | 76 | 11142 | 0.01 | 0.01 | 1.89E-01 | 4.16E-01 | 110 | 0.01 | 0.01 | 2.68E-02 | 0.00 | 7.18E-01 | - | - |
| Participation in study circles | Social | 763 | -0.13 | 0.09 | 1.30E-01 | 3.25E-01 | 97 | 676 | -0.15 | 0.09 | 1.08E-01 | 3.02E-01 | 87 | -0.14 | 0.06 | 2.74E-02 | 0.00 | 8.91E-01 | - | - |
| If you exercise, change in exercise habits during the last year | Physical activity | 10745 | 0.01 | 0.01 | 4.31E-01 | 6.30E-01 | 166 | 10570 | 0.02 | 0.01 | 2.09E-02 | 1.21E-01 | 42 | 0.01 | 0.01 | 2.81E-02 | 0.12 | 2.87E-01 | - | - |
| Pinoresinol intake (ug/day) | Nutrients | 11340 | -0.02 | 0.01 | 3.21E-03 | **3.39E-02** | 23 | 11142 | 0.00 | 0.01 | 9.93E-01 | 9.97E-01 | 242 | -0.01 | 0.01 | 3.68E-02 | 0.77 | 3.75E-02 | - | - |
| Total protein intake (g/day) | Nutrients | 11340 | 0.02 | 0.01 | 3.51E-02 | 1.52E-01 | 56 | 11142 | 0.01 | 0.01 | 4.17E-01 | 6.10E-01 | 166 | 0.01 | 0.01 | 3.88E-02 | 0.00 | 3.61E-01 | - | - |
| Minced meat dishes | Food | 11340 | 0.03 | 0.01 | 3.60E-04 | **5.47E-03** | 16 | 11142 | 0.00 | 0.01 | 5.55E-01 | 7.42E-01 | 181 | 0.01 | 0.01 | 3.96E-02 | 0.89 | 2.93E-03 | - | - |
| Fibre intake (g/day) | Nutrients | 11340 | -0.02 | 0.01 | 1.26E-02 | 7.67E-02 | 40 | 11142 | 0.00 | 0.01 | 6.83E-01 | 8.18E-01 | 203 | -0.01 | 0.01 | 4.04E-02 | 0.54 | 1.39E-01 | - | - |
| Average portion size of potatoes/rice/pasta based on photographic illustration of four sizes (smallest to largest) | Food | 11340 | 0.01 | 0.01 | 5.46E-01 | 7.13E-01 | 185 | 11142 | 0.02 | 0.01 | 2.38E-02 | 1.32E-01 | 43 | 0.01 | 0.01 | 4.23E-02 | 0.26 | 2.45E-01 | - | - |
| Zinc intake (mg/day) | Nutrients | 11340 | -0.01 | 0.01 | 2.86E-01 | 5.11E-01 | 136 | 11142 | -0.02 | 0.01 | 7.23E-02 | 2.41E-01 | 73 | -0.01 | 0.01 | 4.27E-02 | 0.00 | 6.09E-01 | - | - |
| Sour milk, yoghurt (3% fat) | Food | 11340 | -0.01 | 0.01 | 4.22E-01 | 6.28E-01 | 163 | 11142 | -0.02 | 0.01 | 3.98E-02 | 1.73E-01 | 54 | -0.01 | 0.01 | 4.28E-02 | 0.00 | 3.81E-01 | - | - |
| Milk, sour milk (1,5%) | Beverage | 11340 | 0.01 | 0.01 | 1.82E-01 | 3.81E-01 | 116 | 11142 | 0.01 | 0.01 | 1.27E-01 | 3.39E-01 | 91 | 0.01 | 0.01 | 4.30E-02 | 0.00 | 8.97E-01 | - | - |
| Control over own work assignment | Psychosocial | 12134 | -0.01 | 0.01 | 1.35E-01 | 3.25E-01 | 100 | 11915 | -0.01 | 0.01 | 1.76E-01 | 4.05E-01 | 104 | -0.01 | 0.01 | 4.40E-02 | 0.00 | 9.23E-01 | - | - |
| Marital status: Single vs Divorced/separated | Social | 1986 | 0.06 | 0.04 | 1.38E-01 | 3.25E-01 | 102 | 1952 | 0.06 | 0.05 | 1.77E-01 | 4.05E-01 | 105 | 0.06 | 0.03 | 4.48E-02 | 0.00 | 9.50E-01 | - | - |
| Selenium intake (ug/day) | Nutrients | 11340 | 0.01 | 0.01 | 1.33E-01 | 3.25E-01 | 98 | 11142 | 0.01 | 0.01 | 1.85E-01 | 4.13E-01 | 109 | 0.01 | 0.01 | 4.56E-02 | 0.00 | 9.08E-01 | - | - |

| | | | | | | | | | | | | | | | | | | | | |
|---|---|---|---|---|---|---|---|---|---|---|---|---|---|---|---|---|---|---|---|---|
| Enterolactone intake (ug/day) | Nutrients | 11340 | -0.01 | 0.01 | 4.48E-01 | 6.44E-01 | 169 | 11142 | -0.02 | 0.01 | 4.19E-02 | 1.73E-01 | 59 | -0.01 | 0.01 | 4.78E-02 | 0.00 | 3.72E-01 | - | - |
| Work shifts/weekends | Psychosocial | 11896 | 0.01 | 0.02 | 4.97E-01 | 6.67E-01 | 181 | 11705 | 0.04 | 0.02 | 3.44E-02 | 1.67E-01 | 50 | 0.02 | 0.01 | 4.83E-02 | 0.04 | 3.09E-01 | - | - |
| Vitamin B6 intake (mg/day) | Nutrients | 11340 | 0.01 | 0.01 | 2.55E-01 | 4.81E-01 | 129 | 11142 | 0.01 | 0.01 | 1.02E-01 | 2.92E-01 | 85 | 0.01 | 0.01 | 4.99E-02 | 0.00 | 7.23E-01 | - | - |
| White (soft) bread, thin crisp bread | Food | 11340 | 0.00 | 0.01 | 9.10E-01 | 9.33E-01 | 237 | 11142 | 0.02 | 0.01 | 8.69E-03 | 6.81E-02 | 31 | 0.01 | 0.01 | 5.17E-02 | 0.68 | 7.75E-02 | - | - |
| Number of social contacts with the same interests as you | Social | 12225 | 0.00 | 0.01 | 7.83E-01 | 8.82E-01 | 215 | 12027 | -0.02 | 0.01 | 1.32E-02 | 8.75E-02 | 36 | -0.01 | 0.01 | 5.21E-02 | 0.59 | 1.18E-01 | - | - |
| Cholesterol intake (g/day) | Nutrients | 11340 | 0.02 | 0.01 | 4.19E-03 | **4.25E-02** | 24 | 11142 | 0.00 | 0.01 | 8.90E-01 | 9.41E-01 | 230 | 0.01 | 0.01 | 5.38E-02 | 0.78 | 3.38E-02 | - | - |
| Cream, creme fraiche, sour cream | Food | 11340 | 0.01 | 0.01 | 1.75E-01 | 3.71E-01 | 114 | 11142 | 0.01 | 0.01 | 1.74E-01 | 4.05E-01 | 103 | 0.01 | 0.01 | 5.50E-02 | 0.00 | 9.56E-01 | - | - |
| Eicosapentaenoic acid (EPA) intake (g/day) | Nutrients | 11340 | 0.01 | 0.01 | 1.25E-01 | 3.20E-01 | 95 | 11142 | 0.01 | 0.01 | 2.57E-01 | 4.99E-01 | 125 | 0.01 | 0.01 | 5.96E-02 | 0.00 | 7.68E-01 | - | - |
| Patience status | Psychosocial | 6684 | -0.02 | 0.01 | 7.22E-02 | 2.25E-01 | 78 | 6576 | -0.01 | 0.01 | 3.91E-01 | 5.87E-01 | 162 | -0.01 | 0.01 | 6.08E-02 | 0.00 | 5.02E-01 | - | - |
| Docosahexaenoic acid (DHA) intake (g/day) | Nutrients | 11340 | 0.01 | 0.01 | 1.08E-01 | 2.94E-01 | 89 | 11142 | 0.01 | 0.01 | 3.20E-01 | 5.42E-01 | 142 | 0.01 | 0.01 | 6.61E-02 | 0.00 | 6.54E-01 | - | - |
| Salty fish | Food | 11340 | 0.00 | 0.01 | 6.17E-01 | 7.65E-01 | 196 | 11142 | 0.02 | 0.01 | 3.65E-02 | 1.73E-01 | 51 | 0.01 | 0.01 | 6.62E-02 | 0.20 | 2.63E-01 | - | - |
| Oil for cooking | Food | 11340 | 0.01 | 0.01 | 6.58E-02 | 2.16E-01 | 74 | 11142 | 0.01 | 0.01 | 4.59E-01 | 6.59E-01 | 169 | 0.01 | 0.01 | 6.80E-02 | 0.00 | 4.37E-01 | - | - |
| Sweets | Food | 11340 | -0.01 | 0.01 | 1.89E-01 | 3.92E-01 | 117 | 11142 | -0.01 | 0.01 | 2.09E-01 | 4.28E-01 | 117 | -0.01 | 0.01 | 6.90E-02 | 0.00 | 9.64E-01 | - | - |
| Monosaccharides intake (g/day) | Nutrients | 11340 | -0.02 | 0.01 | 7.77E-02 | 2.36E-01 | 80 | 11142 | -0.01 | 0.01 | 4.28E-01 | 6.19E-01 | 168 | -0.01 | 0.01 | 7.08E-02 | 0.00 | 4.89E-01 | - | - |
| Enough time for job assignments | Psychosocial | 12083 | -0.01 | 0.01 | 2.84E-01 | 5.11E-01 | 134 | 11859 | -0.01 | 0.01 | 1.38E-01 | 3.50E-01 | 95 | -0.01 | 0.01 | 7.09E-02 | 0.00 | 7.68E-01 | - | - |
| Breakfast habits: Coffee/tea and wheat buns or rusk for breakfast vs not breakfast at all | Food | 968 | -0.03 | 0.07 | 6.54E-01 | 7.98E-01 | 199 | 935 | -0.15 | 0.07 | 3.31E-02 | 1.64E-01 | 49 | -0.09 | 0.05 | 7.19E-02 | 0.34 | 2.18E-01 | - | - |
| Boiled coffee | Beverage | 11340 | 0.02 | 0.01 | 1.13E-02 | 7.45E-02 | 37 | 11142 | 0.00 | 0.01 | 9.98E-01 | 9.98E-01 | 243 | 0.01 | 0.01 | 7.61E-02 | 0.69 | 7.07E-02 | - | - |
| Confidence status | Psychosocial | 6680 | -0.01 | 0.01 | 1.48E-01 | 3.38E-01 | 106 | 6575 | -0.01 | 0.01 | 3.06E-01 | 5.42E-01 | 136 | -0.01 | 0.01 | 8.04E-02 | 0.00 | 7.65E-01 | - | - |
| Corn flakes | Food | 11340 | -0.01 | 0.01 | 3.42E-01 | 5.62E-01 | 146 | 11142 | -0.01 | 0.01 | 1.31E-01 | 3.45E-01 | 92 | -0.01 | 0.01 | 8.10E-02 | 0.00 | 7.02E-01 | - | - |
| Sausage as main dish | Food | 11340 | 0.01 | 0.01 | 1.64E-01 | 3.60E-01 | 109 | 11142 | 0.01 | 0.01 | 2.85E-01 | 5.29E-01 | 131 | 0.01 | 0.01 | 8.17E-02 | 0.00 | 8.25E-01 | - | - |
| Whole grain soft bread | Food | 11340 | -0.02 | 0.01 | 6.99E-03 | 5.48E-02 | 31 | 11142 | 0.00 | 0.01 | 8.01E-01 | 8.92E-01 | 218 | -0.01 | 0.01 | 8.37E-02 | 0.77 | 3.70E-02 | - | - |
| Brown beans, pea soup | Food | 11340 | -0.01 | 0.01 | 4.26E-01 | 6.28E-01 | 165 | 11142 | -0.01 | 0.01 | 1.09E-01 | 3.02E-01 | 88 | -0.01 | 0.01 | 9.01E-02 | 0.00 | 5.68E-01 | - | - |
| Berries (fresh or frozen) | Food | 11340 | 0.00 | 0.01 | 5.93E-01 | 7.43E-01 | 194 | 11142 | -0.01 | 0.01 | 6.47E-02 | 2.22E-01 | 71 | -0.01 | 0.01 | 9.11E-02 | 0.00 | 3.58E-01 | - | - |
| Banana | Food | 11340 | 0.01 | 0.01 | 1.44E-01 | 3.36E-01 | 104 | 11142 | 0.01 | 0.01 | 3.56E-01 | 5.65E-01 | 153 | 0.01 | 0.01 | 9.23E-02 | 0.00 | 6.96E-01 | - | - |
| Self-employed | Psychosocial | 12032 | -0.04 | 0.03 | 1.20E-01 | 3.13E-01 | 93 | 11835 | -0.02 | 0.03 | 4.20E-01 | 6.11E-01 | 167 | -0.03 | 0.02 | 9.40E-02 | 0.00 | 6.05E-01 | - | - |
| Linoleic acid intake (g/day) | Nutrients | 11340 | 0.01 | 0.01 | 2.11E-01 | 4.28E-01 | 120 | 11142 | 0.01 | 0.01 | 2.74E-01 | 5.16E-01 | 129 | 0.01 | 0.01 | 9.75E-02 | 0.00 | 9.12E-01 | - | - |
| Do you feel important and appreciated in your home? | Psychosocial | 6649 | -0.02 | 0.01 | 6.97E-02 | 2.20E-01 | 77 | 6526 | -0.01 | 0.01 | 5.91E-01 | 7.64E-01 | 187 | -0.01 | 0.01 | 9.89E-02 | 0.00 | 3.55E-01 | - | - |
| Satisfaction with accomodation | Psychosocial | 6694 | -0.01 | 0.01 | 1.98E-01 | 4.09E-01 | 118 | 6583 | -0.01 | 0.01 | 2.95E-01 | 5.38E-01 | 133 | -0.01 | 0.01 | 9.90E-02 | 0.00 | 8.62E-01 | - | - |
| Number of cigarretes smoked per day (in groups) | Tobacco use | 10334 | 0.00 | 0.01 | 7.91E-01 | 8.82E-01 | 218 | 10081 | -0.02 | 0.01 | 4.19E-02 | 1.73E-01 | 58 | -0.01 | 0.01 | 1.04E-01 | 0.36 | 2.10E-01 | - | - |
| Support from others | Social | 12249 | -0.01 | 0.01 | 3.03E-01 | 5.27E-01 | 140 | 12058 | -0.01 | 0.01 | 2.37E-01 | 4.72E-01 | 122 | -0.01 | 0.01 | 1.18E-01 | 0.00 | 9.13E-01 | - | - |
| Possibility to speak with colleagues during breaks | Psychosocial | 12115 | 0.01 | 0.01 | 1.25E-01 | 3.20E-01 | 94 | 11876 | 0.01 | 0.01 | 5.05E-01 | 6.94E-01 | 177 | 0.01 | 0.01 | 1.19E-01 | 0.00 | 5.42E-01 | - | - |
| Cohabitation: Live alone vs Adult and children | Social | 8843 | 0.02 | 0.03 | 3.82E-01 | 6.03E-01 | 154 | 8551 | -0.08 | 0.03 | 2.48E-03 | **2.75E-02** | 21 | -0.03 | 0.02 | 1.25E-01 | 0.87 | 5.94E-03 | - | - |
| High mental demand from job | Psychosocial | 12047 | 0.00 | 0.01 | 8.17E-01 | 8.98E-01 | 221 | 11826 | -0.01 | 0.01 | 5.46E-02 | 2.01E-01 | 66 | -0.01 | 0.01 | 1.28E-01 | 0.30 | 2.32E-01 | - | - |
| Mood status | Psychosocial | 6683 | -0.01 | 0.01 | 4.01E-01 | 6.11E-01 | 159 | 6574 | -0.01 | 0.01 | 1.91E-01 | 4.16E-01 | 111 | -0.01 | 0.01 | 1.29E-01 | 0.00 | 7.41E-01 | - | - |
| Satisfaction with leisure time | Psychosocial | 6689 | -0.01 | 0.01 | 2.17E-01 | 4.35E-01 | 121 | 6563 | -0.01 | 0.01 | 3.73E-01 | 5.78E-01 | 157 | -0.01 | 0.01 | 1.33E-01 | 0.00 | 8.08E-01 | - | - |
| Whole grain intake (g/day) | Food | 11340 | 0.00 | 0.01 | 7.88E-01 | 8.82E-01 | 217 | 11142 | 0.01 | 0.01 | 6.79E-02 | 2.29E-01 | 72 | 0.01 | 0.01 | 1.37E-01 | 0.16 | 2.74E-01 | - | - |
| Ingenuity or creativity demand from job | Psychosocial | 12078 | -0.01 | 0.01 | 4.26E-01 | 6.28E-01 | 164 | 11875 | -0.01 | 0.01 | 1.99E-01 | 4.28E-01 | 113 | -0.01 | 0.01 | 1.42E-01 | 0.00 | 7.26E-01 | - | - |
| Close relationship with anyone | Social | 12253 | 0.01 | 0.01 | 4.74E-01 | 6.55E-01 | 176 | 12060 | -0.02 | 0.01 | 6.73E-03 | 5.64E-02 | 29 | -0.01 | 0.01 | 1.58E-01 | 0.83 | 1.55E-02 | - | - |
| Fitness status | Physical activity | 6680 | 0.01 | 0.01 | 2.39E-01 | 4.68E-01 | 124 | 6572 | 0.01 | 0.01 | 4.13E-01 | 6.08E-01 | 165 | 0.01 | 0.01 | 1.58E-01 | 0.00 | 7.98E-01 | - | - |
| Vitamin B2 intake (ug/day) | Nutrients | 11340 | 0.01 | 0.01 | 2.98E-01 | 5.25E-01 | 138 | 11142 | 0.01 | 0.01 | 3.40E-01 | 5.51E-01 | 150 | 0.01 | 0.01 | 1.58E-01 | 0.00 | 9.48E-01 | - | - |
| Smoking status: Former smokers vs non-smokers | Tobacco use | 8444 | -0.01 | 0.02 | 7.54E-01 | 8.64E-01 | 212 | 8324 | -0.03 | 0.02 | 9.65E-02 | 2.89E-01 | 81 | -0.02 | 0.01 | 1.62E-01 | 0.00 | 3.42E-01 | - | - |
| Vision status | General health | 6682 | -0.01 | 0.01 | 2.97E-01 | 5.25E-01 | 137 | 6579 | -0.01 | 0.01 | 3.49E-01 | 5.58E-01 | 152 | -0.01 | 0.01 | 1.62E-01 | 0.00 | 9.45E-01 | - | - |
| Repetitive job | Psychosocial | 12125 | -0.01 | 0.01 | 1.07E-01 | 2.94E-01 | 88 | 11912 | 0.00 | 0.01 | 7.24E-01 | 8.46E-01 | 208 | -0.01 | 0.01 | 1.64E-01 | 0.00 | 3.76E-01 | - | - |
| Linolenic acid intake (g/day) | Nutrients | 11340 | 0.01 | 0.01 | 3.26E-01 | 5.46E-01 | 145 | 11142 | 0.01 | 0.01 | 3.30E-01 | 5.42E-01 | 148 | 0.01 | 0.01 | 1.67E-01 | 0.00 | 9.95E-01 | - | - |
| Palmitic acid intake (g/day) | Nutrients | 11340 | 0.02 | 0.01 | 3.26E-02 | 1.44E-01 | 55 | 11142 | 0.00 | 0.01 | 8.54E-01 | 9.19E-01 | 225 | 0.01 | 0.01 | 1.68E-01 | 0.63 | 1.00E-01 | - | - |
| Memory status | Psychosocial | 6669 | -0.01 | 0.01 | 3.55E-01 | 5.68E-01 | 152 | 6569 | -0.01 | 0.01 | 3.10E-01 | 5.42E-01 | 138 | -0.01 | 0.01 | 1.70E-01 | 0.00 | 9.49E-01 | - | - |
| High physical demand from job | Physical activity | 12158 | 0.02 | 0.01 | 3.25E-02 | 1.44E-01 | 54 | 11938 | 0.00 | 0.01 | 8.21E-01 | 9.03E-01 | 221 | 0.01 | 0.01 | 1.74E-01 | 0.64 | 9.58E-02 | - | - |
| Average portion size of vegetables based on photographic illustration of four sizes (smallest to largest) | Food | 11340 | -0.02 | 0.01 | 6.81E-02 | 2.20E-01 | 75 | 11142 | 0.00 | 0.01 | 9.46E-01 | 9.70E-01 | 237 | -0.01 | 0.01 | 1.81E-01 | 0.35 | 2.15E-01 | - | - |
| Energy status | Psychosocial | 6681 | -0.01 | 0.01 | 3.92E-01 | 6.10E-01 | 155 | 6565 | -0.01 | 0.01 | 3.02E-01 | 5.42E-01 | 134 | -0.01 | 0.01 | 1.82E-01 | 0.00 | 9.03E-01 | - | - |
| Sour milk, yoghurt (low fat) | Food | 11340 | 0.01 | 0.01 | 3.54E-01 | 5.68E-01 | 150 | 11142 | 0.01 | 0.01 | 3.58E-01 | 5.65E-01 | 154 | 0.01 | 0.01 | 1.92E-01 | 0.00 | 9.93E-01 | - | - |
| Pizza | Food | 11340 | 0.00 | 0.01 | 9.60E-01 | 9.64E-01 | 242 | 11142 | 0.01 | 0.01 | 7.98E-02 | 2.55E-01 | 76 | 0.01 | 0.01 | 2.01E-01 | 0.30 | 2.31E-01 | - | - |
| Frequency of hunting or fishing during leisure time | Physical activity | 11414 | 0.00 | 0.01 | 6.29E-01 | 7.76E-01 | 197 | 11250 | -0.01 | 0.01 | 1.83E-01 | 4.13E-01 | 107 | -0.01 | 0.01 | 2.01E-01 | 0.00 | 5.44E-01 | - | - |
| Satisfaction with economy | Psychosocial | 6688 | -0.01 | 0.01 | 1.75E-01 | 3.71E-01 | 115 | 6572 | 0.00 | 0.01 | 6.58E-01 | 8.04E-01 | 199 | -0.01 | 0.01 | 2.04E-01 | 0.00 | 5.17E-01 | - | - |
| Average portion size of meat/fish based on photographic illustration of four sizes (smallest to largest) | Food | 11340 | 0.01 | 0.01 | 2.32E-01 | 4.58E-01 | 123 | 11142 | 0.01 | 0.01 | 5.55E-01 | 7.42E-01 | 182 | 0.01 | 0.01 | 2.07E-01 | 0.00 | 6.67E-01 | - | - |
| Vitamin A intake (mg/day) | Nutrients | 11340 | 0.01 | 0.01 | 2.82E-01 | 5.11E-01 | 133 | 11142 | 0.01 | 0.01 | 4.76E-01 | 6.67E-01 | 172 | 0.01 | 0.01 | 2.07E-01 | 0.00 | 7.88E-01 | - | - |
| Sometimes physically straining work | Physical activity | 12094 | -0.02 | 0.02 | 1.67E-01 | 3.62E-01 | 112 | 11860 | -0.01 | 0.02 | 7.20E-01 | 8.45E-01 | 207 | -0.01 | 0.01 | 2.17E-01 | 0.00 | 4.73E-01 | - | - |
| Everyday exercise satisfaction | Physical activity | 12237 | 0.01 | 0.01 | 3.95E-01 | 6.10E-01 | 156 | 12045 | 0.01 | 0.01 | 3.71E-01 | 5.78E-01 | 155 | 0.01 | 0.01 | 2.17E-01 | 0.00 | 9.70E-01 | - | - |
| Polyunsaturated fat intake (g/day) | Nutrients | 11340 | 0.01 | 0.01 | 2.18E-01 | 4.35E-01 | 122 | 11142 | 0.00 | 0.01 | 6.31E-01 | 7.92E-01 | 193 | 0.01 | 0.01 | 2.26E-01 | 0.00 | 5.97E-01 | - | - |
| Smoking status: Former occasional smokers vs non-smokers | Tobacco use | 7111 | 0.04 | 0.03 | 1.09E-01 | 2.94E-01 | 90 | 6899 | 0.00 | 0.03 | 9.11E-01 | 9.52E-01 | 232 | 0.02 | 0.02 | 2.26E-01 | 0.10 | 2.91E-01 | - | - |
| Disaccharides intake (g/day) | Nutrients | 11340 | -0.01 | 0.01 | 3.96E-01 | 6.10E-01 | 158 | 11142 | -0.01 | 0.01 | 3.95E-01 | 5.89E-01 | 163 | -0.01 | 0.01 | 2.30E-01 | 0.00 | 9.99E-01 | - | - |
| Sausage, liver pate on bread | Food | 11340 | -0.02 | 0.01 | 4.80E-02 | 1.74E-01 | 67 | 11142 | 0.00 | 0.01 | 7.86E-01 | 8.84E-01 | 216 | -0.01 | 0.01 | 2.38E-01 | 0.61 | 1.07E-01 | - | - |
| Matairesinol intake (ug/day) | Nutrients | 11340 | -0.02 | 0.01 | 1.18E-02 | 7.51E-02 | 38 | 11142 | 0.01 | 0.01 | 3.90E-01 | 5.87E-01 | 161 | -0.01 | 0.01 | 2.44E-01 | 0.83 | 1.67E-02 | - | - |
| Snuff status: Former snuff users vs non-snuff users | Tobacco use | 9464 | 0.05 | 0.03 | 6.55E-02 | 2.16E-01 | 73 | 9346 | -0.01 | 0.03 | 8.37E-01 | 9.12E-01 | 223 | 0.02 | 0.02 | 2.45E-01 | 0.52 | 1.49E-01 | - | - |
| Changed everyday exercise during the last year | Physical activity | 12275 | 0.00 | 0.01 | 5.47E-01 | 7.13E-01 | 186 | 12075 | 0.01 | 0.01 | 3.04E-01 | 5.42E-01 | 135 | 0.01 | 0.01 | 2.49E-01 | 0.00 | 7.60E-01 | - | - |
| Lariciresinol intake (ug/day) | Nutrients | 11340 | -0.02 | 0.01 | 4.28E-02 | 1.64E-01 | 62 | 11142 | 0.00 | 0.01 | 6.95E-01 | 8.27E-01 | 204 | -0.01 | 0.01 | 2.50E-01 | 0.66 | 8.69E-02 | - | - |
| Sum of Lariciresinol, Matairesinol, Pinoresinol, Secoisolariciresinol intake (ug/day) | Nutrients | 11340 | -0.02 | 0.01 | 3.66E-02 | 1.55E-01 | 57 | 11142 | 0.00 | 0.01 | 6.44E-01 | 7.97E-01 | 195 | -0.01 | 0.01 | 2.51E-01 | 0.69 | 7.08E-02 | - | - |
| Frequncy of social contacts with colleagues during leisure time | Psychosocial | 11848 | 0.00 | 0.01 | 8.60E-01 | 9.09E-01 | 230 | 11612 | -0.01 | 0.01 | 1.59E-01 | 3.86E-01 | 100 | -0.01 | 0.01 | 2.63E-01 | 0.00 | 3.83E-01 | - | - |
| Number of people with whom you can speak openly | Social | 12240 | 0.01 | 0.01 | 3.45E-01 | 5.62E-01 | 149 | 12050 | -0.02 | 0.01 | 1.33E-02 | 8.75E-02 | 37 | -0.01 | 0.01 | 2.81E-01 | 0.83 | 1.55E-02 | - | - |
| Contradictory demands in job | Psychosocial | 12045 | 0.00 | 0.01 | 8.13E-01 | 8.98E-01 | 220 | 11826 | 0.01 | 0.01 | 2.06E-01 | 4.28E-01 | 115 | 0.01 | 0.01 | 2.90E-01 | 0.00 | 4.66E-01 | - | - |
| Sum of all lignans intake (ug/day) | Nutrients | 11340 | -0.02 | 0.01 | 1.77E-02 | 1.05E-01 | 41 | 11142 | 0.01 | 0.01 | 3.85E-01 | 5.87E-01 | 159 | -0.01 | 0.01 | 2.91E-01 | 0.81 | 2.18E-02 | - | - |
| Fatty fish (e.g. herring, whitefish, salmon) | Food | 11340 | 0.02 | 0.01 | 5.28E-02 | 1.86E-01 | 69 | 11142 | 0.00 | 0.01 | 6.83E-01 | 8.18E-01 | 202 | 0.01 | 0.01 | 2.91E-01 | 0.64 | 9.41E-02 | - | - |
| People to ask for help apart from the ones at home | Social | 12225 | 0.05 | 0.03 | 4.65E-02 | 1.71E-01 | 66 | 12036 | -0.01 | 0.02 | 6.32E-01 | 7.92E-01 | 194 | 0.02 | 0.02 | 2.91E-01 | 0.68 | 7.94E-02 | - | - |
| Informed of having high blood pressure | General health | 12279 | -0.02 | 0.02 | 3.16E-01 | 5.35E-01 | 142 | 12077 | -0.01 | 0.02 | 6.46E-01 | 7.97E-01 | 196 | -0.02 | 0.02 | 3.02E-01 | 0.00 | 6.98E-01 | - | - |
| Possibility to leave your work for a while to speak with a colleague | Psychosocial | 12061 | 0.00 | 0.01 | 8.45E-01 | 9.04E-01 | 227 | 11800 | -0.01 | 0.01 | 9.80E-02 | 2.90E-01 | 82 | -0.01 | 0.01 | 3.07E-01 | 0.42 | 1.88E-01 | - | - |
| Pancake, waffle, Swedish dumpling | Food | 11340 | 0.00 | 0.01 | 8.95E-01 | 9.27E-01 | 234 | 11142 | 0.01 | 0.01 | 1.92E-01 | 4.16E-01 | 112 | 0.01 | 0.01 | 3.09E-01 | 0.00 | 4.07E-01 | - | - |
| Frequency of walking during leisure time | Physical activity | 11927 | 0.01 | 0.01 | 4.12E-01 | 6.22E-01 | 161 | 11789 | -0.02 | 0.01 | 2.75E-02 | 1.39E-01 | 48 | -0.01 | 0.01 | 3.25E-01 | 0.78 | 3.26E-02 | - | - |
| Mixed frozen vegetables | Food | 5085 | -0.01 | 0.01 | 4.17E-01 | 6.26E-01 | 162 | 4989 | -0.01 | 0.01 | 5.68E-01 | 7.54E-01 | 183 | -0.01 | 0.01 | 3.29E-01 | 0.00 | 8.59E-01 | - | - |
| Salad dressing with oil | Food | 11340 | 0.00 | 0.01 | 7.22E-01 | 8.63E-01 | 203 | 11142 | 0.01 | 0.01 | 8.65E-02 | 2.66E-01 | 79 | 0.01 | 0.01 | 3.33E-01 | 0.53 | 1.44E-01 | - | - |
| White cabbage, lettuce, lettuce cabbage, spinach, borecole | Food | 11340 | 0.00 | 0.01 | 8.24E-01 | 9.01E-01 | 222 | 11142 | 0.01 | 0.01 | 2.64E-01 | 5.04E-01 | 127 | 0.01 | 0.01 | 3.43E-01 | 0.00 | 5.27E-01 | - | - |
| Medioresinol intake (ug/day) | Nutrients | 11340 | -0.02 | 0.01 | 2.07E-02 | 1.12E-01 | 45 | 11142 | 0.01 | 0.01 | 3.35E-01 | 5.46E-01 | 149 | -0.01 | 0.01 | 3.44E-01 | 0.81 | 2.02E-02 | - | - |
| Enterodiol intake (ug/day) | Nutrients | 11340 | 0.00 | 0.01 | 7.35E-01 | 8.63E-01 | 204 | 11142 | -0.01 | 0.01 | 3.22E-01 | 5.42E-01 | 143 | -0.01 | 0.01 | 3.47E-01 | 0.00 | 6.47E-01 | - | - |
| Syringaresinol intake (ug/day) | Nutrients | 11340 | -0.02 | 0.01 | 1.96E-02 | 1.12E-01 | 42 | 11142 | 0.01 | 0.01 | 3.12E-01 | 5.42E-01 | 139 | -0.01 | 0.01 | 3.53E-01 | 0.82 | 1.79E-02 | - | - |
| Folic acid intake (ug/day) | Nutrients | 11340 | 0.00 | 0.01 | 9.66E-01 | 9.66E-01 | 243 | 11142 | 0.01 | 0.01 | 2.08E-01 | 4.28E-01 | 116 | 0.01 | 0.01 | 3.55E-01 | 0.00 | 3.91E-01 | - | - |
| Job demands to work very fast | Psychosocial | 12126 | 0.01 | 0.01 | 4.55E-01 | 6.47E-01 | 170 | 11911 | 0.00 | 0.01 | 5.92E-01 | 7.64E-01 | 188 | 0.00 | 0.01 | 3.64E-01 | 0.00 | 8.84E-01 | - | - |
| Soft cheese | Food | 5085 | 0.00 | 0.01 | 8.96E-01 | 9.27E-01 | 235 | 4989 | 0.01 | 0.01 | 2.76E-01 | 5.16E-01 | 130 | 0.01 | 0.01 | 3.70E-01 | 0.00 | 5.28E-01 | - | - |
| Cookies, pastry | Food | 11340 | 0.00 | 0.01 | 9.28E-01 | 9.40E-01 | 240 | 11142 | -0.01 | 0.01 | 1.84E-01 | 4.13E-01 | 108 | 0.00 | 0.01 | 3.76E-01 | 0.00 | 3.19E-01 | - | - |

| | | | | | | | | | | | | | | | | | | | | |
|---|---|---|---|---|---|---|---|---|---|---|---|---|---|---|---|---|---|---|---|---|
| Satisfaction with work situation | Psychosocial | 6629 | 0.00 | 0.01 | 7.35E-01 | 8.63E-01 | 205 | 6510 | -0.01 | 0.01 | 3.72E-01 | 5.78E-01 | 156 | -0.01 | 0.01 | 3.84E-01 | 0.00 | 6.95E-01 | - | - |
| Tomato, cucumber | Food | 11340 | 0.01 | 0.01 | 5.01E-01 | 6.69E-01 | 182 | 11142 | 0.00 | 0.01 | 5.83E-01 | 7.64E-01 | 185 | 0.00 | 0.01 | 3.88E-01 | 0.00 | 9.32E-01 | - | - |
| Milk, sour milk (3%) | Beverage | 11340 | 0.00 | 0.01 | 9.26E-01 | 9.40E-01 | 239 | 11142 | -0.01 | 0.01 | 2.66E-01 | 5.04E-01 | 128 | 0.00 | 0.01 | 3.94E-01 | 0.00 | 4.71E-01 | - | - |
| Rice | Food | 11340 | 0.00 | 0.01 | 7.76E-01 | 8.81E-01 | 214 | 11142 | 0.01 | 0.01 | 1.37E-01 | 3.50E-01 | 94 | 0.00 | 0.01 | 3.94E-01 | 0.36 | 2.11E-01 | - | - |
| Potassium intake (mg/day) | Nutrients | 11340 | 0.00 | 0.01 | 7.41E-01 | 8.63E-01 | 207 | 11142 | 0.01 | 0.01 | 3.85E-01 | 5.87E-01 | 158 | 0.01 | 0.01 | 3.96E-01 | 0.00 | 7.04E-01 | - | - |
| Light beer | Alcohol | 11340 | 0.01 | 0.01 | 4.46E-01 | 6.44E-01 | 168 | 11142 | 0.00 | 0.01 | 6.65E-01 | 8.08E-01 | 200 | 0.00 | 0.01 | 4.01E-01 | 0.00 | 8.02E-01 | - | - |
| Frequency of engaging in clubs, associations or study circles | Social | 8186 | -0.01 | 0.01 | 4.59E-01 | 6.49E-01 | 172 | 8072 | 0.02 | 0.01 | 5.38E-02 | 2.01E-01 | 64 | 0.01 | 0.01 | 4.04E-01 | 0.72 | 5.88E-02 | - | - |
| Travel to work: Cycle to work vs passive travel to work | Physical activity | 9515 | 0.03 | 0.02 | 7.99E-02 | 2.37E-01 | 82 | 9414 | -0.01 | 0.02 | 5.54E-01 | 7.42E-01 | 180 | 0.01 | 0.01 | 4.08E-01 | 0.63 | 9.83E-02 | - | - |
| Participation in sports or physical exercise associations | Social | 763 | 0.07 | 0.06 | 2.57E-01 | 4.81E-01 | 130 | 676 | 0.00 | 0.07 | 9.62E-01 | 9.78E-01 | 239 | 0.04 | 0.04 | 4.13E-01 | 0.00 | 4.33E-01 | - | - |
| Do you feel important and appreciated outside your home? | Psychosocial | 6692 | -0.01 | 0.01 | 4.86E-01 | 6.67E-01 | 177 | 6584 | 0.00 | 0.01 | 6.46E-01 | 7.97E-01 | 197 | -0.01 | 0.01 | 4.14E-01 | 0.00 | 8.64E-01 | - | - |
| Bregott on bread | Food | 11340 | 0.00 | 0.01 | 7.03E-01 | 8.46E-01 | 201 | 11142 | -0.01 | 0.01 | 1.37E-01 | 3.50E-01 | 93 | 0.00 | 0.01 | 4.30E-01 | 0.42 | 1.88E-01 | - | - |
| Meat stew | Food | 11340 | 0.01 | 0.01 | 1.64E-01 | 3.60E-01 | 111 | 11142 | 0.00 | 0.01 | 7.60E-01 | 8.67E-01 | 212 | 0.00 | 0.01 | 4.42E-01 | 0.30 | 2.31E-01 | - | - |
| Frequent social contacts with colleagues during work | Psychosocial | 12033 | 0.01 | 0.01 | 7.37E-02 | 2.27E-01 | 79 | 11807 | -0.01 | 0.01 | 4.65E-01 | 6.61E-01 | 171 | 0.00 | 0.01 | 4.54E-01 | 0.68 | 7.48E-02 | - | - |
| Beta-sitostanol intake (mg/day) | Nutrients | 11340 | -0.02 | 0.01 | 2.04E-02 | 1.12E-01 | 44 | 11142 | 0.01 | 0.01 | 2.11E-01 | 4.28E-01 | 120 | 0.00 | 0.01 | 4.55E-01 | 0.84 | 1.15E-02 | - | - |
| Number of friends that can come to your home at any time and feel at home | Social | 12224 | 0.01 | 0.01 | 4.36E-01 | 6.34E-01 | 167 | 12029 | -0.01 | 0.01 | 6.46E-02 | 2.22E-01 | 70 | 0.00 | 0.01 | 4.55E-01 | 0.71 | 6.27E-02 | - | - |
| Travel to work: Walk to work vs passive travel to work | Physical activity | 7945 | 0.00 | 0.03 | 9.39E-01 | 9.47E-01 | 241 | 7784 | -0.03 | 0.03 | 2.53E-01 | 4.96E-01 | 124 | -0.02 | 0.02 | 4.62E-01 | 0.00 | 3.80E-01 | - | - |
| Breakfast habits: Porridge w/o sandwich for breakfast vs not breakfast at all | Food | 1678 | 0.02 | 0.05 | 7.44E-01 | 8.63E-01 | 209 | 1615 | -0.07 | 0.05 | 1.77E-01 | 4.05E-01 | 106 | -0.02 | 0.03 | 4.81E-01 | 0.30 | 2.31E-01 | - | - |
| Sedentary or standing work | Physical activity | 12094 | -0.02 | 0.02 | 2.86E-01 | 5.11E-01 | 135 | 11860 | 0.00 | 0.02 | 9.33E-01 | 9.65E-01 | 235 | -0.01 | 0.01 | 4.85E-01 | 0.00 | 4.17E-01 | - | - |
| Liver, kidney | Food | 5085 | 0.00 | 0.01 | 7.03E-01 | 8.46E-01 | 202 | 4989 | 0.01 | 0.01 | 5.46E-01 | 7.41E-01 | 179 | 0.01 | 0.01 | 4.87E-01 | 0.00 | 8.72E-01 | - | - |
| Participation in associations or voluntary organisations | Social | 12238 | 0.01 | 0.02 | 6.36E-01 | 7.81E-01 | 198 | 12034 | -0.02 | 0.02 | 1.46E-01 | 3.64E-01 | 97 | -0.01 | 0.01 | 4.92E-01 | 0.46 | 1.72E-01 | - | - |
| Oatflake, whole wheat, rye or barley porridge | Food | 11340 | -0.01 | 0.01 | 4.66E-01 | 6.50E-01 | 174 | 11142 | 0.00 | 0.01 | 8.19E-01 | 9.03E-01 | 220 | 0.00 | 0.01 | 4.99E-01 | 0.00 | 7.22E-01 | - | - |
| Number of social interactions during a normal week | Social | 12244 | 0.01 | 0.01 | 2.69E-01 | 4.94E-01 | 132 | 12062 | -0.02 | 0.01 | 3.93E-02 | 1.73E-01 | 53 | 0.00 | 0.01 | 5.02E-01 | 0.80 | 2.50E-02 | - | - |
| Steak, chop, e.g. | Food | 11340 | 0.01 | 0.01 | 1.27E-01 | 3.21E-01 | 96 | 11142 | -0.02 | 0.01 | 1.28E-02 | 8.75E-02 | 35 | 0.00 | 0.01 | 5.04E-01 | 0.88 | 4.48E-03 | - | - |
| Hearing status | General health | 6688 | -0.01 | 0.01 | 4.02E-01 | 6.11E-01 | 160 | 6587 | 0.00 | 0.01 | 9.45E-01 | 9.70E-01 | 236 | 0.00 | 0.01 | 5.22E-01 | 0.00 | 5.86E-01 | - | - |
| Years smoking | Tobacco use | 10855 | 0.00 | 0.01 | 7.43E-01 | 8.63E-01 | 208 | 10665 | -0.01 | 0.01 | 2.36E-01 | 4.72E-01 | 121 | 0.00 | 0.01 | 5.46E-01 | 0.13 | 2.84E-01 | - | - |
| Cohabitation: Live alone vs Only children | Social | 1676 | 0.07 | 0.05 | 1.38E-01 | 3.25E-01 | 103 | 1644 | -0.04 | 0.05 | 4.78E-01 | 6.67E-01 | 174 | 0.02 | 0.04 | 5.47E-01 | 0.57 | 1.26E-01 | - | - |
| Receive hugs to comfort and support you | Social | 12200 | 0.02 | 0.02 | 2.55E-01 | 4.81E-01 | 128 | 11997 | -0.01 | 0.02 | 7.60E-01 | 8.67E-01 | 213 | 0.01 | 0.01 | 5.52E-01 | 0.04 | 3.08E-01 | - | - |
| Vitamin E intake (mg/day) | Nutrients | 11340 | 0.00 | 0.01 | 8.58E-01 | 9.09E-01 | 229 | 11142 | 0.01 | 0.01 | 5.12E-01 | 6.99E-01 | 178 | 0.00 | 0.01 | 5.54E-01 | 0.00 | 7.38E-01 | - | - |
| Potato salad | Food | 5085 | -0.01 | 0.01 | 5.57E-01 | 7.13E-01 | 189 | 4989 | 0.02 | 0.01 | 1.56E-01 | 3.82E-01 | 99 | 0.01 | 0.01 | 5.56E-01 | 0.50 | 1.56E-01 | - | - |
| Light but partly physically active work | Physical activity | 12094 | 0.00 | 0.02 | 8.39E-01 | 9.02E-01 | 226 | 11860 | -0.02 | 0.02 | 3.06E-01 | 5.42E-01 | 137 | -0.01 | 0.01 | 5.63E-01 | 0.00 | 3.85E-01 | - | - |
| Rosehip, sweet syrup soup | Food | 11340 | 0.01 | 0.01 | 4.91E-01 | 6.67E-01 | 179 | 11142 | 0.00 | 0.01 | 9.13E-01 | 9.52E-01 | 233 | 0.00 | 0.01 | 5.71E-01 | 0.00 | 6.84E-01 | - | - |
| Fat intake (g/day) | Nutrients | 11340 | 0.01 | 0.01 | 1.37E-01 | 3.25E-01 | 101 | 11142 | -0.01 | 0.01 | 4.94E-01 | 6.82E-01 | 176 | 0.00 | 0.01 | 5.74E-01 | 0.58 | 1.25E-01 | - | - |
| Iron intake (mg/day) | Nutrients | 11340 | -0.01 | 0.01 | 3.17E-01 | 5.35E-01 | 144 | 11142 | 0.00 | 0.01 | 8.15E-01 | 9.03E-01 | 219 | 0.00 | 0.01 | 5.87E-01 | 0.00 | 3.83E-01 | - | - |
| Learn new things at job | Psychosocial | 12107 | 0.00 | 0.01 | 5.63E-01 | 7.17E-01 | 191 | 11900 | 0.00 | 0.01 | 8.54E-01 | 9.19E-01 | 226 | 0.00 | 0.01 | 5.90E-01 | 0.00 | 7.80E-01 | - | - |
| Butter on bread | Food | 11340 | 0.00 | 0.01 | 5.81E-01 | 7.36E-01 | 192 | 11142 | 0.00 | 0.01 | 8.52E-01 | 9.19E-01 | 224 | 0.00 | 0.01 | 5.99E-01 | 0.00 | 8.02E-01 | - | - |
| Smoked fish/meat | Food | 11340 | 0.00 | 0.01 | 6.81E-01 | 8.27E-01 | 200 | 11142 | 0.00 | 0.01 | 7.80E-01 | 8.81E-01 | 215 | 0.00 | 0.01 | 6.25E-01 | 0.00 | 9.24E-01 | - | - |
| Marital status: Single vs Married/partner | Social | 11398 | 0.01 | 0.03 | 8.08E-01 | 8.97E-01 | 219 | 11177 | -0.02 | 0.03 | 3.48E-01 | 5.58E-01 | 151 | -0.01 | 0.02 | 6.26E-01 | 0.00 | 4.02E-01 | - | - |
| Last time a colleague visited you at home | Psychosocial | 11999 | 0.01 | 0.01 | 3.55E-01 | 5.68E-01 | 151 | 11749 | -0.01 | 0.01 | 1.06E-01 | 2.99E-01 | 86 | 0.00 | 0.01 | 6.28E-01 | 0.69 | 7.21E-02 | - | - |
| Breakfast habits: Gruel w/o sandwich for breakfast vs not breakfast at all | Food | 933 | -0.02 | 0.08 | 7.85E-01 | 8.82E-01 | 216 | 873 | -0.04 | 0.09 | 6.72E-01 | 8.12E-01 | 201 | -0.03 | 0.06 | 6.28E-01 | 0.00 | 8.89E-01 | - | - |
| Trans fat intake (g/day) | Nutrients | 11340 | 0.00 | 0.01 | 7.46E-01 | 8.63E-01 | 210 | 11142 | 0.00 | 0.01 | 7.31E-01 | 8.50E-01 | 209 | 0.00 | 0.01 | 6.37E-01 | 0.00 | 9.94E-01 | - | - |
| Low fat milk (0,5%) | Beverage | 11340 | 0.00 | 0.01 | 7.36E-01 | 8.63E-01 | 206 | 11142 | 0.00 | 0.01 | 7.43E-01 | 8.59E-01 | 210 | 0.00 | 0.01 | 6.38E-01 | 0.00 | 9.93E-01 | - | - |
| Margarine on bread | Food | 11340 | 0.00 | 0.01 | 8.49E-01 | 9.04E-01 | 228 | 11142 | 0.00 | 0.01 | 6.53E-01 | 8.01E-01 | 198 | 0.00 | 0.01 | 6.50E-01 | 0.00 | 8.55E-01 | - | - |
| Campesterol intake (mg/day) | Nutrients | 11340 | -0.01 | 0.01 | 4.96E-01 | 6.67E-01 | 180 | 11142 | 0.00 | 0.01 | 9.67E-01 | 9.78E-01 | 240 | 0.00 | 0.01 | 6.51E-01 | 0.00 | 6.10E-01 | - | - |
| Fiber cereals | Food | 11340 | 0.00 | 0.01 | 5.55E-01 | 7.13E-01 | 188 | 11142 | 0.00 | 0.01 | 9.70E-01 | 9.78E-01 | 241 | 0.00 | 0.01 | 6.59E-01 | 0.00 | 6.95E-01 | - | - |
| Apple, pear, peach, orange, mandarin and grapefruit | Food | 11340 | 0.00 | 0.01 | 8.71E-01 | 9.12E-01 | 232 | 11142 | 0.01 | 0.01 | 4.85E-01 | 6.74E-01 | 175 | 0.00 | 0.01 | 7.04E-01 | 0.00 | 5.44E-01 | - | - |
| Campestanol intake (mg/day) | Nutrients | 11340 | -0.02 | 0.01 | 4.30E-02 | 1.64E-01 | 63 | 11142 | 0.01 | 0.01 | 1.38E-01 | 3.50E-01 | 96 | 0.00 | 0.01 | 7.06E-01 | 0.84 | 1.31E-02 | - | - |
| Formic acid intaje (g/day) | Nutrients | 11340 | 0.01 | 0.01 | 4.55E-01 | 6.47E-01 | 171 | 11142 | -0.01 | 0.01 | 2.09E-01 | 4.28E-01 | 118 | 0.00 | 0.01 | 7.16E-01 | 0.50 | 1.57E-01 | - | - |
| Would you say that the number of people that you meet in your everyday life is enough or would you like to meet more or fewer people? | Social | 12248 | 0.00 | 0.01 | 5.58E-01 | 7.13E-01 | 190 | 12037 | 0.00 | 0.01 | 9.25E-01 | 9.60E-01 | 234 | 0.00 | 0.01 | 7.27E-01 | 0.00 | 6.31E-01 | - | - |
| Smoking status: Smokers vs non-smokers | Tobacco use | 8124 | 0.04 | 0.02 | 5.58E-02 | 1.91E-01 | 71 | 7886 | -0.03 | 0.02 | 1.47E-01 | 3.64E-01 | 98 | 0.00 | 0.02 | 7.54E-01 | 0.82 | 1.73E-02 | - | - |
| Cheese 10-17% | Food | 11340 | 0.00 | 0.01 | 8.66E-01 | 9.11E-01 | 231 | 11142 | 0.00 | 0.01 | 7.97E-01 | 8.92E-01 | 217 | 0.00 | 0.01 | 7.63E-01 | 0.00 | 9.52E-01 | - | - |
| Equol intake (ug/day) | Nutrients | 11340 | 0.00 | 0.01 | 8.37E-01 | 9.02E-01 | 225 | 11142 | 0.00 | 0.01 | 8.35E-01 | 9.12E-01 | 222 | 0.00 | 0.01 | 7.70E-01 | 0.00 | 1.00E+00 | - | - |
| Marital status: Single vs Widow/widower | Social | 1277 | -0.12 | 0.12 | 3.10E-01 | 5.34E-01 | 141 | 1249 | 0.16 | 0.12 | 1.67E-01 | 4.03E-01 | 101 | 0.02 | 0.09 | 7.74E-01 | 0.65 | 9.10E-02 | - | - |
| Stigmasterol intake (mg/day) | Nutrients | 11340 | 0.00 | 0.01 | 7.49E-01 | 8.63E-01 | 211 | 11142 | 0.01 | 0.01 | 4.76E-01 | 6.67E-01 | 173 | 0.00 | 0.01 | 7.80E-01 | 0.00 | 4.66E-01 | - | - |
| Lean fish (e.g. perch, bass, cod) | Food | 11340 | -0.01 | 0.01 | 1.17E-01 | 3.10E-01 | 92 | 11142 | 0.02 | 0.01 | 4.96E-02 | 1.91E-01 | 63 | 0.00 | 0.01 | 7.94E-01 | 0.84 | 1.25E-02 | - | - |
| Participation in other association | Social | 763 | -0.04 | 0.06 | 5.07E-01 | 6.73E-01 | 183 | 676 | 0.03 | 0.07 | 6.99E-01 | 8.28E-01 | 205 | -0.01 | 0.04 | 8.10E-01 | 0.00 | 4.65E-01 | - | - |
| Beta-sitosterol intake (mg/day) | Nutrients | 11340 | -0.01 | 0.01 | 4.90E-01 | 6.67E-01 | 178 | 11142 | 0.01 | 0.01 | 3.23E-01 | 5.42E-01 | 144 | 0.00 | 0.01 | 8.31E-01 | 0.29 | 2.35E-01 | - | - |
| Blota (broth + bread) | Food | 5085 | 0.00 | 0.01 | 8.82E-01 | 9.20E-01 | 233 | 4989 | 0.00 | 0.01 | 8.87E-01 | 9.41E-01 | 229 | 0.00 | 0.01 | 8.38E-01 | 0.00 | 9.88E-01 | - | - |
| Root vegetables, carrot | Food | 11340 | 0.01 | 0.01 | 3.17E-01 | 5.35E-01 | 143 | 11142 | -0.01 | 0.01 | 2.04E-01 | 4.28E-01 | 114 | 0.00 | 0.01 | 8.39E-01 | 0.61 | 1.09E-01 | - | - |
| Cheese 28% | Food | 11340 | 0.00 | 0.01 | 5.50E-01 | 7.13E-01 | 187 | 11142 | 0.00 | 0.01 | 7.06E-01 | 8.33E-01 | 206 | 0.00 | 0.01 | 8.77E-01 | 0.00 | 4.91E-01 | - | - |
| Skill demand from job | Psychosocial | 12097 | 0.00 | 0.01 | 5.97E-01 | 7.44E-01 | 195 | 11897 | 0.00 | 0.01 | 7.57E-01 | 8.67E-01 | 211 | 0.00 | 0.01 | 8.78E-01 | 0.00 | 5.53E-01 | - | - |
| Overall state of health compared to other your age | General health | 12082 | -0.02 | 0.01 | 2.77E-02 | 1.28E-01 | 52 | 11912 | 0.02 | 0.01 | 4.71E-02 | 1.87E-01 | 61 | 0.00 | 0.01 | 8.78E-01 | 0.89 | 3.07E-03 | - | - |
| Light and physically active work | Physical activity | 12094 | 0.01 | 0.02 | 7.58E-01 | 8.64E-01 | 213 | 11860 | -0.01 | 0.02 | 6.12E-01 | 7.74E-01 | 192 | 0.00 | 0.01 | 8.89E-01 | 0.00 | 5.64E-01 | - | - |
| Beta-carotene intake (mg/day) | Nutrients | 11340 | 0.01 | 0.01 | 3.03E-01 | 5.27E-01 | 139 | 11142 | -0.01 | 0.01 | 2.44E-01 | 4.82E-01 | 123 | 0.00 | 0.01 | 9.14E-01 | 0.58 | 1.21E-01 | - | - |
| Sugar, honey, marmelade, jam | Food | 11340 | 0.01 | 0.01 | 3.96E-01 | 6.10E-01 | 157 | 11142 | -0.01 | 0.01 | 4.62E-01 | 6.60E-01 | 170 | 0.00 | 0.01 | 9.40E-01 | 0.20 | 2.63E-01 | - | - |
| Whole grain crisp bread | Food | 11340 | -0.01 | 0.01 | 5.32E-01 | 7.03E-01 | 184 | 11142 | 0.00 | 0.01 | 5.94E-01 | 7.64E-01 | 189 | 0.00 | 0.01 | 9.50E-01 | 0.00 | 4.13E-01 | - | - |
| Long-term sickness | General health | 11877 | 0.03 | 0.02 | 2.66E-01 | 4.94E-01 | 131 | 11623 | -0.03 | 0.02 | 2.90E-01 | 5.34E-01 | 132 | 0.00 | 0.02 | 9.51E-01 | 0.58 | 1.25E-01 | - | - |
| Saturated fat intake (g/day) | Nutrients | 11340 | 0.01 | 0.01 | 2.41E-01 | 4.68E-01 | 125 | 11142 | -0.01 | 0.01 | 2.10E-01 | 4.28E-01 | 119 | 0.00 | 0.01 | 9.51E-01 | 0.66 | 8.61E-02 | - | - |
| Frequency of cycling during leisure time | Physical activity | 10553 | 0.00 | 0.01 | 8.30E-01 | 9.01E-01 | 223 | 10339 | 0.00 | 0.01 | 7.65E-01 | 8.69E-01 | 214 | 0.00 | 0.01 | 9.52E-01 | 0.00 | 7.17E-01 | - | - |
| Travel to work: Irregular travel mode to work vs passive travel to work | Physical activity | 8068 | 0.01 | 0.03 | 8.31E-01 | 9.01E-01 | 224 | 7957 | 0.00 | 0.03 | 8.81E-01 | 9.41E-01 | 227 | 0.00 | 0.02 | 9.63E-01 | 0.00 | 7.98E-01 | - | - |
| Mashed potato | Food | 5085 | -0.02 | 0.01 | 1.64E-01 | 3.60E-01 | 110 | 4989 | 0.02 | 0.01 | 1.73E-01 | 4.05E-01 | 102 | 0.00 | 0.01 | 9.64E-01 | 0.74 | 5.16E-02 | - | - |
| Blood based food | Food | 5085 | 0.00 | 0.01 | 9.20E-01 | 9.39E-01 | 238 | 4989 | 0.00 | 0.01 | 9.59E-01 | 9.78E-01 | 238 | 0.00 | 0.01 | 9.70E-01 | 0.00 | 9.15E-01 | - | - |
| Sum of phytosterols intake (mg/day) | Nutrients | 11340 | -0.01 | 0.01 | 3.78E-01 | 6.01E-01 | 153 | 11142 | 0.01 | 0.01 | 4.01E-01 | 5.95E-01 | 164 | 0.00 | 0.01 | 9.79E-01 | 0.32 | 2.24E-01 | - | - |
| Calcium intake (mg/day) | Nutrients | 11340 | 0.00 | 0.01 | 9.08E-01 | 9.33E-01 | 236 | 11142 | 0.00 | 0.01 | 8.85E-01 | 9.41E-01 | 228 | 0.00 | 0.01 | 9.83E-01 | 0.00 | 8.54E-01 | - | - |
| Pentadecanoic acid intake (g/day) | Nutrients | 11340 | 0.01 | 0.01 | 3.43E-01 | 5.62E-01 | 148 | 11142 | -0.01 | 0.01 | 3.29E-01 | 5.42E-01 | 147 | 0.00 | 0.01 | 9.84E-01 | 0.46 | 1.74E-01 | - | - |
| Heptadecanoic acid intake (g/day) | Nutrients | 11340 | 0.01 | 0.01 | 3.43E-01 | 5.62E-01 | 147 | 11142 | -0.01 | 0.01 | 3.29E-01 | 5.42E-01 | 146 | 0.00 | 0.01 | 9.84E-01 | 0.46 | 1.74E-01 | - | - |

**Supplementary Table 16**. Longitudinal association results for triglycerides

| | | Training set | | | | | | Testing set | | | | | | Metanalysis | | | | | R2 | |
|---|---|---|---|---|---|---|---|---|---|---|---|---|---|---|---|---|---|---|---|---|
| **Description** | **Group** | **N** | **Effect estimate** | **S.E.** | **p-value** | **p-value$_{FDR}$** | **p-value rank** | **N** | **Effect estimate** | **S.E.** | **p-value** | **p-value$_{FDR}$** | **p-value rank** | **Effect estimate** | **S.E.** | **p-value** | **$I^2$** | **Q p-value** | **adjusted R2** | **R2 rank** |
| Smoking status: Smokers vs non-smokers | Tobacco use | 6586 | 0.13 | 0.02 | 4.72E-12 | **1.15E-09** | 1 | 6348 | 0.11 | 0.02 | 6.93E-09 | **1.68E-06** | 1 | 0.12 | 0.01 | 2.56E-19 | 0.00 | 3.51E-01 | 0.27 | 2 |
| Years smoking | Tobacco use | 8786 | 0.05 | 0.01 | 1.23E-11 | **1.49E-09** | 2 | 8607 | 0.03 | 0.01 | 2.51E-06 | **2.04E-04** | 3 | 0.04 | 0.01 | 5.98E-16 | 0.64 | 9.50E-02 | 0.25 | 10 |
| Number of cigarretes smoked per day (in groups) | Tobacco use | 8354 | 0.04 | 0.01 | 1.99E-08 | **1.61E-06** | 3 | 8095 | 0.03 | 0.01 | 9.81E-06 | **3.97E-04** | 6 | 0.04 | 0.01 | 1.60E-12 | 0.18 | 2.70E-01 | 0.26 | 5 |
| Number of snuff boxes per week | Tobacco use | 9370 | 0.04 | 0.01 | 2.87E-07 | **1.75E-05** | 4 | 9203 | 0.02 | 0.01 | 9.91E-04 | **1.85E-02** | 13 | 0.03 | 0.01 | 2.76E-09 | 0.47 | 1.71E-01 | 0.26 | 3 |
| Overall state of health during the last year | General health | 9921 | -0.03 | 0.01 | 1.05E-04 | **3.20E-03** | 8 | 9722 | -0.03 | 0.01 | 8.49E-06 | **3.97E-04** | 5 | -0.03 | 0.00 | 3.59E-09 | 0.00 | 8.04E-01 | 0.26 | 6 |
| Snuff status: Snuff users vs non-snuff users | Tobacco use | 8391 | 0.09 | 0.02 | 1.40E-05 | **6.79E-04** | 5 | 8209 | 0.06 | 0.02 | 2.66E-03 | **3.08E-02** | 21 | 0.07 | 0.01 | 2.28E-07 | 0.13 | 2.83E-01 | 0.26 | 4 |
| Feel the need to reduce alcohol consumption | Alcohol | 8949 | 0.05 | 0.02 | 3.27E-02 | 2.14E-01 | 37 | 8844 | 0.12 | 0.02 | 3.84E-07 | **4.67E-05** | 2 | 0.09 | 0.02 | 2.36E-07 | 0.73 | 5.56E-02 | - | - |
| Fiber cereals | Food | 9175 | -0.03 | 0.01 | 1.04E-04 | **3.20E-03** | 7 | 8974 | -0.02 | 0.01 | 8.66E-04 | **1.75E-02** | 12 | -0.03 | 0.01 | 3.61E-07 | 0.00 | 5.96E-01 | 0.27 | 1 |
| Grams of tobacco smoked per week | Tobacco use | 6076 | 0.06 | 0.02 | 2.40E-04 | **5.30E-03** | 9 | 5847 | 0.04 | 0.01 | 2.08E-03 | **2.65E-02** | 17 | 0.05 | 0.01 | 1.95E-06 | 0.00 | 5.54E-01 | 0.25 | 9 |
| Number of cigarretes smoked per day | Tobacco use | 6076 | 0.06 | 0.02 | 2.40E-04 | **5.30E-03** | 10 | 5847 | 0.04 | 0.01 | 2.08E-03 | **2.65E-02** | 18 | 0.05 | 0.01 | 1.95E-06 | 0.00 | 5.54E-01 | 0.25 | 7 |
| Number of cigars smoked per day | Tobacco use | 6076 | 0.06 | 0.02 | 2.40E-04 | **5.30E-03** | 11 | 5847 | 0.04 | 0.01 | 2.08E-03 | **2.65E-02** | 19 | 0.05 | 0.01 | 1.95E-06 | 0.00 | 5.54E-01 | 0.25 | 8 |
| Frequency of picking berries or mushrooms during leisure time | Physical activity | 9340 | -0.03 | 0.01 | 2.31E-05 | **9.36E-04** | 6 | 9184 | -0.01 | 0.01 | 6.14E-02 | 2.26E-01 | 66 | -0.02 | 0.01 | 1.93E-05 | 0.69 | 7.48E-02 | - | - |
| Sour milk, yoghurt (3% fat) | Food | 9175 | -0.02 | 0.01 | 8.57E-04 | **1.74E-02** | 12 | 8974 | -0.02 | 0.01 | 1.06E-02 | 7.82E-02 | 33 | -0.02 | 0.01 | 3.25E-05 | 0.00 | 5.34E-01 | - | - |
| Strong beer | Alcohol | 9175 | 0.02 | 0.01 | 4.15E-03 | 6.05E-02 | 16 | 8974 | 0.02 | 0.01 | 3.85E-03 | **4.07E-02** | 23 | 0.02 | 0.01 | 4.68E-05 | 0.00 | 9.83E-01 | - | - |
| Long-term sickness | General health | 9610 | 0.05 | 0.02 | 1.07E-02 | 1.09E-01 | 24 | 9361 | 0.06 | 0.02 | 1.61E-03 | **2.55E-02** | 15 | 0.06 | 0.01 | 5.31E-05 | 0.00 | 7.15E-01 | - | - |
| White (soft) bread, thin crisp bread | Food | 9175 | 0.01 | 0.01 | 1.35E-01 | 3.87E-01 | 85 | 8974 | 0.03 | 0.01 | 1.01E-04 | **3.50E-03** | 7 | 0.02 | 0.01 | 1.17E-04 | 0.60 | 1.12E-01 | - | - |
| Eat breakfast from 2000 | Food | 1054 | -0.19 | 0.08 | 2.56E-02 | 1.88E-01 | 33 | 967 | -0.26 | 0.08 | 1.68E-03 | **2.55E-02** | 16 | -0.22 | 0.06 | 1.40E-04 | 0.00 | 5.15E-01 | - | - |
| Liquor, spirits | Alcohol | 9175 | 0.01 | 0.01 | 7.21E-02 | 2.97E-01 | 59 | 8974 | 0.03 | 0.01 | 4.17E-04 | **1.27E-02** | 8 | 0.02 | 0.01 | 1.45E-04 | 0.21 | 2.61E-01 | - | - |
| Feel uneasy or guilty because of your way of drinking | Alcohol | 8948 | 0.02 | 0.02 | 4.78E-01 | 7.12E-01 | 163 | 8839 | 0.10 | 0.02 | 7.41E-06 | **3.97E-04** | 4 | 0.06 | 0.02 | 1.80E-04 | 0.85 | 1.03E-02 | - | - |
| Parents or siblings have diabetes | General health | 9838 | 0.06 | 0.02 | 2.93E-03 | **4.74E-02** | 15 | 9654 | 0.04 | 0.02 | 2.16E-02 | 1.17E-01 | 45 | 0.05 | 0.01 | 2.06E-04 | 0.00 | 5.49E-01 | - | - |
| Sleep status | Sleep | 5617 | -0.02 | 0.01 | 6.71E-02 | 2.90E-01 | 56 | 5477 | -0.03 | 0.01 | 1.10E-03 | **1.91E-02** | 14 | -0.02 | 0.01 | 2.73E-04 | 0.00 | 3.83E-01 | - | - |
| Frequency of shoverling snow during leisure time | Physical activity | 9460 | -0.02 | 0.01 | 1.62E-02 | 1.41E-01 | 28 | 9329 | -0.02 | 0.01 | 6.45E-03 | 5.60E-02 | 28 | -0.02 | 0.01 | 2.80E-04 | 0.00 | 9.21E-01 | - | - |
| Participation in associations or voluntary organisations | Social | 9895 | -0.03 | 0.01 | 8.79E-02 | 3.24E-01 | 66 | 9694 | -0.05 | 0.01 | 7.16E-04 | **1.64E-02** | 10 | -0.04 | 0.01 | 2.90E-04 | 0.19 | 2.66E-01 | - | - |
| Ice cream | Food | 9175 | -0.02 | 0.01 | 7.35E-03 | 8.28E-02 | 21 | 8974 | -0.02 | 0.01 | 1.43E-02 | 8.58E-02 | 40 | -0.02 | 0.01 | 2.99E-04 | 0.00 | 7.41E-01 | - | - |
| Years using snuff | Tobacco use | 9127 | 0.02 | 0.01 | 4.00E-02 | 2.21E-01 | 44 | 8924 | 0.02 | 0.01 | 2.38E-03 | **2.89E-02** | 20 | 0.02 | 0.01 | 3.03E-04 | 0.00 | 5.25E-01 | - | - |
| Monounsaturated fat intake (g/day) | Nutrients | 9175 | 0.02 | 0.01 | 1.43E-02 | 1.34E-01 | 26 | 8974 | 0.02 | 0.01 | 8.60E-03 | 7.07E-02 | 29 | 0.02 | 0.01 | 3.28E-04 | 0.00 | 9.62E-01 | - | - |
| Fried potatoes, pommes frites | Food | 9175 | 0.02 | 0.01 | 7.50E-03 | 8.28E-02 | 22 | 8974 | 0.02 | 0.01 | 1.81E-02 | 1.05E-01 | 42 | 0.02 | 0.01 | 3.75E-04 | 0.00 | 7.63E-01 | - | - |
| Parents or siblings had a cerebral hemorrhage/thrombosis or cardiac infarction before the age of 60 | General health | 9828 | 0.04 | 0.02 | 3.56E-02 | 2.19E-01 | 39 | 9638 | 0.05 | 0.02 | 4.87E-03 | **4.38E-02** | 27 | 0.04 | 0.01 | 4.89E-04 | 0.00 | 6.61E-01 | - | - |
| Informed of having high blood pressure | General health | 9930 | 0.03 | 0.02 | 1.56E-01 | 4.08E-01 | 93 | 9739 | 0.06 | 0.02 | 6.75E-04 | **1.64E-02** | 9 | 0.05 | 0.01 | 5.52E-04 | 0.39 | 1.99E-01 | - | - |
| Meat on bread | Food | 9175 | 0.02 | 0.01 | 1.29E-03 | **2.41E-02** | 13 | 8974 | 0.01 | 0.01 | 1.22E-01 | 3.17E-01 | 93 | 0.02 | 0.01 | 7.54E-04 | 0.29 | 2.35E-01 | - | - |
| Average portion size of vegetables based on photographic illustration of four sizes (smallest to largest) | Food | 9175 | -0.02 | 0.01 | 3.47E-02 | 2.19E-01 | 38 | 8974 | -0.02 | 0.01 | 1.15E-02 | 8.22E-02 | 34 | -0.02 | 0.01 | 1.01E-03 | 0.00 | 8.32E-01 | - | - |
| Breakfast habits: Porridge w/o sandwich for breakfast vs not breakfast at all | Food | 1312 | -0.14 | 0.05 | 7.22E-03 | 8.28E-02 | 20 | 1292 | -0.09 | 0.05 | 4.62E-02 | 1.87E-01 | 60 | -0.11 | 0.03 | 1.01E-03 | 0.00 | 5.16E-01 | - | - |
| Total energy intake (kcal/day) | Nutrients | 9175 | -0.02 | 0.01 | 3.88E-02 | 2.21E-01 | 42 | 8974 | -0.02 | 0.01 | 1.23E-02 | 8.46E-02 | 35 | -0.02 | 0.01 | 1.20E-03 | 0.00 | 8.18E-01 | - | - |
| Skill demand from job | Psychosocial | 9779 | -0.01 | 0.01 | 9.87E-02 | 3.29E-01 | 73 | 9581 | -0.02 | 0.01 | 4.26E-03 | **4.31E-02** | 24 | -0.02 | 0.00 | 1.32E-03 | 0.00 | 4.46E-01 | - | - |
| Chips, popcorn, salted nuts | Food | 9175 | 0.01 | 0.01 | 1.52E-01 | 4.08E-01 | 89 | 8974 | 0.02 | 0.01 | 4.54E-03 | **4.38E-02** | 25 | 0.02 | 0.01 | 2.55E-03 | 0.00 | 3.17E-01 | - | - |
| Alcohol intake (g/day) | Alcohol | 9175 | 0.02 | 0.01 | 4.62E-02 | 2.44E-01 | 46 | 8974 | 0.02 | 0.01 | 2.66E-02 | 1.32E-01 | 49 | 0.02 | 0.01 | 2.88E-03 | 0.00 | 9.14E-01 | - | - |
| Fibre intake (g/day) | Nutrients | 9175 | -0.02 | 0.01 | 1.71E-02 | 1.44E-01 | 29 | 8974 | -0.01 | 0.01 | 7.20E-02 | 2.49E-01 | 70 | -0.02 | 0.01 | 3.24E-03 | 0.00 | 6.15E-01 | - | - |
| Bacon | Food | 9175 | 0.02 | 0.01 | 5.79E-03 | 7.81E-02 | 18 | 8974 | 0.01 | 0.01 | 1.51E-01 | 3.46E-01 | 106 | 0.01 | 0.00 | 3.32E-03 | 0.05 | 3.04E-01 | - | - |
| Cookies, pastry | Food | 9175 | -0.01 | 0.01 | 1.42E-01 | 3.99E-01 | 86 | 8974 | -0.02 | 0.01 | 9.02E-03 | 7.07E-02 | 31 | -0.01 | 0.01 | 3.62E-03 | 0.00 | 4.74E-01 | - | - |
| Appetite status | Psychosocial | 5606 | -0.02 | 0.01 | 5.62E-02 | 2.72E-01 | 50 | 5468 | -0.02 | 0.01 | 3.42E-02 | 1.51E-01 | 55 | -0.02 | 0.01 | 4.35E-03 | 0.00 | 9.74E-01 | - | - |
| Carbohydrates intake (g/day) | Nutrients | 9175 | -0.02 | 0.01 | 2.71E-02 | 1.94E-01 | 34 | 8974 | -0.01 | 0.01 | 7.64E-02 | 2.54E-01 | 73 | -0.01 | 0.01 | 5.00E-03 | 0.00 | 7.04E-01 | - | - |
| Beta-sitosterol intake (mg/day) | Nutrients | 9175 | -0.01 | 0.01 | 1.53E-01 | 4.08E-01 | 90 | 8974 | -0.02 | 0.01 | 1.45E-02 | 8.58E-02 | 41 | -0.01 | 0.01 | 5.78E-03 | 0.00 | 5.22E-01 | - | - |
| Teetotaler | Alcohol | 9906 | -0.05 | 0.02 | 3.05E-02 | 2.12E-01 | 35 | 9719 | -0.04 | 0.02 | 8.53E-02 | 2.59E-01 | 80 | -0.05 | 0.02 | 6.03E-03 | 0.00 | 7.47E-01 | - | - |
| Lariciresinol intake (ug/day) | Nutrients | 9175 | -0.02 | 0.01 | 2.54E-02 | 1.88E-01 | 32 | 8974 | -0.01 | 0.01 | 1.02E-01 | 2.86E-01 | 87 | -0.01 | 0.01 | 6.47E-03 | 0.00 | 6.19E-01 | - | - |
| Coffee rolls/buns, rusk | Food | 9175 | -0.01 | 0.01 | 7.95E-02 | 3.12E-01 | 62 | 8974 | -0.01 | 0.01 | 3.89E-02 | 1.63E-01 | 58 | -0.01 | 0.01 | 6.79E-03 | 0.00 | 9.10E-01 | - | - |
| Berries (fresh or frozen) | Food | 9175 | -0.01 | 0.01 | 9.71E-02 | 3.28E-01 | 72 | 8974 | -0.01 | 0.01 | 3.25E-02 | 1.49E-01 | 53 | -0.01 | 0.00 | 7.10E-03 | 0.00 | 7.78E-01 | - | - |
| Breakfast habits: Gruel w/o sandwich for breakfast vs not breakfast at all | Food | 728 | -0.11 | 0.09 | 2.08E-01 | 4.83E-01 | 104 | 706 | -0.19 | 0.08 | 1.42E-02 | 8.58E-02 | 38 | -0.16 | 0.06 | 7.24E-03 | 0.00 | 5.17E-01 | - | - |
| Pancake, waffle, Swedish dumpling | Food | 9175 | -0.02 | 0.01 | 3.97E-02 | 2.21E-01 | 43 | 8974 | -0.01 | 0.01 | 8.25E-02 | 2.59E-01 | 76 | -0.01 | 0.01 | 7.43E-03 | 0.00 | 7.75E-01 | - | - |
| Hamburger | Food | 9175 | 0.01 | 0.01 | 2.27E-01 | 4.89E-01 | 113 | 8974 | 0.02 | 0.01 | 1.37E-02 | 8.58E-02 | 37 | 0.01 | 0.01 | 8.64E-03 | 0.00 | 4.21E-01 | - | - |
| Cohabitation: Live alone vs Only one adult (spouse, partner) | Social | 2988 | -0.08 | 0.03 | 1.00E-02 | 1.06E-01 | 23 | 3030 | -0.03 | 0.03 | 2.52E-01 | 4.74E-01 | 129 | -0.06 | 0.02 | 8.68E-03 | 0.06 | 3.02E-01 | - | - |
| Sodas, soft drinks, juice | Beverage | 9175 | 0.01 | 0.01 | 2.17E-01 | 4.89E-01 | 106 | 8974 | 0.02 | 0.01 | 1.43E-02 | 8.58E-02 | 39 | 0.01 | 0.01 | 8.95E-03 | 0.00 | 4.04E-01 | - | - |
| Sodium intake (mg/day) | Nutrients | 9175 | 0.01 | 0.01 | 1.07E-01 | 3.50E-01 | 74 | 8974 | 0.01 | 0.01 | 3.84E-02 | 1.63E-01 | 57 | 0.01 | 0.01 | 9.01E-03 | 0.00 | 7.89E-01 | - | - |
| Tea | Beverage | 9175 | -0.01 | 0.01 | 1.54E-01 | 4.08E-01 | 91 | 8974 | -0.02 | 0.01 | 3.01E-02 | 1.44E-01 | 51 | -0.01 | 0.01 | 1.07E-02 | 0.00 | 6.38E-01 | - | - |
| Cohabitation: Live alone vs Adult and children | Social | 7105 | -0.06 | 0.03 | 1.86E-02 | 1.51E-01 | 30 | 6811 | -0.03 | 0.03 | 2.41E-01 | 4.62E-01 | 127 | -0.05 | 0.02 | 1.28E-02 | 0.00 | 3.98E-01 | - | - |
| Light beer | Alcohol | 9175 | 0.00 | 0.01 | 9.81E-01 | 9.96E-01 | 239 | 8974 | -0.03 | 0.01 | 7.44E-04 | **1.64E-02** | 11 | -0.01 | 0.01 | 1.49E-02 | 0.82 | 1.95E-02 | - | - |
| Satisfaction with economy | Psychosocial | 5612 | -0.01 | 0.01 | 1.43E-01 | 3.99E-01 | 87 | 5463 | -0.02 | 0.01 | 5.24E-02 | 1.99E-01 | 64 | -0.02 | 0.01 | 1.56E-02 | 0.00 | 8.02E-01 | - | - |
| Satisfaction with work situation | Psychosocial | 5560 | 0.00 | 0.01 | 7.05E-01 | 8.61E-01 | 199 | 5409 | -0.03 | 0.01 | 3.41E-03 | **3.77E-02** | 22 | -0.02 | 0.01 | 1.60E-02 | 0.66 | 8.73E-02 | - | - |
| Exercise during the last three months | Physical activity | 9919 | 0.00 | 0.01 | 6.01E-01 | 8.16E-01 | 179 | 9729 | -0.02 | 0.01 | 4.84E-03 | **4.38E-02** | 26 | -0.01 | 0.00 | 1.68E-02 | 0.60 | 1.14E-01 | - | - |
| Corn flakes | Food | 9175 | -0.02 | 0.01 | 1.52E-02 | 1.36E-01 | 27 | 8974 | -0.01 | 0.01 | 3.30E-01 | 5.50E-01 | 145 | -0.01 | 0.01 | 1.76E-02 | 0.18 | 2.70E-01 | - | - |
| Travel to work: Cycle to work vs passive travel to work | Physical activity | 7640 | -0.03 | 0.02 | 6.41E-02 | 2.83E-01 | 55 | 7528 | -0.03 | 0.02 | 1.40E-01 | 3.31E-01 | 103 | -0.03 | 0.01 | 1.87E-02 | 0.00 | 7.84E-01 | - | - |
| Sum of phytosterols intake (mg/day) | Nutrients | 9175 | -0.01 | 0.01 | 2.91E-01 | 5.27E-01 | 134 | 8974 | -0.02 | 0.01 | 2.60E-02 | 1.32E-01 | 47 | -0.01 | 0.01 | 1.91E-02 | 0.00 | 4.44E-01 | - | - |
| Sum of Lariciresinol, Matairesinol, Pinoresinol, Secoisolariciresinol intake (ug/day) | Nutrients | 9175 | -0.01 | 0.01 | 5.38E-02 | 2.67E-01 | 49 | 8974 | -0.01 | 0.01 | 1.62E-01 | 3.58E-01 | 110 | -0.01 | 0.01 | 1.92E-02 | 0.00 | 6.61E-01 | - | - |
| Rosehip, sweet syrup soup | Food | 9175 | 0.02 | 0.01 | 6.17E-03 | 7.89E-02 | 19 | 8974 | 0.00 | 0.01 | 5.58E-01 | 7.32E-01 | 185 | 0.01 | 0.00 | 2.02E-02 | 0.59 | 1.17E-01 | - | - |
| Milk, sour milk (1,5%) | Beverage | 9175 | 0.01 | 0.01 | 3.58E-01 | 5.91E-01 | 147 | 8974 | 0.02 | 0.01 | 2.14E-02 | 1.17E-01 | 44 | 0.01 | 0.01 | 2.11E-02 | 0.00 | 3.62E-01 | - | - |
| Satisfaction with home and family situation | Psychosocial | 5613 | -0.01 | 0.01 | 1.29E-01 | 3.83E-01 | 81 | 5470 | -0.02 | 0.01 | 9.09E-02 | 2.73E-01 | 81 | -0.02 | 0.01 | 2.31E-02 | 0.00 | 9.77E-01 | - | - |
| Memory status | Psychosocial | 5595 | -0.02 | 0.01 | 1.23E-02 | 1.19E-01 | 25 | 5462 | -0.01 | 0.01 | 4.60E-01 | 6.53E-01 | 170 | -0.01 | 0.01 | 2.55E-02 | 0.45 | 1.76E-01 | - | - |
| Marital status: Single vs Divorced/separated | Social | 1568 | 0.08 | 0.05 | 8.16E-02 | 3.15E-01 | 63 | 1561 | 0.06 | 0.04 | 1.46E-01 | 3.41E-01 | 104 | 0.07 | 0.03 | 2.58E-02 | 0.00 | 6.68E-01 | - | - |
| Root vegetables, carrot | Food | 9175 | -0.01 | 0.01 | 1.70E-01 | 4.25E-01 | 97 | 8974 | -0.01 | 0.01 | 7.97E-02 | 2.58E-01 | 75 | -0.01 | 0.01 | 2.65E-02 | 0.00 | 8.47E-01 | - | - |
| Sour milk, yoghurt (low fat) | Food | 9175 | -0.01 | 0.01 | 2.27E-01 | 4.89E-01 | 112 | 8974 | -0.01 | 0.01 | 5.62E-02 | 2.10E-01 | 65 | -0.01 | 0.00 | 2.67E-02 | 0.00 | 6.57E-01 | - | - |
| Would you say that the number of people that you meet in your everyday life is enough or would you like to meet more or fewer people? | Social | 9907 | -0.01 | 0.01 | 1.19E-01 | 3.75E-01 | 77 | 9705 | -0.01 | 0.01 | 1.16E-01 | 3.06E-01 | 92 | -0.01 | 0.00 | 2.68E-02 | 0.00 | 9.60E-01 | - | - |
| Satisfaction with accomodation | Psychosocial | 5618 | -0.02 | 0.01 | 6.84E-02 | 2.90E-01 | 57 | 5472 | -0.01 | 0.01 | 1.83E-01 | 3.93E-01 | 113 | -0.01 | 0.01 | 2.69E-02 | 0.00 | 6.56E-01 | - | - |
| Enough time for job assignments | Psychosocial | 9764 | -0.01 | 0.01 | 5.05E-02 | 2.55E-01 | 48 | 9556 | -0.01 | 0.01 | 2.67E-01 | 4.92E-01 | 132 | -0.01 | 0.00 | 3.10E-02 | 0.00 | 5.24E-01 | - | - |
| Pinoresinol intake (ug/day) | Nutrients | 9175 | -0.01 | 0.01 | 9.57E-02 | 3.28E-01 | 71 | 8974 | -0.01 | 0.01 | 1.66E-01 | 3.63E-01 | 111 | -0.01 | 0.01 | 3.14E-02 | 0.00 | 7.99E-01 | - | - |
| Vitamin B12 intake (ug/day) | Nutrients | 9175 | 0.01 | 0.01 | 4.28E-01 | 6.58E-01 | 158 | 8974 | 0.02 | 0.01 | 3.16E-02 | 1.48E-01 | 52 | 0.01 | 0.01 | 3.55E-02 | 0.00 | 3.63E-01 | - | - |
| Apple, pear, peach, orange, mandarin and grapefruit | Food | 9175 | -0.02 | 0.01 | 3.20E-02 | 2.14E-01 | 36 | 8974 | -0.01 | 0.01 | 3.90E-01 | 5.98E-01 | 158 | -0.01 | 0.01 | 3.57E-02 | 0.00 | 3.36E-01 | - | - |
| Beta-carotene intake (mg/day) | Nutrients | 9175 | -0.01 | 0.01 | 2.26E-01 | 4.89E-01 | 110 | 8974 | -0.01 | 0.01 | 8.32E-02 | 2.59E-01 | 77 | -0.01 | 0.01 | 3.63E-02 | 0.00 | 7.66E-01 | - | - |
| Wine | Alcohol | 9175 | 0.01 | 0.01 | 2.25E-01 | 4.89E-01 | 108 | 8974 | 0.01 | 0.01 | 8.42E-02 | 2.59E-01 | 79 | 0.01 | 0.01 | 3.68E-02 | 0.00 | 7.54E-01 | - | - |
| Average portion size of potatoes/rice/pasta based on photographic illustration of four sizes (smallest to largest) | Food | 9175 | -0.01 | 0.01 | 2.74E-01 | 5.26E-01 | 123 | 8974 | -0.01 | 0.01 | 7.56E-02 | 2.54E-01 | 72 | -0.01 | 0.01 | 4.10E-02 | 0.00 | 6.72E-01 | - | - |
| Arachidonic acid (ARA) intake (g/day) | Nutrients | 9175 | 0.01 | 0.01 | 6.92E-02 | 2.90E-01 | 58 | 8974 | 0.01 | 0.01 | 2.75E-01 | 5.03E-01 | 133 | 0.01 | 0.01 | 4.13E-02 | 0.00 | 5.65E-01 | - | - |
| Vitamin E intake (mg/day) | Nutrients | 9175 | -0.01 | 0.01 | 3.17E-01 | 5.57E-01 | 138 | 8974 | -0.01 | 0.01 | 6.46E-02 | 2.34E-01 | 67 | -0.01 | 0.01 | 4.18E-02 | 0.00 | 5.99E-01 | - | - |
| Sausage, liver pate on bread | Food | 9175 | 0.01 | 0.01 | 2.09E-01 | 4.83E-01 | 105 | 8974 | 0.01 | 0.01 | 1.14E-01 | 3.05E-01 | 91 | 0.01 | 0.01 | 4.39E-02 | 0.00 | 8.97E-01 | - | - |
| Hearing status | General health | 5610 | -0.01 | 0.01 | 4.13E-01 | 6.52E-01 | 154 | 5474 | -0.02 | 0.01 | 4.93E-02 | 1.93E-01 | 61 | -0.01 | 0.01 | 4.52E-02 | 0.00 | 4.69E-01 | - | - |
| Zinc intake (mg/day) | Nutrients | 9175 | 0.00 | 0.01 | 5.52E-01 | 7.75E-01 | 173 | 8974 | -0.02 | 0.01 | 2.93E-02 | 1.42E-01 | 50 | -0.01 | 0.01 | 4.63E-02 | 0.12 | 2.86E-01 | - | - |
| Enterolactone intake (ug/day) | Nutrients | 9175 | -0.01 | 0.01 | 6.24E-02 | 2.81E-01 | 54 | 8974 | -0.01 | 0.01 | 3.27E-01 | 5.50E-01 | 142 | -0.01 | 0.01 | 4.65E-02 | 0.00 | 4.94E-01 | - | - |
| Frequency of gardening during leisure time | Physical activity | 9395 | -0.01 | 0.01 | 1.35E-01 | 3.87E-01 | 84 | 9239 | -0.01 | 0.01 | 1.88E-01 | 3.98E-01 | 115 | -0.01 | 0.01 | 4.74E-02 | 0.00 | 8.50E-01 | - | - |

| | | | | | | | | | | | | | | | | | | | | |
|---|---|---|---|---|---|---|---|---|---|---|---|---|---|---|---|---|---|---|---|---|
| Mashed potato | Food | 3917 | 0.02 | 0.01 | 1.28E-01 | 3.83E-01 | 80 | 3865 | 0.01 | 0.01 | 2.06E-01 | 4.20E-01 | 119 | 0.01 | 0.01 | 4.91E-02 | 0.00 | 8.26E-01 | - | - |
| Salad dressing with oil | Food | 9175 | -0.01 | 0.01 | 3.26E-01 | 5.65E-01 | 140 | 8974 | -0.01 | 0.01 | 7.75E-02 | 2.54E-01 | 74 | -0.01 | 0.00 | 4.96E-02 | 0.00 | 6.33E-01 | - | - |
| Learn new things at job | Psychosocial | 9790 | -0.01 | 0.01 | 9.08E-02 | 3.25E-01 | 67 | 9587 | -0.01 | 0.01 | 2.78E-01 | 5.04E-01 | 134 | -0.01 | 0.00 | 5.08E-02 | 0.00 | 6.37E-01 | - | - |
| Monosaccharides intake (g/day) | Nutrients | 9175 | -0.01 | 0.01 | 1.98E-01 | 4.77E-01 | 101 | 8974 | -0.01 | 0.01 | 1.47E-01 | 3.41E-01 | 105 | -0.01 | 0.01 | 5.28E-02 | 0.00 | 9.46E-01 | - | - |
| Stigmasterol intake (mg/day) | Nutrients | 9175 | 0.00 | 0.01 | 5.36E-01 | 7.63E-01 | 170 | 8974 | -0.01 | 0.01 | 3.83E-02 | 1.63E-01 | 56 | -0.01 | 0.01 | 5.33E-02 | 0.00 | 3.32E-01 | - | - |
| Iodine intake (ug/day) | Nutrients | 9175 | 0.01 | 0.01 | 3.21E-01 | 5.61E-01 | 139 | 8974 | 0.01 | 0.01 | 9.36E-02 | 2.74E-01 | 83 | 0.01 | 0.01 | 5.78E-02 | 0.00 | 6.57E-01 | - | - |
| High physical demand from job | Physical activity | 9828 | 0.00 | 0.01 | 9.93E-01 | 9.97E-01 | 242 | 9619 | 0.02 | 0.01 | 8.83E-03 | 7.07E-02 | 30 | 0.01 | 0.00 | 5.82E-02 | 0.69 | 7.05E-02 | - | - |
| Control over planning and execution of the workday | Psychosocial | 9813 | 0.00 | 0.01 | 7.09E-01 | 8.61E-01 | 200 | 9603 | -0.02 | 0.01 | 2.43E-02 | 1.29E-01 | 46 | -0.01 | 0.00 | 5.93E-02 | 0.40 | 1.98E-01 | - | - |
| Close relationship with anyone | Social | 9908 | 0.00 | 0.01 | 5.15E-01 | 7.54E-01 | 166 | 9721 | -0.01 | 0.01 | 4.93E-02 | 1.93E-01 | 62 | -0.01 | 0.00 | 6.11E-02 | 0.00 | 3.76E-01 | - | - |
| Folic acid intake (ug/day) | Nutrients | 9175 | -0.01 | 0.01 | 4.27E-01 | 6.58E-01 | 157 | 8974 | -0.01 | 0.01 | 6.86E-02 | 2.45E-01 | 68 | -0.01 | 0.01 | 6.16E-02 | 0.00 | 5.01E-01 | - | - |
| Educational level | Psychosocial | 9915 | -0.01 | 0.01 | 1.62E-01 | 4.13E-01 | 95 | 9729 | -0.01 | 0.01 | 2.13E-01 | 4.29E-01 | 120 | -0.01 | 0.01 | 6.19E-02 | 0.00 | 8.82E-01 | - | - |
| Butter for cooking | Food | 9175 | 0.00 | 0.01 | 6.80E-01 | 8.61E-01 | 192 | 8974 | 0.01 | 0.01 | 3.31E-02 | 1.49E-01 | 54 | 0.01 | 0.01 | 6.63E-02 | 0.25 | 2.47E-01 | - | - |
| Mood status | Psychosocial | 5606 | -0.01 | 0.01 | 2.02E-01 | 4.80E-01 | 102 | 5463 | -0.01 | 0.01 | 2.01E-01 | 4.13E-01 | 118 | -0.01 | 0.01 | 7.08E-02 | 0.00 | 9.52E-01 | - | - |
| Magnesium intake (mg/day) | Nutrients | 9175 | -0.01 | 0.01 | 3.89E-01 | 6.30E-01 | 150 | 8974 | -0.01 | 0.01 | 1.23E-01 | 3.17E-01 | 94 | -0.01 | 0.01 | 8.64E-02 | 0.00 | 6.67E-01 | - | - |
| Self-employed | Psychosocial | 9721 | -0.04 | 0.02 | 8.62E-02 | 3.22E-01 | 65 | 9530 | -0.02 | 0.02 | 4.65E-01 | 6.53E-01 | 173 | -0.03 | 0.02 | 8.73E-02 | 0.00 | 4.57E-01 | - | - |
| Smoked fish/meat | Food | 9175 | 0.01 | 0.01 | 3.81E-01 | 6.21E-01 | 149 | 8974 | 0.01 | 0.01 | 1.36E-01 | 3.31E-01 | 100 | 0.01 | 0.00 | 9.22E-02 | 0.00 | 6.97E-01 | - | - |
| Number of social interactions during a normal week | Social | 9900 | -0.01 | 0.01 | 4.52E-01 | 6.83E-01 | 161 | 9724 | -0.01 | 0.01 | 1.10E-01 | 2.98E-01 | 89 | -0.01 | 0.00 | 9.37E-02 | 0.00 | 5.79E-01 | - | - |
| Oatflake, whole wheat, rye or barley porridge | Food | 9175 | -0.01 | 0.01 | 5.75E-02 | 2.72E-01 | 51 | 8974 | 0.00 | 0.01 | 6.34E-01 | 7.78E-01 | 198 | -0.01 | 0.01 | 9.73E-02 | 0.08 | 2.97E-01 | - | - |
| Rice | Food | 9175 | -0.01 | 0.01 | 1.49E-01 | 4.08E-01 | 88 | 8974 | -0.01 | 0.01 | 3.74E-01 | 5.83E-01 | 156 | -0.01 | 0.01 | 1.01E-01 | 0.00 | 6.71E-01 | - | - |
| Support from others | Social | 9903 | -0.01 | 0.01 | 2.56E-01 | 5.23E-01 | 119 | 9717 | -0.01 | 0.01 | 2.38E-01 | 4.62E-01 | 125 | -0.01 | 0.00 | 1.01E-01 | 0.00 | 9.99E-01 | - | - |
| Ingenuity or creativity demand from job | Psychosocial | 9764 | -0.01 | 0.01 | 2.56E-01 | 5.23E-01 | 118 | 9561 | -0.01 | 0.01 | 2.40E-01 | 4.62E-01 | 126 | -0.01 | 0.00 | 1.02E-01 | 0.00 | 9.97E-01 | - | - |
| Salty fish | Food | 9175 | 0.01 | 0.01 | 1.78E-01 | 4.40E-01 | 98 | 8974 | 0.01 | 0.01 | 3.33E-01 | 5.50E-01 | 147 | 0.01 | 0.00 | 1.03E-01 | 0.00 | 7.61E-01 | - | - |
| Butter on bread | Food | 9175 | 0.00 | 0.01 | 7.34E-01 | 8.74E-01 | 203 | 8974 | 0.02 | 0.01 | 9.35E-03 | 7.10E-02 | 32 | 0.01 | 0.00 | 1.04E-01 | 0.76 | 3.96E-02 | - | - |
| Equol intake (ug/day) | Nutrients | 9175 | -0.01 | 0.01 | 4.67E-01 | 7.01E-01 | 162 | 8974 | -0.01 | 0.01 | 1.38E-01 | 3.31E-01 | 101 | -0.01 | 0.01 | 1.15E-01 | 0.00 | 6.20E-01 | - | - |
| Sausage as main dish | Food | 9175 | 0.00 | 0.01 | 5.56E-01 | 7.76E-01 | 174 | 8974 | 0.01 | 0.01 | 1.06E-01 | 2.92E-01 | 88 | 0.01 | 0.01 | 1.17E-01 | 0.00 | 4.76E-01 | - | - |
| Calcium intake (mg/day) | Nutrients | 9175 | -0.01 | 0.01 | 1.13E-01 | 3.65E-01 | 75 | 8974 | 0.00 | 0.01 | 5.12E-01 | 6.95E-01 | 179 | -0.01 | 0.01 | 1.18E-01 | 0.00 | 4.80E-01 | - | - |
| Vitamin B2 intake (ug/day) | Nutrients | 9175 | -0.01 | 0.01 | 4.21E-02 | 2.27E-01 | 45 | 8974 | 0.00 | 0.01 | 8.18E-01 | 9.02E-01 | 220 | -0.01 | 0.01 | 1.18E-01 | 0.42 | 1.87E-01 | - | - |
| Linoleic acid intake (g/day) | Nutrients | 9175 | 0.00 | 0.01 | 7.38E-01 | 8.74E-01 | 205 | 8974 | -0.01 | 0.01 | 6.98E-02 | 2.46E-01 | 69 | -0.01 | 0.00 | 1.23E-01 | 0.02 | 3.13E-01 | - | - |
| Cheese 28% | Food | 9175 | -0.01 | 0.01 | 2.05E-01 | 4.83E-01 | 103 | 8974 | -0.01 | 0.01 | 3.55E-01 | 5.64E-01 | 152 | -0.01 | 0.01 | 1.23E-01 | 0.00 | 7.71E-01 | - | - |
| Palmitic acid intake (g/day) | Nutrients | 9175 | 0.01 | 0.01 | 4.01E-01 | 6.37E-01 | 152 | 8974 | 0.01 | 0.01 | 1.97E-01 | 4.10E-01 | 117 | 0.01 | 0.01 | 1.30E-01 | 0.00 | 7.80E-01 | - | - |
| Number of social contacts with the same interests as you | Social | 9881 | -0.01 | 0.01 | 3.14E-01 | 5.57E-01 | 137 | 9693 | -0.01 | 0.01 | 2.59E-01 | 4.84E-01 | 130 | -0.01 | 0.00 | 1.31E-01 | 0.00 | 9.57E-01 | - | - |
| Minced meat dishes | Food | 9175 | 0.02 | 0.01 | 1.53E-03 | **2.66E-02** | 14 | 8974 | -0.01 | 0.01 | 3.85E-01 | 5.97E-01 | 157 | 0.01 | 0.01 | 1.36E-01 | 0.88 | 3.42E-03 | - | - |
| Campesterol intake (mg/day) | Nutrients | 9175 | 0.00 | 0.01 | 9.38E-01 | 9.86E-01 | 231 | 8974 | -0.01 | 0.01 | 5.16E-02 | 1.99E-01 | 63 | -0.01 | 0.01 | 1.44E-01 | 0.40 | 1.98E-01 | - | - |
| Vitamin B3 intake (mg/day) | Nutrients | 9175 | 0.01 | 0.01 | 7.76E-02 | 3.09E-01 | 61 | 8974 | 0.00 | 0.01 | 7.35E-01 | 8.58E-01 | 208 | 0.01 | 0.01 | 1.44E-01 | 0.09 | 2.94E-01 | - | - |
| Margarine on bread | Food | 9175 | -0.01 | 0.01 | 2.43E-01 | 5.19E-01 | 114 | 8974 | -0.01 | 0.01 | 3.73E-01 | 5.83E-01 | 155 | -0.01 | 0.01 | 1.49E-01 | 0.00 | 7.87E-01 | - | - |
| Patience status | Psychosocial | 5610 | -0.01 | 0.01 | 2.25E-01 | 4.89E-01 | 109 | 5468 | -0.01 | 0.01 | 4.13E-01 | 6.16E-01 | 163 | -0.01 | 0.01 | 1.55E-01 | 0.00 | 7.32E-01 | - | - |
| Plant based protein intake (g/day) | Nutrients | 9175 | -0.01 | 0.01 | 9.23E-02 | 3.25E-01 | 68 | 8974 | 0.00 | 0.01 | 7.28E-01 | 8.58E-01 | 206 | -0.01 | 0.01 | 1.60E-01 | 0.00 | 3.22E-01 | - | - |
| Receive hugs to comfort and support you | Social | 9866 | -0.01 | 0.02 | 4.94E-01 | 7.32E-01 | 164 | 9666 | -0.02 | 0.02 | 2.14E-01 | 4.29E-01 | 121 | -0.02 | 0.01 | 1.71E-01 | 0.00 | 7.16E-01 | - | - |
| Milk, sour milk (3%) | Beverage | 9175 | 0.00 | 0.01 | 8.31E-01 | 9.39E-01 | 215 | 8974 | 0.01 | 0.01 | 9.85E-02 | 2.81E-01 | 85 | 0.01 | 0.01 | 1.76E-01 | 0.00 | 3.32E-01 | - | - |
| Travel to work: Walk to work vs passive travel to work | Physical activity | 6463 | -0.03 | 0.03 | 3.44E-01 | 5.84E-01 | 143 | 6330 | -0.03 | 0.03 | 3.40E-01 | 5.53E-01 | 149 | -0.03 | 0.02 | 1.79E-01 | 0.00 | 1.00E+00 | - | - |
| Control over own work assignment | Psychosocial | 9807 | 0.00 | 0.01 | 8.49E-01 | 9.46E-01 | 217 | 9600 | -0.01 | 0.01 | 9.58E-02 | 2.77E-01 | 84 | -0.01 | 0.00 | 1.82E-01 | 0.03 | 3.11E-01 | - | - |
| Frequency of cycling during leisure time | Physical activity | 8441 | 0.00 | 0.01 | 7.42E-01 | 8.74E-01 | 206 | 8279 | -0.01 | 0.01 | 1.34E-01 | 3.30E-01 | 98 | -0.01 | 0.01 | 1.85E-01 | 0.00 | 4.39E-01 | - | - |
| Brown beans, pea soup | Food | 9175 | 0.01 | 0.01 | 2.76E-01 | 5.26E-01 | 126 | 8974 | 0.01 | 0.01 | 4.25E-01 | 6.25E-01 | 165 | 0.01 | 0.01 | 1.88E-01 | 0.00 | 7.62E-01 | - | - |
| Satisfaction with leisure time | Psychosocial | 5613 | 0.00 | 0.01 | 7.87E-01 | 9.09E-01 | 210 | 5456 | -0.02 | 0.01 | 4.31E-02 | 1.78E-01 | 59 | -0.01 | 0.01 | 1.95E-01 | 0.60 | 1.15E-01 | - | - |
| Matairesinol intake (ug/day) | Nutrients | 9175 | -0.01 | 0.01 | 1.31E-01 | 3.83E-01 | 83 | 8974 | 0.00 | 0.01 | 7.28E-01 | 8.58E-01 | 205 | -0.01 | 0.01 | 1.97E-01 | 0.00 | 3.91E-01 | - | - |
| Banana | Food | 9175 | 0.00 | 0.01 | 9.54E-01 | 9.88E-01 | 234 | 8974 | -0.01 | 0.01 | 7.28E-02 | 2.49E-01 | 71 | -0.01 | 0.01 | 2.03E-01 | 0.38 | 2.06E-01 | - | - |
| Blota (broth + bread) | Food | 3917 | 0.01 | 0.01 | 5.60E-01 | 7.77E-01 | 175 | 3865 | 0.01 | 0.01 | 2.15E-01 | 4.29E-01 | 122 | 0.01 | 0.01 | 2.05E-01 | 0.00 | 6.02E-01 | - | - |
| Permanent employment | Psychosocial | 9721 | 0.05 | 0.02 | 4.23E-03 | 6.05E-02 | 17 | 9530 | -0.02 | 0.02 | 3.14E-01 | 5.48E-01 | 139 | 0.01 | 0.01 | 2.16E-01 | 0.87 | 5.63E-03 | - | - |
| Saturated fat intake (g/day) | Nutrients | 9175 | 0.00 | 0.01 | 6.69E-01 | 8.56E-01 | 190 | 8974 | 0.01 | 0.01 | 1.93E-01 | 4.05E-01 | 116 | 0.01 | 0.01 | 2.16E-01 | 0.00 | 5.59E-01 | - | - |
| Vitamin D intake (ug/day) | Nutrients | 9175 | 0.02 | 0.01 | 2.40E-02 | 1.88E-01 | 31 | 8974 | 0.00 | 0.01 | 6.32E-01 | 7.78E-01 | 196 | 0.01 | 0.01 | 2.23E-01 | 0.74 | 4.99E-02 | - | - |
| Cohabitation: Live alone vs Other/others | Social | 1000 | -0.09 | 0.08 | 2.24E-01 | 4.89E-01 | 107 | 951 | -0.04 | 0.07 | 5.83E-01 | 7.45E-01 | 190 | -0.06 | 0.05 | 2.24E-01 | 0.00 | 5.82E-01 | - | - |
| Boiled coffee | Beverage | 9175 | 0.00 | 0.01 | 7.17E-01 | 8.67E-01 | 201 | 8974 | 0.01 | 0.01 | 1.85E-01 | 3.94E-01 | 114 | 0.01 | 0.01 | 2.25E-01 | 0.00 | 5.19E-01 | - | - |
| Oil for cooking | Food | 9175 | 0.00 | 0.01 | 8.71E-01 | 9.46E-01 | 223 | 8974 | -0.01 | 0.01 | 1.39E-01 | 3.31E-01 | 102 | -0.01 | 0.00 | 2.37E-01 | 0.00 | 3.66E-01 | - | - |
| Overall state of health compared to other your age | General health | 9750 | -0.01 | 0.01 | 2.79E-01 | 5.26E-01 | 129 | 9578 | 0.00 | 0.01 | 5.52E-01 | 7.32E-01 | 183 | -0.01 | 0.00 | 2.39E-01 | 0.00 | 7.11E-01 | - | - |
| Whole grain intake (g/day) | Food | 9175 | -0.01 | 0.01 | 8.41E-02 | 3.19E-01 | 64 | 8974 | 0.00 | 0.01 | 9.89E-01 | 9.90E-01 | 242 | -0.01 | 0.01 | 2.39E-01 | 0.37 | 2.06E-01 | - | - |
| Medium beer | Alcohol | 9175 | 0.01 | 0.01 | 4.98E-01 | 7.34E-01 | 165 | 8974 | 0.01 | 0.01 | 3.24E-01 | 5.50E-01 | 141 | 0.01 | 0.01 | 2.40E-01 | 0.00 | 8.20E-01 | - | - |
| Marital status: Single vs Widow/widower | Social | 1004 | 0.01 | 0.13 | 9.59E-01 | 9.88E-01 | 235 | 982 | 0.14 | 0.10 | 1.56E-01 | 3.52E-01 | 107 | 0.09 | 0.08 | 2.52E-01 | 0.00 | 4.00E-01 | - | - |
| Potato salad | Food | 3917 | -0.01 | 0.01 | 2.90E-01 | 5.27E-01 | 131 | 3865 | 0.03 | 0.01 | 1.25E-02 | 8.46E-02 | 36 | 0.01 | 0.01 | 2.71E-01 | 0.84 | 1.32E-02 | - | - |
| Phosphate intake (mg/day) | Nutrients | 9175 | -0.01 | 0.01 | 2.75E-01 | 5.26E-01 | 124 | 8974 | 0.00 | 0.01 | 6.73E-01 | 8.15E-01 | 200 | -0.01 | 0.01 | 2.91E-01 | 0.00 | 6.14E-01 | - | - |
| Breakfast habits: Coffee/tea and wheat buns or rusk for breakfast vs not breakfast at all | Food | 754 | -0.09 | 0.08 | 2.55E-01 | 5.23E-01 | 116 | 764 | -0.03 | 0.07 | 6.90E-01 | 8.26E-01 | 203 | -0.06 | 0.05 | 2.93E-01 | 0.00 | 5.54E-01 | - | - |
| Soft whey cheese | Food | 3917 | 0.00 | 0.01 | 9.88E-01 | 9.96E-01 | 241 | 3865 | -0.01 | 0.01 | 1.78E-01 | 3.85E-01 | 112 | -0.01 | 0.01 | 3.11E-01 | 0.00 | 3.73E-01 | - | - |
| Frequency of dancing during leisure time | Physical activity | 8789 | 0.01 | 0.01 | 2.91E-01 | 5.27E-01 | 133 | 8629 | -0.02 | 0.01 | 1.91E-02 | 1.08E-01 | 43 | -0.01 | 0.01 | 3.24E-01 | 0.82 | 1.76E-02 | - | - |
| Travel to work: Irregular travel mode to work vs passive travel to work | Physical activity | 6553 | 0.03 | 0.03 | 2.26E-01 | 4.89E-01 | 111 | 6440 | 0.01 | 0.02 | 8.39E-01 | 9.04E-01 | 223 | 0.02 | 0.02 | 3.30E-01 | 0.00 | 4.55E-01 | - | - |
| Energy status | Psychosocial | 5607 | -0.01 | 0.01 | 2.76E-01 | 5.26E-01 | 125 | 5460 | 0.00 | 0.01 | 7.61E-01 | 8.68E-01 | 213 | -0.01 | 0.01 | 3.35E-01 | 0.00 | 5.53E-01 | - | - |
| Iron intake (mg/day) | Nutrients | 9175 | -0.01 | 0.01 | 1.25E-01 | 3.83E-01 | 79 | 8974 | 0.00 | 0.01 | 8.77E-01 | 9.23E-01 | 231 | -0.01 | 0.01 | 3.41E-01 | 0.32 | 2.26E-01 | - | - |
| Whole grain crisp bread | Food | 9175 | 0.00 | 0.01 | 5.34E-01 | 7.63E-01 | 169 | 8974 | 0.01 | 0.01 | 4.72E-01 | 6.60E-01 | 174 | 0.00 | 0.01 | 3.43E-01 | 0.00 | 9.63E-01 | - | - |
| Marital status: Single vs Married/partner | Social | 9223 | -0.01 | 0.02 | 7.89E-01 | 9.09E-01 | 211 | 8984 | -0.02 | 0.02 | 2.91E-01 | 5.20E-01 | 136 | -0.02 | 0.02 | 3.46E-01 | 0.00 | 5.84E-01 | - | - |
| Eicosapentaenoic acid (EPA) intake (g/day) | Nutrients | 9175 | 0.01 | 0.01 | 1.14E-01 | 3.65E-01 | 76 | 8974 | 0.00 | 0.01 | 8.40E-01 | 9.04E-01 | 225 | 0.00 | 0.01 | 3.49E-01 | 0.40 | 1.97E-01 | - | - |
| Vision status | General health | 5608 | 0.01 | 0.01 | 2.96E-01 | 5.32E-01 | 135 | 5471 | -0.02 | 0.01 | 2.64E-02 | 1.32E-01 | 48 | -0.01 | 0.01 | 3.59E-01 | 0.81 | 2.28E-02 | - | - |
| Blood based food | Food | 3917 | -0.01 | 0.01 | 2.83E-01 | 5.27E-01 | 130 | 3865 | 0.00 | 0.01 | 8.44E-01 | 9.04E-01 | 227 | -0.01 | 0.01 | 3.62E-01 | 0.00 | 5.50E-01 | - | - |
| Docosahexaenoic acid (DHA) intake (g/day) | Nutrients | 9175 | 0.01 | 0.01 | 9.33E-02 | 3.25E-01 | 69 | 8974 | 0.00 | 0.01 | 7.34E-01 | 8.58E-01 | 207 | 0.00 | 0.01 | 3.68E-01 | 0.53 | 1.45E-01 | - | - |
| Polyunsaturated fat intake (g/day) | Nutrients | 9175 | 0.00 | 0.01 | 6.08E-01 | 8.21E-01 | 180 | 8974 | -0.01 | 0.01 | 8.38E-02 | 2.59E-01 | 78 | 0.00 | 0.00 | 3.72E-01 | 0.59 | 1.17E-01 | - | - |
| Low fat margarine on bread | Food | 9175 | 0.00 | 0.01 | 8.24E-01 | 9.35E-01 | 214 | 8974 | 0.01 | 0.01 | 3.19E-01 | 5.50E-01 | 140 | 0.00 | 0.01 | 3.80E-01 | 0.00 | 6.00E-01 | - | - |
| White cabbage, lettuce, lettuce cabbage, spinach, borecole | Food | 9175 | 0.00 | 0.01 | 6.84E-01 | 8.61E-01 | 193 | 8974 | -0.01 | 0.01 | 1.11E-01 | 2.98E-01 | 90 | 0.00 | 0.01 | 3.83E-01 | 0.49 | 1.62E-01 | - | - |
| Fat intake (g/day) | Nutrients | 9175 | 0.01 | 0.01 | 4.49E-01 | 6.81E-01 | 160 | 8974 | 0.00 | 0.01 | 6.30E-01 | 7.78E-01 | 195 | 0.00 | 0.01 | 3.84E-01 | 0.00 | 8.27E-01 | - | - |
| People to ask for help apart from the ones at home | Social | 9885 | 0.02 | 0.02 | 3.47E-01 | 5.86E-01 | 144 | 9698 | 0.01 | 0.02 | 7.73E-01 | 8.71E-01 | 214 | 0.01 | 0.02 | 3.94E-01 | 0.00 | 6.24E-01 | - | - |
| Tiamin intake (mg/day) | Nutrients | 9175 | 0.00 | 0.01 | 6.91E-01 | 8.61E-01 | 195 | 8974 | 0.01 | 0.01 | 4.30E-01 | 6.25E-01 | 167 | 0.00 | 0.01 | 3.97E-01 | 0.00 | 7.99E-01 | - | - |
| Linolenic acid intake (g/day) | Nutrients | 9175 | 0.00 | 0.01 | 5.89E-01 | 8.08E-01 | 177 | 8974 | -0.01 | 0.01 | 9.35E-02 | 2.74E-01 | 82 | 0.00 | 0.01 | 4.01E-01 | 0.58 | 1.21E-01 | - | - |
| Possibility to leave your work for a while to speak with a colleague | Psychosocial | 9739 | -0.01 | 0.01 | 1.63E-01 | 4.13E-01 | 96 | 9501 | 0.00 | 0.01 | 8.49E-01 | 9.05E-01 | 228 | 0.00 | 0.00 | 4.02E-01 | 0.22 | 2.58E-01 | - | - |
| Number of friends that can come to your home at any time and feel at home | Social | 9881 | 0.00 | 0.01 | 6.80E-01 | 8.61E-01 | 191 | 9696 | -0.01 | 0.01 | 4.45E-01 | 6.44E-01 | 168 | 0.00 | 0.00 | 4.02E-01 | 0.00 | 8.20E-01 | - | - |
| Frequency of walking during leisure time | Physical activity | 9647 | 0.00 | 0.01 | 6.65E-01 | 8.54E-01 | 189 | 9505 | -0.01 | 0.01 | 4.62E-01 | 6.53E-01 | 171 | 0.00 | 0.01 | 4.05E-01 | 0.00 | 8.47E-01 | - | - |
| Breakfast habits: Only coffee/tea for breakfast vs not breakfast at all | Food | 618 | 0.06 | 0.13 | 6.58E-01 | 8.53E-01 | 187 | 614 | 0.08 | 0.11 | 4.80E-01 | 6.62E-01 | 176 | 0.07 | 0.08 | 4.07E-01 | 0.00 | 9.24E-01 | - | - |
| Confidence status | Psychosocial | 5606 | 0.00 | 0.01 | 8.80E-01 | 9.50E-01 | 225 | 5467 | -0.01 | 0.01 | 3.39E-01 | 5.53E-01 | 148 | -0.01 | 0.01 | 4.24E-01 | 0.00 | 5.85E-01 | - | - |
| Potassium intake (mg/day) | Nutrients | 9175 | 0.00 | 0.01 | 6.36E-01 | 8.36E-01 | 185 | 8974 | -0.01 | 0.01 | 1.25E-01 | 3.20E-01 | 95 | 0.00 | 0.01 | 4.27E-01 | 0.49 | 1.63E-01 | - | - |
| Heptadecanoic acid intake (g/day) | Nutrients | 9175 | 0.00 | 0.01 | 6.32E-01 | 8.36E-01 | 183 | 8974 | 0.01 | 0.01 | 1.31E-01 | 3.28E-01 | 96 | 0.00 | 0.01 | 4.42E-01 | 0.48 | 1.66E-01 | - | - |
| Pentadecanoic acid intake (g/day) | Nutrients | 9175 | 0.00 | 0.01 | 6.32E-01 | 8.36E-01 | 182 | 8974 | 0.01 | 0.01 | 1.31E-01 | 3.28E-01 | 97 | 0.00 | 0.01 | 4.42E-01 | 0.48 | 1.66E-01 | - | - |
| Soft cheese | Food | 3917 | 0.01 | 0.01 | 3.70E-01 | 6.08E-01 | 148 | 3865 | 0.00 | 0.01 | 8.40E-01 | 9.04E-01 | 224 | 0.01 | 0.01 | 4.48E-01 | 0.00 | 6.05E-01 | - | - |
| Medioresinol intake (ug/day) | Nutrients | 9175 | 0.00 | 0.01 | 8.60E-01 | 9.46E-01 | 220 | 8974 | 0.01 | 0.01 | 2.31E-01 | 4.53E-01 | 124 | 0.00 | 0.01 | 4.55E-01 | 0.00 | 3.41E-01 | - | - |
| Cholesterol intake (g/day) | Nutrients | 9175 | 0.00 | 0.01 | 6.24E-01 | 8.36E-01 | 181 | 8974 | 0.00 | 0.01 | 5.76E-01 | 7.40E-01 | 189 | 0.00 | 0.01 | 4.57E-01 | 0.00 | 9.78E-01 | - | - |

| | | | | | | | | | | | | | | | | | | | |
|---|---|---|---|---|---|---|---|---|---|---|---|---|---|---|---|---|---|---|---|
| Do you feel important and appreciated in your home? | Psychosocial | 5576 | -0.01 | 0.01 | 2.77E-01 | 5.26E-01 | 127 | 5429 | 0.00 | 0.01 | 9.90E-01 | 9.90E-01 | 243 | 0.00 | 0.01 | 4.58E-01 | 0.00 | 4.27E-01 | - | - |
| Fitness status | Physical activity | 5601 | 0.00 | 0.01 | 9.60E-01 | 9.88E-01 | 236 | 5461 | -0.01 | 0.01 | 2.98E-01 | 5.28E-01 | 137 | -0.01 | 0.01 | 4.64E-01 | 0.00 | 4.58E-01 | - | - |
| Meat stew | Food | 9175 | 0.01 | 0.01 | 7.39E-02 | 2.99E-01 | 60 | 8974 | 0.00 | 0.01 | 4.89E-01 | 6.68E-01 | 178 | 0.00 | 0.01 | 4.65E-01 | 0.68 | 7.65E-02 | - | - |
| Participation in other association | Social | 668 | 0.15 | 0.07 | 3.60E-02 | 2.19E-01 | 40 | 609 | -0.07 | 0.07 | 3.05E-01 | 5.36E-01 | 138 | 0.04 | 0.05 | 4.67E-01 | 0.80 | 2.62E-02 | - | - |
| Changed everyday exercise during the last year | Physical activity | 9924 | 0.01 | 0.01 | 2.56E-01 | 5.23E-01 | 117 | 9732 | 0.00 | 0.01 | 9.08E-01 | 9.43E-01 | 234 | 0.00 | 0.00 | 4.83E-01 | 0.00 | 3.67E-01 | - | - |
| Bregott on bread | Food | 9175 | 0.01 | 0.01 | 3.57E-01 | 5.91E-01 | 146 | 8974 | 0.00 | 0.01 | 9.28E-01 | 9.57E-01 | 235 | 0.00 | 0.01 | 4.84E-01 | 0.00 | 5.45E-01 | - | - |
| Animal based protein intake (g/day) | Nutrients | 9175 | 0.01 | 0.01 | 3.95E-01 | 6.36E-01 | 151 | 8974 | 0.00 | 0.01 | 8.94E-01 | 9.32E-01 | 233 | 0.00 | 0.01 | 4.97E-01 | 0.00 | 5.98E-01 | - | - |
| Sum of all lignans intake (ug/day) | Nutrients | 9175 | -0.01 | 0.01 | 2.54E-01 | 5.23E-01 | 115 | 8974 | 0.00 | 0.01 | 8.70E-01 | 9.23E-01 | 229 | 0.00 | 0.01 | 5.05E-01 | 0.00 | 3.47E-01 | - | - |
| Contradictory demands in job | Psychosocial | 9724 | 0.00 | 0.01 | 6.95E-01 | 8.61E-01 | 196 | 9523 | 0.00 | 0.01 | 5.98E-01 | 7.61E-01 | 191 | 0.00 | 0.00 | 5.15E-01 | 0.00 | 9.35E-01 | - | - |
| Shellfish (e.g. shrimps, scallops) | Food | 3917 | 0.00 | 0.01 | 9.51E-01 | 9.88E-01 | 233 | 3865 | 0.01 | 0.01 | 3.29E-01 | 5.50E-01 | 144 | 0.00 | 0.01 | 5.25E-01 | 0.00 | 4.58E-01 | - | - |
| Whole grain soft bread | Food | 9175 | -0.01 | 0.01 | 2.90E-01 | 5.27E-01 | 132 | 8974 | 0.00 | 0.01 | 8.92E-01 | 9.32E-01 | 232 | 0.00 | 0.01 | 5.29E-01 | 0.00 | 3.90E-01 | - | - |
| Repetitive job | Psychosocial | 9804 | 0.01 | 0.01 | 2.78E-01 | 5.26E-01 | 128 | 9596 | 0.00 | 0.01 | 8.44E-01 | 9.04E-01 | 226 | 0.00 | 0.00 | 5.41E-01 | 0.00 | 3.58E-01 | - | - |
| Pizza | Food | 9175 | -0.02 | 0.01 | 3.76E-02 | 2.21E-01 | 41 | 8974 | 0.01 | 0.01 | 2.67E-01 | 4.92E-01 | 131 | 0.00 | 0.01 | 5.53E-01 | 0.81 | 2.25E-02 | - | - |
| Smoking status: Former occasional smokers vs non-smokers | Tobacco use | 5654 | 0.03 | 0.02 | 1.61E-01 | 4.13E-01 | 94 | 5455 | -0.01 | 0.02 | 6.03E-01 | 7.64E-01 | 192 | 0.01 | 0.02 | 5.56E-01 | 0.47 | 1.69E-01 | - | - |
| Fatty fish (e.g. herring, whitefish, salmon) | Food | 9175 | 0.01 | 0.01 | 5.97E-02 | 2.74E-01 | 53 | 8974 | -0.01 | 0.01 | 3.41E-01 | 5.53E-01 | 150 | 0.00 | 0.01 | 5.57E-01 | 0.76 | 4.27E-02 | - | - |
| Secoisolariciresinol intake (ug/day) | Nutrients | 9175 | -0.01 | 0.01 | 4.48E-01 | 6.81E-01 | 159 | 8974 | 0.00 | 0.01 | 9.29E-01 | 9.57E-01 | 236 | 0.00 | 0.01 | 5.58E-01 | 0.00 | 6.25E-01 | - | - |
| Cohabitation: Live alone vs Only children | Social | 1304 | -0.01 | 0.05 | 8.20E-01 | 9.35E-01 | 213 | 1308 | 0.04 | 0.04 | 3.29E-01 | 5.50E-01 | 143 | 0.02 | 0.03 | 5.61E-01 | 0.00 | 4.14E-01 | - | - |
| Formic acid intaje (g/day) | Nutrients | 9175 | 0.00 | 0.01 | 8.55E-01 | 9.46E-01 | 218 | 8974 | 0.01 | 0.01 | 3.32E-01 | 5.50E-01 | 146 | 0.00 | 0.01 | 5.64E-01 | 0.00 | 4.24E-01 | - | - |
| Vitamin A intake (mg/day) | Nutrients | 9175 | 0.00 | 0.01 | 8.72E-01 | 9.46E-01 | 224 | 8974 | 0.01 | 0.01 | 3.55E-01 | 5.64E-01 | 153 | 0.00 | 0.01 | 5.71E-01 | 0.00 | 4.55E-01 | - | - |
| Light but partly physically active work | Physical activity | 9772 | 0.00 | 0.02 | 9.50E-01 | 9.88E-01 | 232 | 9546 | -0.01 | 0.02 | 4.10E-01 | 6.14E-01 | 162 | -0.01 | 0.01 | 5.81E-01 | 0.00 | 5.38E-01 | - | - |
| Cream, creme fraiche, sour cream | Food | 9175 | 0.01 | 0.01 | 2.66E-01 | 5.26E-01 | 121 | 8974 | 0.00 | 0.01 | 7.77E-01 | 8.71E-01 | 217 | 0.00 | 0.00 | 5.84E-01 | 0.02 | 3.13E-01 | - | - |
| Distance to work in kilometers (one way) | Physical activity | 8653 | -0.01 | 0.01 | 4.19E-01 | 6.56E-01 | 155 | 8559 | 0.00 | 0.01 | 9.87E-01 | 9.90E-01 | 240 | 0.00 | 0.01 | 5.85E-01 | 0.00 | 5.52E-01 | - | - |
| Frequency of hunting or fishing during leisure time | Physical activity | 9209 | -0.01 | 0.01 | 4.21E-01 | 6.56E-01 | 156 | 9073 | 0.01 | 0.01 | 1.34E-01 | 3.30E-01 | 99 | 0.00 | 0.01 | 5.94E-01 | 0.62 | 1.06E-01 | - | - |
| Low fat milk (0,5%) | Beverage | 9175 | 0.00 | 0.01 | 9.08E-01 | 9.72E-01 | 227 | 8974 | 0.00 | 0.01 | 5.44E-01 | 7.26E-01 | 182 | 0.00 | 0.01 | 6.04E-01 | 0.00 | 7.38E-01 | - | - |
| Last time a colleague visited you at home | Psychosocial | 9690 | 0.00 | 0.01 | 8.45E-01 | 9.46E-01 | 216 | 9451 | 0.00 | 0.01 | 6.09E-01 | 7.67E-01 | 193 | 0.00 | 0.00 | 6.13E-01 | 0.00 | 8.33E-01 | - | - |
| Beta-sitostanol intake (mg/day) | Nutrients | 9175 | 0.00 | 0.01 | 8.08E-01 | 9.26E-01 | 212 | 8974 | 0.01 | 0.01 | 3.63E-01 | 5.72E-01 | 154 | 0.00 | 0.01 | 6.23E-01 | 0.00 | 4.22E-01 | - | - |
| If you exercise, change in exercise habits during the last year | Physical activity | 8657 | 0.01 | 0.01 | 2.61E-01 | 5.26E-01 | 120 | 8516 | 0.00 | 0.01 | 6.78E-01 | 8.15E-01 | 202 | 0.00 | 0.01 | 6.32E-01 | 0.17 | 2.72E-01 | - | - |
| Selenium intake (ug/day) | Nutrients | 9175 | 0.00 | 0.01 | 9.20E-01 | 9.78E-01 | 228 | 8974 | 0.00 | 0.01 | 5.73E-01 | 7.40E-01 | 188 | 0.00 | 0.01 | 6.33E-01 | 0.00 | 7.52E-01 | - | - |
| Work shifts/weekends | Psychosocial | 9611 | 0.03 | 0.02 | 4.97E-02 | 2.55E-01 | 47 | 9412 | -0.02 | 0.02 | 2.27E-01 | 4.49E-01 | 123 | 0.01 | 0.01 | 6.36E-01 | 0.80 | 2.41E-02 | - | - |
| White meat (poultry) | Food | 9175 | 0.00 | 0.01 | 8.83E-01 | 9.50E-01 | 226 | 8974 | -0.01 | 0.01 | 4.29E-01 | 6.25E-01 | 166 | 0.00 | 0.01 | 6.43E-01 | 0.00 | 5.10E-01 | - | - |
| Brewed (filtered) coffee | Beverage | 9175 | 0.00 | 0.01 | 7.53E-01 | 8.80E-01 | 208 | 8974 | 0.00 | 0.01 | 7.44E-01 | 8.64E-01 | 209 | 0.00 | 0.00 | 6.50E-01 | 0.00 | 9.97E-01 | - | - |
| Job demands to work very fast | Psychosocial | 9801 | 0.00 | 0.01 | 7.27E-01 | 8.74E-01 | 202 | 9596 | 0.00 | 0.01 | 7.77E-01 | 8.71E-01 | 216 | 0.00 | 0.00 | 6.56E-01 | 0.00 | 9.55E-01 | - | - |
| Enterodiol intake (ug/day) | Nutrients | 9175 | 0.00 | 0.01 | 9.64E-01 | 9.88E-01 | 237 | 8974 | 0.00 | 0.01 | 5.73E-01 | 7.40E-01 | 187 | 0.00 | 0.01 | 6.59E-01 | 0.00 | 7.23E-01 | - | - |
| Frequent social contacts with colleagues during work | Psychosocial | 9721 | 0.01 | 0.01 | 1.22E-01 | 3.80E-01 | 78 | 9502 | -0.01 | 0.01 | 3.91E-01 | 5.98E-01 | 159 | 0.00 | 0.00 | 6.62E-01 | 0.66 | 8.65E-02 | - | - |
| Liver, kidney | Food | 3917 | -0.02 | 0.01 | 1.56E-01 | 4.08E-01 | 92 | 3865 | 0.01 | 0.01 | 4.17E-01 | 6.18E-01 | 164 | 0.00 | 0.01 | 6.92E-01 | 0.60 | 1.13E-01 | - | - |
| Margarine for cooking | Food | 9175 | 0.01 | 0.01 | 1.79E-01 | 4.40E-01 | 99 | 8974 | -0.01 | 0.01 | 4.51E-01 | 6.49E-01 | 169 | 0.00 | 0.01 | 7.03E-01 | 0.55 | 1.36E-01 | - | - |
| Steak, chop, e.g. | Food | 9175 | 0.00 | 0.01 | 5.37E-01 | 7.63E-01 | 171 | 8974 | 0.00 | 0.01 | 9.38E-01 | 9.61E-01 | 237 | 0.00 | 0.01 | 7.17E-01 | 0.00 | 6.13E-01 | - | - |
| Sometimes physically straining work | Physical activity | 9772 | -0.02 | 0.02 | 2.68E-01 | 5.26E-01 | 122 | 9546 | 0.01 | 0.01 | 5.57E-01 | 7.32E-01 | 184 | 0.00 | 0.01 | 7.27E-01 | 0.31 | 2.29E-01 | - | - |
| Mixed frozen vegetables | Food | 3917 | 0.01 | 0.01 | 4.01E-01 | 6.37E-01 | 153 | 3865 | 0.00 | 0.01 | 7.54E-01 | 8.64E-01 | 212 | 0.00 | 0.01 | 7.27E-01 | 0.00 | 4.09E-01 | - | - |
| Light and physically active work | Physical activity | 9772 | -0.01 | 0.02 | 5.84E-01 | 8.06E-01 | 176 | 9546 | 0.00 | 0.02 | 9.63E-01 | 9.79E-01 | 239 | 0.00 | 0.01 | 7.29E-01 | 0.00 | 6.70E-01 | - | - |
| Frequncy of social contacts with colleagues during leisure time | Psychosocial | 9571 | 0.00 | 0.01 | 7.85E-01 | 9.09E-01 | 209 | 9344 | 0.00 | 0.01 | 4.65E-01 | 6.53E-01 | 172 | 0.00 | 0.00 | 7.32E-01 | 0.00 | 4.83E-01 | - | - |
| Number of people with whom you can speak openly | Social | 9895 | 0.00 | 0.01 | 9.86E-01 | 9.96E-01 | 240 | 9713 | 0.00 | 0.01 | 6.47E-01 | 7.90E-01 | 199 | 0.00 | 0.00 | 7.49E-01 | 0.00 | 7.42E-01 | - | - |
| Vitamin C intake (mg/day) | Nutrients | 9175 | 0.00 | 0.01 | 6.00E-01 | 8.16E-01 | 178 | 8974 | -0.01 | 0.01 | 3.48E-01 | 5.60E-01 | 151 | 0.00 | 0.01 | 7.55E-01 | 0.06 | 3.03E-01 | - | - |
| Vitamin B6 intake (mg/day) | Nutrients | 9175 | 0.01 | 0.01 | 5.83E-02 | 2.72E-01 | 52 | 8974 | -0.01 | 0.01 | 1.59E-01 | 3.54E-01 | 109 | 0.00 | 0.01 | 7.84E-01 | 0.82 | 1.90E-02 | - | - |
| Everyday exercise satisfaction | Physical activity | 9892 | 0.00 | 0.01 | 9.80E-01 | 9.96E-01 | 238 | 9704 | 0.00 | 0.01 | 7.48E-01 | 8.64E-01 | 210 | 0.00 | 0.00 | 8.04E-01 | 0.00 | 8.36E-01 | - | - |
| Participation in sports or physical exercise associations | Social | 668 | -0.07 | 0.07 | 2.99E-01 | 5.35E-01 | 136 | 609 | 0.05 | 0.07 | 4.76E-01 | 6.61E-01 | 175 | -0.01 | 0.05 | 8.29E-01 | 0.35 | 2.14E-01 | - | - |
| Sedentary or standing work | Physical activity | 9772 | 0.02 | 0.02 | 1.30E-01 | 3.83E-01 | 82 | 9546 | -0.02 | 0.02 | 2.44E-01 | 4.63E-01 | 128 | 0.00 | 0.01 | 8.36E-01 | 0.72 | 5.75E-02 | - | - |
| Frequency of engaging in clubs, associations or study circles | Social | 6577 | 0.00 | 0.01 | 7.35E-01 | 8.74E-01 | 204 | 6453 | 0.00 | 0.01 | 9.63E-01 | 9.79E-01 | 238 | 0.00 | 0.01 | 8.41E-01 | 0.00 | 7.83E-01 | - | - |
| Tomato, cucumber | Food | 9175 | 0.00 | 0.01 | 5.45E-01 | 7.70E-01 | 172 | 8974 | -0.01 | 0.01 | 3.94E-01 | 5.99E-01 | 160 | 0.00 | 0.01 | 8.46E-01 | 0.05 | 3.05E-01 | - | - |
| Pasta | Food | 9175 | 0.00 | 0.01 | 7.00E-01 | 8.61E-01 | 197 | 8974 | 0.00 | 0.01 | 5.40E-01 | 7.24E-01 | 181 | 0.00 | 0.01 | 8.50E-01 | 0.00 | 4.84E-01 | - | - |
| Disaccharides intake (g/day) | Nutrients | 9175 | 0.00 | 0.01 | 8.57E-01 | 9.46E-01 | 219 | 8974 | 0.00 | 0.01 | 6.96E-01 | 8.30E-01 | 204 | 0.00 | 0.01 | 8.75E-01 | 0.00 | 6.89E-01 | - | - |
| High mental demand from job | Psychosocial | 9733 | 0.00 | 0.01 | 9.31E-01 | 9.84E-01 | 230 | 9524 | 0.00 | 0.01 | 7.86E-01 | 8.76E-01 | 218 | 0.00 | 0.00 | 8.91E-01 | 0.00 | 8.03E-01 | - | - |
| Average portion size of meat/fish based on photographic illustration of four sizes (smallest to largest) | Food | 9175 | 0.01 | 0.01 | 1.95E-01 | 4.74E-01 | 100 | 8974 | -0.01 | 0.01 | 1.57E-01 | 3.52E-01 | 108 | 0.00 | 0.01 | 8.93E-01 | 0.73 | 5.55E-02 | - | - |
| Syringaresinol intake (ug/day) | Nutrients | 9175 | -0.01 | 0.01 | 3.51E-01 | 5.88E-01 | 145 | 8974 | 0.00 | 0.01 | 4.83E-01 | 6.64E-01 | 177 | 0.00 | 0.01 | 8.94E-01 | 0.26 | 2.47E-01 | - | - |
| Possibility to speak with colleagues during breaks | Psychosocial | 9790 | 0.00 | 0.01 | 6.54E-01 | 8.53E-01 | 186 | 9565 | 0.00 | 0.01 | 8.06E-01 | 8.95E-01 | 219 | 0.00 | 0.00 | 8.95E-01 | 0.00 | 6.22E-01 | - | - |
| Campestanol intake (mg/day) | Nutrients | 9175 | -0.01 | 0.01 | 3.37E-01 | 5.77E-01 | 142 | 8974 | 0.01 | 0.01 | 2.81E-01 | 5.06E-01 | 135 | 0.00 | 0.01 | 9.03E-01 | 0.52 | 1.50E-01 | - | - |
| Trans fat intake (g/day) | Nutrients | 9175 | 0.00 | 0.01 | 1.00E+00 | 1.00E+00 | 243 | 8974 | 0.00 | 0.01 | 8.76E-01 | 9.23E-01 | 230 | 0.00 | 0.01 | 9.09E-01 | 0.00 | 9.15E-01 | - | - |
| Total protein intake (g/day) | Nutrients | 9175 | 0.00 | 0.01 | 8.66E-01 | 9.46E-01 | 222 | 8974 | 0.00 | 0.01 | 9.89E-01 | 9.90E-01 | 241 | 0.00 | 0.01 | 9.16E-01 | 0.00 | 8.96E-01 | - | - |
| Sweets | Food | 9175 | 0.00 | 0.01 | 9.22E-01 | 9.78E-01 | 229 | 8974 | 0.00 | 0.01 | 8.20E-01 | 9.02E-01 | 221 | 0.00 | 0.01 | 9.26E-01 | 0.00 | 8.19E-01 | - | - |
| Sucrose intake (g/day) | Nutrients | 9175 | 0.00 | 0.01 | 6.87E-01 | 8.61E-01 | 194 | 8974 | 0.00 | 0.01 | 7.76E-01 | 8.71E-01 | 215 | 0.00 | 0.01 | 9.41E-01 | 0.00 | 6.26E-01 | - | - |
| Cheese 10-17% | Food | 9175 | 0.01 | 0.01 | 3.29E-01 | 5.67E-01 | 141 | 8974 | -0.01 | 0.01 | 4.09E-01 | 6.14E-01 | 161 | 0.00 | 0.01 | 9.52E-01 | 0.39 | 2.02E-01 | - | - |
| Boiled or baked potato | Food | 9175 | 0.01 | 0.01 | 9.35E-02 | 3.25E-01 | 70 | 8974 | -0.01 | 0.01 | 9.93E-02 | 2.81E-01 | 86 | 0.00 | 0.01 | 9.58E-01 | 0.82 | 1.87E-02 | - | - |
| Sugar, honey, marmelade, jam | Food | 9175 | 0.00 | 0.01 | 6.60E-01 | 8.53E-01 | 188 | 8974 | 0.00 | 0.01 | 6.24E-01 | 7.78E-01 | 194 | 0.00 | 0.00 | 9.58E-01 | 0.00 | 5.12E-01 | - | - |
| Participation in study circles | Social | 668 | -0.04 | 0.11 | 7.05E-01 | 8.61E-01 | 198 | 609 | 0.04 | 0.10 | 6.75E-01 | 8.15E-01 | 201 | 0.00 | 0.07 | 9.59E-01 | 0.00 | 5.74E-01 | - | - |
| Cambridge physical activity index | Physical activity | 9753 | 0.00 | 0.01 | 5.32E-01 | 7.63E-01 | 168 | 9534 | 0.00 | 0.01 | 5.19E-01 | 7.01E-01 | 180 | 0.00 | 0.00 | 9.74E-01 | 0.00 | 3.70E-01 | - | - |
| Do you feel important and appreciated outside your home? | Psychosocial | 5616 | 0.00 | 0.01 | 6.33E-01 | 8.36E-01 | 184 | 5475 | 0.00 | 0.01 | 6.32E-01 | 7.78E-01 | 197 | 0.00 | 0.01 | 9.74E-01 | 0.00 | 4.99E-01 | - | - |
| Smoking status: Former smokers vs non-smokers | Tobacco use | 6747 | 0.00 | 0.02 | 8.64E-01 | 9.46E-01 | 221 | 6641 | 0.00 | 0.02 | 8.38E-01 | 9.04E-01 | 222 | 0.00 | 0.01 | 9.75E-01 | 0.00 | 7.91E-01 | - | - |
| Lean fish (e.g. perch, bass, cod) | Food | 9175 | 0.00 | 0.01 | 5.21E-01 | 7.59E-01 | 167 | 8974 | 0.00 | 0.01 | 5.65E-01 | 7.38E-01 | 186 | 0.00 | 0.01 | 9.77E-01 | 0.00 | 3.89E-01 | - | - |
| Snuff status: Former snuff users vs non-snuff users | Tobacco use | 7675 | -0.01 | 0.02 | 7.45E-01 | 8.74E-01 | 207 | 7548 | 0.01 | 0.02 | 7.53E-01 | 8.64E-01 | 211 | 0.00 | 0.02 | 9.96E-01 | 0.00 | 6.51E-01 | - | - |

**Supplementary Table 17**. Longitudinal association results for HDL cholesterol

| | | Training set | | | | | | Testing set | | | | | | Metanalysis | | | | |
|---|---|---|---|---|---|---|---|---|---|---|---|---|---|---|---|---|---|---|
| **Description** | **Group** | **N** | **Effect estimate** | **S.E.** | **p-value** | **p-value$_{FDR}$** | **p-value rank** | **N** | **Effect estimate** | **S.E.** | **p-value** | **p-value$_{FDR}$** | **p-value rank** | **Effect estimate** | **S.E.** | **p-value** | **$I^2$** | **Q p-value** |
| Milk, sour milk (1,5%) | Beverage | 1799 | -0.03 | 0.01 | 2.73E-03 | 1.31E-01 | 5 | 1674 | -0.02 | 0.01 | 2.58E-02 | 4.11E-01 | 14 | -0.02 | 0.01 | 2.06E-04 | 0.00 | 6.49E-01 |
| Fitness status | Physical activity | 1232 | 0.03 | 0.01 | 1.09E-02 | 2.37E-01 | 11 | 1185 | 0.03 | 0.01 | 1.43E-02 | 4.11E-01 | 7 | 0.03 | 0.01 | 4.02E-04 | 0.00 | 9.78E-01 |
| Alcohol intake (g/day) | Alcohol | 1799 | 0.03 | 0.01 | 1.07E-03 | 9.55E-02 | 1 | 1674 | 0.02 | 0.01 | 1.52E-01 | 6.37E-01 | 56 | 0.02 | 0.01 | 6.09E-04 | 0.04 | 3.06E-01 |
| Disaccharides intake (g/day) | Nutrients | 1799 | -0.01 | 0.01 | 1.20E-01 | 6.15E-01 | 44 | 1674 | -0.03 | 0.01 | 3.43E-03 | 4.10E-01 | 2 | -0.02 | 0.01 | 1.54E-03 | 0.00 | 3.24E-01 |
| Wine | Alcohol | 1799 | 0.03 | 0.01 | 1.45E-03 | 9.55E-02 | 2 | 1674 | 0.01 | 0.01 | 2.35E-01 | 7.57E-01 | 74 | 0.02 | 0.01 | 1.60E-03 | 0.38 | 2.03E-01 |
| Medium beer | Alcohol | 1799 | 0.03 | 0.01 | 4.32E-03 | 1.62E-01 | 6 | 1674 | 0.01 | 0.01 | 2.06E-01 | 7.15E-01 | 68 | 0.02 | 0.01 | 2.47E-03 | 0.00 | 4.40E-01 |
| Vitamin B3 intake (mg/day) | Nutrients | 1799 | 0.01 | 0.01 | 2.50E-01 | 7.20E-01 | 83 | 1674 | 0.03 | 0.01 | 1.52E-03 | 3.63E-01 | 1 | 0.02 | 0.01 | 2.48E-03 | 0.56 | 1.33E-01 |
| Sucrose intake (g/day) | Nutrients | 1799 | -0.01 | 0.01 | 1.19E-01 | 6.15E-01 | 43 | 1674 | -0.02 | 0.01 | 8.71E-03 | 4.11E-01 | 4 | -0.02 | 0.01 | 3.16E-03 | 0.00 | 4.31E-01 |
| Sour milk, yoghurt (low fat) | Food | 1799 | 0.02 | 0.01 | 3.68E-02 | 4.63E-01 | 19 | 1674 | 0.02 | 0.01 | 7.08E-02 | 5.03E-01 | 31 | 0.02 | 0.01 | 5.77E-03 | 0.00 | 9.06E-01 |
| Secoisolariciresinol intake (ug/day) | Nutrients | 1799 | 0.03 | 0.01 | 1.60E-03 | 9.55E-02 | 4 | 1674 | 0.01 | 0.01 | 4.73E-01 | 9.29E-01 | 119 | 0.02 | 0.01 | 6.04E-03 | 0.66 | 8.48E-02 |
| Cohabitation: Live alone vs Only children | Social | 258 | -0.05 | 0.05 | 3.26E-01 | 7.43E-01 | 105 | 255 | -0.11 | 0.04 | 7.75E-03 | 4.11E-01 | 3 | -0.09 | 0.03 | 6.49E-03 | 0.00 | 3.82E-01 |
| Years smoking | Tobacco use | 1714 | -0.01 | 0.01 | 1.36E-01 | 6.15E-01 | 52 | 1612 | -0.02 | 0.01 | 3.34E-02 | 4.42E-01 | 16 | -0.02 | 0.01 | 1.07E-02 | 0.00 | 6.20E-01 |
| Hearing status | General health | 1238 | 0.02 | 0.01 | 8.96E-02 | 6.15E-01 | 32 | 1188 | 0.02 | 0.01 | 8.00E-02 | 5.24E-01 | 36 | 0.02 | 0.01 | 1.47E-02 | 0.00 | 9.48E-01 |
| Number of cigarretes smoked per day (in groups) | Tobacco use | 1594 | -0.01 | 0.01 | 1.37E-01 | 6.15E-01 | 53 | 1512 | -0.02 | 0.01 | 5.84E-02 | 4.99E-01 | 28 | -0.02 | 0.01 | 1.66E-02 | 0.00 | 7.88E-01 |
| Tea | Beverage | 1799 | -0.01 | 0.01 | 1.19E-01 | 6.15E-01 | 42 | 1674 | -0.02 | 0.01 | 6.62E-02 | 5.03E-01 | 29 | -0.02 | 0.01 | 1.71E-02 | 0.00 | 7.15E-01 |
| Smoking status: Smokers vs non-smokers | Tobacco use | 1273 | -0.03 | 0.03 | 2.87E-01 | 7.20E-01 | 94 | 1217 | -0.06 | 0.03 | 2.40E-02 | 4.11E-01 | 13 | -0.04 | 0.02 | 1.82E-02 | 0.00 | 4.14E-01 |
| Equol intake (ug/day) | Nutrients | 1799 | 0.02 | 0.01 | 2.24E-02 | 3.83E-01 | 14 | 1674 | 0.01 | 0.01 | 3.03E-01 | 8.14E-01 | 89 | 0.02 | 0.01 | 1.86E-02 | 0.00 | 3.89E-01 |
| Strong beer | Alcohol | 1799 | 0.01 | 0.01 | 2.86E-01 | 7.20E-01 | 92 | 1674 | 0.03 | 0.01 | 2.05E-02 | 4.11E-01 | 10 | 0.01 | 0.01 | 2.66E-02 | 0.37 | 2.06E-01 |
| Bacon | Food | 1799 | 0.01 | 0.01 | 4.55E-01 | 8.50E-01 | 128 | 1674 | 0.02 | 0.01 | 1.53E-02 | 4.11E-01 | 8 | 0.01 | 0.01 | 3.14E-02 | 0.45 | 1.77E-01 |
| Light and physically active work | Physical activity | 1910 | 0.03 | 0.02 | 1.92E-01 | 6.39E-01 | 72 | 1786 | 0.04 | 0.02 | 8.12E-02 | 5.24E-01 | 37 | 0.03 | 0.01 | 3.19E-02 | 0.00 | 7.06E-01 |
| Informed of having high blood pressure | General health | 1943 | -0.04 | 0.02 | 7.81E-02 | 6.15E-01 | 29 | 1826 | -0.03 | 0.02 | 2.17E-01 | 7.20E-01 | 72 | -0.04 | 0.02 | 3.35E-02 | 0.00 | 7.37E-01 |
| Frequent social contacts with colleagues during work | Psychosocial | 1900 | 0.02 | 0.01 | 1.38E-02 | 2.74E-01 | 12 | 1784 | 0.00 | 0.01 | 6.31E-01 | 9.71E-01 | 153 | 0.01 | 0.01 | 3.63E-02 | 0.48 | 1.65E-01 |
| Last time a colleague visited you at home | Psychosocial | 1897 | 0.02 | 0.01 | 4.76E-03 | 1.62E-01 | 7 | 1769 | 0.00 | 0.01 | 9.19E-01 | 9.89E-01 | 221 | 0.01 | 0.01 | 3.75E-02 | 0.73 | 5.54E-02 |
| Self-employed | Psychosocial | 1906 | 0.09 | 0.03 | 1.46E-03 | 9.55E-02 | 3 | 1804 | -0.01 | 0.03 | 7.12E-01 | 9.71E-01 | 172 | 0.04 | 0.02 | 3.98E-02 | 0.84 | 1.38E-02 |
| Exercise during the last three months | Physical activity | 1941 | 0.00 | 0.01 | 6.03E-01 | 9.14E-01 | 157 | 1825 | 0.02 | 0.01 | 1.41E-02 | 4.11E-01 | 6 | 0.01 | 0.01 | 4.05E-02 | 0.53 | 1.46E-01 |
| Eicosapentaenoic acid (EPA) intake (g/day) | Nutrients | 1799 | 0.01 | 0.01 | 5.02E-01 | 8.88E-01 | 133 | 1674 | 0.02 | 0.01 | 3.69E-02 | 4.42E-01 | 19 | 0.01 | 0.01 | 4.82E-02 | 0.00 | 3.40E-01 |
| Would you say that the number of people that you meet in your everyday life is enough or would you like to meet more or fewer people? | Social | 1935 | 0.01 | 0.01 | 2.31E-01 | 6.89E-01 | 80 | 1823 | 0.01 | 0.01 | 1.19E-01 | 5.82E-01 | 49 | 0.01 | 0.01 | 5.25E-02 | 0.00 | 7.43E-01 |
| Docosahexaenoic acid (DHA) intake (g/day) | Nutrients | 1799 | 0.01 | 0.01 | 5.88E-01 | 9.14E-01 | 153 | 1674 | 0.02 | 0.01 | 3.57E-02 | 4.42E-01 | 17 | 0.01 | 0.01 | 5.97E-02 | 0.14 | 2.80E-01 |
| Years using snuff | Tobacco use | 1789 | 0.01 | 0.01 | 1.80E-01 | 6.19E-01 | 69 | 1655 | 0.01 | 0.01 | 1.85E-01 | 6.93E-01 | 64 | 0.01 | 0.01 | 6.00E-02 | 0.00 | 9.01E-01 |
| Magnesium intake (mg/day) | Nutrients | 1799 | 0.02 | 0.01 | 2.61E-02 | 3.90E-01 | 16 | 1674 | 0.00 | 0.01 | 7.02E-01 | 9.71E-01 | 168 | 0.01 | 0.01 | 6.01E-02 | 0.36 | 2.10E-01 |
| Repetitive job | Psychosocial | 1912 | -0.02 | 0.01 | 4.47E-02 | 5.34E-01 | 20 | 1799 | -0.01 | 0.01 | 5.66E-01 | 9.29E-01 | 143 | -0.01 | 0.01 | 6.21E-02 | 0.00 | 3.47E-01 |
| Matairesinol intake (ug/day) | Nutrients | 1799 | 0.02 | 0.01 | 4.95E-02 | 5.63E-01 | 21 | 1674 | 0.01 | 0.01 | 5.39E-01 | 9.29E-01 | 136 | 0.01 | 0.01 | 6.33E-02 | 0.00 | 3.73E-01 |
| Learn new things at job | Psychosocial | 1910 | 0.02 | 0.01 | 6.62E-02 | 6.15E-01 | 23 | 1791 | 0.01 | 0.01 | 4.70E-01 | 9.29E-01 | 118 | 0.01 | 0.01 | 6.61E-02 | 0.00 | 4.69E-01 |
| Frequency of cycling during leisure time | Physical activity | 1575 | 0.01 | 0.01 | 4.24E-01 | 8.18E-01 | 124 | 1472 | 0.02 | 0.01 | 6.72E-02 | 5.03E-01 | 30 | 0.01 | 0.01 | 6.70E-02 | 0.00 | 4.24E-01 |
| Sodium intake (mg/day) | Nutrients | 1799 | 0.00 | 0.01 | 6.90E-01 | 9.33E-01 | 176 | 1674 | 0.02 | 0.01 | 2.58E-02 | 4.11E-01 | 15 | 0.01 | 0.01 | 6.76E-02 | 0.44 | 1.80E-01 |
| Fatty fish (e.g. herring, whitefish, salmon) | Food | 1799 | 0.01 | 0.01 | 2.13E-01 | 6.53E-01 | 78 | 1674 | 0.01 | 0.01 | 1.79E-01 | 6.78E-01 | 63 | 0.01 | 0.01 | 6.79E-02 | 0.00 | 8.71E-01 |
| Close relationship with anyone | Social | 1937 | 0.02 | 0.01 | 8.47E-03 | 2.11E-01 | 9 | 1826 | 0.00 | 0.01 | 9.67E-01 | 9.90E-01 | 232 | 0.01 | 0.01 | 6.95E-02 | 0.73 | 5.60E-02 |
| Number of social contacts with the same interests as you | Social | 1936 | 0.01 | 0.01 | 1.50E-01 | 6.15E-01 | 57 | 1821 | 0.01 | 0.01 | 2.63E-01 | 7.68E-01 | 81 | 0.01 | 0.01 | 6.99E-02 | 0.00 | 8.43E-01 |
| Cambridge physical activity index | Physical activity | 1904 | 0.00 | 0.01 | 9.06E-01 | 9.95E-01 | 216 | 1784 | 0.02 | 0.01 | 1.56E-02 | 4.11E-01 | 9 | 0.01 | 0.01 | 7.46E-02 | 0.63 | 1.01E-01 |
| Cohabitation: Live alone vs Other/others | Social | 238 | 0.09 | 0.06 | 1.06E-01 | 6.15E-01 | 36 | 195 | 0.05 | 0.05 | 3.66E-01 | 9.16E-01 | 95 | 0.07 | 0.04 | 7.56E-02 | 0.00 | 5.88E-01 |
| Steak, chop, e.g. | Food | 1799 | 0.00 | 0.01 | 7.76E-01 | 9.38E-01 | 197 | 1674 | 0.02 | 0.01 | 2.23E-02 | 4.11E-01 | 12 | 0.01 | 0.01 | 8.67E-02 | 0.58 | 1.23E-01 |
| Rosehip, sweet syrup soup | Food | 1799 | -0.01 | 0.01 | 3.11E-01 | 7.31E-01 | 100 | 1674 | -0.01 | 0.01 | 1.75E-01 | 6.75E-01 | 62 | -0.01 | 0.01 | 9.55E-02 | 0.00 | 7.64E-01 |
| Vitamin B6 intake (mg/day) | Nutrients | 1799 | 0.00 | 0.01 | 7.33E-01 | 9.35E-01 | 187 | 1674 | 0.02 | 0.01 | 5.45E-02 | 4.99E-01 | 25 | 0.01 | 0.01 | 1.16E-01 | 0.26 | 2.46E-01 |
| Tiamin intake (mg/day) | Nutrients | 1799 | 0.01 | 0.01 | 2.10E-01 | 6.51E-01 | 77 | 1674 | 0.01 | 0.01 | 3.51E-01 | 8.92E-01 | 94 | 0.01 | 0.01 | 1.22E-01 | 0.00 | 8.24E-01 |
| Boiled coffee | Beverage | 1799 | 0.01 | 0.01 | 4.90E-01 | 8.88E-01 | 130 | 1674 | 0.01 | 0.01 | 1.34E-01 | 5.95E-01 | 53 | 0.01 | 0.01 | 1.23E-01 | 0.00 | 5.55E-01 |
| Coffee rolls/buns, rusk | Food | 1799 | -0.01 | 0.01 | 2.99E-01 | 7.31E-01 | 97 | 1674 | -0.01 | 0.01 | 2.63E-01 | 7.68E-01 | 80 | -0.01 | 0.01 | 1.27E-01 | 0.00 | 9.43E-01 |
| Everyday exercise satisfaction | Physical activity | 1939 | 0.02 | 0.01 | 2.13E-02 | 3.83E-01 | 13 | 1822 | 0.00 | 0.01 | 8.63E-01 | 9.74E-01 | 208 | 0.01 | 0.01 | 1.27E-01 | 0.67 | 8.28E-02 |
| Light but partly physically active work | Physical activity | 1910 | -0.01 | 0.02 | 6.08E-01 | 9.14E-01 | 159 | 1786 | -0.04 | 0.02 | 1.02E-01 | 5.42E-01 | 43 | -0.03 | 0.02 | 1.31E-01 | 0.00 | 4.14E-01 |
| Average portion size of meat/fish based on photographic illustration of four sizes (smallest to largest) | Food | 1799 | 0.00 | 0.01 | 7.22E-01 | 9.35E-01 | 184 | 1674 | 0.03 | 0.01 | 9.64E-03 | 4.11E-01 | 5 | 0.01 | 0.01 | 1.32E-01 | 0.78 | 3.26E-02 |
| Energy status | Psychosocial | 1235 | 0.02 | 0.01 | 1.55E-01 | 6.15E-01 | 59 | 1185 | 0.01 | 0.01 | 5.13E-01 | 9.29E-01 | 126 | 0.01 | 0.01 | 1.39E-01 | 0.00 | 6.04E-01 |
| Educational level | Psychosocial | 1938 | 0.02 | 0.01 | 7.04E-02 | 6.15E-01 | 24 | 1828 | 0.00 | 0.01 | 8.23E-01 | 9.74E-01 | 197 | 0.01 | 0.01 | 1.41E-01 | 0.14 | 2.82E-01 |
| Grams of tobacco smoked per week | Tobacco use | 1146 | 0.01 | 0.02 | 6.54E-01 | 9.30E-01 | 166 | 1080 | 0.03 | 0.02 | 9.26E-02 | 5.40E-01 | 39 | 0.02 | 0.01 | 1.44E-01 | 0.00 | 3.44E-01 |
| Number of cigarretes smoked per day | Tobacco use | 1146 | 0.01 | 0.02 | 6.54E-01 | 9.30E-01 | 167 | 1080 | 0.03 | 0.02 | 9.26E-02 | 5.40E-01 | 40 | 0.02 | 0.01 | 1.44E-01 | 0.00 | 3.44E-01 |
| Number of cigars smoked per day | Tobacco use | 1146 | 0.01 | 0.02 | 6.54E-01 | 9.30E-01 | 168 | 1080 | 0.03 | 0.02 | 9.26E-02 | 5.40E-01 | 41 | 0.02 | 0.01 | 1.44E-01 | 0.00 | 3.44E-01 |
| Meat stew | Food | 1799 | 0.01 | 0.01 | 1.73E-01 | 6.19E-01 | 65 | 1674 | 0.01 | 0.01 | 5.23E-01 | 9.29E-01 | 131 | 0.01 | 0.01 | 1.48E-01 | 0.00 | 6.74E-01 |
| Sum of all lignans intake (ug/day) | Nutrients | 1799 | 0.01 | 0.01 | 1.00E-01 | 6.15E-01 | 34 | 1674 | 0.00 | 0.01 | 7.15E-01 | 9.71E-01 | 173 | 0.01 | 0.01 | 1.48E-01 | 0.00 | 3.87E-01 |
| Appetite status | Psychosocial | 1235 | 0.01 | 0.01 | 4.13E-01 | 8.18E-01 | 118 | 1186 | 0.01 | 0.01 | 2.17E-01 | 7.20E-01 | 71 | 0.01 | 0.01 | 1.50E-01 | 0.00 | 7.20E-01 |
| White meat (poultry) | Food | 1799 | 0.01 | 0.01 | 3.05E-01 | 7.31E-01 | 98 | 1674 | 0.01 | 0.01 | 3.17E-01 | 8.42E-01 | 90 | 0.01 | 0.01 | 1.52E-01 | 0.00 | 9.94E-01 |
| Syringaresinol intake (ug/day) | Nutrients | 1799 | 0.01 | 0.01 | 1.34E-01 | 6.15E-01 | 51 | 1674 | 0.00 | 0.01 | 6.36E-01 | 9.71E-01 | 155 | 0.01 | 0.01 | 1.57E-01 | 0.00 | 4.96E-01 |
| Cookies, pastry | Food | 1799 | 0.00 | 0.01 | 6.77E-01 | 9.30E-01 | 173 | 1674 | -0.02 | 0.01 | 9.89E-02 | 5.42E-01 | 42 | -0.01 | 0.01 | 1.60E-01 | 0.00 | 3.35E-01 |
| Job demands to work very fast | Psychosocial | 1910 | 0.00 | 0.01 | 9.81E-01 | 9.95E-01 | 231 | 1795 | 0.02 | 0.01 | 4.49E-02 | 4.88E-01 | 22 | 0.01 | 0.01 | 1.61E-01 | 0.51 | 1.51E-01 |
| Medioresinol intake (ug/day) | Nutrients | 1799 | 0.01 | 0.01 | 1.65E-01 | 6.15E-01 | 64 | 1674 | 0.01 | 0.01 | 5.72E-01 | 9.29E-01 | 147 | 0.01 | 0.01 | 1.62E-01 | 0.00 | 5.89E-01 |
| Beta-sitostanol intake (mg/day) | Nutrients | 1799 | 0.01 | 0.01 | 1.33E-01 | 6.15E-01 | 50 | 1674 | 0.00 | 0.01 | 6.65E-01 | 9.71E-01 | 163 | 0.01 | 0.01 | 1.63E-01 | 0.00 | 4.80E-01 |
| Iron intake (mg/day) | Nutrients | 1799 | 0.01 | 0.01 | 2.47E-01 | 7.19E-01 | 82 | 1674 | 0.01 | 0.01 | 4.31E-01 | 9.29E-01 | 108 | 0.01 | 0.01 | 1.66E-01 | 0.00 | 8.39E-01 |
| Satisfaction with home and family situation | Psychosocial | 1234 | 0.01 | 0.01 | 2.86E-01 | 7.20E-01 | 93 | 1187 | 0.01 | 0.01 | 3.75E-01 | 9.22E-01 | 97 | 0.01 | 0.01 | 1.66E-01 | 0.00 | 9.18E-01 |
| Salty fish | Food | 1799 | 0.00 | 0.01 | 7.46E-01 | 9.35E-01 | 188 | 1674 | 0.02 | 0.01 | 3.63E-02 | 4.42E-01 | 18 | 0.01 | 0.01 | 1.69E-01 | 0.62 | 1.06E-01 |
| Parents or siblings had a cerebral hemorrhage/thrombosis or cardiac infarction before the age of 60 | General health | 1924 | -0.01 | 0.02 | 6.90E-01 | 9.33E-01 | 175 | 1806 | -0.03 | 0.02 | 1.25E-01 | 5.95E-01 | 50 | -0.02 | 0.02 | 1.74E-01 | 0.00 | 4.15E-01 |
| People to ask for help apart from the ones at home | Social | 1933 | -0.03 | 0.03 | 2.64E-01 | 7.20E-01 | 87 | 1822 | -0.02 | 0.03 | 4.31E-01 | 9.29E-01 | 107 | -0.03 | 0.02 | 1.77E-01 | 0.00 | 8.28E-01 |
| Teetotaler | Alcohol | 1934 | 0.00 | 0.03 | 9.75E-01 | 9.95E-01 | 230 | 1825 | -0.06 | 0.03 | 5.53E-02 | 4.99E-01 | 26 | -0.03 | 0.02 | 1.78E-01 | 0.46 | 1.72E-01 |
| High physical demand from job | Physical activity | 1918 | 0.00 | 0.01 | 8.96E-01 | 9.95E-01 | 211 | 1802 | 0.02 | 0.01 | 7.16E-02 | 5.03E-01 | 34 | 0.01 | 0.01 | 1.82E-01 | 0.33 | 2.23E-01 |
| Meat on bread | Food | 1799 | -0.02 | 0.01 | 2.61E-02 | 3.90E-01 | 15 | 1674 | 0.01 | 0.01 | 5.40E-01 | 9.29E-01 | 137 | -0.01 | 0.01 | 1.90E-01 | 0.72 | 5.72E-02 |
| Pentadecanoic acid intake (g/day) | Nutrients | 1799 | 0.00 | 0.01 | 7.55E-01 | 9.35E-01 | 192 | 1674 | -0.02 | 0.01 | 1.07E-01 | 5.42E-01 | 46 | -0.01 | 0.01 | 1.92E-01 | 0.00 | 3.16E-01 |
| Heptadecanoic acid intake (g/day) | Nutrients | 1799 | 0.00 | 0.01 | 7.55E-01 | 9.35E-01 | 193 | 1674 | -0.02 | 0.01 | 1.07E-01 | 5.42E-01 | 47 | -0.01 | 0.01 | 1.92E-01 | 0.00 | 3.16E-01 |
| Vitamin C intake (mg/day) | Nutrients | 1799 | 0.00 | 0.01 | 7.71E-01 | 9.38E-01 | 196 | 1674 | -0.03 | 0.01 | 2.22E-02 | 4.11E-01 | 11 | -0.01 | 0.01 | 1.93E-01 | 0.72 | 5.67E-02 |
| Hamburger | Food | 1799 | -0.02 | 0.01 | 7.84E-02 | 6.15E-01 | 30 | 1674 | 0.00 | 0.01 | 9.09E-01 | 9.87E-01 | 220 | -0.01 | 0.01 | 1.94E-01 | 0.30 | 2.33E-01 |
| Enterodiol intake (ug/day) | Nutrients | 1799 | 0.01 | 0.01 | 1.24E-01 | 6.15E-01 | 46 | 1674 | 0.00 | 0.01 | 8.27E-01 | 9.74E-01 | 198 | 0.01 | 0.01 | 1.96E-01 | 0.00 | 3.89E-01 |
| Sum of Lariciresinol, Matairesinol, Pinoresinol, Secoisolariciresinol intake (ug/day) | Nutrients | 1799 | 0.02 | 0.01 | 7.68E-02 | 6.15E-01 | 27 | 1674 | 0.00 | 0.01 | 9.92E-01 | 9.92E-01 | 239 | 0.01 | 0.01 | 1.98E-01 | 0.32 | 2.24E-01 |
| Frequency of picking berries or mushrooms during leisure time | Physical activity | 1819 | -0.01 | 0.01 | 2.40E-01 | 7.09E-01 | 81 | 1690 | -0.01 | 0.01 | 5.30E-01 | 9.29E-01 | 134 | -0.01 | 0.01 | 2.02E-01 | 0.00 | 7.03E-01 |

| | | | | | | | | | | | | | | | | | | |
|---|---|---|---|---|---|---|---|---|---|---|---|---|---|---|---|---|---|---|
| Sometimes physically straining work | Physical activity | 1910 | 0.01 | 0.02 | 6.60E-01 | 9.30E-01 | 169 | 1786 | 0.03 | 0.02 | 1.73E-01 | 6.75E-01 | 61 | 0.02 | 0.01 | 2.07E-01 | 0.00 | 4.98E-01 |
| Pizza | Food | 1799 | -0.01 | 0.01 | 1.99E-01 | 6.44E-01 | 74 | 1674 | 0.00 | 0.01 | 6.64E-01 | 9.71E-01 | 162 | -0.01 | 0.01 | 2.07E-01 | 0.00 | 6.19E-01 |
| Snuff status: Snuff users vs non-snuff users | Tobacco use | 1623 | 0.01 | 0.02 | 5.17E-01 | 8.88E-01 | 137 | 1530 | 0.03 | 0.03 | 2.46E-01 | 7.68E-01 | 76 | 0.02 | 0.02 | 2.09E-01 | 0.00 | 6.60E-01 |
| Fried potatoes, pommes frites | Food | 1799 | 0.00 | 0.01 | 8.58E-01 | 9.91E-01 | 207 | 1674 | 0.02 | 0.01 | 3.81E-02 | 4.42E-01 | 20 | 0.01 | 0.01 | 2.13E-01 | 0.64 | 9.48E-02 |
| Average portion size of vegetables based on photographic illustration of four sizes (smallest to largest) | Food | 1799 | 0.02 | 0.01 | 7.56E-02 | 6.15E-01 | 26 | 1674 | 0.00 | 0.01 | 9.35E-01 | 9.89E-01 | 225 | 0.01 | 0.01 | 2.18E-01 | 0.39 | 1.99E-01 |
| Formic acid intaje (g/day) | Nutrients | 1799 | 0.00 | 0.01 | 6.91E-01 | 9.33E-01 | 177 | 1674 | -0.01 | 0.01 | 1.60E-01 | 6.38E-01 | 60 | -0.01 | 0.01 | 2.20E-01 | 0.00 | 4.28E-01 |
| Support from others | Social | 1936 | 0.02 | 0.01 | 6.93E-03 | 2.07E-01 | 8 | 1823 | -0.01 | 0.01 | 3.84E-01 | 9.22E-01 | 99 | 0.01 | 0.01 | 2.21E-01 | 0.85 | 1.04E-02 |
| Calcium intake (mg/day) | Nutrients | 1799 | 0.00 | 0.01 | 8.99E-01 | 9.95E-01 | 212 | 1674 | -0.02 | 0.01 | 5.65E-02 | 4.99E-01 | 27 | -0.01 | 0.01 | 2.27E-01 | 0.54 | 1.38E-01 |
| Mashed potato | Food | 651 | -0.01 | 0.02 | 4.15E-01 | 8.18E-01 | 120 | 580 | -0.01 | 0.02 | 3.79E-01 | 9.22E-01 | 98 | -0.01 | 0.01 | 2.30E-01 | 0.00 | 9.86E-01 |
| Iodine intake (ug/day) | Nutrients | 1799 | -0.01 | 0.01 | 2.06E-01 | 6.49E-01 | 76 | 1674 | 0.00 | 0.01 | 6.98E-01 | 9.71E-01 | 166 | -0.01 | 0.01 | 2.35E-01 | 0.00 | 5.63E-01 |
| Sedentary or standing work | Physical activity | 1910 | -0.01 | 0.02 | 5.97E-01 | 9.14E-01 | 155 | 1786 | -0.02 | 0.02 | 2.45E-01 | 7.68E-01 | 75 | -0.02 | 0.01 | 2.35E-01 | 0.00 | 6.38E-01 |
| Campestanol intake (mg/day) | Nutrients | 1799 | 0.01 | 0.01 | 1.54E-01 | 6.15E-01 | 58 | 1674 | 0.00 | 0.01 | 8.40E-01 | 9.74E-01 | 201 | 0.01 | 0.01 | 2.39E-01 | 0.00 | 4.07E-01 |
| Whole grain crisp bread | Food | 1799 | 0.01 | 0.01 | 3.69E-01 | 8.09E-01 | 109 | 1674 | 0.01 | 0.01 | 4.45E-01 | 9.29E-01 | 112 | 0.01 | 0.01 | 2.39E-01 | 0.00 | 9.63E-01 |
| Smoked fish/meat | Food | 1799 | -0.01 | 0.01 | 1.75E-01 | 6.19E-01 | 66 | 1674 | 0.00 | 0.01 | 8.35E-01 | 9.74E-01 | 199 | -0.01 | 0.01 | 2.40E-01 | 0.00 | 4.78E-01 |
| Boiled or baked potato | Food | 1799 | 0.00 | 0.01 | 8.10E-01 | 9.64E-01 | 201 | 1674 | 0.02 | 0.01 | 5.03E-02 | 4.99E-01 | 23 | 0.01 | 0.01 | 2.41E-01 | 0.60 | 1.13E-01 |
| Bregott on bread | Food | 1799 | -0.01 | 0.01 | 1.55E-01 | 6.15E-01 | 60 | 1674 | 0.00 | 0.01 | 8.46E-01 | 9.74E-01 | 206 | -0.01 | 0.01 | 2.42E-01 | 0.00 | 4.04E-01 |
| Long-term sickness | General health | 1889 | -0.03 | 0.03 | 3.23E-01 | 7.43E-01 | 103 | 1765 | -0.02 | 0.03 | 5.17E-01 | 9.29E-01 | 127 | -0.02 | 0.02 | 2.44E-01 | 0.00 | 8.42E-01 |
| If you exercise, change in exercise habits during the last year | Physical activity | 1696 | 0.01 | 0.01 | 1.81E-01 | 6.19E-01 | 70 | 1618 | 0.00 | 0.01 | 8.02E-01 | 9.74E-01 | 190 | 0.01 | 0.01 | 2.50E-01 | 0.00 | 4.67E-01 |
| Overall state of health compared to other your age | General health | 1895 | 0.02 | 0.01 | 7.22E-02 | 6.15E-01 | 25 | 1803 | 0.00 | 0.01 | 7.77E-01 | 9.74E-01 | 183 | 0.01 | 0.01 | 2.51E-01 | 0.50 | 1.57E-01 |
| Lariciresinol intake (ug/day) | Nutrients | 1799 | 0.01 | 0.01 | 1.44E-01 | 6.15E-01 | 55 | 1674 | 0.00 | 0.01 | 8.95E-01 | 9.87E-01 | 215 | 0.01 | 0.01 | 2.52E-01 | 0.00 | 3.61E-01 |
| Sodas, soft drinks, juice | Beverage | 1799 | 0.00 | 0.01 | 9.09E-01 | 9.95E-01 | 217 | 1674 | -0.01 | 0.01 | 1.37E-01 | 5.95E-01 | 55 | -0.01 | 0.01 | 2.52E-01 | 0.00 | 3.39E-01 |
| Marital status: Single vs Married/partner | Social | 1810 | 0.02 | 0.03 | 4.19E-01 | 8.18E-01 | 122 | 1693 | 0.02 | 0.03 | 4.26E-01 | 9.29E-01 | 106 | 0.02 | 0.02 | 2.56E-01 | 0.00 | 9.95E-01 |
| Number of social interactions during a normal week | Social | 1935 | -0.01 | 0.01 | 4.38E-01 | 8.37E-01 | 125 | 1822 | -0.01 | 0.01 | 4.17E-01 | 9.29E-01 | 104 | -0.01 | 0.01 | 2.62E-01 | 0.00 | 9.69E-01 |
| Plant based protein intake (g/day) | Nutrients | 1799 | 0.01 | 0.01 | 2.02E-01 | 6.45E-01 | 75 | 1674 | 0.00 | 0.01 | 7.87E-01 | 9.74E-01 | 186 | 0.01 | 0.01 | 2.62E-01 | 0.00 | 5.07E-01 |
| Whole grain intake (g/day) | Food | 1799 | 0.01 | 0.01 | 2.61E-01 | 7.20E-01 | 84 | 1674 | 0.00 | 0.01 | 6.63E-01 | 9.71E-01 | 161 | 0.01 | 0.01 | 2.64E-01 | 0.00 | 6.51E-01 |
| Marital status: Single vs Divorced/separated | Social | 325 | -0.04 | 0.04 | 3.25E-01 | 7.43E-01 | 104 | 326 | -0.02 | 0.04 | 5.54E-01 | 9.29E-01 | 140 | -0.03 | 0.03 | 2.69E-01 | 0.00 | 7.52E-01 |
| Cohabitation: Live alone vs Adult and children | Social | 1386 | 0.05 | 0.03 | 1.17E-01 | 6.15E-01 | 41 | 1250 | 0.00 | 0.03 | 9.41E-01 | 9.90E-01 | 227 | 0.03 | 0.02 | 2.78E-01 | 0.23 | 2.56E-01 |
| Low fat milk (0,5%) | Beverage | 1799 | 0.02 | 0.01 | 1.42E-01 | 6.15E-01 | 54 | 1674 | 0.00 | 0.01 | 9.35E-01 | 9.89E-01 | 226 | 0.01 | 0.01 | 2.80E-01 | 0.00 | 3.18E-01 |
| Feel the need to reduce alcohol consumption | Alcohol | 1735 | 0.06 | 0.03 | 5.55E-02 | 6.03E-01 | 22 | 1633 | -0.01 | 0.03 | 6.87E-01 | 9.71E-01 | 164 | 0.02 | 0.02 | 2.82E-01 | 0.63 | 1.02E-01 |
| Sleep status | Sleep | 1235 | 0.01 | 0.01 | 2.86E-01 | 7.20E-01 | 91 | 1189 | 0.00 | 0.01 | 6.59E-01 | 9.71E-01 | 159 | 0.01 | 0.01 | 2.84E-01 | 0.00 | 6.66E-01 |
| Breakfast habits: Only coffee/tea for breakfast vs not breakfast at all | Food | 136 | 0.08 | 0.09 | 3.97E-01 | 8.18E-01 | 113 | 141 | 0.07 | 0.11 | 5.18E-01 | 9.29E-01 | 129 | 0.08 | 0.07 | 2.85E-01 | 0.00 | 9.65E-01 |
| Control over planning and execution of the workday | Psychosocial | 1917 | 0.01 | 0.01 | 1.96E-01 | 6.41E-01 | 73 | 1797 | 0.00 | 0.01 | 8.46E-01 | 9.74E-01 | 205 | 0.01 | 0.01 | 2.87E-01 | 0.00 | 4.46E-01 |
| Oil for cooking | Food | 1799 | 0.00 | 0.01 | 9.31E-01 | 9.95E-01 | 222 | 1674 | -0.01 | 0.01 | 1.13E-01 | 5.61E-01 | 48 | -0.01 | 0.01 | 2.93E-01 | 0.30 | 2.33E-01 |
| Soft cheese | Food | 651 | 0.02 | 0.01 | 1.77E-01 | 6.19E-01 | 67 | 580 | 0.00 | 0.02 | 9.51E-01 | 9.90E-01 | 228 | 0.01 | 0.01 | 3.00E-01 | 0.00 | 3.84E-01 |
| Fibre intake (g/day) | Nutrients | 1799 | 0.02 | 0.01 | 1.08E-01 | 6.15E-01 | 37 | 1674 | 0.00 | 0.01 | 8.08E-01 | 9.74E-01 | 194 | 0.01 | 0.01 | 3.01E-01 | 0.37 | 2.09E-01 |
| Sweets | Food | 1799 | -0.01 | 0.01 | 8.77E-02 | 6.15E-01 | 31 | 1674 | 0.01 | 0.01 | 5.97E-01 | 9.58E-01 | 149 | -0.01 | 0.01 | 3.10E-01 | 0.54 | 1.41E-01 |
| Do you feel important and appreciated outside your home? | Psychosocial | 1237 | 0.01 | 0.01 | 5.30E-01 | 8.88E-01 | 141 | 1185 | 0.01 | 0.01 | 4.32E-01 | 9.29E-01 | 109 | 0.01 | 0.01 | 3.19E-01 | 0.00 | 8.97E-01 |
| Skill demand from job | Psychosocial | 1908 | 0.01 | 0.01 | 4.08E-01 | 8.18E-01 | 115 | 1792 | 0.01 | 0.01 | 5.70E-01 | 9.29E-01 | 146 | 0.01 | 0.01 | 3.21E-01 | 0.00 | 8.75E-01 |
| Work shifts/weekends | Psychosocial | 1879 | 0.00 | 0.02 | 9.85E-01 | 9.95E-01 | 234 | 1762 | 0.03 | 0.02 | 1.56E-01 | 6.38E-01 | 58 | 0.01 | 0.01 | 3.25E-01 | 0.04 | 3.07E-01 |
| Light beer | Alcohol | 1799 | 0.02 | 0.01 | 1.28E-01 | 6.15E-01 | 47 | 1674 | 0.00 | 0.01 | 8.70E-01 | 9.74E-01 | 212 | 0.01 | 0.01 | 3.29E-01 | 0.28 | 2.38E-01 |
| Shellfish (e.g. shrimps, scallops) | Food | 651 | 0.01 | 0.02 | 5.32E-01 | 8.88E-01 | 142 | 580 | 0.01 | 0.02 | 4.51E-01 | 9.29E-01 | 114 | 0.01 | 0.01 | 3.30E-01 | 0.00 | 9.17E-01 |
| Frequency of walking during leisure time | Physical activity | 1870 | 0.01 | 0.01 | 5.09E-01 | 8.88E-01 | 135 | 1762 | 0.01 | 0.01 | 4.75E-01 | 9.29E-01 | 120 | 0.01 | 0.01 | 3.31E-01 | 0.00 | 9.61E-01 |
| Vitamin B2 intake (ug/day) | Nutrients | 1799 | 0.00 | 0.01 | 9.05E-01 | 9.95E-01 | 215 | 1674 | -0.01 | 0.01 | 2.00E-01 | 7.14E-01 | 67 | -0.01 | 0.01 | 3.36E-01 | 0.00 | 3.93E-01 |
| Frequency of engaging in clubs, associations or study circles | Social | 1302 | -0.02 | 0.01 | 1.28E-01 | 6.15E-01 | 48 | 1210 | 0.00 | 0.01 | 8.97E-01 | 9.87E-01 | 216 | -0.01 | 0.01 | 3.37E-01 | 0.29 | 2.35E-01 |
| Number of snuff boxes per week | Tobacco use | 1851 | 0.00 | 0.01 | 9.02E-01 | 9.95E-01 | 213 | 1733 | 0.01 | 0.01 | 1.31E-01 | 5.95E-01 | 52 | 0.01 | 0.01 | 3.39E-01 | 0.28 | 2.40E-01 |
| Zinc intake (mg/day) | Nutrients | 1799 | 0.01 | 0.01 | 3.12E-01 | 7.31E-01 | 102 | 1674 | 0.00 | 0.01 | 7.60E-01 | 9.74E-01 | 182 | 0.01 | 0.01 | 3.44E-01 | 0.00 | 6.39E-01 |
| Saturated fat intake (g/day) | Nutrients | 1799 | -0.01 | 0.01 | 5.48E-01 | 8.88E-01 | 145 | 1674 | -0.01 | 0.01 | 4.57E-01 | 9.29E-01 | 116 | -0.01 | 0.01 | 3.45E-01 | 0.00 | 8.79E-01 |
| Average portion size of potatoes/rice/pasta based on photographic illustration of four sizes (smallest to largest) | Food | 1799 | 0.00 | 0.01 | 8.37E-01 | 9.76E-01 | 205 | 1674 | 0.01 | 0.01 | 2.49E-01 | 7.68E-01 | 77 | 0.01 | 0.01 | 3.47E-01 | 0.00 | 4.84E-01 |
| Ingenuity or creativity demand from job | Psychosocial | 1912 | 0.01 | 0.01 | 1.20E-01 | 6.15E-01 | 45 | 1793 | 0.00 | 0.01 | 7.56E-01 | 9.74E-01 | 180 | 0.01 | 0.01 | 3.52E-01 | 0.39 | 2.00E-01 |
| Potassium intake (mg/day) | Nutrients | 1799 | 0.01 | 0.01 | 5.11E-01 | 8.88E-01 | 136 | 1674 | 0.01 | 0.01 | 5.25E-01 | 9.29E-01 | 132 | 0.01 | 0.01 | 3.60E-01 | 0.00 | 9.97E-01 |
| Trans fat intake (g/day) | Nutrients | 1799 | -0.02 | 0.01 | 1.64E-01 | 6.15E-01 | 62 | 1674 | 0.00 | 0.01 | 8.67E-01 | 9.74E-01 | 210 | -0.01 | 0.01 | 3.67E-01 | 0.14 | 2.82E-01 |
| Vision status | General health | 1240 | 0.00 | 0.01 | 9.38E-01 | 9.95E-01 | 224 | 1187 | 0.02 | 0.01 | 1.52E-01 | 6.37E-01 | 57 | 0.01 | 0.01 | 3.67E-01 | 0.20 | 2.64E-01 |
| Salad dressing with oil | Food | 1799 | 0.01 | 0.01 | 1.63E-01 | 6.15E-01 | 61 | 1674 | 0.00 | 0.01 | 8.40E-01 | 9.74E-01 | 203 | 0.01 | 0.01 | 3.70E-01 | 0.15 | 2.77E-01 |
| Potato salad | Food | 651 | -0.01 | 0.02 | 5.28E-01 | 8.88E-01 | 139 | 580 | -0.01 | 0.02 | 5.28E-01 | 9.29E-01 | 133 | -0.01 | 0.01 | 3.72E-01 | 0.00 | 9.64E-01 |
| Total protein intake (g/day) | Nutrients | 1799 | 0.00 | 0.01 | 7.03E-01 | 9.33E-01 | 180 | 1674 | 0.01 | 0.01 | 3.68E-01 | 9.16E-01 | 96 | 0.01 | 0.01 | 3.74E-01 | 0.00 | 6.82E-01 |
| Low fat margarine on bread | Food | 1799 | 0.01 | 0.01 | 1.64E-01 | 6.15E-01 | 63 | 1674 | 0.00 | 0.01 | 8.40E-01 | 9.74E-01 | 202 | 0.01 | 0.01 | 3.82E-01 | 0.18 | 2.70E-01 |
| Folic acid intake (ug/day) | Nutrients | 1799 | 0.02 | 0.01 | 2.78E-02 | 3.91E-01 | 17 | 1674 | -0.01 | 0.01 | 2.79E-01 | 7.95E-01 | 84 | 0.01 | 0.01 | 3.87E-01 | 0.81 | 2.17E-02 |
| Pinoresinol intake (ug/day) | Nutrients | 1799 | 0.01 | 0.01 | 1.32E-01 | 6.15E-01 | 49 | 1674 | 0.00 | 0.01 | 7.25E-01 | 9.74E-01 | 178 | 0.01 | 0.01 | 3.92E-01 | 0.40 | 1.98E-01 |
| Margarine for cooking | Food | 1799 | 0.00 | 0.01 | 6.77E-01 | 9.30E-01 | 174 | 1674 | 0.01 | 0.01 | 1.05E-01 | 5.42E-01 | 45 | 0.01 | 0.01 | 4.03E-01 | 0.52 | 1.47E-01 |
| Corn flakes | Food | 1799 | 0.01 | 0.01 | 5.50E-01 | 8.88E-01 | 147 | 1674 | 0.01 | 0.01 | 5.69E-01 | 9.29E-01 | 145 | 0.01 | 0.01 | 4.09E-01 | 0.00 | 9.92E-01 |
| Brewed (filtered) coffee | Beverage | 1799 | 0.01 | 0.01 | 2.77E-01 | 7.20E-01 | 89 | 1674 | 0.00 | 0.01 | 9.69E-01 | 9.90E-01 | 233 | 0.01 | 0.01 | 4.12E-01 | 0.00 | 4.75E-01 |
| Monosaccharides intake (g/day) | Nutrients | 1799 | 0.00 | 0.01 | 6.70E-01 | 9.30E-01 | 170 | 1674 | -0.02 | 0.01 | 1.02E-01 | 5.42E-01 | 44 | -0.01 | 0.01 | 4.24E-01 | 0.55 | 1.37E-01 |
| Enterolactone intake (ug/day) | Nutrients | 1799 | 0.01 | 0.01 | 1.10E-01 | 6.15E-01 | 38 | 1674 | -0.01 | 0.01 | 5.58E-01 | 9.29E-01 | 141 | 0.01 | 0.01 | 4.27E-01 | 0.56 | 1.32E-01 |
| Contradictory demands in job | Psychosocial | 1898 | 0.00 | 0.01 | 7.32E-01 | 9.35E-01 | 186 | 1782 | -0.01 | 0.01 | 1.37E-01 | 5.95E-01 | 54 | 0.00 | 0.01 | 4.28E-01 | 0.41 | 1.91E-01 |
| Participation in associations or voluntary organisations | Social | 1935 | -0.01 | 0.02 | 7.18E-01 | 9.35E-01 | 183 | 1812 | -0.01 | 0.02 | 4.44E-01 | 9.29E-01 | 111 | -0.01 | 0.01 | 4.29E-01 | 0.00 | 7.63E-01 |
| Mood status | Psychosocial | 1235 | 0.01 | 0.01 | 1.85E-01 | 6.24E-01 | 71 | 1180 | 0.00 | 0.01 | 8.07E-01 | 9.74E-01 | 193 | 0.01 | 0.01 | 4.29E-01 | 0.16 | 2.75E-01 |
| Blood based food | Food | 651 | 0.02 | 0.01 | 2.25E-01 | 6.80E-01 | 79 | 580 | 0.00 | 0.02 | 8.43E-01 | 9.74E-01 | 204 | 0.01 | 0.01 | 4.34E-01 | 0.00 | 3.42E-01 |
| Arachidonic acid (ARA) intake (g/day) | Nutrients | 1799 | -0.01 | 0.01 | 4.13E-01 | 8.18E-01 | 119 | 1674 | 0.02 | 0.01 | 5.24E-02 | 4.99E-01 | 24 | 0.00 | 0.01 | 4.34E-01 | 0.74 | 5.05E-02 |
| Whole grain soft bread | Food | 1799 | 0.01 | 0.01 | 2.76E-01 | 7.20E-01 | 88 | 1674 | 0.00 | 0.01 | 9.69E-01 | 9.90E-01 | 234 | 0.01 | 0.01 | 4.36E-01 | 0.00 | 4.45E-01 |
| Sausage as main dish | Food | 1799 | 0.00 | 0.01 | 9.65E-01 | 9.95E-01 | 228 | 1674 | 0.01 | 0.01 | 1.90E-01 | 6.97E-01 | 65 | 0.00 | 0.01 | 4.38E-01 | 0.11 | 2.89E-01 |
| Do you feel important and appreciated in your home? | Psychosocial | 1226 | 0.00 | 0.01 | 7.53E-01 | 9.35E-01 | 191 | 1179 | 0.01 | 0.01 | 4.38E-01 | 9.29E-01 | 110 | 0.01 | 0.01 | 4.40E-01 | 0.00 | 7.46E-01 |
| Vitamin E intake (mg/day) | Nutrients | 1799 | 0.00 | 0.01 | 9.95E-01 | 9.95E-01 | 239 | 1674 | 0.01 | 0.01 | 2.69E-01 | 7.73E-01 | 83 | 0.00 | 0.01 | 4.51E-01 | 0.00 | 4.18E-01 |
| Feel uneasy or guilty because of your way of drinking | Alcohol | 1734 | 0.03 | 0.03 | 3.84E-01 | 8.18E-01 | 111 | 1633 | 0.00 | 0.03 | 8.66E-01 | 9.74E-01 | 209 | 0.02 | 0.02 | 4.62E-01 | 0.00 | 6.21E-01 |
| Palmitic acid intake (g/day) | Nutrients | 1799 | -0.01 | 0.01 | 5.57E-01 | 8.88E-01 | 150 | 1674 | 0.00 | 0.01 | 6.57E-01 | 9.71E-01 | 157 | 0.00 | 0.01 | 4.63E-01 | 0.00 | 9.49E-01 |
| Pasta | Food | 1799 | 0.00 | 0.01 | 6.45E-01 | 9.30E-01 | 163 | 1674 | 0.01 | 0.01 | 5.68E-01 | 9.29E-01 | 144 | 0.00 | 0.01 | 4.65E-01 | 0.00 | 9.43E-01 |
| Carbohydrates intake (g/day) | Nutrients | 1799 | 0.00 | 0.01 | 7.87E-01 | 9.45E-01 | 199 | 1674 | -0.01 | 0.01 | 4.50E-01 | 9.29E-01 | 113 | 0.00 | 0.01 | 4.74E-01 | 0.00 | 7.18E-01 |
| Blota (broth + bread) | Food | 651 | 0.00 | 0.02 | 9.56E-01 | 9.95E-01 | 227 | 580 | -0.02 | 0.02 | 2.86E-01 | 7.99E-01 | 85 | -0.01 | 0.01 | 4.83E-01 | 0.00 | 4.20E-01 |
| Number of friends that can come to your home at any time and feel at home | Social | 1933 | 0.00 | 0.01 | 6.97E-01 | 9.33E-01 | 178 | 1818 | 0.01 | 0.01 | 5.45E-01 | 9.29E-01 | 138 | 0.00 | 0.01 | 4.83E-01 | 0.00 | 8.74E-01 |
| Breakfast habits: Gruel w/o sandwich for breakfast vs not breakfast at all | Food | 156 | 0.07 | 0.06 | 2.78E-01 | 7.20E-01 | 90 | 154 | -0.05 | 0.10 | 6.30E-01 | 9.71E-01 | 152 | 0.04 | 0.05 | 4.85E-01 | 0.00 | 3.34E-01 |
| Snuff status: Former snuff users vs non-snuff users | Tobacco use | 1452 | 0.08 | 0.03 | 8.83E-03 | 2.11E-01 | 10 | 1342 | -0.05 | 0.03 | 8.45E-02 | 5.31E-01 | 38 | 0.01 | 0.02 | 4.85E-01 | 0.89 | 2.21E-03 |
| Enough time for job assignments | Psychosocial | 1905 | 0.00 | 0.01 | 7.31E-01 | 9.35E-01 | 185 | 1786 | -0.01 | 0.01 | 5.20E-01 | 9.29E-01 | 130 | 0.00 | 0.01 | 4.86E-01 | 0.00 | 8.24E-01 |
| Liquor, spirits | Alcohol | 1799 | 0.01 | 0.01 | 1.79E-01 | 6.19E-01 | 68 | 1674 | 0.00 | 0.01 | 6.34E-01 | 9.71E-01 | 154 | 0.00 | 0.01 | 4.89E-01 | 0.36 | 2.13E-01 |

| | | | | | | | | | | | | | | | | | | |
|---|---|---|---|---|---|---|---|---|---|---|---|---|---|---|---|---|---|---|
| Sugar, honey, marmelade, jam | Food | 1799 | 0.00 | 0.01 | 5.91E-01 | 9.14E-01 | 154 | 1674 | 0.00 | 0.01 | 6.61E-01 | 9.71E-01 | 160 | 0.00 | 0.01 | 4.90E-01 | 0.00 | 9.47E-01 |
| Parents or siblings have diabetes | General health | 1927 | -0.04 | 0.02 | 1.11E-01 | 6.15E-01 | 39 | 1815 | 0.02 | 0.02 | 5.05E-01 | 9.29E-01 | 124 | -0.01 | 0.02 | 4.92E-01 | 0.60 | 1.13E-01 |
| Sour milk, yoghurt (3% fat) | Food | 1799 | 0.00 | 0.01 | 7.11E-01 | 9.35E-01 | 181 | 1674 | -0.01 | 0.01 | 1.60E-01 | 6.38E-01 | 59 | 0.00 | 0.01 | 5.00E-01 | 0.40 | 1.98E-01 |
| Breakfast habits: Porridge w/o sandwich for breakfast vs not breakfast at all | Food | 278 | 0.07 | 0.04 | 1.04E-01 | 6.15E-01 | 35 | 255 | -0.05 | 0.06 | 3.28E-01 | 8.61E-01 | 91 | 0.02 | 0.03 | 5.08E-01 | 0.69 | 7.41E-02 |
| Milk, sour milk (3%) | Beverage | 1799 | 0.00 | 0.01 | 5.28E-01 | 8.88E-01 | 140 | 1674 | 0.00 | 0.01 | 8.16E-01 | 9.74E-01 | 195 | 0.00 | 0.01 | 5.21E-01 | 0.00 | 8.43E-01 |
| Linoleic acid intake (g/day) | Nutrients | 1799 | 0.00 | 0.01 | 8.93E-01 | 9.95E-01 | 210 | 1674 | -0.01 | 0.01 | 2.88E-01 | 7.99E-01 | 86 | 0.00 | 0.01 | 5.23E-01 | 0.00 | 3.89E-01 |
| Phosphate intake (mg/day) | Nutrients | 1799 | 0.01 | 0.01 | 2.89E-01 | 7.20E-01 | 96 | 1674 | 0.00 | 0.01 | 8.47E-01 | 9.74E-01 | 207 | 0.00 | 0.01 | 5.25E-01 | 0.00 | 3.84E-01 |
| Butter on bread | Food | 1799 | 0.00 | 0.01 | 8.48E-01 | 9.84E-01 | 206 | 1674 | -0.01 | 0.01 | 2.62E-01 | 7.68E-01 | 79 | 0.00 | 0.01 | 5.25E-01 | 0.00 | 3.44E-01 |
| Frequncy of social contacts with colleagues during leisure time | Psychosocial | 1860 | 0.01 | 0.01 | 4.65E-01 | 8.61E-01 | 129 | 1745 | 0.00 | 0.01 | 9.00E-01 | 9.87E-01 | 218 | 0.00 | 0.01 | 5.34E-01 | 0.00 | 6.85E-01 |
| Banana | Food | 1799 | -0.01 | 0.01 | 4.03E-01 | 8.18E-01 | 114 | 1674 | 0.00 | 0.01 | 9.92E-01 | 9.92E-01 | 238 | 0.00 | 0.01 | 5.39E-01 | 0.00 | 5.70E-01 |
| Selenium intake (ug/day) | Nutrients | 1799 | 0.00 | 0.01 | 9.74E-01 | 9.95E-01 | 229 | 1674 | 0.01 | 0.01 | 3.46E-01 | 8.90E-01 | 93 | 0.00 | 0.01 | 5.40E-01 | 0.00 | 4.74E-01 |
| Liver, kidney | Food | 651 | 0.01 | 0.01 | 3.42E-01 | 7.72E-01 | 106 | 580 | 0.00 | 0.02 | 8.72E-01 | 9.74E-01 | 214 | 0.01 | 0.01 | 5.45E-01 | 0.00 | 4.53E-01 |
| White (soft) bread, thin crisp bread | Food | 1799 | -0.01 | 0.01 | 5.33E-01 | 8.88E-01 | 143 | 1674 | 0.00 | 0.01 | 8.36E-01 | 9.74E-01 | 200 | 0.00 | 0.01 | 5.55E-01 | 0.00 | 7.74E-01 |
| Oatflake, whole wheat, rye or barley porridge | Food | 1799 | 0.01 | 0.01 | 4.08E-01 | 8.18E-01 | 116 | 1674 | 0.00 | 0.01 | 9.55E-01 | 9.90E-01 | 229 | 0.00 | 0.01 | 5.62E-01 | 0.00 | 5.53E-01 |
| Fiber cereals | Food | 1799 | 0.01 | 0.01 | 3.11E-01 | 7.31E-01 | 101 | 1674 | 0.00 | 0.01 | 8.05E-01 | 9.74E-01 | 192 | 0.00 | 0.01 | 5.63E-01 | 0.00 | 3.85E-01 |
| Mixed frozen vegetables | Food | 651 | 0.01 | 0.01 | 4.91E-01 | 8.88E-01 | 131 | 580 | -0.03 | 0.02 | 7.12E-02 | 5.03E-01 | 32 | -0.01 | 0.01 | 5.63E-01 | 0.71 | 6.49E-02 |
| Frequency of shoverling snow during leisure time | Physical activity | 1838 | 0.00 | 0.01 | 6.71E-01 | 9.30E-01 | 171 | 1727 | 0.00 | 0.01 | 7.15E-01 | 9.71E-01 | 174 | 0.00 | 0.01 | 5.77E-01 | 0.00 | 9.66E-01 |
| Chips, popcorn, salted nuts | Food | 1799 | 0.00 | 0.01 | 8.07E-01 | 9.64E-01 | 200 | 1674 | 0.00 | 0.01 | 6.07E-01 | 9.61E-01 | 151 | 0.00 | 0.01 | 5.80E-01 | 0.00 | 8.92E-01 |
| Cheese 10-17% | Food | 1799 | 0.01 | 0.01 | 2.64E-01 | 7.20E-01 | 86 | 1674 | 0.00 | 0.01 | 7.03E-01 | 9.71E-01 | 170 | 0.00 | 0.01 | 5.93E-01 | 0.10 | 2.93E-01 |
| White cabbage, lettuce, lettuce cabbage, spinach, borecole | Food | 1799 | 0.02 | 0.01 | 3.53E-02 | 4.63E-01 | 18 | 1674 | -0.02 | 0.01 | 1.29E-01 | 5.95E-01 | 51 | 0.00 | 0.01 | 5.98E-01 | 0.85 | 1.10E-02 |
| Soft whey cheese | Food | 651 | 0.01 | 0.01 | 5.24E-01 | 8.88E-01 | 138 | 580 | -0.04 | 0.02 | 3.89E-02 | 4.42E-01 | 21 | -0.01 | 0.01 | 6.00E-01 | 0.77 | 3.55E-02 |
| Linolenic acid intake (g/day) | Nutrients | 1799 | 0.00 | 0.01 | 9.81E-01 | 9.95E-01 | 232 | 1674 | -0.01 | 0.01 | 4.54E-01 | 9.29E-01 | 115 | 0.00 | 0.01 | 6.17E-01 | 0.00 | 5.77E-01 |
| Satisfaction with leisure time | Psychosocial | 1234 | -0.01 | 0.01 | 5.99E-01 | 9.14E-01 | 156 | 1182 | 0.01 | 0.01 | 2.09E-01 | 7.15E-01 | 69 | 0.00 | 0.01 | 6.28E-01 | 0.38 | 2.03E-01 |
| Root vegetables, carrot | Food | 1799 | 0.01 | 0.01 | 1.14E-01 | 6.15E-01 | 40 | 1674 | -0.01 | 0.01 | 3.03E-01 | 8.14E-01 | 88 | 0.00 | 0.01 | 6.35E-01 | 0.70 | 6.75E-02 |
| Receive hugs to comfort and support you | Social | 1926 | 0.01 | 0.02 | 7.02E-01 | 9.33E-01 | 179 | 1807 | 0.01 | 0.02 | 7.81E-01 | 9.74E-01 | 184 | 0.01 | 0.02 | 6.40E-01 | 0.00 | 9.48E-01 |
| Beta-carotene intake (mg/day) | Nutrients | 1799 | 0.02 | 0.01 | 9.91E-02 | 6.15E-01 | 33 | 1674 | -0.01 | 0.01 | 2.63E-01 | 7.68E-01 | 82 | 0.00 | 0.01 | 6.46E-01 | 0.73 | 5.24E-02 |
| Tomato, cucumber | Food | 1799 | 0.01 | 0.01 | 3.06E-01 | 7.31E-01 | 99 | 1674 | 0.00 | 0.01 | 7.19E-01 | 9.71E-01 | 177 | 0.00 | 0.01 | 6.48E-01 | 0.00 | 3.25E-01 |
| Brown beans, pea soup | Food | 1799 | 0.01 | 0.01 | 4.42E-01 | 8.39E-01 | 126 | 1674 | 0.00 | 0.01 | 8.67E-01 | 9.74E-01 | 211 | 0.00 | 0.01 | 6.57E-01 | 0.00 | 5.16E-01 |
| Apple, pear, peach, orange, mandarin and grapefruit | Food | 1799 | 0.00 | 0.01 | 9.83E-01 | 9.95E-01 | 233 | 1674 | -0.01 | 0.01 | 5.02E-01 | 9.29E-01 | 123 | 0.00 | 0.01 | 6.58E-01 | 0.00 | 6.12E-01 |
| Travel to work: Cycle to work vs passive travel to work | Physical activity | 1423 | 0.01 | 0.02 | 5.50E-01 | 8.88E-01 | 146 | 1348 | 0.00 | 0.03 | 9.86E-01 | 9.92E-01 | 236 | 0.01 | 0.02 | 6.60E-01 | 0.00 | 6.85E-01 |
| Possibility to leave your work for a while to speak with a colleague | Psychosocial | 1903 | 0.01 | 0.01 | 2.64E-01 | 7.20E-01 | 85 | 1776 | -0.02 | 0.01 | 7.85E-02 | 5.24E-01 | 35 | 0.00 | 0.01 | 6.60E-01 | 0.76 | 4.15E-02 |
| Breakfast habits: Coffee/tea and wheat buns or rusk for breakfast vs not breakfast at all | Food | 180 | 0.03 | 0.05 | 5.57E-01 | 8.88E-01 | 149 | 175 | -0.01 | 0.07 | 9.22E-01 | 9.89E-01 | 222 | 0.02 | 0.04 | 6.71E-01 | 0.00 | 6.75E-01 |
| Changed everyday exercise during the last year | Physical activity | 1943 | 0.01 | 0.01 | 4.13E-01 | 8.18E-01 | 117 | 1825 | 0.00 | 0.01 | 8.02E-01 | 9.74E-01 | 189 | 0.00 | 0.01 | 6.77E-01 | 0.00 | 4.54E-01 |
| Overall state of health during the last year | General health | 1939 | 0.00 | 0.01 | 6.52E-01 | 9.30E-01 | 165 | 1825 | 0.00 | 0.01 | 9.04E-01 | 9.87E-01 | 219 | 0.00 | 0.01 | 6.85E-01 | 0.00 | 8.17E-01 |
| Travel to work: Walk to work vs passive travel to work | Physical activity | 1288 | 0.05 | 0.03 | 7.68E-02 | 6.15E-01 | 28 | 1225 | -0.04 | 0.03 | 1.93E-01 | 6.98E-01 | 66 | 0.01 | 0.02 | 6.86E-01 | 0.79 | 3.07E-02 |
| Lean fish (e.g. perch, bass, cod) | Food | 1799 | -0.01 | 0.01 | 3.64E-01 | 8.06E-01 | 108 | 1674 | 0.02 | 0.01 | 7.14E-02 | 5.03E-01 | 33 | 0.00 | 0.01 | 6.87E-01 | 0.74 | 4.78E-02 |
| Satisfaction with economy | Psychosocial | 1233 | 0.01 | 0.01 | 3.82E-01 | 8.18E-01 | 110 | 1181 | 0.00 | 0.01 | 7.18E-01 | 9.71E-01 | 176 | 0.00 | 0.01 | 7.02E-01 | 0.00 | 3.87E-01 |
| Animal based protein intake (g/day) | Nutrients | 1799 | 0.00 | 0.01 | 8.69E-01 | 9.93E-01 | 209 | 1674 | 0.01 | 0.01 | 4.69E-01 | 9.29E-01 | 117 | 0.00 | 0.01 | 7.19E-01 | 0.00 | 5.16E-01 |
| Butter for cooking | Food | 1799 | 0.01 | 0.01 | 2.88E-01 | 7.20E-01 | 95 | 1674 | -0.01 | 0.01 | 4.03E-01 | 9.29E-01 | 102 | 0.00 | 0.01 | 7.38E-01 | 0.42 | 1.90E-01 |
| Memory status | Psychosocial | 1231 | 0.00 | 0.01 | 9.19E-01 | 9.95E-01 | 220 | 1183 | 0.00 | 0.01 | 7.11E-01 | 9.71E-01 | 171 | 0.00 | 0.01 | 7.38E-01 | 0.00 | 8.50E-01 |
| Cheese 28% | Food | 1799 | 0.00 | 0.01 | 9.92E-01 | 9.95E-01 | 238 | 1674 | 0.00 | 0.01 | 6.44E-01 | 9.71E-01 | 156 | 0.00 | 0.01 | 7.41E-01 | 0.00 | 7.47E-01 |
| Fat intake (g/day) | Nutrients | 1799 | 0.00 | 0.01 | 6.18E-01 | 9.19E-01 | 160 | 1674 | 0.00 | 0.01 | 9.55E-01 | 9.90E-01 | 230 | 0.00 | 0.01 | 7.46E-01 | 0.00 | 7.01E-01 |
| Polyunsaturated fat intake (g/day) | Nutrients | 1799 | 0.00 | 0.01 | 9.43E-01 | 9.95E-01 | 226 | 1674 | 0.00 | 0.01 | 7.02E-01 | 9.71E-01 | 169 | 0.00 | 0.01 | 7.51E-01 | 0.00 | 8.23E-01 |
| Vitamin B12 intake (ug/day) | Nutrients | 1799 | -0.01 | 0.01 | 5.02E-01 | 8.88E-01 | 134 | 1674 | 0.00 | 0.01 | 7.82E-01 | 9.74E-01 | 185 | 0.00 | 0.01 | 7.56E-01 | 0.00 | 5.12E-01 |
| Distance to work in kilometers (one way) | Physical activity | 1649 | -0.01 | 0.01 | 3.58E-01 | 7.99E-01 | 107 | 1552 | 0.00 | 0.01 | 6.58E-01 | 9.71E-01 | 158 | 0.00 | 0.01 | 7.61E-01 | 0.00 | 3.30E-01 |
| High mental demand from job | Psychosocial | 1895 | 0.00 | 0.01 | 7.77E-01 | 9.38E-01 | 198 | 1779 | 0.01 | 0.01 | 4.79E-01 | 9.29E-01 | 121 | 0.00 | 0.01 | 7.62E-01 | 0.00 | 4.84E-01 |
| Margarine on bread | Food | 1799 | 0.00 | 0.01 | 9.04E-01 | 9.95E-01 | 214 | 1674 | 0.01 | 0.01 | 4.97E-01 | 9.29E-01 | 122 | 0.00 | 0.01 | 7.62E-01 | 0.00 | 5.36E-01 |
| Minced meat dishes | Food | 1799 | 0.00 | 0.01 | 9.18E-01 | 9.95E-01 | 219 | 1674 | 0.01 | 0.01 | 5.76E-01 | 9.30E-01 | 148 | 0.00 | 0.01 | 7.63E-01 | 0.00 | 6.29E-01 |
| Permanent employment | Psychosocial | 1906 | -0.02 | 0.02 | 4.15E-01 | 8.18E-01 | 121 | 1804 | 0.02 | 0.02 | 2.09E-01 | 7.15E-01 | 70 | 0.00 | 0.01 | 7.70E-01 | 0.54 | 1.42E-01 |
| Marital status: Single vs Widow/widower | Social | 220 | 0.20 | 0.13 | 1.45E-01 | 6.15E-01 | 56 | 231 | -0.10 | 0.08 | 2.25E-01 | 7.36E-01 | 73 | -0.02 | 0.07 | 7.82E-01 | 0.72 | 5.97E-02 |
| Smoking status: Former smokers vs non-smokers | Tobacco use | 1357 | 0.01 | 0.02 | 7.51E-01 | 9.35E-01 | 190 | 1257 | -0.02 | 0.02 | 4.09E-01 | 9.29E-01 | 103 | 0.00 | 0.02 | 7.83E-01 | 0.00 | 4.00E-01 |
| Cholesterol intake (g/day) | Nutrients | 1799 | -0.01 | 0.01 | 5.51E-01 | 8.88E-01 | 148 | 1674 | 0.00 | 0.01 | 7.89E-01 | 9.74E-01 | 187 | 0.00 | 0.01 | 7.92E-01 | 0.00 | 5.49E-01 |
| Campesterol intake (mg/day) | Nutrients | 1799 | 0.00 | 0.01 | 9.88E-01 | 9.95E-01 | 237 | 1674 | 0.00 | 0.01 | 6.91E-01 | 9.71E-01 | 165 | 0.00 | 0.01 | 7.94E-01 | 0.00 | 7.63E-01 |
| Frequency of dancing during leisure time | Physical activity | 1665 | 0.00 | 0.01 | 9.87E-01 | 9.95E-01 | 236 | 1541 | 0.00 | 0.01 | 7.18E-01 | 9.71E-01 | 175 | 0.00 | 0.01 | 7.95E-01 | 0.00 | 8.01E-01 |
| Monounsaturated fat intake (g/day) | Nutrients | 1799 | -0.01 | 0.01 | 5.82E-01 | 9.14E-01 | 152 | 1674 | 0.01 | 0.01 | 3.42E-01 | 8.88E-01 | 92 | 0.00 | 0.01 | 8.01E-01 | 0.13 | 2.85E-01 |
| Travel to work: Irregular travel mode to work vs passive travel to work | Physical activity | 1281 | 0.00 | 0.03 | 9.35E-01 | 9.95E-01 | 223 | 1235 | -0.01 | 0.03 | 7.90E-01 | 9.74E-01 | 188 | -0.01 | 0.02 | 8.09E-01 | 0.00 | 8.89E-01 |
| Pancake, waffle, Swedish dumpling | Food | 1799 | 0.00 | 0.01 | 8.16E-01 | 9.66E-01 | 202 | 1674 | 0.00 | 0.01 | 9.33E-01 | 9.89E-01 | 224 | 0.00 | 0.01 | 8.21E-01 | 0.00 | 9.22E-01 |
| Beta-sitosterol intake (mg/day) | Nutrients | 1799 | 0.01 | 0.01 | 4.54E-01 | 8.50E-01 | 127 | 1674 | -0.01 | 0.01 | 2.56E-01 | 7.68E-01 | 78 | 0.00 | 0.01 | 8.27E-01 | 0.45 | 1.79E-01 |
| Smoking status: Former occasional smokers vs non-smokers | Tobacco use | 1080 | 0.01 | 0.03 | 7.64E-01 | 9.37E-01 | 194 | 1033 | -0.02 | 0.03 | 5.18E-01 | 9.29E-01 | 128 | 0.00 | 0.02 | 8.29E-01 | 0.00 | 4.97E-01 |
| Rice | Food | 1799 | -0.01 | 0.01 | 5.43E-01 | 8.88E-01 | 144 | 1674 | 0.00 | 0.01 | 8.05E-01 | 9.74E-01 | 191 | 0.00 | 0.01 | 8.31E-01 | 0.00 | 5.34E-01 |
| Cream, creme fraiche, sour cream | Food | 1799 | 0.00 | 0.01 | 9.42E-01 | 9.95E-01 | 225 | 1674 | 0.00 | 0.01 | 7.00E-01 | 9.71E-01 | 167 | 0.00 | 0.01 | 8.50E-01 | 0.00 | 7.31E-01 |
| Possibility to speak with colleagues during breaks | Psychosocial | 1908 | 0.00 | 0.01 | 8.25E-01 | 9.71E-01 | 203 | 1792 | 0.00 | 0.01 | 9.64E-01 | 9.90E-01 | 231 | 0.00 | 0.01 | 8.50E-01 | 0.00 | 9.01E-01 |
| Control over own work assignment | Psychosocial | 1910 | 0.00 | 0.01 | 5.78E-01 | 9.14E-01 | 151 | 1795 | -0.01 | 0.01 | 4.03E-01 | 9.29E-01 | 101 | 0.00 | 0.01 | 8.51E-01 | 0.00 | 3.24E-01 |
| Stigmasterol intake (mg/day) | Nutrients | 1799 | 0.01 | 0.01 | 3.86E-01 | 8.18E-01 | 112 | 1674 | -0.01 | 0.01 | 5.09E-01 | 9.29E-01 | 125 | 0.00 | 0.01 | 8.51E-01 | 0.13 | 2.83E-01 |
| Cohabitation: Live alone vs Only one adult (spouse, partner) | Social | 615 | 0.01 | 0.03 | 6.44E-01 | 9.30E-01 | 162 | 609 | -0.03 | 0.03 | 4.17E-01 | 9.29E-01 | 105 | 0.00 | 0.02 | 8.52E-01 | 0.00 | 3.60E-01 |
| Satisfaction with work situation | Psychosocial | 1220 | 0.00 | 0.01 | 6.50E-01 | 9.30E-01 | 164 | 1165 | 0.00 | 0.01 | 8.18E-01 | 9.74E-01 | 196 | 0.00 | 0.01 | 8.65E-01 | 0.00 | 6.31E-01 |
| Berries (fresh or frozen) | Food | 1799 | 0.00 | 0.01 | 7.46E-01 | 9.35E-01 | 189 | 1674 | 0.00 | 0.01 | 6.05E-01 | 9.61E-01 | 150 | 0.00 | 0.01 | 8.69E-01 | 0.00 | 5.57E-01 |
| Ice cream | Food | 1799 | 0.00 | 0.01 | 9.30E-01 | 9.95E-01 | 221 | 1674 | 0.00 | 0.01 | 7.59E-01 | 9.74E-01 | 181 | 0.00 | 0.01 | 8.71E-01 | 0.00 | 7.83E-01 |
| Total energy intake (kcal/day) | Nutrients | 1799 | -0.01 | 0.01 | 4.22E-01 | 8.18E-01 | 123 | 1674 | 0.01 | 0.01 | 2.91E-01 | 7.99E-01 | 87 | 0.00 | 0.01 | 8.74E-01 | 0.42 | 1.88E-01 |
| Confidence status | Psychosocial | 1231 | 0.00 | 0.01 | 7.12E-01 | 9.35E-01 | 182 | 1186 | 0.00 | 0.01 | 8.72E-01 | 9.74E-01 | 213 | 0.00 | 0.01 | 8.78E-01 | 0.00 | 7.10E-01 |
| Frequency of hunting or fishing during leisure time | Physical activity | 1776 | 0.00 | 0.01 | 7.64E-01 | 9.37E-01 | 195 | 1685 | 0.00 | 0.01 | 9.00E-01 | 9.87E-01 | 217 | 0.00 | 0.01 | 9.01E-01 | 0.00 | 7.64E-01 |
| Sausage, liver pate on bread | Food | 1799 | 0.00 | 0.01 | 6.75E-01 | 9.30E-01 | 172 | 1674 | 0.01 | 0.01 | 5.59E-01 | 9.29E-01 | 142 | 0.00 | 0.01 | 9.15E-01 | 0.00 | 4.77E-01 |
| Sum of phytosterols intake (mg/day) | Nutrients | 1799 | 0.01 | 0.01 | 4.98E-01 | 8.88E-01 | 132 | 1674 | -0.01 | 0.01 | 3.86E-01 | 9.22E-01 | 100 | 0.00 | 0.01 | 9.27E-01 | 0.17 | 2.73E-01 |
| Patience status | Psychosocial | 1235 | 0.00 | 0.01 | 8.32E-01 | 9.75E-01 | 204 | 1185 | 0.00 | 0.01 | 7.55E-01 | 9.74E-01 | 179 | 0.00 | 0.01 | 9.46E-01 | 0.00 | 7.11E-01 |
| Frequency of gardening during leisure time | Physical activity | 1813 | 0.00 | 0.01 | 9.09E-01 | 9.95E-01 | 218 | 1686 | 0.00 | 0.01 | 9.74E-01 | 9.91E-01 | 235 | 0.00 | 0.01 | 9.51E-01 | 0.00 | 9.20E-01 |
| Vitamin A intake (mg/day) | Nutrients | 1799 | 0.01 | 0.01 | 6.06E-01 | 9.14E-01 | 158 | 1674 | -0.01 | 0.01 | 5.34E-01 | 9.29E-01 | 135 | 0.00 | 0.01 | 9.51E-01 | 0.00 | 4.21E-01 |
| Vitamin D intake (ug/day) | Nutrients | 1799 | 0.00 | 0.01 | 6.19E-01 | 9.19E-01 | 161 | 1674 | 0.01 | 0.01 | 5.48E-01 | 9.29E-01 | 139 | 0.00 | 0.01 | 9.53E-01 | 0.00 | 4.36E-01 |
| Satisfaction with accomodation | Psychosocial | 1235 | 0.00 | 0.01 | 8.68E-01 | 9.93E-01 | 208 | 1186 | 0.00 | 0.01 | 9.24E-01 | 9.89E-01 | 223 | 0.00 | 0.01 | 9.54E-01 | 0.00 | 8.55E-01 |
| Number of people with whom you can speak openly | Social | 1937 | 0.00 | 0.01 | 9.85E-01 | 9.95E-01 | 235 | 1823 | 0.00 | 0.01 | 9.87E-01 | 9.92E-01 | 237 | 0.00 | 0.01 | 9.81E-01 | 0.00 | 9.99E-01 |

**Supplementary Table 18**. Longitudinal association results for LDL cholesterol

| | | Training set | | | | | | Testing set | | | | | | Metanalysis | | | | |
|---|---|---|---|---|---|---|---|---|---|---|---|---|---|---|---|---|---|---|
| **Description** | **Group** | **N** | **Effect estimate** | **S.E.** | **p-value** | **p-value$_{FDR}$** | **p-value rank** | **N** | **Effect estimate** | **S.E.** | **p-value** | **p-value$_{FDR}$** | **p-value rank** | **Effect estimate** | **S.E.** | **p-value** | **$I^2$** | **Q p-value** |
| Snuff status: Snuff users vs non-snuff users | Tobacco use | 1594 | 0.23 | 0.05 | 1.53E-05 | **3.66E-03** | 1 | 1497 | 0.07 | 0.06 | 2.30E-01 | 8.09E-01 | 68 | 0.15 | 0.04 | 7.56E-05 | 0.78 | 3.22E-02 |
| Frequency of picking berries or mushrooms during leisure time | Physical activity | 1781 | -0.02 | 0.02 | 3.10E-01 | 7.40E-01 | 99 | 1658 | -0.10 | 0.02 | 1.61E-05 | **3.84E-03** | 1 | -0.06 | 0.02 | 1.74E-04 | 0.82 | 1.74E-02 |
| Years using snuff | Tobacco use | 1758 | 0.07 | 0.02 | 1.64E-04 | **1.97E-02** | 2 | 1624 | 0.03 | 0.02 | 1.58E-01 | 7.73E-01 | 49 | 0.05 | 0.01 | 1.86E-04 | 0.56 | 1.31E-01 |
| Number of snuff boxes per week | Tobacco use | 1819 | 0.07 | 0.02 | 2.91E-04 | **2.32E-02** | 3 | 1700 | 0.03 | 0.02 | 1.25E-01 | 7.30E-01 | 41 | 0.05 | 0.01 | 2.19E-04 | 0.47 | 1.71E-01 |
| Breakfast habits: Only coffee/tea for breakfast vs not breakfast at all | Food | 135 | 0.72 | 0.23 | 2.69E-03 | 1.61E-01 | 4 | 137 | 0.46 | 0.25 | 7.61E-02 | 6.09E-01 | 29 | 0.60 | 0.17 | 5.32E-04 | 0.00 | 4.47E-01 |
| Frequncy of social contacts with colleagues during leisure time | Psychosocial | 1822 | -0.04 | 0.02 | 7.24E-02 | 7.09E-01 | 21 | 1713 | -0.05 | 0.02 | 2.03E-02 | 4.42E-01 | 11 | -0.04 | 0.01 | 3.63E-03 | 0.00 | 6.84E-01 |
| Feel uneasy or guilty because of your way of drinking | Alcohol | 1694 | 0.14 | 0.06 | 3.02E-02 | 7.09E-01 | 9 | 1601 | 0.10 | 0.07 | 1.16E-01 | 7.18E-01 | 38 | 0.12 | 0.05 | 8.03E-03 | 0.00 | 7.01E-01 |
| Boiled coffee | Beverage | 1763 | 0.03 | 0.02 | 1.06E-01 | 7.09E-01 | 34 | 1642 | 0.04 | 0.02 | 5.66E-02 | 5.88E-01 | 23 | 0.03 | 0.01 | 1.28E-02 | 0.00 | 8.03E-01 |
| Vitamin C intake (mg/day) | Nutrients | 1763 | -0.04 | 0.02 | 9.93E-02 | 7.09E-01 | 32 | 1642 | -0.05 | 0.02 | 6.22E-02 | 6.09E-01 | 24 | -0.04 | 0.02 | 1.35E-02 | 0.00 | 7.50E-01 |
| Trans fat intake (g/day) | Nutrients | 1763 | -0.04 | 0.03 | 1.32E-01 | 7.17E-01 | 43 | 1642 | -0.05 | 0.03 | 6.54E-02 | 6.09E-01 | 25 | -0.05 | 0.02 | 1.83E-02 | 0.00 | 7.46E-01 |
| Smoking status: Former occasional smokers vs non-smokers | Tobacco use | 1058 | 0.13 | 0.08 | 9.49E-02 | 7.09E-01 | 31 | 1012 | 0.12 | 0.07 | 1.00E-01 | 6.84E-01 | 35 | 0.12 | 0.05 | 1.91E-02 | 0.00 | 9.50E-01 |
| Number of social contacts with the same interests as you | Social | 1896 | -0.03 | 0.02 | 1.69E-01 | 7.28E-01 | 52 | 1787 | -0.04 | 0.02 | 5.63E-02 | 5.88E-01 | 22 | -0.03 | 0.01 | 2.06E-02 | 0.00 | 6.71E-01 |
| Learn new things at job | Psychosocial | 1871 | -0.01 | 0.02 | 7.18E-01 | 8.95E-01 | 191 | 1759 | -0.06 | 0.02 | 3.00E-03 | 1.43E-01 | 5 | -0.03 | 0.01 | 2.16E-02 | 0.73 | 5.48E-02 |
| Number of cigarretes smoked per day (in groups) | Tobacco use | 1560 | 0.00 | 0.02 | 8.31E-01 | 9.28E-01 | 214 | 1484 | -0.07 | 0.02 | 5.31E-04 | 5.90E-02 | 2 | -0.03 | 0.01 | 2.32E-02 | 0.86 | 8.38E-03 |
| Ingenuity or creativity demand from job | Psychosocial | 1873 | -0.03 | 0.02 | 1.80E-01 | 7.28E-01 | 55 | 1761 | -0.04 | 0.02 | 8.16E-02 | 6.09E-01 | 32 | -0.03 | 0.01 | 3.03E-02 | 0.00 | 7.07E-01 |
| Educational level | Psychosocial | 1899 | 0.04 | 0.02 | 2.77E-02 | 7.09E-01 | 8 | 1794 | 0.02 | 0.02 | 4.42E-01 | 9.10E-01 | 116 | 0.03 | 0.01 | 3.26E-02 | 0.00 | 3.49E-01 |
| Travel to work: Irregular travel mode to work vs passive travel to work | Physical activity | 1259 | -0.08 | 0.07 | 2.67E-01 | 7.40E-01 | 85 | 1211 | -0.13 | 0.07 | 7.86E-02 | 6.09E-01 | 30 | -0.10 | 0.05 | 4.30E-02 | 0.00 | 6.26E-01 |
| Possibility to leave your work for a while to speak with a colleague | Psychosocial | 1864 | -0.02 | 0.02 | 4.10E-01 | 7.98E-01 | 122 | 1744 | -0.04 | 0.02 | 4.11E-02 | 5.64E-01 | 15 | -0.03 | 0.01 | 4.37E-02 | 0.00 | 3.75E-01 |
| Boiled or baked potato | Food | 1763 | -0.04 | 0.02 | 6.05E-02 | 7.09E-01 | 17 | 1642 | -0.02 | 0.02 | 3.95E-01 | 9.10E-01 | 103 | -0.03 | 0.01 | 5.12E-02 | 0.00 | 5.02E-01 |
| Grams of tobacco smoked per week | Tobacco use | 1122 | 0.08 | 0.04 | 9.21E-02 | 7.09E-01 | 28 | 1056 | 0.05 | 0.04 | 2.88E-01 | 8.29E-01 | 81 | 0.06 | 0.03 | 5.26E-02 | 0.00 | 6.40E-01 |
| Number of cigarretes smoked per day | Tobacco use | 1122 | 0.08 | 0.04 | 9.21E-02 | 7.09E-01 | 29 | 1056 | 0.05 | 0.04 | 2.88E-01 | 8.29E-01 | 82 | 0.06 | 0.03 | 5.26E-02 | 0.00 | 6.40E-01 |
| Number of cigars smoked per day | Tobacco use | 1122 | 0.08 | 0.04 | 9.21E-02 | 7.09E-01 | 30 | 1056 | 0.05 | 0.04 | 2.88E-01 | 8.29E-01 | 83 | 0.06 | 0.03 | 5.26E-02 | 0.00 | 6.40E-01 |
| Potato salad | Food | 640 | -0.06 | 0.03 | 7.00E-02 | 7.09E-01 | 20 | 567 | -0.03 | 0.04 | 4.17E-01 | 9.10E-01 | 107 | -0.05 | 0.03 | 5.55E-02 | 0.00 | 5.92E-01 |
| Energy status | Psychosocial | 1209 | 0.00 | 0.03 | 9.57E-01 | 9.77E-01 | 234 | 1165 | -0.07 | 0.02 | 8.64E-03 | 3.44E-01 | 6 | -0.03 | 0.02 | 5.70E-02 | 0.70 | 6.93E-02 |
| Job demands to work very fast | Psychosocial | 1872 | -0.02 | 0.02 | 2.26E-01 | 7.28E-01 | 73 | 1763 | -0.03 | 0.02 | 1.53E-01 | 7.64E-01 | 48 | -0.03 | 0.01 | 6.27E-02 | 0.00 | 8.40E-01 |
| Tiamin intake (mg/day) | Nutrients | 1763 | 0.03 | 0.02 | 1.95E-01 | 7.28E-01 | 62 | 1642 | 0.03 | 0.02 | 1.84E-01 | 7.87E-01 | 55 | 0.03 | 0.02 | 6.34E-02 | 0.00 | 9.67E-01 |
| Medium beer | Alcohol | 1763 | 0.02 | 0.02 | 1.99E-01 | 7.28E-01 | 63 | 1642 | 0.03 | 0.02 | 1.82E-01 | 7.87E-01 | 53 | 0.03 | 0.02 | 6.66E-02 | 0.00 | 7.92E-01 |
| Whole grain soft bread | Food | 1763 | -0.03 | 0.02 | 8.02E-02 | 7.09E-01 | 26 | 1642 | -0.02 | 0.02 | 4.41E-01 | 9.10E-01 | 115 | -0.03 | 0.01 | 6.85E-02 | 0.00 | 5.60E-01 |
| Sour milk, yoghurt (low fat) | Food | 1763 | 0.05 | 0.02 | 2.12E-02 | 7.09E-01 | 6 | 1642 | 0.00 | 0.02 | 8.93E-01 | 9.83E-01 | 214 | 0.03 | 0.01 | 7.29E-02 | 0.53 | 1.46E-01 |
| Bacon | Food | 1763 | -0.03 | 0.02 | 6.59E-02 | 7.09E-01 | 19 | 1642 | -0.01 | 0.02 | 5.55E-01 | 9.46E-01 | 138 | -0.02 | 0.01 | 7.51E-02 | 0.00 | 4.52E-01 |
| Control over planning and execution of the workday | Psychosocial | 1878 | -0.02 | 0.02 | 4.39E-01 | 8.07E-01 | 130 | 1765 | -0.04 | 0.02 | 7.92E-02 | 6.09E-01 | 31 | -0.03 | 0.01 | 7.56E-02 | 0.00 | 4.69E-01 |
| Vitamin B2 intake (ug/day) | Nutrients | 1763 | 0.03 | 0.02 | 1.16E-01 | 7.09E-01 | 37 | 1642 | 0.02 | 0.02 | 3.72E-01 | 9.09E-01 | 97 | 0.03 | 0.01 | 7.83E-02 | 0.00 | 6.80E-01 |
| Last time a colleague visited you at home | Psychosocial | 1859 | -0.02 | 0.02 | 2.39E-01 | 7.28E-01 | 77 | 1737 | -0.03 | 0.02 | 1.90E-01 | 7.87E-01 | 57 | -0.02 | 0.01 | 7.87E-02 | 0.00 | 9.12E-01 |
| Apple, pear, peach, orange, mandarin and grapefruit | Food | 1763 | -0.04 | 0.02 | 5.50E-02 | 7.09E-01 | 15 | 1642 | -0.01 | 0.02 | 6.73E-01 | 9.77E-01 | 160 | -0.03 | 0.02 | 8.97E-02 | 0.00 | 3.21E-01 |
| Cookies, pastry | Food | 1763 | 0.02 | 0.02 | 1.82E-01 | 7.28E-01 | 56 | 1642 | 0.02 | 0.02 | 3.56E-01 | 9.09E-01 | 90 | 0.02 | 0.01 | 1.07E-01 | 0.00 | 8.53E-01 |
| Sometimes physically straining work | Physical activity | 1871 | -0.08 | 0.04 | 6.22E-02 | 7.09E-01 | 18 | 1755 | -0.02 | 0.04 | 7.24E-01 | 9.77E-01 | 175 | -0.05 | 0.03 | 1.11E-01 | 0.06 | 3.02E-01 |
| Pasta | Food | 1763 | 0.01 | 0.02 | 6.93E-01 | 8.95E-01 | 185 | 1642 | 0.04 | 0.02 | 7.20E-02 | 6.09E-01 | 27 | 0.02 | 0.01 | 1.19E-01 | 0.00 | 3.25E-01 |
| Disaccharides intake (g/day) | Nutrients | 1763 | 0.05 | 0.02 | 2.69E-02 | 7.09E-01 | 7 | 1642 | 0.00 | 0.02 | 9.68E-01 | 9.97E-01 | 232 | 0.02 | 0.02 | 1.20E-01 | 0.60 | 1.15E-01 |
| Vision status | General health | 1214 | -0.02 | 0.03 | 5.15E-01 | 8.44E-01 | 146 | 1167 | -0.04 | 0.03 | 1.16E-01 | 7.18E-01 | 37 | -0.03 | 0.02 | 1.20E-01 | 0.00 | 4.87E-01 |
| Strong beer | Alcohol | 1763 | 0.03 | 0.02 | 1.21E-01 | 7.09E-01 | 38 | 1642 | 0.01 | 0.03 | 6.30E-01 | 9.77E-01 | 150 | 0.02 | 0.01 | 1.22E-01 | 0.00 | 6.16E-01 |
| Cohabitation: Live alone vs Only children | Social | 248 | 0.28 | 0.11 | 1.63E-02 | 7.09E-01 | 5 | 250 | -0.05 | 0.13 | 6.93E-01 | 9.77E-01 | 166 | 0.13 | 0.09 | 1.23E-01 | 0.72 | 5.67E-02 |
| Soft whey cheese | Food | 640 | -0.04 | 0.03 | 1.29E-01 | 7.17E-01 | 42 | 567 | -0.02 | 0.05 | 6.82E-01 | 9.77E-01 | 164 | -0.04 | 0.02 | 1.25E-01 | 0.00 | 7.24E-01 |
| Informed of having high blood pressure | General health | 1904 | -0.07 | 0.05 | 1.85E-01 | 7.28E-01 | 57 | 1791 | -0.05 | 0.06 | 4.19E-01 | 9.10E-01 | 109 | -0.06 | 0.04 | 1.29E-01 | 0.00 | 7.37E-01 |
| Vitamin A intake (mg/day) | Nutrients | 1763 | -0.01 | 0.03 | 7.10E-01 | 8.95E-01 | 188 | 1642 | -0.05 | 0.03 | 7.44E-02 | 6.09E-01 | 28 | -0.03 | 0.02 | 1.32E-01 | 0.05 | 3.05E-01 |
| Monosaccharides intake (g/day) | Nutrients | 1763 | -0.01 | 0.02 | 5.03E-01 | 8.36E-01 | 144 | 1642 | -0.03 | 0.02 | 1.51E-01 | 7.64E-01 | 46 | -0.02 | 0.02 | 1.43E-01 | 0.00 | 5.46E-01 |
| Tea | Beverage | 1763 | -0.04 | 0.02 | 7.32E-02 | 7.09E-01 | 22 | 1642 | 0.00 | 0.02 | 9.31E-01 | 9.85E-01 | 226 | -0.02 | 0.02 | 1.57E-01 | 0.18 | 2.70E-01 |
| Oil for cooking | Food | 1763 | 0.04 | 0.02 | 5.02E-02 | 7.09E-01 | 14 | 1642 | 0.00 | 0.02 | 9.56E-01 | 9.93E-01 | 230 | 0.02 | 0.01 | 1.64E-01 | 0.47 | 1.68E-01 |
| Years smoking | Tobacco use | 1678 | 0.03 | 0.02 | 2.35E-01 | 7.28E-01 | 76 | 1586 | -0.07 | 0.02 | 1.40E-03 | 8.35E-02 | 4 | -0.02 | 0.02 | 1.68E-01 | 0.90 | 1.78E-03 |
| Ice cream | Food | 1763 | -0.01 | 0.02 | 5.74E-01 | 8.53E-01 | 160 | 1642 | -0.03 | 0.02 | 1.73E-01 | 7.87E-01 | 50 | -0.02 | 0.02 | 1.69E-01 | 0.00 | 5.92E-01 |
| Light and physically active work | Physical activity | 1871 | 0.02 | 0.05 | 6.24E-01 | 8.74E-01 | 169 | 1755 | 0.07 | 0.05 | 1.38E-01 | 7.48E-01 | 44 | 0.05 | 0.03 | 1.70E-01 | 0.00 | 4.51E-01 |
| Memory status | Psychosocial | 1204 | 0.00 | 0.03 | 8.71E-01 | 9.51E-01 | 219 | 1163 | -0.05 | 0.02 | 4.12E-02 | 5.64E-01 | 16 | -0.02 | 0.02 | 1.73E-01 | 0.57 | 1.26E-01 |
| Phosphate intake (mg/day) | Nutrients | 1763 | 0.02 | 0.02 | 2.99E-01 | 7.40E-01 | 93 | 1642 | 0.02 | 0.02 | 3.88E-01 | 9.09E-01 | 102 | 0.02 | 0.02 | 1.78E-01 | 0.00 | 9.34E-01 |
| Smoking status: Smokers vs non-smokers | Tobacco use | 1246 | 0.08 | 0.06 | 1.36E-01 | 7.17E-01 | 44 | 1197 | -0.19 | 0.06 | 7.40E-04 | 5.90E-02 | 3 | -0.05 | 0.04 | 1.79E-01 | 0.92 | 5.71E-04 |
| Margarine for cooking | Food | 1763 | -0.01 | 0.02 | 4.91E-01 | 8.26E-01 | 139 | 1642 | -0.02 | 0.02 | 2.26E-01 | 8.09E-01 | 64 | -0.02 | 0.01 | 1.81E-01 | 0.00 | 6.96E-01 |
| Fibre intake (g/day) | Nutrients | 1763 | -0.03 | 0.02 | 2.06E-01 | 7.28E-01 | 64 | 1642 | -0.01 | 0.02 | 5.58E-01 | 9.46E-01 | 140 | -0.02 | 0.02 | 1.82E-01 | 0.00 | 6.86E-01 |
| Fiber cereals | Food | 1763 | 0.02 | 0.02 | 2.90E-01 | 7.40E-01 | 92 | 1642 | 0.02 | 0.02 | 4.31E-01 | 9.10E-01 | 111 | 0.02 | 0.02 | 1.89E-01 | 0.00 | 8.96E-01 |
| Blota (broth + bread) | Food | 640 | -0.07 | 0.04 | 5.65E-02 | 7.09E-01 | 16 | 567 | 0.01 | 0.04 | 8.68E-01 | 9.83E-01 | 206 | -0.03 | 0.03 | 1.90E-01 | 0.49 | 1.62E-01 |
| Pinoresinol intake (ug/day) | Nutrients | 1763 | -0.02 | 0.02 | 3.66E-01 | 7.89E-01 | 111 | 1642 | -0.02 | 0.02 | 3.57E-01 | 9.09E-01 | 92 | -0.02 | 0.01 | 1.98E-01 | 0.00 | 9.52E-01 |
| Marital status: Single vs Widow/widower | Social | 216 | -0.54 | 0.31 | 7.70E-02 | 7.09E-01 | 24 | 225 | -0.05 | 0.25 | 8.43E-01 | 9.83E-01 | 201 | -0.24 | 0.19 | 2.03E-01 | 0.37 | 2.09E-01 |
| Everyday exercise satisfaction | Physical activity | 1900 | -0.02 | 0.02 | 4.28E-01 | 8.07E-01 | 126 | 1788 | -0.02 | 0.02 | 3.15E-01 | 8.84E-01 | 85 | -0.02 | 0.01 | 2.04E-01 | 0.00 | 8.64E-01 |
| Parents or siblings had a cerebral hemorrhage/thrombosis or cardiac infarction before the age of 60 | General health | 1887 | 0.07 | 0.05 | 1.42E-01 | 7.17E-01 | 47 | 1771 | 0.02 | 0.05 | 7.60E-01 | 9.77E-01 | 186 | 0.05 | 0.04 | 2.05E-01 | 0.00 | 4.20E-01 |
| Tomato, cucumber | Food | 1763 | 0.04 | 0.02 | 1.07E-01 | 7.09E-01 | 35 | 1642 | 0.00 | 0.02 | 8.67E-01 | 9.83E-01 | 205 | 0.02 | 0.02 | 2.09E-01 | 0.05 | 3.04E-01 |
| Milk, sour milk (1,5%) | Beverage | 1763 | 0.01 | 0.02 | 6.31E-01 | 8.74E-01 | 171 | 1642 | 0.03 | 0.02 | 1.95E-01 | 7.87E-01 | 59 | 0.02 | 0.01 | 2.14E-01 | 0.00 | 5.44E-01 |
| Liver, kidney | Food | 640 | -0.03 | 0.03 | 2.66E-01 | 7.40E-01 | 84 | 567 | -0.02 | 0.04 | 5.52E-01 | 9.46E-01 | 137 | -0.03 | 0.02 | 2.15E-01 | 0.00 | 8.16E-01 |
| Hearing status | General health | 1212 | -0.01 | 0.03 | 6.73E-01 | 8.84E-01 | 182 | 1168 | -0.03 | 0.03 | 1.88E-01 | 7.87E-01 | 56 | -0.02 | 0.02 | 2.17E-01 | 0.00 | 5.33E-01 |
| White (soft) bread, thin crisp bread | Food | 1763 | 0.00 | 0.02 | 9.76E-01 | 9.91E-01 | 235 | 1642 | 0.04 | 0.02 | 6.94E-02 | 6.09E-01 | 26 | 0.02 | 0.02 | 2.18E-01 | 0.44 | 1.81E-01 |
| Lariciresinol intake (ug/day) | Nutrients | 1763 | -0.03 | 0.02 | 1.92E-01 | 7.28E-01 | 60 | 1642 | -0.01 | 0.02 | 7.04E-01 | 9.77E-01 | 170 | -0.02 | 0.01 | 2.27E-01 | 0.00 | 5.33E-01 |
| Meat on bread | Food | 1763 | 0.03 | 0.02 | 7.87E-02 | 7.09E-01 | 25 | 1642 | 0.00 | 0.02 | 8.17E-01 | 9.83E-01 | 198 | 0.02 | 0.01 | 2.28E-01 | 0.41 | 1.93E-01 |
| Satisfaction with work situation | Psychosocial | 1194 | -0.02 | 0.02 | 3.61E-01 | 7.87E-01 | 109 | 1146 | -0.02 | 0.02 | 4.34E-01 | 9.10E-01 | 113 | -0.02 | 0.02 | 2.30E-01 | 0.00 | 9.34E-01 |
| If you exercise, change in exercise habits during the last year | Physical activity | 1661 | -0.01 | 0.02 | 7.65E-01 | 9.08E-01 | 201 | 1591 | 0.04 | 0.02 | 4.48E-02 | 5.64E-01 | 18 | 0.02 | 0.02 | 2.37E-01 | 0.63 | 9.88E-02 |
| Exercise during the last three months | Physical activity | 1901 | 0.03 | 0.02 | 1.49E-01 | 7.26E-01 | 49 | 1791 | 0.00 | 0.02 | 8.76E-01 | 9.83E-01 | 210 | 0.02 | 0.01 | 2.42E-01 | 0.00 | 3.90E-01 |
| Fried potatoes, pommes frites | Food | 1763 | -0.02 | 0.01 | 2.25E-01 | 7.28E-01 | 71 | 1642 | -0.01 | 0.02 | 7.06E-01 | 9.77E-01 | 171 | -0.01 | 0.01 | 2.44E-01 | 0.00 | 6.07E-01 |
| Vitamin E intake (mg/day) | Nutrients | 1763 | -0.01 | 0.02 | 7.50E-01 | 9.05E-01 | 198 | 1642 | -0.03 | 0.02 | 1.75E-01 | 7.87E-01 | 51 | -0.02 | 0.01 | 2.52E-01 | 0.00 | 4.27E-01 |
| Whole grain intake (g/day) | Food | 1763 | 0.01 | 0.02 | 5.98E-01 | 8.74E-01 | 163 | 1642 | 0.02 | 0.02 | 2.78E-01 | 8.29E-01 | 76 | 0.02 | 0.01 | 2.60E-01 | 0.00 | 6.62E-01 |
| Frequency of dancing during leisure time | Physical activity | 1632 | 0.03 | 0.02 | 1.90E-01 | 7.28E-01 | 59 | 1516 | 0.01 | 0.02 | 8.11E-01 | 9.83E-01 | 196 | 0.02 | 0.02 | 2.72E-01 | 0.00 | 4.51E-01 |
| Marital status: Single vs Divorced/separated | Social | 315 | 0.05 | 0.09 | 6.03E-01 | 8.74E-01 | 164 | 318 | 0.12 | 0.11 | 2.80E-01 | 8.29E-01 | 77 | 0.08 | 0.07 | 2.72E-01 | 0.00 | 6.26E-01 |
| Breakfast habits: Porridge w/o sandwich for breakfast vs not breakfast at all | Food | 268 | -0.18 | 0.11 | 1.10E-01 | 7.09E-01 | 36 | 249 | 0.02 | 0.12 | 8.68E-01 | 9.83E-01 | 207 | -0.09 | 0.08 | 2.74E-01 | 0.29 | 2.35E-01 |
| Snuff status: Former snuff users vs non-snuff users | Tobacco use | 1420 | 0.09 | 0.06 | 1.40E-01 | 7.17E-01 | 46 | 1313 | 0.00 | 0.07 | 9.83E-01 | 9.97E-01 | 235 | 0.05 | 0.05 | 2.83E-01 | 0.03 | 3.11E-01 |

| | | | | | | | | | | | | | | | | | |
|---|---|---|---|---|---|---|---|---|---|---|---|---|---|---|---|---|---|
| Cohabitation: Live alone vs Other/others | Social | 231 | -0.05 | 0.12 | 7.11E-01 | 8.95E-01 | 190 | 191 | -0.24 | 0.18 | 1.83E-01 | 7.87E-01 | 54 | -0.11 | 0.10 | 2.85E-01 | 0.00 | 3.77E-01 |
| Work shifts/weekends | Psychosocial | 1840 | -0.04 | 0.04 | 3.06E-01 | 7.40E-01 | 98 | 1730 | 0.12 | 0.04 | 1.01E-02 | 3.45E-01 | 7 | 0.03 | 0.03 | 2.86E-01 | 0.85 | 1.05E-02 |
| Sum of Lariciresinol, Matairesinol, Pinoresinol, Secoisolariciresinol intake (ug/day) | Nutrients | 1763 | -0.02 | 0.02 | 3.51E-01 | 7.84E-01 | 107 | 1642 | -0.01 | 0.02 | 5.75E-01 | 9.48E-01 | 145 | -0.02 | 0.01 | 2.88E-01 | 0.00 | 8.15E-01 |
| Cheese 28% | Food | 1763 | -0.01 | 0.02 | 5.65E-01 | 8.51E-01 | 158 | 1642 | -0.02 | 0.02 | 3.59E-01 | 9.09E-01 | 93 | -0.02 | 0.02 | 2.93E-01 | 0.00 | 7.96E-01 |
| Control over own work assignment | Psychosocial | 1871 | 0.01 | 0.02 | 5.26E-01 | 8.49E-01 | 148 | 1763 | -0.04 | 0.02 | 3.25E-02 | 5.64E-01 | 12 | -0.01 | 0.01 | 2.97E-01 | 0.74 | 4.84E-02 |
| Frequency of gardening during leisure time | Physical activity | 1777 | 0.01 | 0.02 | 6.26E-01 | 8.74E-01 | 170 | 1655 | -0.04 | 0.02 | 4.72E-02 | 5.64E-01 | 20 | -0.02 | 0.02 | 2.98E-01 | 0.68 | 7.84E-02 |
| Receive hugs to comfort and support you | Social | 1887 | -0.06 | 0.05 | 2.56E-01 | 7.36E-01 | 83 | 1773 | -0.02 | 0.05 | 7.59E-01 | 9.77E-01 | 184 | -0.04 | 0.04 | 3.00E-01 | 0.00 | 5.77E-01 |
| Banana | Food | 1763 | 0.01 | 0.02 | 5.59E-01 | 8.51E-01 | 157 | 1642 | 0.02 | 0.02 | 3.87E-01 | 9.09E-01 | 100 | 0.02 | 0.02 | 3.12E-01 | 0.00 | 7.98E-01 |
| Satisfaction with accomodation | Psychosocial | 1209 | 0.05 | 0.02 | 4.81E-02 | 7.09E-01 | 13 | 1166 | -0.02 | 0.03 | 5.27E-01 | 9.46E-01 | 132 | 0.02 | 0.02 | 3.13E-01 | 0.70 | 6.94E-02 |
| Iodine intake (ug/day) | Nutrients | 1763 | 0.00 | 0.02 | 8.51E-01 | 9.42E-01 | 216 | 1642 | 0.03 | 0.02 | 2.06E-01 | 7.87E-01 | 61 | 0.01 | 0.01 | 3.20E-01 | 0.00 | 4.23E-01 |
| Brown beans, pea soup | Food | 1763 | 0.05 | 0.02 | 3.57E-02 | 7.09E-01 | 11 | 1642 | -0.02 | 0.03 | 4.49E-01 | 9.10E-01 | 118 | 0.02 | 0.02 | 3.20E-01 | 0.75 | 4.54E-02 |
| Secoisolariciresinol intake (ug/day) | Nutrients | 1763 | 0.02 | 0.02 | 2.78E-01 | 7.40E-01 | 88 | 1642 | 0.01 | 0.02 | 7.68E-01 | 9.82E-01 | 187 | 0.01 | 0.01 | 3.28E-01 | 0.00 | 5.81E-01 |
| Overall state of health during the last year | General health | 1901 | 0.01 | 0.02 | 4.92E-01 | 8.26E-01 | 140 | 1790 | -0.04 | 0.02 | 3.73E-02 | 5.64E-01 | 13 | -0.01 | 0.01 | 3.34E-01 | 0.74 | 4.88E-02 |
| Carbohydrates intake (g/day) | Nutrients | 1763 | -0.02 | 0.02 | 2.44E-01 | 7.28E-01 | 80 | 1642 | 0.00 | 0.02 | 8.83E-01 | 9.83E-01 | 211 | -0.01 | 0.01 | 3.43E-01 | 0.00 | 4.88E-01 |
| Medioresinol intake (ug/day) | Nutrients | 1763 | -0.02 | 0.02 | 2.41E-01 | 7.28E-01 | 79 | 1642 | 0.00 | 0.02 | 9.01E-01 | 9.83E-01 | 216 | -0.01 | 0.01 | 3.44E-01 | 0.00 | 4.81E-01 |
| Breakfast habits: Coffee/tea and wheat buns or rusk for breakfast vs not breakfast at all | Food | 177 | -0.16 | 0.13 | 2.20E-01 | 7.28E-01 | 69 | 170 | 0.01 | 0.17 | 9.36E-01 | 9.86E-01 | 227 | -0.10 | 0.10 | 3.51E-01 | 0.00 | 4.21E-01 |
| Self-employed | Psychosocial | 1867 | -0.05 | 0.06 | 4.07E-01 | 7.98E-01 | 120 | 1770 | -0.03 | 0.07 | 6.65E-01 | 9.77E-01 | 158 | -0.04 | 0.05 | 3.68E-01 | 0.00 | 8.01E-01 |
| Magnesium intake (mg/day) | Nutrients | 1763 | 0.01 | 0.02 | 6.47E-01 | 8.74E-01 | 177 | 1642 | 0.02 | 0.02 | 4.17E-01 | 9.10E-01 | 106 | 0.01 | 0.02 | 3.73E-01 | 0.00 | 7.82E-01 |
| Pizza | Food | 1763 | 0.00 | 0.02 | 9.40E-01 | 9.76E-01 | 230 | 1642 | -0.03 | 0.02 | 1.99E-01 | 7.87E-01 | 60 | -0.01 | 0.02 | 3.82E-01 | 0.00 | 3.45E-01 |
| Zinc intake (mg/day) | Nutrients | 1763 | -0.01 | 0.02 | 5.35E-01 | 8.49E-01 | 149 | 1642 | -0.01 | 0.02 | 5.39E-01 | 9.46E-01 | 134 | -0.01 | 0.02 | 3.83E-01 | 0.00 | 9.77E-01 |
| Linoleic acid intake (g/day) | Nutrients | 1763 | 0.03 | 0.02 | 1.37E-01 | 7.17E-01 | 45 | 1642 | -0.01 | 0.02 | 7.37E-01 | 9.77E-01 | 177 | 0.01 | 0.01 | 3.86E-01 | 0.37 | 2.09E-01 |
| Sum of all lignans intake (ug/day) | Nutrients | 1763 | -0.02 | 0.02 | 2.84E-01 | 7.40E-01 | 90 | 1642 | 0.00 | 0.02 | 9.04E-01 | 9.83E-01 | 217 | -0.01 | 0.01 | 3.86E-01 | 0.00 | 5.22E-01 |
| Frequency of cycling during leisure time | Physical activity | 1553 | 0.02 | 0.02 | 4.38E-01 | 8.07E-01 | 128 | 1447 | -0.05 | 0.02 | 4.00E-02 | 5.64E-01 | 14 | -0.01 | 0.02 | 3.88E-01 | 0.76 | 4.33E-02 |
| Beta-sitostanol intake (mg/day) | Nutrients | 1763 | -0.02 | 0.02 | 2.53E-01 | 7.36E-01 | 82 | 1642 | 0.00 | 0.02 | 9.85E-01 | 9.97E-01 | 236 | -0.01 | 0.01 | 3.92E-01 | 0.00 | 4.48E-01 |
| Matairesinol intake (ug/day) | Nutrients | 1763 | -0.02 | 0.02 | 3.56E-01 | 7.87E-01 | 108 | 1642 | -0.01 | 0.02 | 7.99E-01 | 9.83E-01 | 192 | -0.01 | 0.01 | 3.94E-01 | 0.00 | 6.62E-01 |
| Salad dressing with oil | Food | 1763 | -0.02 | 0.02 | 3.06E-01 | 7.40E-01 | 97 | 1642 | 0.00 | 0.02 | 9.06E-01 | 9.83E-01 | 219 | -0.01 | 0.01 | 4.00E-01 | 0.00 | 5.51E-01 |
| Changed everyday exercise during the last year | Physical activity | 1904 | 0.02 | 0.02 | 3.94E-01 | 7.98E-01 | 116 | 1791 | 0.01 | 0.02 | 7.45E-01 | 9.77E-01 | 179 | 0.01 | 0.01 | 4.00E-01 | 0.00 | 7.23E-01 |
| Travel to work: Cycle to work vs passive travel to work | Physical activity | 1397 | 0.00 | 0.05 | 9.78E-01 | 9.91E-01 | 236 | 1320 | -0.07 | 0.06 | 2.29E-01 | 8.09E-01 | 67 | -0.03 | 0.04 | 4.04E-01 | 0.00 | 3.86E-01 |
| Skill demand from job | Psychosocial | 1870 | 0.00 | 0.02 | 8.36E-01 | 9.30E-01 | 215 | 1760 | -0.02 | 0.02 | 3.28E-01 | 8.95E-01 | 86 | -0.01 | 0.01 | 4.11E-01 | 0.00 | 5.69E-01 |
| Chips, popcorn, salted nuts | Food | 1763 | 0.02 | 0.02 | 4.77E-01 | 8.26E-01 | 136 | 1642 | 0.01 | 0.02 | 6.40E-01 | 9.77E-01 | 151 | 0.01 | 0.01 | 4.15E-01 | 0.00 | 8.07E-01 |
| Animal based protein intake (g/day) | Nutrients | 1763 | 0.02 | 0.02 | 2.78E-01 | 7.40E-01 | 87 | 1642 | 0.00 | 0.02 | 1.00E+00 | 1.00E+00 | 239 | 0.01 | 0.01 | 4.16E-01 | 0.00 | 4.72E-01 |
| Possibility to speak with colleagues during breaks | Psychosocial | 1870 | 0.03 | 0.02 | 7.64E-02 | 7.09E-01 | 23 | 1760 | -0.01 | 0.02 | 4.89E-01 | 9.27E-01 | 126 | 0.01 | 0.01 | 4.38E-01 | 0.67 | 8.22E-02 |
| Frequency of shoverling snow during leisure time | Physical activity | 1801 | 0.00 | 0.02 | 8.80E-01 | 9.56E-01 | 220 | 1693 | -0.03 | 0.02 | 2.07E-01 | 7.87E-01 | 62 | -0.01 | 0.02 | 4.39E-01 | 0.02 | 3.13E-01 |
| Frequency of walking during leisure time | Physical activity | 1832 | 0.00 | 0.02 | 9.35E-01 | 9.76E-01 | 228 | 1729 | -0.02 | 0.02 | 3.04E-01 | 8.66E-01 | 84 | -0.01 | 0.01 | 4.41E-01 | 0.00 | 4.94E-01 |
| Feel the need to reduce alcohol consumption | Alcohol | 1695 | 0.06 | 0.06 | 3.24E-01 | 7.52E-01 | 102 | 1602 | 0.01 | 0.07 | 9.28E-01 | 9.85E-01 | 224 | 0.04 | 0.05 | 4.41E-01 | 0.00 | 5.33E-01 |
| Palmitic acid intake (g/day) | Nutrients | 1763 | 0.01 | 0.02 | 4.56E-01 | 8.26E-01 | 132 | 1642 | 0.01 | 0.02 | 7.51E-01 | 9.77E-01 | 182 | 0.01 | 0.01 | 4.43E-01 | 0.00 | 7.93E-01 |
| Campestanol intake (mg/day) | Nutrients | 1763 | -0.03 | 0.02 | 1.85E-01 | 7.28E-01 | 58 | 1642 | 0.01 | 0.02 | 7.50E-01 | 9.77E-01 | 180 | -0.01 | 0.01 | 4.47E-01 | 0.22 | 2.58E-01 |
| Mood status | Psychosocial | 1209 | 0.02 | 0.03 | 3.87E-01 | 7.98E-01 | 114 | 1161 | -0.05 | 0.03 | 5.23E-02 | 5.88E-01 | 21 | -0.01 | 0.02 | 4.50E-01 | 0.75 | 4.68E-02 |
| Arachidonic acid (ARA) intake (g/day) | Nutrients | 1763 | 0.01 | 0.02 | 6.23E-01 | 8.74E-01 | 168 | 1642 | 0.01 | 0.02 | 5.72E-01 | 9.48E-01 | 144 | 0.01 | 0.01 | 4.55E-01 | 0.00 | 9.52E-01 |
| Travel to work: Walk to work vs passive travel to work | Physical activity | 1267 | -0.05 | 0.07 | 4.11E-01 | 7.98E-01 | 123 | 1201 | -0.01 | 0.07 | 8.45E-01 | 9.83E-01 | 202 | -0.04 | 0.05 | 4.61E-01 | 0.00 | 6.77E-01 |
| Do you feel important and appreciated in your home? | Psychosocial | 1201 | 0.01 | 0.03 | 6.59E-01 | 8.75E-01 | 180 | 1160 | -0.04 | 0.03 | 1.51E-01 | 7.64E-01 | 47 | -0.01 | 0.02 | 4.63E-01 | 0.42 | 1.90E-01 |
| Soft cheese | Food | 640 | -0.02 | 0.03 | 6.47E-01 | 8.74E-01 | 175 | 567 | -0.02 | 0.04 | 5.57E-01 | 9.46E-01 | 139 | -0.02 | 0.02 | 4.64E-01 | 0.00 | 8.93E-01 |
| Average portion size of vegetables based on photographic illustration of four sizes (smallest to largest) | Food | 1763 | -0.02 | 0.02 | 2.50E-01 | 7.36E-01 | 81 | 1642 | 0.00 | 0.02 | 8.76E-01 | 9.83E-01 | 209 | -0.01 | 0.02 | 4.68E-01 | 0.00 | 3.65E-01 |
| Light but partly physically active work | Physical activity | 1871 | 0.07 | 0.05 | 1.94E-01 | 7.28E-01 | 61 | 1755 | -0.13 | 0.05 | 1.74E-02 | 4.16E-01 | 10 | -0.03 | 0.04 | 4.70E-01 | 0.85 | 8.95E-03 |
| Pancake, waffle, Swedish dumpling | Food | 1763 | -0.01 | 0.02 | 5.44E-01 | 8.49E-01 | 152 | 1642 | 0.04 | 0.02 | 8.90E-02 | 6.44E-01 | 33 | 0.01 | 0.01 | 4.72E-01 | 0.64 | 9.75E-02 |
| Sausage as main dish | Food | 1763 | -0.01 | 0.02 | 7.11E-01 | 8.95E-01 | 189 | 1642 | 0.04 | 0.02 | 1.03E-01 | 6.84E-01 | 36 | 0.01 | 0.01 | 4.73E-01 | 0.56 | 1.31E-01 |
| Shellfish (e.g. shrimps, scallops) | Food | 640 | 0.03 | 0.03 | 3.98E-01 | 7.98E-01 | 118 | 567 | 0.01 | 0.04 | 8.94E-01 | 9.83E-01 | 215 | 0.02 | 0.03 | 4.73E-01 | 0.00 | 6.40E-01 |
| Low fat milk (0,5%) | Beverage | 1763 | 0.00 | 0.02 | 9.14E-01 | 9.67E-01 | 225 | 1642 | -0.02 | 0.02 | 3.97E-01 | 9.10E-01 | 104 | -0.01 | 0.02 | 4.98E-01 | 0.00 | 6.04E-01 |
| Total protein intake (g/day) | Nutrients | 1763 | 0.01 | 0.02 | 6.11E-01 | 8.74E-01 | 166 | 1642 | 0.01 | 0.02 | 6.73E-01 | 9.77E-01 | 161 | 0.01 | 0.01 | 5.09E-01 | 0.00 | 9.79E-01 |
| Selenium intake (ug/day) | Nutrients | 1763 | 0.01 | 0.02 | 5.75E-01 | 8.53E-01 | 161 | 1642 | 0.01 | 0.02 | 7.19E-01 | 9.77E-01 | 174 | 0.01 | 0.02 | 5.11E-01 | 0.00 | 9.11E-01 |
| Butter on bread | Food | 1763 | -0.02 | 0.02 | 3.27E-01 | 7.52E-01 | 103 | 1642 | 0.04 | 0.02 | 4.33E-02 | 5.64E-01 | 17 | 0.01 | 0.01 | 5.19E-01 | 0.78 | 3.14E-02 |
| Butter for cooking | Food | 1763 | 0.01 | 0.02 | 4.02E-01 | 7.98E-01 | 119 | 1642 | 0.00 | 0.02 | 9.77E-01 | 9.97E-01 | 234 | 0.01 | 0.01 | 5.19E-01 | 0.00 | 5.91E-01 |
| Contradictory demands in job | Psychosocial | 1860 | -0.01 | 0.02 | 6.14E-01 | 8.74E-01 | 167 | 1750 | -0.01 | 0.02 | 6.86E-01 | 9.77E-01 | 165 | -0.01 | 0.01 | 5.20E-01 | 0.00 | 9.54E-01 |
| Satisfaction with home and family situation | Psychosocial | 1208 | -0.01 | 0.03 | 8.07E-01 | 9.24E-01 | 208 | 1168 | -0.02 | 0.03 | 5.18E-01 | 9.45E-01 | 131 | -0.01 | 0.02 | 5.29E-01 | 0.00 | 7.76E-01 |
| Pentadecanoic acid intake (g/day) | Nutrients | 1763 | 0.01 | 0.02 | 6.56E-01 | 8.75E-01 | 178 | 1642 | 0.01 | 0.02 | 6.60E-01 | 9.77E-01 | 154 | 0.01 | 0.01 | 5.32E-01 | 0.00 | 9.71E-01 |
| Heptadecanoic acid intake (g/day) | Nutrients | 1763 | 0.01 | 0.02 | 6.56E-01 | 8.75E-01 | 179 | 1642 | 0.01 | 0.02 | 6.60E-01 | 9.77E-01 | 155 | 0.01 | 0.01 | 5.32E-01 | 0.00 | 9.71E-01 |
| Calcium intake (mg/day) | Nutrients | 1763 | 0.01 | 0.02 | 5.79E-01 | 8.54E-01 | 162 | 1642 | 0.01 | 0.02 | 7.50E-01 | 9.77E-01 | 181 | 0.01 | 0.01 | 5.33E-01 | 0.00 | 8.84E-01 |
| Syringaresinol intake (ug/day) | Nutrients | 1763 | -0.02 | 0.02 | 3.12E-01 | 7.40E-01 | 100 | 1642 | 0.00 | 0.02 | 8.58E-01 | 9.83E-01 | 204 | -0.01 | 0.01 | 5.34E-01 | 0.00 | 4.14E-01 |
| Confidence status | Psychosocial | 1206 | 0.02 | 0.02 | 4.94E-01 | 8.26E-01 | 143 | 1166 | -0.04 | 0.02 | 1.19E-01 | 7.18E-01 | 39 | -0.01 | 0.02 | 5.36E-01 | 0.60 | 1.13E-01 |
| Close relationship with anyone | Social | 1898 | 0.00 | 0.02 | 8.07E-01 | 9.24E-01 | 207 | 1792 | -0.02 | 0.02 | 2.82E-01 | 8.29E-01 | 79 | -0.01 | 0.01 | 5.37E-01 | 0.00 | 3.61E-01 |
| Sugar, honey, marmelade, jam | Food | 1763 | 0.02 | 0.02 | 2.29E-01 | 7.28E-01 | 74 | 1642 | -0.01 | 0.02 | 6.96E-01 | 9.77E-01 | 168 | 0.01 | 0.01 | 5.57E-01 | 0.20 | 2.63E-01 |
| Formic acid intaje (g/day) | Nutrients | 1763 | 0.01 | 0.02 | 7.00E-01 | 8.95E-01 | 187 | 1642 | 0.01 | 0.02 | 6.54E-01 | 9.77E-01 | 153 | 0.01 | 0.01 | 5.59E-01 | 0.00 | 9.34E-01 |
| Lean fish (e.g. perch, bass, cod) | Food | 1763 | 0.00 | 0.02 | 9.56E-01 | 9.77E-01 | 233 | 1642 | 0.02 | 0.02 | 3.82E-01 | 9.09E-01 | 99 | 0.01 | 0.01 | 5.62E-01 | 0.00 | 5.11E-01 |
| Potassium intake (mg/day) | Nutrients | 1763 | -0.01 | 0.02 | 6.47E-01 | 8.74E-01 | 176 | 1642 | -0.01 | 0.02 | 7.26E-01 | 9.77E-01 | 176 | -0.01 | 0.02 | 5.66E-01 | 0.00 | 9.52E-01 |
| Fatty fish (e.g. herring, whitefish, salmon) | Food | 1763 | 0.00 | 0.02 | 9.95E-01 | 9.97E-01 | 238 | 1642 | -0.02 | 0.02 | 3.88E-01 | 9.09E-01 | 101 | -0.01 | 0.02 | 5.68E-01 | 0.00 | 5.16E-01 |
| Vitamin D intake (ug/day) | Nutrients | 1763 | 0.03 | 0.02 | 1.04E-01 | 7.09E-01 | 33 | 1642 | -0.02 | 0.02 | 3.56E-01 | 9.09E-01 | 91 | 0.01 | 0.01 | 5.81E-01 | 0.69 | 7.39E-02 |
| Marital status: Single vs Married/partner | Social | 1777 | 0.04 | 0.06 | 4.93E-01 | 8.26E-01 | 141 | 1659 | -0.09 | 0.06 | 1.41E-01 | 7.48E-01 | 45 | -0.02 | 0.05 | 5.83E-01 | 0.57 | 1.26E-01 |
| Cholesterol intake (g/day) | Nutrients | 1763 | 0.01 | 0.02 | 5.24E-01 | 8.49E-01 | 147 | 1642 | 0.00 | 0.02 | 9.12E-01 | 9.83E-01 | 221 | 0.01 | 0.02 | 5.84E-01 | 0.00 | 7.32E-01 |
| Teetotaler | Alcohol | 1895 | -0.07 | 0.06 | 2.82E-01 | 7.40E-01 | 89 | 1791 | 0.02 | 0.07 | 7.16E-01 | 9.77E-01 | 173 | -0.02 | 0.05 | 5.89E-01 | 0.00 | 3.17E-01 |
| Minced meat dishes | Food | 1763 | 0.00 | 0.02 | 8.64E-01 | 9.50E-01 | 217 | 1642 | -0.02 | 0.02 | 3.30E-01 | 8.95E-01 | 88 | -0.01 | 0.01 | 5.97E-01 | 0.00 | 4.03E-01 |
| Sodas, soft drinks, juice | Beverage | 1763 | 0.01 | 0.02 | 6.91E-01 | 8.95E-01 | 184 | 1642 | -0.02 | 0.02 | 2.57E-01 | 8.29E-01 | 71 | -0.01 | 0.01 | 5.98E-01 | 0.14 | 2.81E-01 |
| Polyunsaturated fat intake (g/day) | Nutrients | 1763 | 0.03 | 0.02 | 1.62E-01 | 7.28E-01 | 51 | 1642 | -0.01 | 0.02 | 4.70E-01 | 9.22E-01 | 121 | 0.01 | 0.01 | 6.03E-01 | 0.55 | 1.37E-01 |
| Plant based protein intake (g/day) | Nutrients | 1763 | -0.03 | 0.02 | 1.21E-01 | 7.09E-01 | 40 | 1642 | 0.02 | 0.02 | 3.47E-01 | 9.09E-01 | 89 | -0.01 | 0.01 | 6.09E-01 | 0.67 | 8.19E-02 |
| Sour milk, yoghurt (3% fat) | Food | 1763 | 0.00 | 0.02 | 8.18E-01 | 9.24E-01 | 211 | 1642 | 0.01 | 0.02 | 6.16E-01 | 9.77E-01 | 149 | 0.01 | 0.02 | 6.12E-01 | 0.00 | 8.28E-01 |
| Saturated fat intake (g/day) | Nutrients | 1763 | 0.01 | 0.02 | 6.39E-01 | 8.74E-01 | 172 | 1642 | 0.01 | 0.02 | 8.17E-01 | 9.83E-01 | 197 | 0.01 | 0.01 | 6.14E-01 | 0.00 | 8.89E-01 |
| Liquor, spirits | Alcohol | 1763 | -0.02 | 0.02 | 3.92E-01 | 7.98E-01 | 115 | 1642 | 0.01 | 0.02 | 8.08E-01 | 9.83E-01 | 195 | -0.01 | 0.01 | 6.15E-01 | 0.00 | 4.62E-01 |
| Frequency of hunting or fishing during leisure time | Physical activity | 1740 | -0.01 | 0.02 | 6.46E-01 | 8.74E-01 | 174 | 1655 | -0.01 | 0.02 | 8.08E-01 | 9.83E-01 | 194 | -0.01 | 0.02 | 6.17E-01 | 0.00 | 8.84E-01 |
| Vitamin B6 intake (mg/day) | Nutrients | 1763 | -0.02 | 0.02 | 4.63E-01 | 8.26E-01 | 134 | 1642 | 0.00 | 0.02 | 9.52E-01 | 9.93E-01 | 229 | -0.01 | 0.02 | 6.21E-01 | 0.00 | 5.85E-01 |
| Cream, creme fraiche, sour cream | Food | 1763 | 0.00 | 0.02 | 9.28E-01 | 9.76E-01 | 227 | 1642 | 0.01 | 0.02 | 5.03E-01 | 9.30E-01 | 129 | 0.01 | 0.01 | 6.21E-01 | 0.00 | 6.45E-01 |
| Monounsaturated fat intake (g/day) | Nutrients | 1763 | 0.02 | 0.02 | 3.96E-01 | 7.98E-01 | 117 | 1642 | 0.00 | 0.02 | 8.50E-01 | 9.83E-01 | 203 | 0.01 | 0.02 | 6.23E-01 | 0.00 | 4.73E-01 |
| Sum of phytosterols intake (mg/day) | Nutrients | 1763 | 0.01 | 0.02 | 7.46E-01 | 9.05E-01 | 195 | 1642 | -0.02 | 0.02 | 2.86E-01 | 8.29E-01 | 80 | -0.01 | 0.01 | 6.33E-01 | 0.02 | 3.13E-01 |
| Beta-sitosterol intake (mg/day) | Nutrients | 1763 | 0.00 | 0.02 | 8.09E-01 | 9.24E-01 | 209 | 1642 | -0.02 | 0.02 | 3.28E-01 | 8.95E-01 | 87 | -0.01 | 0.01 | 6.33E-01 | 0.00 | 3.75E-01 |

| | | | | | | | | | | | | | | | | | | |
|---|---|---|---|---|---|---|---|---|---|---|---|---|---|---|---|---|---|---|
| Sweets | Food | 1763 | 0.02 | 0.02 | 2.90E-01 | 7.40E-01 | 91 | 1642 | -0.01 | 0.02 | 5.43E-01 | 9.46E-01 | 136 | 0.01 | 0.01 | 6.36E-01 | 0.21 | 2.60E-01 |
| Fitness status | Physical activity | 1206 | 0.02 | 0.03 | 5.50E-01 | 8.49E-01 | 155 | 1165 | -0.03 | 0.03 | 2.07E-01 | 7.87E-01 | 63 | -0.01 | 0.02 | 6.40E-01 | 0.42 | 1.89E-01 |
| Alcohol intake (g/day) | Alcohol | 1763 | 0.02 | 0.02 | 3.32E-01 | 7.57E-01 | 105 | 1642 | -0.01 | 0.02 | 6.52E-01 | 9.77E-01 | 152 | 0.01 | 0.02 | 6.45E-01 | 0.00 | 3.34E-01 |
| Cohabitation: Live alone vs Only one adult (spouse, partner) | Social | 598 | 0.10 | 0.08 | 2.25E-01 | 7.28E-01 | 72 | 600 | -0.17 | 0.09 | 4.69E-02 | 5.64E-01 | 19 | -0.03 | 0.06 | 6.52E-01 | 0.81 | 2.21E-02 |
| Iron intake (mg/day) | Nutrients | 1763 | -0.03 | 0.02 | 2.14E-01 | 7.28E-01 | 66 | 1642 | 0.02 | 0.02 | 4.82E-01 | 9.22E-01 | 125 | -0.01 | 0.02 | 6.52E-01 | 0.46 | 1.75E-01 |
| Equol intake (ug/day) | Nutrients | 1763 | 0.03 | 0.02 | 2.34E-01 | 7.28E-01 | 75 | 1642 | -0.01 | 0.02 | 5.42E-01 | 9.46E-01 | 135 | 0.01 | 0.02 | 6.60E-01 | 0.37 | 2.07E-01 |
| Sedentary or standing work | Physical activity | 1871 | 0.01 | 0.04 | 7.60E-01 | 9.08E-01 | 200 | 1755 | 0.01 | 0.05 | 7.59E-01 | 9.77E-01 | 185 | 0.01 | 0.03 | 6.66E-01 | 0.00 | 9.91E-01 |
| Parents or siblings have diabetes | General health | 1888 | -0.10 | 0.05 | 3.93E-02 | 7.09E-01 | 12 | 1779 | 0.08 | 0.05 | 1.20E-01 | 7.18E-01 | 40 | -0.02 | 0.04 | 6.67E-01 | 0.85 | 1.09E-02 |
| Mashed potato | Food | 640 | -0.05 | 0.04 | 1.74E-01 | 7.28E-01 | 54 | 567 | 0.03 | 0.04 | 4.27E-01 | 9.10E-01 | 110 | -0.01 | 0.03 | 6.68E-01 | 0.57 | 1.29E-01 |
| Vitamin B12 intake (ug/day) | Nutrients | 1763 | 0.01 | 0.02 | 7.44E-01 | 9.05E-01 | 194 | 1642 | 0.01 | 0.02 | 7.80E-01 | 9.83E-01 | 189 | 0.01 | 0.01 | 6.68E-01 | 0.00 | 9.93E-01 |
| Sleep status | Sleep | 1209 | 0.05 | 0.02 | 3.40E-02 | 7.09E-01 | 10 | 1169 | -0.04 | 0.02 | 1.35E-01 | 7.48E-01 | 43 | 0.01 | 0.02 | 6.68E-01 | 0.85 | 1.05E-02 |
| Linolenic acid intake (g/day) | Nutrients | 1763 | 0.03 | 0.02 | 1.54E-01 | 7.28E-01 | 50 | 1642 | -0.02 | 0.02 | 3.59E-01 | 9.09E-01 | 94 | 0.01 | 0.01 | 6.71E-01 | 0.63 | 1.01E-01 |
| Average portion size of potatoes/rice/pasta based on photographic illustration of four sizes (smallest to largest) | Food | 1763 | -0.02 | 0.02 | 3.27E-01 | 7.52E-01 | 104 | 1642 | 0.04 | 0.02 | 9.80E-02 | 6.84E-01 | 34 | 0.01 | 0.02 | 6.74E-01 | 0.72 | 6.04E-02 |
| Average portion size of meat/fish based on photographic illustration of four sizes (smallest to largest) | Food | 1763 | -0.03 | 0.02 | 2.18E-01 | 7.28E-01 | 68 | 1642 | 0.02 | 0.02 | 4.79E-01 | 9.22E-01 | 123 | -0.01 | 0.02 | 6.76E-01 | 0.46 | 1.75E-01 |
| Wine | Alcohol | 1763 | 0.02 | 0.02 | 4.25E-01 | 8.07E-01 | 124 | 1642 | -0.04 | 0.02 | 1.30E-01 | 7.40E-01 | 42 | -0.01 | 0.02 | 6.78E-01 | 0.64 | 9.68E-02 |
| Appetite status | Psychosocial | 1209 | -0.03 | 0.02 | 2.16E-01 | 7.28E-01 | 67 | 1166 | 0.02 | 0.03 | 4.93E-01 | 9.27E-01 | 127 | -0.01 | 0.02 | 6.78E-01 | 0.45 | 1.76E-01 |
| Oatflake, whole wheat, rye or barley porridge | Food | 1763 | 0.00 | 0.02 | 8.24E-01 | 9.24E-01 | 213 | 1642 | -0.01 | 0.02 | 7.16E-01 | 9.77E-01 | 172 | -0.01 | 0.01 | 6.82E-01 | 0.00 | 9.04E-01 |
| Cohabitation: Live alone vs Adult and children | Social | 1358 | 0.12 | 0.07 | 8.36E-02 | 7.09E-01 | 27 | 1220 | -0.18 | 0.07 | 1.51E-02 | 4.16E-01 | 8 | -0.02 | 0.05 | 6.87E-01 | 0.89 | 3.08E-03 |
| Meat stew | Food | 1763 | 0.01 | 0.02 | 6.68E-01 | 8.82E-01 | 181 | 1642 | 0.00 | 0.02 | 9.13E-01 | 9.83E-01 | 222 | 0.01 | 0.01 | 6.93E-01 | 0.00 | 8.41E-01 |
| Cambridge physical activity index | Physical activity | 1864 | 0.00 | 0.02 | 9.52E-01 | 9.77E-01 | 232 | 1753 | 0.01 | 0.02 | 5.32E-01 | 9.46E-01 | 133 | 0.01 | 0.01 | 6.94E-01 | 0.00 | 6.25E-01 |
| Folic acid intake (ug/day) | Nutrients | 1763 | 0.02 | 0.02 | 3.73E-01 | 7.97E-01 | 112 | 1642 | -0.01 | 0.02 | 6.99E-01 | 9.77E-01 | 169 | 0.01 | 0.02 | 6.98E-01 | 0.00 | 3.74E-01 |
| Number of people with whom you can speak openly | Social | 1897 | 0.01 | 0.02 | 4.38E-01 | 8.07E-01 | 129 | 1789 | -0.01 | 0.02 | 7.86E-01 | 9.83E-01 | 190 | 0.01 | 0.01 | 7.00E-01 | 0.00 | 4.68E-01 |
| Enterodiol intake (ug/day) | Nutrients | 1763 | 0.00 | 0.02 | 8.23E-01 | 9.24E-01 | 212 | 1642 | -0.02 | 0.02 | 4.18E-01 | 9.10E-01 | 108 | -0.01 | 0.02 | 7.15E-01 | 0.00 | 4.49E-01 |
| Corn flakes | Food | 1763 | 0.02 | 0.02 | 3.77E-01 | 7.97E-01 | 113 | 1642 | -0.01 | 0.02 | 6.66E-01 | 9.77E-01 | 159 | 0.01 | 0.01 | 7.22E-01 | 0.00 | 3.59E-01 |
| Vitamin B3 intake (mg/day) | Nutrients | 1763 | -0.01 | 0.02 | 5.37E-01 | 8.49E-01 | 151 | 1642 | 0.00 | 0.02 | 8.91E-01 | 9.83E-01 | 213 | -0.01 | 0.01 | 7.23E-01 | 0.00 | 6.01E-01 |
| Sucrose intake (g/day) | Nutrients | 1763 | 0.03 | 0.02 | 1.22E-01 | 7.09E-01 | 41 | 1642 | -0.02 | 0.02 | 2.70E-01 | 8.29E-01 | 74 | 0.01 | 0.01 | 7.23E-01 | 0.71 | 6.17E-02 |
| High physical demand from job | Physical activity | 1880 | 0.00 | 0.02 | 8.84E-01 | 9.56E-01 | 221 | 1770 | 0.01 | 0.02 | 5.06E-01 | 9.30E-01 | 130 | 0.01 | 0.01 | 7.26E-01 | 0.00 | 5.59E-01 |
| Permanent employment | Psychosocial | 1867 | 0.05 | 0.04 | 2.23E-01 | 7.28E-01 | 70 | 1770 | -0.03 | 0.05 | 4.47E-01 | 9.10E-01 | 117 | 0.01 | 0.03 | 7.28E-01 | 0.49 | 1.63E-01 |
| Frequent social contacts with colleagues during work | Psychosocial | 1862 | 0.03 | 0.02 | 1.21E-01 | 7.09E-01 | 39 | 1752 | -0.02 | 0.02 | 2.68E-01 | 8.29E-01 | 73 | 0.00 | 0.01 | 7.38E-01 | 0.72 | 6.05E-02 |
| Hamburger | Food | 1763 | 0.03 | 0.02 | 1.44E-01 | 7.17E-01 | 48 | 1642 | -0.06 | 0.02 | 1.65E-02 | 4.16E-01 | 9 | 0.00 | 0.02 | 7.41E-01 | 0.87 | 5.26E-03 |
| Do you feel important and appreciated outside your home? | Psychosocial | 1210 | 0.02 | 0.02 | 4.35E-01 | 8.07E-01 | 127 | 1165 | -0.01 | 0.02 | 7.40E-01 | 9.77E-01 | 178 | 0.01 | 0.02 | 7.51E-01 | 0.00 | 4.31E-01 |
| Repetitive job | Psychosocial | 1873 | 0.00 | 0.02 | 9.37E-01 | 9.76E-01 | 229 | 1767 | 0.01 | 0.02 | 5.90E-01 | 9.53E-01 | 148 | 0.00 | 0.01 | 7.59E-01 | 0.00 | 6.52E-01 |
| Distance to work in kilometers (one way) | Physical activity | 1611 | -0.01 | 0.02 | 6.80E-01 | 8.88E-01 | 183 | 1524 | 0.01 | 0.02 | 4.33E-01 | 9.10E-01 | 112 | 0.00 | 0.01 | 7.65E-01 | 0.00 | 4.05E-01 |
| Margarine on bread | Food | 1763 | 0.00 | 0.02 | 7.96E-01 | 9.24E-01 | 205 | 1642 | 0.00 | 0.02 | 8.84E-01 | 9.83E-01 | 212 | 0.00 | 0.01 | 7.67E-01 | 0.00 | 9.75E-01 |
| Enough time for job assignments | Psychosocial | 1866 | -0.02 | 0.02 | 3.37E-01 | 7.59E-01 | 106 | 1754 | 0.01 | 0.02 | 5.67E-01 | 9.47E-01 | 143 | 0.00 | 0.01 | 7.68E-01 | 0.14 | 2.80E-01 |
| Low fat margarine on bread | Food | 1763 | 0.00 | 0.02 | 8.66E-01 | 9.50E-01 | 218 | 1642 | 0.01 | 0.02 | 8.07E-01 | 9.83E-01 | 193 | 0.00 | 0.01 | 7.72E-01 | 0.00 | 9.49E-01 |
| White cabbage, lettuce, lettuce cabbage, spinach, borecole | Food | 1763 | 0.02 | 0.02 | 3.01E-01 | 7.40E-01 | 96 | 1642 | -0.02 | 0.02 | 4.75E-01 | 9.22E-01 | 122 | 0.00 | 0.02 | 7.75E-01 | 0.33 | 2.21E-01 |
| Campesterol intake (mg/day) | Nutrients | 1763 | 0.01 | 0.02 | 4.75E-01 | 8.26E-01 | 135 | 1642 | -0.02 | 0.02 | 2.29E-01 | 8.09E-01 | 65 | 0.00 | 0.01 | 7.75E-01 | 0.47 | 1.70E-01 |
| Breakfast habits: Gruel w/o sandwich for breakfast vs not breakfast at all | Food | 155 | -0.02 | 0.17 | 8.95E-01 | 9.59E-01 | 223 | 150 | -0.08 | 0.24 | 7.56E-01 | 9.77E-01 | 183 | -0.04 | 0.14 | 7.75E-01 | 0.00 | 8.56E-01 |
| Light beer | Alcohol | 1763 | 0.01 | 0.03 | 6.40E-01 | 8.74E-01 | 173 | 1642 | -0.03 | 0.03 | 3.73E-01 | 9.09E-01 | 98 | -0.01 | 0.02 | 7.78E-01 | 0.00 | 3.34E-01 |
| Sodium intake (mg/day) | Nutrients | 1763 | -0.02 | 0.02 | 3.13E-01 | 7.40E-01 | 101 | 1642 | 0.01 | 0.02 | 4.97E-01 | 9.29E-01 | 128 | 0.00 | 0.01 | 7.89E-01 | 0.29 | 2.35E-01 |
| Frequency of engaging in clubs, associations or study circles | Social | 1273 | -0.03 | 0.02 | 2.76E-01 | 7.40E-01 | 86 | 1188 | 0.02 | 0.02 | 4.59E-01 | 9.21E-01 | 119 | 0.00 | 0.02 | 7.90E-01 | 0.40 | 1.97E-01 |
| Docosahexaenoic acid (DHA) intake (g/day) | Nutrients | 1763 | -0.01 | 0.02 | 7.68E-01 | 9.08E-01 | 202 | 1642 | 0.00 | 0.02 | 9.49E-01 | 9.93E-01 | 228 | 0.00 | 0.02 | 8.00E-01 | 0.00 | 8.69E-01 |
| Brewed (filtered) coffee | Beverage | 1763 | 0.01 | 0.02 | 5.48E-01 | 8.49E-01 | 154 | 1642 | -0.01 | 0.02 | 7.77E-01 | 9.83E-01 | 188 | 0.00 | 0.01 | 8.04E-01 | 0.00 | 5.38E-01 |
| Long-term sickness | General health | 1850 | -0.06 | 0.06 | 3.00E-01 | 7.40E-01 | 94 | 1733 | 0.05 | 0.06 | 4.41E-01 | 9.10E-01 | 114 | -0.01 | 0.04 | 8.08E-01 | 0.38 | 2.04E-01 |
| Berries (fresh or frozen) | Food | 1763 | 0.00 | 0.02 | 9.03E-01 | 9.64E-01 | 224 | 1642 | -0.01 | 0.02 | 6.77E-01 | 9.77E-01 | 162 | 0.00 | 0.01 | 8.21E-01 | 0.00 | 7.11E-01 |
| Fat intake (g/day) | Nutrients | 1763 | 0.01 | 0.02 | 4.89E-01 | 8.26E-01 | 137 | 1642 | -0.01 | 0.02 | 6.78E-01 | 9.77E-01 | 163 | 0.00 | 0.01 | 8.25E-01 | 0.00 | 4.37E-01 |
| Satisfaction with economy | Psychosocial | 1207 | 0.02 | 0.02 | 4.09E-01 | 7.98E-01 | 121 | 1162 | -0.03 | 0.02 | 2.51E-01 | 8.29E-01 | 70 | 0.00 | 0.02 | 8.27E-01 | 0.49 | 1.62E-01 |
| Overall state of health compared to other your age | General health | 1856 | 0.01 | 0.02 | 7.19E-01 | 8.95E-01 | 192 | 1768 | -0.02 | 0.02 | 4.68E-01 | 9.22E-01 | 120 | 0.00 | 0.01 | 8.29E-01 | 0.00 | 4.35E-01 |
| Milk, sour milk (3%) | Beverage | 1763 | 0.01 | 0.02 | 6.98E-01 | 8.95E-01 | 186 | 1642 | 0.00 | 0.02 | 8.71E-01 | 9.83E-01 | 208 | 0.00 | 0.01 | 8.32E-01 | 0.00 | 7.17E-01 |
| Eicosapentaenoic acid (EPA) intake (g/day) | Nutrients | 1763 | -0.01 | 0.02 | 7.99E-01 | 9.24E-01 | 206 | 1642 | 0.00 | 0.02 | 9.74E-01 | 9.97E-01 | 233 | 0.00 | 0.02 | 8.41E-01 | 0.00 | 8.72E-01 |
| Total energy intake (kcal/day) | Nutrients | 1763 | -0.02 | 0.02 | 4.26E-01 | 8.07E-01 | 125 | 1642 | 0.03 | 0.02 | 2.71E-01 | 8.29E-01 | 75 | 0.00 | 0.02 | 8.50E-01 | 0.45 | 1.79E-01 |
| Root vegetables, carrot | Food | 1763 | 0.01 | 0.02 | 5.56E-01 | 8.51E-01 | 156 | 1642 | -0.02 | 0.02 | 3.71E-01 | 9.09E-01 | 96 | 0.00 | 0.01 | 8.70E-01 | 0.11 | 2.90E-01 |
| High mental demand from job | Psychosocial | 1856 | 0.01 | 0.02 | 7.82E-01 | 9.20E-01 | 203 | 1747 | 0.00 | 0.02 | 9.60E-01 | 9.93E-01 | 231 | 0.00 | 0.01 | 8.72E-01 | 0.00 | 8.17E-01 |
| Patience status | Psychosocial | 1209 | 0.01 | 0.03 | 7.41E-01 | 9.05E-01 | 193 | 1165 | 0.00 | 0.02 | 9.17E-01 | 9.83E-01 | 223 | 0.00 | 0.02 | 8.76E-01 | 0.00 | 7.57E-01 |
| Smoked fish/meat | Food | 1763 | -0.01 | 0.02 | 4.53E-01 | 8.26E-01 | 131 | 1642 | 0.02 | 0.02 | 2.60E-01 | 8.29E-01 | 72 | 0.00 | 0.01 | 8.80E-01 | 0.45 | 1.79E-01 |
| Support from others | Social | 1896 | -0.01 | 0.02 | 7.48E-01 | 9.05E-01 | 196 | 1789 | 0.00 | 0.02 | 9.10E-01 | 9.83E-01 | 220 | 0.00 | 0.01 | 8.89E-01 | 0.00 | 7.56E-01 |
| Rosehip, sweet syrup soup | Food | 1763 | 0.01 | 0.02 | 5.37E-01 | 8.49E-01 | 150 | 1642 | -0.02 | 0.02 | 4.01E-01 | 9.10E-01 | 105 | 0.00 | 0.02 | 8.96E-01 | 0.06 | 3.01E-01 |
| Bregott on bread | Food | 1763 | 0.02 | 0.02 | 3.62E-01 | 7.87E-01 | 110 | 1642 | -0.02 | 0.02 | 2.42E-01 | 8.29E-01 | 69 | 0.00 | 0.01 | 8.99E-01 | 0.54 | 1.39E-01 |
| Beta-carotene intake (mg/day) | Nutrients | 1763 | 0.01 | 0.02 | 5.15E-01 | 8.44E-01 | 145 | 1642 | -0.02 | 0.02 | 3.69E-01 | 9.09E-01 | 95 | 0.00 | 0.01 | 9.03E-01 | 0.18 | 2.70E-01 |
| Cheese 10-17% | Food | 1763 | -0.01 | 0.02 | 4.89E-01 | 8.26E-01 | 138 | 1642 | 0.01 | 0.02 | 5.82E-01 | 9.53E-01 | 146 | 0.00 | 0.01 | 9.10E-01 | 0.00 | 3.80E-01 |
| Steak, chop, e.g. | Food | 1763 | 0.01 | 0.02 | 5.45E-01 | 8.49E-01 | 153 | 1642 | -0.01 | 0.02 | 5.87E-01 | 9.53E-01 | 147 | 0.00 | 0.01 | 9.16E-01 | 0.00 | 4.20E-01 |
| Whole grain crisp bread | Food | 1763 | 0.00 | 0.02 | 8.16E-01 | 9.24E-01 | 210 | 1642 | -0.01 | 0.02 | 6.95E-01 | 9.77E-01 | 167 | 0.00 | 0.01 | 9.26E-01 | 0.00 | 6.55E-01 |
| Number of friends that can come to your home at any time and feel at home | Social | 1893 | 0.01 | 0.02 | 4.94E-01 | 8.26E-01 | 142 | 1784 | -0.01 | 0.02 | 5.67E-01 | 9.47E-01 | 142 | 0.00 | 0.01 | 9.27E-01 | 0.00 | 3.75E-01 |
| Blood based food | Food | 640 | 0.00 | 0.03 | 9.43E-01 | 9.76E-01 | 231 | 567 | 0.01 | 0.04 | 8.22E-01 | 9.83E-01 | 199 | 0.00 | 0.02 | 9.31E-01 | 0.00 | 8.27E-01 |
| Mixed frozen vegetables | Food | 640 | 0.00 | 0.03 | 9.14E-01 | 9.67E-01 | 226 | 567 | 0.00 | 0.04 | 9.97E-01 | 1.00E+00 | 238 | 0.00 | 0.03 | 9.32E-01 | 0.00 | 9.47E-01 |
| Sausage, liver pate on bread | Food | 1763 | -0.01 | 0.02 | 7.48E-01 | 9.05E-01 | 197 | 1642 | 0.00 | 0.02 | 8.27E-01 | 9.83E-01 | 200 | 0.00 | 0.01 | 9.37E-01 | 0.00 | 7.03E-01 |
| Satisfaction with leisure time | Psychosocial | 1208 | 0.01 | 0.02 | 6.08E-01 | 8.74E-01 | 165 | 1163 | -0.01 | 0.03 | 6.64E-01 | 9.77E-01 | 156 | 0.00 | 0.02 | 9.49E-01 | 0.00 | 5.03E-01 |
| Salty fish | Food | 1763 | -0.01 | 0.02 | 5.66E-01 | 8.51E-01 | 159 | 1642 | 0.01 | 0.02 | 5.58E-01 | 9.46E-01 | 141 | 0.00 | 0.01 | 9.49E-01 | 0.00 | 4.13E-01 |
| Enterolactone intake (ug/day) | Nutrients | 1763 | 0.01 | 0.02 | 7.55E-01 | 9.07E-01 | 199 | 1642 | -0.01 | 0.02 | 6.64E-01 | 9.77E-01 | 157 | 0.00 | 0.02 | 9.57E-01 | 0.00 | 5.95E-01 |
| Stigmasterol intake (mg/day) | Nutrients | 1763 | 0.02 | 0.02 | 2.40E-01 | 7.28E-01 | 78 | 1642 | -0.03 | 0.02 | 1.77E-01 | 7.87E-01 | 52 | 0.00 | 0.01 | 9.63E-01 | 0.69 | 7.37E-02 |
| Would you say that the number of people that you meet in your everyday life is enough or would you like to meet more or fewer people? | Social | 1895 | 0.00 | 0.02 | 9.87E-01 | 9.95E-01 | 237 | 1789 | 0.00 | 0.02 | 9.31E-01 | 9.85E-01 | 225 | 0.00 | 0.01 | 9.63E-01 | 0.00 | 9.40E-01 |
| Coffee rolls/buns, rusk | Food | 1763 | -0.02 | 0.02 | 4.62E-01 | 8.26E-01 | 133 | 1642 | 0.01 | 0.02 | 4.80E-01 | 9.22E-01 | 124 | 0.00 | 0.01 | 9.74E-01 | 0.04 | 3.08E-01 |
| Number of social interactions during a normal week | Social | 1895 | 0.00 | 0.02 | 8.88E-01 | 9.56E-01 | 222 | 1788 | 0.00 | 0.02 | 9.06E-01 | 9.83E-01 | 218 | 0.00 | 0.01 | 9.84E-01 | 0.00 | 8.55E-01 |
| Smoking status: Former smokers vs non-smokers | Tobacco use | 1330 | 0.06 | 0.05 | 2.09E-01 | 7.28E-01 | 65 | 1235 | -0.07 | 0.05 | 1.93E-01 | 7.87E-01 | 58 | 0.00 | 0.04 | 9.90E-01 | 0.69 | 7.04E-02 |
| Participation in associations or voluntary organisations | Social | 1895 | 0.04 | 0.04 | 3.01E-01 | 7.40E-01 | 95 | 1778 | -0.05 | 0.04 | 2.81E-01 | 8.29E-01 | 78 | 0.00 | 0.03 | 9.94E-01 | 0.55 | 1.35E-01 |
| People to ask for help apart from the ones at home | Social | 1893 | 0.00 | 0.06 | 9.97E-01 | 9.97E-01 | 239 | 1789 | 0.00 | 0.07 | 9.95E-01 | 1.00E+00 | 237 | 0.00 | 0.05 | 9.95E-01 | 0.00 | 9.98E-01 |
| White meat (poultry) | Food | 1763 | 0.01 | 0.02 | 7.94E-01 | 9.24E-01 | 204 | 1642 | -0.01 | 0.02 | 7.95E-01 | 9.83E-01 | 191 | 0.00 | 0.02 | 9.96E-01 | 0.00 | 7.12E-01 |
| Rice | Food | 1763 | -0.03 | 0.02 | 1.72E-01 | 7.28E-01 | 53 | 1642 | 0.02 | 0.02 | 2.29E-01 | 8.09E-01 | 66 | 0.00 | 0.01 | 1.00E+00 | 0.70 | 6.87E-02 |

**Supplementary Table 19**. Longitudinal association results for fasting glucose

| | | Training set | | | | | | Testing set | | | | | | Metanalysis | | | | | R2 | |
|---|---|---|---|---|---|---|---|---|---|---|---|---|---|---|---|---|---|---|---|---|
| **Description** | **Group** | **N** | **Effect estimate** | **S.E.** | **p-value** | **p-value$_{FDR}$** | **p-value rank** | **N** | **Effect estimate** | **S.E.** | **p-value** | **p-value$_{FDR}$** | **p-value rank** | **Effect estimate** | **S.E.** | **p-value** | **$I^2$** | **Q p-value** | **adjusted R2** | **R2 rank** |
| Smoking status: Smokers vs non-smokers | Tobacco use | 8137 | 0.16 | 0.02 | 1.10E-15 | **1.33E-13** | 2 | 7885 | 0.14 | 0.02 | 1.44E-12 | **3.50E-10** | 1 | 0.15 | 0.01 | 1.03E-26 | 0.00 | 5.62E-01 | 0.09 | 1 |
| Number of cigarretes smoked per day (in groups) | Tobacco use | 10341 | 0.06 | 0.01 | 3.86E-16 | **9.38E-14** | 1 | 10078 | 0.05 | 0.01 | 2.55E-09 | **2.07E-07** | 3 | 0.05 | 0.01 | 1.62E-23 | 0.55 | 1.38E-01 | 0.09 | 2 |
| Years smoking | Tobacco use | 10872 | 0.05 | 0.01 | 2.74E-10 | **2.22E-08** | 3 | 10662 | 0.05 | 0.01 | 1.21E-10 | **1.47E-08** | 2 | 0.05 | 0.01 | 1.84E-19 | 0.00 | 8.27E-01 | 0.09 | 3 |
| Parents or siblings have diabetes | General health | 12182 | 0.08 | 0.02 | 2.34E-05 | **1.14E-03** | 5 | 11989 | 0.10 | 0.02 | 2.14E-07 | **1.30E-05** | 4 | 0.09 | 0.01 | 2.95E-11 | 0.00 | 4.27E-01 | 0.09 | 4 |
| Educational level | Psychosocial | 12277 | -0.03 | 0.01 | 7.90E-06 | **4.80E-04** | 4 | 12073 | -0.02 | 0.01 | 1.52E-03 | **3.07E-02** | 12 | -0.03 | 0.01 | 5.80E-08 | 0.00 | 4.34E-01 | 0.09 | 5 |
| Breakfast habits: Only coffee/tea for breakfast vs not breakfast at all | Food | 784 | -0.29 | 0.11 | 6.06E-03 | 8.20E-02 | 17 | 750 | -0.45 | 0.10 | 1.57E-05 | **7.62E-04** | 5 | -0.37 | 0.07 | 4.79E-07 | 0.11 | 2.88E-01 | - | - |
| Grams of tobacco smoked per week | Tobacco use | 7644 | -0.04 | 0.01 | 5.56E-03 | 8.20E-02 | 14 | 7383 | -0.06 | 0.01 | 2.01E-04 | **4.87E-03** | 8 | -0.05 | 0.01 | 4.38E-06 | 0.00 | 5.06E-01 | - | - |
| Number of cigarretes smoked per day | Tobacco use | 7644 | -0.04 | 0.01 | 5.56E-03 | 8.20E-02 | 15 | 7383 | -0.06 | 0.01 | 2.01E-04 | **4.87E-03** | 9 | -0.05 | 0.01 | 4.38E-06 | 0.00 | 5.06E-01 | - | - |
| Number of cigars smoked per day | Tobacco use | 7644 | -0.04 | 0.01 | 5.56E-03 | 8.20E-02 | 16 | 7383 | -0.06 | 0.01 | 2.01E-04 | **4.87E-03** | 10 | -0.05 | 0.01 | 4.38E-06 | 0.00 | 5.06E-01 | - | - |
| Arachidonic acid (ARA) intake (g/day) | Nutrients | 11348 | 0.01 | 0.01 | 5.69E-02 | 3.14E-01 | 44 | 11147 | 0.03 | 0.01 | 7.58E-05 | **2.63E-03** | 7 | 0.02 | 0.01 | 3.94E-05 | 0.58 | 1.21E-01 | - | - |
| Participation in associations or voluntary organisations | Social | 12252 | -0.05 | 0.02 | 2.70E-03 | 5.97E-02 | 11 | 12039 | -0.04 | 0.02 | 1.54E-02 | 1.39E-01 | 27 | -0.04 | 0.01 | 1.22E-04 | 0.00 | 7.49E-01 | - | - |
| Vitamin B12 intake (ug/day) | Nutrients | 11348 | 0.01 | 0.01 | 6.39E-02 | 3.38E-01 | 46 | 11147 | 0.03 | 0.01 | 3.03E-04 | **6.70E-03** | 11 | 0.02 | 0.01 | 1.31E-04 | 0.46 | 1.72E-01 | - | - |
| Frequency of picking berries or mushrooms during leisure time | Physical activity | 11596 | 0.02 | 0.01 | 4.48E-03 | 8.20E-02 | 13 | 11407 | 0.02 | 0.01 | 1.43E-02 | 1.38E-01 | 24 | 0.02 | 0.01 | 1.78E-04 | 0.00 | 8.37E-01 | - | - |
| Sausage, liver pate on bread | Food | 11348 | 0.01 | 0.01 | 1.92E-01 | 5.17E-01 | 90 | 11147 | 0.03 | 0.01 | 6.53E-05 | **2.63E-03** | 6 | 0.02 | 0.01 | 1.95E-04 | 0.74 | 5.18E-02 | - | - |
| Fatty fish (e.g. herring, whitefish, salmon) | Food | 11348 | 0.02 | 0.01 | 1.71E-03 | **4.15E-02** | 10 | 11147 | 0.02 | 0.01 | 4.97E-02 | 2.63E-01 | 46 | 0.02 | 0.01 | 2.97E-04 | 0.00 | 4.36E-01 | - | - |
| Minced meat dishes | Food | 11348 | 0.01 | 0.01 | 5.51E-02 | 3.11E-01 | 43 | 11147 | 0.02 | 0.01 | 2.11E-03 | **3.95E-02** | 13 | 0.02 | 0.01 | 4.30E-04 | 0.00 | 3.90E-01 | - | - |
| Distance to work in kilometers (one way) | Physical activity | 10808 | 0.03 | 0.01 | 1.81E-04 | **7.33E-03** | 6 | 10690 | 0.01 | 0.01 | 3.27E-01 | 7.10E-01 | 112 | 0.02 | 0.01 | 6.88E-04 | 0.71 | 6.26E-02 | - | - |
| Whole grain intake (g/day) | Food | 11348 | -0.02 | 0.01 | 9.67E-03 | 1.04E-01 | 22 | 11147 | -0.02 | 0.01 | 3.05E-02 | 2.00E-01 | 37 | -0.02 | 0.01 | 7.64E-04 | 0.00 | 8.23E-01 | - | - |
| Exercise during the last three months | Physical activity | 12283 | -0.02 | 0.01 | 9.32E-03 | 1.04E-01 | 21 | 12077 | -0.01 | 0.01 | 9.52E-02 | 3.76E-01 | 61 | -0.02 | 0.01 | 2.42E-03 | 0.00 | 5.54E-01 | - | - |
| Monounsaturated fat intake (g/day) | Nutrients | 11348 | 0.02 | 0.01 | 6.41E-03 | 8.20E-02 | 19 | 11147 | 0.01 | 0.01 | 1.30E-01 | 4.50E-01 | 70 | 0.02 | 0.01 | 2.52E-03 | 0.00 | 4.39E-01 | - | - |
| Fiber cereals | Food | 11348 | -0.02 | 0.01 | 1.38E-02 | 1.27E-01 | 26 | 11147 | -0.01 | 0.01 | 7.60E-02 | 3.34E-01 | 55 | -0.02 | 0.01 | 2.64E-03 | 0.00 | 6.78E-01 | - | - |
| Oatflake, whole wheat, rye or barley porridge | Food | 11348 | -0.02 | 0.01 | 7.04E-03 | 8.55E-02 | 20 | 11147 | -0.01 | 0.01 | 1.32E-01 | 4.50E-01 | 71 | -0.02 | 0.01 | 2.76E-03 | 0.00 | 4.51E-01 | - | - |
| Breakfast habits: Porridge w/o sandwich for breakfast vs not breakfast at all | Food | 1683 | -0.12 | 0.04 | 6.12E-03 | 8.20E-02 | 18 | 1620 | -0.06 | 0.04 | 1.59E-01 | 4.84E-01 | 80 | -0.09 | 0.03 | 3.33E-03 | 0.00 | 3.44E-01 | - | - |
| Cookies, pastry | Food | 11348 | -0.02 | 0.01 | 1.93E-02 | 1.56E-01 | 30 | 11147 | -0.01 | 0.01 | 9.47E-02 | 3.76E-01 | 60 | -0.02 | 0.01 | 4.45E-03 | 0.00 | 6.73E-01 | - | - |
| Average portion size of potatoes/rice/pasta based on photographic illustration of four sizes (smallest to largest) | Food | 11348 | -0.02 | 0.01 | 3.21E-03 | 6.49E-02 | 12 | 11147 | -0.01 | 0.01 | 3.18E-01 | 6.96E-01 | 111 | -0.02 | 0.01 | 4.82E-03 | 0.43 | 1.87E-01 | - | - |
| Docosahexaenoic acid (DHA) intake (g/day) | Nutrients | 11348 | 0.02 | 0.01 | 2.82E-02 | 2.08E-01 | 32 | 11147 | 0.01 | 0.01 | 8.23E-02 | 3.51E-01 | 57 | 0.01 | 0.01 | 5.31E-03 | 0.00 | 7.91E-01 | - | - |
| Selenium intake (ug/day) | Nutrients | 11348 | 0.01 | 0.01 | 1.36E-01 | 4.80E-01 | 69 | 11147 | 0.02 | 0.01 | 1.36E-02 | 1.38E-01 | 23 | 0.01 | 0.01 | 5.52E-03 | 0.00 | 4.35E-01 | - | - |
| Vitamin A intake (mg/day) | Nutrients | 11348 | 0.02 | 0.01 | 3.17E-02 | 2.08E-01 | 37 | 11147 | 0.02 | 0.01 | 7.70E-02 | 3.34E-01 | 56 | 0.02 | 0.01 | 5.53E-03 | 0.00 | 8.29E-01 | - | - |
| Tea | Beverage | 11348 | -0.02 | 0.01 | 2.78E-02 | 2.08E-01 | 31 | 11147 | -0.01 | 0.01 | 8.91E-02 | 3.73E-01 | 58 | -0.01 | 0.01 | 5.66E-03 | 0.00 | 7.82E-01 | - | - |
| Liquor, spirits | Alcohol | 11348 | 0.02 | 0.01 | 1.43E-03 | **4.15E-02** | 8 | 11147 | 0.01 | 0.01 | 5.21E-01 | 8.44E-01 | 150 | 0.02 | 0.01 | 5.79E-03 | 0.66 | 8.47E-02 | - | - |
| Carbohydrates intake (g/day) | Nutrients | 11348 | -0.01 | 0.01 | 7.06E-02 | 3.53E-01 | 48 | 11147 | -0.02 | 0.01 | 3.83E-02 | 2.32E-01 | 40 | -0.01 | 0.01 | 6.18E-03 | 0.00 | 7.97E-01 | - | - |
| Soft whey cheese | Food | 5080 | -0.03 | 0.01 | 1.00E-02 | 1.04E-01 | 23 | 4983 | -0.01 | 0.01 | 2.21E-01 | 5.91E-01 | 91 | -0.02 | 0.01 | 7.11E-03 | 0.00 | 3.46E-01 | - | - |
| Informed of having high blood pressure | General health | 12293 | 0.04 | 0.02 | 7.52E-02 | 3.66E-01 | 50 | 12080 | 0.04 | 0.02 | 4.61E-02 | 2.57E-01 | 43 | 0.04 | 0.01 | 7.68E-03 | 0.00 | 8.51E-01 | - | - |
| Vitamin D intake (ug/day) | Nutrients | 11348 | 0.03 | 0.01 | 3.83E-04 | **1.33E-02** | 7 | 11147 | 0.00 | 0.01 | 9.26E-01 | 9.96E-01 | 222 | 0.01 | 0.01 | 8.35E-03 | 0.82 | 1.72E-02 | - | - |
| Sour milk, yoghurt (3% fat) | Food | 11348 | -0.01 | 0.01 | 7.95E-02 | 3.69E-01 | 51 | 11147 | -0.01 | 0.01 | 6.30E-02 | 3.00E-01 | 51 | -0.01 | 0.01 | 1.07E-02 | 0.00 | 8.93E-01 | - | - |
| Average portion size of vegetables based on photographic illustration of four sizes (smallest to largest) | Food | 11348 | -0.02 | 0.01 | 1.67E-02 | 1.40E-01 | 29 | 11147 | -0.01 | 0.01 | 2.47E-01 | 6.38E-01 | 94 | -0.01 | 0.01 | 1.13E-02 | 0.00 | 4.19E-01 | - | - |
| Bacon | Food | 11348 | 0.01 | 0.01 | 8.98E-02 | 3.83E-01 | 57 | 11147 | 0.01 | 0.01 | 6.08E-02 | 2.99E-01 | 48 | 0.01 | 0.01 | 1.17E-02 | 0.00 | 8.43E-01 | - | - |
| Plant based protein intake (g/day) | Nutrients | 11348 | -0.02 | 0.01 | 3.13E-02 | 2.08E-01 | 35 | 11147 | -0.01 | 0.01 | 1.68E-01 | 4.97E-01 | 82 | -0.01 | 0.01 | 1.21E-02 | 0.00 | 6.25E-01 | - | - |
| Eicosapentaenoic acid (EPA) intake (g/day) | Nutrients | 11348 | 0.01 | 0.01 | 6.74E-02 | 3.48E-01 | 47 | 11147 | 0.01 | 0.01 | 1.06E-01 | 3.93E-01 | 65 | 0.01 | 0.01 | 1.47E-02 | 0.00 | 9.24E-01 | - | - |
| Hamburger | Food | 11348 | 0.02 | 0.01 | 4.33E-02 | 2.51E-01 | 42 | 11147 | 0.01 | 0.01 | 1.63E-01 | 4.90E-01 | 81 | 0.01 | 0.01 | 1.52E-02 | 0.00 | 7.08E-01 | - | - |
| Sodas, soft drinks, juice | Beverage | 11348 | 0.02 | 0.01 | 3.71E-02 | 2.26E-01 | 40 | 11147 | 0.01 | 0.01 | 1.91E-01 | 5.51E-01 | 84 | 0.01 | 0.01 | 1.56E-02 | 0.00 | 6.48E-01 | - | - |
| Animal based protein intake (g/day) | Nutrients | 11348 | 0.01 | 0.01 | 4.12E-01 | 7.10E-01 | 138 | 11147 | 0.02 | 0.01 | 8.32E-03 | 1.12E-01 | 18 | 0.01 | 0.01 | 1.60E-02 | 0.45 | 1.76E-01 | - | - |
| Lariciresinol intake (ug/day) | Nutrients | 11348 | -0.01 | 0.01 | 3.77E-01 | 7.10E-01 | 126 | 11147 | -0.02 | 0.01 | 1.20E-02 | 1.38E-01 | 21 | -0.01 | 0.01 | 1.77E-02 | 0.32 | 2.25E-01 | - | - |
| Everyday exercise satisfaction | Physical activity | 12250 | 0.02 | 0.01 | 1.03E-02 | 1.04E-01 | 24 | 12048 | 0.01 | 0.01 | 4.82E-01 | 8.13E-01 | 144 | 0.01 | 0.01 | 1.91E-02 | 0.37 | 2.07E-01 | - | - |
| Fat intake (g/day) | Nutrients | 11348 | 0.02 | 0.01 | 4.22E-02 | 2.50E-01 | 41 | 11147 | 0.01 | 0.01 | 2.18E-01 | 5.90E-01 | 90 | 0.01 | 0.01 | 2.03E-02 | 0.00 | 6.10E-01 | - | - |
| Frequency of engaging in clubs, associations or study circles | Social | 8196 | -0.01 | 0.01 | 2.45E-01 | 6.02E-01 | 97 | 8073 | -0.02 | 0.01 | 3.36E-02 | 2.11E-01 | 38 | -0.01 | 0.01 | 2.13E-02 | 0.00 | 4.51E-01 | - | - |
| Cholesterol intake (g/day) | Nutrients | 11348 | 0.01 | 0.01 | 3.89E-01 | 7.10E-01 | 131 | 11147 | 0.02 | 0.01 | 1.47E-02 | 1.38E-01 | 26 | 0.01 | 0.01 | 2.13E-02 | 0.28 | 2.38E-01 | - | - |
| Apple, pear, peach, orange, mandarin and grapefruit | Food | 11348 | -0.01 | 0.01 | 2.72E-01 | 6.11E-01 | 108 | 11147 | -0.02 | 0.01 | 2.94E-02 | 1.99E-01 | 36 | -0.01 | 0.01 | 2.16E-02 | 0.00 | 4.10E-01 | - | - |
| Pancake, waffle, Swedish dumpling | Food | 11348 | -0.01 | 0.01 | 1.77E-01 | 5.16E-01 | 82 | 11147 | -0.02 | 0.01 | 5.80E-02 | 2.99E-01 | 47 | -0.01 | 0.01 | 2.24E-02 | 0.00 | 6.53E-01 | - | - |
| Smoking status: Former smokers vs non-smokers | Tobacco use | 8461 | 0.03 | 0.02 | 8.06E-02 | 3.69E-01 | 53 | 8317 | 0.03 | 0.02 | 1.38E-01 | 4.54E-01 | 74 | 0.03 | 0.01 | 2.24E-02 | 0.00 | 8.48E-01 | - | - |
| Cambridge physical activity index | Physical activity | 12087 | -0.01 | 0.01 | 1.68E-01 | 5.12E-01 | 80 | 11849 | -0.01 | 0.01 | 6.95E-02 | 3.13E-01 | 54 | -0.01 | 0.01 | 2.44E-02 | 0.00 | 7.23E-01 | - | - |
| Self-employed | Psychosocial | 12046 | -0.02 | 0.02 | 4.99E-01 | 7.73E-01 | 154 | 11840 | -0.06 | 0.03 | 1.33E-02 | 1.38E-01 | 22 | -0.04 | 0.02 | 2.80E-02 | 0.43 | 1.85E-01 | - | - |
| Steak, chop, e.g. | Food | 11348 | 0.01 | 0.01 | 3.93E-01 | 7.10E-01 | 133 | 11147 | 0.02 | 0.01 | 2.12E-02 | 1.63E-01 | 31 | 0.01 | 0.01 | 2.82E-02 | 0.19 | 2.68E-01 | - | - |
| Sum of Lariciresinol, Matairesinol, Pinoresinol, Secoisolariciresinol intake (ug/day) | Nutrients | 11348 | -0.01 | 0.01 | 4.18E-01 | 7.10E-01 | 143 | 11147 | -0.02 | 0.01 | 1.97E-02 | 1.60E-01 | 29 | -0.01 | 0.01 | 2.82E-02 | 0.22 | 2.57E-01 | - | - |
| Feel the need to reduce alcohol consumption | Alcohol | 11114 | 0.05 | 0.02 | 3.64E-02 | 2.26E-01 | 39 | 10991 | 0.02 | 0.02 | 3.46E-01 | 7.31E-01 | 115 | 0.04 | 0.02 | 3.13E-02 | 0.00 | 4.27E-01 | - | - |
| Sugar, honey, marmelade, jam | Food | 11348 | -0.01 | 0.01 | 3.28E-01 | 6.74E-01 | 118 | 11147 | -0.02 | 0.01 | 4.65E-02 | 2.57E-01 | 44 | -0.01 | 0.01 | 3.73E-02 | 0.00 | 4.44E-01 | - | - |
| Number of social interactions during a normal week | Social | 12260 | 0.01 | 0.01 | 6.25E-02 | 3.37E-01 | 45 | 12067 | 0.01 | 0.01 | 2.89E-01 | 6.66E-01 | 105 | 0.01 | 0.01 | 3.79E-02 | 0.00 | 5.93E-01 | - | - |
| Alcohol intake (g/day) | Alcohol | 11348 | 0.01 | 0.01 | 1.40E-01 | 4.81E-01 | 70 | 11147 | 0.01 | 0.01 | 1.46E-01 | 4.66E-01 | 76 | 0.01 | 0.01 | 3.82E-02 | 0.00 | 9.60E-01 | - | - |
| Pinoresinol intake (ug/day) | Nutrients | 11348 | -0.01 | 0.01 | 4.88E-01 | 7.73E-01 | 150 | 11147 | -0.02 | 0.01 | 2.28E-02 | 1.68E-01 | 33 | -0.01 | 0.01 | 3.83E-02 | 0.27 | 2.40E-01 | - | - |
| Last time a colleague visited you at home | Psychosocial | 12011 | 0.02 | 0.01 | 1.46E-02 | 1.27E-01 | 28 | 11753 | 0.00 | 0.01 | 7.16E-01 | 9.66E-01 | 179 | 0.01 | 0.01 | 4.36E-02 | 0.51 | 1.54E-01 | - | - |
| Number of friends that can come to your home at any time and feel at home | Social | 12240 | 0.02 | 0.01 | 3.33E-02 | 2.13E-01 | 38 | 12032 | 0.01 | 0.01 | 4.97E-01 | 8.16E-01 | 148 | 0.01 | 0.01 | 4.50E-02 | 0.00 | 3.24E-01 | - | - |
| Margarine for cooking | Food | 11348 | 0.02 | 0.01 | 1.62E-03 | **4.15E-02** | 9 | 11147 | 0.00 | 0.01 | 6.72E-01 | 9.45E-01 | 173 | 0.01 | 0.01 | 4.61E-02 | 0.84 | 1.32E-02 | - | - |
| Frequent social contacts with colleagues during work | Psychosocial | 12046 | 0.00 | 0.01 | 5.05E-01 | 7.73E-01 | 157 | 11810 | -0.02 | 0.01 | 3.39E-02 | 2.11E-01 | 39 | -0.01 | 0.01 | 5.12E-02 | 0.13 | 2.84E-01 | - | - |
| Milk, sour milk (3%) | Beverage | 11348 | 0.00 | 0.01 | 7.28E-01 | 8.81E-01 | 199 | 11147 | 0.02 | 0.01 | 1.45E-02 | 1.38E-01 | 25 | 0.01 | 0.01 | 5.30E-02 | 0.58 | 1.25E-01 | - | - |
| Cream, creme fraiche, sour cream | Food | 11348 | -0.02 | 0.01 | 1.43E-02 | 1.27E-01 | 27 | 11147 | 0.00 | 0.01 | 7.81E-01 | 9.75E-01 | 194 | -0.01 | 0.01 | 5.36E-02 | 0.58 | 1.25E-01 | - | - |
| Vitamin B3 intake (mg/day) | Nutrients | 11348 | 0.00 | 0.01 | 6.58E-01 | 8.51E-01 | 188 | 11147 | 0.02 | 0.01 | 1.98E-02 | 1.60E-01 | 30 | 0.01 | 0.01 | 5.48E-02 | 0.48 | 1.63E-01 | - | - |
| Fibre intake (g/day) | Nutrients | 11348 | -0.01 | 0.01 | 3.84E-01 | 7.10E-01 | 127 | 11147 | -0.02 | 0.01 | 6.46E-02 | 3.02E-01 | 52 | -0.01 | 0.01 | 5.69E-02 | 0.00 | 4.60E-01 | - | - |
| Overall state of health compared to other your age | General health | 12096 | 0.00 | 0.01 | 9.61E-01 | 9.69E-01 | 241 | 11918 | -0.02 | 0.01 | 5.83E-03 | 8.33E-02 | 17 | -0.01 | 0.01 | 5.87E-02 | 0.75 | 4.46E-02 | - | - |
| Overall state of health during the last year | General health | 12280 | 0.00 | 0.01 | 8.78E-01 | 9.40E-01 | 227 | 12067 | -0.02 | 0.01 | 1.07E-02 | 1.30E-01 | 20 | -0.01 | 0.01 | 5.89E-02 | 0.66 | 8.47E-02 | - | - |
| Control over own work assignment | Psychosocial | 12148 | 0.00 | 0.01 | 7.39E-01 | 8.89E-01 | 202 | 11920 | 0.02 | 0.01 | 1.83E-02 | 1.59E-01 | 28 | 0.01 | 0.01 | 6.22E-02 | 0.55 | 1.38E-01 | - | - |
| Frequency of dancing during leisure time | Physical activity | 10929 | 0.00 | 0.01 | 8.31E-01 | 9.39E-01 | 215 | 10724 | -0.02 | 0.01 | 4.05E-03 | 7.04E-02 | 14 | -0.01 | 0.01 | 6.80E-02 | 0.80 | 2.57E-02 | - | - |
| Enough time for job assignments | Psychosocial | 12099 | 0.01 | 0.01 | 1.35E-01 | 4.80E-01 | 68 | 11863 | 0.01 | 0.01 | 2.87E-01 | 6.66E-01 | 104 | 0.01 | 0.01 | 6.92E-02 | 0.00 | 7.94E-01 | - | - |
| Ingenuity or creativity demand from job | Psychosocial | 12091 | -0.02 | 0.01 | 1.12E-02 | 1.09E-01 | 25 | 11878 | 0.00 | 0.01 | 9.47E-01 | 9.96E-01 | 228 | -0.01 | 0.01 | 7.10E-02 | 0.69 | 7.47E-02 | - | - |
| Frequency of gardening during leisure time | Physical activity | 11654 | 0.01 | 0.01 | 9.32E-02 | 3.84E-01 | 59 | 11466 | 0.01 | 0.01 | 3.95E-01 | 7.57E-01 | 125 | 0.01 | 0.01 | 7.20E-02 | 0.00 | 5.80E-01 | - | - |
| Cohabitation: Live alone vs Adult and children | Social | 8848 | -0.03 | 0.03 | 2.53E-01 | 6.02E-01 | 101 | 8557 | -0.04 | 0.03 | 1.59E-01 | 4.84E-01 | 79 | -0.03 | 0.02 | 7.27E-02 | 0.00 | 7.87E-01 | - | - |
| Vitamin B6 intake (mg/day) | Nutrients | 11348 | 0.00 | 0.01 | 5.84E-01 | 8.09E-01 | 174 | 11147 | 0.02 | 0.01 | 4.23E-02 | 2.45E-01 | 42 | 0.01 | 0.01 | 7.30E-02 | 0.17 | 2.71E-01 | - | - |
| Matairesinol intake (ug/day) | Nutrients | 11348 | -0.01 | 0.01 | 3.49E-01 | 6.89E-01 | 123 | 11147 | -0.01 | 0.01 | 1.07E-01 | 3.93E-01 | 66 | -0.01 | 0.01 | 7.34E-02 | 0.00 | 6.02E-01 | - | - |
| Medium beer | Alcohol | 11348 | 0.00 | 0.01 | 8.08E-01 | 9.22E-01 | 213 | 11147 | 0.02 | 0.01 | 4.36E-03 | 7.07E-02 | 15 | 0.01 | 0.01 | 7.96E-02 | 0.80 | 2.37E-02 | - | - |
| Support from others | Social | 12265 | -0.01 | 0.01 | 1.73E-01 | 5.16E-01 | 81 | 12062 | -0.01 | 0.01 | 2.69E-01 | 6.53E-01 | 99 | -0.01 | 0.01 | 8.03E-02 | 0.00 | 8.81E-01 | - | - |
| Work shifts/weekends | Psychosocial | 11908 | 0.00 | 0.02 | 8.61E-01 | 9.40E-01 | 219 | 11709 | -0.04 | 0.02 | 2.15E-02 | 1.63E-01 | 32 | -0.02 | 0.01 | 8.23E-02 | 0.56 | 1.30E-01 | - | - |
| Average portion size of meat/fish based on photographic illustration of four sizes (smallest to largest) | Food | 11348 | 0.00 | 0.01 | 9.61E-01 | 9.69E-01 | 240 | 11147 | 0.02 | 0.01 | 1.02E-02 | 1.30E-01 | 19 | 0.01 | 0.01 | 8.24E-02 | 0.72 | 5.84E-02 | - | - |
| Feel uneasy or guilty because of your way of drinking | Alcohol | 11111 | 0.03 | 0.02 | 2.54E-01 | 6.02E-01 | 102 | 10982 | 0.03 | 0.02 | 2.01E-01 | 5.68E-01 | 86 | 0.03 | 0.02 | 8.75E-02 | 0.00 | 9.01E-01 | - | - |
| Polyunsaturated fat intake (g/day) | Nutrients | 11348 | 0.01 | 0.01 | 9.72E-02 | 3.94E-01 | 60 | 11147 | 0.01 | 0.01 | 4.64E-01 | 8.05E-01 | 140 | 0.01 | 0.01 | 8.80E-02 | 0.00 | 5.40E-01 | - | - |
| Sausage as main dish | Food | 11348 | 0.01 | 0.01 | 1.81E-01 | 5.16E-01 | 85 | 11147 | 0.01 | 0.01 | 2.96E-01 | 6.66E-01 | 108 | 0.01 | 0.01 | 9.07E-02 | 0.00 | 8.77E-01 | - | - |
| Rice | Food | 11348 | -0.02 | 0.01 | 2.96E-02 | 2.08E-01 | 34 | 11147 | 0.00 | 0.01 | 9.14E-01 | 9.96E-01 | 217 | -0.01 | 0.01 | 9.82E-02 | 0.50 | 1.56E-01 | - | - |

| | | | | | | | | | | | | | | | | | | | | |
|---|---|---|---|---|---|---|---|---|---|---|---|---|---|---|---|---|---|---|---|---|
| Years using snuff | Tobacco use | 11323 | -0.01 | 0.01 | 1.18E-01 | 4.36E-01 | 66 | 11100 | -0.01 | 0.01 | 4.54E-01 | 7.93E-01 | 138 | -0.01 | 0.01 | 1.00E-01 | 0.00 | 5.85E-01 | - | - |
| Breakfast habits: Gruel w/o sandwich for breakfast vs not breakfast at all | Food | 933 | -0.10 | 0.07 | 1.52E-01 | 4.94E-01 | 74 | 873 | -0.06 | 0.07 | 3.95E-01 | 7.57E-01 | 126 | -0.08 | 0.05 | 1.03E-01 | 0.00 | 7.23E-01 | - | - |
| Palmitic acid intake (g/day) | Nutrients | 11348 | 0.01 | 0.01 | 4.18E-01 | 7.10E-01 | 142 | 11147 | 0.01 | 0.01 | 1.35E-01 | 4.50E-01 | 73 | 0.01 | 0.01 | 1.06E-01 | 0.00 | 5.99E-01 | - | - |
| Marital status: Single vs Married/partner | Social | 11413 | -0.04 | 0.02 | 1.25E-01 | 4.53E-01 | 67 | 11182 | -0.02 | 0.02 | 4.81E-01 | 8.13E-01 | 143 | -0.03 | 0.02 | 1.12E-01 | 0.00 | 5.72E-01 | - | - |
| Shellfish (e.g. shrimps, scallops) | Food | 5080 | 0.02 | 0.01 | 1.12E-01 | 4.20E-01 | 65 | 4983 | 0.01 | 0.01 | 5.37E-01 | 8.53E-01 | 152 | 0.01 | 0.01 | 1.13E-01 | 0.00 | 5.34E-01 | - | - |
| Satisfaction with economy | Psychosocial | 6704 | 0.01 | 0.01 | 2.51E-01 | 6.02E-01 | 100 | 6582 | 0.01 | 0.01 | 2.90E-01 | 6.66E-01 | 106 | 0.01 | 0.01 | 1.19E-01 | 0.00 | 9.77E-01 | - | - |
| High mental demand from job | Psychosocial | 12061 | 0.01 | 0.01 | 1.80E-01 | 5.16E-01 | 83 | 11831 | 0.01 | 0.01 | 4.12E-01 | 7.71E-01 | 130 | 0.01 | 0.01 | 1.24E-01 | 0.00 | 7.42E-01 | - | - |
| Sour milk, yoghurt (low fat) | Food | 11348 | -0.01 | 0.01 | 8.65E-02 | 3.82E-01 | 55 | 11147 | 0.00 | 0.01 | 6.86E-01 | 9.53E-01 | 175 | -0.01 | 0.01 | 1.27E-01 | 0.00 | 3.78E-01 | - | - |
| Total protein intake (g/day) | Nutrients | 11348 | 0.00 | 0.01 | 9.44E-01 | 9.63E-01 | 238 | 11147 | 0.02 | 0.01 | 2.50E-02 | 1.74E-01 | 34 | 0.01 | 0.01 | 1.37E-01 | 0.65 | 9.30E-02 | - | - |
| Light and physically active work | Physical activity | 12108 | -0.01 | 0.02 | 5.58E-01 | 7.98E-01 | 170 | 11862 | -0.03 | 0.02 | 1.27E-01 | 4.50E-01 | 68 | -0.02 | 0.01 | 1.39E-01 | 0.00 | 4.88E-01 | - | - |
| Control over planning and execution of the workday | Psychosocial | 12157 | 0.00 | 0.01 | 8.00E-01 | 9.17E-01 | 212 | 11925 | 0.01 | 0.01 | 6.15E-02 | 2.99E-01 | 50 | 0.01 | 0.01 | 1.42E-01 | 0.29 | 2.35E-01 | - | - |
| Would you say that the number of people that you meet in your everyday life is enough or would you like to meet more or fewer people? | Social | 12263 | -0.01 | 0.01 | 1.52E-01 | 4.94E-01 | 75 | 12041 | 0.00 | 0.01 | 6.03E-01 | 9.10E-01 | 161 | -0.01 | 0.01 | 1.64E-01 | 0.00 | 5.38E-01 | - | - |
| Sum of all lignans intake (ug/day) | Nutrients | 11348 | 0.00 | 0.01 | 6.21E-01 | 8.33E-01 | 181 | 11147 | -0.01 | 0.01 | 1.44E-01 | 4.65E-01 | 75 | -0.01 | 0.01 | 1.72E-01 | 0.00 | 4.72E-01 | - | - |
| Chips, popcorn, salted nuts | Food | 11348 | 0.01 | 0.01 | 1.06E-01 | 4.08E-01 | 63 | 11147 | 0.00 | 0.01 | 7.90E-01 | 9.80E-01 | 196 | 0.01 | 0.01 | 1.73E-01 | 0.00 | 3.62E-01 | - | - |
| Breakfast habits: Coffee/tea and wheat buns or rusk for breakfast vs not breakfast at all | Food | 971 | -0.08 | 0.07 | 2.36E-01 | 6.02E-01 | 95 | 938 | -0.05 | 0.06 | 4.54E-01 | 7.93E-01 | 139 | -0.06 | 0.05 | 1.74E-01 | 0.00 | 7.27E-01 | - | - |
| Corn flakes | Food | 11348 | 0.00 | 0.01 | 5.17E-01 | 7.73E-01 | 161 | 11147 | -0.01 | 0.01 | 2.11E-01 | 5.79E-01 | 88 | -0.01 | 0.01 | 1.82E-01 | 0.00 | 6.50E-01 | - | - |
| Close relationship with anyone | Social | 12267 | 0.00 | 0.01 | 9.72E-01 | 9.76E-01 | 242 | 12065 | -0.01 | 0.01 | 4.97E-02 | 2.63E-01 | 45 | -0.01 | 0.01 | 1.82E-01 | 0.52 | 1.50E-01 | - | - |
| Long-term sickness | General health | 11893 | 0.03 | 0.02 | 2.26E-01 | 5.84E-01 | 94 | 11626 | 0.01 | 0.02 | 5.19E-01 | 8.44E-01 | 149 | 0.02 | 0.02 | 1.86E-01 | 0.00 | 7.12E-01 | - | - |
| Job demands to work very fast | Psychosocial | 12139 | 0.01 | 0.01 | 7.11E-02 | 3.53E-01 | 49 | 11913 | 0.00 | 0.01 | 9.95E-01 | 1.00E+00 | 241 | 0.01 | 0.01 | 1.87E-01 | 0.34 | 2.18E-01 | - | - |
| Wine | Alcohol | 11348 | 0.00 | 0.01 | 6.41E-01 | 8.42E-01 | 184 | 11147 | 0.01 | 0.01 | 1.57E-01 | 4.84E-01 | 77 | 0.01 | 0.01 | 1.90E-01 | 0.00 | 4.81E-01 | - | - |
| Ice cream | Food | 11348 | -0.01 | 0.01 | 3.38E-01 | 6.79E-01 | 121 | 11147 | 0.02 | 0.01 | 5.76E-03 | 8.33E-02 | 16 | 0.01 | 0.01 | 2.02E-01 | 0.86 | 8.54E-03 | - | - |
| Memory status | Psychosocial | 6684 | 0.01 | 0.01 | 4.91E-01 | 7.73E-01 | 151 | 6579 | 0.01 | 0.01 | 2.63E-01 | 6.52E-01 | 98 | 0.01 | 0.01 | 2.04E-01 | 0.00 | 7.32E-01 | - | - |
| Banana | Food | 11348 | 0.00 | 0.01 | 8.67E-01 | 9.40E-01 | 221 | 11147 | 0.01 | 0.01 | 9.99E-02 | 3.79E-01 | 64 | 0.01 | 0.01 | 2.08E-01 | 0.13 | 2.83E-01 | - | - |
| White meat (poultry) | Food | 11348 | 0.01 | 0.01 | 3.87E-01 | 7.10E-01 | 129 | 11147 | 0.01 | 0.01 | 3.64E-01 | 7.44E-01 | 119 | 0.01 | 0.01 | 2.11E-01 | 0.00 | 9.44E-01 | - | - |
| Enterodiol intake (ug/day) | Nutrients | 11348 | 0.00 | 0.01 | 7.68E-01 | 9.02E-01 | 207 | 11147 | 0.01 | 0.01 | 1.34E-01 | 4.50E-01 | 72 | 0.01 | 0.01 | 2.12E-01 | 0.00 | 3.78E-01 | - | - |
| Participation in study circles | Social | 763 | 0.07 | 0.08 | 3.67E-01 | 7.10E-01 | 124 | 680 | 0.08 | 0.10 | 3.88E-01 | 7.57E-01 | 122 | 0.07 | 0.06 | 2.14E-01 | 0.00 | 8.98E-01 | - | - |
| Linolenic acid intake (g/day) | Nutrients | 11348 | 0.01 | 0.01 | 8.04E-02 | 3.69E-01 | 52 | 11147 | 0.00 | 0.01 | 9.54E-01 | 9.96E-01 | 231 | 0.01 | 0.01 | 2.20E-01 | 0.36 | 2.12E-01 | - | - |
| Participation in sports or physical exercise associations | Social | 763 | -0.01 | 0.05 | 8.73E-01 | 9.40E-01 | 223 | 680 | -0.13 | 0.07 | 6.76E-02 | 3.10E-01 | 53 | -0.05 | 0.04 | 2.24E-01 | 0.47 | 1.68E-01 | - | - |
| Calcium intake (mg/day) | Nutrients | 11348 | -0.01 | 0.01 | 1.88E-01 | 5.17E-01 | 87 | 11147 | 0.00 | 0.01 | 7.25E-01 | 9.66E-01 | 182 | -0.01 | 0.01 | 2.31E-01 | 0.00 | 5.14E-01 | - | - |
| Light beer | Alcohol | 11348 | 0.01 | 0.01 | 2.16E-01 | 5.70E-01 | 92 | 11147 | 0.00 | 0.01 | 6.59E-01 | 9.34E-01 | 169 | 0.01 | 0.01 | 2.34E-01 | 0.00 | 5.79E-01 | - | - |
| Sodium intake (mg/day) | Nutrients | 11348 | 0.00 | 0.01 | 9.55E-01 | 9.69E-01 | 239 | 11147 | 0.01 | 0.01 | 9.60E-02 | 3.76E-01 | 62 | 0.01 | 0.01 | 2.36E-01 | 0.27 | 2.42E-01 | - | - |
| Frequency of hunting or fishing during leisure time | Physical activity | 11426 | 0.01 | 0.01 | 1.92E-01 | 5.17E-01 | 89 | 11254 | 0.00 | 0.01 | 7.48E-01 | 9.67E-01 | 188 | 0.01 | 0.01 | 2.44E-01 | 0.00 | 5.02E-01 | - | - |
| Saturated fat intake (g/day) | Nutrients | 11348 | 0.01 | 0.01 | 3.93E-01 | 7.10E-01 | 132 | 11147 | 0.01 | 0.01 | 4.30E-01 | 7.88E-01 | 132 | 0.01 | 0.01 | 2.45E-01 | 0.00 | 9.87E-01 | - | - |
| Sleep status | Sleep | 6712 | 0.00 | 0.01 | 8.74E-01 | 9.40E-01 | 224 | 6597 | -0.02 | 0.01 | 1.30E-01 | 4.50E-01 | 69 | -0.01 | 0.01 | 2.45E-01 | 0.00 | 3.24E-01 | - | - |
| Repetitive job | Psychosocial | 12140 | 0.01 | 0.01 | 1.66E-01 | 5.10E-01 | 79 | 11916 | 0.00 | 0.01 | 8.40E-01 | 9.96E-01 | 200 | 0.01 | 0.01 | 2.52E-01 | 0.00 | 4.21E-01 | - | - |
| Vitamin E intake (mg/day) | Nutrients | 11348 | 0.01 | 0.01 | 1.02E-01 | 4.00E-01 | 62 | 11147 | 0.00 | 0.01 | 9.47E-01 | 9.96E-01 | 229 | 0.01 | 0.01 | 2.56E-01 | 0.28 | 2.38E-01 | - | - |
| Syringaresinol intake (ug/day) | Nutrients | 11348 | 0.00 | 0.01 | 6.09E-01 | 8.22E-01 | 180 | 11147 | -0.01 | 0.01 | 2.82E-01 | 6.66E-01 | 102 | -0.01 | 0.01 | 2.66E-01 | 0.00 | 6.69E-01 | - | - |
| Tomato, cucumber | Food | 11348 | 0.00 | 0.01 | 5.74E-01 | 8.09E-01 | 172 | 11147 | 0.01 | 0.01 | 3.06E-01 | 6.76E-01 | 110 | 0.01 | 0.01 | 2.66E-01 | 0.00 | 7.21E-01 | - | - |
| Satisfaction with leisure time | Psychosocial | 6704 | 0.01 | 0.01 | 5.88E-01 | 8.09E-01 | 176 | 6573 | 0.01 | 0.01 | 3.05E-01 | 6.76E-01 | 109 | 0.01 | 0.01 | 2.71E-01 | 0.00 | 7.14E-01 | - | - |
| Vision status | General health | 6698 | 0.00 | 0.01 | 6.56E-01 | 8.51E-01 | 187 | 6589 | -0.01 | 0.01 | 2.60E-01 | 6.52E-01 | 96 | -0.01 | 0.01 | 2.72E-01 | 0.00 | 6.09E-01 | - | - |
| Energy status | Psychosocial | 6697 | 0.01 | 0.01 | 5.46E-01 | 7.89E-01 | 168 | 6575 | 0.01 | 0.01 | 3.41E-01 | 7.31E-01 | 113 | 0.01 | 0.01 | 2.74E-01 | 0.00 | 7.86E-01 | - | - |
| Teetotaler | Alcohol | 12267 | -0.03 | 0.02 | 3.06E-01 | 6.48E-01 | 112 | 12069 | -0.01 | 0.03 | 6.27E-01 | 9.30E-01 | 164 | -0.02 | 0.02 | 2.80E-01 | 0.00 | 7.31E-01 | - | - |
| Possibility to speak with colleagues during breaks | Psychosocial | 12128 | 0.00 | 0.01 | 5.35E-01 | 7.83E-01 | 166 | 11880 | -0.01 | 0.01 | 3.67E-01 | 7.44E-01 | 120 | -0.01 | 0.01 | 2.84E-01 | 0.00 | 8.20E-01 | - | - |
| Frequncy of social contacts with colleagues during leisure time | Psychosocial | 11861 | 0.00 | 0.01 | 6.99E-01 | 8.67E-01 | 196 | 11616 | -0.01 | 0.01 | 2.55E-01 | 6.51E-01 | 95 | -0.01 | 0.01 | 2.87E-01 | 0.00 | 5.75E-01 | - | - |
| Sedentary or standing work | Physical activity | 12108 | 0.01 | 0.02 | 5.05E-01 | 7.73E-01 | 155 | 11862 | 0.01 | 0.02 | 4.02E-01 | 7.57E-01 | 129 | 0.01 | 0.01 | 2.88E-01 | 0.00 | 8.87E-01 | - | - |
| Cohabitation: Live alone vs Only one adult (spouse, partner) | Social | 3672 | -0.02 | 0.03 | 5.16E-01 | 7.73E-01 | 160 | 3744 | -0.03 | 0.03 | 3.92E-01 | 7.57E-01 | 123 | -0.02 | 0.02 | 2.89E-01 | 0.00 | 8.59E-01 | - | - |
| Mood status | Psychosocial | 6699 | 0.01 | 0.01 | 2.41E-01 | 6.02E-01 | 96 | 6584 | 0.00 | 0.01 | 7.62E-01 | 9.69E-01 | 191 | 0.01 | 0.01 | 2.90E-01 | 0.00 | 5.57E-01 | - | - |
| Total energy intake (kcal/day) | Nutrients | 11348 | 0.01 | 0.01 | 2.55E-01 | 6.02E-01 | 103 | 11147 | 0.00 | 0.01 | 7.44E-01 | 9.67E-01 | 187 | 0.01 | 0.01 | 2.93E-01 | 0.00 | 5.87E-01 | - | - |
| Fried potatoes, pommes frites | Food | 11348 | 0.00 | 0.01 | 5.25E-01 | 7.73E-01 | 165 | 11147 | 0.01 | 0.01 | 3.94E-01 | 7.57E-01 | 124 | 0.01 | 0.01 | 2.95E-01 | 0.00 | 8.60E-01 | - | - |
| Brewed (filtered) coffee | Beverage | 11348 | 0.00 | 0.01 | 7.28E-01 | 8.81E-01 | 200 | 11147 | 0.01 | 0.01 | 6.13E-02 | 2.99E-01 | 49 | 0.01 | 0.01 | 2.99E-01 | 0.61 | 1.11E-01 | - | - |
| Cheese 28% | Food | 11348 | -0.01 | 0.01 | 9.14E-02 | 3.83E-01 | 58 | 11147 | 0.00 | 0.01 | 7.60E-01 | 9.69E-01 | 190 | -0.01 | 0.01 | 3.11E-01 | 0.48 | 1.66E-01 | - | - |
| Campestanol intake (mg/day) | Nutrients | 11348 | 0.00 | 0.01 | 5.15E-01 | 7.73E-01 | 159 | 11147 | -0.01 | 0.01 | 4.45E-01 | 7.93E-01 | 134 | -0.01 | 0.01 | 3.18E-01 | 0.00 | 9.17E-01 | - | - |
| Berries (fresh or frozen) | Food | 11348 | 0.00 | 0.01 | 5.51E-01 | 7.92E-01 | 169 | 11147 | -0.02 | 0.01 | 4.10E-02 | 2.43E-01 | 41 | -0.01 | 0.01 | 3.26E-01 | 0.72 | 5.90E-02 | - | - |
| Snuff status: Former snuff users vs non-snuff users | Tobacco use | 9474 | 0.04 | 0.03 | 1.60E-01 | 5.04E-01 | 77 | 9345 | 0.00 | 0.02 | 9.83E-01 | 1.00E+00 | 238 | 0.02 | 0.02 | 3.35E-01 | 0.05 | 3.06E-01 | - | - |
| Beta-sitosterol intake (mg/day) | Nutrients | 11348 | 0.00 | 0.01 | 7.74E-01 | 9.04E-01 | 208 | 11147 | -0.01 | 0.01 | 9.11E-02 | 3.75E-01 | 59 | -0.01 | 0.01 | 3.38E-01 | 0.50 | 1.55E-01 | - | - |
| Root vegetables, carrot | Food | 11348 | -0.01 | 0.01 | 4.13E-01 | 7.10E-01 | 140 | 11147 | 0.00 | 0.01 | 5.97E-01 | 9.06E-01 | 160 | -0.01 | 0.01 | 3.39E-01 | 0.00 | 8.51E-01 | - | - |
| Trans fat intake (g/day) | Nutrients | 11348 | 0.01 | 0.01 | 1.58E-01 | 5.04E-01 | 76 | 11147 | 0.00 | 0.01 | 9.34E-01 | 9.96E-01 | 224 | 0.01 | 0.01 | 3.39E-01 | 0.08 | 2.98E-01 | - | - |
| Smoking status: Former occasional smokers vs non-smokers | Tobacco use | 7116 | 0.03 | 0.03 | 2.67E-01 | 6.11E-01 | 106 | 6889 | 0.01 | 0.03 | 8.15E-01 | 9.96E-01 | 198 | 0.02 | 0.02 | 3.46E-01 | 0.00 | 5.28E-01 | - | - |
| Salty fish | Food | 11348 | 0.00 | 0.01 | 6.66E-01 | 8.52E-01 | 190 | 11147 | 0.01 | 0.01 | 3.61E-01 | 7.44E-01 | 118 | 0.00 | 0.01 | 3.46E-01 | 0.00 | 7.17E-01 | - | - |
| Sometimes physically straining work | Physical activity | 12108 | 0.03 | 0.02 | 1.01E-01 | 4.00E-01 | 61 | 11862 | -0.01 | 0.02 | 7.17E-01 | 9.66E-01 | 180 | 0.01 | 0.01 | 3.51E-01 | 0.49 | 1.63E-01 | - | - |
| Pasta | Food | 11348 | -0.01 | 0.01 | 1.65E-01 | 5.10E-01 | 78 | 11147 | 0.00 | 0.01 | 9.13E-01 | 9.96E-01 | 216 | -0.01 | 0.01 | 3.56E-01 | 0.08 | 2.96E-01 | - | - |
| Monosaccharides intake (g/day) | Nutrients | 11348 | 0.00 | 0.01 | 7.89E-01 | 9.09E-01 | 211 | 11147 | -0.01 | 0.01 | 2.95E-01 | 6.66E-01 | 107 | -0.01 | 0.01 | 3.60E-01 | 0.00 | 5.65E-01 | - | - |
| Beta-carotene intake (mg/day) | Nutrients | 11348 | -0.01 | 0.01 | 3.88E-01 | 7.10E-01 | 130 | 11147 | 0.00 | 0.01 | 6.78E-01 | 9.47E-01 | 174 | 0.00 | 0.01 | 3.63E-01 | 0.00 | 7.64E-01 | - | - |
| Appetite status | Psychosocial | 6699 | 0.00 | 0.01 | 7.02E-01 | 8.67E-01 | 197 | 6585 | 0.01 | 0.01 | 3.85E-01 | 7.57E-01 | 121 | 0.01 | 0.01 | 3.81E-01 | 0.00 | 7.16E-01 | - | - |
| Secoisolariciresinol intake (ug/day) | Nutrients | 11348 | 0.00 | 0.01 | 6.50E-01 | 8.49E-01 | 186 | 11147 | -0.01 | 0.01 | 4.31E-01 | 7.88E-01 | 133 | 0.00 | 0.01 | 3.84E-01 | 0.00 | 7.97E-01 | - | - |
| White (soft) bread, thin crisp bread | Food | 11348 | -0.01 | 0.01 | 8.28E-02 | 3.73E-01 | 54 | 11147 | 0.00 | 0.01 | 5.69E-01 | 8.81E-01 | 157 | 0.00 | 0.01 | 3.89E-01 | 0.61 | 1.08E-01 | - | - |
| Beta-sitostanol intake (mg/day) | Nutrients | 11348 | 0.00 | 0.01 | 5.96E-01 | 8.09E-01 | 179 | 11147 | -0.01 | 0.01 | 4.97E-01 | 8.16E-01 | 147 | 0.00 | 0.01 | 3.94E-01 | 0.00 | 9.00E-01 | - | - |
| Satisfaction with home and family situation | Psychosocial | 6706 | 0.01 | 0.01 | 1.88E-01 | 5.17E-01 | 88 | 6589 | 0.00 | 0.01 | 8.80E-01 | 9.96E-01 | 208 | 0.01 | 0.01 | 3.97E-01 | 0.04 | 3.08E-01 | - | - |
| Blood based food | Food | 5080 | 0.02 | 0.01 | 1.45E-01 | 4.84E-01 | 73 | 4983 | 0.00 | 0.01 | 6.97E-01 | 9.57E-01 | 177 | 0.01 | 0.01 | 4.10E-01 | 0.37 | 2.07E-01 | - | - |
| Meat on bread | Food | 11348 | 0.01 | 0.01 | 4.12E-01 | 7.10E-01 | 139 | 11147 | 0.00 | 0.01 | 7.44E-01 | 9.67E-01 | 186 | 0.00 | 0.01 | 4.11E-01 | 0.00 | 7.47E-01 | - | - |
| Participation in other association | Social | 763 | 0.06 | 0.05 | 2.69E-01 | 6.11E-01 | 107 | 680 | -0.01 | 0.07 | 9.03E-01 | 9.96E-01 | 214 | 0.03 | 0.04 | 4.14E-01 | 0.00 | 4.50E-01 | - | - |
| Frequency of walking during leisure time | Physical activity | 11938 | 0.00 | 0.01 | 5.77E-01 | 8.09E-01 | 173 | 11791 | 0.00 | 0.01 | 5.68E-01 | 8.81E-01 | 156 | 0.00 | 0.01 | 4.25E-01 | 0.00 | 9.82E-01 | - | - |
| High physical demand from job | Physical activity | 12171 | 0.02 | 0.01 | 2.86E-02 | 2.08E-01 | 33 | 11942 | -0.01 | 0.01 | 2.43E-01 | 6.34E-01 | 93 | 0.00 | 0.01 | 4.27E-01 | 0.82 | 1.88E-02 | - | - |
| Equol intake (ug/day) | Nutrients | 11348 | -0.01 | 0.01 | 4.09E-01 | 7.10E-01 | 136 | 11147 | 0.00 | 0.01 | 7.82E-01 | 9.75E-01 | 195 | 0.00 | 0.01 | 4.30E-01 | 0.00 | 7.12E-01 | - | - |
| Light but partly physically active work | Physical activity | 12108 | -0.04 | 0.02 | 3.16E-02 | 2.08E-01 | 36 | 11862 | 0.02 | 0.02 | 2.69E-01 | 6.53E-01 | 100 | -0.01 | 0.01 | 4.33E-01 | 0.81 | 2.22E-02 | - | - |
| Meat stew | Food | 11348 | -0.01 | 0.01 | 2.83E-01 | 6.30E-01 | 109 | 11147 | 0.02 | 0.01 | 2.51E-02 | 1.74E-01 | 35 | 0.00 | 0.01 | 4.49E-01 | 0.82 | 1.80E-02 | - | - |
| Contradictory demands in job | Psychosocial | 12058 | -0.01 | 0.01 | 3.94E-01 | 7.10E-01 | 134 | 11832 | 0.00 | 0.01 | 8.50E-01 | 9.96E-01 | 202 | 0.00 | 0.01 | 4.53E-01 | 0.00 | 6.54E-01 | - | - |
| Skill demand from job | Psychosocial | 12112 | -0.01 | 0.01 | 3.28E-01 | 6.74E-01 | 117 | 11900 | 0.00 | 0.01 | 9.65E-01 | 9.96E-01 | 234 | 0.00 | 0.01 | 4.60E-01 | 0.00 | 5.21E-01 | - | - |
| Iodine intake (ug/day) | Nutrients | 11348 | 0.00 | 0.01 | 7.86E-01 | 9.09E-01 | 209 | 11147 | 0.01 | 0.01 | 1.76E-01 | 5.14E-01 | 83 | 0.00 | 0.01 | 4.65E-01 | 0.27 | 2.41E-01 | - | - |
| Changed everyday exercise during the last year | Physical activity | 12288 | 0.01 | 0.01 | 1.80E-01 | 5.16E-01 | 84 | 12078 | 0.00 | 0.01 | 7.23E-01 | 9.66E-01 | 181 | 0.00 | 0.01 | 4.69E-01 | 0.29 | 2.36E-01 | - | - |
| Butter on bread | Food | 11348 | 0.01 | 0.01 | 1.40E-01 | 4.81E-01 | 71 | 11147 | 0.00 | 0.01 | 5.86E-01 | 8.97E-01 | 158 | 0.00 | 0.01 | 4.75E-01 | 0.49 | 1.62E-01 | - | - |
| Strong beer | Alcohol | 11348 | 0.01 | 0.01 | 3.06E-01 | 6.48E-01 | 111 | 11147 | 0.00 | 0.01 | 9.42E-01 | 9.96E-01 | 226 | 0.00 | 0.01 | 4.80E-01 | 0.00 | 4.56E-01 | - | - |
| Linoleic acid intake (g/day) | Nutrients | 11348 | 0.00 | 0.01 | 5.22E-01 | 7.73E-01 | 163 | 11147 | 0.00 | 0.01 | 7.28E-01 | 9.66E-01 | 183 | 0.00 | 0.01 | 4.82E-01 | 0.00 | 8.50E-01 | - | - |
| Frequency of cycling during leisure time | Physical activity | 10562 | -0.01 | 0.01 | 5.05E-01 | 7.73E-01 | 156 | 10340 | 0.00 | 0.01 | 7.53E-01 | 9.68E-01 | 189 | 0.00 | 0.01 | 4.85E-01 | 0.00 | 8.11E-01 | - | - |
| Whole grain soft bread | Food | 11348 | 0.00 | 0.01 | 6.98E-01 | 8.67E-01 | 195 | 11147 | 0.00 | 0.01 | 5.54E-01 | 8.68E-01 | 155 | 0.00 | 0.01 | 4.91E-01 | 0.00 | 8.70E-01 | - | - |
| Mashed potato | Food | 5080 | 0.01 | 0.01 | 4.22E-01 | 7.13E-01 | 144 | 4983 | 0.00 | 0.01 | 9.22E-01 | 9.96E-01 | 221 | 0.01 | 0.01 | 5.10E-01 | 0.00 | 6.39E-01 | - | - |
| Potato salad | Food | 5080 | -0.02 | 0.01 | 8.97E-02 | 3.83E-01 | 56 | 4983 | 0.01 | 0.01 | 3.97E-01 | 7.57E-01 | 127 | -0.01 | 0.01 | 5.11E-01 | 0.68 | 7.52E-02 | - | - |
| Marital status: Single vs Divorced/separated | Social | 1991 | 0.02 | 0.04 | 5.73E-01 | 8.09E-01 | 171 | 1954 | 0.02 | 0.05 | 7.36E-01 | 9.67E-01 | 184 | 0.02 | 0.03 | 5.15E-01 | 0.00 | 9.28E-01 | - | - |
| Liver, kidney | Food | 5080 | 0.01 | 0.01 | 3.46E-01 | 6.89E-01 | 122 | 4983 | 0.00 | 0.01 | 9.50E-01 | 9.96E-01 | 230 | 0.01 | 0.01 | 5.17E-01 | 0.00 | 4.92E-01 | - | - |

| | | | | | | | | | | | | | | | | | | | | |
|---|---|---|---|---|---|---|---|---|---|---|---|---|---|---|---|---|---|---|---|---|
| Number of social contacts with the same interests as you | Social | 12242 | 0.01 | 0.01 | 3.72E-01 | 7.10E-01 | 125 | 12031 | 0.00 | 0.01 | 1.00E+00 | 1.00E+00 | 243 | 0.00 | 0.01 | 5.20E-01 | 0.00 | 5.35E-01 | - | - |
| Whole grain crisp bread | Food | 11348 | 0.01 | 0.01 | 3.07E-01 | 6.48E-01 | 113 | 11147 | 0.00 | 0.01 | 8.87E-01 | 9.96E-01 | 210 | 0.00 | 0.01 | 5.21E-01 | 0.00 | 4.19E-01 | - | - |
| Sum of phytosterols intake (mg/day) | Nutrients | 11348 | 0.00 | 0.01 | 5.13E-01 | 7.73E-01 | 158 | 11147 | -0.01 | 0.01 | 1.08E-01 | 3.93E-01 | 67 | 0.00 | 0.01 | 5.26E-01 | 0.62 | 1.07E-01 | - | - |
| Soft cheese | Food | 5080 | 0.00 | 0.01 | 8.49E-01 | 9.40E-01 | 217 | 4983 | 0.01 | 0.01 | 2.82E-01 | 6.66E-01 | 103 | 0.01 | 0.01 | 5.29E-01 | 0.00 | 3.72E-01 | - | - |
| Vitamin C intake (mg/day) | Nutrients | 11348 | 0.00 | 0.01 | 8.49E-01 | 9.40E-01 | 218 | 11147 | -0.01 | 0.01 | 4.74E-01 | 8.12E-01 | 142 | 0.00 | 0.01 | 5.29E-01 | 0.00 | 6.96E-01 | - | - |
| Mixed frozen vegetables | Food | 5080 | 0.01 | 0.01 | 3.33E-01 | 6.74E-01 | 120 | 4983 | 0.00 | 0.01 | 9.17E-01 | 9.96E-01 | 218 | 0.00 | 0.01 | 5.29E-01 | 0.00 | 4.57E-01 | - | - |
| Do you feel important and appreciated in your home? | Psychosocial | 6661 | 0.01 | 0.01 | 4.71E-01 | 7.73E-01 | 148 | 6535 | -0.02 | 0.01 | 9.90E-02 | 3.79E-01 | 63 | 0.00 | 0.01 | 5.35E-01 | 0.65 | 9.09E-02 | - | - |
| Stigmasterol intake (mg/day) | Nutrients | 11348 | 0.01 | 0.01 | 4.01E-01 | 7.10E-01 | 135 | 11147 | 0.00 | 0.01 | 9.71E-01 | 9.96E-01 | 236 | 0.00 | 0.01 | 5.60E-01 | 0.00 | 5.45E-01 | - | - |
| Number of people with whom you can speak openly | Social | 12256 | 0.01 | 0.01 | 2.47E-01 | 6.02E-01 | 98 | 12054 | 0.00 | 0.01 | 7.15E-01 | 9.66E-01 | 178 | 0.00 | 0.01 | 5.61E-01 | 0.12 | 2.86E-01 | - | - |
| Smoked fish/meat | Food | 11348 | 0.00 | 0.01 | 7.19E-01 | 8.81E-01 | 198 | 11147 | 0.00 | 0.01 | 6.45E-01 | 9.34E-01 | 167 | 0.00 | 0.01 | 5.63E-01 | 0.00 | 9.32E-01 | - | - |
| Cheese 10-17% | Food | 11348 | -0.01 | 0.01 | 4.98E-01 | 7.73E-01 | 153 | 11147 | 0.00 | 0.01 | 8.98E-01 | 9.96E-01 | 213 | 0.00 | 0.01 | 5.64E-01 | 0.00 | 7.06E-01 | - | - |
| Magnesium intake (mg/day) | Nutrients | 11348 | -0.01 | 0.01 | 1.94E-01 | 5.17E-01 | 91 | 11147 | 0.00 | 0.01 | 5.87E-01 | 8.97E-01 | 159 | 0.00 | 0.01 | 5.72E-01 | 0.40 | 1.97E-01 | - | - |
| Permanent employment | Psychosocial | 12046 | -0.01 | 0.02 | 6.75E-01 | 8.55E-01 | 192 | 11840 | 0.02 | 0.02 | 2.12E-01 | 5.79E-01 | 89 | 0.01 | 0.01 | 5.73E-01 | 0.29 | 2.34E-01 | - | - |
| Iron intake (mg/day) | Nutrients | 11348 | -0.01 | 0.01 | 4.41E-01 | 7.38E-01 | 145 | 11147 | 0.00 | 0.01 | 9.84E-01 | 1.00E+00 | 239 | 0.00 | 0.01 | 5.84E-01 | 0.00 | 5.87E-01 | - | - |
| Pizza | Food | 11348 | 0.01 | 0.01 | 3.85E-01 | 7.10E-01 | 128 | 11147 | 0.00 | 0.01 | 8.60E-01 | 9.96E-01 | 205 | 0.00 | 0.01 | 6.12E-01 | 0.00 | 4.68E-01 | - | - |
| Satisfaction with work situation | Psychosocial | 6645 | -0.01 | 0.01 | 5.23E-01 | 7.73E-01 | 164 | 6521 | 0.00 | 0.01 | 9.56E-01 | 9.96E-01 | 232 | 0.00 | 0.01 | 6.17E-01 | 0.00 | 6.88E-01 | - | - |
| Receive hugs to comfort and support you | Social | 12211 | 0.00 | 0.02 | 9.06E-01 | 9.49E-01 | 232 | 12000 | -0.01 | 0.02 | 5.51E-01 | 8.68E-01 | 154 | -0.01 | 0.01 | 6.20E-01 | 0.00 | 7.25E-01 | - | - |
| Milk, sour milk (1,5%) | Beverage | 11348 | 0.01 | 0.01 | 2.66E-01 | 6.11E-01 | 104 | 11147 | 0.00 | 0.01 | 6.45E-01 | 9.34E-01 | 166 | 0.00 | 0.01 | 6.25E-01 | 0.17 | 2.71E-01 | - | - |
| Parents or siblings had a cerebral hemorrhage/thrombosis or cardiac infarction before the age of 60 | General health | 12159 | 0.00 | 0.02 | 9.87E-01 | 9.87E-01 | 243 | 11968 | 0.01 | 0.02 | 4.92E-01 | 8.16E-01 | 145 | 0.01 | 0.01 | 6.25E-01 | 0.00 | 6.29E-01 | - | - |
| Marital status: Single vs Widow/widower | Social | 1282 | -0.07 | 0.11 | 4.97E-01 | 7.73E-01 | 152 | 1252 | 0.00 | 0.12 | 9.72E-01 | 9.96E-01 | 237 | -0.04 | 0.08 | 6.33E-01 | 0.00 | 6.29E-01 | - | - |
| Disaccharides intake (g/day) | Nutrients | 11348 | 0.00 | 0.01 | 8.75E-01 | 9.40E-01 | 226 | 11147 | 0.00 | 0.01 | 6.12E-01 | 9.18E-01 | 162 | 0.00 | 0.01 | 6.43E-01 | 0.00 | 7.95E-01 | - | - |
| People to ask for help apart from the ones at home | Social | 12238 | 0.00 | 0.02 | 8.75E-01 | 9.40E-01 | 225 | 12040 | 0.01 | 0.02 | 6.16E-01 | 9.18E-01 | 163 | 0.01 | 0.02 | 6.43E-01 | 0.00 | 8.03E-01 | - | - |
| Learn new things at job | Psychosocial | 12119 | 0.00 | 0.01 | 5.41E-01 | 7.87E-01 | 167 | 11905 | 0.00 | 0.01 | 9.87E-01 | 1.00E+00 | 240 | 0.00 | 0.01 | 6.49E-01 | 0.00 | 6.83E-01 | - | - |
| Eat breakfast from 2000 | Food | 1191 | -0.10 | 0.08 | 2.21E-01 | 5.76E-01 | 93 | 1069 | 0.06 | 0.09 | 4.97E-01 | 8.16E-01 | 146 | -0.03 | 0.06 | 6.53E-01 | 0.43 | 1.84E-01 | - | - |
| Cohabitation: Live alone vs Other/others | Social | 1262 | 0.08 | 0.07 | 2.66E-01 | 6.11E-01 | 105 | 1225 | -0.06 | 0.09 | 5.25E-01 | 8.45E-01 | 151 | 0.03 | 0.06 | 6.53E-01 | 0.31 | 2.30E-01 | - | - |
| Folic acid intake (ug/day) | Nutrients | 11348 | 0.00 | 0.01 | 6.37E-01 | 8.42E-01 | 182 | 11147 | 0.00 | 0.01 | 8.77E-01 | 9.96E-01 | 207 | 0.00 | 0.01 | 6.54E-01 | 0.00 | 8.31E-01 | - | - |
| Pentadecanoic acid intake (g/day) | Nutrients | 11348 | -0.01 | 0.01 | 3.07E-01 | 6.48E-01 | 114 | 11147 | 0.00 | 0.01 | 6.60E-01 | 9.34E-01 | 171 | 0.00 | 0.01 | 6.61E-01 | 0.04 | 3.07E-01 | - | - |
| Heptadecanoic acid intake (g/day) | Nutrients | 11348 | -0.01 | 0.01 | 3.07E-01 | 6.48E-01 | 115 | 11147 | 0.00 | 0.01 | 6.60E-01 | 9.34E-01 | 170 | 0.00 | 0.01 | 6.61E-01 | 0.04 | 3.07E-01 | - | - |
| Possibility to leave your work for a while to speak with a colleague | Psychosocial | 12074 | 0.00 | 0.01 | 8.86E-01 | 9.41E-01 | 229 | 11804 | 0.00 | 0.01 | 6.38E-01 | 9.34E-01 | 165 | 0.00 | 0.01 | 6.69E-01 | 0.00 | 8.07E-01 | - | - |
| Medioresinol intake (ug/day) | Nutrients | 11348 | 0.00 | 0.01 | 8.97E-01 | 9.44E-01 | 231 | 11147 | -0.01 | 0.01 | 4.54E-01 | 7.93E-01 | 137 | 0.00 | 0.01 | 6.72E-01 | 0.00 | 5.28E-01 | - | - |
| Phosphate intake (mg/day) | Nutrients | 11348 | -0.01 | 0.01 | 1.43E-01 | 4.81E-01 | 72 | 11147 | 0.01 | 0.01 | 3.46E-01 | 7.31E-01 | 114 | 0.00 | 0.01 | 6.81E-01 | 0.65 | 9.03E-02 | - | - |
| Vitamin B2 intake (ug/day) | Nutrients | 11348 | -0.01 | 0.01 | 2.49E-01 | 6.02E-01 | 99 | 11147 | 0.00 | 0.01 | 5.37E-01 | 8.53E-01 | 153 | 0.00 | 0.01 | 6.82E-01 | 0.35 | 2.15E-01 | - | - |
| Travel to work: Walk to work vs passive travel to work | Physical activity | 7953 | -0.02 | 0.03 | 4.11E-01 | 7.10E-01 | 137 | 7787 | 0.01 | 0.03 | 7.77E-01 | 9.75E-01 | 193 | -0.01 | 0.02 | 6.96E-01 | 0.00 | 4.38E-01 | - | - |
| Enterolactone intake (ug/day) | Nutrients | 11348 | -0.01 | 0.01 | 4.51E-01 | 7.50E-01 | 146 | 11147 | 0.00 | 0.01 | 8.19E-01 | 9.96E-01 | 199 | 0.00 | 0.01 | 6.98E-01 | 0.00 | 4.92E-01 | - | - |
| Butter for cooking | Food | 11348 | -0.01 | 0.01 | 4.86E-01 | 7.73E-01 | 149 | 11147 | 0.01 | 0.01 | 2.08E-01 | 5.79E-01 | 87 | 0.00 | 0.01 | 7.14E-01 | 0.48 | 1.64E-01 | - | - |
| Bregott on bread | Food | 11348 | 0.00 | 0.01 | 8.26E-01 | 9.38E-01 | 214 | 11147 | 0.00 | 0.01 | 7.70E-01 | 9.75E-01 | 192 | 0.00 | 0.01 | 7.18E-01 | 0.00 | 9.52E-01 | - | - |
| Travel to work: Irregular travel mode to work vs passive travel to work | Physical activity | 8081 | -0.01 | 0.03 | 7.50E-01 | 8.93E-01 | 204 | 7958 | 0.02 | 0.03 | 4.00E-01 | 7.57E-01 | 128 | 0.01 | 0.02 | 7.20E-01 | 0.00 | 4.09E-01 | - | - |
| Brown beans, pea soup | Food | 11348 | 0.00 | 0.01 | 6.70E-01 | 8.52E-01 | 191 | 11147 | 0.00 | 0.01 | 9.57E-01 | 9.96E-01 | 233 | 0.00 | 0.01 | 7.28E-01 | 0.00 | 8.00E-01 | - | - |
| Oil for cooking | Food | 11348 | 0.00 | 0.01 | 9.19E-01 | 9.59E-01 | 233 | 11147 | 0.00 | 0.01 | 6.92E-01 | 9.56E-01 | 176 | 0.00 | 0.01 | 7.29E-01 | 0.00 | 8.29E-01 | - | - |
| Sucrose intake (g/day) | Nutrients | 11348 | 0.00 | 0.01 | 6.97E-01 | 8.67E-01 | 194 | 11147 | -0.01 | 0.01 | 3.61E-01 | 7.44E-01 | 117 | 0.00 | 0.01 | 7.31E-01 | 0.00 | 3.52E-01 | - | - |
| Coffee rolls/buns, rusk | Food | 11348 | 0.00 | 0.01 | 7.87E-01 | 9.09E-01 | 210 | 11147 | -0.01 | 0.01 | 4.46E-01 | 7.93E-01 | 135 | 0.00 | 0.01 | 7.39E-01 | 0.00 | 4.61E-01 | - | - |
| Low fat margarine on bread | Food | 11348 | 0.01 | 0.01 | 4.16E-01 | 7.10E-01 | 141 | 11147 | -0.01 | 0.01 | 1.95E-01 | 5.57E-01 | 85 | 0.00 | 0.01 | 7.61E-01 | 0.56 | 1.34E-01 | - | - |
| Fitness status | Physical activity | 6696 | 0.01 | 0.01 | 5.86E-01 | 8.09E-01 | 175 | 6582 | 0.00 | 0.01 | 8.96E-01 | 9.96E-01 | 212 | 0.00 | 0.01 | 7.62E-01 | 0.00 | 6.38E-01 | - | - |
| Patience status | Psychosocial | 6700 | 0.01 | 0.01 | 5.90E-01 | 8.09E-01 | 177 | 6586 | 0.00 | 0.01 | 8.96E-01 | 9.96E-01 | 211 | 0.00 | 0.01 | 7.65E-01 | 0.00 | 6.40E-01 | - | - |
| Rosehip, sweet syrup soup | Food | 11348 | 0.00 | 0.01 | 5.94E-01 | 8.09E-01 | 178 | 11147 | 0.00 | 0.01 | 8.56E-01 | 9.96E-01 | 203 | 0.00 | 0.01 | 7.91E-01 | 0.00 | 6.20E-01 | - | - |
| Formic acid intaje (g/day) | Nutrients | 11348 | 0.00 | 0.01 | 6.41E-01 | 8.42E-01 | 185 | 11147 | 0.00 | 0.01 | 9.06E-01 | 9.96E-01 | 215 | 0.00 | 0.01 | 7.98E-01 | 0.00 | 6.85E-01 | - | - |
| Tiamin intake (mg/day) | Nutrients | 11348 | 0.00 | 0.01 | 8.72E-01 | 9.40E-01 | 222 | 11147 | 0.00 | 0.01 | 8.57E-01 | 9.96E-01 | 204 | 0.00 | 0.01 | 8.09E-01 | 0.00 | 9.85E-01 | - | - |
| Zinc intake (mg/day) | Nutrients | 11348 | -0.01 | 0.01 | 3.10E-01 | 6.48E-01 | 116 | 11147 | 0.01 | 0.01 | 4.29E-01 | 7.88E-01 | 131 | 0.00 | 0.01 | 8.47E-01 | 0.38 | 2.03E-01 | - | - |
| Campesterol intake (mg/day) | Nutrients | 11348 | 0.01 | 0.01 | 1.09E-01 | 4.16E-01 | 64 | 11147 | -0.01 | 0.01 | 1.59E-01 | 4.84E-01 | 78 | 0.00 | 0.01 | 8.49E-01 | 0.78 | 3.37E-02 | - | - |
| Margarine on bread | Food | 11348 | 0.00 | 0.01 | 6.64E-01 | 8.52E-01 | 189 | 11147 | 0.01 | 0.01 | 4.72E-01 | 8.12E-01 | 141 | 0.00 | 0.01 | 8.56E-01 | 0.00 | 4.12E-01 | - | - |
| Frequency of shoverling snow during leisure time | Physical activity | 11745 | 0.00 | 0.01 | 7.28E-01 | 8.81E-01 | 201 | 11574 | 0.00 | 0.01 | 9.18E-01 | 9.96E-01 | 219 | 0.00 | 0.01 | 8.59E-01 | 0.00 | 7.53E-01 | - | - |
| If you exercise, change in exercise habits during the last year | Physical activity | 10759 | 0.00 | 0.01 | 7.55E-01 | 8.94E-01 | 205 | 10578 | 0.00 | 0.01 | 9.42E-01 | 9.96E-01 | 227 | 0.00 | 0.01 | 8.63E-01 | 0.00 | 7.87E-01 | - | - |
| Hearing status | General health | 6704 | 0.00 | 0.01 | 6.96E-01 | 8.67E-01 | 193 | 6596 | 0.00 | 0.01 | 8.65E-01 | 9.96E-01 | 206 | 0.00 | 0.01 | 8.69E-01 | 0.00 | 6.94E-01 | - | - |
| Salad dressing with oil | Food | 11348 | 0.00 | 0.01 | 9.30E-01 | 9.61E-01 | 235 | 11147 | 0.00 | 0.01 | 8.86E-01 | 9.96E-01 | 209 | 0.00 | 0.01 | 8.71E-01 | 0.00 | 9.66E-01 | - | - |
| Cohabitation: Live alone vs Only children | Social | 1678 | 0.03 | 0.04 | 5.19E-01 | 7.73E-01 | 162 | 1643 | -0.06 | 0.06 | 2.80E-01 | 6.66E-01 | 101 | -0.01 | 0.04 | 8.81E-01 | 0.36 | 2.12E-01 | - | - |
| Travel to work: Cycle to work vs passive travel to work | Physical activity | 9513 | -0.02 | 0.02 | 1.87E-01 | 5.17E-01 | 86 | 9415 | 0.02 | 0.02 | 2.63E-01 | 6.52E-01 | 97 | 0.00 | 0.01 | 8.87E-01 | 0.66 | 8.45E-02 | - | - |
| Do you feel important and appreciated outside your home? | Psychosocial | 6707 | 0.01 | 0.01 | 2.85E-01 | 6.30E-01 | 110 | 6595 | -0.01 | 0.01 | 3.57E-01 | 7.44E-01 | 116 | 0.00 | 0.01 | 8.94E-01 | 0.49 | 1.60E-01 | - | - |
| Boiled coffee | Beverage | 11348 | 0.01 | 0.01 | 3.30E-01 | 6.74E-01 | 119 | 11147 | -0.01 | 0.01 | 2.29E-01 | 6.05E-01 | 92 | 0.00 | 0.01 | 8.96E-01 | 0.58 | 1.23E-01 | - | - |
| Lean fish (e.g. perch, bass, cod) | Food | 11348 | 0.00 | 0.01 | 9.35E-01 | 9.62E-01 | 236 | 11147 | 0.00 | 0.01 | 8.05E-01 | 9.93E-01 | 197 | 0.00 | 0.01 | 9.14E-01 | 0.00 | 8.13E-01 | - | - |
| Sweets | Food | 11348 | 0.00 | 0.01 | 8.83E-01 | 9.41E-01 | 228 | 11147 | 0.00 | 0.01 | 9.96E-01 | 1.00E+00 | 242 | 0.00 | 0.01 | 9.18E-01 | 0.00 | 9.16E-01 | - | - |
| White cabbage, lettuce, lettuce cabbage, spinach, borecole | Food | 11348 | 0.00 | 0.01 | 9.24E-01 | 9.60E-01 | 234 | 11147 | 0.00 | 0.01 | 9.66E-01 | 9.96E-01 | 235 | 0.00 | 0.01 | 9.22E-01 | 0.00 | 9.72E-01 | - | - |
| Potassium intake (mg/day) | Nutrients | 11348 | 0.00 | 0.01 | 7.58E-01 | 8.94E-01 | 206 | 11147 | 0.00 | 0.01 | 6.54E-01 | 9.34E-01 | 168 | 0.00 | 0.01 | 9.33E-01 | 0.00 | 5.91E-01 | - | - |
| Boiled or baked potato | Food | 11348 | 0.00 | 0.01 | 8.43E-01 | 9.40E-01 | 216 | 11147 | 0.00 | 0.01 | 9.22E-01 | 9.96E-01 | 220 | 0.00 | 0.01 | 9.39E-01 | 0.00 | 8.36E-01 | - | - |
| Snuff status: Snuff users vs non-snuff users | Tobacco use | 10361 | -0.01 | 0.02 | 6.39E-01 | 8.42E-01 | 183 | 10202 | 0.01 | 0.02 | 6.61E-01 | 9.34E-01 | 172 | 0.00 | 0.02 | 9.65E-01 | 0.00 | 5.22E-01 | - | - |
| Number of snuff boxes per week | Tobacco use | 11621 | 0.00 | 0.01 | 8.94E-01 | 9.44E-01 | 230 | 11437 | 0.00 | 0.01 | 9.36E-01 | 9.96E-01 | 225 | 0.00 | 0.01 | 9.67E-01 | 0.00 | 8.81E-01 | - | - |
| Blota (broth + bread) | Food | 5080 | 0.00 | 0.01 | 8.64E-01 | 9.40E-01 | 220 | 4983 | 0.00 | 0.01 | 8.49E-01 | 9.96E-01 | 201 | 0.00 | 0.01 | 9.86E-01 | 0.00 | 7.99E-01 | - | - |
| Satisfaction with accomodation | Psychosocial | 6710 | -0.01 | 0.01 | 4.58E-01 | 7.58E-01 | 147 | 6593 | 0.01 | 0.01 | 4.48E-01 | 7.93E-01 | 136 | 0.00 | 0.01 | 9.91E-01 | 0.11 | 2.89E-01 | - | - |
| Confidence status | Psychosocial | 6696 | 0.00 | 0.01 | 7.47E-01 | 8.93E-01 | 203 | 6585 | 0.00 | 0.01 | 7.42E-01 | 9.67E-01 | 185 | 0.00 | 0.01 | 9.95E-01 | 0.00 | 6.45E-01 | - | - |
| Low fat milk (0,5%) | Beverage | 11348 | 0.00 | 0.01 | 9.38E-01 | 9.62E-01 | 237 | 11147 | 0.00 | 0.01 | 9.33E-01 | 9.96E-01 | 223 | 0.00 | 0.01 | 9.98E-01 | 0.00 | 9.09E-01 | - | - |

**Supplementary Table 20**. Longitudinal association results for 2h glucose

| | | Training set | | | | | | Testing set | | | | | | Metanalysis | | | | | R2 | |
|---|---|---|---|---|---|---|---|---|---|---|---|---|---|---|---|---|---|---|---|---|
| **Description** | **Group** | **N** | **Effect estimate** | **S.E.** | **p-value** | **p-value$_{FDR}$** | **p-value rank** | **N** | **Effect estimate** | **S.E.** | **p-value** | **p-value$_{FDR}$** | **p-value rank** | **Effect estimate** | **S.E.** | **p-value** | **$I^2$** | **Q p-value** | **adjusted R2** | **R2 rank** |
| Informed of having high blood pressure | General health | 11515 | 0.18 | 0.04 | 3.69E-06 | **4.49E-04** | 2 | 11276 | 0.23 | 0.04 | 1.23E-08 | **1.50E-06** | 2 | 0.20 | 0.03 | 2.88E-13 | 0.00 | 4.28E-01 | 0.18 | 2 |
| Parents or siblings have diabetes | General health | 11411 | 0.13 | 0.04 | 6.12E-04 | **2.97E-02** | 5 | 11191 | 0.22 | 0.04 | 4.28E-09 | **1.04E-06** | 1 | 0.17 | 0.03 | 4.77E-11 | 0.67 | 8.21E-02 | 0.18 | 3 |
| Average portion size of potatoes/rice/pasta based on photographic illustration of four sizes (smallest to largest) | Food | 10673 | -0.06 | 0.02 | 2.95E-05 | **1.79E-03** | 4 | 10406 | -0.08 | 0.02 | 3.29E-07 | **2.67E-05** | 3 | -0.07 | 0.01 | 5.19E-11 | 0.00 | 4.93E-01 | 0.18 | 7 |
| Exercise during the last three months | Physical activity | 11509 | -0.06 | 0.01 | 2.59E-06 | **4.49E-04** | 1 | 11277 | -0.05 | 0.01 | 7.13E-04 | **1.84E-02** | 9 | -0.06 | 0.01 | 9.89E-09 | 0.00 | 3.97E-01 | 0.18 | 4 |
| Cambridge physical activity index | Physical activity | 11333 | -0.06 | 0.01 | 1.50E-05 | **1.21E-03** | 3 | 11072 | -0.04 | 0.01 | 2.60E-03 | **4.52E-02** | 14 | -0.05 | 0.01 | 1.98E-07 | 0.00 | 3.78E-01 | 0.18 | 6 |
| Secoisolariciresinol intake (ug/day) | Nutrients | 10673 | -0.05 | 0.01 | 1.20E-03 | **3.63E-02** | 8 | 10406 | -0.05 | 0.01 | 6.14E-04 | **1.84E-02** | 8 | -0.05 | 0.01 | 2.44E-06 | 0.00 | 8.64E-01 | 0.19 | 1 |
| Educational level | Psychosocial | 11504 | -0.05 | 0.01 | 1.15E-03 | **3.63E-02** | 7 | 11277 | -0.04 | 0.01 | 3.24E-03 | **4.92E-02** | 16 | -0.04 | 0.01 | 1.16E-05 | 0.00 | 8.72E-01 | 0.18 | 5 |
| Grams of tobacco smoked per week | Tobacco use | 7206 | -0.07 | 0.03 | 1.46E-02 | 1.98E-01 | 15 | 6935 | -0.11 | 0.03 | 1.91E-04 | **7.75E-03** | 4 | -0.09 | 0.02 | 1.28E-05 | 0.00 | 3.57E-01 | - | - |
| Number of cigarretes smoked per day | Tobacco use | 7206 | -0.07 | 0.03 | 1.46E-02 | 1.98E-01 | 16 | 6935 | -0.11 | 0.03 | 1.91E-04 | **7.75E-03** | 5 | -0.09 | 0.02 | 1.28E-05 | 0.00 | 3.57E-01 | - | - |
| Number of cigars smoked per day | Tobacco use | 7206 | -0.07 | 0.03 | 1.46E-02 | 1.98E-01 | 17 | 6935 | -0.11 | 0.03 | 1.91E-04 | **7.75E-03** | 6 | -0.09 | 0.02 | 1.28E-05 | 0.00 | 3.57E-01 | - | - |
| Average portion size of vegetables based on photographic illustration of four sizes (smallest to largest) | Food | 10673 | -0.05 | 0.01 | 8.75E-04 | **3.54E-02** | 6 | 10406 | -0.04 | 0.02 | 5.82E-03 | 7.07E-02 | 20 | -0.05 | 0.01 | 1.65E-05 | 0.00 | 7.15E-01 | - | - |
| Average portion size of meat/fish based on photographic illustration of four sizes (smallest to largest) | Food | 10673 | -0.05 | 0.02 | 1.83E-03 | **4.95E-02** | 9 | 10406 | -0.05 | 0.02 | 3.07E-03 | **4.92E-02** | 15 | -0.05 | 0.01 | 1.72E-05 | 0.00 | 9.39E-01 | 0.18 | 8 |
| Teetotaler | Alcohol | 11488 | 0.09 | 0.05 | 4.02E-02 | 3.05E-01 | 32 | 11269 | 0.17 | 0.05 | 5.58E-04 | **1.84E-02** | 7 | 0.13 | 0.03 | 1.12E-04 | 0.17 | 2.71E-01 | - | - |
| Brewed (filtered) coffee | Beverage | 10673 | -0.04 | 0.01 | 3.54E-03 | 8.61E-02 | 10 | 10406 | -0.03 | 0.01 | 1.58E-02 | 1.39E-01 | 27 | -0.04 | 0.01 | 1.62E-04 | 0.00 | 7.41E-01 | - | - |
| Years using snuff | Tobacco use | 10591 | -0.03 | 0.01 | 5.30E-02 | 3.90E-01 | 33 | 10358 | -0.05 | 0.02 | 1.13E-03 | **2.49E-02** | 11 | -0.04 | 0.01 | 2.48E-04 | 0.00 | 3.33E-01 | - | - |
| High mental demand from job | Psychosocial | 11308 | 0.03 | 0.01 | 3.66E-02 | 2.87E-01 | 31 | 11048 | 0.04 | 0.01 | 8.68E-03 | 9.98E-02 | 21 | 0.03 | 0.01 | 8.62E-04 | 0.00 | 6.94E-01 | - | - |
| Pancake, waffle, Swedish dumpling | Food | 10673 | -0.03 | 0.01 | 6.21E-02 | 4.08E-01 | 35 | 10406 | -0.04 | 0.01 | 5.24E-03 | 6.70E-02 | 19 | -0.03 | 0.01 | 1.01E-03 | 0.00 | 4.92E-01 | - | - |
| Snuff status: Snuff users vs non-snuff users | Tobacco use | 9678 | -0.08 | 0.04 | 3.63E-02 | 2.87E-01 | 30 | 9511 | -0.10 | 0.04 | 1.10E-02 | 1.11E-01 | 24 | -0.09 | 0.03 | 1.05E-03 | 0.00 | 7.33E-01 | - | - |
| Total energy intake (kcal/day) | Nutrients | 10673 | -0.02 | 0.02 | 2.18E-01 | 7.01E-01 | 74 | 10406 | -0.05 | 0.02 | 2.22E-03 | **4.14E-02** | 13 | -0.03 | 0.01 | 2.55E-03 | 0.44 | 1.82E-01 | - | - |
| Hamburger | Food | 10673 | 0.03 | 0.01 | 2.26E-02 | 2.75E-01 | 20 | 10406 | 0.03 | 0.01 | 5.16E-02 | 2.95E-01 | 42 | 0.03 | 0.01 | 2.78E-03 | 0.00 | 8.32E-01 | - | - |
| Meat on bread | Food | 10673 | 0.03 | 0.01 | 2.64E-02 | 2.87E-01 | 22 | 10406 | 0.03 | 0.01 | 5.28E-02 | 2.95E-01 | 43 | 0.03 | 0.01 | 3.25E-03 | 0.00 | 8.92E-01 | - | - |
| Marital status: Single vs Married/partner | Social | 10683 | -0.06 | 0.05 | 1.66E-01 | 6.04E-01 | 67 | 10441 | -0.13 | 0.05 | 5.23E-03 | 6.70E-02 | 18 | -0.10 | 0.03 | 3.25E-03 | 0.05 | 3.04E-01 | - | - |
| Breakfast habits: Only coffee/tea for breakfast vs not breakfast at all | Food | 723 | -0.34 | 0.20 | 9.14E-02 | 4.40E-01 | 49 | 690 | -0.45 | 0.18 | 1.50E-02 | 1.39E-01 | 26 | -0.40 | 0.13 | 3.28E-03 | 0.00 | 6.87E-01 | - | - |
| Overall state of health compared to other your age | General health | 11329 | -0.02 | 0.01 | 8.74E-02 | 4.40E-01 | 47 | 11124 | -0.03 | 0.01 | 2.66E-02 | 1.96E-01 | 33 | -0.03 | 0.01 | 5.55E-03 | 0.00 | 7.00E-01 | - | - |
| Snuff status: Former snuff users vs non-snuff users | Tobacco use | 8865 | -0.04 | 0.05 | 3.46E-01 | 7.61E-01 | 106 | 8719 | -0.14 | 0.05 | 3.57E-03 | 5.10E-02 | 17 | -0.09 | 0.03 | 6.78E-03 | 0.51 | 1.52E-01 | - | - |
| Cohabitation: Live alone vs Adult and children | Social | 8314 | -0.10 | 0.05 | 3.56E-02 | 2.87E-01 | 29 | 8017 | -0.09 | 0.05 | 8.90E-02 | 3.91E-01 | 55 | -0.09 | 0.04 | 7.11E-03 | 0.00 | 8.01E-01 | - | - |
| Cohabitation: Live alone vs Only children | Social | 1558 | -0.13 | 0.08 | 1.07E-01 | 4.78E-01 | 54 | 1524 | -0.18 | 0.09 | 3.31E-02 | 2.24E-01 | 36 | -0.16 | 0.06 | 8.33E-03 | 0.00 | 6.68E-01 | - | - |
| Skill demand from job | Psychosocial | 11355 | -0.03 | 0.01 | 3.02E-02 | 2.87E-01 | 24 | 11113 | -0.02 | 0.01 | 1.38E-01 | 4.96E-01 | 66 | -0.03 | 0.01 | 9.71E-03 | 0.00 | 6.44E-01 | - | - |
| Distance to work in kilometers (one way) | Physical activity | 10151 | 0.03 | 0.01 | 1.46E-02 | 1.98E-01 | 18 | 10009 | 0.02 | 0.01 | 2.37E-01 | 6.66E-01 | 86 | 0.03 | 0.01 | 1.01E-02 | 0.00 | 3.90E-01 | - | - |
| Lean fish (e.g. perch, bass, cod) | Food | 10673 | -0.03 | 0.01 | 1.70E-02 | 2.18E-01 | 19 | 10406 | -0.02 | 0.01 | 2.19E-01 | 6.47E-01 | 82 | -0.03 | 0.01 | 1.01E-02 | 0.00 | 4.44E-01 | - | - |
| Parents or siblings had a cerebral hemorrhage/thrombosis or cardiac infarction before the age of 60 | General health | 11388 | 0.09 | 0.04 | 1.00E-02 | 1.74E-01 | 14 | 11174 | 0.04 | 0.04 | 3.16E-01 | 7.55E-01 | 101 | 0.06 | 0.02 | 1.11E-02 | 0.16 | 2.76E-01 | - | - |
| Smoking status: Former smokers vs non-smokers | Tobacco use | 7962 | -0.03 | 0.04 | 3.60E-01 | 7.61E-01 | 112 | 7775 | -0.09 | 0.04 | 9.04E-03 | 9.98E-02 | 22 | -0.06 | 0.03 | 1.26E-02 | 0.30 | 2.31E-01 | - | - |
| Work shifts/weekends | Psychosocial | 11162 | -0.03 | 0.03 | 2.70E-01 | 7.27E-01 | 88 | 10936 | -0.07 | 0.03 | 1.77E-02 | 1.48E-01 | 29 | -0.05 | 0.02 | 1.41E-02 | 0.00 | 3.66E-01 | - | - |
| Mood status | Psychosocial | 6333 | 0.02 | 0.02 | 2.56E-01 | 7.27E-01 | 85 | 6179 | 0.04 | 0.02 | 2.12E-02 | 1.67E-01 | 30 | 0.03 | 0.01 | 1.51E-02 | 0.00 | 4.02E-01 | - | - |
| Travel to work: Cycle to work vs passive travel to work | Physical activity | 8938 | -0.09 | 0.04 | 7.55E-03 | 1.41E-01 | 13 | 8804 | -0.03 | 0.04 | 4.55E-01 | 8.10E-01 | 134 | -0.06 | 0.03 | 1.54E-02 | 0.45 | 1.77E-01 | - | - |
| Cookies, pastry | Food | 10673 | -0.02 | 0.01 | 9.18E-02 | 4.40E-01 | 50 | 10406 | -0.02 | 0.01 | 8.64E-02 | 3.91E-01 | 52 | -0.02 | 0.01 | 1.62E-02 | 0.00 | 9.73E-01 | - | - |
| Sodas, soft drinks, juice | Beverage | 10673 | 0.03 | 0.01 | 6.38E-02 | 4.08E-01 | 36 | 10406 | 0.02 | 0.02 | 1.25E-01 | 4.75E-01 | 64 | 0.03 | 0.01 | 1.64E-02 | 0.00 | 8.58E-01 | - | - |
| Would you say that the number of people that you meet in your everyday life is enough or would you like to meet more or fewer people? | Social | 11488 | -0.02 | 0.01 | 1.37E-01 | 5.56E-01 | 60 | 11243 | -0.03 | 0.01 | 5.72E-02 | 3.09E-01 | 45 | -0.02 | 0.01 | 1.67E-02 | 0.00 | 7.49E-01 | - | - |
| Possibility to leave your work for a while to speak with a colleague | Psychosocial | 11320 | 0.02 | 0.01 | 1.78E-01 | 6.18E-01 | 70 | 11034 | 0.03 | 0.01 | 4.25E-02 | 2.71E-01 | 38 | 0.02 | 0.01 | 1.73E-02 | 0.00 | 6.07E-01 | - | - |
| Frequency of cycling during leisure time | Physical activity | 9898 | 0.00 | 0.01 | 8.19E-01 | 9.59E-01 | 204 | 9666 | -0.05 | 0.01 | 1.68E-03 | **3.41E-02** | 12 | -0.02 | 0.01 | 1.79E-02 | 0.77 | 3.77E-02 | - | - |
| Light beer | Alcohol | 10673 | -0.01 | 0.02 | 3.57E-01 | 7.61E-01 | 111 | 10406 | -0.03 | 0.01 | 2.47E-02 | 1.88E-01 | 32 | -0.02 | 0.01 | 2.43E-02 | 0.00 | 3.65E-01 | - | - |
| Fitness status | Physical activity | 6330 | -0.04 | 0.02 | 3.01E-02 | 2.87E-01 | 23 | 6174 | -0.02 | 0.02 | 3.09E-01 | 7.55E-01 | 99 | -0.03 | 0.01 | 2.43E-02 | 0.00 | 4.14E-01 | - | - |
| Apple, pear, peach, orange, mandarin and grapefruit | Food | 10673 | -0.03 | 0.02 | 6.38E-02 | 4.08E-01 | 37 | 10406 | -0.02 | 0.02 | 1.94E-01 | 6.13E-01 | 77 | -0.02 | 0.01 | 2.56E-02 | 0.00 | 7.11E-01 | - | - |
| Smoking status: Former occasional smokers vs non-smokers | Tobacco use | 6706 | -0.08 | 0.05 | 9.06E-02 | 4.40E-01 | 48 | 6476 | -0.07 | 0.05 | 1.47E-01 | 4.96E-01 | 72 | -0.08 | 0.03 | 2.62E-02 | 0.00 | 8.73E-01 | - | - |
| Cheese 28% | Food | 10673 | -0.03 | 0.01 | 3.22E-02 | 2.87E-01 | 27 | 10406 | -0.01 | 0.01 | 3.46E-01 | 7.56E-01 | 110 | -0.02 | 0.01 | 2.86E-02 | 0.00 | 4.08E-01 | - | - |
| Repetitive job | Psychosocial | 11379 | 0.03 | 0.01 | 3.14E-02 | 2.87E-01 | 26 | 11127 | 0.01 | 0.01 | 3.73E-01 | 7.69E-01 | 118 | 0.02 | 0.01 | 3.07E-02 | 0.00 | 3.85E-01 | - | - |
| Fiber cereals | Food | 10673 | -0.03 | 0.01 | 6.74E-02 | 4.08E-01 | 38 | 10406 | -0.02 | 0.01 | 2.67E-01 | 6.94E-01 | 93 | -0.02 | 0.01 | 3.74E-02 | 0.00 | 6.20E-01 | - | - |
| Satisfaction with accomodation | Psychosocial | 6343 | 0.03 | 0.02 | 1.65E-01 | 6.04E-01 | 66 | 6185 | 0.03 | 0.02 | 1.24E-01 | 4.75E-01 | 63 | 0.03 | 0.01 | 3.83E-02 | 0.00 | 9.21E-01 | - | - |
| Butter for cooking | Food | 10673 | 0.01 | 0.01 | 7.06E-01 | 9.53E-01 | 179 | 10406 | 0.04 | 0.01 | 1.23E-02 | 1.20E-01 | 25 | 0.02 | 0.01 | 4.16E-02 | 0.56 | 1.33E-01 | - | - |
| Banana | Food | 10673 | 0.02 | 0.01 | 2.12E-01 | 7.01E-01 | 73 | 10406 | 0.02 | 0.01 | 1.06E-01 | 4.43E-01 | 58 | 0.02 | 0.01 | 4.29E-02 | 0.00 | 7.84E-01 | - | - |
| Participation in associations or voluntary organisations | Social | 11477 | -0.02 | 0.03 | 4.71E-01 | 8.62E-01 | 132 | 11238 | -0.06 | 0.03 | 3.29E-02 | 2.24E-01 | 35 | -0.04 | 0.02 | 4.49E-02 | 0.05 | 3.06E-01 | - | - |
| Ingenuity or creativity demand from job | Psychosocial | 11334 | -0.01 | 0.01 | 5.82E-01 | 9.21E-01 | 151 | 11089 | -0.03 | 0.01 | 2.13E-02 | 1.67E-01 | 31 | -0.02 | 0.01 | 4.53E-02 | 0.37 | 2.06E-01 | - | - |
| Everyday exercise satisfaction | Physical activity | 11479 | 0.02 | 0.01 | 1.01E-01 | 4.62E-01 | 53 | 11251 | 0.02 | 0.01 | 2.37E-01 | 6.66E-01 | 85 | 0.02 | 0.01 | 4.53E-02 | 0.00 | 7.68E-01 | - | - |
| Years smoking | Tobacco use | 10177 | -0.02 | 0.02 | 2.72E-01 | 7.27E-01 | 90 | 9956 | -0.03 | 0.02 | 8.37E-02 | 3.91E-01 | 51 | -0.02 | 0.01 | 4.59E-02 | 0.00 | 6.43E-01 | - | - |
| Root vegetables, carrot | Food | 10673 | -0.01 | 0.01 | 4.31E-01 | 8.26E-01 | 126 | 10406 | -0.03 | 0.01 | 4.46E-02 | 2.71E-01 | 40 | -0.02 | 0.01 | 4.80E-02 | 0.00 | 3.88E-01 | - | - |
| Beta-carotene intake (mg/day) | Nutrients | 10673 | -0.01 | 0.01 | 3.63E-01 | 7.61E-01 | 115 | 10406 | -0.03 | 0.01 | 6.07E-02 | 3.21E-01 | 46 | -0.02 | 0.01 | 4.89E-02 | 0.00 | 4.94E-01 | - | - |
| Disaccharides intake (g/day) | Nutrients | 10673 | 0.03 | 0.01 | 3.06E-02 | 2.87E-01 | 25 | 10406 | 0.01 | 0.01 | 5.54E-01 | 8.54E-01 | 157 | 0.02 | 0.01 | 5.02E-02 | 0.16 | 2.75E-01 | - | - |
| Vitamin A intake (mg/day) | Nutrients | 10673 | 0.04 | 0.02 | 3.51E-02 | 2.87E-01 | 28 | 10406 | 0.01 | 0.02 | 5.27E-01 | 8.54E-01 | 150 | 0.02 | 0.01 | 5.16E-02 | 0.05 | 3.05E-01 | - | - |
| Long-term sickness | General health | 11148 | 0.08 | 0.04 | 7.15E-02 | 4.08E-01 | 41 | 10858 | 0.04 | 0.04 | 3.52E-01 | 7.56E-01 | 113 | 0.06 | 0.03 | 5.17E-02 | 0.00 | 5.64E-01 | - | - |
| Overall state of health during the last year | General health | 11502 | -0.01 | 0.01 | 3.23E-01 | 7.61E-01 | 100 | 11265 | -0.02 | 0.01 | 7.68E-02 | 3.73E-01 | 50 | -0.02 | 0.01 | 5.19E-02 | 0.00 | 5.66E-01 | - | - |
| Marital status: Single vs Divorced/separated | Social | 1851 | -0.01 | 0.07 | 8.57E-01 | 9.59E-01 | 217 | 1803 | -0.20 | 0.08 | 1.00E-02 | 1.06E-01 | 23 | -0.10 | 0.05 | 5.46E-02 | 0.67 | 8.40E-02 | - | - |
| Low fat margarine on bread | Food | 10673 | 0.01 | 0.01 | 4.95E-01 | 8.78E-01 | 136 | 10406 | 0.03 | 0.01 | 4.93E-02 | 2.92E-01 | 41 | 0.02 | 0.01 | 6.23E-02 | 0.00 | 3.55E-01 | - | - |
| Lariciresinol intake (ug/day) | Nutrients | 10673 | -0.01 | 0.01 | 3.56E-01 | 7.61E-01 | 110 | 10406 | -0.03 | 0.01 | 8.68E-02 | 3.91E-01 | 53 | -0.02 | 0.01 | 6.33E-02 | 0.00 | 5.63E-01 | - | - |
| Number of social interactions during a normal week | Social | 11484 | 0.02 | 0.01 | 1.08E-01 | 4.78E-01 | 55 | 11269 | 0.01 | 0.01 | 3.17E-01 | 7.55E-01 | 102 | 0.02 | 0.01 | 6.48E-02 | 0.00 | 6.78E-01 | - | - |
| Patience status | Psychosocial | 6334 | 0.03 | 0.02 | 7.20E-02 | 4.08E-01 | 42 | 6177 | 0.02 | 0.02 | 4.23E-01 | 7.91E-01 | 130 | 0.03 | 0.01 | 6.60E-02 | 0.00 | 4.80E-01 | - | - |
| Eat breakfast from 2000 | Food | 1133 | -0.15 | 0.15 | 3.18E-01 | 7.61E-01 | 99 | 1014 | -0.24 | 0.15 | 1.12E-01 | 4.45E-01 | 61 | -0.20 | 0.11 | 6.63E-02 | 0.00 | 6.89E-01 | - | - |
| Soft cheese | Food | 4726 | 0.03 | 0.02 | 9.28E-02 | 4.40E-01 | 51 | 4623 | 0.02 | 0.02 | 3.96E-01 | 7.89E-01 | 122 | 0.03 | 0.01 | 7.44E-02 | 0.00 | 5.47E-01 | - | - |
| Cholesterol intake (g/day) | Nutrients | 10673 | -0.03 | 0.02 | 6.99E-02 | 4.08E-01 | 40 | 10406 | -0.01 | 0.02 | 4.88E-01 | 8.23E-01 | 144 | -0.02 | 0.01 | 7.47E-02 | 0.00 | 4.42E-01 | - | - |
| Sleep status | Sleep | 6345 | 0.04 | 0.02 | 2.46E-02 | 2.85E-01 | 21 | 6188 | 0.00 | 0.02 | 8.03E-01 | 9.61E-01 | 203 | 0.02 | 0.01 | 7.74E-02 | 0.50 | 1.58E-01 | - | - |
| Number of snuff boxes per week | Tobacco use | 10872 | -0.01 | 0.01 | 4.89E-01 | 8.78E-01 | 134 | 10673 | -0.03 | 0.01 | 7.42E-02 | 3.68E-01 | 49 | -0.02 | 0.01 | 8.08E-02 | 0.00 | 4.31E-01 | - | - |
| Whole grain intake (g/day) | Food | 10673 | -0.02 | 0.01 | 2.45E-01 | 7.25E-01 | 80 | 10406 | -0.02 | 0.01 | 1.97E-01 | 6.14E-01 | 78 | -0.02 | 0.01 | 8.30E-02 | 0.00 | 9.18E-01 | - | - |
| Vitamin B12 intake (ug/day) | Nutrients | 10673 | 0.02 | 0.01 | 1.10E-01 | 4.79E-01 | 56 | 10406 | 0.01 | 0.01 | 4.03E-01 | 7.91E-01 | 124 | 0.02 | 0.01 | 8.33E-02 | 0.00 | 6.18E-01 | - | - |
| Contradictory demands in job | Psychosocial | 11309 | -0.01 | 0.01 | 3.63E-01 | 7.61E-01 | 114 | 11049 | -0.02 | 0.01 | 1.44E-01 | 4.96E-01 | 70 | -0.02 | 0.01 | 9.41E-02 | 0.00 | 6.86E-01 | - | - |
| Soft whey cheese | Food | 4726 | -0.02 | 0.02 | 2.24E-01 | 7.01E-01 | 77 | 4623 | -0.02 | 0.02 | 2.50E-01 | 6.76E-01 | 90 | -0.02 | 0.01 | 9.44E-02 | 0.00 | 9.73E-01 | - | - |
| Vitamin B3 intake (mg/day) | Nutrients | 10673 | -0.01 | 0.01 | 3.27E-01 | 7.61E-01 | 101 | 10406 | -0.02 | 0.01 | 1.83E-01 | 5.85E-01 | 76 | -0.02 | 0.01 | 1.03E-01 | 0.00 | 7.94E-01 | - | - |
| Satisfaction with home and family situation | Psychosocial | 6339 | 0.03 | 0.02 | 1.18E-01 | 4.95E-01 | 57 | 6181 | 0.01 | 0.02 | 4.63E-01 | 8.10E-01 | 138 | 0.02 | 0.01 | 1.05E-01 | 0.00 | 5.54E-01 | - | - |
| Strong beer | Alcohol | 10673 | -0.01 | 0.02 | 3.86E-01 | 7.94E-01 | 118 | 10406 | -0.02 | 0.02 | 1.61E-01 | 5.29E-01 | 73 | -0.02 | 0.01 | 1.10E-01 | 0.00 | 6.84E-01 | - | - |
| Satisfaction with leisure time | Psychosocial | 6337 | -0.02 | 0.02 | 2.65E-01 | 7.27E-01 | 86 | 6167 | 0.06 | 0.02 | 7.58E-04 | **1.84E-02** | 10 | 0.02 | 0.01 | 1.11E-01 | 0.90 | 1.52E-03 | - | - |
| White meat (poultry) | Food | 10673 | -0.01 | 0.01 | 5.92E-01 | 9.21E-01 | 156 | 10406 | -0.02 | 0.01 | 9.02E-02 | 3.91E-01 | 56 | -0.02 | 0.01 | 1.12E-01 | 0.00 | 4.28E-01 | - | - |
| Sucrose intake (g/day) | Nutrients | 10673 | 0.02 | 0.01 | 1.35E-01 | 5.55E-01 | 59 | 10406 | 0.01 | 0.01 | 4.69E-01 | 8.10E-01 | 140 | 0.02 | 0.01 | 1.15E-01 | 0.00 | 5.97E-01 | - | - |
| Fried potatoes, pommes frites | Food | 10673 | 0.02 | 0.01 | 8.23E-02 | 4.39E-01 | 45 | 10406 | 0.01 | 0.01 | 6.33E-01 | 8.82E-01 | 172 | 0.02 | 0.01 | 1.16E-01 | 0.00 | 3.78E-01 | - | - |
| Sum of Lariciresinol, Matairesinol, Pinoresinol, Secoisolariciresinol intake (ug/day) | Nutrients | 10673 | -0.01 | 0.01 | 4.58E-01 | 8.51E-01 | 130 | 10406 | -0.02 | 0.01 | 1.41E-01 | 4.96E-01 | 67 | -0.02 | 0.01 | 1.18E-01 | 0.00 | 5.94E-01 | - | - |

| | | | | | | | | | | | | | | | | | | | | |
|---|---|---|---|---|---|---|---|---|---|---|---|---|---|---|---|---|---|---|---|---|
| Liver, kidney | Food | 4726 | 0.02 | 0.02 | 2.24E-01 | 7.01E-01 | 76 | 4623 | 0.02 | 0.02 | 3.25E-01 | 7.56E-01 | 104 | 0.02 | 0.01 | 1.18E-01 | 0.00 | 9.21E-01 | - | - |
| Berries (fresh or frozen) | Food | 10673 | -0.01 | 0.01 | 4.96E-01 | 8.78E-01 | 137 | 10406 | -0.02 | 0.01 | 1.42E-01 | 4.96E-01 | 69 | -0.01 | 0.01 | 1.29E-01 | 0.00 | 5.71E-01 | - | - |
| Frequency of shoverling snow during leisure time | Physical activity | 11010 | 0.00 | 0.01 | 8.80E-01 | 9.72E-01 | 220 | 10817 | -0.03 | 0.01 | 4.41E-02 | 2.71E-01 | 39 | -0.02 | 0.01 | 1.29E-01 | 0.44 | 1.83E-01 | - | - |
| Support from others | Social | 11489 | -0.04 | 0.01 | 5.91E-03 | 1.20E-01 | 12 | 11261 | 0.01 | 0.01 | 5.11E-01 | 8.40E-01 | 147 | -0.01 | 0.01 | 1.30E-01 | 0.83 | 1.68E-02 | - | - |
| Boiled coffee | Beverage | 10673 | -0.02 | 0.02 | 3.12E-01 | 7.61E-01 | 98 | 10406 | -0.02 | 0.02 | 2.59E-01 | 6.85E-01 | 92 | -0.02 | 0.01 | 1.31E-01 | 0.00 | 9.28E-01 | - | - |
| Satisfaction with economy | Psychosocial | 6338 | 0.03 | 0.02 | 1.18E-01 | 4.95E-01 | 58 | 6176 | 0.01 | 0.02 | 5.73E-01 | 8.64E-01 | 161 | 0.02 | 0.01 | 1.33E-01 | 0.00 | 4.80E-01 | - | - |
| Close relationship with anyone | Social | 11494 | -0.01 | 0.01 | 4.18E-01 | 8.20E-01 | 124 | 11267 | -0.02 | 0.01 | 2.19E-01 | 6.47E-01 | 81 | -0.01 | 0.01 | 1.50E-01 | 0.00 | 7.57E-01 | - | - |
| Light but partly physically active work | Physical activity | 11349 | 0.00 | 0.04 | 9.10E-01 | 9.77E-01 | 225 | 11084 | 0.07 | 0.04 | 5.34E-02 | 2.95E-01 | 44 | 0.04 | 0.03 | 1.52E-01 | 0.41 | 1.93E-01 | - | - |
| Frequency of walking during leisure time | Physical activity | 11193 | 0.02 | 0.01 | 1.99E-01 | 6.82E-01 | 71 | 11012 | 0.01 | 0.01 | 4.62E-01 | 8.10E-01 | 136 | 0.01 | 0.01 | 1.53E-01 | 0.00 | 7.04E-01 | - | - |
| Sweets | Food | 10673 | 0.03 | 0.01 | 7.25E-02 | 4.08E-01 | 43 | 10406 | 0.00 | 0.01 | 8.31E-01 | 9.71E-01 | 208 | 0.01 | 0.01 | 1.53E-01 | 0.19 | 2.68E-01 | - | - |
| Sour milk, yoghurt (low fat) | Food | 10673 | 0.01 | 0.01 | 4.59E-01 | 8.51E-01 | 131 | 10406 | 0.02 | 0.01 | 2.11E-01 | 6.45E-01 | 79 | 0.01 | 0.01 | 1.61E-01 | 0.00 | 7.04E-01 | - | - |
| Milk, sour milk (1,5%) | Beverage | 10673 | 0.02 | 0.01 | 1.55E-01 | 5.98E-01 | 63 | 10406 | 0.01 | 0.01 | 5.98E-01 | 8.79E-01 | 165 | 0.01 | 0.01 | 1.67E-01 | 0.00 | 5.33E-01 | - | - |
| Memory status | Psychosocial | 6321 | 0.03 | 0.02 | 6.98E-02 | 4.08E-01 | 39 | 6170 | 0.00 | 0.02 | 8.92E-01 | 9.79E-01 | 220 | 0.02 | 0.01 | 1.68E-01 | 0.29 | 2.36E-01 | - | - |
| High physical demand from job | Physical activity | 11404 | -0.02 | 0.01 | 2.47E-01 | 7.25E-01 | 82 | 11152 | -0.01 | 0.01 | 4.52E-01 | 8.10E-01 | 133 | -0.01 | 0.01 | 1.76E-01 | 0.00 | 7.86E-01 | - | - |
| Possibility to speak with colleagues during breaks | Psychosocial | 11365 | 0.01 | 0.01 | 3.48E-01 | 7.61E-01 | 108 | 11095 | 0.01 | 0.01 | 3.41E-01 | 7.56E-01 | 108 | 0.01 | 0.01 | 1.82E-01 | 0.00 | 9.83E-01 | - | - |
| Oatflake, whole wheat, rye or barley porridge | Food | 10673 | -0.02 | 0.01 | 2.25E-01 | 7.01E-01 | 78 | 10406 | -0.01 | 0.01 | 5.31E-01 | 8.54E-01 | 151 | -0.01 | 0.01 | 1.91E-01 | 0.00 | 6.92E-01 | - | - |
| Sausage, liver pate on bread | Food | 10673 | -0.01 | 0.01 | 7.08E-01 | 9.53E-01 | 180 | 10406 | 0.03 | 0.01 | 2.75E-02 | 1.96E-01 | 34 | 0.01 | 0.01 | 2.02E-01 | 0.70 | 6.62E-02 | - | - |
| Receive hugs to comfort and support you | Social | 11439 | -0.04 | 0.04 | 2.97E-01 | 7.61E-01 | 93 | 11203 | -0.03 | 0.04 | 4.58E-01 | 8.10E-01 | 135 | -0.03 | 0.03 | 2.06E-01 | 0.00 | 8.48E-01 | - | - |
| Participation in other association | Social | 724 | 0.02 | 0.11 | 8.45E-01 | 9.59E-01 | 212 | 643 | -0.24 | 0.12 | 4.07E-02 | 2.68E-01 | 37 | -0.10 | 0.08 | 2.06E-01 | 0.62 | 1.04E-01 | - | - |
| White cabbage, lettuce, lettuce cabbage, spinach, borecole | Food | 10673 | -0.02 | 0.01 | 2.12E-01 | 7.01E-01 | 72 | 10406 | -0.01 | 0.01 | 6.00E-01 | 8.79E-01 | 166 | -0.01 | 0.01 | 2.09E-01 | 0.00 | 6.14E-01 | - | - |
| Cohabitation: Live alone vs Other/others | Social | 1164 | 0.00 | 0.12 | 9.80E-01 | 9.88E-01 | 241 | 1134 | 0.23 | 0.13 | 6.88E-02 | 3.56E-01 | 47 | 0.11 | 0.09 | 2.12E-01 | 0.43 | 1.85E-01 | - | - |
| Magnesium intake (mg/day) | Nutrients | 10673 | -0.01 | 0.02 | 5.83E-01 | 9.21E-01 | 153 | 10406 | -0.02 | 0.02 | 2.38E-01 | 6.66E-01 | 87 | -0.01 | 0.01 | 2.23E-01 | 0.00 | 6.50E-01 | - | - |
| Steak, chop, e.g. | Food | 10673 | -0.04 | 0.01 | 5.06E-03 | 1.12E-01 | 11 | 10406 | 0.02 | 0.02 | 2.49E-01 | 6.76E-01 | 89 | -0.01 | 0.01 | 2.23E-01 | 0.87 | 5.50E-03 | - | - |
| Milk, sour milk (3%) | Beverage | 10673 | 0.02 | 0.01 | 1.58E-01 | 5.99E-01 | 64 | 10406 | 0.00 | 0.01 | 7.77E-01 | 9.54E-01 | 198 | 0.01 | 0.01 | 2.25E-01 | 0.00 | 4.39E-01 | - | - |
| Travel to work: Irregular travel mode to work vs passive travel to work | Physical activity | 7573 | -0.09 | 0.05 | 7.38E-02 | 4.08E-01 | 44 | 7440 | 0.00 | 0.05 | 9.28E-01 | 9.79E-01 | 226 | -0.04 | 0.04 | 2.31E-01 | 0.43 | 1.84E-01 | - | - |
| Tea | Beverage | 10673 | 0.01 | 0.01 | 3.50E-01 | 7.61E-01 | 109 | 10406 | 0.01 | 0.01 | 4.62E-01 | 8.10E-01 | 137 | 0.01 | 0.01 | 2.37E-01 | 0.00 | 8.99E-01 | - | - |
| Number of cigarretes smoked per day (in groups) | Tobacco use | 9681 | 0.00 | 0.01 | 8.32E-01 | 9.59E-01 | 207 | 9435 | 0.02 | 0.01 | 1.46E-01 | 4.96E-01 | 71 | 0.01 | 0.01 | 2.40E-01 | 0.00 | 3.78E-01 | - | - |
| Bacon | Food | 10673 | 0.01 | 0.01 | 5.83E-01 | 9.21E-01 | 152 | 10406 | 0.02 | 0.01 | 2.69E-01 | 6.94E-01 | 94 | 0.01 | 0.01 | 2.45E-01 | 0.00 | 6.79E-01 | - | - |
| Beta-sitosterol intake (mg/day) | Nutrients | 10673 | 0.00 | 0.01 | 8.14E-01 | 9.59E-01 | 203 | 10406 | -0.02 | 0.01 | 1.63E-01 | 5.29E-01 | 75 | -0.01 | 0.01 | 2.51E-01 | 0.00 | 4.08E-01 | - | - |
| Vision status | General health | 6333 | -0.02 | 0.02 | 3.61E-01 | 7.61E-01 | 113 | 6181 | -0.01 | 0.02 | 4.81E-01 | 8.17E-01 | 143 | -0.02 | 0.01 | 2.52E-01 | 0.00 | 8.87E-01 | - | - |
| Participation in study circles | Social | 724 | -0.03 | 0.17 | 8.40E-01 | 9.59E-01 | 211 | 643 | 0.30 | 0.17 | 7.25E-02 | 3.67E-01 | 48 | 0.13 | 0.12 | 2.56E-01 | 0.50 | 1.59E-01 | - | - |
| Frequency of hunting or fishing during leisure time | Physical activity | 10711 | -0.01 | 0.02 | 6.16E-01 | 9.26E-01 | 160 | 10515 | -0.02 | 0.02 | 2.73E-01 | 6.99E-01 | 95 | -0.01 | 0.01 | 2.62E-01 | 0.00 | 6.61E-01 | - | - |
| Vitamin B2 intake (ug/day) | Nutrients | 10673 | 0.02 | 0.01 | 2.66E-01 | 7.27E-01 | 87 | 10406 | 0.01 | 0.01 | 6.47E-01 | 8.82E-01 | 178 | 0.01 | 0.01 | 2.65E-01 | 0.00 | 6.50E-01 | - | - |
| Frequency of picking berries or mushrooms during leisure time | Physical activity | 10874 | 0.01 | 0.01 | 5.57E-01 | 9.20E-01 | 147 | 10660 | 0.01 | 0.02 | 3.21E-01 | 7.56E-01 | 103 | 0.01 | 0.01 | 2.65E-01 | 0.00 | 7.66E-01 | - | - |
| Control over own work assignment | Psychosocial | 11386 | 0.01 | 0.01 | 6.94E-01 | 9.53E-01 | 176 | 11129 | 0.02 | 0.01 | 2.44E-01 | 6.73E-01 | 88 | 0.01 | 0.01 | 2.73E-01 | 0.00 | 5.76E-01 | - | - |
| Enough time for job assignments | Psychosocial | 11338 | 0.00 | 0.01 | 7.38E-01 | 9.56E-01 | 187 | 11084 | 0.02 | 0.01 | 2.21E-01 | 6.47E-01 | 83 | 0.01 | 0.01 | 2.73E-01 | 0.00 | 5.23E-01 | - | - |
| Mashed potato | Food | 4726 | 0.02 | 0.02 | 2.75E-01 | 7.27E-01 | 92 | 4623 | 0.01 | 0.02 | 7.04E-01 | 9.01E-01 | 190 | 0.01 | 0.01 | 2.89E-01 | 0.00 | 6.48E-01 | - | - |
| Blota (broth + bread) | Food | 4726 | 0.01 | 0.02 | 4.46E-01 | 8.41E-01 | 129 | 4623 | 0.02 | 0.02 | 4.71E-01 | 8.10E-01 | 141 | 0.02 | 0.01 | 2.98E-01 | 0.00 | 9.05E-01 | - | - |
| Do you feel important and appreciated outside your home? | Psychosocial | 6342 | 0.03 | 0.02 | 1.70E-01 | 6.08E-01 | 68 | 6187 | 0.00 | 0.02 | 9.17E-01 | 9.79E-01 | 225 | 0.01 | 0.01 | 2.98E-01 | 0.00 | 3.69E-01 | - | - |
| Whole grain crisp bread | Food | 10673 | 0.01 | 0.01 | 5.23E-01 | 8.99E-01 | 141 | 10406 | 0.01 | 0.01 | 4.20E-01 | 7.91E-01 | 128 | 0.01 | 0.01 | 3.07E-01 | 0.00 | 8.98E-01 | - | - |
| Self-employed | Psychosocial | 11291 | 0.04 | 0.05 | 3.47E-01 | 7.61E-01 | 107 | 11058 | -0.12 | 0.05 | 1.60E-02 | 1.39E-01 | 28 | -0.03 | 0.03 | 3.14E-01 | 0.82 | 1.72E-02 | - | - |
| Cream, creme fraiche, sour cream | Food | 10673 | -0.03 | 0.01 | 5.80E-02 | 4.08E-01 | 34 | 10406 | 0.01 | 0.01 | 6.51E-01 | 8.82E-01 | 179 | -0.01 | 0.01 | 3.19E-01 | 0.64 | 9.39E-02 | - | - |
| Satisfaction with work situation | Psychosocial | 6281 | -0.01 | 0.02 | 7.81E-01 | 9.57E-01 | 198 | 6119 | 0.03 | 0.02 | 9.18E-02 | 3.91E-01 | 57 | 0.01 | 0.01 | 3.19E-01 | 0.48 | 1.65E-01 | - | - |
| Rosehip, sweet syrup soup | Food | 10673 | 0.01 | 0.01 | 3.03E-01 | 7.61E-01 | 96 | 10406 | 0.01 | 0.01 | 7.13E-01 | 9.07E-01 | 191 | 0.01 | 0.01 | 3.22E-01 | 0.00 | 6.43E-01 | - | - |
| Breakfast habits: Coffee/tea and wheat buns or rusk for breakfast vs not breakfast at all | Food | 896 | -0.10 | 0.12 | 4.04E-01 | 8.00E-01 | 122 | 859 | -0.07 | 0.12 | 5.72E-01 | 8.64E-01 | 160 | -0.08 | 0.08 | 3.23E-01 | 0.00 | 8.44E-01 | - | - |
| Vitamin B6 intake (mg/day) | Nutrients | 10673 | 0.01 | 0.01 | 6.50E-01 | 9.29E-01 | 169 | 10406 | 0.01 | 0.02 | 3.43E-01 | 7.56E-01 | 109 | 0.01 | 0.01 | 3.23E-01 | 0.00 | 7.19E-01 | - | - |
| Alcohol intake (g/day) | Alcohol | 10673 | -0.01 | 0.01 | 3.28E-01 | 7.61E-01 | 102 | 10406 | -0.01 | 0.01 | 6.96E-01 | 8.95E-01 | 189 | -0.01 | 0.01 | 3.30E-01 | 0.00 | 6.89E-01 | - | - |
| Travel to work: Walk to work vs passive travel to work | Physical activity | 7455 | -0.09 | 0.05 | 9.41E-02 | 4.40E-01 | 52 | 7283 | 0.02 | 0.05 | 7.43E-01 | 9.21E-01 | 196 | -0.04 | 0.04 | 3.33E-01 | 0.49 | 1.60E-01 | - | - |
| Brown beans, pea soup | Food | 10673 | 0.00 | 0.01 | 8.51E-01 | 9.59E-01 | 213 | 10406 | 0.02 | 0.01 | 2.33E-01 | 6.66E-01 | 84 | 0.01 | 0.01 | 3.34E-01 | 0.00 | 4.68E-01 | - | - |
| Permanent employment | Psychosocial | 11291 | 0.01 | 0.03 | 7.56E-01 | 9.56E-01 | 192 | 11058 | 0.03 | 0.03 | 3.03E-01 | 7.51E-01 | 98 | 0.02 | 0.02 | 3.45E-01 | 0.00 | 6.06E-01 | - | - |
| Appetite status | Psychosocial | 6334 | -0.02 | 0.02 | 2.46E-01 | 7.25E-01 | 81 | 6182 | 0.00 | 0.02 | 8.81E-01 | 9.79E-01 | 217 | -0.01 | 0.01 | 3.55E-01 | 0.00 | 4.74E-01 | - | - |
| Sum of phytosterols intake (mg/day) | Nutrients | 10673 | 0.00 | 0.01 | 9.13E-01 | 9.77E-01 | 226 | 10406 | -0.02 | 0.01 | 1.63E-01 | 5.29E-01 | 74 | -0.01 | 0.01 | 3.67E-01 | 0.13 | 2.85E-01 | - | - |
| Calcium intake (mg/day) | Nutrients | 10673 | 0.01 | 0.01 | 3.63E-01 | 7.61E-01 | 116 | 10406 | 0.01 | 0.01 | 7.25E-01 | 9.13E-01 | 193 | 0.01 | 0.01 | 3.71E-01 | 0.00 | 6.98E-01 | - | - |
| Chips, popcorn, salted nuts | Food | 10673 | 0.01 | 0.01 | 7.21E-01 | 9.53E-01 | 184 | 10406 | -0.02 | 0.01 | 1.09E-01 | 4.45E-01 | 59 | -0.01 | 0.01 | 3.74E-01 | 0.47 | 1.68E-01 | - | - |
| Medioresinol intake (ug/day) | Nutrients | 10673 | 0.01 | 0.01 | 3.01E-01 | 7.61E-01 | 95 | 10406 | 0.00 | 0.01 | 8.38E-01 | 9.75E-01 | 209 | 0.01 | 0.01 | 3.78E-01 | 0.00 | 5.62E-01 | - | - |
| Feel the need to reduce alcohol consumption | Alcohol | 10418 | -0.02 | 0.05 | 7.43E-01 | 9.56E-01 | 188 | 10277 | 0.07 | 0.05 | 1.15E-01 | 4.50E-01 | 62 | 0.03 | 0.03 | 3.79E-01 | 0.45 | 1.77E-01 | - | - |
| Wine | Alcohol | 10673 | -0.02 | 0.01 | 2.70E-01 | 7.27E-01 | 89 | 10406 | 0.00 | 0.01 | 9.08E-01 | 9.79E-01 | 223 | -0.01 | 0.01 | 3.83E-01 | 0.00 | 4.93E-01 | - | - |
| Carbohydrates intake (g/day) | Nutrients | 10673 | 0.01 | 0.01 | 3.07E-01 | 7.61E-01 | 97 | 10406 | 0.00 | 0.01 | 8.63E-01 | 9.79E-01 | 213 | 0.01 | 0.01 | 3.96E-01 | 0.00 | 5.52E-01 | - | - |
| If you exercise, change in exercise habits during the last year | Physical activity | 10086 | -0.02 | 0.01 | 8.30E-02 | 4.39E-01 | 46 | 9886 | 0.01 | 0.01 | 5.85E-01 | 8.67E-01 | 164 | -0.01 | 0.01 | 3.96E-01 | 0.61 | 1.08E-01 | - | - |
| Coffee rolls/buns, rusk | Food | 10673 | 0.01 | 0.01 | 7.19E-01 | 9.53E-01 | 182 | 10406 | 0.01 | 0.01 | 4.11E-01 | 7.91E-01 | 126 | 0.01 | 0.01 | 4.03E-01 | 0.00 | 7.44E-01 | - | - |
| Liquor, spirits | Alcohol | 10673 | 0.00 | 0.01 | 8.37E-01 | 9.59E-01 | 210 | 10406 | 0.01 | 0.01 | 3.33E-01 | 7.56E-01 | 105 | 0.01 | 0.01 | 4.09E-01 | 0.00 | 5.84E-01 | - | - |
| Enterodiol intake (ug/day) | Nutrients | 10673 | 0.02 | 0.01 | 2.75E-01 | 7.27E-01 | 91 | 10406 | 0.00 | 0.01 | 9.75E-01 | 9.79E-01 | 242 | 0.01 | 0.01 | 4.22E-01 | 0.00 | 4.59E-01 | - | - |
| Frequency of gardening during leisure time | Physical activity | 10924 | 0.00 | 0.01 | 8.61E-01 | 9.59E-01 | 218 | 10702 | -0.01 | 0.01 | 3.38E-01 | 7.56E-01 | 107 | -0.01 | 0.01 | 4.27E-01 | 0.00 | 5.72E-01 | - | - |
| Job demands to work very fast | Psychosocial | 11376 | 0.01 | 0.01 | 6.98E-01 | 9.53E-01 | 177 | 11126 | -0.02 | 0.01 | 1.32E-01 | 4.93E-01 | 65 | -0.01 | 0.01 | 4.36E-01 | 0.45 | 1.78E-01 | - | - |
| Pasta | Food | 10673 | -0.01 | 0.01 | 6.57E-01 | 9.29E-01 | 172 | 10406 | -0.01 | 0.01 | 5.12E-01 | 8.40E-01 | 148 | -0.01 | 0.01 | 4.36E-01 | 0.00 | 8.82E-01 | - | - |
| Polyunsaturated fat intake (g/day) | Nutrients | 10673 | 0.00 | 0.01 | 7.52E-01 | 9.56E-01 | 191 | 10406 | -0.01 | 0.01 | 4.31E-01 | 7.95E-01 | 131 | -0.01 | 0.01 | 4.37E-01 | 0.00 | 7.35E-01 | - | - |
| Minced meat dishes | Food | 10673 | -0.01 | 0.01 | 6.11E-01 | 9.26E-01 | 158 | 10406 | -0.01 | 0.01 | 5.67E-01 | 8.64E-01 | 159 | -0.01 | 0.01 | 4.45E-01 | 0.00 | 9.68E-01 | - | - |
| Bregott on bread | Food | 10673 | 0.00 | 0.01 | 9.46E-01 | 9.77E-01 | 233 | 10406 | -0.02 | 0.01 | 2.54E-01 | 6.77E-01 | 91 | -0.01 | 0.01 | 4.49E-01 | 0.00 | 3.91E-01 | - | - |
| Margarine on bread | Food | 10673 | 0.01 | 0.01 | 5.20E-01 | 8.99E-01 | 140 | 10406 | 0.01 | 0.01 | 6.75E-01 | 8.92E-01 | 182 | 0.01 | 0.01 | 4.51E-01 | 0.00 | 8.82E-01 | - | - |
| Trans fat intake (g/day) | Nutrients | 10673 | 0.00 | 0.02 | 9.50E-01 | 9.77E-01 | 234 | 10406 | -0.02 | 0.02 | 3.16E-01 | 7.55E-01 | 100 | -0.01 | 0.01 | 4.51E-01 | 0.00 | 5.06E-01 | - | - |
| Campestanol intake (mg/day) | Nutrients | 10673 | 0.01 | 0.01 | 3.29E-01 | 7.61E-01 | 103 | 10406 | 0.00 | 0.01 | 9.41E-01 | 9.79E-01 | 233 | 0.01 | 0.01 | 4.54E-01 | 0.00 | 5.28E-01 | - | - |
| Medium beer | Alcohol | 10673 | 0.01 | 0.01 | 6.41E-01 | 9.29E-01 | 167 | 10406 | 0.01 | 0.01 | 5.51E-01 | 8.54E-01 | 156 | 0.01 | 0.01 | 4.54E-01 | 0.00 | 9.12E-01 | - | - |
| Sour milk, yoghurt (3% fat) | Food | 10673 | -0.02 | 0.01 | 1.43E-01 | 5.70E-01 | 61 | 10406 | 0.01 | 0.01 | 6.75E-01 | 8.92E-01 | 183 | -0.01 | 0.01 | 4.54E-01 | 0.43 | 1.85E-01 | - | - |
| Equol intake (ug/day) | Nutrients | 10673 | 0.01 | 0.01 | 5.90E-01 | 9.21E-01 | 155 | 10406 | 0.01 | 0.01 | 6.14E-01 | 8.82E-01 | 169 | 0.01 | 0.01 | 4.60E-01 | 0.00 | 9.85E-01 | - | - |
| Do you feel important and appreciated in your home? | Psychosocial | 6297 | 0.01 | 0.02 | 7.19E-01 | 9.53E-01 | 183 | 6129 | 0.01 | 0.02 | 5.04E-01 | 8.39E-01 | 146 | 0.01 | 0.01 | 4.67E-01 | 0.00 | 8.30E-01 | - | - |
| Vitamin D intake (ug/day) | Nutrients | 10673 | 0.02 | 0.01 | 1.60E-01 | 5.99E-01 | 65 | 10406 | -0.01 | 0.01 | 6.95E-01 | 8.95E-01 | 188 | 0.01 | 0.01 | 4.68E-01 | 0.37 | 2.06E-01 | - | - |
| Selenium intake (ug/day) | Nutrients | 10673 | -0.01 | 0.01 | 5.84E-01 | 9.21E-01 | 154 | 10406 | -0.01 | 0.01 | 6.38E-01 | 8.82E-01 | 173 | -0.01 | 0.01 | 4.71E-01 | 0.00 | 9.67E-01 | - | - |
| Boiled or baked potato | Food | 10673 | -0.01 | 0.01 | 4.40E-01 | 8.36E-01 | 128 | 10406 | 0.00 | 0.01 | 8.31E-01 | 9.71E-01 | 207 | -0.01 | 0.01 | 4.83E-01 | 0.00 | 6.99E-01 | - | - |
| Campesterol intake (mg/day) | Nutrients | 10673 | 0.01 | 0.01 | 6.37E-01 | 9.29E-01 | 166 | 10406 | -0.02 | 0.01 | 1.41E-01 | 4.96E-01 | 68 | -0.01 | 0.01 | 4.83E-01 | 0.47 | 1.68E-01 | - | - |
| Frequncy of social contacts with colleagues during leisure time | Psychosocial | 11128 | 0.00 | 0.01 | 8.01E-01 | 9.59E-01 | 201 | 10861 | -0.02 | 0.01 | 2.12E-01 | 6.45E-01 | 80 | -0.01 | 0.01 | 4.85E-01 | 0.12 | 2.87E-01 | - | - |
| Sodium intake (mg/day) | Nutrients | 10673 | 0.01 | 0.01 | 6.93E-01 | 9.53E-01 | 175 | 10406 | 0.01 | 0.01 | 5.55E-01 | 8.54E-01 | 158 | 0.01 | 0.01 | 4.87E-01 | 0.00 | 8.83E-01 | - | - |
| Energy status | Psychosocial | 6332 | 0.01 | 0.02 | 6.17E-01 | 9.26E-01 | 161 | 6168 | 0.01 | 0.02 | 6.43E-01 | 8.82E-01 | 175 | 0.01 | 0.01 | 4.96E-01 | 0.00 | 9.78E-01 | - | - |
| Docosahexaenoic acid (DHA) intake (g/day) | Nutrients | 10673 | 0.00 | 0.01 | 9.73E-01 | 9.88E-01 | 239 | 10406 | -0.01 | 0.01 | 3.50E-01 | 7.56E-01 | 112 | -0.01 | 0.01 | 5.01E-01 | 0.00 | 5.16E-01 | - | - |
| Salad dressing with oil | Food | 10673 | -0.01 | 0.01 | 3.94E-01 | 7.97E-01 | 120 | 10406 | 0.00 | 0.01 | 9.36E-01 | 9.79E-01 | 230 | -0.01 | 0.01 | 5.05E-01 | 0.00 | 5.90E-01 | - | - |
| Rice | Food | 10673 | 0.00 | 0.01 | 9.00E-01 | 9.77E-01 | 222 | 10406 | 0.01 | 0.01 | 4.23E-01 | 7.91E-01 | 129 | 0.01 | 0.01 | 5.15E-01 | 0.00 | 6.29E-01 | - | - |
| Margarine for cooking | Food | 10673 | 0.01 | 0.01 | 4.32E-01 | 8.26E-01 | 127 | 10406 | -0.02 | 0.01 | 8.69E-02 | 3.91E-01 | 54 | -0.01 | 0.01 | 5.21E-01 | 0.68 | 7.64E-02 | - | - |

| | | | | | | | | | | | | | | | | | | | | |
|---|---|---|---|---|---|---|---|---|---|---|---|---|---|---|---|---|---|---|---|---|
| Folic acid intake (ug/day) | Nutrients | 10673 | -0.01 | 0.01 | 6.47E-01 | 9.29E-01 | 168 | 10406 | -0.01 | 0.02 | 6.63E-01 | 8.91E-01 | 181 | -0.01 | 0.01 | 5.27E-01 | 0.00 | 9.90E-01 | - | - |
| Number of friends that can come to your home at any time and feel at home | Social | 11467 | 0.00 | 0.01 | 9.17E-01 | 9.77E-01 | 228 | 11236 | 0.01 | 0.01 | 4.32E-01 | 7.95E-01 | 132 | 0.01 | 0.01 | 5.32E-01 | 0.00 | 6.25E-01 | - | - |
| Vitamin E intake (mg/day) | Nutrients | 10673 | 0.00 | 0.01 | 8.28E-01 | 9.59E-01 | 205 | 10406 | -0.02 | 0.01 | 2.77E-01 | 7.02E-01 | 96 | -0.01 | 0.01 | 5.42E-01 | 0.00 | 3.55E-01 | - | - |
| Sedentary or standing work | Physical activity | 11349 | 0.03 | 0.03 | 4.04E-01 | 8.00E-01 | 121 | 11084 | 0.00 | 0.03 | 9.90E-01 | 9.90E-01 | 243 | 0.01 | 0.02 | 5.46E-01 | 0.00 | 5.64E-01 | - | - |
| Beta-sitostanol intake (mg/day) | Nutrients | 10673 | 0.01 | 0.01 | 3.68E-01 | 7.65E-01 | 117 | 10406 | 0.00 | 0.01 | 9.45E-01 | 9.79E-01 | 234 | 0.01 | 0.01 | 5.53E-01 | 0.00 | 4.97E-01 | - | - |
| Cheese 10-17% | Food | 10673 | 0.02 | 0.01 | 1.52E-01 | 5.94E-01 | 62 | 10406 | -0.01 | 0.01 | 5.46E-01 | 8.54E-01 | 155 | 0.01 | 0.01 | 5.56E-01 | 0.52 | 1.50E-01 | - | - |
| Potassium intake (mg/day) | Nutrients | 10673 | -0.01 | 0.02 | 5.76E-01 | 9.21E-01 | 150 | 10406 | 0.00 | 0.02 | 8.07E-01 | 9.62E-01 | 204 | -0.01 | 0.01 | 5.69E-01 | 0.00 | 8.27E-01 | - | - |
| Smoking status: Smokers vs non-smokers | Tobacco use | 7604 | -0.02 | 0.04 | 5.62E-01 | 9.21E-01 | 148 | 7374 | -0.01 | 0.04 | 8.25E-01 | 9.71E-01 | 205 | -0.02 | 0.03 | 5.69E-01 | 0.00 | 8.06E-01 | - | - |
| Butter on bread | Food | 10673 | -0.01 | 0.01 | 5.25E-01 | 8.99E-01 | 142 | 10406 | 0.00 | 0.01 | 8.87E-01 | 9.79E-01 | 219 | -0.01 | 0.01 | 5.76E-01 | 0.00 | 7.38E-01 | - | - |
| Fibre intake (g/day) | Nutrients | 10673 | 0.00 | 0.01 | 7.61E-01 | 9.56E-01 | 193 | 10406 | -0.01 | 0.02 | 6.31E-01 | 8.82E-01 | 171 | -0.01 | 0.01 | 5.80E-01 | 0.00 | 8.97E-01 | - | - |
| Cohabitation: Live alone vs Only one adult (spouse, partner) | Social | 3391 | -0.02 | 0.06 | 7.06E-01 | 9.53E-01 | 178 | 3460 | -0.02 | 0.06 | 6.93E-01 | 8.95E-01 | 186 | -0.02 | 0.04 | 5.85E-01 | 0.00 | 9.92E-01 | - | - |
| Participation in sports or physical exercise associations | Social | 724 | 0.01 | 0.11 | 9.42E-01 | 9.77E-01 | 231 | 643 | -0.10 | 0.12 | 4.09E-01 | 7.91E-01 | 125 | -0.04 | 0.08 | 6.10E-01 | 0.00 | 5.14E-01 | - | - |
| Number of people with whom you can speak openly | Social | 11483 | -0.01 | 0.01 | 4.05E-01 | 8.00E-01 | 123 | 11256 | 0.00 | 0.01 | 8.98E-01 | 9.79E-01 | 222 | 0.00 | 0.01 | 6.12E-01 | 0.00 | 5.01E-01 | - | - |
| Shellfish (e.g. shrimps, scallops) | Food | 4726 | 0.00 | 0.02 | 9.50E-01 | 9.77E-01 | 235 | 4623 | 0.02 | 0.02 | 4.15E-01 | 7.91E-01 | 127 | 0.01 | 0.01 | 6.13E-01 | 0.00 | 5.20E-01 | - | - |
| Frequent social contacts with colleagues during work | Psychosocial | 11294 | 0.00 | 0.01 | 8.30E-01 | 9.59E-01 | 206 | 11034 | -0.01 | 0.01 | 3.60E-01 | 7.62E-01 | 115 | 0.00 | 0.01 | 6.27E-01 | 0.00 | 4.21E-01 | - | - |
| People to ask for help apart from the ones at home | Social | 11463 | -0.03 | 0.04 | 5.46E-01 | 9.14E-01 | 145 | 11243 | 0.00 | 0.04 | 9.38E-01 | 9.79E-01 | 231 | -0.02 | 0.03 | 6.29E-01 | 0.00 | 7.11E-01 | - | - |
| Linoleic acid intake (g/day) | Nutrients | 10673 | -0.01 | 0.01 | 5.99E-01 | 9.26E-01 | 157 | 10406 | 0.00 | 0.01 | 9.16E-01 | 9.79E-01 | 224 | 0.00 | 0.01 | 6.53E-01 | 0.00 | 7.69E-01 | - | - |
| Feel uneasy or guilty because of your way of drinking | Alcohol | 10416 | 0.00 | 0.04 | 9.93E-01 | 9.93E-01 | 243 | 10272 | 0.03 | 0.04 | 5.41E-01 | 8.54E-01 | 153 | 0.01 | 0.03 | 6.64E-01 | 0.00 | 6.67E-01 | - | - |
| Syringaresinol intake (ug/day) | Nutrients | 10673 | 0.01 | 0.01 | 5.18E-01 | 8.99E-01 | 139 | 10406 | 0.00 | 0.01 | 9.63E-01 | 9.79E-01 | 238 | 0.00 | 0.01 | 6.68E-01 | 0.00 | 6.27E-01 | - | - |
| Fatty fish (e.g. herring, whitefish, salmon) | Food | 10673 | 0.02 | 0.01 | 2.56E-01 | 7.27E-01 | 84 | 10406 | -0.01 | 0.01 | 5.77E-01 | 8.66E-01 | 162 | 0.00 | 0.01 | 6.69E-01 | 0.30 | 2.34E-01 | - | - |
| Pinoresinol intake (ug/day) | Nutrients | 10673 | 0.00 | 0.01 | 9.44E-01 | 9.77E-01 | 232 | 10406 | -0.01 | 0.01 | 4.95E-01 | 8.29E-01 | 145 | 0.00 | 0.01 | 6.69E-01 | 0.00 | 5.91E-01 | - | - |
| Ice cream | Food | 10673 | -0.01 | 0.01 | 6.79E-01 | 9.53E-01 | 173 | 10406 | 0.00 | 0.01 | 8.53E-01 | 9.79E-01 | 211 | 0.00 | 0.01 | 6.73E-01 | 0.00 | 8.66E-01 | - | - |
| Eicosapentaenoic acid (EPA) intake (g/day) | Nutrients | 10673 | 0.00 | 0.01 | 9.68E-01 | 9.88E-01 | 238 | 10406 | -0.01 | 0.01 | 5.21E-01 | 8.49E-01 | 149 | 0.00 | 0.01 | 6.77E-01 | 0.00 | 6.24E-01 | - | - |
| Blood based food | Food | 4726 | 0.03 | 0.02 | 1.75E-01 | 6.16E-01 | 69 | 4623 | -0.02 | 0.02 | 3.66E-01 | 7.64E-01 | 116 | 0.01 | 0.01 | 6.84E-01 | 0.60 | 1.14E-01 | - | - |
| Tomato, cucumber | Food | 10673 | 0.00 | 0.01 | 8.33E-01 | 9.59E-01 | 208 | 10406 | -0.01 | 0.01 | 7.23E-01 | 9.13E-01 | 192 | 0.00 | 0.01 | 6.90E-01 | 0.00 | 9.16E-01 | - | - |
| Frequency of engaging in clubs, associations or study circles | Social | 7691 | -0.02 | 0.02 | 3.45E-01 | 7.61E-01 | 105 | 7550 | 0.01 | 0.02 | 6.94E-01 | 8.95E-01 | 187 | 0.00 | 0.01 | 6.95E-01 | 0.00 | 3.44E-01 | - | - |
| Light and physically active work | Physical activity | 11349 | -0.01 | 0.03 | 6.91E-01 | 9.53E-01 | 174 | 11084 | 0.00 | 0.03 | 8.79E-01 | 9.79E-01 | 216 | -0.01 | 0.02 | 6.97E-01 | 0.00 | 8.65E-01 | - | - |
| Learn new things at job | Psychosocial | 11359 | -0.01 | 0.01 | 6.25E-01 | 9.26E-01 | 164 | 11117 | 0.00 | 0.01 | 9.62E-01 | 9.79E-01 | 237 | 0.00 | 0.01 | 7.02E-01 | 0.00 | 7.57E-01 | - | - |
| Sugar, honey, marmelade, jam | Food | 10673 | 0.01 | 0.01 | 6.55E-01 | 9.29E-01 | 170 | 10406 | 0.00 | 0.01 | 9.29E-01 | 9.79E-01 | 227 | 0.00 | 0.01 | 7.04E-01 | 0.00 | 8.03E-01 | - | - |
| Smoked fish/meat | Food | 10673 | 0.00 | 0.01 | 9.53E-01 | 9.77E-01 | 237 | 10406 | 0.01 | 0.01 | 6.44E-01 | 8.82E-01 | 176 | 0.00 | 0.01 | 7.14E-01 | 0.00 | 7.74E-01 | - | - |
| Sausage as main dish | Food | 10673 | 0.00 | 0.01 | 9.22E-01 | 9.77E-01 | 229 | 10406 | 0.01 | 0.01 | 5.45E-01 | 8.54E-01 | 154 | 0.00 | 0.01 | 7.25E-01 | 0.00 | 6.15E-01 | - | - |
| Number of social contacts with the same interests as you | Social | 11468 | 0.01 | 0.01 | 7.11E-01 | 9.53E-01 | 181 | 11236 | -0.01 | 0.01 | 3.91E-01 | 7.89E-01 | 120 | 0.00 | 0.01 | 7.36E-01 | 0.00 | 3.83E-01 | - | - |
| Animal based protein intake (g/day) | Nutrients | 10673 | 0.00 | 0.01 | 9.87E-01 | 9.92E-01 | 242 | 10406 | 0.01 | 0.01 | 6.42E-01 | 8.82E-01 | 174 | 0.00 | 0.01 | 7.37E-01 | 0.00 | 7.48E-01 | - | - |
| Whole grain soft bread | Food | 10673 | 0.01 | 0.01 | 5.49E-01 | 9.14E-01 | 146 | 10406 | 0.00 | 0.01 | 8.70E-01 | 9.79E-01 | 214 | 0.00 | 0.01 | 7.52E-01 | 0.00 | 5.93E-01 | - | - |
| Iodine intake (ug/day) | Nutrients | 10673 | 0.01 | 0.01 | 6.13E-01 | 9.26E-01 | 159 | 10406 | 0.00 | 0.01 | 9.31E-01 | 9.79E-01 | 228 | 0.00 | 0.01 | 7.62E-01 | 0.00 | 6.79E-01 | - | - |
| Salty fish | Food | 10673 | 0.00 | 0.01 | 7.35E-01 | 9.56E-01 | 186 | 10406 | 0.00 | 0.01 | 9.34E-01 | 9.79E-01 | 229 | 0.00 | 0.01 | 7.64E-01 | 0.00 | 8.60E-01 | - | - |
| Tiamin intake (mg/day) | Nutrients | 10673 | 0.00 | 0.01 | 7.63E-01 | 9.56E-01 | 194 | 10406 | -0.01 | 0.01 | 4.73E-01 | 8.10E-01 | 142 | 0.00 | 0.01 | 7.71E-01 | 0.00 | 4.71E-01 | - | - |
| Frequency of dancing during leisure time | Physical activity | 10244 | 0.01 | 0.01 | 5.33E-01 | 9.02E-01 | 143 | 10020 | -0.02 | 0.01 | 2.98E-01 | 7.46E-01 | 97 | 0.00 | 0.01 | 7.80E-01 | 0.28 | 2.38E-01 | - | - |
| Hearing status | General health | 6339 | 0.02 | 0.02 | 2.18E-01 | 7.01E-01 | 75 | 6188 | -0.02 | 0.02 | 4.02E-01 | 7.91E-01 | 123 | 0.00 | 0.01 | 7.86E-01 | 0.53 | 1.43E-01 | - | - |
| Matairesinol intake (ug/day) | Nutrients | 10673 | 0.00 | 0.01 | 9.77E-01 | 9.88E-01 | 240 | 10406 | -0.01 | 0.01 | 6.80E-01 | 8.93E-01 | 185 | 0.00 | 0.01 | 7.88E-01 | 0.00 | 7.53E-01 | - | - |
| Enterolactone intake (ug/day) | Nutrients | 10673 | 0.00 | 0.01 | 9.01E-01 | 9.77E-01 | 224 | 10406 | 0.00 | 0.01 | 7.99E-01 | 9.61E-01 | 202 | 0.00 | 0.01 | 7.90E-01 | 0.00 | 9.25E-01 | - | - |
| Total protein intake (g/day) | Nutrients | 10673 | 0.00 | 0.01 | 9.15E-01 | 9.77E-01 | 227 | 10406 | 0.01 | 0.01 | 6.25E-01 | 8.82E-01 | 170 | 0.00 | 0.01 | 7.91E-01 | 0.00 | 6.72E-01 | - | - |
| Changed everyday exercise during the last year | Physical activity | 11514 | -0.02 | 0.01 | 2.33E-01 | 7.15E-01 | 79 | 11277 | 0.02 | 0.01 | 1.10E-01 | 4.45E-01 | 60 | 0.00 | 0.01 | 7.92E-01 | 0.74 | 4.79E-02 | - | - |
| Monounsaturated fat intake (g/day) | Nutrients | 10673 | 0.00 | 0.02 | 8.96E-01 | 9.77E-01 | 221 | 10406 | -0.01 | 0.02 | 6.10E-01 | 8.82E-01 | 167 | 0.00 | 0.01 | 7.92E-01 | 0.00 | 6.48E-01 | - | - |
| Confidence status | Psychosocial | 6331 | -0.01 | 0.02 | 7.48E-01 | 9.56E-01 | 190 | 6178 | 0.00 | 0.02 | 9.75E-01 | 9.79E-01 | 241 | 0.00 | 0.01 | 8.02E-01 | 0.00 | 8.38E-01 | - | - |
| Arachidonic acid (ARA) intake (g/day) | Nutrients | 10673 | 0.00 | 0.01 | 7.75E-01 | 9.57E-01 | 195 | 10406 | 0.00 | 0.01 | 9.51E-01 | 9.79E-01 | 235 | 0.00 | 0.01 | 8.04E-01 | 0.00 | 8.77E-01 | - | - |
| Iron intake (mg/day) | Nutrients | 10673 | 0.00 | 0.02 | 8.37E-01 | 9.59E-01 | 209 | 10406 | -0.01 | 0.02 | 5.83E-01 | 8.67E-01 | 163 | 0.00 | 0.01 | 8.13E-01 | 0.00 | 5.92E-01 | - | - |
| Fat intake (g/day) | Nutrients | 10673 | 0.00 | 0.01 | 8.57E-01 | 9.59E-01 | 216 | 10406 | 0.00 | 0.01 | 8.79E-01 | 9.79E-01 | 215 | 0.00 | 0.01 | 8.14E-01 | 0.00 | 9.85E-01 | - | - |
| Plant based protein intake (g/day) | Nutrients | 10673 | 0.00 | 0.01 | 7.84E-01 | 9.57E-01 | 199 | 10406 | 0.00 | 0.01 | 9.68E-01 | 9.79E-01 | 239 | 0.00 | 0.01 | 8.23E-01 | 0.00 | 8.70E-01 | - | - |
| Monosaccharides intake (g/day) | Nutrients | 10673 | 0.00 | 0.02 | 9.01E-01 | 9.77E-01 | 223 | 10406 | 0.00 | 0.02 | 8.49E-01 | 9.79E-01 | 210 | 0.00 | 0.01 | 8.24E-01 | 0.00 | 9.61E-01 | - | - |
| Linolenic acid intake (g/day) | Nutrients | 10673 | 0.00 | 0.01 | 9.52E-01 | 9.77E-01 | 236 | 10406 | 0.00 | 0.01 | 7.34E-01 | 9.19E-01 | 194 | 0.00 | 0.01 | 8.45E-01 | 0.00 | 7.76E-01 | - | - |
| Corn flakes | Food | 10673 | 0.01 | 0.01 | 5.34E-01 | 9.02E-01 | 144 | 10406 | -0.01 | 0.01 | 3.68E-01 | 7.64E-01 | 117 | 0.00 | 0.01 | 8.49E-01 | 0.14 | 2.81E-01 | - | - |
| Sometimes physically straining work | Physical activity | 11349 | 0.02 | 0.03 | 5.64E-01 | 9.21E-01 | 149 | 11084 | -0.01 | 0.03 | 7.49E-01 | 9.24E-01 | 197 | 0.00 | 0.02 | 8.50E-01 | 0.00 | 5.28E-01 | - | - |
| Zinc intake (mg/day) | Nutrients | 10673 | 0.00 | 0.02 | 7.81E-01 | 9.57E-01 | 197 | 10406 | 0.00 | 0.02 | 9.70E-01 | 9.79E-01 | 240 | 0.00 | 0.01 | 8.64E-01 | 0.00 | 8.25E-01 | - | - |
| Vitamin C intake (mg/day) | Nutrients | 10673 | -0.01 | 0.01 | 3.43E-01 | 7.61E-01 | 104 | 10406 | 0.01 | 0.01 | 4.65E-01 | 8.10E-01 | 139 | 0.00 | 0.01 | 8.65E-01 | 0.29 | 2.36E-01 | - | - |
| Oil for cooking | Food | 10673 | -0.01 | 0.01 | 4.92E-01 | 8.78E-01 | 135 | 10406 | 0.01 | 0.01 | 6.47E-01 | 8.82E-01 | 177 | 0.00 | 0.01 | 8.67E-01 | 0.00 | 4.19E-01 | - | - |
| Pentadecanoic acid intake (g/day) | Nutrients | 10673 | -0.01 | 0.01 | 6.21E-01 | 9.26E-01 | 162 | 10406 | 0.00 | 0.01 | 7.90E-01 | 9.59E-01 | 200 | 0.00 | 0.01 | 8.68E-01 | 0.00 | 5.91E-01 | - | - |
| Heptadecanoic acid intake (g/day) | Nutrients | 10673 | -0.01 | 0.01 | 6.21E-01 | 9.26E-01 | 163 | 10406 | 0.00 | 0.01 | 7.90E-01 | 9.59E-01 | 199 | 0.00 | 0.01 | 8.68E-01 | 0.00 | 5.91E-01 | - | - |
| Pizza | Food | 10673 | 0.01 | 0.01 | 4.72E-01 | 8.62E-01 | 133 | 10406 | -0.01 | 0.02 | 3.37E-01 | 7.56E-01 | 106 | 0.00 | 0.01 | 8.69E-01 | 0.29 | 2.35E-01 | - | - |
| Formic acid intaje (g/day) | Nutrients | 10673 | 0.00 | 0.01 | 7.32E-01 | 9.56E-01 | 185 | 10406 | 0.00 | 0.01 | 8.82E-01 | 9.79E-01 | 218 | 0.00 | 0.01 | 8.88E-01 | 0.00 | 7.29E-01 | - | - |
| Phosphate intake (mg/day) | Nutrients | 10673 | 0.01 | 0.01 | 6.57E-01 | 9.29E-01 | 171 | 10406 | 0.00 | 0.02 | 7.93E-01 | 9.59E-01 | 201 | 0.00 | 0.01 | 8.96E-01 | 0.00 | 6.18E-01 | - | - |
| Meat stew | Food | 10673 | -0.02 | 0.01 | 2.98E-01 | 7.61E-01 | 94 | 10406 | 0.01 | 0.01 | 3.95E-01 | 7.89E-01 | 121 | 0.00 | 0.01 | 8.98E-01 | 0.44 | 1.81E-01 | - | - |
| Palmitic acid intake (g/day) | Nutrients | 10673 | 0.00 | 0.01 | 8.10E-01 | 9.59E-01 | 202 | 10406 | 0.01 | 0.01 | 6.76E-01 | 8.92E-01 | 184 | 0.00 | 0.01 | 9.03E-01 | 0.00 | 6.40E-01 | - | - |
| Breakfast habits: Porridge w/o sandwich for breakfast vs not breakfast at all | Food | 1559 | -0.01 | 0.08 | 9.27E-01 | 9.77E-01 | 230 | 1482 | 0.00 | 0.09 | 9.56E-01 | 9.79E-01 | 236 | -0.01 | 0.06 | 9.17E-01 | 0.00 | 9.83E-01 | - | - |
| Sum of all lignans intake (ug/day) | Nutrients | 10673 | 0.00 | 0.01 | 7.45E-01 | 9.56E-01 | 189 | 10406 | -0.01 | 0.01 | 6.53E-01 | 8.82E-01 | 180 | 0.00 | 0.01 | 9.34E-01 | 0.00 | 5.83E-01 | - | - |
| Marital status: Single vs Widow/widower | Social | 1178 | -0.25 | 0.21 | 2.48E-01 | 7.25E-01 | 83 | 1148 | 0.18 | 0.19 | 3.55E-01 | 7.56E-01 | 114 | -0.01 | 0.14 | 9.35E-01 | 0.54 | 1.39E-01 | - | - |
| Breakfast habits: Gruel w/o sandwich for breakfast vs not breakfast at all | Food | 856 | 0.02 | 0.13 | 8.67E-01 | 9.62E-01 | 219 | 802 | -0.01 | 0.14 | 9.39E-01 | 9.79E-01 | 232 | 0.01 | 0.10 | 9.45E-01 | 0.00 | 8.65E-01 | - | - |
| Stigmasterol intake (mg/day) | Nutrients | 10673 | 0.01 | 0.01 | 4.98E-01 | 8.78E-01 | 138 | 10406 | -0.01 | 0.01 | 5.37E-01 | 8.54E-01 | 152 | 0.00 | 0.01 | 9.61E-01 | 0.00 | 3.60E-01 | - | - |
| Saturated fat intake (g/day) | Nutrients | 10673 | 0.00 | 0.01 | 7.76E-01 | 9.57E-01 | 196 | 10406 | 0.00 | 0.01 | 8.25E-01 | 9.71E-01 | 206 | 0.00 | 0.01 | 9.62E-01 | 0.00 | 7.21E-01 | - | - |
| White (soft) bread, thin crisp bread | Food | 10673 | 0.00 | 0.01 | 7.93E-01 | 9.59E-01 | 200 | 10406 | 0.00 | 0.01 | 7.38E-01 | 9.20E-01 | 195 | 0.00 | 0.01 | 9.62E-01 | 0.00 | 6.73E-01 | - | - |
| Mixed frozen vegetables | Food | 4726 | 0.00 | 0.02 | 8.55E-01 | 9.59E-01 | 215 | 4623 | 0.00 | 0.02 | 8.95E-01 | 9.79E-01 | 221 | 0.00 | 0.01 | 9.68E-01 | 0.00 | 8.24E-01 | - | - |
| Control over planning and execution of the workday | Psychosocial | 11394 | -0.01 | 0.01 | 6.37E-01 | 9.29E-01 | 165 | 11135 | 0.01 | 0.01 | 6.10E-01 | 8.82E-01 | 168 | 0.00 | 0.01 | 9.85E-01 | 0.00 | 4.87E-01 | - | - |
| Low fat milk (0,5%) | Beverage | 10673 | 0.00 | 0.01 | 8.51E-01 | 9.59E-01 | 214 | 10406 | 0.00 | 0.01 | 8.60E-01 | 9.79E-01 | 212 | 0.00 | 0.01 | 9.92E-01 | 0.00 | 7.97E-01 | - | - |
| Potato salad | Food | 4726 | 0.02 | 0.02 | 4.23E-01 | 8.21E-01 | 125 | 4623 | -0.02 | 0.02 | 3.47E-01 | 7.56E-01 | 111 | 0.00 | 0.01 | 9.98E-01 | 0.35 | 2.16E-01 | - | - |
| Last time a colleague visited you at home | Psychosocial | 11262 | 0.01 | 0.01 | 3.91E-01 | 7.97E-01 | 119 | 10985 | -0.01 | 0.01 | 3.81E-01 | 7.78E-01 | 119 | 0.00 | 0.01 | 1.00E+00 | 0.34 | 2.20E-01 | - | - |

**Supplementary Table 21**. Summary of significant top 5 lifestyle variables regarding variance explained for all the cardiometabolic traits in linear mixed model analyses

| Description | Group | BMI | SBP | DBP | Tot Chol | Trigl | HDL-C | LDL-C | F Glu | 2h Glu | N of tentative associations |
|---|---|---|---|---|---|---|---|---|---|---|---|
| Years smoking | Tobacco use | + | - | - | + | + | - | | + | - | 8 |
| Informed of having high blood pressure | General health | + | + | + | | + | - | | + | + | 7 |
| Exercise during the last three months | Physical activity | - | | - | | - | + | - | - | - | 7 |
| Overall state of health during the last year | General health | - | - | - | | - | + | | - | - | 7 |
| Number of cigarretes smoked per day (in groups) | Tobacco use | + | | - | + | + | - | | + | - | 7 |
| Educational level | Psychosocial | - | - | - | - | | + | - | - | | 7 |
| Fitness status | Physical activity | - | - | - | | - | + | | - | | 6 |
| Smoking status: Smokers vs non-smokers | Tobacco use | | | - | + | + | - | | + | - | 6 |
| Smoking status: Former smokers vs non-smokers | Tobacco use | + | - | - | | + | | | + | - | 6 |
| Travel to work: Cycle to work vs passive travel to work | Physical activity | - | - | | - | - | + | | | | 5 |
| Frequency of hunting or fishing during leisure time | Physical activity | | + | + | + | | | | - | - | 5 |
| Brewed (filtered) coffee | Beverage | - | - | - | | - | | | | - | 5 |
| Boiled coffee | Beverage | + | | | + | + | | + | | - | 5 |
| Secoisolariciresinol intake (ug/day) | Nutrients | - | | - | | - | + | | | - | 5 |
| Snuff status: Snuff users vs non-snuff users | Tobacco use | | - | - | | + | + | | | - | 5 |
| Participation in associations or voluntary organisations | Social | | - | - | | - | | | - | - | 5 |
| Alcohol intake (g/day) | Alcohol | - | | | + | | + | | | - | 4 |
| Parents or siblings have diabetes | General health | + | | | | | - | | + | + | 4 |
| Tea | Beverage | - | | | - | | | - | | + | 4 |
| Strong beer | Alcohol | | | | + | + | + | | | - | 4 |
| Average portion size of meat/fish based on photographic illustration of four sizes (smallest to largest) | Food | + | - | | | | | + | | - | 4 |
| Cambridge physical activity index | Physical activity | | | | | - | + | | - | - | 4 |
| Self rate of overall health | General health | | - | - | - | - | | | | | 4 |
| High mental demand from job | Psychosocial | | + | + | + | | | | | | 3 |
| Marital status: Single vs Divorced/separated | Social | - | - | - | | | | | | | 3 |
| Fibre intake (g/day) | Nutrients | - | | | | - | | | - | | 3 |
| Frequency of picking berries or mushrooms during leisure time | Physical activity | | | | + | | | | - | - | 3 |
| Fiber cereals | Food | | | | - | - | | | - | | 3 |
| Apple, pear, peach, orange, mandarin and grapefruit | Food | | | | | - | | | - | - | 3 |
| Wine | Alcohol | - | | | + | | + | | | | 3 |
| Liquor, spirits | Alcohol | + | | | + | | + | | | | 3 |
| Average portion size of vegetables based on photographic illustration of four sizes (smallest to largest) | Food | - | | - | | | | | | - | 3 |
| Overall state of health compared to other your age | General health | | - | | - | | | | | - | 3 |
| Parents or siblings had a cerebral hemorrhage/thrombosis or cardiac infarction before the age of 60 | General health | | + | + | + | | | | | | 3 |
| Teetotaler | Alcohol | + | | | | | | | - | + | 3 |
| Energy status | Psychosocial | | | | | - | + | | | - | 3 |
| Monounsaturated fat intake (g/day) | Nutrients | | | | + | + | | + | | | 3 |
| Cohabitation: Live alone vs Only children | Social | | - | - | - | | | | | | 3 |
| Repetitive job | Psychosocial | | + | + | | | | | | | 2 |
| Marital status: Single vs Married/partner | Social | | - | - | | | | | | | 2 |
| Formic acid intaje (g/day) | Nutrients | | | | + | | | + | | | 2 |
| Palmitic acid intake (g/day) | Nutrients | | | | + | | | + | | | 2 |
| Whole grain intake (g/day) | Food | | + | | | | | | - | | 2 |
| Frequency of walking during leisure time | Physical activity | | | + | | | + | | | | 2 |
| Frequency of cycling during leisure time | Physical activity | | | | - | - | | | | | 2 |
| Whole grain crisp bread | Food | | + | + | | | | | | | 2 |
| Banana | Food | | | | - | | | - | | | 2 |
| Bacon | Food | | | | + | | + | | | | 2 |
| Butter for cooking | Food | | | | | | + | + | | | 2 |
| Margarine for cooking | Food | | | | + | + | | | | | 2 |
| Low fat milk (0,5%) | Beverage | + | | | - | | | | | | 2 |
| Medium beer | Alcohol | - | | | | | + | | | | 2 |
| Feel the need to reduce alcohol consumption | Alcohol | | | + | | + | | | | | 2 |
| Cholesterol intake (g/day) | Nutrients | | - | | | | + | | | | 2 |
| Sum of Lariciresinol, Matairesinol, Pinoresinol, Secoisolariciresinol intake (ug/day) | Nutrients | | | | | | | | - | - | 2 |
| Lariciresinol intake (ug/day) | Nutrients | | | | | | | | - | - | 2 |

| | | | | | | | | | | | |
|---|---|---|---|---|---|---|---|---|---|---|---|
| Appetite status | Psychosocial | | | | | - | | | - | | 2 |
| Sleep status | Sleep | - | | | | - | | | | | 2 |
| Satisfaction with economy | Psychosocial | | | + | | - | | | | | 2 |
| Vision status | General health | | | | | - | | | | + | 2 |
| Average portion size of potatoes/rice/pasta based on photographic illustration of four sizes (smallest to largest) | Food | + | | - | | | | | | | 2 |
| Plant based protein intake (g/day) | Nutrients | | | | - | | | - | | | 2 |
| Cohabitation: Live alone vs Adult and children | Social | | | - | | - | | | | | 2 |
| Excellent health | General health | | | - | - | | | | | | 2 |
| Long-term sickness | General health | | | | | + | - | | | | 2 |
| Years using snuff | Tobacco use | + | | | | | + | | | | 2 |
| Participation in sports or physical exercise associations | Social | - | | | | | | | | - | 2 |
| Trans fat intake (g/day) | Nutrients | | | | + | | | + | | | 2 |
| Permanent employment | Psychosocial | - | | | | | | | | | 1 |
| Job demands to work very fast | Psychosocial | - | | | | | | | | | 1 |
| Frequncy of social contacts with colleagues during leisure time | Psychosocial | | | + | | | | | | | 1 |
| High physical demand from job | Physical activity | | + | | | | | | | | 1 |
| Ingenuity or creativity demand from job | Psychosocial | | | | | | | | | - | 1 |
| Contradictory demands in job | Psychosocial | | - | | | | | | | | 1 |
| Learn new things at job | Psychosocial | | - | | | | | | | | 1 |
| Skill demand from job | Psychosocial | | | | | | | | | - | 1 |
| Possibility to speak with colleagues during breaks | Psychosocial | - | | | | | | | | | 1 |
| Vitamin C intake (mg/day) | Nutrients | | | | | - | | | | | 1 |
| Beta-sitostanol intake (mg/day) | Nutrients | | + | | | | | | | | 1 |
| Beta-sitosterol intake (mg/day) | Nutrients | | | | | | | | - | | 1 |
| Disaccharides intake (g/day) | Nutrients | | | | | | - | | | | 1 |
| Total energy intake (kcal/day) | Nutrients | | | - | | | | | | | 1 |
| Arachidonic acid (ARA) intake (g/day) | Nutrients | + | | | | | | | | | 1 |
| Folic acid intake (ug/day) | Nutrients | | | | | - | | | | | 1 |
| Phosphate intake (mg/day) | Nutrients | | | - | | | | | | | 1 |
| Travel to work: Irregular travel mode to work vs passive travel to work | Physical activity | - | | | | | | | | | 1 |
| Frequency of shoverling snow during leisure time | Physical activity | | | | | | | | | - | 1 |
| Everyday exercise satisfaction | Physical activity | - | | | | | | | | | 1 |
| If you exercise, change in exercise habits during the last year | Physical activity | | | | - | | | | | | 1 |
| Bregott on bread | Food | | | | + | | | | | | 1 |
| Whole grain soft bread | Food | | | | | | | | - | | 1 |
| Coffee rolls/buns, rusk | Food | | | | | | - | | | | 1 |
| Oatflake, whole wheat, rye or barley porridge | Food | | | | | | | | - | | 1 |
| Rosehip, sweet syrup soup | Food | | | | | | | | | + | 1 |
| Low fat margarine on bread | Food | | + | | | | | | | | 1 |
| Fried potatoes, pommes frites | Food | | | | | + | | | | | 1 |
| Pizza | Food | | | | | + | | | | | 1 |
| Sausage as main dish | Food | + | | | | | | | | | 1 |
| Hamburger | Food | | | | | | | | | + | 1 |
| White meat (poultry) | Food | | - | | | | | | | | 1 |
| Sugar, honey, marmelade, jam | Food | | | | | | - | | | | 1 |
| Cookies, pastry | Food | | | | | | - | | | | 1 |
| Milk, sour milk (3%) | Beverage | | | | + | | | | | | 1 |
| Cream, creme fraiche, sour cream | Food | | | | | | | + | | | 1 |
| Iodine intake (ug/day) | Nutrients | | | - | | | | | | | 1 |
| Carbohydrates intake (g/day) | Nutrients | | | | | | - | | | | 1 |
| Breakfast habits: Gruel w/o sandwich for breakfast vs not breakfast at all | Food | - | | | | | | | | | 1 |
| Medioresinol intake (ug/day) | Nutrients | | + | | | | | | | | 1 |
| Pinoresinol intake (ug/day) | Nutrients | | | | | | | | - | | 1 |
| Sum of all lignans intake (ug/day) | Nutrients | | + | | | | | | | | 1 |
| Syringaresinol intake (ug/day) | Nutrients | | + | | | | | | | | 1 |
| Mood status | Psychosocial | | | | | - | | | | | 1 |
| Satisfaction with work situation | Psychosocial | | | | | - | | | | | 1 |
| Magnesium intake (mg/day) | Nutrients | | | | | | | | | - | 1 |
| Monosaccharides intake (g/day) | Nutrients | | | | | | | | | - | 1 |

| | | | | | | | | | | | |
|---|---|---|---|---|---|---|---|---|---|---|---|
| Sodium intake (mg/day) | Nutrients | + | | | | | | | | | 1 |
| Animal based protein intake (g/day) | Nutrients | + | | | | | | | | | 1 |
| Sucrose intake (g/day) | Nutrients | | | | | | - | | | | 1 |
| Selenium intake (ug/day) | Nutrients | | | - | | | | | | | 1 |
| Physical limitation to participate in steneous activities: running, lifting heavy objects, taking part in physically demanding sports | General health | | | | + | | | | | | 1 |
| Physical limitation to participate in moderately demanding activities: walking up several stairs | General health | + | | | | | | | | | 1 |
| Physical limitation to participate in moderately demanding activities: bending down or kneeling | General health | + | | | | | | | | | 1 |
| For how much of the time during the last four weeks have you felt tired? | General health | | | | | | - | | | | 1 |
| Work shifts/weekends | Psychosocial | + | | | | | | | | | 1 |
| Snore during sleep | Sleep | + | | | | | | | | | 1 |
| Number of cigarretes smoked per day | Tobacco use | - | | | | | | | | | 1 |
| Smoking status: Former occasional smokers vs non-smokers | Tobacco use | | - | | | | | | | | 1 |
| Number of snuff boxes per week | Tobacco use | | - | | | | | | | | 1 |
| Snuff status: Former snuff users vs non-snuff users | Tobacco use | | | | + | | | | | | 1 |
| Participation in study circles | Social | + | | | | | | | | | 1 |
| Participation in other association | Social | + | | | | | | | | | 1 |
| Number of friends that can come to your home at any time and feel at home | Social | | | | + | | | | | | 1 |
| Number of social contacts with the same interests as you | Social | | | | | | | | | - | 1 |
| Frequency of engaging in clubs, associations or study circles | Social | | - | | | | | | | | 1 |
| Receive hugs to comfort and support you | Social | | | | | - | | | | | 1 |

BMI: Body Mass Index; SBP: Systolic blood pressure; DBP: Diastolic blood pressure; Tot Chol: Total cholesterol; Trigl: Triglycerides; HDL-C: HDL cholesterol; LDL-C: LDL cholesterol; F Glu: Fasting glucose; 2h Glu: 2h glucose

**Supplementary Table 22**. Number of tentative signals ranked among the top 5 regarding variance explained in linear mixed model analyses that are shared between the different cardiometabolic traits

| | BMI | SBP | DBP | Tot Chol | Trigl | HDL-C | LDL-C | F Glu | 2h Glu |
|---|---|---|---|---|---|---|---|---|---|
| **BMI** | 45 | | | | | | | | |
| **SBP** | 10 | 35 | | | | | | | |
| **DBP** | 13 | 18 | 34 | | | | | | |
| **Tot Chol** | 10 | 9 | 10 | 36 | | | | | |
| **Trigl** | 13 | 10 | 16 | 11 | 38 | | | | |
| **HDL-C** | 15 | 8 | 11 | 10 | 14 | 32 | | | |
| **LDL-C** | 5 | 2 | 2 | 9 | 3 | 3 | 13 | | |
| **F Glu** | 11 | 9 | 11 | 7 | 14 | 10 | 2 | 26 | |
| **2h Glu** | 16 | 10 | 13 | 10 | 17 | 13 | 4 | 16 | 37 |

**Supplementary Table 23**. Summary of significant top 5 lifestyle variables regarding variance explained for all the cardiometabolic traits in longitudinal analyses

| Description | Group | BMI | SBP | DBP | Tot Chol | Trigl | F glu | 2h glu | N of tentative associations |
|---|---|---|---|---|---|---|---|---|---|
| Informed of having high blood pressure | General health | + | + | + | | | | + | 4 |
| Number of cigarretes smoked per day (in groups) | Tobacco use | + | | | | + | + | | 3 |
| Number of cigarretes smoked per day | Tobacco use | + | | | + | + | | | 3 |
| Smoking status: Smokers vs non-smokers | Tobacco use | + | | | | + | + | | 3 |
| Snuff status: Snuff users vs non-snuff users | Tobacco use | + | | | + | + | | | 3 |
| Educational level | Psychosocial | | - | | | | - | - | 3 |
| Parents or siblings have diabetes | General health | | | | | | + | + | 2 |
| Fiber cereals | Food | - | | | | - | | | 2 |
| Overall state of health during the last year | General health | - | | | | - | | | 2 |
| Parents or siblings had a cerebral hemorrhage/thrombosis or cardiac infarction before the age of 60 | General health | | + | + | | | | | 2 |
| Grams of tobacco smoked per week | Tobacco use | | | | + | + | | | 2 |
| Number of cigars smoked per day | Tobacco use | | | | + | + | | | 2 |
| Number of snuff boxes per week | Tobacco use | | | | + | + | | | 2 |
| Years using snuff | Tobacco use | + | | | + | | | | 2 |
| Arachidonic acid (ARA) intake (g/day) | Nutrients | + | | | | | | | 1 |
| Enterodiol intake (ug/day) | Nutrients | + | | | | | | | 1 |
| Equol intake (ug/day) | Nutrients | + | | | | | | | 1 |
| Matairesinol intake (ug/day) | Nutrients | - | | | | | | | 1 |
| Secoisolariciresinol intake (ug/day) | Nutrients | | | | | | | - | 1 |
| Monounsaturated fat intake (g/day) | Nutrients | | | | + | | | | 1 |
| Tiamin intake (mg/day) | Nutrients | + | | | | | | | 1 |
| Alcohol intake (g/day) | Alcohol | | | | + | | | | 1 |
| Permanent employment | Psychosocial | | | | + | | | | 1 |
| High physical demand from job | Physical activity | + | | | | | | | 1 |
| Repetitive job | Psychosocial | + | | | | | | | 1 |
| Marital status: Single vs Married/partner | Social | - | | | | | | | 1 |
| Travel to work: Cycle to work vs passive travel to work | Physical activity | - | | | | | | | 1 |
| Frequency of picking berries or mushrooms during leisure time | Physical activity | | | | - | | | | 1 |
| Changed everyday exercise during the last year | Physical activity | + | | | | | | | 1 |
| Exercise during the last three months | Physical activity | | | | | | | - | 1 |
| If you exercise, change in exercise habits during the last year | Physical activity | + | | | | | | | 1 |
| Oatflake, whole wheat, rye or barley porridge | Food | | | - | | | | | 1 |
| Sour milk, yoghurt (3% fat) | Food | - | | | | | | | 1 |
| Tomato, cucumber | Food | + | | | | | | | 1 |
| Fried potatoes, pommes frites | Food | + | | | | | | | 1 |
| Chips, popcorn, salted nuts | Food | | | | + | | | | 1 |
| Brewed (filtered) coffee | Beverage | | | | + | | | | 1 |
| Strong beer | Alcohol | | | | + | | | | 1 |
| Teetotaler | Alcohol | | | | - | | | | 1 |
| Average portion size of meat/fish based on photographic illustration of four sizes (smallest to largest) | Food | | | | | | | - | 1 |
| Breakfast habits: Only coffee/tea for breakfast vs not breakfast at all | Food | | | | + | | | | 1 |
| Satisfaction with home and family situation | Psychosocial | - | | | | | | | 1 |
| Sleep status | Sleep | - | | | | | | | 1 |
| Satisfaction with accomodation | Psychosocial | - | | | | | | | 1 |
| Satisfaction with economy | Psychosocial | - | | | | | | | 1 |
| Fitness status | Physical activity | - | | | | | | | 1 |
| Cambridge physical activity index | Physical activity | | | | | | | - | 1 |

| | | | | | | | | | |
|---|---|---|---|---|---|---|---|---|---|
| Average portion size of potatoes/rice/pasta based on photographic illustration of four sizes (smallest to largest) | Food | | | | | | | - | 1 |
| Cohabitation: Live alone vs Only one adult (spouse, partner) | Social | - | | | | | | | 1 |
| Cohabitation: Live alone vs Adult and children | Social | - | | | | | | | 1 |
| Work shifts/weekends | Psychosocial | + | | | | | | | 1 |
| Years smoking | Tobacco use | | | | | | + | | 1 |
| Close relationship with anyone | Social | - | | | | | | | 1 |

BMI: Body Mass Index; SBP: Systolic blood pressure; DBP: Diastolic blood pressure; Tot Chol: Total cholesterol; Trigl: Triglycerides; F Glu: Fasting glucose; 2h Glu: 2h glucose

**Supplementary Table 24**. Number of tentative signals ranked among the top 5 regarding variance explained in longitudinal analyses that are shared between the different cardiometabolic traits

| | BMI | SBP | DBP | Tot Chol | Trigl | HDL-C | LDL-C | F Glu | 2h Glu |
|---|---|---|---|---|---|---|---|---|---|
| **BMI** | 31 | | | | | | | | |
| **SBP** | 1 | 3 | | | | | | | |
| **DBP** | 1 | 2 | 3 | | | | | | |
| **Tot Chol** | 3 | 0 | 0 | 15 | | | | | |
| **Trigl** | 6 | 0 | 0 | 5 | 9 | | | | |
| **HDL-C** | 0 | 0 | 0 | 0 | 0 | 0 | | | |
| **LDL-C** | 0 | 0 | 0 | 0 | 0 | 0 | 0 | | |
| **F Glu** | 2 | 1 | 0 | 0 | 2 | 0 | 0 | 5 | |
| **2h Glu** | 1 | 2 | 1 | 0 | 0 | 0 | 0 | 2 | 8 |

**Supplementary Table 25**. Reference for the numerical labels in the figures

| Number | Description |
|---|---|
| 1 | Fitness status |
| 2 | Physical limitation to participate in moderately demanding activities: bending down or kneeling |
| 3 | Physical limitation to participate in moderately demanding activities: walking up several stairs |
| 4 | Physical limitation to participate in steneous activities: running, lifting heavy objects, taking part in physically demanding sports |
| 5 | Self rate of overall health |
| 6 | Physical limitation to participate in moderately demanding activities: walking more than 2 km |
| 7 | Everyday exercise satisfaction |
| 8 | Overall state of health during the last year |
| 9 | Excellent health |
| 10 | Informed of having high blood pressure |
| 11 | Exercise during the last three months |
| 12 | Energy status |
| 13 | Overall state of health compared to other your age |
| 14 | Physical limitation to participate in moderately demanding activities: moving a table, vacuuming, walking in the forest or gardening |
| 15 | Pain during the last four weeks |
| 16 | How much has the pain during the last four weeks disturbed your normal work? |
| 17 | As healthy as anyone |
| 18 | Cambridge physical activity index |
| 19 | Average portion size of meat/fish based on photographic illustration of four sizes (smallest to largest) |
| 20 | For how much of the time during the last four weeks have you felt really alert and strong? |
| 21 | Travel to work: Cycle to work vs passive travel to work |
| 22 | Physical limitation that made you do less than you wanted during the last four weeks |
| 23 | For how much of the time during the last four weeks have you felt worn out? |
| 24 | Frequency of walking during leisure time |
| 25 | Secoisolariciresinol intake (ug/day) |
| 26 | For how much of the time during the last four weeks have you felt full of energy? |
| 27 | Fibre intake (g/day) |
| 28 | Average portion size of potatoes/rice/pasta based on photographic illustration of four sizes (smallest to largest) |
| 29 | Animal based protein intake (g/day) |
| 30 | Arachidonic acid (ARA) intake (g/day) |
| 31 | For how much of the time during the last four weeks have you felt tired? |
| 32 | Physical limitation that made you not being able to perform certain work tasks or other activities during the last four weeks |
| 33 | Sodium intake (mg/day) |
| 34 | Frequency of cycling during leisure time |
| 35 | Sum of Lariciresinol, Matairesinol, Pinoresinol, Secoisolariciresinol intake (ug/day) |
| 36 | Get sick more often than other people |
| 37 | Sausage as main dish |
| 38 | Physical limitation that limited your ability to perform certain work tasks or other activities during the last four weeks |
| 39 | Vitamin B3 intake (mg/day) |
| 40 | Long-term sickness |
| 41 | Vitamin D intake (ug/day) |
| 42 | Pinoresinol intake (ug/day) |
| 43 | Participation in sports or physical exercise associations |
| 44 | Lariciresinol intake (ug/day) |
| 45 | Parents or siblings have diabetes |
| 46 | Monounsaturated fat intake (g/day) |
| 47 | Physical limitation to participate in moderately demanding activities: lifting or carrying grocery bags |
| 48 | Folic acid intake (ug/day) |
| 49 | Educational level |
| 50 | Plant based protein intake (g/day) |
| 51 | Worsen in health in the future |
| 52 | Total protein intake (g/day) |
| 53 | Changed everyday exercise during the last year |
| 54 | Steak, chop, e.g. |
| 55 | Monosaccharides intake (g/day) |
| 56 | Meat stew |
| 57 | Wine |
| 58 | Bacon |
| 59 | If you exercise, change in exercise habits during the last year |
| 60 | Whole grain intake (g/day) |
| 61 | Cholesterol intake (g/day) |
| 62 | Breakfast habits: Porridge w/o sandwich for breakfast vs not breakfast at all |
| 63 | Carbohydrates intake (g/day) |
| 64 | Matairesinol intake (ug/day) |
| 65 | Permanent employment |
| 66 | Extent to what your physical and emotional health disrupted your usual social life during the last four weeks |
| 67 | Stigmasterol intake (mg/day) |
| 68 | For how much of the time during the last four weeks has your physical health or your emotional problems limited your ability to interact with others? |
| 69 | Minced meat dishes |
| 70 | Vitamin B12 intake (ug/day) |
| 71 | Physical limitation that reduced the normal time spent at work or in other activities during the last four weeks |
| 72 | Emotional problems that made you do less than you wanted during the last four weeks |
| 73 | Breakfast habits: Gruel w/o sandwich for breakfast vs not breakfast at all |

74 Vitamin C intake (mg/day)
75 Tea
76 Sum of all lignans intake (ug/day)
77 Smoking status: Former smokers vs non-smokers
78 Hamburger
79 Beta-sitosterol intake (mg/day)
80 Fiber cereals
81 Fat intake (g/day)
82 Satisfaction with leisure time
83 Oatflake, whole wheat, rye or barley porridge
84 White cabbage, lettuce, lettuce cabbage, spinach, borecole
85 Root vegetables, carrot
86 Beta-carotene intake (mg/day)
87 Salad dressing with oil
88 Eat breakfast from 2000
89 Participation in other association
90 Frequency of dancing during leisure time
91 Palmitic acid intake (g/day)
92 Frequency of picking berries or mushrooms during leisure time
93 Magnesium intake (mg/day)
94 Distance to work in kilometers (one way)
95 Tomato, cucumber
96 Selenium intake (ug/day)
97 Pancake, waffle, Swedish dumpling
98 Alcohol intake (g/day)
99 Frequency of hunting or fishing during leisure time
100 Sum of phytosterols intake (mg/day)
101 Low fat milk (0,5%)
102 Fried potatoes, pommes frites
103 Salty fish
104 Average portion size of vegetables based on photographic illustration of four sizes (smallest to largest)
105 Self rate of overall health compared to a year ago
106 Apple, pear, peach, orange, mandarin and grapefruit
107 Boiled coffee
108 Possibility to speak with colleagues during breaks
109 Margarine for cooking
110 For how much of the time during the last four weeks have you felt gloomy and sad?
111 Sucrose intake (g/day)
112 Sausage, liver pate on bread
113 Sour milk, yoghurt (3% fat)
114 White meat (poultry)
115 Syringaresinol intake (ug/day)
116 Marital status: Single vs Divorced/separated
117 Years using snuff
118 Light and physically active work
119 For how much of the time during the last four weeks have you felt happy?
120 Travel to work: Walk to work vs passive travel to work
121 For how much of the time during the last four weeks have you felt so depressed that nothing could cheer you up?
122 Berries (fresh or frozen)
123 Vitamin B6 intake (mg/day)
124 Brewed (filtered) coffee
125 Pizza
126 Boiled or baked potato
127 Beta-sitostanol intake (mg/day)
128 Eicosapentaenoic acid (EPA) intake (g/day)
129 Breakfast habits: Coffee/tea and wheat buns or rusk for breakfast vs not breakfast at all
130 Job demands to work very fast
131 Participation in study circles
132 Mashed potato
133 Docosahexaenoic acid (DHA) intake (g/day)
134 Years smoking
135 Campestanol intake (mg/day)
136 Banana
137 Mixed frozen vegetables
138 Sedentary or standing work
139 Work shifts/weekends
140 Brown beans, pea soup
141 Satisfaction with economy
142 Vitamin A intake (mg/day)
143 Equol intake (ug/day)
144 Total energy intake (kcal/day)
145 Medioresinol intake (ug/day)
146 Medium beer
147 Patience status
148 Number of cigarretes smoked per day (in groups)
149 Liquor, spirits

150 Saturated fat intake (g/day)
151 Parents or siblings had a cerebral hemorrhage/thrombosis or cardiac infarction before the age of 60
152 For how much of the time during the last four weeks have you felt calm and serene?
153 Travel to work: Irregular travel mode to work vs passive travel to work
154 Frequent social contacts with colleagues during work
155 Sugar, honey, marmelade, jam
156 Low fat margarine on bread
157 Zinc intake (mg/day)
158 Repetitive job
159 Number of cigarretes smoked per day
160 Enterodiol intake (ug/day)
161 Receive hugs to comfort and support you
162 Sleep status
163 Snuff status: Former snuff users vs non-snuff users
164 Pasta
165 Pentadecanoic acid intake (g/day)
166 Heptadecanoic acid intake (g/day)
167 Teetotaler
168 Mood status
169 Whole grain soft bread
170 Whole grain crisp bread
171 Hearing status
172 Milk, sour milk (3%)
173 Milk, sour milk (1,5%)
174 Frequency of gardening during leisure time
175 Polyunsaturated fat intake (g/day)
176 Soft cheese
177 Would you say that the number of people that you meet in your everyday life is enough or would you like to meet more or fewer people?
178 Satisfaction with work situation
179 Skill demand from job
180 Vitamin B2 intake (ug/day)
181 Cream, creme fraiche, sour cream
182 Light beer
183 Disaccharides intake (g/day)
184 Appetite status
185 Support from others
186 Soft whey cheese
187 Blota (broth + bread)
188 Breakfast habits: Only coffee/tea for breakfast vs not breakfast at all
189 Phosphate intake (mg/day)
190 Control over own work assignment
191 Iodine intake (ug/day)
192 Number of friends that can come to your home at any time and feel at home
193 Number of social interactions during a normal week
194 Coffee rolls/buns, rusk
195 Snuff status: Snuff users vs non-snuff users
196 Participation in associations or voluntary organisations
197 Smoked fish/meat
198 Lean fish (e.g. perch, bass, cod)
199 Do you feel important and appreciated outside your home?
200 Do you feel important and appreciated in your home?
201 Grams of tobacco smoked per week
202 Learn new things at job
203 Confidence status
204 Cohabitation: Live alone vs Only children
205 White (soft) bread, thin crisp bread
206 Cheese 28%
207 Liver, kidney
208 Light but partly physically active work
209 Cookies, pastry
210 Fatty fish (e.g. herring, whitefish, salmon)
211 Smoking status: Former occasional smokers vs non-smokers
212 For how much of the time during the last four weeks have you felt very nervous?
213 Rosehip, sweet syrup soup
214 Oil for cooking
215 Control over planning and execution of the workday
216 Calcium intake (mg/day)
217 Sodas, soft drinks, juice
218 Satisfaction with home and family situation
219 Satisfaction with accomodation
220 Feel uneasy or guilty because of your way of drinking
221 Sweets
222 Number of social contacts with the same interests as you
223 Frequncy of social contacts with colleagues during leisure time
224 Marital status: Single vs Married/partner
225 Last time a colleague visited you at home

226 Ice cream
227 Marital status: Single vs Widow/widower
228 Linolenic acid intake (g/day)
229 Iron intake (mg/day)
230 Corn flakes
231 Potato salad
232 Self-employed
233 Trans fat intake (g/day)
234 Sour milk, yoghurt (low fat)
235 Campesterol intake (mg/day)
236 Frequency of shoverling snow during leisure time
237 Possibility to leave your work for a while to speak with a colleague
238 Strong beer
239 Bregott on bread
240 Vitamin E intake (mg/day)
241 Meat on bread
242 Enough time for job assignments
243 Memory status
244 People to ask for help apart from the ones at home
245 Frequency of engaging in clubs, associations or study circles
246 Sometimes physically straining work
247 High physical demand from job
248 Shellfish (e.g. shrimps, scallops)
249 Cohabitation: Live alone vs Other/others
250 Enterolactone intake (ug/day)
251 Smoking status: Smokers vs non-smokers
252 Potassium intake (mg/day)
253 Butter on bread
254 Cheese 10-17%
255 Feel the need to reduce alcohol consumption
256 Number of snuff boxes per week
257 Ingenuity or creativity demand from job
258 Butter for cooking
259 Formic acid intaje (g/day)
260 Number of people with whom you can speak openly
261 Tiamin intake (mg/day)
262 Rice
263 Chips, popcorn, salted nuts
264 High mental demand from job
265 Blood based food
266 Vision status
267 Close relationship with anyone
268 Margarine on bread
269 Number of cigars smoked per day
270 Cohabitation: Live alone vs Only one adult (spouse, partner)
271 Cohabitation: Live alone vs Adult and children
272 Linoleic acid intake (g/day)
273 Contradictory demands in job
274 Time spent in a week in moderately steneous activities
275 Amount of exercise during the last 12 months
276 Frequency of alcohol consumption
277 Amount of alcohol drunk in a day
278 Frequency of drinking six or more glasses at the same occasion
279 Times during last year that you felt guilty because of your drinking
280 Risk of sleeping while sitting and reading
281 Risk of sleeping while watching TV
282 Risk of sleeping while sitting inactive in a public place
283 Risk of sleeping as a passenger in a car for one hour without break
284 Risk of sleeping while lying down resting in the afternoon
285 Risk of sleeping while sitting still after having lunch
286 Snore during sleep
287 Breath-holds during sleep
